# A genome-wide association study identified 10 novel genomic loci associated with intrinsic capacity

**DOI:** 10.1101/2025.02.05.25321753

**Authors:** Melkamu Bedimo Beyene, Renuka Visvanathan, Robel Alemu, Beben Benyamin, Rudrarup Bhattacharjee, Habtamu B. Beyene, Olga Theou, Matteo Cesari, John R. Beard, Azmeraw T. Amare

**Affiliations:** Adelaide Medical School, Faculty of Health and Medical Sciences, University of Adelaide, Adelaide, SA, Australia; Adelaide Geriatrics Training and Research with Aged Care Centre (GTRAC), Faculty of Health and Medical Sciences, University of Adelaide and the Basil Hetzel Institute, Central Adelaide Local Health Network Woodville, SA, 5011, Australia; Aged and Extended Care Services, The Queen Elizabeth Hospital, Central Adelaide Local Health Network, Woodville, SA, 5011, Australia; University of California, Los Angeles (UCLA), CA, USA; Broad Institute of MIT and Harvard, Medical and Population Genetics Program, Cambridge, MA, USA; Australian Centre for Precision Health, Allied Health and Human Performance, University of South Australia, Adelaide, 5000, Australia; South Australian Health and Medical Research Institute, Adelaide, 5000, Australia; Adelaide Centre for Epigenetics, School of Biomedicine, The University of Adelaide, Adelaide, SA, 5005, Australia; South Australian Immunogenomics Research Institute (SAiGENCI), The University of Adelaide, Adelaide, SA, 5005, Australia; Metabolomics Laboratory, Baker Heart and Diabetes Institute, Melbourne; Department of Cardiovascular Research, Translation and Implementation, La Trobe University, Bundoora, Australia; Physiotherapy and Geriatric Medicine, Dalhousie University, Halifax, Nova Scotia, Canada; Ageing and Health Unit, Department of Maternal, Newborn, Child & Adolescent Health and Ageing, World Health Organization, Geneva, Switzerland; Department of Clinical Sciences and Community Health, University of Milan, Milan, Italy; Robert N. Butler Columbia Aging Centre, Columbia University, NY, USA

## Abstract

In 2015, the World Health Organization introduced the concept of intrinsic capacity (IC), a composite of all the physical and mental attributes that contribute to healthy aging. While substantial evidence supports the biological basis of IC and its subdomains, the extent of genetic influence on IC remains largely unexplored, with no studies currently available. Understanding the genetic basis of IC is crucial to advancing our knowledge and identifying interventions that promote healthy aging. To investigate the genetic basis of IC, we used data from the UK Biobank (UKB; N=44,631) and the Canadian longitudinal study on aging (CLSA; N=13,085). We estimated SNP-based heritability (h^2^_SNP_) at 25.2% in UKB and 19.5% in CLSA. A Genome-Wide Association Study (GWAS) identified 38 independent SNPs for IC across 10 genomic loci and 4,289 candidate SNPs, mapped to 197 genes. Post-GWAS analysis revealed the role of these genes on cellular processes such as cell proliferation, immune function, metabolism, and neurodegeneration, with high expressions in muscle, heart, brain, adipose, and tibial nerve tissues. Of the 52 traits tested, 23 showed significant genetic correlations with IC, and a higher genetic loading for IC was associated with higher IC scores. This study is the first to identify genetic variants and pathways associated with IC, providing a foundation for future research on healthy aging.

## Main

Aging is a complex biological process characterized by a gradual decline in biological functions, ultimately impacting health and well-being. These changes increase the risk of chronic conditions, but declines in functioning occur even in the absence of disease. This process is driven by the interplay between genetic, environmental, and lifestyle factors and their interactions, resulting in considerable variability in healthy aging trajectories among individuals. As the global population is aging rapidly, a better understanding of the determinants of healthy aging is becoming increasingly important and could lead to novel and personalized health promotion interventions [1, 2].

In 2015, the World Health Organization proposed a framework for healthy aging focused on building and maintaining the functional ability of older adults. Central to this framework was the concept of intrinsic capacity (IC), which refers to the composite of all the physical and mental capacities an individual can draw upon across the lifespan [3]. Unlike disease and disability, which are relatively late and dichotomous measures of poor health in older adults, IC provides a holistic and continuous framing of health that mirrors recent work on the complex and dynamic biological changes that drive aging. IC is operationalized through five key domains: cognitive, psychological, sensory, locomotor, and vitality capacities [3–8]. IC has been identified as a powerful predictor of many subsequent outcomes, including mortality, care dependence, and a range of chronic diseases, even after accounting for personal characteristics and multimorbidity.

While research on IC has expanded significantly since it was conceptualized, most studies have primarily focused on defining its operational framework and examining associations with different health outcomes, sociodemographic, behavioral, and lifestyle choices, and selected biological markers [9]. To date, no study has specifically explored the genetic basis of IC across the genome, with only one candidate gene study documenting an association with the *APOE4* genotype [10]. This represents a paucity of evidence about the genetics of IC, particularly given the potential for biological factors to inform targeted interventions for maintaining IC and promoting healthy aging. The identification of genetic variants associated with IC has the potential to provide valuable insight into the biological pathways underlying aging and functional ability and for planning personalized interventions targeted to promote healthy aging.

In this study, we address the critical gap in understanding the genetic basis of IC by leveraging data from the United Kingdom Biobank (UKB) and the Canadian Longitudinal Study on Aging (CLSA). We performed a comprehensive analysis that included estimating h^2^_SNP_ and conducting GWAS and post-GWAS functional characterization to identify genetic variants associated with IC. The other analyses included pathway analysis, gene-based and gene set analysis, genetic correlation, and polygenic score analyses.

## Results

### Sample characteristics

After applying quality control (QC) filtering, data from 57,716 individuals were analyzed, comprising 44,631 participants from the UKB with a mean (SD) age of 56 (7.6) years and 13,085 participants from the CLSA with a mean (SD) age of 61 (9.5) years. Just over half of the participants from the UKB (N=24,142; 54.1%) and CLSA (N=6,645; 50.8%) were female. Most participants (94.6% of UKB and 92.1% of CLSA) were of European ancestry. The IC scores in both cohorts have an approximate normal distribution with a mean (SD) of 0.000014 (0.64) in the UK biobank and 0.0005 (0.60) in CLSA. A total of ∼8.9 million SNPs, including 5.8 million common SNPs across both cohorts, were included in the final analysis.

### Heritability (h^2^_SNP_)

The h^2^_SNP_ of IC was estimated at 25.2% (95% CI: 23.2 - 27.2%) in the UKB and 19.5% (95% CI: 14.2 - 24.8%) in the CLSA. These estimates reflect the proportion of total phenotypic variance in IC attributable to additive genetic effects. The breakdown of the total phenotypic variance (V_IC_) into genetic (V_G_) and environmental components (Ve) is provided in Supplementary Table 1.

### Genome-wide Wide Association Study (GWAS)

Initially conducting GWAS in both cohorts using a mixed linear model method (fastGWA_mlm), we performed an inverse variance weighted fixed effects GWAS Meta-analysis. The Meta GWAS has identified 38 independent genome-wide significant signals (P < 5×10^-8^) across 10 genomic loci on chromosomes 1, 4, 10, 13, 17, and 20, near or within the *PTP4A2, PRPF3, LCORL, RN7SL89P, ANAPC10, HK1, DLEU1, MAPT, SCN4A,* and *STAU1* genes, respectively (Figure 1). The loci with the strongest association, rs9891103, P=6.50×10^-14,^ located near the *MAPT* gene, contains 18 of the 38 independent significant SNPs. Details of the 10 genomic loci, lead SNPs, and nearest genes are presented in Table 1 and Figure 1. Regional plots of the 10 loci, illustrating the genomic context and patterns of association, are provided in Supplementary Figure 1.

**Figure 1:**
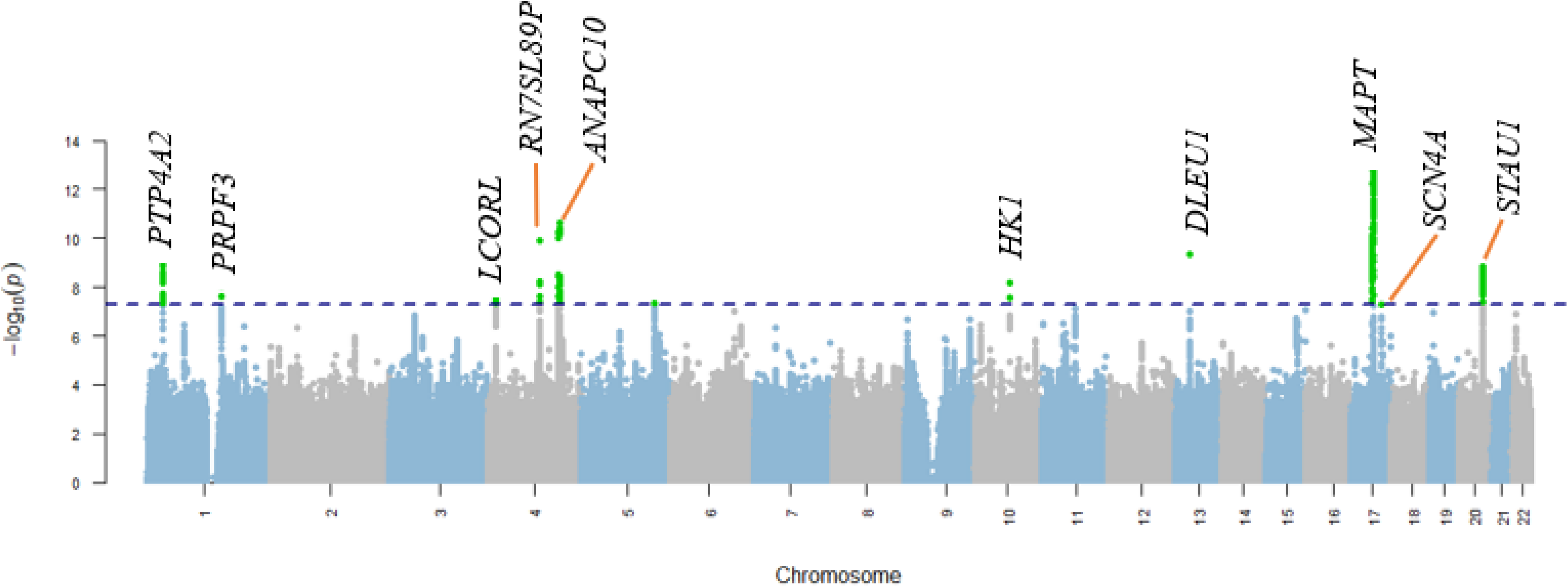
Manhattan plot for the meta-analysis of GWAS summary statistics for IC from the UKB and CLSA GWAS analyses on IC. Each data point represents a SNP, with their positions along the chromosomes (x-axis) plotted against their respective −log10 transformed P-values (y-axis). The plot highlights significant genomic loci associated with IC, with notable peaks surpassing the genome-wide significance threshold cutoff (P < 5×10^-8^) (horizontal blue line).

**Table 1:**
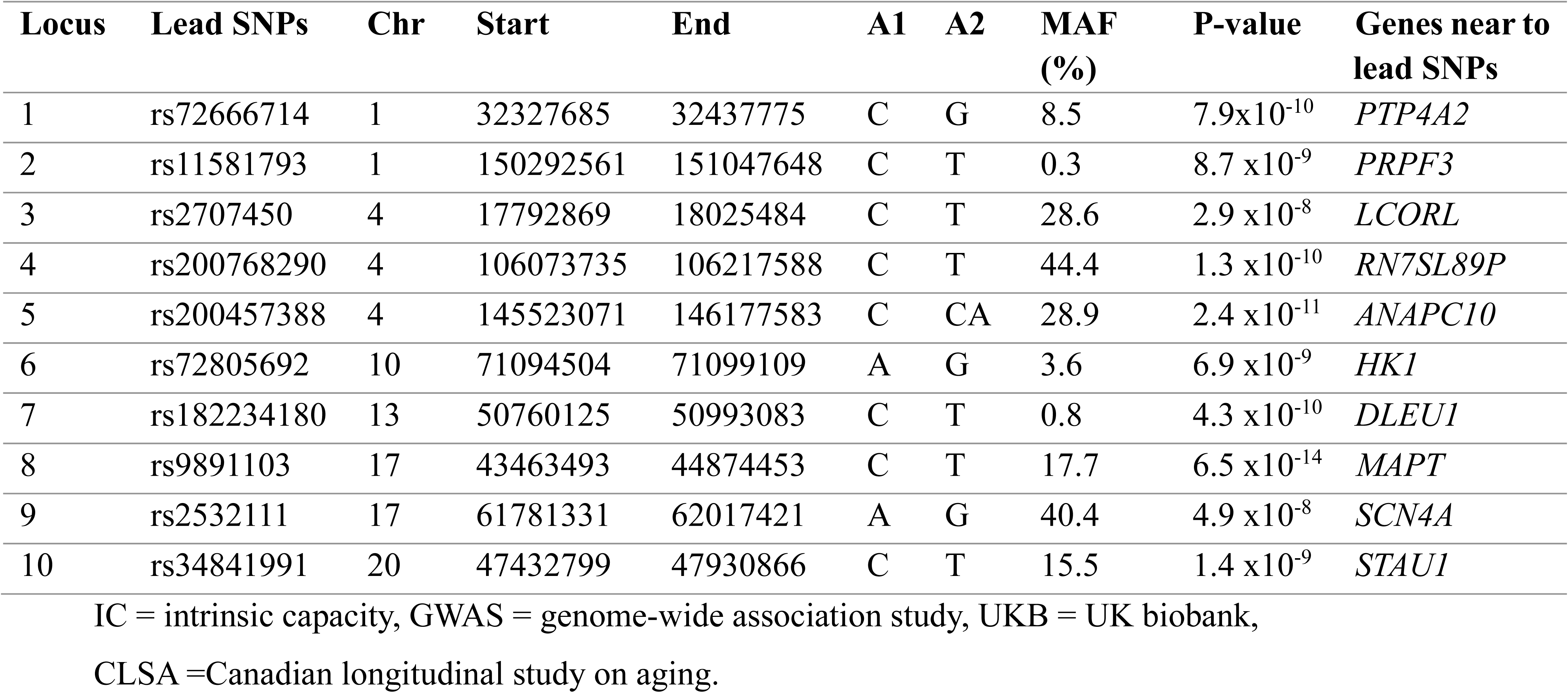
Ten genomic loci associated with IC, lead SNPs, and nearest genes.

The LD-based annotation of independent significant SNPs identified 4,289 candidate SNPs mapped to 197 genes. Among the candidate SNPs, 813 SNPs have been reported for their association with phenotypes related to IC domains and ageing. These include certain biomarkers, cognitive and psychological traits, heart and lung function measures, grip strength, mobility and physical activity, body composition traits, neurodegenerative diseases (e.g., Alzheimer’s and Parkinson’s disease), and cancer. These findings suggest potential pleiotropy, highlighting shared genetic pathways underlying IC and related traits. The full details of the candidate SNPs associated traits are provided in Supplementary Table 2.

### Gene-based tests and gene set analysis

Using MAGMA gene annotation, the 8.9 million GWAS SNPs were mapped to 19,327 protein-coding genes. Of these, 18 genes were significantly associated with IC after Bonferroni correction (P-value cut off ≤ 2.6 x 10^-6^). The gene-based Manhattan plot (Supplementary Figure 2) highlights the 18 genes significantly associated with IC, and the full list of the gene-based associations can be found in Supplementary Table 3.

A gene expression analysis of each of the 197 prioritized genes identified varying expression patterns across 54 tissue types in the GTEx v8 dataset, with seven genes *(MCL1, JTB, COX6B1, DDX5, PSMC5, PSMD4,* and *PTP4A2)* showing consistently high expression in all 54 tissues (Figure 2). Notably, in the heatmap, some of the genes have high expression in tissues relevant to IC domains, including *MAPT, SEMA6C,* and *SUCLA2* in muscle, *MAPT, PREX1,* and *CRHR1* in the brain, *RAB34, PFKM,* and *SLC25A4* in the heart, and *ICAM2* and *CTSK* in adipose and nerve tissues (Figure 2). The tissue specificity analysis showed a significant over-representation of our prioritized genes set in DEGs for pancreas, skeletal muscle, left ventricle, liver, whole blood, and two brain regions—the Putamen basal ganglia and Hippocampus (Figure 3) and the gene set enrichment analysis (GSEA) in GWAS catalog gene sets found a statistically significant over-representation of our prioritized genes in 38 curated gene sets from previous GWAS studies for different phenotypes and diseases (Figure 4).

**Figure 2:**
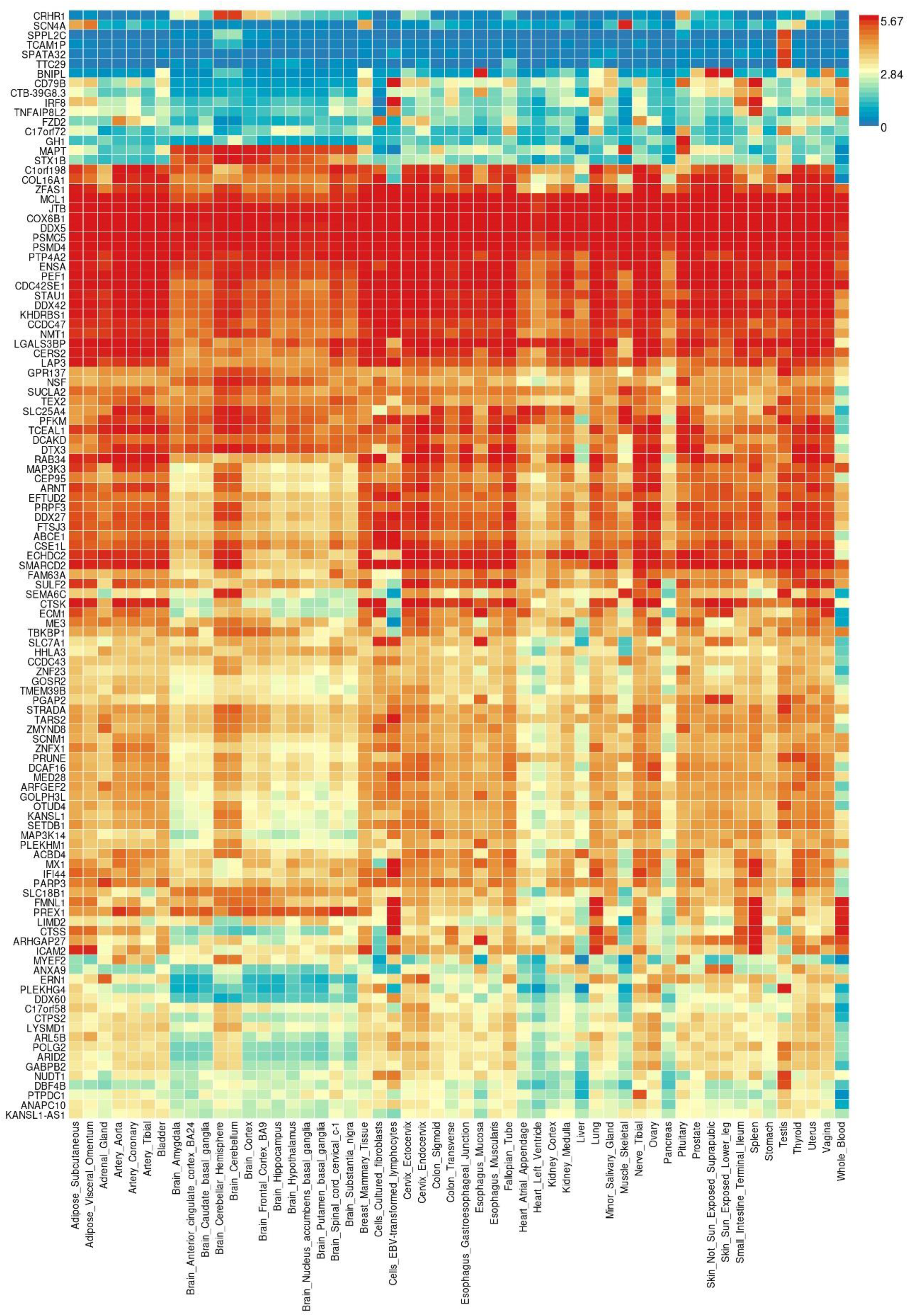
Heatmap showing gene expression pattern – average log 2(RPKM) per tissue for prioritized genes. The color intensity represents the level of gene expression (red shows the highest expression and blue are least expression). *RPKM = Read Per Kilobase per Million*

**Figure 3:**
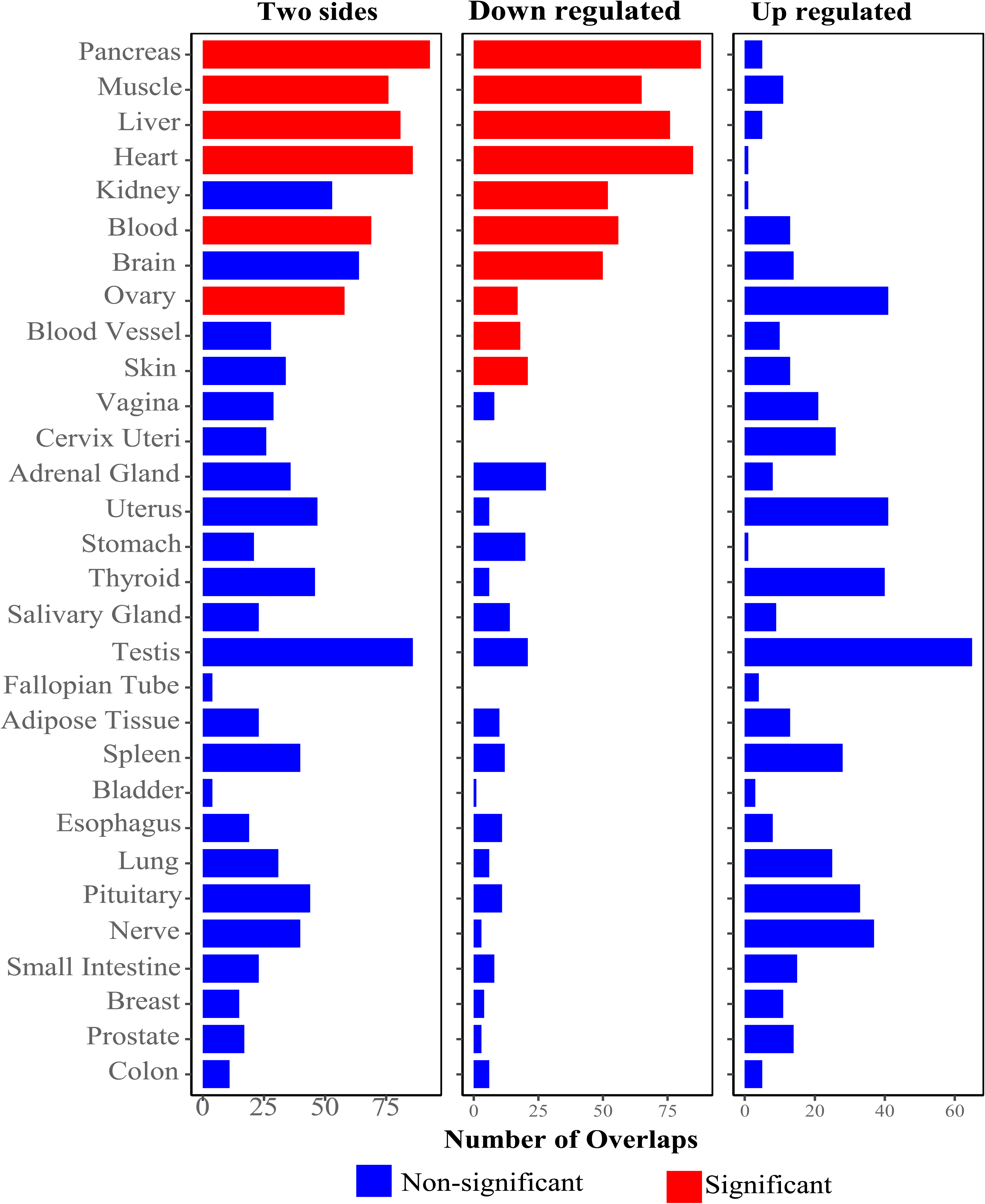
Bar diagram of tissue specificity analysis for our prioritized genes set in differentially expressed gene sets (DEGs) for 30 general tissue types in the GTEx v8 database. Each bar shows the DEGs for the tissues, and the height of the bars shows the number of overlaps. The DEG sets in which our prioritized gene set has a significant over-representation (Bonferroni-corrected P-value < 0.05) are highlighted in red.

**Figure 4:**
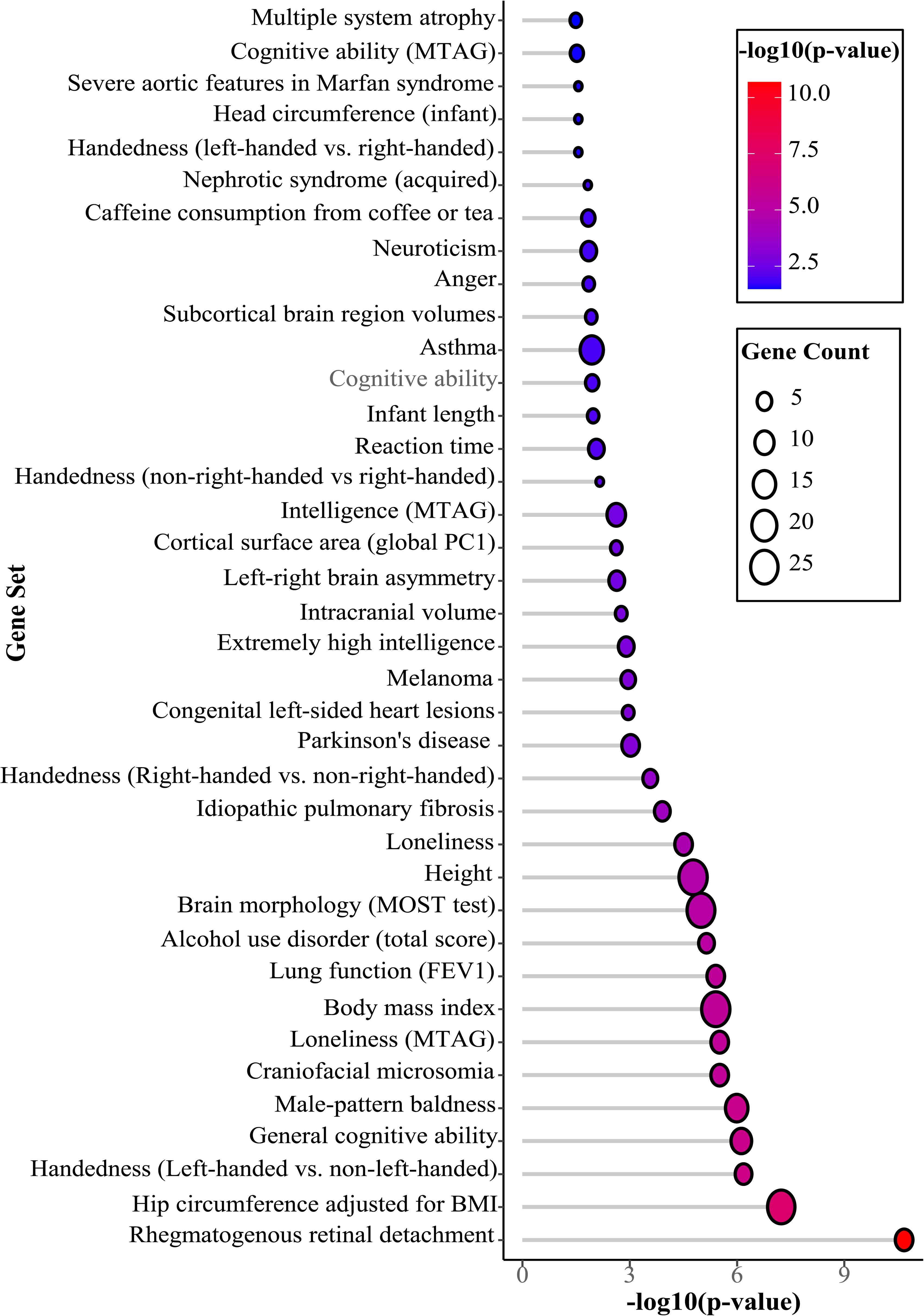
Lollipop plot for GSEA results shows the overlap of our 197 prioritized genes set with gene sets associated with different traits or diseases from the GWAS catalog. On the x-axis are the −log10 adjusted P-values, while the y-axis represents the gene sets ranked by level of significance, with stronger associations at the bottom. The length of the grey horizontal line indicates the −log10 adjusted P-value, with longer lines signifying greater enrichment significance. Circle sizes in the legend reflect the number of overlapping genes between our prioritized genes and the GWAS Catalog gene sets.

### Pathways Analysis

Pathway enrichment analysis performed on 197 prioritized genes identified 88 suggestive pathways (6 from Panther pathways and 82 from reactome pathways) at a nominal significance threshold (P < 0.05), although they were not significant after correction for multiple testing. Among these nominally significant pathways include the cell cycle, synaptic vesicle trafficking, mRNA splicing, Alzheimer’s disease-presenilin pathway, coenzyme A biosynthesis, ubiquitin-proteasome pathway, transcriptional regulation by RUNX1, programmed cell death/apoptosis, class B/2 (secretin family receptors), TCF-dependent signaling in response to WNT, C-type lectin receptors (CLRs), DC42 GTPase cycle, and RAC1 GTPase cycle (Figure 5, supplementary Table 4).

**Figure 5:**
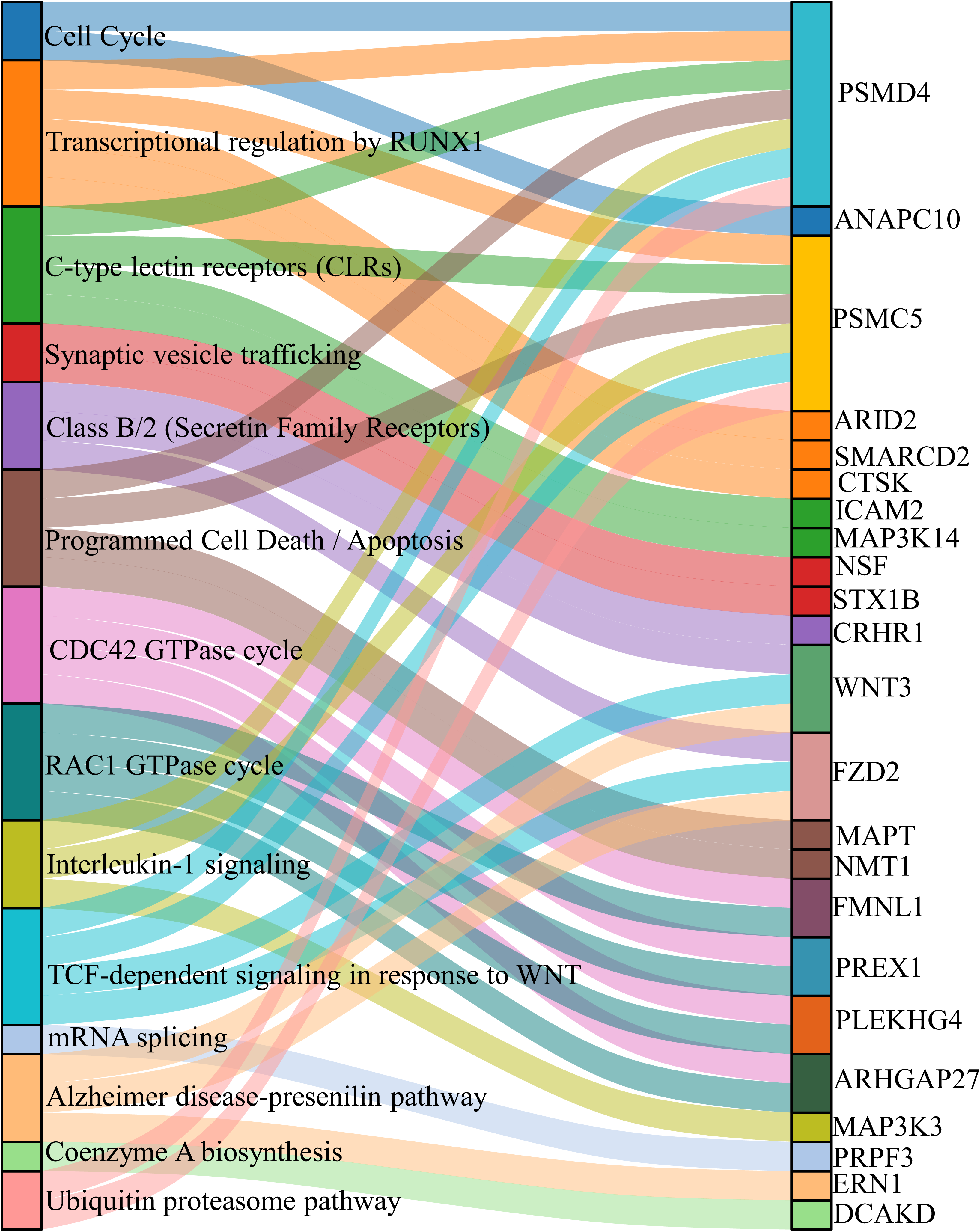
Sankey diagram for selected pathways (left) related to IC-associated genes (right) that showed significant overrepresentation at nominal P-value threshold (P<0.05).

### Polygenic score analysis

The polygenic score (PGS) for IC was significantly associated with the IC score (P < 9.75×10^-16^). A decile-based comparison revealed a linear trend of association, indicating that individuals with greater genetic loading for IC variants are more likely to have a higher IC score. For example, compared to individuals in the first decile (with a low genetic loading for IC variants), those in the tenth decile (with a higher genetic loading) had on average 0.19 increased IC score (95%CI: 0.13-0.25) (Supplementary figure 3).

### Genetic correlation analysis

A genetic correlation analysis was performed for 52 selected phenotypic traits related to physical, cognitive, and metabolic functions and selected for their potential relevance to IC. Of these, 23 traits demonstrated statistically significant genetic correlations with IC, indicating shared genetic architectures (Figure 6). Notably, the strongest genetic correlations were observed with traits associated with vitality, including forced vital capacity (FVC), forced expiratory volume in 1 second (FEV1), and hand grip strength.

**Figure 6:**
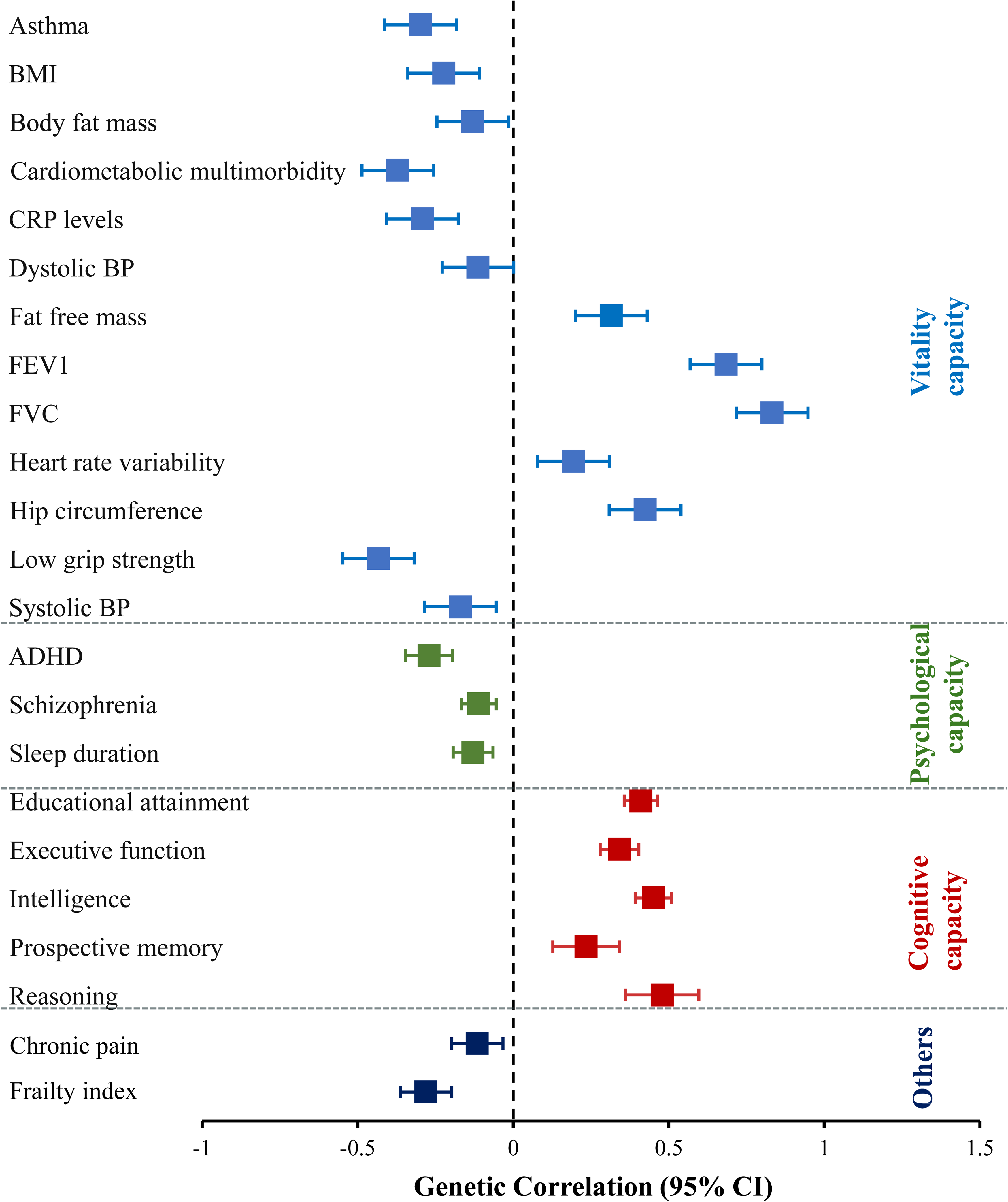
Box plots showing the results of the genetic correlation analyses and 95% confidence intervals between IC and 23 traits with statistically significant associations. The horizontal axis represents genetic correlations (r) ranging from zero (at the centre) to negative correlations on the left, and positive correlations on the right. The vertical axis lists the traits that significantly correlate with IC, organized by categories based on the domains they define. *BP: blood pressure, FVC: Forced vital capacity, FEV1: Forced expiratory volume in one second, CRP: C-reactive protein, BMI: Body mass index, ADHD: Attention deficit/hyperreactivity disorder*.

**Figure 7:**
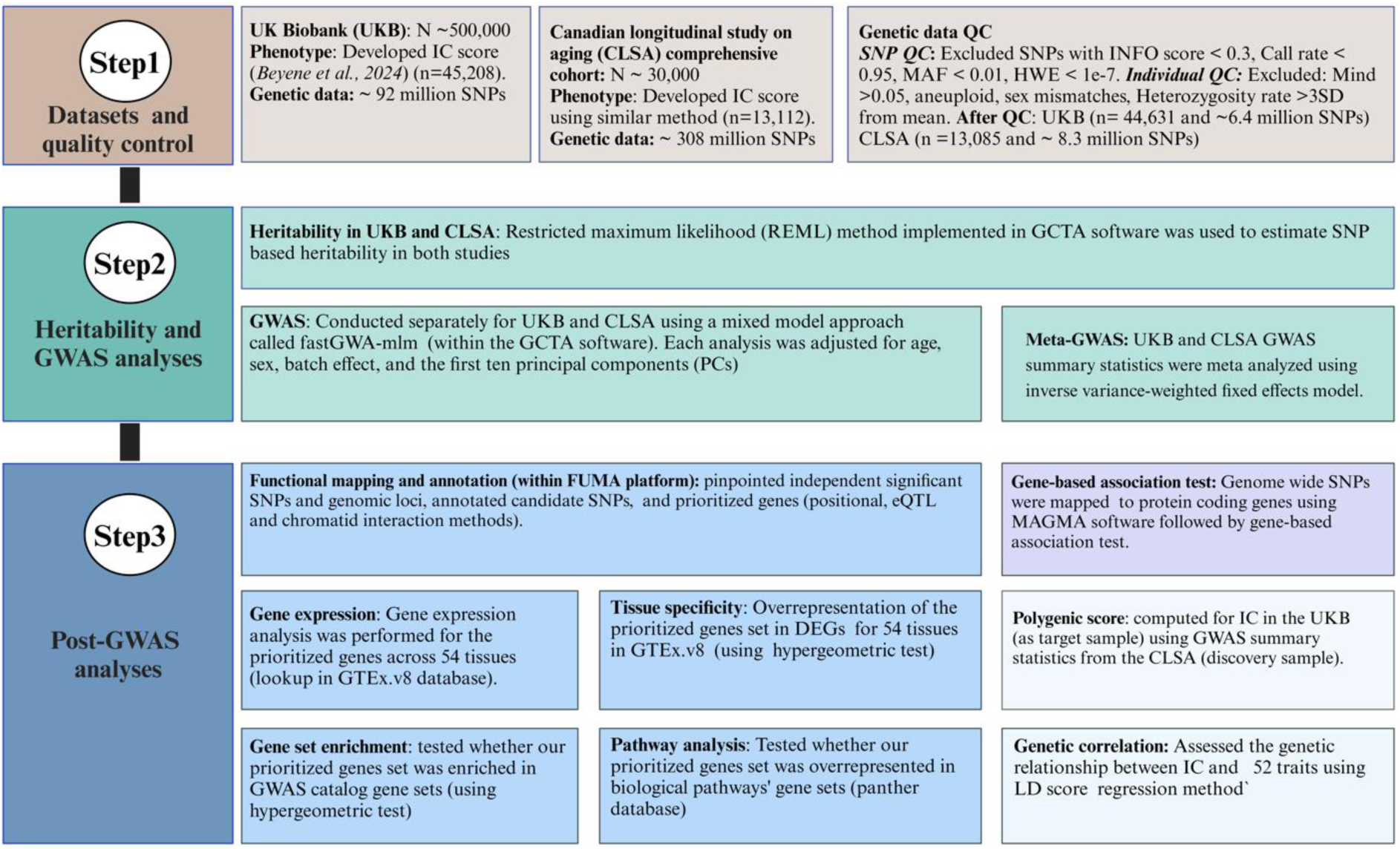
Data analysis flow chart. This chart visualizes an integrated pipeline from data preprocessing and quality control (step 1) through heritability and GWAS (step 2) and various post-GWAS analyses (step 3). *SNP = single nucleotide polymorphisms, IC = intrinsic capacity, QC = quality control, Mind = missing call rate, SD = standard deviation, INFO =imputation quality score, MAF = minor allele frequency, HWE = Hardy Weinberg equilibrium, GWAS = genome-wide association study, Meta-GWAS = Genome-wide association meta-analysis, FUMA = Functional mapping and annotation, eQTL = expression quantitative trait loci, GTEx.v8 = Genotype tissue expression project version 8, DEGs = differentially expressed gene sets, LD = linkage disequilibrium*.

## Discussion

This study presents the first GWAS for intrinsic capacity, providing novel insights into the genetic architecture of this complex trait, with key findings that genetic factors accounted for about a quarter of the total phenotypic variability in IC. We identified 38 independent SNPs across 10 genomic loci, which map to 197 genes expressed in tissues relevant to IC and are significantly overrepresented in gene sets and biological pathways related to IC. The PGS analysis revealed a significant association between PGS for IC and IC scores, implying the polygenic nature of this trait, and genetic correlation analyses found a strong genetic overlap between IC and 23 traits related to IC subdomains and associated phenotypes, highlighting potential pleiotropy.

Our findings indicated that common genetic factors accounted for about a quarter of the total phenotypic variability in IC (h^2^_SNP_ estimates ranging from 19.6% to 25.2%), with the remaining (three-quarters) variability related to environmental factors, gene-to-environment interaction effects, rare genetic variants’ effects, and stochastic variations. These estimates are comparable to previously reported heritability estimates for traits related to IC domains, including 11%-31% for cognitive function measures, 23.9% for grip strength [11], 9%-16% for functional decline [12], 22%-37% for depression [13], 25%-29% for unilateral hearing difficulty [14], and 32%-36% for FEV1 [15].

Our analysis of genes near the lead variants of IC loci and significant genes from the gene-based test revealed a plausible list of genes involved in biological processes with potential implications for IC, aligning with previous studies. For instance, several genes have been implicated in physiological processes and metabolism, such as *PTP4A2* (near rs72666714), and other Protein tyrosine phosphatases (*PTP*s) play roles in a wide range of fundamental physiological processes [16], while *HK1*(near rs72805692) is central to glucose metabolism and energy production [17]. Similarly, *LCORL* and *GPX1* are metabolic regulators [18, 19] and *MAPT* is linked to glucose metabolism and mitochondrial function [20]. Other genes, such as *RNF123* and *SCARF2*, are associated with lung function (e.g., FEV1 and FVC) and inflammatory markers (e.g., Glycan), with *SCARF2* emerging as a potential therapeutic target for chronic obstructive pulmonary disease [21, 22]. Our findings, together with these supporting evidence, highlight the relevance of the identified genes for vitality capacity-related processes, which in turn suggests a central role that vitality capacity may have for IC - supporting a working definition of vitality as an underlying physiological determinant of IC [23].

Intriguingly, some of the genes are also implicated in psychological and cognitive functioning and neurodegenerative disease phenotypes, such as gene *STAU1 -* critical for synaptic function and neuronal plasticity [24], and *MAPT, ARFGEF2, SCN4A,* and *GOSR2* are involved in vesicle trafficking, neuronal functions, and neurological conditions such as epilepsy and seizures [25–28]. Specifically, *MAPT* (which encodes tau protein), the nearest gene to the top hit intron variant rs9891103 in this study, has been implicated in more than 26 tauopathies, neurodegenerative diseases, psychiatric traits, cognitive and brain structure phenotypes [25, 29–32] and *ARHGAP27* has been implicated in various neuropsychiatric conditions [33]. Studies also showed that mutations in *PRPF3* have been discovered to cause visual impairment during old age [34] and Tauopathies (related to the *MAPT* gene) have been studied to have associations with hearing impairment [35] which implies that some of the genes associated with IC in this study may be because of their role in the sensory function. Furthermore, genes such as *MAPT* [29, 36] and *SCN4A* [37, 38] have roles in muscle, bone (osteoblast and osteoclast), and motor functions. As such, their association with IC, supported by previous findings, may imply the role of these genes in locomotive capacity. Taken together, our findings, consistent with previous studies, suggest that the genes identified in this study and implicated in neuropsychological and cognitive functions may have critical roles in the biology of IC.

Additionally, several genes are involved in programmed cell death (apoptosis) and cellular senescence – hallmarks of biological aging. *PTP4A2* (generally *PTP*s) have been reported as emerging regulators of apoptosis [39], *HK1* encodes a protein that inhibits TNF-induced apoptosis at the mitochondria [40] and *STAU1* is involved in amplifying pro-apoptotic activation during the unfolded protein response [41]. Genes such as *AL136115.1* and *CDHR4* are directly implicated in cellular senescence [42, 43]. These findings suggest a potential common biological mechanism underlying IC and the aging process, further supporting the role of these pathways in maintaining functional capacity across the lifespan.

The gene expression and tissue specificity findings also support the top-hit results discussed above. This is implied by the expression of the prioritized genes in tissues related to IC domains and their involvement in aging-related processes such as apoptosis, cellular senescence, telomere shortening, metabolism, and motor and cognitive functions. Seven of the prioritized genes (*MCL1*, *JTB, COX6B1*, *DDX5*, *PSMC5*, *PSMD4*, and *PTP4A2*) showed consistently high expression across all 54 tissues. Of these, *MCL1, JTB, PSMC5,* and *PSMD4* are involved in apoptosis [44–46]; *DDX5* slows telomere shortening and has anti-aging roles [47, 48]; *JTB* and *COX6B1* support metabolism and energy production [45, 49] with potential implications for vitality capacity. The *NSF* and *STX1B* are key for vesicular transport and neurotransmitter release [50, 51], *PSMC5* is related to cognitive deficits and motor impairments [52], and *SEMA6C* supports neuronal development and synaptic plasticity [53] - all of which are critical for cognitive functioning.

Specifically, some of the prioritized genes were expressed in tissues relevant to IC domains such as muscle (*MAPT*, *SEMA6C*, and *SUCLA2),* brain *(MAPT, PREX1, SEMA6C, and CRHR1),* heart *(RAB34, PFKM, and SLC25A4),* adipose tissue *(ICAM2, CTSK), and* nerve tissue *(IFI44, PTPDC1, and CTSK)*. *MAPT* and *SEMA6C* have been discussed above for having roles in neuronal development and synaptic plasticity. Their expression in muscle and brain tissues emphasizes their involvement in motor and cognitive function aspects of IC. The *CTSK*, expressed in the nerve tissue, has been linked to neuronal plasticity and cognitive function [54].

Similarly, *SUCLA2* has roles in muscle function and energy metabolism – and hence expression in muscle here [55] and *PREX1* is involved in processes such as axon guidance, synaptic plasticity, and neuronal migration [56], which aligns with our finding that it is expressed in brain tissues*. CRHR1* has been reported for its associations with cognitive functioning and mental health [57].

*PFKM* enhances glycolysis and oxidative phosphorylation [58], and *SLC25A4* maintains mitochondrial energy balance [59] with mutations linked to cardiomyopathy and exercise intolerance [60]. The *ICAM2* and *CTSK* genes, expressed in adipose tissues, have in previous studies been linked to inflammation, adipocyte differentiation, and obesity [61–63]. The expression of these genes in heart and adipose tissues in our study is consistent with these findings, and it may imply their potential roles in vitality capacity.

Tissue specificity analyses also supported the findings from tissue expression analysis that a significant over-representation of our prioritized genes was found in tissues potentially relevant to IC – pancreas, muscle, heart, blood, liver, and brain, and the GSEA of our prioritized genes set in the GWAS catalog gene sets showed significant enrichment in various curated gene sets for traits related to IC domains [5, 9] both highlighting the relevance of the genetic variants we identified for IC.

The pathways identified in this study, with suggestive enrichment, have been implicated in biological aging and likely provide important insight into the biological processes underlying IC. For example, enriched pathways such as cell cycle (*PSMD4, ANAPC10*), TCF-dependent signaling in response to the WNT pathway (*PSMC5, WNT3, PSMD4, FZD2*), ubiquitin-proteasome pathway (*PSMC5, PSMD4*), programmed cell death/apoptosis (*PSMC5, PSMD4, MAPT, NMT1*) modulate cell proliferation, cellular turnover, and immune function, all of which play key roles in the biology of aging [64–67]. Other enriched pathways, such as coenzyme A biosynthesis (*DCAKD*), class B/2 secretin family receptors (*CRHR1, WNT3, FZD2*), and C-type lectin receptors pathway (*ICAM2, PSMC5, MAP3K14, PSMD4*) are involved in metabolism and immune response functions [68–70] with potential implications for IC domains, mainly the vitality capacity. Likewise, the synaptic vesicle trafficking pathway (*NSF, STX1B*), the cornerstone of learning and memory function [71], and Alzheimer’s disease-presenilin pathway (*ERN1, WNT3, FZD2*), a key part of the molecular mechanism of Alzheimer’s disease [72] – have roles for cognitive function, and neuronal health, and are linked to neurodegeneration [73, 74].

The PGS analysis findings implied polygenicity of IC, revealing a significant association between IC and PGS for IC, with PGS explaining a small (0.3%) but statistically significant variability in IC. This is not special for IC as existing evidence suggests that for complex traits, the PGS explains only a small amount of variability in the phenotype [75]. It is consistent with findings for some complex traits – such as 0.7-2.1% for major depression and anxiety[76], and 0.4% - 2.1% for frailty in the ELSA cohort [77] but lower than that for traits like educational attainment (up to 12%) or cognitive performance (7-10%) [78].

The results of the genetic correlation analysis indicated how IC was interrelated with multiple traits at the genetic level, suggesting that genetic variants influencing IC also affect these other traits. The strongest genetic correlations were found between IC and traits such as FVC, FEV1, and hand-grip strength, and more than 50% of the significant correlations were with vitality-related phenotypes. These findings imply the central position of the vitality domain for IC and align with the recent definition of vitality as an underlying physiological determinant of IC [23]. Positive correlations with cognitive traits (e.g., intelligence and educational attainment) underscore that genetic factors contributing to cognitive capacity may also contribute positively to maintaining IC, whereas negative correlations with frailty and cardiometabolic multimorbidity, however, may imply that genetic factors that increase the risk of frailty and cardiometabolic multimorbidity will decrease IC, supporting evidence that physical decline accelerates aging-related deterioration [9].

The main strength of this study lies in the use of data from two big international cohort studies with detailed phenotypic and genetic data and the breadth of genetic analyses implemented, ranging from heritability testing and GWAS analyses up to various post-GWAS analyses. However, the study has certain limitations that should be considered when interpreting the findings. First, the UKB and CLSA study cohorts are of predominantly European genetic ancestry, limiting the generalizability of results to other populations. Second, IC is a complex trait encompassing multiple domains, and the genetic relationships within these domains are not well understood, complicating biological interpretation. Third, the lack of standardized IC measurement across studies may pose challenges for replication and external validity. This may be the case for our study between UKB and CLSA cohorts, but we minimized the possible difference between cohorts by using similar or closely related variables, confirming no significant differences in IC scores using appropriate tests and performing a meta-GWAS analysis. Lastly, biological interpretation from gene expression, gene set, and pathway analyses are inherently constrained by the specific databases utilized. Despite these limitations, our study provides foundational insights into the genetic architecture of IC.

In conclusion, this study provides the first comprehensive genetic analysis of IC, demonstrating its heritability and identifying novel genetic variants, genes, and pathways associated with this complex trait. GWAS revealed several distinct genomic loci, many overlapping with IC domains and related traits, suggesting the validity of our findings. Gene expression analyses highlighted the involvement of GWAS-prioritized genes in tissues critical to IC, such as muscle, brain, heart, lung, and adipose, while gene set and pathway analyses implicated significant biological pathways relevant to IC. Overall, these results enhance our understanding of the genetic underpinnings of IC and offer a foundation for exploring biological mechanisms that may serve as a baseline for future research aimed at developing personalized interventions to maintain or improve IC in aging populations. Future research should validate these findings in ancestrally diverse populations and conduct functional studies to elucidate the roles of these genes and pathways. Investigating gene-environment interactions, including the influence of lifestyle, socioeconomic status, and behavioral factors, is essential to understanding how genetic variants affect IC. Additionally, Mendelian randomization studies could establish causal links between genetic variants and IC, helping to identify modifiable factors and inform strategies to mitigate aging-related declines in IC.

## Methods

### Study Sample

We analysed data from two large international cohort studies, the UKB (N∼500,000) and the CLSA (N∼50,000). The UKB recruited adults aged 40-69 years from 22 centers across the United Kingdom (UK) between 2006 and 2010, with 94.6% of participants self-identifying as having European genetic ancestry [79, 80]. The CLSA recruited adults aged 45-85 years from all 10 provinces in Canada between 2011 and 2015, with 92.09% self-identified as being of European genetic ancestry [81–83]. Both studies collected comprehensive phenotypic data, including questionnaires, physical assessments, and biomarkers, along with genetic data. The CLSA data has genetic information for the comprehensive cohort only, and we included only the comprehensive cohort participants in this study. For the comprehensive cohort, the participants were recruited from within 25-50 kms of 11 data centres located in 7 provinces.

### Development of IC score

The IC score was developed and validated separately for each of the two cohorts. For UKB participants (N=44,631), it was previously reported [5] and using the same methodology, IC scores were computed for the CLSA participants (N = 13,112) – which were participants with full responses to 14 question items used to measure IC. A detailed description of the approach used to generate the IC score in CLSA is presented in Supplementary File 1. Briefly, these IC scores were derived using factor analysis, specifically through a bifactor model within a structural equation model framework. This approach was chosen due to its superior goodness-of-fit statistics compared to conventional methods [9].

### Genetic data quality control

Quality control was performed separately on the imputed genetic dataset of the UKB (released in July 2017) [79] and the CLSA (released in September 2018) [81] using consistent SNP and individual-based filtering criteria. First, SNPs with an imputation information score below 0.3, missing call rates exceeding 5%, minor allele frequencies below 1%, and Hardy-Weinberg equilibrium P < 1×10^-7^ were removed. Then, individuals with SNP missing rates exceeding 5%, sex mismatches between reported and genetically inferred sex, chromosome aneuploidy (ambiguous genetically imputed sex), and heterozygous/homozygous rate deviating significantly from the mean (>3SD) were excluded.

After applying these QC filters, the UKB dataset retained 6.4 million SNPs and 44,631 individuals, while the CLSA dataset retained 8.3 million SNPs and 13,085 individuals, all of whom had IC scores and were included in the genetic analysis. Full details of the QC parameters and pipeline are available in Supplementary File 2.

### Genetic data analyses

We performed a comprehensive suite of genetic data analyses to explore the genetic architecture of IC, encompassing SNP-based heritability estimation (h^2^_SNP_), GWAS, post-GWAS analyses such as functional mapping and annotation, gene-based and gene set analyses, pathway analysis, polygenic score (PGS) calculation, and genetic correlation analyses (Figure 6). Each analytical method is detailed below:

#### SNP-based heritability (h^2^SNP)

The SNP-based heritability of IC was estimated for both cohorts using the restricted maximum likelihood (REML) method implemented in GCTA software [84]. This approach partitions the total phenotypic variance into genetic and environmental components, specifically quantifying the proportion attributable to additive genetic effects of SNPs across the genome [84]. The REML framework leverages genome-wide SNP data to provide robust heritability estimates [84, 85]. The statistical significance of h^2^SNP was evaluated using a likelihood ratio test (LRT), contrasting the null hypothesis (h^2^SNP =0) with the alternative hypothesis (h^2^SNP ^≠^0).

#### Genome-Wide Association Study (GWAS)

We conducted GWAS using a linear mixed model (fastGWA-mlm) implemented in the GCTA software to identify genetic variants associated with IC. This approach adjusts for fixed effects (age, sex, batch effect, and the first ten principal components) and incorporates random effects to account for relatedness among individuals [86]. Population structure was controlled using PCs, while a sparse genetic relationship matrix (GRM), derived from an initial dense GRM with a cutoff of 0.05, was used to model relatedness [84]. The GWAS analysis was performed separately for each cohort, applying a stringent genome-wide significance threshold of P < 5×10^-8^ to correct for multiple testing. Subsequently, a genome-wide meta-analysis (Meta-GWAS) was performed using the summary statistics from the cohort-specific GWAS results. Meta-analysis was conducted using an inverse variance-weighted fixed effects model in Metal Software [84]. Meta-GWAS results provided a comprehensive set of summary statistics, which are used for downstream functional mapping and pathway analyses, described in subsequent sections

#### Post-GWAS analyses

##### Functional mapping and annotation

Functional mapping and annotation of candidate SNP were performed using FUMA (Functional Mapping and Annotation) software (Version 1.6.1) [87], leveraging 1000 Genomes Phase 3 reference panel. Independent SNPs were first identified - defined as significant SNPs with r^2^ < 0.6 in linkage disequilibrium (LD) with one another. These independent SNPs were then used to annotate candidate SNPs (those with LD with independent significant SNPs; r^2^ ≥0.6) and delineate genomic loci (by first assigning independent significant SNPs with r>0.1 into the same locus and then assigning independent significant SNPs which are closer than 250kb into same genomic locus). Gene mapping for the candidate SNPs was subsequently conducted using three complementary strategies within FUMA: (1) Positional mapping, which links SNPs to genes based on physical proximity; (2) Expression Quantitative Trait Loci (eQTL), which connects SNPs to genes whose expression levels they influence, using tissue-specific eQTL data; and (3) Chromatin interaction mappings, which identifies SNP-gene connections through long-range chromatin interactions, using data from chromatin conformation studies. These approaches generated a comprehensive list of prioritized genes associated with all candidate SNPs, which were then used as inputs for subsequent gene expression, tissue specificity, and gene set and pathway enrichment analyses.

Gene-based and gene-set analyses: The gene-based test was performed using Multi-marker Analysis of Genomic Annotation (MAGMA, version 1.08), integrated within the FUMA framework [88]. SNPs from the GWAS were mapped to genes based on a 10kb window on either side of a gene, resulting in the identification of 19,327 genes [87]. A genome-wide significance threshold for the gene-based test was set at p = 0.05/19327 = 2.6×10^-6^. All the other gene-based analyses (i.e., gene expression, tissue specificity, gene set enrichment, and pathway analyses) were conducted using the prioritized genes through FUMA’s SNP2GENE annotation.

The gene expression analysis examined the normalized gene expression values measured in reads per kilobase per million (RPKM) for each of the prioritized genes individually across 54 tissue types in the Genotype-Tissue Expression database version 8 (GTEx.v8).

A hypergeometric test with the Benjamini-Hochberg procedure for multiple test adjustment was used for tissue specificity analysis, gene set enrichment analysis (GSEA), and pathway analyses to test the overrepresentation (significant overlap) of our GWAS prioritized genes in differentially expressed gene sets (DEGs), GWAS catalog gene sets, and gene sets for biological pathways respectively [87, 89, 90]. In tissue specificity analysis, this method tested whether the prioritized genes were significantly enriched in differentially expressed gene sets (DEGs) for various tissue types [87]. DEGs were pre-calculated in the GTEx database (including all genes in the database) using a two-sided t-test comparing the expression of each gene in every tissue to all other tissues. The genes were then considered differentially expressed in each tissue if they met a Bonferroni corrected P-value of ≤ 0.05 and showed a log fold change ≥ 0.58 (equivalent to a 1.5-fold change). In GSEA, it tested if our GWAS prioritized gene set overlapped significantly with GWAS Catalog gene sets (i.e., with gene sets constructed for different traits in previous GWAS studies) [89]. In pathway analysis implemented in the PANTHER platform (incorporating panther and reactome pathways), it was used to test for enrichment of our GWAS prioritized genes set in gene sets built for different biological pathways [90].

##### Polygenic score analysis (PGS)

To assess the cumulative effect of multiple genetic variants on IC, a PGS was computed for IC in the CLSA cohort (as the target sample) using GWAS summary statistics from the UKB cohort (as the discovery sample) [91]. The association of this PGS score with the IC score was tested using a linear regression model and a decile-based stratified analysis was performed to examine patterns of IC across PGS deciles[92–94].

##### Genetic correlation analysis

To evaluate the genetic relationship of IC with other related traits, a genetic correlation analysis was performed using the LD score (LDSC) regression method [95] that utilized our GWAS summary statistics and LD scores derived from the 1000 Genomes project European genetic ancestry reference panel. A total of 52 traits were included in the analysis, selected based on a literature review of phenotypes likely associated with IC and its domains [9]. The summary statistics for these traits were obtained from the GWAS catalog [24] and details of the specific studies from which the GWAS summary statistics for each trait were obtained are provided in supplementary file 3.

## Supporting information

List of traits tested for their genetic correlation with IC

GWAS catalog reported SNPs from the IC candidate SNPs

MAGMA gene based test for all gene mapped from the whole GWAS SNPs

List of panther and reactome pathways where the IC prioritized genes have shown statistically signficant enrichment

Development of IC score in the Canadian longitudinal study of aging

partiotion of variation in IC due to genetic varaints tested and other factors along with SNP based heritability estimate

Data management and analysis codes

## Data Availability

Data, protocols, and other metadata of the UKB and CLSA are available to the scientific community upon request as per their respective data-sharing policies https://www.ukbiobank.ac.uk/enable-your-research/apply-for-access and https://www.clsa-elcv.ca/data-access/. Data for the GWAS summary statistics used for genetic correlation analysis were obtained from the GWAS catalog https://www.ebi.ac.uk/gwas/, and access links for all the 52 summary statistics are available in Supplementary file 3.

## Code Availability

The following codes and software we used for data analysis are freely available: Plink2.0 https://www.cog-genomics.org/plink/2.0/ GRM calculation https://yanglab.westlake.edu.cn/software/gcta/#MakingaGRM LDSC Heritability estimation and genetic correlation analysis https://github.com/bulik/ldsc GCTA https://yanglab.westlake.edu.cn/software/gcta/#Overview fastGWA mixed linear model https://yanglab.westlake.edu.cn/software/gcta/#fastGWA Meta GWAS analysis https://genome.sph.umich.edu/wiki/METAL_Documentation FUMA https://fuma.ctglab.nl/ Bioconductor package for leftover between genome builds https://www.bioconductor.org/install/ polygenic score https://github.com/getian107/PRScsx Panther pathway analysis https://pantherdb.org/ R and Rstudio https://www.r-project.org/ and https://posit.co/download/rstudio-desktop/. Details of customized data analysis codes (in Linux format) for quality control, GRM calculation, heritability estimation, GWAS, Meta GWAS, polygenic score, and genetic correlation analyses can be made available upon request and/or can be found in online supplementary file 2.

## Acknowledgments

MB Beyene received postgraduate scholarship support from the University of Adelaide (The University Adelaide Research Scholarship) and AT Amare is currently supported by the National Health and Medical Research Council (NHMRC) Emerging Leadership (EL1) Investigator Grant (APP2008000). Access to the UK Biobank data was partially supported by funding from the Hospital Research Foundation to Professor Visvanathan.

The funding agencies had no role in the design or conduct of the study, data collection, data analysis, data interpretation, and preparing, reviewing, or approving the manuscript.

This research was made possible using the data/biospecimens collected by the CLSA and the UK biobank. Funding for the CLSA is provided by the Government of Canada through the Canadian Institutes of Health Research (CIHR) under grant reference: LSA 94473 and the Canada Foundation for Innovation, as well as the following provinces, Newfoundland, Nova Scotia, Quebec, Ontario, Manitoba, Alberta, and British Columbia. This research has been conducted using the CLSA dataset [Comprehensive baseline v7.0 and GEN 3 data sets] under Application Number [2304009], and the UK biobank under application ID 70215. The CLSA is led by Drs. Parminder Raina, Christina Wolfson and Susan Kirkland.

The authors declare that authors alone are responsible for the views expressed in this article and do not necessarily represent the views, decisions, or policies of the institutions with which they are affiliated nor the views of the CLSA and UKB.

## Contributions

M.B. designed the study, data analysis, and methodology, wrote the initial draft, and contributed significantly to the subsequent revision of the article. A.A and R.V supervised the study, secured funding, contributed significantly to the design and analysis of the study, and drafting of the article as well as subsequent critical revision of the paper for its intellectual content. R.A., R.B, and H.B contributed significantly to the design of the study, drawing the graphs, drafting the article, and subsequent revision of the manuscript. J.B, M.C, B.B, O.T, Y.S, J.T contributed to the design of the study, contributed to the writeup of the paper, and critically revised the article for its intellectual content. All Authors critically reviewed and provided final approval of the manuscript submission.

---

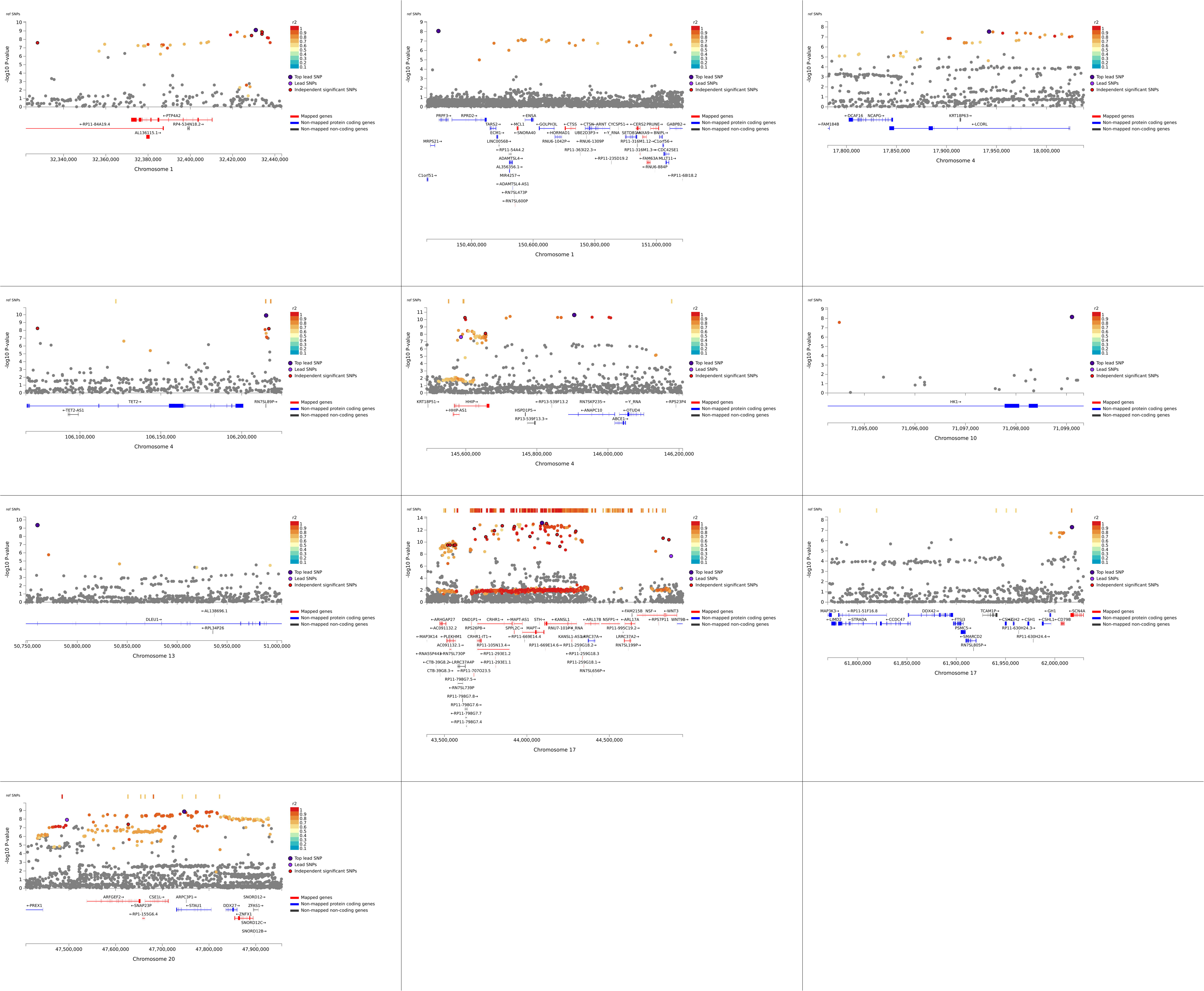
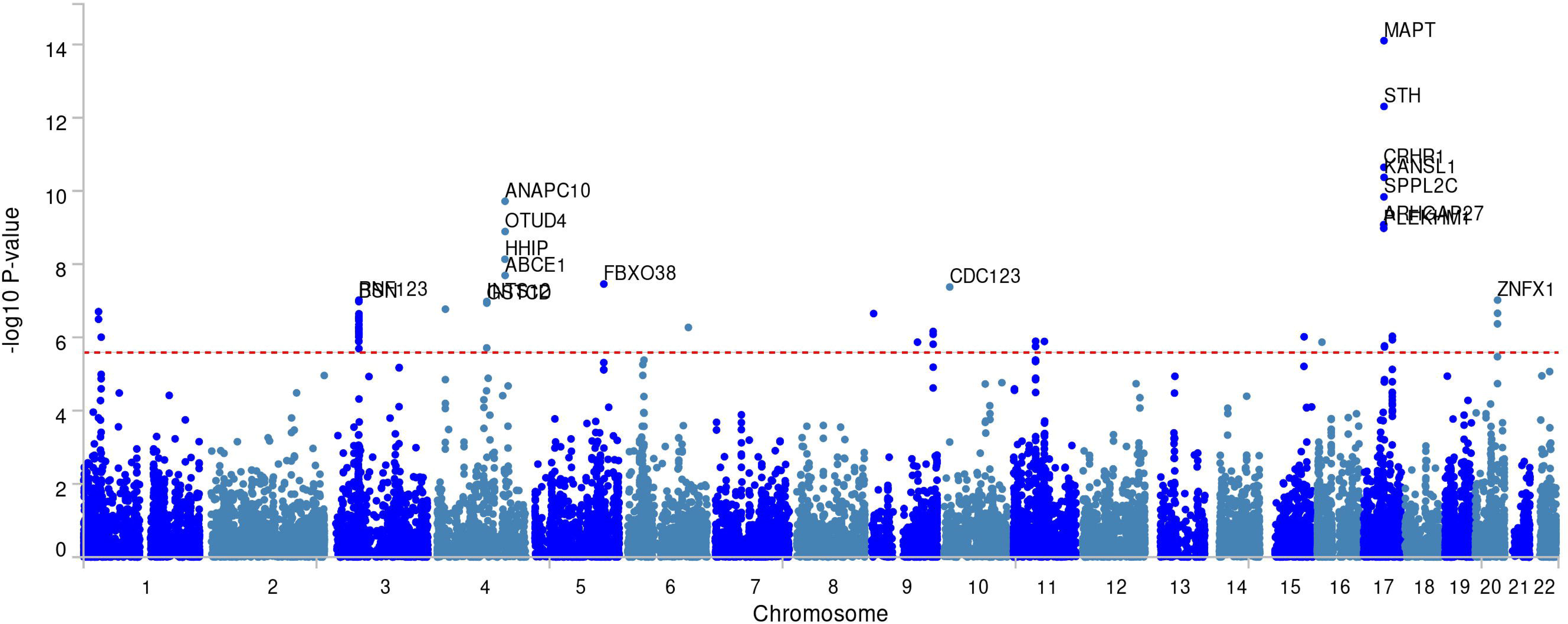
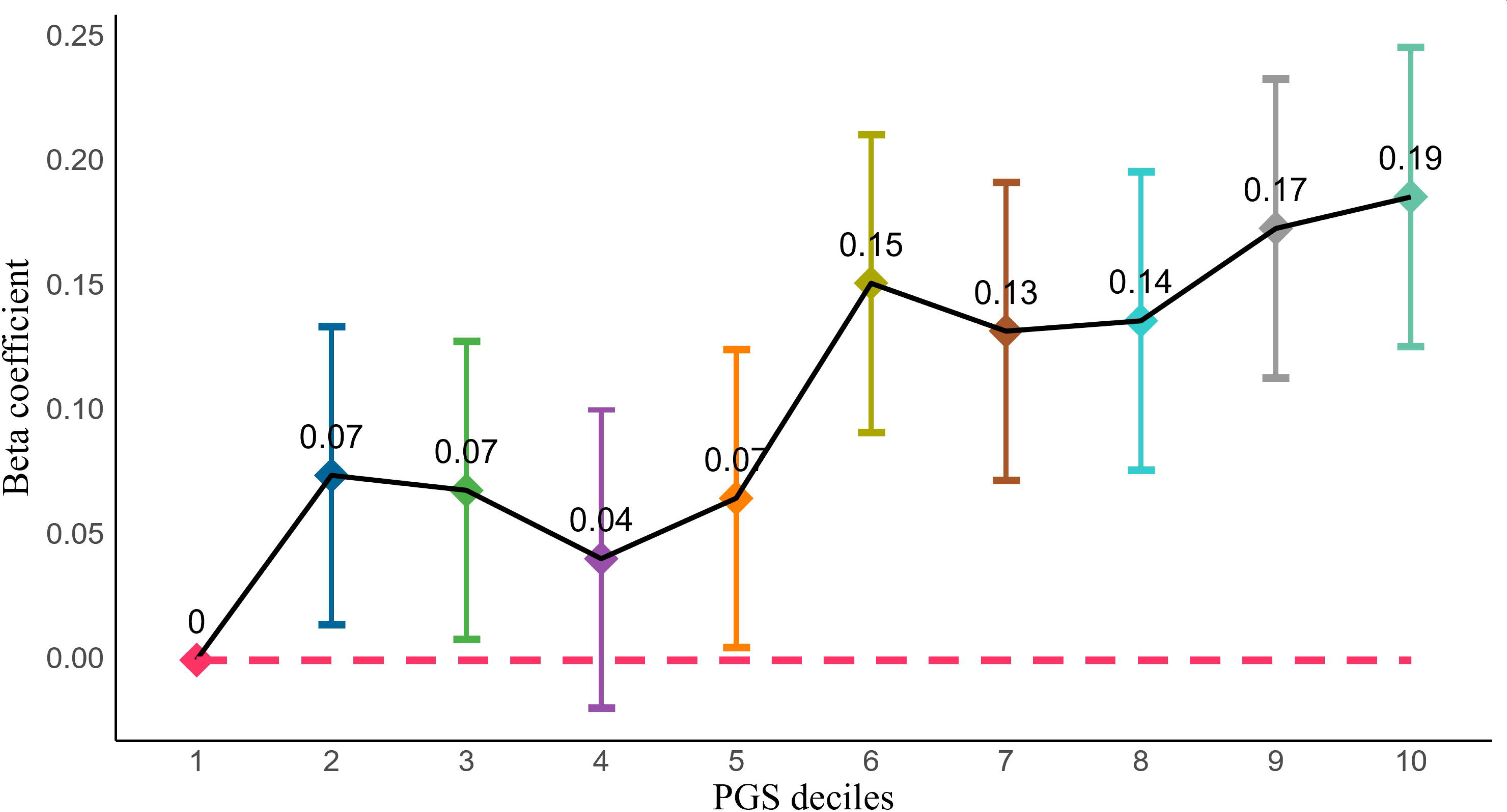

