## Supplementary material for "A genome-wide association study identified 10 novel genomic loci associated with intrinsic capacity": List of traits tested for their genetic correlation with IC

| No | trait | year | Ancestery | N | Study access link |
|---|---|---|---|---|---|
| 1 | Low_grip_strength | 2021 | European | 256,523 | https://pubmed.ncbi.nlm.nih.gov/33510174/ |
| 2 | FEV1 | 2023 | European | 475,645 | https://www.nature.com/articles/s41588-023-01314-0 |
| 3 | Hemoglobin_concentration (UKB) | 2024 | European | 174,488 | https://www.nature.com/articles/s42003-024-05874-7 |
| 4 | BMI | 2024 | European | 650,000 | https://www.nature.com/articles/s41588-024-01940-2 |
| 5 | Hip_circumference_BMIadjusted | 2021 | European | 219,872 | https://www.nature.com/articles/s41598-021-89176-6 |
| 6 | Body_fat_mass | 2024 | European | 337,196 | https://www.nature.com/articles/s41598-024-54291-7 |
| 7 | FVC | 2024 | European | 255,647 | https://pubmed.ncbi.nlm.nih.gov/38165527/ |
| 8 | ADHD | 2023 | European | 225,534 | https://pubmed.ncbi.nlm.nih.gov/36702997/ |
| 9 | Alzheimer's_disease | 2022 | European | 788,989 | https://www.nature.com/articles/s41588-022-01024-z |
| 10 | Anxiety_panic_disorder | 2019 | European | 10,240 | https://www.nature.com/articles/s41380-019-0590-2 |
| 11 | Depression_MDD | 2019 | European | 807,553 | https://www.nature.com/articles/s41593-018-0326-7 |
| 12 | Bipolar | 2021 | European | 413,466 | https://www.nature.com/articles/s41588-021-00857-4 |
| 13 | Schizophrenia | 2018 | European and east Asian | 185,864 | https://pubmed.ncbi.nlm.nih.gov/29483656/ |
| 14 | Alcohol_use_disorder | 2024 | European | 118,022 | https://www.nature.com/articles/s41562-024-01909-5 |
| 15 | Neuroticism | 2024 | European | 174,488 | https://pubmed.ncbi.nlm.nih.gov/38351177/ |
| 16 | Reasoning_verbal_numerical | 2016 | European | 36,035 | https://www.nature.com/articles/mp201645 |
| 17 | Intelligence | 2018 | European | 269,867 | https://www.nature.com/articles/s41588-018-0152-6 |
| 18 | Reaction_time | 2018 | European | 300,486 | https://www.nature.com/articles/s41467-018-04362-x |
| 19 | Executive_function | 2022 | European | 427,037 | https://www.sciencedirect.com/science/article/pii/S0006322322014056?via%3Dihub |
| 20 | Memory_performance | 2016 | European | 112, 067 | https://www.nature.com/articles/mp201645 |
| 21 | Prospective_memory | 2022 | European | 427,037 | https://www.sciencedirect.com/science/article/pii/S0006322322014056?via%3Dihub |
| 22 | Eudcational_attainment | 2024 | European (766,345) and East Asian (165,232) | 931,577 | https://www.nature.com/articles/s41562-023-01781-9 |
| 23 | Frailty_index | 2021 | European | 175,226 | https://pubmed.ncbi.nlm.nih.gov/34431594/ |
| 24 | Duration_mod_int_PA | 2022 | European | 88411 | https://pubmed.ncbi.nlm.nih.gov/35043453/ |
| 25 | Walking_pace_slow | 2021 | European | 428255 | https://pubmed.ncbi.nlm.nih.gov/34662886/ |
| 26 | Duration_of_walks | 2021 | European | 372,854 | https://pubmed.ncbi.nlm.nih.gov/34662886/ |
| 27 | Fat_free_mass | 2024 | European | 337739 | https://pubmed.ncbi.nlm.nih.gov/38538606/ |
| 28 | Bone_density | 2024 | European | 115532 | https://pubmed.ncbi.nlm.nih.gov/38965376/ |
| 29 | Hearing_difficulty_background_noise | 2021 | European | 447071 | https://pubmed.ncbi.nlm.nih.gov/34737426/ |
| 30 | Visual_impairment (progressive visual loss) | 2024 | European | 80058 | https://pubmed.ncbi.nlm.nih.gov/38965376/ |
| 31 | Retinal_detachment | 2019 | European | 364210 | https://pubmed.ncbi.nlm.nih.gov/31816047/ |
| 32 | Age-related hearing impairment | 2020 | European | 330759 | https://pubmed.ncbi.nlm.nih.gov/32986727/ |
| 33 | Cardiometabolic_multimorbidity | 2024 | European | 367,147 | https://pubmed.ncbi.nlm.nih.gov/38409652/ |
| 34 | Parkinson_disease | 2023 | asian and 1497 Hispanic) | 611,485 | https://www.nature.com/articles/s41588-023-01584-8 |
| 35 | Cancer (UKB data field 2453) | 2021 | European | 454,736 | https://pubmed.ncbi.nlm.nih.gov/34737426/ |
| 36 | Multiple_system_atrophy | 2024 | European | 8016 | https://pubmed.ncbi.nlm.nih.gov/38701790/ |
| 37 | Dementias | 2018 | European | 403339 | https://pubmed.ncbi.nlm.nih.gov/30104761/ |
| 38 | Systolic_BP | 2024 | European | 1028980 | https://pubmed.ncbi.nlm.nih.gov/38689001/ |
| 39 | Dystolic_BP | 2024 | European | 1028980 | https://pubmed.ncbi.nlm.nih.gov/38689001/ |
| 40 | Essential_Hypertension | 2021 | European | 331754 | https://pubmed.ncbi.nlm.nih.gov/34662886/ |
| 41 | Total_lipid_LDL | 2024 | European | 93799 | https://pubmed.ncbi.nlm.nih.gov/39278973/ |
| 42 | Total_lipid_HDL | 2024 | European | 93799 | https://pubmed.ncbi.nlm.nih.gov/39278973/ |
| 43 | fasting_glucose | 2023 | European, 3153 Hispanic, 2405 South Asian | 129665 | https://pubmed.ncbi.nlm.nih.gov/39280063/ |
| 44 | Telomere_length | 2022 | European | 902 | https://pubmed.ncbi.nlm.nih.gov/35681050/ |
| 45 | fasting_insulin | 2023 | European, 1142 Hispanic, 2155 South Asian | 104140 | https://pubmed.ncbi.nlm.nih.gov/39280063/ |
| 46 | Heart_rate_variability | 2023 | European | 46075 | https://pubmed.ncbi.nlm.nih.gov/37803156/ |
| 47 | Asthma | 2023 | European | 381782 | https://pubmed.ncbi.nlm.nih.gov/37377600/ |
| 48 | CRP_levels (UKB) | 2024 | European | 174488 | https://pubmed.ncbi.nlm.nih.gov/38351177/ |
| 49 | Chronic_pain | 2024 | European | 360249 | https://pubmed.ncbi.nlm.nih.gov/38965376/ |
| 50 | Insulin_resistance | 2023 | European | 53287 | https://pubmed.ncbi.nlm.nih.gov/37291194/ |
| 51 | Sleep_duration | 2018 | European | 91105 | https://pubmed.ncbi.nlm.nih.gov/30531941/ |
| 52 | kidney_function_GFR | 2024 | European | 406504 | https://pubmed.ncbi.nlm.nih.gov/39256582/ |
