## Supplementary material for "A genome-wide association study identified 10 novel genomic loci associated with intrinsic capacity": GWAS catalog reported SNPs from the IC candidate SNPs

| Genomic Locus | IndSigSNP | chr | bp | snp | Date Added To Catalog | PMID | FirstAuth | Date | Journal | Link | Study | Trait | InitialN | Replicatio | Region | ReportedGene | MappedGene | UpGene | DownGene | SNP_Gene_ID | UpGeneDi | DownGen | Strongest | SNPs | marged | SNP_ID_cu | Context | intergenic | RiskAF | P | Pmlog | Ptext | OrBeta | 95CI | Platform | CNV |
|---|---|---|---|---|---|---|---|---|---|---|---|---|---|---|---|---|---|---|---|---|---|---|---|---|---|---|---|---|---|---|---|---|---|---|---|---|
| 1 | 1:32327685 | 1 | 32356815 | rs34701865 | 24/08/2021 | 3.4E+07 | Christakoudi S | 21/05/2021 | Sci Rep | www.ncbi.nlm.nih.gov/pubmed/34021172 | GWAS of allometric body-shape indices in UK Biobank identifies loci suggesting associations with morphogenesis, organogenesis, adrenal cell renewal and cancer. | Hip circumference adjusted for BMI | 186,825 B | NA | 1p35.2 | RP11-84A19.4 | SPOCD1 - PTP4A | ENSG00000134668 | ENSG00000184007 | | 75163 | 15207 | rs34701865-T | rs34701865 | 0 | 34701865 | intron_variant | 1 | NR | 1.00E-08 | 8 | | 0.036605 | [0.024-0.0 | Affymetrix [323123] (imputed) | N |
| 1 | 1:32327685 | 1 | 32397127 | rs72665000 | 17/04/2020 | 3.2E+07 | Zhu Z | 24/10/2019 | J Allergy Clin Immunol | www.ncbi.nlm.nih.gov/pubmed/31669095 | Shared Genetic and Experimental Links between Obesity-Related Traits and Asthma Subtypes in UK Biobank. | Waist circumference adjusted for body mass index | 457,690 E | NA | 1p35.2 | NR | PTP4A2 | | | ENSG00000184007 | NA | NA | rs72665000-? | rs72665000 | 0 | 72665000 | intron_variant | 0 | NR | 8.00E-10 | 9.09691 | | NA | | Affymetrix [~ 8270000] (imputed) | N |
| 2 | 1:150292561 | 1 | 1.51E+08 | rs11580946 | 29/06/2021 | 3.3E+07 | Sinnott-Armstrong N | 18/01/2021 | Nat Genet | www.ncbi.nlm.nih.gov/pubmed/33462484 | Genetics of 35 blood and urine biomarkers in the UK Biobank. | Alanine aminotransferase levels | 342,387 E | NA | 1q21.2 | NR | MCL1 | | | ENSG00000143384 | NA | NA | rs11580946-A | rs11580946 | 0 | 11580946 | missense_variant | 0 | NR | 1.00E-14 | 14 | | 0.0737 | [0.055-0.0 | Affymetrix [9000000] (imputed) | N |
| 2 | 1:150292561 | 1 | 1.51E+08 | rs11580946 | 29/06/2021 | 3.3E+07 | Sinnott-Armstrong N | 18/01/2021 | Nat Genet | www.ncbi.nlm.nih.gov/pubmed/33462484 | Genetics of 35 blood and urine biomarkers in the UK Biobank. | Aspartate aminotransferase to alanine aminotransferase ratio | 341,165 E | NA | 1q21.2 | NR | MCL1 | | | ENSG00000143384 | NA | NA | rs11580946-A | rs11580946 | 0 | 11580946 | missense_variant | 0 | NR | 2.00E-09 | 8.69897 | | 0.0573 | [0.039-0.0 | Affymetrix [9000000] (imputed) | N |
| 2 | 1:150292561 | 1 | 1.51E+08 | rs11580946 | 29/06/2021 | 3.3E+07 | Sinnott-Armstrong N | 18/01/2021 | Nat Genet | www.ncbi.nlm.nih.gov/pubmed/33462484 | Genetics of 35 blood and urine biomarkers in the UK Biobank. | Gamma glutamyl transferase levels | 342,339 E | NA | 1q21.2 | NR | MCL1 | | | ENSG00000143384 | NA | NA | rs11580946-A | rs11580946 | 0 | 11580946 | missense_variant | 0 | NR | 1.00E-19 | 19 | | 0.0864 | [0.068-0.1 | Affymetrix [9000000] (imputed) | N |
| 2 | 1:150292561 | 1 | 1.51E+08 | rs11580946 | 21/09/2021 | 3.3E+07 | Surendran P | 23/11/2020 | Nat Genet | www.ncbi.nlm.nih.gov/pubmed/33230300 | Discovery of rare variants associated with blood pressure regulation through meta-analysis of 1.3 million individuals. | Pulse pressure | 810,865 E | 354,096 E | 1q21.2 | | MCL1 | | | ENSG00000143384 | NA | NA | rs11580946-A | rs11580946 | 0 | 11580946 | missense_variant | 0 | 0.015 | 3.00E-09 | 8.522879 | | 0.388206 | [0.26-0.52 | Affymetrix, Illumina [242866] (imputed) | N |
| 2 | 1:150292561 | 1 | 1.51E+08 | rs11580946 | 21/09/2021 | 3.3E+07 | Surendran P | 23/11/2020 | Nat Genet | www.ncbi.nlm.nih.gov/pubmed/33230300 | Discovery of rare variants associated with blood pressure regulation through meta-analysis of 1.3 million individuals. | Systolic blood pressure | 810,865 E | 354,096 E | 1q21.2 | | MCL1 | | | ENSG00000143384 | NA | NA | rs11580946-A | rs11580946 | 0 | 11580946 | missense_variant | 0 | 0.0147 | 2.00E-08 | 7.69897 | | 0.523103 | [0.34-0.7] | Affymetrix, Illumina [247314] (imputed) | N |
| 2 | 1:150292561 | 1 | 1.51E+08 | rs11580946 | 8/02/2019 | 3.1E+07 | Kichaev G | 27/12/2018 | Am J Hum Genet | www.ncbi.nlm.nih.gov/pubmed/30595370 | Leveraging Polygenic Functional Enrichment to Improve GWAS Power. | Systolic blood pressure | approxima | NA | 1q21.2 | | MCL1 | | | ENSG00000143384 | NA | NA | rs11580946-? | rs11580946 | 0 | 11580946 | missense_variant | 0 | NR | 1.00E-09 | 9 | | NA | | NR [~ 8900000] (imputed) | N |
| 2 | 1:150292561 | 1 | 1.5E+08 | rs11581793 | 22/03/2021 | 3.3E+07 | Nielsen JB | 18/12/2020 | Nat Commun | www.ncbi.nlm.nih.gov/pubmed/33339817 | Loss-of-function genomic variants highlight potential therapeutic targets for cardiovascular disease. | Alanine aminotransferase levels | 76,081 Eur | NA | 1q21.2 | MRPS21, PRPF3 | MRPS21 - PRPF3 | ENSG00000266472 | ENSG00000117360 | | 11134 | 1366 | rs11581793-? | rs11581793 | 0 | 11581793 | regulatory_region_variant | 1 | | 2.00E-08 | 7.69897 | | 0.1441 | [0.094-0.1 | Illumina [31500000] (imputed) | N |
| 2 | 1:150292561 | 1 | 1.5E+08 | rs187799739 | 21/05/2019 | 3.1E+07 | Kichaev G | 27/12/2018 | Am J Hum Genet | www.ncbi.nlm.nih.gov/pubmed/30595370 | Leveraging Polygenic Functional Enrichment to Improve GWAS Power. | Height | approxima | NA | 1q21.2 | | RPRD2 | | | ENSG00000163125 | NA | NA | rs187799739-? | rs187799739 | 0 | 1.88E+08 | intron_variant | 0 | NR | 1.00E-33 | 33 | | NA | | NR [~ 8900000] (imputed) | N |
| 2 | 1:150292561 | 1 | 1.5E+08 | rs187799739 | 8/02/2019 | 3.1E+07 | Kichaev G | 27/12/2018 | Am J Hum Genet | www.ncbi.nlm.nih.gov/pubmed/30595370 | Leveraging Polygenic Functional Enrichment to Improve GWAS Power. | Lung function (FVC) | approxima | NA | 1q21.2 | | RPRD2 | | | ENSG00000163125 | NA | NA | rs187799739-? | rs187799739 | 0 | 1.88E+08 | intron_variant | 0 | NR | 8.00E-12 | 11.09691 | | NA | | NR [~ 8900000] (imputed) | N |
| 2 | 1:150292561 | 1 | 1.51E+08 | rs190596489 | 16/11/2018 | 3E+07 | Emilsson V | 2/08/2018 | Science | www.ncbi.nlm.nih.gov/pubmed/30072576 | Co-regulatory networks of human serum proteins link genetics to disease. | Blood protein levels | 3,200 Euro | NA | 1q21.2 | MCL1 | MCL1 - ENSA | ENSG00000143384 | ENSG00000143420 | | 9829 | 11284 | rs190596489-A | rs190596489 | 0 | 1.91E+08 | intergenic_variant | 1 | 0.992969 | 2.00E-07 | 6.69897 | (MCL1, 10 | 0.778233 | [0.48-1.07 | Illumina [> 1000000] (imputed) | N |
| 2 | 1:150292561 | 1 | 1.51E+08 | rs28521412 | 8/02/2019 | 3.1E+07 | Hubel C | 28/12/2018 | Am J Med Genet B Neuropsychiatr Genet | www.ncbi.nlm.nih.gov/pubmed/30593698 | Genomics of body fat percentage may contribute to sex bias in anorexia nervosa. | Fat-free mass | 70,700 Eur | NA | 1q21.3 | NR | CERS2 - ANXA9 | ENSG00000143418 | ENSG00000143412 | | 7241 | 5 | rs28521412-? | rs28521412 | 0 | 28521412 | intergenic_variant | 1 | NR | 1.00E-06 | 6 | (female) | 0.48049 | [0.29-0.67 | Affymetrix [7794483] (imputed) | N |
| 2 | 1:150292561 | 1 | 1.51E+08 | rs28521412 | 8/02/2019 | 3.1E+07 | Hubel C | 28/12/2018 | Am J Med Genet B Neuropsychiatr Genet | www.ncbi.nlm.nih.gov/pubmed/30593698 | Genomics of body fat percentage may contribute to sex bias in anorexia nervosa. | Fat-free mass | 70,700 Eur | NA | 1q21.3 | NR | CERS2 - ANXA9 | ENSG00000143418 | ENSG00000143412 | | 7241 | 5 | rs28521412-? | rs28521412 | 0 | 28521412 | intergenic_variant | 1 | NR | 2.00E-06 | 5.69897 | | 0.41951 | [0.25-0.59 | Affymetrix [7794483] (imputed) | N |
| 2 | 1:150292561 | 1 | 1.51E+08 | rs28521412 | 24/08/2021 | 3.4E+07 | Christakoudi S | 21/05/2021 | Sci Rep | www.ncbi.nlm.nih.gov/pubmed/34021172 | GWAS of allometric body-shape indices in UK Biobank identifies loci suggesting associations with morphogenesis, organogenesis, adrenal cell renewal and cancer. | Hip circumference adjusted for BMI | 219,872 B | NA | 1q21.3 | ANXA9 | CERS2 - ANXA9 | ENSG00000143418 | ENSG00000143412 | | 7241 | 5 | rs28521412-A | rs28521412 | 0 | 28521412 | intergenic_variant | 1 | NR | 1.00E-16 | 16 | | 0.094603 | [0.072-0.1 | Affymetrix [321016] (imputed) | N |
| 2 | 1:150292561 | 1 | 1.51E+08 | rs6684154 | 13/08/2021 | 3.4E+07 | Ward LD | 27/07/2021 | Nat Commun | www.ncbi.nlm.nih.gov/pubmed/34315874 | GWAS of serum ALT and AST reveals an association of SLC30A10 Thr95Ile with hypermanganesemia symptoms. | Alanine aminotransferase levels | 387,859 B | NA | 1q21.3 | PRUNE1 | PRUNE1 | | | ENSG00000143363 | NA | NA | rs6684154-? | rs6684154 | 0 | 6684154 | intron_variant | 0 | NR | 3.00E-15 | 14.52288 | | 0.0142 | [0.011-0.0 | Affymetrix [21347608] (imputed) | N |
| 2 | 1:150292561 | 1 | 1.51E+08 | rs6684154 | 13/08/2021 | 3.4E+07 | Ward LD | 27/07/2021 | Nat Commun | www.ncbi.nlm.nih.gov/pubmed/34315874 | GWAS of serum ALT and AST reveals an association of SLC30A10 Thr95Ile with hypermanganesemia symptoms. | Aspartate aminotransferase levels | 386,570 B | NA | 1q21.3 | PRUNE1 | PRUNE1 | | | ENSG00000143363 | NA | NA | rs6684154-? | rs6684154 | 0 | 6684154 | intron_variant | 0 | NR | 3.00E-08 | 7.522879 | | 0.0061 | [0.0037-0. | Affymetrix [21347608] (imputed) | N |
| 2 | 1:150292561 | 1 | 1.51E+08 | rs6684154 | 9/06/2021 | 3.4E+07 | Pazoki R | 10/05/2021 | Nat Commun | www.ncbi.nlm.nih.gov/pubmed/33972514 | Genetic analysis in European ancestry individuals identifies 517 loci associated with liver enzymes. | Liver enzyme levels (alanine transaminase) | 437,267 E | 315,572 E | 1q21.3 | PRUNE1 | PRUNE1 | | | ENSG00000143363 | NA | NA | rs6684154-C | rs6684154 | 0 | 6684154 | intron_variant | 0 | 0.984478 | 1.00E-21 | 21 | | 0.015071 | [0.012-0.0 | Affymetrix [13995440] (imputed) | N |
| 2 | 1:150292561 | 1 | 1.51E+08 | rs72704611 | 17/04/2020 | 3.2E+07 | Zhu Z | 24/10/2019 | J Allergy Clin Immunol | www.ncbi.nlm.nih.gov/pubmed/31669095 | Shared Genetic and Experimental Links between Obesity-Related Traits and Asthma Subtypes in UK Biobank. | Waist-to-hip ratio adjusted for BMI | 457,690 E | NA | 1q21.3 | NR | CTSS | | | ENSG00000163131 | NA | NA | rs72704611-? | rs72704611 | 0 | 72704611 | intron_variant | 0 | NR | 1.00E-08 | 8 | | NA | | Affymetrix [~ 8270000] (imputed) | N |
| 3 | 4:17942560 | 4 | 17792869 | rs12640250 | 8/01/2020 | 3E+07 | Mahajan A | 8/10/2018 | Nat Genet | www.ncbi.nlm.nih.gov/pubmed/30297969 | Fine-mapping type 2 diabetes loci to single-variant resolution using high-density imputation and islet-specific epigenome maps. | Type 2 diabetes | 74,124 Eur | NA | 4p15.31 | LCORL | FAM184B - DCAF | ENSG00000047662 | ENSG00000163257 | | 9625 | 9409 | rs12640250-C | rs12640250 | 0 | 12640250 | intergenic_variant | 1 | 0.7149 | 4.00E-08 | 7.39794 | | 1.04 | [1.03-1.05 | Affymetrix, Illumina [~ 27000000] (imputed) | N |
| 3 | 4:17942560 | 4 | 18015156 | rs16896276 | 26/01/2021 | 3.2E+07 | Fan Q | 19/03/2020 | Commun Biol | www.ncbi.nlm.nih.gov/pubmed/32193507 | Genome-wide association meta-analysis of corneal curvature identifies novel loci and shared genetic influences across axial length and refractive error. | Corneal curvature | 29,580 Eur | 88,218 Eur | 4p15.31 | | LCORL | | | ENSG00000178177 | NA | NA | rs16896276-A | rs16896276 | 0 | 16896276 | intron_variant | 0 | 0.26 | 1.00E-31 | 31 | | 0.013 | mm increa | Affymetrix, Illumina [8936601] (imputed) | N |
| 3 | 4:17942560 | 4 | 18015156 | rs16896276 | 26/01/2021 | 3.2E+07 | Fan Q | 19/03/2020 | Commun Biol | www.ncbi.nlm.nih.gov/pubmed/32193507 | Genome-wide association meta-analysis of corneal curvature identifies novel loci and shared genetic influences across axial length and refractive error. | Corneal curvature | 29,580 Eur | NA | 4p15.31 | | LCORL | | | ENSG00000178177 | NA | NA | rs16896276-A | rs16896276 | 0 | 16896276 | intron_variant | 0 | 0.27 | 1.00E-07 | 7 | | 0.014 | mm increa | Affymetrix, Illumina [8936601] (imputed) | N |
| 3 | 4:17942560 | 4 | 18015156 | rs16896276 | 26/01/2021 | 3.2E+07 | Fan Q | 19/03/2020 | Commun Biol | www.ncbi.nlm.nih.gov/pubmed/32193507 | Genome-wide association meta-analysis of corneal curvature identifies novel loci and shared genetic influences across axial length and refractive error. | Corneal curvature | 35,422 ind | NA | 4p15.31 | | LCORL | | | ENSG00000178177 | NA | NA | rs16896276-A | rs16896276 | 0 | 16896276 | intron_variant | 0 | NR | 9.00E-09 | 8.045757 | | 0.0143 | [0.0096-0. | Affymetrix, Illumina [8936601] (imputed) | N |
| 3 | 4:17942560 | 4 | 18015156 | rs16896276 | 16/11/2022 | 3.5E+07 | Knol MJ | 15/11/2021 | Hum Mol Genet | www.ncbi.nlm.nih.gov/pubmed/34791242 | Genetic architecture of orbital telorism. | Orbital telorism | 34,130 Eur | NA | 4p15.31 | | LCORL | | | ENSG00000178177 | NA | NA | rs16896276-? | rs16896276 | 0 | 16896276 | intron_variant | 0 | NR | 4.00E-09 | 8.39794 | | 0.1431 | [0.095-0.1 | Affymetrix, Illumina [NR] (imputed) | N |
| 3 | 4:17942560 | 4 | 18015156 | rs16896276 | 17/04/2020 | 3.2E+07 | Zhu Z | 24/10/2019 | J Allergy Clin Immunol | www.ncbi.nlm.nih.gov/pubmed/31669095 | Shared Genetic and Experimental Links between Obesity-Related Traits and Asthma Subtypes in UK Biobank. | Waist circumference adjusted for body mass index | 457,690 E | NA | 4p15.31 | NR | LCORL | | | ENSG00000178177 | NA | NA | rs16896276-? | rs16896276 | 0 | 16896276 | intron_variant | 0 | NR | 9.00E-20 | 19.04576 | | NA | | Affymetrix [~ 8270000] (imputed) | N |
| 3 | 4:17942560 | 4 | 18025484 | rs2011603 | 19/10/2018 | 3E+07 | Lee JJ | 23/07/2018 | Nat Genet | www.ncbi.nlm.nih.gov/pubmed/30038396 | Gene discovery and polygenic prediction from a genome-wide association study of educational attainment in 1.1 million individuals. | Educational attainment (years of education) | up to 1,13 | NA | 4p15.31 | | LCORL - NA | ENSG00000178177 | | | 1985 | NA | rs2011603-A | rs2011603 | 0 | 2011603 | intergenic_variant | 1 | 0.7359 | 1.00E-08 | 8 | | 0.0091 | [0.006-0.0 | Affymetrix, Illumina [10000000] (imputed) | N |
| 3 | 4:17942560 | 4 | 18025484 | rs2011603 | 7/03/2019 | 3.1E+07 | Giri A | 21/12/2018 | Nat Genet | www.ncbi.nlm.nih.gov/pubmed/30578418 | Trans-ethnic association study of blood pressure determinants in over 750,000 individuals. | Pulse pressure | 365,998 E | 299,024 E | 4p15.31 | intergenic | LCORL - NA | ENSG00000178177 | | | 1985 | NA | rs2011603-A | rs2011603 | 0 | 2011603 | intergenic_variant | 1 | 0.7256 | 2.00E-11 | 10.69897 | | 0.1432 | [0.1-0.18] | Affymetrix [NR] (imputed) | N |
| 3 | 4:17942560 | 4 | 18025484 | rs2011603 | 14/09/2020 | 3.3E+07 | Vujkovic M | 15/06/2020 | Nat Genet | www.ncbi.nlm.nih.gov/pubmed/32541925 | Discovery of 318 new risk loci for type 2 diabetes and related vascular outcomes among 1.4 million participants in a multi-ancestry meta-analysis. | Type 2 diabetes | 148,726 E | NA | 4p15.31 | NR | LCORL - NA | ENSG00000178177 | | | 1985 | NA | rs2011603-G | rs2011603 | 0 | 2011603 | intergenic_variant | 1 | 0.2752 | 2.00E-13 | 12.69897 | | 0.0383 | [0.028-0.0 | Affymetrix, Illumina [1198787] (imputed) | N |
| 3 | 4:17942560 | 4 | 17998426 | rs2061456 | 15/01/2020 | 3.1E+07 | Plotnikov D | 16/05/2019 | Br J Ophthalmol | www.ncbi.nlm.nih.gov/pubmed/31097437 | Association between birth weight and refractive error in adulthood: a Mendelian randomisation study. | Birth weight | 188,039 E | NA | 4p15.31 | NR | LCORL | | | ENSG00000178177 | NA | NA | rs2061456-A | rs2061456 | 0 | 2061456 | intron_variant | 0 | 0.25 | 4.00E-25 | 24.39794 | | 0.038 | [0.03-0.04 | Affymetrix [up to 10400000] (imputed) | N |
| 3 | 4:17942560 | 4 | 17998426 | rs2061456 | 17/07/2019 | 2.9E+07 | Tachmazidou I | 1/06/2017 | Am J Hum Genet | www.ncbi.nlm.nih.gov/pubmed/28552196 | Whole-Genome Sequencing Coupled to Imputation Discovers Genetic Signals for Anthropometric Traits. | Height | 1,249 who | 205,003 E | 4p15.31 | LCORL | LCORL | | | ENSG00000178177 | NA | NA | rs2061456-A | rs2061456 | 0 | 2061456 | intron_variant | 0 | 0.261 | 1.00E-19 | 19 | | 0.0656 | [0.051-0.0 | Affymetrix, Illumina, Perlegen [15844966] (imputed) | N |
| 3 | 4:17942560 | 4 | 17998426 | rs2061456 | 5/08/2022 | 3.6E+07 | Chiou JS | 13/07/2022 | BMC Med | www.ncbi.nlm.nih.gov/pubmed/35831902 | Your height affects your health: genetic determinants and health-related outcomes in Taiwan. | Height | 67,452 Ha | NA | 4p15.31 | | LCORL | | | ENSG00000178177 | NA | NA | rs2061456-A | rs2061456 | 0 | 2061456 | intron_variant | 0 | NR | 9.00E-57 | 56.04576 | | 0.08 | [0.07-0.09 | Affymetrix [NR] (imputed) | N |
| 3 | 4:17942560 | 4 | 17812615 | rs2074974 | 24/08/2021 | 3.4E+07 | Christakoudi S | 21/05/2021 | Sci Rep | www.ncbi.nlm.nih.gov/pubmed/34021172 | GWAS of allometric body-shape indices in UK Biobank identifies loci suggesting associations with morphogenesis, organogenesis, adrenal cell renewal and cancer. | Hip circumference adjusted for BMI | 219,872 B | NA | 4p15.31 | NCAPG | NCAPG | | | ENSG00000109805 | NA | NA | rs2074974-A | rs2074974 | 0 | 2074974 | 5_prime_UTR_variant | 0 | NR | 1.00E-11 | 11 | | 0.021246 | [0.015-0.0 | Affymetrix [321016] (imputed) | N |
| 3 | 4:17942560 | 4 | 17917781 | rs2174633 | 28/01/2019 | 3.1E+07 | Morris JA | 31/12/2018 | Nat Genet | www.ncbi.nlm.nih.gov/pubmed/30598549 | An atlas of genetic influences on osteoporosis in humans and mice. | Heel bone mineral density | 426,824 B | NA | 4p15.31 | LCORL | LCORL | | | ENSG00000178177 | NA | NA | rs2174633-A | rs2174633 | 0 | 2174633 | intron_variant | 0 | 0.265235 | 5.00E-13 | 12.30103 | | 0.015139 | [0.011-0.0 | NR [13737936] (imputed) | N |
| 3 | 4:17942560 | 4 | 17917781 | rs2174633 | 8/02/2019 | 3.1E+07 | Kichaev G | 27/12/2018 | Am J Hum Genet | www.ncbi.nlm.nih.gov/pubmed/30595370 | Leveraging Polygenic Functional Enrichment to Improve GWAS Power. | Heel bone mineral density | approxima | NA | 4p15.31 | | LCORL | | | ENSG00000178177 | NA | NA | rs2174633-? | rs2174633 | 0 | 2174633 | intron_variant | 0 | NR | 2.00E-18 | 17.69897 | | NA | | NR [~ 8900000] (imputed) | N |
| 3 | 4:17942560 | 4 | 17917781 | rs2174633 | 18/10/2018 | 3E+07 | Kim SK | 26/07/2018 | PLoS One | www.ncbi.nlm.nih.gov/pubmed/30048462 | Identification of 613 new loci associated with heel bone mineral density and a polygenic risk score for bone mineral density, osteoporosis and fracture. | Heel bone mineral density | 394,929 E | NA | 4p15.31 | | LCORL | | | ENSG00000178177 | NA | NA | rs2174633-? | rs2174633 | 0 | 2174633 | intron_variant | 0 | NR | 2.00E-12 | 11.69897 | | 0.016269 | [0.012-0.0 | Affymetrix [20259828] (imputed) | N |
| 3 | 4:17942560 | 4 | 17917781 | rs2174633 | 5/08/2019 | 3.1E+07 | Warrington NM | 1/05/2019 | Nat Genet | www.ncbi.nlm.nih.gov/pubmed/31043758 | Maternal and fetal genetic effects on birth weight and their relevance to cardio-metabolic risk factors. | Offspring birth weight | 210,267 E | NR | 4p15.31 | LCORL, DCAF16 | LCORL | | | ENSG00000178177 | NA | NA | rs2174633-A | rs2174633 | 0 | 2174633 | intron_variant | 0 | 0.267868 | 4.00E-12 | 11.39794 | | 0.023908 | [0.017-0.0 | Affymetrix, Illumina [14869762] (imputed) | N |
| 3 | 4:17942560 | 4 | 17972372 | rs2320299 | 21/09/2021 | 3.3E+07 | Surendran P | 23/11/2020 | Nat Genet | www.ncbi.nlm.nih.gov/pubmed/33230300 | Discovery of rare variants associated with blood pressure regulation through meta-analysis of 1.3 million individuals. | Diastolic blood pressure | 810,865 E | 354,096 E | 4p15.31 | | LCORL | | | ENSG00000178177 | NA | NA | rs2320299-A | rs2320299 | 0 | 2320299 | intron_variant | 0 | 0.7285 | 6.00E-11 | 10.22185 | | 0.104971 | [0.074-0.1 | Affymetrix, Illumina [242866] (imputed) | N |
| 3 | 4:17942560 | 4 | 17972372 | rs2320299 | 21/09/2021 | 3.3E+07 | Surendran P | 23/11/2020 | Nat Genet | www.ncbi.nlm.nih.gov/pubmed/33230300 | Discovery of rare variants associated with blood pressure regulation through meta-analysis of 1.3 million individuals. | Hypertension | 810,865 E | 354,096 E | 4p15.31 | | LCORL | | | ENSG00000178177 | NA | NA | rs2320299-A | rs2320299 | 0 | 2320299 | intron_variant | 0 | 0.7265 | 1.00E-13 | 13 | | 7.44408 | z score inc | Affymetrix, Illumina [239843] (imputed) | N |
| 3 | 4:17942560 | 4 | 17972372 | rs2320299 | 21/09/2021 | 3.3E+07 | Surendran P | 23/11/2020 | Nat Genet | www.ncbi.nlm.nih.gov/pubmed/33230300 | Discovery of rare variants associated with blood pressure regulation through meta-analysis of 1.3 million individuals. | Pulse pressure | 810,865 E | 354,096 E | 4p15.31 | | LCORL | | | ENSG00000178177 | NA | NA | rs2320299-A | rs2320299 | 0 | 2320299 | intron_variant | 0 | 0.7285 | 8.00E-14 | 13.09691 | | 0.134267 | [0.099-0.1 | Affymetrix, Illumina [242866] (imputed) | N |
| 3 | 4:17942560 | 4 | 17972372 | rs2320299 | 21/09/2021 | 3.3E+07 | Surendran P | 23/11/2020 | Nat Genet | www.ncbi.nlm.nih.gov/pubmed/33230300 | Discovery of rare variants associated with blood pressure regulation through meta-analysis of 1.3 million individuals. | Systolic blood pressure | 810,865 E | 354,096 E | 4p15.31 | | LCORL | | | ENSG00000178177 | NA | NA | rs2320299-A | rs2320299 | 0 | 2320299 | intron_variant | 0 | 0.7286 | 9.00E-21 | 20.04576 | | 0.237589 | [0.19-0.29 | Affymetrix, Illumina [242865] (imputed) | N |
| 3 | 4:17942560 | 4 | 18022834 | rs2610989 | 20/06/2022 | 3.5E+07 | Helgeland O | 21/03/2022 | Nat Metab | www.ncbi.nlm.nih.gov/pubmed/35315439 | Characterization of the genetic architecture of infant and early childhood body mass index. | BMI at 1 year old | 24,551 Eur | NA | 4p15.31 | | LCORL | | | ENSG00000178177 | NA | NA | rs2610989-T | rs2610989 | 0 | 2610989 | intron_variant | 0 | 0.261018 | 2.00E-06 | 5.69897 | | 0.054456 | [0.033-0.0 | Illumina [9200000] (imputed) | N |
| 3 | 4:17942560 | 4 | 18022834 | rs2610989 | 20/06/2022 | 3.5E+07 | Helgeland O | 21/03/2022 | Nat Metab | www.ncbi.nlm.nih.gov/pubmed/35315439 | Characterization of the genetic architecture of infant and early childhood body mass index. | BMI at 1.5 years old | 21,871 Eur | NA | 4p15.31 | | LCORL | | | ENSG00000178177 | NA | NA | rs2610989-T | rs2610989 | 0 | 2610989 | intron_variant | 0 | 0.261342 | 6.00E-08 | 7.221849 | | 0.059748 | [0.039-0.0 | Illumina [9200000] (imputed) | N |
| 3 | 4:17942560 | 4 | 18022834 | rs2610989 | 20/06/2022 | 3.5E+07 | Helgeland O | 21/03/2022 | Nat Metab | www.ncbi.nlm.nih.gov/pubmed/35315439 | Characterization of the genetic architecture of infant and early childhood body mass index. | BMI at 2 years old | 21,831 Eur | NA | 4p15.31 | | LCORL | | | ENSG00000178177 | NA | NA | rs2610989-T | rs2610989 | 0 | 2610989 | intron_variant | 0 | 0.261779 | 1.00E-06 | 6 | | 0.061031 | [0.037-0.0 | Illumina [9200000] (imputed) | N |
| 3 | 4:17942560 | 4 | 18022834 | rs2610989 | 20/06/2022 | 3.5E+07 | Helgeland O | 21/03/2022 | Nat Metab | www.ncbi.nlm.nih.gov/pubmed/35315439 | Characterization of the genetic architecture of infant and early childhood body mass index. | BMI at 5 years old | 16,902 Eur | NA | 4p15.31 | | LCORL | | | ENSG00000178177 | NA | NA | rs2610989-T | rs2610989 | 0 | 2610989 | intron_variant | 0 | 0.262489 | 2.00E-06 | 5.69897 | | 0.065286 | [0.039-0.0 | Illumina [9200000] (imputed) | N |

| | | | | | | | | | | | | | | | | | | | | | | | | | | | | | | | | | | | | |
|---|---|---|---|---|---|---|---|---|---|---|---|---|---|---|---|---|---|---|---|---|---|---|---|---|---|---|---|---|---|---|---|---|---|---|---|---|
| 3 | 4:17942560 | 4 | 18022834 | rs2610989 | 20/06/2022 | 3.5E+07 | Helgeland O | 21/03/2022 | Nat Metab | www.ncbi.nlm.nih.gov/pubmed/35315439 | Characterization of the genetic architecture of infant and early childhood body mass index. | BMI at 6 months old | 11,095 Eur | NA | 4p15.31 | | LCORL | | | ENSG00000178177 | NA | NA | rs2610989-T | rs2610989 | 0 | 2610989 | intron_variant | 0 | 0.261584 | 4.00E-06 | 5.39794 | | 0.048107 | [0.028-0.0 | Illumina [9200000] (imputed) | N |
| 3 | 4:17942560 | 4 | 18022834 | rs2610989 | 20/06/2022 | 3.5E+07 | Helgeland O | 21/03/2022 | Nat Metab | www.ncbi.nlm.nih.gov/pubmed/35315439 | Characterization of the genetic architecture of infant and early childhood body mass index. | BMI at 8 months old | 24,302 Eur | NA | 4p15.31 | | LCORL | | | ENSG00000178177 | NA | NA | rs2610989-T | rs2610989 | 0 | 2610989 | intron_variant | 0 | 0.261319 | 3.00E-06 | 5.522879 | | 0.051667 | [0.031-0.0 | Illumina [9200000] (imputed) | N |
| 3 | 4:17942560 | 4 | 18008232 | rs2610990 | 1/07/2022 | 3.5E+07 | Okbay A | 31/03/2022 | Nat Genet | www.ncbi.nlm.nih.gov/pubmed/35361970 | Polygenic prediction of educational attainment within and between families from genome-wide association analyses in 3 million individuals. | Educational attainment | 3,037,499 | NA | 4p15.31 | | LCORL | | | ENSG00000178177 | NA | NA | rs2610990-A | rs2610990 | 0 | 2610990 | intron_variant | 0 | 0.2583 | 3.00E-22 | 21.52288 | | 0.011579 | [0.0092-0. | Illumina [10675380] (imputed) | N |
| 3 | 4:17942560 | 4 | 18008232 | rs2610990 | 24/08/2021 | 3.4E+07 | Christakoudi S | 21/05/2021 | Sci Rep | www.ncbi.nlm.nih.gov/pubmed/34021172 | GWAS of allometric body-shape indices in UK Biobank identifies loci suggesting associations with morphogenesis, organogenesis, adrenal cell renewal and cancer. | Hip circumference adjusted for BMI | 219,872 B | NA | 4p15.31 | LCORL | LCORL | | | ENSG00000178177 | NA | NA | rs2610990-A | rs2610990 | 0 | 2610990 | intron_variant | 0 | NR | 2.00E-19 | 18.69897 | | 0.028798 | [0.023-0.0 | Affymetrix [321016] (imputed) | N |
| 3 | 4:17942560 | 4 | 18008232 | rs2610990 | 29/11/2018 | 3E+07 | Evangelou E | 17/09/2018 | Nat Genet | www.ncbi.nlm.nih.gov/pubmed/30224653 | Genetic analysis of over 1 million people identifies 535 new loci associated with blood pressure traits. | Systolic blood pressure | 757,601 E | 249,262 E | 4p15.31 | LCORL | LCORL | | | ENSG00000178177 | NA | NA | rs2610990-A | rs2610990 | 0 | 2610990 | intron_variant | 0 | 0.2607 | 4.00E-20 | 19.39794 | | 0.2573 | [0.2-0.31] | Affymetrix, Illumina [~ 7100000] (imputed) | N |
| 3 | 4:17942560 | 4 | 17942560 | rs2707450 | 16/10/2019 | 3.2E+07 | Akiyama M | 27/09/2019 | Nat Commun | www.ncbi.nlm.nih.gov/pubmed/31562340 | Characterizing rare and low-frequency height-associated variants in the Japanese population. | Height | 159,095 Ja | 32,692 Jap | 4p15.31 | LCORL | LCORL | | | ENSG00000178177 | NA | NA | rs2707450-? | rs2707450 | 0 | 2707450 | intron_variant | 0 | NR | 5.00E-19 | 18.30103 | | 0.030342 | [0.024-0.0 | Illumina [27896057] (imputed) | N |
| 3 | 4:17942560 | 4 | 17942560 | rs2707450 | 7/03/2019 | 3.1E+07 | Giri A | 21/12/2018 | Nat Genet | www.ncbi.nlm.nih.gov/pubmed/30578418 | Trans-ethnic association study of blood pressure determinants in over 750,000 individuals. | Systolic blood pressure | 365,998 E | 299,024 E | 4p15.31 | LCORL | LCORL | | | ENSG00000178177 | NA | NA | rs2707450-T | rs2707450 | 0 | 2707450 | intron_variant | 0 | 0.7357 | 2.00E-12 | 11.69897 | | 0.2125 | [0.15-0.27 | Affymetrix [NR] (imputed) | N |
| 3 | 4:17942560 | 4 | 17946432 | rs2724475 | 24/10/2012 | 2.1E+07 | Lango Allen H | 29/09/2010 | Nature | www.ncbi.nlm.nih.gov/pubmed/20881960 | Hundreds of variants clustered in genomic loci and biological pathways affect human height. | Height | 133,653 E | 50,074 Eur | 4p15.31 | LCORL | LCORL | | | ENSG00000178177 | NA | NA | rs2724475-? | rs2724475 | 0 | 2724475 | intron_variant | 0 | NR | 2.00E-09 | 8.69897 | (Condition | NA | [NR] | Affymetrix, Illumina [2834208] (imputed) | N |
| 3 | 4:17942560 | 4 | 17946432 | rs2724475 | 2/07/2019 | 3.1E+07 | Wojcik GL | 19/06/2019 | Nature | www.ncbi.nlm.nih.gov/pubmed/31217584 | Genetic analyses of diverse populations improves discovery for complex traits. | Height | 17,286 Afri | NA | 4p15.31 | NR | LCORL | | | ENSG00000178177 | NA | NA | rs2724475-? | rs2724475 | 0 | 2724475 | intron_variant | 0 | NR | 2.00E-26 | 25.69897 | | 0.062986 | [0.051-0.0 | Illumina [34656550] (imputed) | N |
| 3 | 4:17942560 | 4 | 17946432 | rs2724475 | 26/04/2022 | 3.5E+07 | Fernandez-Rhodes L | 11/03/2022 | HGG Adv | www.ncbi.nlm.nih.gov/pubmed/35399580 | Ancestral diversity improves discovery and fine-mapping of genetic loci for anthropometric traits-The Hispanic/Latino Anthropometry Consortium. | Height | 59,771 His | 8,110 Hisp | 4p15.31 | | LCORL | | | ENSG00000178177 | NA | NA | rs2724475-T | rs2724475 | 0 | 2724475 | intron_variant | 0 | 0.3383 | 6.00E-11 | 10.22185 | | 0.0498 | [0.038-0.0 | Affymetrix, Illumina [8927938] (imputed) | N |
| 3 | 4:17942560 | 4 | 17946432 | rs2724475 | 14/12/2020 | 3.2E+07 | Ruth KS | 10/02/2020 | Nat Med | www.ncbi.nlm.nih.gov/pubmed/32042192 | Using human genetics to understand the disease impacts of testosterone in men and women. | Sex hormone-binding globulin levels | 370,125 E | NA | 4p15.31 | NR | LCORL | | | ENSG00000178177 | NA | NA | rs2724475-T | rs2724475 | 0 | 2724475 | intron_variant | 0 | 0.261619 | 5.00E-13 | 12.30103 | | 0.007659 | [0.0056-0. | Affymetrix [16582965] (imputed) | N |
| 3 | 4:17942560 | 4 | 17946432 | rs2724475 | 14/12/2020 | 3.2E+07 | Ruth KS | 10/02/2020 | Nat Med | www.ncbi.nlm.nih.gov/pubmed/32042192 | Using human genetics to understand the disease impacts of testosterone in men and women. | Sex hormone-binding globulin levels adjusted for BMI | 368,929 E | NA | 4p15.31 | NR | LCORL | | | ENSG00000178177 | NA | NA | rs2724475-T | rs2724475 | 0 | 2724475 | intron_variant | 0 | 0.261619 | 5.00E-14 | 13.30103 | | 0.007273 | [0.0054-0. | Affymetrix [16582909] (imputed) | N |
| 3 | 4:17942560 | 4 | 17969698 | rs2724485 | 24/08/2022 | 3.6E+07 | Plotnikov D | 1/06/2022 | Invest Ophthalmol Vis Sci | www.ncbi.nlm.nih.gov/pubmed/35762941 | High Blood Pressure and Intraocular Pressure: A Mendelian Randomization Study. | Systolic blood pressure | 526,001 E | NA | 4p15.31 | | LCORL | | | ENSG00000178177 | NA | NA | rs2724485-T | rs2724485 | 0 | 2724485 | intron_variant | 0 | NR | 2.00E-10 | 9.69897 | | 0.2561 | [0.18-0.34 | NR [NR] (imputed) | N |
| 3 | 4:17942560 | 4 | 17903654 | rs4144829 | 5/08/2019 | 3.1E+07 | Warrington NM | 1/05/2019 | Nat Genet | www.ncbi.nlm.nih.gov/pubmed/31043758 | Maternal and fetal genetic effects on birth weight and their relevance to cardio-metabolic risk factors. | Birth weight | 298,142 E | NR | 4p15.31 | LCORL, DCAF16 | LCORL | | | ENSG00000178177 | NA | NA | rs4144829-C | rs4144829 | 0 | 4144829 | intron_variant | 0 | 0.267008 | 4.00E-34 | 33.39794 | | 0.035507 | [0.03-0.04 | Affymetrix, Illumina [15144367] (imputed) | N |
| 3 | 4:17942560 | 4 | 17903654 | rs4144829 | 13/08/2021 | 3.4E+07 | Liu Y | 15/06/2021 | Elife | www.ncbi.nlm.nih.gov/pubmed/34128465 | Genetic architecture of 11 organ traits derived from abdominal MRI using deep learning. | Lung volume | 32,860 Eur | NA | 4p15.31 | NCAPG | LCORL | | | ENSG00000178177 | NA | NA | rs4144829-? | rs4144829 | 0 | 4144829 | intron_variant | 0 | NR | 1.00E-09 | 9 | | 0.0466 | NR L decre | Affymetrix [9390170] (imputed) | N |
| 3 | 4:17942560 | 4 | 17903654 | rs4144829 | 24/08/2021 | 3.4E+07 | Christakoudi S | 21/05/2021 | Sci Rep | www.ncbi.nlm.nih.gov/pubmed/34021172 | GWAS of allometric body-shape indices in UK Biobank identifies loci suggesting associations with morphogenesis, organogenesis, adrenal cell renewal and cancer. | Waist circumference adjusted for body mass index | 186,825 B | NA | 4p15.31 | LCORL | LCORL | | | ENSG00000178177 | NA | NA | rs4144829-C | rs4144829 | 0 | 4144829 | intron_variant | 0 | NR | 3.00E-09 | 8.522879 | | 0.020853 | [0.014-0.0 | Affymetrix [321114] (imputed) | N |
| 3 | 4:17942560 | 4 | 17854055 | rs6842303 | 16/12/2019 | 3.2E+07 | Helgeland O | 1/10/2019 | Nat Commun | www.ncbi.nlm.nih.gov/pubmed/31575865 | Genome-wide association study reveals dynamic role of genetic variation in infant and early childhood growth. | BMI at 1 year old | 6,970 Nor | 3,839 Nor | 4p15.31 | LCORL | LCORL | | | ENSG00000178177 | NA | NA | rs6842303-T | rs6842303 | 0 | 6842303 | intron_variant | 0 | 0.27934 | 2.00E-08 | 7.69897 | | 0.086 | [0.057-0.1 | Illumina [~ 8500000] (imputed) | N |
| 3 | 4:17942560 | 4 | 17854055 | rs6842303 | 16/12/2019 | 3.2E+07 | Helgeland O | 1/10/2019 | Nat Commun | www.ncbi.nlm.nih.gov/pubmed/31575865 | Genome-wide association study reveals dynamic role of genetic variation in infant and early childhood growth. | BMI at 1.5 years old | 6,800 Nor | 3,780 Nor | 4p15.31 | LCORL | LCORL | | | ENSG00000178177 | NA | NA | rs6842303-T | rs6842303 | 0 | 6842303 | intron_variant | 0 | 0.27934 | 8.00E-09 | 8.09691 | | 0.089 | [0.06-0.11 | Illumina [~ 8500000] (imputed) | N |
| 3 | 4:17942560 | 4 | 17993410 | rs724577 | 27/01/2020 | 3.2E+07 | Yang XL | 11/10/2019 | Front Genet | www.ncbi.nlm.nih.gov/pubmed/31681408 | Three Novel Loci for Infant Head Circumference Identified by a Joint Association Analysis. | Birth length (MTAG) | up to 182, | NA | 4p15.31 | NR | LCORL | | | ENSG00000178177 | NA | NA | rs724577-A | rs724577 | 0 | 724577 | intron_variant | 0 | 0.358 | 4.00E-16 | 15.39794 | | 0.049361 | [0.037-0.0 | NR [1977400] (imputed) | N |
| 3 | 4:17942560 | 4 | 17993410 | rs724577 | 28/02/2013 | 2.3E+07 | Horikoshi M | 2/12/2012 | Nat Genet | www.ncbi.nlm.nih.gov/pubmed/23202124 | New loci associated with birth weight identify genetic links between intrauterine growth and adult height and metabolism. | Birth weight | Up to 26,8 | Up to 42,5 | 4p15.31 | LCORL | LCORL | | | ENSG00000178177 | NA | NA | rs724577-C | rs724577 | 0 | 724577 | intron_variant | 0 | 0.73 | 5.00E-11 | 10.30103 | | 0.042 | [0.030-0.0 | Affymetrix, Illumina [~ 2700000] (imputed) | N |
| 3 | 4:17942560 | 4 | 17993410 | rs724577 | 27/01/2020 | 3.2E+07 | Yang XL | 11/10/2019 | Front Genet | www.ncbi.nlm.nih.gov/pubmed/31681408 | Three Novel Loci for Infant Head Circumference Identified by a Joint Association Analysis. | Birth weight (MTAG) | up to 182, | NA | 4p15.31 | NR | LCORL | | | ENSG00000178177 | NA | NA | rs724577-A | rs724577 | 0 | 724577 | intron_variant | 0 | 0.27 | 7.00E-17 | 16.1549 | | 0.035029 | [0.027-0.0 | NR [1977400] (imputed) | N |
| 3 | 4:17942560 | 4 | 17993410 | rs724577 | 8/11/2011 | 2.2E+07 | N'Diaye A | 6/10/2011 | PLoS Genet | www.ncbi.nlm.nih.gov/pubmed/21998595 | Identification, replication, and fine-mapping of Loci associated with adult height in individuals of african ancestry. | Height | 20,427 Afri | Up to 16,4 | 4p15.31 | LCORL | LCORL | | | ENSG00000178177 | NA | NA | rs724577-A | rs724577 | 0 | 724577 | intron_variant | 0 | 0.31 | 1.00E-15 | 15 | | 0.067 | [0.051-0.0 | Affymetrix, Illumina [3310998] (imputed) | N |
| 3 | 4:17942560 | 4 | 17993410 | rs724577 | 2/07/2019 | 3.1E+07 | Wojcik GL | 19/06/2019 | Nature | www.ncbi.nlm.nih.gov/pubmed/31217584 | Genetic analyses of diverse populations improves discovery for complex traits. | Height | 17,286 Afri | NA | 4p15.31 | NR | LCORL | | | ENSG00000178177 | NA | NA | rs724577-? | rs724577 | 0 | 724577 | intron_variant | 0 | NR | 3.00E-26 | 25.52288 | | 0.062901 | [0.051-0.0 | Illumina [34656550] (imputed) | N |
| 3 | 4:17942560 | 4 | 17871012 | rs73242122 | 1/07/2022 | 3.5E+07 | Okbay A | 31/03/2022 | Nat Genet | www.ncbi.nlm.nih.gov/pubmed/35361970 | Polygenic prediction of educational attainment within and between families from genome-wide association analyses in 3 million individuals. | Educational attainment | 3,037,499 | NA | 4p15.31 | | LCORL | | | ENSG00000178177 | NA | NA | rs73242122-C | rs73242122 | 0 | 73242122 | intron_variant | 0 | 0.7774 | 7.00E-08 | 7.154902 | | 0.006775 | [0.0043-0. | Illumina [10675380] (imputed) | N |
| 3 | 4:17942560 | 4 | 17798334 | rs7679066 | 24/08/2021 | 3.4E+07 | Christakoudi S | 21/05/2021 | Sci Rep | www.ncbi.nlm.nih.gov/pubmed/34021172 | GWAS of allometric body-shape indices in UK Biobank identifies loci suggesting associations with morphogenesis, organogenesis, adrenal cell renewal and cancer. | Hip circumference adjusted for BMI | 186,825 B | NA | 4p15.31 | DCAF16 | FAM184B - DCAF | ENSG00000047662 | ENSG00000163257 | | 15090 | 3944 | rs7679066-G | rs7679066 | 0 | 7679066 | intergenic_variant | 1 | NR | 2.00E-14 | 13.69897 | | 0.025864 | [0.019-0.0 | Affymetrix [323123] (imputed) | N |
| 3 | 4:17942560 | 4 | 17919811 | rs925098 | 15/01/2018 | 2.8E+07 | Horikoshi M | 13/10/2016 | Nature | www.ncbi.nlm.nih.gov/pubmed/27680694 | Genome-wide associations for birth weight and correlations with adult disease. | Birth weight | 133,903 E | 9,799 Euro | 4p15.31 | LCORL | LCORL | | | ENSG00000178177 | NA | NA | rs925098-G | rs925098 | 0 | 925098 | intron_variant | 0 | 0.28 | 1.00E-15 | 15 | | 0.0322 | [0.024-0.0 | Affymetrix, Illumina [at least 22434434] (imputed) | N |
| 3 | 4:17942560 | 4 | 17919811 | rs925098 | 25/11/2011 | 2.2E+07 | Carty CL | 21/10/2011 | Hum Mol Genet | www.ncbi.nlm.nih.gov/pubmed/22021425 | Genome-wide association study of body height in African Americans: the Women's Health Initiative SNP Health Association Resource (SHARe). | Height | 8,149 Afric | Up to 20,8 | 4p15.31 | LCORL | LCORL | | | ENSG00000178177 | NA | NA | rs925098-G | rs925098 | 0 | 925098 | intron_variant | 0 | 0.35 | 3.00E-14 | 13.52288 | | 0.056 | [0.036-0.0 | Affymetrix [855034] | N |
| 3 | 4:17942560 | 4 | 17919811 | rs925098 | 2/07/2019 | 3.1E+07 | Wojcik GL | 19/06/2019 | Nature | www.ncbi.nlm.nih.gov/pubmed/31217584 | Genetic analyses of diverse populations improves discovery for complex traits. | Height | 17,286 Afri | NA | 4p15.31 | NR | LCORL | | | ENSG00000178177 | NA | NA | rs925098-? | rs925098 | 0 | 925098 | intron_variant | 0 | NR | 2.00E-23 | 22.69897 | | 0.058421 | [0.047-0.0 | Illumina [34656550] (imputed) | N |
| 3 | 4:17942560 | 4 | 17919811 | rs925098 | 1/07/2021 | 3.4E+07 | Graff M | 5/03/2021 | Am J Hum Genet | www.ncbi.nlm.nih.gov/pubmed/33713608 | Discovery and fine-mapping of height loci via high-density imputation of GWASs in individuals of African ancestry. | Height | 41,389 Afri | 11,364 Afri | 4p15.31 | LCORL | LCORL | | | ENSG00000178177 | NA | NA | rs925098-G | rs925098 | 0 | 925098 | intron_variant | 0 | | 2.00E-59 | 58.69897 | | NA | | Affymetrix, Illumina [17972582] (imputed) | N |
| 3 | 4:17942560 | 4 | 17919811 | rs925098 | 1/07/2021 | 3.4E+07 | Graff M | 5/03/2021 | Am J Hum Genet | www.ncbi.nlm.nih.gov/pubmed/33713608 | Discovery and fine-mapping of height loci via high-density imputation of GWASs in individuals of African ancestry. | Height | 25,369 Afri | 8,449 Afric | 4p15.31 | LCORL | LCORL | | | ENSG00000178177 | NA | NA | rs925098-G | rs925098 | 0 | 925098 | intron_variant | 0 | 0.351 | 6.00E-08 | 7.221849 | (AA) | 0.046 | [0.028-0.0 | Affymetrix, Illumina [19885954] (imputed) | N |
| 3 | 4:17942560 | 4 | 17919811 | rs925098 | 1/07/2021 | 3.4E+07 | Graff M | 5/03/2021 | Am J Hum Genet | www.ncbi.nlm.nih.gov/pubmed/33713608 | Discovery and fine-mapping of height loci via high-density imputation of GWASs in individuals of African ancestry. | Height | 16,021 Afri | 2,915 Afric | 4p15.31 | LCORL | LCORL | | | ENSG00000178177 | NA | NA | rs925098-G | rs925098 | 0 | 925098 | intron_variant | 0 | 0.347 | 1.00E-07 | 7 | (AA) | 0.059 | [0.037-0.0 | Affymetrix, Illumina [16055004] (imputed) | N |
| 3 | 4:17942560 | 4 | 17919811 | rs925098 | 1/07/2021 | 3.4E+07 | Graff M | 5/03/2021 | Am J Hum Genet | www.ncbi.nlm.nih.gov/pubmed/33713608 | Discovery and fine-mapping of height loci via high-density imputation of GWASs in individuals of African ancestry. | Height | 41,389 Afri | 11,364 Afri | 4p15.31 | LCORL | LCORL | | | ENSG00000178177 | NA | NA | rs925098-G | rs925098 | 0 | 925098 | intron_variant | 0 | 0.35 | 3.00E-13 | 12.52288 | (AA) | 0.05 | [0.036-0.0 | Affymetrix, Illumina [17972582] (imputed) | N |
| 3 | 4:17942560 | 4 | 17919811 | rs925098 | 14/12/2020 | 3.2E+07 | Ruth KS | 10/02/2020 | Nat Med | www.ncbi.nlm.nih.gov/pubmed/32042192 | Using human genetics to understand the disease impacts of testosterone in men and women. | Sex hormone-binding globulin levels | 189,473 E | NA | 4p15.31 | NR | LCORL | | | ENSG00000178177 | NA | NA | rs925098-G | rs925098 | 0 | 925098 | intron_variant | 0 | 0.264724 | 8.00E-09 | 8.09691 | | 0.00981 | [0.0067-0. | Affymetrix [16585866] (imputed) | N |
| 3 | 4:17942560 | 4 | 17919811 | rs925098 | 13/06/2022 | 3.5E+07 | Haas CB | 22/02/2022 | Endocrinology | www.ncbi.nlm.nih.gov/pubmed/35192695 | Cross-ancestry genome-wide association studies of sex hormone concentrations in pre- and postmenopausal women. | Sex hormone-binding globulin levels | 196,901 E | NA | 4p15.31 | | LCORL | | | ENSG00000178177 | NA | NA | rs925098-G | rs925098 | 0 | 925098 | intron_variant | 0 | 0.264593 | 1.00E-06 | 6 | | 0.017685 | [0.011-0.0 | NR [NR] (imputed) | N |
| 3 | 4:17942560 | 4 | 17919811 | rs925098 | 13/06/2022 | 3.5E+07 | Haas CB | 22/02/2022 | Endocrinology | www.ncbi.nlm.nih.gov/pubmed/35192695 | Cross-ancestry genome-wide association studies of sex hormone concentrations in pre- and postmenopausal women. | Sex hormone-binding globulin levels | 4,522 Afric | NA | 4p15.31 | | LCORL | | | ENSG00000178177 | NA | NA | rs925098-? | rs925098 | 0 | 925098 | intron_variant | 0 | NR | 2.00E-07 | 6.69897 | | 5.22 | z score dec | NR [NR] (imputed) | N |
| 3 | 4:17942560 | 4 | 17919811 | rs925098 | 14/12/2020 | 3.2E+07 | Ruth KS | 10/02/2020 | Nat Med | www.ncbi.nlm.nih.gov/pubmed/32042192 | Using human genetics to understand the disease impacts of testosterone in men and women. | Sex hormone-binding globulin levels adjusted for BMI | 188,908 E | NA | 4p15.31 | NR | LCORL | | | ENSG00000178177 | NA | NA | rs925098-G | rs925098 | 0 | 925098 | intron_variant | 0 | 0.264724 | 3.00E-09 | 8.522879 | | 0.00885 | [0.0061-0. | Affymetrix [16585854] (imputed) | N |
| 3 | 4:17942560 | 4 | 17901679 | rs979532 | 24/08/2021 | 3.4E+07 | Christakoudi S | 21/05/2021 | Sci Rep | www.ncbi.nlm.nih.gov/pubmed/34021172 | GWAS of allometric body-shape indices in UK Biobank identifies loci suggesting associations with morphogenesis, organogenesis, adrenal cell renewal and cancer. | Hip circumference adjusted for BMI | 186,825 B | NA | 4p15.31 | LCORL | LCORL | | | ENSG00000178177 | NA | NA | rs979532-C | rs979532 | 0 | 979532 | intron_variant | 0 | NR | 2.00E-21 | 20.69897 | | 0.033283 | [0.026-0.0 | Affymetrix [323123] (imputed) | N |
| 3 | 4:17942560 | 4 | 17901679 | rs979532 | 8/02/2019 | 3.1E+07 | Kichaev G | 27/12/2018 | Am J Hum Genet | www.ncbi.nlm.nih.gov/pubmed/30595370 | Leveraging Polygenic Functional Enrichment to Improve GWAS Power. | Systolic blood pressure | approxima | NA | 4p15.31 | | LCORL | | | ENSG00000178177 | NA | NA | rs979532-? | rs979532 | 0 | 979532 | intron_variant | 0 | NR | 4.00E-12 | 11.39794 | | NA | | NR [~ 8900000] (imputed) | N |
| 5 | 4:106215109 | 4 | 1.06E+08 | rs200768290 | 29/06/2021 | 3.3E+07 | de la Fuente J | 7/09/2020 | Nat Hum Behav | www.ncbi.nlm.nih.gov/pubmed/32895543 | A general dimension of genetic sharing across diverse cognitive traits inferred from molecular data. | Verbal-numerical reasoning | 171,304 B | NA | 4q24 | | RN7SL89P - PPA2 | ENSG00000243383 | ENSG00000138777 | | 6 | 75125 | rs200768290-? | rs200768290 | 0 | 2.01E+08 | regulatory_region_variant | 1 | NR | 8.00E-16 | 15.09691 | | NA | | NR [7857346] (imputed) | N |
| 5 | 4:106216800 | 4 | 1.06E+08 | rs34104813 | 14/12/2021 | 3.5E+07 | Sakaue S | 30/09/2021 | Nat Genet | www.ncbi.nlm.nih.gov/pubmed/34594039 | A cross-population atlas of genetic associations for 220 human phenotypes. | Neutrophil count | 349,856 E | NA | 4q24 | | TET2 | | | ENSG00000168769 | NA | NA | rs34104813-T | rs34104813 | 0 | 34104813 | intron_variant | 0 | NR | 2.00E-21 | 20.69897 | | 0.0214 | [0.017-0.0 | Affymetrix, Illumina [20526770] (imputed) | N |
| 5 | 4:106216800 | 4 | 1.06E+08 | rs34104813 | 24/08/2022 | 3.6E+07 | Plotnikov D | 1/06/2022 | Invest Ophthalmol Vis Sci | www.ncbi.nlm.nih.gov/pubmed/35762941 | High Blood Pressure and Intraocular Pressure: A Mendelian Randomization Study. | Pulse pressure | 526,001 E | NA | 4q24 | | TET2 | | | ENSG00000168769 | NA | NA | rs34104813-T | rs34104813 | 0 | 34104813 | intron_variant | 0 | NR | 5.00E-09 | 8.30103 | | 0.1511 | [0.1-0.2] u | NR [NR] (imputed) | N |
| 5 | 4:106215109 | 4 | 1.06E+08 | rs57692580 | 2/09/2019 | 3.1E+07 | Adams MJ | 1/07/2019 | Int J Epidemiol | www.ncbi.nlm.nih.gov/pubmed/31263887 | Factors associated with sharing e-mail information and mental health survey participation in large population cohorts. | Mental health study participation (completed survey) | 122,647 B | NA | 4q24 | RN7SL89P | TET2 - RN7SL89P | ENSG00000168769 | ENSG00000243383 | | 13503 | 345 | rs57692580-A | rs57692580 | 0 | 57692580 | regulatory_region_variant | 1 | 0.39 | 3.00E-08 | 7.522879 | | 0.006151 | [0.004-0.0 | NR [16367095] (imputed) | N |
| 5 | 4:106216800 | 4 | 1.06E+08 | rs62332762 | 18/02/2021 | 3.3E+07 | Said MA | 8/12/2020 | J Am Heart Assoc | www.ncbi.nlm.nih.gov/pubmed/33287642 | Associations of Observational and Genetically Determined Caffeine Intake With Coronary Artery Disease and Diabetes Mellitus. | Caffeine consumption from coffee or tea | 362,316 E | NA | 4q24 | TET2 | TET2 | | | ENSG00000168769 | NA | NA | rs62332762-C | rs62332762 | 0 | 62332762 | intron_variant | 0 | 0.597585 | 1.00E-14 | 14 | | 0.01828 | [0.014-0.0 | Affymetrix [19400838] (imputed) | N |
| 6 | 4:145586459 | 4 | 1.46E+08 | rs11100862 | 30/07/2021 | 3.3E+07 | Lundtoft C | 26/10/2020 | PLoS Genet | www.ncbi.nlm.nih.gov/pubmed/33104735 | Function of multiple sclerosis-protective HLA class I alleles revealed by genome-wide protein-quantitative trait loci mapping of interferon signalling. | interferon-related traits | 303 Europ | NA | 4q31.21 | NR | KRT18P51 - HHIP | ENSG00000250504 | ENSG00000248890 | | 63516 | 6423 | rs11100862-A | rs11100862 | 0 | 11100862 | intron_variant | 1 | 0.452 | 4.00E-06 | 5.39794 | (IFNGR1_c | NA | | Illumina [5084123] (imputed) | N |
| 6 | 4:145905213 | 4 | 1.46E+08 | rs11733300 | 24/08/2021 | 3.4E+07 | Christakoudi S | 21/05/2021 | Sci Rep | www.ncbi.nlm.nih.gov/pubmed/34021172 | GWAS of allometric body-shape indices in UK Biobank identifies loci suggesting associations with morphogenesis, organogenesis, adrenal cell renewal and cancer. | Hip circumference adjusted for BMI | 186,825 B | NA | 4q31.21 | OTUD4 | OTUD4 - RPS23P | ENSG00000164164 | ENSG00000240992 | | 32085 | 57136 | rs11733300-A | rs11733300 | 0 | 11733300 | intergenic_variant | 1 | NR | 2.00E-17 | 16.69897 | | 0.026175 | [0.02-0.03 | Affymetrix [323123] (imputed) | N |
| 6 | 4:145597759 | 4 | 1.46E+08 | rs11946517 | 28/01/2019 | 3.1E+07 | Morris JA | 31/12/2018 | Nat Genet | www.ncbi.nlm.nih.gov/pubmed/30598549 | An atlas of genetic influences on osteoporosis in humans and mice. | Heel bone mineral density | 426,824 B | NA | 4q31.21 | HHIP | HHIP | | | ENSG00000164161 | NA | NA | rs11946517-G | rs11946517 | 0 | 11946517 | intron_variant | 0 | 0.416446 | 6.00E-14 | 13.22185 | | 0.014091 | [0.01-0.01 | NR [13737936] (imputed) | N |
| 6 | 4:145597759 | 4 | 1.46E+08 | rs11946517 | 8/02/2019 | 3.1E+07 | Kichaev G | 27/12/2018 | Am J Hum Genet | www.ncbi.nlm.nih.gov/pubmed/30595370 | Leveraging Polygenic Functional Enrichment to Improve GWAS Power. | Heel bone mineral density | approxima | NA | 4q31.21 | | HHIP | | | ENSG00000164161 | NA | NA | rs11946517-? | rs11946517 | 0 | 11946517 | intron_variant | 0 | NR | 2.00E-22 | 21.69897 | | NA | | NR [~ 8900000] (imputed) | N |

| | | | | | | | | | | | | | | | | | | | | | | | | | | | | | | | | | | | | |
|---|---|---|---|---|---|---|---|---|---|---|---|---|---|---|---|---|---|---|---|---|---|---|---|---|---|---|---|---|---|---|---|---|---|---|---|---|
| 6 | 4:145597759 | 4 | 1.46E+08 | rs11946517 | 24/08/2021 | 3.4E+07 | Christakoudi S | 21/05/2021 | Sci Rep | www.ncbi.nlm.nih.gov/pubmed/34021172 | GWAS of allometric body-shape indices in UK Biobank identifies loci suggesting associations with morphogenesis, organogenesis, adrenal cell renewal and cancer. | Hip circumference adjusted for BMI | 186,825 B | NA | 4q31.21 | HHIP | HHIP | | | ENSG00000164161 | NA | NA | rs11946517-G | rs11946517 | 0 | 11946517 | intron_variant | 0 | NR | 1.00E-36 | 36 | | 0.039349 | [0.033-0.0 | Affymetrix [323123] (imputed) | N |
| 6 | 4:145586459 | 4 | 1.46E+08 | rs12507427 | 1/08/2019 | 3.1E+07 | Styrkarsdottir U | 3/05/2019 | Nat Commun | www.ncbi.nlm.nih.gov/pubmed/31053729 | GWAS of bone size yields twelve loci that also affect height, BMD, osteoarthritis or fractures. | Femoral neck size | 28,954 Eu | 9,383 Euro | 4q31.21 | HHIP | HHIP, HHIP-AS1 | | | ENSG00000164161, E | NA | NA | rs12507427-A | rs12507427 | 0 | 12507427 | 5_prime_UTR_variant | 0 | 0.431 | 8.00E-14 | 13.09691 | | 0.054 | [0.007-0.0 | Illumina [33400000] (imputed) | N |
| 6 | 4:145586459 | 4 | 1.46E+08 | rs12507427 | 1/08/2019 | 3.1E+07 | Styrkarsdottir U | 3/05/2019 | Nat Commun | www.ncbi.nlm.nih.gov/pubmed/31053729 | GWAS of bone size yields twelve loci that also affect height, BMD, osteoarthritis or fractures. | Hip bone size | 28,900 Eu | 7,912 Euro | 4q31.21 | HHIP | HHIP, HHIP-AS1 | | | ENSG00000164161, E | NA | NA | rs12507427-A | rs12507427 | 0 | 12507427 | 5_prime_UTR_variant | 0 | 0.43146 | 5.00E-09 | 8.30103 | | 0.043 | NR unit inc | Illumina [33400000] (imputed) | N |
| 6 | 4:145586459 | 4 | 1.46E+08 | rs13125694 | 16/10/2019 | 3.2E+07 | Akiyama M | 27/09/2019 | Nat Commun | www.ncbi.nlm.nih.gov/pubmed/31562340 | Characterizing rare and low-frequency height-associated variants in the Japanese population. | Height | 159,095 Ja | 32,692 Jap | 4q31.21 | HHIP-AS1 | HHIP-AS1 | | | ENSG00000248890 | NA | NA | rs13125694-? | rs13125694 | 0 | 13125694 | intron_variant | 0 | NR | 1.00E-29 | 29 | | 0.044578 | [0.037-0.0 | Illumina [27896057] (imputed) | N |
| 6 | 4:145586459 | 4 | 1.46E+08 | rs13125694 | 5/08/2022 | 3.6E+07 | Chiou JS | 13/07/2022 | BMC Med | www.ncbi.nlm.nih.gov/pubmed/35831902 | Your height affects your health: genetic determinants and health-related outcomes in Taiwan. | Height | 67,452 Ha | NA | 4q31.21 | | HHIP-AS1 | | | ENSG00000248890 | NA | NA | rs13125694-T | rs13125694 | 0 | 13125694 | intron_variant | 0 | NR | 5.00E-21 | 20.30103 | | 0.057 | [0.045-0.0 | Affymetrix [NR] (imputed) | N |
| 6 | 4:145586459 | 4 | 1.46E+08 | rs13125694 | 14/12/2021 | 3.5E+07 | Sakaue S | 30/09/2021 | Nat Genet | www.ncbi.nlm.nih.gov/pubmed/34594039 | A cross-population atlas of genetic associations for 220 human phenotypes. | Height | 165,056 E | NA | 4q31.21 | | HHIP-AS1 | | | ENSG00000248890 | NA | NA | rs13125694-T | rs13125694 | 0 | 13125694 | intron_variant | 0 | 0.191937 | 6.00E-27 | 26.22185 | | 0.029849 | [0.024-0.0 | Illumina [13479295] (imputed) | N |
| 6 | 4:145586459 | 4 | 1.46E+08 | rs13125694 | 24/08/2021 | 3.4E+07 | Christakoudi S | 21/05/2021 | Sci Rep | www.ncbi.nlm.nih.gov/pubmed/34021172 | GWAS of allometric body-shape indices in UK Biobank identifies loci suggesting associations with morphogenesis, organogenesis, adrenal cell renewal and cancer. | Hip circumference adjusted for BMI | 186,825 B | NA | 4q31.21 | HHIP-AS1 | HHIP-AS1 | | | ENSG00000248890 | NA | NA | rs13125694-T | rs13125694 | 0 | 13125694 | intron_variant | 0 | NR | 6.00E-54 | 53.22185 | | 0.047896 | [0.042-0.0 | Affymetrix [323123] (imputed) | N |
| 6 | 4:145586459 | 4 | 1.46E+08 | rs13125694 | 24/08/2021 | 3.4E+07 | Christakoudi S | 21/05/2021 | Sci Rep | www.ncbi.nlm.nih.gov/pubmed/34021172 | GWAS of allometric body-shape indices in UK Biobank identifies loci suggesting associations with morphogenesis, organogenesis, adrenal cell renewal and cancer. | Hip index | 186,825 B | NA | 4q31.21 | HHIP-AS1 | HHIP-AS1 | | | ENSG00000248890 | NA | NA | rs13125694-T | rs13125694 | 0 | 13125694 | intron_variant | 0 | NR | 5.00E-12 | 11.30103 | | 0.022372 | [0.016-0.0 | Affymetrix [323858] (imputed) | N |
| 6 | 4:145586459 | 4 | 1.46E+08 | rs13146972 | 26/04/2022 | 3.5E+07 | Fernandez-Rhodes L | 11/03/2022 | HGG Adv | www.ncbi.nlm.nih.gov/pubmed/35399580 | Ancestral diversity improves discovery and fine-mapping of genetic loci for anthropometric traits-The Hispanic/Latino Anthropometry Consortium. | Height | 59,771 His | 8,110 Hisp | 4q31.21 | | HHIP-AS1, HHIP | | | ENSG00000248890, E | NA | NA | rs13146972-T | rs13146972 | 0 | 13146972 | non_coding_transcript_exon | 0 | 0.4319 | 6.00E-14 | 13.22185 | | 0.0406 | [0.03-0.05 | Affymetrix, Illumina [8927938] (imputed) | N |
| 6 | 4:145655389 | 4 | 1.46E+08 | rs1389029 | 4/08/2022 | 3.6E+07 | Fan CC | 3/05/2022 | Nat Commun | www.ncbi.nlm.nih.gov/pubmed/35505052 | Multivariate genome-wide association study on tissue-sensitive diffusion metrics highlights pathways that shape the human brain. | Whole brain restricted isotropic diffusion (multivariate analysis) | 23,543 Eu | 11,555 Eu | 4q31.21 | | HHIP | | | ENSG00000164161 | NA | NA | rs1389029-? | rs1389029 | 0 | 1389029 | intron_variant | 0 | NR | 1.00E-22 | 22 | (EA) | NA | | NR [NR] (imputed) | N |
| 6 | 4:145655389 | 4 | 1.46E+08 | rs1492819 | 24/08/2021 | 3.4E+07 | Christakoudi S | 21/05/2021 | Sci Rep | www.ncbi.nlm.nih.gov/pubmed/34021172 | GWAS of allometric body-shape indices in UK Biobank identifies loci suggesting associations with morphogenesis, organogenesis, adrenal cell renewal and cancer. | Waist circumference adjusted for body mass index | 186,825 B | NA | 4q31.21 | HHIP | HHIP | | | ENSG00000164161 | NA | NA | rs1492819-T | rs1492819 | 0 | 1492819 | intron_variant | 0 | NR | 2.00E-08 | 7.69897 | | 0.017354 | [0.011-0.0 | Affymetrix [321114] (imputed) | N |
| 6 | 4:145655389 | 4 | 1.46E+08 | rs1492820 | 12/09/2008 | 1.8E+07 | Lettre G | 6/04/2008 | Nat Genet | www.ncbi.nlm.nih.gov/pubmed/18391950 | Identification of ten loci associated with height highlights new biological pathways in human growth. | Height | 15,821 Eu | Up to 17,8 | 4q31.21 | HHIP | HHIP | | | ENSG00000164161 | NA | NA | rs1492820-G | rs1492820 | 0 | 1492820 | intron_variant | 0 | 0.48 | 1.00E-11 | 11 | | 0.29 | [0.21-0.37 | Affymetrix, Illumina [2260683] (imputed) | N |
| 6 | 4:145655389 | 4 | 1.46E+08 | rs1492820 | 2/07/2019 | 3.1E+07 | Wojcik GL | 19/06/2019 | Nature | www.ncbi.nlm.nih.gov/pubmed/31217584 | Genetic analyses of diverse populations improves discovery for complex traits. | Height | 17,286 Afr | NA | 4q31.21 | NR | HHIP | | | ENSG00000164161 | NA | NA | rs1492820-? | rs1492820 | 0 | 1492820 | intron_variant | 0 | NR | 2.00E-06 | 5.69897 | | 0.027213 | [0.016-0.0 | Illumina [34656550] (imputed) | N |
| 6 | 4:145586459 | 4 | 1.46E+08 | rs17776795 | 15/01/2020 | 3.1E+07 | Plotnikov D | 16/05/2019 | Br J Ophthalmol | www.ncbi.nlm.nih.gov/pubmed/31097437 | Association between birth weight and refractive error in adulthood: a Mendelian randomisation study. | Birth weight | 188,039 E | NA | 4q31.21 | NR | KRT18P51 - HHIP | ENSG00000250504 | ENSG00000248890 | | 50701 | 19238 | rs17776795-A | rs17776795 | 0 | 17776795 | intron_variant | 1 | 0.42 | 6.00E-19 | 18.22185 | | 0.029 | [0.023-0.0 | Affymetrix [up to 10400000] (imputed) | N |
| 6 | 4:145905213 | 4 | 1.46E+08 | rs200457388 | 7/02/2020 | 3.2E+07 | Hernandez Cordero AI | 21/11/2019 | Am J Hum Genet | www.ncbi.nlm.nih.gov/pubmed/31761296 | Genome-wide Associations Reveal Human-Mouse Genetic Convergence and Modifiers of Myogenesis, CPNE1 and STC2. | Appendicular lean mass | 181,862 E | NA | 4q31.21 | NR | ANAPC10 | | | ENSG00000164162 | NA | NA | rs200457388-C | rs200457388 | 0 | 2E+08 | intron_variant | 0 | 0.423616 | 1.00E-12 | 12 | | 0.04922 | [0.036-0.0 | Affymetrix [17914406] (imputed) | N |
| 6 | 4:145905213 | 4 | 1.46E+08 | rs200457388 | 24/08/2021 | 3.4E+07 | Christakoudi S | 21/05/2021 | Sci Rep | www.ncbi.nlm.nih.gov/pubmed/34021172 | GWAS of allometric body-shape indices in UK Biobank identifies loci suggesting associations with morphogenesis, organogenesis, adrenal cell renewal and cancer. | Hip circumference adjusted for BMI | 186,825 B | NA | 4q31.21 | ANAPC10 | ANAPC10 | | | ENSG00000164162 | NA | NA | rs200457388-C | rs200457388 | 0 | 2E+08 | intron_variant | 0 | NR | 4.00E-31 | 30.39794 | | 0.035944 | [0.03-0.04 | Affymetrix [323123] (imputed) | N |
| 6 | 4:145905213 | 4 | 1.46E+08 | rs200457388 | 24/08/2021 | 3.4E+07 | Christakoudi S | 21/05/2021 | Sci Rep | www.ncbi.nlm.nih.gov/pubmed/34021172 | GWAS of allometric body-shape indices in UK Biobank identifies loci suggesting associations with morphogenesis, organogenesis, adrenal cell renewal and cancer. | Waist-hip index | 219,872 B | NA | 4q31.21 | ANAPC10 | ANAPC10 | | | ENSG00000164162 | NA | NA | rs200457388-C | rs200457388 | 0 | 2E+08 | intron_variant | 0 | NR | 4.00E-09 | 8.39794 | | 0.01677 | [0.011-0.0 | Affymetrix [319769] (imputed) | N |
| 6 | 4:145905213 | 4 | 1.46E+08 | rs200457388 | 24/08/2021 | 3.4E+07 | Christakoudi S | 21/05/2021 | Sci Rep | www.ncbi.nlm.nih.gov/pubmed/34021172 | GWAS of allometric body-shape indices in UK Biobank identifies loci suggesting associations with morphogenesis, organogenesis, adrenal cell renewal and cancer. | Waist-to-hip ratio adjusted for BMI | 219,872 B | NA | 4q31.21 | ANAPC10 | ANAPC10 | | | ENSG00000164162 | NA | NA | rs200457388-C | rs200457388 | 0 | 2E+08 | intron_variant | 0 | NR | 4.00E-10 | 9.39794 | | 0.017821 | [0.012-0.0 | Affymetrix [319769] (imputed) | N |
| 6 | 4:145586459 | 4 | 1.46E+08 | rs2131354 | 17/07/2019 | 2.9E+07 | Tachmazidou I | 1/06/2017 | Am J Hum Genet | www.ncbi.nlm.nih.gov/pubmed/28552196 | Whole-Genome Sequencing Coupled to Imputation Discovers Genetic Signals for Anthropometric Traits. | Height | 1,249 who | 205,003 E | 4q31.21 | HHIP | HHIP | | | ENSG00000164161 | NA | NA | rs2131354-A | rs2131354 | 0 | 2131354 | intron_variant | 0 | 0.5368 | 2.00E-19 | 18.69897 | | 0.0599 | [0.047-0.0 | Affymetrix, Illumina, Perlegen [15844966] (imputed) | N |
| 6 | 4:145586459 | 4 | 1.46E+08 | rs2131354 | 18/05/2021 | 3.3E+07 | Hebbar P | 9/09/2020 | Hum Genet | www.ncbi.nlm.nih.gov/pubmed/32902719 | Genome-wide landscape establishes novel association signals for metabolic traits in the Arab population. | Height | 2,732 Kuw | NA | 4q31.21 | HHIP | HHIP | | | ENSG00000164161 | NA | NA | rs2131354-? | rs2131354 | 0 | 2131354 | intron_variant | 0 | NR | 8.00E-08 | 7.09691 | | 5.358 | NR unit inc | Illumina [up to 1520000] (imputed) | N |
| 6 | 4:145586459 | 4 | 1.46E+08 | rs2131354 | 17/07/2019 | 2.9E+07 | Tachmazidou I | 1/06/2017 | Am J Hum Genet | www.ncbi.nlm.nih.gov/pubmed/28552196 | Whole-Genome Sequencing Coupled to Imputation Discovers Genetic Signals for Anthropometric Traits. | Hip circumference adjusted for BMI | 1,247 who | 205,905 E | 4q31.21 | HHIP | HHIP | | | ENSG00000164161 | NA | NA | rs2131354-A | rs2131354 | 0 | 2131354 | intron_variant | 0 | 0.5421 | 2.00E-09 | 8.69897 | | 0.0516 | [0.035-0.0 | Affymetrix, Illumina, Perlegen [15844966] (imputed) | N |
| 6 | 4:145586459 | 4 | 1.46E+08 | rs2131354 | 5/08/2019 | 3.1E+07 | Warrington NM | 1/05/2019 | Nat Genet | www.ncbi.nlm.nih.gov/pubmed/31043758 | Maternal and fetal genetic effects on birth weight and their relevance to cardio-metabolic risk factors. | Offspring birth weight | 210,267 E | NR | 4q31.21 | LOC646576, HHI | HHIP | | | ENSG00000164161 | NA | NA | rs2131354-A | rs2131354 | 0 | 2131354 | intron_variant | 0 | 0.526791 | 3.00E-16 | 15.52288 | | 0.025738 | [0.02-0.03 | Affymetrix, Illumina [14869762] (imputed) | N |
| 6 | 4:145586459 | 4 | 1.46E+08 | rs2131354 | 4/08/2022 | 3.6E+07 | Fan CC | 3/05/2022 | Nat Commun | www.ncbi.nlm.nih.gov/pubmed/35505052 | Multivariate genome-wide association study on tissue-sensitive diffusion metrics highlights pathways that shape the human brain. | Whole brain restricted directional diffusion (multivariate analysis) | 23,543 Eu | 11,555 Eu | 4q31.21 | | HHIP | | | ENSG00000164161 | NA | NA | rs2131354-? | rs2131354 | 0 | 2131354 | intron_variant | 0 | NR | 3.00E-19 | 18.52288 | (EA) | NA | | NR [NR] (imputed) | N |
| 6 | 4:145905213 | 4 | 1.46E+08 | rs4576021 | 7/02/2020 | 3.2E+07 | Hernandez Cordero AI | 21/11/2019 | Am J Hum Genet | www.ncbi.nlm.nih.gov/pubmed/31761296 | Genome-wide Associations Reveal Human-Mouse Genetic Convergence and Modifiers of Myogenesis, CPNE1 and STC2. | Appendicular lean mass | 181,862 E | NA | 4q31.21 | NR | HHIP - ANAPC10 | ENSG00000164161 | ENSG00000164162 | | 47346 | 39291 | rs4576021-T | rs4576021 | 0 | 4576021 | intron_variant | 1 | 0.42047 | 2.00E-11 | 10.69897 | | 0.046526 | [0.033-0.0 | Affymetrix [17914406] (imputed) | N |
| 6 | 4:145905213 | 4 | 1.46E+08 | rs4576021 | 18/10/2018 | 3E+07 | Kim SK | 26/07/2018 | PLoS One | www.ncbi.nlm.nih.gov/pubmed/30048462 | Identification of 613 new loci associated with heel bone mineral density and a polygenic risk score for bone mineral density, osteoporosis and fracture. | Heel bone mineral density | 394,929 E | NA | 4q31.21 | | HHIP - ANAPC10 | ENSG00000164161 | ENSG00000164162 | | 47346 | 39291 | rs4576021-? | rs4576021 | 0 | 4576021 | intron_variant | 1 | NR | 1.00E-23 | 23 | | 0.020726 | [0.017-0.0 | Affymetrix [20259828] (imputed) | N |
| 6 | 4:145655389 | 4 | 1.46E+08 | rs55817790 | 24/08/2021 | 3.4E+07 | Christakoudi S | 21/05/2021 | Sci Rep | www.ncbi.nlm.nih.gov/pubmed/34021172 | GWAS of allometric body-shape indices in UK Biobank identifies loci suggesting associations with morphogenesis, organogenesis, adrenal cell renewal and cancer. | Waist-hip index | 219,872 B | NA | 4q31.21 | HHIP | HHIP | | | ENSG00000164161 | NA | NA | rs55817790-A | rs55817790 | 0 | 55817790 | intron_variant | 0 | NR | 4.00E-08 | 7.39794 | | 0.015691 | [0.01-0.02 | Affymetrix [319769] (imputed) | N |
| 6 | 4:145655389 | 4 | 1.46E+08 | rs55817790 | 24/08/2021 | 3.4E+07 | Christakoudi S | 21/05/2021 | Sci Rep | www.ncbi.nlm.nih.gov/pubmed/34021172 | GWAS of allometric body-shape indices in UK Biobank identifies loci suggesting associations with morphogenesis, organogenesis, adrenal cell renewal and cancer. | Waist-to-hip ratio adjusted for BMI | 219,872 B | NA | 4q31.21 | HHIP | HHIP | | | ENSG00000164161 | NA | NA | rs55817790-A | rs55817790 | 0 | 55817790 | intron_variant | 0 | NR | 3.00E-09 | 8.522879 | | 0.016937 | [0.011-0.0 | Affymetrix [319769] (imputed) | N |
| 6 | 4:145905213 | 4 | 1.46E+08 | rs6537315 | 28/05/2020 | 3.2E+07 | Hysi PG | 30/03/2020 | Nat Genet | www.ncbi.nlm.nih.gov/pubmed/32231278 | Meta-analysis of 542,934 subjects of European ancestry identifies new genes and mechanisms predisposing to refractive error and myopia. | Refractive error | 542,934 E | NA | 4q31.21 | HHIP, OTUD4 | HHIP - ANAPC10 | ENSG00000164161 | ENSG00000164162 | | 58289 | 28348 | rs6537315-? | rs6537315 | 0 | 6537315 | intron_variant | 1 | | 5.00E-09 | 8.30103 | | NA | | Affymetrix, Illumina [NR] (imputed) | N |
| 6 | 4:145905213 | 4 | 1.46E+08 | rs67365433 | 7/02/2020 | 3.2E+07 | Hernandez Cordero AI | 21/11/2019 | Am J Hum Genet | www.ncbi.nlm.nih.gov/pubmed/31761296 | Genome-wide Associations Reveal Human-Mouse Genetic Convergence and Modifiers of Myogenesis, CPNE1 and STC2. | Appendicular lean mass | 181,862 E | NA | 4q31.21 | NR | ANAPC10 | | | ENSG00000164162 | NA | NA | rs67365433-A | rs67365433 | 0 | 67365433 | intron_variant | 0 | 0.423698 | 1.00E-12 | 12 | | 0.049167 | [0.036-0.0 | Affymetrix [17914406] (imputed) | N |
| 6 | 4:145905213 | 4 | 1.46E+08 | rs67365433 | 24/08/2021 | 3.4E+07 | Christakoudi S | 21/05/2021 | Sci Rep | www.ncbi.nlm.nih.gov/pubmed/34021172 | GWAS of allometric body-shape indices in UK Biobank identifies loci suggesting associations with morphogenesis, organogenesis, adrenal cell renewal and cancer. | Waist-hip index | 186,825 B | NA | 4q31.21 | ANAPC10 | ANAPC10 | | | ENSG00000164162 | NA | NA | rs67365433-A | rs67365433 | 0 | 67365433 | intron_variant | 0 | NR | 2.00E-09 | 8.69897 | | 0.018861 | [0.013-0.0 | Affymetrix [127266] (imputed) | N |
| 6 | 4:145905213 | 4 | 1.46E+08 | rs67365433 | 24/08/2021 | 3.4E+07 | Christakoudi S | 21/05/2021 | Sci Rep | www.ncbi.nlm.nih.gov/pubmed/34021172 | GWAS of allometric body-shape indices in UK Biobank identifies loci suggesting associations with morphogenesis, organogenesis, adrenal cell renewal and cancer. | Waist-to-hip ratio adjusted for BMI | 186,825 B | NA | 4q31.21 | ANAPC10 | ANAPC10 | | | ENSG00000164162 | NA | NA | rs67365433-A | rs67365433 | 0 | 67365433 | intron_variant | 0 | NR | 2.00E-10 | 9.69897 | | 0.020219 | [0.014-0.0 | Affymetrix [323858] (imputed) | N |
| 6 | 4:145655389 | 4 | 1.46E+08 | rs6812830 | 14/02/2022 | 3.5E+07 | Chambers T | 25/01/2022 | Mol Psychiatry | www.ncbi.nlm.nih.gov/pubmed/35079123 | Genetic common variants associated with cerebellar volume and their overlap with mental disorders: a study on 33,265 individuals from the UK-Biobank. | Total cerebellar volume (excluding Crus I vermis) | 33,265 Eu | NA | 4q31.21 | | HHIP | | | ENSG00000164161 | NA | NA | rs6812830-A | rs6812830 | 0 | 6812830 | intron_variant | 0 | NR | 2.00E-08 | 7.69897 | | 0.0306 | [0.02-0.04 | Affymetrix [6193476] (imputed) | N |
| 6 | 4:145655389 | 4 | 1.46E+08 | rs6817771 | 26/04/2022 | 3.5E+07 | Fernandez-Rhodes L | 11/03/2022 | HGG Adv | www.ncbi.nlm.nih.gov/pubmed/35399580 | Ancestral diversity improves discovery and fine-mapping of genetic loci for anthropometric traits-The Hispanic/Latino Anthropometry Consortium. | Height | 59,771 His | 8,110 Hisp | 4q31.21 | | HHIP | | | ENSG00000164161 | NA | NA | rs6817771-C | rs6817771 | 0 | 6817771 | intron_variant | 0 | 0.4102 | 4.00E-10 | 9.39794 | | 0.0363 | [0.025-0.0 | Affymetrix, Illumina [8927938] (imputed) | N |
| 6 | 4:145655389 | 4 | 1.46E+08 | rs6827641 | 4/08/2022 | 3.6E+07 | Fan CC | 3/05/2022 | Nat Commun | www.ncbi.nlm.nih.gov/pubmed/35505052 | Multivariate genome-wide association study on tissue-sensitive diffusion metrics highlights pathways that shape the human brain. | Whole brain free water diffusion (multivariate analysis) | 23,543 Eu | 11,555 Eu | 4q31.21 | | HHIP | | | ENSG00000164161 | NA | NA | rs6827641-? | rs6827641 | 0 | 6827641 | intron_variant | 0 | NR | 3.00E-17 | 16.52288 | (EA) | NA | | NR [NR] (imputed) | N |
| 6 | 4:145586459 | 4 | 1.46E+08 | rs6845999 | 5/08/2019 | 3.1E+07 | Warrington NM | 1/05/2019 | Nat Genet | www.ncbi.nlm.nih.gov/pubmed/31043758 | Maternal and fetal genetic effects on birth weight and their relevance to cardio-metabolic risk factors. | Birth weight | 298,142 E | NR | 4q31.21 | LOC646576, HHI | HHIP-AS1 | | | ENSG00000248890 | NA | NA | rs6845999-T | rs6845999 | 0 | 6845999 | intron_variant | 0 | 0.431122 | 2.00E-24 | 23.69897 | | 0.026255 | [0.021-0.0 | Affymetrix, Illumina [15144367] (imputed) | N |
| 6 | 4:145586459 | 4 | 1.46E+08 | rs6845999 | 15/07/2015 | 2.5E+07 | He M | 26/11/2014 | Hum Mol Genet | www.ncbi.nlm.nih.gov/pubmed/25429064 | Meta-analysis of genome-wide association studies of adult height in East Asians identifies 17 novel loci. | Height | 36,227 Ea | 57,699 Ea | 4q31.21 | HHIP | HHIP-AS1 | | | ENSG00000248890 | NA | NA | rs6845999-T | rs6845999 | 0 | 6845999 | intron_variant | 0 | 0.22 | 4.00E-14 | 13.39794 | | 0.037 | [0.021-0.0 | Affymetrix, Illumina [2704730] (imputed) | N |
| 6 | 4:145586459 | 4 | 1.46E+08 | rs6845999 | 2/07/2019 | 3.1E+07 | Wojcik GL | 19/06/2019 | Nature | www.ncbi.nlm.nih.gov/pubmed/31217584 | Genetic analyses of diverse populations improves discovery for complex traits. | Height | 17,286 Afr | NA | 4q31.21 | NR | HHIP-AS1 | | | ENSG00000248890 | NA | NA | rs6845999-? | rs6845999 | 0 | 6845999 | intron_variant | 0 | NR | 3.00E-08 | 7.522879 | | 0.036422 | [0.024-0.0 | Illumina [34656550] (imputed) | N |
| 6 | 4:145586459 | 4 | 1.46E+08 | rs6845999 | 26/04/2022 | 3.5E+07 | Fernandez-Rhodes L | 11/03/2022 | HGG Adv | www.ncbi.nlm.nih.gov/pubmed/35399580 | Ancestral diversity improves discovery and fine-mapping of genetic loci for anthropometric traits-The Hispanic/Latino Anthropometry Consortium. | Height | 59,771 His | 8,110 Hisp | 4q31.21 | | HHIP-AS1 | | | ENSG00000248890 | NA | NA | rs6845999-T | rs6845999 | 0 | 6845999 | intron_variant | 0 | 0.4323 | 3.00E-16 | 15.52288 | | 0.0431 | [0.033-0.0 | Affymetrix, Illumina [8927938] (imputed) | N |
| 6 | 4:145586459 | 4 | 1.46E+08 | rs6845999 | 5/08/2022 | 3.6E+07 | Chiou JS | 13/07/2022 | BMC Med | www.ncbi.nlm.nih.gov/pubmed/35831902 | Your height affects your health: genetic determinants and health-related outcomes in Taiwan. | Height | 67,452 Ha | NA | 4q31.21 | | HHIP-AS1 | | | ENSG00000248890 | NA | NA | rs6845999-T | rs6845999 | 0 | 6845999 | intron_variant | 0 | NR | 7.00E-22 | 21.1549 | | 0.058 | [0.046-0.0 | Affymetrix [NR] (imputed) | N |
| 6 | 4:145655389 | 4 | 1.46E+08 | rs6854783 | 17/09/2008 | 1.8E+07 | Weedon MN | 6/04/2008 | Nat Genet | www.ncbi.nlm.nih.gov/pubmed/18391952 | Genome-wide association analysis identifies 20 loci that influence adult height. | Height | 13,665 Eu | 16,482 Eu | 4q31.21 | HHIP | HHIP | | | ENSG00000164161 | NA | NA | rs6854783-A | rs6854783 | 0 | 6854783 | intron_variant | 0 | 0.43 | 2.00E-09 | 8.69897 | | 0.06 | [0.03-0.08 | Affymetrix [402951] | N |

| | | | | | | | | | | | | | | | | | | | | | | | | | | | | | | | | | | | | |
|---|---|---|---|---|---|---|---|---|---|---|---|---|---|---|---|---|---|---|---|---|---|---|---|---|---|---|---|---|---|---|---|---|---|---|---|---|
| 6 | 4:145586459 | 4 | 1.46E+08 | rs7675744 | 24/08/2021 | 3.4E+07 | Christakoudi S | 21/05/2021 | Sci Rep | www.ncbi.nlm.nih.gov/pubmed/34021172 | GWAS of allometric body-shape indices in UK Biobank identifies loci suggesting associations with morphogenesis, organogenesis, adrenal cell renewal and cancer. | Hip circumference adjusted for BMI | 219,872 B | NA | 4q31.21 | HHIP-AS1, HHIP | HHIP-AS1, HHIP | | | ENSG00000248890, E | NA | NA | rs7675744-C | rs7675744 | 0 | 7675744 | non_coding_transcript_exon | 0 | NR | 2.00E-50 | 49.69897 | | 0.04237 | [0.037-0.0 | Affymetrix [321016] (imputed) | N |
| 6 | 4:145586459 | 4 | 1.46E+08 | rs7692554 | 25/03/2022 | 3.4E+07 | Lin E | 16/07/2021 | Hum Mol Genet | www.ncbi.nlm.nih.gov/pubmed/34270706 | Genome-wide association study in the Taiwan biobank identifies four novel genes for human height: NABP2, RASA2, RNF41 and SLC39A5. | Height | 14,571 Tai | 20,506 Tai | 4q31.21 | HHIP-LOC10537 | KRT18P51 - HHIP | ENSG00000250504 | ENSG00000248890 | | 42201 | 27735 | rs7692554-? | rs7692554 | 0 | 7692554 | intron_variant | 1 | NR | 4.00E-08 | 7.39794 | | 0.44 | [0.28-0.6] | Affymetrix [5682219] (imputed) | N |
| 6 | 4:145586459 | 4 | 1.46E+08 | rs77362667 | 25/03/2022 | 3.4E+07 | Lin E | 16/07/2021 | Hum Mol Genet | www.ncbi.nlm.nih.gov/pubmed/34270706 | Genome-wide association study in the Taiwan biobank identifies four novel genes for human height: NABP2, RASA2, RNF41 and SLC39A5. | Height | 14,571 Tai | 20,506 Tai | 4q31.21 | HHIP | HHIP | | | ENSG00000164161 | NA | NA | rs77362667-? | rs77362667 | 0 | 77362667 | intron_variant | 0 | NR | 3.00E-08 | 7.522879 | | 0.45 | [0.29-0.61 | Affymetrix [5682219] (imputed) | N |
| 6 | 4:145905213 | 4 | 1.46E+08 | rs789351 | 24/08/2021 | 3.4E+07 | Christakoudi S | 21/05/2021 | Sci Rep | www.ncbi.nlm.nih.gov/pubmed/34021172 | GWAS of allometric body-shape indices in UK Biobank identifies loci suggesting associations with morphogenesis, organogenesis, adrenal cell renewal and cancer. | Hip circumference adjusted for BMI | 219,872 B | NA | 4q31.21 | ANAPC10 | ANAPC10 | | | ENSG00000164162 | NA | NA | rs789351-T | rs789351 | 0 | 789351 | intron_variant | 0 | NR | 3.00E-27 | 26.52288 | | 0.030705 | [0.025-0.0 | Affymetrix [321016] (imputed) | N |
| 6 | 4:145905213 | 4 | 1.46E+08 | rs789351 | 4/11/2019 | 3E+07 | Pulit SL | 14/09/2018 | Hum Mol Genet | www.ncbi.nlm.nih.gov/pubmed/30239722 | Meta-analysis of genome-wide association studies for body fat distribution in 694,649 individuals of European ancestry. | Waist-hip ratio | 697,734 E | NA | 4q31.21 | NA | ANAPC10 | | | ENSG00000164162 | NA | NA | rs789351-T | rs789351 | 0 | 789351 | intron_variant | 0 | 0.4202 | 8.00E-13 | 12.09691 | | 0.0123 | [0.009-0.0 | NR [~ 27400000] (imputed) | N |
| 6 | 4:145586459 | 4 | 1.46E+08 | rs982903 | 17/04/2020 | 3.2E+07 | Zhu Z | 24/10/2019 | J Allergy Clin Immunol | www.ncbi.nlm.nih.gov/pubmed/31669095 | Shared Genetic and Experimental Links between Obesity-Related Traits and Asthma Subtypes in UK Biobank. | Waist circumference adjusted for body mass index | 457,690 E | NA | 4q31.21 | NR | HHIP | | | ENSG00000164161 | NA | NA | rs982903-? | rs982903 | 0 | 982903 | intron_variant | 0 | NR | 3.00E-13 | 12.52288 | | NA | | Affymetrix [~ 8270000] (imputed) | N |
| 7 | 10:71099109 | 10 | 71094504 | rs17476364 | 21/07/2020 | 3.2E+07 | Richardson TG | 23/03/2020 | PLoS Med | www.ncbi.nlm.nih.gov/pubmed/32203549 | Evaluating the relationship between circulating lipoprotein lipids and apolipoproteins with risk of coronary heart disease: A multivariable Mendelian randomisation analysis. | Apolipoprotein B levels | 439,214 E | NA | 10q22.1 | HK1 | HK1 | | | ENSG00000156515 | NA | NA | rs17476364-T | rs17476364 | 0 | 17476364 | intron_variant | 0 | 0.891606 | 3.00E-10 | 9.522879 | | 0.021129 | [0.015-0.0 | Affymetrix [NR] (imputed) | N |
| 7 | 10:71099109 | 10 | 71094504 | rs17476364 | 10/06/2021 | 3.4E+07 | Chen J | 31/05/2021 | Nat Genet | www.ncbi.nlm.nih.gov/pubmed/34059833 | The trans-ancestral genomic architecture of glycemic traits. | Glycated hemoglobin levels | 146,806 E | NA | 10q22.1 | HK1 | HK1 | | | ENSG00000156515 | NA | NA | rs17476364-T | rs17476364 | 0 | 17476364 | intron_variant | 0 | 0.9011 | 1E-314 | 314 | | 0.0858 | [0.081-0.0 | Affymetrix, Illumina [33811881] (imputed) | N |
| 7 | 10:71099109 | 10 | 71094504 | rs17476364 | 30/08/2017 | 2.8E+07 | Astle WJ | 17/11/2016 | Cell | www.ncbi.nlm.nih.gov/pubmed/27863252 | The Allelic Landscape of Human Blood Cell Trait Variation and Links to Common Complex Disease. | Hematocrit | 173,039 E | NA | 10q22.1 | HK1 | HK1 | | | ENSG00000156515 | NA | NA | rs17476364-C | rs17476364 | 0 | 17476364 | intron_variant | 0 | 0.1103 | ####### | 158.0969 | | 0.151778 | [0.14-0.16 | Affymetrix [~ 2500000] (imputed) | N |
| 7 | 10:71099109 | 10 | 71094504 | rs17476364 | 22/03/2021 | 3.2E+07 | Hodonsky CJ | 14/03/2020 | BMC Genomics | www.ncbi.nlm.nih.gov/pubmed/32171239 | Ancestry-specific associations identified in genome-wide combined-phenotype study of red blood cell traits emphasize benefits of diversity in genomics. | Hematocrit | 16,184 Afri | NA | 10q22.1 | | HK1 | | | ENSG00000156515 | NA | NA | rs17476364-? | rs17476364 | 0 | 17476364 | intron_variant | 0 | 0.08 | 4.00E-26 | 25.39794 | | 0.3706 | [0.3-0.44] | Affymetrix, Illumina [21000000] (imputed) | N |
| 7 | 10:71099109 | 10 | 71094504 | rs17476364 | 22/03/2021 | 3.2E+07 | Hodonsky CJ | 14/03/2020 | BMC Genomics | www.ncbi.nlm.nih.gov/pubmed/32171239 | Ancestry-specific associations identified in genome-wide combined-phenotype study of red blood cell traits emphasize benefits of diversity in genomics. | Hematocrit | 29,383 Eur | NA | 10q22.1 | | HK1 | | | ENSG00000156515 | NA | NA | rs17476364-? | rs17476364 | 0 | 17476364 | intron_variant | 0 | 0.0946 | 1.00E-17 | 17 | | 0.361 | [0.28-0.44 | Affymetrix, Illumina [21000000] (imputed) | N |
| 7 | 10:71099109 | 10 | 71094504 | rs17476364 | 22/03/2021 | 3.2E+07 | Hodonsky CJ | 14/03/2020 | BMC Genomics | www.ncbi.nlm.nih.gov/pubmed/32171239 | Ancestry-specific associations identified in genome-wide combined-phenotype study of red blood cell traits emphasize benefits of diversity in genomics. | Hematocrit | 20,704 His | NA | 10q22.1 | | HK1 | | | ENSG00000156515 | NA | NA | rs17476364-? | rs17476364 | 0 | 17476364 | intron_variant | 0 | 0.0651 | 7.00E-09 | 8.154902 | | 0.3816 | [0.25-0.51 | Affymetrix, Illumina [21000000] (imputed) | N |
| 7 | 10:71099109 | 10 | 71094504 | rs17476364 | 18/09/2020 | 3.3E+07 | Vuckovic D | 1/09/2020 | Cell | www.ncbi.nlm.nih.gov/pubmed/32888494 | The Polygenic and Monogenic Basis of Blood Traits and Diseases. | Hematocrit | 408,112 B | NA | 10q22.1 | HK1 | HK1 | | | ENSG00000156515 | NA | NA | rs17476364-C | rs17476364 | 0 | 17476364 | intron_variant | 0 | 0.108758 | 5E-372 | 371.301 | | 0.147147 | [0.14-0.15 | Affymetrix [93095623] (imputed) | N |
| 7 | 10:71099109 | 10 | 71094504 | rs17476364 | 14/12/2021 | 3.5E+07 | Sakaue S | 30/09/2021 | Nat Genet | www.ncbi.nlm.nih.gov/pubmed/34594039 | A cross-population atlas of genetic associations for 220 human phenotypes. | Hematocrit | 350,475 E | NA | 10q22.1 | | HK1 | | | ENSG00000156515 | NA | NA | rs17476364-C | rs17476364 | 0 | 17476364 | intron_variant | 0 | NR | ####### | 282 | | 0.1115 | [0.11-0.12 | Affymetrix, Illumina [20538534] (imputed) | N |
| 7 | 10:71099109 | 10 | 71094504 | rs17476364 | 18/09/2020 | 3.3E+07 | Vuckovic D | 1/09/2020 | Cell | www.ncbi.nlm.nih.gov/pubmed/32888494 | The Polygenic and Monogenic Basis of Blood Traits and Diseases. | Hemoglobin | 408,112 B | NA | 10q22.1 | HK1 | HK1 | | | ENSG00000156515 | NA | NA | rs17476364-C | rs17476364 | 0 | 17476364 | intron_variant | 0 | 0.108778 | 1E-368 | 368 | | 0.146551 | [0.14-0.15 | Affymetrix [93095623] (imputed) | N |
| 7 | 10:71099109 | 10 | 71094504 | rs17476364 | 14/12/2021 | 3.5E+07 | Sakaue S | 30/09/2021 | Nat Genet | www.ncbi.nlm.nih.gov/pubmed/34594039 | A cross-population atlas of genetic associations for 220 human phenotypes. | Hemoglobin | 350,474 E | NA | 10q22.1 | | HK1 | | | ENSG00000156515 | NA | NA | rs17476364-C | rs17476364 | 0 | 17476364 | intron_variant | 0 | NR | ####### | 288.5229 | | 0.1098 | [0.1-0.12] | Affymetrix, Illumina [20538521] (imputed) | N |
| 7 | 10:71099109 | 10 | 71094504 | rs17476364 | 14/12/2021 | 3.5E+07 | Sakaue S | 30/09/2021 | Nat Genet | www.ncbi.nlm.nih.gov/pubmed/34594039 | A cross-population atlas of genetic associations for 220 human phenotypes. | Hemoglobin A1c levels | 344,182 E | NA | 10q22.1 | | HK1 | | | ENSG00000156515 | NA | NA | rs17476364-C | rs17476364 | 0 | 17476364 | intron_variant | 0 | NR | 4E-1745 | 1744.398 | | 0.3256 | [0.32-0.33 | Affymetrix, Illumina [20525742] (imputed) | N |
| 7 | 10:71099109 | 10 | 71094504 | rs17476364 | 30/08/2017 | 2.8E+07 | Astle WJ | 17/11/2016 | Cell | www.ncbi.nlm.nih.gov/pubmed/27863252 | The Allelic Landscape of Human Blood Cell Trait Variation and Links to Common Complex Disease. | Hemoglobin concentration | 172,925 E | NA | 10q22.1 | HK1 | HK1 | | | ENSG00000156515 | NA | NA | rs17476364-C | rs17476364 | 0 | 17476364 | intron_variant | 0 | 0.1103 | ####### | 156 | | 0.151392 | [0.14-0.16 | Affymetrix [~ 2500000] (imputed) | N |
| 7 | 10:71099109 | 10 | 71094504 | rs17476364 | 22/03/2021 | 3.2E+07 | Hodonsky CJ | 14/03/2020 | BMC Genomics | www.ncbi.nlm.nih.gov/pubmed/32171239 | Ancestry-specific associations identified in genome-wide combined-phenotype study of red blood cell traits emphasize benefits of diversity in genomics. | Hemoglobin concentration | 29,379 Eur | NA | 10q22.1 | | HK1 | | | ENSG00000156515 | NA | NA | rs17476364-? | rs17476364 | 0 | 17476364 | intron_variant | 0 | 0.0946 | 9.00E-19 | 18.04576 | | 0.126 | [0.098-0.1 | Affymetrix, Illumina [21000000] (imputed) | N |
| 7 | 10:71099109 | 10 | 71094504 | rs17476364 | 22/03/2021 | 3.2E+07 | Hodonsky CJ | 14/03/2020 | BMC Genomics | www.ncbi.nlm.nih.gov/pubmed/32171239 | Ancestry-specific associations identified in genome-wide combined-phenotype study of red blood cell traits emphasize benefits of diversity in genomics. | Hemoglobin concentration | 20,692 His | NA | 10q22.1 | | HK1 | | | ENSG00000156515 | NA | NA | rs17476364-? | rs17476364 | 0 | 17476364 | intron_variant | 0 | 0.0651 | 6.00E-10 | 9.221849 | | 0.14 | [0.096-0.1 | Affymetrix, Illumina [21000000] (imputed) | N |
| 7 | 10:71099109 | 10 | 71094504 | rs17476364 | 22/03/2021 | 3.2E+07 | Hodonsky CJ | 14/03/2020 | BMC Genomics | www.ncbi.nlm.nih.gov/pubmed/32171239 | Ancestry-specific associations identified in genome-wide combined-phenotype study of red blood cell traits emphasize benefits of diversity in genomics. | Hemoglobin concentration | 16,186 Afri | NA | 10q22.1 | | HK1 | | | ENSG00000156515 | NA | NA | rs17476364-? | rs17476364 | 0 | 17476364 | intron_variant | 0 | 0.08 | 1.00E-27 | 27 | | 0.1301 | [0.11-0.15 | Affymetrix, Illumina [21000000] (imputed) | N |
| 7 | 10:71099109 | 10 | 71094504 | rs17476364 | 30/08/2017 | 2.8E+07 | Astle WJ | 17/11/2016 | Cell | www.ncbi.nlm.nih.gov/pubmed/27863252 | The Allelic Landscape of Human Blood Cell Trait Variation and Links to Common Complex Disease. | High light scatter reticulocyte count | 170,761 E | NA | 10q22.1 | HK1 | HK1 | | | ENSG00000156515 | NA | NA | rs17476364-C | rs17476364 | 0 | 17476364 | intron_variant | 0 | 0.1104 | 3.00E-69 | 68.52288 | | 0.100903 | [0.09-0.11 | Affymetrix [~ 2500000] (imputed) | N |
| 7 | 10:71099109 | 10 | 71094504 | rs17476364 | 18/09/2020 | 3.3E+07 | Vuckovic D | 1/09/2020 | Cell | www.ncbi.nlm.nih.gov/pubmed/32888494 | The Polygenic and Monogenic Basis of Blood Traits and Diseases. | High light scatter reticulocyte count | 408,112 B | NA | 10q22.1 | HK1 | HK1 | | | ENSG00000156515 | NA | NA | rs17476364-C | rs17476364 | 0 | 17476364 | intron_variant | 0 | 0.108757 | ####### | 194.2218 | | 0.106568 | [0.1-0.114 | Affymetrix [93095623] (imputed) | N |
| 7 | 10:71099109 | 10 | 71094504 | rs17476364 | 30/08/2017 | 2.8E+07 | Astle WJ | 17/11/2016 | Cell | www.ncbi.nlm.nih.gov/pubmed/27863252 | The Allelic Landscape of Human Blood Cell Trait Variation and Links to Common Complex Disease. | High light scatter reticulocyte percentage of red cells | 170,763 E | NA | 10q22.1 | HK1 | HK1 | | | ENSG00000156515 | NA | NA | rs17476364-C | rs17476364 | 0 | 17476364 | intron_variant | 0 | 0.1104 | 3.00E-53 | 52.52288 | | 0.08805 | [0.077-0.0 | Affymetrix [~ 2500000] (imputed) | N |
| 7 | 10:71099109 | 10 | 71094504 | rs17476364 | 18/09/2020 | 3.3E+07 | Vuckovic D | 1/09/2020 | Cell | www.ncbi.nlm.nih.gov/pubmed/32888494 | The Polygenic and Monogenic Basis of Blood Traits and Diseases. | High light scatter reticulocyte percentage of red cells | 408,112 B | NA | 10q22.1 | HK1 | HK1 | | | ENSG00000156515 | NA | NA | rs17476364-C | rs17476364 | 0 | 17476364 | intron_variant | 0 | 0.108749 | ####### | 151.699 | | 0.094088 | [0.087-0.1 | Affymetrix [93095623] (imputed) | N |
| 7 | 10:71099109 | 10 | 71094504 | rs17476364 | 18/09/2020 | 3.3E+07 | Vuckovic D | 1/09/2020 | Cell | www.ncbi.nlm.nih.gov/pubmed/32888494 | The Polygenic and Monogenic Basis of Blood Traits and Diseases. | Immature fraction of reticulocytes | 408,112 B | NA | 10q22.1 | HK1 | HK1 | | | ENSG00000156515 | NA | NA | rs17476364-C | rs17476364 | 0 | 17476364 | intron_variant | 0 | 0.10878 | 6.00E-40 | 39.22185 | | 0.047658 | [0.041-0.0 | Affymetrix [93095623] (imputed) | N |
| 7 | 10:71099109 | 10 | 71094504 | rs17476364 | 22/03/2021 | 3.4E+07 | Bell S | 3/02/2021 | Commun Biol | www.ncbi.nlm.nih.gov/pubmed/33536631 | A genome-wide meta-analysis yields 46 new loci associating with biomarkers of iron homeostasis. | Iron status biomarkers (ferritin levels) | 246,139 E | NA | 10q22.1 | HK1 | HK1 | | | ENSG00000156515 | NA | NA | rs17476364-C | rs17476364 | 0 | 17476364 | intron_variant | 0 | 0.108 | 4.00E-14 | 13.39794 | | 0.043 | [0.032-0.0 | Affymetrix, Illumina [40000000] (imputed) | N |
| 7 | 10:71099109 | 10 | 71094504 | rs17476364 | 21/07/2020 | 3.2E+07 | Richardson TG | 23/03/2020 | PLoS Med | www.ncbi.nlm.nih.gov/pubmed/32203549 | Evaluating the relationship between circulating lipoprotein lipids and apolipoproteins with risk of coronary heart disease: A multivariable Mendelian randomisation analysis. | LDL cholesterol levels | 440,546 E | NA | 10q22.1 | HK1 | HK1 | | | ENSG00000156515 | NA | NA | rs17476364-T | rs17476364 | 0 | 17476364 | intron_variant | 0 | 0.891668 | 9.00E-11 | 10.04576 | | 0.021785 | [0.015-0.0 | Affymetrix [NR] (imputed) | N |
| 7 | 10:71099109 | 10 | 71094504 | rs17476364 | 17/11/2022 | 3.5E+07 | Graham SE | 9/12/2021 | Nature | www.ncbi.nlm.nih.gov/pubmed/34887591 | The power of genetic diversity in genome-wide association studies of lipids. | Low density lipoprotein cholesterol levels | 40,963 Sou | NA | 10q22.1 | | HK1 | | | ENSG00000156515 | NA | NA | rs17476364-? | rs17476364 | 0 | 17476364 | intron_variant | 0 | NR | 1.00E-26 | 26 | | NA | | NR [52000000] (imputed) | N |
| 7 | 10:71099109 | 10 | 71094504 | rs17476364 | 17/11/2022 | 3.5E+07 | Graham SE | 9/12/2021 | Nature | www.ncbi.nlm.nih.gov/pubmed/34887591 | The power of genetic diversity in genome-wide association studies of lipids. | Low density lipoprotein cholesterol levels | 1,320,016 | NA | 10q22.1 | | HK1 | | | ENSG00000156515 | NA | NA | rs17476364-C | rs17476364 | 0 | 17476364 | intron_variant | 0 | 0.098856 | 3.00E-23 | 22.52288 | | 0.026846 | [0.022-0.0 | NR [52000000] (imputed) | N |
| 7 | 10:71099109 | 10 | 71094504 | rs17476364 | 30/08/2017 | 2.8E+07 | Astle WJ | 17/11/2016 | Cell | www.ncbi.nlm.nih.gov/pubmed/27863252 | The Allelic Landscape of Human Blood Cell Trait Variation and Links to Common Complex Disease. | Mean corpuscular hemoglobin | 172,332 E | NA | 10q22.1 | HK1 | HK1 | | | ENSG00000156515 | NA | NA | rs17476364-C | rs17476364 | 0 | 17476364 | intron_variant | 0 | 0.1103 | 4.00E-44 | 43.39794 | | 0.078982 | [0.068-0.0 | Affymetrix [~ 2500000] (imputed) | N |
| 7 | 10:71099109 | 10 | 71094504 | rs17476364 | 18/09/2020 | 3.3E+07 | Vuckovic D | 1/09/2020 | Cell | www.ncbi.nlm.nih.gov/pubmed/32888494 | The Polygenic and Monogenic Basis of Blood Traits and Diseases. | Mean corpuscular hemoglobin | 408,112 B | NA | 10q22.1 | HK1 | HK1 | | | ENSG00000156515 | NA | NA | rs17476364-C | rs17476364 | 0 | 17476364 | intron_variant | 0 | 0.108777 | ####### | 109.2218 | | 0.079404 | [0.072-0.0 | Affymetrix [93095623] (imputed) | N |
| 7 | 10:71099109 | 10 | 71094504 | rs17476364 | 21/09/2020 | 3.3E+07 | Chen MH | 1/09/2020 | Cell | www.ncbi.nlm.nih.gov/pubmed/32888493 | Trans-ethnic and Ancestry-Specific Blood-Cell Genetics in 746,667 Individuals from 5 Global Populations. | Mean corpuscular hemoglobin | 486,823 E | NA | 10q22.1 | NR | HK1 | | | ENSG00000156515 | NA | NA | rs17476364-C | rs17476364 | 0 | 17476364 | intron_variant | 0 | 0.108072 | ####### | 147.1549 | | 0.080275 | [0.074-0.0 | Affymetrix, Illumina [50266654] (imputed) | N |
| 7 | 10:71099109 | 10 | 71094504 | rs17476364 | 21/09/2020 | 3.3E+07 | Chen MH | 1/09/2020 | Cell | www.ncbi.nlm.nih.gov/pubmed/32888493 | Trans-ethnic and Ancestry-Specific Blood-Cell Genetics in 746,667 Individuals from 5 Global Populations. | Mean corpuscular hemoglobin | 630,125 Af | NA | 10q22.1 | NR | HK1 | | | ENSG00000156515 | NA | NA | rs17476364-C | rs17476364 | 0 | 17476364 | intron_variant | 0 | 0.104331 | ####### | 144 | | NA | | Affymetrix, Illumina [20825802] (imputed) | N |
| 7 | 10:71099109 | 10 | 71094504 | rs17476364 | 8/02/2019 | 3.1E+07 | Kichaev G | 27/12/2018 | Am J Hum Genet | www.ncbi.nlm.nih.gov/pubmed/30595370 | Leveraging Polygenic Functional Enrichment to Improve GWAS Power. | Mean corpuscular hemoglobin | approxima | NA | 10q22.1 | | HK1 | | | ENSG00000156515 | NA | NA | rs17476364-? | rs17476364 | 0 | 17476364 | intron_variant | 0 | NR | ####### | 127.301 | | NA | | NR [~ 8900000] (imputed) | N |
| 7 | 10:71099109 | 10 | 71094504 | rs17476364 | 14/12/2021 | 3.5E+07 | Sakaue S | 30/09/2021 | Nat Genet | www.ncbi.nlm.nih.gov/pubmed/34594039 | A cross-population atlas of genetic associations for 220 human phenotypes. | Mean corpuscular hemoglobin | 350,472 E | NA | 10q22.1 | | HK1 | | | ENSG00000156515 | NA | NA | rs17476364-C | rs17476364 | 0 | 17476364 | intron_variant | 0 | NR | 3.00E-78 | 77.52288 | | 0.0707 | [0.063-0.0 | Affymetrix, Illumina [20535878] (imputed) | N |
| 7 | 10:71099109 | 10 | 71094504 | rs17476364 | 30/08/2017 | 2.8E+07 | Astle WJ | 17/11/2016 | Cell | www.ncbi.nlm.nih.gov/pubmed/27863252 | The Allelic Landscape of Human Blood Cell Trait Variation and Links to Common Complex Disease. | Mean corpuscular volume | 172,433 E | NA | 10q22.1 | HK1 | HK1 | | | ENSG00000156515 | NA | NA | rs17476364-C | rs17476364 | 0 | 17476364 | intron_variant | 0 | 0.1103 | 7.00E-55 | 54.1549 | | 0.088114 | [0.077-0.0 | Affymetrix [~ 2500000] (imputed) | N |
| 7 | 10:71099109 | 10 | 71094504 | rs17476364 | 22/03/2021 | 3.2E+07 | Hodonsky CJ | 14/03/2020 | BMC Genomics | www.ncbi.nlm.nih.gov/pubmed/32171239 | Ancestry-specific associations identified in genome-wide combined-phenotype study of red blood cell traits emphasize benefits of diversity in genomics. | Mean corpuscular volume | 8,556 Afric | NA | 10q22.1 | | HK1 | | | ENSG00000156515 | NA | NA | rs17476364-? | rs17476364 | 0 | 17476364 | intron_variant | 0 | 0.08 | 1.00E-06 | 6 | | 0.3633 | [0.22-0.51 | Affymetrix, Illumina [21000000] (imputed) | N |
| 7 | 10:71099109 | 10 | 71094504 | rs17476364 | 18/09/2020 | 3.3E+07 | Vuckovic D | 1/09/2020 | Cell | www.ncbi.nlm.nih.gov/pubmed/32888494 | The Polygenic and Monogenic Basis of Blood Traits and Diseases. | Mean corpuscular volume | 408,112 B | NA | 10q22.1 | HK1 | HK1 | | | ENSG00000156515 | NA | NA | rs17476364-C | rs17476364 | 0 | 17476364 | intron_variant | 0 | 0.108777 | ####### | 142 | | 0.094037 | [0.088-0.1 | Affymetrix [93095623] (imputed) | N |
| 7 | 10:71099109 | 10 | 71094504 | rs17476364 | 21/09/2020 | 3.3E+07 | Chen MH | 1/09/2020 | Cell | www.ncbi.nlm.nih.gov/pubmed/32888493 | Trans-ethnic and Ancestry-Specific Blood-Cell Genetics in 746,667 Individuals from 5 Global Populations. | Mean corpuscular volume | 544,127 E | NA | 10q22.1 | NR | HK1 | | | ENSG00000156515 | NA | NA | rs17476364-C | rs17476364 | 0 | 17476364 | intron_variant | 0 | 0.10674 | ####### | 211 | | 0.091405 | [0.086-0.0 | Affymetrix, Illumina [50615308] (imputed) | N |
| 7 | 10:71099109 | 10 | 71094504 | rs17476364 | 21/09/2020 | 3.3E+07 | Chen MH | 1/09/2020 | Cell | www.ncbi.nlm.nih.gov/pubmed/32888493 | Trans-ethnic and Ancestry-Specific Blood-Cell Genetics in 746,667 Individuals from 5 Global Populations. | Mean corpuscular volume | 696,882 Af | NA | 10q22.1 | NR | HK1 | | | ENSG00000156515 | NA | NA | rs17476364-C | rs17476364 | 0 | 17476364 | intron_variant | 0 | 0.102938 | ####### | 210 | | NA | | Affymetrix, Illumina [24246363] (imputed) | N |
| 7 | 10:71099109 | 10 | 71094504 | rs17476364 | 12/11/2021 | 3.4E+07 | Thompson A | 8/06/2021 | Hum Mol Genet | www.ncbi.nlm.nih.gov/pubmed/34104963 | Assessing the impact of alcohol consumption on the genetic contribution to mean corpuscular volume. | Mean corpuscular volume | 362,595 B | NA | 10q22.1 | | HK1 | | | ENSG00000156515 | NA | NA | rs17476364-T | rs17476364 | 0 | 17476364 | intron_variant | 0 | 0.89 | 2.00E-92 | 91.69897 | | 0.36 | [-0.39--0.3 | Affymetrix [NR] (imputed) | N |
| 7 | 10:71099109 | 10 | 71094504 | rs17476364 | 14/12/2021 | 3.5E+07 | Sakaue S | 30/09/2021 | Nat Genet | www.ncbi.nlm.nih.gov/pubmed/34594039 | A cross-population atlas of genetic associations for 220 human phenotypes. | Mean corpuscular volume | 350,473 E | NA | 10q22.1 | | HK1 | | | ENSG00000156515 | NA | NA | rs17476364-C | rs17476364 | 0 | 17476364 | intron_variant | 0 | NR | 1.00E-98 | 98 | | 0.0798 | [0.072-0.0 | Affymetrix, Illumina [20535844] (imputed) | N |
| 7 | 10:71099109 | 10 | 71094504 | rs17476364 | 18/09/2020 | 3.3E+07 | Vuckovic D | 1/09/2020 | Cell | www.ncbi.nlm.nih.gov/pubmed/32888494 | The Polygenic and Monogenic Basis of Blood Traits and Diseases. | Mean reticulocyte volume | 408,112 B | NA | 10q22.1 | HK1 | HK1 | | | ENSG00000156515 | NA | NA | rs17476364-C | rs17476364 | 0 | 17476364 | intron_variant | 0 | 0.108783 | 1.00E-59 | 59 | | 0.058846 | [0.052-0.0 | Affymetrix [93095623] (imputed) | N |
| 7 | 10:71099109 | 10 | 71094504 | rs17476364 | 18/09/2020 | 3.3E+07 | Vuckovic D | 1/09/2020 | Cell | www.ncbi.nlm.nih.gov/pubmed/32888494 | The Polygenic and Monogenic Basis of Blood Traits and Diseases. | Mean spheric corpuscular volume | 408,112 B | NA | 10q22.1 | HK1 | HK1 | | | ENSG00000156515 | NA | NA | rs17476364-C | rs17476364 | 0 | 17476364 | intron_variant | 0 | 0.108696 | ####### | 158.5229 | | 0.097074 | [0.09-0.10 | Affymetrix [93095623] (imputed) | N |
| 7 | 10:71099109 | 10 | 71094504 | rs17476364 | 17/11/2022 | 3.5E+07 | Graham SE | 9/12/2021 | Nature | www.ncbi.nlm.nih.gov/pubmed/34887591 | The power of genetic diversity in genome-wide association studies of lipids. | Non-HDL cholesterol levels | 1,320,016 | NA | 10q22.1 | | HK1 | | | ENSG00000156515 | NA | NA | rs17476364-C | rs17476364 | 0 | 17476364 | intron_variant | 0 | 0.099411 | 2.00E-15 | 14.69897 | | 0.02536 | [0.02-0.03 | NR [52000000] (imputed) | N |
| 7 | 10:71099109 | 10 | 71094504 | rs17476364 | 17/11/2022 | 3.5E+07 | Graham SE | 9/12/2021 | Nature | www.ncbi.nlm.nih.gov/pubmed/34887591 | The power of genetic diversity in genome-wide association studies of lipids. | Non-HDL cholesterol levels | 40,963 Sou | NA | 10q22.1 | | HK1 | | | ENSG00000156515 | NA | NA | rs17476364-? | rs17476364 | 0 | 17476364 | intron_variant | 0 | NR | 1.00E-17 | 17 | | NA | | NR [52000000] (imputed) | N |

| | | | | | | | | | | | | | | | | | | | | | | | | | | | | | | | | | | | | |
|---|---|---|---|---|---|---|---|---|---|---|---|---|---|---|---|---|---|---|---|---|---|---|---|---|---|---|---|---|---|---|---|---|---|---|---|---|
| 7 | 10:71099109 | 10 | 71094504 | rs17476364 | 13/12/2021 | 3.4E+07 | Cade BE | 26/08/2021 | Genome Med | www.ncbi.nlm.nih.gov/pubmed/34446064 | Whole-genome association analyses of sleep-disordered breathing phenotypes in the NHLBI TOPMed program. | Obstructive sleep apnea trait (minimum oxyhemoglobin saturation across sleep episode) | 5,173 Euro | NA | 10q22.1 | | HK1 | | | ENSG00000156515 | NA | NA | rs17476364-C | rs17476364 | 0 | 17476364 | intron_variant | 0 | NR | 2.00E-08 | 7.69897 | | 1.129 | [0.74-1.52 | Affymetrix, Illumina [22105437] (imputed) | N |
| 7 | 10:71099109 | 10 | 71094504 | rs17476364 | 21/09/2020 | 3.3E+07 | Chen MH | 1/09/2020 | Cell | www.ncbi.nlm.nih.gov/pubmed/32888493 | Trans-ethnic and Ancestry-Specific Blood-Cell Genetics in 746,667 Individuals from 5 Global Populations. | Platelet count | 542,827 E | NA | 10q22.1 | NR | HK1 | | | ENSG00000156515 | NA | NA | rs17476364-C | rs17476364 | 0 | 17476364 | intron_variant | 0 | 0.106471 | 5.00E-11 | 10.30103 | | 0.019617 | [0.014-0.0 | Affymetrix, Illumina [50241410] (imputed) | N |
| 7 | 10:71099109 | 10 | 71094504 | rs17476364 | 18/09/2020 | 3.3E+07 | Vuckovic D | 1/09/2020 | Cell | www.ncbi.nlm.nih.gov/pubmed/32888494 | The Polygenic and Monogenic Basis of Blood Traits and Diseases. | Plateletcrit | 408,112 B | NA | 10q22.1 | HK1 | HK1 | | | ENSG00000156515 | NA | NA | rs17476364-C | rs17476364 | 0 | 17476364 | intron_variant | 0 | 0.108814 | 1.00E-09 | 9 | | 0.021964 | [0.015-0.0 | Affymetrix [93095623] (imputed) | N |
| 7 | 10:71099109 | 10 | 71094504 | rs17476364 | 30/08/2017 | 2.8E+07 | Astle WJ | 17/11/2016 | Cell | www.ncbi.nlm.nih.gov/pubmed/27863252 | The Allelic Landscape of Human Blood Cell Trait Variation and Links to Common Complex Disease. | Red blood cell count | 172,952 E | NA | 10q22.1 | HK1 | HK1 | | | ENSG00000156515 | NA | NA | rs17476364-C | rs17476364 | 0 | 17476364 | intron_variant | 0 | 0.1103 | 2.00E-48 | 47.69897 | | 0.083301 | [0.072-0.0 | Affymetrix [~ 2500000] (imputed) | N |
| 7 | 10:71099109 | 10 | 71094504 | rs17476364 | 21/09/2020 | 3.3E+07 | Chen MH | 1/09/2020 | Cell | www.ncbi.nlm.nih.gov/pubmed/32888493 | Trans-ethnic and Ancestry-Specific Blood-Cell Genetics in 746,667 Individuals from 5 Global Populations. | Red blood cell count | 727,624 A | NA | 10q22.1 | NR | HK1 | | | ENSG00000156515 | NA | NA | rs17476364-C | rs17476364 | 0 | 17476364 | intron_variant | 0 | 0.10294 | ####### | 141 | | NA | | Affymetrix, Illumina [24332387] (imputed) | N |
| 7 | 10:71099109 | 10 | 71094504 | rs17476364 | 21/09/2020 | 3.3E+07 | Chen MH | 1/09/2020 | Cell | www.ncbi.nlm.nih.gov/pubmed/32888493 | Trans-ethnic and Ancestry-Specific Blood-Cell Genetics in 746,667 Individuals from 5 Global Populations. | Red blood cell count | 545,203 E | NA | 10q22.1 | NR | HK1 | | | ENSG00000156515 | NA | NA | rs17476364-C | rs17476364 | 0 | 17476364 | intron_variant | 0 | 0.106734 | ####### | 145.0969 | | 0.077124 | [0.071-0.0 | Affymetrix, Illumina [50628390] (imputed) | N |
| 7 | 10:71099109 | 10 | 71094504 | rs17476364 | 8/02/2019 | 3.1E+07 | Kichaev G | 27/12/2018 | Am J Hum Genet | www.ncbi.nlm.nih.gov/pubmed/30595370 | Leveraging Polygenic Functional Enrichment to Improve GWAS Power. | Red blood cell count | approxima | NA | 10q22.1 | | HK1 | | | ENSG00000156515 | NA | NA | rs17476364-? | rs17476364 | 0 | 17476364 | intron_variant | 0 | NR | ####### | 124.301 | | NA | | NR [~ 8900000] (imputed) | N |
| 7 | 10:71099109 | 10 | 71094504 | rs17476364 | 18/09/2020 | 3.3E+07 | Vuckovic D | 1/09/2020 | Cell | www.ncbi.nlm.nih.gov/pubmed/32888494 | The Polygenic and Monogenic Basis of Blood Traits and Diseases. | Red blood cell count | 408,112 B | NA | 10q22.1 | HK1 | HK1 | | | ENSG00000156515 | NA | NA | rs17476364-C | rs17476364 | 0 | 17476364 | intron_variant | 0 | 0.10876 | ####### | 115 | | 0.081053 | [0.074-0.0 | Affymetrix [93095623] (imputed) | N |
| 7 | 10:71099109 | 10 | 71094504 | rs17476364 | 14/12/2021 | 3.5E+07 | Sakaue S | 30/09/2021 | Nat Genet | www.ncbi.nlm.nih.gov/pubmed/34594039 | A cross-population atlas of genetic associations for 220 human phenotypes. | Red blood cell count | 350,475 E | NA | 10q22.1 | | HK1 | | | ENSG00000156515 | NA | NA | rs17476364-C | rs17476364 | 0 | 17476364 | intron_variant | 0 | NR | 2.00E-87 | 86.69897 | | 0.0646 | [0.058-0.0 | Affymetrix, Illumina [20538708] (imputed) | N |
| 7 | 10:71099109 | 10 | 71094504 | rs17476364 | 30/08/2017 | 2.8E+07 | Astle WJ | 17/11/2016 | Cell | www.ncbi.nlm.nih.gov/pubmed/27863252 | The Allelic Landscape of Human Blood Cell Trait Variation and Links to Common Complex Disease. | Red cell distribution width | 171,529 E | NA | 10q22.1 | HK1 | HK1 | | | ENSG00000156515 | NA | NA | rs17476364-C | rs17476364 | 0 | 17476364 | intron_variant | 0 | 0.1103 | 2.00E-11 | 10.69897 | | 0.03798 | [0.027-0.0 | Affymetrix [~ 2500000] (imputed) | N |
| 7 | 10:71099109 | 10 | 71094504 | rs17476364 | 21/09/2020 | 3.3E+07 | Chen MH | 1/09/2020 | Cell | www.ncbi.nlm.nih.gov/pubmed/32888493 | Trans-ethnic and Ancestry-Specific Blood-Cell Genetics in 746,667 Individuals from 5 Global Populations. | Red cell distribution width | 563,352 A | NA | 10q22.1 | NR | HK1 | | | ENSG00000156515 | NA | NA | rs17476364-T | rs17476364 | 0 | 17476364 | intron_variant | 0 | 0.896569 | 1.00E-32 | 32 | | NA | | Affymetrix, Illumina [22063850] (imputed) | N |
| 7 | 10:71099109 | 10 | 71094504 | rs17476364 | 21/09/2020 | 3.3E+07 | Chen MH | 1/09/2020 | Cell | www.ncbi.nlm.nih.gov/pubmed/32888493 | Trans-ethnic and Ancestry-Specific Blood-Cell Genetics in 746,667 Individuals from 5 Global Populations. | Red cell distribution width | 531,774 E | NA | 10q22.1 | NR | HK1 | | | ENSG00000156515 | NA | NA | rs17476364-T | rs17476364 | 0 | 17476364 | intron_variant | 0 | 0.892832 | 6.00E-36 | 35.22185 | | 0.038198 | [0.032-0.0 | Affymetrix, Illumina [49860007] (imputed) | N |
| 7 | 10:71099109 | 10 | 71094504 | rs17476364 | 18/09/2020 | 3.3E+07 | Vuckovic D | 1/09/2020 | Cell | www.ncbi.nlm.nih.gov/pubmed/32888494 | The Polygenic and Monogenic Basis of Blood Traits and Diseases. | Red cell distribution width | 408,112 B | NA | 10q22.1 | HK1 | HK1 | | | ENSG00000156515 | NA | NA | rs17476364-C | rs17476364 | 0 | 17476364 | intron_variant | 0 | 0.108735 | 3.00E-23 | 22.52288 | | 0.035448 | [0.028-0.0 | Affymetrix [93095623] (imputed) | N |
| 7 | 10:71099109 | 10 | 71094504 | rs17476364 | 8/02/2019 | 3.1E+07 | Kichaev G | 27/12/2018 | Am J Hum Genet | www.ncbi.nlm.nih.gov/pubmed/30595370 | Leveraging Polygenic Functional Enrichment to Improve GWAS Power. | Red cell distribution width | approxima | NA | 10q22.1 | | HK1 | | | ENSG00000156515 | NA | NA | rs17476364-? | rs17476364 | 0 | 17476364 | intron_variant | 0 | NR | 2.00E-25 | 24.69897 | | NA | | NR [~ 8900000] (imputed) | N |
| 7 | 10:71099109 | 10 | 71094504 | rs17476364 | 30/08/2017 | 2.8E+07 | Astle WJ | 17/11/2016 | Cell | www.ncbi.nlm.nih.gov/pubmed/27863252 | The Allelic Landscape of Human Blood Cell Trait Variation and Links to Common Complex Disease. | Reticulocyte count | 170,641 E | NA | 10q22.1 | HK1 | HK1 | | | ENSG00000156515 | NA | NA | rs17476364-C | rs17476364 | 0 | 17476364 | intron_variant | 0 | 0.1104 | 2.00E-96 | 95.69897 | | 0.119759 | [0.11-0.13 | Affymetrix [~ 2500000] (imputed) | N |
| 7 | 10:71099109 | 10 | 71094504 | rs17476364 | 18/09/2020 | 3.3E+07 | Vuckovic D | 1/09/2020 | Cell | www.ncbi.nlm.nih.gov/pubmed/32888494 | The Polygenic and Monogenic Basis of Blood Traits and Diseases. | Reticulocyte count | 408,112 B | NA | 10q22.1 | HK1 | HK1 | | | ENSG00000156515 | NA | NA | rs17476364-C | rs17476364 | 0 | 17476364 | intron_variant | 0 | 0.108807 | ####### | 224.0969 | | 0.115151 | [0.11-0.12 | Affymetrix [93095623] (imputed) | N |
| 7 | 10:71099109 | 10 | 71094504 | rs17476364 | 30/08/2017 | 2.8E+07 | Astle WJ | 17/11/2016 | Cell | www.ncbi.nlm.nih.gov/pubmed/27863252 | The Allelic Landscape of Human Blood Cell Trait Variation and Links to Common Complex Disease. | Reticulocyte fraction of red cells | 170,690 E | NA | 10q22.1 | HK1 | HK1 | | | ENSG00000156515 | NA | NA | rs17476364-C | rs17476364 | 0 | 17476364 | intron_variant | 0 | 0.1104 | 3.00E-72 | 71.52288 | | 0.1033 | [0.092-0.1 | Affymetrix [~ 2500000] (imputed) | N |
| 7 | 10:71099109 | 10 | 71094504 | rs17476364 | 18/09/2020 | 3.3E+07 | Vuckovic D | 1/09/2020 | Cell | www.ncbi.nlm.nih.gov/pubmed/32888494 | The Polygenic and Monogenic Basis of Blood Traits and Diseases. | Reticulocyte fraction of red cells | 408,112 B | NA | 10q22.1 | HK1 | HK1 | | | ENSG00000156515 | NA | NA | rs17476364-C | rs17476364 | 0 | 17476364 | intron_variant | 0 | 0.108802 | ####### | 168.1549 | | 0.099627 | [0.093-0.1 | Affymetrix [93095623] (imputed) | N |
| 7 | 10:71099109 | 10 | 71094504 | rs17476364 | 14/12/2021 | 3.5E+07 | Sakaue S | 30/09/2021 | Nat Genet | www.ncbi.nlm.nih.gov/pubmed/34594039 | A cross-population atlas of genetic associations for 220 human phenotypes. | Total bilirubin levels | 342,829 E | NA | 10q22.1 | | HK1 | | | ENSG00000156515 | NA | NA | rs17476364-C | rs17476364 | 0 | 17476364 | intron_variant | 0 | NR | ####### | 111.699 | | 0.0828 | [0.076-0.0 | Affymetrix, Illumina [20535842] (imputed) | N |
| 7 | 10:71099109 | 10 | 71094504 | rs17476364 | 17/11/2022 | 3.5E+07 | Graham SE | 9/12/2021 | Nature | www.ncbi.nlm.nih.gov/pubmed/34887591 | The power of genetic diversity in genome-wide association studies of lipids. | Total cholesterol levels | 40,963 So | NA | 10q22.1 | | HK1 | | | ENSG00000156515 | NA | NA | rs17476364-? | rs17476364 | 0 | 17476364 | intron_variant | 0 | NR | 4.00E-36 | 35.39794 | | NA | | NR [52000000] (imputed) | N |
| 7 | 10:71099109 | 10 | 71094504 | rs17476364 | 17/11/2022 | 3.5E+07 | Graham SE | 9/12/2021 | Nature | www.ncbi.nlm.nih.gov/pubmed/34887591 | The power of genetic diversity in genome-wide association studies of lipids. | Total cholesterol levels | 1,320,016 | NA | 10q22.1 | | HK1 | | | ENSG00000156515 | NA | NA | rs17476364-C | rs17476364 | 0 | 17476364 | intron_variant | 0 | 0.098904 | 2.00E-29 | 28.69897 | | 0.030331 | [0.026-0.0 | NR [52000000] (imputed) | N |
| 7 | 10:71099109 | 10 | 71099109 | rs72805692 | 1/03/2022 | 3.5E+07 | Downie CG | 24/12/2021 | Diabetologia | www.ncbi.nlm.nih.gov/pubmed/34951656 | Multi-ethnic GWAS and fine-mapping of glycaemic traits identify novel loci in the PAGE Study. | Glycated haemoglobin HbA1c levels adjusted for BMI | 23,357 Eur | NA | 10q22.1 | | HK1 | | | ENSG00000156515 | NA | NA | rs72805692-A | rs72805692 | 0 | 72805692 | intron_variant | 0 | 0.083 | 5.00E-44 | 43.30103 | | 0.2647 | [0.23-0.3] | Affymetrix, Illumina [23060038] (imputed) | N |
| 7 | 10:71099109 | 10 | 71099109 | rs72805692 | 1/03/2022 | 3.5E+07 | Downie CG | 24/12/2021 | Diabetologia | www.ncbi.nlm.nih.gov/pubmed/34951656 | Multi-ethnic GWAS and fine-mapping of glycaemic traits identify novel loci in the PAGE Study. | Glycated haemoglobin HbA1c levels adjusted for BMI | 9,228 Euro | NA | 10q22.1 | | HK1 | | | ENSG00000156515 | NA | NA | rs72805692-A | rs72805692 | 0 | 72805692 | intron_variant | 0 | 0.1039 | 3.00E-25 | 24.52288 | | 0.2682 | [0.22-0.32 | Affymetrix, Illumina [23060038] (imputed) | N |
| 7 | 10:71099109 | 10 | 71099109 | rs72805692 | 1/03/2022 | 3.5E+07 | Downie CG | 24/12/2021 | Diabetologia | www.ncbi.nlm.nih.gov/pubmed/34951656 | Multi-ethnic GWAS and fine-mapping of glycaemic traits identify novel loci in the PAGE Study. | Glycated haemoglobin HbA1c levels adjusted for BMI | 10,693 His | NA | 10q22.1 | | HK1 | | | ENSG00000156515 | NA | NA | rs72805692-A | rs72805692 | 0 | 72805692 | intron_variant | 0 | 0.0635 | 3.00E-20 | 19.52288 | | 0.2628 | [0.21-0.32 | Affymetrix, Illumina [23060038] (imputed) | N |
| 7 | 10:71099109 | 10 | 71099109 | rs72805692 | 12/08/2019 | 3.1E+07 | Moon JY | 18/06/2019 | Diabetes Care | www.ncbi.nlm.nih.gov/pubmed/31213470 | A Genome-Wide Association Study Identifies Blood Disorder-Related Variants Influencing Hemoglobin A1c With Implications for Glycemic Status in U.S. Hispanics/Latinos. | Glycated hemoglobin levels | 9,636 Hisp | 4,777 Hisp | 10q22.1 | HK1 | HK1 | | | ENSG00000156515 | NA | NA | rs72805692-G | rs72805692 | 0 | 72805692 | intron_variant | 0 | 0.066 | 6.00E-26 | 25.22185 | | 0.1 | [0.08-0.12 | Illumina [11510031] (imputed) | N |
| 7 | 10:71099109 | 10 | 71099109 | rs72805692 | 10/06/2021 | 3.4E+07 | Chen J | 31/05/2021 | Nat Genet | www.ncbi.nlm.nih.gov/pubmed/34059833 | The trans-ancestral genomic architecture of glycemic traits. | Glycated hemoglobin levels | 15,248 His | NA | 10q22.1 | HK1 | HK1 | | | ENSG00000156515 | NA | NA | rs72805692-A | rs72805692 | 0 | 72805692 | intron_variant | 0 | 0.9293 | 2.00E-22 | 21.69897 | | 0.0859 | [0.069-0.1 | Affymetrix, Illumina [29199641] (imputed) | N |
| 7 | 10:71099109 | 10 | 71099109 | rs72805692 | 21/07/2020 | 3.2E+07 | Richardson TG | 23/03/2020 | PLoS Med | www.ncbi.nlm.nih.gov/pubmed/32203549 | Evaluating the relationship between circulating lipoprotein lipids and apolipoproteins with risk of coronary heart disease: A multivariable Mendelian randomisation analysis. | HDL cholesterol levels | 403,943 E | NA | 10q22.1 | HK1 | HK1 | | | ENSG00000156515 | NA | NA | rs72805692-A | rs72805692 | 0 | 72805692 | intron_variant | 0 | 0.884869 | 9.00E-10 | 9.045757 | | 0.018309 | [0.012-0.0 | Affymetrix [NR] (imputed) | N |
| 7 | 10:71099109 | 10 | 71099109 | rs72805692 | 28/01/2022 | 3.4E+07 | Hu Y | 21/04/2021 | Am J Hum Genet | www.ncbi.nlm.nih.gov/pubmed/33887194 | Whole-genome sequencing association analysis of quantitative red blood cell phenotypes: The NHLBI TOPMed program. | Hematocrit | 33,324 Am | NA | 10q22.1 | | HK1 | | | ENSG00000156515 | NA | NA | rs72805692-G | rs72805692 | 0 | 72805692 | intron_variant | 0 | NR | 5.00E-26 | 25.30103 | | 0.39 | [0.31-0.47 | Illumina [110396782] | N |
| 7 | 10:71099109 | 10 | 71099109 | rs72805692 | 28/01/2022 | 3.4E+07 | Hu Y | 21/04/2021 | Am J Hum Genet | www.ncbi.nlm.nih.gov/pubmed/33887194 | Whole-genome sequencing association analysis of quantitative red blood cell phenotypes: The NHLBI TOPMed program. | Hemoglobin | 33,325 Am | NA | 10q22.1 | | HK1 | | | ENSG00000156515 | NA | NA | rs72805692-G | rs72805692 | 0 | 72805692 | intron_variant | 0 | NR | 3.00E-25 | 24.52288 | | 0.13 | [0.11-0.15 | Illumina [110351653] | N |
| 7 | 10:71099109 | 10 | 71099109 | rs72805692 | 1/07/2019 | 3.1E+07 | Wojcik GL | 19/06/2019 | Nature | www.ncbi.nlm.nih.gov/pubmed/31217584 | Genetic analyses of diverse populations improves discovery for complex traits. | Hemoglobin A1c levels | 559 Africar | NA | 10q22.1 | HK1 | HK1 | | | ENSG00000156515 | NA | NA | rs72805692-? | rs72805692 | 0 | 72805692 | intron_variant | 0 | NR | 1.00E-21 | 21 | | 0.265 | [0.21-0.32 | Illumina [22871999] (imputed) | N |
| 7 | 10:71099109 | 10 | 71099109 | rs72805692 | 1/07/2019 | 3.1E+07 | Wojcik GL | 19/06/2019 | Nature | www.ncbi.nlm.nih.gov/pubmed/31217584 | Genetic analyses of diverse populations improves discovery for complex traits. | Hemoglobin A1c levels | 559 Africar | NA | 10q22.1 | HK1 | HK1 | | | ENSG00000156515 | NA | NA | rs72805692-? | rs72805692 | 0 | 72805692 | intron_variant | 0 | NR | 6.00E-22 | 21.22185 | (Hispanic/ | 0.268372 | [0.21-0.32 | Illumina [22871999] (imputed) | N |
| 7 | 10:71099109 | 10 | 71099109 | rs72805692 | 21/09/2020 | 3.3E+07 | Chen MH | 1/09/2020 | Cell | www.ncbi.nlm.nih.gov/pubmed/32888493 | Trans-ethnic and Ancestry-Specific Blood-Cell Genetics in 746,667 Individuals from 5 Global Populations. | Hemoglobin concentration | 563,946 E | NA | 10q22.1 | NR | HK1 | | | ENSG00000156515 | NA | NA | rs72805692-G | rs72805692 | 0 | 72805692 | intron_variant | 0 | 0.113894 | 2E-482 | 481.699 | | 0.136402 | [0.13-0.14 | Affymetrix, Illumina [50638016] (imputed) | N |
| 7 | 10:71099109 | 10 | 71099109 | rs72805692 | 27/07/2021 | 3.4E+07 | Page GP | 20/05/2021 | J Clin Invest | www.ncbi.nlm.nih.gov/pubmed/34014839 | Multi-ancestry genome-wide association study identifies 27 loci associated with measures of hemolysis following blood storage. | hemolysis of donated blood (osmotic) | Up to 7,58 | NA | 10q22.1 | HK1 | HK1 | | | ENSG00000156515 | NA | NA | rs72805692-? | rs72805692 | 0 | 72805692 | intron_variant | 0 | NR | 5.00E-11 | 10.30103 | | 1.8 | NR unit inc | NR [14100000] (imputed) | N |
| 7 | 10:71099109 | 10 | 71099109 | rs72805692 | 27/07/2021 | 3.4E+07 | Page GP | 20/05/2021 | J Clin Invest | www.ncbi.nlm.nih.gov/pubmed/34014839 | Multi-ancestry genome-wide association study identifies 27 loci associated with measures of hemolysis following blood storage. | hemolysis of donated blood (osmotic) | Up to 7,58 | NA | 10q22.1 | HK1 | HK1 | | | ENSG00000156515 | NA | NA | rs72805692-? | rs72805692 | 0 | 72805692 | intron_variant | 0 | NR | 5.00E-08 | 7.30103 | | NA | | NR [14100000] (imputed) | N |
| 7 | 10:71099109 | 10 | 71099109 | rs72805692 | 30/08/2017 | 2.8E+07 | Astle WJ | 17/11/2016 | Cell | www.ncbi.nlm.nih.gov/pubmed/27863252 | The Allelic Landscape of Human Blood Cell Trait Variation and Links to Common Complex Disease. | Immature fraction of reticulocytes | 170,548 E | NA | 10q22.1 | HK1 | HK1 | | | ENSG00000156515 | NA | NA | rs72805692-G | rs72805692 | 0 | 72805692 | intron_variant | 0 | 0.1169 | 1.00E-11 | 11 | | 0.037438 | [0.027-0.0 | Affymetrix [~ 2500000] (imputed) | N |
| 7 | 10:71099109 | 10 | 71099109 | rs72805692 | 28/01/2022 | 3.4E+07 | Hu Y | 21/04/2021 | Am J Hum Genet | www.ncbi.nlm.nih.gov/pubmed/33887194 | Whole-genome sequencing association analysis of quantitative red blood cell phenotypes: The NHLBI TOPMed program. | Mean corpuscular volume | 22,267 Am | NA | 10q22.1 | | HK1 | | | ENSG00000156515 | NA | NA | rs72805692-G | rs72805692 | 0 | 72805692 | intron_variant | 0 | NR | 3.00E-09 | 8.522879 | | 0.39 | [0.25-0.53 | Illumina [97506867] | N |
| 7 | 10:71099109 | 10 | 71099109 | rs72805692 | 22/02/2022 | 3.5E+07 | Miao Z | 24/08/2021 | HGG Adv | www.ncbi.nlm.nih.gov/pubmed/35047847 | Identification of 90 NAFLD GWAS loci and establishment of NAFLD PRS and causal role of NAFLD in coronary artery disease. | Nonalcoholic fatty liver disease (imputed) | 28,396 Eur | NA | 10q22.1 | | HK1 | | | ENSG00000156515 | NA | NA | rs72805692-? | rs72805692 | 0 | 72805692 | intron_variant | 0 | NR | 3.00E-09 | 8.522879 | | 0.008334 | [0.0056-0. | NR [NR] | N |
| 7 | 10:71099109 | 10 | 71099109 | rs72805692 | 15/03/2022 | 3.5E+07 | Zekavat SM | 8/11/2021 | Circulation | www.ncbi.nlm.nih.gov/pubmed/34743558 | Deep Learning of the Retina Enables Phenome- and Genome-wide Analyses of the Microvasculature. | Retinal vascular fractal density | 38,932 Bri | NA | 10q22.1 | | HK1 | | | ENSG00000156515 | NA | NA | rs72805692-G | rs72805692 | 0 | 72805692 | intron_variant | 0 | 0.11873 | 2.00E-07 | 6.69897 | | 0.054727 | [0.034-0.0 | Affymetrix [15580782] (imputed) | N |
| 8 | 13:50760125 | 13 | 50993083 | rs143280072 | 24/08/2021 | 3.4E+07 | Christakoudi S | 21/05/2021 | Sci Rep | www.ncbi.nlm.nih.gov/pubmed/34021172 | GWAS of allometric body-shape indices in UK Biobank identifies loci suggesting associations with morphogenesis, organogenesis, adrenal cell renewal and cancer. | Hip circumference adjusted for BMI | 219,872 B | NA | 13q14.3 | DLEU1 | DLEU1 | | | ENSG00000176124 | NA | NA | rs143280072-C | rs143280072 | 0 | 1.43E+08 | intron_variant | 0 | NR | 2.00E-12 | 11.69897 | | 0.058605 | [0.042-0.0 | Affymetrix [321016] (imputed) | N |
| 8 | 13:50760125 | 13 | 50993083 | rs143280072 | 24/08/2021 | 3.4E+07 | Christakoudi S | 21/05/2021 | Sci Rep | www.ncbi.nlm.nih.gov/pubmed/34021172 | GWAS of allometric body-shape indices in UK Biobank identifies loci suggesting associations with morphogenesis, organogenesis, adrenal cell renewal and cancer. | Hip circumference adjusted for BMI | 186,825 B | NA | 13q14.3 | DLEU1 | DLEU1 | | | ENSG00000176124 | NA | NA | rs143280072-C | rs143280072 | 0 | 1.43E+08 | intron_variant | 0 | NR | 2.00E-22 | 21.69897 | | 0.088197 | [0.07-0.10 | Affymetrix [323123] (imputed) | N |
| 8 | 13:50760125 | 13 | 50760125 | rs182234180 | 18/09/2020 | 3.3E+07 | Vuckovic D | 1/09/2020 | Cell | www.ncbi.nlm.nih.gov/pubmed/32888494 | The Polygenic and Monogenic Basis of Blood Traits and Diseases. | Lymphocyte count | 408,112 B | NA | 13q14.2 | DLEU1 | DLEU1 | | | ENSG00000176124 | NA | NA | rs182234180-T | rs182234180 | 0 | 1.82E+08 | non_coding_transcript_exon_ | 0 | 0.030056 | 3.00E-18 | 17.52288 | | 0.057718 | [0.045-0.0 | Affymetrix [93095623] (imputed) | N |
| 8 | 13:50760125 | 13 | 50760125 | rs182234180 | 18/09/2020 | 3.3E+07 | Vuckovic D | 1/09/2020 | Cell | www.ncbi.nlm.nih.gov/pubmed/32888494 | The Polygenic and Monogenic Basis of Blood Traits and Diseases. | Lymphocyte percentage of white cells | 408,112 B | NA | 13q14.2 | DLEU1 | DLEU1 | | | ENSG00000176124 | NA | NA | rs182234180-T | rs182234180 | 0 | 1.82E+08 | non_coding_transcript_exon_ | 0 | 0.03006 | 1.00E-09 | 9 | | 0.040107 | [0.027-0.0 | Affymetrix [93095623] (imputed) | N |
| 8 | 13:50760125 | 13 | 50771235 | rs2703087 | 24/08/2021 | 3.4E+07 | Christakoudi S | 21/05/2021 | Sci Rep | www.ncbi.nlm.nih.gov/pubmed/34021172 | GWAS of allometric body-shape indices in UK Biobank identifies loci suggesting associations with morphogenesis, organogenesis, adrenal cell renewal and cancer. | Hip circumference adjusted for BMI | 219,872 B | NA | 13q14.2 | DLEU1 | DLEU1 | | | ENSG00000176124 | NA | NA | rs2703087-A | rs2703087 | 0 | 2703087 | intron_variant | 0 | NR | 1.00E-09 | 9 | | 0.044012 | [0.03-0.05 | Affymetrix [321016] (imputed) | N |
| 8 | 13:50760125 | 13 | 50771235 | rs2703087 | 24/08/2021 | 3.4E+07 | Christakoudi S | 21/05/2021 | Sci Rep | www.ncbi.nlm.nih.gov/pubmed/34021172 | GWAS of allometric body-shape indices in UK Biobank identifies loci suggesting associations with morphogenesis, organogenesis, adrenal cell renewal and cancer. | Hip circumference adjusted for BMI | 186,825 B | NA | 13q14.2 | DLEU1 | DLEU1 | | | ENSG00000176124 | NA | NA | rs2703087-A | rs2703087 | 0 | 2703087 | intron_variant | 0 | NR | 7.00E-22 | 21.1549 | | 0.075745 | [0.06-0.09 | Affymetrix [323123] (imputed) | N |
| 9 | 17:44118848 | 17 | 43932789 | rs10445365 | 31/05/2022 | 3.4E+07 | Smith SM | 19/04/2021 | Nat Neurosci | www.ncbi.nlm.nih.gov/pubmed/33875891 | An expanded set of genome-wide association studies of brain imaging phenotypes in UK Biobank. | IDP dMRI ProbtrackX L2 atr l | 20,860 Bri | 10,495 Bri | 17q21.31 | | MAPT-AS1, MAPT-AS1, MAPT-AS1 | | | ENSG00000264589, E | NA | NA | rs10445365-T | rs10445365 | 0 | 10445365 | intron_variant | 0 | 0.184 | 3.00E-15 | 14.52288 | | 0.103 | [0.078-0.1 | Affymetrix [17103079] (imputed) | N |
| 9 | 17:44118848 | 17 | 43932789 | rs10445365 | 31/05/2022 | 3.4E+07 | Smith SM | 19/04/2021 | Nat Neurosci | www.ncbi.nlm.nih.gov/pubmed/33875891 | An expanded set of genome-wide association studies of brain imaging phenotypes in UK Biobank. | IDP dMRI ProbtrackX MD atr l | 20,860 Bri | 10,495 Bri | 17q21.31 | | MAPT-AS1, MAPT-AS1, MAPT-AS1 | | | ENSG00000264589, E | NA | NA | rs10445365-T | rs10445365 | 0 | 10445365 | intron_variant | 0 | 0.184 | 2.00E-16 | 15.69897 | | 0.106 | [0.081-0.1 | Affymetrix [17103079] (imputed) | N |
| 9 | 17:44118848 | 17 | 43932789 | rs10445365 | 31/05/2022 | 3.4E+07 | Smith SM | 19/04/2021 | Nat Neurosci | www.ncbi.nlm.nih.gov/pubmed/33875891 | An expanded set of genome-wide association studies of brain imaging phenotypes in UK Biobank. | IDP dMRI TBSS ICVF Anterior corona radiata R | 20,859 Bri | 10,496 Bri | 17q21.31 | | MAPT-AS1, MAPT-AS1, MAPT-AS1 | | | ENSG00000264589, E | NA | NA | rs10445365-T | rs10445365 | 0 | 10445365 | intron_variant | 0 | 0.184 | 2.00E-19 | 18.69897 | | 0.117 | [0.092-0.1 | Affymetrix [17103079] (imputed) | N |

| | | | | | | | | | | | | | | | | | | | | | | | | | | | | | | | | | | | | |
|---|---|---|---|---|---|---|---|---|---|---|---|---|---|---|---|---|---|---|---|---|---|---|---|---|---|---|---|---|---|---|---|---|---|---|---|---|
| 9 | 17:44118848 | 17 | 43932789 | rs10445365 | 31/05/2022 | 3.4E+07 | Smith SM | 19/04/2021 | Nat Neurosci | www.ncbi.nlm.nih.gov/pubmed/33875891 | An expanded set of genome-wide association studies of brain imaging phenotypes in UK Biobank. | IDP dMRI TBSS ICVF Anterior limb of internal capsule R | 20,859 Brit | 10,496 Brit | 17q21.31 | | MAPT-AS1, MAPT-AS1, MAPT-AS1 | | | ENSG00000264589, E | NA | NA | rs10445365-T | rs10445365 | 0 | 10445365 | intron_variant | 0 | 0.184 | 2.00E-14 | 13.69897 | | 0.1 | [0.075-0.1 | Affymetrix [17103079] (imputed) | N |
| 9 | 17:44118848 | 17 | 43932789 | rs10445365 | 31/05/2022 | 3.4E+07 | Smith SM | 19/04/2021 | Nat Neurosci | www.ncbi.nlm.nih.gov/pubmed/33875891 | An expanded set of genome-wide association studies of brain imaging phenotypes in UK Biobank. | IDP dMRI TBSS ICVF Retrolenticular part of internal capsule L | 20,859 Brit | 10,496 Brit | 17q21.31 | | MAPT-AS1, MAPT-AS1, MAPT-AS1 | | | ENSG00000264589, E | NA | NA | rs10445365-T | rs10445365 | 0 | 10445365 | intron_variant | 0 | 0.184 | 4.00E-15 | 14.39794 | | 0.102 | [0.077-0.1 | Affymetrix [17103079] (imputed) | N |
| 9 | 17:44118848 | 17 | 43932797 | rs10445366 | 27/07/2021 | 3.4E+07 | Mills MC | 1/07/2021 | Nat Hum Behav | www.ncbi.nlm.nih.gov/pubmed/34211149 | Identification of 371 genetic variants for age at first sex and birth linked to externalising behaviour. | Age at first birth | 542,901 Eu | NA | 17q21.31 | MAPT-AS1 | MAPT-AS1, MAPT-AS1, MAPT-AS1 | | | ENSG00000264589, E | NA | NA | rs10445366-C | rs10445366 | 0 | 10445366 | intron_variant | 0 | 0.1843 | 7.00E-11 | 10.1549 | | 0.0886 | [0.062-0.1 | NR [9744772] (imputed) | N |
| 9 | 17:44118848 | 17 | 43932798 | rs10445367 | 14/12/2021 | 3.5E+07 | Sakaue S | 30/09/2021 | Nat Genet | www.ncbi.nlm.nih.gov/pubmed/34594039 | A cross-population atlas of genetic associations for 220 human phenotypes. | Neutrophil count | 349,856 Eu | NA | 17q21.31 | | MAPT-AS1, MAPT-AS1, MAPT-AS1 | | | ENSG00000264589, E | NA | NA | rs10445367-T | rs10445367 | 0 | 10445367 | intron_variant | 0 | NR | 1.00E-18 | 18 | | 0.0282 | [0.022-0.0 | Affymetrix, Illumina [20526770] (imputed) | N |
| 9 | 17:44118848 | 17 | 43932798 | rs10445367 | 5/02/2020 | 3.2E+07 | Zhao B | 30/10/2019 | Mol Psychiatry | www.ncbi.nlm.nih.gov/pubmed/31666681 | Large-scale GWAS reveals genetic architecture of brain white matter microstructure and genetic overlap with cognitive and mental health traits (nÔÇë=ÔÇë17,706). | White matter microstructure (fractional anisotropy) | 17,706 Eur | NA | 17q21.31 | NR | MAPT-AS1, MAPT-AS1, MAPT-AS1 | | | ENSG00000264589, E | NA | NA | rs10445367-? | rs10445367 | 0 | 10445367 | intron_variant | 0 | | 1.00E-09 | 9 | (Genu of co | NA | | NR [8955960] (imputed) | N |
| 9 | 17:44118848 | 17 | 43933171 | rs10445368 | 23/10/2020 | 3.3E+07 | van der Meer D | 14/07/2020 | Nat Commun | www.ncbi.nlm.nih.gov/pubmed/32665545 | Understanding the genetic determinants of the brain with MOSTest. | Cortical surface area (MOSTest) | 26,502 Eur | NA | 17q21.31 | VCAN | MAPT-AS1, MAPT-AS1, MAPT-AS1 | | | ENSG00000264589, E | NA | NA | rs10445368-? | rs10445368 | 0 | 10445368 | intron_variant | 0 | | 2.00E-38 | 37.69897 | | NA | | NR [7428630] (imputed) | N |
| 9 | 17:44118848 | 17 | 43950976 | rs10491140 | 18/10/2018 | 3E+07 | Kim SK | 26/07/2018 | PLoS One | www.ncbi.nlm.nih.gov/pubmed/30048462 | Identification of 613 new loci associated with heel bone mineral density and a polygenic risk score for bone mineral density, osteoporosis and fracture. | Heel bone mineral density | 394,929 Eu | NA | 17q21.31 | | MAPT-AS1, MAPT-AS1, MAPT-AS1 | | | ENSG00000264589, E | NA | NA | rs10491140-? | rs10491140 | 0 | 10491140 | intron_variant | 0 | NR | 3.00E-26 | 25.52288 | | 0.025714 | [0.021-0.0 | Affymetrix [20259828] (imputed) | N |
| 9 | 17:44118848 | 17 | 44073889 | rs1052553 | 26/04/2018 | 2.6E+07 | Onengut-Gumuscu S | 1/04/2015 | Nat Genet | www.ncbi.nlm.nih.gov/pubmed/25751624 | Fine mapping of type 1 diabetes susceptibility loci and evidence for colocalization of causal variants with lymphoid gene enhancers. | Type 1 diabetes | 6,683 Euro | NA | 17q21.31 | Intergenic | MAPT, MAPT, MAPT | | | ENSG00000277956, E | NA | NA | rs1052553-A | rs1052553 | 0 | 1052553 | synonymous_variant | 0 | 0.76 | 8.00E-08 | 7.09691 | | 1.123596 | | Illumina [135870] | N |
| 9 | 17:44118848 | 17 | 44073889 | rs1052553 | 7/07/2021 | 3.4E+07 | Robertson CC | 14/06/2021 | Nat Genet | www.ncbi.nlm.nih.gov/pubmed/34127860 | Fine-mapping, trans-ancestral and genomic analyses identify causal variants, cells, genes and drug targets for type 1 diabetes. | Type 1 diabetes | 20,065 Eur | NA | 17q21.31 | MAPT | MAPT, MAPT, MAPT | | | ENSG00000277956, E | NA | NA | rs1052553-G | rs1052553 | 0 | 1052553 | synonymous_variant | 0 | 0.232 | 2.00E-15 | 14.69897 | | 0.879 | | Illumina [715631] (imputed) | N |
| 9 | 17:44118848 | 17 | 44073889 | rs1052553 | 7/07/2021 | 3.4E+07 | Robertson CC | 14/06/2021 | Nat Genet | www.ncbi.nlm.nih.gov/pubmed/34127860 | Fine-mapping, trans-ancestral and genomic analyses identify causal variants, cells, genes and drug targets for type 1 diabetes. | Type 1 diabetes | 20,065 Eur | NA | 17q21.31 | MAPT | MAPT, MAPT, MAPT | | | ENSG00000277956, E | NA | NA | rs1052553-G | rs1052553 | 0 | 1052553 | synonymous_variant | 0 | 0.232 | 2.00E-15 | 14.69897 | | 0.879 | | Illumina [715631] (imputed) | N |
| 9 | 17:43564222 | 17 | 43513441 | rs11012 | 5/02/2010 | 2E+07 | Edwards TL | 13/01/2010 | Ann Hum Genet | www.ncbi.nlm.nih.gov/pubmed/20070850 | Genome-wide association study confirms SNPs in SNCA and the MAPT region as common risk factors for Parkinson disease. | Parkinson's disease | 1,752 Euro | NA | 17q21.31 | MAPT, PLEKHM1, | PLEKHM1 | | | ENSG00000277111 | NA | NA | rs11012-T | rs11012 | 0 | 11012 | 3_prime_UTR_variant | 0 | NR | 6.00E-08 | 7.221849 | | 1.43 | [1.27-1.61 | Illumina [495715] (imputed) | N |
| 9 | 17:44118848 | 17 | 43839951 | rs11079718 | 18/09/2020 | 3.3E+07 | Vuckovic D | 1/09/2020 | Cell | www.ncbi.nlm.nih.gov/pubmed/32888494 | The Polygenic and Monogenic Basis of Blood Traits and Diseases. | Reticulocyte count | 408,112 Bi | NA | 17q21.31 | RP11-105N13.4, | LINC02210-CRHR1, LINC02210-CRHR1 | | | ENSG00000282456, E | NA | NA | rs11079718-T | rs11079718 | 0 | 11079718 | intron_variant | 0 | 0.223963 | 5.00E-45 | 44.30103 | | 0.0378 | [0.033-0.0 | Affymetrix [93095623] (imputed) | N |
| 9 | 17:44118848 | 17 | 43857989 | rs111433752 | 13/12/2016 | 2.7E+07 | Smith DJ | 12/04/2016 | Mol Psychiatry | www.ncbi.nlm.nih.gov/pubmed/27067015 | Genome-wide analysis of over 106ÔÇë000 individuals identifies 9 neuroticism-associated loci. | Neuroticism | 91,370 Eur | NA | 17q21.31 | Intergenic | LINC02210-CRHR1, LINC02210-CRHR1 | | | ENSG00000282456, E | NA | NA | rs111433752-T | rs111433752 | 0 | 1.11E+08 | intron_variant | 0 | 0.79 | 9.00E-12 | 11.04576 | | 0.12 | [0.085-0.1 | Affymetrix [7207648] (imputed) | N |
| 9 | 17:44118848 | 17 | 44183403 | rs111676341 | 13/01/2021 | 3.3E+07 | Demange PA | 7/01/2021 | Nat Genet | www.ncbi.nlm.nih.gov/pubmed/33414549 | Investigating the genetic architecture of noncognitive skills using GWAS-by-subtraction. | Cognitive aspects of educational attainment | 257,700 Eu | NA | 17q21.31 | NR | KANSL1, KANSL1, KANSL1 | | | ENSG00000120071, E | NA | NA | rs111676341-A | rs111676341 | 0 | 1.12E+08 | intron_variant | 0 | 0.240557 | 2.00E-11 | 10.69897 | | 0.049242 | [0.035-0.0 | Affymetrix, Illumina, Perlegen [7305956] (imputed) | N |
| 9 | 17:44118848 | 17 | 44019107 | rs111825734 | 31/05/2022 | 3.4E+07 | Smith SM | 19/04/2021 | Nat Neurosci | www.ncbi.nlm.nih.gov/pubmed/33875891 | An expanded set of genome-wide association studies of brain imaging phenotypes in UK Biobank. | IDP dMRI TBSS MD Anterior corona radiata R | 20,860 Brit | 10,496 Brit | 17q21.31 | | MAPT, MAPT, MAPT | | | ENSG00000277956, E | NA | NA | rs111825734-G | rs111825734 | 0 | 1.12E+08 | intron_variant | 0 | 0.232 | 9.00E-15 | 14.04576 | | 0.09 | [0.066-0.1 | Affymetrix [17103079] (imputed) | N |
| 9 | 17:44118848 | 17 | 43666492 | rs112010353 | 12/11/2018 | 3E+07 | Lee JJ | 23/07/2018 | Nat Genet | www.ncbi.nlm.nih.gov/pubmed/30038396 | Gene discovery and polygenic prediction from a genome-wide association study of educational attainment in 1.1 million individuals. | Self-reported math ability | 564,698 Eu | NA | 17q21.31 | Intergenic | DND1P1 - MAPK8 | ENSG00000282614 | ENSG00000263503 | | 2197 | 11743 | rs112010353-C | rs112010353 | 0 | 1.12E+08 | intergenic_variant | 1 | 0.8237 | 2.00E-08 | 7.69897 | | 0.0143 | [0.0092-0. | Affymetrix, Illumina [10000000] (imputed) | N |
| 9 | 17:44118848 | 17 | 43785098 | rs112123127 | 31/05/2022 | 3.4E+07 | Smith SM | 19/04/2021 | Nat Neurosci | www.ncbi.nlm.nih.gov/pubmed/33875891 | An expanded set of genome-wide association studies of brain imaging phenotypes in UK Biobank. | IDP dMRI TBSS MD Anterior limb of internal capsule R | 20,860 Brit | 10,496 Brit | 17q21.31 | | LINC02210-CRHR1, LINC02210-CRHR1 | | | ENSG00000282456, E | NA | NA | rs112123127-T | rs112123127 | 0 | 1.12E+08 | intron_variant | 0 | 0.221 | 8.00E-14 | 13.09691 | | 0.088 | [0.064-0.1 | Affymetrix [17103079] (imputed) | N |
| 9 | 17:43564222 | 17 | 43563304 | rs112275793 | 14/12/2021 | 3.5E+07 | Sakaue S | 30/09/2021 | Nat Genet | www.ncbi.nlm.nih.gov/pubmed/34594039 | A cross-population atlas of genetic associations for 220 human phenotypes. | Aspartate aminotransferase levels | 342,990 Eu | NA | 17q21.31 | | PLEKHM1 | | | ENSG00000277111 | NA | NA | rs112275793-C | rs112275793 | 0 | 1.12E+08 | intron_variant | 0 | NR | 2.00E-19 | 18.69897 | | 0.0277 | [0.022-0.0 | Affymetrix, Illumina [20538469] (imputed) | N |
| 9 | 17:44118848 | 17 | 43669491 | rs112327620 | 14/12/2021 | 3.5E+07 | Sakaue S | 30/09/2021 | Nat Genet | www.ncbi.nlm.nih.gov/pubmed/34594039 | A cross-population atlas of genetic associations for 220 human phenotypes. | Hemoglobin | 350,474 Eu | NA | 17q21.31 | | DND1P1 - MAPK8 | ENSG00000282614 | ENSG00000263503 | | 5196 | 8744 | rs112327620-G | rs112327620 | 0 | 1.12E+08 | regulatory_region_variant | 1 | NR | 3.00E-63 | 62.52288 | | 0.0388 | [0.034-0.0 | Affymetrix, Illumina [20538521] (imputed) | N |
| 9 | 17:44118848 | 17 | 44126673 | rs112333322 | 22/01/2019 | 3E+07 | Nagel M | 2/03/2018 | Nat Commun | www.ncbi.nlm.nih.gov/pubmed/29500382 | Item-level analyses reveal genetic heterogeneity in neuroticism. | Experiencing mood swings | 373,733 Eu | NA | 17q21.31 | NR | KANSL1, KANSL1, KANSL1 | | | ENSG00000120071, E | NA | NA | rs112333322-A | rs112333322 | 0 | 1.12E+08 | intron_variant | 0 | 0.214965 | 4.00E-19 | 18.39794 | | 8.94 | z score dec | NR [10847151] (imputed) | N |
| 9 | 17:44118848 | 17 | 44066172 | rs112385572 | 3/12/2018 | 2.8E+07 | Hagenaars SP | 14/02/2017 | PLoS Genet | www.ncbi.nlm.nih.gov/pubmed/28196072 | Genetic prediction of male pattern baldness. | Male-pattern baldness | 52,874 British ancestr | | 17q21.31 | CRHR1, KANSL1, | MAPT, MAPT, MAPT | | | ENSG00000277956, E | NA | NA | rs112385572-G | rs112385572 | 0 | 1.12E+08 | intron_variant | 0 | NR | 8.00E-29 | 28.09691 | | 0.076825 | NR unit dec | NR [NR] (imputed) | N |
| 9 | 17:44118848 | 17 | 44019103 | rs112568425 | 18/09/2020 | 3.3E+07 | Vuckovic D | 1/09/2020 | Cell | www.ncbi.nlm.nih.gov/pubmed/32888494 | The Polygenic and Monogenic Basis of Blood Traits and Diseases. | Immature fraction of reticulocytes | 408,112 Bi | NA | 17q21.31 | MAPT | MAPT, MAPT, MAPT | | | ENSG00000277956, E | NA | NA | rs112568425-G | rs112568425 | 0 | 1.13E+08 | intron_variant | 0 | 0.279718 | 1.00E-14 | 14 | | 0.020362 | [0.015-0.0 | Affymetrix [93095623] (imputed) | N |
| 9 | 17:44118848 | 17 | 44072984 | rs112572874 | 24/10/2022 | 3.4E+07 | Harrison S | 28/07/2021 | Sci Adv | www.ncbi.nlm.nih.gov/pubmed/34321204 | Testosterone and socioeconomic position: Mendelian randomization in 306,248 men and women in UK Biobank. | Free testosterone levels | 148,248 Bi | NA | 17q21.31 | | MAPT, MAPT, MAPT | | | ENSG00000277956, E | NA | NA | rs112572874-G | rs112572874 | 0 | 1.13E+08 | intron_variant | 0 | 0.23 | 6.00E-09 | 8.221849 | | 0 | [0-0] nmol/ | Affymetrix [NR] (imputed) | N |
| 9 | 17:44118848 | 17 | 44348634 | rs112665297 | 5/02/2020 | 3.2E+07 | Zhao B | 30/10/2019 | Mol Psychiatry | www.ncbi.nlm.nih.gov/pubmed/31666681 | Large-scale GWAS reveals genetic architecture of brain white matter microstructure and genetic overlap with cognitive and mental health traits (nÔÇë=ÔÇë17,706). | White matter microstructure (axial diusivities) | 17,706 Eur | NA | 17q21.31 | NR | MAPK8IP1P1 - AR | ENSG00000282723 | ENSG00000228696 | | 26224 | 3516 | rs112665297-? | rs112665297 | 0 | 1.13E+08 | intergenic_variant | 1 | | 1.00E-11 | 11 | (Inferior fro | NA | | NR [8955960] (imputed) | N |
| 9 | 17:44118848 | 17 | 43695263 | rs112997627 | 24/08/2021 | 3.4E+07 | Simcoe M | 10/03/2021 | Sci Adv | www.ncbi.nlm.nih.gov/pubmed/33692100 | Genome-wide association study in almost 195,000 individuals identifies 50 previously unidentified genetic loci for eye color. | Eye color | 157,485 Eu | 35,501 Eur | 17q21.31 | | RPS26P8 - LINC0 | ENSG00000282238 | ENSG00000204650 | | 8979 | 2431 | rs112997627-T | rs112997627 | 0 | 1.13E+08 | intergenic_variant | 1 | 0.801 | 3.00E-08 | 7.522879 | (EA) | 5.56 | NR z score | Illumina [11532091] (imputed) | N |
| 9 | 17:44118848 | 17 | 43569245 | rs113322852 | 13/01/2021 | 3.3E+07 | Demange PA | 7/01/2021 | Nat Genet | www.ncbi.nlm.nih.gov/pubmed/33414549 | Investigating the genetic architecture of noncognitive skills using GWAS-by-subtraction. | Cognitive aspects of educational attainment | 257,700 Eu | NA | 17q21.31 | NR | PLEKHM1 - LRRC | ENSG00000277111 | ENSG00000282331 | | 1130 | 14862 | rs113322852-T | rs113322852 | 0 | 1.13E+08 | regulatory_region_variant | 1 | 0.198807 | 2.00E-12 | 11.69897 | | 0.055314 | [0.04-0.07 | Affymetrix, Illumina, Perlegen [7305956] (imputed) | N |
| 9 | 17:44118848 | 17 | 43569245 | rs113322852 | 18/03/2019 | 3.1E+07 | Baselmans BML | 14/01/2019 | Nat Genet | www.ncbi.nlm.nih.gov/pubmed/30643256 | Multivariate genome-wide analyses of the well-being spectrum. | Neuroticism | 523,783 Eu | 59,206 Eur | 17q21.31 | NR | PLEKHM1 - LRRC | ENSG00000277111 | ENSG00000282331 | | 1130 | 14862 | rs113322852-A | rs113322852 | 0 | 1.13E+08 | regulatory_region_variant | 1 | 0.190285 | 3.00E-19 | 18.52288 | | 0.021022 | [0.016-0.0 | NR [NR] | N |
| 9 | 17:44118848 | 17 | 43850932 | rs113518470 | 11/05/2017 | 2.7E+07 | Pickrell JK | 16/05/2016 | Nat Genet | www.ncbi.nlm.nih.gov/pubmed/27182965 | Detection and interpretation of shared genetic influences on 42 human traits. | Joint mobility (Beighton score) | 64,143 Eur | NA | 17q21.31 | LRRC37A3, CRHF | LINC02210-CRHR1, LINC02210-CRHR1 | | | ENSG00000282456, E | NA | NA | rs113518470-? | rs113518470 | 0 | 1.14E+08 | intron_variant | 0 | NR | 6.00E-11 | 10.22185 | | 0.09 | [0.116-0.0 | Illumina [12039492] (imputed) | N |
| 9 | 17:44118848 | 17 | 43991272 | rs113568679 | 15/03/2022 | 3.5E+07 | Kulminski AM | 6/11/2021 | Geroscience | www.ncbi.nlm.nih.gov/pubmed/34743297 | Pleiotropic predisposition to Alzheimer's disease and educational attainment: insights from the summary statistics analysis. | Alzheimer's disease or educational attainment (pleiotropy) | (see Lamb | NA | 17q21.31 | | MAPT, MAPT, MAPT | | | ENSG00000277956, E | NA | NA | rs113568679-A | rs113568679 | 1 | 17563986 | intron_variant | 0 | 0.7295 | 7.00E-09 | 8.154902 | | 43.90281 | z score inc | NR [6814003] | N |
| 9 | 17:44091886 | 17 | 44228169 | rs114974956 | 6/12/2019 | 3.2E+07 | Alliey-Rodriguez N | 17/09/2019 | Transl Psychiatry | www.ncbi.nlm.nih.gov/pubmed/31530798 | NRXN1 is associated with enlargement of the temporal horns of the lateral ventricles in psychosis. | Ventral diencephalon volume | 87 Europe | NA | 17q21.31 | KANSL1 | KANSL1, KANSL1, KANSL1 | | | ENSG00000120071, E | NA | NA | rs114974956-? | rs114974956 | 1 | 1918793 | intron_variant | 0 | | 6.00E-06 | 5.221849 | | NA | | Illumina [4322238] (imputed) | N |
| 9 | 17:44118848 | 17 | 44699851 | rs116956554 | 8/10/2020 | 3.3E+07 | Zhou W | 7/08/2020 | Nat Commun | www.ncbi.nlm.nih.gov/pubmed/32769997 | GWAS of thyroid stimulating hormone highlights pleiotropic effects and inverse association with thyroid cancer. | Thyroid stimulating hormone levels | 119,715 Eu | NA | 17q21.31 | NSFP1 | NSF, NSF, NSF | | | ENSG00000278174, E | NA | NA | rs116956554-A | rs116956554 | 0 | 1.17E+08 | intron_variant | 0 | 0.179849 | 3.00E-12 | 11.52288 | | 0.0471 | [0.034-0.0 | Illumina [22400000] (imputed) | N |
| 9 | 17:44118848 | 17 | 44051588 | rs117124984 | 25/06/2019 | 3.1E+07 | Jansen PR | 25/02/2019 | Nat Genet | www.ncbi.nlm.nih.gov/pubmed/30804565 | Genome-wide analysis of insomnia in 1,331,010 individuals identifies new risk loci and functional pathways. | Daytime nap | 386,577 Eu | NA | 17q21.31 | NR | MAPT, MAPT, MAPT | | | ENSG00000277956, E | NA | NA | rs117124984-C | rs117124984 | 0 | 1.17E+08 | 5_prime_UTR_variant | 0 | 0.7853 | 3.00E-13 | 12.52288 | | 1.099626 | [1.07-1.13 | Affymetrix, Illumina [10858887] (imputed) | N |
| 9 | 17:44118848 | 17 | 44051589 | rs118087478 | 28/01/2020 | 3.2E+07 | Zhao B | 1/11/2019 | Nat Genet | www.ncbi.nlm.nih.gov/pubmed/31676860 | Genome-wide association analysis of 19,629 individuals identifies variants influencing regional brain volumes and refines their genetic co-architecture with cognitive and mental health traits. | Brain region volumes | 19,629 Brit | NA | 17q21.31 | NR | MAPT, MAPT, MAPT | | | ENSG00000277956, E | NA | NA | rs118087478-? | rs118087478 | 0 | 1.18E+08 | 5_prime_UTR_variant | 0 | NR | 8.00E-14 | 13.09691 | (total brain | NA | | NR [9227495] (imputed) | N |
| 9 | 17:44118848 | 17 | 44015446 | rs12150229 | 25/06/2019 | 3.1E+07 | Jansen PR | 25/02/2019 | Nat Genet | www.ncbi.nlm.nih.gov/pubmed/30804565 | Genome-wide analysis of insomnia in 1,331,010 individuals identifies new risk loci and functional pathways. | Ease of getting up in the morning | 385,949 Eu | NA | 17q21.31 | NR | MAPT, MAPT, MAPT | | | ENSG00000277956, E | NA | NA | rs12150229-G | rs12150229 | 0 | 12150229 | intron_variant | 0 | 0.2236 | 4.00E-09 | 8.39794 | | 0.0122 | [0.0081-0. | Affymetrix, Illumina [10862568] (imputed) | N |
| 9 | 17:44118848 | 17 | 43826637 | rs12150672 | 11/05/2017 | 2.8E+07 | van Rooij FJA | 22/12/2016 | Am J Hum Genet | www.ncbi.nlm.nih.gov/pubmed/28017375 | Genome-wide Trans-ethnic Meta-analysis Identifies Seven Genetic Loci Influencing Erythrocyte Traits and a Role for RBPMS in Erythropoiesis. | Red blood cell count | up to 40,25 | 16,389 ind | 17q21.31 | NR | LINC02210-CRHR1, LINC02210-CRHR1 | | | ENSG00000282456, E | NA | NA | rs12150672-? | rs12150672 | 0 | 12150672 | intron_variant | 0 | NR | 4.00E-09 | 8.39794 | | NA | | NR [2500000] (imputed) | N |
| 9 | 17:44118848 | 17 | 43923654 | rs12185233 | 4/11/2019 | 3E+07 | Pulit SL | 14/09/2018 | Hum Mol Genet | www.ncbi.nlm.nih.gov/pubmed/30239722 | Meta-analysis of genome-wide association studies for body fat distribution in 694,649 individuals of European ancestry. | Waist-to-hip ratio adjusted for BMI | 379,501 Eu | NA | 17q21.31 | NR | SPPL2C, SPPL2C, SPPL2C, MAPT-AS1, MAPT-AS1, MAPT-AS1 | | | ENSG00000277106, E | NA | NA | rs12185233-C | rs12185233 | 0 | 12185233 | missense_variant | 0 | 0.2133 | 2.00E-14 | 13.69897 | | 0.0159 | [0.012-0.0 | NR [~ 27400000] (imputed) | N |
| 9 | 17:44118848 | 17 | 43923683 | rs12185268 | 8/07/2016 | 2.6E+07 | Kouri N | 16/06/2015 | Nat Commun | www.ncbi.nlm.nih.gov/pubmed/26077951 | Genome-wide association study of corticobasal degeneration identifies risk variants shared with progressive supranuclear palsy. | Corticobasal degeneration | 152 cases, | NA | 17q21.31 | MAPT | MAPT-AS1, MAPT-AS1, MAPT-AS1, SPPL2C, SPPL2C, SPPL2C | | | ENSG00000264589, E | NA | NA | rs12185268-? | rs12185268 | 0 | 12185268 | missense_variant | 0 | NR | 1.00E-09 | 9 | | 3.46 | [2.27-5.28 | Illumina [533898] (imputed) | N |
| 9 | 17:44118848 | 17 | 43923683 | rs12185268 | 20/07/2011 | 2.2E+07 | Do CB | 23/06/2011 | PLoS Genet | www.ncbi.nlm.nih.gov/pubmed/21738487 | Web-based genome-wide association study identifies two novel loci and a substantial genetic component for Parkinson's disease. | Parkinson's disease | 3,426 Euro | NA | 17q21.31 | MAPT | MAPT-AS1, MAPT-AS1, MAPT-AS1, SPPL2C, SPPL2C, SPPL2C | | | ENSG00000264589, E | NA | NA | rs12185268-A | rs12185268 | 0 | 12185268 | missense_variant | 0 | 0.79 | 3.00E-14 | 13.52288 | | 1.3 | [1.22-1.39 | Illumina [522782] | N |
| 9 | 17:44118848 | 17 | 43924073 | rs12373123 | 24/01/2022 | 3.4E+07 | He L | 26/02/2021 | Transl Psychiatry | www.ncbi.nlm.nih.gov/pubmed/33637690 | Exome-wide age-of-onset analysis reveals exonic variants in ERN1 and SPPL2C associated with Alzheimer's disease. | Alzheimer's disease (age of onset) in APOE e4 non-carriers | 7,185 Euro | 5,416 Euro | 17q21.31 | | MAPT-AS1, MAPT-AS1, MAPT-AS1, SPPL2C, SPPL2C, SPPL2C | | | ENSG00000264589, E | NA | NA | rs12373123-? | rs12373123 | 0 | 12373123 | missense_variant | 0 | NR | 7.00E-08 | 7.154902 | | 0.861 | | NR [98334] | N |
| 9 | 17:44118848 | 17 | 43924073 | rs12373123 | 17/06/2020 | 3.2E+07 | Oskarsson GR | 23/04/2020 | Commun Biol | www.ncbi.nlm.nih.gov/pubmed/32327693 | Predicted loss and gain of function mutations in ACO1 are associated with erythropoiesis. | Hemoglobin levels | 684,122 Eu | NA | 17q21.31 | SPPL2C | MAPT-AS1, MAPT-AS1, MAPT-AS1, SPPL2C, SPPL2C, SPPL2C | | | ENSG00000264589, E | NA | NA | rs12373123-C | rs12373123 | 0 | 12373123 | missense_variant | 0 | | 2.00E-61 | 60.69897 | | 0.044 | [NR] unit in | Affymetrix, Illumina [43000000] (imputed) | N |
| 9 | 17:44118848 | 17 | 43924219 | rs12373124 | 15/08/2012 | 2.3E+07 | Li R | 31/05/2012 | PLoS Genet | www.ncbi.nlm.nih.gov/pubmed/22693459 | Six novel susceptibility Loci for early-onset androgenetic alopecia and their unexpected association with common diseases. | Male-pattern baldness | 3,891 Euro | NA | 17q21.31 | intergenic | SPPL2C, SPPL2C, SPPL2C, MAPT-AS1, MAPT-AS1, MAPT-AS1 | | | ENSG00000277106, E | NA | NA | rs12373124-T | rs12373124 | 0 | 12373124 | synonymous_variant | 0 | 0.438 | 5.00E-10 | 9.30103 | | 1.33 | [1.21-1.45 | Affymetrix, Illumina [2391230] (imputed) | N |
| 9 | 17:44118848 | 17 | 43924219 | rs12373124 | 3/12/2018 | 2.8E+07 | Hagenaars SP | 14/02/2017 | PLoS Genet | www.ncbi.nlm.nih.gov/pubmed/28196072 | Genetic prediction of male pattern baldness. | Male-pattern baldness | 52,874 British ancestr | | 17q21.31 | NR | SPPL2C, SPPL2C, SPPL2C, MAPT-AS1, MAPT-AS1, MAPT-AS1 | | | ENSG00000277106, E | NA | NA | rs12373124-C | rs12373124 | 0 | 12373124 | synonymous_variant | 0 | NR | 1.00E-27 | 27 | | 0.08 | NR unit dec | NR [NR] (imputed) | N |
| 9 | 17:44118848 | 17 | 43924200 | rs12373142 | 1/05/2019 | 3.1E+07 | Sakornsakolpat P | 25/02/2019 | Nat Genet | www.ncbi.nlm.nih.gov/pubmed/30804561 | Genetic landscape of chronic obstructive pulmonary disease identifies heterogeneous cell-type and phenotype associations. | Chronic obstructive pulmonary disease | 35,735 cas | NA | 17q21.31 | SPPL2C | MAPT-AS1, MAPT-AS1, MAPT-AS1, SPPL2C, SPPL2C, SPPL2C | | | ENSG00000264589, E | NA | NA | rs12373142-G | rs12373142 | 0 | 12373142 | missense_variant | 0 | 0.22 | 1.00E-09 | 9 | | 1.08 | [1.05-1.10 | Affymetrix, Illumina [6224355] (imputed) | N |
| 9 | 17:44118848 | 17 | 43924200 | rs12373142 | 1/06/2021 | 3.4E+07 | Moll M | 28/04/2021 | Am J Physiol Lung Cell Mol Physiol | www.ncbi.nlm.nih.gov/pubmed/33909500 | A Systematic Analysis of Protein-altering Exonic Variants in Chronic Obstructive Pulmonary Disease. | Chronic obstructive pulmonary disease | up to 31,4 | NA | 17q21.31 | SPPL2C | MAPT-AS1, MAPT-AS1, MAPT-AS1, SPPL2C, SPPL2C, SPPL2C | | | ENSG00000264589, E | NA | NA | rs12373142-G | rs12373142 | 0 | 12373142 | missense_variant | 0 | 0.22 | 2.00E-10 | 9.69897 | | 1.08 | [1.05 -1.1] | Illumina [at least 109036] | N |
| 9 | 17:43924337 | 17 | 43924337 | rs12373168 | 5/09/2022 | 3.6E+07 | Rao S | 28/06/2022 | Neuropsychobiology | www.ncbi.nlm.nih.gov/pubmed/35764056 | Genetic Relationships between Attention-Deficit/Hyperactivity Disorder, Autism Spectrum Disorder, and Intelligence. | Attention deficit hyperactivity disorder or autism spectrum disorder or intelligence (pleiotropy) | 20,183 Eur | NA | 17q21.31 | | MAPT-AS1, MAPT-AS1, MAPT-AS1, SPPL2C, SPPL2C, SPPL2C | | | ENSG00000264589, E | NA | NA | rs12373168-? | rs12373168 | 0 | 12373168 | 3_prime_UTR_variant | 0 | NR | 5.00E-10 | 9.30103 | | NA | | NR [6307410] (imputed) | N |

| | | | | | | | | | | | | | | | | | | | | | | | | | | | | | | | | | | | | |
|---|---|---|---|---|---|---|---|---|---|---|---|---|---|---|---|---|---|---|---|---|---|---|---|---|---|---|---|---|---|---|---|---|---|---|---|---|
| 9 | 17:43520118 | 17 | 43495420 | rs12936645 | 3/08/2020 | 3.2E+07 | Han X | 30/04/2020 | JAMA Ophthalmol | www.ncbi.nlm.nih.gov/pubmed/32352494 | Association of Myopia and Intraocular Pressure With Retinal Detachment in European Descent Participants of the UK Biobank Cohort: A Mendelian Randomization Study. | Spherical equivalent | 95,827 Eur | NA | 17q21.31 | NR | ARHGAP27 | | | ENSG00000159314 | NA | NA | rs12936645-T | rs12936645 | 0 | 12936645 | intron_variant | 0 | | 5.00E-09 | 8.30103 | | 0.09 | [0.051-0.1 | NR [NR] | N |
| 9 | 17:43564222 | 17 | 43507297 | rs12949256 | 17/06/2020 | 3.2E+07 | Oskarsson GR | 23/04/2020 | Commun Biol | www.ncbi.nlm.nih.gov/pubmed/32327693 | Predicted loss and gain of function mutations in ACO1 are associated with erythropoiesis. | Hemoglobin levels | 684,122 Eu | NA | 17q21.31 | ARHGAP27 | ARHGAP27 | | | ENSG00000159314 | NA | NA | rs12949256-T | rs12949256 | 0 | 12949256 | missense_variant | 0 | | 3.00E-51 | 50.52288 | | 0.043 | [NR] unit in | Affymetrix, Illumina [43000000] (imputed) | N |
| 9 | 17:43803189 | 17 | 43803189 | rs1358071 | 31/05/2022 | 3.4E+07 | Smith SM | 19/04/2021 | Nat Neurosci | www.ncbi.nlm.nih.gov/pubmed/33875891 | An expanded set of genome-wide association studies of brain imaging phenotypes in UK Biobank. | IDP dMRI TBSS L1 Superior longitudinal fasciculus R | 20,860 Brit | 10,496 Brit | 17q21.31 | | LINC02210-CRHR1, LINC02210-CRHR1 | | | ENSG00000282456, E | NA | NA | rs1358071-A | rs1358071 | 0 | 1358071 | intron_variant | 0 | 0.731 | 4.00E-16 | 15.39794 | | 0.09 | [0.068-0.1 | Affymetrix [17103079] (imputed) | N |
| 9 | 17:44118848 | 17 | 44787312 | rs1378358 | 20/07/2020 | 3.2E+07 | Shin J | 20/03/2020 | Cereb Cortex | www.ncbi.nlm.nih.gov/pubmed/32198502 | Global and Regional Development of the Human Cerebral Cortex: Molecular Architecture and Occupational Aptitudes. | Cortical surface area (global PC1) | 23,784 Eur | 19,512 Eur | 17q21.31 | NSF | NSF, NSF, NSF | | | ENSG00000278174, E | NA | NA | rs1378358-? | rs1378358 | 0 | 1378358 | intron_variant | 0 | NR | 1.00E-31 | 31 | | 2.0049 | [1.67-2.34 | Affymetrix, Illumina [NR] (imputed) | N |
| 9 | 17:44118848 | 17 | 44787312 | rs1378358 | 22/01/2019 | 3E+07 | Nagel M | 2/03/2018 | Nat Commun | www.ncbi.nlm.nih.gov/pubmed/29500382 | Item-level analyses reveal genetic heterogeneity in neuroticism. | Feeling miserable | 376,097 Eu | NA | 17q21.31 | NR | NSF, NSF, NSF | | | ENSG00000278174, E | NA | NA | rs1378358-T | rs1378358 | 0 | 1378358 | intron_variant | 0 | 0.206287 | 2.00E-11 | 10.69897 | | 6.69 | z score inc | NR [10847151] (imputed) | N |
| 9 | 17:44118848 | 17 | 44787312 | rs1378358 | 22/01/2019 | 3E+07 | Nagel M | 2/03/2018 | Nat Commun | www.ncbi.nlm.nih.gov/pubmed/29500382 | Item-level analyses reveal genetic heterogeneity in neuroticism. | Feeling worry | 372,869 Eu | NA | 17q21.31 | GOSR2 | NSF, NSF, NSF | | | ENSG00000278174, E | NA | NA | rs1378358-T | rs1378358 | 0 | 1378358 | intron_variant | 0 | 0.206287 | 2.00E-08 | 7.69897 | | 5.619 | unit increa | NR [10847151] (imputed) | N |
| 9 | 17:44118848 | 17 | 44787312 | rs1378358 | 5/03/2021 | 3.3E+07 | Tadros R | 25/01/2021 | Nat Genet | www.ncbi.nlm.nih.gov/pubmed/33495596 | Shared genetic pathways contribute to risk of hypertrophic and dilated cardiomyopathies with opposite directions of effect. | Hypertrophic cardiomyopathy | 1,733 Euro | 2,694 Euro | 17q21.31 | NSF | NSF, NSF, NSF | | | ENSG00000278174, E | NA | NA | rs1378358-T | rs1378358 | 0 | 1378358 | intron_variant | 0 | 0.2314 | 7.00E-16 | 15.1549 | | 1.281355 | 1.21-1.36 | Illumina [6530233] (imputed) | N |
| 9 | 17:44118848 | 17 | 44787312 | rs1378358 | 5/03/2021 | 3.3E+07 | Tadros R | 25/01/2021 | Nat Genet | www.ncbi.nlm.nih.gov/pubmed/33495596 | Shared genetic pathways contribute to risk of hypertrophic and dilated cardiomyopathies with opposite directions of effect. | Hypertrophic cardiomyopathy (MTAG) | 1,733 Euro | NA | 17q21.31 | NSF | NSF, NSF, NSF | | | ENSG00000278174, E | NA | NA | rs1378358-T | rs1378358 | 0 | 1378358 | intron_variant | 0 | 0.2314 | 5.00E-13 | 12.30103 | | 1.337631 | 1.22-1.47 | Illumina [6530233] (imputed) | N |
| 9 | 17:44118848 | 17 | 44787312 | rs1378358 | 22/01/2019 | 3E+07 | Nagel M | 2/03/2018 | Nat Commun | www.ncbi.nlm.nih.gov/pubmed/29500382 | Item-level analyses reveal genetic heterogeneity in neuroticism. | Neurociticism | 380,506 Eu | NA | 17q21.31 | NR | NSF, NSF, NSF | | | ENSG00000278174, E | NA | NA | rs1378358-T | rs1378358 | 0 | 1378358 | intron_variant | 0 | 0.206287 | 2.00E-27 | 26.69897 | | 10.86 | z score inc | NR [10847151] (imputed) | N |
| 9 | 17:44118848 | 17 | 44787312 | rs1378358 | 18/03/2019 | 3.1E+07 | Baselmans BML | 14/01/2019 | Nat Genet | www.ncbi.nlm.nih.gov/pubmed/30643256 | Multivariate genome-wide analyses of the well-being spectrum. | Neuroticism | 523,783 Eu | 59,206 Eur | 17q21.31 | NR | NSF, NSF, NSF | | | ENSG00000278174, E | NA | NA | rs1378358-T | rs1378358 | 0 | 1378358 | intron_variant | 0 | 0.217293 | 2.00E-18 | 17.69897 | | 0.019632 | [0.015-0.0 | NR [NR] | N |
| 9 | 17:44118848 | 17 | 44335579 | rs139077859 | 1/02/2022 | 3.5E+07 | Hanscombe KB | 9/11/2021 | Genome Med | www.ncbi.nlm.nih.gov/pubmed/34753499 | The genetic case for cardiorespiratory fitness as a clinical vital sign and the routine prescription of physical activity in healthcare. | Cardiorespiratory fitness (vo2 max) | 70,783 Eur | NA | 17q21.31 | | MAPK8IP1P1 - AR | ENSG00000282723 | ENSG00000228696 | | 13169 | 16571 | rs139077859-G | rs139077859 | 0 | 1.39E+08 | intergenic_variant | 1 | 0.79 | 2.00E-08 | 7.69897 | | 0.033 | [0.021-0.0 | Affymetrix [8918919] (imputed) | N |
| 9 | 17:44118848 | 17 | 44335579 | rs139077859 | 31/05/2022 | 3.4E+07 | Smith SM | 19/04/2021 | Nat Neurosci | www.ncbi.nlm.nih.gov/pubmed/33875891 | An expanded set of genome-wide association studies of brain imaging phenotypes in UK Biobank. | IDP dMRI TBSS L1 Superior longitudinal fasciculus L | 20,860 Brit | 10,496 Brit | 17q21.31 | | MAPK8IP1P1 - AR | ENSG00000282723 | ENSG00000228696 | | 13169 | 16571 | rs139077859-A | rs139077859 | 0 | 1.39E+08 | intergenic_variant | 1 | 0.206 | 5.00E-14 | 13.30103 | | 0.093 | [0.069-0.1 | Affymetrix [17103079] (imputed) | N |
| 9 | 17:44118848 | 17 | 43933790 | rs142955985 | 14/12/2021 | 3.5E+07 | Sakaue S | 30/09/2021 | Nat Genet | www.ncbi.nlm.nih.gov/pubmed/34594039 | A cross-population atlas of genetic associations for 220 human phenotypes. | Eosinophil counts | 349,856 Eu | NA | 17q21.31 | | MAPT-AS1, MAPT-AS1, MAPT-AS1 | | | ENSG00000264589, E | NA | NA | rs142955985-T | rs142955985 | 0 | 1.43E+08 | intron_variant | 0 | NR | 2.00E-24 | 23.69897 | | 0.0218 | [0.018-0.0 | Affymetrix, Illumina [20529668] (imputed) | N |
| 9 | 17:44118848 | 17 | 43685698 | rs143246821 | 1/11/2018 | 3E+07 | Wyss AB | 30/07/2018 | Nat Commun | www.ncbi.nlm.nih.gov/pubmed/30061609 | Multiethnic meta-analysis identifies ancestry-specific and cross-ancestry loci for pulmonary function. | Lung function (FEV1) | 60,552 European ance | | 17q21.31 | LOC644172, CRH | MAPK8IP1P2 - RP | ENSG00000263503 | ENSG00000282238 | | 5992 | 239 | rs143246821-A | rs143246821 | 1 | 444967 | upstream_gene_variant | 1 | 0.79 | 9.00E-10 | 9.045757 | (EA) | 30.58 | [20.8-40.3 | Affymetrix, Illumina [10900000] (imputed) | N |
| 9 | 17:44118848 | 17 | 43685698 | rs143246821 | 1/11/2018 | 3E+07 | Wyss AB | 30/07/2018 | Nat Commun | www.ncbi.nlm.nih.gov/pubmed/30061609 | Multiethnic meta-analysis identifies ancestry-specific and cross-ancestry loci for pulmonary function. | Lung function (FVC) | 60,552 European ance | | 17q21.31 | | MAPK8IP1P2 - RP | ENSG00000263503 | ENSG00000282238 | | 5992 | 239 | rs143246821-? | rs143246821 | 1 | 444967 | upstream_gene_variant | 1 | NR | 7.00E-08 | 7.154902 | (EA) | 30.54 | [19.45-41. | Affymetrix, Illumina [10900000] (imputed) | N |
| 9 | 17:44118848 | 17 | 44132458 | rs143364530 | 15/03/2022 | 3.5E+07 | Kulminski AM | 6/11/2021 | Geroscience | www.ncbi.nlm.nih.gov/pubmed/34743297 | Pleiotropic predisposition to Alzheimer's disease and educational attainment: insights from the summary statistics analysis. | Alzheimer's disease or educational attainment (pleiotropy) | (see Lamb | NA | 17q21.31 | | KANSL1, KANSL1, KANSL1 | | | ENSG00000120071, E | NA | NA | rs143364530-A | rs143364530 | 1 | 62063686 | intron_variant | 0 | 0.278 | 3.00E-08 | 7.522879 | | 40.49891 | z score inc | NR [6814003] | N |
| 9 | 17:44118848 | 17 | 43546057 | rs143982995 | 15/03/2022 | 3.5E+07 | Kulminski AM | 6/11/2021 | Geroscience | www.ncbi.nlm.nih.gov/pubmed/34743297 | Pleiotropic predisposition to Alzheimer's disease and educational attainment: insights from the summary statistics analysis. | Alzheimer's disease or educational attainment (pleiotropy) | (see Lamb | NA | 17q21.31 | | PLEKHM1 | | | ENSG00000277111 | NA | NA | rs143982995-T | rs143982995 | 1 | 62065399 | intron_variant | 0 | 0.222 | 6.00E-07 | 6.221849 | | 34.3468 | z score inc | NR [6814003] | N |
| 9 | 17:43564222 | 17 | 43564222 | rs144733372 | 3/12/2018 | 2.8E+07 | Hagenaars SP | 14/02/2017 | PLoS Genet | www.ncbi.nlm.nih.gov/pubmed/28196072 | Genetic prediction of male pattern baldness. | Male-pattern baldness | 52,874 British ancestry | | 17q21.31 | ACBD4, ARHGAP2 | PLEKHM1 | | | ENSG00000277111 | NA | NA | rs144733372-G | rs144733372 | 1 | 9899111 | intron_variant | 0 | NR | 2.00E-22 | 21.69897 | | 0.076714 | NR unit de | NR [NR] (imputed) | N |
| 9 | 17:43564222 | 17 | 43564222 | rs144733372 | 10/07/2018 | 3E+07 | Amare AT | 6/03/2018 | Front Psychiatry | www.ncbi.nlm.nih.gov/pubmed/29559929 | Association of the Polygenic Scores for Personality Traits and Response to Selective Serotonin Reuptake Inhibitors in Patients with Major Depressive Disorder. | Remission after SSRI treatment in MDD or neuroticism | 865 individ | NA | 17q21.31 | PLEKHM1 | PLEKHM1 | | | ENSG00000277111 | NA | NA | rs144733372-G | rs144733372 | 1 | 9899111 | intron_variant | 0 | NR | 3.00E-11 | 10.52288 | | NA | | NR [NR] (imputed) | N |
| 9 | 17:44118848 | 17 | 43569909 | rs146746174 | 12/11/2018 | 3E+07 | Lee JJ | 23/07/2018 | Nat Genet | www.ncbi.nlm.nih.gov/pubmed/30038396 | Gene discovery and polygenic prediction from a genome-wide association study of educational attainment in 1.1 million individuals. | Cognitive performance | 257,841 Eu | NA | 17q21.31 | Intergenic | PLEKHM1 - LRRC | ENSG00000277111 | ENSG00000282331 | | 1794 | 14198 | rs146746174-T | rs146746174 | 1 | 62065449 | intergenic_variant | 1 | 0.815 | 1.00E-12 | 12 | | 0.0263 | [0.019-0.0 | Affymetrix, Illumina [10100000] (imputed) | N |
| 9 | 17:44118848 | 17 | 43569909 | rs146746174 | 12/11/2018 | 3E+07 | Lee JJ | 23/07/2018 | Nat Genet | www.ncbi.nlm.nih.gov/pubmed/30038396 | Gene discovery and polygenic prediction from a genome-wide association study of educational attainment in 1.1 million individuals. | Cognitive performance (MTAG) | 402,382 Eu | NA | 17q21.31 | Intergenic | PLEKHM1 - LRRC | ENSG00000277111 | ENSG00000282331 | | 1794 | 14198 | rs146746174-T | rs146746174 | 1 | 62065449 | intergenic_variant | 1 | 0.815 | 1.00E-23 | 23 | (MTAG) | 0.0297 | [0.024-0.0 | Affymetrix, Illumina [7100000] (imputed) | N |
| 9 | 17:44118848 | 17 | 43569909 | rs146746174 | 12/11/2018 | 3E+07 | Lee JJ | 23/07/2018 | Nat Genet | www.ncbi.nlm.nih.gov/pubmed/30038396 | Gene discovery and polygenic prediction from a genome-wide association study of educational attainment in 1.1 million individuals. | Self-reported math ability (MTAG) | 670,471 Eu | NA | 17q21.31 | Intergenic | PLEKHM1 - LRRC | ENSG00000277111 | ENSG00000282331 | | 1794 | 14198 | rs146746174-T | rs146746174 | 1 | 62065449 | intergenic_variant | 1 | 0.8153 | 3.00E-15 | 14.52288 | (MTAG) | 0.0182 | [0.014-0.0 | Affymetrix, Illumina [7100000] (imputed) | N |
| 9 | 17:44118848 | 17 | 44335635 | rs147431626 | 5/02/2020 | 3.2E+07 | Zhao B | 30/10/2019 | Mol Psychiatry | www.ncbi.nlm.nih.gov/pubmed/31666681 | Large-scale GWAS reveals genetic architecture of brain white matter microstructure and genetic overlap with cognitive and mental health traits (nÔÇë=ÔÇë17,706). | White matter microstructure (radial diusivities) | 17,706 Eur | NA | 17q21.31 | NR | MAPK8IP1P1 - AR | ENSG00000282723 | ENSG00000228696 | | 13225 | 16515 | rs147431626-? | rs147431626 | 0 | 1.47E+08 | intergenic_variant | 1 | | 4.00E-13 | 12.39794 | (Anterior li | NA | | NR [8955960] (imputed) | N |
| 9 | 17:44118848 | 17 | 43549608 | rs149366495 | 4/05/2021 | 3.3E+07 | Hofer E | 22/09/2020 | Nat Commun | www.ncbi.nlm.nih.gov/pubmed/32963231 | Genetic correlations and genome-wide associations of cortical structure in general population samples of 22,824 adults. | Cortical volume | 22,824 Eur | NA | 17q21.31 | PLEKHM1 | PLEKHM1 | | | ENSG00000277111 | NA | NA | rs149366495-? | rs149366495 | 0 | 1.49E+08 | intron_variant | 0 | | 8.00E-13 | 12.09691 | (Global) | NA | | Affymetrix, Illumina [NR] (imputed) | N |
| 9 | 17:44118848 | 17 | 43687072 | rs150180355 | 15/03/2022 | 3.5E+07 | Kulminski AM | 6/11/2021 | Geroscience | www.ncbi.nlm.nih.gov/pubmed/34743297 | Pleiotropic predisposition to Alzheimer's disease and educational attainment: insights from the summary statistics analysis. | Alzheimer's disease or educational attainment (pleiotropy) | (see Lamb | NA | 17q21.31 | | RPS26P8 - LINC0 | ENSG00000282238 | ENSG00000204650 | | 788 | 10622 | rs150180355-A | rs150180355 | 1 | 2949997 | downstream_gene_variant | 1 | 0.2519 | 4.00E-07 | 6.39794 | | 35.50565 | z score inc | NR [6814003] | N |
| 9 | 17:44874453 | 17 | 44874453 | rs1563304 | 17/10/2018 | 3E+07 | Nielsen JB | 30/07/2018 | Nat Genet | www.ncbi.nlm.nih.gov/pubmed/30061737 | Biobank-driven genomic discovery yields new insight into atrial fibrillation biology. | Atrial fibrillation | 60,620 Eur | NA | 17q21.31 | WNT3 | WNT3 | | | ENSG00000108379 | NA | NA | rs1563304-T | rs1563304 | 0 | 1563304 | intron_variant | 0 | 0.178 | 3.00E-12 | 11.52288 | | 1.07 | [1.05-1.09 | Illumina [34740186] (imputed) | N |
| 9 | 17:44874453 | 17 | 44874453 | rs1563304 | 22/01/2019 | 3E+07 | Nagel M | 2/03/2018 | Nat Commun | www.ncbi.nlm.nih.gov/pubmed/29500382 | Item-level analyses reveal genetic heterogeneity in neuroticism. | Experiencing mood swings | 373,733 Eu | NA | 17q21.31 | MAPT | WNT3 | | | ENSG00000108379 | NA | NA | rs1563304-T | rs1563304 | 0 | 1563304 | intron_variant | 0 | 0.172485 | 2.00E-10 | 9.69897 | | 6.37 | z score inc | NR [10847151] (imputed) | N |
| 9 | 17:44874453 | 17 | 44874453 | rs1563304 | 22/01/2019 | 3E+07 | Nagel M | 2/03/2018 | Nat Commun | www.ncbi.nlm.nih.gov/pubmed/29500382 | Item-level analyses reveal genetic heterogeneity in neuroticism. | Feeling hurt | 372,047 Eu | NA | 17q21.31 | MAPT | WNT3 | | | ENSG00000108379 | NA | NA | rs1563304-T | rs1563304 | 0 | 1563304 | intron_variant | 0 | 0.172485 | 1.00E-11 | 11 | | 6.76 | z score inc | NR [10847151] (imputed) | N |
| 9 | 17:44874453 | 17 | 44874453 | rs1563304 | 31/10/2018 | 3E+07 | Nagel M | 25/06/2018 | Nat Genet | www.ncbi.nlm.nih.gov/pubmed/29942085 | Meta-analysis of genome-wide association studies for neuroticism in 449,484 individuals identifies novel genetic loci and pathways. | Neuroticism | 449,484 Eu | NA | 17q21.31 | NR | WNT3 | | | ENSG00000108379 | NA | NA | rs1563304-T | rs1563304 | 0 | 1563304 | intron_variant | 0 | NR | 5.00E-12 | 11.30103 | | 6.9 | z score inc | Affymetrix [10847151] (imputed) | N |
| 9 | 17:44874453 | 17 | 44874453 | rs1563304 | 1/02/2018 | 2.9E+07 | Luciano M | 18/12/2017 | Nat Genet | www.ncbi.nlm.nih.gov/pubmed/29255261 | Association analysis in over 329,000 individuals identifies 116 independent variants influencing neuroticism. | Neuroticism | 329,821 Eu | 122,867 Eu | 17q21.31 | intergenic | WNT3 | | | ENSG00000108379 | NA | NA | rs1563304-T | rs1563304 | 0 | 1563304 | intron_variant | 0 | 0.17248 | 3.00E-10 | 9.522879 | | 6.281 | unit increa | Illumina [18485882] (imputed) | N |
| 9 | 17:44874453 | 17 | 44874453 | rs1563304 | 25/06/2019 | 3.1E+07 | Jansen PR | 25/02/2019 | Nat Genet | www.ncbi.nlm.nih.gov/pubmed/30804565 | Genome-wide analysis of insomnia in 1,331,010 individuals identifies new risk loci and functional pathways. | Snoring | 359,916 Eu | NA | 17q21.31 | NR | WNT3 | | | ENSG00000108379 | NA | NA | rs1563304-T | rs1563304 | 0 | 1563304 | intron_variant | 0 | 0.1782 | 4.00E-08 | 7.39794 | | 1.04 | [1.03-1.05 | Affymetrix, Illumina [10862568] (imputed) | N |
| 9 | 17:44118848 | 17 | 43675489 | rs1614886 | 31/05/2022 | 3.4E+07 | Smith SM | 19/04/2021 | Nat Neurosci | www.ncbi.nlm.nih.gov/pubmed/33875891 | An expanded set of genome-wide association studies of brain imaging phenotypes in UK Biobank. | IDP dMRI TBSS MD Anterior limb of internal capsule L | 20,860 Brit | 10,496 Brit | 17q21.31 | | DND1P1 - MAPK8 | ENSG00000282614 | ENSG00000263503 | | 11194 | 2746 | rs1614886-T | rs1614886 | 0 | 1614886 | regulatory_region_variant | 1 | 0.258 | 3.00E-18 | 17.52288 | | 0.1 | [0.076-0.1 | Affymetrix [17103079] (imputed) | N |
| 9 | 17:43750921 | 17 | 43756969 | rs1635288 | 31/05/2022 | 3.4E+07 | Smith SM | 19/04/2021 | Nat Neurosci | www.ncbi.nlm.nih.gov/pubmed/33875891 | An expanded set of genome-wide association studies of brain imaging phenotypes in UK Biobank. | aparc-a2009s lh area S-collat-transv-ant | 21,282 Brit | 10,686 Brit | 17q21.31 | | LINC02210-CRHR1, LINC02210-CRHR1 | | | ENSG00000282456, E | NA | NA | rs1635288-G | rs1635288 | 0 | 1635288 | intron_variant | 0 | 0.754 | 7.00E-11 | 10.1549 | | 0.073 | [0.051-0.0 | Affymetrix [17103079] (imputed) | N |
| 9 | 17:43750921 | 17 | 43751913 | rs1635291 | 30/03/2020 | 3.2E+07 | Thompson A | 15/01/2020 | Sci Adv | www.ncbi.nlm.nih.gov/pubmed/31998841 | Functional validity, role, and implications of heavy alcohol consumption genetic loci. | Alcohol consumption (heavy vs. light/non-drinkers) | 21,967 Brit | 47,967 Eur | 17q21.31 | CRHR1 | LINC02210-CRHR1, LINC02210-CRHR1 | | | ENSG00000282456, E | NA | NA | rs1635291-A | rs1635291 | 0 | 1635291 | intron_variant | 0 | 0.754 | 7.00E-09 | 8.154902 | | 5.803 | NR z score | Affymetrix [11141077] (imputed) | N |
| 9 | 17:43750921 | 17 | 43744344 | rs1635298 | 1/06/2021 | 3.3E+07 | Bigdeli TB | 10/11/2020 | Schizophr Bull | www.ncbi.nlm.nih.gov/pubmed/33169155 | Genome-Wide Association Studies of Schizophrenia and Bipolar Disorder in a Diverse Cohort of US Veterans. | Schizophrenia | 37,759 Eur | NA | 17q21.31 | CRHR1 | LINC02210-CRHR1, LINC02210-CRHR1 | | | ENSG00000282456, E | NA | NA | rs1635298-A | rs1635298 | 0 | 1635298 | intron_variant | 0 | NR | 3.00E-08 | 7.522879 | | 1.07 | [1.05-1.09 | Affymetrix [up to 49134253] (imputed) | N |
| 9 | 17:44118848 | 17 | 44790203 | rs169201 | 4/05/2021 | 3.3E+07 | Hofer E | 22/09/2020 | Nat Commun | www.ncbi.nlm.nih.gov/pubmed/32963231 | Genetic correlations and genome-wide associations of cortical structure in general population samples of 22,824 adults. | Cortical volume | 22,824 Eur | NA | 17q21.31 | NSF | NSF, NSF, NSF | | | ENSG00000278174, E | NA | NA | rs169201-? | rs169201 | 0 | 169201 | intron_variant | 0 | | 2.00E-13 | 12.69897 | (Global) | NA | | Affymetrix, Illumina [NR] (imputed) | N |
| 9 | 17:44118848 | 17 | 43908152 | rs16940672 | 29/01/2020 | 3.2E+07 | Hill WD | 16/12/2019 | Nat Commun | www.ncbi.nlm.nih.gov/pubmed/31844048 | Genome-wide analysis identifies molecular systems and 149 genetic loci associated with income. | Household income (MTAG) | 505,541 Bi | NA | 17q21.31 | CRHR1 | MAPT-AS1, MAPT-AS1, MAPT-AS1, LINC02210-CRHR1, LINC | | | ENSG00000264589, E | NA | NA | rs16940672-T | rs16940672 | 0 | 16940672 | non_coding_transcript_exon | 0 | 0.218 | 2.00E-16 | 15.69897 | | 0.020259 | [0.015-0.0 | Affymetrix [up to 18485882] (imputed) | N |
| 9 | 17:44091886 | 17 | 44186063 | rs16940904 | 31/08/2022 | 3.6E+07 | Borges MC | 13/06/2022 | BMC Med | www.ncbi.nlm.nih.gov/pubmed/35692035 | Role of circulating polyunsaturated fatty acids on cardiovascular diseases risk: analysis using Mendelian randomization and fatty acid genetic association data from over 114,000 UK Biobank participants. | Circulating docosahexaenoic acid levels | 114,999 Eu | NA | 17q21.31 | | KANSL1, KANSL1, KANSL1 | | | ENSG00000120071, E | NA | NA | rs16940904-C | rs16940904 | 0 | 16940904 | intron_variant | 0 | 0.77 | 2.00E-16 | 15.69897 | | 0.04 | [0.031-0.0 | NR [12321876] (imputed) | N |
| 9 | 17:44091886 | 17 | 44186063 | rs16940904 | 27/04/2022 | 3.5E+07 | Richardson TG | 25/02/2022 | PLoS Biol | www.ncbi.nlm.nih.gov/pubmed/35213538 | Characterising metabolomic signatures of lipid-modifying therapies through drug target mendelian randomisation. | Docosahexaenoic acid levels | 115,006 Eu | NA | 17q21.31 | | KANSL1, KANSL1, KANSL1 | | | ENSG00000120071, E | NA | NA | rs16940904-C | rs16940904 | 0 | 16940904 | intron_variant | 0 | 0.773434 | 1.00E-16 | 16 | | 0.038987 | [0.03-0.04 | NR [11722792] (imputed) | N |
| 9 | 17:44091886 | 17 | 44186063 | rs16940904 | 27/04/2022 | 3.5E+07 | Richardson TG | 25/02/2022 | PLoS Biol | www.ncbi.nlm.nih.gov/pubmed/35213538 | Characterising metabolomic signatures of lipid-modifying therapies through drug target mendelian randomisation. | Omega-3 fatty acid levels | 115,006 Eu | NA | 17q21.31 | | KANSL1, KANSL1, KANSL1 | | | ENSG00000120071, E | NA | NA | rs16940904-C | rs16940904 | 0 | 16940904 | intron_variant | 0 | 0.773434 | 4.00E-13 | 12.39794 | | 0.034508 | [0.025-0.0 | NR [11722792] (imputed) | N |
| 9 | 17:44091886 | 17 | 44186063 | rs16940904 | 27/04/2022 | 3.5E+07 | Richardson TG | 25/02/2022 | PLoS Biol | www.ncbi.nlm.nih.gov/pubmed/35213538 | Characterising metabolomic signatures of lipid-modifying therapies through drug target mendelian randomisation. | Ratio of docosahexaenoic acid to total fatty acid levels | 115,006 Eu | NA | 17q21.31 | | KANSL1, KANSL1, KANSL1 | | | ENSG00000120071, E | NA | NA | rs16940904-C | rs16940904 | 0 | 16940904 | intron_variant | 0 | 0.773434 | 4.00E-16 | 15.39794 | | 0.038921 | [0.03-0.04 | NR [11722792] (imputed) | N |
| 9 | 17:44091886 | 17 | 44186063 | rs16940904 | 27/04/2022 | 3.5E+07 | Richardson TG | 25/02/2022 | PLoS Biol | www.ncbi.nlm.nih.gov/pubmed/35213538 | Characterising metabolomic signatures of lipid-modifying therapies through drug target mendelian randomisation. | Ratio of omega-3 fatty acids to total fatty acids | 115,006 Eu | NA | 17q21.31 | | KANSL1, KANSL1, KANSL1 | | | ENSG00000120071, E | NA | NA | rs16940904-C | rs16940904 | 0 | 16940904 | intron_variant | 0 | 0.773434 | 5.00E-18 | 17.30103 | | 0.041183 | [0.032-0.0 | NR [11722792] (imputed) | N |
| 9 | 17:44091886 | 17 | 44186063 | rs16940904 | 31/08/2022 | 3.6E+07 | Borges MC | 13/06/2022 | BMC Med | www.ncbi.nlm.nih.gov/pubmed/35692035 | Role of circulating polyunsaturated fatty acids on cardiovascular diseases risk: analysis using Mendelian randomization and fatty acid genetic association data from over 114,000 UK Biobank participants. | Total omega-3 fatty acid levels | 114,999 Eu | NA | 17q21.31 | | KANSL1, KANSL1, KANSL1 | | | ENSG00000120071, E | NA | NA | rs16940904-C | rs16940904 | 0 | 16940904 | intron_variant | 0 | 0.77 | 3.00E-13 | 12.52288 | | 0.036 | [0.026-0.0 | NR [12321876] (imputed) | N |

| | | | | | | | | | | | | | | | | | | | | | | | | | | | | | | | | | | | | |
|---|---|---|---|---|---|---|---|---|---|---|---|---|---|---|---|---|---|---|---|---|---|---|---|---|---|---|---|---|---|---|---|---|---|---|---|---|
| 9 | 17:44118848 | 17 | 43672118 | rs1724417 | 4/08/2022 | 3.6E+07 | May-Wilson S | 18/05/2022 | Nat Commun | www.ncbi.nlm.nih.gov/pubmed/35585065 | Large-scale GWAS of food liking reveals genetic determinants and genetic correlations with distinct neurophysiological traits. | Baked/steamed fish liking | 158,335 Eu | NA | 17q21.31 | | DND1P1 - MAPK8 | ENSG00000282614 | ENSG00000263503 | | 7823 | 6117 | rs1724417-G | rs1724417 | 0 | 1724417 | downstream_gene_variant | 1 | 0.1726 | 2.00E-08 | 7.69897 | | 0.02836 | [0.018-0.0 | Affymetrix [11353280] (imputed) | N |
| 9 | 17:44118848 | 17 | 43672118 | rs1724417 | 4/08/2022 | 3.6E+07 | May-Wilson S | 18/05/2022 | Nat Commun | www.ncbi.nlm.nih.gov/pubmed/35585065 | Large-scale GWAS of food liking reveals genetic determinants and genetic correlations with distinct neurophysiological traits. | F-fish liking (derived food-liking factor) | 155,344 Eu | NA | 17q21.31 | | DND1P1 - MAPK8 | ENSG00000282614 | ENSG00000263503 | | 7823 | 6117 | rs1724417-G | rs1724417 | 0 | 1724417 | downstream_gene_variant | 1 | 0.1727 | 9.00E-14 | 13.04576 | | 0.10025 | [0.074-0.1 | Affymetrix [9578548] (imputed) | N |
| 9 | 17:44118848 | 17 | 43672118 | rs1724417 | 4/08/2022 | 3.6E+07 | May-Wilson S | 18/05/2022 | Nat Commun | www.ncbi.nlm.nih.gov/pubmed/35585065 | Large-scale GWAS of food liking reveals genetic determinants and genetic correlations with distinct neurophysiological traits. | F-oily fish liking (derived food-liking factor) | 158,547 Eu | NA | 17q21.31 | | DND1P1 - MAPK8 | ENSG00000282614 | ENSG00000263503 | | 7823 | 6117 | rs1724417-G | rs1724417 | 0 | 1724417 | downstream_gene_variant | 1 | 0.1727 | 5.00E-09 | 8.30103 | | 0.10055 | [0.067-0.1 | Affymetrix [9609308] (imputed) | N |
| 9 | 17:44118848 | 17 | 43672118 | rs1724417 | 4/08/2022 | 3.6E+07 | May-Wilson S | 18/05/2022 | Nat Commun | www.ncbi.nlm.nih.gov/pubmed/35585065 | Large-scale GWAS of food liking reveals genetic determinants and genetic correlations with distinct neurophysiological traits. | F-seafood liking (derived food-liking factor) | 156,740 Eu | NA | 17q21.31 | | DND1P1 - MAPK8 | ENSG00000282614 | ENSG00000263503 | | 7823 | 6117 | rs1724417-G | rs1724417 | 0 | 1724417 | downstream_gene_variant | 1 | 0.1727 | 2.00E-11 | 10.69897 | | 0.08833 | [0.063-0.1 | Affymetrix [9618964] (imputed) | N |
| 9 | 17:44118848 | 17 | 43662020 | rs1724429 | 14/09/2020 | 3.2E+07 | Zhou H | 25/05/2020 | Nat Neurosci | www.ncbi.nlm.nih.gov/pubmed/32451486 | Genome-wide meta-analysis of problematic alcohol use in 435,563 individuals yields insights into biology and relationships with other traits. | Problematic alcohol use (MTAG) | 972,915 Eu | NA | 17q21.31 | NR | RDM1P1 - DND1P | ENSG00000266504 | ENSG00000282614 | | 25584 | 1217 | rs1724429-G | rs1724429 | 0 | 1724429 | intron_variant | 1 | 0.2 | 7.00E-16 | 15.1549 | | 0.024308 | [0.018-0.0 | Affymetrix [10613246] (imputed) | N |
| 9 | 17:44118848 | 17 | 43656176 | rs1724438 | 21/09/2020 | 3.3E+07 | Chen MH | 1/09/2020 | Cell | www.ncbi.nlm.nih.gov/pubmed/32888493 | Trans-ethnic and Ancestry-Specific Blood-Cell Genetics in 746,667 Individuals from 5 Global Populations. | Eosinophil counts | 474,237 Eu | NA | 17q21.31 | NR | RDM1P1 - DND1P | ENSG00000266504 | ENSG00000282614 | | 19740 | 7061 | rs1724438-A | rs1724438 | 0 | 1724438 | intron_variant | 1 | 0.206005 | 6.00E-44 | 43.22185 | | 0.035496 | [0.03-0.04 | Affymetrix, Illumina [50000174] (imputed) | N |
| 9 | 17:44118848 | 17 | 43656176 | rs1724438 | 21/09/2020 | 3.3E+07 | Chen MH | 1/09/2020 | Cell | www.ncbi.nlm.nih.gov/pubmed/32888493 | Trans-ethnic and Ancestry-Specific Blood-Cell Genetics in 746,667 Individuals from 5 Global Populations. | Eosinophil counts | 583,850 Af | NA | 17q21.31 | NR | RDM1P1 - DND1P | ENSG00000266504 | ENSG00000282614 | | 19740 | 7061 | rs1724438-A | rs1724438 | 0 | 1724438 | intron_variant | 1 | 0.198341 | 4.00E-41 | 40.39794 | | NA | | Affymetrix, Illumina [19872809] (imputed) | N |
| 9 | 17:44118848 | 17 | 43656176 | rs1724438 | 31/05/2022 | 3.4E+07 | Smith SM | 19/04/2021 | Nat Neurosci | www.ncbi.nlm.nih.gov/pubmed/33875891 | An expanded set of genome-wide association studies of brain imaging phenotypes in UK Biobank. | IDP dMRI TBSS MD Cingulum cingulate gyrus R | 20,860 Brit | 10,496 Brit | 17q21.31 | | RDM1P1 - DND1P | ENSG00000266504 | ENSG00000282614 | | 19740 | 7061 | rs1724438-A | rs1724438 | 0 | 1724438 | intron_variant | 1 | 0.205 | 4.00E-11 | 10.39794 | | 0.081 | [0.057-0.1 | Affymetrix [17103079] (imputed) | N |
| 9 | 17:44118848 | 17 | 43830938 | rs17426174 | 17/11/2022 | 3.6E+07 | Baselmans B | 6/10/2021 | Biol Psychiatry Glob Open Sci | www.ncbi.nlm.nih.gov/pubmed/36324656 | The Genetic and Neural Substrates of Externalizing Behavior. | Disruptive behavior (multivariate analysis) | 523,150 Eu | NA | 17q21.31 | | LINC02210-CRHR1, LINC02210-CRHR1 | | | ENSG00000282456, E | NA | NA | rs17426174-? | rs17426174 | 0 | 17426174 | intron_variant | 0 | NR | 1.00E-13 | 13 | | NA | | Affymetrix [NR] | N |
| 9 | 17:44118848 | 17 | 43830938 | rs17426174 | 23/02/2018 | 2.9E+07 | Hill WD | 11/01/2018 | Mol Psychiatry | www.ncbi.nlm.nih.gov/pubmed/29326435 | A combined analysis of genetically correlated traits identifies 187 loci and a role for neurogenesis and myelination in intelligence. | Intelligence (MTAG) | 120,934 Br | NA | 17q21.31 | RP11-105N13.4 | LINC02210-CRHR1, LINC02210-CRHR1 | | | ENSG00000282456, E | NA | NA | rs17426174-C | rs17426174 | 0 | 17426174 | intron_variant | 0 | NR | 6.00E-11 | 10.22185 | | 0.023149 | [0.016-0.0 | Affymetrix [NR] | N |
| 9 | 17:44118848 | 17 | 43811072 | rs17563683 | 30/08/2017 | 2.8E+07 | Astle WJ | 17/11/2016 | Cell | www.ncbi.nlm.nih.gov/pubmed/27863252 | The Allelic Landscape of Human Blood Cell Trait Variation and Links to Common Complex Disease. | Hematocrit | 173,039 Eu | NA | 17q21.31 | CRHR1, RP11-10 | LINC02210-CRHR1, LINC02210-CRHR1 | | | ENSG00000282456, E | NA | NA | rs17563683-G | rs17563683 | 0 | 17563683 | intron_variant | 0 | 0.2333 | 2.00E-28 | 27.69897 | | 0.045156 | [0.037-0.0 | Affymetrix [~ 2500000] (imputed) | N |
| 9 | 17:44118848 | 17 | 43811072 | rs17563683 | 30/08/2017 | 2.8E+07 | Astle WJ | 17/11/2016 | Cell | www.ncbi.nlm.nih.gov/pubmed/27863252 | The Allelic Landscape of Human Blood Cell Trait Variation and Links to Common Complex Disease. | Hemoglobin concentration | 172,925 Eu | NA | 17q21.31 | CRHR1, RP11-10 | LINC02210-CRHR1, LINC02210-CRHR1 | | | ENSG00000282456, E | NA | NA | rs17563683-G | rs17563683 | 0 | 17563683 | intron_variant | 0 | 0.2333 | 3.00E-26 | 25.52288 | | 0.043442 | [0.035-0.0 | Affymetrix [~ 2500000] (imputed) | N |
| 9 | 17:44118848 | 17 | 43811072 | rs17563683 | 17/06/2020 | 3.2E+07 | Oskarsson GR | 23/04/2020 | Commun Biol | www.ncbi.nlm.nih.gov/pubmed/32327693 | Predicted loss and gain of function mutations in ACO1 are associated with erythropoiesis. | Hemoglobin levels | 684,122 Eu | NA | 17q21.31 | MGC57346-CRH | LINC02210-CRHR1, LINC02210-CRHR1 | | | ENSG00000282456, E | NA | NA | rs17563683-G | rs17563683 | 0 | 17563683 | intron_variant | 0 | | 4.00E-61 | 60.39794 | | 0.043 | [NR] unit in | Affymetrix, Illumina [43000000] (imputed) | N |
| 9 | 17:44118848 | 17 | 43991272 | rs17563986 | 12/01/2018 | 2.9E+07 | Lam M | 28/11/2017 | Cell Rep | www.ncbi.nlm.nih.gov/pubmed/29186694 | Large-Scale Cognitive GWAS Meta-Analysis Reveals Tissue-Specific Neural Expression and Potential Nootropic Drug Targets. | Cognitive ability | 107,207 Eu | NA | 17q21.31 | MAPT | MAPT, MAPT, MAPT | | | ENSG00000277956, E | NA | NA | rs17563986-? | rs17563986 | 0 | 17563986 | intron_variant | 0 | NR | 8.00E-08 | 7.09691 | | 5.368 | z score inc | Affymetrix, Illumina [7357080] (imputed) | N |
| 9 | 17:44118848 | 17 | 43991272 | rs17563986 | 12/01/2018 | 2.9E+07 | Lam M | 28/11/2017 | Cell Rep | www.ncbi.nlm.nih.gov/pubmed/29186694 | Large-Scale Cognitive GWAS Meta-Analysis Reveals Tissue-Specific Neural Expression and Potential Nootropic Drug Targets. | Cognitive ability (MTAG) | 436,124 Eu | NA | 17q21.31 | MAPT | MAPT, MAPT, MAPT | | | ENSG00000277956, E | NA | NA | rs17563986-? | rs17563986 | 0 | 17563986 | intron_variant | 0 | NR | 5.00E-12 | 11.30103 | | 6.905 | z score inc | Affymetrix, Illumina, Perlegen [7333576] (imputed) | N |
| 9 | 17:44118848 | 17 | 44006601 | rs17564829 | 14/05/2018 | 2.3E+07 | Liu JZ | 1/10/2012 | Nat Genet | www.ncbi.nlm.nih.gov/pubmed/22961000 | Dense fine-mapping study identifies new susceptibility loci for primary biliary cirrhosis. | Primary biliary cirrhosis | 2,861 Britis | NA | 17q21.31 | CRHR1, MAPT | MAPT, MAPT, MAPT | | | ENSG00000277956, E | NA | NA | rs17564829-G | rs17564829 | 0 | 17564829 | intron_variant | 0 | 0.24 | 2.00E-09 | 8.69897 | | 1.25 | [1.16-1.35 | Illumina [237619] (imputed) | N |
| 9 | 17:44118848 | 17 | 44187492 | rs17577094 | 5/12/2014 | 2.5E+07 | Vacic V | 19/05/2014 | Hum Mol Genet | www.ncbi.nlm.nih.gov/pubmed/24842889 | Genome-wide mapping of IBD segments in an Ashkenazi PD cohort identifies associated haplotypes. | Parkinson's disease | 1,130 Ashk | 306 Ashke | 17q21.31 | MAPT | KANSL1, KANSL1, KANSL1 | | | ENSG00000120071, E | NA | NA | rs17577094-? | rs17577094 | 0 | 17577094 | intron_variant | 0 | 0.74 | 8.00E-09 | 8.09691 | | 1.56 | [1.39-1.79 | Affymetrix, Illumina [1069161] (imputed) | N |
| 9 | 17:44118848 | 17 | 44192923 | rs17577369 | 22/01/2019 | 3E+07 | Nagel M | 2/03/2018 | Nat Commun | www.ncbi.nlm.nih.gov/pubmed/29500382 | Item-level analyses reveal genetic heterogeneity in neuroticism. | Feeling miserable | 376,097 Eu | NA | 17q21.31 | NR | KANSL1, KANSL1, KANSL1 | | | ENSG00000120071, E | NA | NA | rs17577369-A | rs17577369 | 0 | 17577369 | intron_variant | 0 | 0.2157 | 4.00E-12 | 11.39794 | | 6.96 | z score dec | NR [10847151] (imputed) | N |
| 9 | 17:44118848 | 17 | 43516402 | rs17631303 | 2/08/2013 | 2.4E+07 | Couch FJ | 27/03/2013 | PLoS Genet | www.ncbi.nlm.nih.gov/pubmed/23544013 | Genome-wide association study in BRCA1 mutation carriers identifies novel loci associated with breast and ovarian cancer risk. | Ovarian cancer in BRCA1 mutation carriers | 683 Europ | 1,706 Euro | 17q21.31 | NR | PLEKHM1 | | | ENSG00000277111 | NA | NA | rs17631303-? | rs17631303 | 0 | 17631303 | intron_variant | 0 | NR | 1.00E-08 | 8 | | NA | | Illumina [2568349] (imputed) | N |
| 9 | 17:44118848 | 17 | 43516402 | rs17631303 | 2/08/2013 | 2.4E+07 | Couch FJ | 27/03/2013 | PLoS Genet | www.ncbi.nlm.nih.gov/pubmed/23544013 | Genome-wide association study in BRCA1 mutation carriers identifies novel loci associated with breast and ovarian cancer risk. | Ovarian cancer in BRCA1 mutation carriers | 683 Europ | 1,706 Euro | 17q21.31 | intergenic | PLEKHM1 | | | ENSG00000277111 | NA | NA | rs17631303-G | rs17631303 | 0 | 17631303 | intron_variant | 0 | 0.19 | 1.00E-08 | 8 | | 1.27 | [1.17-1.38 | Illumina [2568349] (imputed) | N |
| 9 | 17:44118848 | 17 | 43994648 | rs17649553 | 9/05/2015 | 2.5E+07 | Nalls MA | 27/07/2014 | Nat Genet | www.ncbi.nlm.nih.gov/pubmed/25064009 | Large-scale meta-analysis of genome-wide association data identifies six new risk loci for Parkinson's disease. | Parkinson's disease | 13,708 Eur | 5,353 Euro | 17q21.31 | MAPT | MAPT, MAPT, MAPT | | | ENSG00000277956, E | NA | NA | rs17649553-C | rs17649553 | 0 | 17649553 | intron_variant | 0 | 0.774 | 2.00E-48 | 47.69897 | | 1.3 | [1.27-1.34 | Illumina [7893274] (imputed) | N |
| 9 | 17:44118848 | 17 | 43994648 | rs17649553 | 24/10/2017 | 2.9E+07 | Chang D | 11/09/2017 | Nat Genet | www.ncbi.nlm.nih.gov/pubmed/28892059 | A meta-analysis of genome-wide association studies identifies 17 new Parkinson's disease risk loci. | Parkinson's disease | 20,184 Eur | 5,851 Euro | 17q21.31 | ARHGAP27, CRH | MAPT, MAPT, MAPT | | | ENSG00000277956, E | NA | NA | rs17649553-C | rs17649553 | 0 | 17649553 | intron_variant | 0 | 0.779 | 1.00E-68 | 68 | | 1.282051 | [1.25-1.32 | Illumina [11933700] (imputed) | N |
| 9 | 17:44118848 | 17 | 44037491 | rs17650842 | 22/01/2019 | 3E+07 | Nagel M | 2/03/2018 | Nat Commun | www.ncbi.nlm.nih.gov/pubmed/29500382 | Item-level analyses reveal genetic heterogeneity in neuroticism. | Irritable mood | 366,726 Eu | NA | 17q21.31 | NR | MAPT, MAPT, MAPT | | | ENSG00000277956, E | NA | NA | rs17650842-A | rs17650842 | 0 | 17650842 | intron_variant | 0 | 0.21663 | 2.00E-12 | 11.69897 | | 7.01 | z score dec | NR [~ 10847151] (imputed) | N |
| 9 | 17:44118848 | 17 | 44098967 | rs17652520 | 29/01/2020 | 3.2E+07 | Hill WD | 16/12/2019 | Nat Commun | www.ncbi.nlm.nih.gov/pubmed/31844048 | Genome-wide analysis identifies molecular systems and 149 genetic loci associated with income. | Household income | 286,301 Br | NA | 17q21.31 | MAPT | MAPT, MAPT, MAPT | | | ENSG00000277956, E | NA | NA | rs17652520-A | rs17652520 | 0 | 17652520 | intron_variant | 0 | NR | 5.00E-12 | 11.30103 | | 0.013847 | [0.0099-0. | Affymetrix [18485882] (imputed) | N |
| 9 | 17:44118848 | 17 | 44098967 | rs17652520 | 4/08/2022 | 3.6E+07 | May-Wilson S | 18/05/2022 | Nat Commun | www.ncbi.nlm.nih.gov/pubmed/35585065 | Large-scale GWAS of food liking reveals genetic determinants and genetic correlations with distinct neurophysiological traits. | Lager liking | 156,958 Eu | NA | 17q21.31 | | MAPT, MAPT, MAPT | | | ENSG00000277956, E | NA | NA | rs17652520-A | rs17652520 | 0 | 17652520 | intron_variant | 0 | 0.2287 | 8.00E-10 | 9.09691 | | 0.02614 | [0.018-0.0 | Affymetrix [11353280] (imputed) | N |
| 9 | 17:44118848 | 17 | 44098967 | rs17652520 | 21/06/2019 | 3.1E+07 | Wu Y | 23/04/2019 | Nat Commun | www.ncbi.nlm.nih.gov/pubmed/31015401 | Genome-wide association study of medication-use and associated disease in the UK Biobank. | Medication use (anilides) | 83,218 Eur | NA | 17q21.31 | MAPT | MAPT, MAPT, MAPT | | | ENSG00000277956, E | NA | NA | rs17652520-A | rs17652520 | 0 | 17652520 | intron_variant | 0 | 0.231386 | 8.00E-13 | 12.09691 | | 0.05571 | [0.04-0.07 | Affymetrix [7288503] (imputed) | N |
| 9 | 17:44118848 | 17 | 44098967 | rs17652520 | 1/02/2018 | 2.9E+07 | Luciano M | 18/12/2017 | Nat Genet | www.ncbi.nlm.nih.gov/pubmed/29255261 | Association analysis in over 329,000 individuals identifies 116 independent variants influencing neuroticism. | Neuroticism | 329,821 Eu | 122,867 Eu | 17q21.31 | intergenic | MAPT, MAPT, MAPT | | | ENSG00000277956, E | NA | NA | rs17652520-A | rs17652520 | 0 | 17652520 | intron_variant | 0 | 0.2216 | 1.00E-25 | 25 | | 10.472 | unit increa | Illumina [18485882] (imputed) | N |
| 9 | 17:44118848 | 17 | 44098967 | rs17652520 | 8/02/2019 | 3.1E+07 | Kichaev G | 27/12/2018 | Am J Hum Genet | www.ncbi.nlm.nih.gov/pubmed/30595370 | Leveraging Polygenic Functional Enrichment to Improve GWAS Power. | Neuroticism | approxima | NA | 17q21.31 | | MAPT, MAPT, MAPT | | | ENSG00000277956, E | NA | NA | rs17652520-? | rs17652520 | 0 | 17652520 | intron_variant | 0 | NR | 2.00E-27 | 26.69897 | | NA | | NR [~ 8900000] (imputed) | N |
| 9 | 17:44118848 | 17 | 44192957 | rs17661015 | 22/01/2019 | 3E+07 | Nagel M | 2/03/2018 | Nat Commun | www.ncbi.nlm.nih.gov/pubmed/29500382 | Item-level analyses reveal genetic heterogeneity in neuroticism. | Feeling hurt | 372,047 Eu | NA | 17q21.31 | NR | KANSL1, KANSL1, KANSL1 | | | ENSG00000120071, E | NA | NA | rs17661015-T | rs17661015 | 0 | 17661015 | intron_variant | 0 | 0.215131 | 3.00E-28 | 27.52288 | | 11.02 | z score dec | NR [10847151] (imputed) | N |
| 9 | 17:44118848 | 17 | 44192957 | rs17661015 | 22/01/2019 | 3E+07 | Nagel M | 2/03/2018 | Nat Commun | www.ncbi.nlm.nih.gov/pubmed/29500382 | Item-level analyses reveal genetic heterogeneity in neuroticism. | Irritable mood | 366,726 Eu | NA | 17q21.31 | NR | KANSL1, KANSL1, KANSL1 | | | ENSG00000120071, E | NA | NA | rs17661015-T | rs17661015 | 0 | 17661015 | intron_variant | 0 | 0.215131 | 4.00E-12 | 11.39794 | | 6.93 | z score dec | NR [~ 10847151] (imputed) | N |
| 9 | 17:44118848 | 17 | 44357351 | rs17665188 | 22/01/2019 | 3E+07 | Nagel M | 2/03/2018 | Nat Commun | www.ncbi.nlm.nih.gov/pubmed/29500382 | Item-level analyses reveal genetic heterogeneity in neuroticism. | Experiencing mood swings | 373,733 Eu | NA | 17q21.31 | NR | ARL17B, ARL17B, ARL17B | | | ENSG00000275748, E | NA | NA | rs17665188-T | rs17665188 | 0 | 17665188 | intron_variant | 0 | 0.213949 | 1.00E-18 | 18 | | 8.79 | z score dec | NR [10847151] (imputed) | N |
| 9 | 17:44118848 | 17 | 43906828 | rs17689882 | 3/11/2015 | 2.6E+07 | Hibar DP | 21/01/2015 | Nature | www.ncbi.nlm.nih.gov/pubmed/25607358 | Common genetic variants influence human subcortical brain structures. | Subcortical brain region volumes | up to 13,1 | up to 15,1 | 17q21.31 | CRHR1 | CRHR1, CRHR1, CRHR1, LINC02210-CRHR1, LINC02210-C | | | ENSG00000276191, E | NA | NA | rs17689882-A | rs17689882 | 0 | 17689882 | intron_variant | 0 | 0.22 | 8.00E-09 | 8.09691 | (Intracrani | 13460.47 | [8891.61-1 | Affymetrix, Illumina [NR] (imputed) | N |
| 9 | 17:44118848 | 17 | 43927290 | rs17769552 | 13/09/2022 | 3.6E+07 | Hoglund J | 22/07/2022 | Front Immunol | www.ncbi.nlm.nih.gov/pubmed/35935937 | Gene-Based Variant Analysis of Whole-Exome Sequencing in Relation to Eosinophil Count. | Eosinophil counts | 365,954 Br | NA | 17q21.31 | | MAPT-AS1, MAPT-AS1, MAPT-AS1 | | | ENSG00000264589, E | NA | NA | rs17769552-A | rs17769552 | 0 | 17769552 | intron_variant | 0 | 0.198 | 1.00E-17 | 17 | | 0.038 | [0.03-0.04 | Affymetrix [6661079] (imputed) | N |
| 9 | 17:44118848 | 17 | 43927290 | rs17769552 | 9/07/2018 | 3E+07 | Tikkanen E | 24/04/2018 | Sci Rep | www.ncbi.nlm.nih.gov/pubmed/29691431 | Biological Insights Into Muscular Strength: Genetic Findings in the UK Biobank. | Hand grip strength | 334,825 Br | NA | 17q21.31 | SPPL2C | MAPT-AS1, MAPT-AS1, MAPT-AS1 | | | ENSG00000264589, E | NA | NA | rs17769552-A | rs17769552 | 0 | 17769552 | intron_variant | 0 | 0.2235 | 2.00E-08 | 7.69897 | | 0.002 | [0.0014-0. | Affymetrix [15275733] (imputed) | N |
| 9 | 17:44118848 | 17 | 44219831 | rs1819040 | 14/03/2022 | 3.4E+07 | COVID-19 Host Genetics Initiative | 8/07/2021 | Nature | www.ncbi.nlm.nih.gov/pubmed/34237774 | Mapping the human genetic architecture of COVID-19. | COVID-19 (hospitalized vs population) | 790 Africar | NA | 17q21.31 | | KANSL1, KANSL1, KANSL1 | | | ENSG00000120071, E | NA | NA | rs1819040-A | rs1819040 | 0 | 1819040 | intron_variant | 0 | 0.1855 | 2.00E-10 | 9.69897 | | 0.879018 | [0.844849 | Affymetrix, Illumina [NR] (imputed) | N |
| 9 | 17:44118848 | 17 | 44219831 | rs1819040 | 14/03/2022 | 3.4E+07 | COVID-19 Host Genetics Initiative | 8/07/2021 | Nature | www.ncbi.nlm.nih.gov/pubmed/34237774 | Mapping the human genetic architecture of COVID-19. | SARS-CoV-2 infection | 2,544 Afric | NA | 17q21.31 | | KANSL1, KANSL1, KANSL1 | | | ENSG00000120071, E | NA | NA | rs1819040-A | rs1819040 | 0 | 1819040 | intron_variant | 0 | 0.184 | 5.00E-06 | 5.30103 | | 0.957168 | [0.939337 | Affymetrix, Illumina, Perlegen [NR] (imputed) | N |
| 9 | 17:44118848 | 17 | 44359783 | rs184837332 | 12/11/2021 | 3.4E+07 | Thompson A | 8/06/2021 | Hum Mol Genet | www.ncbi.nlm.nih.gov/pubmed/34104963 | Assessing the impact of alcohol consumption on the genetic contribution to mean corpuscular volume. | Mean corpuscular volume | 362,595 Br | NA | 17q21.31 | | ARL17B, ARL17B, ARL17B | | | ENSG00000275748, E | NA | NA | rs184837332-G | rs184837332 | 0 | 1.85E+08 | intron_variant | 0 | 0.775 | 9.00E-14 | 13.04576 | | 0.1 | [0.07-0.13 | Affymetrix [NR] (imputed) | N |
| 9 | 17:44118848 | 17 | 44408004 | rs1863114 | 17/06/2020 | 3.2E+07 | Oskarsson GR | 23/04/2020 | Commun Biol | www.ncbi.nlm.nih.gov/pubmed/32327693 | Predicted loss and gain of function mutations in ACO1 are associated with erythropoiesis. | Hemoglobin levels | 684,122 Eu | NA | 17q21.31 | LRRC37A | ARL17B, ARL17B, ARL17B, LRRC37A, LRRC37A | | | ENSG00000275748, E | NA | NA | rs1863114-C | rs1863114 | 0 | 1863114 | missense_variant | 0 | | 2.00E-50 | 49.69897 | | 0.045 | [NR] unit in | Affymetrix, Illumina [43000000] (imputed) | N |
| 9 | 17:44118848 | 17 | 43977827 | rs1864325 | 11/04/2013 | 2.3E+07 | Estrada K | 15/04/2012 | Nat Genet | www.ncbi.nlm.nih.gov/pubmed/22504420 | Genome-wide meta-analysis identifies 56 bone mineral density loci and reveals 14 loci associated with risk of fracture. | Lumbar spine bone mineral density | Up to 32,9 | Up to 50,9 | 17q21.31 | MAPT | MAPT, MAPT, MAPT | | | ENSG00000277956, E | NA | NA | rs1864325-T | rs1864325 | 0 | 1864325 | intron_variant | 0 | 0.22 | 5.00E-11 | 10.30103 | | 0.04 | [NR] unit d | Affymetrix, Illumina [~ 2500000] (imputed) | N |
| 9 | 17:44118848 | 17 | 43682323 | rs186806998 | 1/11/2018 | 3E+07 | Wyss AB | 30/07/2018 | Nat Commun | www.ncbi.nlm.nih.gov/pubmed/30061609 | Multiethnic meta-analysis identifies ancestry-specific and cross-ancestry loci for pulmonary function. | Lung function (FEV1) | 60,552 European ance | | 17q21.31 | CRHR1 | MAPK8IP1P2 - RP | ENSG00000263503 | ENSG00000282238 | | 2617 | 3614 | rs186806998-T | rs186806998 | 1 | 56383763 | upstream_gene_variant | 1 | 0.79 | 1.00E-09 | 9 | (EA) | 36.39 | [24.71-48. | Affymetrix, Illumina [10900000] (imputed) | N |
| 9 | 17:44118848 | 17 | 43682323 | rs186806998 | 1/11/2018 | 3E+07 | Wyss AB | 30/07/2018 | Nat Commun | www.ncbi.nlm.nih.gov/pubmed/30061609 | Multiethnic meta-analysis identifies ancestry-specific and cross-ancestry loci for pulmonary function. | Lung function (FEV1) | 60,552 European ance | | 17q21.31 | LOC644172, CRH | MAPK8IP1P2 - RP | ENSG00000263503 | ENSG00000282238 | | 2617 | 3614 | rs186806998-T | rs186806998 | 1 | 56383763 | upstream_gene_variant | 1 | 0.82 | 3.00E-10 | 9.522879 | | 29.5 | [20.29-38. | Affymetrix, Illumina [10900000] (imputed) | N |
| 9 | 17:44118848 | 17 | 43682323 | rs186806998 | 1/11/2018 | 3E+07 | Wyss AB | 30/07/2018 | Nat Commun | www.ncbi.nlm.nih.gov/pubmed/30061609 | Multiethnic meta-analysis identifies ancestry-specific and cross-ancestry loci for pulmonary function. | Lung function (FVC) | 60,552 European ance | | 17q21.31 | | MAPK8IP1P2 - RP | ENSG00000263503 | ENSG00000282238 | | 2617 | 3614 | rs186806998-? | rs186806998 | 1 | 56383763 | upstream_gene_variant | 1 | NR | 1.00E-08 | 8 | | 30.52 | [20.01-41. | Affymetrix, Illumina [10900000] (imputed) | N |
| 9 | 17:44118848 | 17 | 43911443 | rs1876829 | 22/09/2019 | 3.1E+07 | Lotta LA | 1/12/2018 | JAMA | www.ncbi.nlm.nih.gov/pubmed/30575882 | Association of Genetic Variants Related to Gluteofemoral vs Abdominal Fat Distribution With Type 2 Diabetes, Coronary Disease, and Cardiovascular Risk Factors. | Waist-hip ratio | 663,598 Eu | NA | 17q21.31 | NR | LINC02210-CRHR1, LINC02210-CRHR1, MAPT-AS1, MAPT-A | | | ENSG00000282456, E | NA | NA | rs1876829-C | rs1876829 | 0 | 1876829 | intron_variant | 0 | 0.22 | 5.00E-08 | 7.30103 | | NA | | Affymetrix [2446094] (imputed) | N |
| 9 | 17:44118848 | 17 | 43911443 | rs1876829 | 22/09/2019 | 3.1E+07 | Lotta LA | 1/12/2018 | JAMA | www.ncbi.nlm.nih.gov/pubmed/30575882 | Association of Genetic Variants Related to Gluteofemoral vs Abdominal Fat Distribution With Type 2 Diabetes, Coronary Disease, and Cardiovascular Risk Factors. | Waist-to-hip ratio adjusted for BMI | 660,648 Eu | NA | 17q21.31 | NR | LINC02210-CRHR1, LINC02210-CRHR1, MAPT-AS1, MAPT-A | | | ENSG00000282456, E | NA | NA | rs1876829-C | rs1876829 | 0 | 1876829 | intron_variant | 0 | 0.22 | 7.00E-17 | 16.1549 | | 0.019 | [0.015-0.0 | Affymetrix [2446094] (imputed) | N |
| 9 | 17:44118848 | 17 | 43567337 | rs1879586 | 4/08/2017 | 2.8E+07 | Phelan CM | 27/03/2017 | Nat Genet | www.ncbi.nlm.nih.gov/pubmed/28346442 | Identification of 12 new susceptibility loci for different histotypes of epithelial ovarian cancer. | Epithelial ovarian cancer | 16,924 Eur | NA | 17q21.31 | NR | PLEKHM1 | | | ENSG00000277111 | NA | NA | rs1879586-G | rs1879586 | 0 | 1879586 | intron_variant | 0 | 0.181254 | 2.00E-19 | 18.69897 | | 1.161137 | [1.123966 | Illumina [11595112] (imputed) | N |
| 9 | 17:44118848 | 17 | 43567337 | rs1879586 | 7/08/2017 | 2.8E+07 | Phelan CM | 27/03/2017 | Nat Genet | www.ncbi.nlm.nih.gov/pubmed/28346442 | Identification of 12 new susceptibility loci for different histotypes of epithelial ovarian cancer. | High-grade serous ovarian cancer | 13,037 Eur | NA | 17q21.31 | NR | PLEKHM1 | | | ENSG00000277111 | NA | NA | rs1879586-G | rs1879586 | 0 | 1879586 | intron_variant | 0 | 0.181254 | 3.00E-13 | 12.52288 | | 1.158558 | [1.113784 | Illumina [11595112] (imputed) | N |
| 9 | 17:44118848 | 17 | 43567337 | rs1879586 | 3/08/2017 | 2.8E+07 | Phelan CM | 27/03/2017 | Nat Genet | www.ncbi.nlm.nih.gov/pubmed/28346442 | Identification of 12 new susceptibility loci for different histotypes of epithelial ovarian cancer. | Invasive epithelial ovarian cancer | 22,406 Eur | NA | 17q21.31 | NR | PLEKHM1 | | | ENSG00000277111 | NA | NA | rs1879586-G | rs1879586 | 0 | 1879586 | intron_variant | 0 | 0.181254 | 2.00E-13 | 12.69897 | | 1.133901 | [1.096609 | Illumina [11595112] (imputed) | N |
| 9 | 17:44118848 | 17 | 43567337 | rs1879586 | 3/08/2017 | 2.8E+07 | Phelan CM | 27/03/2017 | Nat Genet | www.ncbi.nlm.nih.gov/pubmed/28346442 | Identification of 12 new susceptibility loci for different histotypes of epithelial ovarian cancer. | Ovarian cancer in BRCA1 mutation carriers | 2,933 Euro | NA | 17q21.31 | NR | PLEKHM1 | | | ENSG00000277111 | NA | NA | rs1879586-G | rs1879586 | 0 | 1879586 | intron_variant | 0 | 0.181254 | 7.00E-06 | 5.154902 | | 1.180455 | [1.098309 | Illumina [11595112] (imputed) | N |

| | | | | | | | | | | | | | | | | | | | | | | | | | | | | | | | | | | | | |
|---|---|---|---|---|---|---|---|---|---|---|---|---|---|---|---|---|---|---|---|---|---|---|---|---|---|---|---|---|---|---|---|---|---|---|---|---|
| 9 | 17:44118848 | 17 | 43567337 | rs1879586 | 7/08/2017 | 2.8E+07 | Phelan CM | 27/03/2017 | Nat Genet | www.ncbi.nlm.nih.gov/pubmed/28346442 | Identification of 12 new susceptibility loci for different histotypes of epithelial ovarian cancer. | Serous invasive ovarian cancer | 14,049 Eu | NA | 17q21.31 | NR | PLEKHM1 | | | ENSG00000277111 | NA | NA | rs1879586-G | rs1879586 | 0 | 1879586 | intron_variant | 0 | 0.181254 | 3.00E-12 | 11.52288 | | 1.147209 | [1.104061- | Illumina [11595112] (imputed) | N |
| 9 | 17:44118848 | 17 | 44248769 | rs1881193 | 22/01/2019 | 3E+07 | Nagel M | 2/03/2018 | Nat Commun | www.ncbi.nlm.nih.gov/pubmed/29500382 | Item-level analyses reveal genetic heterogeneity in neuroticism. | Experiencing mood swings | 373,733 Eu | NA | 17q21.31 | NR | KANSL1, KANSL1, KANSL1 | | | ENSG00000120071, E | NA | NA | rs1881193-T | rs1881193 | 0 | 1881193 | missense_variant | 0 | 0.21737 | 1.00E-18 | 18 | | 8.84 | z score dec | NR [10847151] (imputed) | N |
| 9 | 17:44091886 | 17 | 44228169 | rs1918793 | 31/05/2022 | 3.4E+07 | Smith SM | 19/04/2021 | Nat Neurosci | www.ncbi.nlm.nih.gov/pubmed/33875891 | An expanded set of genome-wide association studies of brain imaging phenotypes in UK Biobank. | IDP dMRI TBSS L3 Genu of corpus callosum | 20,860 Brit | 10,496 Brit | 17q21.31 | | KANSL1, KANSL1, KANSL1 | | | ENSG00000120071, E | NA | NA | rs1918793-T | rs1918793 | 0 | 1918793 | intron_variant | 0 | 0.227 | 2.00E-15 | 14.69897 | | 0.093 | [0.069-0.1 | Affymetrix [17103079] (imputed) | N |
| 9 | 17:44118848 | 17 | 44218138 | rs1918800 | 31/05/2022 | 3.4E+07 | Smith SM | 19/04/2021 | Nat Neurosci | www.ncbi.nlm.nih.gov/pubmed/33875891 | An expanded set of genome-wide association studies of brain imaging phenotypes in UK Biobank. | ThalamNuclei lh volume VAmc | 21,282 Brit | 10,686 Brit | 17q21.31 | | KANSL1, KANSL1, KANSL1 | | | ENSG00000120071, E | NA | NA | rs1918800-T | rs1918800 | 0 | 1918800 | intron_variant | 0 | 0.182 | 9.00E-11 | 10.04576 | | 0.087 | [0.062-0.1 | Affymetrix [17103079] (imputed) | N |
| 9 | 17:44118848 | 17 | 43991515 | rs192818565 | 25/02/2017 | 2.7E+07 | Okbay A | 26/05/2016 | Nature | www.ncbi.nlm.nih.gov/pubmed/27225129 | Genome-wide association study identifies 74 loci associated with educational attainment. | Educational attainment (college completion) | 280,007 Eu | NA | 17q21.31 | NR | MAPT, MAPT, MAPT | | | ENSG00000277956, E | NA | NA | rs192818565-T | rs192818565 | 1 | 62056842 | intron_variant | 0 | 0.8097 | 5.00E-11 | 10.30103 | | 1.053487 | [NR] | Affymetrix, Illumina, Perlegen [~ 9300000] (imputed) | N |
| 9 | 17:44118848 | 17 | 43991515 | rs192818565 | 24/02/2017 | 2.7E+07 | Okbay A | 26/05/2016 | Nature | www.ncbi.nlm.nih.gov/pubmed/27225129 | Genome-wide association study identifies 74 loci associated with educational attainment. | Educational attainment (years of education) | 405,072 Eu | NA | 17q21.31 | NR | MAPT, MAPT, MAPT | | | ENSG00000277956, E | NA | NA | rs192818565-T | rs192818565 | 1 | 62056842 | intron_variant | 0 | 0.7988 | 2.00E-12 | 11.69897 | | 0.020457 | NR unit inc | Affymetrix, Illumina, Perlegen [~ 9300000] (imputed) | N |
| 9 | 17:44118848 | 17 | 44142332 | rs193236081 | 17/03/2017 | 2.7E+07 | Okbay A | 18/04/2016 | Nat Genet | www.ncbi.nlm.nih.gov/pubmed/27089181 | Genetic variants associated with subjective well-being, depressive symptoms, and neuroticism identified through genome-wide analyses. | Neuroticism | 170,911 Eu | NA | 17q21.31 | NR | KANSL1, KANSL1, KANSL1 | | | ENSG00000120071, E | NA | NA | rs193236081-T | rs193236081 | 1 | 77184207 | intron_variant | 0 | 0.774 | 6.00E-11 | 10.22185 | | 0.028413 | [0.02-0.03 | Affymetrix [up to 6544775] (imputed) | N |
| 9 | 17:44118848 | 17 | 43539437 | rs1971631 | 3/10/2022 | 3.4E+07 | Merino J | 23/08/2021 | Nat Hum Behav | www.ncbi.nlm.nih.gov/pubmed/34426670 | Genetic analysis of dietary intake identifies new loci and functional links with metabolic traits. | Dietary macronutrient intake (multi-trait analysis) | 282,271 Eu | NA | 17q21.31 | | PLEKHM1 | | | ENSG00000277111 | NA | NA | rs1971631-C | rs1971631 | 0 | 1971631 | intron_variant | 0 | 0.825 | 2.00E-10 | 9.69897 | | NA | | Affymetrix [11800000] (imputed) | N |
| 9 | 17:44118848 | 17 | 44056767 | rs1981997 | 1/11/2013 | 2.4E+07 | Fingerlin TE | 14/04/2013 | Nat Genet | www.ncbi.nlm.nih.gov/pubmed/23583980 | Genome-wide association study identifies multiple susceptibility loci for pulmonary fibrosis. | Interstitial lung disease | 1,161 Euro | 876 Europ | 17q21.31 | MAPT | MAPT, MAPT, MAPT | | | ENSG00000277956, E | NA | NA | rs1981997-G | rs1981997 | 0 | 1981997 | non_coding_transcript_exon | 0 | 0.77 | 9.00E-14 | 13.04576 | | 1.41 | [1.28-1.56 | Illumina [439828] | N |
| 9 | 17:44118848 | 17 | 44083402 | rs1991556 | 24/09/2019 | 3.1E+07 | Evangelou E | 29/07/2019 | Nat Hum Behav | www.ncbi.nlm.nih.gov/pubmed/31358974 | New alcohol-related genes suggest shared genetic mechanisms with neuropsychiatric disorders. | Alcohol consumption | 480,842 Eu | NA | 17q21.31 | MAPT, STH | MAPT, MAPT, MAPT | | | ENSG00000277956, E | NA | NA | rs1991556-A | rs1991556 | 0 | 1991556 | intron_variant | 0 | 0.22 | 5.00E-23 | 22.30103 | | 0.012 | [0.01-0.01 | Affymetrix, Illumina [~ 7100000] (imputed) | N |
| 9 | 17:44118848 | 17 | 44083402 | rs1991556 | 8/02/2019 | 3.1E+07 | Kichaev G | 27/12/2018 | Am J Hum Genet | www.ncbi.nlm.nih.gov/pubmed/30595370 | Leveraging Polygenic Functional Enrichment to Improve GWAS Power. | Lung function (FVC) | approxima | NA | 17q21.31 | | MAPT, MAPT, MAPT | | | ENSG00000277956, E | NA | NA | rs1991556-? | rs1991556 | 0 | 1991556 | intron_variant | 0 | NR | 1.00E-53 | 53 | | NA | | NR [~ 8900000] (imputed) | N |
| 9 | 17:44118848 | 17 | 44083402 | rs1991556 | 18/01/2019 | 3.1E+07 | Doherty A | 10/12/2018 | Nat Commun | www.ncbi.nlm.nih.gov/pubmed/30531941 | GWAS identifies 14 loci for device-measured physical activity and sleep duration. | Sleep duration | 91,105 Eu | NA | 17q21.31 | STH | MAPT, MAPT, MAPT | | | ENSG00000277956, E | NA | NA | rs1991556-G | rs1991556 | 0 | 1991556 | intron_variant | 0 | 0.774711 | 3.00E-09 | 8.522879 | | 0.032805 | [0.022-0.0 | NR [9926106] (imputed) | N |
| 9 | 17:44118848 | 17 | 44819565 | rs199443 | 22/01/2019 | 3E+07 | Nagel M | 2/03/2018 | Nat Commun | www.ncbi.nlm.nih.gov/pubmed/29500382 | Item-level analyses reveal genetic heterogeneity in neuroticism. | Feeling fed-up | 374,971 Eu | NA | 17q21.31 | NR | NSF, NSF, NSF | | | ENSG00000278174, E | NA | NA | rs199443-T | rs199443 | 0 | 199443 | intron_variant | 0 | 0.20675 | 4.00E-13 | 12.39794 | | 7.26 | z score inc | NR [10847151] (imputed) | N |
| 9 | 17:44118848 | 17 | 44819565 | rs199443 | 27/09/2016 | 2.7E+07 | Dong J | 1/11/2015 | Medicine (Baltimore) | www.ncbi.nlm.nih.gov/pubmed/26632684 | Genome-wide Meta-analysis on the Sense of Smell Among US Older Adults. | Sense of smell | 6,252 Euro | NA | 17q21.31 | NSF | NSF, NSF, NSF | | | ENSG00000278174, E | NA | NA | rs199443-T | rs199443 | 0 | 199443 | intron_variant | 0 | 0.2 | 3.00E-06 | 5.522879 | (Ajusted fo | 0.03 | [0.01-0.05 | Affymetrix, Illumina [2543887] (imputed) | N |
| 9 | 17:44118848 | 17 | 44819565 | rs199443 | 27/09/2016 | 2.7E+07 | Dong J | 1/11/2015 | Medicine (Baltimore) | www.ncbi.nlm.nih.gov/pubmed/26632684 | Genome-wide Meta-analysis on the Sense of Smell Among US Older Adults. | Sense of smell | 6,252 Euro | NA | 17q21.31 | NSF | NSF, NSF, NSF | | | ENSG00000278174, E | NA | NA | rs199443-T | rs199443 | 0 | 199443 | intron_variant | 0 | 0.2 | 4.00E-06 | 5.39794 | | 0.03 | [0.01-0.05 | Affymetrix, Illumina [2543887] (imputed) | N |
| 9 | 17:44118848 | 17 | 44812188 | rs199447 | 31/10/2018 | 3E+07 | Nagel M | 25/06/2018 | Nat Genet | www.ncbi.nlm.nih.gov/pubmed/29942085 | Meta-analysis of genome-wide association studies for neuroticism in 449,484 individuals identifies novel genetic loci and pathways. | Neuroticism | 449,484 Eu | NA | 17q21.31 | NR | NSF, NSF, NSF | | | ENSG00000278174, E | NA | NA | rs199447-T | rs199447 | 0 | 199447 | intron_variant | 0 | NR | 2.00E-26 | 25.69897 | | 10.659 | z score inc | Affymetrix [10847151] (imputed) | N |
| 9 | 17:44118848 | 17 | 44797919 | rs199456 | 28/01/2019 | 3.1E+07 | Gao XR | 7/12/2018 | Hum Mol Genet | www.ncbi.nlm.nih.gov/pubmed/30535121 | Genome-wide association analyses identify 139 loci associated with macular thickness in the UK Biobank cohort. | Macular thickness | 59,814 Eu | 8,609 Euro | 17q21.31 | NSF | NSF, NSF, NSF | | | ENSG00000278174, E | NA | NA | rs199456-? | rs199456 | 0 | 199456 | intron_variant | 0 | NR | 3.00E-28 | 27.52288 | | 0.86 | [0.7-1.02] | Affymetrix [11100000] (imputed) | N |
| 9 | 17:44874453 | 17 | 44866602 | rs199497 | 19/08/2022 | 3.6E+07 | Carcel-Marquez J | 8/07/2022 | Front Cardiovasc Med | www.ncbi.nlm.nih.gov/pubmed/35872910 | A Polygenic Risk Score Based on a Cardioembolic Stroke Multitrait Analysis Improves a Clinical Prediction Model for This Stroke Subtype. | Atrial fibrillation (MTAG) | 1,030,836 | NA | 17q21.31 | | WNT3 | | | ENSG00000108379 | NA | NA | rs199497-T | rs199497 | 0 | 199497 | intron_variant | 0 | NR | 1.00E-10 | 10 | | 0.012 | [0.0081-0. | Illumina [6808676] (imputed) | N |
| 9 | 17:44874453 | 17 | 44866602 | rs199497 | 12/11/2018 | 3E+07 | Lee JJ | 23/07/2018 | Nat Genet | www.ncbi.nlm.nih.gov/pubmed/30038396 | Gene discovery and polygenic prediction from a genome-wide association study of educational attainment in 1.1 million individuals. | Cognitive performance (MTAG) | 402,382 Eu | NA | 17q21.31 | Intergenic | WNT3 | | | ENSG00000108379 | NA | NA | rs199497-T | rs199497 | 0 | 199497 | intron_variant | 0 | 0.8232 | 6.00E-10 | 9.221849 | (MTAG) | 0.0189 | [0.013-0.0 | Affymetrix, Illumina [7100000] (imputed) | N |
| 9 | 17:44874453 | 17 | 44866602 | rs199497 | 2/05/2019 | 3.1E+07 | Hill WD | 13/03/2019 | Mol Psychiatry | www.ncbi.nlm.nih.gov/pubmed/30867560 | Genetic contributions to two special factors of neuroticism are associated with affluence, higher intelligence, better health, and longer life. | General factor of neuroticism | 270,059 Bi | NA | 17q21.31 | WNT3 | WNT3 | | | ENSG00000108379 | NA | NA | rs199497-T | rs199497 | 0 | 199497 | intron_variant | 0 | NR | 2.00E-10 | 9.69897 | | 0.010905 | [0.0076-0. | Affymetrix [18485882] (imputed) | N |
| 9 | 17:44874453 | 17 | 44866602 | rs199497 | 27/09/2022 | 3.5E+07 | Trubetskoy V | 8/04/2022 | Nature | www.ncbi.nlm.nih.gov/pubmed/35396580 | Mapping genomic loci implicates genes and synaptic biology in schizophrenia. | Schizophrenia | 53,386 Eu | 1,979 Euro | 17q21.31 | | WNT3 | | | ENSG00000108379 | NA | NA | rs199497-T | rs199497 | 0 | 199497 | intron_variant | 0 | 0.866 | 7.00E-06 | 5.154902 | | 1.05559 | [1.03-1.08 | Affymetrix [7585078] (imputed) | N |
| 9 | 17:44118848 | 17 | 44865498 | rs199499 | 30/08/2022 | 3.2E+07 | Cole JB | 19/03/2020 | Nat Commun | www.ncbi.nlm.nih.gov/pubmed/32193382 | Comprehensive genomic analysis of dietary habits in UK Biobank identifies hundreds of genetic associations. | Alcohol consumption (drinks per month) (UKB data field 1578, 4424) | 449,210 Eu | NA | 17q21.31 | | WNT3 | | | ENSG00000108379 | NA | NA | rs199499-C | rs199499 | 0 | 199499 | intron_variant | 0 | 0.794081 | 2.00E-17 | 16.69897 | | 0.023427 | [0.018-0.0 | NR [NR] (imputed) | N |
| 9 | 17:44118848 | 17 | 44865498 | rs199499 | 3/02/2016 | 2.6E+07 | Jun G | 17/03/2015 | Mol Psychiatry | www.ncbi.nlm.nih.gov/pubmed/25778476 | A novel Alzheimer disease locus located near the gene encoding tau protein. | Alzheimer's disease in APOE e4- carriers | 7,184 case | 718 Europ | 17q21.31 | WNT3 | WNT3 | | | ENSG00000108379 | NA | NA | rs199499-C | rs199499 | 0 | 199499 | intron_variant | 0 | 0.81 | 6.00E-06 | 5.221849 | | 1.162791 | [1.09-1.23 | NR [NR] | N |
| 9 | 17:44118848 | 17 | 44862162 | rs199503 | 15/03/2022 | 3.5E+07 | Kulminski AM | 6/11/2021 | Geroscience | www.ncbi.nlm.nih.gov/pubmed/34743297 | Pleiotropic predisposition to Alzheimer's disease and educational attainment: insights from the summary statistics analysis. | Alzheimer's disease or educational attainment (pleiotropy) | (see Lamb | NA | 17q21.31 | | WNT3 | | | ENSG00000108379 | NA | NA | rs199503-A | rs199503 | 0 | 199503 | intron_variant | 0 | 0.2481 | 2.00E-08 | 7.69897 | | 42.19048 | z score inc | NR [6814003] | N |
| 9 | 17:44118848 | 17 | 44862162 | rs199503 | 27/04/2021 | 3.4E+07 | Peyrot WJ | 8/03/2021 | Nat Genet | www.ncbi.nlm.nih.gov/pubmed/33686288 | Identifying loci with different allele frequencies among cases of eight psychiatric disorders using CC-GWAS. | Schizophrenia vs autism spectrum disorder (ordinary least squares (OLS)) | 40,675 sch | NA | 17q21.31 | | WNT3 | | | ENSG00000108379 | NA | NA | rs199503-? | rs199503 | 0 | 199503 | intron_variant | 0 | NR | 3.00E-09 | 8.522879 | | 0.0179 | NR unit dec | NR [4484088] (imputed) | N |
| 9 | 17:44118848 | 17 | 44862162 | rs199503 | 24/08/2021 | 3.4E+07 | Christakoudi S | 21/05/2021 | Sci Rep | www.ncbi.nlm.nih.gov/pubmed/34021172 | GWAS of allometric body-shape indices in UK Biobank identifies loci suggesting associations with morphogenesis, organogenesis, adrenal cell renewal and cancer. | Waist-hip index | 219,872 Bi | NA | 17q21.31 | WNT3 | WNT3 | | | ENSG00000108379 | NA | NA | rs199503-A | rs199503 | 0 | 199503 | intron_variant | 0 | NR | 1.00E-10 | 10 | | 0.022195 | [0.015-0.0 | Affymetrix [319769] (imputed) | N |
| 9 | 17:44118848 | 17 | 44862162 | rs199503 | 24/08/2021 | 3.4E+07 | Christakoudi S | 21/05/2021 | Sci Rep | www.ncbi.nlm.nih.gov/pubmed/34021172 | GWAS of allometric body-shape indices in UK Biobank identifies loci suggesting associations with morphogenesis, organogenesis, adrenal cell renewal and cancer. | Waist-to-hip ratio adjusted for BMI | 219,872 Bi | NA | 17q21.31 | WNT3 | WNT3 | | | ENSG00000108379 | NA | NA | rs199503-A | rs199503 | 0 | 199503 | intron_variant | 0 | NR | 1.00E-10 | 10 | | 0.022073 | [0.015-0.0 | Affymetrix [319769] (imputed) | N |
| 9 | 17:44861003 | 17 | 44861003 | rs199504 | 4/05/2021 | 3.3E+07 | Hofer E | 22/09/2020 | Nat Commun | www.ncbi.nlm.nih.gov/pubmed/32963231 | Genetic correlations and genome-wide associations of cortical structure in general population samples of 22,824 adults. | Cortical thickness | 22,163 Eu | 22,363 Eu | 17q21.31 | WNT3 | WNT3 | | | ENSG00000108379 | NA | NA | rs199504-? | rs199504 | 0 | 199504 | intron_variant | 0 | | 6.00E-13 | 12.22185 | (Superior t | NA | | Affymetrix, Illumina [NR] (imputed) | N |
| 9 | 17:44861003 | 17 | 44859410 | rs199505 | 31/10/2018 | 3E+07 | Nagel M | 25/06/2018 | Nat Genet | www.ncbi.nlm.nih.gov/pubmed/29942085 | Meta-analysis of genome-wide association studies for neuroticism in 449,484 individuals identifies novel genetic loci and pathways. | Depressed affect | 357,957 Eu | NA | 17q21.31 | NR | WNT3 | | | ENSG00000108379 | NA | NA | rs199505-? | rs199505 | 0 | 199505 | intron_variant | 0 | NR | 1.00E-18 | 18 | | 0.02469 | [0.019-0.0 | Affymetrix [10847151] (imputed) | N |
| 9 | 17:44861003 | 17 | 44857929 | rs199510 | 2/11/2021 | 3.4E+07 | Karlsson Linner R | 26/08/2021 | Nat Neurosci | www.ncbi.nlm.nih.gov/pubmed/34446935 | Multivariate analysis of 1.5 million people identifies genetic associations with traits related to self-regulation and addiction. | Externalizing behaviour (multivariate analysis) | up to 53,2 | NA | 17q21.31 | | WNT3 | | | ENSG00000108379 | NA | NA | rs199510-A | rs199510 | 0 | 199510 | intron_variant | 0 | 0.238 | 3.00E-10 | 9.522879 | | 0.01 | [0.0061-0. | NR [NR] (imputed) | N |
| 9 | 17:44861003 | 17 | 44857352 | rs199512 | 8/10/2019 | 3.2E+07 | Wiberg A | 5/09/2019 | Brain | www.ncbi.nlm.nih.gov/pubmed/31504236 | Handedness, language areas and neuropsychiatric diseases: insights from brain imaging and genetics. | Handedness (Left-handed vs. non-left-handed) | 38,322 Eu | NA | 17q21.31 | WNT3, MAPT, MA | WNT3 | | | ENSG00000108379 | NA | NA | rs199512-? | rs199512 | 0 | 199512 | intron_variant | 0 | NR | 5.00E-08 | 7.30103 | | NA | | NR [~ 11000000] (imputed) | N |
| 9 | 17:44861003 | 17 | 44857352 | rs199512 | 8/10/2019 | 3.2E+07 | Wiberg A | 5/09/2019 | Brain | www.ncbi.nlm.nih.gov/pubmed/31504236 | Handedness, language areas and neuropsychiatric diseases: insights from brain imaging and genetics. | Handedness (left-handed vs. right-handed) | 38,322 Eu | NA | 17q21.31 | WNT3, MAPT, MA | WNT3 | | | ENSG00000108379 | NA | NA | rs199512-C | rs199512 | 0 | 199512 | intron_variant | 0 | 0.791 | 4.00E-09 | 8.39794 | | 1.06 | [1.04-1.07 | NR [~ 11000000] (imputed) | N |
| 9 | 17:44861003 | 17 | 44857352 | rs199512 | 8/10/2019 | 3.2E+07 | Wiberg A | 5/09/2019 | Brain | www.ncbi.nlm.nih.gov/pubmed/31504236 | Handedness, language areas and neuropsychiatric diseases: insights from brain imaging and genetics. | Handedness (non-right-handed vs right-handed) | 44,631 Eu | NA | 17q21.31 | WNT3, MAPT, MA | WNT3 | | | ENSG00000108379 | NA | NA | rs199512-? | rs199512 | 0 | 199512 | intron_variant | 0 | NR | 5.00E-08 | 7.30103 | | NA | | NR [~ 11000000] (imputed) | N |
| 9 | 17:44861003 | 17 | 44856641 | rs199515 | 6/04/2022 | 3.5E+07 | Bellenguez C | 4/04/2022 | Nat Genet | www.ncbi.nlm.nih.gov/pubmed/35379992 | New insights into the genetic etiology of Alzheimer's disease and related dementias. | Alzheimer's disease | 39,106 Eu | 25,392 cas | 17q21.31 | | WNT3 | | | ENSG00000108379 | NA | NA | rs199515-G | rs199515 | 0 | 199515 | intron_variant | 0 | 0.2194 | 9.00E-13 | 12.04576 | | 0.94 | [0.93-0.96 | Affymetrix, Illumina [21101114] (imputed) | N |
| 9 | 17:44861003 | 17 | 44856641 | rs199515 | 20/04/2012 | 2.2E+07 | Pankratz N | 1/03/2012 | Ann Neurol | www.ncbi.nlm.nih.gov/pubmed/22451204 | Meta-analysis of Parkinson's disease: identification of a novel locus, RIT2. | Parkinson's disease | 4,238 Euro | 3,738 Euro | 17q21.31 | WNT3 | WNT3 | | | ENSG00000108379 | NA | NA | rs199515-? | rs199515 | 0 | 199515 | intron_variant | 0 | NR | 3.00E-17 | 16.52288 | | 1.32 | [NR] | Illumina [2500000] (imputed) | N |
| 9 | 17:44118848 | 17 | 44847834 | rs199525 | 22/01/2019 | 3E+07 | Nagel M | 2/03/2018 | Nat Commun | www.ncbi.nlm.nih.gov/pubmed/29500382 | Item-level analyses reveal genetic heterogeneity in neuroticism. | Feeling guilty | 373,380 Eu | NA | 17q21.31 | GOSR2 | WNT3 | | | ENSG00000108379 | NA | NA | rs199525-T | rs199525 | 0 | 199525 | intron_variant | 0 | 0.207235 | 4.00E-08 | 7.39794 | | 5.51 | z score dec | NR [10847151] (imputed) | N |
| 9 | 17:44118848 | 17 | 44847834 | rs199525 | 24/08/2021 | 3.4E+07 | Christakoudi S | 21/05/2021 | Sci Rep | www.ncbi.nlm.nih.gov/pubmed/34021172 | GWAS of allometric body-shape indices in UK Biobank identifies loci suggesting associations with morphogenesis, organogenesis, adrenal cell renewal and cancer. | Hip circumference adjusted for BMI | 219,872 Bi | NA | 17q21.31 | WNT3 | WNT3 | | | ENSG00000108379 | NA | NA | rs199525-G | rs199525 | 0 | 199525 | intron_variant | 0 | NR | 2.00E-08 | 7.69897 | | 0.019361 | [0.013-0.0 | Affymetrix [321016] (imputed) | N |
| 9 | 17:44118848 | 17 | 44847834 | rs199525 | 17/04/2019 | 3.1E+07 | Klein M | 1/03/2019 | Am J Psychiatry | www.ncbi.nlm.nih.gov/pubmed/30818988 | Genetic Markers of ADHD-Related Variations in Intracranial Volume. | Intracranial volume | 24,024 Eu | NA | 17q21.31 | NR | WNT3 | | | ENSG00000108379 | NA | NA | rs199525-T | rs199525 | 0 | 199525 | intron_variant | 0 | NR | 2.00E-20 | 19.69897 | | 9.262 | NR z score | NR [9186920] (imputed) | N |
| 9 | 17:44118848 | 17 | 44847834 | rs199525 | 1/11/2018 | 3E+07 | Wyss AB | 30/07/2018 | Nat Commun | www.ncbi.nlm.nih.gov/pubmed/30061609 | Multiethnic meta-analysis identifies ancestry-specific and cross-ancestry loci for pulmonary function. | Lung function (FEV1) | 60,552 European ance | | 17q21.31 | WNT3 | WNT3 | | | ENSG00000108379 | NA | NA | rs199525-T | rs199525 | 0 | 199525 | intron_variant | 0 | 0.8 | 1.00E-09 | 9 | | 18.85 | [12.81-24. | Affymetrix, Illumina [10900000] (imputed) | N |
| 9 | 17:44118848 | 17 | 44847834 | rs199525 | 1/11/2018 | 3E+07 | Wyss AB | 30/07/2018 | Nat Commun | www.ncbi.nlm.nih.gov/pubmed/30061609 | Multiethnic meta-analysis identifies ancestry-specific and cross-ancestry loci for pulmonary function. | Lung function (FEV1) | 60,552 European ance | | 17q21.31 | WNT3 | WNT3 | | | ENSG00000108379 | NA | NA | rs199525-T | rs199525 | 0 | 199525 | intron_variant | 0 | 0.79 | 2.00E-08 | 7.69897 | (EA) | 19.16 | [12.46-25. | Affymetrix, Illumina [10900000] (imputed) | N |
| 9 | 17:44118848 | 17 | 44847834 | rs199525 | 1/11/2018 | 3E+07 | Wyss AB | 30/07/2018 | Nat Commun | www.ncbi.nlm.nih.gov/pubmed/30061609 | Multiethnic meta-analysis identifies ancestry-specific and cross-ancestry loci for pulmonary function. | Lung function (FVC) | 60,552 European ance | | 17q21.31 | WNT3 | WNT3 | | | ENSG00000108379 | NA | NA | rs199525-T | rs199525 | 0 | 199525 | intron_variant | 0 | 0.8 | 8.00E-09 | 8.09691 | | 20.32 | [13.42-27. | Affymetrix, Illumina [10900000] (imputed) | N |
| 9 | 17:44118848 | 17 | 44847834 | rs199525 | 1/11/2018 | 3E+07 | Wyss AB | 30/07/2018 | Nat Commun | www.ncbi.nlm.nih.gov/pubmed/30061609 | Multiethnic meta-analysis identifies ancestry-specific and cross-ancestry loci for pulmonary function. | Lung function (FVC) | 60,552 European ance | | 17q21.31 | WNT3 | WNT3 | | | ENSG00000108379 | NA | NA | rs199525-T | rs199525 | 0 | 199525 | intron_variant | 0 | 0.79 | 3.00E-07 | 6.522879 | (EA) | 20.03 | [12.41-27. | Affymetrix, Illumina [10900000] (imputed) | N |
| 9 | 17:44118848 | 17 | 44843136 | rs199528 | 4/10/2019 | 3.1E+07 | Brazel DM | 6/12/2018 | Biol Psychiatry | www.ncbi.nlm.nih.gov/pubmed/30679032 | Exome Chip Meta-analysis Fine Maps Causal Variants and Elucidates the Genetic Architecture of Rare Coding Variants in Smoking and Alcohol┬áUse. | Alcohol consumption (drinks per week) | up to 357,8 | NA | 17q21.31 | WNT3 | WNT3 | | | ENSG00000108379 | NA | NA | rs199528-T | rs199528 | 0 | 199528 | intron_variant | 0 | 0.223 | 5.00E-21 | 20.30103 | | 0.0286 | [0.023-0.0 | Illumina [~ 250000] (imputed) | N |
| 9 | 17:44118848 | 17 | 44828931 | rs199533 | 24/01/2022 | 3.4E+07 | He L | 26/02/2021 | Transl Psychiatry | www.ncbi.nlm.nih.gov/pubmed/33637690 | Exome-wide age-of-onset analysis reveals exonic variants in ERN1 and SPPL2C associated with Alzheimer's disease. | Alzheimer's disease (age of onset) (adjusted for APOE e4 carrier status) | 10,216 Eu | 9,261 Euro | 17q21.31 | | NSF, NSF, NSF | | | ENSG00000278174, E | NA | NA | rs199533-? | rs199533 | 0 | 199533 | synonymous_variant | 0 | NR | 4.00E-07 | 6.39794 | | 0.896 | | NR [110450] | N |
| 9 | 17:44118848 | 17 | 44828931 | rs199533 | 24/01/2022 | 3.4E+07 | He L | 26/02/2021 | Transl Psychiatry | www.ncbi.nlm.nih.gov/pubmed/33637690 | Exome-wide age-of-onset analysis reveals exonic variants in ERN1 and SPPL2C associated with Alzheimer's disease. | Alzheimer's disease (age of onset) in APOE e4 non-carriers | 7,185 Euro | 5,416 Euro | 17q21.31 | | NSF, NSF, NSF | | | ENSG00000278174, E | NA | NA | rs199533-? | rs199533 | 0 | 199533 | synonymous_variant | 0 | NR | 1.00E-07 | 7 | | 0.859 | | NR [98334] | N |
| 9 | 17:44118848 | 17 | 44828931 | rs199533 | 13/11/2020 | 3.3E+07 | Rashkin SR | 4/09/2020 | Nat Commun | www.ncbi.nlm.nih.gov/pubmed/32887889 | Pan-cancer study detects genetic risk variants and shared genetic basis in two large cohorts. | Breast cancer | 17,881 Eu | NA | 17q21.31 | | NSF, NSF, NSF | | | ENSG00000278174, E | NA | NA | rs199533-G | rs199533 | 0 | 199533 | synonymous_variant | 0 | NR | 5.00E-08 | 7.30103 | | 1.078 | NR | Affymetrix [7846216] (imputed) | N |

| | | | | | | | | | | | | | | | | | | | | | | | | | | | | | | | | | | | | |
|---|---|---|---|---|---|---|---|---|---|---|---|---|---|---|---|---|---|---|---|---|---|---|---|---|---|---|---|---|---|---|---|---|---|---|---|---|
| 9 | 17:44118848 | 17 | 44828931 | rs199533 | 13/11/2020 | 3.3E+07 | Rashkin SR | 4/09/2020 | Nat Commun | www.ncbi.nlm.nih.gov/pubmed/32887889 | Pan-cancer study detects genetic risk variants and shared genetic basis in two large cohorts. | Cancer (pleiotropy) | 64,962 Eu | NA | 17q21.31 | | NSF, NSF, NSF | | | ENSG00000278174, E | NA | NA | rs199533-G | rs199533 | 0 | 199533 | synonymous_variant | 0 | NR | 4.00E-07 | 6.39794 | | 1.06 | [1.036-1.0 | Affymetrix [7846216] (imputed) | N |
| 9 | 17:44118848 | 17 | 44828931 | rs199533 | 10/12/2009 | 2E+07 | Simon-Sanchez J | 15/11/2009 | Nat Genet | www.ncbi.nlm.nih.gov/pubmed/19915575 | Genome-wide association study reveals genetic risk underlying Parkinson's disease. | Parkinson's disease | 1,713 Euro | 3,361 Euro | 17q21.31 | NSF | NSF, NSF, NSF | | | ENSG00000278174, E | NA | NA | rs199533-C | rs199533 | 0 | 199533 | synonymous_variant | 0 | 0.83 | 1.00E-14 | 14 | | 1.28 | [NR] | Illumina [463185] | N |
| 9 | 17:44118848 | 17 | 44828931 | rs199533 | 22/09/2010 | 2.1E+07 | Hamza TH | 15/08/2010 | Nat Genet | www.ncbi.nlm.nih.gov/pubmed/20711177 | Common genetic variation in the HLA region is associated with late-onset sporadic Parkinson's disease. | Parkinson's disease | 2,000 Euro | Up to 1,44 | 17q21.31 | MAPT | NSF, NSF, NSF | | | ENSG00000278174, E | NA | NA | rs199533-C | rs199533 | 0 | 199533 | synonymous_variant | 0 | 0.78 | 1.00E-06 | 6 | | 1.35 | [1.19-1.52 | Illumina [811597] | N |
| 9 | 17:44118848 | 17 | 44828931 | rs199533 | 1/02/2022 | 3.5E+07 | Hanscombe KB | 9/11/2021 | Genome Med | www.ncbi.nlm.nih.gov/pubmed/34753499 | The genetic case for cardiorespiratory fitness as a clinical vital sign and the routine prescription of physical activity in healthcare. | Physical activity | 89,683 Eu | NA | 17q21.31 | | NSF, NSF, NSF | | | ENSG00000278174, E | NA | NA | rs199533-G | rs199533 | 0 | 199533 | synonymous_variant | 0 | 0.79 | 4.00E-08 | 7.39794 | | 0.25 | [0.16-0.34 | Affymetrix [8918919] (imputed) | N |
| 9 | 17:44118848 | 17 | 44822662 | rs199535 | 4/05/2021 | 3.3E+07 | Hofer E | 22/09/2020 | Nat Commun | www.ncbi.nlm.nih.gov/pubmed/32963231 | Genetic correlations and genome-wide associations of cortical structure in general population samples of 22,824 adults. | Cortical surface area | 18,617 Eu | 22,363 Eu | 17q21.31 | NSF | NSF, NSF, NSF | | | ENSG00000278174, E | NA | NA | rs199535-? | rs199535 | 0 | 199535 | intron_variant | 0 | | 8.00E-18 | 17.09691 | (Fusiform) | NA | | Affymetrix, Illumina [NR] (imputed) | N |
| 9 | 17:44118848 | 17 | 44822662 | rs199535 | 3/10/2022 | 3.4E+07 | Merino J | 23/08/2021 | Nat Hum Behav | www.ncbi.nlm.nih.gov/pubmed/34426670 | Genetic analysis of dietary intake identifies new loci and functional links with metabolic traits. | Dietary macronutrient intake (multi-trait analysis) | 282,271 E | NA | 17q21.31 | | NSF, NSF, NSF | | | ENSG00000278174, E | NA | NA | rs199535-A | rs199535 | 0 | 199535 | intron_variant | 0 | 0.79 | 1.00E-10 | 10 | | NA | | Affymetrix [11800000] (imputed) | N |
| 9 | 17:44118848 | 17 | 44625866 | rs199913382 | 17/06/2020 | 3.2E+07 | Oskarsson GR | 23/04/2020 | Commun Biol | www.ncbi.nlm.nih.gov/pubmed/32327693 | Predicted loss and gain of function mutations in ACO1 are associated with erythropoiesis. | Hemoglobin levels | 684,122 E | NA | 17q21.31 | LRRC37A2 | ARL17A, LRRC37A2 | | | ENSG00000185829, E | NA | NA | rs199913382-C | rs199913382 | 0 | 2E+08 | missense_variant | 0 | | 3.00E-54 | 53.52288 | | 0.043 | [NR] unit in | Affymetrix, Illumina [43000000] (imputed) | N |
| 9 | 17:44118848 | 17 | 44214888 | rs2077551 | 9/03/2020 | 3.2E+07 | Allen RJ | 11/11/2019 | Am J Respir Crit Care Med | www.ncbi.nlm.nih.gov/pubmed/31710517 | Genome-Wide Association Study of Susceptibility to Idiopathic Pulmonary Fibrosis. | Idiopathic pulmonary fibrosis | 2,668 Euro | 1,456 Euro | 17q21.31 | MAPT | KANSL1, KANSL1, KANSL1 | | | ENSG00000120071, E | NA | NA | rs2077551-T | rs2077551 | 0 | 2077551 | intron_variant | 0 | 0.814 | 3.00E-16 | 15.52288 | | 1.408451 | [1.3-1.54] | Affymetrix, Illumina [10790934] (imputed) | N |
| 9 | 17:44118848 | 17 | 44214888 | rs2077551 | 26/08/2022 | 3.6E+07 | Allen RJ | 10/06/2022 | Thorax | www.ncbi.nlm.nih.gov/pubmed/35688625 | Genome-wide association study across five cohorts identifies five novel loci associated with idiopathic pulmonary fibrosis. | Idiopathic pulmonary fibrosis | 4,125 Euro | NA | 17q21.31 | | KANSL1, KANSL1, KANSL1 | | | ENSG00000120071, E | NA | NA | rs2077551-T | rs2077551 | 0 | 2077551 | intron_variant | 0 | 0.807 | 2.00E-20 | 19.69897 | | 1.42 | [1.32-1.53 | Illumina [7554248] (imputed) | N |
| 9 | 17:44118848 | 17 | 43919105 | rs2106785 | 22/01/2019 | 3E+07 | Nagel M | 2/03/2018 | Nat Commun | www.ncbi.nlm.nih.gov/pubmed/29500382 | Item-level analyses reveal genetic heterogeneity in neuroticism. | Irritable mood | 366,726 E | NA | 17q21.31 | ACBD4 | MAPT-AS1, MAPT-AS1, MAPT-AS1 | | | ENSG00000264589, E | NA | NA | rs2106785-T | rs2106785 | 0 | 2106785 | non_coding_transcript_exon_ | 0 | 0.213768 | 3.00E-13 | 12.52288 | | 7.29 | z score inc | NR [~ 10847151] (imputed) | N |
| 9 | 17:44118848 | 17 | 43919096 | rs2106786 | 28/01/2020 | 3.2E+07 | Zhao B | 1/11/2019 | Nat Genet | www.ncbi.nlm.nih.gov/pubmed/31676860 | Genome-wide association analysis of 19,629 individuals identifies variants influencing regional brain volumes and refines their genetic co-architecture with cognitive and mental health traits. | Brain region volumes | 19,629 Bri | NA | 17q21.31 | NR | MAPT-AS1, MAPT-AS1, MAPT-AS1 | | | ENSG00000264589, E | NA | NA | rs2106786-? | rs2106786 | 0 | 2106786 | non_coding_transcript_exon | 0 | NR | 3.00E-08 | 7.522879 | (right pallid | NA | | NR [9227495] (imputed) | N |
| 9 | 17:44118848 | 17 | 43919096 | rs2106786 | 30/08/2017 | 2.8E+07 | Astle WJ | 17/11/2016 | Cell | www.ncbi.nlm.nih.gov/pubmed/27863252 | The Allelic Landscape of Human Blood Cell Trait Variation and Links to Common Complex Disease. | Red blood cell count | 172,952 E | NA | 17q21.31 | SPPL2C | MAPT-AS1, MAPT-AS1, MAPT-AS1 | | | ENSG00000264589, E | NA | NA | rs2106786-G | rs2106786 | 0 | 2106786 | non_coding_transcript_exon | 0 | 0.2242 | 3.00E-36 | 35.52288 | | 0.053745 | [0.045-0.0 | Affymetrix [~ 2500000] (imputed) | N |
| 9 | 17:44118848 | 17 | 43953719 | rs2214258 | 1/02/2018 | 2.9E+07 | Luciano M | 18/12/2017 | Nat Genet | www.ncbi.nlm.nih.gov/pubmed/29255261 | Association analysis in over 329,000 individuals identifies 116 independent variants influencing neuroticism. | Neuroticism | 329,821 E | 122,867 E | 17q21.31 | intergenic | MAPT-AS1, MAPT-AS1, MAPT-AS1 | | | ENSG00000264589, E | NA | NA | rs2214258-C | rs2214258 | 0 | 2214258 | intron_variant | 0 | 0.21533 | 2.00E-25 | 24.69897 | | 10.415 | unit increa | Illumina [18485882] (imputed) | N |
| 9 | 17:44118848 | 17 | 44222019 | rs2222746 | 10/12/2020 | 3.2E+07 | Hollis B | 24/03/2020 | Nat Commun | www.ncbi.nlm.nih.gov/pubmed/32210231 | Genomic analysis of male puberty timing highlights shared genetic basis with hair colour and lifespan. | Male puberty timing (age at voice breaking MTAG) | 205,354 E | NA | 17q21.31 | | KANSL1, KANSL1, KANSL1 | | | ENSG00000120071, E | NA | NA | rs2222746-T | rs2222746 | 0 | 2222746 | intron_variant | 0 | NR | 9.00E-28 | 27.04576 | | 0.047722 | [0.039-0.0 | NR [7400000] (imputed) | N |
| 9 | 17:44118848 | 17 | 44222019 | rs2222746 | 10/12/2020 | 3.2E+07 | Hollis B | 24/03/2020 | Nat Commun | www.ncbi.nlm.nih.gov/pubmed/32210231 | Genomic analysis of male puberty timing highlights shared genetic basis with hair colour and lifespan. | Male puberty timing (early vs. average onset facial hair) | 13,226 Eu | NA | 17q21.31 | | KANSL1, KANSL1, KANSL1 | | | ENSG00000120071, E | NA | NA | rs2222746-T | rs2222746 | 0 | 2222746 | intron_variant | 0 | NR | 4.00E-06 | 5.39794 | | 0.00525 | [0.003-0.0 | NR [7400000] (imputed) | N |
| 9 | 17:44118848 | 17 | 44222019 | rs2222746 | 10/12/2020 | 3.2E+07 | Hollis B | 24/03/2020 | Nat Commun | www.ncbi.nlm.nih.gov/pubmed/32210231 | Genomic analysis of male puberty timing highlights shared genetic basis with hair colour and lifespan. | Male puberty timing (late vs. average onset facial hair) | 26,066 Eu | NA | 17q21.31 | | KANSL1, KANSL1, KANSL1 | | | ENSG00000120071, E | NA | NA | rs2222746-T | rs2222746 | 0 | 2222746 | intron_variant | 0 | NR | 2.00E-30 | 29.69897 | | 0.016454 | [0.014-0.0 | NR [7400000] (imputed) | N |
| 9 | 17:44118848 | 17 | 44222019 | rs2222746 | 10/12/2020 | 3.2E+07 | Hollis B | 24/03/2020 | Nat Commun | www.ncbi.nlm.nih.gov/pubmed/32210231 | Genomic analysis of male puberty timing highlights shared genetic basis with hair colour and lifespan. | Male puberty timing (late vs. average onset voice breaking) | 11,334 Eu | NA | 17q21.31 | | KANSL1, KANSL1, KANSL1 | | | ENSG00000120071, E | NA | NA | rs2222746-T | rs2222746 | 0 | 2222746 | intron_variant | 0 | NR | 2.00E-11 | 10.69897 | | 0.006902 | [0.0049-0. | NR [7400000] (imputed) | N |
| 9 | 17:44118848 | 17 | 44354549 | rs2261201 | 22/01/2019 | 3E+07 | Nagel M | 2/03/2018 | Nat Commun | www.ncbi.nlm.nih.gov/pubmed/29500382 | Item-level analyses reveal genetic heterogeneity in neuroticism. | Irritable mood | 366,726 E | NA | 17q21.31 | NR | ARL17B, ARL17B, ARL17B | | | ENSG00000275748, E | NA | NA | rs2261201-C | rs2261201 | 0 | 2261201 | intron_variant | 0 | 0.214631 | 5.00E-12 | 11.30103 | | 6.91 | z score dec | NR [~ 10847151] (imputed) | N |
| 9 | 17:44118848 | 17 | 44304884 | rs2261497 | 31/05/2022 | 3.4E+07 | Smith SM | 19/04/2021 | Nat Neurosci | www.ncbi.nlm.nih.gov/pubmed/33875891 | An expanded set of genome-wide association studies of brain imaging phenotypes in UK Biobank. | aparc-a2009s lh area S-orbital-H-Shaped | 21,282 Bri | 10,685 Bri | 17q21.31 | | KANSL1 - MAPK8 | ENSG00000278458 | ENSG00000282723 | | 2129 | 16088 | rs2261497-T | rs2261497 | 0 | 2261497 | TF_binding_site_variant | 1 | 0.258 | 5.00E-10 | 9.30103 | | 0.071 | [0.049-0.0 | Affymetrix [17103079] (imputed) | N |
| 9 | 17:44118848 | 17 | 43919068 | rs2316766 | 21/07/2020 | 3.2E+07 | Grasby KL | 1/03/2020 | Science | www.ncbi.nlm.nih.gov/pubmed/32193296 | The genetic architecture of the human cerebral cortex. | Cortical thickness | 33,992 Eu | 14,727 Eu | 17q21.31 | NR | MAPT-AS1, MAPT-AS1, MAPT-AS1 | | | ENSG00000264589, E | NA | NA | rs2316766-T | rs2316766 | 0 | 2316766 | non_coding_transcript_exon | 0 | 0.2061 | 3.00E-07 | 6.522879 | (average) | 0.003726 | [0.0023-0. | Affymetrix, Illumina [NR] | N |
| 9 | 17:44118848 | 17 | 43733983 | rs241033 | 24/08/2022 | 3.6E+07 | Plotnikov D | 1/06/2022 | Invest Ophthalmol Vis Sci | www.ncbi.nlm.nih.gov/pubmed/35762941 | High Blood Pressure and Intraocular Pressure: A Mendelian Randomization Study. | Systolic blood pressure | 526,001 E | NA | 17q21.31 | | LINC02210-CRHR1, LINC02210-CRHR1 | | | ENSG00000282456, E | NA | NA | rs241033-A | rs241033 | 0 | 241033 | intron_variant | 0 | NR | 5.00E-10 | 9.30103 | | 0.2944 | [0.2-0.39] | NR [NR] (imputed) | N |
| 9 | 17:44118848 | 17 | 43731719 | rs241036 | 22/01/2019 | 3E+07 | Nagel M | 2/03/2018 | Nat Commun | www.ncbi.nlm.nih.gov/pubmed/29500382 | Item-level analyses reveal genetic heterogeneity in neuroticism. | Experiencing mood swings | 373,733 E | NA | 17q21.31 | NR | LINC02210-CRHR1, LINC02210-CRHR1, LINC02210, LINC( | | | ENSG00000282456, E | NA | NA | rs241036-A | rs241036 | 0 | 241036 | non_coding_transcript_exon | 0 | 0.217056 | 8.00E-20 | 19.09691 | | 9.11 | z score dec | NR [10847151] (imputed) | N |
| 9 | 17:44118848 | 17 | 43731719 | rs241036 | 8/02/2019 | 3.1E+07 | Kichaev G | 27/12/2018 | Am J Hum Genet | www.ncbi.nlm.nih.gov/pubmed/30595370 | Leveraging Polygenic Functional Enrichment to Improve GWAS Power. | Menarche (age at onset) | approxima | NA | 17q21.31 | | LINC02210-CRHR1, LINC02210-CRHR1, LINC02210, LINC( | | | ENSG00000282456, E | NA | NA | rs241036-? | rs241036 | 0 | 241036 | non_coding_transcript_exon | 0 | NR | 7.00E-13 | 12.1549 | | NA | | NR [~ 8900000] (imputed) | N |
| 9 | 17:44118848 | 17 | 44327370 | rs2458185 | 4/08/2022 | 3.6E+07 | May-Wilson S | 18/05/2022 | Nat Commun | www.ncbi.nlm.nih.gov/pubmed/35585065 | Large-scale GWAS of food liking reveals genetic determinants and genetic correlations with distinct neurophysiological traits. | Salami liking | 154,673 E | NA | 17q21.31 | | MAPK8IP1P1 - AR | ENSG00000282723 | ENSG00000228696 | | 4960 | 24780 | rs2458185-A | rs2458185 | 0 | 2458185 | downstream_gene_variant | 1 | 0.2116 | 1.00E-16 | 16 | | 0.03706 | [0.028-0.0 | Affymetrix [11353280] (imputed) | N |
| 9 | 17:44118848 | 17 | 43665468 | rs2458198 | 31/05/2022 | 3.4E+07 | Smith SM | 19/04/2021 | Nat Neurosci | www.ncbi.nlm.nih.gov/pubmed/33875891 | An expanded set of genome-wide association studies of brain imaging phenotypes in UK Biobank. | aparc-Desikan lh area TotalSurface | 21,282 Bri | 10,686 Bri | 17q21.31 | | DND1P1 - MAPK8 | ENSG00000282614 | ENSG00000263503 | | 1173 | 12767 | rs2458198-G | rs2458198 | 0 | 2458198 | intron_variant | 1 | 0.228 | 3.00E-22 | 21.52288 | | 0.112 | [0.088-0.1 | Affymetrix [17103079] (imputed) | N |
| 9 | 17:44118848 | 17 | 43665468 | rs2458198 | 31/05/2022 | 3.4E+07 | Smith SM | 19/04/2021 | Nat Neurosci | www.ncbi.nlm.nih.gov/pubmed/33875891 | An expanded set of genome-wide association studies of brain imaging phenotypes in UK Biobank. | aparc-Desikan rh area TotalSurface | 21,282 Bri | 10,686 Bri | 17q21.31 | | DND1P1 - MAPK8 | ENSG00000282614 | ENSG00000263503 | | 1173 | 12767 | rs2458198-G | rs2458198 | 0 | 2458198 | intron_variant | 1 | 0.228 | 3.00E-24 | 23.52288 | | 0.118 | [0.094-0.1 | Affymetrix [17103079] (imputed) | N |
| 9 | 17:44118848 | 17 | 43665468 | rs2458198 | 31/05/2022 | 3.4E+07 | Smith SM | 19/04/2021 | Nat Neurosci | www.ncbi.nlm.nih.gov/pubmed/33875891 | An expanded set of genome-wide association studies of brain imaging phenotypes in UK Biobank. | aparc-pial rh area TotalSurface | 21,282 Bri | 10,686 Bri | 17q21.31 | | DND1P1 - MAPK8 | ENSG00000282614 | ENSG00000263503 | | 1173 | 12767 | rs2458198-G | rs2458198 | 0 | 2458198 | intron_variant | 1 | 0.228 | 1.00E-12 | 12 | | 0.082 | [0.058-0.1 | Affymetrix [17103079] (imputed) | N |
| 9 | 17:44118848 | 17 | 43665468 | rs2458198 | 31/05/2022 | 3.4E+07 | Smith SM | 19/04/2021 | Nat Neurosci | www.ncbi.nlm.nih.gov/pubmed/33875891 | An expanded set of genome-wide association studies of brain imaging phenotypes in UK Biobank. | IDP T1 SIENAX white normalised volume | 22,138 Bri | 11,086 Bri | 17q21.31 | | DND1P1 - MAPK8 | ENSG00000282614 | ENSG00000263503 | | 1173 | 12767 | rs2458198-G | rs2458198 | 0 | 2458198 | intron_variant | 1 | 0.228 | 4.00E-12 | 11.39794 | | 0.079 | [0.057-0.1 | Affymetrix [17103079] (imputed) | N |
| 9 | 17:44118848 | 17 | 43665468 | rs2458198 | 31/05/2022 | 3.4E+07 | Smith SM | 19/04/2021 | Nat Neurosci | www.ncbi.nlm.nih.gov/pubmed/33875891 | An expanded set of genome-wide association studies of brain imaging phenotypes in UK Biobank. | IDP T1 SIENAX white unnormalised volume | 22,138 Bri | 11,086 Bri | 17q21.31 | | DND1P1 - MAPK8 | ENSG00000282614 | ENSG00000263503 | | 1173 | 12767 | rs2458198-G | rs2458198 | 0 | 2458198 | intron_variant | 1 | 0.228 | 2.00E-12 | 11.69897 | | 0.08 | [0.058-0.1 | Affymetrix [17103079] (imputed) | N |
| 9 | 17:44118848 | 17 | 43671737 | rs2463523 | 13/11/2020 | 3.3E+07 | Rashkin SR | 4/09/2020 | Nat Commun | www.ncbi.nlm.nih.gov/pubmed/32887889 | Pan-cancer study detects genetic risk variants and shared genetic basis in two large cohorts. | Breast cancer | 17,881 Eu | NA | 17q21.31 | | DND1P1 - MAPK8 | ENSG00000282614 | ENSG00000263503 | | 7442 | 6498 | rs2463523-A | rs2463523 | 0 | 2463523 | regulatory_region_variant | 1 | NR | 3.00E-10 | 9.522879 | | 1.0887 | NR | Affymetrix [7846216] (imputed) | N |
| 9 | 17:44118848 | 17 | 43671737 | rs2463523 | 27/04/2022 | 3.5E+07 | Richardson TG | 25/02/2022 | PLoS Biol | www.ncbi.nlm.nih.gov/pubmed/35213538 | Characterising metabolomic signatures of lipid-modifying therapies through drug target mendelian randomisation. | Ratio of omega-6 fatty acids to omega-3 fatty acids | 115,006 E | NA | 17q21.31 | | DND1P1 - MAPK8 | ENSG00000282614 | ENSG00000263503 | | 7442 | 6498 | rs2463523-A | rs2463523 | 0 | 2463523 | regulatory_region_variant | 1 | 0.76521 | 3.00E-17 | 16.52288 | | 0.040145 | [0.031-0.0 | NR [11722792] (imputed) | N |
| 9 | 17:44118848 | 17 | 44285531 | rs2469933 | 4/10/2019 | 3.1E+07 | Brazel DM | 6/12/2018 | Biol Psychiatry | www.ncbi.nlm.nih.gov/pubmed/30679032 | Exome Chip Meta-analysis Fine Maps Causal Variants and Elucidates the Genetic Architecture of Rare Coding Variants in Smoking and Alcohol┬áUse. | Alcohol consumption (drinks per week) | up to 357,8 | NA | 17q21.31 | KIAA1267 | KANSL1, KANSL1, KANSL1 | | | ENSG00000120071, E | NA | NA | rs2469933-A | rs2469933 | 0 | 2469933 | intron_variant | 0 | 0.225 | 2.00E-22 | 21.69897 | | 0.0295 | [0.024-0.0 | Illumina [~ 250000] (imputed) | N |
| 9 | 17:44091886 | 17 | 44257783 | rs2532252 | 31/05/2022 | 3.4E+07 | Smith SM | 19/04/2021 | Nat Neurosci | www.ncbi.nlm.nih.gov/pubmed/33875891 | An expanded set of genome-wide association studies of brain imaging phenotypes in UK Biobank. | aparc-Desikan rh area TotalSurface | 21,282 Bri | 10,686 Bri | 17q21.31 | | KANSL1, KANSL1, KANSL1 | | | ENSG00000120071, E | NA | NA | rs2532252-G | rs2532252 | 0 | 2532252 | intron_variant | 0 | 0.23 | 2.00E-24 | 23.69897 | | 0.118 | [0.094-0.1 | Affymetrix [17103079] (imputed) | N |
| 9 | 17:44118848 | 17 | 44256296 | rs2532253 | 18/09/2020 | 3.3E+07 | Vuckovic D | 1/09/2020 | Cell | www.ncbi.nlm.nih.gov/pubmed/32888494 | The Polygenic and Monogenic Basis of Blood Traits and Diseases. | Red cell distribution width | 408,112 B | NA | 17q21.31 | KANSL1 | KANSL1, KANSL1, KANSL1 | | | ENSG00000120071, E | NA | NA | rs2532253-A | rs2532253 | 0 | 2532253 | intron_variant | 0 | 0.196735 | 2.00E-18 | 17.69897 | | 0.03246 | [0.025-0.0 | Affymetrix [93095623] (imputed) | N |
| 9 | 17:44118848 | 17 | 44252468 | rs2532263 | 30/11/2017 | 2.9E+07 | Michailidou K | 23/10/2017 | Nature | www.ncbi.nlm.nih.gov/pubmed/29059683 | Association analysis identifies 65 new breast cancer risk loci. | Breast cancer | 76,192 Eu | 46,785 Eu | 17q21.31 | KANSL1 | KANSL1, KANSL1, KANSL1 | | | ENSG00000120071, E | NA | NA | rs2532263-G | rs2532263 | 0 | 2532263 | intron_variant | 0 | 0.81 | 7.00E-13 | 12.1549 | (EA) | 1.052632 | [1.03-1.08 | Illumina [~ 11800000] (imputed) | N |
| 9 | 17:44118848 | 17 | 44252468 | rs2532263 | 2/05/2019 | 3.1E+07 | Hill WD | 13/03/2019 | Mol Psychiatry | www.ncbi.nlm.nih.gov/pubmed/30867560 | Genetic contributions to two special factors of neuroticism are associated with affluence, higher intelligence, better health, and longer life. | General factor of neuroticism | 270,059 B | NA | 17q21.31 | KANSL1 | KANSL1, KANSL1, KANSL1 | | | ENSG00000120071, E | NA | NA | rs2532263-G | rs2532263 | 0 | 2532263 | intron_variant | 0 | NR | 4.00E-20 | 19.39794 | | 0.015798 | [0.012-0.0 | Affymetrix [18485882] (imputed) | N |
| 9 | 17:44118848 | 17 | 44229986 | rs2532300 | 7/06/2022 | 3.5E+07 | Kousathanas A | 7/03/2022 | Nature | www.ncbi.nlm.nih.gov/pubmed/35255492 | Whole genome sequencing reveals host factors underlying critical Covid-19. | COVID-19 (critical illness vs population or mild symptoms) | 5,989 Euro | NA | 17q21.31 | | KANSL1, KANSL1, KANSL1 | | | ENSG00000120071, E | NA | NA | rs2532300-T | rs2532300 | 0 | 2532300 | intron_variant | 0 | NR | 2.00E-08 | 7.69897 | | NA | | Illumina [NR] | N |
| 9 | 17:44118848 | 17 | 44229986 | rs2532300 | 7/06/2022 | 3.5E+07 | Kousathanas A | 7/03/2022 | Nature | www.ncbi.nlm.nih.gov/pubmed/35255492 | Whole genome sequencing reveals host factors underlying critical Covid-19. | COVID-19 (critical illness vs population or mild symptoms) | 5,989 Euro | NA | 17q21.31 | | KANSL1, KANSL1, KANSL1 | | | ENSG00000120071, E | NA | NA | rs2532300-T | rs2532300 | 0 | 2532300 | intron_variant | 0 | 0.77 | 4.00E-09 | 8.39794 | | 1.2 | [1.1-1.22] | Illumina [8121457] | N |
| 9 | 17:44118848 | 17 | 44339473 | rs2532349 | 13/09/2019 | 2.6E+07 | Wain LV | 27/09/2015 | Lancet Respir Med | www.ncbi.nlm.nih.gov/pubmed/26423011 | Novel insights into the genetics of smoking behaviour, lung function, and chronic obstructive pulmonary disease (UK BiLEVE): a genetic association study in UK Biobank. | Lung function in never smokers (low FEV1 vs high FEV1) | 9,745 Euro | NA | 17q21.31 | KANSL1 | MAPK8IP1P1 - AR | ENSG00000282723 | ENSG00000228696 | | 17063 | 12677 | rs2532349-G | rs2532349 | 0 | 2532349 | intergenic_variant | 1 | 0.242 | 2.00E-10 | 9.69897 | | 1.22 | [1.15-1.29 | Affymetrix [28509962] (imputed) | N |
| 9 | 17:44118848 | 17 | 44317210 | rs2532384 | 4/08/2022 | 3.6E+07 | May-Wilson S | 18/05/2022 | Nat Commun | www.ncbi.nlm.nih.gov/pubmed/35585065 | Large-scale GWAS of food liking reveals genetic determinants and genetic correlations with distinct neurophysiological traits. | Goat cheese liking | 150,875 E | NA | 17q21.31 | | KANSL1 - MAPK8 | ENSG00000278458 | ENSG00000282723 | | 14455 | 3762 | rs2532384-A | rs2532384 | 0 | 2532384 | upstream_gene_variant | 1 | 0.2237 | 2.00E-17 | 16.69897 | | 0.03695 | [0.028-0.0 | Affymetrix [11353280] (imputed) | N |
| 9 | 17:44118848 | 17 | 44313757 | rs2532386 | 26/11/2020 | 3.3E+07 | Bi W | 23/06/2020 | Am J Hum Genet | www.ncbi.nlm.nih.gov/pubmed/32589924 | A Fast and Accurate Method for Genome-Wide Time-to-Event Data Analysis and Its Application to UK Biobank. | Osteoarthrosis (time to event) | 29,071 Bri | NA | 17q21.31 | KANSL1, LRRC37 | KANSL1 - MAPK8 | ENSG00000278458 | ENSG00000282723 | | 11002 | 7215 | rs2532386-? | rs2532386 | 0 | 2532386 | regulatory_region_variant | 1 | NR | 4.00E-08 | 7.39794 | | NA | | NR [~ 24000000] (imputed) | N |
| 9 | 17:44118848 | 17 | 44313522 | rs2532387 | 4/08/2022 | 3.6E+07 | May-Wilson S | 18/05/2022 | Nat Commun | www.ncbi.nlm.nih.gov/pubmed/35585065 | Large-scale GWAS of food liking reveals genetic determinants and genetic correlations with distinct neurophysiological traits. | F-sharp flavour liking (derived food-liking factor) | 152,547 E | NA | 17q21.31 | | KANSL1 - MAPK8 | ENSG00000278458 | ENSG00000282723 | | 10767 | 7450 | rs2532387-T | rs2532387 | 0 | 2532387 | regulatory_region_variant | 1 | 0.2169 | 5.00E-25 | 24.30103 | | 0.11162 | [0.09-0.13 | Affymetrix [9605914] (imputed) | N |
| 9 | 17:44118848 | 17 | 44313522 | rs2532387 | 13/12/2021 | 3.4E+07 | Kachuri L | 26/08/2021 | Am J Hum Genet | www.ncbi.nlm.nih.gov/pubmed/34469753 | Genetic determinants of blood-cell traits influence susceptibility to childhood acute lymphoblastic leukemia. | Lymphocyte-to-monocyte ratio | 234,184 E | 100,284 E | 17q21.31 | | KANSL1 - MAPK8 | ENSG00000278458 | ENSG00000282723 | | 10767 | 7450 | rs2532387-G | rs2532387 | 0 | 2532387 | regulatory_region_variant | 1 | 0.781 | 1.00E-19 | 19 | | NA | | Affymetrix [10369434] (imputed) | N |
| 9 | 17:44118848 | 17 | 44311107 | rs2532389 | 13/11/2020 | 3.3E+07 | Rashkin SR | 4/09/2020 | Nat Commun | www.ncbi.nlm.nih.gov/pubmed/32887889 | Pan-cancer study detects genetic risk variants and shared genetic basis in two large cohorts. | Cancer (pleiotropy) | 64,962 Eu | NA | 17q21.31 | | KANSL1 - MAPK8 | ENSG00000278458 | ENSG00000282723 | | 8352 | 9865 | rs2532389-G | rs2532389 | 0 | 2532389 | intergenic_variant | 1 | NR | 1.00E-09 | 9 | | 1.074 | [1.05-1.09 | Affymetrix [7846216] (imputed) | N |
| 9 | 17:44118848 | 17 | 44307193 | rs2532395 | 4/08/2022 | 3.6E+07 | May-Wilson S | 18/05/2022 | Nat Commun | www.ncbi.nlm.nih.gov/pubmed/35585065 | Large-scale GWAS of food liking reveals genetic determinants and genetic correlations with distinct neurophysiological traits. | Brussel sprout liking | 159,493 E | NA | 17q21.31 | | KANSL1 - MAPK8 | ENSG00000278458 | ENSG00000282723 | | 4438 | 13779 | rs2532395-T | rs2532395 | 0 | 2532395 | upstream_gene_variant | 1 | 0.2126 | 1.00E-22 | 22 | | 0.04292 | [0.034-0.0 | Affymetrix [11353280] (imputed) | N |
| 9 | 17:44118848 | 17 | 44304130 | rs2532402 | 4/08/2022 | 3.6E+07 | May-Wilson S | 18/05/2022 | Nat Commun | www.ncbi.nlm.nih.gov/pubmed/35585065 | Large-scale GWAS of food liking reveals genetic determinants and genetic correlations with distinct neurophysiological traits. | Horseradish liking | 152,847 E | NA | 17q21.31 | | KANSL1 - MAPK8 | ENSG00000278458 | ENSG00000282723 | | 1375 | 16842 | rs2532402-G | rs2532402 | 0 | 2532402 | upstream_gene_variant | 1 | 0.2226 | 1.00E-14 | 14 | | 0.03378 | [0.025-0.0 | Affymetrix [11353280] (imputed) | N |

| | | | | | | | | | | | | | | | | | | | | | | | | | | | | | | | | | | | | |
|---|---|---|---|---|---|---|---|---|---|---|---|---|---|---|---|---|---|---|---|---|---|---|---|---|---|---|---|---|---|---|---|---|---|---|---|---|
| 9 | 17:44118848 | 17 | 44304130 | rs2532402 | 3/02/2022 | 3.5E+07 | Qi G | 18/01/2022 | Genet Epidemiol | www.ncbi.nlm.nih.gov/pubmed/35043453 | Genome-wide association studies of 27 accelerometry-derived physical activity measurements identified novel loci and genetic mechanisms. | Moderate to vigorous physical activity duration | 88,411 Eur | NA | 17q21.31 | | KANSL1 - MAPK8I | ENSG00000278458 | ENSG00000282723 | | 1375 | 16842 | rs2532402-G | rs2532402 | 0 | 2532402 | upstream_gene_variant | 1 | 0.221 | 2.00E-10 | 9.69897 | | 0.035 | unit increa | Affymetrix [8951705] (imputed) | N |
| 9 | 17:44118848 | 17 | 44304130 | rs2532402 | 3/02/2022 | 3.5E+07 | Qi G | 18/01/2022 | Genet Epidemiol | www.ncbi.nlm.nih.gov/pubmed/35043453 | Genome-wide association studies of 27 accelerometry-derived physical activity measurements identified novel loci and genetic mechanisms. | Physical activity (Total log acceleration) | 88,411 Eur | NA | 17q21.31 | | KANSL1 - MAPK8I | ENSG00000278458 | ENSG00000282723 | | 1375 | 16842 | rs2532402-G | rs2532402 | 0 | 2532402 | upstream_gene_variant | 1 | 0.221 | 2.00E-12 | 11.69897 | | 0.04 | unit increa | Affymetrix [8951705] (imputed) | N |
| 9 | 17:44091886 | 17 | 44324539 | rs2668639 | 31/05/2022 | 3.4E+07 | Smith SM | 19/04/2021 | Nat Neurosci | www.ncbi.nlm.nih.gov/pubmed/33875891 | An expanded set of genome-wide association studies of brain imaging phenotypes in UK Biobank. | IDP dMRI ProbtrackX L1 slf r | 20,860 Brit | 10,495 Brit | 17q21.31 | | MAPK8IP1P1 - AR | ENSG00000282723 | ENSG00000228696 | | 2129 | 27611 | rs2668639-C | rs2668639 | 0 | 2668639 | intergenic_variant | 1 | 0.773 | 3.00E-13 | 12.52288 | | 0.085 | [0.061-0.1 | Affymetrix [17103079] (imputed) | N |
| 9 | 17:44091886 | 17 | 44324539 | rs2668639 | 31/05/2022 | 3.4E+07 | Smith SM | 19/04/2021 | Nat Neurosci | www.ncbi.nlm.nih.gov/pubmed/33875891 | An expanded set of genome-wide association studies of brain imaging phenotypes in UK Biobank. | IDP dMRI TBSS L1 Superior longitudinal fasciculus R | 20,860 Brit | 10,496 Brit | 17q21.31 | | MAPK8IP1P1 - AR | ENSG00000282723 | ENSG00000228696 | | 2129 | 27611 | rs2668639-C | rs2668639 | 0 | 2668639 | intergenic_variant | 1 | 0.773 | 3.00E-15 | 14.52288 | | 0.092 | [0.068-0.1 | Affymetrix [17103079] (imputed) | N |
| 9 | 17:44118848 | 17 | 43657440 | rs2668674 | 5/03/2021 | 3.3E+07 | Tadros R | 25/01/2021 | Nat Genet | www.ncbi.nlm.nih.gov/pubmed/33495596 | Shared genetic pathways contribute to risk of hypertrophic and dilated cardiomyopathies with opposite directions of effect. | Left ventricle wall thickness | 19,260 Eur | NA | 17q21.31 | ARL17A | RDM1P1 - DND1F | ENSG00000266504 | ENSG00000282614 | | 21004 | 5797 | rs2668674-G | rs2668674 | 0 | 2668674 | intron_variant | 1 | 0.1991 | 2.00E-11 | 10.69897 | | 0.0418 | [0.03-0.05 | Affymetrix [~ 9500000] (imputed) | N |
| 9 | 17:44118848 | 17 | 43657440 | rs2668674 | 5/03/2021 | 3.3E+07 | Tadros R | 25/01/2021 | Nat Genet | www.ncbi.nlm.nih.gov/pubmed/33495596 | Shared genetic pathways contribute to risk of hypertrophic and dilated cardiomyopathies with opposite directions of effect. | Left ventricular mass | 19,260 Eur | NA | 17q21.31 | ARL17A | RDM1P1 - DND1F | ENSG00000266504 | ENSG00000282614 | | 21004 | 5797 | rs2668674-G | rs2668674 | 0 | 2668674 | intron_variant | 1 | 0.1991 | 2.00E-06 | 5.69897 | | 0.7099 | [0.42-1] un | Affymetrix [~ 9500000] (imputed) | N |
| 9 | 17:44118848 | 17 | 43657440 | rs2668674 | 5/03/2021 | 3.3E+07 | Tadros R | 25/01/2021 | Nat Genet | www.ncbi.nlm.nih.gov/pubmed/33495596 | Shared genetic pathways contribute to risk of hypertrophic and dilated cardiomyopathies with opposite directions of effect. | Left ventricular mass to end-diastolic volume ratio | 19,260 Eur | NA | 17q21.31 | ARL17A | RDM1P1 - DND1F | ENSG00000266504 | ENSG00000282614 | | 21004 | 5797 | rs2668674-G | rs2668674 | 0 | 2668674 | intron_variant | 1 | 0.1991 | 4.00E-07 | 6.39794 | | 0.0048 | [0.003-0.0 | Affymetrix [~ 9500000] (imputed) | N |
| 9 | 17:44118848 | 17 | 43659975 | rs2668680 | 28/03/2019 | 3.1E+07 | Liu M | 14/01/2019 | Nat Genet | www.ncbi.nlm.nih.gov/pubmed/30643251 | Association studies of up to 1.2 million individuals yield new insights into the genetic etiology of tobacco and alcohol use. | Alcohol consumption (drinks per week) (MTAG) | up to 1,039 | NA | 17q21.31 | NR | RDM1P1 - DND1F | ENSG00000266504 | ENSG00000282614 | | 23539 | 3262 | rs2668680-C | rs2668680 | 0 | 2668680 | intron_variant | 1 | 0.177 | 4.00E-19 | 18.39794 | | 0.017489 | [0.014-0.0 | NR [~ 9732723] (imputed) | N |
| 9 | 17:44118848 | 17 | 44287373 | rs2668690 | 27/04/2022 | 3.5E+07 | Richardson TG | 25/02/2022 | PLoS Biol | www.ncbi.nlm.nih.gov/pubmed/35213538 | Characterising metabolomic signatures of lipid-modifying therapies through drug target mendelian randomisation. | Phospholipids to total lipids ratio in small HDL | 115,082 Eu | NA | 17q21.31 | | KANSL1, KANSL1, KANSL1 | | | ENSG00000120071, E | NA | NA | rs2668690-G | rs2668690 | 0 | 2668690 | intron_variant | 0 | 0.808457 | 1.00E-10 | 10 | | 0.034313 | [0.024-0.0 | NR [11722792] (imputed) | N |
| 9 | 17:44118848 | 17 | 44287373 | rs2668690 | 27/04/2022 | 3.5E+07 | Richardson TG | 25/02/2022 | PLoS Biol | www.ncbi.nlm.nih.gov/pubmed/35213538 | Characterising metabolomic signatures of lipid-modifying therapies through drug target mendelian randomisation. | Phospholipids to total lipids ratio in very large HDL | 115,056 Eu | NA | 17q21.31 | | KANSL1, KANSL1, KANSL1 | | | ENSG00000120071, E | NA | NA | rs2668690-G | rs2668690 | 0 | 2668690 | intron_variant | 0 | 0.808461 | 3.00E-08 | 7.522879 | | 0.028132 | [0.018-0.0 | NR [11722792] (imputed) | N |
| 9 | 17:43520118 | 17 | 43520118 | rs2684641 | 26/10/2022 | 3.6E+07 | Hatoum AS | 14/07/2022 | Biol Psychiatry | www.ncbi.nlm.nih.gov/pubmed/36150907 | Genome-wide Association Study Shows That Executive Functioning Is Influenced by GABAergic Processes and Is a Neurocognitive Genetic Correlate of Psychiatric Disorders. | Common executive function | 427,037 Eu | NA | 17q21.31 | | PLEKHM1 | | | ENSG00000277111 | NA | NA | rs2684641-? | rs2684641 | 0 | 2684641 | intron_variant | 0 | NR | 1.00E-10 | 10 | | NA | | Affymetrix [7391068] (imputed) | N |
| 9 | 17:44118848 | 17 | 44333889 | rs2696422 | 31/05/2022 | 3.4E+07 | Smith SM | 19/04/2021 | Nat Neurosci | www.ncbi.nlm.nih.gov/pubmed/33875891 | An expanded set of genome-wide association studies of brain imaging phenotypes in UK Biobank. | IDP SWI T2star left putamen | 20,043 Brit | 10,013 Brit | 17q21.31 | | MAPK8IP1P1 - AR | ENSG00000282723 | ENSG00000228696 | | 11479 | 18261 | rs2696422-C | rs2696422 | 0 | 2696422 | intergenic_variant | 1 | 0.221 | 1.00E-15 | 15 | | 0.096 | [0.072-0.1 | Affymetrix [17103079] (imputed) | N |
| 9 | 17:44118848 | 17 | 44283571 | rs2696455 | 27/04/2022 | 3.5E+07 | Richardson TG | 25/02/2022 | PLoS Biol | www.ncbi.nlm.nih.gov/pubmed/35213538 | Characterising metabolomic signatures of lipid-modifying therapies through drug target mendelian randomisation. | Average diameter for HDL particles | 115,082 Eu | NA | 17q21.31 | | KANSL1, KANSL1, KANSL1 | | | ENSG00000120071, E | NA | NA | rs2696455-C | rs2696455 | 0 | 2696455 | intron_variant | 0 | 0.804129 | 4.00E-08 | 7.39794 | | 0.026668 | [0.017-0.0 | NR [11722792] (imputed) | N |
| 9 | 17:44118848 | 17 | 44283571 | rs2696455 | 31/05/2022 | 3.4E+07 | Smith SM | 19/04/2021 | Nat Neurosci | www.ncbi.nlm.nih.gov/pubmed/33875891 | An expanded set of genome-wide association studies of brain imaging phenotypes in UK Biobank. | IDP dMRI TBSS MD Anterior limb of internal capsule L | 20,860 Brit | 10,496 Brit | 17q21.31 | | KANSL1, KANSL1, KANSL1 | | | ENSG00000120071, E | NA | NA | rs2696455-T | rs2696455 | 0 | 2696455 | intron_variant | 0 | 0.195 | 1.00E-18 | 18 | | 0.114 | [0.089-0.1 | Affymetrix [17103079] (imputed) | N |
| 9 | 17:44118848 | 17 | 44284057 | rs2696457 | 22/01/2019 | 3E+07 | Nagel M | 2/03/2018 | Nat Commun | www.ncbi.nlm.nih.gov/pubmed/29500382 | Item-level analyses reveal genetic heterogeneity in neuroticism. | Feeling worry | 372,869 Eu | NA | 17q21.31 | NR | KANSL1, KANSL1, KANSL1 | | | ENSG00000120071, E | NA | NA | rs2696457-T | rs2696457 | 0 | 2696457 | intron_variant | 0 | 0.215657 | 6.00E-10 | 9.221849 | | 6.186 | unit increa | NR [10847151] (imputed) | N |
| 9 | 17:44118848 | 17 | 44364649 | rs2696497 | 22/01/2019 | 3E+07 | Nagel M | 2/03/2018 | Nat Commun | www.ncbi.nlm.nih.gov/pubmed/29500382 | Item-level analyses reveal genetic heterogeneity in neuroticism. | Feeling fed-up | 374,971 Eu | NA | 17q21.31 | NR | ARL17B, ARL17B, ARL17B | | | ENSG00000275748, E | NA | NA | rs2696497-T | rs2696497 | 0 | 2696497 | intron_variant | 0 | 0.21384 | 2.00E-15 | 14.69897 | | 7.92 | z score inc | NR [10847151] (imputed) | N |
| 9 | 17:44118848 | 17 | 44363929 | rs2696500 | 22/01/2019 | 3E+07 | Nagel M | 2/03/2018 | Nat Commun | www.ncbi.nlm.nih.gov/pubmed/29500382 | Item-level analyses reveal genetic heterogeneity in neuroticism. | Neurociticism | 380,506 Eu | NA | 17q21.31 | NR | ARL17B, ARL17B, ARL17B | | | ENSG00000275748, E | NA | NA | rs2696500-T | rs2696500 | 0 | 2696500 | 3_prime_UTR_variant | 0 | 0.214048 | 4.00E-29 | 28.39794 | | 11.21 | z score inc | NR [10847151] (imputed) | N |
| 9 | 17:44118848 | 17 | 44357885 | rs2696518 | 31/05/2022 | 3.4E+07 | Smith SM | 19/04/2021 | Nat Neurosci | www.ncbi.nlm.nih.gov/pubmed/33875891 | An expanded set of genome-wide association studies of brain imaging phenotypes in UK Biobank. | IDP dMRI ProbtrackX L1 ptr l | 20,860 Brit | 10,495 Brit | 17q21.31 | | ARL17B, ARL17B, ARL17B | | | ENSG00000275748, E | NA | NA | rs2696518-G | rs2696518 | 0 | 2696518 | intron_variant | 0 | 0.204 | 1.00E-09 | 9 | | 0.077 | [0.052-0.1 | Affymetrix [17103079] (imputed) | N |
| 9 | 17:44118848 | 17 | 44357885 | rs2696518 | 31/05/2022 | 3.4E+07 | Smith SM | 19/04/2021 | Nat Neurosci | www.ncbi.nlm.nih.gov/pubmed/33875891 | An expanded set of genome-wide association studies of brain imaging phenotypes in UK Biobank. | IDP dMRI ProbtrackX L1 str r | 20,860 Brit | 10,495 Brit | 17q21.31 | | ARL17B, ARL17B, ARL17B | | | ENSG00000275748, E | NA | NA | rs2696518-G | rs2696518 | 0 | 2696518 | intron_variant | 0 | 0.204 | 2.00E-13 | 12.69897 | | 0.093 | [0.068-0.1 | Affymetrix [17103079] (imputed) | N |
| 9 | 17:44118848 | 17 | 44356793 | rs2696524 | 5/02/2020 | 3.2E+07 | Zhao B | 30/10/2019 | Mol Psychiatry | www.ncbi.nlm.nih.gov/pubmed/31666681 | Large-scale GWAS reveals genetic architecture of brain white matter microstructure and genetic overlap with cognitive and mental health traits (nÔÇë=ÔÇë17,706). | White matter microstructure (fractional anisotropy) | 17,706 Eur | NA | 17q21.31 | NR | ARL17B, ARL17B, ARL17B | | | ENSG00000275748, E | NA | NA | rs2696524-? | rs2696524 | 0 | 2696524 | intron_variant | 0 | | 5.00E-11 | 10.30103 | (Fornix (cr | NA | | NR [8955960] (imputed) | N |
| 9 | 17:44118848 | 17 | 44355602 | rs2696532 | 22/01/2019 | 3E+07 | Nagel M | 2/03/2018 | Nat Commun | www.ncbi.nlm.nih.gov/pubmed/29500382 | Item-level analyses reveal genetic heterogeneity in neuroticism. | Feeling guilty | 373,380 Eu | NA | 17q21.31 | NR | ARL17B, ARL17B, ARL17B | | | ENSG00000275748, E | NA | NA | rs2696532-A | rs2696532 | 0 | 2696532 | intron_variant | 0 | 0.212157 | 4.00E-08 | 7.39794 | | 5.49 | z score dec | NR [10847151] (imputed) | N |
| 9 | 17:44118848 | 17 | 44347218 | rs2696557 | 22/01/2019 | 3E+07 | Nagel M | 2/03/2018 | Nat Commun | www.ncbi.nlm.nih.gov/pubmed/29500382 | Item-level analyses reveal genetic heterogeneity in neuroticism. | Feeling nervous | 373,121 Eu | NA | 17q21.31 | NR | MAPK8IP1P1 - AR | ENSG00000282723 | ENSG00000228696 | | 24808 | 4932 | rs2696557-A | rs2696557 | 0 | 2696557 | intergenic_variant | 1 | 0.214201 | 3.00E-15 | 14.52288 | | 7.91 | z score inc | NR [10847151] (imputed) | N |
| 9 | 17:44118848 | 17 | 44347218 | rs2696557 | 31/05/2022 | 3.4E+07 | Smith SM | 19/04/2021 | Nat Neurosci | www.ncbi.nlm.nih.gov/pubmed/33875891 | An expanded set of genome-wide association studies of brain imaging phenotypes in UK Biobank. | IDP dMRI TBSS ICVF Superior fronto-occipital fasciculus R | 20,859 Brit | 10,496 Brit | 17q21.31 | | MAPK8IP1P1 - AR | ENSG00000282723 | ENSG00000228696 | | 24808 | 4932 | rs2696557-A | rs2696557 | 0 | 2696557 | intergenic_variant | 1 | 0.221 | 8.00E-27 | 26.09691 | | 0.128 | [0.1-0.15] ( | Affymetrix [17103079] (imputed) | N |
| 9 | 17:44118848 | 17 | 44221836 | rs2696589 | 1/02/2018 | 2.9E+07 | Luciano M | 18/12/2017 | Nat Genet | www.ncbi.nlm.nih.gov/pubmed/29255261 | Association analysis in over 329,000 individuals identifies 116 independent variants influencing neuroticism. | Neuroticism | 329,821 Eu | 122,867 Eu | 17q21.31 | intergenic | KANSL1, KANSL1, KANSL1 | | | ENSG00000120071, E | NA | NA | rs2696589-T | rs2696589 | 0 | 2696589 | intron_variant | 0 | 0.19528 | 3.00E-25 | 24.52288 | | 10.371 | unit increa | Illumina [18485882] (imputed) | N |
| 9 | 17:44118848 | 17 | 44318663 | rs2696591 | 4/08/2022 | 3.6E+07 | Fan CC | 3/05/2022 | Nat Commun | www.ncbi.nlm.nih.gov/pubmed/35505052 | Multivariate genome-wide association study on tissue-sensitive diffusion metrics highlights pathways that shape the human brain. | Whole brain restricted directional diffusion (multivariate analysis) | 23,543 Eur | 11,555 Eur | 17q21.31 | | KANSL1 - MAPK8I | ENSG00000278458 | ENSG00000282723 | | 15908 | 2309 | rs2696591-? | rs2696591 | 0 | 2696591 | upstream_gene_variant | 1 | NR | 8.00E-96 | 95.09691 | | NA | | NR [NR] (imputed) | N |
| 9 | 17:44091886 | 17 | 44318863 | rs2696606 | 31/05/2022 | 3.4E+07 | Smith SM | 19/04/2021 | Nat Neurosci | www.ncbi.nlm.nih.gov/pubmed/33875891 | An expanded set of genome-wide association studies of brain imaging phenotypes in UK Biobank. | aseg global volume BrainSegNotVentSurf | 21,282 Brit | 10,686 Brit | 17q21.31 | | KANSL1 - MAPK8I | ENSG00000278458 | ENSG00000282723 | | 16108 | 2109 | rs2696606-C | rs2696606 | 0 | 2696606 | intergenic_variant | 1 | 0.228 | 5.00E-14 | 13.30103 | | 0.087 | [0.063-0.1 | Affymetrix [17103079] (imputed) | N |
| 9 | 17:44091886 | 17 | 44318863 | rs2696606 | 31/05/2022 | 3.4E+07 | Smith SM | 19/04/2021 | Nat Neurosci | www.ncbi.nlm.nih.gov/pubmed/33875891 | An expanded set of genome-wide association studies of brain imaging phenotypes in UK Biobank. | aseg global volume SupraTentorialNotVent | 21,282 Brit | 10,686 Brit | 17q21.31 | | KANSL1 - MAPK8I | ENSG00000278458 | ENSG00000282723 | | 16108 | 2109 | rs2696606-C | rs2696606 | 0 | 2696606 | intergenic_variant | 1 | 0.228 | 1.00E-12 | 12 | | 0.082 | [0.058-0.1 | Affymetrix [17103079] (imputed) | N |
| 9 | 17:44091886 | 17 | 44318863 | rs2696606 | 31/05/2022 | 3.4E+07 | Smith SM | 19/04/2021 | Nat Neurosci | www.ncbi.nlm.nih.gov/pubmed/33875891 | An expanded set of genome-wide association studies of brain imaging phenotypes in UK Biobank. | BA-exvivo rh area V2 | 21,281 Brit | 10,686 Brit | 17q21.31 | | KANSL1 - MAPK8I | ENSG00000278458 | ENSG00000282723 | | 16108 | 2109 | rs2696606-C | rs2696606 | 0 | 2696606 | intergenic_variant | 1 | 0.228 | 2.00E-16 | 15.69897 | | 0.095 | [0.071-0.1 | Affymetrix [17103079] (imputed) | N |
| 9 | 17:44091886 | 17 | 44325635 | rs2696618 | 31/05/2022 | 3.4E+07 | Smith SM | 19/04/2021 | Nat Neurosci | www.ncbi.nlm.nih.gov/pubmed/33875891 | An expanded set of genome-wide association studies of brain imaging phenotypes in UK Biobank. | aparc-DKTatlas lh area fusiform | 21,282 Brit | 10,686 Brit | 17q21.31 | | MAPK8IP1P1 - AR | ENSG00000282723 | ENSG00000228696 | | 3225 | 26515 | rs2696618-G | rs2696618 | 0 | 2696618 | intergenic_variant | 1 | 0.772 | 6.00E-24 | 23.22185 | | 0.117 | [0.093-0.1 | Affymetrix [17103079] (imputed) | N |
| 9 | 17:44118848 | 17 | 44326864 | rs2696625 | 1/02/2022 | 3.5E+07 | Hanscombe KB | 9/11/2021 | Genome Med | www.ncbi.nlm.nih.gov/pubmed/34753499 | The genetic case for cardiorespiratory fitness as a clinical vital sign and the routine prescription of physical activity in healthcare. | Physical activity | 89,683 Eur | NA | 17q21.31 | | MAPK8IP1P1 - AR | ENSG00000282723 | ENSG00000228696 | | 4454 | 25286 | rs2696625-A | rs2696625 | 0 | 2696625 | intergenic_variant | 1 | 0.77 | 5.00E-12 | 11.30103 | | 0.31 | [0.22-0.4] ( | Affymetrix [8918919] (imputed) | N |
| 9 | 17:44118848 | 17 | 44326864 | rs2696625 | 18/01/2019 | 3.1E+07 | Doherty A | 10/12/2018 | Nat Commun | www.ncbi.nlm.nih.gov/pubmed/30531941 | GWAS identifies 14 loci for device-measured physical activity and sleep duration. | Physical activity (overall physical activity time) | 91,105 Eur | NA | 17q21.31 | KANSL1-AS1 | MAPK8IP1P1 - AR | ENSG00000282723 | ENSG00000228696 | | 4454 | 25286 | rs2696625-A | rs2696625 | 0 | 2696625 | intergenic_variant | 1 | 0.77 | 3.00E-12 | 11.52288 | | 0.037 | [0.027-0.0 | NR [9926106] (imputed) | N |
| 9 | 17:44118848 | 17 | 44327482 | rs2696630 | 31/05/2022 | 3.4E+07 | Smith SM | 19/04/2021 | Nat Neurosci | www.ncbi.nlm.nih.gov/pubmed/33875891 | An expanded set of genome-wide association studies of brain imaging phenotypes in UK Biobank. | aparc-Desikan lh area fusiform | 21,282 Brit | 10,686 Brit | 17q21.31 | | MAPK8IP1P1 - AR | ENSG00000282723 | ENSG00000228696 | | 5072 | 24668 | rs2696630-A | rs2696630 | 0 | 2696630 | intergenic_variant | 1 | 0.229 | 1.00E-23 | 23 | | 0.116 | [0.092-0.1 | Affymetrix [17103079] (imputed) | N |
| 9 | 17:44118848 | 17 | 44245766 | rs2696658 | 23/10/2020 | 3.3E+07 | van der Meer D | 14/07/2020 | Nat Commun | www.ncbi.nlm.nih.gov/pubmed/32665545 | Understanding the genetic determinants of the brain with MOSTest. | Brain morphology (MOSTest) | 26,502 Eur | NA | 17q21.31 | KIF18B, C1QL1, D | KANSL1, KANSL1, KANSL1 | | | ENSG00000120071, E | NA | NA | rs2696658-? | rs2696658 | 0 | 2696658 | intron_variant | 0 | | 2.00E-65 | 64.69897 | | NA | | NR [7428630] (imputed) | N |
| 9 | 17:44118848 | 17 | 44318184 | rs2696668 | 22/01/2019 | 3E+07 | Nagel M | 2/03/2018 | Nat Commun | www.ncbi.nlm.nih.gov/pubmed/29500382 | Item-level analyses reveal genetic heterogeneity in neuroticism. | Feeling hurt | 372,047 Eu | NA | 17q21.31 | NR | KANSL1 - MAPK8I | ENSG00000278458 | ENSG00000282723 | | 15429 | 2788 | rs2696668-A | rs2696668 | 0 | 2696668 | intergenic_variant | 1 | 0.215905 | 1.00E-26 | 26 | | 10.68 | z score dec | NR [10847151] (imputed) | N |
| 9 | 17:44118848 | 17 | 44310101 | rs2696682 | 23/11/2022 | 3.5E+07 | van der Meer D | 11/02/2022 | Biol Psychiatry | www.ncbi.nlm.nih.gov/pubmed/35164939 | Boosting Schizophrenia Genetics by Utilizing Genetic Overlap With Brain Morphology. | Brain morphology (MOSTest) | 33,735 wh | NA | 17q21.31 | | KANSL1 - MAPK8I | ENSG00000278458 | ENSG00000282723 | | 7346 | 10871 | rs2696682-? | rs2696682 | 0 | 2696682 | intergenic_variant | 1 | NR | ####### | 122 | | NA | | NR [9061072] (imputed) | N |
| 9 | 17:44118848 | 17 | 44257788 | rs2696697 | 18/08/2022 | 3.6E+07 | Gouveia C | 19/05/2022 | Sci Rep | www.ncbi.nlm.nih.gov/pubmed/35589863 | Genome-wide association of polygenic risk extremes for Alzheimer's disease in the UK Biobank. | AlzheimerÔÇÖs disease polygenic risk score (upper quantile vs lower quantile) | 18,892 Eur | NA | 17q21.31 | | KANSL1, KANSL1, KANSL1 | | | ENSG00000120071, E | NA | NA | rs2696697-? | rs2696697 | 0 | 2696697 | intron_variant | 0 | NR | 5.00E-16 | 15.30103 | | 0.87 | [0.84-0.9] | NR [NR] | N |
| 9 | 17:44118848 | 17 | 44269546 | rs2732596 | 29/01/2020 | 3.2E+07 | Hill WD | 16/12/2019 | Nat Commun | www.ncbi.nlm.nih.gov/pubmed/31844048 | Genome-wide analysis identifies molecular systems and 149 genetic loci associated with income. | Household income | 286,301 Bi | NA | 17q21.31 | KANSL1 | KANSL1, KANSL1, KANSL1 | | | ENSG00000120071, E | NA | NA | rs2732596-T | rs2732596 | 0 | 2732596 | intron_variant | 0 | NR | 5.00E-11 | 10.30103 | | 0.013165 | [0.0092-0. | Affymetrix [18485882] (imputed) | N |
| 9 | 17:44118848 | 17 | 44364209 | rs2732614 | 27/09/2016 | 2.7E+07 | Dong J | 1/11/2015 | Medicine (Baltimore) | www.ncbi.nlm.nih.gov/pubmed/26632684 | Genome-wide Meta-analysis on the Sense of Smell Among US Older Adults. | Sense of smell | 6,252 Euro | NA | 17q21.31 | KIAA1267, LRRC3 | ARL17B, ARL17B, ARL17B | | | ENSG00000275748, E | NA | NA | rs2732614-T | rs2732614 | 0 | 2732614 | 3_prime_UTR_variant | 0 | 0.2 | 7.00E-06 | 5.154902 | (Ajusted fo | 0.03 | [0.01-0.05 | Affymetrix, Illumina [2543887] (imputed) | N |
| 9 | 17:44118848 | 17 | 44364209 | rs2732614 | 27/09/2016 | 2.7E+07 | Dong J | 1/11/2015 | Medicine (Baltimore) | www.ncbi.nlm.nih.gov/pubmed/26632684 | Genome-wide Meta-analysis on the Sense of Smell Among US Older Adults. | Sense of smell | 6,252 Euro | NA | 17q21.31 | KIAA1267, LRRC3 | ARL17B, ARL17B, ARL17B | | | ENSG00000275748, E | NA | NA | rs2732614-T | rs2732614 | 0 | 2732614 | 3_prime_UTR_variant | 0 | 0.2 | 8.00E-06 | 5.09691 | | 0.03 | [0.01-0.05 | Affymetrix, Illumina [2543887] (imputed) | N |
| 9 | 17:44118848 | 17 | 44289232 | rs2732631 | 28/01/2019 | 3.1E+07 | Gao XR | 7/12/2018 | Hum Mol Genet | www.ncbi.nlm.nih.gov/pubmed/30535121 | Genome-wide association analyses identify 139 loci associated with macular thickness in the UK Biobank cohort. | Macular thickness | 59,814 Eur | 8,609 Euro | 17q21.31 | KANSL1 | KANSL1, KANSL1, KANSL1 | | | ENSG00000120071, E | NA | NA | rs2732631-? | rs2732631 | 0 | 2732631 | intron_variant | 0 | NR | 3.00E-35 | 34.52288 | | 0.959 | [0.8-1.12] ( | Affymetrix [11100000] (imputed) | N |
| 9 | 17:44118848 | 17 | 44344988 | rs2732650 | 31/05/2022 | 3.4E+07 | Smith SM | 19/04/2021 | Nat Neurosci | www.ncbi.nlm.nih.gov/pubmed/33875891 | An expanded set of genome-wide association studies of brain imaging phenotypes in UK Biobank. | IDP dMRI TBSS MD Anterior limb of internal capsule R | 20,860 Brit | 10,496 Brit | 17q21.31 | | MAPK8IP1P1 - AR | ENSG00000282723 | ENSG00000228696 | | 22578 | 7162 | rs2732650-G | rs2732650 | 0 | 2732650 | intron_variant | 1 | 0.224 | 1.00E-14 | 14 | | 0.092 | [0.068-0.1 | Affymetrix [17103079] (imputed) | N |
| 9 | 17:44118848 | 17 | 44344988 | rs2732650 | 31/05/2022 | 3.4E+07 | Smith SM | 19/04/2021 | Nat Neurosci | www.ncbi.nlm.nih.gov/pubmed/33875891 | An expanded set of genome-wide association studies of brain imaging phenotypes in UK Biobank. | IDP dMRI TBSS MD Cingulum cingulate gyrus L | 20,860 Brit | 10,496 Brit | 17q21.31 | | MAPK8IP1P1 - AR | ENSG00000282723 | ENSG00000228696 | | 22578 | 7162 | rs2732650-G | rs2732650 | 0 | 2732650 | intron_variant | 1 | 0.224 | 3.00E-10 | 9.522879 | | 0.075 | [0.051-0.0 | Affymetrix [17103079] (imputed) | N |
| 9 | 17:44118848 | 17 | 44344988 | rs2732650 | 21/01/2020 | 3.2E+07 | Abdellaoui A | 13/09/2019 | Hum Mol Genet | www.ncbi.nlm.nih.gov/pubmed/31518406 | Phenome-wide investigation of health outcomes associated with genetic predisposition to loneliness. | Loneliness | 511,280 Eu | NA | 17q21.31 | RP11-259G18.1 | MAPK8IP1P1 - AR | ENSG00000282723 | ENSG00000228696 | | 22578 | 7162 | rs2732650-? | rs2732650 | 0 | 2732650 | intron_variant | 1 | NR | 3.00E-08 | 7.522879 | | 0.02 | [0.012-0.0 | Affymetrix, Illumina [up to 14113458] (imputed) | N |
| 9 | 17:44118848 | 17 | 44346407 | rs2732655 | 5/02/2020 | 3.2E+07 | Zhao B | 30/10/2019 | Mol Psychiatry | www.ncbi.nlm.nih.gov/pubmed/31666681 | Large-scale GWAS reveals genetic architecture of brain white matter microstructure and genetic overlap with cognitive and mental health traits (nÔÇë=ÔÇë17,706). | White matter microstructure (fractional anisotropy) | 17,706 Eur | NA | 17q21.31 | NR | MAPK8IP1P1 - AR | ENSG00000282723 | ENSG00000228696 | | 23997 | 5743 | rs2732655-? | rs2732655 | 0 | 2732655 | intergenic_variant | 1 | | 1.00E-08 | 8 | (Genu of co | NA | | NR [8955960] (imputed) | N |

| | | | | | | | | | | | | | | | | | | | | | | | | | | | | | | | | | | | | |
|---|---|---|---|---|---|---|---|---|---|---|---|---|---|---|---|---|---|---|---|---|---|---|---|---|---|---|---|---|---|---|---|---|---|---|---|---|
| 9 | 17:44118848 | 17 | 44338503 | rs2732689 | 4/08/2022 | 3.6E+07 | May-Wilson S | 18/05/2022 | Nat Commun | www.ncbi.nlm.nih.gov/pubmed/35585065 | Large-scale GWAS of food liking reveals genetic determinants and genetic correlations with distinct neurophysiological traits. | F-acquired taste liking (derived food-liking factor) | 156,639 Eu | NA | 17q21.31 | | MAPK8IP1P1 - AR | ENSG00000282723 | ENSG00000228696 | | 16093 | 13647 | rs2732689-A | rs2732689 | 0 | 2732689 | upstream_gene_variant | 1 | 0.2365 | 7.00E-18 | 17.1549 | | 0.09423 | [0.073-0.1 | Affymetrix [9389114] (imputed) | N |
| 9 | 17:44118848 | 17 | 44354904 | rs2732699 | 30/11/2017 | 2.9E+07 | Michailidou K | 23/10/2017 | Nature | www.ncbi.nlm.nih.gov/pubmed/29059683 | Association analysis identifies 65 new breast cancer risk loci. | Breast cancer | 76,192 Eu | 46,785 Eu | 17q21.31 | NR | ARL17B, ARL17B, ARL17B | | | ENSG00000275748, E | NA | NA | rs2732699-C | rs2732699 | 0 | 2732699 | intron_variant | 0 | 0.8401 | 1.00E-10 | 10 | (EA) | 0.0595 | [0.041-0.0 | Illumina [~ 11800000] (imputed) | N |
| 9 | 17:44118848 | 17 | 44353728 | rs2732702 | 31/05/2022 | 3.4E+07 | Smith SM | 19/04/2021 | Nat Neurosci | www.ncbi.nlm.nih.gov/pubmed/33875891 | An expanded set of genome-wide association studies of brain imaging phenotypes in UK Biobank. | aparc-Desikan lh area TotalSurface | 21,282 Brit | 10,686 Brit | 17q21.31 | | ARL17B, ARL17B, ARL17B | | | ENSG00000275748, E | NA | NA | rs2732702-G | rs2732702 | 0 | 2732702 | intron_variant | 0 | 0.225 | 1.00E-22 | 22 | | 0.116 | [0.092-0.1 | Affymetrix [17103079] (imputed) | N |
| 9 | 17:44118848 | 17 | 44351686 | rs2732706 | 14/01/2022 | 3.5E+07 | Bonfiglio F | 8/12/2021 | Cell Genom | www.ncbi.nlm.nih.gov/pubmed/34957435 | GWAS of stool frequency provides insights into gastrointestinal motility and irritable bowel syndrome. | Stool frequency | 167,875 Eu | NA | 17q21.31 | | MAPK8IP1P1 - AR | ENSG00000282723 | ENSG00000228696 | | 29276 | 464 | rs2732706-T | rs2732706 | 0 | 2732706 | downstream_gene_variant | 1 | 0.221 | 4.00E-12 | 11.39794 | | 0.024 | [0.018-0.0 | Affymetrix, Illumina [8817117] (imputed) | N |
| 9 | 17:44118848 | 17 | 44351387 | rs2732708 | 22/01/2019 | 3E+07 | Nagel M | 2/03/2018 | Nat Commun | www.ncbi.nlm.nih.gov/pubmed/29500382 | Item-level analyses reveal genetic heterogeneity in neuroticism. | Feeling miserable | 376,097 Eu | NA | 17q21.31 | NR | MAPK8IP1P1 - AR | ENSG00000282723 | ENSG00000228696 | | 28977 | 763 | rs2732708-T | rs2732708 | 0 | 2732708 | downstream_gene_variant | 1 | 0.214039 | 3.00E-11 | 10.52288 | | 6.64 | z score inc | NR [10847151] (imputed) | N |
| 9 | 17:44118848 | 17 | 44351387 | rs2732708 | 1/02/2018 | 2.9E+07 | Luciano M | 18/12/2017 | Nat Genet | www.ncbi.nlm.nih.gov/pubmed/29255261 | Association analysis in over 329,000 individuals identifies 116 independent variants influencing neuroticism. | Neuroticism | 329,821 Eu | 122,867 Eu | 17q21.31 | intergenic | MAPK8IP1P1 - AR | ENSG00000282723 | ENSG00000228696 | | 28977 | 763 | rs2732708-T | rs2732708 | 0 | 2732708 | downstream_gene_variant | 1 | 0.21404 | 2.00E-23 | 22.69897 | | 9.955 | unit increa | Illumina [18485882] (imputed) | N |
| 9 | 17:44118848 | 17 | 44321478 | rs2732714 | 31/05/2022 | 3.4E+07 | Smith SM | 19/04/2021 | Nat Neurosci | www.ncbi.nlm.nih.gov/pubmed/33875891 | An expanded set of genome-wide association studies of brain imaging phenotypes in UK Biobank. | IDP T1 SIENAX brain-normalised volume | 22,138 Brit | 11,086 Brit | 17q21.31 | | MAPK8IP1P1, MAPK8IP1P1 | | | ENSG00000262500, E | NA | NA | rs2732714-G | rs2732714 | 0 | 2732714 | non_coding_transcript_exon | 0 | 0.194 | 5.00E-13 | 12.30103 | | 0.09 | [0.066-0.1 | Affymetrix [17103079] (imputed) | N |
| 9 | 17:44118848 | 17 | 44321478 | rs2732714 | 31/05/2022 | 3.4E+07 | Smith SM | 19/04/2021 | Nat Neurosci | www.ncbi.nlm.nih.gov/pubmed/33875891 | An expanded set of genome-wide association studies of brain imaging phenotypes in UK Biobank. | IDP T1 SIENAX brain-unnormalised volume | 22,138 Brit | 11,086 Brit | 17q21.31 | | MAPK8IP1P1, MAPK8IP1P1 | | | ENSG00000262500, E | NA | NA | rs2732714-G | rs2732714 | 0 | 2732714 | non_coding_transcript_exon | 0 | 0.194 | 3.00E-13 | 12.52288 | | 0.091 | [0.067-0.1 | Affymetrix [17103079] (imputed) | N |
| 9 | 17:44118848 | 17 | 44321478 | rs2732714 | 31/05/2022 | 3.4E+07 | Smith SM | 19/04/2021 | Nat Neurosci | www.ncbi.nlm.nih.gov/pubmed/33875891 | An expanded set of genome-wide association studies of brain imaging phenotypes in UK Biobank. | IDP T1 SIENAX white normalised volume | 22,138 Brit | 11,086 Brit | 17q21.31 | | MAPK8IP1P1, MAPK8IP1P1 | | | ENSG00000262500, E | NA | NA | rs2732714-G | rs2732714 | 0 | 2732714 | non_coding_transcript_exon | 0 | 0.194 | 3.00E-12 | 11.52288 | | 0.087 | [0.063-0.1 | Affymetrix [17103079] (imputed) | N |
| 9 | 17:44118848 | 17 | 44321478 | rs2732714 | 31/05/2022 | 3.4E+07 | Smith SM | 19/04/2021 | Nat Neurosci | www.ncbi.nlm.nih.gov/pubmed/33875891 | An expanded set of genome-wide association studies of brain imaging phenotypes in UK Biobank. | IDP T1 SIENAX white unnormalised volume | 22,138 Brit | 11,086 Brit | 17q21.31 | | MAPK8IP1P1, MAPK8IP1P1 | | | ENSG00000262500, E | NA | NA | rs2732714-G | rs2732714 | 0 | 2732714 | non_coding_transcript_exon | 0 | 0.194 | 2.00E-12 | 11.69897 | | 0.088 | [0.064-0.1 | Affymetrix [17103079] (imputed) | N |
| 9 | 17:44296846 | 17 | 43688317 | rs28768976 | 27/07/2021 | 3.3E+07 | Harper AR | 25/01/2021 | Nat Genet | www.ncbi.nlm.nih.gov/pubmed/33495597 | Common genetic variants and modifiable risk factors underpin hypertrophic cardiomyopathy susceptibility and expressivity. | Hypertrophic cardiomyopathy | 233 Africar | 1,643 Euro | 17q21.31 | SPPL2C | RPS26P8 - LINC0 | ENSG00000282238 | ENSG00000204650 | | 2033 | 9377 | rs28768976-A | rs28768976 | 0 | 28768976 | downstream_gene_variant | 1 | 0.772 | 4.00E-12 | 11.39794 | | 0.255 | [0.18-0.33 | Affymetrix [8590397] (imputed) | N |
| 9 | 17:44296846 | 17 | 43688317 | rs28768976 | 27/07/2021 | 3.3E+07 | Harper AR | 25/01/2021 | Nat Genet | www.ncbi.nlm.nih.gov/pubmed/33495597 | Common genetic variants and modifiable risk factors underpin hypertrophic cardiomyopathy susceptibility and expressivity. | Hypertrophic cardiomyopathy (sarcomere negative) | 170 Africar | NA | 17q21.31 | SPPL2C | RPS26P8 - LINC0 | ENSG00000282238 | ENSG00000204650 | | 2033 | 9377 | rs28768976-A | rs28768976 | 0 | 28768976 | downstream_gene_variant | 1 | 0.770677 | 2.00E-09 | 8.69897 | | 0.268112 | [0.18-0.36 | Affymetrix [8590397] (imputed) | N |
| 9 | 17:44296846 | 17 | 43688317 | rs28768976 | 1/02/2018 | 2.9E+07 | Luciano M | 18/12/2017 | Nat Genet | www.ncbi.nlm.nih.gov/pubmed/29255261 | Association analysis in over 329,000 individuals identifies 116 independent variants influencing neuroticism. | Neuroticism | 329,821 Eu | 122,867 Eu | 17q21.31 | intergenic | RPS26P8 - LINC0 | ENSG00000282238 | ENSG00000204650 | | 2033 | 9377 | rs28768976-A | rs28768976 | 0 | 28768976 | downstream_gene_variant | 1 | 0.77211 | 1.00E-25 | 25 | | 10.453 | unit decrea | Illumina [18485882] (imputed) | N |
| 9 | 17:44118848 | 17 | 43714850 | rs2942168 | 20/04/2015 | 2.1E+07 | Nalls MA | 1/02/2011 | Lancet | www.ncbi.nlm.nih.gov/pubmed/21292315 | Imputation of sequence variants for identification of genetic risks for Parkinson's disease: a meta-analysis of genome-wide association studies. | Parkinson's disease | 5,333 Euro | 7,053 case | 17q21.31 | MAPT | LINC02210-CRHR1, LINC02210-CRHR1, LINC02210, LINC0 | | | ENSG00000282456, E | NA | NA | rs2942168-G | rs2942168 | 0 | 2942168 | non_coding_transcript_exon | 0 | 0.78 | 1.00E-28 | 28 | | 1.27 | [1.23-1.31 | Illumina [7689524] (imputed) | N |
| 9 | 17:44118848 | 17 | 44332351 | rs2950015 | 18/09/2020 | 3.3E+07 | Vuckovic D | 1/09/2020 | Cell | www.ncbi.nlm.nih.gov/pubmed/32888494 | The Polygenic and Monogenic Basis of Blood Traits and Diseases. | Hematocrit | 408,112 Bi | NA | 17q21.31 | RP11-259G18.3 | MAPK8IP1P1 - AR | ENSG00000282723 | ENSG00000228696 | | 9941 | 19799 | rs2950015-G | rs2950015 | 0 | 2950015 | regulatory_region_variant | 1 | 0.227742 | 2.00E-55 | 54.69897 | | 0.04169 | [0.036-0.0 | Affymetrix [93095623] (imputed) | N |
| 9 | 17:44118848 | 17 | 44332351 | rs2950015 | 18/09/2020 | 3.3E+07 | Vuckovic D | 1/09/2020 | Cell | www.ncbi.nlm.nih.gov/pubmed/32888494 | The Polygenic and Monogenic Basis of Blood Traits and Diseases. | Hemoglobin | 408,112 Bi | NA | 17q21.31 | RP11-259G18.3 | MAPK8IP1P1 - AR | ENSG00000282723 | ENSG00000228696 | | 9941 | 19799 | rs2950015-G | rs2950015 | 0 | 2950015 | regulatory_region_variant | 1 | 0.227758 | 6.00E-48 | 47.22185 | | 0.04 | [0.035-0.0 | Affymetrix [93095623] (imputed) | N |
| 9 | 17:44118848 | 17 | 44343004 | rs2950706 | 5/08/2019 | 3.1E+07 | Ward J | 5/06/2019 | Mol Psychiatry | www.ncbi.nlm.nih.gov/pubmed/31168069 | The genomic basis of mood instability: identification of 46 loci in 363,705 UK Biobank participants, genetic correlation with psychiatric disorders, and association with gene expression and function. | Mood instability | 157,039 Eu | NA | 17q21.31 | CRHR1, MAPT, W | MAPK8IP1P1 - AR | ENSG00000282723 | ENSG00000228696 | | 20594 | 9146 | rs2950706-? | rs2950706 | 0 | 2950706 | intergenic_variant | 1 | NR | 7.00E-20 | 19.1549 | | 0.013745 | [0.011-0.0 | Affymetrix [NR] (imputed) | N |
| 9 | 17:44118848 | 17 | 44368212 | rs2957297 | 17/09/2018 | 3E+07 | Davies G | 29/05/2018 | Nat Commun | www.ncbi.nlm.nih.gov/pubmed/29844566 | Study of 300,486 individuals identifies 148 independent genetic loci influencing general cognitive function. | General cognitive ability | up to 300,4 | NA | 17q21.31 | NR | ARL17B, ARL17B, ARL17B | | | ENSG00000275748, E | NA | NA | rs2957297-? | rs2957297 | 0 | 2957297 | intron_variant | 0 | NR | 2.00E-07 | 6.69897 | | 5.231 | z-score de | Affymetrix, Illumina [at least 2500000] (imputed) | N |
| 9 | 17:44118848 | 17 | 44331214 | rs2957316 | 14/12/2021 | 3.5E+07 | Sakaue S | 30/09/2021 | Nat Genet | www.ncbi.nlm.nih.gov/pubmed/34594039 | A cross-population atlas of genetic associations for 220 human phenotypes. | Interstitial lung disease | 2,267 Euro | NA | 17q21.31 | | MAPK8IP1P1 - AR | ENSG00000282723 | ENSG00000228696 | | 8804 | 20936 | rs2957316-C | rs2957316 | 0 | 2957316 | TF_binding_site_variant | 1 | NR | 3.00E-09 | 8.522879 | | 0.2644 | [0.18-0.35 | Affymetrix, Illumina [25843170] (imputed) | N |
| 9 | 17:44118848 | 17 | 44331214 | rs2957316 | 14/12/2021 | 3.5E+07 | Sakaue S | 30/09/2021 | Nat Genet | www.ncbi.nlm.nih.gov/pubmed/34594039 | A cross-population atlas of genetic associations for 220 human phenotypes. | Pulmonary fibrosis | 1,566 Euro | NA | 17q21.31 | | MAPK8IP1P1 - AR | ENSG00000282723 | ENSG00000228696 | | 8804 | 20936 | rs2957316-C | rs2957316 | 0 | 2957316 | TF_binding_site_variant | 1 | NR | 5.00E-08 | 7.30103 | | 0.2809 | [0.18-0.38 | Affymetrix, Illumina [25843031] (imputed) | N |
| 9 | 17:43564222 | 17 | 43511435 | rs34465449 | 24/09/2021 | 3.3E+07 | Wendt FR | 21/12/2020 | Nat Hum Behav | www.ncbi.nlm.nih.gov/pubmed/33349686 | Multivariate genome-wide analysis of education, socioeconomic status and brain phenome. | Neuroticism conditioned on cognitive performance (multi-trait conditioning and joint analysis) | 170,911 Eu | NA | 17q21.31 | | ARHGAP27 | | | ENSG00000159314 | NA | NA | rs34465449-? | rs34465449 | 0 | 34465449 | intron_variant | 0 | NR | 1.00E-12 | 12 | | NA | | NR [NR] | N |
| 9 | 17:43520118 | 17 | 43520675 | rs35329349 | 14/12/2021 | 3.5E+07 | Sakaue S | 30/09/2021 | Nat Genet | www.ncbi.nlm.nih.gov/pubmed/34594039 | A cross-population atlas of genetic associations for 220 human phenotypes. | White blood cell count | 350,470 Eu | NA | 17q21.31 | | PLEKHM1 | | | ENSG00000277111 | NA | NA | rs35329349-C | rs35329349 | 0 | 35329349 | intron_variant | 0 | NR | 1.00E-10 | 10 | | 0.0207 | [0.014-0.0 | Affymetrix, Illumina [20538670] (imputed) | N |
| 9 | 17:43564222 | 17 | 43515927 | rs35354512 | 24/09/2021 | 3.3E+07 | Wendt FR | 21/12/2020 | Nat Hum Behav | www.ncbi.nlm.nih.gov/pubmed/33349686 | Multivariate genome-wide analysis of education, socioeconomic status and brain phenome. | Neuroticism conditioned on highest math class (multi-trait conditioning and joint analysis) | 170,911 Eu | NA | 17q21.31 | | PLEKHM1 | | | ENSG00000277111 | NA | NA | rs35354512-? | rs35354512 | 0 | 35354512 | intron_variant | 0 | NR | 1.00E-12 | 12 | | NA | | NR [NR] | N |
| 9 | 17:43564222 | 17 | 43515927 | rs35354512 | 24/09/2021 | 3.3E+07 | Wendt FR | 21/12/2020 | Nat Hum Behav | www.ncbi.nlm.nih.gov/pubmed/33349686 | Multivariate genome-wide analysis of education, socioeconomic status and brain phenome. | Neuroticism conditioned on self-rated math ability (multi-trait conditioning and joint analysis) | 170,911 Eu | NA | 17q21.31 | | PLEKHM1 | | | ENSG00000277111 | NA | NA | rs35354512-? | rs35354512 | 0 | 35354512 | intron_variant | 0 | NR | 5.00E-12 | 11.30103 | | NA | | NR [NR] | N |
| 9 | 17:43556807 | 17 | 43499328 | rs35519908 | 5/02/2020 | 3.2E+07 | Zhao B | 30/10/2019 | Mol Psychiatry | www.ncbi.nlm.nih.gov/pubmed/31666681 | Large-scale GWAS reveals genetic architecture of brain white matter microstructure and genetic overlap with cognitive and mental health traits (nÔÇë=ÔÇë17,706). | White matter microstructure (mean diusivities) | 17,706 Eu | NA | 17q21.31 | NR | ARHGAP27 | | | ENSG00000159314 | NA | NA | rs35519908-? | rs35519908 | 0 | 35519908 | intron_variant | 0 | | 3.00E-09 | 8.522879 | (Cingulate | NA | | NR [8955960] (imputed) | N |
| 9 | 17:44118848 | 17 | 44192590 | rs35524223 | 16/06/2017 | 2.8E+07 | Wain LV | 6/02/2017 | Nat Genet | www.ncbi.nlm.nih.gov/pubmed/28166213 | Genome-wide association analyses for lung function and chronic obstructive pulmonary disease identify new loci and potential druggable targets. | Lung function (FEV1) | 48,943 Eu | 57,176 Eu | 17q21.31 | KANSL1 | KANSL1, KANSL1, KANSL1 | | | ENSG00000120071, E | NA | NA | rs35524223-A | rs35524223 | 0 | 35524223 | intron_variant | 0 | 0.22 | 1.00E-13 | 13 | | 0.061 | [0.045-0.0 | Affymetrix [up to 27624732] (imputed) | N |
| 9 | 17:44118848 | 17 | 44192590 | rs35524223 | 16/06/2017 | 2.8E+07 | Wain LV | 6/02/2017 | Nat Genet | www.ncbi.nlm.nih.gov/pubmed/28166213 | Genome-wide association analyses for lung function and chronic obstructive pulmonary disease identify new loci and potential druggable targets. | Lung function (FVC) | 48,943 Eu | 57,176 Eu | 17q21.31 | KANSL1 | KANSL1, KANSL1, KANSL1 | | | ENSG00000120071, E | NA | NA | rs35524223-A | rs35524223 | 0 | 35524223 | intron_variant | 0 | 0.2166 | 7.00E-13 | 12.1549 | | 0.059 | [0.043-0.0 | Affymetrix [27624732] (imputed) | N |
| 9 | 17:44118848 | 17 | 44248837 | rs35643216 | 17/06/2020 | 3.2E+07 | Oskarsson GR | 23/04/2020 | Commun Biol | www.ncbi.nlm.nih.gov/pubmed/32327693 | Predicted loss and gain of function mutations in ACO1 are associated with erythropoiesis. | Hemoglobin levels | 684,122 Eu | NA | 17q21.31 | KANSL1 | KANSL1, KANSL1, KANSL1 | | | ENSG00000120071, E | NA | NA | rs35643216-C | rs35643216 | 0 | 35643216 | missense_variant | 0 | | 2.00E-56 | 55.69897 | | 0.046 | [NR] unit in | Affymetrix, Illumina [43000000] (imputed) | N |
| 9 | 17:44118848 | 17 | 44353414 | rs36050210 | 31/05/2022 | 3.4E+07 | Smith SM | 19/04/2021 | Nat Neurosci | www.ncbi.nlm.nih.gov/pubmed/33875891 | An expanded set of genome-wide association studies of brain imaging phenotypes in UK Biobank. | IDP dMRI TBSS L3 Anterior limb of internal capsule R | 20,860 Brit | 10,496 Brit | 17q21.31 | | ARL17B, ARL17B, ARL17B | | | ENSG00000275748, E | NA | NA | rs36050210-G | rs36050210 | 0 | 36050210 | intron_variant | 0 | 0.224 | 2.00E-18 | 17.69897 | | 0.105 | [0.081-0.1 | Affymetrix [17103079] (imputed) | N |
| 9 | 17:44296846 | 17 | 43705601 | rs365825 | 11/05/2017 | 2.7E+07 | Pickrell JK | 16/05/2016 | Nat Genet | www.ncbi.nlm.nih.gov/pubmed/27182965 | Detection and interpretation of shared genetic influences on 42 human traits. | Parkinson's disease | 9,619 Euro | NA | 17q21.31 | CRHR1, LRRC37A | LINC02210-CRHR1, LINC02210-CRHR1, LINC02210, LINC0 | | | ENSG00000282456, E | NA | NA | rs365825-? | rs365825 | 0 | 365825 | intron_variant | 0 | NR | 4.00E-32 | 31.39794 | | 1.272265 | [1.22-1.33 | Illumina [12438219] (imputed) | N |
| 9 | 17:44118848 | 17 | 44611782 | rs369234167 | 31/05/2022 | 3.4E+07 | Smith SM | 19/04/2021 | Nat Neurosci | www.ncbi.nlm.nih.gov/pubmed/33875891 | An expanded set of genome-wide association studies of brain imaging phenotypes in UK Biobank. | aseg global volume BrainSegNotVent | 21,282 Brit | 10,686 Brit | 17q21.31 | | LRRC37A2, ARL17A | | | ENSG00000277221, E | NA | NA | rs369234167-C | rs369234167 | 0 | 3.69E+08 | intron_variant | 0 | 0.199 | 4.00E-14 | 13.39794 | | 0.099 | [0.074-0.1 | Affymetrix [17103079] (imputed) | N |
| 9 | 17:44118848 | 17 | 44086726 | rs370558872 | 1/07/2022 | 3.5E+07 | Okbay A | 31/03/2022 | Nat Genet | www.ncbi.nlm.nih.gov/pubmed/35361970 | Polygenic prediction of educational attainment within and between families from genome-wide association analyses in 3 million individuals. | Educational attainment | 3,037,499 | NA | 17q21.31 | | MAPT, MAPT, MAPT | | | ENSG00000277956, E | NA | NA | rs370558872-T | rs370558872 | 0 | 3.71E+08 | intron_variant | 0 | 0.7764 | 4.00E-58 | 57.39794 | | 0.023336 | [0.02-0.02 | Illumina [10675380] (imputed) | N |
| 9 | 17:44118848 | 17 | 44355304 | rs371894855 | 31/05/2022 | 3.4E+07 | Smith SM | 19/04/2021 | Nat Neurosci | www.ncbi.nlm.nih.gov/pubmed/33875891 | An expanded set of genome-wide association studies of brain imaging phenotypes in UK Biobank. | IDP dMRI TBSS ICVF Posterior limb of internal capsule R | 20,859 Brit | 10,496 Brit | 17q21.31 | | ARL17B, ARL17B, ARL17B | | | ENSG00000275748, E | NA | NA | rs371894855-T | rs371894855 | 0 | 3.72E+08 | intron_variant | 0 | 0.204 | 2.00E-20 | 19.69897 | | 0.117 | [0.092-0.1 | Affymetrix [17103079] (imputed) | N |
| 9 | 17:44183317 | 17 | 44183317 | rs371996525 | 28/03/2022 | 3.4E+07 | Naqvi S | 5/04/2021 | Nat Genet | www.ncbi.nlm.nih.gov/pubmed/33821002 | Shared heritability of human face and brain shape. | Brain shape (segment 1) | 19,670 Eu | NA | 17q21.31 | | KANSL1, KANSL1, KANSL1 | | | ENSG00000120071, E | NA | NA | rs371996525-? | rs371996525 | 0 | 3.72E+08 | intron_variant | 0 | NR | 4.00E-48 | 47.39794 | | NA | | NR [9705931] (imputed) | N |
| 9 | 17:43564222 | 17 | 43557054 | rs375822897 | 18/10/2018 | 3E+07 | Kim SK | 26/07/2018 | PLoS One | www.ncbi.nlm.nih.gov/pubmed/30048462 | Identification of 613 new loci associated with heel bone mineral density and a polygenic risk score for bone mineral density, osteoporosis and fracture. | Heel bone mineral density | 394,929 Eu | NA | 17q21.31 | | PLEKHM1 | | | ENSG00000277111 | NA | NA | rs375822897-? | rs375822897 | 0 | 3.76E+08 | intron_variant | 0 | NR | 2.00E-24 | 23.69897 | | 0.027821 | [0.022-0.0 | Affymetrix [20259828] (imputed) | N |
| 9 | 17:44118848 | 17 | 44057595 | rs3785884 | 21/09/2020 | 3.3E+07 | Chen MH | 1/09/2020 | Cell | www.ncbi.nlm.nih.gov/pubmed/32888493 | Trans-ethnic and Ancestry-Specific Blood-Cell Genetics in 746,667 Individuals from 5 Global Populations. | Hematocrit | 562,259 Eu | NA | 17q21.31 | NR | MAPT, MAPT, MAPT | | | ENSG00000277956, E | NA | NA | rs3785884-A | rs3785884 | 0 | 3785884 | non_coding_transcript_exon | 0 | 0.23073 | 1.00E-96 | 96 | | 0.045836 | [0.042-0.0 | Affymetrix, Illumina [50640577] (imputed) | N |
| 9 | 17:44118848 | 17 | 44057595 | rs3785884 | 21/09/2020 | 3.3E+07 | Chen MH | 1/09/2020 | Cell | www.ncbi.nlm.nih.gov/pubmed/32888493 | Trans-ethnic and Ancestry-Specific Blood-Cell Genetics in 746,667 Individuals from 5 Global Populations. | Hemoglobin concentration | 563,946 Eu | NA | 17q21.31 | NR | MAPT, MAPT, MAPT | | | ENSG00000277956, E | NA | NA | rs3785884-A | rs3785884 | 0 | 3785884 | non_coding_transcript_exon_ | 0 | 0.230491 | 1.00E-93 | 93 | | 0.045117 | [0.041-0.0 | Affymetrix, Illumina [50638016] (imputed) | N |
| 9 | 17:44118848 | 17 | 43686419 | rs385199 | 31/05/2022 | 3.4E+07 | Smith SM | 19/04/2021 | Nat Neurosci | www.ncbi.nlm.nih.gov/pubmed/33875891 | An expanded set of genome-wide association studies of brain imaging phenotypes in UK Biobank. | aparc-pial lh area TotalSurface | 21,282 Brit | 10,686 Brit | 17q21.31 | | RPS26P8 - LINC0 | ENSG00000282238 | ENSG00000204650 | | 135 | 11275 | rs385199-C | rs385199 | 0 | 385199 | intergenic_variant | 1 | 0.227 | 1.00E-10 | 10 | | 0.075 | [0.051-0.0 | Affymetrix [17103079] (imputed) | N |
| 9 | 17:44118848 | 17 | 43686419 | rs385199 | 15/04/2021 | 3.4E+07 | Dashti HS | 10/02/2021 | Nat Commun | www.ncbi.nlm.nih.gov/pubmed/33568662 | Genetic determinants of daytime napping and effects on cardiometabolic health. | Daytime nap | 452,633 Eu | 541,333 Eu | 17q21.31 | CRHR1, PLEKHM | RPS26P8 - LINC0 | ENSG00000282238 | ENSG00000204650 | | 135 | 11275 | rs385199-A | rs385199 | 0 | 385199 | intergenic_variant | 1 | | 1.00E-72 | 72 | | 0.0185 | [0.017-0.0 | Affymetrix [13304133] (imputed) | N |
| 9 | 17:44118848 | 17 | 43687268 | rs391338 | 31/05/2022 | 3.4E+07 | Smith SM | 19/04/2021 | Nat Neurosci | www.ncbi.nlm.nih.gov/pubmed/33875891 | An expanded set of genome-wide association studies of brain imaging phenotypes in UK Biobank. | IDP dMRI TBSS L3 Anterior limb of internal capsule R | 20,860 Brit | 10,496 Brit | 17q21.31 | | RPS26P8 - LINC0 | ENSG00000282238 | ENSG00000204650 | | 984 | 10426 | rs391338-A | rs391338 | 0 | 391338 | regulatory_region_variant | 1 | 0.221 | 5.00E-17 | 16.30103 | | 0.099 | [0.075-0.1 | Affymetrix [17103079] (imputed) | N |
| 9 | 17:44296846 | 17 | 43719143 | rs393152 | 8/07/2016 | 2.6E+07 | Kouri N | 16/06/2015 | Nat Commun | www.ncbi.nlm.nih.gov/pubmed/26077951 | Genome-wide association study of corticobasal degeneration identifies risk variants shared with progressive supranuclear palsy. | Corticobasal degeneration | 152 cases, | 67 cases, 4 | 17q21.31 | MAPT | LINC02210, LINC02210, LINC02210-CRHR1, LINC02210-C | | | ENSG00000282171, E | NA | NA | rs393152-? | rs393152 | 0 | 393152 | non_coding_transcript_exon | 0 | NR | 1.00E-12 | 12 | | 3.7 | [NR] | Illumina [533898] (imputed) | N |
| 9 | 17:44296846 | 17 | 43719143 | rs393152 | 10/12/2009 | 2E+07 | Simon-Sanchez J | 15/11/2009 | Nat Genet | www.ncbi.nlm.nih.gov/pubmed/19915575 | Genome-wide association study reveals genetic risk underlying Parkinson's disease. | Parkinson's disease | 1,713 Euro | 3,361 Euro | 17q21.31 | MAPT, C17orf69, | LINC02210, LINC02210, LINC02210-CRHR1, LINC02210-C | | | ENSG00000282171, E | NA | NA | rs393152-A | rs393152 | 0 | 393152 | non_coding_transcript_exon | 0 | 0.82 | 2.00E-16 | 15.69897 | | 1.3 | [NR] | Illumina [463185] | N |
| 9 | 17:44861003 | 17 | 44859144 | rs415430 | 14/09/2011 | 2.2E+07 | Liu X | 3/08/2011 | BMC Med Genet | www.ncbi.nlm.nih.gov/pubmed/21812969 | Genome-wide association study identifies candidate genes for Parkinson's disease in an Ashkenazi Jewish population. | Parkinson's disease | 268 Ashke | 1,782 Euro | 17q21.31 | WNT3 | WNT3 | | | ENSG00000108379 | NA | NA | rs415430-T | rs415430 | 0 | 415430 | intron_variant | 0 | 0.786 | 7.00E-07 | 6.154902 | | NA | | Illumina [525124] | N |
| 9 | 17:44118848 | 17 | 44021717 | rs4327091 | 24/08/2022 | 3.6E+07 | Plotnikov D | 1/06/2022 | Invest Ophthalmol Vis Sci | www.ncbi.nlm.nih.gov/pubmed/35762941 | High Blood Pressure and Intraocular Pressure: A Mendelian Randomization Study. | Systolic blood pressure | 526,001 Eu | NA | 17q21.31 | | MAPT, MAPT, MAPT | | | ENSG00000277956, E | NA | NA | rs4327091-A | rs4327091 | 0 | 4327091 | intron_variant | 0 | NR | 7.00E-11 | 10.1549 | | 0.31 | [0.22-0.4] | NR [NR] (imputed) | N |

| | | | | | | | | | | | | | | | | | | | | | | | | | | | | | | | | | | | | |
|---|---|---|---|---|---|---|---|---|---|---|---|---|---|---|---|---|---|---|---|---|---|---|---|---|---|---|---|---|---|---|---|---|---|---|---|---|
| 9 | 17:44118848 | 17 | 43686420 | rs440778 | 31/05/2022 | 3.4E+07 | Smith SM | 19/04/2021 | Nat Neurosci | www.ncbi.nlm.nih.gov/pubmed/33875891 | An expanded set of genome-wide association studies of brain imaging phenotypes in UK Biobank. | aseg global volume BrainSeg | 21,282 Brit | 10,686 Brit | 17q21.31 | | RPS26P8 - LINC0 | ENSG00000282238 | ENSG00000204650 | | 136 | 11274 | rs440778-G | rs440778 | 0 | 440778 | intergenic_variant | 1 | 0.223 | 4.00E-13 | 12.39794 | | 0.085 | [0.061-0.1 | Affymetrix [17103079] (imputed) | N |
| 9 | 17:44118848 | 17 | 43686420 | rs440778 | 31/05/2022 | 3.4E+07 | Smith SM | 19/04/2021 | Nat Neurosci | www.ncbi.nlm.nih.gov/pubmed/33875891 | An expanded set of genome-wide association studies of brain imaging phenotypes in UK Biobank. | aseg global volume BrainSegNotVent | 21,282 Brit | 10,686 Brit | 17q21.31 | | RPS26P8 - LINC0 | ENSG00000282238 | ENSG00000204650 | | 136 | 11274 | rs440778-G | rs440778 | 0 | 440778 | intergenic_variant | 1 | 0.223 | 1.00E-13 | 13 | | 0.087 | [0.063-0.1 | Affymetrix [17103079] (imputed) | N |
| 9 | 17:44118848 | 17 | 43686420 | rs440778 | 31/05/2022 | 3.4E+07 | Smith SM | 19/04/2021 | Nat Neurosci | www.ncbi.nlm.nih.gov/pubmed/33875891 | An expanded set of genome-wide association studies of brain imaging phenotypes in UK Biobank. | aseg global volume BrainSegNotVentSurf | 21,282 Brit | 10,686 Brit | 17q21.31 | | RPS26P8 - LINC0 | ENSG00000282238 | ENSG00000204650 | | 136 | 11274 | rs440778-G | rs440778 | 0 | 440778 | intergenic_variant | 1 | 0.223 | 8.00E-14 | 13.09691 | | 0.087 | [0.063-0.1 | Affymetrix [17103079] (imputed) | N |
| 9 | 17:44118848 | 17 | 43686420 | rs440778 | 31/05/2022 | 3.4E+07 | Smith SM | 19/04/2021 | Nat Neurosci | www.ncbi.nlm.nih.gov/pubmed/33875891 | An expanded set of genome-wide association studies of brain imaging phenotypes in UK Biobank. | aseg global volume SupraTentorial | 21,282 Brit | 10,686 Brit | 17q21.31 | | RPS26P8 - LINC0 | ENSG00000282238 | ENSG00000204650 | | 136 | 11274 | rs440778-G | rs440778 | 0 | 440778 | intergenic_variant | 1 | 0.223 | 5.00E-12 | 11.30103 | | 0.081 | [0.057-0.1 | Affymetrix [17103079] (imputed) | N |
| 9 | 17:44118848 | 17 | 43686420 | rs440778 | 31/05/2022 | 3.4E+07 | Smith SM | 19/04/2021 | Nat Neurosci | www.ncbi.nlm.nih.gov/pubmed/33875891 | An expanded set of genome-wide association studies of brain imaging phenotypes in UK Biobank. | aseg global volume SupraTentorialNotVent | 21,282 Brit | 10,686 Brit | 17q21.31 | | RPS26P8 - LINC0 | ENSG00000282238 | ENSG00000204650 | | 136 | 11274 | rs440778-G | rs440778 | 0 | 440778 | intergenic_variant | 1 | 0.223 | 2.00E-12 | 11.69897 | | 0.082 | [0.058-0.1 | Affymetrix [17103079] (imputed) | N |
| 9 | 17:44118848 | 17 | 43686420 | rs440778 | 31/05/2022 | 3.4E+07 | Smith SM | 19/04/2021 | Nat Neurosci | www.ncbi.nlm.nih.gov/pubmed/33875891 | An expanded set of genome-wide association studies of brain imaging phenotypes in UK Biobank. | aseg lh volume CerebralWhiteMatter | 21,282 Brit | 10,686 Brit | 17q21.31 | | RPS26P8 - LINC0 | ENSG00000282238 | ENSG00000204650 | | 136 | 11274 | rs440778-G | rs440778 | 0 | 440778 | intergenic_variant | 1 | 0.223 | 9.00E-14 | 13.04576 | | 0.087 | [0.063-0.1 | Affymetrix [17103079] (imputed) | N |
| 9 | 17:44118848 | 17 | 43686420 | rs440778 | 31/05/2022 | 3.4E+07 | Smith SM | 19/04/2021 | Nat Neurosci | www.ncbi.nlm.nih.gov/pubmed/33875891 | An expanded set of genome-wide association studies of brain imaging phenotypes in UK Biobank. | aseg rh volume CerebralWhiteMatter | 21,282 Brit | 10,686 Brit | 17q21.31 | | RPS26P8 - LINC0 | ENSG00000282238 | ENSG00000204650 | | 136 | 11274 | rs440778-G | rs440778 | 0 | 440778 | intergenic_variant | 1 | 0.223 | 1.00E-15 | 15 | | 0.093 | [0.069-0.1 | Affymetrix [17103079] (imputed) | N |
| 9 | 17:44118848 | 17 | 43686420 | rs440778 | 31/05/2022 | 3.4E+07 | Smith SM | 19/04/2021 | Nat Neurosci | www.ncbi.nlm.nih.gov/pubmed/33875891 | An expanded set of genome-wide association studies of brain imaging phenotypes in UK Biobank. | IDP T1 SIENAX brain-normalised volume | 22,138 Brit | 11,086 Brit | 17q21.31 | | RPS26P8 - LINC0 | ENSG00000282238 | ENSG00000204650 | | 136 | 11274 | rs440778-G | rs440778 | 0 | 440778 | intergenic_variant | 1 | 0.223 | 1.00E-12 | 12 | | 0.081 | [0.059-0.1 | Affymetrix [17103079] (imputed) | N |
| 9 | 17:44118848 | 17 | 43686420 | rs440778 | 31/05/2022 | 3.4E+07 | Smith SM | 19/04/2021 | Nat Neurosci | www.ncbi.nlm.nih.gov/pubmed/33875891 | An expanded set of genome-wide association studies of brain imaging phenotypes in UK Biobank. | IDP T1 SIENAX brain-unnormalised volume | 22,138 Brit | 11,086 Brit | 17q21.31 | | RPS26P8 - LINC0 | ENSG00000282238 | ENSG00000204650 | | 136 | 11274 | rs440778-G | rs440778 | 0 | 440778 | intergenic_variant | 1 | 0.223 | 1.00E-12 | 12 | | 0.081 | [0.059-0.1 | Affymetrix [17103079] (imputed) | N |
| 9 | 17:44118848 | 17 | 44205690 | rs4471723 | 22/01/2019 | 3E+07 | Nagel M | 2/03/2018 | Nat Commun | www.ncbi.nlm.nih.gov/pubmed/29500382 | Item-level analyses reveal genetic heterogeneity in neuroticism. | Feeling guilty | 373,380 Eu | NA | 17q21.31 | NR | KANSL1, KANSL1, KANSL1 | | | ENSG00000120071, E | NA | NA | rs4471723-T | rs4471723 | 0 | 4471723 | intron_variant | 0 | 0.219254 | 5.00E-09 | 8.30103 | | 5.84 | z score inc | NR [10847151] (imputed) | N |
| 9 | 17:44118848 | 17 | 44205690 | rs4471723 | 4/08/2022 | 3.6E+07 | Fan CC | 3/05/2022 | Nat Commun | www.ncbi.nlm.nih.gov/pubmed/35505052 | Multivariate genome-wide association study on tissue-sensitive diffusion metrics highlights pathways that shape the human brain. | Whole brain restricted isotropic diffusion (multivariate analysis) | 23,543 Eur | 11,555 Eur | 17q21.31 | | KANSL1, KANSL1, KANSL1 | | | ENSG00000120071, E | NA | NA | rs4471723-? | rs4471723 | 0 | 4471723 | intron_variant | 0 | NR | ####### | 104 | | NA | | NR [NR] (imputed) | N |
| 9 | 17:44118848 | 17 | 44192618 | rs4606752 | 30/08/2017 | 2.8E+07 | Astle WJ | 17/11/2016 | Cell | www.ncbi.nlm.nih.gov/pubmed/27863252 | The Allelic Landscape of Human Blood Cell Trait Variation and Links to Common Complex Disease. | High light scatter reticulocyte count | 170,761 Eu | NA | 17q21.31 | KANSL1 | KANSL1, KANSL1, KANSL1 | | | ENSG00000120071, E | NA | NA | rs4606752-G | rs4606752 | 0 | 4606752 | intron_variant | 0 | 0.2267 | 2.00E-16 | 15.69897 | | 0.034705 | [0.026-0.0 | Affymetrix [~ 2500000] (imputed) | N |
| 9 | 17:44118848 | 17 | 44192618 | rs4606752 | 30/08/2017 | 2.8E+07 | Astle WJ | 17/11/2016 | Cell | www.ncbi.nlm.nih.gov/pubmed/27863252 | The Allelic Landscape of Human Blood Cell Trait Variation and Links to Common Complex Disease. | Reticulocyte count | 170,641 Eu | NA | 17q21.31 | KANSL1 | KANSL1, KANSL1, KANSL1 | | | ENSG00000120071, E | NA | NA | rs4606752-G | rs4606752 | 0 | 4606752 | intron_variant | 0 | 0.2266 | 1.00E-17 | 17 | | 0.036083 | [0.028-0.0 | Affymetrix [~ 2500000] (imputed) | N |
| 9 | 17:43556807 | 17 | 43471489 | rs4763 | 30/11/2017 | 2.9E+07 | Michailidou K | 23/10/2017 | Nature | www.ncbi.nlm.nih.gov/pubmed/29059683 | Association analysis identifies 65 new breast cancer risk loci. | Breast cancer | 76,192 Eur | 46,785 Eur | 17q21.31 | NR | ARHGAP27 | | | ENSG00000159314 | NA | NA | rs4763-A | rs4763 | 0 | 4763 | 3_prime_UTR_variant | 0 | 0.1745 | 2.00E-08 | 7.69897 | (EA) | 0.0464 | [0.03-0.06 | Illumina [~ 11800000] (imputed) | N |
| 9 | 17:43556807 | 17 | 43471489 | rs4763 | 5/02/2020 | 3.2E+07 | Zhao B | 30/10/2019 | Mol Psychiatry | www.ncbi.nlm.nih.gov/pubmed/31666681 | Large-scale GWAS reveals genetic architecture of brain white matter microstructure and genetic overlap with cognitive and mental health traits (nÔÇë=ÔÇë17,706). | White matter microstructure (axial diusivities) | 17,706 Eur | NA | 17q21.31 | NR | ARHGAP27 | | | ENSG00000159314 | NA | NA | rs4763-? | rs4763 | 0 | 4763 | 3_prime_UTR_variant | 0 | | 9.00E-10 | 9.045757 | (Superior l | NA | | NR [8955960] (imputed) | N |
| 9 | 17:44118848 | 17 | 44787313 | rs538628 | 4/05/2021 | 3.3E+07 | Hofer E | 22/09/2020 | Nat Commun | www.ncbi.nlm.nih.gov/pubmed/32963231 | Genetic correlations and genome-wide associations of cortical structure in general population samples of 22,824 adults. | Cortical surface area | 18,617 Eur | 22,363 Eur | 17q21.31 | NSF | NSF, NSF, NSF | | | ENSG00000278174, E | NA | NA | rs538628-? | rs538628 | 0 | 538628 | intron_variant | 0 | | 1.00E-43 | 43 | (Global) | NA | | Affymetrix, Illumina [NR] (imputed) | N |
| 9 | 17:44118848 | 17 | 44787313 | rs538628 | 22/01/2019 | 3E+07 | Nagel M | 2/03/2018 | Nat Commun | www.ncbi.nlm.nih.gov/pubmed/29500382 | Item-level analyses reveal genetic heterogeneity in neuroticism. | Feeling nervous | 373,121 Eu | NA | 17q21.31 | NR | NSF, NSF, NSF | | | ENSG00000278174, E | NA | NA | rs538628-C | rs538628 | 0 | 538628 | intron_variant | 0 | 0.206782 | 9.00E-13 | 12.04576 | | 7.14 | z score inc | NR [10847151] (imputed) | N |
| 9 | 17:44118848 | 17 | 44787313 | rs538628 | 3/12/2018 | 2.8E+07 | Hagenaars SP | 14/02/2017 | PLoS Genet | www.ncbi.nlm.nih.gov/pubmed/28196072 | Genetic prediction of male pattern baldness. | Male-pattern baldness | 52,874 British ancestr | | 17q21.31 | ARL17A, LRRC37 | NSF, NSF, NSF | | | ENSG00000278174, E | NA | NA | rs538628-C | rs538628 | 0 | 538628 | intron_variant | 0 | NR | 4.00E-27 | 26.39794 | | 0.080084 | NR unit de | NR [NR] (imputed) | N |
| 9 | 17:44118848 | 17 | 44284057 | rs548785130 | 5/02/2020 | 3.2E+07 | Zhao B | 30/10/2019 | Mol Psychiatry | www.ncbi.nlm.nih.gov/pubmed/31666681 | Large-scale GWAS reveals genetic architecture of brain white matter microstructure and genetic overlap with cognitive and mental health traits (nÔÇë=ÔÇë17,706). | White matter microstructure (radial diusivities) | 17,706 Eur | NA | 17q21.31 | NR | KANSL1, KANSL1, KANSL1 | | | ENSG00000120071, E | NA | NA | rs548785130-? | rs548785130 | 1 | 2696457 | intron_variant | 0 | | 6.00E-12 | 11.22185 | (Fornix (cre | NA | | NR [8955960] (imputed) | N |
| 9 | 17:44118848 | 17 | 44338694 | rs550530305 | 5/02/2020 | 3.2E+07 | Zhao B | 30/10/2019 | Mol Psychiatry | www.ncbi.nlm.nih.gov/pubmed/31666681 | Large-scale GWAS reveals genetic architecture of brain white matter microstructure and genetic overlap with cognitive and mental health traits (nÔÇë=ÔÇë17,706). | White matter microstructure (axial diusivities) | 17,706 Eur | NA | 17q21.31 | NR | MAPK8IP1P1 - AR | ENSG00000282723 | ENSG00000228696 | | 16284 | 13456 | rs550530305-? | rs550530305 | 1 | 2732687 | intergenic_variant | 1 | | 2.00E-08 | 7.69897 | (Anterior c | NA | | NR [8955960] (imputed) | N |
| 9 | 17:44118848 | 17 | 44324572 | rs555203107 | 17/12/2018 | 2.9E+07 | Pilling LC | 28/09/2017 | PLoS One | www.ncbi.nlm.nih.gov/pubmed/28957414 | Red blood cell distribution width: Genetic evidence for aging pathways in 116,666 volunteers. | Red cell distribution width | 116,666 B | NA | 17q21.31 | NR | MAPK8IP1P1 - AR | ENSG00000282723 | ENSG00000228696 | | 2162 | 27578 | rs555203107-G | rs555203107 | 1 | 62073112 | downstream_gene_variant | 1 | NR | 8.00E-28 | 27.09691 | | 0.05169 | [0.042-0.0 | Affymetrix [16832071] (imputed) | N |
| 9 | 17:44118848 | 17 | 43844560 | rs55657917 | 16/08/2018 | 3E+07 | Klimentidis YC | 13/06/2018 | Int J Obes (Lond) | www.ncbi.nlm.nih.gov/pubmed/29899525 | Genome-wide association study of habitual physical activity in over 377,000 UK Biobank participants identifies multiple variants including CADM2 and APOE. | Accelerometer-based physical activity measurement (average acceleration) | 91,084 Eur | NA | 17q21.31 | CRHR1 | LINC02210-CRHR1, LINC02210-CRHR1 | | | ENSG00000282456, E | NA | NA | rs55657917-T | rs55657917 | 0 | 55657917 | intron_variant | 0 | 0.78 | 5.00E-12 | 11.30103 | | 0.3 | milli-gravit | Affymetrix [11800000] (imputed) | N |
| 9 | 17:44118848 | 17 | 43844560 | rs55657917 | 22/01/2019 | 3E+07 | Nagel M | 2/03/2018 | Nat Commun | www.ncbi.nlm.nih.gov/pubmed/29500382 | Item-level analyses reveal genetic heterogeneity in neuroticism. | Experiencing mood swings | 373,733 Eu | NA | 17q21.31 | HOXB2 | LINC02210-CRHR1, LINC02210-CRHR1 | | | ENSG00000282456, E | NA | NA | rs55657917-T | rs55657917 | 0 | 55657917 | intron_variant | 0 | 0.213403 | 3.00E-20 | 19.52288 | | 9.23 | z score dec | NR [10847151] (imputed) | N |
| 9 | 17:44118848 | 17 | 43844560 | rs55657917 | 22/01/2019 | 3E+07 | Nagel M | 2/03/2018 | Nat Commun | www.ncbi.nlm.nih.gov/pubmed/29500382 | Item-level analyses reveal genetic heterogeneity in neuroticism. | Feeling hurt | 372,047 Eu | NA | 17q21.31 | HOXB2 | LINC02210-CRHR1, LINC02210-CRHR1 | | | ENSG00000282456, E | NA | NA | rs55657917-T | rs55657917 | 0 | 55657917 | intron_variant | 0 | 0.213403 | 7.00E-29 | 28.1549 | | 11.16 | z score dec | NR [10847151] (imputed) | N |
| 9 | 17:44118848 | 17 | 43844560 | rs55657917 | 18/01/2019 | 3.1E+07 | Doherty A | 10/12/2018 | Nat Commun | www.ncbi.nlm.nih.gov/pubmed/30531941 | GWAS identifies 14 loci for device-measured physical activity and sleep duration. | Physical activity (overall physical activity time) | 91,105 Eur | NA | 17q21.31 | LINC02210-CRH | LINC02210-CRHR1, LINC02210-CRHR1 | | | ENSG00000282456, E | NA | NA | rs55657917-T | rs55657917 | 0 | 55657917 | intron_variant | 0 | 0.779584 | 8.00E-12 | 11.09691 | | 0.036815 | [0.026-0.0 | NR [9926106] (imputed) | N |
| 9 | 17:44118848 | 17 | 44164323 | rs55692232 | 14/09/2020 | 3.2E+07 | Zhou H | 25/05/2020 | Nat Neurosci | www.ncbi.nlm.nih.gov/pubmed/32451486 | Genome-wide meta-analysis of problematic alcohol use in 435,563 individuals yields insights into biology and relationships with other traits. | Alcohol consumption (drinks per week) (MTAG) | 972,915 Eu | NA | 17q21.31 | NR | KANSL1, KANSL1, KANSL1 | | | ENSG00000120071, E | NA | NA | rs55692232-A | rs55692232 | 0 | 55692232 | intron_variant | 0 | 0.205 | 7.00E-21 | 20.1549 | | 0.020043 | [0.016-0.0 | Affymetrix [NR] (imputed) | N |
| 9 | 17:44118848 | 17 | 44517766 | rs557236423 | 31/05/2022 | 3.4E+07 | Smith SM | 19/04/2021 | Nat Neurosci | www.ncbi.nlm.nih.gov/pubmed/33875891 | An expanded set of genome-wide association studies of brain imaging phenotypes in UK Biobank. | aseg global volume BrainSeg | 21,282 Brit | 10,686 Brit | 17q21.31 | | NSFP1, NSFP1, NSFP1 | | | ENSG00000260075, E | NA | NA | rs557236423-A | rs557236423 | 0 | 5.57E+08 | intron_variant | 0 | 0.259 | 1.00E-13 | 13 | | 0.093 | [0.068-0.1 | Affymetrix [17103079] (imputed) | N |
| 9 | 17:44118848 | 17 | 44034575 | rs55746658 | 10/03/2022 | 3.5E+07 | Yang Y | 4/11/2021 | Circ Genom Precis Med | www.ncbi.nlm.nih.gov/pubmed/34732054 | Identification of Functional Genetic Determinants of Cardiac Troponin T and I in a Multiethnic Population and Causal Associations With Atrial Fibrillation. | High-sensitivity cardiac troponin I concentration (MTAG) | 12,730 Eur | NA | 17q21.31 | | MAPT, MAPT, MAPT | | | ENSG00000277956, E | NA | NA | rs55746658-? | rs55746658 | 0 | 55746658 | intron_variant | 0 | NR | 2.00E-06 | 5.69897 | | 4.72 | Z score de | Affymetrix, Illumina [NR] (imputed) | N |
| 9 | 17:44118848 | 17 | 44034575 | rs55746658 | 10/03/2022 | 3.5E+07 | Yang Y | 4/11/2021 | Circ Genom Precis Med | www.ncbi.nlm.nih.gov/pubmed/34732054 | Identification of Functional Genetic Determinants of Cardiac Troponin T and I in a Multiethnic Population and Causal Associations With Atrial Fibrillation. | High-sensitivity cardiac troponin T levels (MTAG) | 18,590 Eur | NA | 17q21.31 | | MAPT, MAPT, MAPT | | | ENSG00000277956, E | NA | NA | rs55746658-? | rs55746658 | 0 | 55746658 | intron_variant | 0 | NR | 2.00E-06 | 5.69897 | | 4.72 | Z score de | Affymetrix, Illumina [NR] (imputed) | N |
| 9 | 17:44118848 | 17 | 43679193 | rs55826010 | 31/05/2022 | 3.4E+07 | Smith SM | 19/04/2021 | Nat Neurosci | www.ncbi.nlm.nih.gov/pubmed/33875891 | An expanded set of genome-wide association studies of brain imaging phenotypes in UK Biobank. | aparc-a2009s lh thickness S-orbital-H-Shaped | 21,282 Brit | 10,685 Brit | 17q21.31 | | MAPK8IP1P2, MAPK8IP1P2 | | | ENSG00000282351, E | NA | NA | rs55826010-A | rs55826010 | 0 | 55826010 | non_coding_transcript_exon | 0 | 0.237 | 7.00E-09 | 8.154902 | | 0.067 | [0.045-0.0 | Affymetrix [17103079] (imputed) | N |
| 9 | 17:44118848 | 17 | 43757450 | rs55974014 | 9/01/2021 | 3.3E+07 | Cuellar-Partida G | 28/09/2020 | Nat Hum Behav | www.ncbi.nlm.nih.gov/pubmed/32989287 | Genome-wide association study identifies 48 common genetic variants associated with handedness. | Left-handedness | 1,766,671 | NA | 17q21.31 | CRHR1 | LINC02210-CRHR1, LINC02210-CRHR1 | | | ENSG00000282456, E | NA | NA | rs55974014-A | rs55974014 | 0 | 55974014 | intron_variant | 0 | 0.2142 | 3.00E-30 | 29.52288 | | 0.94989 | [NR] | Illumina [12493444] (imputed) | N |
| 9 | 17:44118848 | 17 | 44013966 | rs56072903 | 4/08/2022 | 3.6E+07 | May-Wilson S | 18/05/2022 | Nat Commun | www.ncbi.nlm.nih.gov/pubmed/35585065 | Large-scale GWAS of food liking reveals genetic determinants and genetic correlations with distinct neurophysiological traits. | F-small fish liking (derived food-liking factor) | 150,746 Eu | NA | 17q21.31 | | MAPT, MAPT, MAPT | | | ENSG00000277956, E | NA | NA | rs56072903-C | rs56072903 | 0 | 56072903 | intron_variant | 0 | 0.2032 | 6.00E-12 | 11.22185 | | 0.08537 | [0.061-0.1 | Affymetrix [9605657] (imputed) | N |
| 9 | 17:44118848 | 17 | 44013966 | rs56072903 | 4/08/2022 | 3.6E+07 | May-Wilson S | 18/05/2022 | Nat Commun | www.ncbi.nlm.nih.gov/pubmed/35585065 | Large-scale GWAS of food liking reveals genetic determinants and genetic correlations with distinct neurophysiological traits. | Herring liking | 150,609 Eu | NA | 17q21.31 | | MAPT, MAPT, MAPT | | | ENSG00000277956, E | NA | NA | rs56072903-C | rs56072903 | 0 | 56072903 | intron_variant | 0 | 0.2024 | 1.00E-13 | 13 | | 0.03465 | [0.025-0.0 | Affymetrix [11353280] (imputed) | N |
| 9 | 17:44118848 | 17 | 44013966 | rs56072903 | 4/08/2022 | 3.6E+07 | May-Wilson S | 18/05/2022 | Nat Commun | www.ncbi.nlm.nih.gov/pubmed/35585065 | Large-scale GWAS of food liking reveals genetic determinants and genetic correlations with distinct neurophysiological traits. | Mackerel liking | 156,695 Eu | NA | 17q21.31 | | MAPT, MAPT, MAPT | | | ENSG00000277956, E | NA | NA | rs56072903-C | rs56072903 | 0 | 56072903 | intron_variant | 0 | 0.203 | 2.00E-08 | 7.69897 | | 0.02588 | [0.017-0.0 | Affymetrix [11353280] (imputed) | N |
| 9 | 17:44118848 | 17 | 44077851 | rs56108300 | 21/09/2020 | 3.3E+07 | Chen MH | 1/09/2020 | Cell | www.ncbi.nlm.nih.gov/pubmed/32888493 | Trans-ethnic and Ancestry-Specific Blood-Cell Genetics in 746,667 Individuals from 5 Global Populations. | Neutrophil count | 519,288 Eu | NA | 17q21.31 | NR | MAPT, MAPT, MAPT | | | ENSG00000277956, E | NA | NA | rs56108300-A | rs56108300 | 0 | 56108300 | intron_variant | 0 | 0.224125 | 1.00E-24 | 24 | | 0.023682 | [0.019-0.0 | Affymetrix, Illumina [50250472] (imputed) | N |
| 9 | 17:44118848 | 17 | 43512318 | rs56168933 | 20/07/2020 | 3.2E+07 | Shin J | 20/03/2020 | Cereb Cortex | www.ncbi.nlm.nih.gov/pubmed/32198502 | Global and Regional Development of the Human Cerebral Cortex: Molecular Architecture and Occupational Aptitudes. | Cortical surface area (global PC1) | 23,784 Eur | 19,512 Eur | 17q21.31 | ARHGAP27 | ARHGAP27 - PLEI | ENSG00000159314 | ENSG00000277111 | | 531 | 948 | rs56168933-? | rs56168933 | 0 | 56168933 | intergenic_variant | 1 | NR | 8.00E-28 | 27.09691 | | 2.0304 | [1.67-2.39 | Affymetrix, Illumina [NR] (imputed) | N |
| 9 | 17:43564222 | 17 | 43555253 | rs56192752 | 19/10/2018 | 3E+07 | Lee JJ | 23/07/2018 | Nat Genet | www.ncbi.nlm.nih.gov/pubmed/30038396 | Gene discovery and polygenic prediction from a genome-wide association study of educational attainment in 1.1 million individuals. | Educational attainment (years of education) | up to 1,13 | NA | 17q21.31 | | PLEKHM1 | | | ENSG00000277111 | NA | NA | rs56192752-A | rs56192752 | 0 | 56192752 | 3_prime_UTR_variant | 0 | 0.8218 | 9.00E-31 | 30.04576 | (condition | 0.0218 | [0.018-0.0 | Affymetrix, Illumina [10000000] (imputed) | N |
| 9 | 17:43564222 | 17 | 43555253 | rs56192752 | 24/09/2021 | 3.3E+07 | Wendt FR | 21/12/2020 | Nat Hum Behav | www.ncbi.nlm.nih.gov/pubmed/33349686 | Multivariate genome-wide analysis of education, socioeconomic status and brain phenome. | Neuroticism conditioned on average household income before tax (multi-trait conditioning and joint analysis) | 170,911 Eu | NA | 17q21.31 | | PLEKHM1 | | | ENSG00000277111 | NA | NA | rs56192752-? | rs56192752 | 0 | 56192752 | 3_prime_UTR_variant | 0 | NR | 4.00E-08 | 7.39794 | | NA | | NR [NR] | N |
| 9 | 17:43564222 | 17 | 43555253 | rs56192752 | 24/09/2021 | 3.3E+07 | Wendt FR | 21/12/2020 | Nat Hum Behav | www.ncbi.nlm.nih.gov/pubmed/33349686 | Multivariate genome-wide analysis of education, socioeconomic status and brain phenome. | Neuroticism conditioned on educational attainment (multi-trait conditioning and joint analysis) | 170,911 Eu | NA | 17q21.31 | | PLEKHM1 | | | ENSG00000277111 | NA | NA | rs56192752-? | rs56192752 | 0 | 56192752 | 3_prime_UTR_variant | 0 | NR | 1.00E-10 | 10 | | NA | | NR [NR] | N |
| 9 | 17:43564222 | 17 | 43555253 | rs56192752 | 24/09/2021 | 3.3E+07 | Wendt FR | 21/12/2020 | Nat Hum Behav | www.ncbi.nlm.nih.gov/pubmed/33349686 | Multivariate genome-wide analysis of education, socioeconomic status and brain phenome. | Neuroticism conditioned on Townsend deprivation index (multi-trait conditioning and joint analysis) | 170,911 Eu | NA | 17q21.31 | | PLEKHM1 | | | ENSG00000277111 | NA | NA | rs56192752-? | rs56192752 | 0 | 56192752 | 3_prime_UTR_variant | 0 | NR | 7.00E-14 | 13.1549 | | NA | | NR [NR] | N |
| 9 | 17:44118848 | 17 | 44041107 | rs56280951 | 22/01/2019 | 3E+07 | Nagel M | 2/03/2018 | Nat Commun | www.ncbi.nlm.nih.gov/pubmed/29500382 | Item-level analyses reveal genetic heterogeneity in neuroticism. | Feeling miserable | 376,097 Eu | NA | 17q21.31 | ACBD4 | MAPT, MAPT, MAPT | | | ENSG00000277956, E | NA | NA | rs56280951-A | rs56280951 | 0 | 56280951 | intron_variant | 0 | 0.21432 | 8.00E-13 | 12.09691 | | 7.17 | z score inc | NR [10847151] (imputed) | N |
| 9 | 17:44118848 | 17 | 43853922 | rs56303031 | 8/02/2019 | 3.1E+07 | Kichaev G | 27/12/2018 | Am J Hum Genet | www.ncbi.nlm.nih.gov/pubmed/30595370 | Leveraging Polygenic Functional Enrichment to Improve GWAS Power. | Heel bone mineral density | approxima | NA | 17q21.31 | | LINC02210-CRHR1, LINC02210-CRHR1 | | | ENSG00000282456, E | NA | NA | rs56303031-? | rs56303031 | 0 | 56303031 | intron_variant | 0 | NR | 6.00E-24 | 23.22185 | | NA | | NR [~ 8900000] (imputed) | N |

| | | | | | | | | | | | | | | | | | | | | | | | | | | | | | | | | | | | | |
|---|---|---|---|---|---|---|---|---|---|---|---|---|---|---|---|---|---|---|---|---|---|---|---|---|---|---|---|---|---|---|---|---|---|---|---|---|
| 9 | 17:44118848 | 17 | 43871982 | rs56319902 | 19/10/2018 | 3E+07 | Lee JJ | 23/07/2018 | Nat Genet | www.ncbi.nlm.nih.gov/pubmed/30038396 | Gene discovery and polygenic prediction from a genome-wide association study of educational attainment in 1.1 million individuals. | Educational attainment (years of education) | up to 1,13 | NA | 17q21.31 | | CRHR1, CRHR1, CRHR1, LINC02210-CRHR1, LINC02210-C | | | ENSG00000276191, E | NA | NA | rs56319902-T | rs56319902 | 0 | 56319902 | intron_variant | 0 | 0.2155 | 6.00E-33 | 32.22185 | | 0.0206 | [0.017-0.0 | Affymetrix, Illumina [10000000] (imputed) | N |
| 9 | 17:44118848 | 17 | 43871982 | rs56319902 | 7/03/2022 | 3.5E+07 | van der Meer D | 15/12/2021 | Sci Adv | www.ncbi.nlm.nih.gov/pubmed/34910505 | The genetic architecture of human cortical folding. | Vertex-wise sulcal depth | 8,072 indiv | NA | 17q21.31 | | CRHR1, CRHR1, CRHR1, LINC02210-CRHR1, LINC02210-C | | | ENSG00000276191, E | NA | NA | rs56319902-T | rs56319902 | 0 | 56319902 | intron_variant | 0 | 0.24 | 2.00E-15 | 14.69897 | | NA | | NR [13131314] (imputed) | N |
| 9 | 17:44118848 | 17 | 43871982 | rs56319902 | 7/03/2022 | 3.5E+07 | van der Meer D | 15/12/2021 | Sci Adv | www.ncbi.nlm.nih.gov/pubmed/34910505 | The genetic architecture of human cortical folding. | Vertex-wise sulcal depth | 5,199 indiv | NA | 17q21.31 | | CRHR1, CRHR1, CRHR1, LINC02210-CRHR1, LINC02210-C | | | ENSG00000276191, E | NA | NA | rs56319902-T | rs56319902 | 0 | 56319902 | intron_variant | 0 | 0.24 | 3.00E-06 | 5.522879 | | NA | | Affymetrix [9061022] (imputed) | N |
| 9 | 17:44118848 | 17 | 43871982 | rs56319902 | 7/03/2022 | 3.5E+07 | van der Meer D | 15/12/2021 | Sci Adv | www.ncbi.nlm.nih.gov/pubmed/34910505 | The genetic architecture of human cortical folding. | Vertex-wise sulcal depth | 33,748 Eur | NA | 17q21.31 | | CRHR1, CRHR1, CRHR1, LINC02210-CRHR1, LINC02210-C | | | ENSG00000276191, E | NA | NA | rs56319902-T | rs56319902 | 0 | 56319902 | intron_variant | 0 | 0.24 | ####### | 230 | | 32.43 | z score inc | Affymetrix [9061022] (imputed) | N |
| 9 | 17:44118848 | 17 | 44326712 | rs56368550 | 4/08/2022 | 3.6E+07 | May-Wilson S | 18/05/2022 | Nat Commun | www.ncbi.nlm.nih.gov/pubmed/35585065 | Large-scale GWAS of food liking reveals genetic determinants and genetic correlations with distinct neurophysiological traits. | Salad dressing liking | 159,310 Eu | NA | 17q21.31 | | MAPK8IP1P1 - AR | ENSG00000282723 | ENSG00000228696 | | 4302 | 25438 | rs56368550-C | rs56368550 | 0 | 56368550 | downstream_gene_variant | 1 | 0.1989 | 1.00E-09 | 9 | | 0.02772 | [0.019-0.0 | Affymetrix [11353280] (imputed) | N |
| 9 | 17:43803189 | 17 | 43798787 | rs564681262 | 5/02/2020 | 3.2E+07 | Zhao B | 30/10/2019 | Mol Psychiatry | www.ncbi.nlm.nih.gov/pubmed/31666681 | Large-scale GWAS reveals genetic architecture of brain white matter microstructure and genetic overlap with cognitive and mental health traits (nÔÇë=ÔÇë17,706). | White matter microstructure (axial diusivities) | 17,706 Eur | NA | 17q21.31 | NR | LINC02210-CRHR1, LINC02210-CRHR1 | | | ENSG00000282456, E | NA | NA | rs564681262-? | rs564681262 | 1 | 7214673 | intron_variant | 0 | | 2.00E-08 | 7.69897 | (Inferior fro | NA | | NR [8955960] (imputed) | N |
| 9 | 17:44118848 | 17 | 43668850 | rs564850730 | 4/08/2022 | 3.6E+07 | May-Wilson S | 18/05/2022 | Nat Commun | www.ncbi.nlm.nih.gov/pubmed/35585065 | Large-scale GWAS of food liking reveals genetic determinants and genetic correlations with distinct neurophysiological traits. | Shellfish liking | 155,491 Eu | NA | 17q21.31 | | DND1P1 - MAPK8 | ENSG00000282614 | ENSG00000263503 | | 4555 | 9385 | rs564850730-C | rs564850730 | 0 | 5.65E+08 | upstream_gene_variant | 1 | 0.1708 | 5.00E-11 | 10.30103 | | 0.03361 | [0.024-0.0 | Affymetrix [11353280] (imputed) | N |
| 9 | 17:44118848 | 17 | 44364335 | rs578102530 | 30/08/2022 | 3.2E+07 | Cole JB | 19/03/2020 | Nat Commun | www.ncbi.nlm.nih.gov/pubmed/32193382 | Comprehensive genomic analysis of dietary habits in UK Biobank identifies hundreds of genetic associations. | Alcohol consumption (drinks per month) (UKB data field 1578, 4424) | 449,210 Eu | NA | 17q21.31 | | ARL17B, ARL17B, ARL17B | | | ENSG00000275748, E | NA | NA | rs578102530-G | rs578102530 | 0 | 5.78E+08 | splice_region_variant | 0 | 0.812702 | 8.00E-23 | 22.09691 | | 0.028136 | [0.023-0.0 | NR [NR] (imputed) | N |
| 9 | 17:43850645 | 17 | 43850645 | rs60814418 | 1/07/2022 | 3.5E+07 | Okbay A | 31/03/2022 | Nat Genet | www.ncbi.nlm.nih.gov/pubmed/35361970 | Polygenic prediction of educational attainment within and between families from genome-wide association analyses in 3 million individuals. | Educational attainment | 3,037,499 | NA | 17q21.31 | | LINC02210-CRHR1, LINC02210-CRHR1 | | | ENSG00000282456, E | NA | NA | rs60814418-T | rs60814418 | 0 | 60814418 | intron_variant | 0 | 0.2242 | 5.00E-72 | 71.30103 | | 0.022627 | [0.02-0.02 | Illumina [10675380] (imputed) | N |
| 9 | 17:44118848 | 17 | 43785349 | rs61667602 | 14/09/2022 | 3.6E+07 | COVID-19 Host Genetics Initiative | 3/08/2022 | Nature | www.ncbi.nlm.nih.gov/pubmed/35922517 | A first update on mapping the human genetic architecture of COVID-19. | COVID-19 (critical illness vs population) | 188 African | NA | 17q21.31 | | LINC02210-CRHR1, LINC02210-CRHR1 | | | ENSG00000282456, E | NA | NA | rs61667602-C | rs61667602 | 0 | 61667602 | intron_variant | 0 | 0.195 | 6.00E-08 | 7.221849 | | 0.89 | [0.85-0.93 | Affymetrix, Illumina [NR] (imputed) | N |
| 9 | 17:44118848 | 17 | 43785349 | rs61667602 | 14/09/2022 | 3.6E+07 | COVID-19 Host Genetics Initiative | 3/08/2022 | Nature | www.ncbi.nlm.nih.gov/pubmed/35922517 | A first update on mapping the human genetic architecture of COVID-19. | COVID-19 (hospitalized vs population) | 2,113 Afric | NA | 17q21.31 | | LINC02210-CRHR1, LINC02210-CRHR1 | | | ENSG00000282456, E | NA | NA | rs61667602-C | rs61667602 | 0 | 61667602 | intron_variant | 0 | 0.189 | 4.00E-12 | 11.39794 | | 0.91 | [0.89-0.94 | Affymetrix, Illumina [NR] (imputed) | N |
| 9 | 17:44118848 | 17 | 43785349 | rs61667602 | 18/09/2019 | 3.1E+07 | Gelernter J | 8/04/2019 | Biol Psychiatry | www.ncbi.nlm.nih.gov/pubmed/31151762 | Genome-wide Association Study of Maximum Habitual Alcohol Intake in >140,000 U.S. European and African American Veterans Yields Novel Risk Loci. | Maximum habitual alcohol consumption | 126,936 Eu | NA | 17q21.31 | CRHR1 | LINC02210-CRHR1, LINC02210-CRHR1 | | | ENSG00000282456, E | NA | NA | rs61667602-? | rs61667602 | 0 | 61667602 | intron_variant | 0 | NR | 1.00E-13 | 13 | | 7.439 | NR z score | Affymetrix [up to 6007810] (imputed) | N |
| 9 | 17:44118848 | 17 | 43744203 | rs62053943 | 24/08/2021 | 3.4E+07 | Christakoudi S | 21/05/2021 | Sci Rep | www.ncbi.nlm.nih.gov/pubmed/34021172 | GWAS of allometric body-shape indices in UK Biobank identifies loci suggesting associations with morphogenesis, organogenesis, adrenal cell renewal and cancer. | A body shape index | 219,872 Bi | NA | 17q21.31 | CRHR1, RP11-10 | LINC02210-CRHR1, LINC02210-CRHR1 | | | ENSG00000282456, E | NA | NA | rs62053943-T | rs62053943 | 0 | 62053943 | intron_variant | 0 | NR | 1.00E-10 | 10 | | 0.025622 | [0.018-0.0 | Affymetrix [321169] (imputed) | N |
| 9 | 17:44118848 | 17 | 43744203 | rs62053943 | 2/05/2019 | 3.1E+07 | Hill WD | 13/03/2019 | Mol Psychiatry | www.ncbi.nlm.nih.gov/pubmed/30867560 | Genetic contributions to two special factors of neuroticism are associated with affluence, higher intelligence, better health, and longer life. | General factor of neuroticism | 270,059 Bi | NA | 17q21.31 | RP11-105N13.4 | LINC02210-CRHR1, LINC02210-CRHR1 | | | ENSG00000282456, E | NA | NA | rs62053943-C | rs62053943 | 0 | 62053943 | intron_variant | 0 | NR | 4.00E-14 | 13.39794 | | 0.012948 | [0.0096-0. | Affymetrix [18485882] (imputed) | N |
| 9 | 17:44118848 | 17 | 43744203 | rs62053943 | 27/01/2020 | 3.2E+07 | Bandres-Ciga S | 29/10/2019 | Mov Disord | www.ncbi.nlm.nih.gov/pubmed/31660654 | The Genetic Architecture of Parkinson Disease in Spain: Characterizing Population-Specific Risk, Differential Haplotype Structures, and Providing Etiologic Insight. | Parkinson's disease | 4,639 Spar | NA | 17q21.31 | CRHR1 | LINC02210-CRHR1, LINC02210-CRHR1 | | | ENSG00000282456, E | NA | NA | rs62053943-T | rs62053943 | 0 | 62053943 | intron_variant | 0 | 0.17489 | 8.00E-09 | 8.09691 | | 0.2957 | [0.2-0.4] u | Illumina [9911207] (imputed) | N |
| 9 | 17:44118848 | 17 | 43744203 | rs62053943 | 19/12/2019 | 3.2E+07 | Nalls MA | 1/12/2019 | Lancet Neurol | www.ncbi.nlm.nih.gov/pubmed/31701892 | Identification of novel risk loci, causal insights, and heritable risk for Parkinson's disease: a meta-analysis of genome-wide association studies. | Parkinson's disease or first degree relation to individual with Parkinson's disease | 15,056 Eur | 22,632 Eur | 17q21.31 | CRHR1 | LINC02210-CRHR1, LINC02210-CRHR1 | | | ENSG00000282456, E | NA | NA | rs62053943-T | rs62053943 | 0 | 62053943 | intron_variant | 0 | 0.1552 | 4.00E-68 | 67.39794 | | 0.27 | [0.24-0.3] | NR [7784415] (imputed) | N |
| 9 | 17:44118848 | 17 | 43744203 | rs62053943 | 5/02/2020 | 3.2E+07 | Zhao B | 30/10/2019 | Mol Psychiatry | www.ncbi.nlm.nih.gov/pubmed/31666681 | Large-scale GWAS reveals genetic architecture of brain white matter microstructure and genetic overlap with cognitive and mental health traits (nÔÇë=ÔÇë17,706). | White matter microstructure (radial diusivities) | 17,706 Eur | NA | 17q21.31 | NR | LINC02210-CRHR1, LINC02210-CRHR1 | | | ENSG00000282456, E | NA | NA | rs62053943-? | rs62053943 | 0 | 62053943 | intron_variant | 0 | | 2.00E-08 | 7.69897 | (Anterior li | NA | | NR [8955960] (imputed) | N |
| 9 | 17:44118848 | 17 | 43950441 | rs62055489 | 15/12/2021 | 3.5E+07 | Shadrin AA | 21/09/2021 | Neuroimage | www.ncbi.nlm.nih.gov/pubmed/34560273 | Vertex-wise multivariate genome-wide association study identifies 780 unique genetic loci associated with cortical morphology. | Cortical surface area | 35,657 Wh | NA | 17q21.31 | NR | MAPT-AS1, MAPT-AS1, MAPT-AS1 | | | ENSG00000264589, E | NA | NA | rs62055489-A | rs62055489 | 0 | 62055489 | intron_variant | 0 | 0.2195 | 1.00E-17 | 17 | | NA | | NR [9056863] (imputed) | N |
| 9 | 17:44118848 | 17 | 43964539 | rs62055544 | 22/01/2019 | 3E+07 | Nagel M | 2/03/2018 | Nat Commun | www.ncbi.nlm.nih.gov/pubmed/29500382 | Item-level analyses reveal genetic heterogeneity in neuroticism. | Feeling fed-up | 374,971 Eu | NA | 17q21.31 | NR | MAPT-AS1, MAPT-AS1, MAPT-AS1 | | | ENSG00000264589, E | NA | NA | rs62055544-A | rs62055544 | 0 | 62055544 | intron_variant | 0 | 0.216596 | 5.00E-16 | 15.30103 | | 8.1 | z score dec | NR [10847151] (imputed) | N |
| 9 | 17:44118848 | 17 | 43964567 | rs62055546 | 18/03/2019 | 3.1E+07 | Karlsson Linner R | 14/01/2019 | Nat Genet | www.ncbi.nlm.nih.gov/pubmed/30643258 | Genome-wide association analyses of risk tolerance and risky behaviors in over 1 million individuals identify hundreds of loci and shared genetic influences. | Alcohol consumption (drinks per week) | 414,343 Eu | NA | 17q21.31 | MAPT | MAPT-AS1, MAPT-AS1, MAPT-AS1 | | | ENSG00000264589, E | NA | NA | rs62055546-A | rs62055546 | 0 | 62055546 | intron_variant | 0 | 0.7779 | 8.00E-25 | 24.09691 | | 0.027622 | [0.022-0.0 | Affymetrix [11514936] (imputed) | N |
| 9 | 17:44118848 | 17 | 43754850 | rs62055691 | 4/10/2019 | 3.1E+07 | Brazel DM | 6/12/2018 | Biol Psychiatry | www.ncbi.nlm.nih.gov/pubmed/30679032 | Exome Chip Meta-analysis Fine Maps Causal Variants and Elucidates the Genetic Architecture of Rare Coding Variants in Smoking and Alcohol┬áUse. | Alcohol consumption (drinks per week) | up to 357,8 | NA | 17q21.31 | CRHR1 | LINC02210-CRHR1, LINC02210-CRHR1 | | | ENSG00000282456, E | NA | NA | rs62055691-C | rs62055691 | 0 | 62055691 | intron_variant | 0 | 0.225 | 2.00E-23 | 22.69897 | | 0.0303 | [0.024-0.0 | Illumina [~ 250000] (imputed) | N |
| 9 | 17:44118848 | 17 | 43758125 | rs62055696 | 1/02/2022 | 3.5E+07 | Hanscombe KB | 9/11/2021 | Genome Med | www.ncbi.nlm.nih.gov/pubmed/34753499 | The genetic case for cardiorespiratory fitness as a clinical vital sign and the routine prescription of physical activity in healthcare. | Physical activity | 89,683 Eur | NA | 17q21.31 | | LINC02210-CRHR1, LINC02210-CRHR1 | | | ENSG00000282456, E | NA | NA | rs62055696-A | rs62055696 | 0 | 62055696 | intron_variant | 0 | 0.78 | 1.00E-11 | 11 | | 0.309 | [0.22-0.4] | Affymetrix [8918919] (imputed) | N |
| 9 | 17:44118848 | 17 | 43758125 | rs62055696 | 1/02/2022 | 3.5E+07 | Hanscombe KB | 9/11/2021 | Genome Med | www.ncbi.nlm.nih.gov/pubmed/34753499 | The genetic case for cardiorespiratory fitness as a clinical vital sign and the routine prescription of physical activity in healthcare. | Physical activity | 50,331 Eur | NA | 17q21.31 | | LINC02210-CRHR1, LINC02210-CRHR1 | | | ENSG00000282456, E | NA | NA | rs62055696-A | rs62055696 | 0 | 62055696 | intron_variant | 0 | 0.78 | 1.00E-09 | 9 | | 0.358 | [0.24-0.47 | Affymetrix [8918919] (imputed) | N |
| 9 | 17:44118848 | 17 | 43758787 | rs62055701 | 22/01/2019 | 3E+07 | Nagel M | 2/03/2018 | Nat Commun | www.ncbi.nlm.nih.gov/pubmed/29500382 | Item-level analyses reveal genetic heterogeneity in neuroticism. | Irritable mood | 366,726 Eu | NA | 17q21.31 | NR | LINC02210-CRHR1, LINC02210-CRHR1 | | | ENSG00000282456, E | NA | NA | rs62055701-A | rs62055701 | 0 | 62055701 | intron_variant | 0 | 0.214491 | 6.00E-13 | 12.22185 | | 7.19 | z score inc | NR [~ 10847151] (imputed) | N |
| 9 | 17:44118848 | 17 | 43838720 | rs62055888 | 14/12/2021 | 3.5E+07 | Sakaue S | 30/09/2021 | Nat Genet | www.ncbi.nlm.nih.gov/pubmed/34594039 | A cross-population atlas of genetic associations for 220 human phenotypes. | Hematocrit | 350,475 Eu | NA | 17q21.31 | | LINC02210-CRHR1, LINC02210-CRHR1 | | | ENSG00000282456, E | NA | NA | rs62055888-T | rs62055888 | 0 | 62055888 | intron_variant | 0 | NR | 2.00E-54 | 53.69897 | | 0.0364 | [0.032-0.0 | Affymetrix, Illumina [20538534] (imputed) | N |
| 9 | 17:44118848 | 17 | 43848750 | rs62055935 | 22/01/2019 | 3E+07 | Nagel M | 2/03/2018 | Nat Commun | www.ncbi.nlm.nih.gov/pubmed/29500382 | Item-level analyses reveal genetic heterogeneity in neuroticism. | Feeling nervous | 373,121 Eu | NA | 17q21.31 | NR | LINC02210-CRHR1, LINC02210-CRHR1 | | | ENSG00000282456, E | NA | NA | rs62055935-T | rs62055935 | 0 | 62055935 | intron_variant | 0 | 0.214475 | 2.00E-15 | 14.69897 | | 7.97 | z score dec | NR [10847151] (imputed) | N |
| 9 | 17:44118848 | 17 | 43975415 | rs62056789 | 27/05/2022 | 3.5E+07 | Wendt FR | 18/02/2022 | Mol Psychiatry | www.ncbi.nlm.nih.gov/pubmed/35181757 | Using phenotype risk scores to enhance gene discovery for generalized anxiety disorder and posttraumatic stress disorder. | Generalized anxiety disorder (mental health questionnaire or predicted) | 489,579 Eu | NA | 17q21.31 | | MAPT-IT1, MAPT-IT1, MAPT-IT1, MAPT, MAPT, MAPT | | | ENSG00000281284, E | NA | NA | rs62056789-A | rs62056789 | 0 | 62056789 | non_coding_transcript_exon | 0 | NR | 4.00E-13 | 12.39794 | | 0.017556 | [0.013-0.0 | Affymetrix [NR] (imputed) | N |
| 9 | 17:44118848 | 17 | 43975415 | rs62056789 | 27/05/2022 | 3.5E+07 | Wendt FR | 18/02/2022 | Mol Psychiatry | www.ncbi.nlm.nih.gov/pubmed/35181757 | Using phenotype risk scores to enhance gene discovery for generalized anxiety disorder and posttraumatic stress disorder. | Generalized anxiety disorder (phenotype risk score) | 489,579 Eu | NA | 17q21.31 | | MAPT-IT1, MAPT-IT1, MAPT-IT1, MAPT, MAPT, MAPT | | | ENSG00000281284, E | NA | NA | rs62056789-A | rs62056789 | 0 | 62056789 | non_coding_transcript_exon_ | 0 | NR | 1.00E-11 | 11 | | 0.016372 | [0.012-0.0 | Affymetrix [NR] (imputed) | N |
| 9 | 17:44118848 | 17 | 43975415 | rs62056789 | 29/01/2020 | 3.2E+07 | Hill WD | 16/12/2019 | Nat Commun | www.ncbi.nlm.nih.gov/pubmed/31844048 | Genome-wide analysis identifies molecular systems and 149 genetic loci associated with income. | Household income | 286,301 Bi | NA | 17q21.31 | MAPT | MAPT-IT1, MAPT-IT1, MAPT-IT1, MAPT, MAPT, MAPT | | | ENSG00000281284, E | NA | NA | rs62056789-C | rs62056789 | 0 | 62056789 | non_coding_transcript_exon | 0 | NR | 3.00E-11 | 10.52288 | | 0.013291 | [0.0094-0. | Affymetrix [18485882] (imputed) | N |
| 9 | 17:44118848 | 17 | 43975415 | rs62056789 | 29/01/2020 | 3.2E+07 | Hill WD | 16/12/2019 | Nat Commun | www.ncbi.nlm.nih.gov/pubmed/31844048 | Genome-wide analysis identifies molecular systems and 149 genetic loci associated with income. | Household income (MTAG) | 505,541 Bi | NA | 17q21.31 | MAPT | MAPT-IT1, MAPT-IT1, MAPT-IT1, MAPT, MAPT, MAPT | | | ENSG00000281284, E | NA | NA | rs62056789-C | rs62056789 | 0 | 62056789 | non_coding_transcript_exon | 0 | 0.3822 | 3.00E-12 | 11.52288 | | 0.017144 | [0.012-0.0 | Affymetrix [up to 18485882] (imputed) | N |
| 9 | 17:44118848 | 17 | 43975415 | rs62056789 | 27/05/2022 | 3.5E+07 | Wendt FR | 18/02/2022 | Mol Psychiatry | www.ncbi.nlm.nih.gov/pubmed/35181757 | Using phenotype risk scores to enhance gene discovery for generalized anxiety disorder and posttraumatic stress disorder. | Post-traumatic stress disorder (mental health questionnaire or predicted) | 497,803 Eu | NA | 17q21.31 | | MAPT-IT1, MAPT-IT1, MAPT-IT1, MAPT, MAPT, MAPT | | | ENSG00000281284, E | NA | NA | rs62056789-A | rs62056789 | 0 | 62056789 | non_coding_transcript_exon_ | 0 | NR | 1.00E-19 | 19 | | 0.021979 | [0.017-0.0 | Affymetrix [NR] (imputed) | N |
| 9 | 17:44118848 | 17 | 43975415 | rs62056789 | 27/05/2022 | 3.5E+07 | Wendt FR | 18/02/2022 | Mol Psychiatry | www.ncbi.nlm.nih.gov/pubmed/35181757 | Using phenotype risk scores to enhance gene discovery for generalized anxiety disorder and posttraumatic stress disorder. | Post-traumatic stress disorder (phenotype risk score) | 497,803 Eu | NA | 17q21.31 | | MAPT-IT1, MAPT-IT1, MAPT-IT1, MAPT, MAPT, MAPT | | | ENSG00000281284, E | NA | NA | rs62056789-A | rs62056789 | 0 | 62056789 | non_coding_transcript_exon_ | 0 | NR | 1.00E-16 | 16 | | 0.020087 | [0.015-0.0 | Affymetrix [NR] (imputed) | N |
| 9 | 17:44118848 | 17 | 43790005 | rs62056934 | 7/03/2022 | 3.5E+07 | van der Meer D | 15/12/2021 | Sci Adv | www.ncbi.nlm.nih.gov/pubmed/34910505 | The genetic architecture of human cortical folding. | Vertex-wise cortical surface area | 8,072 indiv | NA | 17q21.31 | | LINC02210-CRHR1, LINC02210-CRHR1 | | | ENSG00000282456, E | NA | NA | rs62056934-G | rs62056934 | 0 | 62056934 | intron_variant | 0 | 0.24 | 9.00E-12 | 11.04576 | | NA | | NR [13131314] (imputed) | N |
| 9 | 17:44118848 | 17 | 43790005 | rs62056934 | 7/03/2022 | 3.5E+07 | van der Meer D | 15/12/2021 | Sci Adv | www.ncbi.nlm.nih.gov/pubmed/34910505 | The genetic architecture of human cortical folding. | Vertex-wise cortical surface area | 33,748 Eur | NA | 17q21.31 | | LINC02210-CRHR1, LINC02210-CRHR1 | | | ENSG00000282456, E | NA | NA | rs62056934-G | rs62056934 | 0 | 62056934 | intron_variant | 0 | 0.24 | ####### | 139.5229 | | 25.21 | z score inc | Affymetrix [9061022] (imputed) | N |
| 9 | 17:44118848 | 17 | 43790005 | rs62056934 | 7/03/2022 | 3.5E+07 | van der Meer D | 15/12/2021 | Sci Adv | www.ncbi.nlm.nih.gov/pubmed/34910505 | The genetic architecture of human cortical folding. | Vertex-wise cortical thickness | 33,748 Eur | NA | 17q21.31 | | LINC02210-CRHR1, LINC02210-CRHR1 | | | ENSG00000282456, E | NA | NA | rs62056934-G | rs62056934 | 0 | 62056934 | intron_variant | 0 | 0.24 | ####### | 202.5229 | | 30.42 | z score inc | Affymetrix [9061022] (imputed) | N |
| 9 | 17:44118848 | 17 | 43856639 | rs62057061 | 31/10/2018 | 3E+07 | Nagel M | 25/06/2018 | Nat Genet | www.ncbi.nlm.nih.gov/pubmed/29942085 | Meta-analysis of genome-wide association studies for neuroticism in 449,484 individuals identifies novel genetic loci and pathways. | Depressed affect | 357,957 Eu | NA | 17q21.31 | NR | LINC02210-CRHR1, LINC02210-CRHR1 | | | ENSG00000282456, E | NA | NA | rs62057061-G | rs62057061 | 0 | 62057061 | intron_variant | 0 | NR | 2.00E-22 | 21.69897 | | 0.02689 | [0.021-0.0 | Affymetrix [10847151] (imputed) | N |
| 9 | 17:44118848 | 17 | 43859640 | rs62057070 | 21/07/2020 | 3.2E+07 | Grasby KL | 1/03/2020 | Science | www.ncbi.nlm.nih.gov/pubmed/32193296 | The genetic architecture of the human cerebral cortex. | Cortical surface area | 33,992 Eur | 14,727 Eur | 17q21.31 | NR | LINC02210-CRHR1, LINC02210-CRHR1 | | | ENSG00000282456, E | NA | NA | rs62057070-A | rs62057070 | 0 | 62057070 | intron_variant | 0 | 0.7896 | 5.00E-13 | 12.30103 | (Fusiform) | 17.64018 | [12.86-22. | Affymetrix, Illumina [NR] | N |
| 9 | 17:44118848 | 17 | 43896032 | rs62057107 | 12/11/2018 | 3E+07 | Lee JJ | 23/07/2018 | Nat Genet | www.ncbi.nlm.nih.gov/pubmed/30038396 | Gene discovery and polygenic prediction from a genome-wide association study of educational attainment in 1.1 million individuals. | Educational attainment (MTAG) | 1,311,438 | NA | 17q21.31 | Intergenic | LINC02210-CRHR1, LINC02210-CRHR1, CRHR1, CRHR1, C | | | ENSG00000282456, E | NA | NA | rs62057107-T | rs62057107 | 0 | 62057107 | intron_variant | 0 | 0.2192 | 5.00E-38 | 37.30103 | (MTAG) | 0.0209 | [0.018-0.0 | Affymetrix, Illumina [7100000] (imputed) | N |
| 9 | 17:44118848 | 17 | 43896032 | rs62057107 | 12/11/2018 | 3E+07 | Lee JJ | 23/07/2018 | Nat Genet | www.ncbi.nlm.nih.gov/pubmed/30038396 | Gene discovery and polygenic prediction from a genome-wide association study of educational attainment in 1.1 million individuals. | Highest math class taken (MTAG) | 811,539 Eu | NA | 17q21.31 | Intergenic | LINC02210-CRHR1, LINC02210-CRHR1, CRHR1, CRHR1, C | | | ENSG00000282456, E | NA | NA | rs62057107-T | rs62057107 | 0 | 62057107 | intron_variant | 0 | 0.2219 | 4.00E-28 | 27.39794 | (MTAG) | 0.022 | [0.018-0.0 | Affymetrix, Illumina [7100000] (imputed) | N |
| 9 | 17:44118848 | 17 | 43903842 | rs62057151 | 22/01/2019 | 3E+07 | Nagel M | 2/03/2018 | Nat Commun | www.ncbi.nlm.nih.gov/pubmed/29500382 | Item-level analyses reveal genetic heterogeneity in neuroticism. | Feeling worry | 372,869 Eu | NA | 17q21.31 | NR | LINC02210-CRHR1, LINC02210-CRHR1, CRHR1, CRHR1, C | | | ENSG00000282456, E | NA | NA | rs62057151-T | rs62057151 | 0 | 62057151 | non_coding_transcript_exon | 0 | 0.223812 | 1.00E-09 | 9 | | 6.049 | unit increa | NR [10847151] (imputed) | N |
| 9 | 17:44118848 | 17 | 44243870 | rs62060891 | 15/12/2021 | 3.5E+07 | Shadrin AA | 21/09/2021 | Neuroimage | www.ncbi.nlm.nih.gov/pubmed/34560273 | Vertex-wise multivariate genome-wide association study identifies 780 unique genetic loci associated with cortical morphology. | Cortical thickness | 35,657 Wh | NA | 17q21.31 | NR | KANSL1, KANSL1, KANSL1 | | | ENSG00000120071, E | NA | NA | rs62060891-C | rs62060891 | 0 | 62060891 | intron_variant | 0 | 0.2215 | 1.00E-17 | 17 | | NA | | NR [9056863] (imputed) | N |
| 9 | 17:44118848 | 17 | 44273653 | rs62060954 | 27/04/2022 | 3.5E+07 | Richardson TG | 25/02/2022 | PLoS Biol | www.ncbi.nlm.nih.gov/pubmed/35213538 | Characterising metabolomic signatures of lipid-modifying therapies through drug target mendelian randomisation. | Degree of unsaturation | 115,006 Eu | NA | 17q21.31 | | KANSL1, KANSL1, KANSL1, KANSL1-AS1, KANSL1-AS1, KANS | | | ENSG00000120071, E | NA | NA | rs62060954-T | rs62060954 | 0 | 62060954 | non_coding_transcript_exon | 0 | 0.839881 | 5.00E-10 | 9.30103 | | 0.036063 | [0.025-0.0 | NR [11722792] (imputed) | N |
| 9 | 17:44118848 | 17 | 44018399 | rs62061733 | 8/02/2019 | 3.1E+07 | Kichaev G | 27/12/2018 | Am J Hum Genet | www.ncbi.nlm.nih.gov/pubmed/30595370 | Leveraging Polygenic Functional Enrichment to Improve GWAS Power. | Eosinophil counts | approxima | NA | 17q21.31 | | MAPT, MAPT, MAPT | | | ENSG00000277956, E | NA | NA | rs62061733-? | rs62061733 | 0 | 62061733 | intron_variant | 0 | NR | 3.00E-29 | 28.52288 | | NA | | NR [~ 8900000] (imputed) | N |
| 9 | 17:44118848 | 17 | 44018399 | rs62061733 | 22/01/2019 | 3E+07 | Nagel M | 2/03/2018 | Nat Commun | www.ncbi.nlm.nih.gov/pubmed/29500382 | Item-level analyses reveal genetic heterogeneity in neuroticism. | Feeling hurt | 372,047 Eu | NA | 17q21.31 | NR | MAPT, MAPT, MAPT | | | ENSG00000277956, E | NA | NA | rs62061733-A | rs62061733 | 0 | 62061733 | intron_variant | 0 | 0.22111 | 2.00E-28 | 27.69897 | | 11.07 | z score dec | NR [10847151] (imputed) | N |

| | | | | | | | | | | | | | | | | | | | | | | | | | | | | | | | | | | | | |
|---|---|---|---|---|---|---|---|---|---|---|---|---|---|---|---|---|---|---|---|---|---|---|---|---|---|---|---|---|---|---|---|---|---|---|---|---|
| 9 | 17:44118848 | 17 | 44018488 | rs62061734 | 30/11/2017 | 2.9E+07 | Michailidou K | 23/10/2017 | Nature | www.ncbi.nlm.nih.gov/pubmed/29059683 | Association analysis identifies 65 new breast cancer risk loci. | Breast cancer | 76,192 Eur | 46,785 Eur | 17q21.31 | NR | MAPT, MAPT, MAPT | | | ENSG00000277956, E | NA | NA | rs62061734-C | rs62061734 | 0 | 62061734 | intron_variant | 0 | 0.2103 | 8.00E-12 | 11.09691 | (EA) | 0.0526 | [0.038-0.0 | Illumina [~ 11800000] (imputed) | N |
| 9 | 17:44118848 | 17 | 44018488 | rs62061734 | 25/06/2019 | 3.1E+07 | Jansen PR | 25/02/2019 | Nat Genet | www.ncbi.nlm.nih.gov/pubmed/30804565 | Genome-wide analysis of insomnia in 1,331,010 individuals identifies new risk loci and functional pathways. | Sleep duration | 384,317 E | NA | 17q21.31 | | MAPT, MAPT, MAPT | | | ENSG00000277956, E | NA | NA | rs62061734-C | rs62061734 | 0 | 62061734 | intron_variant | 0 | 0.2205 | 2.00E-09 | 8.69897 | | 0.01775 | [0.012-0.0 | Affymetrix, Illumina [10862567] (imputed) | N |
| 9 | 17:44118848 | 17 | 44096553 | rs62062288 | 14/12/2018 | 3E+07 | Sanchez-Roige S | 19/10/2018 | Am J Psychiatry | www.ncbi.nlm.nih.gov/pubmed/30336701 | Genome-Wide Association Study Meta-Analysis of the Alcohol Use Disorders Identification Test (AUDIT) in Two Population-Based Cohorts. | Alcohol use disorder (consumption score) | 121,604 B | NA | 17q21.31 | MAPT | MAPT, MAPT, MAPT | | | ENSG00000277956, E | NA | NA | rs62062288-A | rs62062288 | 0 | 62062288 | intron_variant | 0 | NR | 2.00E-09 | 8.69897 | | 0.008 | [0.006-0.0 | NR [16213998] | N |
| 9 | 17:44118848 | 17 | 44096553 | rs62062288 | 7/09/2021 | 3.4E+07 | Dao C | 16/04/2021 | Addiction | www.ncbi.nlm.nih.gov/pubmed/33861876 | The Impact of Removing Former Drinkers from Genome-wide Association Studies of AUDIT-C. | Alcohol use disorder (consumption score) | 5,135 Euro | NR | 17q21.31 | MAPT | MAPT, MAPT, MAPT | | | ENSG00000277956, E | NA | NA | rs62062288-? | rs62062288 | 0 | 62062288 | intron_variant | 0 | NR | 2.00E-14 | 13.69897 | | 0.097 | [0.072-0.1 | NR [7682250] (imputed) | N |
| 9 | 17:44118848 | 17 | 44096553 | rs62062288 | 14/12/2018 | 3E+07 | Sanchez-Roige S | 19/10/2018 | Am J Psychiatry | www.ncbi.nlm.nih.gov/pubmed/30336701 | Genome-Wide Association Study Meta-Analysis of the Alcohol Use Disorders Identification Test (AUDIT) in Two Population-Based Cohorts. | Alcohol use disorder (total score) | 141,932 E | NA | 17q21.31 | CRHR1, DND1P1, | MAPT, MAPT, MAPT | | | ENSG00000277956, E | NA | NA | rs62062288-A | rs62062288 | 0 | 62062288 | intron_variant | 0 | NR | 5.00E-10 | 9.30103 | | 6.21 | [NR] z scor | NR [16213998] | N |
| 9 | 17:44118848 | 17 | 44096553 | rs62062288 | 18/03/2019 | 3.1E+07 | Karlsson Linner R | 14/01/2019 | Nat Genet | www.ncbi.nlm.nih.gov/pubmed/30643258 | Genome-wide association analyses of risk tolerance and risky behaviors in over 1 million individuals identify hundreds of loci and shared genetic influences. | Automobile speeding propensity | 404,291 E | NA | 17q21.31 | KANSL1 | MAPT, MAPT, MAPT | | | ENSG00000277956, E | NA | NA | rs62062288-A | rs62062288 | 0 | 62062288 | intron_variant | 0 | 0.2196 | 1.00E-12 | 12 | | 0.019352 | [0.014-0.0 | Affymetrix [11514463] (imputed) | N |
| 9 | 17:44118848 | 17 | 44096553 | rs62062288 | 22/01/2019 | 3E+07 | Nagel M | 2/03/2018 | Nat Commun | www.ncbi.nlm.nih.gov/pubmed/29500382 | Item-level analyses reveal genetic heterogeneity in neuroticism. | Feeling guilty | 373,380 E | NA | 17q21.31 | NR | MAPT, MAPT, MAPT | | | ENSG00000277956, E | NA | NA | rs62062288-A | rs62062288 | 0 | 62062288 | intron_variant | 0 | 0.211505 | 7.00E-10 | 9.154902 | | 6.17 | z score inc | NR [10847151] (imputed) | N |
| 9 | 17:44118848 | 17 | 44096553 | rs62062288 | 22/01/2019 | 3E+07 | Nagel M | 2/03/2018 | Nat Commun | www.ncbi.nlm.nih.gov/pubmed/29500382 | Item-level analyses reveal genetic heterogeneity in neuroticism. | Feeling worry | 372,869 E | NA | 17q21.31 | NR | MAPT, MAPT, MAPT | | | ENSG00000277956, E | NA | NA | rs62062288-A | rs62062288 | 0 | 62062288 | intron_variant | 0 | 0.211505 | 1.00E-09 | 9 | | 6.079 | unit increa | NR [10847151] (imputed) | N |
| 9 | 17:44118848 | 17 | 44096553 | rs62062288 | 22/01/2019 | 3E+07 | Nagel M | 2/03/2018 | Nat Commun | www.ncbi.nlm.nih.gov/pubmed/29500382 | Item-level analyses reveal genetic heterogeneity in neuroticism. | Neurociticism | 380,506 E | NA | 17q21.31 | STH | MAPT, MAPT, MAPT | | | ENSG00000277956, E | NA | NA | rs62062288-A | rs62062288 | 0 | 62062288 | intron_variant | 0 | 0.211505 | 6.00E-32 | 31.22185 | | 11.77 | z score inc | NR [10847151] (imputed) | N |
| 9 | 17:44118848 | 17 | 44096553 | rs62062288 | 29/05/2020 | 3.2E+07 | Cai N | 30/03/2020 | Nat Genet | www.ncbi.nlm.nih.gov/pubmed/32231276 | Minimal phenotyping yields genome-wide association signals of low specificity for major depression. | Neuroticism | 274,107 B | NA | 17q21.31 | NR | MAPT, MAPT, MAPT | | | ENSG00000277956, E | NA | NA | rs62062288-A | rs62062288 | 0 | 62062288 | intron_variant | 0 | 0.2181 | 7.00E-22 | 21.1549 | | 0.01863 | [0.015-0.0 | NR [8968716] (imputed) | N |
| 9 | 17:44118848 | 17 | 44096553 | rs62062288 | 18/03/2019 | 3.1E+07 | Karlsson Linner R | 14/01/2019 | Nat Genet | www.ncbi.nlm.nih.gov/pubmed/30643258 | Genome-wide association analyses of risk tolerance and risky behaviors in over 1 million individuals identify hundreds of loci and shared genetic influences. | Risk-taking tendency (4-domain principal component model) | 315,894 E | NA | 17q21.31 | KANSL1 | MAPT, MAPT, MAPT | | | ENSG00000277956, E | NA | NA | rs62062288-A | rs62062288 | 0 | 62062288 | intron_variant | 0 | 0.2207 | 1.00E-29 | 29 | | 0.035171 | [0.029-0.0 | Affymetrix [~ 11515000] (imputed) | N |
| 9 | 17:44118848 | 17 | 44096553 | rs62062288 | 22/01/2019 | 3E+07 | Nagel M | 2/03/2018 | Nat Commun | www.ncbi.nlm.nih.gov/pubmed/29500382 | Item-level analyses reveal genetic heterogeneity in neuroticism. | Worry too long after an embarrassing experience | 367,725 E | NA | 17q21.31 | NR | MAPT, MAPT, MAPT | | | ENSG00000277956, E | NA | NA | rs62062288-A | rs62062288 | 0 | 62062288 | intron_variant | 0 | 0.211505 | 4.00E-08 | 7.39794 | | 5.5 | z score inc | NR [10847151] (imputed) | N |
| 9 | 17:44118848 | 17 | 44027366 | rs62062797 | 5/02/2020 | 3.2E+07 | Zhao B | 30/10/2019 | Mol Psychiatry | www.ncbi.nlm.nih.gov/pubmed/31666681 | Large-scale GWAS reveals genetic architecture of brain white matter microstructure and genetic overlap with cognitive and mental health traits (nÔÇë=ÔÇë17,706). | White matter microstructure (fractional anisotropy) | 17,706 Eur | NA | 17q21.31 | NR | MAPT, MAPT, MAPT | | | ENSG00000277956, E | NA | NA | rs62062797-? | rs62062797 | 0 | 62062797 | intron_variant | 0 | | 5.00E-11 | 10.30103 | (Anterior li | NA | | NR [8955960] (imputed) | N |
| 9 | 17:44118848 | 17 | 44038785 | rs62063281 | 4/08/2022 | 3.6E+07 | May-Wilson S | 18/05/2022 | Nat Commun | www.ncbi.nlm.nih.gov/pubmed/35585065 | Large-scale GWAS of food liking reveals genetic determinants and genetic correlations with distinct neurophysiological traits. | F-strong vegetable liking (derived food-liking factor) | 147,344 E | NA | 17q21.31 | | MAPT, MAPT, MAPT | | | ENSG00000277956, E | NA | NA | rs62063281-G | rs62063281 | 0 | 62063281 | intron_variant | 0 | 0.2192 | 8.00E-13 | 12.09691 | | 0.08345 | [0.061-0.1 | Affymetrix [9601173] (imputed) | N |
| 9 | 17:44118848 | 17 | 44038785 | rs62063281 | 18/03/2019 | 3.1E+07 | Karlsson Linner R | 14/01/2019 | Nat Genet | www.ncbi.nlm.nih.gov/pubmed/30643258 | Genome-wide association analyses of risk tolerance and risky behaviors in over 1 million individuals identify hundreds of loci and shared genetic influences. | Number of sexual partners | 370,711 E | NA | 17q21.31 | STH | MAPT, MAPT, MAPT | | | ENSG00000277956, E | NA | NA | rs62063281-A | rs62063281 | 0 | 62063281 | intron_variant | 0 | 0.7784 | 4.00E-15 | 14.39794 | | 0.02236 | [0.017-0.0 | Affymetrix [11515109] (imputed) | N |
| 9 | 17:44118848 | 17 | 44038785 | rs62063281 | 8/02/2019 | 3.1E+07 | Tachmazidou I | 21/01/2019 | Nat Genet | www.ncbi.nlm.nih.gov/pubmed/30664745 | Identification of new therapeutic targets for osteoarthritis through genome-wide analyses of UK Biobank data. | Osteoarthritis (hip) | 15,704 Eur | NA | 17q21.31 | NR | MAPT, MAPT, MAPT | | | ENSG00000277956, E | NA | NA | rs62063281-G | rs62063281 | 0 | 62063281 | intron_variant | 0 | 0.22 | 5.00E-12 | 11.30103 | | 1.1 | [1.07-1.13 | Affymetrix, Illumina [up to 17000000] (imputed) | N |
| 9 | 17:44118848 | 17 | 44038785 | rs62063281 | 26/11/2020 | 3.3E+07 | Bi W | 23/06/2020 | Am J Hum Genet | www.ncbi.nlm.nih.gov/pubmed/32589924 | A Fast and Accurate Method for Genome-Wide Time-to-Event Data Analysis and Its Application to UK Biobank. | Osteoarthrosis (time to event) | 29,071 Brit | NA | 17q21.31 | MAPT | MAPT, MAPT, MAPT | | | ENSG00000277956, E | NA | NA | rs62063281-? | rs62063281 | 0 | 62063281 | intron_variant | 0 | NR | 3.00E-08 | 7.522879 | | NA | | NR [~ 24000000] (imputed) | N |
| 9 | 17:44118848 | 17 | 44038785 | rs62063281 | 17/11/2022 | 3.6E+07 | Baselmans B | 6/10/2021 | Biol Psychiatry Glob Open Sci | www.ncbi.nlm.nih.gov/pubmed/36324656 | The Genetic and Neural Substrates of Externalizing Behavior. | Risk-taking behavior (multivariate analysis) | 1,506,537 | NA | 17q21.31 | | MAPT, MAPT, MAPT | | | ENSG00000277956, E | NA | NA | rs62063281-? | rs62063281 | 0 | 62063281 | intron_variant | 0 | NR | 3.00E-26 | 25.52288 | | NA | | NR [NR] | N |
| 9 | 17:44118848 | 17 | 44061023 | rs62063786 | 17/06/2020 | 3.2E+07 | Oskarsson GR | 23/04/2020 | Commun Biol | www.ncbi.nlm.nih.gov/pubmed/32327693 | Predicted loss and gain of function mutations in ACO1 are associated with erythropoiesis. | Hemoglobin levels | 684,122 E | NA | 17q21.31 | MAPT | MAPT, MAPT, MAPT | | | ENSG00000277956, E | NA | NA | rs62063786-A | rs62063786 | 0 | 62063786 | missense_variant | 0 | | 1.00E-61 | 61 | | 0.044 | [NR] unit in | Affymetrix, Illumina [43000000] (imputed) | N |
| 9 | 17:44118848 | 17 | 43654468 | rs62064364 | 31/05/2022 | 3.4E+07 | Smith SM | 19/04/2021 | Nat Neurosci | www.ncbi.nlm.nih.gov/pubmed/33875891 | An expanded set of genome-wide association studies of brain imaging phenotypes in UK Biobank. | IDP dMRI TBSS L3 Anterior limb of internal capsule L | 20,860 Brit | 10,496 Brit | 17q21.31 | | RDM1P1 - DND1F | ENSG00000266504 | ENSG00000282614 | | 18032 | 8769 | rs62064364-T | rs62064364 | 0 | 62064364 | intron_variant | 1 | 0.22 | 2.00E-28 | 27.69897 | | 0.132 | [0.11-0.16 | Affymetrix [17103079] (imputed) | N |
| 9 | 17:44118848 | 17 | 43654468 | rs62064364 | 28/01/2019 | 3.1E+07 | Gao XR | 7/12/2018 | Hum Mol Genet | www.ncbi.nlm.nih.gov/pubmed/30535121 | Genome-wide association analyses identify 139 loci associated with macular thickness in the UK Biobank cohort. | Macular thickness | 59,814 Eur | 8,609 Euro | 17q21.31 | LRRC37A4P, MAF | RDM1P1 - DND1F | ENSG00000266504 | ENSG00000282614 | | 18032 | 8769 | rs62064364-? | rs62064364 | 0 | 62064364 | intron_variant | 1 | NR | 4.00E-35 | 34.39794 | | 0.978 | [0.81-1.14 | Affymetrix [11100000] (imputed) | N |
| 9 | 17:44118848 | 17 | 43654468 | rs62064364 | 5/02/2020 | 3.2E+07 | Zhao B | 30/10/2019 | Mol Psychiatry | www.ncbi.nlm.nih.gov/pubmed/31666681 | Large-scale GWAS reveals genetic architecture of brain white matter microstructure and genetic overlap with cognitive and mental health traits (nÔÇë=ÔÇë17,706). | White matter microstructure (axial diusivities) | 17,706 Eur | NA | 17q21.31 | NR | RDM1P1 - DND1F | ENSG00000266504 | ENSG00000282614 | | 18032 | 8769 | rs62064364-? | rs62064364 | 0 | 62064364 | intron_variant | 1 | | 5.00E-10 | 9.30103 | (External c | NA | | NR [8955960] (imputed) | N |
| 9 | 17:44118848 | 17 | 43666001 | rs62064395 | 31/05/2022 | 3.4E+07 | Smith SM | 19/04/2021 | Nat Neurosci | www.ncbi.nlm.nih.gov/pubmed/33875891 | An expanded set of genome-wide association studies of brain imaging phenotypes in UK Biobank. | IDP dMRI ProbtrackX MD str l | 20,860 Brit | 10,495 Brit | 17q21.31 | | DND1P1 - MAPK8 | ENSG00000282614 | ENSG00000263503 | | 1706 | 12234 | rs62064395-C | rs62064395 | 0 | 62064395 | intergenic_variant | 1 | 0.233 | 4.00E-14 | 13.39794 | | 0.088 | [0.064-0.1 | Affymetrix [17103079] (imputed) | N |
| 9 | 17:44118848 | 17 | 43666001 | rs62064395 | 31/05/2022 | 3.4E+07 | Smith SM | 19/04/2021 | Nat Neurosci | www.ncbi.nlm.nih.gov/pubmed/33875891 | An expanded set of genome-wide association studies of brain imaging phenotypes in UK Biobank. | IDP dMRI TBSS L2 Genu of corpus callosum | 20,860 Brit | 10,496 Brit | 17q21.31 | | DND1P1 - MAPK8 | ENSG00000282614 | ENSG00000263503 | | 1706 | 12234 | rs62064395-C | rs62064395 | 0 | 62064395 | intergenic_variant | 1 | 0.233 | 3.00E-15 | 14.52288 | | 0.092 | [0.068-0.1 | Affymetrix [17103079] (imputed) | N |
| 9 | 17:44118848 | 17 | 43666001 | rs62064395 | 31/05/2022 | 3.4E+07 | Smith SM | 19/04/2021 | Nat Neurosci | www.ncbi.nlm.nih.gov/pubmed/33875891 | An expanded set of genome-wide association studies of brain imaging phenotypes in UK Biobank. | IDP dMRI TBSS L3 Genu of corpus callosum | 20,860 Brit | 10,496 Brit | 17q21.31 | | DND1P1 - MAPK8 | ENSG00000282614 | ENSG00000263503 | | 1706 | 12234 | rs62064395-C | rs62064395 | 0 | 62064395 | intergenic_variant | 1 | 0.233 | 5.00E-15 | 14.30103 | | 0.091 | [0.067-0.1 | Affymetrix [17103079] (imputed) | N |
| 9 | 17:44118848 | 17 | 43666001 | rs62064395 | 31/05/2022 | 3.4E+07 | Smith SM | 19/04/2021 | Nat Neurosci | www.ncbi.nlm.nih.gov/pubmed/33875891 | An expanded set of genome-wide association studies of brain imaging phenotypes in UK Biobank. | IDP dMRI TBSS MD Anterior corona radiata L | 20,860 Brit | 10,496 Brit | 17q21.31 | | DND1P1 - MAPK8 | ENSG00000282614 | ENSG00000263503 | | 1706 | 12234 | rs62064395-C | rs62064395 | 0 | 62064395 | intergenic_variant | 1 | 0.233 | 1.00E-16 | 16 | | 0.096 | [0.072-0.1 | Affymetrix [17103079] (imputed) | N |
| 9 | 17:44118848 | 17 | 43473307 | rs62064597 | 24/08/2022 | 3.6E+07 | Plotnikov D | 1/06/2022 | Invest Ophthalmol Vis Sci | www.ncbi.nlm.nih.gov/pubmed/35762941 | High Blood Pressure and Intraocular Pressure: A Mendelian Randomization Study. | Systolic blood pressure | 526,001 E | NA | 17q21.31 | | ARHGAP27 | | | ENSG00000159314 | NA | NA | rs62064597-C | rs62064597 | 0 | 62064597 | intron_variant | 0 | NR | 6.00E-10 | 9.221849 | | 0.2971 | [0.2-0.39] | NR [NR] (imputed) | N |
| 9 | 17:43556807 | 17 | 43474668 | rs62064598 | 2/05/2019 | 3.1E+07 | Hill WD | 13/03/2019 | Mol Psychiatry | www.ncbi.nlm.nih.gov/pubmed/30867560 | Genetic contributions to two special factors of neuroticism are associated with affluence, higher intelligence, better health, and longer life. | General factor of neuroticism | 270,059 B | NA | 17q21.31 | ENSG000002673 | ARHGAP27 | | | ENSG00000159314 | NA | NA | rs62064598-C | rs62064598 | 0 | 62064598 | intron_variant | 0 | NR | 3.00E-15 | 14.52288 | | 0.013541 | [0.01-0.01 | Affymetrix [18485882] (imputed) | N |
| 9 | 17:43556807 | 17 | 43488792 | rs62064641 | 8/02/2019 | 3.1E+07 | Kichaev G | 27/12/2018 | Am J Hum Genet | www.ncbi.nlm.nih.gov/pubmed/30595370 | Leveraging Polygenic Functional Enrichment to Improve GWAS Power. | White blood cell count | approxima | NA | 17q21.31 | | ARHGAP27 | | | ENSG00000159314 | NA | NA | rs62064641-? | rs62064641 | 0 | 62064641 | intron_variant | 0 | NR | 1.00E-13 | 13 | | NA | | NR [~ 8900000] (imputed) | N |
| 9 | 17:43556807 | 17 | 43488792 | rs62064641 | 31/10/2018 | 3E+07 | Nagel M | 25/06/2018 | Nat Genet | www.ncbi.nlm.nih.gov/pubmed/29942085 | Meta-analysis of genome-wide association studies for neuroticism in 449,484 individuals identifies novel genetic loci and pathways. | Worry | 348,219 E | NA | 17q21.31 | NR | ARHGAP27 | | | ENSG00000159314 | NA | NA | rs62064641-C | rs62064641 | 0 | 62064641 | intron_variant | 0 | NR | 2.00E-11 | 10.69897 | | 0.02055 | [0.015-0.0 | Affymetrix [10847151] (imputed) | N |
| 9 | 17:44118848 | 17 | 44082528 | rs62064667 | 31/05/2022 | 3.4E+07 | Smith SM | 19/04/2021 | Nat Neurosci | www.ncbi.nlm.nih.gov/pubmed/33875891 | An expanded set of genome-wide association studies of brain imaging phenotypes in UK Biobank. | IDP dMRI TBSS L1 Anterior corona radiata L | 20,860 Brit | 10,496 Brit | 17q21.31 | | MAPT, MAPT, MAPT | | | ENSG00000277956, E | NA | NA | rs62064667-T | rs62064667 | 0 | 62064667 | intron_variant | 0 | 0.224 | 2.00E-13 | 12.69897 | | 0.087 | [0.063-0.1 | Affymetrix [17103079] (imputed) | N |
| 9 | 17:44118848 | 17 | 43573061 | rs62065452 | 31/05/2022 | 3.4E+07 | Smith SM | 19/04/2021 | Nat Neurosci | www.ncbi.nlm.nih.gov/pubmed/33875891 | An expanded set of genome-wide association studies of brain imaging phenotypes in UK Biobank. | aparc-Desikan rh area lateraloccipital | 21,282 Brit | 10,686 Brit | 17q21.31 | | PLEKHM1 - LRRC | ENSG00000277111 | ENSG00000282331 | | 4946 | 11046 | rs62065452-C | rs62065452 | 0 | 62065452 | intergenic_variant | 1 | 0.223 | 3.00E-14 | 13.52288 | | 0.088 | [0.064-0.1 | Affymetrix [17103079] (imputed) | N |
| 9 | 17:44118848 | 17 | 43573061 | rs62065452 | 31/05/2022 | 3.4E+07 | Smith SM | 19/04/2021 | Nat Neurosci | www.ncbi.nlm.nih.gov/pubmed/33875891 | An expanded set of genome-wide association studies of brain imaging phenotypes in UK Biobank. | aparc-Desikan rh volume fusiform | 21,282 Brit | 10,686 Brit | 17q21.31 | | PLEKHM1 - LRRC | ENSG00000277111 | ENSG00000282331 | | 4946 | 11046 | rs62065452-C | rs62065452 | 0 | 62065452 | intergenic_variant | 1 | 0.223 | 6.00E-10 | 9.221849 | | 0.072 | [0.048-0.0 | Affymetrix [17103079] (imputed) | N |
| 9 | 17:44118848 | 17 | 43573061 | rs62065452 | 31/05/2022 | 3.4E+07 | Smith SM | 19/04/2021 | Nat Neurosci | www.ncbi.nlm.nih.gov/pubmed/33875891 | An expanded set of genome-wide association studies of brain imaging phenotypes in UK Biobank. | aparc-DKTatlas rh area lateraloccipital | 21,282 Brit | 10,686 Brit | 17q21.31 | | PLEKHM1 - LRRC | ENSG00000277111 | ENSG00000282331 | | 4946 | 11046 | rs62065452-C | rs62065452 | 0 | 62065452 | intergenic_variant | 1 | 0.223 | 2.00E-14 | 13.69897 | | 0.089 | [0.065-0.1 | Affymetrix [17103079] (imputed) | N |
| 9 | 17:44118848 | 17 | 43573419 | rs62065453 | 22/01/2019 | 3E+07 | Nagel M | 2/03/2018 | Nat Commun | www.ncbi.nlm.nih.gov/pubmed/29500382 | Item-level analyses reveal genetic heterogeneity in neuroticism. | Feeling nervous | 373,121 E | NA | 17q21.31 | NR | PLEKHM1 - LRRC | ENSG00000277111 | ENSG00000282331 | | 5304 | 10688 | rs62065453-T | rs62065453 | 0 | 62065453 | intron_variant | 1 | 0.217174 | 6.00E-15 | 14.22185 | | 7.81 | z score inc | NR [10847151] (imputed) | N |
| 9 | 17:44118848 | 17 | 43573419 | rs62065453 | 22/01/2019 | 3E+07 | Nagel M | 2/03/2018 | Nat Commun | www.ncbi.nlm.nih.gov/pubmed/29500382 | Item-level analyses reveal genetic heterogeneity in neuroticism. | Irritable mood | 366,726 E | NA | 17q21.31 | NR | PLEKHM1 - LRRC | ENSG00000277111 | ENSG00000282331 | | 5304 | 10688 | rs62065453-T | rs62065453 | 0 | 62065453 | intron_variant | 1 | 0.217174 | 3.00E-12 | 11.52288 | | 6.99 | z score inc | NR [~ 10847151] (imputed) | N |
| 9 | 17:44118848 | 17 | 43573419 | rs62065453 | 9/06/2021 | 3.4E+07 | Pazoki R | 10/05/2021 | Nat Commun | www.ncbi.nlm.nih.gov/pubmed/33972514 | Genetic analysis in European ancestry individuals identifies 517 loci associated with liver enzymes. | Liver enzyme levels (alkaline phosphatase) | 437,438 E | 315,572 E | 17q21.31 | PLEKHM1 | PLEKHM1 - LRRC | ENSG00000277111 | ENSG00000282331 | | 5304 | 10688 | rs62065453-C | rs62065453 | 0 | 62065453 | intron_variant | 1 | 0.774564 | 2.00E-17 | 16.69897 | | 0.002322 | [0.0018-0. | Affymetrix [13995440] (imputed) | N |
| 9 | 17:44118848 | 17 | 43573419 | rs62065453 | 1/02/2018 | 2.9E+07 | Luciano M | 18/12/2017 | Nat Genet | www.ncbi.nlm.nih.gov/pubmed/29255261 | Association analysis in over 329,000 individuals identifies 116 independent variants influencing neuroticism. | Neuroticism | 329,821 E | 122,867 E | 17q21.31 | intergenic | PLEKHM1 - LRRC | ENSG00000277111 | ENSG00000282331 | | 5304 | 10688 | rs62065453-T | rs62065453 | 0 | 62065453 | intron_variant | 1 | 0.21717 | 2.00E-24 | 23.69897 | | 10.227 | unit increa | Illumina [18485882] (imputed) | N |
| 9 | 17:44118848 | 17 | 43688042 | rs62066084 | 4/08/2022 | 3.6E+07 | Fan CC | 3/05/2022 | Nat Commun | www.ncbi.nlm.nih.gov/pubmed/35505052 | Multivariate genome-wide association study on tissue-sensitive diffusion metrics highlights pathways that shape the human brain. | Whole brain free water diffusion (multivariate analysis) | 23,543 Eur | 11,555 Eur | 17q21.31 | | RPS26P8 - LINC0 | ENSG00000282238 | ENSG00000204650 | | 1758 | 9652 | rs62066084-? | rs62066084 | 0 | 62066084 | downstream_gene_variant | 1 | NR | 4.00E-57 | 56.39794 | | NA | | NR [NR] (imputed) | N |
| 9 | 17:44118848 | 17 | 44308053 | rs62070949 | 31/05/2022 | 3.4E+07 | Smith SM | 19/04/2021 | Nat Neurosci | www.ncbi.nlm.nih.gov/pubmed/33875891 | An expanded set of genome-wide association studies of brain imaging phenotypes in UK Biobank. | aparc-Desikan rh area fusiform | 21,282 Brit | 10,686 Brit | 17q21.31 | | KANSL1 - MAPK8 | ENSG00000278458 | ENSG00000282723 | | 5298 | 12919 | rs62070949-G | rs62070949 | 0 | 62070949 | intergenic_variant | 1 | 0.157 | 5.00E-26 | 25.30103 | | 0.155 | [0.13-0.18 | Affymetrix [17103079] (imputed) | N |
| 9 | 17:44118848 | 17 | 44308053 | rs62070949 | 31/05/2022 | 3.4E+07 | Smith SM | 19/04/2021 | Nat Neurosci | www.ncbi.nlm.nih.gov/pubmed/33875891 | An expanded set of genome-wide association studies of brain imaging phenotypes in UK Biobank. | aparc-DKTatlas rh area fusiform | 21,282 Brit | 10,686 Brit | 17q21.31 | | KANSL1 - MAPK8 | ENSG00000278458 | ENSG00000282723 | | 5298 | 12919 | rs62070949-G | rs62070949 | 0 | 62070949 | intergenic_variant | 1 | 0.157 | 3.00E-24 | 23.52288 | | 0.15 | [0.12-0.18 | Affymetrix [17103079] (imputed) | N |
| 9 | 17:44118848 | 17 | 44308053 | rs62070949 | 31/05/2022 | 3.4E+07 | Smith SM | 19/04/2021 | Nat Neurosci | www.ncbi.nlm.nih.gov/pubmed/33875891 | An expanded set of genome-wide association studies of brain imaging phenotypes in UK Biobank. | aparc-pial rh area fusiform | 21,282 Brit | 10,686 Brit | 17q21.31 | | KANSL1 - MAPK8 | ENSG00000278458 | ENSG00000282723 | | 5298 | 12919 | rs62070949-G | rs62070949 | 0 | 62070949 | intergenic_variant | 1 | 0.157 | 3.00E-18 | 17.52288 | | 0.128 | [0.099-0.1 | Affymetrix [17103079] (imputed) | N |
| 9 | 17:44118848 | 17 | 44316491 | rs62073099 | 4/08/2022 | 3.6E+07 | May-Wilson S | 18/05/2022 | Nat Commun | www.ncbi.nlm.nih.gov/pubmed/35585065 | Large-scale GWAS of food liking reveals genetic determinants and genetic correlations with distinct neurophysiological traits. | F-bitter food liking (derived food-liking factor) | 157,485 E | NA | 17q21.31 | | KANSL1 - MAPK8 | ENSG00000278458 | ENSG00000282723 | | 13736 | 4481 | rs62073099-C | rs62073099 | 0 | 62073099 | upstream_gene_variant | 1 | 0.1997 | 2.00E-10 | 9.69897 | | 0.02904 | [0.02-0.03 | Affymetrix [11353280] (imputed) | N |
| 9 | 17:44118848 | 17 | 44316491 | rs62073099 | 4/08/2022 | 3.6E+07 | May-Wilson S | 18/05/2022 | Nat Commun | www.ncbi.nlm.nih.gov/pubmed/35585065 | Large-scale GWAS of food liking reveals genetic determinants and genetic correlations with distinct neurophysiological traits. | F-strong flavour liking (derived food-liking factor) | 153,924 E | NA | 17q21.31 | | KANSL1 - MAPK8 | ENSG00000278458 | ENSG00000282723 | | 13736 | 4481 | rs62073099-C | rs62073099 | 0 | 62073099 | upstream_gene_variant | 1 | 0.1996 | 1.00E-16 | 16 | | 0.09416 | [0.072-0.1 | Affymetrix [9535166] (imputed) | N |
| 9 | 17:44118848 | 17 | 44388912 | rs62073233 | 27/04/2022 | 3.5E+07 | Richardson TG | 25/02/2022 | PLoS Biol | www.ncbi.nlm.nih.gov/pubmed/35213538 | Characterising metabolomic signatures of lipid-modifying therapies through drug target mendelian randomisation. | Glucose levels | 114,870 E | NA | 17q21.31 | | LRRC37A, LRRC37A, ARL17B, ARL17B, ARL17B | | | ENSG00000176681, E | NA | NA | rs62073233-G | rs62073233 | 0 | 62073233 | intron_variant | 0 | 0.78721 | 2.00E-09 | 8.69897 | | 0.033992 | [0.023-0.0 | NR [11722792] (imputed) | N |

| | | | | | | | | | | | | | | | | | | | | | | | | | | | | | | | | | | | | |
|---|---|---|---|---|---|---|---|---|---|---|---|---|---|---|---|---|---|---|---|---|---|---|---|---|---|---|---|---|---|---|---|---|---|---|---|---|
| 9 | 17:44118848 | 17 | 43696159 | rs675600 | 30/08/2017 | 2.8E+07 | Astle WJ | 17/11/2016 | Cell | www.ncbi.nlm.nih.gov/pubmed/27863252 | The Allelic Landscape of Human Blood Cell Trait Variation and Links to Common Complex Disease. | Granulocyte count | 169,822 E | NA | 17q21.31 | CRHR1-IT1, CRH | RPS26P8 - LINC0 | ENSG00000282238 | ENSG00000204650 | | 9875 | 1535 | rs675600-T | rs675600 | 0 | 675600 | intergenic_variant | 1 | 0.2046 | 5.00E-09 | 8.30103 | | 0.026739 | [0.018-0.0 | Affymetrix [~ 2500000] (imputed) | N |
| 9 | 17:44118848 | 17 | 43696159 | rs675600 | 30/08/2017 | 2.8E+07 | Astle WJ | 17/11/2016 | Cell | www.ncbi.nlm.nih.gov/pubmed/27863252 | The Allelic Landscape of Human Blood Cell Trait Variation and Links to Common Complex Disease. | Myeloid white cell count | 169,219 E | NA | 17q21.31 | CRHR1-IT1, CRH | RPS26P8 - LINC0 | ENSG00000282238 | ENSG00000204650 | | 9875 | 1535 | rs675600-T | rs675600 | 0 | 675600 | intergenic_variant | 1 | 0.2046 | 3.00E-09 | 8.522879 | | 0.027331 | [0.018-0.0 | Affymetrix [~ 2500000] (imputed) | N |
| 9 | 17:44118848 | 17 | 43696159 | rs675600 | 30/08/2017 | 2.8E+07 | Astle WJ | 17/11/2016 | Cell | www.ncbi.nlm.nih.gov/pubmed/27863252 | The Allelic Landscape of Human Blood Cell Trait Variation and Links to Common Complex Disease. | Neutrophil count | 170,702 E | NA | 17q21.31 | CRHR1-IT1, CRH | RPS26P8 - LINC0 | ENSG00000282238 | ENSG00000204650 | | 9875 | 1535 | rs675600-T | rs675600 | 0 | 675600 | intergenic_variant | 1 | 0.2045 | 3.00E-11 | 10.52288 | | 0.030358 | [0.021-0.0 | Affymetrix [~ 2500000] (imputed) | N |
| 9 | 17:44118848 | 17 | 43696159 | rs675600 | 30/08/2017 | 2.8E+07 | Astle WJ | 17/11/2016 | Cell | www.ncbi.nlm.nih.gov/pubmed/27863252 | The Allelic Landscape of Human Blood Cell Trait Variation and Links to Common Complex Disease. | Sum basophil neutrophil counts | 170,143 E | NA | 17q21.31 | CRHR1-IT1, CRH | RPS26P8 - LINC0 | ENSG00000282238 | ENSG00000204650 | | 9875 | 1535 | rs675600-T | rs675600 | 0 | 675600 | intergenic_variant | 1 | 0.2046 | 2.00E-11 | 10.69897 | | 0.030561 | [0.022-0.0 | Affymetrix [~ 2500000] (imputed) | N |
| 9 | 17:44118848 | 17 | 43696159 | rs675600 | 30/08/2017 | 2.8E+07 | Astle WJ | 17/11/2016 | Cell | www.ncbi.nlm.nih.gov/pubmed/27863252 | The Allelic Landscape of Human Blood Cell Trait Variation and Links to Common Complex Disease. | Sum neutrophil eosinophil counts | 170,384 E | NA | 17q21.31 | CRHR1-IT1, CRH | RPS26P8 - LINC0 | ENSG00000282238 | ENSG00000204650 | | 9875 | 1535 | rs675600-T | rs675600 | 0 | 675600 | intergenic_variant | 1 | 0.2045 | 6.00E-09 | 8.221849 | | 0.026524 | [0.018-0.0 | Affymetrix [~ 2500000] (imputed) | N |
| 9 | 17:44118848 | 17 | 44860021 | rs70600 | 22/01/2019 | 3E+07 | Nagel M | 2/03/2018 | Nat Commun | www.ncbi.nlm.nih.gov/pubmed/29500382 | Item-level analyses reveal genetic heterogeneity in neuroticism. | Irritable mood | 366,726 E | NA | 17q21.31 | NR | WNT3 | | | ENSG00000108379 | NA | NA | rs70600-T | rs70600 | 0 | 70600 | intron_variant | 0 | 0.20718 | 3.00E-11 | 10.52288 | | 6.65 | z score inc | NR [~ 10847151] (imputed) | N |
| 9 | 17:44118848 | 17 | 44332793 | rs71375338 | 18/09/2020 | 3.3E+07 | Vuckovic D | 1/09/2020 | Cell | www.ncbi.nlm.nih.gov/pubmed/32888494 | The Polygenic and Monogenic Basis of Blood Traits and Diseases. | Neutrophil count | 408,112 B | NA | 17q21.31 | RP11-259G18.3 | MAPK8IP1P1 - AR | ENSG00000282723 | ENSG00000228696 | | 10383 | 19357 | rs71375338-A | rs71375338 | 0 | 71375338 | intergenic_variant | 1 | 0.268548 | 3.00E-20 | 19.52288 | | 0.023167 | [0.018-0.0 | Affymetrix [93095623] (imputed) | N |
| 9 | 17:44118848 | 17 | 44332793 | rs71375338 | 21/09/2020 | 3.3E+07 | Chen MH | 1/09/2020 | Cell | www.ncbi.nlm.nih.gov/pubmed/32888493 | Trans-ethnic and Ancestry-Specific Blood-Cell Genetics in 746,667 Individuals from 5 Global Populations. | Neutrophil count | 627,215 A | NA | 17q21.31 | NR | MAPK8IP1P1 - AR | ENSG00000282723 | ENSG00000228696 | | 10383 | 19357 | rs71375338-A | rs71375338 | 0 | 71375338 | intergenic_variant | 1 | 0.259541 | 3.00E-22 | 21.52288 | | NA | | Affymetrix, Illumina [23114412] (imputed) | N |
| 9 | 17:44118848 | 17 | 44332793 | rs71375338 | 13/12/2021 | 3.4E+07 | Kachuri L | 26/08/2021 | Am J Hum Genet | www.ncbi.nlm.nih.gov/pubmed/34469753 | Genetic determinants of blood-cell traits influence susceptibility to childhood acute lymphoblastic leukemia. | Neutrophil-to-lymphocyte ratio | 234,502 E | 100,442 E | 17q21.31 | | MAPK8IP1P1 - AR | ENSG00000282723 | ENSG00000228696 | | 10383 | 19357 | rs71375338-G | rs71375338 | 0 | 71375338 | intergenic_variant | 1 | 0.73 | 3.00E-16 | 15.52288 | | NA | | Affymetrix [10369434] (imputed) | N |
| 9 | 17:43520118 | 17 | 43484598 | rs73984391 | 31/05/2022 | 3.4E+07 | Smith SM | 19/04/2021 | Nat Neurosci | www.ncbi.nlm.nih.gov/pubmed/33875891 | An expanded set of genome-wide association studies of brain imaging phenotypes in UK Biobank. | IDP dMRI TBSS L3 Anterior corona radiata L | 20,860 Brit | 10,496 Brit | 17q21.31 | | ARHGAP27 | | | ENSG00000159314 | NA | NA | rs73984391-T | rs73984391 | 0 | 73984391 | intron_variant | 0 | 0.182 | 2.00E-10 | 9.69897 | | 0.081 | [0.056-0.1 | Affymetrix [17103079] (imputed) | N |
| 9 | 17:44118848 | 17 | 44260845 | rs74693982 | 31/05/2022 | 3.4E+07 | Smith SM | 19/04/2021 | Nat Neurosci | www.ncbi.nlm.nih.gov/pubmed/33875891 | An expanded set of genome-wide association studies of brain imaging phenotypes in UK Biobank. | IDP dMRI ProbtrackX L1 slf l | 20,860 Brit | 10,495 Brit | 17q21.31 | | KANSL1, KANSL1, KANSL1 | | | ENSG00000120071, E | NA | NA | rs74693982-A | rs74693982 | 0 | 74693982 | intron_variant | 0 | 0.19 | 9.00E-09 | 8.045757 | | 0.076 | [0.051-0.1 | Affymetrix [17103079] (imputed) | N |
| 9 | 17:44118848 | 17 | 43810873 | rs75022332 | 22/01/2019 | 3E+07 | Nagel M | 2/03/2018 | Nat Commun | www.ncbi.nlm.nih.gov/pubmed/29500382 | Item-level analyses reveal genetic heterogeneity in neuroticism. | Worry too long after an embarrassing experience | 367,725 E | NA | 17q21.31 | ACBD4 | LINC02210-CRHR1, LINC02210-CRHR1 | | | ENSG00000282456, E | NA | NA | rs75022332-A | rs75022332 | 0 | 75022332 | non_coding_transcript_exon | 0 | 0.213608 | 2.00E-08 | 7.69897 | | 5.62 | z score dec | NR [10847151] (imputed) | N |
| 9 | 17:44118848 | 17 | 44357654 | rs75067210 | 4/08/2022 | 3.6E+07 | May-Wilson S | 18/05/2022 | Nat Commun | www.ncbi.nlm.nih.gov/pubmed/35585065 | Large-scale GWAS of food liking reveals genetic determinants and genetic correlations with distinct neurophysiological traits. | Beans liking | 159,014 E | NA | 17q21.31 | | ARL17B, ARL17B, ARL17B | | | ENSG00000275748, E | NA | NA | rs75067210-T | rs75067210 | 0 | 75067210 | intron_variant | 0 | 0.1805 | 1.00E-09 | 9 | | 0.09898 | [0.067-0.1 | Affymetrix [9618863] (imputed) | N |
| 9 | 17:44118848 | 17 | 44357654 | rs75067210 | 4/08/2022 | 3.6E+07 | May-Wilson S | 18/05/2022 | Nat Commun | www.ncbi.nlm.nih.gov/pubmed/35585065 | Large-scale GWAS of food liking reveals genetic determinants and genetic correlations with distinct neurophysiological traits. | Broad bean liking | 158,921 E | NA | 17q21.31 | | ARL17B, ARL17B, ARL17B | | | ENSG00000275748, E | NA | NA | rs75067210-T | rs75067210 | 0 | 75067210 | intron_variant | 0 | 0.1805 | 3.00E-10 | 9.522879 | | 0.03108 | [0.021-0.0 | Affymetrix [11353280] (imputed) | N |
| 9 | 17:44118848 | 17 | 43973233 | rs76324150 | 28/05/2020 | 3.2E+07 | Hysi PG | 30/03/2020 | Nat Genet | www.ncbi.nlm.nih.gov/pubmed/32231278 | Meta-analysis of 542,934 subjects of European ancestry identifies new genes and mechanisms predisposing to refractive error and myopia. | Refractive error | 542,934 E | NA | 17q21.31 | ARHGAP27, MAPT | MAPT-IT1, MAPT-IT1, MAPT-IT1, MAPT, MAPT, MAPT | | | ENSG00000281284, E | NA | NA | rs76324150-? | rs76324150 | 0 | 76324150 | non_coding_transcript_exon | 0 | | 8.00E-16 | 15.09691 | | NA | | Affymetrix, Illumina [NR] (imputed) | N |
| 9 | 17:44118848 | 17 | 43758887 | rs76539714 | 24/10/2022 | 3.4E+07 | Harrison S | 28/07/2021 | Sci Adv | www.ncbi.nlm.nih.gov/pubmed/34321204 | Testosterone and socioeconomic position: Mendelian randomization in 306,248 men and women in UK Biobank. | Albumin levels | 158,000 B | NA | 17q21.31 | | LINC02210-CRHR1, LINC02210-CRHR1 | | | ENSG00000282456, E | NA | NA | rs76539714-G | rs76539714 | 0 | 76539714 | intron_variant | 0 | 0.23 | 4.00E-10 | 9.39794 | | 0.09 | [0.061-0.1 | Affymetrix [NR] (imputed) | N |
| 9 | 17:44118848 | 17 | 43758887 | rs76539714 | 31/05/2022 | 3.4E+07 | Smith SM | 19/04/2021 | Nat Neurosci | www.ncbi.nlm.nih.gov/pubmed/33875891 | An expanded set of genome-wide association studies of brain imaging phenotypes in UK Biobank. | aparc-Desikan lh area fusiform | 21,282 Brit | 10,686 Brit | 17q21.31 | | LINC02210-CRHR1, LINC02210-CRHR1 | | | ENSG00000282456, E | NA | NA | rs76539714-G | rs76539714 | 0 | 76539714 | intron_variant | 0 | 0.224 | 1.00E-22 | 22 | | 0.114 | [0.09-0.13 | Affymetrix [17103079] (imputed) | N |
| 9 | 17:44118848 | 17 | 43758887 | rs76539714 | 31/05/2022 | 3.4E+07 | Smith SM | 19/04/2021 | Nat Neurosci | www.ncbi.nlm.nih.gov/pubmed/33875891 | An expanded set of genome-wide association studies of brain imaging phenotypes in UK Biobank. | aparc-DKTatlas lh area fusiform | 21,282 Brit | 10,686 Brit | 17q21.31 | | LINC02210-CRHR1, LINC02210-CRHR1 | | | ENSG00000282456, E | NA | NA | rs76539714-G | rs76539714 | 0 | 76539714 | intron_variant | 0 | 0.224 | 4.00E-23 | 22.39794 | | 0.116 | [0.092-0.1 | Affymetrix [17103079] (imputed) | N |
| 9 | 17:44118848 | 17 | 43758887 | rs76539714 | 31/05/2022 | 3.4E+07 | Smith SM | 19/04/2021 | Nat Neurosci | www.ncbi.nlm.nih.gov/pubmed/33875891 | An expanded set of genome-wide association studies of brain imaging phenotypes in UK Biobank. | IDP dMRI TBSS L1 External capsule R | 20,860 Brit | 10,496 Brit | 17q21.31 | | LINC02210-CRHR1, LINC02210-CRHR1 | | | ENSG00000282456, E | NA | NA | rs76539714-G | rs76539714 | 0 | 76539714 | intron_variant | 0 | 0.224 | 6.00E-15 | 14.22185 | | 0.092 | [0.068-0.1 | Affymetrix [17103079] (imputed) | N |
| 9 | 17:44118848 | 17 | 44189858 | rs76640332 | 30/08/2017 | 2.8E+07 | Astle WJ | 17/11/2016 | Cell | www.ncbi.nlm.nih.gov/pubmed/27863252 | The Allelic Landscape of Human Blood Cell Trait Variation and Links to Common Complex Disease. | Lymphocyte percentage of white cells | 171,748 E | NA | 17q21.31 | KANSL1 | KANSL1, KANSL1, KANSL1 | | | ENSG00000120071, E | NA | NA | rs76640332-A | rs76640332 | 0 | 76640332 | intron_variant | 0 | 0.222 | 5.00E-13 | 12.30103 | | 0.030675 | [0.022-0.0 | Affymetrix [~ 2500000] (imputed) | N |
| 9 | 17:44118848 | 17 | 43776741 | rs76761706 | 22/01/2019 | 3E+07 | Nagel M | 2/03/2018 | Nat Commun | www.ncbi.nlm.nih.gov/pubmed/29500382 | Item-level analyses reveal genetic heterogeneity in neuroticism. | Neurociticism | 380,506 E | NA | 17q21.31 | NR | LINC02210-CRHR1, LINC02210-CRHR1 | | | ENSG00000282456, E | NA | NA | rs76761706-T | rs76761706 | 0 | 76761706 | intron_variant | 0 | 0.215553 | 7.00E-32 | 31.1549 | | 11.76 | z score dec | NR [10847151] (imputed) | N |
| 9 | 17:44118848 | 17 | 43658409 | rs769375 | 31/05/2022 | 3.4E+07 | Smith SM | 19/04/2021 | Nat Neurosci | www.ncbi.nlm.nih.gov/pubmed/33875891 | An expanded set of genome-wide association studies of brain imaging phenotypes in UK Biobank. | IDP dMRI TBSS L3 Superior fronto-occipital fasciculus L | 20,860 Brit | 10,496 Brit | 17q21.31 | | LRRC37A4P, LRRC37A4P | | | ENSG00000214425, E | NA | NA | rs769375-A | rs769375 | 0 | 769375 | intron_variant | 0 | 0.211 | 7.00E-11 | 10.1549 | | 0.078 | [0.054-0.1 | Affymetrix [17103079] (imputed) | N |
| 9 | 17:44118848 | 17 | 43810896 | rs77804065 | 22/01/2019 | 3E+07 | Nagel M | 2/03/2018 | Nat Commun | www.ncbi.nlm.nih.gov/pubmed/29500382 | Item-level analyses reveal genetic heterogeneity in neuroticism. | Feeling guilty | 373,380 E | NA | 17q21.31 | ACBD4 | LINC02210-CRHR1, LINC02210-CRHR1 | | | ENSG00000282456, E | NA | NA | rs77804065-T | rs77804065 | 0 | 77804065 | splice_region_variant | 0 | 0.217158 | 6.00E-10 | 9.221849 | | 6.18 | z score inc | NR [10847151] (imputed) | N |
| 9 | 17:44118848 | 17 | 43810896 | rs77804065 | 1/03/2018 | 2.9E+07 | Meng W | 31/01/2018 | EBioMedicine | www.ncbi.nlm.nih.gov/pubmed/29397368 | A Genome-Wide Association Study Finds Genetic Associations with Broadly-Defined Headache in UK Biobank (N=223,773). | Headache | 74,461 Brit | NA | 17q21.31 | LINC02210, CRH | LINC02210-CRHR1, LINC02210-CRHR1 | | | ENSG00000282456, E | NA | NA | rs77804065-T | rs77804065 | 0 | 77804065 | splice_region_variant | 0 | NR | 6.00E-15 | 14.22185 | | 0.013 | [0.0097-0. | Affymetrix [9304965] (imputed) | N |
| 9 | 17:44118848 | 17 | 43810896 | rs77804065 | 18/09/2019 | 3.1E+07 | Gelernter J | 8/04/2019 | Biol Psychiatry | www.ncbi.nlm.nih.gov/pubmed/31151762 | Genome-wide Association Study of Maximum Habitual Alcohol Intake in >140,000 U.S. European and African American Veterans Yields Novel Risk Loci. | Maximum habitual alcohol consumption | 126,936 E | NA | 17q21.31 | CRHR1 | LINC02210-CRHR1, LINC02210-CRHR1 | | | ENSG00000282456, E | NA | NA | rs77804065-T | rs77804065 | 0 | 77804065 | splice_region_variant | 0 | NR | 2.00E-12 | 11.69897 | (EA) | 0.0711 | NR unit de | Affymetrix [up to 6007810] (imputed) | N |
| 9 | 17:44118848 | 17 | 43810896 | rs77804065 | 31/10/2018 | 3E+07 | Nagel M | 25/06/2018 | Nat Genet | www.ncbi.nlm.nih.gov/pubmed/29942085 | Meta-analysis of genome-wide association studies for neuroticism in 449,484 individuals identifies novel genetic loci and pathways. | Neuroticism | 449,484 E | NA | 17q21.31 | NR | LINC02210-CRHR1, LINC02210-CRHR1 | | | ENSG00000282456, E | NA | NA | rs77804065-T | rs77804065 | 0 | 77804065 | splice_region_variant | 0 | NR | 1.00E-31 | 31 | | 11.693 | z score inc | Affymetrix [10847151] (imputed) | N |
| 9 | 17:44118848 | 17 | 43810896 | rs77804065 | 1/02/2018 | 2.9E+07 | Luciano M | 18/12/2017 | Nat Genet | www.ncbi.nlm.nih.gov/pubmed/29255261 | Association analysis in over 329,000 individuals identifies 116 independent variants influencing neuroticism. | Neuroticism | 329,821 E | 122,867 E | 17q21.31 | ARHGAP27, CRH | LINC02210-CRHR1, LINC02210-CRHR1 | | | ENSG00000282456, E | NA | NA | rs77804065-T | rs77804065 | 0 | 77804065 | splice_region_variant | 0 | 0.21716 | 8.00E-26 | 25.09691 | | 10.503 | unit increa | Illumina [18485882] (imputed) | N |
| 9 | 17:44118848 | 17 | 44051612 | rs77875796 | 29/04/2022 | 3.5E+07 | Ko H | 6/10/2021 | Brain | www.ncbi.nlm.nih.gov/pubmed/34613391 | Genome-wide association study of occupational attainment as a proxy for cognitive reserve. | Occupational attainment | 248,847 E | NA | 17q21.31 | | MAPT, MAPT, MAPT | | | ENSG00000277956, E | NA | NA | rs77875796-G | rs77875796 | 0 | 77875796 | 5_prime_UTR_variant | 0 | 0.241 | 5.00E-09 | 8.30103 | | 0.049 | [0.033-0.0 | Affymetrix [9575249] (imputed) | N |
| 9 | 17:44118848 | 17 | 44339607 | rs77917260 | 31/05/2022 | 3.4E+07 | Smith SM | 19/04/2021 | Nat Neurosci | www.ncbi.nlm.nih.gov/pubmed/33875891 | An expanded set of genome-wide association studies of brain imaging phenotypes in UK Biobank. | IDP dMRI TBSS L1 External capsule L | 20,860 Brit | 10,496 Brit | 17q21.31 | | MAPK8IP1P1 - AR | ENSG00000282723 | ENSG00000228696 | | 17197 | 12543 | rs77917260-T | rs77917260 | 0 | 77917260 | regulatory_region_variant | 1 | 0.212 | 1.00E-16 | 16 | | 0.1 | [0.076-0.1 | Affymetrix [17103079] (imputed) | N |
| 9 | 17:44118848 | 17 | 44290850 | rs78826239 | 31/05/2022 | 3.4E+07 | Smith SM | 19/04/2021 | Nat Neurosci | www.ncbi.nlm.nih.gov/pubmed/33875891 | An expanded set of genome-wide association studies of brain imaging phenotypes in UK Biobank. | aparc-a2009s rh area G-oc-temp-lat-fusifor | 21,282 Brit | 10,686 Brit | 17q21.31 | | KANSL1, KANSL1, KANSL1 | | | ENSG00000120071, E | NA | NA | rs78826239-A | rs78826239 | 0 | 78826239 | intron_variant | 0 | 0.202 | 2.00E-11 | 10.69897 | | 0.084 | [0.059-0.1 | Affymetrix [17103079] (imputed) | N |
| 9 | 17:44118848 | 17 | 44189910 | rs78929339 | 14/12/2021 | 3.5E+07 | Sakaue S | 30/09/2021 | Nat Genet | www.ncbi.nlm.nih.gov/pubmed/34594039 | A cross-population atlas of genetic associations for 220 human phenotypes. | Medication use (anilides) | 83,218 Eur | NA | 17q21.31 | | KANSL1, KANSL1, KANSL1 | | | ENSG00000120071, E | NA | NA | rs78929339-G | rs78929339 | 0 | 78929339 | intron_variant | 0 | NR | 9.00E-13 | 12.04576 | | 0.0559 | [0.041-0.0 | Affymetrix, Illumina [15248280] (imputed) | N |
| 9 | 17:44118848 | 17 | 43828764 | rs79172804 | 31/05/2022 | 3.4E+07 | Smith SM | 19/04/2021 | Nat Neurosci | www.ncbi.nlm.nih.gov/pubmed/33875891 | An expanded set of genome-wide association studies of brain imaging phenotypes in UK Biobank. | IDP dMRI TBSS ICVF Superior fronto-occipital fasciculus L | 20,859 Brit | 10,496 Brit | 17q21.31 | | LINC02210-CRHR1, LINC02210-CRHR1 | | | ENSG00000282456, E | NA | NA | rs79172804-A | rs79172804 | 0 | 79172804 | intron_variant | 0 | 0.189 | 2.00E-23 | 22.69897 | | 0.126 | [0.1-0.15] | Affymetrix [17103079] (imputed) | N |
| 9 | 17:44118848 | 17 | 43828764 | rs79172804 | 31/05/2022 | 3.4E+07 | Smith SM | 19/04/2021 | Nat Neurosci | www.ncbi.nlm.nih.gov/pubmed/33875891 | An expanded set of genome-wide association studies of brain imaging phenotypes in UK Biobank. | IDP dMRI TBSS ICVF Superior fronto-occipital fasciculus R | 20,859 Brit | 10,496 Brit | 17q21.31 | | LINC02210-CRHR1, LINC02210-CRHR1 | | | ENSG00000282456, E | NA | NA | rs79172804-A | rs79172804 | 0 | 79172804 | intron_variant | 0 | 0.189 | 3.00E-27 | 26.52288 | | 0.137 | [0.11-0.16 | Affymetrix [17103079] (imputed) | N |
| 9 | 17:44118848 | 17 | 43828764 | rs79172804 | 31/05/2022 | 3.4E+07 | Smith SM | 19/04/2021 | Nat Neurosci | www.ncbi.nlm.nih.gov/pubmed/33875891 | An expanded set of genome-wide association studies of brain imaging phenotypes in UK Biobank. | IDP dMRI TBSS L1 Superior fronto-occipital fasciculus R | 20,860 Brit | 10,496 Brit | 17q21.31 | | LINC02210-CRHR1, LINC02210-CRHR1 | | | ENSG00000282456, E | NA | NA | rs79172804-A | rs79172804 | 0 | 79172804 | intron_variant | 0 | 0.189 | 8.00E-13 | 12.09691 | | 0.091 | [0.066-0.1 | Affymetrix [17103079] (imputed) | N |
| 9 | 17:44118848 | 17 | 43828764 | rs79172804 | 31/05/2022 | 3.4E+07 | Smith SM | 19/04/2021 | Nat Neurosci | www.ncbi.nlm.nih.gov/pubmed/33875891 | An expanded set of genome-wide association studies of brain imaging phenotypes in UK Biobank. | IDP dMRI TBSS L2 Superior fronto-occipital fasciculus L | 20,860 Brit | 10,496 Brit | 17q21.31 | | LINC02210-CRHR1, LINC02210-CRHR1 | | | ENSG00000282456, E | NA | NA | rs79172804-A | rs79172804 | 0 | 79172804 | intron_variant | 0 | 0.189 | 1.00E-11 | 11 | | 0.086 | [0.061-0.1 | Affymetrix [17103079] (imputed) | N |
| 9 | 17:44118848 | 17 | 43828764 | rs79172804 | 31/05/2022 | 3.4E+07 | Smith SM | 19/04/2021 | Nat Neurosci | www.ncbi.nlm.nih.gov/pubmed/33875891 | An expanded set of genome-wide association studies of brain imaging phenotypes in UK Biobank. | IDP dMRI TBSS MD Superior corona radiata L | 20,860 Brit | 10,496 Brit | 17q21.31 | | LINC02210-CRHR1, LINC02210-CRHR1 | | | ENSG00000282456, E | NA | NA | rs79172804-A | rs79172804 | 0 | 79172804 | intron_variant | 0 | 0.189 | 2.00E-12 | 11.69897 | | 0.089 | [0.064-0.1 | Affymetrix [17103079] (imputed) | N |
| 9 | 17:44118848 | 17 | 43828764 | rs79172804 | 31/05/2022 | 3.4E+07 | Smith SM | 19/04/2021 | Nat Neurosci | www.ncbi.nlm.nih.gov/pubmed/33875891 | An expanded set of genome-wide association studies of brain imaging phenotypes in UK Biobank. | IDP dMRI TBSS MD Superior fronto-occipital fasciculus L | 20,860 Brit | 10,496 Brit | 17q21.31 | | LINC02210-CRHR1, LINC02210-CRHR1 | | | ENSG00000282456, E | NA | NA | rs79172804-A | rs79172804 | 0 | 79172804 | intron_variant | 0 | 0.189 | 3.00E-13 | 12.52288 | | 0.092 | [0.067-0.1 | Affymetrix [17103079] (imputed) | N |
| 9 | 17:44118848 | 17 | 43828764 | rs79172804 | 31/05/2022 | 3.4E+07 | Smith SM | 19/04/2021 | Nat Neurosci | www.ncbi.nlm.nih.gov/pubmed/33875891 | An expanded set of genome-wide association studies of brain imaging phenotypes in UK Biobank. | IDP dMRI TBSS MD Superior fronto-occipital fasciculus R | 20,860 Brit | 10,496 Brit | 17q21.31 | | LINC02210-CRHR1, LINC02210-CRHR1 | | | ENSG00000282456, E | NA | NA | rs79172804-A | rs79172804 | 0 | 79172804 | intron_variant | 0 | 0.189 | 2.00E-18 | 17.69897 | | 0.111 | [0.086-0.1 | Affymetrix [17103079] (imputed) | N |
| 9 | 17:44118848 | 17 | 44207932 | rs79301522 | 22/01/2019 | 3E+07 | Nagel M | 2/03/2018 | Nat Commun | www.ncbi.nlm.nih.gov/pubmed/29500382 | Item-level analyses reveal genetic heterogeneity in neuroticism. | Neurociticism | 380,506 E | NA | 17q21.31 | NR | KANSL1, KANSL1, KANSL1 | | | ENSG00000120071, E | NA | NA | rs79301522-A | rs79301522 | 0 | 79301522 | intron_variant | 0 | 0.217029 | 1.00E-30 | 30 | | 11.51 | z score dec | NR [10847151] (imputed) | N |
| 9 | 17:44118848 | 17 | 43940021 | rs79412431 | 27/03/2019 | 3.1E+07 | Shrine N | 25/02/2019 | Nat Genet | www.ncbi.nlm.nih.gov/pubmed/30804560 | New genetic signals for lung function highlight pathways and chronic obstructive pulmonary disease associations across multiple ancestries. | FEV1 | 321,047 E | 79,005 Eur | 17q21.31 | MAPT-AS1 | MAPT-AS1, MAPT-AS1, MAPT-AS1 | | | ENSG00000264589, E | NA | NA | rs79412431-A | rs79412431 | 0 | 79412431 | intron_variant | 0 | 0.2161 | 3.00E-49 | 48.52288 | | 0.0426 | [0.037-0.0 | Affymetrix [19819130] (imputed) | N |
| 9 | 17:44118848 | 17 | 43940021 | rs79412431 | 27/03/2019 | 3.1E+07 | Shrine N | 25/02/2019 | Nat Genet | www.ncbi.nlm.nih.gov/pubmed/30804560 | New genetic signals for lung function highlight pathways and chronic obstructive pulmonary disease associations across multiple ancestries. | Lung function (FVC) | 321,047 E | 79,005 Eur | 17q21.31 | MAPT-AS1 | MAPT-AS1, MAPT-AS1, MAPT-AS1 | | | ENSG00000264589, E | NA | NA | rs79412431-A | rs79412431 | 0 | 79412431 | intron_variant | 0 | 0.2161 | 3.00E-46 | 45.52288 | | 0.0415 | [0.036-0.0 | Affymetrix [19819130] (imputed) | N |
| 9 | 17:44118848 | 17 | 43940021 | rs79412431 | 27/03/2019 | 3.1E+07 | Shrine N | 25/02/2019 | Nat Genet | www.ncbi.nlm.nih.gov/pubmed/30804560 | New genetic signals for lung function highlight pathways and chronic obstructive pulmonary disease associations across multiple ancestries. | Peak expiratory flow | 321,047 E | 24,218 Eur | 17q21.31 | MAPT-AS1 | MAPT-AS1, MAPT-AS1, MAPT-AS1 | | | ENSG00000264589, E | NA | NA | rs79412431-A | rs79412431 | 0 | 79412431 | intron_variant | 0 | 0.2161 | 6.00E-15 | 14.22185 | | 0.0239 | [0.018-0.0 | Affymetrix [19819130] (imputed) | N |
| 9 | 17:44118848 | 17 | 43897722 | rs79600142 | 21/07/2020 | 3.2E+07 | Grasby KL | 1/03/2020 | Science | www.ncbi.nlm.nih.gov/pubmed/32193296 | The genetic architecture of the human cerebral cortex. | Cortical surface area | 33,992 Eur | 14,727 Eur | 17q21.31 | NR | MAPT-AS1, MAPT-AS1, MAPT-AS1, CRHR1, CRHR1, CRHR1, L | | | ENSG00000264589, E | NA | NA | rs79600142-T | rs79600142 | 0 | 79600142 | intron_variant | 0 | 0.79 | 1.00E-43 | 43 | (total) | 1414.626 | [1214.65-1 | Affymetrix, Illumina [NR] | N |
| 9 | 17:44118848 | 17 | 43463493 | rs79724577 | 31/05/2022 | 3.4E+07 | Smith SM | 19/04/2021 | Nat Neurosci | www.ncbi.nlm.nih.gov/pubmed/33875891 | An expanded set of genome-wide association studies of brain imaging phenotypes in UK Biobank. | IDP dMRI ProbtrackX MD atr l | 20,860 Brit | 10,495 Brit | 17q21.31 | | RNA5SP443 - AR | ENSG00000199953 | ENSG00000159314 | | 58630 | 7775 | rs79724577-C | rs79724577 | 0 | 79724577 | regulatory_region_variant | 1 | 0.181 | 9.00E-13 | 12.04576 | | 0.091 | [0.066-0.1 | Affymetrix [17103079] (imputed) | N |
| 9 | 17:44118848 | 17 | 43463493 | rs79724577 | 31/05/2022 | 3.4E+07 | Smith SM | 19/04/2021 | Nat Neurosci | www.ncbi.nlm.nih.gov/pubmed/33875891 | An expanded set of genome-wide association studies of brain imaging phenotypes in UK Biobank. | IDP dMRI TBSS ICVF Anterior corona radiata R | 20,859 Brit | 10,496 Brit | 17q21.31 | | RNA5SP443 - AR | ENSG00000199953 | ENSG00000159314 | | 58630 | 7775 | rs79724577-C | rs79724577 | 0 | 79724577 | regulatory_region_variant | 1 | 0.181 | 1.00E-14 | 14 | | 0.099 | [0.074-0.1 | Affymetrix [17103079] (imputed) | N |
| 9 | 17:44118848 | 17 | 43463493 | rs79724577 | 31/05/2022 | 3.4E+07 | Smith SM | 19/04/2021 | Nat Neurosci | www.ncbi.nlm.nih.gov/pubmed/33875891 | An expanded set of genome-wide association studies of brain imaging phenotypes in UK Biobank. | IDP dMRI TBSS ICVF Retrolenticular part of internal capsule L | 20,859 Brit | 10,496 Brit | 17q21.31 | | RNA5SP443 - AR | ENSG00000199953 | ENSG00000159314 | | 58630 | 7775 | rs79724577-C | rs79724577 | 0 | 79724577 | regulatory_region_variant | 1 | 0.181 | 3.00E-13 | 12.52288 | | 0.093 | [0.068-0.1 | Affymetrix [17103079] (imputed) | N |
| 9 | 17:44118848 | 17 | 43849415 | rs79730878 | 28/01/2019 | 3.1E+07 | Morris JA | 31/12/2018 | Nat Genet | www.ncbi.nlm.nih.gov/pubmed/30598549 | An atlas of genetic influences on osteoporosis in humans and mice. | Heel bone mineral density | 426,824 B | NA | 17q21.31 | CRHR1-IT1-CRH | LINC02210-CRHR1, LINC02210-CRHR1 | | | ENSG00000282456, E | NA | NA | rs79730878-T | rs79730878 | 0 | 79730878 | intron_variant | 0 | 0.771203 | 6.00E-20 | 19.22185 | | 0.020214 | [0.016-0.0 | NR [13737936] (imputed) | N |

| | | | | | | | | | | | | | | | | | | | | | | | | | | | | | | | | | | | | |
|---|---|---|---|---|---|---|---|---|---|---|---|---|---|---|---|---|---|---|---|---|---|---|---|---|---|---|---|---|---|---|---|---|---|---|---|---|
| 9 | 17:44118848 | 17 | 43657257 | rs79742625 | 20/05/2019 | 3.1E+07 | Dudding T | 5/03/2019 | Nat Commun | www.ncbi.nlm.nih.gov/pubmed/30837455 | Genome wide analysis for mouth ulcers identifies associations at immune regulatory loci. | Mouth ulcers | 47,079 Brit | 98,298 cas | 17q21.31 | | RDM1P1 - DND1F | ENSG00000266504 | ENSG00000282614 | | 20821 | 5980 | rs79742625-T | rs79742625 | 0 | 79742625 | intron_variant | 1 | 0.192862 | 3.00E-18 | 17.52288 | | 1.060499 | [1.04-1.08 | Affymetrix, Illumina [9851866] (imputed) | N |
| 9 | 17:44118848 | 17 | 44013475 | rs79857651 | 22/01/2019 | 3E+07 | Nagel M | 2/03/2018 | Nat Commun | www.ncbi.nlm.nih.gov/pubmed/29500382 | Item-level analyses reveal genetic heterogeneity in neuroticism. | Experiencing mood swings | 373,733 Eu | NA | 17q21.31 | NR | MAPT, MAPT, MAPT | | | ENSG00000277956, E | NA | NA | rs79857651-A | rs79857651 | 0 | 79857651 | intron_variant | 0 | 0.215594 | 9.00E-20 | 19.04576 | | 9.1 | z score inc | NR [10847151] (imputed) | N |
| 9 | 17:44118848 | 17 | 44161470 | rs80028338 | 4/08/2022 | 3.6E+07 | May-Wilson S | 18/05/2022 | Nat Commun | www.ncbi.nlm.nih.gov/pubmed/35585065 | Large-scale GWAS of food liking reveals genetic determinants and genetic correlations with distinct neurophysiological traits. | F-savoury food liking (derived food-liking factor) | 157,915 Eu | NA | 17q21.31 | | KANSL1, KANSL1, KANSL1 | | | ENSG00000120071, E | NA | NA | rs80028338-C | rs80028338 | 0 | 80028338 | intron_variant | 0 | 0.2051 | 1.00E-12 | 12 | | 0.10169 | [0.074-0.1 | Affymetrix [9551341] (imputed) | N |
| 9 | 17:44118848 | 17 | 44161470 | rs80028338 | 5/02/2020 | 3.2E+07 | Zhao B | 30/10/2019 | Mol Psychiatry | www.ncbi.nlm.nih.gov/pubmed/31666681 | Large-scale GWAS reveals genetic architecture of brain white matter microstructure and genetic overlap with cognitive and mental health traits (nÔÇë=ÔÇë17,706). | White matter microstructure (fractional anisotropy) | 17,706 Eur | NA | 17q21.31 | NR | KANSL1, KANSL1, KANSL1 | | | ENSG00000120071, E | NA | NA | rs80028338-? | rs80028338 | 0 | 80028338 | intron_variant | 0 | | 4.00E-08 | 7.39794 | (Average a | NA | | NR [8955960] (imputed) | N |
| 9 | 17:44118848 | 17 | 44224940 | rs80103986 | 2/02/2018 | 2.9E+07 | Willems SM | 12/07/2017 | Nat Commun | www.ncbi.nlm.nih.gov/pubmed/29313844 | Large-scale GWAS identifies multiple loci for hand grip strength providing biological insights into muscular fitness. | Hand grip strength | 142,035 Eu | 53,145 Eur | 17q21.31 | KANSL1 | KANSL1, KANSL1, KANSL1 | | | ENSG00000120071, E | NA | NA | rs80103986-A | rs80103986 | 0 | 80103986 | intron_variant | 0 | 0.805 | 1.00E-09 | 9 | | 0.171 | [0.12-0.23 | Affymetrix [at least 17000000] (imputed) | N |
| 9 | 17:44118848 | 17 | 43776742 | rs80143279 | 7/07/2020 | 3.2E+07 | Nagel M | 23/01/2020 | Acta Psychiatr Scand | www.ncbi.nlm.nih.gov/pubmed/31972866 | Genome-wide association study of the sensitivity to environmental stress and adversity (SESA) neuroticism cluster. | Sensitivity to environmental stress and adversity | 351,827 Eu | NA | 17q21.31 | NR | LINC02210-CRHR1, LINC02210-CRHR1 | | | ENSG00000282456, E | NA | NA | rs80143279-C | rs80143279 | 0 | 80143279 | intron_variant | 0 | NR | 8.00E-22 | 21.09691 | | 0.02682 | NR unit inc | NR [7259579] | N |
| 9 | 17:44118848 | 17 | 44081064 | rs8070723 | 5/04/2022 | 3.5E+07 | Pasman JA | 2/12/2021 | Behav Genet | www.ncbi.nlm.nih.gov/pubmed/34855049 | Genetic Risk for Smoking: Disentangling Interplay Between Genes and Socioeconomic Status. | Lifetime smoking | 272,943 Eu | NA | 17q21.31 | | MAPT, MAPT, MAPT | | | ENSG00000277956, E | NA | NA | rs8070723-? | rs8070723 | 0 | 8070723 | intron_variant | 0 | NR | 5.00E-08 | 7.30103 | | NA | | NR [1300000] | N |
| 9 | 17:44118848 | 17 | 44081064 | rs8070723 | 5/04/2022 | 3.5E+07 | Pasman JA | 2/12/2021 | Behav Genet | www.ncbi.nlm.nih.gov/pubmed/34855049 | Genetic Risk for Smoking: Disentangling Interplay Between Genes and Socioeconomic Status. | Lifetime smoking (without educational attainment) | 272,943 Eu | NA | 17q21.31 | | MAPT, MAPT, MAPT | | | ENSG00000277956, E | NA | NA | rs8070723-? | rs8070723 | 0 | 8070723 | intron_variant | 0 | NR | 5.00E-10 | 9.30103 | | NA | | NR [1300000] | N |
| 9 | 17:44118848 | 17 | 44081064 | rs8070723 | 3/01/2011 | 2.1E+07 | Spencer CC | 2/11/2010 | Hum Mol Genet | www.ncbi.nlm.nih.gov/pubmed/21044948 | Dissection of the genetics of Parkinson's disease identifies an additional association 5' of SNCA and multiple associated haplotypes at 17q21. | Parkinson's disease | 1,705 Euro | 1,039 Euro | 17q21.31 | MAPT | MAPT, MAPT, MAPT | | | ENSG00000277956, E | NA | NA | rs8070723-? | rs8070723 | 0 | 8070723 | intron_variant | 0 | 0.76 | 7.00E-12 | 11.1549 | | 1.3 | [1.19-1.43 | Illumina [532616] | N |
| 9 | 17:44118848 | 17 | 44081064 | rs8070723 | 14/07/2011 | 2.2E+07 | Hoglinger GU | 19/06/2011 | Nat Genet | www.ncbi.nlm.nih.gov/pubmed/21685912 | Identification of common variants influencing risk of the tauopathy progressive supranuclear palsy. | Progressive supranuclear palsy | 1,069 Euro | 1,051 Euro | 17q21.31 | MAPT | MAPT, MAPT, MAPT | | | ENSG00000277956, E | NA | NA | rs8070723-? | rs8070723 | 0 | 8070723 | intron_variant | 0 | 0.23 | ####### | 117.699 | | 5.11 | [4.43-5.91 | Illumina [531451] | N |
| 9 | 17:44118848 | 17 | 44081064 | rs8070723 | 4/11/2019 | 3E+07 | Pulit SL | 14/09/2018 | Hum Mol Genet | www.ncbi.nlm.nih.gov/pubmed/30239722 | Meta-analysis of genome-wide association studies for body fat distribution in 694,649 individuals of European ancestry. | Waist-hip ratio | 381,152 Eu | NA | 17q21.31 | NR | MAPT, MAPT, MAPT | | | ENSG00000277956, E | NA | NA | rs8070723-A | rs8070723 | 0 | 8070723 | intron_variant | 0 | 0.7848 | 4.00E-15 | 14.39794 | | 0.0162 | [0.012-0.0 | NR [~ 27400000] (imputed) | N |
| 9 | 17:44118848 | 17 | 43893716 | rs8072451 | 3/11/2015 | 2.6E+07 | Hibar DP | 21/01/2015 | Nature | www.ncbi.nlm.nih.gov/pubmed/25607358 | Common genetic variants influence human subcortical brain structures. | Subcortical brain region volumes | up to 13,1 | up to 15,1 | 17q21.31 | MAPT, GRN, CRHI | CRHR1, CRHR1, CRHR1, LINC02210-CRHR1, LINC02210-C | | | ENSG00000276191, E | NA | NA | rs8072451-? | rs8072451 | 0 | 8072451 | intron_variant | 0 | NR | 1.00E-08 | 8 | (Intracrani | 14489.99 | [9539.28-1 | Affymetrix, Illumina [NR] (imputed) | N |
| 9 | 17:44091886 | 17 | 44162597 | rs8080583 | 12/01/2018 | 2.9E+07 | Lam M | 28/11/2017 | Cell Rep | www.ncbi.nlm.nih.gov/pubmed/29186694 | Large-Scale Cognitive GWAS Meta-Analysis Reveals Tissue-Specific Neural Expression and Potential Nootropic Drug Targets. | Cognitive ability | 107,207 Eu | NA | 17q21.31 | KANSL1 | KANSL1, KANSL1, KANSL1 | | | ENSG00000120071, E | NA | NA | rs8080583-? | rs8080583 | 0 | 8080583 | intron_variant | 0 | NR | 1.00E-08 | 8 | | 5.717 | z score dec | Affymetrix, Illumina [7357080] (imputed) | N |
| 9 | 17:44118848 | 17 | 44065740 | rs919462 | 12/01/2018 | 2.9E+07 | Pirastu N | 17/11/2017 | Nat Commun | www.ncbi.nlm.nih.gov/pubmed/29146897 | GWAS for male-pattern baldness identifies 71 susceptibility loci explaining 38% of the risk. | Male-pattern baldness | 25,662 Brit | 13,367 Eur | 17q21.31 | WNT3, MAPT, PLE | MAPT, MAPT, MAPT | | | ENSG00000277956, E | NA | NA | rs919462-C | rs919462 | 0 | 919462 | intron_variant | 0 | NR | 1.00E-26 | 26 | | 0.168393 | [0.14-0.2] | Affymetrix [27512692] (imputed) | N |
| 9 | 17:44091886 | 17 | 44187257 | rs9303525 | 1/06/2012 | 2.3E+07 | Ikram MA | 15/04/2012 | Nat Genet | www.ncbi.nlm.nih.gov/pubmed/22504418 | Common variants at 6q22 and 17q21 are associated with intracranial volume. | Intracranial volume | 8,175 Euro | 1,752 Euro | 17q21.31 | MAPT, GRN, CRHI | KANSL1, KANSL1, KANSL1 | | | ENSG00000120071, E | NA | NA | rs9303525-G | rs9303525 | 0 | 9303525 | intron_variant | 0 | 0.22 | 8.00E-15 | 14.09691 | | 14.9 | [NR] ml de | Affymetrix, Illumina [2229753] (imputed) | N |
| 9 | 17:44118848 | 17 | 44150152 | rs974295 | 29/01/2020 | 3.2E+07 | Hill WD | 16/12/2019 | Nat Commun | www.ncbi.nlm.nih.gov/pubmed/31844048 | Genome-wide analysis identifies molecular systems and 149 genetic loci associated with income. | Household income | 286,301 Br | NA | 17q21.31 | KANSL1 | KANSL1, KANSL1, KANSL1 | | | ENSG00000120071, E | NA | NA | rs974295-T | rs974295 | 0 | 974295 | intron_variant | 0 | NR | 3.00E-11 | 10.52288 | | 0.013331 | [0.0094-0. | Affymetrix [18485882] (imputed) | N |
| 9 | 17:44826056 | 17 | 44826056 | rs9896243 | 31/10/2018 | 3E+07 | Nagel M | 25/06/2018 | Nat Genet | www.ncbi.nlm.nih.gov/pubmed/29942085 | Meta-analysis of genome-wide association studies for neuroticism in 449,484 individuals identifies novel genetic loci and pathways. | Worry | 348,219 Eu | NA | 17q21.31 | NR | NSF, NSF, NSF | | | ENSG00000278174, E | NA | NA | rs9896243-G | rs9896243 | 0 | 9896243 | intron_variant | 0 | NR | 2.00E-11 | 10.69897 | | 0.01934 | [0.014-0.0 | Affymetrix [10847151] (imputed) | N |
| 9 | 17:44118848 | 17 | 43822772 | rs9906974 | 18/09/2020 | 3.3E+07 | Vuckovic D | 1/09/2020 | Cell | www.ncbi.nlm.nih.gov/pubmed/32888494 | The Polygenic and Monogenic Basis of Blood Traits and Diseases. | Eosinophil percentage of white cells | 408,112 Br | NA | 17q21.31 | RP11-105N13.4, | LINC02210-CRHR1, LINC02210-CRHR1 | | | ENSG00000282456, E | NA | NA | rs9906974-C | rs9906974 | 0 | 9906974 | intron_variant | 0 | 0.218668 | 1.00E-45 | 45 | | 0.038293 | [0.033-0.0 | Affymetrix [93095623] (imputed) | N |
| 10 | 17:62017421 | 17 | 61996255 | rs2005172 | 7/02/2020 | 3.2E+07 | Hernandez Cordero AI | 21/11/2019 | Am J Hum Genet | www.ncbi.nlm.nih.gov/pubmed/31761296 | Genome-wide Associations Reveal Human-Mouse Genetic Convergence and Modifiers of Myogenesis, CPNE1 and STC2. | Appendicular lean mass | 85,750 Eur | NA | 17q23.3 | NR | GH1 - CD79B | ENSG00000259384 | ENSG00000007312 | | 56 | 9845 | rs2005172-C | rs2005172 | 0 | 2005172 | intron_variant | 1 | 0.641109 | 2.00E-15 | 14.69897 | | 0.080328 | [0.06-0.1] | Affymetrix [21375087] (imputed) | N |
| 10 | 17:62017421 | 17 | 61996255 | rs2005172 | 4/11/2020 | 3.3E+07 | Pei YF | 23/10/2020 | Commun Biol | www.ncbi.nlm.nih.gov/pubmed/33097823 | The genetic architecture of appendicular lean mass characterized by association analysis in the UK Biobank study. | Appendicular lean mass | 450,243 Eu | NA | 17q23.3 | GH1 | GH1 - CD79B | ENSG00000259384 | ENSG00000007312 | | 56 | 9845 | rs2005172-A | rs2005172 | 0 | 2005172 | intron_variant | 1 | 0.3603 | ####### | 127.699 | | 0.048 | [0.044-0.0 | Affymetrix [18391927] (imputed) | N |
| 10 | 17:62017421 | 17 | 61996255 | rs2005172 | 8/02/2019 | 3.1E+07 | Hubel C | 28/12/2018 | Am J Med Genet B Neuropsychiatr Genet | www.ncbi.nlm.nih.gov/pubmed/30593698 | Genomics of body fat percentage may contribute to sex bias in anorexia nervosa. | Fat-free mass | 70,700 Eur | NA | 17q23.3 | NR | GH1 - CD79B | ENSG00000259384 | ENSG00000007312 | | 56 | 9845 | rs2005172-? | rs2005172 | 0 | 2005172 | intron_variant | 1 | NR | 2.00E-15 | 14.69897 | (male) | 0.29124 | [0.22-0.36 | Affymetrix [7794483] (imputed) | N |
| 10 | 17:62017421 | 17 | 61996255 | rs2005172 | 8/02/2019 | 3.1E+07 | Hubel C | 28/12/2018 | Am J Med Genet B Neuropsychiatr Genet | www.ncbi.nlm.nih.gov/pubmed/30593698 | Genomics of body fat percentage may contribute to sex bias in anorexia nervosa. | Fat-free mass | 70,700 Eur | NA | 17q23.3 | NR | GH1 - CD79B | ENSG00000259384 | ENSG00000007312 | | 56 | 9845 | rs2005172-? | rs2005172 | 0 | 2005172 | intron_variant | 1 | NR | 8.00E-14 | 13.09691 | (female) | 0.18928 | [0.14-0.24 | Affymetrix [7794483] (imputed) | N |
| 10 | 17:62017421 | 17 | 61996255 | rs2005172 | 8/02/2019 | 3.1E+07 | Hubel C | 28/12/2018 | Am J Med Genet B Neuropsychiatr Genet | www.ncbi.nlm.nih.gov/pubmed/30593698 | Genomics of body fat percentage may contribute to sex bias in anorexia nervosa. | Fat-free mass | 70,700 Eur | NA | 17q23.3 | NR | GH1 - CD79B | ENSG00000259384 | ENSG00000007312 | | 56 | 9845 | rs2005172-? | rs2005172 | 0 | 2005172 | intron_variant | 1 | NR | 3.00E-26 | 25.52288 | | 0.2464 | [0.2-0.29] | Affymetrix [7794483] (imputed) | N |
| 10 | 17:62017421 | 17 | 61996255 | rs2005172 | 14/12/2021 | 3.5E+07 | Sakaue S | 30/09/2021 | Nat Genet | www.ncbi.nlm.nih.gov/pubmed/34594039 | A cross-population atlas of genetic associations for 220 human phenotypes. | Weight | 360,116 Eu | NA | 17q23.3 | | GH1 - CD79B | ENSG00000259384 | ENSG00000007312 | | 56 | 9845 | rs2005172-C | rs2005172 | 0 | 2005172 | intron_variant | 1 | NR | 3.00E-16 | 15.52288 | | 0.014 | [0.011-0.0 | Affymetrix, Illumina [20539548] (imputed) | N |
| 10 | 17:62017421 | 17 | 62007498 | rs2070776 | 24/09/2015 | 2.5E+07 | Wood AR | 5/10/2014 | Nat Genet | www.ncbi.nlm.nih.gov/pubmed/25282103 | Defining the role of common variation in the genomic and biological architecture of adult human height. | Height | 253,288 Eu | 80,067 Eur | 17q23.3 | CD79B | CD79B | | | ENSG00000007312 | NA | NA | rs2070776-A | rs2070776 | 0 | 2070776 | stop_gained | 0 | 0.346 | 6.00E-41 | 40.22185 | | 0.042 | [0.036-0.0 | Affymetrix, Illumina, Perlegen [2550858] (imputed) | N |
| 10 | 17:62017421 | 17 | 62007498 | rs2070776 | 24/10/2012 | 2.1E+07 | Lango Allen H | 29/09/2010 | Nature | www.ncbi.nlm.nih.gov/pubmed/20881960 | Hundreds of variants clustered in genomic loci and biological pathways affect human height. | Height | 133,653 Eu | 50,074 Eur | 17q23.3 | GH region | CD79B | | | ENSG00000007312 | NA | NA | rs2070776-? | rs2070776 | 0 | 2070776 | stop_gained | 0 | NR | 9.00E-09 | 8.045757 | (Condition | NA | [NR] | Affymetrix, Illumina [2834208] (imputed) | N |
| 10 | 17:62017421 | 17 | 62007498 | rs2070776 | 2/07/2019 | 3.1E+07 | Wojcik GL | 19/06/2019 | Nature | www.ncbi.nlm.nih.gov/pubmed/31217584 | Genetic analyses of diverse populations improves discovery for complex traits. | Height | 17,286 Afri | NA | 17q23.3 | NR | CD79B | | | ENSG00000007312 | NA | NA | rs2070776-? | rs2070776 | 0 | 2070776 | stop_gained | 0 | NR | 8.00E-06 | 5.09691 | | 0.025638 | [0.014-0.0 | Illumina [34656550] (imputed) | N |
| 10 | 17:62017421 | 17 | 62007498 | rs2070776 | 5/08/2022 | 3.6E+07 | Chiou JS | 13/07/2022 | BMC Med | www.ncbi.nlm.nih.gov/pubmed/35831902 | Your height affects your health: genetic determinants and health-related outcomes in Taiwan. | Height | 67,452 Ha | NA | 17q23.3 | | CD79B | | | ENSG00000007312 | NA | NA | rs2070776-G | rs2070776 | 0 | 2070776 | stop_gained | 0 | NR | 7.00E-20 | 19.1549 | | 0.046 | [0.036-0.0 | Affymetrix [NR] (imputed) | N |
| 10 | 17:62017421 | 17 | 62008318 | rs2320125 | 5/08/2022 | 3.6E+07 | Chiou JS | 13/07/2022 | BMC Med | www.ncbi.nlm.nih.gov/pubmed/35831902 | Your height affects your health: genetic determinants and health-related outcomes in Taiwan. | Height | 67,452 Ha | NA | 17q23.3 | | CD79B | | | ENSG00000007312 | NA | NA | rs2320125-C | rs2320125 | 0 | 2320125 | intron_variant | 0 | NR | 5.00E-20 | 19.30103 | | 0.046 | [0.036-0.0 | Affymetrix [NR] (imputed) | N |
| 10 | 17:62017421 | 17 | 62008318 | rs2320125 | 22/09/2019 | 3.1E+07 | Lotta LA | 1/12/2018 | JAMA | www.ncbi.nlm.nih.gov/pubmed/30575882 | Association of Genetic Variants Related to Gluteofemoral vs Abdominal Fat Distribution With Type 2 Diabetes, Coronary Disease, and Cardiovascular Risk Factors. | Waist-hip ratio | 663,598 Eu | NA | 17q23.3 | NR | CD79B | | | ENSG00000007312 | NA | NA | rs2320125-T | rs2320125 | 0 | 2320125 | intron_variant | 0 | 0.35 | 5.00E-08 | 7.30103 | | NA | | Affymetrix [2446094] (imputed) | N |
| 10 | 17:62017421 | 17 | 62008318 | rs2320125 | 17/04/2020 | 3.2E+07 | Zhu Z | 24/10/2019 | J Allergy Clin Immunol | www.ncbi.nlm.nih.gov/pubmed/31669095 | Shared Genetic and Experimental Links between Obesity-Related Traits and Asthma Subtypes in UK Biobank. | Waist-to-hip ratio adjusted for BMI | 457,690 Eu | NA | 17q23.3 | NR | CD79B | | | ENSG00000007312 | NA | NA | rs2320125-? | rs2320125 | 0 | 2320125 | intron_variant | 0 | NR | 1.00E-08 | 8 | | NA | | Affymetrix [~ 8270000] (imputed) | N |
| 10 | 17:62017421 | 17 | 62008318 | rs2320125 | 4/11/2019 | 3E+07 | Pulit SL | 14/09/2018 | Hum Mol Genet | www.ncbi.nlm.nih.gov/pubmed/30239722 | Meta-analysis of genome-wide association studies for body fat distribution in 694,649 individuals of European ancestry. | Waist-to-hip ratio adjusted for BMI | 379,501 Eu | NA | 17q23.3 | NR | CD79B | | | ENSG00000007312 | NA | NA | rs2320125-T | rs2320125 | 0 | 2320125 | intron_variant | 0 | 0.3502 | 5.00E-12 | 11.30103 | | 0.013 | [0.0093-0. | NR [~ 27400000] (imputed) | N |
| 10 | 17:62017421 | 17 | 62008318 | rs2320125 | 22/09/2019 | 3.1E+07 | Lotta LA | 1/12/2018 | JAMA | www.ncbi.nlm.nih.gov/pubmed/30575882 | Association of Genetic Variants Related to Gluteofemoral vs Abdominal Fat Distribution With Type 2 Diabetes, Coronary Disease, and Cardiovascular Risk Factors. | Waist-to-hip ratio adjusted for BMI | 660,648 Eu | NA | 17q23.3 | NR | CD79B | | | ENSG00000007312 | NA | NA | rs2320125-T | rs2320125 | 0 | 2320125 | intron_variant | 0 | 0.35 | 2.00E-10 | 9.69897 | | 0.012 | [0.0083-0. | Affymetrix [2446094] (imputed) | N |
| 10 | 17:62017421 | 17 | 62017421 | rs2532111 | 17/07/2019 | 2.9E+07 | Tachmazidou I | 1/06/2017 | Am J Hum Genet | www.ncbi.nlm.nih.gov/pubmed/28552196 | Whole-Genome Sequencing Coupled to Imputation Discovers Genetic Signals for Anthropometric Traits. | Height | 1,249 who | 205,003 Eu | 17q23.3 | SCN4A | SCN4A | | | ENSG00000007314 | NA | NA | rs2532111-A | rs2532111 | 0 | 2532111 | 3_prime_UTR_variant | 0 | 0.3539 | 7.00E-09 | 8.154902 | | 0.0389 | [0.026-0.0 | Affymetrix, Illumina, Perlegen [15844966] (imputed) | N |
| 10 | 17:62017421 | 17 | 62017421 | rs2532111 | 24/08/2021 | 3.4E+07 | Christakoudi S | 21/05/2021 | Sci Rep | www.ncbi.nlm.nih.gov/pubmed/34021172 | GWAS of allometric body-shape indices in UK Biobank identifies loci suggesting associations with morphogenesis, organogenesis, adrenal cell renewal and cancer. | Hip circumference adjusted for BMI | 186,825 Br | NA | 17q23.3 | SCN4A | SCN4A | | | ENSG00000007314 | NA | NA | rs2532111-A | rs2532111 | 0 | 2532111 | 3_prime_UTR_variant | 0 | NR | 5.00E-14 | 13.30103 | | 0.024271 | [0.018-0.0 | Affymetrix [323123] (imputed) | N |
| 10 | 17:62017421 | 17 | 62017421 | rs2532111 | 8/02/2019 | 3.1E+07 | Kichaev G | 27/12/2018 | Am J Hum Genet | www.ncbi.nlm.nih.gov/pubmed/30595370 | Leveraging Polygenic Functional Enrichment to Improve GWAS Power. | Waist-hip ratio | approxima | NA | 17q23.3 | | SCN4A | | | ENSG00000007314 | NA | NA | rs2532111-? | rs2532111 | 0 | 2532111 | 3_prime_UTR_variant | 0 | NR | 6.00E-10 | 9.221849 | | NA | | NR [~ 8900000] (imputed) | N |
| 11 | 20:47627095 | 20 | 47648856 | rs1022688 | 21/01/2019 | 3E+07 | Day FR | 3/07/2018 | Nat Commun | www.ncbi.nlm.nih.gov/pubmed/29970889 | Elucidating the genetic basis of social interaction and isolation. | Loneliness | 80,134 Eur | NA | 20q13.13 | PREX1, ARFGEF2, | ARFGEF2 | | | ENSG00000124198 | NA | NA | rs1022688-G | rs1022688 | 0 | 1022688 | 3_prime_UTR_variant | 0 | 0.67 | 3.00E-08 | 7.522879 | | 0.0048 | [0.003-0.0 | NR [NR] (imputed) | N |
| 11 | 20:47627095 | 20 | 47648856 | rs1022688 | 21/01/2019 | 3E+07 | Day FR | 3/07/2018 | Nat Commun | www.ncbi.nlm.nih.gov/pubmed/29970889 | Elucidating the genetic basis of social interaction and isolation. | Loneliness (MTAG) | 487,647 Eu | NA | 20q13.13 | PREX1, ARFGEF2, | ARFGEF2 | | | ENSG00000124198 | NA | NA | rs1022688-G | rs1022688 | 0 | 1022688 | 3_prime_UTR_variant | 0 | 0.669916 | 3.00E-09 | 8.522879 | | 0.012942 | [0.0087-0. | NR [NR] (imputed) | N |
| 11 | 20:47746974 | 20 | 47817815 | rs13036715 | 30/11/2018 | 3E+07 | Baselmans BML | 2/10/2018 | Sci Rep | www.ncbi.nlm.nih.gov/pubmed/30279531 | A genetic perspective on the relationship between eudaimonic -and hedonic well-being. | Eudaimonic well-being | 108,154 Br | NA | 20q13.13 | NR | STAU1 - DDX27 | ENSG00000124214 | ENSG00000124228 | | 12911 | 18069 | rs13036715-A | rs13036715 | 0 | 13036715 | intergenic_variant | 1 | 0.241067 | 3.00E-07 | 6.522879 | | 5.11717 | [NR] z scor | Affymetrix [~ 8600000] (imputed) | N |
| 11 | 20:47746974 | 20 | 47849216 | rs13038866 | 17/04/2020 | 3.2E+07 | Zhu Z | 24/10/2019 | J Allergy Clin Immunol | www.ncbi.nlm.nih.gov/pubmed/31669095 | Shared Genetic and Experimental Links between Obesity-Related Traits and Asthma Subtypes in UK Biobank. | Waist circumference adjusted for body mass index | 457,690 Eu | NA | 20q13.13 | NR | DDX27 | | | ENSG00000124228 | NA | NA | rs13038866-? | rs13038866 | 0 | 13038866 | intron_variant | 0 | NR | 3.00E-12 | 11.52288 | | NA | | Affymetrix [~ 8270000] (imputed) | N |
| 11 | 20:47746974 | 20 | 47800829 | rs13041213 | 23/02/2018 | 2.9E+07 | Hill WD | 11/01/2018 | Mol Psychiatry | www.ncbi.nlm.nih.gov/pubmed/29326435 | A combined analysis of genetically correlated traits identifies 187 loci and a role for neurogenesis and myelination in intelligence. | Intelligence (MTAG) | 120,934 Br | NA | 20q13.13 | STAU1 | STAU1 | | | ENSG00000124214 | NA | NA | rs13041213-A | rs13041213 | 0 | 13041213 | intron_variant | 0 | NR | 7.00E-12 | 11.1549 | | 0.024944 | [0.018-0.0 | Affymetrix [NR] | N |
| 11 | 20:47627095 | 20 | 47705496 | rs1538482 | 8/02/2019 | 3.1E+07 | Hubel C | 28/12/2018 | Am J Med Genet B Neuropsychiatr Genet | www.ncbi.nlm.nih.gov/pubmed/30593698 | Genomics of body fat percentage may contribute to sex bias in anorexia nervosa. | Body fat percentage | 70,700 Eur | NA | 20q13.13 | NR | CSE1L | | | ENSG00000124207 | NA | NA | rs1538482-? | rs1538482 | 0 | 1538482 | intron_variant | 0 | NR | 3.00E-10 | 9.522879 | | 0.14468 | [0.1-0.19] | Affymetrix [7794483] (imputed) | N |
| 11 | 20:47627095 | 20 | 47705496 | rs1538482 | 8/02/2019 | 3.1E+07 | Hubel C | 28/12/2018 | Am J Med Genet B Neuropsychiatr Genet | www.ncbi.nlm.nih.gov/pubmed/30593698 | Genomics of body fat percentage may contribute to sex bias in anorexia nervosa. | Body fat percentage | 70,700 Eur | NA | 20q13.13 | NR | CSE1L | | | ENSG00000124207 | NA | NA | rs1538482-? | rs1538482 | 0 | 1538482 | intron_variant | 0 | NR | 8.00E-06 | 5.09691 | (male) | 0.12586 | [0.071-0.1 | Affymetrix [7794483] (imputed) | N |
| 11 | 20:47627095 | 20 | 47705496 | rs1538482 | 17/03/2017 | 2.7E+07 | Okbay A | 18/04/2016 | Nat Genet | www.ncbi.nlm.nih.gov/pubmed/27089181 | Genetic variants associated with subjective well-being, depressive symptoms, and neuroticism identified through genome-wide analyses. | Positive affect | 180,281 Ko | NA | 20q13.13 | NR | CSE1L | | | ENSG00000124207 | NA | NA | rs1538482-T | rs1538482 | 0 | 1538482 | intron_variant | 0 | 0.6958 | 1.00E-06 | 6 | | 0.0177 | [0.011-0.0 | Affymetrix, Illumina, Perlegen [at least 5170349] (imputed) | N |
| 11 | 20:47627095 | 20 | 47705496 | rs1538482 | 24/09/2021 | 3.3E+07 | Wendt FR | 21/12/2020 | Nat Hum Behav | www.ncbi.nlm.nih.gov/pubmed/33349686 | Multivariate genome-wide analysis of education, socioeconomic status and brain phenome. | Subjective wellbeing conditioned on cognitive performance (multi-trait conditioning and joint analysis) | 298,420 Eu | NA | 20q13.13 | | CSE1L | | | ENSG00000124207 | NA | NA | rs1538482-? | rs1538482 | 0 | 1538482 | intron_variant | 0 | NR | 9.00E-11 | 10.04576 | | NA | | NR [NR] | N |

| | | | | | | | | | | | | | | | | | | | | | | | | | | | | | | | | | | | | |
|---|---|---|---|---|---|---|---|---|---|---|---|---|---|---|---|---|---|---|---|---|---|---|---|---|---|---|---|---|---|---|---|---|---|---|---|---|
| 11 | 20:47627095 | 20 | 47603241 | rs1569750 | 17/11/2022 | 3.5E+07 | Graham SE | 9/12/2021 | Nature | www.ncbi.nlm.nih.gov/pubmed/34887591 | The power of genetic diversity in genome-wide association studies of lipids. | Non-HDL cholesterol levels | 1,320,016 | NA | 20q13.13 | | ARFGEF2 | | | ENSG00000124198 | NA | NA | rs1569750-G | rs1569750 | 0 | 1569750 | intron_variant | 0 | 0.303776 | 5.00E-10 | 9.30103 | | 0.012993 | [0.0095-0. | NR [52000000] (imputed) | N |
| 11 | 20:47746974 | 20 | 47772264 | rs17450430 | 24/09/2015 | 2.5E+07 | Wood AR | 5/10/2014 | Nat Genet | www.ncbi.nlm.nih.gov/pubmed/25282103 | Defining the role of common variation in the genomic and biological architecture of adult human height. | Height | 253,288 Eu | 80,067 Eur | 20q13.13 | STAU1 | STAU1 | | | ENSG00000124214 | NA | NA | rs17450430-A | rs17450430 | 0 | 17450430 | intron_variant | 0 | 0.761 | 2.00E-24 | 23.69897 | | 0.035 | [0.029-0.0 | Affymetrix, Illumina, Perlegen [2550858] (imputed) | N |
| 11 | 20:47627095 | 20 | 47701765 | rs1885163 | 17/11/2022 | 3.5E+07 | Graham SE | 9/12/2021 | Nature | www.ncbi.nlm.nih.gov/pubmed/34887591 | The power of genetic diversity in genome-wide association studies of lipids. | Non-HDL cholesterol levels | 40,963 Sou | NA | 20q13.13 | | CSE1L | | | ENSG00000124207 | NA | NA | rs1885163-? | rs1885163 | 0 | 1885163 | intron_variant | 0 | NR | 1.00E-11 | 11 | | NA | | NR [52000000] (imputed) | N |
| 11 | 20:47746974 | 20 | 47552467 | rs189331311 | 24/08/2021 | 3.4E+07 | Christakoudi S | 21/05/2021 | Sci Rep | www.ncbi.nlm.nih.gov/pubmed/34021172 | GWAS of allometric body-shape indices in UK Biobank identifies loci suggesting associations with morphogenesis, organogenesis, adrenal cell renewal and cancer. | Waist circumference adjusted for body mass index | 186,825 Br | NA | 20q13.13 | ARFGEF2 | ARFGEF2 | | | ENSG00000124198 | NA | NA | rs189331311-C | rs189331311 | 0 | 1.89E+08 | intron_variant | 0 | NR | 5.00E-08 | 7.30103 | | 0.019805 | [0.013-0.0 | Affymetrix [321114] (imputed) | N |
| 11 | 20:47746974 | 20 | 47701024 | rs2075677 | 18/03/2019 | 3.1E+07 | Baselmans BML | 14/01/2019 | Nat Genet | www.ncbi.nlm.nih.gov/pubmed/30643256 | Multivariate genome-wide analyses of the well-being spectrum. | Positive affect | 410,603 Eu | NA | 20q13.13 | NR | CSE1L | | | ENSG00000124207 | NA | NA | rs2075677-G | rs2075677 | 0 | 2075677 | intron_variant | 0 | 0.238398 | 2.00E-14 | 13.69897 | | 0.018965 | [0.014-0.0 | NR [NR] | N |
| 11 | 20:47746974 | 20 | 47701024 | rs2075677 | 17/03/2017 | 2.7E+07 | Okbay A | 18/04/2016 | Nat Genet | www.ncbi.nlm.nih.gov/pubmed/27089181 | Genetic variants associated with subjective well-being, depressive symptoms, and neuroticism identified through genome-wide analyses. | Subjective well-being | 298,420 Ko | NA | 20q13.13 | CSE1L | CSE1L | | | ENSG00000124207 | NA | NA | rs2075677-A | rs2075677 | 0 | 2075677 | intron_variant | 0 | 0.7603 | 2.00E-08 | 7.69897 | | 0.0175 | [0.011-0.0 | Affymetrix, Illumina, Perlegen [at least 5170349] (imputed) | N |
| 11 | 20:47746974 | 20 | 47701024 | rs2075677 | 7/07/2022 | 3.6E+07 | Kim S | 19/05/2022 | Nat Hum Behav | www.ncbi.nlm.nih.gov/pubmed/35589828 | Shared genetic architectures of subjective well-being in East Asian and European ancestry populations. | Subjective well-being | 110,919 Ko | NA | 20q13.13 | | CSE1L | | | ENSG00000124207 | NA | NA | rs2075677-A | rs2075677 | 0 | 2075677 | intron_variant | 0 | NR | 4.00E-10 | 9.39794 | | 0.013317 | unit increa | NR [NR] (imputed) | N |
| 11 | 20:47627095 | 20 | 47582884 | rs2295027 | 17/11/2022 | 3.5E+07 | Graham SE | 9/12/2021 | Nature | www.ncbi.nlm.nih.gov/pubmed/34887591 | The power of genetic diversity in genome-wide association studies of lipids. | Low density lipoprotein cholesterol levels | 40,963 Sou | NA | 20q13.13 | | ARFGEF2 | | | ENSG00000124198 | NA | NA | rs2295027-? | rs2295027 | 0 | 2295027 | intron_variant | 0 | NR | 1.00E-12 | 12 | | NA | | NR [52000000] (imputed) | N |
| 11 | 20:47627095 | 20 | 47582884 | rs2295027 | 17/11/2022 | 3.5E+07 | Graham SE | 9/12/2021 | Nature | www.ncbi.nlm.nih.gov/pubmed/34887591 | The power of genetic diversity in genome-wide association studies of lipids. | Low density lipoprotein cholesterol levels | 1,320,016 | NA | 20q13.13 | | ARFGEF2 | | | ENSG00000124198 | NA | NA | rs2295027-A | rs2295027 | 0 | 2295027 | intron_variant | 0 | 0.322025 | 6.00E-11 | 10.22185 | | 0.011545 | [0.0086-0. | NR [52000000] (imputed) | N |
| 11 | 20:47627095 | 20 | 47582884 | rs2295027 | 17/11/2022 | 3.5E+07 | Graham SE | 9/12/2021 | Nature | www.ncbi.nlm.nih.gov/pubmed/34887591 | The power of genetic diversity in genome-wide association studies of lipids. | Total cholesterol levels | 1,320,016 | NA | 20q13.13 | | ARFGEF2 | | | ENSG00000124198 | NA | NA | rs2295027-A | rs2295027 | 0 | 2295027 | intron_variant | 0 | 0.321731 | 3.00E-11 | 10.52288 | | 0.011737 | [0.0089-0. | NR [52000000] (imputed) | N |
| 11 | 20:47746974 | 20 | 47903019 | rs237743 | 24/10/2012 | 2.1E+07 | Lango Allen H | 29/09/2010 | Nature | www.ncbi.nlm.nih.gov/pubmed/20881960 | Hundreds of variants clustered in genomic loci and biological pathways affect human height. | Height | 133,653 Eu | 50,074 Eur | 20q13.13 | ZNFX1 | ZFAS1 | | | ENSG00000177410 | NA | NA | rs237743-A | rs237743 | 0 | 237743 | intron_variant | 0 | 0.21 | 1.00E-20 | 20 | | 0.041 | [NR] unit in | Affymetrix, Illumina [2834208] (imputed) | N |
| 11 | 20:47746974 | 20 | 47851919 | rs238150 | 7/07/2022 | 3.6E+07 | Kim S | 19/05/2022 | Nat Hum Behav | www.ncbi.nlm.nih.gov/pubmed/35589828 | Shared genetic architectures of subjective well-being in East Asian and European ancestry populations. | Subjective well-being | 110,919 Ko | NA | 20q13.13 | | DDX27 | | | ENSG00000124228 | NA | NA | rs238150-T | rs238150 | 0 | 238150 | non_coding_transcript_exon | 0 | 0.81 | 3.00E-11 | 10.52288 | | 0.015 | [0.011-0.0 | NR [NR] (imputed) | N |
| 11 | 20:47746974 | 20 | 47851919 | rs238150 | 7/07/2022 | 3.6E+07 | Kim S | 19/05/2022 | Nat Hum Behav | www.ncbi.nlm.nih.gov/pubmed/35589828 | Shared genetic architectures of subjective well-being in East Asian and European ancestry populations. | Subjective well-being | 563,176 Eu | NA | 20q13.13 | | DDX27 | | | ENSG00000124228 | NA | NA | rs238150-T | rs238150 | 0 | 238150 | non_coding_transcript_exon | 0 | 0.784 | 5.00E-12 | 11.30103 | | 0.016 | [0.012-0.0 | NR [NR] (imputed) | N |
| 11 | 20:47746974 | 20 | 47844341 | rs238164 | 16/10/2019 | 3.2E+07 | Akiyama M | 27/09/2019 | Nat Commun | www.ncbi.nlm.nih.gov/pubmed/31562340 | Characterizing rare and low-frequency height-associated variants in the Japanese population. | Height | 159,095 Ja | 32,692 Jap | 20q13.13 | DDX27 | DDX27 | | | ENSG00000124228 | NA | NA | rs238164-? | rs238164 | 0 | 238164 | intron_variant | 0 | NR | 2.00E-09 | 8.69897 | | 0.029782 | [0.02-0.03 | Illumina [27896057] (imputed) | N |
| 11 | 20:47627095 | 20 | 47780105 | rs2426139 | 15/12/2021 | 3.5E+07 | Shadrin AA | 21/09/2021 | Neuroimage | www.ncbi.nlm.nih.gov/pubmed/34560273 | Vertex-wise multivariate genome-wide association study identifies 780 unique genetic loci associated with cortical morphology. | Cortical thickness | 35,657 Wh | NA | 20q13.13 | NR | STAU1 | | | ENSG00000124214 | NA | NA | rs2426139-A | rs2426139 | 0 | 2426139 | intron_variant | 0 | 0.3008 | 5.00E-08 | 7.30103 | | NA | | NR [9056863] (imputed) | N |
| 11 | 20:47495794 | 20 | 47493894 | rs34116853 | 12/11/2018 | 3E+07 | Lee JJ | 23/07/2018 | Nat Genet | www.ncbi.nlm.nih.gov/pubmed/30038396 | Gene discovery and polygenic prediction from a genome-wide association study of educational attainment in 1.1 million individuals. | Highest math class taken (MTAG) | 811,539 Eu | NA | 20q13.13 | Intergenic | PREX1 - ARFGEF2 | ENSG00000124126 | ENSG00000124198 | | 49358 | 44354 | rs34116853-A | rs34116853 | 0 | 34116853 | regulatory_region_variant | 1 | 0.6796 | 1.00E-12 | 12 | (MTAG) | 0.0125 | [0.009-0.0 | Affymetrix, Illumina [7100000] (imputed) | N |
| 11 | 20:47495794 | 20 | 47493894 | rs34116853 | 12/11/2018 | 3E+07 | Lee JJ | 23/07/2018 | Nat Genet | www.ncbi.nlm.nih.gov/pubmed/30038396 | Gene discovery and polygenic prediction from a genome-wide association study of educational attainment in 1.1 million individuals. | Self-reported math ability | 564,698 Eu | NA | 20q13.13 | Intergenic | PREX1 - ARFGEF2 | ENSG00000124126 | ENSG00000124198 | | 49358 | 44354 | rs34116853-A | rs34116853 | 0 | 34116853 | regulatory_region_variant | 1 | 0.6805 | 1.00E-09 | 9 | | 0.0127 | [0.0086-0. | Affymetrix, Illumina [10000000] (imputed) | N |
| 11 | 20:47495794 | 20 | 47493894 | rs34116853 | 12/11/2018 | 3E+07 | Lee JJ | 23/07/2018 | Nat Genet | www.ncbi.nlm.nih.gov/pubmed/30038396 | Gene discovery and polygenic prediction from a genome-wide association study of educational attainment in 1.1 million individuals. | Self-reported math ability (MTAG) | 670,471 Eu | NA | 20q13.13 | Intergenic | PREX1 - ARFGEF2 | ENSG00000124126 | ENSG00000124198 | | 49358 | 44354 | rs34116853-A | rs34116853 | 0 | 34116853 | regulatory_region_variant | 1 | 0.6805 | 9.00E-14 | 13.04576 | (MTAG) | 0.0143 | [0.011-0.0 | Affymetrix, Illumina [7100000] (imputed) | N |
| 11 | 20:47627095 | 20 | 47768988 | rs348258 | 21/01/2020 | 3.2E+07 | Abdellaoui A | 13/09/2019 | Hum Mol Genet | www.ncbi.nlm.nih.gov/pubmed/31518406 | Phenome-wide investigation of health outcomes associated with genetic predisposition to loneliness. | Loneliness | 511,280 Eu | NA | 20q13.13 | STAU1 | STAU1 | | | ENSG00000124214 | NA | NA | rs348258-? | rs348258 | 0 | 348258 | intron_variant | 0 | NR | 1.00E-08 | 8 | | 0.02 | [0.014-0.0 | Affymetrix, Illumina [up to 14113458] (imputed) | N |
| 11 | 20:47627095 | 20 | 47786190 | rs348276 | 12/01/2018 | 2.9E+07 | Lam M | 28/11/2017 | Cell Rep | www.ncbi.nlm.nih.gov/pubmed/29186694 | Large-Scale Cognitive GWAS Meta-Analysis Reveals Tissue-Specific Neural Expression and Potential Nootropic Drug Targets. | Cognitive ability | 107,207 Eu | NA | 20q13.13 | STAU1 | STAU1 | | | ENSG00000124214 | NA | NA | rs348276-? | rs348276 | 0 | 348276 | intron_variant | 0 | NR | 1.00E-08 | 8 | | 5.699 | z score inc | Affymetrix, Illumina [7357080] (imputed) | N |
| 11 | 20:47627095 | 20 | 47786190 | rs348276 | 4/09/2019 | 3.1E+07 | Lam M | 1/08/2019 | Am J Hum Genet | www.ncbi.nlm.nih.gov/pubmed/31374203 | Pleiotropic Meta-Analysis of Cognition, Education, and Schizophrenia Differentiates Roles of Early Neurodevelopmental and Adult Synaptic Pathways. | Cognitive ability, years of educational attainment or schizophrenia (pleiotropy) | 328,917 in | NA | 20q13.13 | STAU1 | STAU1 | | | ENSG00000124214 | NA | NA | rs348276-? | rs348276 | 0 | 348276 | intron_variant | 0 | NR | 5.00E-09 | 8.30103 | | 0.012521 | [0.0083-0. | NR [7306098] | N |
| 11 | 20:47627095 | 20 | 47786190 | rs348276 | 24/09/2021 | 3.3E+07 | Wendt FR | 21/12/2020 | Nat Hum Behav | www.ncbi.nlm.nih.gov/pubmed/33349686 | Multivariate genome-wide analysis of education, socioeconomic status and brain phenome. | Subjective wellbeing conditioned on average household income before tax (multi-trait conditioning and joint analysis) | 298,420 Eu | NA | 20q13.13 | | STAU1 | | | ENSG00000124214 | NA | NA | rs348276-? | rs348276 | 0 | 348276 | intron_variant | 0 | NR | 9.00E-10 | 9.045757 | | NA | | NR [NR] | N |
| 11 | 20:47627095 | 20 | 47786190 | rs348276 | 24/09/2021 | 3.3E+07 | Wendt FR | 21/12/2020 | Nat Hum Behav | www.ncbi.nlm.nih.gov/pubmed/33349686 | Multivariate genome-wide analysis of education, socioeconomic status and brain phenome. | Subjective wellbeing conditioned on Townsend deprivation index (multi-trait conditioning and joint analysis) | 298,420 Eu | NA | 20q13.13 | | STAU1 | | | ENSG00000124214 | NA | NA | rs348276-? | rs348276 | 0 | 348276 | intron_variant | 0 | NR | 2.00E-10 | 9.69897 | | NA | | NR [NR] | N |
| 11 | 20:47746974 | 20 | 47746974 | rs34841991 | 30/11/2018 | 3E+07 | Baselmans BML | 2/10/2018 | Sci Rep | www.ncbi.nlm.nih.gov/pubmed/30279531 | A genetic perspective on the relationship between eudaimonic -and hedonic well-being. | Hedonic well-being | 221,575 Bi | NA | 20q13.13 | NR | STAU1 | | | ENSG00000124214 | NA | NA | rs34841991-C | rs34841991 | 0 | 34841991 | intron_variant | 0 | 0.24 | 2.00E-10 | 9.69897 | | 0.022 | [0.014-0.0 | Affymetrix [~ 8600000] (imputed) | N |
| 11 | 20:47746974 | 20 | 47806092 | rs3795075 | 14/12/2021 | 3.5E+07 | Sakaue S | 30/09/2021 | Nat Genet | www.ncbi.nlm.nih.gov/pubmed/34594039 | A cross-population atlas of genetic associations for 220 human phenotypes. | Height | 165,056 Ea | NA | 20q13.13 | | STAU1 - DDX27 | ENSG00000124214 | ENSG00000124228 | | 1188 | 29792 | rs3795075-A | rs3795075 | 0 | 3795075 | intergenic_variant | 1 | 0.089743 | 1.00E-08 | 8 | | 0.020327 | [0.013-0.0 | Illumina [13479295] (imputed) | N |
| 11 | 20:47495794 | 20 | 47440220 | rs4809728 | 18/09/2020 | 3.3E+07 | Vuckovic D | 1/09/2020 | Cell | www.ncbi.nlm.nih.gov/pubmed/32888494 | The Polygenic and Monogenic Basis of Blood Traits and Diseases. | Mean reticulocyte volume | 408,112 Bi | NA | 20q13.13 | PREX1 | PREX1 | | | ENSG00000124126 | NA | NA | rs4809728-G | rs4809728 | 0 | 4809728 | intron_variant | 0 | 0.341007 | 7.00E-12 | 11.1549 | | 0.016283 | [0.012-0.0 | Affymetrix [93095623] (imputed) | N |
| 11 | 20:47495794 | 20 | 47495794 | rs4810883 | 17/09/2018 | 3E+07 | Davies G | 29/05/2018 | Nat Commun | www.ncbi.nlm.nih.gov/pubmed/29844566 | Study of 300,486 individuals identifies 148 independent genetic loci influencing general cognitive function. | General cognitive ability | up to 300,4 | NA | 20q13.13 | ARFGEF2 | PREX1 - ARFGEF2 | ENSG00000124126 | ENSG00000124198 | | 51258 | 42454 | rs4810883-T | rs4810883 | 0 | 4810883 | regulatory_region_variant | 1 | NR | 1.00E-08 | 8 | | 5.69 | z-score inc | Affymetrix, Illumina [at least 2500000] (imputed) | N |
| 11 | 20:47495794 | 20 | 47495794 | rs4810883 | 21/09/2020 | 3.3E+07 | Chen MH | 1/09/2020 | Cell | www.ncbi.nlm.nih.gov/pubmed/32888493 | Trans-ethnic and Ancestry-Specific Blood-Cell Genetics in 746,667 Individuals from 5 Global Populations. | Red blood cell count | 545,203 Eu | NA | 20q13.13 | NR | PREX1 - ARFGEF2 | ENSG00000124126 | ENSG00000124198 | | 51258 | 42454 | rs4810883-T | rs4810883 | 0 | 4810883 | regulatory_region_variant | 1 | 0.329032 | 3.00E-13 | 12.52288 | | 0.014533 | [0.011-0.0 | Affymetrix, Illumina [50628390] (imputed) | N |
| 11 | 20:47495794 | 20 | 47495794 | rs4810883 | 21/09/2020 | 3.3E+07 | Chen MH | 1/09/2020 | Cell | www.ncbi.nlm.nih.gov/pubmed/32888493 | Trans-ethnic and Ancestry-Specific Blood-Cell Genetics in 746,667 Individuals from 5 Global Populations. | Red blood cell count | 727,624 Af | NA | 20q13.13 | NR | PREX1 - ARFGEF2 | ENSG00000124126 | ENSG00000124198 | | 51258 | 42454 | rs4810883-T | rs4810883 | 0 | 4810883 | regulatory_region_variant | 1 | 0.302151 | 1.00E-11 | 11 | | NA | | Affymetrix, Illumina [24332387] (imputed) | N |
| 11 | 20:47495794 | 20 | 47495794 | rs4810883 | 18/09/2020 | 3.3E+07 | Vuckovic D | 1/09/2020 | Cell | www.ncbi.nlm.nih.gov/pubmed/32888494 | The Polygenic and Monogenic Basis of Blood Traits and Diseases. | Red blood cell count | 408,112 Bi | NA | 20q13.13 | | PREX1 - ARFGEF2 | ENSG00000124126 | ENSG00000124198 | | 51258 | 42454 | rs4810883-T | rs4810883 | 0 | 4810883 | regulatory_region_variant | 1 | 0.329974 | 3.00E-10 | 9.522879 | | 0.014927 | [0.01-0.02 | Affymetrix [93095623] (imputed) | N |
| 11 | 20:47495794 | 20 | 47477706 | rs6012542 | 23/02/2018 | 2.9E+07 | Hill WD | 11/01/2018 | Mol Psychiatry | www.ncbi.nlm.nih.gov/pubmed/29326435 | A combined analysis of genetically correlated traits identifies 187 loci and a role for neurogenesis and myelination in intelligence. | Intelligence (MTAG) | 120,934 Bi | NA | 20q13.13 | PREX1 | PREX1 - ARFGEF2 | ENSG00000124126 | ENSG00000124198 | | 33170 | 60542 | rs6012542-T | rs6012542 | 0 | 6012542 | regulatory_region_variant | 1 | NR | 8.00E-12 | 11.09691 | | 0.021212 | [0.015-0.0 | Affymetrix [NR] | N |
| 11 | 20:47627095 | 20 | 47541517 | rs6019535 | 5/09/2022 | 3.6E+07 | Rao S | 28/06/2022 | Neuropsychobiology | www.ncbi.nlm.nih.gov/pubmed/35764056 | Genetic Relationships between Attention-Deficit/Hyperactivity Disorder, Autism Spectrum Disorder, and Intelligence. | Attention deficit hyperactivity disorder or autism spectrum disorder or intelligence (pleiotropy) | 20,183 Eur | NA | 20q13.13 | | ARFGEF2 | | | ENSG00000124198 | NA | NA | rs6019535-? | rs6019535 | 0 | 6019535 | intron_variant | 0 | NR | 6.00E-20 | 19.22185 | | NA | | NR [6307410] (imputed) | N |
| 11 | 20:47627095 | 20 | 47541517 | rs6019535 | 12/09/2018 | 3E+07 | Savage JE | 25/06/2018 | Nat Genet | www.ncbi.nlm.nih.gov/pubmed/29942086 | Genome-wide association meta-analysis in 269,867 individuals identifies new genetic and functional links to intelligence. | Intelligence | 269,867 Eu | NA | 20q13.13 | NR | ARFGEF2 | | | ENSG00000124198 | NA | NA | rs6019535-A | rs6019535 | 0 | 6019535 | intron_variant | 0 | NR | 3.00E-17 | 16.52288 | | 8.436 | z-score inc | Affymetrix, Illumina [9295118] (imputed) | N |
| 11 | 20:47627095 | 20 | 47541517 | rs6019535 | 17/11/2022 | 3.5E+07 | Graham SE | 9/12/2021 | Nature | www.ncbi.nlm.nih.gov/pubmed/34887591 | The power of genetic diversity in genome-wide association studies of lipids. | Total cholesterol levels | 40,963 Sou | NA | 20q13.13 | | ARFGEF2 | | | ENSG00000124198 | NA | NA | rs6019535-? | rs6019535 | 0 | 6019535 | intron_variant | 0 | NR | 8.00E-15 | 14.09691 | | NA | | NR [52000000] (imputed) | N |
| 11 | 20:47627095 | 20 | 47685815 | rs6019624 | 17/09/2018 | 3E+07 | Davies G | 29/05/2018 | Nat Commun | www.ncbi.nlm.nih.gov/pubmed/29844566 | Study of 300,486 individuals identifies 148 independent genetic loci influencing general cognitive function. | Reaction time | 330,069 Eu | NA | 20q13.13 | NR | CSE1L | | | ENSG00000124207 | NA | NA | rs6019624-? | rs6019624 | 0 | 6019624 | intron_variant | 0 | NR | 1.00E-06 | 6 | | 0.008534 | [0.0051-0. | Affymetrix [805426] (imputed) | N |
| 11 | 20:47627095 | 20 | 47666027 | rs6090940 | 18/03/2019 | 3.1E+07 | Baselmans BML | 14/01/2019 | Nat Genet | www.ncbi.nlm.nih.gov/pubmed/30643256 | Multivariate genome-wide analyses of the well-being spectrum. | Life satisfaction | 80,852 Eur | NA | 20q13.13 | NR | CSE1L | | | ENSG00000124207 | NA | NA | rs6090940-T | rs6090940 | 0 | 6090940 | intron_variant | 0 | 0.322627 | 3.00E-08 | 7.522879 | | 0.017017 | [0.011-0.0 | NR [NR] | N |
| 11 | 20:47495794 | 20 | 47446556 | rs6095319 | 26/10/2022 | 3.6E+07 | Hatoum AS | 14/07/2022 | Biol Psychiatry | www.ncbi.nlm.nih.gov/pubmed/36150907 | Genome-wide Association Study Shows That Executive Functioning Is Influenced by GABAergic Processes and Is a Neurocognitive Genetic Correlate of Psychiatric Disorders. | Common executive function | 427,037 Eu | NA | 20q13.13 | | PREX1 - ARFGEF2 | ENSG00000124126 | ENSG00000124198 | | 2020 | 91692 | rs6095319-? | rs6095319 | 0 | 6095319 | intron_variant | 1 | NR | 2.00E-14 | 13.69897 | | NA | | Affymetrix [7391068] (imputed) | N |
| 11 | 20:47627095 | 20 | 47532536 | rs6095360 | 30/11/2018 | 3E+07 | Baselmans BML | 2/10/2018 | Sci Rep | www.ncbi.nlm.nih.gov/pubmed/30279531 | A genetic perspective on the relationship between eudaimonic -and hedonic well-being. | Eudaimonic well-being | 108,154 Bi | NA | 20q13.13 | NR | PREX1 - ARFGEF2 | ENSG00000124126 | ENSG00000124198 | | 88000 | 5712 | rs6095360-G | rs6095360 | 0 | 6095360 | intergenic_variant | 1 | 0.324609 | 9.00E-07 | 6.045757 | | 4.91741 | [NR] z scor | Affymetrix [~ 8600000] (imputed) | N |
| 11 | 20:47627095 | 20 | 47532536 | rs6095360 | 6/02/2019 | 3E+07 | Coleman JRI | 8/03/2018 | Mol Psychiatry | www.ncbi.nlm.nih.gov/pubmed/29520040 | Biological annotation of genetic loci associated with intelligence in a meta-analysis of 87,740 individuals. | Extremely high intelligence | 1,247 Euro | NA | 20q13.13 | ARFGEF2, CSE1L | PREX1 - ARFGEF2 | ENSG00000124126 | ENSG00000124198 | | 88000 | 5712 | rs6095360-A | rs6095360 | 0 | 6095360 | intergenic_variant | 1 | 0.693 | 7.00E-10 | 9.154902 | | 6.16 | NR z-score | Illumina [up to 12595966] (imputed) | N |
| 11 | 20:47627095 | 20 | 47532536 | rs6095360 | 23/02/2018 | 2.9E+07 | Hill WD | 11/01/2018 | Mol Psychiatry | www.ncbi.nlm.nih.gov/pubmed/29326435 | A combined analysis of genetically correlated traits identifies 187 loci and a role for neurogenesis and myelination in intelligence. | Intelligence (MTAG) | 120,934 Bi | NA | 20q13.13 | ARFGEF2 | PREX1 - ARFGEF2 | ENSG00000124126 | ENSG00000124198 | | 88000 | 5712 | rs6095360-A | rs6095360 | 0 | 6095360 | intergenic_variant | 1 | NR | 7.00E-16 | 15.1549 | | 0.025381 | [0.019-0.0 | Affymetrix [NR] | N |
| 11 | 20:47627095 | 20 | 47627095 | rs6095395 | 7/03/2022 | 3.5E+07 | van der Meer D | 15/12/2021 | Sci Adv | www.ncbi.nlm.nih.gov/pubmed/34910505 | The genetic architecture of human cortical folding. | Vertex-wise cortical thickness | 33,748 Eur | NA | 20q13.13 | | ARFGEF2 | | | ENSG00000124198 | NA | NA | rs6095395-G | rs6095395 | 0 | 6095395 | intron_variant | 0 | 0.3 | 1.00E-09 | 9 | | 6.05 | z score inc | Affymetrix [9061022] (imputed) | N |
| 11 | 20:47627095 | 20 | 47678201 | rs6095417 | 17/09/2018 | 3E+07 | Davies G | 29/05/2018 | Nat Commun | www.ncbi.nlm.nih.gov/pubmed/29844566 | Study of 300,486 individuals identifies 148 independent genetic loci influencing general cognitive function. | General cognitive ability | up to 300,4 | NA | 20q13.13 | CSE1L | CSE1L | | | ENSG00000124207 | NA | NA | rs6095417-A | rs6095417 | 0 | 6095417 | intron_variant | 0 | NR | 1.00E-14 | 14 | | 7.724 | z-score inc | Affymetrix, Illumina [at least 2500000] (imputed) | N |
| 11 | 20:47495794 | 20 | 47481726 | rs67021138 | 12/11/2018 | 3E+07 | Lee JJ | 23/07/2018 | Nat Genet | www.ncbi.nlm.nih.gov/pubmed/30038396 | Gene discovery and polygenic prediction from a genome-wide association study of educational attainment in 1.1 million individuals. | Educational attainment (MTAG) | 1,311,438 | NA | 20q13.13 | Intergenic | PREX1 - ARFGEF2 | ENSG00000124126 | ENSG00000124198 | | 37190 | 56522 | rs67021138-T | rs67021138 | 0 | 67021138 | TF_binding_site_variant | 1 | 0.6814 | 9.00E-10 | 9.045757 | (MTAG) | 0.0086 | [0.0059-0. | Affymetrix, Illumina [7100000] (imputed) | N |

| | | | | | | | | | | | | | | | | | | | | | | | | | | | | | | | | | | | | |
|---|---|---|---|---|---|---|---|---|---|---|---|---|---|---|---|---|---|---|---|---|---|---|---|---|---|---|---|---|---|---|---|---|---|---|---|---|
| 11 | 20:47627095 | 20 | 47652254 | rs707533 | 24/09/2021 | 3.3E+07 | Wendt FR | 21/12/2020 | Nat Hum Behav | www.ncbi.nlm.nih.gov/pubmed/33349686 | Multivariate genome-wide analysis of education, socioeconomic status and brain phenome. | Subjective wellbeing conditioned on educational attainment (multi-trait conditioning and joint analysis) | 298,420 Eu | NA | 20q13.13 | | ARFGEF2 | | | ENSG00000124198 | NA | NA | rs707533-? | rs707533 | 0 | 707533 | 3_prime_UTR_variant | 0 | NR | 9.00E-11 | 10.04576 | | NA | | NR [NR] | N |
| 11 | 20:47627095 | 20 | 47652254 | rs707533 | 24/09/2021 | 3.3E+07 | Wendt FR | 21/12/2020 | Nat Hum Behav | www.ncbi.nlm.nih.gov/pubmed/33349686 | Multivariate genome-wide analysis of education, socioeconomic status and brain phenome. | Subjective wellbeing conditioned on highest math class (multi-trait conditioning and joint analysis) | 298,420 Eu | NA | 20q13.13 | | ARFGEF2 | | | ENSG00000124198 | NA | NA | rs707533-? | rs707533 | 0 | 707533 | 3_prime_UTR_variant | 0 | NR | 1.00E-10 | 10 | | NA | | NR [NR] | N |
| 11 | 20:47627095 | 20 | 47652254 | rs707533 | 24/09/2021 | 3.3E+07 | Wendt FR | 21/12/2020 | Nat Hum Behav | www.ncbi.nlm.nih.gov/pubmed/33349686 | Multivariate genome-wide analysis of education, socioeconomic status and brain phenome. | Subjective wellbeing conditioned on self-rated math ability (multi-trait conditioning and joint analysis) | 298,420 Eu | NA | 20q13.13 | | ARFGEF2 | | | ENSG00000124198 | NA | NA | rs707533-? | rs707533 | 0 | 707533 | 3_prime_UTR_variant | 0 | NR | 7.00E-11 | 10.1549 | | NA | | NR [NR] | N |
| 11 | 20:47746974 | 20 | 47701309 | rs7264419 | 18/10/2022 | 3.5E+07 | Maihofer AX | 28/09/2021 | Biol Psychiatry | www.ncbi.nlm.nih.gov/pubmed/34865855 | Enhancing Discovery of Genetic Variants for Posttraumatic Stress Disorder Through Integration of Quantitative Phenotypes and Trauma Exposure Information. | Post-traumatic stress disorder (MTAG) | 217,491 Eu | NA | 20q13.13 | | CSE1L | | | ENSG00000124207 | NA | NA | rs7264419-A | rs7264419 | 0 | 7264419 | intron_variant | 0 | NR | 5.00E-09 | 8.30103 | | 5.85 | Z score dec | Affymetrix, Illumina [NR] (imputed) | N |
| 11 | 20:47495794 | 20 | 47447870 | rs7267607 | 21/09/2020 | 3.3E+07 | Chen MH | 1/09/2020 | Cell | www.ncbi.nlm.nih.gov/pubmed/32888493 | Trans-ethnic and Ancestry-Specific Blood-Cell Genetics in 746,667 Individuals from 5 Global Populations. | Mean corpuscular hemoglobin | 486,823 Eu | NA | 20q13.13 | NR | PREX1 - ARFGEF2 | ENSG00000124126 | ENSG00000124198 | | 3334 | 90378 | rs7267607-C | rs7267607 | 0 | 7267607 | intron_variant | 1 | 0.341038 | 1.00E-22 | 22 | | 0.019896 | [0.016-0.0 | Affymetrix, Illumina [50266654] (imputed) | N |
| 11 | 20:47746974 | 20 | 47762249 | rs8122757 | 8/02/2019 | 3.1E+07 | Kichaev G | 27/12/2018 | Am J Hum Genet | www.ncbi.nlm.nih.gov/pubmed/30595370 | Leveraging Polygenic Functional Enrichment to Improve GWAS Power. | Lung function (FVC) | approxima | NA | 20q13.13 | | STAU1 | | | ENSG00000124214 | NA | NA | rs8122757-? | rs8122757 | 0 | 8122757 | intron_variant | 0 | NR | 7.00E-11 | 10.1549 | | NA | | NR [~ 8900000] (imputed) | N |
