## Supplementary material for "A genome-wide association study identified 10 novel genomic loci associated with intrinsic capacity": MAGMA gene based test for all gene mapped from the whole GWAS SNPs

| GENE | CHR | START | STOP | NSNPS | NPARAM | N | ZSTAT | P | Gene | |
|---|---|---|---|---|---|---|---|---|---|---|
| ENSG00000159314 | 17 | 43461275 | 43521787 | 178 | 14 | 47193 | 5.9669 | 1.21E-09 | ARHGAP27 | GENE = Ensemble Gene ID |
| ENSG00000225190 | 17 | 43503266 | 43578115 | 212 | 10 | 45401 | 5.7189 | 5.36E-09 | PLEKHM1 | CHR = Chromsome |
| ENSG00000124207 | 20 | 47652849 | 47723489 | 219 | 19 | 49975 | 5.4487 | 2.54E-08 | CSE1L | NSNPS = Number of SNPs |
| ENSG00000212673 | 1 | 32369174 | 32390745 | 37 | 11 | 43536 | 5.4262 | 2.88E-08 | AL136115.1 | NPARAM = number of parameters |
| ENSG00000164068 | 3 | 49716932 | 49768962 | 85 | 16 | 54019 | 5.2441 | 7.85E-08 | RNF123 | N= Sample Size |
| ENSG00000184007 | 1 | 32362022 | 32420457 | 97 | 21 | 40984 | 5.0562 | 2.14E-07 | PTP4A2 | ZSTAT = Z-Statistic |
| ENSG00000176095 | 3 | 49751727 | 49833975 | 155 | 15 | 53536 | 5.0498 | 2.21E-07 | IP6K1 | P= P-value |
| ENSG00000176020 | 3 | 49744267 | 49771349 | 52 | 9 | 54619 | 5.0495 | 2.22E-07 | AMIGO3 | Gene = Gene name |
| ENSG00000173540 | 3 | 49744277 | 49771384 | 52 | 9 | 54619 | 5.0495 | 2.22E-07 | GMPPB | |
| ENSG00000108433 | 17 | 44990483 | 45115003 | 461 | 42 | 48872 | 4.9383 | 3.94E-07 | GOSR2 | |
| ENSG00000164161 | 4 | 1.46E+08 | 1.46E+08 | 236 | 24 | 39924 | 4.862 | 5.81E-07 | HHIP | |
| ENSG00000262633 | 17 | 44990499 | 45134520 | 513 | 45 | 48439 | 4.8552 | 6.01E-07 | RP11-156P1.2 | |
| ENSG00000179673 | 17 | 45045523 | 45066614 | 106 | 14 | 49366 | 4.7287 | 1.13E-06 | RPRML | |
| ENSG00000124198 | 20 | 47528427 | 47663230 | 391 | 29 | 46471 | 4.6958 | 1.33E-06 | ARFGEF2 | |
| ENSG00000244486 | 22 | 20768874 | 20802146 | 122 | 24 | 49669 | 4.6557 | 1.61E-06 | SCARF2 | |
| ENSG00000233276 | 3 | 49384609 | 49406033 | 37 | 9 | 51646 | 4.6051 | 2.06E-06 | GPX1 | |
| ENSG00000187492 | 3 | 49818165 | 49847268 | 51 | 12 | 50136 | 4.6036 | 2.08E-06 | CDHR4 | |
| ENSG00000138780 | 4 | 1.07E+08 | 1.07E+08 | 406 | 22 | 46928 | 4.567 | 2.47E-06 | GSTCD | |
| ENSG00000007312 | 17 | 61996100 | 62019714 | 65 | 11 | 49972 | 4.5557 | 2.61E-06 | CD79B | |
| ENSG00000185614 | 3 | 49830687 | 49852463 | 31 | 8 | 49711 | 4.5543 | 2.63E-06 | FAM212A | |
| ENSG00000138785 | 4 | 106593784 | 106827143 | 607 | 33 | 44272 | 4.5465 | 2.73E-06 | INTS12 | |
| ENSG00000145868 | 5 | 147753498 | 147832399 | 154 | 20 | 42649 | 4.5338 | 2.90E-06 | FBXO38 | |
| ENSG00000173531 | 3 | 49711380 | 49736934 | 31 | 9 | 51358 | 4.5227 | 3.05E-06 | MST1 | |
| ENSG00000182179 | 3 | 49832640 | 49861379 | 44 | 11 | 49348 | 4.5009 | 3.38E-06 | UBA7 | |
| ENSG00000254318 | 8 | 94136324 | 94189079 | 145 | 26 | 55922 | 4.4246 | 4.83E-06 | C8orf87 | |
| ENSG00000188483 | 9 | 131927835 | 131950540 | 44 | 12 | 40546 | 4.4029 | 5.34E-06 | IER5L | |
| ENSG00000107679 | 10 | 124124212 | 124201867 | 231 | 25 | 53876 | 4.3845 | 5.81E-06 | PLEKHA1 | |
| ENSG00000183763 | 3 | 49856034 | 49904007 | 76 | 14 | 48094 | 4.3706 | 6.20E-06 | TRAIP | |
| ENSG00000124214 | 20 | 47719878 | 47814904 | 248 | 24 | 45658 | 4.3139 | 8.02E-06 | STAU1 | |
| ENSG00000169218 | 1 | 38066951 | 38110595 | 128 | 26 | 48615 | 4.2532 | 1.05E-05 | RSPO1 | |
| ENSG00000145022 | 3 | 49439639 | 49463908 | 42 | 7 | 47725 | 4.237 | 1.13E-05 | TCTA | |
| ENSG00000164061 | 3 | 49581922 | 49718978 | 210 | 21 | 47165 | 4.2026 | 1.32E-05 | BSN | |
| ENSG00000151465 | 10 | 12227964 | 12302588 | 220 | 29 | 46127 | 4.1921 | 1.38E-05 | CDC123 | |
| ENSG00000258708 | 14 | 37631093 | 37653016 | 47 | 10 | 38856 | 4.1506 | 1.66E-05 | SLC25A21-AS1 | |
| ENSG00000164142 | 4 | 152320368 | 152594784 | 676 | 32 | 51331 | 4.1365 | 1.76E-05 | FAM160A1 | |

| | | | | | | | | | |
|---|---|---|---|---|---|---|---|---|---|
| ENSG00000052841 | 11 | 43370482 | 43526483 | 384 | 52 | 53247 | 4.1161 | 1.93E-05 | *TTC17* |
| ENSG00000198860 | 1 | 184010811 | 184053346 | 67 | 13 | 51308 | 4.0906 | 2.15E-05 | *TSEN15* |
| ENSG00000143520 | 1 | 152311211 | 152342482 | 46 | 7 | 27514 | 4.0891 | 2.17E-05 | *FLG2* |
| ENSG00000145029 | 3 | 49450379 | 49476759 | 52 | 10 | 43977 | 4.0716 | 2.33E-05 | *NICN1* |
| ENSG00000185920 | 9 | 98195262 | 98289339 | 242 | 32 | 47598 | 4.0434 | 2.63E-05 | *PTCH1* |
| ENSG00000067560 | 3 | 49386578 | 49460431 | 150 | 15 | 46944 | 4.038 | 2.70E-05 | *RHOA* |
| ENSG00000179344 | 6 | 32617244 | 32646160 | 1001 | 33 | 45126 | 4.0365 | 2.71E-05 | *HLA-DQB1* |
| ENSG00000143631 | 1 | 152264651 | 152307679 | 126 | 11 | 25951 | 4.0009 | 3.16E-05 | *FLG* |
| ENSG00000176124 | 13 | 50646307 | 51307372 | 1778 | 187 | 49605 | 4.0001 | 3.17E-05 | *DLEU1* |
| ENSG00000145020 | 3 | 49444211 | 49470186 | 44 | 8 | 42779 | 3.9895 | 3.31E-05 | *AMT* |
| ENSG00000066336 | 11 | 47366411 | 47410127 | 144 | 19 | 43439 | 3.965 | 3.67E-05 | *SPI1* |
| ENSG00000118007 | 3 | 136045077 | 136481220 | 797 | 58 | 48806 | 3.9447 | 3.99E-05 | *STAG1* |
| ENSG00000138813 | 4 | 100422161 | 100473460 | 249 | 34 | 42542 | 3.938 | 4.11E-05 | *C4orf17* |
| ENSG00000167130 | 9 | 131833379 | 131862717 | 56 | 10 | 40593 | 3.9364 | 4.14E-05 | *DOLPP1* |
| ENSG00000156218 | 15 | 84312838 | 84718594 | 1166 | 80 | 41849 | 3.9153 | 4.51E-05 | *ADAMTSL3* |
| ENSG00000145331 | 4 | 100457866 | 100495189 | 173 | 17 | 44549 | 3.8734 | 5.37E-05 | *TRMT10A* |
| ENSG00000151338 | 14 | 37657118 | 38031566 | 851 | 59 | 43723 | 3.8696 | 5.45E-05 | *MIPOL1* |
| ENSG00000164162 | 4 | 145878264 | 146029693 | 175 | 29 | 40493 | 3.8623 | 5.62E-05 | *ANAPC10* |
| ENSG00000108379 | 17 | 44829872 | 44920520 | 163 | 40 | 38495 | 3.844 | 6.05E-05 | *WNT3* |
| ENSG00000236699 | 4 | 106463777 | 106639250 | 510 | 62 | 43501 | 3.8331 | 6.33E-05 | *ARHGEF38* |
| ENSG00000205177 | 11 | 33709807 | 33732347 | 91 | 26 | 45135 | 3.8247 | 6.55E-05 | *C11orf91* |
| ENSG00000143536 | 1 | 152371719 | 152396739 | 49 | 8 | 34182 | 3.8101 | 6.95E-05 | *CRNN* |
| ENSG00000187045 | 22 | 37451476 | 37515603 | 229 | 40 | 49995 | 3.8018 | 7.18E-05 | *TMPRSS6* |
| ENSG00000131508 | 5 | 138896016 | 139018018 | 256 | 25 | 50365 | 3.798 | 7.29E-05 | *UBE2D2* |
| ENSG00000166181 | 11 | 43323513 | 43376079 | 102 | 25 | 49752 | 3.7975 | 7.31E-05 | *API5* |
| ENSG00000111011 | 12 | 122979190 | 123021547 | 80 | 13 | 51477 | 3.7802 | 7.83E-05 | *RSRC2* |
| ENSG00000165915 | 11 | 47418683 | 47448047 | 78 | 11 | 47316 | 3.7755 | 7.98E-05 | *SLC39A13* |
| ENSG00000173898 | 11 | 66442719 | 66506697 | 67 | 27 | 49227 | 3.757 | 8.60E-05 | *SPTBN2* |
| ENSG00000138138 | 10 | 89501269 | 89611100 | 229 | 17 | 50926 | 3.7497 | 8.85E-05 | *ATAD1* |
| ENSG00000072958 | 19 | 16298389 | 16356160 | 215 | 15 | 30438 | 3.7387 | 9.25E-05 | *AP1M1* |
| ENSG00000173402 | 3 | 49496146 | 49583048 | 154 | 16 | 45590 | 3.7365 | 9.33E-05 | *DAG1* |
| ENSG00000118508 | 6 | 146854829 | 146886101 | 50 | 14 | 42103 | 3.7295 | 9.59E-05 | *RAB32* |
| ENSG00000119383 | 9 | 131863229 | 131921225 | 154 | 11 | 37403 | 3.7291 | 9.61E-05 | *PPP2R4* |
| ENSG00000164330 | 5 | 158112928 | 158536769 | 892 | 116 | 49529 | 3.7269 | 9.69E-05 | *EBF1* |
| ENSG00000176720 | 2 | 242488136 | 242523546 | 139 | 31 | 41398 | 3.7223 | 9.87E-05 | *BOK* |
| ENSG00000114054 | 3 | 135959148 | 136066738 | 178 | 26 | 47799 | 3.7217 | 9.90E-05 | *PCCB* |

| | | | | | | | | | |
|---|---|---|---|---|---|---|---|---|---|
| ENSG00000163257 | 4 | 17792278 | 17822381 | 73 | 11 | 41727 | 3.6983 | 0.00010851 | *DCAF16* |
| ENSG00000109610 | 4 | 24781534 | 24812464 | 81 | 15 | 41096 | 3.6759 | 0.00011849 | *SOD3* |
| ENSG00000163877 | 1 | 37990050 | 38029903 | 90 | 16 | 51047 | 3.6733 | 0.00011971 | *SNIP1* |
| ENSG00000165916 | 11 | 47430320 | 47457993 | 87 | 10 | 45651 | 3.6733 | 0.00011972 | *PSMC3* |
| ENSG00000033030 | 12 | 122947417 | 122995518 | 97 | 15 | 49705 | 3.6578 | 0.00012719 | *ZCCHC8* |
| ENSG00000140600 | 15 | 84105980 | 84297495 | 489 | 91 | 45958 | 3.6555 | 0.00012833 | *SH3GL3* |
| ENSG00000185002 | 6 | 117188375 | 117263326 | 274 | 19 | 45292 | 3.6413 | 0.00013564 | *RFX6* |
| ENSG00000168944 | 5 | 122670579 | 122769286 | 347 | 22 | 52545 | 3.6079 | 0.00015434 | *CEP120* |
| ENSG00000085063 | 11 | 33709807 | 33767991 | 215 | 40 | 46049 | 3.6055 | 0.00015575 | *CD59* |
| ENSG00000167194 | 16 | 30024655 | 30049057 | 46 | 6 | 50479 | 3.6032 | 0.00015718 | *C16orf92* |
| ENSG00000197915 | 1 | 152174558 | 152206669 | 117 | 10 | 27858 | 3.5815 | 0.00017078 | *HRNR* |
| ENSG00000171812 | 1 | 36550837 | 36600821 | 89 | 20 | 47106 | 3.5799 | 0.00017186 | *COL8A2* |
| ENSG00000136490 | 17 | 61763262 | 61788532 | 36 | 8 | 40639 | 3.5774 | 0.00017351 | *LIMD2* |
| ENSG00000243156 | 22 | 18260415 | 18517325 | 1149 | 113 | 47941 | 3.5723 | 0.0001769 | *MICAL3* |
| ENSG00000073969 | 17 | 44658035 | 44844830 | 76 | 13 | 28771 | 3.5642 | 0.00018251 | *NSF* |
| ENSG00000063761 | 14 | 78256426 | 78411355 | 748 | 78 | 46972 | 3.56 | 0.00018539 | *ADCK1* |
| ENSG00000259384 | 17 | 61984560 | 62006179 | 63 | 15 | 48937 | 3.5451 | 0.00019621 | *GH1* |
| ENSG00000108387 | 17 | 56587611 | 56628179 | 76 | 14 | 53640 | 3.5446 | 0.00019659 | *Sep-04* |
| ENSG00000154813 | 3 | 16289485 | 16316479 | 98 | 13 | 54026 | 3.5316 | 0.00020655 | *DPH3* |
| ENSG00000136463 | 17 | 61668231 | 61695725 | 16 | 7 | 49928 | 3.5289 | 0.00020866 | *TACO1* |
| ENSG00000009950 | 7 | 72997524 | 73048873 | 156 | 21 | 55480 | 3.5175 | 0.00021779 | *MLXIPL* |
| ENSG00000149926 | 16 | 30025748 | 30074299 | 85 | 10 | 47939 | 3.5125 | 0.00022192 | *FAM57B* |
| ENSG00000174579 | 3 | 135857764 | 135926083 | 86 | 21 | 50017 | 3.51 | 0.00022405 | *MSL2* |
| ENSG00000134697 | 1 | 38022417 | 38071536 | 128 | 19 | 49642 | 3.5081 | 0.00022566 | *GNL2* |
| ENSG00000167565 | 19 | 40936751 | 40960612 | 94 | 14 | 49229 | 3.4993 | 0.00023321 | *SERTAD3* |
| ENSG00000166797 | 15 | 64354758 | 64396217 | 109 | 16 | 35404 | 3.4971 | 0.00023514 | *FAM96A* |
| ENSG00000122375 | 10 | 88404314 | 88436605 | 184 | 25 | 48371 | 3.4969 | 0.00023534 | *OPN4* |
| ENSG00000095321 | 9 | 131847089 | 131883468 | 81 | 9 | 37898 | 3.4939 | 0.00023799 | *CRAT* |
| ENSG00000124201 | 20 | 47844483 | 47904963 | 260 | 19 | 36653 | 3.4933 | 0.0002385 | *ZNFX1* |
| ENSG00000136425 | 15 | 78386948 | 78433886 | 151 | 25 | 47327 | 3.4796 | 0.00025113 | *CIB2* |
| ENSG00000115252 | 2 | 182994763 | 183397919 | 1465 | 112 | 48372 | 3.479 | 0.00025165 | *PDE1A* |
| ENSG00000184347 | 5 | 168078745 | 168738133 | 2519 | 284 | 46297 | 3.46 | 0.00027007 | *SLIT3* |
| ENSG00000196735 | 6 | 32585956 | 32624839 | 2064 | 48 | 43781 | 3.4574 | 0.00027268 | *HLA-DQA1* |
| ENSG00000138823 | 4 | 100474918 | 100555156 | 397 | 39 | 47627 | 3.4524 | 0.00027784 | *MTTP* |
| ENSG00000140521 | 15 | 89849534 | 89888092 | 84 | 13 | 52413 | 3.4494 | 0.0002809 | *POLG* |
| ENSG00000088827 | 20 | 3657617 | 3697775 | 161 | 34 | 50724 | 3.442 | 0.00028872 | *SIGLEC1* |

| | | | | | | | | | |
|---|---|---|---|---|---|---|---|---|---|
| ENSG00000186367 | 5 | 129073772 | 129112425 | 113 | 19 | 47362 | 3.4399 | 0.00029095 | *KIAA1024L* |
| ENSG00000007314 | 17 | 62005914 | 62060278 | 136 | 28 | 40745 | 3.4394 | 0.00029148 | *SCN4A* |
| ENSG00000160305 | 21 | 47868812 | 47999926 | 560 | 50 | 48118 | 3.4375 | 0.00029353 | *DIP2A* |
| ENSG00000198909 | 17 | 61689775 | 61783663 | 129 | 12 | 42559 | 3.4338 | 0.00029764 | *MAP3K3* |
| ENSG00000140525 | 15 | 89777180 | 89870492 | 201 | 16 | 52207 | 3.4207 | 0.00031231 | *FANCI* |
| ENSG00000180900 | 8 | 144863090 | 144907549 | 110 | 27 | 45654 | 3.4135 | 0.00032063 | *SCRIB* |
| ENSG00000123600 | 2 | 172163919 | 172301312 | 307 | 39 | 50815 | 3.4097 | 0.00032512 | *METTL8* |
| ENSG00000149927 | 16 | 30006830 | 30044591 | 79 | 10 | 42700 | 3.409 | 0.000326 | *DOC2A* |
| ENSG00000164062 | 3 | 49701435 | 49731396 | 42 | 8 | 44151 | 3.4069 | 0.0003285 | *APEH* |
| ENSG00000099910 | 22 | 20773528 | 20860170 | 211 | 33 | 47162 | 3.405 | 0.00033078 | *KLHL22* |
| ENSG00000160310 | 21 | 48045079 | 48095036 | 186 | 28 | 53222 | 3.4043 | 0.00033168 | *PRMT2* |
| ENSG00000163879 | 1 | 38012520 | 38042458 | 83 | 17 | 51646 | 3.3987 | 0.00033852 | *DNALI1* |
| ENSG00000141741 | 17 | 37874749 | 37897040 | 39 | 9 | 30912 | 3.3879 | 0.00035212 | *MIEN1* |
| ENSG00000181013 | 17 | 56608948 | 56631729 | 49 | 12 | 48751 | 3.3868 | 0.00035361 | *C17orf47* |
| ENSG00000196782 | 4 | 140627907 | 141085338 | 1221 | 156 | 43329 | 3.385 | 0.00035591 | *MAML3* |
| ENSG00000172660 | 17 | 34126459 | 34201619 | 215 | 21 | 46291 | 3.3511 | 0.00040252 | *TAF15* |
| ENSG00000165609 | 10 | 12197324 | 12248143 | 146 | 26 | 49031 | 3.3495 | 0.00040475 | *NUDT5* |
| ENSG00000129965 | 11 | 2143768 | 2192439 | 147 | 33 | 45158 | 3.3468 | 0.00040878 | *INS-IGF2* |
| ENSG00000149932 | 16 | 29942206 | 29994373 | 76 | 8 | 48568 | 3.3444 | 0.00041234 | *TMEM219* |
| ENSG00000165671 | 5 | 176550026 | 176737216 | 484 | 38 | 39705 | 3.3378 | 0.0004223 | *NSD1* |
| ENSG00000134571 | 11 | 47342957 | 47384253 | 127 | 23 | 39070 | 3.3344 | 0.00042737 | *MYBPC3* |
| ENSG00000047662 | 4 | 17620929 | 17793135 | 506 | 32 | 43320 | 3.3224 | 0.00044625 | *FAM184B* |
| ENSG00000179950 | 8 | 144888514 | 144922029 | 61 | 17 | 49119 | 3.3213 | 0.00044804 | *PUF60* |
| ENSG00000167244 | 11 | 2140342 | 2180833 | 133 | 27 | 47671 | 3.3212 | 0.00044812 | *IGF2* |
| ENSG00000149922 | 16 | 30087114 | 30113208 | 36 | 11 | 53218 | 3.3149 | 0.00045837 | *TBX6* |
| ENSG00000115112 | 2 | 121964163 | 122052783 | 217 | 44 | 38648 | 3.3134 | 0.00046083 | *TFCP2L1* |
| ENSG00000109805 | 4 | 17802525 | 17856485 | 122 | 12 | 42657 | 3.3037 | 0.00047713 | *NCAPG* |
| ENSG00000173914 | 11 | 66422469 | 66455392 | 37 | 16 | 49186 | 3.298 | 0.00048685 | *RBM4B* |
| ENSG00000181690 | 8 | 57063463 | 57133883 | 142 | 23 | 53688 | 3.2978 | 0.0004873 | *PLAG1* |
| ENSG00000169064 | 3 | 166948075 | 167109134 | 451 | 44 | 51253 | 3.2932 | 0.00049522 | *ZBBX* |
| ENSG00000090238 | 16 | 30093635 | 30118236 | 37 | 13 | 50905 | 3.2858 | 0.00050839 | *YPEL3* |
| ENSG00000141736 | 17 | 37834167 | 37896679 | 114 | 17 | 35063 | 3.2709 | 0.00053596 | *ERBB2* |
| ENSG00000119705 | 14 | 78164414 | 78237447 | 328 | 29 | 51792 | 3.2708 | 0.0005362 | *SLIRP* |
| ENSG00000167216 | 18 | 44487455 | 44637658 | 472 | 46 | 54010 | 3.2686 | 0.00054044 | *KATNAL2* |
| ENSG00000156787 | 8 | 124044208 | 124174393 | 347 | 50 | 51399 | 3.2685 | 0.00054054 | *TBC1D31* |
| ENSG00000124228 | 20 | 47825884 | 47870614 | 163 | 16 | 39296 | 3.2609 | 0.00055526 | *DDX27* |

| | | | | | | | | | |
|---|---|---|---|---|---|---|---|---|---|
| ENSG00000100603 | 14 | 78173942 | 78237550 | 293 | 26 | 52404 | 3.26 | 0.000557 | *SNW1* |
| ENSG00000125686 | 17 | 37550538 | 37617539 | 140 | 13 | 32800 | 3.2599 | 0.00055735 | *MED1* |
| ENSG00000132671 | 20 | 23006057 | 23027314 | 65 | 10 | 48057 | 3.2568 | 0.00056344 | *SSTR4* |
| ENSG00000167967 | 16 | 2263567 | 2295743 | 72 | 16 | 45670 | 3.252 | 0.00057298 | *E4F1* |
| ENSG00000117245 | 1 | 20980507 | 21054510 | 264 | 67 | 43184 | 3.2487 | 0.0005797 | *KIF17* |
| ENSG00000092199 | 14 | 21667295 | 21747653 | 207 | 24 | 52842 | 3.2472 | 0.00058265 | *HNRNPC* |
| ENSG00000150750 | 11 | 111116707 | 111167126 | 156 | 23 | 50690 | 3.2453 | 0.00058666 | *C11orf53* |
| ENSG00000164164 | 4 | 146021990 | 146111313 | 96 | 24 | 39459 | 3.2409 | 0.00059572 | *OTUD4* |
| ENSG00000187456 | 17 | 34235070 | 34267777 | 105 | 10 | 50096 | 3.2391 | 0.00059961 | *RDM1* |
| ENSG00000268208 | 14 | 78217711 | 78238033 | 92 | 15 | 53171 | 3.2214 | 0.00063774 | *AC008372.1* |
| ENSG00000089225 | 12 | 114781736 | 114856247 | 254 | 50 | 49802 | 3.2191 | 0.00064289 | *TBX5* |
| ENSG00000071967 | 2 | 172368757 | 172424643 | 234 | 33 | 47499 | 3.2189 | 0.0006434 | *CYBRD1* |
| ENSG00000011304 | 19 | 787075 | 822327 | 158 | 33 | 46459 | 3.2187 | 0.00064381 | *PTBP1* |
| ENSG00000018510 | 2 | 178247372 | 178418564 | 431 | 26 | 48407 | 3.216 | 0.00065001 | *AGPS* |
| ENSG00000116044 | 2 | 178082323 | 178267425 | 442 | 47 | 42465 | 3.2159 | 0.00065017 | *NFE2L2* |
| ENSG00000172653 | 17 | 34171955 | 34205895 | 92 | 13 | 49039 | 3.2124 | 0.00065823 | *C17orf66* |
| ENSG00000197734 | 14 | 78217173 | 78246085 | 133 | 19 | 52531 | 3.2114 | 0.00066037 | *C14orf178* |
| ENSG00000167258 | 17 | 37607764 | 37731160 | 275 | 20 | 33563 | 3.2108 | 0.00066192 | *CDK12* |
| ENSG00000178726 | 20 | 23016270 | 23040378 | 59 | 13 | 45192 | 3.202 | 0.00068227 | *THBD* |
| ENSG00000176058 | 9 | 140076069 | 140108645 | 73 | 11 | 55173 | 3.2003 | 0.00068642 | *TPRN* |
| ENSG00000149930 | 16 | 29974962 | 30013582 | 65 | 9 | 42106 | 3.1948 | 0.00069966 | *TAOK2* |
| ENSG00000111885 | 6 | 119488374 | 119680926 | 814 | 55 | 50959 | 3.1899 | 0.00071151 | *MAN1A1* |
| ENSG00000198682 | 10 | 89409370 | 89517462 | 361 | 45 | 43025 | 3.1899 | 0.00071157 | *PAPSS2* |
| ENSG00000198657 | 11 | 124283807 | 124304789 | 112 | 10 | 50713 | 3.1855 | 0.00072262 | *OR8B4* |
| ENSG00000204922 | 11 | 62427745 | 62451159 | 36 | 7 | 54056 | 3.1829 | 0.00072916 | *C11orf83* |
| ENSG00000054116 | 1 | 36592173 | 36625098 | 57 | 10 | 42505 | 3.1761 | 0.00074629 | *TRAPPC3* |
| ENSG00000198945 | 6 | 130324844 | 130472594 | 445 | 51 | 40541 | 3.1738 | 0.00075231 | *L3MBTL3* |
| ENSG00000060718 | 1 | 103332023 | 103584052 | 1239 | 62 | 45903 | 3.173 | 0.00075433 | *COL11A1* |
| ENSG00000028528 | 15 | 64376322 | 64448289 | 148 | 18 | 37722 | 3.1721 | 0.00075671 | *SNX1* |
| ENSG00000173237 | 11 | 66732748 | 66754475 | 21 | 13 | 54219 | 3.1685 | 0.00076602 | *C11orf86* |
| ENSG00000108306 | 17 | 37405384 | 37568776 | 383 | 21 | 31875 | 3.1666 | 0.00077121 | *FBXL20* |
| ENSG00000178177 | 4 | 17832822 | 18033499 | 543 | 28 | 35685 | 3.166 | 0.00077275 | *LCORL* |
| ENSG00000145979 | 6 | 13256774 | 13338815 | 395 | 35 | 38014 | 3.1599 | 0.00078906 | *TBC1D7* |
| ENSG00000107249 | 9 | 3814127 | 4358392 | 2432 | 250 | 45241 | 3.1545 | 0.0008039 | *GLIS3* |
| ENSG00000161570 | 17 | 34188495 | 34217797 | 87 | 14 | 49220 | 3.1522 | 0.00081023 | *CCL5* |
| ENSG00000105559 | 19 | 49330354 | 49381889 | 215 | 28 | 35290 | 3.1518 | 0.00081139 | *PLEKHA4* |

| | | | | | | | | | |
|---|---|---|---|---|---|---|---|---|---|
| ENSG00000077147 | 10 | 98267866 | 98357209 | 253 | 21 | 44109 | 3.151 | 0.0008137 | *TM9SF3* |
| ENSG00000178965 | 1 | 75023795 | 75149422 | 232 | 32 | 39499 | 3.1506 | 0.00081467 | *C1orf173* |
| ENSG00000162063 | 16 | 2469395 | 2518855 | 123 | 7 | 46203 | 3.1488 | 0.00081983 | *CCNF* |
| ENSG00000111452 | 12 | 131428452 | 131636014 | 977 | 128 | 44601 | 3.1427 | 0.00083701 | *GPR133* |
| ENSG00000152620 | 5 | 36182694 | 36252381 | 125 | 22 | 35057 | 3.1397 | 0.00084561 | *NADK2* |
| ENSG00000160285 | 21 | 47598055 | 47658738 | 272 | 37 | 44615 | 3.1331 | 0.00086476 | *LSS* |
| ENSG00000149925 | 16 | 30054411 | 30091778 | 48 | 10 | 43901 | 3.1261 | 0.00088581 | *ALDOA* |
| ENSG00000169194 | 5 | 131981955 | 132006802 | 55 | 14 | 40648 | 3.1239 | 0.00089221 | *IL13* |
| ENSG00000164188 | 5 | 36238536 | 36312216 | 132 | 26 | 33123 | 3.1215 | 0.00089953 | *RANBP3L* |
| ENSG00000176101 | 9 | 140073099 | 140094822 | 44 | 4 | 54856 | 3.1185 | 0.00090899 | *SSNA1* |
| ENSG00000066379 | 6 | 30016676 | 30042686 | 167 | 22 | 42247 | 3.1172 | 0.00091287 | *ZNRD1* |
| ENSG00000108389 | 17 | 56556898 | 56605266 | 74 | 14 | 54923 | 3.1089 | 0.00093887 | *MTMR4* |
| ENSG00000105976 | 7 | 116302444 | 116448440 | 280 | 52 | 42203 | 3.1069 | 0.00094523 | *MET* |
| ENSG00000135698 | 16 | 82171403 | 82213831 | 397 | 22 | 40732 | 3.1033 | 0.00095704 | *MPHOSPH6* |
| ENSG00000214756 | 11 | 62422781 | 62445968 | 41 | 6 | 52067 | 3.0992 | 0.00097007 | *METTL12* |
| ENSG00000116871 | 1 | 36611180 | 36656450 | 69 | 14 | 44928 | 3.0958 | 0.00098143 | *MAP7D1* |
| ENSG00000146535 | 7 | 2757746 | 2893958 | 530 | 39 | 52278 | 3.0932 | 0.00099018 | *GNA12* |
| ENSG00000154814 | 3 | 16296706 | 16401806 | 394 | 34 | 50898 | 3.0914 | 0.00099591 | *OXNAD1* |
| ENSG00000162194 | 11 | 62420287 | 62449727 | 48 | 7 | 52840 | 3.0907 | 0.00099828 | *C11orf48* |
| ENSG00000168769 | 4 | 106057032 | 106210973 | 370 | 50 | 45766 | 3.0876 | 0.0010088 | *TET2* |
| ENSG00000176248 | 9 | 140059236 | 140092989 | 59 | 7 | 49937 | 3.0837 | 0.0010221 | *ANAPC2* |
| ENSG00000124613 | 6 | 27332394 | 27381683 | 274 | 15 | 49433 | 3.0819 | 0.0010285 | *ZNF391* |
| ENSG00000088986 | 12 | 120897653 | 120946296 | 172 | 23 | 40549 | 3.0785 | 0.0010402 | *DYNLL1* |
| ENSG00000184207 | 16 | 2251998 | 2274808 | 63 | 11 | 42312 | 3.0739 | 0.0010563 | *PGP* |
| ENSG00000114455 | 3 | 108005376 | 108107132 | 337 | 38 | 38787 | 3.0555 | 0.0011235 | *HHLA2* |
| ENSG00000162191 | 11 | 62433970 | 62456567 | 35 | 7 | 52633 | 3.0546 | 0.0011267 | *UBXN1* |
| ENSG00000109046 | 17 | 25611102 | 25650657 | 106 | 16 | 48254 | 3.0533 | 0.0011316 | *WSB1* |
| ENSG00000124564 | 6 | 25823294 | 25892514 | 238 | 30 | 42801 | 3.0523 | 0.0011355 | *SLC17A3* |
| ENSG00000204414 | 17 | 61976957 | 62006198 | 89 | 18 | 42110 | 3.0514 | 0.0011389 | *CSHL1* |
| ENSG00000090013 | 19 | 40943696 | 40981747 | 142 | 25 | 46375 | 3.0498 | 0.001145 | *BLVRB* |
| ENSG00000064545 | 19 | 19219978 | 19259322 | 103 | 21 | 50707 | 3.0462 | 0.0011587 | *TMEM161A* |
| ENSG00000248672 | 2 | 160618362 | 160771221 | 584 | 65 | 38134 | 3.0455 | 0.0011613 | *LY75-CD302* |
| ENSG00000054219 | 2 | 160618362 | 160771260 | 584 | 65 | 38134 | 3.0455 | 0.0011613 | *LY75* |
| ENSG00000167272 | 12 | 121006567 | 121029201 | 85 | 14 | 42775 | 3.0443 | 0.0011661 | *POP5* |
| ENSG00000166839 | 15 | 65194101 | 65261042 | 209 | 34 | 42043 | 3.0431 | 0.0011708 | *ANKDD1A* |
| ENSG00000099917 | 22 | 20840200 | 20951919 | 304 | 31 | 44925 | 3.0391 | 0.0011866 | *MED15* |

| | | | | | | | | | |
|---|---|---|---|---|---|---|---|---|---|
| ENSG00000185697 | 8 | 67464410 | 67536482 | 96 | 14 | 37459 | 3.038 | 0.0011907 | *MYBL1* |
| ENSG00000173933 | 11 | 66374097 | 66444153 | 86 | 22 | 47518 | 3.0362 | 0.0011978 | *RBM4* |
| ENSG00000110768 | 11 | 18333842 | 18398591 | 133 | 21 | 42651 | 3.0332 | 0.0012097 | *GTF2H1* |
| ENSG00000273003 | 11 | 64771654 | 64818042 | 136 | 20 | 35136 | 3.0322 | 0.001214 | *RP11-399J13.3* |
| ENSG00000110025 | 11 | 64784880 | 64818044 | 103 | 13 | 31987 | 3.0301 | 0.0012223 | *SNX15* |
| ENSG00000213465 | 11 | 64771585 | 64799656 | 80 | 18 | 38883 | 3.03 | 0.0012228 | *ARL2* |
| ENSG00000160299 | 21 | 47734036 | 47875682 | 595 | 51 | 43425 | 3.0249 | 0.0012435 | *PCNT* |
| ENSG00000196659 | 2 | 178403726 | 178427742 | 58 | 8 | 46926 | 3.0236 | 0.0012489 | *TTC30B* |
| ENSG00000167207 | 16 | 50717514 | 50776988 | 117 | 19 | 30284 | 3.0203 | 0.0012627 | *NOD2* |
| ENSG00000250722 | 5 | 42789982 | 42897494 | 353 | 37 | 44907 | 3.0115 | 0.0013 | *SEPP1* |
| ENSG00000120071 | 17 | 44097282 | 44312733 | 1030 | 8 | 22900 | 3.009 | 0.0013103 | *KANSL1* |
| ENSG00000246705 | 12 | 14917270 | 14940936 | 78 | 15 | 56935 | 3.0079 | 0.0013153 | *H2AFJ* |
| ENSG00000175792 | 3 | 127773621 | 127882757 | 272 | 15 | 54612 | 3.0075 | 0.0013169 | *RUVBL1* |
| ENSG00000179256 | 12 | 14947584 | 14977116 | 80 | 15 | 55981 | 3.0054 | 0.0013261 | *SMCO3* |
| ENSG00000148219 | 9 | 119177504 | 120187348 | 3458 | 291 | 46574 | 2.9995 | 0.0013522 | *ASTN2* |
| ENSG00000110756 | 11 | 18290223 | 18353745 | 179 | 23 | 44350 | 2.9988 | 0.0013553 | *HPS5* |
| ENSG00000198794 | 15 | 75239560 | 75323837 | 159 | 26 | 31401 | 2.996 | 0.0013676 | *SCAMP5* |
| ENSG00000183032 | 14 | 37137636 | 37652071 | 1547 | 163 | 42172 | 2.9875 | 0.0014065 | *SLC25A21* |
| ENSG00000111786 | 12 | 120889471 | 120917596 | 90 | 17 | 42651 | 2.9811 | 0.0014362 | *SRSF9* |
| ENSG00000173612 | 6 | 117103248 | 117160200 | 146 | 20 | 48873 | 2.9798 | 0.0014421 | *GPRC6A* |
| ENSG00000120129 | 5 | 172185093 | 172208198 | 105 | 25 | 43074 | 2.9776 | 0.0014524 | *DUSP1* |
| ENSG00000248643 | 11 | 66374097 | 66423940 | 64 | 17 | 46582 | 2.976 | 0.00146 | *RBM14-RBM4* |
| ENSG00000188820 | 6 | 116772533 | 116794934 | 79 | 17 | 44715 | 2.972 | 0.0014795 | *FAM26F* |
| ENSG00000102753 | 13 | 50263447 | 50377057 | 342 | 40 | 49084 | 2.972 | 0.0014795 | *KPNA3* |
| ENSG00000182685 | 16 | 2249254 | 2271951 | 71 | 13 | 40281 | 2.9713 | 0.0014826 | *BRICD5* |
| ENSG00000183114 | 1 | 20868932 | 20891512 | 97 | 21 | 49215 | 2.9671 | 0.0015031 | *FAM43B* |
| ENSG00000184716 | 15 | 44076360 | 44102419 | 33 | 6 | 31477 | 2.965 | 0.0015136 | *SERINC4* |
| ENSG00000076321 | 1 | 173674080 | 173765840 | 130 | 19 | 50675 | 2.9647 | 0.001515 | *KLHL20* |
| ENSG00000185863 | 9 | 140055310 | 140076861 | 33 | 8 | 44159 | 2.9627 | 0.0015247 | *TMEM210* |
| ENSG00000084092 | 4 | 57819536 | 57854989 | 132 | 25 | 55388 | 2.9614 | 0.001531 | *NOA1* |
| ENSG00000256762 | 17 | 44066616 | 44087060 | 124 | 5 | 22356 | 2.9612 | 0.0015323 | *STH* |
| ENSG00000110871 | 12 | 120931077 | 120982237 | 183 | 22 | 38955 | 2.9608 | 0.001534 | *COQ5* |
| ENSG00000169372 | 12 | 94061151 | 94298616 | 630 | 97 | 50958 | 2.9604 | 0.0015361 | *CRADD* |
| ENSG00000131378 | 3 | 16345081 | 16565533 | 800 | 126 | 46896 | 2.9595 | 0.0015405 | *RFTN1* |
| ENSG00000102886 | 16 | 30106131 | 30135177 | 42 | 16 | 47105 | 2.9543 | 0.0015671 | *GDPD3* |
| ENSG00000121101 | 17 | 56624039 | 56779416 | 380 | 34 | 44077 | 2.9464 | 0.0016072 | *TEX14* |

| | | | | | | | | | |
|---|---|---|---|---|---|---|---|---|---|
| ENSG00000084463 | 12 | 14929410 | 14966474 | 96 | 18 | 56148 | 2.9464 | 0.0016074 | *WBP11* |
| ENSG00000174837 | 19 | 6877577 | 6950470 | 452 | 40 | 39361 | 2.9431 | 0.0016249 | *EMR1* |
| ENSG00000106153 | 7 | 56159262 | 56184269 | 109 | 16 | 53481 | 2.9427 | 0.0016267 | *CHCHD2* |
| ENSG00000140259 | 15 | 44086690 | 44127000 | 63 | 7 | 30124 | 2.9413 | 0.0016343 | *MFAP1* |
| ENSG00000255432 | 11 | 62407166 | 62442650 | 57 | 9 | 52035 | 2.941 | 0.0016358 | *RP11-831H9.11* |
| ENSG00000099331 | 19 | 17176591 | 17335346 | 677 | 91 | 44144 | 2.9383 | 0.0016501 | *MYO9B* |
| ENSG00000186479 | 5 | 63792084 | 63918139 | 285 | 44 | 48718 | 2.9379 | 0.0016521 | *RGS7BP* |
| ENSG00000187713 | 9 | 140088534 | 140110090 | 46 | 12 | 55227 | 2.9365 | 0.0016598 | *TMEM203* |
| ENSG00000087053 | 11 | 95556046 | 95668479 | 330 | 32 | 36060 | 2.9334 | 0.0016762 | *MTMR2* |
| ENSG00000223572 | 15 | 43975084 | 44001420 | 28 | 11 | 35240 | 2.9326 | 0.0016807 | *CKMT1A* |
| ENSG00000092470 | 15 | 44109161 | 44170617 | 101 | 11 | 33053 | 2.923 | 0.0017331 | *WDR76* |
| ENSG00000125245 | 13 | 99896968 | 99923998 | 46 | 14 | 45508 | 2.923 | 0.0017336 | *GPR18* |
| ENSG00000137259 | 6 | 26023320 | 26043796 | 53 | 15 | 48427 | 2.9226 | 0.0017355 | *HIST1H2AB* |
| ENSG00000128602 | 7 | 128818713 | 128863386 | 139 | 25 | 50530 | 2.9216 | 0.0017414 | *SMO* |
| ENSG00000007237 | 17 | 9803926 | 10111868 | 1244 | 196 | 44653 | 2.9212 | 0.0017437 | *GAS7* |
| ENSG00000171724 | 16 | 77812427 | 78024004 | 985 | 166 | 49048 | 2.9128 | 0.0017908 | *VAT1L* |
| ENSG00000140264 | 15 | 44059285 | 44104787 | 68 | 9 | 29923 | 2.9124 | 0.0017932 | *SERF2* |
| ENSG00000157005 | 3 | 187376694 | 187398187 | 85 | 15 | 48958 | 2.9123 | 0.0017939 | *SST* |
| ENSG00000185290 | 7 | 56172374 | 56194093 | 101 | 18 | 54345 | 2.9063 | 0.0018289 | *NUPR1L* |
| ENSG00000239306 | 11 | 66374053 | 66404818 | 42 | 14 | 47559 | 2.9048 | 0.0018376 | *RBM14* |
| ENSG00000204619 | 6 | 30024486 | 30048110 | 176 | 22 | 40974 | 2.9047 | 0.0018381 | *PPP1R11* |
| ENSG00000088538 | 3 | 50702672 | 51431629 | 1382 | 31 | 49018 | 2.902 | 0.0018539 | *DOCK3* |
| ENSG00000102935 | 16 | 49511435 | 49901830 | 1258 | 179 | 46998 | 2.8992 | 0.0018708 | *ZNF423* |
| ENSG00000132842 | 5 | 77286349 | 77600579 | 825 | 53 | 44957 | 2.8962 | 0.0018886 | *AP3B1* |
| ENSG00000172247 | 11 | 47601216 | 47626211 | 37 | 12 | 44347 | 2.8959 | 0.0018904 | *C1QTNF4* |
| ENSG00000169592 | 16 | 29996615 | 30027114 | 56 | 8 | 35567 | 2.8941 | 0.001901 | *INO80E* |
| ENSG00000022840 | 12 | 120961283 | 121025397 | 240 | 25 | 43426 | 2.8921 | 0.0019131 | *RNF10* |
| ENSG00000124678 | 6 | 35075848 | 35126387 | 188 | 18 | 40072 | 2.8886 | 0.0019346 | *TCP11* |
| ENSG00000262560 | 15 | 44054815 | 44101331 | 67 | 9 | 30384 | 2.8852 | 0.001956 | *RP11-296A16.1* |
| ENSG00000188211 | 11 | 17363273 | 17408888 | 155 | 22 | 46510 | 2.8832 | 0.001968 | *NCR3LG1* |
| ENSG00000178718 | 15 | 75236757 | 75259805 | 47 | 12 | 29654 | 2.8791 | 0.0019939 | *RPP25* |
| ENSG00000020633 | 1 | 25216002 | 25301612 | 235 | 41 | 40283 | 2.8783 | 0.001999 | *RUNX3* |
| ENSG00000137727 | 11 | 110437766 | 110593912 | 448 | 40 | 43999 | 2.8779 | 0.0020014 | *ARHGAP20* |
| ENSG00000175073 | 8 | 67530722 | 67589452 | 72 | 16 | 41057 | 2.8668 | 0.0020731 | *VCPIP1* |
| ENSG00000149923 | 16 | 30077299 | 30106698 | 38 | 9 | 49096 | 2.8665 | 0.0020755 | *PPP4C* |
| ENSG00000167220 | 18 | 44623774 | 44686891 | 226 | 22 | 55988 | 2.8617 | 0.0021068 | *HDHD2* |

| | | | | | | | | | |
|---|---|---|---|---|---|---|---|---|---|
| ENSG00000148719 | 10 | 74082588 | 74124988 | 113 | 9 | 24473 | 2.861 | 0.0021114 | *DNAJB12* |
| ENSG00000120334 | 1 | 173758688 | 173803858 | 54 | 12 | 49784 | 2.8609 | 0.0021119 | *CENPL* |
| ENSG00000156381 | 14 | 102963179 | 102986136 | 90 | 16 | 47751 | 2.8594 | 0.002122 | *ANKRD9* |
| ENSG00000117593 | 1 | 173783641 | 173837684 | 90 | 16 | 47951 | 2.8593 | 0.0021229 | *DARS2* |
| ENSG00000170948 | 19 | 8933074 | 8964016 | 93 | 13 | 51078 | 2.8553 | 0.0021499 | *MBD3L1* |
| ENSG00000185294 | 17 | 43912256 | 43934438 | 162 | 7 | 25592 | 2.8504 | 0.0021831 | *SPPL2C* |
| ENSG00000128886 | 15 | 44054798 | 44079741 | 41 | 7 | 28714 | 2.8487 | 0.002195 | *ELL3* |
| ENSG00000215474 | 18 | 44736293 | 44785554 | 140 | 21 | 54355 | 2.8472 | 0.0022054 | *SKOR2* |
| ENSG00000136574 | 8 | 11524468 | 11627511 | 371 | 65 | 40593 | 2.8454 | 0.0022181 | *GATA4* |
| ENSG00000140455 | 15 | 63786793 | 63896839 | 250 | 28 | 44825 | 2.8437 | 0.0022298 | *USP3* |
| ENSG00000242028 | 15 | 44078340 | 44105241 | 34 | 6 | 31828 | 2.8435 | 0.002231 | *HYPK* |
| ENSG00000158445 | 20 | 47970414 | 48109184 | 488 | 73 | 48840 | 2.8415 | 0.0022452 | *KCNB1* |
| ENSG00000204618 | 6 | 30028043 | 30053664 | 194 | 23 | 39554 | 2.8402 | 0.0022543 | *RNF39* |
| ENSG00000117013 | 1 | 41239684 | 41316124 | 234 | 58 | 48713 | 2.8397 | 0.0022578 | *KCNQ4* |
| ENSG00000130349 | 6 | 107339407 | 107382546 | 198 | 25 | 29498 | 2.8387 | 0.0022649 | *C6orf203* |
| ENSG00000162753 | 1 | 173459603 | 173582233 | 292 | 22 | 32066 | 2.8385 | 0.0022666 | *SLC9C2* |
| ENSG00000155052 | 2 | 124772864 | 125682864 | 3130 | 222 | 48752 | 2.8363 | 0.0022816 | *CNTNAP5* |
| ENSG00000257218 | 12 | 120874241 | 120909389 | 117 | 20 | 42818 | 2.8352 | 0.0022899 | *GATC* |
| ENSG00000130779 | 12 | 122745979 | 122917179 | 350 | 34 | 41678 | 2.8325 | 0.0023095 | *CLIP1* |
| ENSG00000183454 | 16 | 9842376 | 10286611 | 2037 | 107 | 46192 | 2.8324 | 0.0023098 | *GRIN2A* |
| ENSG00000147655 | 8 | 108901544 | 109105913 | 612 | 69 | 41508 | 2.8313 | 0.0023181 | *RSPO2* |
| ENSG00000122386 | 16 | 3152561 | 3180518 | 53 | 11 | 50451 | 2.8308 | 0.0023217 | *ZNF205* |
| ENSG00000115233 | 2 | 162154549 | 162278228 | 211 | 25 | 48303 | 2.8289 | 0.0023355 | *PSMD14* |
| ENSG00000125851 | 20 | 17196752 | 17475223 | 1150 | 117 | 46689 | 2.8276 | 0.0023453 | *PCSK2* |
| ENSG00000129951 | 19 | 802518 | 831967 | 115 | 26 | 46016 | 2.8213 | 0.0023917 | *LPPR3* |
| ENSG00000146530 | 7 | 12360511 | 12453567 | 431 | 42 | 46455 | 2.8186 | 0.0024115 | *VWDE* |
| ENSG00000164163 | 4 | 146009084 | 146060331 | 51 | 16 | 37368 | 2.817 | 0.0024237 | *ABCE1* |
| ENSG00000174945 | 7 | 2709156 | 2825134 | 534 | 71 | 48618 | 2.8124 | 0.002459 | *AMZ1* |
| ENSG00000151379 | 2 | 17987763 | 18008368 | 82 | 17 | 34856 | 2.8114 | 0.002466 | *MSGN1* |
| ENSG00000167004 | 15 | 44028590 | 44075477 | 82 | 11 | 29876 | 2.8072 | 0.0024984 | *PDIA3* |
| ENSG00000158079 | 9 | 96783076 | 96882138 | 183 | 26 | 48696 | 2.8065 | 0.002504 | *PTPDC1* |
| ENSG00000213865 | 8 | 67569850 | 67607799 | 46 | 17 | 46602 | 2.8053 | 0.0025136 | *C8orf44* |
| ENSG00000173992 | 11 | 66350292 | 66383490 | 46 | 16 | 45605 | 2.8052 | 0.0025146 | *CCS* |
| ENSG00000170855 | 12 | 120871764 | 120894215 | 67 | 15 | 43830 | 2.803 | 0.0025317 | *TRIAP1* |
| ENSG00000146352 | 6 | 123307116 | 123404072 | 256 | 31 | 38161 | 2.8027 | 0.002534 | *CLVS2* |
| ENSG00000006071 | 11 | 17404432 | 17508449 | 430 | 62 | 45914 | 2.8002 | 0.0025535 | *ABCC8* |

| | | | | | | | | | |
|---|---|---|---|---|---|---|---|---|---|
| ENSG00000102891 | 16 | 56588961 | 56612869 | 115 | 28 | 46661 | 2.799 | 0.0025628 | *MT4* |
| ENSG00000153976 | 17 | 13389006 | 13515244 | 556 | 77 | 41977 | 2.7974 | 0.0025755 | *HS3ST3A1* |
| ENSG00000174547 | 11 | 66192546 | 66244209 | 68 | 16 | 52493 | 2.7974 | 0.002576 | *MRPL11* |
| ENSG00000176681 | 17 | 44360099 | 44425160 | 25 | 3 | 18783 | 2.7957 | 0.0025896 | *LRRC37A* |
| ENSG00000100601 | 14 | 78128747 | 78184363 | 301 | 28 | 51921 | 2.7943 | 0.0026003 | *ALKBH1* |
| ENSG00000196126 | 6 | 32536546 | 32567625 | 548 | 26 | 39798 | 2.7938 | 0.0026049 | *HLA-DRB1* |
| ENSG00000092621 | 1 | 120192421 | 120296838 | 236 | 42 | 45195 | 2.7933 | 0.0026088 | *PHGDH* |
| ENSG00000119401 | 9 | 119439581 | 119473579 | 93 | 17 | 46975 | 2.7915 | 0.0026229 | *TRIM32* |
| ENSG00000172059 | 2 | 10172976 | 10204963 | 118 | 22 | 52116 | 2.7905 | 0.0026314 | *KLF11* |
| ENSG00000160714 | 1 | 154511053 | 154541504 | 73 | 13 | 47203 | 2.7855 | 0.002672 | *UBE2Q1* |
| ENSG00000169594 | 15 | 83914655 | 83963466 | 78 | 11 | 41236 | 2.7822 | 0.0026996 | *BNC1* |
| ENSG00000183831 | 1 | 173568700 | 173649001 | 96 | 20 | 44580 | 2.7806 | 0.0027125 | *ANKRD45* |
| ENSG00000104805 | 19 | 49393307 | 49436629 | 190 | 29 | 42116 | 2.7801 | 0.0027174 | *NUCB1* |
| ENSG00000196666 | 11 | 47598198 | 47620746 | 33 | 14 | 45452 | 2.7799 | 0.002719 | *FAM180B* |
| ENSG00000162062 | 16 | 2500081 | 2524964 | 27 | 13 | 45589 | 2.7794 | 0.0027227 | *C16orf59* |
| ENSG00000197019 | 19 | 40917499 | 40941932 | 56 | 15 | 43651 | 2.7793 | 0.0027238 | *SERTAD1* |
| ENSG00000168061 | 11 | 64798373 | 64822300 | 68 | 10 | 30744 | 2.7788 | 0.0027281 | *SAC3D1* |
| ENSG00000111780 | 12 | 120866001 | 120907809 | 140 | 23 | 44488 | 2.7786 | 0.0027294 | *AL021546.6* |
| ENSG00000173894 | 17 | 77741931 | 77771782 | 58 | 24 | 49494 | 2.7781 | 0.0027339 | *CBX2* |
| ENSG00000161395 | 17 | 37817375 | 37863050 | 118 | 11 | 36030 | 2.7777 | 0.0027376 | *PGAP3* |
| ENSG00000140478 | 15 | 75565176 | 75596816 | 24 | 14 | 36354 | 2.7774 | 0.0027396 | *GOLGA6D* |
| ENSG00000100324 | 22 | 39785746 | 39843065 | 102 | 11 | 46329 | 2.7717 | 0.002788 | *TAB1* |
| ENSG00000171532 | 17 | 37749789 | 37776030 | 50 | 12 | 36965 | 2.7703 | 0.0027998 | *NEUROD2* |
| ENSG00000164485 | 6 | 137454968 | 137504785 | 156 | 22 | 39071 | 2.7696 | 0.0028062 | *IL22RA2* |
| ENSG00000137309 | 6 | 34194650 | 34224008 | 64 | 13 | 44456 | 2.7675 | 0.0028243 | *HMGA1* |
| ENSG00000124693 | 6 | 26021817 | 26042288 | 56 | 15 | 48342 | 2.7672 | 0.0028268 | *HIST1H3B* |
| ENSG00000096654 | 6 | 27408522 | 27450897 | 152 | 11 | 52663 | 2.7669 | 0.0028299 | *ZNF184* |
| ENSG00000141738 | 17 | 37884180 | 37913544 | 47 | 11 | 35380 | 2.7668 | 0.0028307 | *GRB7* |
| ENSG00000118900 | 16 | 4886666 | 4942361 | 219 | 31 | 50766 | 2.7662 | 0.0028358 | *UBN1* |
| ENSG00000090432 | 1 | 20815943 | 20844654 | 82 | 16 | 51706 | 2.7653 | 0.0028431 | *MUL1* |
| ENSG00000002549 | 4 | 17568815 | 17619595 | 168 | 8 | 50908 | 2.7653 | 0.0028439 | *LAP3* |
| ENSG00000102780 | 13 | 42604176 | 42840714 | 923 | 82 | 45377 | 2.7583 | 0.0029053 | *DGKH* |
| ENSG00000197361 | 15 | 63879552 | 63904627 | 55 | 14 | 45851 | 2.7581 | 0.0029067 | *FBXL22* |
| ENSG00000184709 | 9 | 140053210 | 140074503 | 28 | 9 | 41861 | 2.7545 | 0.0029395 | *LRRC26* |
| ENSG00000198053 | 20 | 1865154 | 1930543 | 271 | 42 | 44013 | 2.7525 | 0.002957 | *SIRPA* |
| ENSG00000183044 | 16 | 8758422 | 8888432 | 784 | 129 | 44418 | 2.7515 | 0.0029659 | *ABAT* |

| | | | | | | | | | |
|---|---|---|---|---|---|---|---|---|---|
| ENSG00000113578 | 5 | 141961743 | 142087617 | 402 | 76 | 38974 | 2.7508 | 0.0029725 | *FGF1* |
| ENSG00000156427 | 5 | 170836660 | 170894627 | 183 | 35 | 51070 | 2.7401 | 0.0030706 | *FGF18* |
| ENSG00000187616 | 9 | 136369708 | 136403734 | 100 | 24 | 51350 | 2.7382 | 0.0030887 | *TMEM8C* |
| ENSG00000266996 | 18 | 44532731 | 44554607 | 38 | 11 | 53398 | 2.7313 | 0.0031543 | *TCEB3CL2* |
| ENSG00000184304 | 14 | 30035687 | 30671104 | 1714 | 195 | 44146 | 2.7293 | 0.0031731 | *PRKD1* |
| ENSG00000237289 | 15 | 43875252 | 43907099 | 20 | 4 | 24950 | 2.7282 | 0.0031842 | *CKMT1B* |
| ENSG00000228696 | 17 | 44342150 | 44449130 | 83 | 4 | 17714 | 2.7279 | 0.0031874 | *ARL17B* |
| ENSG00000185252 | 22 | 20738405 | 20772745 | 128 | 20 | 43031 | 2.7256 | 0.0032091 | *ZNF74* |
| ENSG00000176495 | 11 | 59121885 | 59142921 | 74 | 9 | 55577 | 2.7253 | 0.0032125 | *OR5AN1* |
| ENSG00000083799 | 16 | 50765961 | 50845846 | 152 | 21 | 32908 | 2.7235 | 0.0032292 | *CYLD* |
| ENSG00000268488 | 15 | 44065041 | 44085231 | 31 | 5 | 29438 | 2.7233 | 0.0032319 | *AC018512.1* |
| ENSG00000249240 | 15 | 65124082 | 65233118 | 334 | 34 | 37913 | 2.7229 | 0.003236 | *AC069368.3* |
| ENSG00000183476 | 15 | 78360150 | 78407251 | 157 | 35 | 46689 | 2.7166 | 0.0032976 | *SH2D7* |
| ENSG00000138674 | 4 | 83729814 | 83832319 | 246 | 16 | 34831 | 2.7156 | 0.0033081 | *SEC31A* |
| ENSG00000189099 | 4 | 152188323 | 152222605 | 109 | 12 | 36243 | 2.7121 | 0.0033434 | *PRSS48* |
| ENSG00000124529 | 6 | 26017124 | 26037480 | 58 | 15 | 47907 | 2.712 | 0.0033442 | *HIST1H4B* |
| ENSG00000242866 | 15 | 43881596 | 44020458 | 170 | 20 | 26083 | 2.7103 | 0.0033612 | *STRC* |
| ENSG00000135535 | 6 | 109677717 | 109713762 | 118 | 15 | 43956 | 2.7092 | 0.0033726 | *CD164* |
| ENSG00000166268 | 12 | 70209084 | 70362877 | 678 | 74 | 45881 | 2.7078 | 0.003387 | *MYRFL* |
| ENSG00000132694 | 1 | 156894632 | 157025162 | 327 | 28 | 51417 | 2.7065 | 0.0034002 | *ARHGEF11* |
| ENSG00000186868 | 17 | 43961748 | 44115700 | 919 | 9 | 20191 | 2.7028 | 0.0034381 | *MAPT* |
| ENSG00000163602 | 3 | 72410976 | 72506069 | 142 | 35 | 51361 | 2.7016 | 0.0034499 | *RYBP* |
| ENSG00000196167 | 11 | 111154114 | 111185770 | 113 | 20 | 46647 | 2.7004 | 0.0034624 | *COLCA1* |
| ENSG00000176540 | 11 | 48377037 | 48398017 | 38 | 9 | 43326 | 2.7003 | 0.003464 | *OR4C5* |
| ENSG00000145604 | 5 | 36142091 | 36194421 | 137 | 31 | 32858 | 2.6996 | 0.0034708 | *SKP2* |
| ENSG00000137413 | 6 | 42008251 | 42065199 | 240 | 49 | 49880 | 2.699 | 0.0034775 | *TAF8* |
| ENSG00000170791 | 8 | 57114245 | 57141357 | 30 | 16 | 52351 | 2.6978 | 0.0034902 | *CHCHD7* |
| ENSG00000157578 | 21 | 40767770 | 40827731 | 184 | 16 | 50998 | 2.6967 | 0.0035015 | *LCA5L* |
| ENSG00000176160 | 17 | 56487528 | 56575745 | 147 | 20 | 53300 | 2.6966 | 0.0035026 | *HSF5* |
| ENSG00000128944 | 15 | 40664922 | 40696447 | 84 | 14 | 52701 | 2.696 | 0.0035089 | *KNSTRN* |
| ENSG00000268175 | 3 | 149679066 | 149700535 | 52 | 14 | 43690 | 2.6952 | 0.003517 | *AC117395.1* |
| ENSG00000197912 | 16 | 89547325 | 89634176 | 306 | 19 | 52318 | 2.6938 | 0.0035326 | *SPG7* |
| ENSG00000204977 | 13 | 50560024 | 50604617 | 91 | 11 | 48787 | 2.6921 | 0.0035502 | *TRIM13* |
| ENSG00000186714 | 11 | 32613792 | 32826204 | 746 | 43 | 51001 | 2.6915 | 0.0035567 | *CCDC73* |
| ENSG00000131876 | 15 | 101811715 | 101845487 | 93 | 18 | 44155 | 2.6909 | 0.0035625 | *SNRPA1* |
| ENSG00000160298 | 21 | 47710095 | 47753789 | 202 | 26 | 41878 | 2.6882 | 0.0035916 | *C21orf58* |

| | | | | | | | | | |
|---|---|---|---|---|---|---|---|---|---|
| ENSG00000115380 | 2 | 56083102 | 56161274 | 271 | 42 | 40943 | 2.687 | 0.0036051 | *EFEMP1* |
| ENSG00000112742 | 6 | 80703604 | 80762244 | 248 | 16 | 53732 | 2.6847 | 0.0036298 | *TTK* |
| ENSG00000106638 | 7 | 72973262 | 73003121 | 73 | 15 | 53546 | 2.6826 | 0.0036525 | *TBL2* |
| ENSG00000055917 | 2 | 20438452 | 20561995 | 295 | 29 | 53086 | 2.6824 | 0.0036553 | *PUM2* |
| ENSG00000110002 | 11 | 123976069 | 124028428 | 174 | 36 | 47364 | 2.6799 | 0.0036821 | *VWA5A* |
| ENSG00000073331 | 4 | 113196665 | 113373776 | 550 | 93 | 46456 | 2.6768 | 0.0037161 | *ALPK1* |
| ENSG00000189043 | 7 | 10961578 | 10989883 | 107 | 21 | 38042 | 2.6747 | 0.0037403 | *NDUFA4* |
| ENSG00000155367 | 1 | 113242616 | 113268099 | 58 | 18 | 51492 | 2.6717 | 0.0037734 | *PPM1J* |
| ENSG00000124782 | 6 | 7097830 | 7262213 | 365 | 56 | 37359 | 2.6699 | 0.0037937 | *RREB1* |
| ENSG00000123908 | 8 | 141531264 | 141655718 | 408 | 64 | 44980 | 2.6699 | 0.0037941 | *AGO2* |
| ENSG00000157657 | 9 | 116628562 | 116828871 | 624 | 76 | 47509 | 2.6685 | 0.0038098 | *ZNF618* |
| ENSG00000109819 | 4 | 23746664 | 23915712 | 589 | 121 | 48214 | 2.6684 | 0.0038102 | *PPARGC1A* |
| ENSG00000081800 | 7 | 122743585 | 122850040 | 275 | 35 | 54646 | 2.6677 | 0.0038191 | *SLC13A1* |
| ENSG00000171451 | 18 | 65163819 | 65194217 | 85 | 19 | 51667 | 2.6673 | 0.0038234 | *DSEL* |
| ENSG00000166503 | 15 | 83774320 | 83886770 | 200 | 19 | 52493 | 2.6658 | 0.0038398 | *HDGFRP3* |
| ENSG00000240694 | 8 | 26352202 | 26381608 | 120 | 19 | 42025 | 2.6632 | 0.0038698 | *PNMA2* |
| ENSG00000140955 | 16 | 84214744 | 84240774 | 199 | 28 | 53393 | 2.6618 | 0.0038863 | *ADAD2* |
| ENSG00000164924 | 8 | 101918753 | 101975616 | 160 | 23 | 49061 | 2.6609 | 0.0038971 | *YWHAZ* |
| ENSG00000049239 | 1 | 9284834 | 9341396 | 222 | 38 | 48999 | 2.6592 | 0.0039162 | *H6PD* |
| ENSG00000223638 | 19 | 56260380 | 56284541 | 82 | 17 | 47865 | 2.6566 | 0.0039468 | *RFPL4A* |
| ENSG00000198535 | 15 | 62349176 | 62373116 | 80 | 9 | 48225 | 2.6562 | 0.0039514 | *C2CD4A* |
| ENSG00000186687 | 5 | 130496503 | 130551119 | 145 | 15 | 33395 | 2.6528 | 0.0039918 | *LYRM7* |
| ENSG00000268080 | 8 | 94231867 | 94252388 | 50 | 12 | 55670 | 2.6518 | 0.0040028 | *AC016885.1* |
| ENSG00000128245 | 22 | 32330447 | 32363590 | 135 | 24 | 45318 | 2.6516 | 0.0040052 | *YWHAH* |
| ENSG00000138653 | 4 | 115738919 | 116045032 | 835 | 75 | 32720 | 2.6493 | 0.0040327 | *NDST4* |
| ENSG00000011105 | 12 | 3176521 | 3405730 | 1089 | 152 | 48806 | 2.6484 | 0.0040434 | *TSPAN9* |
| ENSG00000175175 | 17 | 56823230 | 57068983 | 430 | 37 | 47258 | 2.6479 | 0.0040499 | *PPM1E* |
| ENSG00000187486 | 11 | 17397406 | 17420878 | 93 | 16 | 45013 | 2.6459 | 0.0040741 | *KCNJ11* |
| ENSG00000002822 | 7 | 1845429 | 2282878 | 2053 | 79 | 43041 | 2.6454 | 0.0040798 | *MAD1L1* |
| ENSG00000164776 | 7 | 56138440 | 56170689 | 124 | 19 | 54428 | 2.6436 | 0.004102 | *PHKG1* |
| ENSG00000185085 | 11 | 62404320 | 62430774 | 41 | 10 | 49929 | 2.6429 | 0.00411 | *INTS5* |
| ENSG00000036448 | 8 | 1983155 | 2123475 | 1231 | 136 | 41903 | 2.6425 | 0.0041151 | *MYOM2* |
| ENSG00000061455 | 5 | 122414816 | 122539960 | 517 | 60 | 50766 | 2.6387 | 0.004161 | *PRDM6* |
| ENSG00000178187 | 5 | 178358192 | 178403434 | 188 | 25 | 49121 | 2.6374 | 0.0041768 | *ZNF454* |
| ENSG00000141744 | 17 | 37814234 | 37836728 | 81 | 8 | 35325 | 2.6356 | 0.0041997 | *PNMT* |
| ENSG00000108384 | 17 | 56759934 | 56821703 | 138 | 20 | 42838 | 2.6346 | 0.0042116 | *RAD51C* |

| | | | | | | | | | |
|---|---|---|---|---|---|---|---|---|---|
| ENSG00000149948 | 12 | 66207911 | 66370075 | 283 | 58 | 40484 | 2.634 | 0.0042189 | *HMGA2* |
| ENSG00000174165 | 11 | 66278108 | 66323709 | 96 | 12 | 51906 | 2.6333 | 0.0042284 | *ZDHHC24* |
| ENSG00000204478 | 1 | 13726907 | 13757803 | 1 | 1 | 12885 | 2.6323 | 0.004241 | *PRAMEF20* |
| ENSG00000163875 | 1 | 37948176 | 37990375 | 134 | 20 | 49799 | 2.6273 | 0.0043031 | *MEAF6* |
| ENSG00000143226 | 1 | 161465220 | 161503803 | 129 | 25 | 46451 | 2.6269 | 0.0043088 | *FCGR2A* |
| ENSG00000267228 | 18 | 44651027 | 44712652 | 233 | 21 | 55111 | 2.6238 | 0.0043475 | *IER3IP1* |
| ENSG00000036549 | 1 | 78018101 | 78159104 | 269 | 29 | 33468 | 2.6231 | 0.0043563 | *ZZZ3* |
| ENSG00000183396 | 3 | 48648192 | 48669288 | 23 | 8 | 45802 | 2.6226 | 0.0043629 | *TMEM89* |
| ENSG00000189410 | 1 | 21036225 | 21069330 | 108 | 25 | 47347 | 2.622 | 0.004371 | *SH2D5* |
| ENSG00000267179 | 19 | 12025890 | 12100105 | 270 | 19 | 36198 | 2.6203 | 0.0043929 | *ZNF763* |
| ENSG00000167522 | 16 | 89324038 | 89566969 | 624 | 67 | 44886 | 2.6112 | 0.0045112 | *ANKRD11* |
| ENSG00000136404 | 15 | 83766159 | 83823606 | 89 | 16 | 50617 | 2.611 | 0.0045136 | *TM6SF1* |
| ENSG00000223417 | 11 | 89634279 | 89664935 | 23 | 6 | 46700 | 2.611 | 0.0045142 | *TRIM49D1* |
| ENSG00000164548 | 7 | 23534399 | 23581660 | 123 | 22 | 52630 | 2.6106 | 0.0045187 | *TRA2A* |
| ENSG00000125388 | 4 | 2955335 | 3052474 | 339 | 31 | 48548 | 2.6102 | 0.0045245 | *GRK4* |
| ENSG00000164270 | 5 | 147820595 | 148066798 | 728 | 80 | 45034 | 2.6099 | 0.0045288 | *HTR4* |
| ENSG00000197054 | 19 | 12025890 | 12101196 | 273 | 19 | 36105 | 2.6068 | 0.0045695 | *ZNF763* |
| ENSG00000173638 | 21 | 46903486 | 46974325 | 285 | 41 | 49503 | 2.6056 | 0.0045855 | *SLC19A1* |
| ENSG00000138050 | 2 | 39953200 | 40016407 | 176 | 20 | 48177 | 2.605 | 0.0045938 | *THUMPD2* |
| ENSG00000114439 | 3 | 107231783 | 107540171 | 645 | 65 | 42159 | 2.6029 | 0.004622 | *BBX* |
| ENSG00000085644 | 16 | 3169778 | 3202806 | 69 | 9 | 43815 | 2.6025 | 0.0046267 | *ZNF213* |
| ENSG00000268280 | 5 | 55214468 | 55234569 | 47 | 19 | 43851 | 2.6024 | 0.0046288 | *AC008914.1* |
| ENSG00000123810 | 19 | 41850326 | 41880078 | 82 | 12 | 52062 | 2.6008 | 0.0046501 | *B9D2* |
| ENSG00000256162 | 6 | 131138546 | 131168275 | 115 | 20 | 49833 | 2.6 | 0.0046618 | *SMLR1* |
| ENSG00000109686 | 4 | 152013903 | 152256784 | 582 | 56 | 34822 | 2.5986 | 0.0046801 | *SH3D19* |
| ENSG00000173702 | 3 | 124614289 | 124682663 | 239 | 46 | 52606 | 2.598 | 0.0046879 | *MUC13* |
| ENSG00000168152 | 4 | 83811837 | 83851122 | 85 | 10 | 33270 | 2.5964 | 0.0047098 | *THAP9* |
| ENSG00000196914 | 11 | 120197787 | 120370645 | 347 | 30 | 50876 | 2.5961 | 0.0047151 | *ARHGEF12* |
| ENSG00000118579 | 4 | 17606254 | 17645728 | 155 | 11 | 45261 | 2.5946 | 0.0047355 | *MED28* |
| ENSG00000116254 | 1 | 6151853 | 6250183 | 351 | 49 | 33972 | 2.5942 | 0.00474 | *CHD5* |
| ENSG00000132394 | 3 | 127862297 | 128137485 | 575 | 34 | 42564 | 2.5935 | 0.0047503 | *EEFSEC* |
| ENSG00000213903 | 14 | 24770656 | 24797242 | 68 | 16 | 44238 | 2.593 | 0.0047568 | *LTB4R* |
| ENSG00000167491 | 19 | 19486635 | 19629740 | 274 | 24 | 44526 | 2.5926 | 0.0047633 | *GATAD2A* |
| ENSG00000136854 | 9 | 130364544 | 130467460 | 129 | 25 | 42397 | 2.5914 | 0.004779 | *STXBP1* |
| ENSG00000268091 | 12 | 131504221 | 131524769 | 136 | 23 | 41006 | 2.5905 | 0.0047915 | *AC078925.1* |
| ENSG00000028277 | 19 | 42580263 | 42710737 | 158 | 13 | 53167 | 2.5882 | 0.0048232 | *POU2F2* |

| | | | | | | | | | |
|---|---|---|---|---|---|---|---|---|---|
| ENSG00000137944 | 1 | 89391456 | 89468636 | 159 | 18 | 44632 | 2.5882 | 0.0048243 | *CCBL2* |
| ENSG00000169989 | 4 | 153680506 | 153710916 | 81 | 15 | 53700 | 2.5877 | 0.0048313 | *TIGD4* |
| ENSG00000188596 | 12 | 96873349 | 97279333 | 1248 | 70 | 52091 | 2.5876 | 0.0048317 | *C12orf55* |
| ENSG00000109919 | 11 | 47628867 | 47674175 | 55 | 13 | 39747 | 2.5867 | 0.0048456 | *MTCH2* |
| ENSG00000185897 | 19 | 35839362 | 35861387 | 57 | 22 | 43142 | 2.5864 | 0.0048494 | *FFAR3* |
| ENSG00000221944 | 2 | 233402779 | 233425226 | 81 | 15 | 52258 | 2.5851 | 0.0048674 | *TIGD1* |
| ENSG00000077254 | 1 | 78151672 | 78235537 | 141 | 26 | 40697 | 2.583 | 0.0048966 | *USP33* |
| ENSG00000156509 | 8 | 101135588 | 101168028 | 58 | 8 | 53892 | 2.583 | 0.0048976 | *FBXO43* |
| ENSG00000144559 | 3 | 11821916 | 11898393 | 165 | 29 | 44129 | 2.5808 | 0.004928 | *TAMM41* |
| ENSG00000116863 | 1 | 36544476 | 36569533 | 29 | 11 | 47059 | 2.58 | 0.0049396 | *ADPRHL2* |
| ENSG00000182791 | 11 | 66347640 | 66370554 | 30 | 13 | 45726 | 2.5788 | 0.0049571 | *CCDC87* |
| ENSG00000145107 | 3 | 196036213 | 196075374 | 158 | 21 | 49245 | 2.5777 | 0.0049727 | *TM4SF19* |
| ENSG00000204540 | 6 | 31072527 | 31117869 | 619 | 63 | 41430 | 2.5736 | 0.0050318 | *PSORS1C1* |
| ENSG00000147689 | 8 | 124181200 | 124232314 | 165 | 18 | 53840 | 2.5726 | 0.0050472 | *FAM83A* |
| ENSG00000178573 | 16 | 79609740 | 79644611 | 120 | 36 | 49231 | 2.571 | 0.0050702 | *MAF* |
| ENSG00000174032 | 13 | 45957451 | 46002590 | 106 | 16 | 31997 | 2.5698 | 0.0050876 | *SLC25A30* |
| ENSG00000167965 | 16 | 2244249 | 2269417 | 67 | 12 | 39017 | 2.5686 | 0.0051061 | *MLST8* |
| ENSG00000109906 | 11 | 113920315 | 114131398 | 664 | 121 | 48559 | 2.5685 | 0.0051069 | *ZBTB16* |
| ENSG00000074800 | 1 | 8911061 | 8949308 | 79 | 11 | 37625 | 2.5672 | 0.0051269 | *ENO1* |
| ENSG00000177202 | 19 | 49100000 | 49120970 | 66 | 17 | 45454 | 2.5671 | 0.0051275 | *SPACA4* |
| ENSG00000120088 | 17 | 43689267 | 43923194 | 1188 | 14 | 21433 | 2.5632 | 0.005185 | *CRHR1* |
| ENSG00000128254 | 22 | 32319507 | 32351504 | 111 | 25 | 45370 | 2.5632 | 0.0051856 | *C22orf24* |
| ENSG00000115816 | 2 | 37418755 | 37468856 | 147 | 28 | 52951 | 2.5621 | 0.0052015 | *CEBPZ* |
| ENSG00000256349 | 11 | 66266550 | 66311084 | 103 | 9 | 52066 | 2.5594 | 0.0052428 | *CTD-3074O7.11* |
| ENSG00000186113 | 11 | 55553032 | 55573976 | 80 | 15 | 53770 | 2.558 | 0.0052636 | *OR5D14* |
| ENSG00000178057 | 3 | 49047892 | 49070928 | 21 | 10 | 44868 | 2.5555 | 0.005302 | *NDUFAF3* |
| ENSG00000111785 | 12 | 107158373 | 107293090 | 245 | 34 | 51882 | 2.5552 | 0.0053067 | *RIC8B* |
| ENSG00000173467 | 7 | 16889029 | 16931611 | 215 | 41 | 30771 | 2.554 | 0.0053244 | *AGR3* |
| ENSG00000141627 | 18 | 46560039 | 46997717 | 1349 | 56 | 45379 | 2.5523 | 0.0053501 | *DYM* |
| ENSG00000232423 | 1 | 12988181 | 13127751 | 16 | 6 | 42974 | 2.5523 | 0.005351 | *PRAMEF6* |
| ENSG00000149547 | 11 | 125429112 | 125464575 | 83 | 15 | 53822 | 2.5521 | 0.0053539 | *EI24* |
| ENSG00000198252 | 14 | 53186898 | 53251716 | 127 | 16 | 54172 | 2.5518 | 0.0053578 | *STYX* |
| ENSG00000129003 | 15 | 62134588 | 62362672 | 750 | 49 | 42015 | 2.5507 | 0.005375 | *VPS13C* |
| ENSG00000198231 | 17 | 61840963 | 61906677 | 147 | 12 | 31696 | 2.5494 | 0.0053956 | *DDX42* |
| ENSG00000131668 | 9 | 96703905 | 96727654 | 74 | 12 | 51437 | 2.5461 | 0.005447 | *BARX1* |
| ENSG00000113522 | 5 | 131881711 | 131990313 | 160 | 14 | 41533 | 2.5457 | 0.0054531 | *RAD50* |

| | | | | | | | | | |
|---|---|---|---|---|---|---|---|---|---|
| ENSG00000174483 | 11 | 66268077 | 66311098 | 101 | 9 | 51957 | 2.5432 | 0.0054925 | *BBS1* |
| ENSG00000108375 | 17 | 56419861 | 56504956 | 174 | 27 | 51018 | 2.542 | 0.0055118 | *RNF43* |
| ENSG00000147669 | 8 | 101152812 | 101176230 | 48 | 7 | 53837 | 2.5415 | 0.0055182 | *POLR2K* |
| ENSG00000111775 | 12 | 120865893 | 120888545 | 73 | 16 | 47999 | 2.5412 | 0.0055232 | *COX6A1* |
| ENSG00000188566 | 9 | 140090147 | 140121461 | 72 | 16 | 55236 | 2.5401 | 0.0055411 | *NDOR1* |
| ENSG00000172769 | 11 | 58159937 | 58180882 | 123 | 5 | 52916 | 2.5395 | 0.0055509 | *OR5B3* |
| ENSG00000063177 | 19 | 49108585 | 49132793 | 54 | 11 | 41144 | 2.5385 | 0.0055672 | *RPL18* |
| ENSG00000196811 | 2 | 233394437 | 233421113 | 87 | 16 | 52453 | 2.5368 | 0.0055932 | *CHRNG* |
| ENSG00000173653 | 11 | 66600306 | 66624017 | 30 | 14 | 47593 | 2.5368 | 0.0055936 | *RCE1* |
| ENSG00000106443 | 7 | 11003499 | 11219250 | 638 | 48 | 44376 | 2.5365 | 0.0055988 | *PHF14* |
| ENSG00000196839 | 20 | 43238163 | 43290874 | 145 | 34 | 44991 | 2.5358 | 0.0056101 | *ADA* |
| ENSG00000125695 | 17 | 61770205 | 61839457 | 155 | 14 | 34068 | 2.5353 | 0.005617 | *RP11-51F16.8* |
| ENSG00000174516 | 11 | 66224216 | 66254808 | 46 | 8 | 52371 | 2.5351 | 0.0056205 | *PELI3* |
| ENSG00000164049 | 3 | 48403709 | 48452666 | 100 | 7 | 50283 | 2.5322 | 0.0056676 | *FBXW12* |
| ENSG00000162545 | 1 | 20798884 | 20822713 | 47 | 13 | 48135 | 2.5309 | 0.0056886 | *CAMK2N1* |
| ENSG00000187048 | 1 | 47384849 | 47417156 | 85 | 10 | 34997 | 2.5307 | 0.0056913 | *CYP4A11* |
| ENSG00000164050 | 3 | 48435261 | 48481594 | 58 | 8 | 53504 | 2.5305 | 0.0056953 | *PLXNB1* |
| ENSG00000148516 | 10 | 31597424 | 31828742 | 253 | 32 | 40523 | 2.53 | 0.0057032 | *ZEB1* |
| ENSG00000164076 | 3 | 49885421 | 49917655 | 51 | 11 | 38665 | 2.5298 | 0.0057059 | *CAMKV* |
| ENSG00000167968 | 16 | 2275817 | 2298712 | 47 | 15 | 48967 | 2.5244 | 0.0057951 | *DNASE1L2* |
| ENSG00000230301 | 3 | 97973088 | 97994200 | 164 | 8 | 22351 | 2.5241 | 0.0058003 | *OR5H6* |
| ENSG00000101440 | 20 | 32772375 | 32867150 | 152 | 14 | 39326 | 2.5233 | 0.0058122 | *ASIP* |
| ENSG00000104450 | 8 | 101160134 | 101281506 | 259 | 23 | 53601 | 2.5229 | 0.0058194 | *SPAG1* |
| ENSG00000129595 | 5 | 111468138 | 111765013 | 952 | 106 | 46152 | 2.5218 | 0.0058386 | *EPB41L4A* |
| ENSG00000196661 | 11 | 124256278 | 124277264 | 66 | 8 | 46393 | 2.5209 | 0.0058531 | *OR8B3* |
| ENSG00000075399 | 16 | 89763542 | 89797394 | 126 | 34 | 36986 | 2.5207 | 0.0058569 | *VPS9D1* |
| ENSG00000196341 | 11 | 124169708 | 124190733 | 71 | 11 | 39409 | 2.5204 | 0.0058617 | *OR8D1* |
| ENSG00000197249 | 14 | 94833084 | 94867030 | 168 | 32 | 45107 | 2.5188 | 0.0058883 | *SERPINA1* |
| ENSG00000172057 | 17 | 38067294 | 38093854 | 75 | 11 | 44059 | 2.5181 | 0.0058997 | *ORMDL3* |
| ENSG00000164144 | 4 | 153691089 | 153849615 | 398 | 34 | 51329 | 2.518 | 0.0059007 | *ARFIP1* |
| ENSG00000197841 | 19 | 35215060 | 35243777 | 83 | 18 | 51897 | 2.5178 | 0.0059049 | *ZNF181* |
| ENSG00000142046 | 19 | 41846816 | 41899988 | 124 | 16 | 51372 | 2.5156 | 0.0059419 | *TMEM91* |
| ENSG00000084093 | 4 | 57764075 | 57812010 | 165 | 22 | 51157 | 2.5132 | 0.0059814 | *REST* |
| ENSG00000101384 | 20 | 10608332 | 10664694 | 175 | 44 | 45297 | 2.5128 | 0.0059885 | *JAG1* |
| ENSG00000186075 | 17 | 38014417 | 38044149 | 98 | 9 | 43276 | 2.5122 | 0.0059995 | *ZPBP2* |
| ENSG00000064726 | 15 | 83675174 | 83746106 | 203 | 24 | 53516 | 2.5103 | 0.0060312 | *BTBD1* |

| | | | | | | | | | |
|---|---|---|---|---|---|---|---|---|---|
| ENSG00000189308 | 4 | 83821126 | 83944079 | 306 | 16 | 34177 | 2.5078 | 0.0060741 | *LIN54* |
| ENSG00000213809 | 12 | 10514952 | 10554473 | 111 | 15 | 35989 | 2.504 | 0.0061406 | *KLRK1* |
| ENSG00000171643 | 5 | 76135826 | 76227475 | 229 | 37 | 31751 | 2.5039 | 0.0061418 | *S100Z* |
| ENSG00000272658 | 14 | 24768161 | 24791252 | 56 | 12 | 46595 | 2.5029 | 0.0061583 | *LTB4R2* |
| ENSG00000118402 | 6 | 80614529 | 80667297 | 203 | 21 | 37036 | 2.5023 | 0.00617 | *ELOVL4* |
| ENSG00000198865 | 5 | 42746903 | 42812462 | 215 | 19 | 52835 | 2.5014 | 0.0061854 | *CCDC152* |
| ENSG00000070193 | 5 | 44293646 | 44399808 | 200 | 20 | 44311 | 2.5013 | 0.0061867 | *FGF10* |
| ENSG00000132825 | 20 | 58501894 | 58525352 | 33 | 14 | 52210 | 2.5008 | 0.0061959 | *PPP1R3D* |
| ENSG00000100519 | 14 | 53163890 | 53205305 | 74 | 11 | 55606 | 2.4979 | 0.006247 | *PSMC6* |
| ENSG00000145569 | 5 | 14571884 | 14625116 | 94 | 23 | 47543 | 2.4971 | 0.00626 | *FAM105A* |
| ENSG00000113209 | 5 | 140504800 | 140527703 | 60 | 18 | 51074 | 2.495 | 0.0062984 | *PCDHB5* |
| ENSG00000266173 | 17 | 61770192 | 61829330 | 132 | 13 | 33899 | 2.4934 | 0.0063266 | *STRADA* |
| ENSG00000185100 | 14 | 105180523 | 105223662 | 161 | 30 | 47335 | 2.4911 | 0.006367 | *ADSSL1* |
| ENSG00000169621 | 2 | 68684693 | 68868004 | 581 | 32 | 43887 | 2.4904 | 0.0063791 | *APLF* |
| ENSG00000273331 | 3 | 196032953 | 196075244 | 166 | 22 | 49485 | 2.4903 | 0.0063814 | *TM4SF19* |
| ENSG00000131351 | 19 | 17150539 | 17196435 | 184 | 21 | 51482 | 2.4897 | 0.006393 | *HAUS8* |
| ENSG00000105523 | 19 | 49094067 | 49128111 | 116 | 24 | 42494 | 2.4893 | 0.0063989 | *FAM83E* |
| ENSG00000100599 | 14 | 92970118 | 93165339 | 701 | 126 | 48104 | 2.4891 | 0.0064035 | *RIN3* |
| ENSG00000166037 | 11 | 95513129 | 95575857 | 230 | 20 | 34307 | 2.4883 | 0.0064176 | *CEP57* |
| ENSG00000163406 | 3 | 121602936 | 121672949 | 261 | 11 | 53144 | 2.4875 | 0.0064328 | *SLC15A2* |
| ENSG00000269601 | 11 | 120319880 | 120340047 | 43 | 12 | 49899 | 2.4871 | 0.0064389 | *AP000758.1* |
| ENSG00000255408 | 5 | 140170783 | 140401929 | 516 | 42 | 53965 | 2.4834 | 0.0065073 | *PCDHA3* |
| ENSG00000204967 | 5 | 140176659 | 140401929 | 504 | 42 | 53905 | 2.4817 | 0.0065374 | *PCDHA4* |
| ENSG00000167378 | 19 | 44078521 | 44110287 | 80 | 22 | 50938 | 2.4812 | 0.0065475 | *IRGQ* |
| ENSG00000120327 | 5 | 140592931 | 140615858 | 80 | 14 | 46808 | 2.4809 | 0.0065531 | *PCDHB14* |
| ENSG00000120952 | 1 | 12906941 | 12931764 | 104 | 8 | 37192 | 2.4803 | 0.0065629 | *PRAMEF2* |
| ENSG00000204969 | 5 | 140164444 | 140401929 | 534 | 42 | 54089 | 2.4792 | 0.0065839 | *PCDHA2* |
| ENSG00000164209 | 5 | 110063837 | 110110857 | 120 | 23 | 41227 | 2.479 | 0.0065866 | *SLC25A46* |
| ENSG00000185097 | 1 | 611059 | 632053 | 1 | 1 | 42765 | 2.4785 | 0.006596 | *OR4F16* |
| ENSG00000129514 | 14 | 38049189 | 38079245 | 75 | 18 | 48019 | 2.4785 | 0.0065973 | *FOXA1* |
| ENSG00000254986 | 11 | 66237484 | 66287130 | 105 | 10 | 52209 | 2.4781 | 0.0066035 | *DPP3* |
| ENSG00000156515 | 10 | 71019740 | 71171638 | 608 | 101 | 45003 | 2.4752 | 0.0066575 | *HK1* |
| ENSG00000174720 | 4 | 113548120 | 113588748 | 104 | 22 | 47062 | 2.475 | 0.0066624 | *LARP7* |
| ENSG00000198574 | 1 | 162355056 | 162391928 | 142 | 17 | 52143 | 2.4728 | 0.0067023 | *SH2D1B* |
| ENSG00000173991 | 17 | 37810440 | 37832808 | 74 | 10 | 41120 | 2.4704 | 0.0067482 | *TCAP* |
| ENSG00000204970 | 5 | 140155876 | 140401929 | 555 | 43 | 54113 | 2.4676 | 0.0068016 | *PCDHA1* |

| | | | | | | | | | |
|---|---|---|---|---|---|---|---|---|---|
| ENSG00000204965 | 5 | 140191222 | 140401929 | 460 | 39 | 53652 | 2.4664 | 0.0068233 | *PCDHA5* |
| ENSG00000130227 | 8 | 21767180 | 21874096 | 273 | 23 | 39294 | 2.466 | 0.0068313 | *XPO7* |
| ENSG00000164024 | 4 | 99906771 | 99993964 | 247 | 25 | 51178 | 2.4657 | 0.0068364 | *METAP1* |
| ENSG00000149089 | 11 | 34864641 | 34948046 | 482 | 24 | 31058 | 2.4643 | 0.0068636 | *APIP* |
| ENSG00000047315 | 4 | 57833888 | 57907334 | 273 | 36 | 54372 | 2.4606 | 0.0069347 | *POLR2B* |
| ENSG00000073578 | 5 | 208356 | 266815 | 356 | 9 | 23225 | 2.46 | 0.006946 | *SDHA* |
| ENSG00000077458 | 11 | 95492106 | 95533573 | 137 | 16 | 35017 | 2.4569 | 0.0070079 | *FAM76B* |
| ENSG00000168685 | 5 | 35842797 | 35889705 | 179 | 17 | 53307 | 2.4566 | 0.0070137 | *IL7R* |
| ENSG00000149929 | 16 | 29993645 | 30017757 | 50 | 9 | 34997 | 2.4565 | 0.0070144 | *HIRIP3* |
| ENSG00000122367 | 10 | 88418206 | 88505825 | 362 | 58 | 46050 | 2.4563 | 0.007018 | *LDB3* |
| ENSG00000162747 | 1 | 161582986 | 161611753 | 41 | 12 | 38291 | 2.4562 | 0.00702 | *FCGR3B* |
| ENSG00000163646 | 3 | 150633950 | 150700786 | 208 | 39 | 42033 | 2.4547 | 0.0070501 | *CLRN1* |
| ENSG00000100577 | 14 | 77777227 | 77807940 | 117 | 27 | 53380 | 2.4541 | 0.0070622 | *GSTZ1* |
| ENSG00000081842 | 5 | 140197563 | 140401929 | 441 | 37 | 53613 | 2.4496 | 0.0071514 | *PCDHA6* |
| ENSG00000122547 | 7 | 36182758 | 36351152 | 699 | 97 | 48455 | 2.4494 | 0.007154 | *EEPD1* |
| ENSG00000084636 | 1 | 32107848 | 32179920 | 156 | 31 | 38212 | 2.4478 | 0.0071866 | *COL16A1* |
| ENSG00000204963 | 5 | 140203969 | 140401929 | 431 | 36 | 53522 | 2.4476 | 0.0071899 | *PCDHA7* |
| ENSG00000175348 | 11 | 8958841 | 8996558 | 134 | 15 | 44922 | 2.4433 | 0.0072761 | *TMEM9B* |
| ENSG00000184599 | 1 | 113253041 | 113279857 | 50 | 16 | 51808 | 2.4433 | 0.0072772 | *FAM19A3* |
| ENSG00000174236 | 12 | 27839428 | 27860566 | 45 | 10 | 52789 | 2.4415 | 0.0073141 | *REP15* |
| ENSG00000162723 | 1 | 159911282 | 159934044 | 73 | 18 | 48771 | 2.4408 | 0.0073275 | *SLAMF9* |
| ENSG00000184445 | 12 | 123001793 | 123120943 | 238 | 26 | 50708 | 2.4403 | 0.0073379 | *KNTC1* |
| ENSG00000160294 | 21 | 47645047 | 47716211 | 311 | 28 | 46064 | 2.4401 | 0.0073424 | *MCM3AP* |
| ENSG00000204962 | 5 | 140210907 | 140401929 | 423 | 35 | 53583 | 2.4379 | 0.0073864 | *PCDHA8* |
| ENSG00000227839 | 1 | 50449990 | 50471874 | 18 | 8 | 49212 | 2.4375 | 0.0073937 | *AL645730.2* |
| ENSG00000112576 | 6 | 41892671 | 42028095 | 466 | 85 | 47283 | 2.4374 | 0.0073957 | *CCND3* |
| ENSG00000074054 | 2 | 122085352 | 122417163 | 856 | 31 | 35559 | 2.4359 | 0.0074272 | *CLASP1* |
| ENSG00000134882 | 13 | 99843028 | 100048688 | 591 | 49 | 41818 | 2.4351 | 0.0074435 | *UBAC2* |
| ENSG00000148053 | 9 | 87273466 | 87648505 | 1121 | 131 | 49375 | 2.4338 | 0.0074697 | *NTRK2* |
| ENSG00000104804 | 19 | 49374222 | 49411990 | 191 | 18 | 35598 | 2.4337 | 0.0074726 | *TULP2* |
| ENSG00000123106 | 12 | 28276182 | 28742883 | 1388 | 40 | 42166 | 2.433 | 0.0074869 | *CCDC91* |
| ENSG00000255298 | 11 | 124124723 | 124145763 | 176 | 8 | 52810 | 2.4325 | 0.0074972 | *OR8G5* |
| ENSG00000103707 | 15 | 65284845 | 65331977 | 144 | 27 | 39758 | 2.4305 | 0.007538 | *MTFMT* |
| ENSG00000135074 | 5 | 156812542 | 157012783 | 803 | 65 | 46868 | 2.4301 | 0.0075468 | *ADAM19* |
| ENSG00000168060 | 11 | 64802290 | 64836021 | 88 | 17 | 38273 | 2.4297 | 0.0075559 | *NAALADL1* |
| ENSG00000204293 | 11 | 124242291 | 124263258 | 91 | 6 | 52340 | 2.4279 | 0.0075941 | *OR8B2* |

| | | | | | | | | | |
|---|---|---|---|---|---|---|---|---|---|
| ENSG00000179172 | 1 | 12897261 | 12918578 | 71 | 5 | 38716 | 2.4274 | 0.0076046 | *HNRNPCL1* |
| ENSG00000172464 | 11 | 56398942 | 56419947 | 74 | 16 | 56073 | 2.4271 | 0.0076103 | *OR5AP2* |
| ENSG00000204961 | 5 | 140217048 | 140401929 | 409 | 34 | 53691 | 2.4236 | 0.0076842 | *PCDHA9* |
| ENSG00000135541 | 6 | 135594670 | 135828914 | 453 | 63 | 42805 | 2.4204 | 0.0077518 | *AHI1* |
| ENSG00000178149 | 3 | 49042921 | 49069726 | 22 | 9 | 43445 | 2.42 | 0.0077609 | *DALRD3* |
| ENSG00000161572 | 17 | 34251522 | 34280703 | 140 | 17 | 48752 | 2.4196 | 0.0077693 | *LYZL6* |
| ENSG00000055609 | 7 | 151822010 | 152143090 | 387 | 35 | 32583 | 2.4194 | 0.0077732 | *KMT2C* |
| ENSG00000114861 | 3 | 70993844 | 71643140 | 1433 | 171 | 42500 | 2.4193 | 0.0077749 | *FOXP1* |
| ENSG00000146285 | 6 | 108015308 | 108155521 | 511 | 61 | 44276 | 2.4178 | 0.0078071 | *SCML4* |
| ENSG00000134240 | 1 | 120280619 | 120321528 | 109 | 21 | 52590 | 2.4177 | 0.007809 | *HMGCS2* |
| ENSG00000197451 | 5 | 177621508 | 177648164 | 116 | 27 | 50722 | 2.4173 | 0.007819 | *HNRNPAB* |
| ENSG00000065371 | 3 | 123677368 | 123721025 | 82 | 24 | 42192 | 2.4172 | 0.00782 | *ROPN1* |
| ENSG00000168781 | 15 | 43815660 | 43892430 | 50 | 13 | 41788 | 2.4172 | 0.0078207 | *PPIP5K1* |
| ENSG00000149541 | 11 | 62372768 | 62399647 | 34 | 7 | 51635 | 2.4171 | 0.0078225 | *B3GAT3* |
| ENSG00000178394 | 5 | 63246183 | 63268334 | 53 | 13 | 53232 | 2.4163 | 0.0078388 | *HTR1A* |
| ENSG00000174106 | 12 | 65553351 | 65652107 | 137 | 22 | 45018 | 2.4146 | 0.0078757 | *LEMD3* |
| ENSG00000185158 | 17 | 30324891 | 30390523 | 113 | 13 | 51751 | 2.4116 | 0.0079417 | *LRRC37B* |
| ENSG00000250120 | 5 | 140225595 | 140401929 | 396 | 33 | 53715 | 2.4113 | 0.0079474 | *PCDHA10* |
| ENSG00000156110 | 10 | 75900960 | 76479061 | 1045 | 43 | 35472 | 2.4108 | 0.0079588 | *ADK* |
| ENSG00000100888 | 14 | 21843353 | 21934285 | 187 | 24 | 41743 | 2.4107 | 0.0079615 | *CHD8* |
| ENSG00000204538 | 6 | 31095313 | 31117127 | 275 | 40 | 40433 | 2.4103 | 0.0079705 | *PSORS1C2* |
| ENSG00000251664 | 5 | 140245058 | 140401929 | 364 | 30 | 53869 | 2.41 | 0.0079752 | *PCDHA12* |
| ENSG00000127663 | 19 | 4959125 | 5163606 | 697 | 74 | 48335 | 2.4086 | 0.0080079 | *KDM4B* |
| ENSG00000164082 | 3 | 51731086 | 51762629 | 33 | 16 | 49837 | 2.407 | 0.0080412 | *GRM2* |
| ENSG00000100580 | 14 | 77791364 | 77853452 | 186 | 28 | 46566 | 2.407 | 0.0080415 | *TMED8* |
| ENSG00000103449 | 16 | 51159886 | 51195278 | 69 | 17 | 40486 | 2.4061 | 0.0080615 | *SALL1* |
| ENSG00000248383 | 5 | 140296302 | 140401929 | 277 | 24 | 53272 | 2.4018 | 0.0081568 | *PCDHAC1* |
| ENSG00000105723 | 19 | 42724338 | 42756777 | 44 | 6 | 52433 | 2.4008 | 0.0081798 | *GSK3A* |
| ENSG00000249034 | 5 | 140230341 | 140253224 | 43 | 11 | 53257 | 2.4005 | 0.0081858 | *AC005609.1* |
| ENSG00000255819 | 12 | 10514952 | 10572745 | 147 | 18 | 37842 | 2.4004 | 0.0081884 | *KLRC4-KLRK1* |
| ENSG00000166762 | 15 | 43910701 | 43970316 | 99 | 13 | 22565 | 2.3987 | 0.0082271 | *CATSPER2* |
| ENSG00000142609 | 1 | 1843396 | 1945276 | 365 | 43 | 45168 | 2.3983 | 0.0082361 | *C1orf222* |
| ENSG00000239389 | 5 | 140251793 | 140401929 | 355 | 29 | 53907 | 2.3976 | 0.008252 | *PCDHA13* |
| ENSG00000249158 | 5 | 140238689 | 140401929 | 384 | 31 | 54022 | 2.3957 | 0.0082934 | *PCDHA11* |
| ENSG00000100473 | 14 | 31333720 | 31374271 | 93 | 31 | 50224 | 2.3956 | 0.0082966 | *COCH* |
| ENSG00000204957 | 19 | 42736927 | 42759125 | 35 | 5 | 50320 | 2.3955 | 0.008298 | *AC006486.1* |

| | | | | | | | | | |
|---|---|---|---|---|---|---|---|---|---|
| ENSG00000100583 | 14 | 77833032 | 77867840 | 95 | 14 | 42114 | 2.3952 | 0.008306 | *SAMD15* |
| ENSG00000087074 | 19 | 49365649 | 49389314 | 109 | 13 | 29502 | 2.3947 | 0.0083165 | *PPP1R15A* |
| ENSG00000164105 | 4 | 174281120 | 174308683 | 37 | 8 | 30702 | 2.3939 | 0.008335 | *SAP30* |
| ENSG00000147443 | 8 | 21756384 | 21781371 | 79 | 20 | 43644 | 2.3939 | 0.008335 | *DOK2* |
| ENSG00000128283 | 22 | 37946454 | 37975412 | 96 | 19 | 38697 | 2.3937 | 0.0083393 | *CDC42EP1* |
| ENSG00000176009 | 11 | 8949119 | 8974580 | 102 | 12 | 48790 | 2.3937 | 0.0083405 | *ASCL3* |
| ENSG00000149489 | 11 | 62369194 | 62392592 | 33 | 5 | 47363 | 2.3929 | 0.0083589 | *ROM1* |
| ENSG00000166869 | 16 | 23755948 | 23780272 | 63 | 12 | 46399 | 2.3929 | 0.0083589 | *CHP2* |
| ENSG00000140479 | 15 | 101830818 | 102075405 | 1140 | 178 | 44046 | 2.3924 | 0.0083684 | *PCSK6* |
| ENSG00000137959 | 1 | 79075607 | 79121830 | 90 | 23 | 52093 | 2.3923 | 0.0083712 | *IFI44L* |
| ENSG00000197786 | 11 | 58115597 | 58136542 | 59 | 5 | 48604 | 2.3917 | 0.0083848 | *OR5B17* |
| ENSG00000108395 | 17 | 57049999 | 57194282 | 235 | 27 | 54422 | 2.3915 | 0.0083896 | *TRIM37* |
| ENSG00000163239 | 1 | 154464695 | 154530623 | 214 | 14 | 49441 | 2.391 | 0.0084002 | *TDRD10* |
| ENSG00000140522 | 15 | 89743100 | 89774982 | 94 | 17 | 51891 | 2.391 | 0.0084013 | *RLBP1* |
| ENSG00000165792 | 14 | 21447929 | 21475189 | 116 | 18 | 47790 | 2.3901 | 0.0084214 | *METTL17* |
| ENSG00000108272 | 17 | 34938228 | 34967235 | 73 | 14 | 40343 | 2.39 | 0.0084244 | *DHRS11* |
| ENSG00000204913 | 17 | 38087727 | 38111000 | 84 | 13 | 52799 | 2.3888 | 0.0084518 | *LRRC3C* |
| ENSG00000151445 | 14 | 77883018 | 77934295 | 145 | 17 | 41042 | 2.3882 | 0.0084658 | *VIPAS39* |
| ENSG00000188763 | 7 | 72838109 | 72860450 | 33 | 11 | 51777 | 2.3863 | 0.0085092 | *FZD9* |
| ENSG00000048342 | 4 | 15461489 | 15613180 | 365 | 52 | 48061 | 2.385 | 0.0085393 | *CC2D2A* |
| ENSG00000120137 | 5 | 167965500 | 168016605 | 84 | 14 | 49833 | 2.3846 | 0.0085492 | *PANK3* |
| ENSG00000135902 | 2 | 233380703 | 233411377 | 110 | 21 | 53176 | 2.3844 | 0.0085528 | *CHRND* |
| ENSG00000171735 | 1 | 6835384 | 7839766 | 3059 | 340 | 47578 | 2.3832 | 0.0085803 | *CAMTA1* |
| ENSG00000128335 | 22 | 36612256 | 36646000 | 174 | 27 | 40044 | 2.383 | 0.0085851 | *APOL2* |
| ENSG00000138621 | 15 | 75305896 | 75419803 | 308 | 29 | 34550 | 2.3805 | 0.0086451 | *PPCDC* |
| ENSG00000155463 | 14 | 23225731 | 23251007 | 80 | 22 | 42324 | 2.3804 | 0.0086462 | *OXA1L* |
| ENSG00000253903 | 8 | 26933694 | 26954766 | 113 | 19 | 44984 | 2.379 | 0.008681 | *RP11-521M14.2* |
| ENSG00000106991 | 9 | 130567291 | 130627035 | 138 | 36 | 35678 | 2.3763 | 0.0087445 | *ENG* |
| ENSG00000196227 | 20 | 58498819 | 58533735 | 56 | 14 | 51434 | 2.376 | 0.0087506 | *FAM217B* |
| ENSG00000126858 | 17 | 30459473 | 30590393 | 236 | 27 | 44534 | 2.3756 | 0.0087604 | *RHOT1* |
| ENSG00000164404 | 5 | 132186873 | 132212576 | 62 | 11 | 46586 | 2.3744 | 0.0087891 | *GDF9* |
| ENSG00000104177 | 15 | 48421625 | 48480714 | 9 | 5 | 44346 | 2.3732 | 0.0088181 | *MYEF2* |
| ENSG00000175104 | 11 | 36498577 | 36541822 | 100 | 19 | 49243 | 2.3714 | 0.0088599 | *TRAF6* |
| ENSG00000054523 | 1 | 10260863 | 10451661 | 727 | 36 | 54059 | 2.3701 | 0.0088924 | *KIF1B* |
| ENSG00000090487 | 15 | 65245362 | 65292648 | 179 | 21 | 41920 | 2.3682 | 0.0089366 | *SPG21* |
| ENSG00000143258 | 1 | 161119240 | 161145513 | 50 | 13 | 53565 | 2.3676 | 0.0089528 | *USP21* |

| | | | | | | | | | |
|---|---|---|---|---|---|---|---|---|---|
| ENSG00000162040 | 16 | 1951464 | 1978441 | 123 | 12 | 47548 | 2.3668 | 0.0089721 | *HS3ST6* |
| ENSG00000179399 | 13 | 92040929 | 93529490 | 4673 | 326 | 43889 | 2.3664 | 0.008982 | *GPC5* |
| ENSG00000196074 | 20 | 58428618 | 58518710 | 213 | 13 | 50643 | 2.3663 | 0.0089829 | *SYCP2* |
| ENSG00000240204 | 7 | 129132320 | 129162773 | 93 | 24 | 38459 | 2.3646 | 0.0090242 | *SMKR1* |
| ENSG00000186513 | 11 | 57947906 | 57968990 | 53 | 9 | 46990 | 2.3645 | 0.0090262 | *OR9Q2* |
| ENSG00000073605 | 17 | 38050848 | 38086107 | 95 | 9 | 40281 | 2.3638 | 0.0090439 | *GSDMB* |
| ENSG00000156113 | 10 | 78619359 | 79408353 | 2271 | 246 | 45275 | 2.3631 | 0.0090608 | *KCNMA1* |
| ENSG00000124299 | 19 | 33867856 | 34022700 | 740 | 76 | 52299 | 2.3629 | 0.0090674 | *PEPD* |
| ENSG00000149100 | 11 | 32595344 | 32637808 | 159 | 24 | 51935 | 2.3604 | 0.0091271 | *EIF3M* |
| ENSG00000063176 | 19 | 49112548 | 49143974 | 72 | 13 | 38087 | 2.3603 | 0.0091299 | *SPHK2* |
| ENSG00000167914 | 17 | 38109226 | 38144019 | 148 | 19 | 53895 | 2.3593 | 0.0091541 | *GSDMA* |
| ENSG00000107798 | 10 | 90963326 | 91184314 | 650 | 81 | 36929 | 2.3564 | 0.0092267 | *LIPA* |
| ENSG00000188549 | 15 | 40613653 | 40643168 | 91 | 23 | 43826 | 2.3559 | 0.0092389 | *C15orf52* |
| ENSG00000135744 | 1 | 230828269 | 230860043 | 162 | 22 | 35090 | 2.3539 | 0.0092892 | *AGT* |
| ENSG00000147324 | 8 | 8630864 | 8761155 | 654 | 64 | 46002 | 2.3532 | 0.009306 | *MFHAS1* |
| ENSG00000164405 | 5 | 132192252 | 132213723 | 56 | 11 | 46996 | 2.3526 | 0.0093201 | *UQCRQ* |
| ENSG00000268643 | 19 | 42728735 | 42769284 | 69 | 7 | 52329 | 2.3521 | 0.0093329 | *AC006486.9* |
| ENSG00000103375 | 16 | 25217052 | 25250261 | 125 | 19 | 47394 | 2.3521 | 0.009333 | *AQP8* |
| ENSG00000197837 | 12 | 14910933 | 14934065 | 95 | 13 | 55991 | 2.3504 | 0.0093763 | *HIST4H4* |
| ENSG00000164366 | 5 | 186986 | 228330 | 224 | 17 | 30607 | 2.3502 | 0.0093811 | *CCDC127* |
| ENSG00000213906 | 14 | 24764940 | 24791259 | 64 | 13 | 43764 | 2.3498 | 0.0093914 | *LTB4R2* |
| ENSG00000105058 | 19 | 16286191 | 16312857 | 87 | 15 | 44498 | 2.3495 | 0.0093982 | *FAM32A* |
| ENSG00000256453 | 5 | 140040379 | 140063171 | 43 | 9 | 54501 | 2.3492 | 0.009407 | *DND1* |
| ENSG00000105568 | 19 | 52683292 | 52740687 | 277 | 22 | 56177 | 2.3489 | 0.0094154 | *PPP2R1A* |
| ENSG00000166444 | 11 | 8704898 | 8942498 | 765 | 76 | 45704 | 2.3478 | 0.009443 | *ST5* |
| ENSG00000105722 | 19 | 42741724 | 42769309 | 54 | 5 | 52809 | 2.3464 | 0.0094775 | *ERF* |
| ENSG00000204219 | 1 | 23697554 | 23761233 | 157 | 29 | 32505 | 2.3462 | 0.0094823 | *TCEA3* |
| ENSG00000134294 | 12 | 46741972 | 46776650 | 56 | 12 | 35766 | 2.3455 | 0.009502 | *SLC38A2* |
| ENSG00000235878 | 21 | 47602391 | 47623673 | 102 | 21 | 46094 | 2.3453 | 0.009505 | *AP001468.1* |
| ENSG00000230055 | 17 | 36876488 | 36901297 | 60 | 18 | 44612 | 2.3448 | 0.0095175 | *CISD3* |
| ENSG00000172037 | 3 | 49148547 | 49180551 | 34 | 16 | 48382 | 2.3421 | 0.0095869 | *LAMB2* |
| ENSG00000185250 | 6 | 109701418 | 109772374 | 209 | 21 | 40358 | 2.341 | 0.0096164 | *PPIL6* |
| ENSG00000198553 | 13 | 50579390 | 50605058 | 52 | 8 | 51110 | 2.3405 | 0.00963 | *KCNRG* |
| ENSG00000170745 | 2 | 18049114 | 18552882 | 2150 | 203 | 43250 | 2.3397 | 0.0096487 | *KCNS3* |
| ENSG00000196226 | 6 | 26033455 | 26053885 | 58 | 13 | 42019 | 2.3393 | 0.0096599 | *HIST1H2BB* |
| ENSG00000119917 | 10 | 91077651 | 91110728 | 162 | 13 | 29575 | 2.3387 | 0.0096758 | *IFIT3* |

| | | | | | | | | | |
|---|---|---|---|---|---|---|---|---|---|
| ENSG00000186472 | 7 | 82373329 | 82802246 | 1422 | 107 | 45743 | 2.3385 | 0.0096804 | *PCLO* |
| ENSG00000124574 | 6 | 43385104 | 43428168 | 118 | 22 | 47202 | 2.3358 | 0.0097503 | *ABCC10* |
| ENSG00000159873 | 22 | 29158662 | 29195283 | 142 | 21 | 39804 | 2.3346 | 0.0097822 | *CCDC117* |
| ENSG00000160844 | 7 | 99788283 | 99879855 | 188 | 22 | 47718 | 2.333 | 0.0098249 | *GATS* |
| ENSG00000120314 | 5 | 140034261 | 140063709 | 53 | 9 | 53998 | 2.3322 | 0.0098441 | *WDR55* |
| ENSG00000149499 | 11 | 62359690 | 62390237 | 44 | 6 | 47844 | 2.3314 | 0.0098663 | *EML3* |
| ENSG00000125384 | 14 | 52771023 | 52805324 | 139 | 24 | 34282 | 2.3313 | 0.0098684 | *PTGER2* |
| ENSG00000058262 | 3 | 127760484 | 127800526 | 118 | 6 | 54710 | 2.3313 | 0.0098694 | *SEC61A1* |
| ENSG00000146733 | 7 | 56068744 | 56129297 | 196 | 27 | 53041 | 2.3306 | 0.0098866 | *PSPH* |
| ENSG00000166411 | 15 | 78413840 | 78474291 | 171 | 27 | 45346 | 2.3303 | 0.0098953 | *IDH3A* |
| ENSG00000185278 | 1 | 173827220 | 173882687 | 156 | 20 | 42099 | 2.3303 | 0.0098965 | *ZBTB37* |
| ENSG00000152518 | 2 | 43439541 | 43463748 | 53 | 17 | 41516 | 2.3294 | 0.0099197 | *ZFP36L2* |
| ENSG00000168229 | 14 | 52724431 | 52753442 | 119 | 22 | 51592 | 2.3288 | 0.0099335 | *PTGDR* |
| ENSG00000268465 | 19 | 48959094 | 48985688 | 70 | 11 | 27721 | 2.3275 | 0.0099685 | *CTC-273B12.7* |
| ENSG00000249915 | 5 | 261736 | 363971 | 373 | 31 | 37398 | 2.3274 | 0.0099713 | *PDCD6* |
| ENSG00000257207 | 2 | 111150511 | 111240652 | 2 | 1 | 43140 | 2.3273 | 0.0099758 | *LIMS3* |
| ENSG00000141750 | 17 | 37356789 | 37392125 | 69 | 19 | 46809 | 2.3269 | 0.0099844 | *STAC2* |
| ENSG00000170445 | 5 | 140042758 | 140081609 | 74 | 9 | 54424 | 2.3264 | 0.0099993 | *HARS* |
| ENSG00000168517 | 17 | 43228067 | 43257407 | 57 | 16 | 48555 | 2.3263 | 0.010002 | *HEXIM2* |
| ENSG00000055813 | 2 | 56401258 | 56623308 | 763 | 112 | 48842 | 2.3257 | 0.010016 | *CCDC85A* |
| ENSG00000240065 | 6 | 32801913 | 32837362 | 283 | 38 | 51629 | 2.3242 | 0.010057 | *PSMB9* |
| ENSG00000167625 | 19 | 42714423 | 42742353 | 28 | 6 | 52202 | 2.3241 | 0.010059 | *ZNF526* |
| ENSG00000056661 | 17 | 36880150 | 36916070 | 99 | 25 | 46728 | 2.3237 | 0.01007 | *PCGF2* |
| ENSG00000164053 | 3 | 48478114 | 48517115 | 66 | 7 | 48601 | 2.3234 | 0.010078 | *ATRIP* |
| ENSG00000248333 | 1 | 1560603 | 1600473 | 42 | 6 | 36140 | 2.3226 | 0.010101 | *CDK11B* |
| ENSG00000268156 | 14 | 77872741 | 77893115 | 51 | 11 | 43697 | 2.3221 | 0.010113 | *FKSG61* |
| ENSG00000101082 | 20 | 35230721 | 35284619 | 68 | 30 | 48584 | 2.32 | 0.010172 | *SLA2* |
| ENSG00000135912 | 2 | 219565568 | 219630139 | 135 | 7 | 46811 | 2.3195 | 0.010184 | *TTLL4* |
| ENSG00000237524 | 2 | 127646472 | 127669673 | 83 | 13 | 31958 | 2.3191 | 0.010194 | *AC114783.1* |
| ENSG00000204869 | 19 | 46533006 | 46590376 | 246 | 28 | 52944 | 2.3189 | 0.010199 | *IGFL4* |
| ENSG00000164051 | 3 | 48463574 | 48491866 | 58 | 8 | 48828 | 2.3189 | 0.0102 | *CCDC51* |
| ENSG00000043143 | 5 | 133850003 | 133928918 | 231 | 39 | 46869 | 2.3183 | 0.010218 | *JADE2* |
| ENSG00000232112 | 3 | 48471667 | 48495616 | 50 | 7 | 48018 | 2.3174 | 0.01024 | *TMA7* |
| ENSG00000148154 | 9 | 114649046 | 114707649 | 359 | 48 | 44877 | 2.3172 | 0.010246 | *UGCG* |
| ENSG00000161955 | 17 | 7451609 | 7474925 | 66 | 18 | 46283 | 2.317 | 0.010253 | *TNFSF13* |
| ENSG00000104055 | 15 | 43514793 | 43569055 | 83 | 10 | 31763 | 2.3154 | 0.010294 | *TGM5* |

| | | | | | | | | | |
|---|---|---|---|---|---|---|---|---|---|
| ENSG00000146966 | 7 | 140208220 | 140383793 | 425 | 45 | 39022 | 2.3146 | 0.010317 | *DENND2A* |
| ENSG00000135929 | 2 | 219636472 | 219690016 | 113 | 8 | 44398 | 2.3117 | 0.010398 | *CYP27A1* |
| ENSG00000228146 | 16 | 3184220 | 3209964 | 75 | 8 | 39541 | 2.3116 | 0.010399 | *CASP16* |
| ENSG00000151090 | 3 | 24148651 | 24546773 | 1280 | 199 | 47745 | 2.3113 | 0.010407 | *THRB* |
| ENSG00000108666 | 17 | 30641584 | 30687042 | 79 | 15 | 42989 | 2.3096 | 0.010455 | *C17orf75* |
| ENSG00000133112 | 13 | 45901008 | 45925505 | 84 | 13 | 30257 | 2.3089 | 0.010475 | *TPT1* |
| ENSG00000139629 | 12 | 51735031 | 51796651 | 162 | 41 | 46835 | 2.3085 | 0.010486 | *GALNT6* |
| ENSG00000179348 | 3 | 128188270 | 128222028 | 121 | 29 | 52152 | 2.3084 | 0.010487 | *GATA2* |
| ENSG00000162073 | 16 | 3009246 | 3033490 | 66 | 16 | 46975 | 2.3084 | 0.010488 | *PAQR4* |
| ENSG00000179546 | 1 | 23506993 | 23531222 | 104 | 8 | 49931 | 2.3076 | 0.010512 | *HTR1D* |
| ENSG00000174957 | 11 | 55934094 | 55955032 | 65 | 11 | 55371 | 2.3063 | 0.010548 | *OR5J2* |
| ENSG00000204175 | 10 | 46984087 | 47015643 | 6 | 4 | 44856 | 2.3053 | 0.010574 | *GPRIN2* |
| ENSG00000257545 | 12 | 106879736 | 107178696 | 715 | 78 | 49444 | 2.3042 | 0.010607 | *RP11-144F15.1* |
| ENSG00000166821 | 15 | 90210995 | 90244014 | 105 | 16 | 45568 | 2.3038 | 0.010616 | *PEX11A* |
| ENSG00000052850 | 11 | 44271994 | 44341716 | 250 | 51 | 49269 | 2.3028 | 0.010645 | *ALX4* |
| ENSG00000109133 | 4 | 41927137 | 41972589 | 172 | 15 | 50992 | 2.3026 | 0.01065 | *TMEM33* |
| ENSG00000197309 | 11 | 124045923 | 124066952 | 92 | 14 | 49834 | 2.3021 | 0.010665 | *OR10D3* |
| ENSG00000176029 | 11 | 8931623 | 8964553 | 125 | 13 | 50060 | 2.2987 | 0.010761 | *C11orf16* |
| ENSG00000225697 | 3 | 48653156 | 48682926 | 36 | 6 | 47488 | 2.2986 | 0.010763 | *SLC26A6* |
| ENSG00000112855 | 5 | 140061011 | 140088889 | 58 | 6 | 55612 | 2.2986 | 0.010764 | *HARS2* |
| ENSG00000145808 | 5 | 128785958 | 129084376 | 892 | 70 | 43083 | 2.2973 | 0.010802 | *ADAMTS19* |
| ENSG00000160307 | 21 | 48008875 | 48035121 | 162 | 19 | 41668 | 2.2959 | 0.010842 | *S100B* |
| ENSG00000218739 | 2 | 37413631 | 37453336 | 117 | 20 | 52811 | 2.2946 | 0.010879 | *CEBPZ-AS1* |
| ENSG00000142409 | 19 | 56588732 | 56642649 | 101 | 22 | 40861 | 2.294 | 0.010894 | *ZNF787* |
| ENSG00000136169 | 13 | 50008429 | 50079138 | 228 | 26 | 44500 | 2.2935 | 0.010909 | *SETDB2* |
| ENSG00000129933 | 19 | 19421490 | 19479563 | 163 | 18 | 40239 | 2.293 | 0.010923 | *MAU2* |
| ENSG00000167972 | 16 | 2315882 | 2400747 | 159 | 32 | 44603 | 2.2926 | 0.010935 | *ABCA3* |
| ENSG00000072364 | 5 | 132201071 | 132309326 | 288 | 20 | 44695 | 2.2925 | 0.010938 | *AFF4* |
| ENSG00000138231 | 3 | 137869854 | 137903791 | 86 | 4 | 52733 | 2.2924 | 0.010942 | *DBR1* |
| ENSG00000073711 | 3 | 135674515 | 135876733 | 371 | 34 | 42622 | 2.2921 | 0.010949 | *PPP2R3A* |
| ENSG00000213516 | 1 | 89435139 | 89468643 | 72 | 16 | 41396 | 2.2917 | 0.010962 | *RBMXL1* |
| ENSG00000125827 | 20 | 7947995 | 8010476 | 174 | 29 | 34720 | 2.2908 | 0.010987 | *TMX4* |
| ENSG00000233041 | 15 | 40633234 | 40658635 | 78 | 23 | 49935 | 2.289 | 0.011039 | *PHGR1* |
| ENSG00000163145 | 4 | 15331442 | 15457790 | 397 | 40 | 32752 | 2.288 | 0.011068 | *C1QTNF7* |
| ENSG00000141200 | 17 | 51890239 | 51912573 | 102 | 14 | 41731 | 2.2877 | 0.011076 | *KIF2B* |
| ENSG00000176692 | 16 | 86590857 | 86612539 | 85 | 17 | 46977 | 2.2854 | 0.011144 | *FOXC2* |

| | | | | | | | | | |
|---|---|---|---|---|---|---|---|---|---|
| ENSG00000151067 | 12 | 2069952 | 2812108 | 2243 | 253 | 46008 | 2.2851 | 0.011154 | *CACNA1C* |
| ENSG00000186577 | 6 | 34204157 | 34227247 | 62 | 10 | 37813 | 2.2832 | 0.01121 | *C6orf1* |
| ENSG00000198898 | 7 | 116441124 | 116572103 | 265 | 32 | 45944 | 2.283 | 0.011216 | *CAPZA2* |
| ENSG00000128268 | 22 | 39843349 | 39898199 | 91 | 26 | 43507 | 2.2817 | 0.011252 | *MGAT3* |
| ENSG00000204539 | 6 | 31072867 | 31098223 | 363 | 36 | 40955 | 2.2812 | 0.011267 | *CDSN* |
| ENSG00000183621 | 10 | 31099136 | 31330866 | 800 | 57 | 47401 | 2.281 | 0.011275 | *ZNF438* |
| ENSG00000100321 | 22 | 39735930 | 39791593 | 155 | 23 | 37737 | 2.28 | 0.011304 | *SYNGR1* |
| ENSG00000197894 | 4 | 99982132 | 100019952 | 220 | 20 | 45908 | 2.2797 | 0.011314 | *ADH5* |
| ENSG00000171462 | 6 | 43408090 | 43434370 | 54 | 14 | 44775 | 2.2787 | 0.011341 | *DLK2* |
| ENSG00000187372 | 5 | 140583509 | 140606993 | 69 | 16 | 43598 | 2.2787 | 0.011343 | *PCDHB13* |
| ENSG00000008300 | 3 | 48663902 | 48710348 | 55 | 7 | 48650 | 2.2774 | 0.011382 | *CELSR3* |
| ENSG00000133067 | 1 | 202153029 | 202298909 | 389 | 48 | 42478 | 2.2773 | 0.011383 | *LGR6* |
| ENSG00000154269 | 6 | 131939582 | 132078553 | 289 | 45 | 32929 | 2.2773 | 0.011384 | *ENPP3* |
| ENSG00000105576 | 19 | 12800008 | 12844825 | 42 | 22 | 46999 | 2.2772 | 0.011388 | *TNPO2* |
| ENSG00000124126 | 20 | 47230790 | 47454420 | 816 | 66 | 47433 | 2.2748 | 0.01146 | *PREX1* |
| ENSG00000134258 | 1 | 117676209 | 117763556 | 276 | 38 | 37348 | 2.2743 | 0.011474 | *VTCN1* |
| ENSG00000254093 | 8 | 10612473 | 10707394 | 554 | 39 | 46060 | 2.2741 | 0.011481 | *PINX1* |
| ENSG00000132692 | 1 | 156601182 | 156639324 | 70 | 23 | 42271 | 2.2727 | 0.011523 | *BCAN* |
| ENSG00000172324 | 11 | 59179416 | 59200449 | 69 | 12 | 50639 | 2.2723 | 0.011534 | *OR5A2* |
| ENSG00000184330 | 1 | 153379000 | 153405701 | 53 | 9 | 43934 | 2.2705 | 0.011588 | *S100A7A* |
| ENSG00000160284 | 21 | 47571062 | 47614390 | 294 | 32 | 40192 | 2.2705 | 0.011589 | *SPATC1L* |
| ENSG00000143222 | 1 | 161112566 | 161138646 | 52 | 13 | 53614 | 2.2699 | 0.011608 | *UFC1* |
| ENSG00000158796 | 1 | 161080764 | 161112478 | 51 | 8 | 54820 | 2.2692 | 0.01163 | *DEDD* |
| ENSG00000178971 | 17 | 8120191 | 8161362 | 127 | 24 | 46112 | 2.2687 | 0.011642 | *CTC1* |
| ENSG00000111371 | 12 | 46566846 | 46673800 | 230 | 53 | 44082 | 2.2683 | 0.011657 | *SLC38A1* |
| ENSG00000173715 | 11 | 66501922 | 66620987 | 128 | 32 | 49546 | 2.267 | 0.011696 | *C11orf80* |
| ENSG00000173210 | 5 | 148511046 | 148650105 | 359 | 40 | 49301 | 2.2668 | 0.011702 | *ABLIM3* |
| ENSG00000136938 | 9 | 100735643 | 100788225 | 93 | 20 | 42588 | 2.2652 | 0.011749 | *ANP32B* |
| ENSG00000206181 | 18 | 44548943 | 44571988 | 69 | 14 | 51406 | 2.2627 | 0.011827 | *TCEB3B* |
| ENSG00000213689 | 3 | 48496445 | 48519044 | 26 | 6 | 51958 | 2.2626 | 0.01183 | *TREX1* |
| ENSG00000168394 | 6 | 32802986 | 32831755 | 235 | 32 | 50457 | 2.2618 | 0.011856 | *TAP1* |
| ENSG00000130881 | 19 | 33658509 | 33708694 | 170 | 32 | 29780 | 2.2599 | 0.011913 | *LRP3* |
| ENSG00000241878 | 22 | 32004477 | 32068418 | 251 | 28 | 50794 | 2.2599 | 0.011914 | *PISD* |
| ENSG00000165917 | 11 | 47449308 | 47480730 | 86 | 9 | 39294 | 2.2592 | 0.011935 | *RAPSN* |
| ENSG00000173418 | 20 | 19987760 | 20024299 | 62 | 14 | 46219 | 2.259 | 0.011941 | *NAA20* |
| ENSG00000204923 | 2 | 68676551 | 68704390 | 56 | 7 | 40586 | 2.2586 | 0.011955 | *FBXO48* |

| | | | | | | | | | |
|---|---|---|---|---|---|---|---|---|---|
| ENSG00000108292 | 17 | 36851795 | 36896056 | 61 | 18 | 45865 | 2.2584 | 0.01196 | *MLLT6* |
| ENSG00000116489 | 1 | 113151795 | 113224241 | 139 | 20 | 50601 | 2.2582 | 0.011965 | *CAPZA1* |
| ENSG00000102882 | 16 | 30115426 | 30144827 | 45 | 15 | 43762 | 2.2568 | 0.012011 | *MAPK3* |
| ENSG00000181450 | 1 | 227741244 | 227875144 | 368 | 26 | 46543 | 2.2563 | 0.012026 | *ZNF678* |
| ENSG00000165555 | 14 | 77850364 | 77899860 | 108 | 14 | 42250 | 2.2558 | 0.012042 | *NOXRED1* |
| ENSG00000164509 | 5 | 55137207 | 55228678 | 267 | 38 | 41717 | 2.2549 | 0.012071 | *IL31RA* |
| ENSG00000185090 | 1 | 38249474 | 38276809 | 59 | 13 | 44465 | 2.2544 | 0.012085 | *MANEAL* |
| ENSG00000089597 | 11 | 62382298 | 62424104 | 50 | 10 | 50566 | 2.254 | 0.012099 | *GANAB* |
| ENSG00000258724 | 8 | 10572909 | 10707357 | 721 | 48 | 46829 | 2.2526 | 0.012142 | *SOX7* |
| ENSG00000161405 | 17 | 37911198 | 38030441 | 299 | 17 | 42794 | 2.2524 | 0.012147 | *IKZF3* |
| ENSG00000156535 | 6 | 74395508 | 74548040 | 500 | 42 | 50388 | 2.2514 | 0.012179 | *CD109* |
| ENSG00000115963 | 2 | 151314709 | 151405525 | 210 | 36 | 53881 | 2.2506 | 0.012204 | *RND3* |
| ENSG00000146007 | 5 | 140068265 | 140096248 | 59 | 5 | 56445 | 2.2504 | 0.012211 | *ZMAT2* |
| ENSG00000145879 | 5 | 147681982 | 147705485 | 85 | 9 | 26834 | 2.2498 | 0.012232 | *SPINK7* |
| ENSG00000135747 | 1 | 247098849 | 247252113 | 800 | 63 | 45010 | 2.2495 | 0.012241 | *ZNF670* |
| ENSG00000188279 | 10 | 49189649 | 49217818 | 8 | 6 | 43904 | 2.2494 | 0.012243 | *FAM25C* |
| ENSG00000171056 | 8 | 10571278 | 10707357 | 728 | 49 | 46866 | 2.2489 | 0.012259 | *SOX7* |
| ENSG00000173120 | 11 | 66876740 | 67035558 | 168 | 32 | 46210 | 2.2488 | 0.012263 | *KDM2A* |
| ENSG00000186567 | 19 | 45155545 | 45197631 | 106 | 24 | 39228 | 2.2487 | 0.012266 | *CEACAM19* |
| ENSG00000135862 | 1 | 182982595 | 183124727 | 605 | 16 | 29691 | 2.2485 | 0.012272 | *LAMC1* |
| ENSG00000106560 | 7 | 150372785 | 150400729 | 83 | 17 | 54666 | 2.2483 | 0.012279 | *GIMAP2* |
| ENSG00000176125 | 7 | 100476346 | 100497339 | 52 | 12 | 42693 | 2.248 | 0.012289 | *UFSP1* |
| ENSG00000173626 | 6 | 116806152 | 116876773 | 227 | 22 | 44574 | 2.2471 | 0.012316 | *TRAPPC3L* |
| ENSG00000128519 | 7 | 122624759 | 122645754 | 75 | 11 | 52789 | 2.2459 | 0.012354 | *TAS2R16* |
| ENSG00000141524 | 17 | 76096539 | 76138488 | 148 | 42 | 47862 | 2.2456 | 0.012364 | *TMC6* |
| ENSG00000166452 | 11 | 8922686 | 8951631 | 100 | 14 | 50295 | 2.245 | 0.012385 | *AKIP1* |
| ENSG00000238091 | 2 | 114194992 | 114215429 | 30 | 5 | 39759 | 2.2432 | 0.012441 | *AC016745.1* |
| ENSG00000114739 | 3 | 38485342 | 38544633 | 127 | 13 | 33802 | 2.2432 | 0.012442 | *ACVR2B* |
| ENSG00000106089 | 7 | 73103536 | 73144002 | 75 | 21 | 51374 | 2.2426 | 0.012462 | *STX1A* |
| ENSG00000169855 | 3 | 78636390 | 79826965 | 3048 | 188 | 46071 | 2.2424 | 0.012468 | *ROBO1* |
| ENSG00000174080 | 11 | 66320934 | 66346312 | 73 | 10 | 43168 | 2.2411 | 0.012509 | *CTSF* |
| ENSG00000088826 | 20 | 4091627 | 4178394 | 426 | 73 | 44494 | 2.2409 | 0.012518 | *SMOX* |
| ENSG00000185087 | 15 | 98970391 | 99067611 | 341 | 43 | 44689 | 2.2405 | 0.01253 | *FAM169B* |
| ENSG00000205029 | 11 | 55596228 | 55617214 | 82 | 16 | 50833 | 2.2404 | 0.012534 | *OR5D16* |
| ENSG00000185053 | 8 | 13937373 | 15105848 | 6869 | 326 | 40974 | 2.2398 | 0.012553 | *SGCZ* |
| ENSG00000070087 | 3 | 149672691 | 149778575 | 313 | 38 | 48009 | 2.2392 | 0.012571 | *PFN2* |

| | | | | | | | | | |
|---|---|---|---|---|---|---|---|---|---|
| ENSG00000170502 | 4 | 88333734 | 88390606 | 187 | 24 | 49152 | 2.2375 | 0.012626 | *NUDT9* |
| ENSG00000116721 | 1 | 12841546 | 12866777 | 90 | 17 | 39164 | 2.2374 | 0.012631 | *PRAMEF1* |
| ENSG00000164171 | 5 | 52275156 | 52400609 | 539 | 47 | 54179 | 2.2372 | 0.012638 | *ITGA2* |
| ENSG00000000971 | 1 | 196611008 | 196726634 | 291 | 27 | 41985 | 2.2368 | 0.012649 | *CFH* |
| ENSG00000128228 | 22 | 21986550 | 22008587 | 70 | 20 | 46316 | 2.2364 | 0.012662 | *SDF2L1* |
| ENSG00000160410 | 19 | 41072757 | 41107305 | 106 | 27 | 46407 | 2.2363 | 0.012665 | *SHKBP1* |
| ENSG00000241839 | 15 | 65124088 | 65170206 | 153 | 17 | 38071 | 2.2351 | 0.012704 | *PLEKHO2* |
| ENSG00000196793 | 10 | 44041792 | 44080066 | 118 | 11 | 52939 | 2.235 | 0.012707 | *ZNF239* |
| ENSG00000226763 | 19 | 44090769 | 44128649 | 68 | 15 | 52435 | 2.235 | 0.01271 | *SRRM5* |
| ENSG00000172554 | 2 | 936554 | 1381385 | 2235 | 125 | 40336 | 2.2346 | 0.012723 | *SNTG2* |
| ENSG00000035664 | 15 | 64189235 | 64374232 | 631 | 73 | 36997 | 2.2339 | 0.012744 | *DAPK2* |
| ENSG00000108588 | 17 | 61812610 | 61863711 | 126 | 11 | 31752 | 2.2327 | 0.012784 | *CCDC47* |
| ENSG00000197061 | 6 | 26094104 | 26114518 | 71 | 13 | 51340 | 2.2321 | 0.012806 | *HIST1H4C* |
| ENSG00000136305 | 14 | 24764302 | 24790636 | 62 | 13 | 43325 | 2.2319 | 0.012811 | *CIDEB* |
| ENSG00000004939 | 17 | 42315753 | 42355509 | 83 | 26 | 44794 | 2.2318 | 0.012815 | *SLC4A1* |
| ENSG00000178035 | 3 | 49051758 | 49076841 | 28 | 10 | 42797 | 2.2306 | 0.012855 | *IMPDH2* |
| ENSG00000174374 | 7 | 74431226 | 74500064 | 53 | 15 | 41760 | 2.23 | 0.012873 | *WBSCR16* |
| ENSG00000134049 | 18 | 44671413 | 44712745 | 155 | 15 | 55310 | 2.2282 | 0.012934 | *IER3IP1* |
| ENSG00000181752 | 11 | 55916870 | 55937793 | 55 | 11 | 55959 | 2.2277 | 0.01295 | *OR8K5* |
| ENSG00000121858 | 3 | 172213298 | 172251297 | 197 | 25 | 48216 | 2.2274 | 0.012961 | *TNFSF10* |
| ENSG00000214708 | 17 | 30458245 | 30480154 | 45 | 7 | 41311 | 2.2273 | 0.012963 | *AC090616.2* |
| ENSG00000127564 | 16 | 3008025 | 3040540 | 103 | 22 | 44668 | 2.2268 | 0.01298 | *PKMYT1* |
| ENSG00000114316 | 3 | 49305264 | 49388145 | 119 | 18 | 45892 | 2.2256 | 0.013019 | *USP4* |
| ENSG00000100591 | 14 | 77914213 | 77945817 | 108 | 13 | 38149 | 2.2248 | 0.013047 | *AHSA1* |
| ENSG00000184990 | 14 | 105209437 | 105244831 | 109 | 20 | 42938 | 2.2247 | 0.013052 | *SIVA1* |
| ENSG00000132840 | 5 | 78355540 | 78395289 | 122 | 17 | 34512 | 2.2241 | 0.013072 | *BHMT2* |
| ENSG00000138663 | 4 | 83945600 | 84006971 | 87 | 21 | 46875 | 2.2232 | 0.0131 | *COPS4* |
| ENSG00000082781 | 3 | 124470795 | 124630265 | 637 | 58 | 52996 | 2.2228 | 0.013115 | *ITGB5* |
| ENSG00000248871 | 17 | 7442416 | 7474918 | 105 | 23 | 46063 | 2.2212 | 0.013167 | *TNFSF12-TNFSF13* |
| ENSG00000233404 | 2 | 102498955 | 102521150 | 45 | 13 | 43781 | 2.2203 | 0.013198 | *FLJ20373* |
| ENSG00000141141 | 17 | 35959787 | 36013493 | 73 | 16 | 37541 | 2.2197 | 0.01322 | *DDX52* |
| ENSG00000115850 | 2 | 136535410 | 136604750 | 107 | 16 | 21665 | 2.2196 | 0.013224 | *LCT* |
| ENSG00000005955 | 17 | 34890737 | 34956278 | 152 | 20 | 39043 | 2.2195 | 0.013225 | *GGNBP2* |
| ENSG00000082213 | 5 | 31522373 | 31565165 | 163 | 31 | 48460 | 2.2193 | 0.013233 | *C5orf22* |
| ENSG00000142669 | 1 | 26595667 | 26618007 | 70 | 9 | 45541 | 2.2179 | 0.01328 | *SH3BGRL3* |
| ENSG00000204909 | 5 | 147690766 | 147729412 | 137 | 12 | 26149 | 2.2179 | 0.013281 | *SPINK9* |

| | | | | | | | | | |
|---|---|---|---|---|---|---|---|---|---|
| ENSG00000163885 | 3 | 126103782 | 126165399 | 251 | 49 | 49808 | 2.2175 | 0.013296 | *CCDC37* |
| ENSG00000164151 | 5 | 5410777 | 5500347 | 169 | 26 | 49804 | 2.2173 | 0.0133 | *KIAA0947* |
| ENSG00000181220 | 7 | 149159885 | 149204908 | 66 | 12 | 51603 | 2.2169 | 0.013314 | *ZNF746* |
| ENSG00000160570 | 19 | 42692750 | 42734292 | 44 | 6 | 53431 | 2.2168 | 0.013317 | *DEDD2* |
| ENSG00000187720 | 15 | 71379291 | 72085722 | 2385 | 234 | 46222 | 2.2167 | 0.013321 | *THSD4* |
| ENSG00000197467 | 10 | 71551644 | 71734031 | 861 | 181 | 51669 | 2.2165 | 0.013329 | *COL13A1* |
| ENSG00000234298 | 18 | 44538658 | 44560534 | 47 | 11 | 51069 | 2.2163 | 0.013334 | *TCEB3CL* |
| ENSG00000128578 | 7 | 129064274 | 129138240 | 164 | 18 | 38261 | 2.2154 | 0.013365 | *STRIP2* |
| ENSG00000111783 | 12 | 106966685 | 107166581 | 461 | 44 | 50015 | 2.215 | 0.01338 | *RFX4* |
| ENSG00000268361 | 19 | 44048844 | 44107188 | 202 | 33 | 47973 | 2.2146 | 0.013395 | *L34079.2* |
| ENSG00000164638 | 7 | 5304000 | 5356501 | 256 | 54 | 42865 | 2.2142 | 0.013408 | *SLC29A4* |
| ENSG00000196757 | 19 | 12025883 | 12071588 | 130 | 15 | 34769 | 2.2135 | 0.01343 | *ZNF700* |
| ENSG00000136682 | 2 | 114185268 | 114263766 | 106 | 10 | 43086 | 2.2111 | 0.013514 | *CBWD2* |
| ENSG00000173273 | 8 | 9403424 | 9649856 | 1009 | 33 | 52566 | 2.2111 | 0.013516 | *TNKS* |
| ENSG00000178921 | 17 | 8140936 | 8183809 | 146 | 25 | 41832 | 2.2109 | 0.013521 | *PFAS* |
| ENSG00000164591 | 5 | 150030436 | 150068927 | 64 | 7 | 30048 | 2.2105 | 0.013537 | *MYOZ3* |
| ENSG00000198756 | 1 | 183888796 | 184016863 | 370 | 57 | 44131 | 2.2084 | 0.013607 | *COLGALT2* |
| ENSG00000205795 | 2 | 10186907 | 10231071 | 177 | 32 | 52480 | 2.2084 | 0.013608 | *CYS1* |
| ENSG00000250091 | 12 | 124400971 | 124429531 | 58 | 7 | 54155 | 2.208 | 0.013622 | *DNAH10OS* |
| ENSG00000003509 | 2 | 37448774 | 37490546 | 147 | 25 | 52444 | 2.2065 | 0.013675 | *NDUFAF7* |
| ENSG00000117481 | 1 | 46795849 | 46840824 | 243 | 7 | 24555 | 2.206 | 0.013692 | *NSUN4* |
| ENSG00000213672 | 3 | 48691364 | 48733797 | 65 | 10 | 45045 | 2.2058 | 0.0137 | *NCKIPSD* |
| ENSG00000105732 | 19 | 42562629 | 42595701 | 28 | 12 | 49736 | 2.2058 | 0.0137 | *ZNF574* |
| ENSG00000117335 | 1 | 207915402 | 207978858 | 154 | 17 | 50410 | 2.2045 | 0.013746 | *CD46* |
| ENSG00000239697 | 17 | 7442208 | 7474925 | 106 | 23 | 45751 | 2.2035 | 0.013779 | *TNFSF12* |
| ENSG00000196071 | 1 | 248090493 | 248274224 | 537 | 56 | 41231 | 2.2034 | 0.013783 | *OR2L13* |
| ENSG00000198075 | 2 | 108984367 | 109014513 | 75 | 12 | 31051 | 2.2033 | 0.013787 | *SULT1C4* |
| ENSG00000100811 | 14 | 100694635 | 100759129 | 152 | 22 | 51596 | 2.2028 | 0.013806 | *YY1* |
| ENSG00000112343 | 6 | 25953030 | 25997384 | 105 | 18 | 41098 | 2.2013 | 0.013857 | *TRIM38* |
| ENSG00000126226 | 13 | 113821891 | 113873029 | 93 | 12 | 32484 | 2.2007 | 0.013878 | *PCID2* |
| ENSG00000070081 | 11 | 17219700 | 17381521 | 377 | 35 | 44000 | 2.2002 | 0.013895 | *NUCB2* |
| ENSG00000104517 | 8 | 103255240 | 103435069 | 570 | 52 | 53258 | 2.2001 | 0.013901 | *UBR5* |
| ENSG00000181634 | 9 | 117536915 | 117578406 | 79 | 22 | 47529 | 2.1992 | 0.013932 | *TNFSF15* |
| ENSG00000083099 | 6 | 90267863 | 90358472 | 221 | 33 | 41746 | 2.1979 | 0.013976 | *LYRM2* |
| ENSG00000159176 | 1 | 201442658 | 201488584 | 167 | 34 | 43145 | 2.1974 | 0.013996 | *CSRP1* |
| ENSG00000150457 | 13 | 21537171 | 21645686 | 301 | 36 | 40811 | 2.1967 | 0.014022 | *LATS2* |

| | | | | | | | | | |
|---|---|---|---|---|---|---|---|---|---|
| ENSG00000117601 | 1 | 173862947 | 173896516 | 85 | 13 | 36313 | 2.1965 | 0.014029 | *SERPINC1* |
| ENSG00000182093 | 21 | 40742170 | 40810454 | 176 | 18 | 49866 | 2.1962 | 0.014038 | *WRB* |
| ENSG00000136485 | 17 | 61617822 | 61681639 | 72 | 21 | 46658 | 2.1961 | 0.014042 | *DCAF7* |
| ENSG00000227450 | 17 | 36895613 | 36916969 | 71 | 20 | 47357 | 2.1959 | 0.014048 | *CTB-58E17.5* |
| ENSG00000178243 | 9 | 138225095 | 138248408 | 62 | 13 | 46835 | 2.1944 | 0.014105 | *C9orf62* |
| ENSG00000187741 | 16 | 89793957 | 89893065 | 672 | 39 | 35769 | 2.194 | 0.014116 | *FANCA* |
| ENSG00000075945 | 1 | 169880467 | 170064349 | 651 | 27 | 53859 | 2.1935 | 0.014136 | *KIFAP3* |
| ENSG00000168350 | 14 | 100602756 | 100636500 | 108 | 22 | 47363 | 2.1926 | 0.014169 | *DEGS2* |
| ENSG00000205884 | 8 | 11821446 | 11842108 | 98 | 21 | 42938 | 2.1919 | 0.014193 | *DEFB136* |
| ENSG00000221819 | 16 | 90085316 | 90106309 | 89 | 16 | 36813 | 2.1904 | 0.014248 | *C16orf3* |
| ENSG00000158850 | 1 | 161131100 | 161157287 | 47 | 13 | 52142 | 2.1902 | 0.014253 | *B4GALT3* |
| ENSG00000159495 | 15 | 43558478 | 43604453 | 68 | 7 | 26505 | 2.1893 | 0.014289 | *TGM7* |
| ENSG00000163499 | 2 | 219844911 | 219868143 | 17 | 9 | 30913 | 2.1865 | 0.014391 | *CRYBA2* |
| ENSG00000151725 | 4 | 185605772 | 185665287 | 368 | 11 | 47922 | 2.1861 | 0.014404 | *CENPU* |
| ENSG00000183530 | 22 | 32062242 | 32156126 | 265 | 29 | 50110 | 2.1858 | 0.014415 | *PRR14L* |
| ENSG00000161133 | 22 | 20694868 | 20755048 | 113 | 27 | 45609 | 2.1837 | 0.014494 | *USP41* |
| ENSG00000140323 | 15 | 40640436 | 40673257 | 86 | 19 | 52049 | 2.1835 | 0.014498 | *DISP2* |
| ENSG00000145087 | 3 | 120616919 | 121153608 | 2014 | 54 | 42905 | 2.1833 | 0.014506 | *STXBP5L* |
| ENSG00000129103 | 7 | 56121695 | 56158363 | 122 | 20 | 55621 | 2.1827 | 0.014528 | *SUMF2* |
| ENSG00000003147 | 7 | 8142814 | 8312317 | 740 | 122 | 43579 | 2.1813 | 0.01458 | *ICA1* |
| ENSG00000177311 | 3 | 141033055 | 141178634 | 338 | 54 | 38886 | 2.1812 | 0.014583 | *ZBTB38* |
| ENSG00000150045 | 12 | 9970077 | 10007606 | 98 | 20 | 47343 | 2.1812 | 0.014585 | *KLRF1* |
| ENSG00000118785 | 4 | 88886819 | 88914562 | 93 | 18 | 39438 | 2.1811 | 0.014587 | *SPP1* |
| ENSG00000204361 | 11 | 114539108 | 114589362 | 180 | 27 | 51012 | 2.1801 | 0.014626 | *NXPE2* |
| ENSG00000149451 | 20 | 3638612 | 3672893 | 140 | 40 | 46476 | 2.1783 | 0.01469 | *ADAM33* |
| ENSG00000119537 | 18 | 60984959 | 61044743 | 104 | 20 | 42591 | 2.1777 | 0.014715 | *KDSR* |
| ENSG00000140527 | 15 | 90224028 | 90296869 | 253 | 18 | 50066 | 2.1769 | 0.014743 | *WDR93* |
| ENSG00000160472 | 19 | 55878204 | 55899612 | 74 | 19 | 40506 | 2.1762 | 0.014769 | *TMEM190* |
| ENSG00000137801 | 15 | 39863280 | 39901667 | 84 | 13 | 42806 | 2.1761 | 0.014775 | *THBS1* |
| ENSG00000157734 | 15 | 64433914 | 64459680 | 56 | 11 | 45833 | 2.1755 | 0.014796 | *SNX22* |
| ENSG00000183807 | 6 | 117063363 | 117096886 | 114 | 22 | 50675 | 2.1735 | 0.014872 | *FAM162B* |
| ENSG00000087087 | 7 | 100462733 | 100496285 | 85 | 16 | 44259 | 2.1732 | 0.014883 | *SRRT* |
| ENSG00000128272 | 22 | 39905700 | 39928691 | 36 | 8 | 51178 | 2.1722 | 0.014921 | *ATF4* |
| ENSG00000139405 | 12 | 113613331 | 113640173 | 67 | 11 | 54106 | 2.1722 | 0.014922 | *RITA1* |
| ENSG00000215568 | 22 | 17432826 | 17499112 | 291 | 31 | 53689 | 2.1677 | 0.015092 | *GAB4* |
| ENSG00000087263 | 16 | 56475402 | 56523012 | 119 | 19 | 43159 | 2.1675 | 0.015099 | *OGFOD1* |

| | | | | | | | | | |
|---|---|---|---|---|---|---|---|---|---|
| ENSG00000185437 | 21 | 40807781 | 40897433 | 414 | 32 | 42197 | 2.1672 | 0.015112 | *SH3BGR* |
| ENSG00000187051 | 22 | 39915098 | 39938860 | 52 | 10 | 41408 | 2.1671 | 0.015114 | *RPS19BP1* |
| ENSG00000077549 | 1 | 19655267 | 19822066 | 660 | 62 | 51441 | 2.1663 | 0.015144 | *CAPZB* |
| ENSG00000168795 | 9 | 37428111 | 37475396 | 86 | 20 | 47761 | 2.1652 | 0.015186 | *ZBTB5* |
| ENSG00000231880 | 11 | 47589277 | 47609823 | 24 | 9 | 41642 | 2.1648 | 0.015201 | *KBTBD4* |
| ENSG00000105737 | 19 | 42492473 | 42583650 | 70 | 20 | 53067 | 2.1643 | 0.015219 | *GRIK5* |
| ENSG00000204536 | 6 | 31100216 | 31136015 | 395 | 43 | 45874 | 2.1632 | 0.015262 | *CCHCR1* |
| ENSG00000112320 | 6 | 107801162 | 107991357 | 406 | 71 | 38628 | 2.1629 | 0.015273 | *SOBP* |
| ENSG00000087152 | 17 | 42259173 | 42287481 | 61 | 16 | 45488 | 2.1628 | 0.015278 | *ATXN7L3* |
| ENSG00000186452 | 12 | 51226703 | 51291667 | 188 | 14 | 49847 | 2.1622 | 0.0153 | *TMPRSS12* |
| ENSG00000086589 | 5 | 150060356 | 150090669 | 29 | 12 | 47205 | 2.1619 | 0.015314 | *RBM22* |
| ENSG00000247270 | 16 | 66506775 | 66529747 | 42 | 11 | 35175 | 2.1609 | 0.015351 | *AC132186.1* |
| ENSG00000131116 | 19 | 44101371 | 44134026 | 64 | 12 | 53031 | 2.1606 | 0.015362 | *ZNF428* |
| ENSG00000103335 | 16 | 88771751 | 88861619 | 529 | 92 | 44586 | 2.1592 | 0.015418 | *PIEZO1* |
| ENSG00000075673 | 13 | 25244549 | 25295921 | 266 | 36 | 48804 | 2.1583 | 0.015454 | *ATP12A* |
| ENSG00000143382 | 1 | 150511884 | 150543413 | 79 | 27 | 45367 | 2.1582 | 0.015455 | *ADAMTSL4* |
| ENSG00000105428 | 19 | 5445426 | 5466867 | 78 | 22 | 47520 | 2.1578 | 0.015471 | *ZNRF4* |
| ENSG00000162882 | 2 | 42984229 | 43029733 | 205 | 19 | 42674 | 2.1571 | 0.015498 | *HAAO* |
| ENSG00000143224 | 1 | 161126200 | 161157803 | 56 | 14 | 53011 | 2.1552 | 0.015575 | *PPOX* |
| ENSG00000145283 | 4 | 87734621 | 87780416 | 77 | 18 | 46719 | 2.1547 | 0.015592 | *SLC10A6* |
| ENSG00000174576 | 11 | 66178475 | 66204178 | 47 | 16 | 47270 | 2.1542 | 0.015613 | *NPAS4* |
| ENSG00000100219 | 22 | 29180543 | 29206585 | 76 | 15 | 37911 | 2.1541 | 0.015615 | *XBP1* |
| ENSG00000129351 | 19 | 10754937 | 10813093 | 108 | 13 | 52890 | 2.1532 | 0.015651 | *ILF3* |
| ENSG00000161082 | 19 | 3214701 | 3307074 | 305 | 53 | 42697 | 2.152 | 0.0157 | *CELF5* |
| ENSG00000146731 | 7 | 56109323 | 56141682 | 99 | 18 | 54776 | 2.152 | 0.015701 | *CCT6A* |
| ENSG00000223609 | 11 | 5243908 | 5266600 | 59 | 12 | 45821 | 2.1513 | 0.015726 | *HBD* |
| ENSG00000092850 | 1 | 36539676 | 36563876 | 19 | 11 | 50195 | 2.1511 | 0.015734 | *TEKT2* |
| ENSG00000196449 | 1 | 38258616 | 38283857 | 38 | 9 | 43943 | 2.1509 | 0.015741 | *YRDC* |
| ENSG00000178172 | 5 | 147572357 | 147604700 | 78 | 5 | 51953 | 2.1495 | 0.015799 | *SPINK6* |
| ENSG00000174307 | 1 | 201424620 | 201448365 | 84 | 23 | 35885 | 2.149 | 0.015817 | *PHLDA3* |
| ENSG00000113248 | 5 | 140615147 | 140637799 | 72 | 11 | 41220 | 2.148 | 0.015858 | *PCDHB15* |
| ENSG00000181264 | 11 | 120185838 | 120214391 | 58 | 13 | 49701 | 2.1475 | 0.015877 | *TMEM136* |
| ENSG00000187475 | 6 | 26097640 | 26118364 | 67 | 11 | 51640 | 2.1471 | 0.015891 | *HIST1H1T* |
| ENSG00000145692 | 5 | 78397602 | 78438108 | 132 | 19 | 41233 | 2.147 | 0.015898 | *BHMT* |
| ENSG00000198099 | 4 | 100034808 | 100088949 | 333 | 14 | 48292 | 2.1462 | 0.015927 | *ADH4* |
| ENSG00000137310 | 6 | 31116319 | 31144936 | 308 | 36 | 48767 | 2.1459 | 0.015941 | *TCF19* |

| | | | | | | | | | |
|---|---|---|---|---|---|---|---|---|---|
| ENSG00000124749 | 6 | 55911388 | 56268892 | 1633 | 103 | 48256 | 2.1454 | 0.015962 | *COL21A1* |
| ENSG00000140632 | 16 | 4843204 | 4907343 | 224 | 34 | 51706 | 2.1448 | 0.015985 | *GLYR1* |
| ENSG00000079819 | 6 | 131150487 | 131394462 | 770 | 66 | 41301 | 2.1445 | 0.015995 | *EPB41L2* |
| ENSG00000204314 | 6 | 32106136 | 32132150 | 71 | 22 | 45152 | 2.1443 | 0.016004 | *PRRT1* |
| ENSG00000172548 | 5 | 156877027 | 156911725 | 115 | 18 | 47179 | 2.1433 | 0.016046 | *NIPAL4* |
| ENSG00000167797 | 11 | 67263968 | 67286120 | 27 | 13 | 46535 | 2.1425 | 0.016078 | *CDK2AP2* |
| ENSG00000141371 | 17 | 58459790 | 58518782 | 57 | 12 | 38198 | 2.1422 | 0.016088 | *C17orf64* |
| ENSG00000163935 | 3 | 52927588 | 53090766 | 398 | 34 | 52898 | 2.1415 | 0.016116 | *SFMBT1* |
| ENSG00000157933 | 1 | 2150134 | 2251558 | 276 | 58 | 46801 | 2.1408 | 0.016145 | *SKI* |
| ENSG00000100068 | 22 | 25737385 | 25811344 | 326 | 50 | 24011 | 2.1407 | 0.016149 | *LRP5L* |
| ENSG00000125821 | 20 | 18558537 | 18754561 | 863 | 55 | 51272 | 2.1385 | 0.01624 | *DTD1* |
| ENSG00000000457 | 1 | 169808772 | 169873408 | 229 | 20 | 56438 | 2.1361 | 0.016337 | *SCYL3* |
| ENSG00000269402 | 17 | 30496379 | 30516885 | 41 | 10 | 42475 | 2.136 | 0.01634 | *AC116407.2* |
| ENSG00000268126 | 5 | 147688550 | 147711891 | 107 | 10 | 25251 | 2.1351 | 0.016376 | *AC091948.1* |
| ENSG00000088782 | 20 | 128111 | 149804 | 97 | 8 | 32050 | 2.1342 | 0.016412 | *DEFB127* |
| ENSG00000104880 | 19 | 7449999 | 7547363 | 506 | 43 | 45563 | 2.1331 | 0.01646 | *ARHGEF18* |
| ENSG00000089012 | 20 | 1599798 | 1648425 | 205 | 32 | 47822 | 2.1323 | 0.01649 | *SIRPG* |
| ENSG00000198885 | 2 | 96981069 | 97004091 | 26 | 11 | 38662 | 2.1309 | 0.016548 | *ITPRIPL1* |
| ENSG00000213714 | 20 | 55098302 | 55121576 | 72 | 17 | 34502 | 2.13 | 0.016587 | *FAM209B* |
| ENSG00000114416 | 3 | 180575929 | 180710541 | 295 | 28 | 46078 | 2.1294 | 0.01661 | *FXR1* |
| ENSG00000166526 | 7 | 99651656 | 99690171 | 66 | 14 | 52898 | 2.1294 | 0.01661 | *ZNF3* |
| ENSG00000189409 | 1 | 1557474 | 1580639 | 10 | 6 | 37567 | 2.1286 | 0.016643 | *MMP23B* |
| ENSG00000203740 | 1 | 170105142 | 170146931 | 169 | 22 | 46489 | 2.1286 | 0.016644 | *METTL11B* |
| ENSG00000105705 | 19 | 19376827 | 19441653 | 175 | 21 | 41238 | 2.1285 | 0.016646 | *SUGP1* |
| ENSG00000219545 | 7 | 7670342 | 8053689 | 1751 | 223 | 47095 | 2.1282 | 0.01666 | *RPA3-AS1* |
| ENSG00000184730 | 16 | 28495970 | 28520291 | 49 | 11 | 44391 | 2.1271 | 0.016704 | *APOBR* |
| ENSG00000232629 | 6 | 32713875 | 32741311 | 361 | 19 | 31316 | 2.1266 | 0.016725 | *HLA-DQB2* |
| ENSG00000179407 | 3 | 128171282 | 128196091 | 107 | 25 | 42959 | 2.125 | 0.016791 | *DNAJB8* |
| ENSG00000096093 | 6 | 52275106 | 52397892 | 477 | 61 | 43819 | 2.1247 | 0.016806 | *EFHC1* |
| ENSG00000135315 | 6 | 84823960 | 84947353 | 223 | 28 | 46603 | 2.1233 | 0.016864 | *KIAA1009* |
| ENSG00000100027 | 22 | 22041833 | 22100123 | 220 | 39 | 48290 | 2.1228 | 0.016886 | *YPEL1* |
| ENSG00000100522 | 14 | 53231912 | 53268386 | 60 | 10 | 52262 | 2.1206 | 0.016978 | *GNPNAT1* |
| ENSG00000166963 | 15 | 43793156 | 43833818 | 47 | 10 | 39624 | 2.1205 | 0.016982 | *MAP1A* |
| ENSG00000124145 | 20 | 43943928 | 43987064 | 145 | 26 | 46960 | 2.1204 | 0.016987 | *SDC4* |
| ENSG00000110429 | 11 | 33752485 | 33806089 | 192 | 25 | 43943 | 2.1203 | 0.016991 | *FBXO3* |
| ENSG00000105443 | 19 | 48962289 | 48995571 | 90 | 16 | 30794 | 2.1186 | 0.01706 | *CYTH2* |

| | | | | | | | | | |
|---|---|---|---|---|---|---|---|---|---|
| ENSG00000160838 | 1 | 156880442 | 156912886 | 93 | 12 | 45999 | 2.1183 | 0.017075 | *LRRC71* |
| ENSG00000064489 | 19 | 19246376 | 19312967 | 246 | 25 | 50457 | 2.1175 | 0.017107 | *MEF2BNB-MEF2B* |
| ENSG00000213999 | 19 | 19246377 | 19313400 | 246 | 25 | 50457 | 2.1175 | 0.017107 | *MEF2B* |
| ENSG00000224389 | 6 | 31972539 | 32013195 | 66 | 25 | 38866 | 2.1174 | 0.017113 | *C4B* |
| ENSG00000163453 | 4 | 57886939 | 57986551 | 536 | 75 | 50604 | 2.1171 | 0.017124 | *IGFBP7* |
| ENSG00000073050 | 19 | 44037192 | 44094625 | 222 | 32 | 47115 | 2.117 | 0.01713 | *XRCC1* |
| ENSG00000166823 | 15 | 90281892 | 90304541 | 87 | 10 | 52172 | 2.1166 | 0.017148 | *MESP1* |
| ENSG00000204655 | 6 | 29614758 | 29650149 | 230 | 30 | 44065 | 2.1161 | 0.017169 | *MOG* |
| ENSG00000234465 | 19 | 44070952 | 44098116 | 89 | 19 | 51692 | 2.1154 | 0.017196 | *PINLYP* |
| ENSG00000143924 | 2 | 42386490 | 42569688 | 581 | 38 | 43555 | 2.1152 | 0.017205 | *EML4* |
| ENSG00000203724 | 1 | 197861777 | 197886497 | 48 | 19 | 46438 | 2.1143 | 0.017245 | *C1orf53* |
| ENSG00000236294 | 7 | 113081127 | 113101457 | 41 | 10 | 35427 | 2.1127 | 0.017312 | *TSRM* |
| ENSG00000166578 | 12 | 113623246 | 113668899 | 101 | 12 | 54139 | 2.1124 | 0.017324 | *IQCD* |
| ENSG00000026559 | 20 | 49610193 | 49649666 | 74 | 22 | 36465 | 2.1115 | 0.017365 | *KCNG1* |
| ENSG00000146729 | 7 | 56009486 | 56077874 | 240 | 32 | 52372 | 2.1113 | 0.017375 | *GBAS* |
| ENSG00000170345 | 14 | 75735477 | 75758933 | 82 | 24 | 38146 | 2.1106 | 0.017403 | *FOS* |
| ENSG00000182628 | 17 | 57177312 | 57242630 | 94 | 15 | 46968 | 2.1101 | 0.017426 | *SKA2* |
| ENSG00000161217 | 3 | 195931093 | 196024828 | 319 | 34 | 48013 | 2.1085 | 0.017492 | *PCYT1A* |
| ENSG00000178662 | 2 | 166316157 | 166555917 | 549 | 72 | 51069 | 2.1083 | 0.017503 | *CSRNP3* |
| ENSG00000173171 | 1 | 155168490 | 155193615 | 38 | 11 | 47427 | 2.107 | 0.01756 | *MTX1* |
| ENSG00000151065 | 12 | 2045220 | 2123701 | 298 | 19 | 54471 | 2.1061 | 0.0176 | *DCP1B* |
| ENSG00000115970 | 2 | 43383800 | 43833185 | 1288 | 75 | 48240 | 2.1059 | 0.017605 | *THADA* |
| ENSG00000183791 | 18 | 44544573 | 44566449 | 52 | 11 | 51379 | 2.1056 | 0.01762 | *TCEB3C* |
| ENSG00000164406 | 5 | 132198014 | 132220738 | 60 | 10 | 41747 | 2.1044 | 0.017672 | *LEAP2* |
| ENSG00000230873 | 6 | 17092489 | 17141603 | 125 | 24 | 46546 | 2.103 | 0.017735 | *STMND1* |
| ENSG00000213619 | 11 | 47576888 | 47616114 | 49 | 15 | 43034 | 2.1025 | 0.017753 | *NDUFS3* |
| ENSG00000170260 | 7 | 148926742 | 148962700 | 118 | 21 | 49382 | 2.1015 | 0.017799 | *ZNF212* |
| ENSG00000248713 | 4 | 100547686 | 100585805 | 138 | 26 | 47442 | 2.1012 | 0.017812 | *RP11-766F14.2* |
| ENSG00000205832 | 16 | 4596491 | 4660715 | 165 | 23 | 37346 | 2.1009 | 0.017825 | *C16orf96* |
| ENSG00000087085 | 7 | 100477615 | 100504594 | 76 | 13 | 43701 | 2.1006 | 0.017836 | *ACHE* |
| ENSG00000160271 | 9 | 135963107 | 136049301 | 264 | 44 | 47546 | 2.1006 | 0.01784 | *RALGDS* |
| ENSG00000092969 | 1 | 218509577 | 218627961 | 270 | 49 | 40173 | 2.1 | 0.017862 | *TGFB2* |
| ENSG00000196344 | 4 | 100323418 | 100366894 | 177 | 36 | 43984 | 2.0998 | 0.017872 | *ADH7* |
| ENSG00000137460 | 4 | 153847504 | 153910848 | 177 | 44 | 47475 | 2.0998 | 0.017874 | *FHDC1* |
| ENSG00000138741 | 4 | 122790182 | 122882909 | 185 | 30 | 51733 | 2.0986 | 0.017928 | *TRPC3* |
| ENSG00000182324 | 19 | 48948766 | 48979367 | 101 | 16 | 26574 | 2.0983 | 0.017939 | *KCNJ14* |

| | | | | | | | | | |
|---|---|---|---|---|---|---|---|---|---|
| ENSG00000132256 | 11 | 5674425 | 5969849 | 1770 | 116 | 50602 | 2.0976 | 0.01797 | *TRIM5* |
| ENSG00000114737 | 3 | 50633921 | 50659262 | 32 | 7 | 43722 | 2.0967 | 0.018008 | *CISH* |
| ENSG00000117592 | 1 | 173436405 | 173467946 | 64 | 14 | 46385 | 2.0963 | 0.018029 | *PRDX6* |
| ENSG00000197479 | 5 | 140569183 | 140592618 | 81 | 15 | 42768 | 2.0963 | 0.018029 | *PCDHB11* |
| ENSG00000132600 | 16 | 68334877 | 68402466 | 227 | 21 | 50247 | 2.0959 | 0.018046 | *PRMT7* |
| ENSG00000088882 | 20 | 2764715 | 2791283 | 101 | 23 | 43596 | 2.0956 | 0.018059 | *CPXM1* |
| ENSG00000129038 | 15 | 74208330 | 74254478 | 187 | 35 | 42999 | 2.0951 | 0.018083 | *LOXL1* |
| ENSG00000158805 | 16 | 89776808 | 89817311 | 257 | 29 | 37065 | 2.0944 | 0.018111 | *ZNF276* |
| ENSG00000143469 | 1 | 210101538 | 210347636 | 474 | 34 | 44245 | 2.0943 | 0.018116 | *SYT14* |
| ENSG00000170153 | 4 | 141770961 | 142144031 | 1140 | 101 | 45840 | 2.0939 | 0.018135 | *RNF150* |
| ENSG00000268861 | 19 | 7435850 | 7545131 | 545 | 45 | 45210 | 2.0917 | 0.018233 | *CTD-2207O23.3* |
| ENSG00000148057 | 9 | 86227964 | 86269045 | 127 | 19 | 47183 | 2.0912 | 0.018256 | *IDNK* |
| ENSG00000067208 | 1 | 92964253 | 93267961 | 770 | 30 | 39254 | 2.0908 | 0.018272 | *EVI5* |
| ENSG00000153443 | 16 | 4648884 | 4675028 | 97 | 13 | 35703 | 2.0907 | 0.018276 | *UBALD1* |
| ENSG00000155275 | 4 | 8427867 | 8505258 | 373 | 37 | 43657 | 2.0905 | 0.018285 | *TRMT44* |
| ENSG00000237847 | 1 | 16989498 | 17010007 | 8 | 4 | 31697 | 2.0904 | 0.01829 | *FAM231A* |
| ENSG00000166831 | 15 | 65022091 | 65077786 | 153 | 20 | 34794 | 2.0901 | 0.018303 | *RBPMS2* |
| ENSG00000178795 | 11 | 76917603 | 77022732 | 425 | 38 | 38306 | 2.0887 | 0.018369 | *GDPD4* |
| ENSG00000168356 | 3 | 38877260 | 39002052 | 358 | 37 | 43119 | 2.0882 | 0.018389 | *SCN11A* |
| ENSG00000243232 | 5 | 140335820 | 140401936 | 120 | 26 | 53408 | 2.0879 | 0.018402 | *PCDHAC2* |
| ENSG00000113262 | 5 | 178395328 | 178433207 | 203 | 37 | 44295 | 2.0875 | 0.01842 | *GRM6* |
| ENSG00000075336 | 18 | 71805746 | 71836197 | 140 | 11 | 33348 | 2.0868 | 0.018454 | *TIMM21* |
| ENSG00000255730 | 19 | 41846758 | 41940906 | 251 | 17 | 51879 | 2.0867 | 0.018456 | *CTC-435M10.3* |
| ENSG00000204516 | 6 | 31452658 | 31488901 | 478 | 40 | 51217 | 2.0865 | 0.018467 | *MICB* |
| ENSG00000128610 | 7 | 121931448 | 121960745 | 70 | 21 | 45898 | 2.0864 | 0.01847 | *FEZF1* |
| ENSG00000153827 | 2 | 230618554 | 230797955 | 269 | 31 | 54347 | 2.086 | 0.018492 | *TRIP12* |
| ENSG00000111834 | 6 | 116927642 | 116964148 | 149 | 20 | 51766 | 2.0858 | 0.018496 | *RSPH4A* |
| ENSG00000113141 | 5 | 140016643 | 140052064 | 77 | 11 | 53958 | 2.0857 | 0.018503 | *IK* |
| ENSG00000164187 | 5 | 36088514 | 36162063 | 115 | 27 | 40214 | 2.0844 | 0.018562 | *LMBRD2* |
| ENSG00000172320 | 11 | 59200617 | 59221667 | 88 | 12 | 42257 | 2.0842 | 0.018571 | *OR5A1* |
| ENSG00000226104 | 19 | 10741715 | 10762093 | 49 | 7 | 55775 | 2.0842 | 0.018571 | *AC011475.1* |
| ENSG00000178188 | 16 | 28847921 | 28895526 | 91 | 7 | 26261 | 2.0838 | 0.018591 | *SH2B1* |
| ENSG00000136877 | 9 | 130546876 | 130586606 | 109 | 17 | 39724 | 2.0836 | 0.018596 | *FPGS* |
| ENSG00000176177 | 22 | 40129049 | 40299868 | 253 | 29 | 52001 | 2.0828 | 0.018633 | *ENTHD1* |
| ENSG00000139218 | 12 | 46302914 | 46395903 | 137 | 25 | 48243 | 2.0826 | 0.018644 | *SCAF11* |
| ENSG00000143155 | 1 | 168138171 | 168179950 | 131 | 15 | 39655 | 2.0825 | 0.018646 | *TIPRL* |

| | | | | | | | | | |
|---|---|---|---|---|---|---|---|---|---|
| ENSG00000178403 | 4 | 113424672 | 113447328 | 67 | 16 | 53955 | 2.0813 | 0.018704 | *NEUROG2* |
| ENSG00000129467 | 14 | 24777555 | 24814299 | 125 | 24 | 36883 | 2.0808 | 0.018726 | *ADCY4* |
| ENSG00000267942 | 12 | 66281518 | 66327967 | 83 | 22 | 36580 | 2.0805 | 0.01874 | *AC090673.2* |
| ENSG00000047936 | 6 | 117599463 | 117757018 | 596 | 75 | 48405 | 2.0802 | 0.018752 | *ROS1* |
| ENSG00000121892 | 4 | 39814483 | 39989576 | 579 | 53 | 43864 | 2.08 | 0.018765 | *PDS5A* |
| ENSG00000130270 | 19 | 1772074 | 1822275 | 148 | 39 | 42316 | 2.08 | 0.018765 | *ATP8B3* |
| ENSG00000115944 | 2 | 42550686 | 42662228 | 259 | 26 | 51883 | 2.0795 | 0.018788 | *COX7A2L* |
| ENSG00000117318 | 1 | 23874409 | 23896285 | 54 | 16 | 33745 | 2.0791 | 0.018806 | *ID3* |
| ENSG00000047932 | 6 | 117629374 | 117933691 | 1135 | 81 | 49808 | 2.0781 | 0.01885 | *GOPC* |
| ENSG00000104765 | 8 | 26230414 | 26373152 | 592 | 99 | 46112 | 2.078 | 0.018854 | *BNIP3L* |
| ENSG00000126231 | 13 | 113802968 | 113836694 | 66 | 7 | 25254 | 2.0775 | 0.018877 | *PROZ* |
| ENSG00000171681 | 12 | 14508610 | 14661697 | 451 | 40 | 47768 | 2.0773 | 0.018888 | *ATF7IP* |
| ENSG00000164707 | 7 | 135355985 | 135424006 | 242 | 33 | 51848 | 2.0761 | 0.018944 | *SLC13A4* |
| ENSG00000141258 | 17 | 2230792 | 2294352 | 213 | 35 | 37844 | 2.076 | 0.018949 | *SGSM2* |
| ENSG00000167971 | 16 | 2217184 | 2256526 | 67 | 18 | 41054 | 2.0754 | 0.018973 | *CASKIN1* |
| ENSG00000162409 | 1 | 57100995 | 57191008 | 345 | 31 | 45615 | 2.0754 | 0.018975 | *PRKAA2* |
| ENSG00000136147 | 13 | 50059746 | 50113123 | 252 | 23 | 37609 | 2.075 | 0.018995 | *PHF11* |
| ENSG00000129282 | 17 | 34948001 | 34975407 | 84 | 16 | 39225 | 2.0735 | 0.019063 | *MRM1* |
| ENSG00000178691 | 17 | 30254037 | 30338064 | 239 | 12 | 51723 | 2.0729 | 0.019092 | *SUZ12* |
| ENSG00000110721 | 11 | 67810326 | 67898911 | 150 | 22 | 54229 | 2.0725 | 0.019107 | *CHKA* |
| ENSG00000185745 | 10 | 91142303 | 91173745 | 98 | 19 | 32176 | 2.0722 | 0.019124 | *IFIT1* |
| ENSG00000198732 | 14 | 70310848 | 70509083 | 780 | 112 | 43878 | 2.0718 | 0.019141 | *SMOC1* |
| ENSG00000123064 | 12 | 113584979 | 113633284 | 101 | 13 | 54507 | 2.0714 | 0.01916 | *DDX54* |
| ENSG00000119771 | 2 | 23598088 | 23941481 | 1336 | 158 | 46437 | 2.071 | 0.019178 | *KLHL29* |
| ENSG00000119242 | 12 | 124393207 | 124467378 | 162 | 15 | 50315 | 2.0706 | 0.019198 | *CCDC92* |
| ENSG00000170915 | 6 | 52216219 | 52282575 | 279 | 48 | 43718 | 2.0704 | 0.019209 | *PAQR8* |
| ENSG00000272741 | 3 | 195965101 | 196055093 | 274 | 22 | 47452 | 2.07 | 0.019227 | *RP11-447L10.1* |
| ENSG00000130182 | 16 | 3128891 | 3159318 | 85 | 16 | 45613 | 2.07 | 0.019227 | *ZSCAN10* |
| ENSG00000047579 | 6 | 15513032 | 15673289 | 400 | 34 | 43528 | 2.069 | 0.019275 | *DTNBP1* |
| ENSG00000066827 | 8 | 135480031 | 135735292 | 893 | 92 | 47918 | 2.0688 | 0.01928 | *ZFAT* |
| ENSG00000167785 | 19 | 8910380 | 8953004 | 134 | 15 | 55582 | 2.0685 | 0.019295 | *ZNF558* |
| ENSG00000009954 | 7 | 72844728 | 72946608 | 173 | 23 | 52228 | 2.068 | 0.019322 | *BAZ1B* |
| ENSG00000188620 | 10 | 124885478 | 124907257 | 39 | 16 | 45147 | 2.0672 | 0.019357 | *HMX3* |
| ENSG00000184005 | 1 | 76530404 | 77110286 | 1968 | 197 | 41075 | 2.0666 | 0.019386 | *ST6GALNAC3* |
| ENSG00000100335 | 22 | 39885437 | 39924137 | 70 | 18 | 49411 | 2.0658 | 0.019423 | *MIEF1* |
| ENSG00000196176 | 6 | 26011907 | 26032278 | 58 | 16 | 43523 | 2.0652 | 0.019452 | *HIST1H4A* |

| | | | | | | | | | |
|---|---|---|---|---|---|---|---|---|---|
| ENSG00000186654 | 22 | 45054593 | 45143561 | 496 | 89 | 43873 | 2.0646 | 0.019482 | *PRR5* |
| ENSG00000176547 | 11 | 48336472 | 48357507 | 74 | 11 | 32630 | 2.064 | 0.01951 | *OR4C3* |
| ENSG00000130167 | 19 | 11396824 | 11447672 | 148 | 23 | 35474 | 2.0638 | 0.019516 | *TSPAN16* |
| ENSG00000121152 | 2 | 96991525 | 97049583 | 73 | 15 | 32865 | 2.0637 | 0.019523 | *NCAPH* |
| ENSG00000165392 | 8 | 30881317 | 31041285 | 384 | 46 | 50240 | 2.0621 | 0.0196 | *WRN* |
| ENSG00000087008 | 4 | 8358009 | 8452450 | 420 | 52 | 45723 | 2.0606 | 0.019671 | *ACOX3* |
| ENSG00000176472 | 19 | 44019649 | 44050282 | 111 | 19 | 47598 | 2.0596 | 0.019717 | *ZNF575* |
| ENSG00000180957 | 22 | 28237657 | 28326122 | 170 | 23 | 50063 | 2.0596 | 0.019717 | *PITPNB* |
| ENSG00000163283 | 2 | 233233244 | 233257599 | 104 | 27 | 49614 | 2.0595 | 0.019723 | *ALPP* |
| ENSG00000125977 | 20 | 32666104 | 32710138 | 64 | 11 | 42256 | 2.0591 | 0.01974 | *EIF2S2* |
| ENSG00000188636 | 22 | 44878452 | 44904178 | 94 | 16 | 40079 | 2.059 | 0.019748 | *LDOC1L* |
| ENSG00000188191 | 7 | 578834 | 777287 | 694 | 95 | 47944 | 2.0589 | 0.01975 | *PRKAR1B* |
| ENSG00000104112 | 15 | 51963550 | 52023223 | 168 | 27 | 52453 | 2.0585 | 0.019769 | *SCG3* |
| ENSG00000090060 | 14 | 96957770 | 97043448 | 171 | 20 | 53746 | 2.0584 | 0.019778 | *PAPOLA* |
| ENSG00000171806 | 1 | 169751670 | 169774107 | 102 | 17 | 55119 | 2.0582 | 0.019786 | *METTL18* |
| ENSG00000187049 | 11 | 61149159 | 61176335 | 22 | 8 | 40640 | 2.0579 | 0.019801 | *TMEM216* |
| ENSG00000131650 | 16 | 3003945 | 3028384 | 74 | 14 | 46722 | 2.0576 | 0.019816 | *KREMEN2* |
| ENSG00000126266 | 19 | 35832445 | 35853367 | 99 | 34 | 42634 | 2.0574 | 0.019822 | *FFAR1* |
| ENSG00000186952 | 5 | 109614934 | 110084657 | 1267 | 68 | 37166 | 2.0568 | 0.019852 | *TMEM232* |
| ENSG00000105516 | 19 | 49123287 | 49150695 | 54 | 10 | 33331 | 2.0555 | 0.019914 | *DBP* |
| ENSG00000136231 | 7 | 23339828 | 23520086 | 461 | 66 | 43299 | 2.0542 | 0.019976 | *IGF2BP3* |
| ENSG00000166794 | 15 | 64438011 | 64465404 | 55 | 11 | 43996 | 2.0536 | 0.02001 | *PPIB* |
| ENSG00000204946 | 7 | 148949262 | 149004393 | 179 | 27 | 47495 | 2.0533 | 0.020024 | *ZNF783* |
| ENSG00000204491 | 1 | 13464689 | 13487522 | 3 | 1 | 44440 | 2.0528 | 0.020047 | *PRAMEF18* |
| ENSG00000181513 | 17 | 43199967 | 43231548 | 74 | 20 | 50792 | 2.052 | 0.020085 | *ACBD4* |
| ENSG00000172667 | 3 | 178725011 | 178800067 | 178 | 24 | 52264 | 2.0511 | 0.020127 | *ZMAT3* |
| ENSG00000079277 | 1 | 47013090 | 47092515 | 167 | 25 | 42141 | 2.0503 | 0.020165 | *MKNK1* |
| ENSG00000048392 | 8 | 103206730 | 103261346 | 196 | 26 | 52077 | 2.0502 | 0.020172 | *RRM2B* |
| ENSG00000168334 | 3 | 39214701 | 39244087 | 61 | 18 | 51386 | 2.0498 | 0.020192 | *XIRP1* |
| ENSG00000174564 | 3 | 136655072 | 136739927 | 170 | 16 | 38806 | 2.0497 | 0.020196 | *IL20RB* |
| ENSG00000172232 | 19 | 815097 | 842017 | 106 | 31 | 46538 | 2.0497 | 0.020196 | *AZU1* |
| ENSG00000174989 | 12 | 117338761 | 117478953 | 340 | 28 | 30281 | 2.0491 | 0.020225 | *FBXW8* |
| ENSG00000142959 | 1 | 45239257 | 45263377 | 54 | 10 | 44684 | 2.0491 | 0.020227 | *BEST4* |
| ENSG00000143951 | 2 | 63338518 | 64064977 | 1745 | 69 | 43569 | 2.049 | 0.020232 | *WDPCP* |
| ENSG00000154511 | 1 | 93297724 | 93437057 | 243 | 14 | 41305 | 2.0488 | 0.020241 | *FAM69A* |
| ENSG00000123178 | 13 | 50476842 | 50520626 | 74 | 10 | 49192 | 2.0487 | 0.020244 | *SPRYD7* |

| | | | | | | | | | |
|---|---|---|---|---|---|---|---|---|---|
| ENSG00000180596 | 6 | 26105101 | 26134154 | 106 | 13 | 54940 | 2.0487 | 0.020245 | *HIST1H2BC* |
| ENSG00000197321 | 10 | 29736267 | 30035710 | 1337 | 144 | 51293 | 2.0484 | 0.02026 | *SVIL* |
| ENSG00000261787 | 8 | 67848736 | 67884825 | 42 | 13 | 43775 | 2.0482 | 0.020269 | *TCF24* |
| ENSG00000183733 | 2 | 70994442 | 71027775 | 133 | 18 | 42072 | 2.0479 | 0.020284 | *FIGLA* |
| ENSG00000131943 | 19 | 30181721 | 30216364 | 120 | 20 | 27243 | 2.0471 | 0.020322 | *C19orf12* |
| ENSG00000120833 | 12 | 93953590 | 93987263 | 101 | 20 | 45172 | 2.0469 | 0.020334 | *SOCS2* |
| ENSG00000135577 | 6 | 142369467 | 142419936 | 128 | 15 | 46410 | 2.0467 | 0.020345 | *NMBR* |
| ENSG00000103199 | 16 | 4788240 | 4827625 | 180 | 23 | 48061 | 2.0463 | 0.020361 | *ZNF500* |
| ENSG00000136048 | 12 | 102261129 | 102415908 | 616 | 84 | 41637 | 2.0453 | 0.020412 | *DRAM1* |
| ENSG00000117480 | 1 | 46849937 | 46889520 | 104 | 17 | 37906 | 2.0447 | 0.02044 | *FAAH* |
| ENSG00000130695 | 1 | 26550691 | 26615299 | 189 | 15 | 46483 | 2.0447 | 0.020442 | *CEP85* |
| ENSG00000186815 | 12 | 113648855 | 113746390 | 210 | 19 | 55531 | 2.0431 | 0.020519 | *TPCN1* |
| ENSG00000176555 | 11 | 48317775 | 48338704 | 92 | 11 | 31645 | 2.0428 | 0.020537 | *OR4S1* |
| ENSG00000068745 | 3 | 48715436 | 48787786 | 119 | 16 | 46781 | 2.041 | 0.020627 | *IP6K2* |
| ENSG00000133640 | 12 | 85420092 | 85667002 | 365 | 30 | 45279 | 2.0404 | 0.020653 | *LRRIQ1* |
| ENSG00000178125 | 8 | 67866334 | 67978839 | 134 | 13 | 34355 | 2.0404 | 0.020656 | *PPP1R42* |
| ENSG00000170091 | 5 | 173462607 | 173680504 | 798 | 120 | 42426 | 2.0401 | 0.02067 | *NSG2* |
| ENSG00000069018 | 5 | 135538999 | 135742730 | 564 | 47 | 50214 | 2.0383 | 0.020759 | *TRPC7* |
| ENSG00000104883 | 19 | 7531761 | 7572335 | 176 | 39 | 48483 | 2.036 | 0.020878 | *PEX11G* |
| ENSG00000158815 | 8 | 21889909 | 21916320 | 77 | 32 | 43596 | 2.0359 | 0.02088 | *FGF17* |
| ENSG00000086991 | 11 | 89047524 | 89332779 | 679 | 79 | 39345 | 2.0348 | 0.020934 | *NOX4* |
| ENSG00000203747 | 1 | 161501549 | 161610917 | 168 | 45 | 35591 | 2.0343 | 0.020958 | *FCGR3A* |
| ENSG00000185842 | 1 | 225073964 | 225596996 | 1731 | 72 | 37607 | 2.0344 | 0.020958 | *DNAH14* |
| ENSG00000214435 | 10 | 104619273 | 104671656 | 185 | 18 | 49500 | 2.0337 | 0.020992 | *AS3MT* |
| ENSG00000185264 | 22 | 37377163 | 37413882 | 207 | 23 | 52298 | 2.0331 | 0.021022 | *TEX33* |
| ENSG00000110455 | 11 | 44077475 | 44115772 | 143 | 27 | 53008 | 2.0326 | 0.021049 | *ACCS* |
| ENSG00000171016 | 15 | 55821088 | 55891145 | 284 | 33 | 42999 | 2.0322 | 0.021066 | *PYGO1* |
| ENSG00000115884 | 2 | 20390558 | 20435194 | 175 | 29 | 50799 | 2.0319 | 0.021083 | *SDC1* |
| ENSG00000197980 | 3 | 156533270 | 156773918 | 767 | 53 | 46305 | 2.0316 | 0.021098 | *LEKR1* |
| ENSG00000168488 | 16 | 28824356 | 28858558 | 77 | 7 | 27567 | 2.0314 | 0.021105 | *ATXN2L* |
| ENSG00000103061 | 16 | 68308406 | 68354849 | 105 | 17 | 49094 | 2.0302 | 0.021169 | *SLC7A6OS* |
| ENSG00000158125 | 2 | 31547187 | 31647581 | 368 | 71 | 48342 | 2.0298 | 0.021188 | *XDH* |
| ENSG00000065609 | 6 | 84252599 | 84429410 | 440 | 48 | 50100 | 2.0292 | 0.021217 | *SNAP91* |
| ENSG00000131165 | 16 | 89700839 | 89734253 | 134 | 25 | 42518 | 2.0289 | 0.021235 | *CHMP1A* |
| ENSG00000103056 | 16 | 68382231 | 68492591 | 261 | 34 | 47974 | 2.0285 | 0.021254 | *SMPD3* |
| ENSG00000163793 | 2 | 27488289 | 27514367 | 42 | 10 | 40783 | 2.0283 | 0.021265 | *DNAJC5G* |

| | | | | | | | | | |
|---|---|---|---|---|---|---|---|---|---|
| ENSG00000126070 | 1 | 36386319 | 36548101 | 110 | 29 | 51838 | 2.0279 | 0.021287 | *AGO3* |
| ENSG00000010244 | 17 | 30667136 | 30718905 | 91 | 16 | 40794 | 2.0271 | 0.021327 | *ZNF207* |
| ENSG00000133116 | 13 | 33580207 | 33650282 | 257 | 30 | 44971 | 2.0267 | 0.021346 | *KL* |
| ENSG00000204482 | 6 | 31543901 | 31566686 | 108 | 31 | 43379 | 2.026 | 0.021384 | *LST1* |
| ENSG00000158793 | 1 | 161077876 | 161105235 | 52 | 9 | 53961 | 2.0251 | 0.021428 | *NIT1* |
| ENSG00000185056 | 5 | 173390782 | 173443143 | 218 | 20 | 39734 | 2.025 | 0.021436 | *C5orf47* |
| ENSG00000015413 | 16 | 89669716 | 89714839 | 173 | 33 | 48080 | 2.0248 | 0.021442 | *DPEP1* |
| ENSG00000115828 | 2 | 37561717 | 37610465 | 180 | 41 | 46705 | 2.0231 | 0.021529 | *QPCT* |
| ENSG00000170956 | 19 | 42290369 | 42325591 | 53 | 6 | 30994 | 2.0228 | 0.021546 | *CEACAM3* |
| ENSG00000196132 | 20 | 62773144 | 62883604 | 384 | 72 | 46024 | 2.0224 | 0.021566 | *MYT1* |
| ENSG00000160741 | 1 | 153910145 | 153941101 | 47 | 10 | 49364 | 2.0215 | 0.021611 | *CRTC2* |
| ENSG00000172774 | 11 | 57972217 | 57993195 | 96 | 13 | 46086 | 2.0214 | 0.02162 | *OR1S1* |
| ENSG00000153879 | 19 | 33854236 | 33883592 | 109 | 17 | 48727 | 2.0209 | 0.021647 | *CEBPG* |
| ENSG00000185565 | 3 | 115511235 | 117726095 | 7123 | 432 | 44395 | 2.0207 | 0.021653 | *LSAMP* |
| ENSG00000104093 | 15 | 51729908 | 51925030 | 515 | 22 | 55031 | 2.0196 | 0.021712 | *DMXL2* |
| ENSG00000196532 | 6 | 26035639 | 26056097 | 60 | 14 | 40531 | 2.0191 | 0.021738 | *HIST1H3C* |
| ENSG00000115194 | 2 | 27466552 | 27508685 | 88 | 16 | 41916 | 2.0181 | 0.021792 | *SLC30A3* |
| ENSG00000170727 | 8 | 145476055 | 145525082 | 57 | 7 | 51694 | 2.0177 | 0.02181 | *BOP1* |
| ENSG00000080802 | 7 | 135036547 | 135204875 | 365 | 47 | 54538 | 2.0176 | 0.021814 | *CNOT4* |
| ENSG00000188848 | 4 | 42102955 | 42164895 | 206 | 28 | 40672 | 2.0173 | 0.021834 | *BEND4* |
| ENSG00000057935 | 2 | 42711709 | 42994087 | 730 | 50 | 45333 | 2.0171 | 0.021841 | *MTA3* |
| ENSG00000161813 | 12 | 50776166 | 50883787 | 87 | 33 | 41141 | 2.0157 | 0.021918 | *LARP4* |
| ENSG00000066923 | 7 | 99765186 | 99829111 | 162 | 12 | 46842 | 2.0156 | 0.021922 | *STAG3* |
| ENSG00000125753 | 19 | 45999837 | 46040241 | 123 | 22 | 52182 | 2.015 | 0.021954 | *VASP* |
| ENSG00000197263 | 11 | 124179134 | 124200184 | 75 | 13 | 42311 | 2.0144 | 0.021982 | *OR8D2* |
| ENSG00000122971 | 12 | 121153538 | 121187811 | 121 | 23 | 42832 | 2.014 | 0.022006 | *ACADS* |
| ENSG00000186094 | 1 | 48988527 | 50499585 | 2425 | 118 | 37620 | 2.0135 | 0.022029 | *AGBL4* |
| ENSG00000152822 | 6 | 146338782 | 146768734 | 818 | 91 | 43046 | 2.0133 | 0.022041 | *GRM1* |
| ENSG00000089101 | 20 | 20023158 | 20351346 | 826 | 130 | 51325 | 2.0124 | 0.022089 | *C20orf26* |
| ENSG00000007392 | 16 | 228968 | 289462 | 144 | 24 | 46485 | 2.0117 | 0.022124 | *LUC7L* |
| ENSG00000134352 | 5 | 55220923 | 55300821 | 157 | 23 | 41332 | 2.0106 | 0.022186 | *IL6ST* |
| ENSG00000179195 | 12 | 124446392 | 124509986 | 132 | 16 | 48748 | 2.0105 | 0.022192 | *ZNF664* |
| ENSG00000100095 | 22 | 26555440 | 26789562 | 954 | 143 | 43380 | 2.0093 | 0.022253 | *SEZ6L* |
| ENSG00000090263 | 7 | 140695854 | 140725028 | 68 | 10 | 49500 | 2.009 | 0.02227 | *MRPS33* |
| ENSG00000112667 | 6 | 43183367 | 43207222 | 84 | 17 | 44460 | 2.0086 | 0.02229 | *DNPH1* |
| ENSG00000178952 | 16 | 28843732 | 28867729 | 49 | 6 | 25919 | 2.0085 | 0.022295 | *TUFM* |

| | | | | | | | | | |
|---|---|---|---|---|---|---|---|---|---|
| ENSG00000169231 | 1 | 155155379 | 155188842 | 52 | 13 | 48569 | 2.0084 | 0.022302 | *THBS3* |
| ENSG00000221838 | 7 | 99689172 | 99717968 | 64 | 11 | 51647 | 2.008 | 0.022322 | *AP4M1* |
| ENSG00000152582 | 5 | 35607946 | 35824713 | 728 | 66 | 44210 | 2.0076 | 0.022343 | *SPEF2* |
| ENSG00000145780 | 5 | 114846608 | 114890591 | 88 | 19 | 52254 | 2.0064 | 0.022405 | *FEM1C* |
| ENSG00000164175 | 5 | 33934721 | 33994835 | 87 | 28 | 29558 | 2.0062 | 0.022415 | *SLC45A2* |
| ENSG00000138347 | 10 | 69855912 | 69981774 | 561 | 53 | 41721 | 2.0059 | 0.022432 | *MYPN* |
| ENSG00000259316 | 15 | 64462547 | 64675980 | 426 | 34 | 44084 | 2.0057 | 0.022446 | *CTD-2116N17.1* |
| ENSG00000172046 | 3 | 49135479 | 49168371 | 32 | 11 | 43681 | 2.0054 | 0.022462 | *USP19* |
| ENSG00000119335 | 9 | 131435703 | 131468679 | 25 | 10 | 53803 | 2.003 | 0.02259 | *SET* |
| ENSG00000168090 | 7 | 99676577 | 99699823 | 48 | 9 | 50512 | 2.0022 | 0.022631 | *COPS6* |
| ENSG00000168079 | 8 | 27717399 | 27860244 | 640 | 88 | 48027 | 2.0019 | 0.022647 | *SCARA5* |
| ENSG00000135870 | 1 | 173890352 | 174001435 | 231 | 19 | 34800 | 2.0018 | 0.02265 | *RC3H1* |
| ENSG00000145191 | 3 | 183842826 | 184412546 | 2008 | 241 | 47718 | 2.0017 | 0.022658 | *EIF2B5* |
| ENSG00000163870 | 3 | 127281912 | 127327094 | 46 | 17 | 54196 | 2.0013 | 0.022678 | *TPRA1* |
| ENSG00000123892 | 11 | 87836410 | 87918635 | 346 | 36 | 50747 | 2.0003 | 0.022732 | *RAB38* |
| ENSG00000010704 | 6 | 26077509 | 26108571 | 106 | 21 | 41094 | 1.9993 | 0.022787 | *HFE* |
| ENSG00000121022 | 8 | 67945314 | 68006018 | 90 | 10 | 34727 | 1.999 | 0.022804 | *COPS5* |
| ENSG00000131188 | 5 | 176863446 | 176893283 | 69 | 11 | 33924 | 1.9982 | 0.022849 | *PRR7* |
| ENSG00000061794 | 12 | 27853706 | 27919228 | 187 | 18 | 54217 | 1.9977 | 0.022872 | *MRPS35* |
| ENSG00000225996 | 1 | 150511897 | 150534367 | 61 | 22 | 45465 | 1.9964 | 0.022943 | *AL356356.1* |
| ENSG00000185567 | 14 | 105393581 | 105454694 | 177 | 27 | 46419 | 1.9962 | 0.022955 | *AHNAK2* |
| ENSG00000248405 | 22 | 45088113 | 45268586 | 1213 | 177 | 43083 | 1.9954 | 0.022998 | *PRR5-ARHGAP8* |
| ENSG00000188603 | 16 | 28467983 | 28516896 | 67 | 14 | 38993 | 1.9952 | 0.023008 | *CLN3* |
| ENSG00000164080 | 3 | 51565160 | 51712683 | 118 | 35 | 52109 | 1.9952 | 0.023013 | *RAD54L2* |
| ENSG00000103044 | 16 | 69129467 | 69162622 | 59 | 18 | 55069 | 1.995 | 0.023023 | *HAS3* |
| ENSG00000271810 | 1 | 113235236 | 113264055 | 74 | 19 | 53037 | 1.9939 | 0.023082 | *RP11-426L16.10* |
| ENSG00000253350 | 8 | 105592961 | 106441488 | 2505 | 205 | 44151 | 1.9933 | 0.023116 | *RP11-127H5.1* |
| ENSG00000175711 | 17 | 80890031 | 81019686 | 700 | 86 | 51063 | 1.9931 | 0.023126 | *B3GNTL1* |
| ENSG00000115183 | 2 | 159815146 | 160099170 | 940 | 108 | 39846 | 1.9917 | 0.023203 | *TANC1* |
| ENSG00000168806 | 15 | 43609974 | 43632803 | 35 | 8 | 35956 | 1.9913 | 0.023222 | *LCMT2* |
| ENSG00000150961 | 4 | 119633978 | 119769838 | 527 | 23 | 48390 | 1.9911 | 0.023235 | *SEC24D* |
| ENSG00000137171 | 6 | 42998515 | 43052837 | 140 | 19 | 38789 | 1.9909 | 0.023245 | *KLC4* |
| ENSG00000142937 | 1 | 45230923 | 45254451 | 45 | 9 | 41892 | 1.9908 | 0.023254 | *RPS8* |
| ENSG00000162552 | 1 | 22433798 | 22480462 | 169 | 30 | 47129 | 1.9904 | 0.023271 | *WNT4* |
| ENSG00000126249 | 19 | 34885289 | 34927073 | 93 | 20 | 38754 | 1.9904 | 0.023271 | *PDCD2L* |
| ENSG00000268242 | 11 | 11737060 | 11757257 | 65 | 18 | 49453 | 1.9899 | 0.023301 | *AC131935.1* |

| | | | | | | | | | |
|---|---|---|---|---|---|---|---|---|---|
| ENSG00000163915 | 3 | 185420316 | 185457575 | 49 | 17 | 48466 | 1.9878 | 0.023415 | *C3orf65* |
| ENSG00000171517 | 1 | 85267285 | 85368896 | 388 | 74 | 52030 | 1.9878 | 0.023419 | *LPAR3* |
| ENSG00000168591 | 17 | 42254338 | 42279099 | 48 | 12 | 41375 | 1.9871 | 0.023455 | *TMUB2* |
| ENSG00000164306 | 4 | 185560767 | 185626117 | 305 | 21 | 47344 | 1.9865 | 0.023491 | *PRIMPOL* |
| ENSG00000120328 | 5 | 140578269 | 140601696 | 68 | 16 | 41651 | 1.9864 | 0.023496 | *PCDHB12* |
| ENSG00000141579 | 17 | 80777311 | 80808454 | 137 | 18 | 47122 | 1.9858 | 0.023526 | *ZNF750* |
| ENSG00000081665 | 19 | 19886648 | 19942560 | 165 | 22 | 43185 | 1.9856 | 0.023538 | *ZNF506* |
| ENSG00000176349 | 7 | 1868222 | 1899567 | 189 | 18 | 44478 | 1.9847 | 0.023592 | *AC110781.3* |
| ENSG00000241484 | 22 | 45088355 | 45268665 | 1211 | 172 | 43097 | 1.9841 | 0.023621 | *ARHGAP8* |
| ENSG00000168928 | 16 | 75227994 | 75251083 | 180 | 27 | 47920 | 1.9841 | 0.023623 | *CTRB2* |
| ENSG00000125447 | 17 | 73222694 | 73268444 | 91 | 15 | 40655 | 1.984 | 0.023629 | *GGA3* |
| ENSG00000108175 | 10 | 80818792 | 81086276 | 951 | 180 | 42702 | 1.983 | 0.023685 | *ZMIZ1* |
| ENSG00000180573 | 6 | 26114373 | 26149344 | 93 | 15 | 53081 | 1.9829 | 0.023687 | *HIST1H2AC* |
| ENSG00000143570 | 1 | 153921575 | 153950188 | 41 | 9 | 49224 | 1.9826 | 0.023704 | *SLC39A1* |
| ENSG00000112234 | 6 | 99306420 | 99405849 | 228 | 24 | 52624 | 1.9824 | 0.023719 | *FBXL4* |
| ENSG00000100865 | 14 | 102798956 | 102839253 | 90 | 22 | 46660 | 1.9823 | 0.023725 | *CINP* |
| ENSG00000100991 | 20 | 33580207 | 33690674 | 347 | 23 | 50019 | 1.9823 | 0.023725 | *TRPC4AP* |
| ENSG00000150995 | 3 | 4525032 | 4899524 | 1288 | 193 | 41148 | 1.982 | 0.02374 | *ITPR1* |
| ENSG00000092964 | 8 | 26361791 | 26525694 | 757 | 76 | 44053 | 1.9814 | 0.023775 | *DPYSL2* |
| ENSG00000166250 | 11 | 122933035 | 123075989 | 670 | 82 | 45903 | 1.9806 | 0.023817 | *CLMP* |
| ENSG00000115827 | 2 | 172280727 | 172351562 | 234 | 18 | 50381 | 1.9804 | 0.023829 | *DCAF17* |
| ENSG00000163659 | 3 | 156381024 | 156434559 | 136 | 17 | 56580 | 1.9802 | 0.023841 | *TIPARP* |
| ENSG00000158062 | 1 | 26597819 | 26654854 | 155 | 15 | 41487 | 1.98 | 0.023851 | *UBXN11* |
| ENSG00000254636 | 10 | 124204169 | 124226868 | 87 | 14 | 51735 | 1.9796 | 0.023877 | *ARMS2* |
| ENSG00000171115 | 7 | 150137718 | 150186480 | 98 | 23 | 46238 | 1.9794 | 0.023884 | *GIMAP8* |
| ENSG00000164754 | 8 | 117848174 | 117897105 | 146 | 19 | 31805 | 1.9787 | 0.023927 | *RAD21* |
| ENSG00000187079 | 11 | 12685969 | 12976298 | 593 | 74 | 46517 | 1.9785 | 0.023939 | *TEAD1* |
| ENSG00000172365 | 11 | 58179738 | 58200786 | 106 | 6 | 53091 | 1.9774 | 0.024 | *OR5B2* |
| ENSG00000007968 | 1 | 23822922 | 23867712 | 119 | 21 | 34016 | 1.9771 | 0.024014 | *E2F2* |
| ENSG00000151846 | 13 | 25660300 | 25683389 | 58 | 11 | 54440 | 1.9765 | 0.02405 | *PABPC3* |
| ENSG00000166748 | 15 | 86675227 | 87582283 | 4153 | 302 | 46341 | 1.9762 | 0.024068 | *AGBL1* |
| ENSG00000109458 | 4 | 144247915 | 144405721 | 355 | 48 | 46799 | 1.9761 | 0.02407 | *GAB1* |
| ENSG00000261832 | 16 | 28467974 | 28513333 | 58 | 12 | 37663 | 1.9754 | 0.024109 | *CLN3* |
| ENSG00000119673 | 14 | 74024324 | 74052357 | 135 | 29 | 27629 | 1.9749 | 0.024141 | *ACOT2* |
| ENSG00000189079 | 12 | 46113448 | 46311823 | 214 | 37 | 46689 | 1.9745 | 0.024163 | *ARID2* |
| ENSG00000171604 | 5 | 139016884 | 139073467 | 111 | 18 | 49225 | 1.9744 | 0.024169 | *CXXC5* |

| | | | | | | | | | |
|---|---|---|---|---|---|---|---|---|---|
| ENSG00000147481 | 8 | 50812349 | 51716678 | 3019 | 115 | 42709 | 1.9738 | 0.024203 | *SNTG1* |
| ENSG00000126777 | 14 | 56015790 | 56178244 | 572 | 26 | 53437 | 1.9733 | 0.024233 | *KTN1* |
| ENSG00000213413 | 7 | 99805864 | 99829113 | 81 | 9 | 45405 | 1.9731 | 0.02424 | *PVRIG* |
| ENSG00000070476 | 3 | 126146444 | 126204762 | 234 | 31 | 45815 | 1.9729 | 0.024255 | *ZXDC* |
| ENSG00000169641 | 1 | 23400516 | 23514301 | 274 | 26 | 44798 | 1.9728 | 0.024259 | *LUZP1* |
| ENSG00000136156 | 13 | 48797294 | 48847063 | 79 | 14 | 41027 | 1.9721 | 0.024299 | *ITM2B* |
| ENSG00000110435 | 11 | 34927376 | 35052138 | 603 | 33 | 31619 | 1.9718 | 0.024319 | *PDHX* |
| ENSG00000174099 | 12 | 65662423 | 65892024 | 426 | 40 | 44255 | 1.9717 | 0.024324 | *MSRB3* |
| ENSG00000125124 | 16 | 56490748 | 56564195 | 183 | 24 | 38724 | 1.9712 | 0.024348 | *BBS2* |
| ENSG00000125818 | 20 | 1083906 | 1170596 | 213 | 43 | 49264 | 1.97 | 0.024417 | *PSMF1* |
| ENSG00000087077 | 7 | 100454760 | 100481076 | 66 | 12 | 42941 | 1.9699 | 0.024422 | *TRIP6* |
| ENSG00000169228 | 5 | 176718199 | 176740745 | 57 | 14 | 39960 | 1.9694 | 0.024455 | *RAB24* |
| ENSG00000132005 | 19 | 14062350 | 14127851 | 59 | 26 | 46463 | 1.969 | 0.024476 | *RFX1* |
| ENSG00000172362 | 11 | 58196593 | 58217646 | 72 | 7 | 53766 | 1.9688 | 0.02449 | *OR5B12* |
| ENSG00000143546 | 1 | 153352508 | 153373664 | 65 | 6 | 54035 | 1.9686 | 0.024497 | *S100A8* |
| ENSG00000141013 | 16 | 90076037 | 90121383 | 194 | 32 | 36884 | 1.9683 | 0.024519 | *GAS8* |
| ENSG00000104218 | 8 | 67964661 | 68118498 | 207 | 21 | 37719 | 1.9681 | 0.024526 | *CSPP1* |
| ENSG00000095752 | 19 | 55865757 | 55891831 | 96 | 21 | 42248 | 1.9676 | 0.024556 | *IL11* |
| ENSG00000139117 | 12 | 39030624 | 39311232 | 746 | 69 | 33750 | 1.9669 | 0.024599 | *CPNE8* |
| ENSG00000181163 | 5 | 170804120 | 170848141 | 159 | 12 | 55034 | 1.9667 | 0.024608 | *NPM1* |
| ENSG00000168917 | 3 | 136527489 | 136584734 | 180 | 9 | 55054 | 1.9663 | 0.024635 | *SLC35G2* |
| ENSG00000169118 | 15 | 64447716 | 64658442 | 424 | 34 | 44343 | 1.9659 | 0.024654 | *CSNK1G1* |
| ENSG00000168830 | 6 | 87637024 | 87736349 | 297 | 32 | 39805 | 1.9656 | 0.024675 | *HTR1E* |
| ENSG00000162374 | 1 | 50503686 | 50679458 | 218 | 43 | 41226 | 1.9655 | 0.024678 | *ELAVL4* |
| ENSG00000157045 | 16 | 15121710 | 15159921 | 110 | 13 | 45957 | 1.9655 | 0.024678 | *NTAN1* |
| ENSG00000111832 | 6 | 116882530 | 116928838 | 127 | 19 | 53719 | 1.9646 | 0.024729 | *RWDD1* |
| ENSG00000180855 | 19 | 12530521 | 12561926 | 280 | 11 | 47583 | 1.9639 | 0.02477 | *ZNF443* |
| ENSG00000153283 | 3 | 111001566 | 111394597 | 1228 | 93 | 38207 | 1.9639 | 0.024773 | *CD96* |
| ENSG00000213347 | 5 | 176718462 | 176749758 | 69 | 17 | 40225 | 1.9636 | 0.024788 | *MXD3* |
| ENSG00000163749 | 4 | 77224154 | 77353021 | 258 | 26 | 43803 | 1.9633 | 0.024804 | *CCDC158* |
| ENSG00000270316 | 10 | 104604029 | 104671656 | 229 | 22 | 49294 | 1.9629 | 0.024826 | *C10orf32-ASMT* |
| ENSG00000182752 | 9 | 118906083 | 119174601 | 513 | 107 | 46740 | 1.9629 | 0.024827 | *PAPPA* |
| ENSG00000196800 | 5 | 147539296 | 147564961 | 158 | 6 | 49120 | 1.9627 | 0.02484 | *SPINK14* |
| ENSG00000137845 | 15 | 58877403 | 59052177 | 320 | 36 | 37272 | 1.9627 | 0.024843 | *ADAM10* |
| ENSG00000224186 | 5 | 134358970 | 134701744 | 843 | 139 | 41795 | 1.9624 | 0.024857 | *C5orf66* |
| ENSG00000122042 | 13 | 30328508 | 30434821 | 299 | 25 | 53511 | 1.9617 | 0.024899 | *UBL3* |

| | | | | | | | | | |
|---|---|---|---|---|---|---|---|---|---|
| ENSG00000269783 | 10 | 99617889 | 99641294 | 55 | 13 | 39211 | 1.961 | 0.024939 | *GOLGA7B* |
| ENSG00000124496 | 6 | 42182669 | 42429789 | 659 | 93 | 39371 | 1.9609 | 0.024945 | *TRERF1* |
| ENSG00000118564 | 4 | 15596162 | 15693302 | 308 | 24 | 55254 | 1.9604 | 0.024977 | *FBXL5* |
| ENSG00000105220 | 19 | 34840385 | 34903061 | 155 | 27 | 39058 | 1.9597 | 0.025015 | *GPI* |
| ENSG00000100678 | 14 | 70500934 | 70665787 | 677 | 83 | 52625 | 1.9594 | 0.025034 | *SLC8A3* |
| ENSG00000069399 | 19 | 45240962 | 45273301 | 117 | 26 | 41366 | 1.9584 | 0.025093 | *BCL3* |
| ENSG00000053371 | 1 | 19620459 | 19648640 | 106 | 24 | 53283 | 1.9581 | 0.025109 | *AKR7A2* |
| ENSG00000172461 | 6 | 96453860 | 96673488 | 860 | 80 | 47345 | 1.9569 | 0.025179 | *FUT9* |
| ENSG00000196620 | 4 | 69502348 | 69546346 | 101 | 23 | 48235 | 1.9567 | 0.02519 | *UGT2B15* |
| ENSG00000130706 | 20 | 60867149 | 60893918 | 101 | 25 | 48787 | 1.9566 | 0.025194 | *ADRM1* |
| ENSG00000214325 | 16 | 75718367 | 75744089 | 65 | 10 | 43633 | 1.9561 | 0.025228 | *AC025287.1* |
| ENSG00000151117 | 11 | 18704669 | 18736332 | 106 | 28 | 48778 | 1.9557 | 0.025248 | *TMEM86A* |
| ENSG00000170923 | 19 | 9202945 | 9223982 | 119 | 9 | 48687 | 1.9554 | 0.025267 | *OR7G2* |
| ENSG00000157837 | 12 | 121190313 | 121352174 | 582 | 41 | 34712 | 1.9549 | 0.025297 | *SPPL3* |
| ENSG00000170381 | 7 | 82983222 | 83288326 | 1195 | 76 | 39297 | 1.9549 | 0.0253 | *SEMA3E* |
| ENSG00000165156 | 8 | 124250696 | 124297781 | 26 | 17 | 46523 | 1.9543 | 0.025332 | *ZHX1* |
| ENSG00000205302 | 5 | 122100691 | 122175803 | 305 | 25 | 44113 | 1.9542 | 0.025337 | *SNX2* |
| ENSG00000163704 | 3 | 9977226 | 10004078 | 65 | 12 | 38087 | 1.9535 | 0.025381 | *PRRT3* |
| ENSG00000143578 | 1 | 153930010 | 153956839 | 41 | 10 | 48816 | 1.9535 | 0.025382 | *CREB3L4* |
| ENSG00000166508 | 7 | 99680351 | 99709563 | 67 | 10 | 50561 | 1.9506 | 0.025552 | *MCM7* |
| ENSG00000178234 | 7 | 151712759 | 151829425 | 147 | 23 | 33070 | 1.9505 | 0.025557 | *GALNT11* |
| ENSG00000214944 | 5 | 72911983 | 73247818 | 1105 | 120 | 47539 | 1.9503 | 0.025571 | *ARHGEF28* |
| ENSG00000038210 | 4 | 25152263 | 25290714 | 450 | 52 | 43707 | 1.9497 | 0.025604 | *PI4K2B* |
| ENSG00000157216 | 1 | 54681105 | 54889152 | 631 | 111 | 44003 | 1.9484 | 0.025685 | *SSBP3* |
| ENSG00000122033 | 13 | 27999776 | 28034739 | 136 | 18 | 42562 | 1.9483 | 0.025688 | *MTIF3* |
| ENSG00000179144 | 7 | 150201918 | 150228161 | 134 | 20 | 48585 | 1.9483 | 0.025692 | *GIMAP7* |
| ENSG00000184897 | 3 | 129023615 | 129045120 | 40 | 8 | 41856 | 1.9482 | 0.025697 | *H1FX* |
| ENSG00000054967 | 11 | 73077309 | 73118519 | 85 | 18 | 42521 | 1.9479 | 0.025712 | *RELT* |
| ENSG00000233670 | 17 | 10715792 | 10751762 | 98 | 23 | 49380 | 1.9478 | 0.025722 | *PIRT* |
| ENSG00000103994 | 15 | 42695021 | 42793321 | 191 | 28 | 40837 | 1.9468 | 0.025778 | *ZNF106* |
| ENSG00000218357 | 22 | 47847048 | 47892860 | 141 | 23 | 50234 | 1.9462 | 0.025817 | *LL22NC03-75H12.2* |
| ENSG00000003249 | 16 | 90061273 | 90096536 | 151 | 32 | 37572 | 1.9456 | 0.025854 | *DBNDD1* |
| ENSG00000005001 | 16 | 2892728 | 2918171 | 98 | 15 | 47083 | 1.945 | 0.025888 | *PRSS22* |
| ENSG00000172772 | 11 | 58024264 | 58045732 | 108 | 10 | 44252 | 1.9446 | 0.025911 | *OR10W1* |
| ENSG00000174595 | 7 | 130407401 | 130428888 | 78 | 18 | 41848 | 1.944 | 0.025948 | *KLF14* |
| ENSG00000175727 | 12 | 122506628 | 122641894 | 294 | 15 | 54578 | 1.9439 | 0.025956 | *MLXIP* |

| | | | | | | | | | |
|---|---|---|---|---|---|---|---|---|---|
| ENSG00000196934 | 22 | 21727663 | 21753446 | 5 | 3 | 42635 | 1.9436 | 0.025972 | *RIMBP3B* |
| ENSG00000170921 | 17 | 61076917 | 61515060 | 807 | 64 | 47429 | 1.9434 | 0.025987 | *TANC2* |
| ENSG00000234745 | 6 | 31311649 | 31334965 | 1022 | 84 | 38590 | 1.9425 | 0.026036 | *HLA-B* |
| ENSG00000240682 | 3 | 128836258 | 128890136 | 57 | 16 | 36909 | 1.9424 | 0.026047 | *ISY1* |
| ENSG00000174695 | 5 | 82338665 | 82383682 | 122 | 13 | 38980 | 1.9421 | 0.026061 | *TMEM167A* |
| ENSG00000255394 | 8 | 11608896 | 11630309 | 137 | 25 | 44060 | 1.9421 | 0.026062 | *C8orf49* |
| ENSG00000203663 | 1 | 248191474 | 248212607 | 35 | 7 | 42687 | 1.9421 | 0.026065 | *OR2L2* |
| ENSG00000108278 | 17 | 34832473 | 34865154 | 70 | 13 | 34681 | 1.9417 | 0.026088 | *ZNHIT3* |
| ENSG00000103064 | 16 | 68288433 | 68345722 | 125 | 19 | 49725 | 1.9416 | 0.026094 | *SLC7A6* |
| ENSG00000197889 | 10 | 14991438 | 15040049 | 241 | 21 | 48523 | 1.9414 | 0.026108 | *MEIG1* |
| ENSG00000196943 | 14 | 24759068 | 24788330 | 78 | 14 | 40326 | 1.9405 | 0.02616 | *NOP9* |
| ENSG00000222004 | 7 | 26667490 | 26696889 | 77 | 10 | 36670 | 1.9398 | 0.0262 | *C7orf71* |
| ENSG00000106290 | 7 | 99694693 | 99727464 | 62 | 13 | 50424 | 1.9393 | 0.026233 | *TAF6* |
| ENSG00000163482 | 2 | 219526749 | 219577439 | 78 | 12 | 34508 | 1.9384 | 0.026289 | *STK36* |
| ENSG00000105227 | 19 | 40889675 | 40929273 | 79 | 22 | 45664 | 1.938 | 0.02631 | *PRX* |
| ENSG00000144668 | 3 | 37483606 | 37875005 | 1147 | 137 | 46306 | 1.9374 | 0.026347 | *ITGA9* |
| ENSG00000166884 | 11 | 59214411 | 59235464 | 109 | 13 | 39469 | 1.9369 | 0.02638 | *OR4D6* |
| ENSG00000167363 | 17 | 80683451 | 80719073 | 135 | 17 | 47922 | 1.9367 | 0.026392 | *FN3K* |
| ENSG00000119973 | 10 | 120342916 | 120365160 | 106 | 17 | 51946 | 1.9364 | 0.026412 | *PRLHR* |
| ENSG00000153250 | 2 | 161118662 | 161360305 | 678 | 67 | 47975 | 1.9358 | 0.026446 | *RBMS1* |
| ENSG00000183379 | 14 | 74862596 | 74902805 | 111 | 19 | 50069 | 1.9352 | 0.026485 | *SYNDIG1L* |
| ENSG00000164244 | 5 | 126843301 | 126900781 | 195 | 15 | 45751 | 1.935 | 0.026498 | *PRRC1* |
| ENSG00000126653 | 17 | 28432539 | 28523493 | 158 | 14 | 35059 | 1.9347 | 0.026514 | *NSRP1* |
| ENSG00000177303 | 17 | 73486342 | 73521664 | 146 | 24 | 40371 | 1.9343 | 0.026537 | *CASKIN2* |
| ENSG00000065665 | 10 | 12161636 | 12221960 | 191 | 20 | 42294 | 1.9336 | 0.026584 | *SEC61A2* |
| ENSG00000260851 | 16 | 66493715 | 66561215 | 120 | 17 | 31482 | 1.932 | 0.026681 | *RP11-403P17.5* |
| ENSG00000077348 | 19 | 41882279 | 41913384 | 87 | 8 | 52353 | 1.9319 | 0.026686 | *EXOSC5* |
| ENSG00000114738 | 3 | 50638951 | 50696720 | 73 | 13 | 43035 | 1.9319 | 0.026689 | *MAPKAPK3* |
| ENSG00000176678 | 16 | 86599974 | 86625303 | 106 | 23 | 44086 | 1.9314 | 0.026717 | *FOXL1* |
| ENSG00000198837 | 1 | 153891977 | 153929172 | 61 | 8 | 51532 | 1.9311 | 0.026734 | *DENND4B* |
| ENSG00000114735 | 3 | 50596583 | 50632366 | 41 | 8 | 45695 | 1.9305 | 0.026774 | *HEMK1* |
| ENSG00000162639 | 1 | 109180912 | 109214148 | 79 | 15 | 34467 | 1.93 | 0.026802 | *HENMT1* |
| ENSG00000105698 | 19 | 35749881 | 35780724 | 90 | 14 | 52349 | 1.9288 | 0.026877 | *USF2* |
| ENSG00000107187 | 9 | 139078096 | 139106955 | 100 | 27 | 50979 | 1.9286 | 0.026892 | *LHX3* |
| ENSG00000111412 | 12 | 117143593 | 117185875 | 150 | 29 | 50940 | 1.9279 | 0.026931 | *C12orf49* |
| ENSG00000107338 | 9 | 37909131 | 38079210 | 535 | 43 | 48937 | 1.9276 | 0.026955 | *SHB* |

| | | | | | | | | | |
|---|---|---|---|---|---|---|---|---|---|
| ENSG00000153002 | 3 | 148498889 | 148587974 | 319 | 54 | 38346 | 1.9275 | 0.026961 | *CPB1* |
| ENSG00000267795 | 16 | 4828398 | 4856492 | 90 | 14 | 51567 | 1.927 | 0.026991 | *SMIM22* |
| ENSG00000179604 | 17 | 71269763 | 71318314 | 146 | 31 | 49292 | 1.9265 | 0.027019 | *CDC42EP4* |
| ENSG00000146066 | 5 | 175805748 | 175826772 | 55 | 13 | 53203 | 1.926 | 0.027053 | *HIGD2A* |
| ENSG00000044090 | 6 | 42995355 | 43031683 | 80 | 14 | 36042 | 1.9259 | 0.027056 | *CUL7* |
| ENSG00000100354 | 22 | 40430821 | 40741812 | 588 | 39 | 37023 | 1.9259 | 0.027059 | *TNRC6B* |
| ENSG00000142227 | 19 | 48814766 | 48843810 | 113 | 19 | 54659 | 1.9258 | 0.027066 | *EMP3* |
| ENSG00000157703 | 7 | 138269030 | 138396097 | 480 | 95 | 45253 | 1.9252 | 0.027099 | *SVOPL* |
| ENSG00000221970 | 7 | 144005218 | 144026150 | 1 | 1 | 43959 | 1.9252 | 0.0271 | *OR2A1* |
| ENSG00000184160 | 4 | 3758075 | 3780251 | 95 | 22 | 38813 | 1.9251 | 0.027106 | *ADRA2C* |
| ENSG00000103168 | 16 | 84201458 | 84230669 | 191 | 32 | 50791 | 1.9239 | 0.027186 | *TAF1C* |
| ENSG00000153975 | 6 | 116946781 | 116999957 | 197 | 22 | 50211 | 1.9236 | 0.027204 | *ZUFSP* |
| ENSG00000103528 | 16 | 19169293 | 19289652 | 286 | 61 | 44777 | 1.9223 | 0.027285 | *SYT17* |
| ENSG00000204475 | 6 | 31546672 | 31570762 | 117 | 33 | 40159 | 1.9222 | 0.027293 | *NCR3* |
| ENSG00000108671 | 17 | 30761279 | 30820336 | 75 | 15 | 43025 | 1.9216 | 0.027328 | *PSMD11* |
| ENSG00000197887 | 11 | 57960674 | 57981653 | 70 | 9 | 49344 | 1.9208 | 0.027382 | *OR1S2* |
| ENSG00000175309 | 5 | 177625498 | 177669792 | 205 | 39 | 46785 | 1.9201 | 0.027425 | *PHYKPL* |
| ENSG00000139800 | 13 | 100605218 | 100634163 | 72 | 17 | 43910 | 1.9186 | 0.027517 | *ZIC5* |
| ENSG00000140320 | 15 | 40721920 | 40770441 | 71 | 23 | 38539 | 1.9182 | 0.027542 | *BAHD1* |
| ENSG00000139174 | 12 | 42842140 | 42994157 | 517 | 76 | 44846 | 1.9178 | 0.027566 | *PRICKLE1* |
| ENSG00000181718 | 11 | 55989490 | 56010737 | 74 | 10 | 56401 | 1.9176 | 0.027578 | *OR5T2* |
| ENSG00000159461 | 16 | 56385364 | 56469450 | 246 | 21 | 36774 | 1.9176 | 0.027582 | *AMFR* |
| ENSG00000180988 | 11 | 5831544 | 5852578 | 157 | 18 | 50942 | 1.9172 | 0.027609 | *OR52N2* |
| ENSG00000140374 | 15 | 76497696 | 76613813 | 274 | 23 | 35101 | 1.917 | 0.02762 | *ETFA* |
| ENSG00000187664 | 19 | 19356450 | 19383605 | 82 | 14 | 38909 | 1.9151 | 0.027737 | *HAPLN4* |
| ENSG00000166819 | 15 | 90197596 | 90232658 | 64 | 23 | 44574 | 1.9147 | 0.027765 | *PLIN1* |
| ENSG00000249709 | 19 | 12626184 | 12672356 | 112 | 35 | 40573 | 1.9141 | 0.027805 | *ZNF564* |
| ENSG00000213927 | 9 | 34651877 | 34674045 | 61 | 11 | 52091 | 1.9141 | 0.027806 | *CCL27* |
| ENSG00000103126 | 16 | 327440 | 412673 | 370 | 49 | 45442 | 1.914 | 0.027809 | *AXIN1* |
| ENSG00000158315 | 1 | 39341479 | 39417471 | 249 | 38 | 36349 | 1.9139 | 0.027819 | *RHBDL2* |
| ENSG00000131711 | 5 | 71393061 | 71515395 | 289 | 65 | 47679 | 1.9134 | 0.027852 | *MAP1B* |
| ENSG00000197919 | 9 | 21430440 | 21451315 | 78 | 16 | 40861 | 1.9132 | 0.027859 | *IFNA1* |
| ENSG00000142961 | 1 | 47063387 | 47092563 | 61 | 10 | 31753 | 1.913 | 0.027872 | *MOB3C* |
| ENSG00000007341 | 1 | 113056140 | 113173447 | 286 | 29 | 49190 | 1.9128 | 0.027889 | *ST7L* |
| ENSG00000039139 | 5 | 13680440 | 13954652 | 1057 | 80 | 50452 | 1.9124 | 0.027914 | *DNAH5* |
| ENSG00000122756 | 9 | 34541430 | 34600121 | 182 | 33 | 51086 | 1.9123 | 0.027916 | *CNTFR* |

| | | | | | | | | | |
|---|---|---|---|---|---|---|---|---|---|
| ENSG00000143543 | 1 | 153936745 | 153960164 | 34 | 7 | 47892 | 1.912 | 0.027938 | *JTB* |
| ENSG00000143375 | 1 | 151472986 | 151521168 | 54 | 21 | 44043 | 1.912 | 0.027939 | *CGN* |
| ENSG00000160867 | 5 | 176503887 | 176535145 | 127 | 23 | 42679 | 1.911 | 0.028004 | *FGFR4* |
| ENSG00000101425 | 20 | 36878551 | 36975907 | 306 | 65 | 44920 | 1.9108 | 0.028017 | *BPI* |
| ENSG00000089916 | 14 | 76608259 | 76730685 | 449 | 40 | 43118 | 1.9107 | 0.028022 | *GPATCH2L* |
| ENSG00000269094 | 17 | 36874086 | 36894451 | 33 | 13 | 42957 | 1.9106 | 0.028025 | *AC006449.1* |
| ENSG00000056291 | 4 | 72887521 | 73023784 | 190 | 21 | 48068 | 1.9105 | 0.028034 | *NPFFR2* |
| ENSG00000109618 | 4 | 25111627 | 25172204 | 149 | 28 | 44822 | 1.9102 | 0.028054 | *SEPSECS* |
| ENSG00000126261 | 19 | 34909257 | 34970853 | 149 | 21 | 35101 | 1.9101 | 0.028063 | *UBA2* |
| ENSG00000174437 | 12 | 110708561 | 110798898 | 226 | 20 | 51609 | 1.9096 | 0.028094 | *ATP2A2* |
| ENSG00000196098 | 3 | 98062698 | 98083663 | 46 | 10 | 40024 | 1.9089 | 0.02814 | *OR5K4* |
| ENSG00000168101 | 16 | 4733695 | 4755860 | 77 | 11 | 53168 | 1.9084 | 0.028168 | *NUDT16L1* |
| ENSG00000113194 | 5 | 175864629 | 175947075 | 205 | 28 | 45038 | 1.9082 | 0.028183 | *FAF2* |
| ENSG00000071054 | 2 | 102303312 | 102521149 | 428 | 42 | 48167 | 1.9076 | 0.028222 | *MAP4K4* |
| ENSG00000183090 | 4 | 144488455 | 144631828 | 349 | 33 | 47556 | 1.9073 | 0.028239 | *FREM3* |
| ENSG00000021645 | 14 | 78698734 | 80340762 | 4494 | 354 | 43039 | 1.9072 | 0.02825 | *NRXN3* |
| ENSG00000116497 | 1 | 33272368 | 33334476 | 105 | 15 | 53940 | 1.9071 | 0.028254 | *S100PBP* |
| ENSG00000137070 | 9 | 34640699 | 34671889 | 72 | 14 | 54377 | 1.907 | 0.028258 | *IL11RA* |
| ENSG00000233232 | 16 | 28457693 | 28491868 | 19 | 7 | 32329 | 1.9067 | 0.028281 | *NPIPB7* |
| ENSG00000205744 | 19 | 6457218 | 6492568 | 111 | 25 | 40018 | 1.9066 | 0.028288 | *DENND1C* |
| ENSG00000198366 | 6 | 26010718 | 26031186 | 53 | 14 | 42450 | 1.9065 | 0.028296 | *HIST1H3A* |
| ENSG00000105767 | 19 | 44116520 | 44153991 | 85 | 18 | 44154 | 1.9064 | 0.028298 | *CADM4* |
| ENSG00000263203 | 16 | 69142757 | 69164331 | 46 | 16 | 55672 | 1.9062 | 0.028314 | *CHTF8* |
| ENSG00000100379 | 22 | 37437779 | 37469430 | 68 | 20 | 47532 | 1.906 | 0.028328 | *KCTD17* |
| ENSG00000153896 | 19 | 35238981 | 35274134 | 84 | 18 | 56798 | 1.9054 | 0.028366 | *ZNF599* |
| ENSG00000165304 | 9 | 36562859 | 36687678 | 212 | 21 | 43505 | 1.9051 | 0.028383 | *MELK* |
| ENSG00000189339 | 1 | 1582939 | 1634167 | 75 | 16 | 34234 | 1.905 | 0.02839 | *SLC35E2B* |
| ENSG00000176927 | 17 | 28246218 | 28445470 | 306 | 22 | 34469 | 1.9046 | 0.028413 | *EFCAB5* |
| ENSG00000233493 | 19 | 55880613 | 55905966 | 105 | 25 | 42410 | 1.9044 | 0.028432 | *TMEM238* |
| ENSG00000144224 | 2 | 136489189 | 136552625 | 93 | 13 | 19612 | 1.9038 | 0.028465 | *UBXN4* |
| ENSG00000167751 | 19 | 51354824 | 51393823 | 216 | 38 | 45498 | 1.9028 | 0.028532 | *KLK2* |
| ENSG00000145103 | 3 | 121696170 | 121751051 | 120 | 23 | 49863 | 1.9021 | 0.028576 | *ILDR1* |
| ENSG00000135324 | 6 | 84733475 | 84810600 | 205 | 26 | 39729 | 1.902 | 0.028585 | *MRAP2* |
| ENSG00000132530 | 17 | 6648766 | 6688966 | 158 | 27 | 38471 | 1.9003 | 0.028696 | *XAF1* |
| ENSG00000113758 | 5 | 176873609 | 176911402 | 77 | 15 | 32294 | 1.8996 | 0.02874 | *DBN1* |
| ENSG00000243317 | 7 | 135337244 | 135388166 | 206 | 29 | 48275 | 1.8992 | 0.028766 | *C7orf73* |

| | | | | | | | | | |
|---|---|---|---|---|---|---|---|---|---|
| ENSG00000182667 | 11 | 131230373 | 132216716 | 3333 | 311 | 47038 | 1.8989 | 0.028787 | *NTM* |
| ENSG00000155640 | 10 | 98731041 | 98755582 | 23 | 7 | 43344 | 1.8985 | 0.028818 | *C10orf12* |
| ENSG00000166246 | 16 | 4774273 | 4809397 | 140 | 12 | 49749 | 1.898 | 0.02885 | *C16orf71* |
| ENSG00000166803 | 15 | 64647193 | 64689886 | 77 | 16 | 42807 | 1.8979 | 0.028857 | *KIAA0101* |
| ENSG00000183657 | 3 | 171499580 | 171537714 | 143 | 18 | 53447 | 1.8972 | 0.028898 | *PP13439* |
| ENSG00000123610 | 2 | 152204106 | 152246560 | 126 | 19 | 46997 | 1.897 | 0.028914 | *TNFAIP6* |
| ENSG00000072694 | 1 | 161541101 | 161658444 | 164 | 41 | 38707 | 1.8965 | 0.02895 | *FCGR2B* |
| ENSG00000135299 | 6 | 90132889 | 90353553 | 526 | 59 | 46068 | 1.8957 | 0.028998 | *ANKRD6* |
| ENSG00000171346 | 17 | 39659995 | 39688781 | 74 | 11 | 49819 | 1.8951 | 0.029038 | *KRT15* |
| ENSG00000148926 | 11 | 10316227 | 10338944 | 60 | 11 | 48991 | 1.8946 | 0.029075 | *ADM* |
| ENSG00000111348 | 12 | 15084951 | 15124662 | 94 | 19 | 50095 | 1.8941 | 0.029106 | *ARHGDIB* |
| ENSG00000239789 | 7 | 56009512 | 56034192 | 86 | 18 | 54062 | 1.8938 | 0.029125 | *MRPS17* |
| ENSG00000136014 | 12 | 95900336 | 95955266 | 224 | 47 | 47548 | 1.8929 | 0.029184 | *USP44* |
| ENSG00000161956 | 17 | 7455192 | 7485287 | 88 | 21 | 46062 | 1.8929 | 0.029184 | *SENP3* |
| ENSG00000114120 | 3 | 140650672 | 140708775 | 170 | 11 | 45459 | 1.8922 | 0.029232 | *SLC25A36* |
| ENSG00000248098 | 19 | 41874215 | 41940910 | 183 | 11 | 51643 | 1.8916 | 0.029275 | *BCKDHA* |
| ENSG00000179958 | 16 | 30424940 | 30451396 | 29 | 15 | 39066 | 1.8907 | 0.029332 | *DCTPP1* |
| ENSG00000266728 | 17 | 25940390 | 25977803 | 103 | 11 | 52865 | 1.8904 | 0.029349 | *AC015688.3* |
| ENSG00000166546 | 16 | 66451200 | 66537432 | 204 | 37 | 43804 | 1.8904 | 0.02935 | *BEAN1* |
| ENSG00000183206 | 18 | 14497338 | 14553584 | 176 | 22 | 40735 | 1.8895 | 0.029415 | *POTEC* |
| ENSG00000183579 | 22 | 29269580 | 29463475 | 502 | 62 | 40957 | 1.8893 | 0.029423 | *ZNRF3* |
| ENSG00000088543 | 3 | 50585462 | 50618458 | 46 | 10 | 45980 | 1.8889 | 0.02945 | *C3orf18* |
| ENSG00000164741 | 8 | 12930870 | 13383167 | 2598 | 173 | 47012 | 1.8889 | 0.029456 | *DLC1* |
| ENSG00000154957 | 17 | 11870756 | 11910827 | 92 | 14 | 53911 | 1.8888 | 0.029461 | *ZNF18* |
| ENSG00000197557 | 2 | 178467720 | 178493694 | 142 | 18 | 36569 | 1.8881 | 0.029509 | *TTC30A* |
| ENSG00000144061 | 2 | 110869888 | 110972643 | 196 | 17 | 23247 | 1.8881 | 0.02951 | *NPHP1* |
| ENSG00000169288 | 4 | 78773674 | 78883944 | 300 | 25 | 37308 | 1.888 | 0.029514 | *MRPL1* |
| ENSG00000071462 | 7 | 73087355 | 73129491 | 122 | 14 | 53052 | 1.8877 | 0.029531 | *WBSCR22* |
| ENSG00000060709 | 12 | 130870682 | 131210826 | 1528 | 217 | 42461 | 1.8876 | 0.029542 | *RIMBP2* |
| ENSG00000163586 | 2 | 88412510 | 88437635 | 79 | 20 | 37923 | 1.8875 | 0.029547 | *FABP1* |
| ENSG00000163491 | 3 | 27141576 | 27420951 | 957 | 53 | 43090 | 1.8873 | 0.029561 | *NEK10* |
| ENSG00000163681 | 3 | 57731177 | 57924895 | 328 | 32 | 51904 | 1.8866 | 0.029606 | *SLMAP* |
| ENSG00000138750 | 4 | 77025812 | 77079668 | 438 | 14 | 44029 | 1.8865 | 0.029612 | *NUP54* |
| ENSG00000184408 | 7 | 119903722 | 120400385 | 972 | 94 | 40199 | 1.8864 | 0.029619 | *KCND2* |
| ENSG00000155903 | 3 | 141195889 | 141344184 | 267 | 31 | 43006 | 1.886 | 0.029645 | *RASA2* |
| ENSG00000172893 | 11 | 71129239 | 71173914 | 89 | 12 | 25907 | 1.8858 | 0.02966 | *DHCR7* |

| | | | | | | | | | |
|---|---|---|---|---|---|---|---|---|---|
| ENSG00000076641 | 8 | 81870045 | 82034303 | 414 | 73 | 34335 | 1.8854 | 0.029686 | *PAG1* |
| ENSG00000166529 | 7 | 99637390 | 99672661 | 61 | 10 | 51998 | 1.8854 | 0.029687 | *ZSCAN21* |
| ENSG00000183873 | 3 | 38579548 | 38701164 | 338 | 78 | 50527 | 1.8853 | 0.029692 | *SCN5A* |
| ENSG00000128482 | 17 | 19304438 | 19330589 | 16 | 7 | 48867 | 1.8853 | 0.029695 | *RNF112* |
| ENSG00000124813 | 6 | 45285894 | 45642086 | 1102 | 115 | 45602 | 1.885 | 0.029713 | *RUNX2* |
| ENSG00000155366 | 1 | 113233728 | 113260056 | 71 | 16 | 53289 | 1.8848 | 0.029731 | *RHOC* |
| ENSG00000177200 | 16 | 53078945 | 53373062 | 808 | 41 | 50706 | 1.8842 | 0.029766 | *CHD9* |
| ENSG00000088992 | 12 | 117466728 | 117547284 | 293 | 38 | 41965 | 1.8842 | 0.02977 | *TESC* |
| ENSG00000270672 | 7 | 142364104 | 142385550 | 90 | 8 | 38698 | 1.8841 | 0.029776 | *MTRNR2L6* |
| ENSG00000203804 | 1 | 150523480 | 150543969 | 47 | 16 | 47276 | 1.8837 | 0.0298 | *ADAMTSL4-AS1* |
| ENSG00000253207 | 8 | 114602539 | 114639187 | 107 | 16 | 36297 | 1.8837 | 0.029801 | *RP11-67H2.1* |
| ENSG00000214517 | 11 | 73872144 | 73975748 | 165 | 33 | 40481 | 1.883 | 0.029853 | *PPME1* |
| ENSG00000187116 | 19 | 54808353 | 54834409 | 78 | 12 | 42548 | 1.8824 | 0.02989 | *LILRA5* |
| ENSG00000136930 | 9 | 127105745 | 127187723 | 125 | 21 | 46497 | 1.8803 | 0.030032 | *PSMB7* |
| ENSG00000142208 | 14 | 105225686 | 105272088 | 107 | 27 | 41968 | 1.8797 | 0.030071 | *AKT1* |
| ENSG00000204264 | 6 | 32798494 | 32822480 | 196 | 33 | 50356 | 1.8794 | 0.030096 | *PSMB8* |
| ENSG00000187186 | 9 | 34654160 | 34676109 | 71 | 11 | 52855 | 1.8794 | 0.030096 | *RP11-195F19.5* |
| ENSG00000053438 | 20 | 36139617 | 36162092 | 37 | 9 | 47635 | 1.8794 | 0.030096 | *NNAT* |
| ENSG00000048162 | 5 | 175800949 | 175825976 | 64 | 16 | 53545 | 1.879 | 0.030123 | *NOP16* |
| ENSG00000212719 | 17 | 21418051 | 21487722 | 49 | 11 | 45959 | 1.8787 | 0.030145 | *C17orf51* |
| ENSG00000214921 | 17 | 42315758 | 42337236 | 47 | 13 | 39979 | 1.8782 | 0.030174 | *AC003102.1* |
| ENSG00000214026 | 11 | 1958508 | 2015752 | 147 | 30 | 42771 | 1.8778 | 0.030202 | *MRPL23* |
| ENSG00000168096 | 16 | 4736513 | 4794379 | 219 | 25 | 49860 | 1.8778 | 0.030205 | *ANKS3* |
| ENSG00000183258 | 5 | 176928578 | 176954470 | 43 | 14 | 36917 | 1.8766 | 0.03029 | *DDX41* |
| ENSG00000103042 | 16 | 58689013 | 58729008 | 169 | 26 | 46252 | 1.8765 | 0.030292 | *SLC38A7* |
| ENSG00000155254 | 10 | 99462930 | 99487907 | 54 | 15 | 38521 | 1.8761 | 0.03032 | *MARVELD1* |
| ENSG00000115825 | 2 | 37467645 | 37561951 | 402 | 37 | 49078 | 1.8757 | 0.03035 | *PRKD3* |
| ENSG00000185189 | 8 | 144905764 | 144934200 | 53 | 15 | 40300 | 1.8753 | 0.030375 | *NRBP2* |
| ENSG00000205268 | 8 | 66619745 | 66764557 | 429 | 37 | 47644 | 1.8751 | 0.030389 | *PDE7A* |
| ENSG00000167904 | 8 | 56598983 | 56695966 | 293 | 30 | 49811 | 1.8747 | 0.030415 | *TMEM68* |
| ENSG00000184719 | 10 | 90023621 | 90354287 | 846 | 83 | 45239 | 1.8734 | 0.030506 | *RNLS* |
| ENSG00000188315 | 3 | 49296035 | 49325342 | 44 | 13 | 44797 | 1.8733 | 0.030511 | *C3orf62* |
| ENSG00000149480 | 11 | 62350686 | 62379312 | 40 | 6 | 45930 | 1.8728 | 0.030549 | *MTA2* |
| ENSG00000106546 | 7 | 17328246 | 17395776 | 95 | 26 | 51399 | 1.8727 | 0.030557 | *AHR* |
| ENSG00000074410 | 15 | 63603577 | 63684360 | 232 | 44 | 41492 | 1.8725 | 0.030566 | *CA12* |
| ENSG00000170959 | 11 | 30841916 | 31401357 | 927 | 67 | 45234 | 1.8724 | 0.030576 | *DCDC1* |

| | | | | | | | | | |
|---|---|---|---|---|---|---|---|---|---|
| ENSG00000134940 | 11 | 125531417 | 125561018 | 80 | 13 | 53492 | 1.872 | 0.030605 | *ACRV1* |
| ENSG00000076003 | 2 | 136587196 | 136643996 | 63 | 10 | 22180 | 1.8711 | 0.030664 | *MCM6* |
| ENSG00000112232 | 6 | 62379865 | 63006132 | 1847 | 89 | 46546 | 1.8707 | 0.030694 | *KHDRBS2* |
| ENSG00000135775 | 1 | 230768235 | 230839728 | 272 | 48 | 43849 | 1.8706 | 0.030701 | *COG2* |
| ENSG00000204010 | 10 | 91127813 | 91154962 | 69 | 16 | 35981 | 1.8704 | 0.030715 | *IFIT1B* |
| ENSG00000196663 | 14 | 102819300 | 102978818 | 532 | 35 | 50579 | 1.8703 | 0.030721 | *TECPR2* |
| ENSG00000196296 | 16 | 28879726 | 28925830 | 67 | 10 | 35217 | 1.87 | 0.030742 | *ATP2A1* |
| ENSG00000168269 | 5 | 169522901 | 169546727 | 128 | 19 | 43568 | 1.8692 | 0.030795 | *FOXI1* |
| ENSG00000141556 | 17 | 80699940 | 80910724 | 881 | 65 | 45982 | 1.8691 | 0.030806 | *TBCD* |
| ENSG00000151617 | 4 | 148392069 | 148476106 | 188 | 22 | 32600 | 1.8689 | 0.030818 | *EDNRA* |
| ENSG00000137574 | 8 | 56675701 | 56748007 | 245 | 16 | 53754 | 1.8687 | 0.030835 | *TGS1* |
| ENSG00000148842 | 10 | 104668050 | 104859978 | 550 | 36 | 49578 | 1.8654 | 0.031062 | *CNNM2* |
| ENSG00000168961 | 17 | 25946824 | 25986586 | 129 | 12 | 53435 | 1.8649 | 0.031101 | *LGALS9* |
| ENSG00000089091 | 20 | 18354011 | 18457925 | 364 | 45 | 44224 | 1.8643 | 0.031137 | *DZANK1* |
| ENSG00000145194 | 3 | 183957438 | 184020819 | 149 | 26 | 51042 | 1.8639 | 0.031171 | *ECE2* |
| ENSG00000185499 | 1 | 155148300 | 155172707 | 36 | 10 | 46532 | 1.8637 | 0.031179 | *MUC1* |
| ENSG00000135638 | 2 | 73133389 | 73172020 | 74 | 18 | 41836 | 1.8636 | 0.031186 | *EMX1* |
| ENSG00000172508 | 11 | 67172439 | 67203078 | 52 | 14 | 44107 | 1.8631 | 0.031226 | *CARNS1* |
| ENSG00000129270 | 17 | 34073268 | 34132711 | 167 | 19 | 48608 | 1.8623 | 0.031278 | *MMP28* |
| ENSG00000128928 | 15 | 40687686 | 40738146 | 136 | 15 | 39379 | 1.8614 | 0.031341 | *IVD* |
| ENSG00000116176 | 16 | 1261651 | 1285257 | 139 | 26 | 54766 | 1.8608 | 0.031387 | *TPSG1* |
| ENSG00000153767 | 3 | 120451484 | 120511916 | 150 | 23 | 39854 | 1.8598 | 0.03146 | *GTF2E1* |
| ENSG00000111644 | 12 | 6737241 | 6766626 | 54 | 15 | 40652 | 1.8594 | 0.031483 | *ACRBP* |
| ENSG00000013573 | 12 | 31216779 | 31267725 | 270 | 11 | 45056 | 1.8593 | 0.031489 | *DDX11* |
| ENSG00000112029 | 6 | 153281664 | 153314714 | 141 | 16 | 50943 | 1.8582 | 0.031572 | *FBXO5* |
| ENSG00000049618 | 6 | 157089063 | 157541913 | 1133 | 115 | 48051 | 1.8575 | 0.031622 | *ARID1B* |
| ENSG00000187013 | 17 | 59479112 | 59500641 | 63 | 11 | 39783 | 1.857 | 0.031653 | *C17orf82* |
| ENSG00000162869 | 2 | 48657737 | 48752525 | 306 | 38 | 50719 | 1.8566 | 0.031684 | *PPP1R21* |
| ENSG00000257923 | 7 | 101448959 | 101937249 | 1345 | 109 | 45906 | 1.8562 | 0.03171 | *CUX1* |
| ENSG00000125744 | 19 | 45978547 | 46010319 | 117 | 21 | 54074 | 1.856 | 0.031725 | *RTN2* |
| ENSG00000074582 | 2 | 219513487 | 219538166 | 62 | 9 | 35577 | 1.8557 | 0.031745 | *BCS1L* |
| ENSG00000163017 | 2 | 74109441 | 74156992 | 173 | 26 | 40682 | 1.8555 | 0.031762 | *ACTG2* |
| ENSG00000067836 | 16 | 4836969 | 4862951 | 74 | 18 | 51063 | 1.8554 | 0.031773 | *ROGDI* |
| ENSG00000204301 | 6 | 32152620 | 32201844 | 309 | 53 | 42149 | 1.8552 | 0.031781 | *NOTCH4* |
| ENSG00000143032 | 1 | 91167096 | 91192794 | 60 | 20 | 51917 | 1.855 | 0.031801 | *BARHL2* |
| ENSG00000065308 | 6 | 52352200 | 52451713 | 428 | 61 | 44351 | 1.8547 | 0.031822 | *TRAM2* |

| | | | | | | | | | |
|---|---|---|---|---|---|---|---|---|---|
| ENSG00000198089 | 22 | 31874674 | 32024572 | 583 | 42 | 50621 | 1.8542 | 0.031853 | *SFI1* |
| ENSG00000157766 | 15 | 89336674 | 89428585 | 307 | 59 | 41629 | 1.8539 | 0.031876 | *ACAN* |
| ENSG00000139329 | 12 | 91486406 | 91515608 | 107 | 15 | 37039 | 1.8538 | 0.031886 | *LUM* |
| ENSG00000173020 | 11 | 67023881 | 67064027 | 42 | 15 | 44804 | 1.8533 | 0.031922 | *ADRBK1* |
| ENSG00000166619 | 20 | 36110874 | 36166333 | 82 | 14 | 45619 | 1.8531 | 0.031936 | *BLCAP* |
| ENSG00000183196 | 16 | 75500949 | 75539282 | 116 | 26 | 39355 | 1.8528 | 0.031957 | *CHST6* |
| ENSG00000226479 | 2 | 120968854 | 120990984 | 45 | 8 | 49553 | 1.8517 | 0.032038 | *TMEM185B* |
| ENSG00000186834 | 17 | 43214684 | 43239468 | 50 | 13 | 48272 | 1.8516 | 0.032039 | *HEXIM1* |
| ENSG00000149782 | 11 | 64008995 | 64046622 | 72 | 20 | 45418 | 1.8514 | 0.032054 | *PLCB3* |
| ENSG00000105697 | 19 | 35761619 | 35786046 | 73 | 14 | 53494 | 1.8514 | 0.032059 | *HAMP* |
| ENSG00000137842 | 15 | 43405477 | 43487344 | 94 | 25 | 43812 | 1.8512 | 0.032071 | *TMEM62* |
| ENSG00000167969 | 16 | 2279396 | 2312301 | 70 | 17 | 49819 | 1.8501 | 0.032153 | *ECI1* |
| ENSG00000174652 | 19 | 9513272 | 9556254 | 198 | 11 | 51797 | 1.8497 | 0.032176 | *ZNF266* |
| ENSG00000042980 | 8 | 24141553 | 24226531 | 325 | 25 | 43216 | 1.8497 | 0.03218 | *ADAM28* |
| ENSG00000185122 | 8 | 145505280 | 145548385 | 87 | 9 | 48510 | 1.8496 | 0.032187 | *HSF1* |
| ENSG00000153147 | 4 | 144424616 | 144488639 | 131 | 11 | 53735 | 1.8493 | 0.032206 | *SMARCA5* |
| ENSG00000176953 | 16 | 28952128 | 28988418 | 52 | 16 | 39032 | 1.8492 | 0.032217 | *NFATC2IP* |
| ENSG00000059691 | 4 | 152581656 | 152692175 | 288 | 18 | 38914 | 1.8489 | 0.032239 | *PET112* |
| ENSG00000142910 | 1 | 32032116 | 32063288 | 53 | 23 | 43398 | 1.8488 | 0.032247 | *TINAGL1* |
| ENSG00000153721 | 6 | 154698638 | 154841793 | 668 | 118 | 46093 | 1.8486 | 0.032258 | *CNKSR3* |
| ENSG00000137877 | 15 | 42130345 | 42196275 | 229 | 40 | 45170 | 1.8482 | 0.032288 | *SPTBN5* |
| ENSG00000196233 | 10 | 98582017 | 98750800 | 242 | 20 | 39112 | 1.8474 | 0.032344 | *LCOR* |
| ENSG00000144741 | 3 | 66109285 | 66448540 | 732 | 88 | 43364 | 1.8472 | 0.032362 | *SLC25A26* |
| ENSG00000163481 | 2 | 219518587 | 219547134 | 62 | 9 | 32691 | 1.8465 | 0.032413 | *RNF25* |
| ENSG00000186564 | 1 | 47891689 | 47916363 | 57 | 12 | 39041 | 1.8454 | 0.032487 | *FOXD2* |
| ENSG00000112655 | 6 | 43034006 | 43139457 | 283 | 42 | 42557 | 1.845 | 0.032518 | *PTK7* |
| ENSG00000185619 | 4 | 689537 | 774428 | 384 | 54 | 49546 | 1.845 | 0.032522 | *PCGF3* |
| ENSG00000108592 | 17 | 61886793 | 61917372 | 60 | 6 | 26376 | 1.8449 | 0.032523 | *FTSJ3* |
| ENSG00000176623 | 8 | 87470486 | 87536586 | 243 | 20 | 51686 | 1.8448 | 0.032534 | *RMDN1* |
| ENSG00000178033 | 6 | 116822809 | 116855955 | 118 | 12 | 46689 | 1.8443 | 0.032566 | *FAM26E* |
| ENSG00000112578 | 6 | 41878926 | 41910784 | 57 | 17 | 49213 | 1.8441 | 0.032586 | *BYSL* |
| ENSG00000118733 | 1 | 102258130 | 102472586 | 782 | 101 | 48760 | 1.8435 | 0.032626 | *OLFM3* |
| ENSG00000091831 | 6 | 151967826 | 152460754 | 1707 | 164 | 39229 | 1.8428 | 0.032676 | *ESR1* |
| ENSG00000124587 | 6 | 42921608 | 42956958 | 155 | 20 | 43788 | 1.8426 | 0.032695 | *PEX6* |
| ENSG00000140623 | 16 | 4817670 | 4848522 | 116 | 17 | 50024 | 1.8421 | 0.032728 | *Sep-12* |
| ENSG00000136631 | 1 | 150029369 | 150127505 | 145 | 15 | 45312 | 1.8416 | 0.032769 | *VPS45* |

| | | | | | | | | | |
|---|---|---|---|---|---|---|---|---|---|
| ENSG00000147955 | 9 | 34624719 | 34647806 | 51 | 13 | 52771 | 1.8415 | 0.032775 | *SIGMAR1* |
| ENSG00000213930 | 9 | 34628130 | 34661032 | 69 | 15 | 54881 | 1.8413 | 0.032789 | *GALT* |
| ENSG00000150054 | 10 | 28329922 | 28633415 | 1286 | 86 | 39222 | 1.8411 | 0.032805 | *MPP7* |
| ENSG00000065060 | 6 | 34749857 | 34860915 | 359 | 24 | 33144 | 1.8406 | 0.032841 | *UHRF1BP1* |
| ENSG00000147432 | 8 | 42542519 | 42602550 | 281 | 13 | 32341 | 1.8402 | 0.032869 | *CHRNB3* |
| ENSG00000136807 | 9 | 130537958 | 130563066 | 64 | 10 | 34640 | 1.8401 | 0.032879 | *CDK9* |
| ENSG00000213949 | 5 | 52073730 | 52265040 | 722 | 102 | 50020 | 1.8398 | 0.0329 | *ITGA1* |
| ENSG00000175416 | 5 | 175809456 | 175853570 | 117 | 25 | 50758 | 1.8391 | 0.032948 | *CLTB* |
| ENSG00000150667 | 15 | 39882232 | 40085031 | 706 | 55 | 37143 | 1.8391 | 0.032949 | *FSIP1* |
| ENSG00000112218 | 6 | 97232002 | 97295353 | 236 | 24 | 50546 | 1.839 | 0.032956 | *GPR63* |
| ENSG00000187134 | 10 | 4924796 | 5035475 | 559 | 65 | 48073 | 1.8382 | 0.033017 | *AKR1C1* |
| ENSG00000124233 | 20 | 43825638 | 43848413 | 60 | 9 | 50681 | 1.8378 | 0.033044 | *SEMG1* |
| ENSG00000198055 | 5 | 176820205 | 176879902 | 102 | 25 | 37360 | 1.8369 | 0.033115 | *GRK6* |
| ENSG00000196834 | 2 | 131207028 | 131277239 | 17 | 10 | 32364 | 1.8357 | 0.0332 | *POTEI* |
| ENSG00000204140 | 6 | 35738794 | 35771452 | 105 | 22 | 31818 | 1.8355 | 0.033219 | *CLPSL1* |
| ENSG00000170788 | 10 | 82085861 | 82126511 | 134 | 16 | 36410 | 1.8353 | 0.033231 | *DYDC1* |
| ENSG00000189233 | 8 | 27869481 | 27951388 | 236 | 53 | 47145 | 1.8347 | 0.033273 | *NUGGC* |
| ENSG00000124812 | 6 | 49791970 | 49854809 | 184 | 14 | 51007 | 1.8345 | 0.033291 | *CRISP1* |
| ENSG00000058673 | 1 | 203754782 | 203833252 | 263 | 22 | 48427 | 1.8336 | 0.033359 | *ZC3H11A* |
| ENSG00000161180 | 22 | 21977005 | 22001616 | 62 | 17 | 37061 | 1.8329 | 0.033406 | *CCDC116* |
| ENSG00000057657 | 6 | 106524195 | 106567814 | 87 | 27 | 51218 | 1.8328 | 0.033417 | *PRDM1* |
| ENSG00000116031 | 2 | 71047347 | 71072952 | 194 | 16 | 47600 | 1.8323 | 0.033456 | *CD207* |
| ENSG00000184571 | 22 | 25105001 | 25180687 | 349 | 41 | 52642 | 1.8319 | 0.033485 | *PIWIL3* |
| ENSG00000167822 | 11 | 55894247 | 55915194 | 97 | 10 | 55467 | 1.8305 | 0.033586 | *OR8J3* |
| ENSG00000152778 | 10 | 91164343 | 91190758 | 95 | 18 | 28741 | 1.8305 | 0.03359 | *IFIT5* |
| ENSG00000135835 | 1 | 180872290 | 180930750 | 171 | 47 | 47818 | 1.8296 | 0.033657 | *KIAA1614* |
| ENSG00000124466 | 19 | 43954939 | 43979812 | 69 | 18 | 45372 | 1.8286 | 0.033733 | *LYPD3* |
| ENSG00000173846 | 1 | 45255897 | 45281662 | 63 | 10 | 49726 | 1.8283 | 0.033749 | *PLK3* |
| ENSG00000196335 | 7 | 23739786 | 23882132 | 520 | 43 | 49414 | 1.8265 | 0.033889 | *STK31* |
| ENSG00000141376 | 17 | 58744814 | 59480199 | 969 | 92 | 37556 | 1.8259 | 0.033933 | *BCAS3* |
| ENSG00000269755 | 19 | 12533917 | 12605630 | 367 | 17 | 47571 | 1.8256 | 0.033952 | *CTD-3105H18.18* |
| ENSG00000169291 | 1 | 154432248 | 154484589 | 118 | 17 | 42421 | 1.8252 | 0.033983 | *SHE* |
| ENSG00000065320 | 17 | 8914859 | 9157317 | 986 | 144 | 46961 | 1.8251 | 0.033993 | *NTN1* |
| ENSG00000175318 | 15 | 72442148 | 72500126 | 93 | 18 | 51233 | 1.8239 | 0.034082 | *GRAMD2* |
| ENSG00000138767 | 4 | 78624541 | 78750769 | 249 | 22 | 35899 | 1.8232 | 0.034136 | *CNOT6L* |
| ENSG00000176601 | 2 | 135712061 | 135815038 | 224 | 22 | 24260 | 1.823 | 0.034152 | *MAP3K19* |

| | | | | | | | | | |
|---|---|---|---|---|---|---|---|---|---|
| ENSG00000132581 | 17 | 26965374 | 26999207 | 32 | 14 | 49325 | 1.8228 | 0.034166 | *SDF2* |
| ENSG00000105717 | 19 | 19662522 | 19739725 | 140 | 21 | 47379 | 1.8221 | 0.034221 | *PBX4* |
| ENSG00000184708 | 22 | 31822963 | 31902094 | 235 | 30 | 48080 | 1.8216 | 0.034257 | *EIF4ENIF1* |
| ENSG00000120329 | 5 | 140672196 | 140693612 | 88 | 16 | 45856 | 1.8207 | 0.034327 | *SLC25A2* |
| ENSG00000221988 | 6 | 32111218 | 32144011 | 87 | 23 | 46945 | 1.8205 | 0.034339 | *PPT2* |
| ENSG00000067369 | 15 | 43689407 | 43812926 | 184 | 19 | 38582 | 1.8202 | 0.034361 | *TP53BP1* |
| ENSG00000169442 | 1 | 26634448 | 26657014 | 72 | 12 | 40846 | 1.8199 | 0.03439 | *CD52* |
| ENSG00000132361 | 17 | 2582680 | 2625957 | 79 | 21 | 43110 | 1.8181 | 0.034528 | *CLUH* |
| ENSG00000166947 | 15 | 43388423 | 43523481 | 148 | 30 | 42206 | 1.8174 | 0.03458 | *EPB42* |
| ENSG00000123636 | 2 | 160165490 | 160483203 | 1023 | 87 | 37824 | 1.8165 | 0.03465 | *BAZ2B* |
| ENSG00000256235 | 5 | 150147508 | 150186296 | 117 | 13 | 22958 | 1.8157 | 0.034708 | *SMIM3* |
| ENSG00000131233 | 1 | 39320175 | 39357289 | 108 | 15 | 32624 | 1.8156 | 0.034716 | *GJA9* |
| ENSG00000135930 | 2 | 233404762 | 233458354 | 151 | 15 | 52258 | 1.8153 | 0.034741 | *EIF4E2* |
| ENSG00000175619 | 11 | 48228344 | 48249314 | 87 | 14 | 32258 | 1.8148 | 0.03478 | *OR4B1* |
| ENSG00000092067 | 14 | 23576513 | 23598825 | 68 | 22 | 44899 | 1.8147 | 0.034785 | *CEBPE* |
| ENSG00000123179 | 13 | 50224859 | 50275611 | 203 | 31 | 45482 | 1.8142 | 0.034824 | *EBPL* |
| ENSG00000269636 | 5 | 150147867 | 150186297 | 116 | 13 | 23044 | 1.8132 | 0.034904 | *AC010441.1* |
| ENSG00000162511 | 1 | 31195316 | 31240667 | 104 | 34 | 45665 | 1.8126 | 0.034943 | *LAPTM5* |
| ENSG00000158517 | 7 | 74178309 | 74213659 | 32 | 13 | 39809 | 1.8123 | 0.034974 | *NCF1* |
| ENSG00000170832 | 17 | 58246455 | 58509831 | 244 | 33 | 40009 | 1.812 | 0.034991 | *USP32* |
| ENSG00000188467 | 15 | 48403169 | 48444869 | 7 | 4 | 48825 | 1.8104 | 0.035118 | *SLC24A5* |
| ENSG00000256870 | 12 | 101539271 | 101614185 | 284 | 35 | 50578 | 1.8097 | 0.035169 | *SLC5A8* |
| ENSG00000112651 | 6 | 43011767 | 43037544 | 69 | 11 | 36537 | 1.8093 | 0.035201 | *MRPL2* |
| ENSG00000130751 | 19 | 47513077 | 47559033 | 115 | 34 | 43441 | 1.8087 | 0.035247 | *NPAS1* |
| ENSG00000228075 | 18 | 54804293 | 54827531 | 95 | 20 | 46479 | 1.8082 | 0.035291 | *BOD1L2* |
| ENSG00000166359 | 19 | 33612996 | 33676701 | 221 | 36 | 30754 | 1.8079 | 0.035308 | *WDR88* |
| ENSG00000172208 | 11 | 48256608 | 48277622 | 88 | 11 | 31731 | 1.8078 | 0.035316 | *OR4X2* |
| ENSG00000105700 | 19 | 18658572 | 18690197 | 96 | 12 | 33559 | 1.8076 | 0.035335 | *KXD1* |
| ENSG00000185372 | 5 | 180541357 | 180562304 | 52 | 12 | 28924 | 1.8075 | 0.035341 | *OR2V1* |
| ENSG00000171345 | 17 | 39669869 | 39694560 | 64 | 6 | 51475 | 1.8075 | 0.035345 | *KRT19* |
| ENSG00000188095 | 15 | 90293822 | 90331982 | 203 | 15 | 52092 | 1.8073 | 0.035359 | *MESP2* |
| ENSG00000170786 | 8 | 57202569 | 57243335 | 115 | 16 | 54442 | 1.807 | 0.035378 | *SDR16C5* |
| ENSG00000122543 | 7 | 5909458 | 5935993 | 159 | 35 | 45497 | 1.8057 | 0.035482 | *OCM* |
| ENSG00000233050 | 8 | 11911898 | 11939256 | 1 | 1 | 42786 | 1.8048 | 0.03555 | *DEFB130* |
| ENSG00000166997 | 7 | 99707236 | 99733134 | 44 | 12 | 48821 | 1.8045 | 0.03558 | *CNPY4* |
| ENSG00000169856 | 15 | 53039637 | 53093273 | 138 | 31 | 37561 | 1.8044 | 0.035586 | *ONECUT1* |

| | | | | | | | | | |
|---|---|---|---|---|---|---|---|---|---|
| ENSG00000079432 | 19 | 42762689 | 42809949 | 63 | 7 | 52018 | 1.804 | 0.035615 | *CIC* |
| ENSG00000133665 | 10 | 82094501 | 82137829 | 183 | 15 | 33121 | 1.8038 | 0.035629 | *DYDC2* |
| ENSG00000254772 | 11 | 62317073 | 62352401 | 59 | 10 | 42120 | 1.8038 | 0.035629 | *EEF1G* |
| ENSG00000257242 | 12 | 92368756 | 92546690 | 596 | 116 | 47514 | 1.8029 | 0.035706 | *C12orf79* |
| ENSG00000111817 | 6 | 116565336 | 116772424 | 553 | 55 | 38659 | 1.8023 | 0.035749 | *DSE* |
| ENSG00000212864 | 9 | 140104707 | 140126033 | 53 | 13 | 55410 | 1.8022 | 0.035757 | *RNF208* |
| ENSG00000106366 | 7 | 100760370 | 100792547 | 113 | 25 | 47466 | 1.8019 | 0.035781 | *SERPINE1* |
| ENSG00000186117 | 11 | 55568854 | 55589952 | 83 | 13 | 52949 | 1.8018 | 0.035786 | *OR5L1* |
| ENSG00000124444 | 19 | 44090544 | 44115309 | 42 | 13 | 53540 | 1.8015 | 0.035812 | *ZNF576* |
| ENSG00000116783 | 1 | 74653947 | 75020112 | 728 | 102 | 40476 | 1.8005 | 0.035887 | *TNNI3K* |
| ENSG00000138074 | 2 | 27412455 | 27445826 | 96 | 14 | 42124 | 1.7999 | 0.035935 | *SLC5A6* |
| ENSG00000272305 | 3 | 52993135 | 53143469 | 412 | 38 | 47302 | 1.7998 | 0.035948 | *RP11-894J14.5* |
| ENSG00000105507 | 19 | 48523210 | 48557310 | 199 | 21 | 50692 | 1.7996 | 0.035964 | *CABP5* |
| ENSG00000124157 | 20 | 43839941 | 43863099 | 51 | 8 | 48559 | 1.7995 | 0.035966 | *SEMG2* |
| ENSG00000146094 | 5 | 176918908 | 176948275 | 47 | 17 | 38640 | 1.7992 | 0.035997 | *DOK3* |
| ENSG00000213889 | 19 | 45982035 | 46015768 | 105 | 20 | 54738 | 1.7989 | 0.036017 | *PPM1N* |
| ENSG00000182362 | 21 | 47696251 | 47727665 | 185 | 17 | 43310 | 1.7982 | 0.036073 | *YBEY* |
| ENSG00000134369 | 1 | 201582411 | 201806102 | 555 | 98 | 45403 | 1.7981 | 0.036078 | *NAV1* |
| ENSG00000100784 | 14 | 91326799 | 91536980 | 533 | 54 | 51311 | 1.7979 | 0.036094 | *RPS6KA5* |
| ENSG00000142303 | 19 | 8635126 | 8685620 | 126 | 38 | 43295 | 1.7978 | 0.036106 | *ADAMTS10* |
| ENSG00000172478 | 2 | 241815465 | 241846306 | 227 | 17 | 53371 | 1.7975 | 0.036132 | *C2orf54* |
| ENSG00000100749 | 14 | 97253641 | 97408059 | 434 | 46 | 48730 | 1.7969 | 0.036173 | *VRK1* |
| ENSG00000241399 | 2 | 160615364 | 160664753 | 195 | 24 | 45491 | 1.7968 | 0.036186 | *CD302* |
| ENSG00000135587 | 6 | 109751966 | 109775122 | 62 | 16 | 41582 | 1.7966 | 0.036197 | *SMPD2* |
| ENSG00000037280 | 5 | 180018506 | 180086624 | 360 | 61 | 48497 | 1.7951 | 0.036317 | *FLT4* |
| ENSG00000196189 | 1 | 156107157 | 156157543 | 114 | 24 | 39261 | 1.7947 | 0.036351 | *SEMA4A* |
| ENSG00000157036 | 3 | 38527618 | 38593652 | 148 | 20 | 40572 | 1.7946 | 0.036356 | *EXOG* |
| ENSG00000072121 | 14 | 68184091 | 68293307 | 288 | 36 | 41302 | 1.7945 | 0.036363 | *ZFYVE26* |
| ENSG00000168124 | 16 | 3244247 | 3265188 | 83 | 13 | 46323 | 1.7943 | 0.036383 | *OR1F1* |
| ENSG00000244687 | 20 | 48687661 | 48742496 | 124 | 16 | 45738 | 1.7943 | 0.036384 | *UBE2V1* |
| ENSG00000137033 | 9 | 6205805 | 6267983 | 215 | 33 | 37310 | 1.7942 | 0.036389 | *IL33* |
| ENSG00000166192 | 15 | 72396599 | 72443311 | 86 | 13 | 50666 | 1.7938 | 0.036422 | *SENP8* |
| ENSG00000184995 | 9 | 21470841 | 21492312 | 64 | 13 | 45226 | 1.7935 | 0.036446 | *IFNE* |
| ENSG00000145626 | 5 | 35941112 | 36011130 | 186 | 29 | 42661 | 1.7931 | 0.03648 | *UGT3A1* |
| ENSG00000010295 | 12 | 6637541 | 6675239 | 95 | 18 | 42340 | 1.7925 | 0.03653 | *IFFO1* |
| ENSG00000124713 | 6 | 42918496 | 42941618 | 104 | 11 | 47163 | 1.792 | 0.036564 | *GNMT* |

| | | | | | | | | | |
|---|---|---|---|---|---|---|---|---|---|
| ENSG00000181499 | 11 | 123803492 | 123824580 | 93 | 16 | 54180 | 1.792 | 0.036569 | *OR6T1* |
| ENSG00000115159 | 2 | 157281802 | 157480247 | 467 | 46 | 46369 | 1.7915 | 0.03661 | *GPD2* |
| ENSG00000268857 | 2 | 157342597 | 157362655 | 43 | 13 | 49896 | 1.7912 | 0.036628 | *AC011308.1* |
| ENSG00000109758 | 4 | 3433614 | 3461211 | 88 | 26 | 45773 | 1.7901 | 0.036718 | *HGFAC* |
| ENSG00000173156 | 11 | 66814289 | 66849484 | 64 | 19 | 43233 | 1.7897 | 0.036754 | *RHOD* |
| ENSG00000101407 | 20 | 36601409 | 36671870 | 121 | 19 | 38410 | 1.7891 | 0.0368 | *TTI1* |
| ENSG00000122970 | 12 | 110552140 | 110666602 | 265 | 24 | 48086 | 1.7888 | 0.036823 | *IFT81* |
| ENSG00000259399 | 20 | 35192956 | 35250787 | 67 | 26 | 43710 | 1.7887 | 0.036832 | *TGIF2-C20orf24* |
| ENSG00000121853 | 3 | 172152923 | 172176246 | 89 | 16 | 39459 | 1.7883 | 0.036862 | *GHSR* |
| ENSG00000215262 | 8 | 36631842 | 36803646 | 341 | 33 | 41925 | 1.7882 | 0.036872 | *KCNU1* |
| ENSG00000000460 | 1 | 169621245 | 169833221 | 1103 | 56 | 51870 | 1.787 | 0.036966 | *C1orf112* |
| ENSG00000255641 | 12 | 10554911 | 10598600 | 66 | 13 | 43327 | 1.7864 | 0.037015 | *NKG2-E* |
| ENSG00000168477 | 6 | 31998931 | 32093111 | 274 | 54 | 35428 | 1.7863 | 0.037021 | *TNXB* |
| ENSG00000171951 | 2 | 224451658 | 224477221 | 44 | 13 | 36548 | 1.7863 | 0.037029 | *SCG2* |
| ENSG00000175294 | 11 | 65774223 | 65803988 | 96 | 19 | 41405 | 1.786 | 0.037046 | *CATSPER1* |
| ENSG00000120370 | 1 | 170491270 | 170532587 | 64 | 16 | 37644 | 1.7857 | 0.037075 | *GORAB* |
| ENSG00000180475 | 11 | 57985354 | 58006390 | 94 | 14 | 41166 | 1.7856 | 0.037081 | *OR10Q1* |
| ENSG00000115392 | 2 | 58376378 | 58478507 | 266 | 38 | 45016 | 1.7851 | 0.037119 | *FANCL* |
| ENSG00000234284 | 5 | 178440753 | 178472065 | 123 | 9 | 45992 | 1.7851 | 0.037121 | *ZNF879* |
| ENSG00000215183 | 9 | 35742987 | 35766610 | 33 | 12 | 52620 | 1.784 | 0.037209 | *MSMP* |
| ENSG00000272636 | 17 | 0 | 41427 | 128 | 16 | 49399 | 1.784 | 0.03721 | *DOC2B* |
| ENSG00000186451 | 3 | 57084469 | 57119460 | 118 | 21 | 47107 | 1.784 | 0.037211 | *SPATA12* |
| ENSG00000177728 | 17 | 73427240 | 73506171 | 260 | 28 | 35403 | 1.784 | 0.037213 | *KIAA0195* |
| ENSG00000213585 | 5 | 133297606 | 133350824 | 138 | 20 | 55232 | 1.7834 | 0.037257 | *VDAC1* |
| ENSG00000177932 | 5 | 178477416 | 178520538 | 146 | 17 | 46756 | 1.7832 | 0.037279 | *ZNF354C* |
| ENSG00000197272 | 16 | 28500683 | 28533372 | 77 | 13 | 47950 | 1.7832 | 0.03728 | *IL27* |
| ENSG00000061918 | 4 | 156670144 | 156738743 | 242 | 48 | 49323 | 1.783 | 0.037294 | *GUCY1B3* |
| ENSG00000163217 | 2 | 69082613 | 69108649 | 56 | 10 | 54559 | 1.7821 | 0.037364 | *BMP10* |
| ENSG00000164756 | 8 | 117952512 | 118198953 | 518 | 57 | 42763 | 1.7821 | 0.037368 | *SLC30A8* |
| ENSG00000196557 | 16 | 1193241 | 1281771 | 347 | 75 | 49226 | 1.7815 | 0.037419 | *CACNA1H* |
| ENSG00000205981 | 3 | 180691497 | 180717562 | 33 | 9 | 46691 | 1.7812 | 0.037439 | *DNAJC19* |
| ENSG00000205030 | 11 | 55584695 | 55605630 | 73 | 16 | 49989 | 1.7809 | 0.037468 | *OR5L2* |
| ENSG00000125970 | 20 | 32571452 | 32706114 | 192 | 22 | 45741 | 1.7808 | 0.037477 | *RALY* |
| ENSG00000167889 | 17 | 74854538 | 74956475 | 585 | 83 | 43470 | 1.7803 | 0.037515 | *MGAT5B* |
| ENSG00000185009 | 10 | 75871524 | 75920821 | 122 | 14 | 43436 | 1.7801 | 0.037529 | *AP3M1* |
| ENSG00000112337 | 6 | 25902982 | 25940946 | 129 | 14 | 40475 | 1.7795 | 0.037581 | *SLC17A2* |

| | | | | | | | | | |
|---|---|---|---|---|---|---|---|---|---|
| ENSG00000260230 | 9 | 111882573 | 111939571 | 242 | 33 | 51184 | 1.7793 | 0.037594 | *FRRS1L* |
| ENSG00000155629 | 10 | 98343069 | 98490271 | 421 | 71 | 45680 | 1.7792 | 0.037607 | *PIK3AP1* |
| ENSG00000177971 | 15 | 75921426 | 75951047 | 51 | 13 | 38786 | 1.779 | 0.037618 | *IMP3* |
| ENSG00000040608 | 22 | 20218938 | 20280769 | 124 | 37 | 44583 | 1.7788 | 0.037634 | *RTN4R* |
| ENSG00000171885 | 18 | 24422002 | 24455782 | 112 | 29 | 46628 | 1.7766 | 0.037816 | *AQP4* |
| ENSG00000125864 | 20 | 17464550 | 17559865 | 351 | 80 | 47777 | 1.7763 | 0.037838 | *BFSP1* |
| ENSG00000140006 | 14 | 64053757 | 64118579 | 169 | 17 | 32580 | 1.7754 | 0.037916 | *WDR89* |
| ENSG00000224361 | 2 | 23719913 | 23757214 | 130 | 24 | 45505 | 1.7751 | 0.03794 | *AC011239.1* |
| ENSG00000131748 | 17 | 37783318 | 37829737 | 124 | 18 | 38054 | 1.775 | 0.037945 | *STARD3* |
| ENSG00000136141 | 13 | 47117303 | 47337175 | 674 | 79 | 47294 | 1.7743 | 0.038004 | *LRCH1* |
| ENSG00000268069 | 12 | 48168706 | 48189787 | 81 | 17 | 53064 | 1.774 | 0.038029 | *AC004466.1* |
| ENSG00000179588 | 16 | 88509725 | 88613424 | 446 | 71 | 44688 | 1.7735 | 0.038073 | *ZFPM1* |
| ENSG00000172199 | 11 | 56133100 | 56154029 | 85 | 12 | 49724 | 1.7732 | 0.038094 | *OR8U1* |
| ENSG00000127084 | 9 | 95699733 | 95808518 | 369 | 49 | 48374 | 1.7728 | 0.03813 | *FGD3* |
| ENSG00000205189 | 8 | 81387854 | 81448500 | 164 | 26 | 41321 | 1.7712 | 0.038267 | *ZBTB10* |
| ENSG00000162755 | 1 | 161058151 | 161080136 | 56 | 17 | 52985 | 1.771 | 0.038283 | *KLHDC9* |
| ENSG00000156140 | 4 | 73136686 | 73444516 | 615 | 56 | 47505 | 1.7708 | 0.0383 | *ADAMTS3* |
| ENSG00000163959 | 3 | 195928358 | 195980049 | 213 | 32 | 50766 | 1.7703 | 0.038342 | *SLC51A* |
| ENSG00000125445 | 17 | 73247755 | 73272454 | 60 | 9 | 34211 | 1.7703 | 0.038342 | *MRPS7* |
| ENSG00000078814 | 20 | 33553206 | 33600240 | 141 | 17 | 48895 | 1.77 | 0.038366 | *MYH7B* |
| ENSG00000132004 | 19 | 12788867 | 12817457 | 23 | 11 | 48361 | 1.7694 | 0.038412 | *FBXW9* |
| ENSG00000254997 | 11 | 71249466 | 71270653 | 101 | 9 | 48337 | 1.7693 | 0.038421 | *KRTAP5-9* |
| ENSG00000101079 | 20 | 35270169 | 35384481 | 133 | 45 | 46337 | 1.769 | 0.038447 | *NDRG3* |
| ENSG00000164683 | 8 | 80666245 | 80690098 | 52 | 14 | 52008 | 1.7687 | 0.038474 | *HEY1* |
| ENSG00000112367 | 6 | 110002499 | 110156631 | 395 | 47 | 32489 | 1.768 | 0.038531 | *FIG4* |
| ENSG00000111653 | 12 | 6749446 | 6782314 | 38 | 13 | 42312 | 1.7677 | 0.038558 | *ING4* |
| ENSG00000161860 | 19 | 12999600 | 13040090 | 94 | 15 | 45483 | 1.7674 | 0.038583 | *SYCE2* |
| ENSG00000255508 | 11 | 62317075 | 62369003 | 79 | 12 | 44296 | 1.7673 | 0.038591 | *MIR3654* |
| ENSG00000178591 | 20 | 57891 | 87735 | 48 | 22 | 41406 | 1.7673 | 0.038591 | *DEFB125* |
| ENSG00000078369 | 1 | 1706729 | 1832495 | 199 | 31 | 44048 | 1.7672 | 0.038599 | *GNB1* |
| ENSG00000130948 | 9 | 98987588 | 99074434 | 280 | 41 | 39166 | 1.7671 | 0.038608 | *HSD17B3* |
| ENSG00000171223 | 19 | 12892310 | 12914124 | 48 | 15 | 40441 | 1.7671 | 0.038609 | *JUNB* |
| ENSG00000128923 | 15 | 59053391 | 59164099 | 282 | 34 | 41470 | 1.767 | 0.038618 | *FAM63B* |
| ENSG00000126814 | 14 | 61428169 | 61458076 | 32 | 15 | 45823 | 1.7668 | 0.038634 | *TRMT5* |
| ENSG00000101638 | 18 | 44249081 | 44349220 | 395 | 75 | 46425 | 1.7667 | 0.038642 | *ST8SIA5* |
| ENSG00000204542 | 6 | 31069000 | 31090336 | 335 | 29 | 46815 | 1.766 | 0.038694 | *C6orf15* |

| | | | | | | | | | |
|---|---|---|---|---|---|---|---|---|---|
| ENSG00000205409 | 11 | 5852158 | 5873182 | 148 | 7 | 55690 | 1.766 | 0.038699 | *OR52E6* |
| ENSG00000250305 | 8 | 12793151 | 12899012 | 721 | 75 | 49219 | 1.7659 | 0.038704 | *KIAA1456* |
| ENSG00000237388 | 11 | 48500269 | 48521332 | 79 | 13 | 29380 | 1.7657 | 0.03872 | *OR4A47* |
| ENSG00000175985 | 14 | 69941409 | 70005215 | 181 | 34 | 46572 | 1.7654 | 0.038745 | *PLEKHD1* |
| ENSG00000163947 | 3 | 56751446 | 57123357 | 1263 | 142 | 44376 | 1.7651 | 0.038777 | *ARHGEF3* |
| ENSG00000112365 | 6 | 109773797 | 109814440 | 106 | 23 | 38784 | 1.765 | 0.038782 | *ZBTB24* |
| ENSG00000113525 | 5 | 131867136 | 131902530 | 41 | 11 | 34168 | 1.7649 | 0.038791 | *IL5* |
| ENSG00000196475 | 4 | 80317508 | 80339372 | 23 | 10 | 47652 | 1.7646 | 0.038814 | *GK2* |
| ENSG00000125788 | 20 | 113010 | 136392 | 72 | 15 | 36611 | 1.7644 | 0.038829 | *DEFB126* |
| ENSG00000147457 | 8 | 23091150 | 23129512 | 155 | 24 | 37529 | 1.7644 | 0.038833 | *CHMP7* |
| ENSG00000149201 | 11 | 86075778 | 86144151 | 279 | 40 | 43117 | 1.7641 | 0.038859 | *CCDC81* |
| ENSG00000214114 | 1 | 39318636 | 39357289 | 117 | 16 | 33391 | 1.7628 | 0.038971 | *MYCBP* |
| ENSG00000068097 | 17 | 58110555 | 58166292 | 75 | 6 | 33422 | 1.7624 | 0.039 | *HEATR6* |
| ENSG00000167105 | 17 | 48338767 | 48368844 | 74 | 29 | 47291 | 1.7623 | 0.039011 | *TMEM92* |
| ENSG00000134533 | 12 | 15250717 | 15511609 | 600 | 59 | 52008 | 1.7615 | 0.039074 | *RERG* |
| ENSG00000214999 | 17 | 7972800 | 7994883 | 66 | 21 | 46065 | 1.7615 | 0.039075 | *AC129492.6* |
| ENSG00000134532 | 12 | 23672440 | 24113966 | 1242 | 189 | 49263 | 1.7601 | 0.039196 | *SOX5* |
| ENSG00000146385 | 6 | 132863832 | 132884860 | 98 | 20 | 43105 | 1.7597 | 0.03923 | *TAAR8* |
| ENSG00000166398 | 19 | 34735442 | 34856491 | 318 | 32 | 37038 | 1.7592 | 0.03927 | *KIAA0355* |
| ENSG00000168646 | 17 | 63514681 | 63567765 | 145 | 32 | 44006 | 1.7592 | 0.039274 | *AXIN2* |
| ENSG00000143256 | 1 | 161060346 | 161097901 | 80 | 18 | 53471 | 1.7588 | 0.039308 | *PFDN2* |
| ENSG00000081087 | 6 | 108352613 | 108497058 | 402 | 52 | 38997 | 1.7584 | 0.03934 | *OSTM1* |
| ENSG00000178913 | 5 | 140688057 | 140710330 | 79 | 14 | 47106 | 1.7584 | 0.039343 | *TAF7* |
| ENSG00000229859 | 11 | 60960779 | 60990570 | 29 | 6 | 49732 | 1.7581 | 0.039369 | *PGA3* |
| ENSG00000258864 | 5 | 112152910 | 112213279 | 224 | 27 | 41615 | 1.7578 | 0.039388 | *CTC-554D6.1* |
| ENSG00000111181 | 12 | 289243 | 333736 | 230 | 47 | 40382 | 1.7578 | 0.039388 | *SLC6A12* |
| ENSG00000161800 | 12 | 50360706 | 50436919 | 120 | 15 | 35654 | 1.7573 | 0.039434 | *RACGAP1* |
| ENSG00000149557 | 11 | 125305646 | 125376213 | 172 | 25 | 53877 | 1.7571 | 0.039448 | *FEZ1* |
| ENSG00000143006 | 1 | 53915072 | 53943161 | 82 | 12 | 37575 | 1.7571 | 0.039451 | *DMRTB1* |
| ENSG00000113211 | 5 | 140519683 | 140542868 | 86 | 15 | 43523 | 1.7566 | 0.039491 | *PCDHB6* |
| ENSG00000177106 | 11 | 684438 | 737727 | 199 | 40 | 46630 | 1.7566 | 0.039492 | *EPS8L2* |
| ENSG00000171014 | 11 | 123800250 | 123821344 | 96 | 15 | 54156 | 1.7557 | 0.039569 | *OR4D5* |
| ENSG00000160161 | 19 | 19639057 | 19667468 | 62 | 11 | 45184 | 1.7556 | 0.039582 | *CILP2* |
| ENSG00000124610 | 6 | 26007260 | 26028040 | 53 | 14 | 39687 | 1.7554 | 0.039599 | *HIST1H1A* |
| ENSG00000143614 | 1 | 153767201 | 153905451 | 290 | 26 | 47264 | 1.755 | 0.039626 | *GATAD2B* |
| ENSG00000163703 | 3 | 9965506 | 9997097 | 84 | 23 | 38002 | 1.7543 | 0.039693 | *CRELD1* |

| | | | | | | | | | |
|---|---|---|---|---|---|---|---|---|---|
| ENSG00000173452 | 7 | 19748933 | 19823221 | 336 | 27 | 33711 | 1.754 | 0.039717 | *TMEM196* |
| ENSG00000213123 | 3 | 196008090 | 196055170 | 151 | 14 | 48132 | 1.7538 | 0.039729 | *TCTEX1D2* |
| ENSG00000066855 | 8 | 66546969 | 66693496 | 429 | 35 | 49423 | 1.7537 | 0.039739 | *MTFR1* |
| ENSG00000112081 | 6 | 36552145 | 36583377 | 161 | 13 | 53362 | 1.7536 | 0.03975 | *SRSF3* |
| ENSG00000137142 | 9 | 38398991 | 38434444 | 196 | 27 | 40910 | 1.7533 | 0.039772 | *IGFBPL1* |
| ENSG00000164484 | 6 | 130676879 | 130774208 | 339 | 31 | 39206 | 1.7533 | 0.039778 | *TMEM200A* |
| ENSG00000171150 | 2 | 46916091 | 47000268 | 322 | 34 | 47267 | 1.7531 | 0.039789 | *SOCS5* |
| ENSG00000152932 | 5 | 57868048 | 58165213 | 1025 | 102 | 49824 | 1.7524 | 0.039853 | *RAB3C* |
| ENSG00000176410 | 7 | 73085299 | 73107783 | 87 | 8 | 52335 | 1.7521 | 0.039875 | *DNAJC30* |
| ENSG00000164099 | 4 | 119191193 | 119284158 | 267 | 35 | 44209 | 1.7521 | 0.039878 | *PRSS12* |
| ENSG00000205810 | 12 | 10554911 | 10583194 | 51 | 12 | 46350 | 1.7519 | 0.039894 | *KLRC3* |
| ENSG00000182173 | 17 | 73502141 | 73530820 | 146 | 21 | 39946 | 1.7511 | 0.039961 | *TSEN54* |
| ENSG00000125434 | 17 | 8181081 | 8208661 | 100 | 19 | 46683 | 1.7504 | 0.040024 | *SLC25A35* |
| ENSG00000064999 | 6 | 34847042 | 35069179 | 611 | 59 | 37975 | 1.7501 | 0.040047 | *ANKS1A* |
| ENSG00000221968 | 11 | 61630991 | 61669523 | 90 | 17 | 45474 | 1.7498 | 0.040076 | *FADS3* |
| ENSG00000002726 | 7 | 150511715 | 150568592 | 295 | 32 | 36732 | 1.7496 | 0.040093 | *AOC1* |
| ENSG00000181698 | 11 | 56033029 | 56054146 | 74 | 13 | 54428 | 1.7485 | 0.040184 | *OR5T1* |
| ENSG00000105755 | 19 | 44000871 | 44041396 | 135 | 17 | 51511 | 1.7482 | 0.040219 | *ETHE1* |
| ENSG00000146216 | 6 | 43201418 | 43265997 | 191 | 29 | 39268 | 1.7478 | 0.040247 | *TTBK1* |
| ENSG00000108344 | 17 | 38127050 | 38164213 | 144 | 14 | 48331 | 1.7477 | 0.040257 | *PSMD3* |
| ENSG00000055130 | 7 | 148385006 | 148508128 | 324 | 35 | 52411 | 1.7475 | 0.040277 | *CUL1* |
| ENSG00000132912 | 5 | 150078002 | 150148671 | 121 | 10 | 28352 | 1.7463 | 0.04038 | *DCTN4* |
| ENSG00000113643 | 5 | 167903450 | 167956304 | 251 | 19 | 49963 | 1.7451 | 0.040483 | *RARS* |
| ENSG00000204644 | 6 | 29630169 | 29658887 | 281 | 26 | 39120 | 1.745 | 0.040494 | *ZFP57* |
| ENSG00000118804 | 4 | 77162886 | 77242752 | 315 | 28 | 46580 | 1.7444 | 0.040541 | *FAM47E-STBD1* |
| ENSG00000128052 | 4 | 55934644 | 56001756 | 180 | 38 | 49076 | 1.7442 | 0.040562 | *KDR* |
| ENSG00000096717 | 10 | 69634427 | 69688147 | 162 | 20 | 36528 | 1.7439 | 0.040589 | *SIRT1* |
| ENSG00000197982 | 1 | 38262651 | 38285126 | 39 | 10 | 42008 | 1.7433 | 0.040643 | *C1orf122* |
| ENSG00000138100 | 2 | 27495260 | 27540307 | 70 | 15 | 41094 | 1.743 | 0.040666 | *TRIM54* |
| ENSG00000214290 | 11 | 111159565 | 111189460 | 114 | 21 | 43214 | 1.7429 | 0.040672 | *COLCA2* |
| ENSG00000123444 | 11 | 47583749 | 47610567 | 34 | 11 | 41035 | 1.7429 | 0.04068 | *KBTBD4* |
| ENSG00000070770 | 16 | 58181811 | 58241824 | 194 | 18 | 51359 | 1.7421 | 0.040744 | *CSNK2A2* |
| ENSG00000204446 | 9 | 89753559 | 89784471 | 132 | 12 | 55566 | 1.7415 | 0.040794 | *C9orf170* |
| ENSG00000160097 | 1 | 33317869 | 33348083 | 52 | 11 | 54948 | 1.741 | 0.040842 | *FNDC5* |
| ENSG00000184640 | 17 | 75266651 | 75506678 | 781 | 117 | 42299 | 1.7409 | 0.040849 | *Sep-09* |
| ENSG00000268313 | 1 | 205672497 | 205694153 | 55 | 18 | 42182 | 1.7408 | 0.040861 | *AC119673.1* |

| | | | | | | | | | |
|---|---|---|---|---|---|---|---|---|---|
| ENSG00000181689 | 11 | 56075783 | 56096721 | 86 | 10 | 55703 | 1.7404 | 0.040892 | *OR8K3* |
| ENSG00000087076 | 19 | 49306274 | 49349935 | 133 | 29 | 43343 | 1.7397 | 0.04096 | *HSD17B14* |
| ENSG00000259030 | 1 | 74653919 | 75019666 | 727 | 101 | 40453 | 1.7389 | 0.041026 | *FPGT-TNNI3K* |
| ENSG00000261796 | 3 | 128796418 | 128889879 | 98 | 22 | 38412 | 1.7389 | 0.041028 | *ISY1-RAB43* |
| ENSG00000175229 | 11 | 65799423 | 65826651 | 63 | 9 | 36042 | 1.7386 | 0.041056 | *GAL3ST3* |
| ENSG00000163286 | 2 | 233261553 | 233285424 | 88 | 18 | 47863 | 1.7378 | 0.041125 | *ALPPL2* |
| ENSG00000165623 | 10 | 13253767 | 13286334 | 206 | 44 | 47829 | 1.7362 | 0.041263 | *UCMA* |
| ENSG00000182185 | 14 | 68276496 | 69206935 | 2478 | 182 | 42444 | 1.7362 | 0.041265 | *RAD51B* |
| ENSG00000070182 | 14 | 65203002 | 65356601 | 560 | 79 | 40864 | 1.7361 | 0.041269 | *SPTB* |
| ENSG00000152154 | 2 | 39882122 | 39955103 | 118 | 14 | 41909 | 1.7358 | 0.041298 | *TMEM178A* |
| ENSG00000205838 | 5 | 34828938 | 34909561 | 252 | 43 | 43814 | 1.7358 | 0.0413 | *TTC23L* |
| ENSG00000129353 | 19 | 10703133 | 10765235 | 197 | 26 | 48212 | 1.7354 | 0.041339 | *SLC44A2* |
| ENSG00000272414 | 4 | 77162874 | 77242282 | 313 | 28 | 46652 | 1.735 | 0.041367 | *FAM47E-STBD1* |
| ENSG00000008083 | 6 | 15236527 | 15532252 | 930 | 112 | 52831 | 1.7337 | 0.041481 | *JARID2* |
| ENSG00000162636 | 1 | 109092711 | 109197522 | 222 | 15 | 33932 | 1.7335 | 0.0415 | *FAM102B* |
| ENSG00000130939 | 1 | 10082890 | 10251297 | 423 | 39 | 41268 | 1.733 | 0.041551 | *UBE4B* |
| ENSG00000078043 | 18 | 44378353 | 44510123 | 397 | 31 | 53176 | 1.7324 | 0.041603 | *PIAS2* |
| ENSG00000161664 | 17 | 42237815 | 42266451 | 66 | 10 | 33676 | 1.7314 | 0.041693 | *ASB16* |
| ENSG00000260234 | 3 | 150590784 | 150669530 | 236 | 54 | 43261 | 1.7308 | 0.041741 | *RP11-166N6.3* |
| ENSG00000144821 | 3 | 108089216 | 108258169 | 396 | 57 | 45994 | 1.7297 | 0.04184 | *MYH15* |
| ENSG00000133805 | 11 | 10319860 | 10539126 | 915 | 123 | 47018 | 1.7296 | 0.041851 | *AMPD3* |
| ENSG00000142686 | 1 | 36169476 | 36195073 | 22 | 11 | 50895 | 1.7284 | 0.041958 | *C1orf216* |
| ENSG00000173421 | 3 | 49225861 | 49305537 | 119 | 17 | 46359 | 1.7279 | 0.042 | *CCDC36* |
| ENSG00000124562 | 6 | 34715183 | 34751571 | 154 | 11 | 33399 | 1.7269 | 0.042089 | *SNRPC* |
| ENSG00000170893 | 3 | 129683148 | 129706781 | 74 | 14 | 37642 | 1.7261 | 0.042164 | *TRH* |
| ENSG00000214309 | 7 | 99714317 | 99736118 | 39 | 11 | 47187 | 1.7258 | 0.042195 | *MBLAC1* |
| ENSG00000100209 | 22 | 29128019 | 29163503 | 109 | 22 | 44792 | 1.7257 | 0.042199 | *HSCB* |
| ENSG00000117525 | 1 | 94984781 | 95017356 | 47 | 13 | 51632 | 1.7257 | 0.042204 | *F3* |
| ENSG00000007402 | 3 | 50390233 | 50551675 | 225 | 33 | 41466 | 1.7242 | 0.042332 | *CACNA2D2* |
| ENSG00000140386 | 15 | 76630526 | 77207785 | 1276 | 45 | 41862 | 1.7242 | 0.042332 | *SCAPER* |
| ENSG00000259305 | 8 | 124228431 | 124289627 | 30 | 19 | 49453 | 1.7237 | 0.042382 | *ZHX1-C8ORF76* |
| ENSG00000258572 | 14 | 95972473 | 95994248 | 76 | 22 | 49295 | 1.7235 | 0.042404 | *RP11-1070N10.3* |
| ENSG00000164494 | 6 | 107463761 | 107790768 | 803 | 90 | 42030 | 1.7234 | 0.04241 | *PDSS2* |
| ENSG00000175414 | 5 | 175782471 | 175838866 | 136 | 29 | 52203 | 1.7232 | 0.042426 | *ARL10* |
| ENSG00000141140 | 17 | 34841477 | 34909284 | 149 | 18 | 33786 | 1.7231 | 0.042435 | *MYO19* |
| ENSG00000175591 | 11 | 72919343 | 72957397 | 67 | 30 | 44522 | 1.7224 | 0.0425 | *P2RY2* |

| | | | | | | | | | |
|---|---|---|---|---|---|---|---|---|---|
| ENSG00000185432 | 12 | 51307255 | 51336300 | 93 | 18 | 46749 | 1.722 | 0.042532 | *METTL7A* |
| ENSG00000112182 | 6 | 90626248 | 91016627 | 848 | 140 | 46082 | 1.722 | 0.042535 | *BACH2* |
| ENSG00000167644 | 19 | 38784801 | 38805649 | 74 | 13 | 54302 | 1.7216 | 0.042573 | *C19orf33* |
| ENSG00000254647 | 11 | 2171009 | 2192571 | 54 | 15 | 38614 | 1.7206 | 0.042661 | *INS* |
| ENSG00000100023 | 22 | 21996559 | 22064304 | 309 | 39 | 48031 | 1.72 | 0.042714 | *PPIL2* |
| ENSG00000013375 | 6 | 83860869 | 83913655 | 92 | 19 | 46274 | 1.7196 | 0.042748 | *PGM3* |
| ENSG00000213420 | 7 | 99757229 | 99784995 | 53 | 10 | 48077 | 1.7197 | 0.042748 | *GPC2* |
| ENSG00000096060 | 6 | 35531362 | 35706360 | 412 | 46 | 48192 | 1.7195 | 0.04276 | *FKBP5* |
| ENSG00000153246 | 2 | 160778519 | 160929121 | 535 | 57 | 37396 | 1.7189 | 0.042817 | *PLA2R1* |
| ENSG00000066933 | 15 | 72104632 | 72420918 | 583 | 40 | 48791 | 1.7185 | 0.042849 | *MYO9A* |
| ENSG00000120519 | 4 | 147165127 | 147453123 | 664 | 60 | 44252 | 1.7185 | 0.042854 | *SLC10A7* |
| ENSG00000105607 | 19 | 12991840 | 13035021 | 100 | 16 | 45930 | 1.7173 | 0.042966 | *GCDH* |
| ENSG00000095932 | 19 | 3464405 | 3490540 | 77 | 20 | 40733 | 1.717 | 0.042991 | *C19orf77* |
| ENSG00000007202 | 17 | 26931458 | 26982472 | 41 | 16 | 50255 | 1.717 | 0.042993 | *KIAA0100* |
| ENSG00000258728 | 9 | 34636642 | 34667110 | 68 | 14 | 54867 | 1.7169 | 0.042999 | *GALT* |
| ENSG00000115355 | 2 | 55504978 | 55657057 | 544 | 35 | 37834 | 1.7166 | 0.043027 | *CCDC88A* |
| ENSG00000076685 | 10 | 104835940 | 104963056 | 360 | 27 | 47706 | 1.7163 | 0.04305 | *NT5C2* |
| ENSG00000105373 | 19 | 48238779 | 48270315 | 184 | 20 | 38911 | 1.7151 | 0.04316 | *GLTSCR2* |
| ENSG00000182095 | 7 | 5336421 | 5475045 | 535 | 107 | 43911 | 1.7148 | 0.043187 | *TNRC18* |
| ENSG00000118496 | 6 | 146104638 | 146145889 | 97 | 16 | 51512 | 1.7143 | 0.043233 | *FBXO30* |
| ENSG00000073670 | 17 | 42826399 | 42869214 | 107 | 17 | 54277 | 1.7142 | 0.043247 | *ADAM11* |
| ENSG00000124102 | 20 | 43793517 | 43815185 | 76 | 5 | 52641 | 1.714 | 0.043269 | *PI3* |
| ENSG00000198523 | 6 | 118859461 | 118891893 | 94 | 15 | 54801 | 1.7139 | 0.043272 | *PLN* |
| ENSG00000076555 | 12 | 109544400 | 109716031 | 572 | 84 | 43381 | 1.7139 | 0.043275 | *ACACB* |
| ENSG00000236761 | 6 | 132019581 | 132042206 | 24 | 9 | 41082 | 1.7137 | 0.04329 | *CTAGE9* |
| ENSG00000106636 | 7 | 44230567 | 44263893 | 122 | 16 | 55951 | 1.7137 | 0.043293 | *YKT6* |
| ENSG00000177042 | 11 | 685428 | 715028 | 122 | 22 | 48315 | 1.7135 | 0.043315 | *TMEM80* |
| ENSG00000144366 | 2 | 189146396 | 189470653 | 799 | 63 | 47005 | 1.7134 | 0.043318 | *GULP1* |
| ENSG00000172780 | 3 | 128796412 | 128851644 | 53 | 16 | 39907 | 1.713 | 0.043354 | *RAB43* |
| ENSG00000102996 | 16 | 58049470 | 58090805 | 86 | 20 | 47743 | 1.7128 | 0.043373 | *MMP15* |
| ENSG00000221983 | 19 | 18672540 | 18698360 | 88 | 11 | 30815 | 1.7126 | 0.043395 | *UBA52* |
| ENSG00000087269 | 4 | 2929660 | 2975112 | 120 | 21 | 42478 | 1.7117 | 0.043472 | *NOP14* |
| ENSG00000079689 | 6 | 25642464 | 25712011 | 267 | 38 | 49411 | 1.711 | 0.04354 | *SCGN* |
| ENSG00000116731 | 1 | 14016693 | 14161574 | 463 | 51 | 46530 | 1.7106 | 0.043579 | *PRDM2* |
| ENSG00000163629 | 4 | 87505468 | 87746324 | 748 | 34 | 53618 | 1.71 | 0.043633 | *PTPN13* |
| ENSG00000172680 | 8 | 57015501 | 57036541 | 81 | 15 | 50635 | 1.71 | 0.043633 | *MOS* |

| | | | | | | | | | |
|---|---|---|---|---|---|---|---|---|---|
| ENSG00000221813 | 7 | 143691022 | 143712068 | 55 | 5 | 36141 | 1.7096 | 0.043673 | *OR6B1* |
| ENSG00000107874 | 10 | 104173002 | 104202418 | 52 | 15 | 52507 | 1.7094 | 0.043688 | *CUEDC2* |
| ENSG00000215462 | 13 | 49997495 | 50030554 | 90 | 18 | 42515 | 1.709 | 0.043729 | *AL136218.1* |
| ENSG00000106771 | 9 | 111767432 | 111892225 | 532 | 48 | 51667 | 1.7085 | 0.04377 | *TMEM245* |
| ENSG00000100906 | 14 | 35860717 | 35883955 | 99 | 22 | 52925 | 1.7076 | 0.043851 | *NFKBIA* |
| ENSG00000134684 | 1 | 33230840 | 33293754 | 124 | 15 | 53329 | 1.7075 | 0.043862 | *YARS* |
| ENSG00000071243 | 7 | 120580803 | 120627270 | 98 | 14 | 31976 | 1.7069 | 0.043924 | *ING3* |
| ENSG00000066468 | 10 | 123227848 | 123367972 | 286 | 66 | 45072 | 1.7068 | 0.043926 | *FGFR2* |
| ENSG00000172932 | 11 | 67046018 | 67079956 | 42 | 15 | 37339 | 1.7061 | 0.043991 | *ANKRD13D* |
| ENSG00000072518 | 11 | 63596400 | 63688491 | 151 | 16 | 30988 | 1.7053 | 0.044065 | *MARK2* |
| ENSG00000167632 | 8 | 140732586 | 141478678 | 2436 | 237 | 43938 | 1.7052 | 0.044079 | *TRAPPC9* |
| ENSG00000105339 | 8 | 142117377 | 142215907 | 246 | 54 | 47779 | 1.7045 | 0.044146 | *DENND3* |
| ENSG00000178031 | 9 | 18463892 | 18920948 | 2079 | 238 | 50242 | 1.7032 | 0.044263 | *ADAMTSL1* |
| ENSG00000255012 | 11 | 56370031 | 56390978 | 71 | 13 | 50115 | 1.7032 | 0.044266 | *OR5M1* |
| ENSG00000162654 | 1 | 89636831 | 89674615 | 94 | 18 | 46554 | 1.7028 | 0.044299 | *GBP4* |
| ENSG00000162745 | 1 | 161942982 | 162003644 | 187 | 44 | 44824 | 1.7023 | 0.044346 | *OLFML2B* |
| ENSG00000128191 | 22 | 20057755 | 20109400 | 193 | 22 | 52670 | 1.702 | 0.044378 | *DGCR8* |
| ENSG00000160868 | 7 | 99344604 | 99391888 | 49 | 15 | 36457 | 1.7015 | 0.044426 | *CYP3A4* |
| ENSG00000198723 | 19 | 7547862 | 7583336 | 124 | 28 | 50202 | 1.701 | 0.044467 | *C19orf45* |
| ENSG00000087884 | 11 | 77522155 | 77639478 | 269 | 22 | 50109 | 1.701 | 0.044471 | *AAMDC* |
| ENSG00000169371 | 15 | 75880424 | 75928810 | 95 | 10 | 29446 | 1.7006 | 0.044508 | *SNUPN* |
| ENSG00000164663 | 6 | 41747634 | 41873099 | 289 | 32 | 48888 | 1.7002 | 0.044546 | *USP49* |
| ENSG00000171587 | 21 | 41372926 | 42229065 | 3524 | 307 | 43713 | 1.7002 | 0.044548 | *DSCAM* |
| ENSG00000196963 | 5 | 140550980 | 140575793 | 105 | 15 | 43549 | 1.7001 | 0.044558 | *PCDHB16* |
| ENSG00000181027 | 19 | 47239303 | 47290245 | 164 | 26 | 52076 | 1.6993 | 0.044629 | *FKRP* |
| ENSG00000164403 | 5 | 132147833 | 132176590 | 34 | 9 | 45076 | 1.6993 | 0.044634 | *SHROOM1* |
| ENSG00000149016 | 11 | 62332517 | 62369649 | 51 | 10 | 46663 | 1.6991 | 0.044653 | *TUT1* |
| ENSG00000085719 | 8 | 87487059 | 87583726 | 315 | 33 | 49387 | 1.6986 | 0.044697 | *CPNE3* |
| ENSG00000167005 | 16 | 56453045 | 56496111 | 102 | 17 | 39649 | 1.6983 | 0.044723 | *NUDT21* |
| ENSG00000213892 | 19 | 45192421 | 45223986 | 136 | 33 | 50313 | 1.6971 | 0.044837 | *CEACAM16* |
| ENSG00000173457 | 11 | 64001956 | 64024413 | 53 | 12 | 42138 | 1.697 | 0.044853 | *PPP1R14B* |
| ENSG00000144655 | 3 | 39173346 | 39206053 | 109 | 24 | 41042 | 1.6965 | 0.044891 | *CSRNP1* |
| ENSG00000090006 | 19 | 41088789 | 41145725 | 183 | 23 | 45205 | 1.6963 | 0.044913 | *LTBP4* |
| ENSG00000197921 | 1 | 2450184 | 2471684 | 60 | 15 | 32992 | 1.6963 | 0.044916 | *HES5* |
| ENSG00000182368 | 9 | 45717107 | 45738274 | 1 | 1 | 42668 | 1.6959 | 0.04495 | *FAM27A* |
| ENSG00000204807 | 9 | 45723559 | 45744896 | 1 | 1 | 42668 | 1.6959 | 0.04495 | *FAM27E2* |

| | | | | | | | | | |
|---|---|---|---|---|---|---|---|---|---|
| ENSG00000134982 | 5 | 112033195 | 112191936 | 454 | 42 | 40610 | 1.6957 | 0.04497 | *APC* |
| ENSG00000183775 | 5 | 143540396 | 143875249 | 672 | 53 | 51392 | 1.6956 | 0.044981 | *KCTD16* |
| ENSG00000137216 | 6 | 44084651 | 44133256 | 178 | 28 | 42853 | 1.6953 | 0.045006 | *TMEM63B* |
| ENSG00000170606 | 5 | 132377654 | 132452141 | 224 | 18 | 52368 | 1.6949 | 0.045044 | *HSPA4* |
| ENSG00000108924 | 17 | 53332373 | 53412426 | 191 | 41 | 42717 | 1.6949 | 0.04505 | *HLF* |
| ENSG00000100664 | 14 | 103789881 | 103821362 | 122 | 20 | 49186 | 1.6948 | 0.045055 | *EIF5* |
| ENSG00000172459 | 11 | 56421138 | 56442163 | 75 | 14 | 52374 | 1.6948 | 0.045061 | *OR5AR1* |
| ENSG00000269099 | 13 | 25581541 | 25601675 | 125 | 8 | 53676 | 1.6948 | 0.045061 | *LSP1* |
| ENSG00000085491 | 1 | 108666658 | 108753471 | 320 | 35 | 51205 | 1.6942 | 0.045115 | *SLC25A24* |
| ENSG00000251283 | 4 | 157545646 | 157573606 | 120 | 17 | 54324 | 1.6941 | 0.045128 | *RP11-171N4.2* |
| ENSG00000100605 | 14 | 93393259 | 93592665 | 646 | 73 | 46344 | 1.6937 | 0.045162 | *ITPK1* |
| ENSG00000163012 | 2 | 187682562 | 187723935 | 176 | 9 | 30146 | 1.6933 | 0.045203 | *ZSWIM2* |
| ENSG00000170889 | 19 | 54694610 | 54762862 | 212 | 45 | 36189 | 1.6933 | 0.045203 | *RPS9* |
| ENSG00000167208 | 16 | 50690211 | 50725264 | 53 | 14 | 38423 | 1.6931 | 0.045216 | *SNX20* |
| ENSG00000145041 | 3 | 51423298 | 51544010 | 109 | 35 | 50336 | 1.693 | 0.045224 | *VPRBP* |
| ENSG00000213996 | 19 | 19365173 | 19394200 | 100 | 18 | 41197 | 1.6925 | 0.045273 | *TM6SF2* |
| ENSG00000178252 | 3 | 49034495 | 49063386 | 26 | 9 | 37051 | 1.692 | 0.045324 | *WDR6* |
| ENSG00000099814 | 14 | 105321617 | 105373107 | 202 | 21 | 33770 | 1.6913 | 0.045387 | *CEP170B* |
| ENSG00000151702 | 11 | 128546430 | 128693162 | 474 | 80 | 44269 | 1.6912 | 0.045404 | *FLI1* |
| ENSG00000024422 | 19 | 48206600 | 48256391 | 187 | 37 | 48294 | 1.6911 | 0.045405 | *EHD2* |
| ENSG00000162341 | 11 | 68806365 | 68868072 | 186 | 15 | 40789 | 1.6905 | 0.045466 | *TPCN2* |
| ENSG00000188396 | 1 | 45261585 | 45282957 | 49 | 8 | 51067 | 1.6904 | 0.045474 | *TCTEX1D4* |
| ENSG00000054965 | 11 | 73101532 | 73319234 | 383 | 38 | 45340 | 1.6904 | 0.045477 | *FAM168A* |
| ENSG00000156466 | 8 | 97144562 | 97183020 | 79 | 17 | 37988 | 1.6903 | 0.045485 | *GDF6* |
| ENSG00000101084 | 20 | 35224137 | 35250960 | 31 | 14 | 44487 | 1.689 | 0.045612 | *C20orf24* |
| ENSG00000072832 | 4 | 5739811 | 5904785 | 798 | 128 | 46528 | 1.6888 | 0.045627 | *CRMP1* |
| ENSG00000117122 | 1 | 17290997 | 17317330 | 80 | 17 | 41683 | 1.6886 | 0.045649 | *MFAP2* |
| ENSG00000103671 | 15 | 64669947 | 64757502 | 160 | 26 | 41138 | 1.6884 | 0.045666 | *TRIP4* |
| ENSG00000135114 | 12 | 121448095 | 121487045 | 154 | 28 | 51351 | 1.6884 | 0.045671 | *OASL* |
| ENSG00000125740 | 19 | 45961253 | 45988437 | 125 | 21 | 48485 | 1.6883 | 0.045681 | *FOSB* |
| ENSG00000203710 | 1 | 207659492 | 207823992 | 375 | 48 | 37003 | 1.6881 | 0.045698 | *CR1* |
| ENSG00000103657 | 15 | 63890817 | 64136141 | 602 | 43 | 45654 | 1.688 | 0.045704 | *HERC1* |
| ENSG00000141665 | 18 | 71730588 | 71825100 | 342 | 22 | 39247 | 1.688 | 0.04571 | *FBXO15* |
| ENSG00000184574 | 12 | 6718001 | 6755613 | 54 | 16 | 38866 | 1.6868 | 0.045825 | *LPAR5* |
| ENSG00000160753 | 1 | 155280687 | 155310905 | 25 | 9 | 44956 | 1.6866 | 0.045838 | *RUSC1* |
| ENSG00000125319 | 17 | 42209274 | 42249844 | 77 | 10 | 34738 | 1.6866 | 0.045845 | *C17orf53* |

| | | | | | | | | | |
|---|---|---|---|---|---|---|---|---|---|
| ENSG00000166407 | 11 | 8235851 | 8300263 | 307 | 64 | 36819 | 1.6861 | 0.045885 | *LMO1* |
| ENSG00000112031 | 6 | 153298497 | 153333820 | 120 | 15 | 55101 | 1.686 | 0.045897 | *MTRF1L* |
| ENSG00000205238 | 7 | 102181679 | 102212755 | 1 | 1 | 43848 | 1.6859 | 0.04591 | *SPDYE2* |
| ENSG00000156076 | 12 | 65434406 | 65525346 | 204 | 29 | 34678 | 1.6857 | 0.045928 | *WIF1* |
| ENSG00000140365 | 15 | 75618232 | 75644268 | 54 | 18 | 43433 | 1.6855 | 0.045947 | *COMMD4* |
| ENSG00000023902 | 1 | 150111373 | 150146916 | 69 | 11 | 42553 | 1.685 | 0.04599 | *PLEKHO1* |
| ENSG00000166402 | 11 | 8030791 | 8137659 | 355 | 71 | 46199 | 1.6849 | 0.046006 | *TUB* |
| ENSG00000232258 | 16 | 8609502 | 8632304 | 161 | 24 | 48094 | 1.6847 | 0.04602 | *TMEM114* |
| ENSG00000133639 | 12 | 92526286 | 92549673 | 60 | 17 | 55563 | 1.684 | 0.046093 | *BTG1* |
| ENSG00000143384 | 1 | 150537032 | 150562066 | 71 | 14 | 41468 | 1.6839 | 0.046097 | *MCL1* |
| ENSG00000127328 | 12 | 70122461 | 70226984 | 327 | 35 | 40867 | 1.6825 | 0.046236 | *RAB3IP* |
| ENSG00000112210 | 6 | 57043607 | 57097078 | 69 | 16 | 49721 | 1.6824 | 0.046248 | *RAB23* |
| ENSG00000128311 | 22 | 37396900 | 37425681 | 136 | 20 | 48705 | 1.682 | 0.04628 | *TST* |
| ENSG00000225399 | 3 | 49287518 | 49308744 | 24 | 7 | 49596 | 1.6819 | 0.046293 | *RP11-3B7.1* |
| ENSG00000127423 | 1 | 26148414 | 26195903 | 115 | 7 | 43007 | 1.6819 | 0.046297 | *AUNIP* |
| ENSG00000186994 | 19 | 8377468 | 8418146 | 192 | 23 | 48682 | 1.6814 | 0.046344 | *KANK3* |
| ENSG00000065802 | 2 | 239325383 | 239370891 | 184 | 21 | 39082 | 1.6813 | 0.046355 | *ASB1* |
| ENSG00000125741 | 19 | 46020685 | 46115470 | 278 | 56 | 43639 | 1.6812 | 0.046363 | *OPA3* |
| ENSG00000222033 | 2 | 171558961 | 171581077 | 53 | 16 | 46686 | 1.6807 | 0.046415 | *LINC01124* |
| ENSG00000166211 | 12 | 101859199 | 101890775 | 121 | 25 | 36768 | 1.6805 | 0.046434 | *SPIC* |
| ENSG00000256391 | 6 | 166297330 | 166319725 | 186 | 23 | 46639 | 1.6804 | 0.04644 | *SDIM1* |
| ENSG00000170325 | 11 | 129759601 | 129882730 | 284 | 50 | 40537 | 1.6803 | 0.046452 | *PRDM10* |
| ENSG00000188379 | 9 | 21374254 | 21395396 | 34 | 11 | 45789 | 1.6802 | 0.04646 | *IFNA2* |
| ENSG00000171992 | 5 | 149970642 | 150048782 | 160 | 35 | 41103 | 1.6801 | 0.046466 | *SYNPO* |
| ENSG00000110900 | 12 | 31069362 | 31159534 | 342 | 78 | 46946 | 1.68 | 0.046476 | *TSPAN11* |
| ENSG00000249931 | 15 | 32674983 | 32705396 | 13 | 8 | 40746 | 1.6799 | 0.046484 | *GOLGA8K* |
| ENSG00000115556 | 2 | 219462488 | 219511907 | 112 | 11 | 43861 | 1.6793 | 0.046544 | *PLCD4* |
| ENSG00000180549 | 9 | 139914626 | 139937462 | 58 | 12 | 54436 | 1.6787 | 0.046605 | *FUT7* |
| ENSG00000131018 | 6 | 152432819 | 152968936 | 1981 | 242 | 47663 | 1.6778 | 0.04669 | *SYNE1* |
| ENSG00000112541 | 6 | 165730776 | 166410091 | 2763 | 284 | 46954 | 1.6778 | 0.046691 | *PDE10A* |
| ENSG00000154114 | 11 | 120884781 | 120971484 | 150 | 15 | 54362 | 1.6776 | 0.046708 | *TBCEL* |
| ENSG00000204616 | 6 | 30060674 | 30090883 | 255 | 39 | 39312 | 1.6776 | 0.04671 | *TRIM31* |
| ENSG00000162645 | 1 | 89561815 | 89626139 | 199 | 16 | 51580 | 1.6775 | 0.046722 | *GBP2* |
| ENSG00000166428 | 14 | 105381153 | 105409574 | 90 | 11 | 49771 | 1.6771 | 0.046759 | *PLD4* |
| ENSG00000136040 | 12 | 94532499 | 94711451 | 694 | 120 | 49242 | 1.6762 | 0.046845 | *PLXNC1* |
| ENSG00000165071 | 8 | 133687253 | 133782958 | 316 | 28 | 42875 | 1.6762 | 0.046851 | *TMEM71* |

| | | | | | | | | | |
|---|---|---|---|---|---|---|---|---|---|
| ENSG00000109107 | 17 | 26890133 | 26914282 | 19 | 10 | 53626 | 1.6761 | 0.046858 | *ALDOC* |
| ENSG00000169282 | 3 | 155745490 | 156266545 | 1551 | 141 | 45652 | 1.6759 | 0.046874 | *KCNAB1* |
| ENSG00000119121 | 9 | 77327411 | 77513010 | 382 | 61 | 46103 | 1.6753 | 0.04694 | *TRPM6* |
| ENSG00000142611 | 1 | 2975732 | 3365185 | 1409 | 217 | 41670 | 1.6746 | 0.047009 | *PRDM16* |
| ENSG00000165672 | 10 | 120917215 | 120948345 | 75 | 13 | 35229 | 1.6744 | 0.047021 | *PRDX3* |
| ENSG00000007908 | 1 | 169681781 | 169743846 | 434 | 26 | 48450 | 1.674 | 0.047062 | *SELE* |
| ENSG00000178104 | 1 | 144826157 | 145086186 | 69 | 29 | 32107 | 1.6736 | 0.047101 | *PDE4DIP* |
| ENSG00000248592 | 3 | 52857137 | 52941578 | 186 | 17 | 51985 | 1.6733 | 0.047133 | *TMEM110-MUSTN1* |
| ENSG00000164691 | 6 | 159445500 | 159476184 | 134 | 12 | 30387 | 1.6733 | 0.047134 | *TAGAP* |
| ENSG00000070785 | 1 | 45306450 | 45462282 | 458 | 25 | 50088 | 1.673 | 0.047165 | *EIF2B3* |
| ENSG00000169230 | 5 | 176720775 | 176743960 | 56 | 15 | 38830 | 1.6726 | 0.047203 | *PRELID1* |
| ENSG00000043039 | 11 | 129235835 | 129332171 | 374 | 79 | 46730 | 1.6724 | 0.047222 | *BARX2* |
| ENSG00000034533 | 3 | 130722719 | 130756493 | 59 | 8 | 49973 | 1.6721 | 0.047253 | *ASTE1* |
| ENSG00000169018 | 15 | 68560141 | 68598203 | 71 | 15 | 49034 | 1.672 | 0.047259 | *FEM1B* |
| ENSG00000078687 | 17 | 75990249 | 76114916 | 298 | 44 | 46330 | 1.6719 | 0.047271 | *TNRC6C* |
| ENSG00000146383 | 6 | 132881461 | 132902498 | 124 | 24 | 47898 | 1.6719 | 0.047273 | *TAAR6* |
| ENSG00000135362 | 11 | 36307838 | 36496754 | 820 | 124 | 44423 | 1.6716 | 0.047304 | *PRR5L* |
| ENSG00000164106 | 4 | 174295852 | 174337531 | 80 | 17 | 38641 | 1.6714 | 0.047324 | *SCRG1* |
| ENSG00000101343 | 20 | 20005012 | 20046690 | 113 | 15 | 52485 | 1.6712 | 0.047341 | *CRNKL1* |
| ENSG00000106415 | 7 | 7998425 | 8143902 | 551 | 53 | 49046 | 1.6711 | 0.047349 | *GLCCI1* |
| ENSG00000196172 | 19 | 23911997 | 23951693 | 94 | 16 | 44009 | 1.6708 | 0.047382 | *ZNF681* |
| ENSG00000070759 | 1 | 45799555 | 45966872 | 300 | 26 | 52517 | 1.6705 | 0.047407 | *TESK2* |
| ENSG00000113532 | 5 | 100132639 | 100248970 | 273 | 23 | 45759 | 1.6698 | 0.04748 | *ST8SIA4* |
| ENSG00000113119 | 5 | 140009012 | 140034993 | 64 | 10 | 53907 | 1.6693 | 0.047524 | *TMCO6* |
| ENSG00000196275 | 7 | 74200483 | 74277847 | 36 | 13 | 42210 | 1.6685 | 0.04761 | *GTF2IRD2* |
| ENSG00000138085 | 2 | 27424895 | 27450046 | 74 | 12 | 42953 | 1.6681 | 0.047651 | *ATRAID* |
| ENSG00000106633 | 7 | 44173872 | 44247769 | 262 | 47 | 51344 | 1.6678 | 0.047676 | *GCK* |
| ENSG00000114126 | 3 | 141653277 | 141878386 | 875 | 37 | 54251 | 1.6678 | 0.047679 | *TFDP2* |
| ENSG00000153060 | 16 | 10711358 | 10798802 | 531 | 60 | 38506 | 1.6676 | 0.047702 | *TEKT5* |
| ENSG00000197472 | 1 | 247098849 | 247181395 | 372 | 43 | 41414 | 1.6672 | 0.04774 | *ZNF695* |
| ENSG00000172031 | 1 | 92485539 | 92539093 | 151 | 19 | 47772 | 1.6666 | 0.047793 | *EPHX4* |
| ENSG00000108961 | 17 | 8181815 | 8203410 | 74 | 17 | 45508 | 1.6666 | 0.047796 | *RANGRF* |
| ENSG00000213512 | 1 | 89587434 | 89651723 | 150 | 18 | 51972 | 1.6665 | 0.047812 | *GBP7* |
| ENSG00000164022 | 4 | 107226701 | 107280383 | 154 | 16 | 52775 | 1.6663 | 0.047829 | *AIMP1* |
| ENSG00000102910 | 16 | 48268207 | 48407033 | 236 | 16 | 38006 | 1.6659 | 0.047865 | *LONP2* |
| ENSG00000119523 | 9 | 101968708 | 101994238 | 64 | 9 | 55782 | 1.6657 | 0.047886 | *ALG2* |

| | | | | | | | | | |
|---|---|---|---|---|---|---|---|---|---|
| ENSG00000130203 | 19 | 45399011 | 45422650 | 81 | 22 | 46774 | 1.6654 | 0.047917 | *APOE* |
| ENSG00000196436 | 16 | 74401776 | 74435978 | 21 | 9 | 43482 | 1.6652 | 0.047933 | *NPIPB15* |
| ENSG00000139675 | 13 | 53181605 | 53227919 | 170 | 12 | 54549 | 1.6651 | 0.047947 | *HNRNPA1L2* |
| ENSG00000211584 | 12 | 48137699 | 48186536 | 182 | 30 | 51968 | 1.665 | 0.047961 | *SLC48A1* |
| ENSG00000142273 | 19 | 45271126 | 45313891 | 115 | 22 | 38951 | 1.6641 | 0.048049 | *CBLC* |
| ENSG00000198912 | 1 | 3795689 | 3826857 | 143 | 22 | 46769 | 1.6639 | 0.048063 | *C1orf174* |
| ENSG00000144840 | 3 | 120395528 | 120471840 | 181 | 29 | 41164 | 1.6638 | 0.048075 | *RABL3* |
| ENSG00000144026 | 2 | 95803075 | 95841158 | 49 | 14 | 49003 | 1.6636 | 0.048093 | *ZNF514* |
| ENSG00000073008 | 19 | 45137098 | 45176850 | 95 | 22 | 40995 | 1.6632 | 0.048132 | *PVR* |
| ENSG00000147364 | 8 | 346428 | 431225 | 463 | 52 | 49006 | 1.663 | 0.048154 | *FBXO25* |
| ENSG00000188782 | 1 | 26507052 | 26539459 | 97 | 20 | 46935 | 1.6629 | 0.048164 | *CATSPER4* |
| ENSG00000249087 | 1 | 23685490 | 23708332 | 60 | 11 | 35242 | 1.6626 | 0.048194 | *C1orf213* |
| ENSG00000196748 | 6 | 35734371 | 35757329 | 89 | 15 | 31415 | 1.6626 | 0.048197 | *CLPSL2* |
| ENSG00000167895 | 17 | 76116851 | 76149049 | 139 | 32 | 48035 | 1.6615 | 0.048305 | *TMC8* |
| ENSG00000268432 | 5 | 157150003 | 157171727 | 80 | 14 | 47783 | 1.6613 | 0.048322 | *AC026407.1* |
| ENSG00000166848 | 16 | 75671684 | 75805770 | 336 | 40 | 41561 | 1.6613 | 0.048324 | *TERF2IP* |
| ENSG00000110536 | 11 | 47576982 | 47605013 | 30 | 11 | 43265 | 1.6606 | 0.048402 | *PTPMT1* |
| ENSG00000137996 | 1 | 100721763 | 100768325 | 129 | 11 | 31045 | 1.6604 | 0.048413 | *RTCA* |
| ENSG00000109255 | 4 | 56451396 | 56512865 | 228 | 27 | 47514 | 1.6604 | 0.048418 | *NMU* |
| ENSG00000198128 | 1 | 248213984 | 248234922 | 57 | 9 | 41671 | 1.6602 | 0.048442 | *OR2L3* |
| ENSG00000149187 | 11 | 47477496 | 47597121 | 163 | 29 | 42841 | 1.66 | 0.048461 | *CELF1* |
| ENSG00000235931 | 10 | 61705188 | 61730671 | 102 | 18 | 51683 | 1.6583 | 0.048627 | *C10orf40* |
| ENSG00000205517 | 19 | 11485017 | 11540018 | 117 | 19 | 34691 | 1.6582 | 0.048639 | *RGL3* |
| ENSG00000226288 | 11 | 4598021 | 4619135 | 82 | 14 | 50727 | 1.658 | 0.048658 | *OR52I2* |
| ENSG00000125966 | 20 | 33804457 | 33874801 | 143 | 22 | 43412 | 1.6579 | 0.048667 | *MMP24* |
| ENSG00000167771 | 11 | 63668693 | 63694316 | 28 | 7 | 31371 | 1.6578 | 0.048681 | *RCOR2* |
| ENSG00000163221 | 1 | 153336184 | 153358125 | 57 | 6 | 52594 | 1.6576 | 0.048694 | *S100A12* |
| ENSG00000188730 | 7 | 49803257 | 49971546 | 403 | 47 | 48397 | 1.6567 | 0.048794 | *VWC2* |
| ENSG00000268413 | 17 | 42325297 | 42365825 | 90 | 28 | 47334 | 1.6554 | 0.048924 | *AC003043.1* |
| ENSG00000165355 | 14 | 39856873 | 39911704 | 153 | 16 | 45313 | 1.655 | 0.04896 | *FBXO33* |
| ENSG00000173599 | 11 | 66605704 | 66735847 | 254 | 39 | 51912 | 1.6546 | 0.049 | *PC* |
| ENSG00000157388 | 3 | 53518683 | 53857760 | 885 | 149 | 42299 | 1.6542 | 0.049038 | *CACNA1D* |
| ENSG00000152219 | 11 | 30334598 | 30369774 | 115 | 14 | 47559 | 1.6541 | 0.04905 | *ARL14EP* |
| ENSG00000083312 | 5 | 72102139 | 72222560 | 253 | 25 | 51029 | 1.6541 | 0.049052 | *TNPO1* |
| ENSG00000148798 | 10 | 105026920 | 105060108 | 77 | 14 | 48015 | 1.654 | 0.049065 | *INA* |
| ENSG00000155363 | 1 | 113205763 | 113253368 | 106 | 19 | 52934 | 1.6536 | 0.0491 | *MOV10* |

| | | | | | | | | | |
|---|---|---|---|---|---|---|---|---|---|
| ENSG00000115956 | 2 | 68582305 | 68634585 | 332 | 23 | 39594 | 1.6533 | 0.049134 | *PLEK* |
| ENSG00000213533 | 3 | 52860235 | 52941612 | 182 | 17 | 51863 | 1.6531 | 0.049155 | *TMEM110* |
| ENSG00000168575 | 8 | 42263993 | 42407069 | 326 | 54 | 39984 | 1.6528 | 0.049184 | *SLC20A2* |
| ENSG00000268870 | 19 | 12492763 | 12561896 | 399 | 15 | 40791 | 1.6528 | 0.049189 | *CTD-3105H18.16* |
| ENSG00000255692 | 12 | 120127573 | 120148816 | 53 | 10 | 35126 | 1.6526 | 0.049209 | *RP1-127H14.3* |
| ENSG00000110697 | 11 | 67249239 | 67283734 | 40 | 16 | 44152 | 1.6525 | 0.049214 | *PITPNM1* |
| ENSG00000113212 | 5 | 140542243 | 140565957 | 87 | 12 | 44102 | 1.6524 | 0.049222 | *PCDHB7* |
| ENSG00000140451 | 15 | 65097831 | 65127867 | 120 | 20 | 40840 | 1.6522 | 0.049249 | *PIF1* |
| ENSG00000102858 | 16 | 4656494 | 4750975 | 422 | 50 | 46022 | 1.6516 | 0.049308 | *MGRN1* |
| ENSG00000155066 | 2 | 95930201 | 95967056 | 43 | 12 | 51874 | 1.6514 | 0.049329 | *PROM2* |
| ENSG00000171791 | 18 | 60780579 | 60997361 | 580 | 133 | 41875 | 1.6512 | 0.04935 | *BCL2* |
| ENSG00000125484 | 9 | 135535422 | 135580342 | 153 | 13 | 53278 | 1.6511 | 0.049363 | *GTF3C4* |
| ENSG00000170759 | 10 | 32287938 | 32355359 | 186 | 19 | 54702 | 1.6506 | 0.049407 | *KIF5B* |
| ENSG00000048462 | 16 | 12048964 | 12071925 | 98 | 15 | 33912 | 1.6502 | 0.049448 | *TNFRSF17* |
| ENSG00000115568 | 2 | 219492639 | 219534378 | 90 | 11 | 38316 | 1.6502 | 0.049451 | *ZNF142* |
| ENSG00000158092 | 3 | 136571050 | 136678665 | 299 | 12 | 51979 | 1.6499 | 0.049477 | *NCK1* |
| ENSG00000111640 | 12 | 6633093 | 6657537 | 69 | 12 | 42598 | 1.6499 | 0.049484 | *GAPDH* |
| ENSG00000213203 | 7 | 150403645 | 150431372 | 88 | 13 | 46758 | 1.6497 | 0.049505 | *GIMAP1* |
| ENSG00000176473 | 14 | 100832755 | 101006640 | 318 | 47 | 40134 | 1.6497 | 0.049505 | *WDR25* |
| ENSG00000143556 | 1 | 153420220 | 153443177 | 53 | 9 | 48571 | 1.6495 | 0.049524 | *S100A7* |
| ENSG00000186710 | 12 | 113577663 | 113607081 | 80 | 13 | 50318 | 1.6493 | 0.049547 | *CCDC42B* |
| ENSG00000181961 | 11 | 55100627 | 55121707 | 87 | 15 | 42528 | 1.6483 | 0.049645 | *OR4A16* |
| ENSG00000114200 | 3 | 165480692 | 165565260 | 325 | 34 | 42934 | 1.647 | 0.049776 | *BCHE* |
| ENSG00000160233 | 21 | 45865369 | 45888739 | 98 | 14 | 52773 | 1.647 | 0.049781 | *LRRC3* |
| ENSG00000188324 | 12 | 55678072 | 55699016 | 46 | 13 | 42648 | 1.6455 | 0.049935 | *OR6C6* |
| ENSG00000165416 | 13 | 53216844 | 53285044 | 222 | 12 | 54887 | 1.645 | 0.049982 | *SUGT1* |
| ENSG00000107929 | 10 | 845484 | 987564 | 384 | 29 | 47371 | 1.6447 | 0.050017 | *LARP4B* |
| ENSG00000213888 | 22 | 31732875 | 31754670 | 53 | 17 | 49779 | 1.6447 | 0.050019 | *AC005003.1* |
| ENSG00000164111 | 4 | 122579110 | 122628268 | 230 | 27 | 38475 | 1.6446 | 0.050025 | *ANXA5* |
| ENSG00000135973 | 2 | 105848200 | 105869924 | 55 | 12 | 45418 | 1.6441 | 0.050082 | *GPR45* |
| ENSG00000167774 | 19 | 8363167 | 8396263 | 157 | 18 | 38356 | 1.6437 | 0.05012 | *NDUFA7* |
| ENSG00000267855 | 19 | 8363490 | 8396280 | 157 | 18 | 38356 | 1.6437 | 0.05012 | *NDUFA7* |
| ENSG00000106261 | 7 | 99603204 | 99649312 | 81 | 15 | 51519 | 1.6427 | 0.050226 | *ZKSCAN1* |
| ENSG00000151498 | 11 | 134113389 | 134145749 | 83 | 15 | 49430 | 1.6425 | 0.050242 | *ACAD8* |
| ENSG00000168256 | 17 | 40153400 | 40187659 | 45 | 11 | 47281 | 1.6421 | 0.050282 | *NKIRAS2* |
| ENSG00000233927 | 19 | 8376042 | 8398224 | 112 | 17 | 46692 | 1.6421 | 0.050285 | *RPS28* |

| | | | | | | | | | |
|---|---|---|---|---|---|---|---|---|---|
| ENSG00000177614 | 1 | 230447392 | 230571475 | 506 | 71 | 44708 | 1.6411 | 0.050384 | *PGBD5* |
| ENSG00000106034 | 7 | 120618731 | 120947498 | 946 | 76 | 46163 | 1.641 | 0.050402 | *CPED1* |
| ENSG00000125246 | 13 | 100248919 | 100559387 | 1016 | 135 | 45545 | 1.6405 | 0.050446 | *CLYBL* |
| ENSG00000204788 | 9 | 69367902 | 69390597 | 1 | 1 | 42767 | 1.6399 | 0.05051 | *CR769776.1* |
| ENSG00000269430 | 21 | 45869814 | 45892078 | 78 | 13 | 52289 | 1.6391 | 0.050601 | *LRRC3DN* |
| ENSG00000179115 | 19 | 13023293 | 13054851 | 61 | 11 | 48123 | 1.639 | 0.050609 | *FARSA* |
| ENSG00000157999 | 7 | 6061007 | 6086017 | 114 | 19 | 48319 | 1.6387 | 0.050637 | *ANKRD61* |
| ENSG00000175467 | 11 | 65719160 | 65757299 | 116 | 16 | 48169 | 1.6386 | 0.050648 | *SART1* |
| ENSG00000244411 | 11 | 71228313 | 71249210 | 70 | 7 | 44488 | 1.6385 | 0.050657 | *KRTAP5-7* |
| ENSG00000165322 | 10 | 32084365 | 32227742 | 311 | 25 | 54638 | 1.6385 | 0.050662 | *ARHGAP12* |
| ENSG00000175390 | 11 | 7981798 | 8033409 | 176 | 40 | 44474 | 1.6383 | 0.050682 | *EIF3F* |
| ENSG00000165475 | 13 | 20967806 | 21109996 | 525 | 77 | 44588 | 1.6377 | 0.05074 | *CRYL1* |
| ENSG00000136720 | 2 | 128984290 | 129086151 | 350 | 57 | 41327 | 1.6374 | 0.050777 | *HS6ST1* |
| ENSG00000169689 | 17 | 79966578 | 79991983 | 50 | 12 | 47192 | 1.6373 | 0.050784 | *STRA13* |
| ENSG00000196781 | 9 | 84188598 | 84314220 | 456 | 99 | 47469 | 1.6372 | 0.05079 | *TLE1* |
| ENSG00000231738 | 12 | 85398094 | 85440055 | 51 | 13 | 48320 | 1.6368 | 0.050831 | *TSPAN19* |
| ENSG00000149554 | 11 | 125485036 | 125556150 | 201 | 20 | 51916 | 1.6362 | 0.050896 | *CHEK1* |
| ENSG00000204267 | 6 | 32779610 | 32816557 | 341 | 52 | 44179 | 1.6359 | 0.050931 | *TAP2* |
| ENSG00000122188 | 1 | 203724304 | 203755361 | 103 | 14 | 45394 | 1.6357 | 0.050951 | *LAX1* |
| ENSG00000019995 | 10 | 126620692 | 126686758 | 166 | 19 | 41351 | 1.6357 | 0.050956 | *ZRANB1* |
| ENSG00000204531 | 6 | 31122119 | 31158508 | 452 | 47 | 43851 | 1.6354 | 0.050984 | *POU5F1* |
| ENSG00000258386 | 14 | 32466108 | 32497891 | 92 | 18 | 45059 | 1.6352 | 0.051 | *RP11-187E13.2* |
| ENSG00000049192 | 5 | 64434563 | 64787747 | 1020 | 81 | 48480 | 1.6349 | 0.051031 | *ADAMTS6* |
| ENSG00000176567 | 11 | 48275413 | 48296330 | 95 | 12 | 30535 | 1.6346 | 0.05107 | *OR4X1* |
| ENSG00000157895 | 12 | 121430225 | 121464305 | 139 | 21 | 51843 | 1.6327 | 0.05127 | *C12orf43* |
| ENSG00000157510 | 5 | 148641434 | 148731365 | 232 | 52 | 44905 | 1.6322 | 0.05132 | *AFAP1L1* |
| ENSG00000111012 | 12 | 58146117 | 58172769 | 27 | 5 | 42615 | 1.632 | 0.051339 | *CYP27B1* |
| ENSG00000152611 | 5 | 35894397 | 35948881 | 197 | 23 | 46192 | 1.6319 | 0.051348 | *CAPSL* |
| ENSG00000103196 | 16 | 84843590 | 84964374 | 670 | 115 | 48273 | 1.6318 | 0.051365 | *CRISPLD2* |
| ENSG00000146147 | 6 | 53784780 | 54141078 | 886 | 91 | 40073 | 1.6317 | 0.051375 | *MLIP* |
| ENSG00000216588 | 19 | 45106940 | 45150081 | 143 | 24 | 47577 | 1.6317 | 0.051375 | *IGSF23* |
| ENSG00000131100 | 22 | 18064902 | 18121584 | 302 | 21 | 40543 | 1.6315 | 0.051393 | *ATP6V1E1* |
| ENSG00000178093 | 19 | 19613227 | 19636838 | 59 | 9 | 44333 | 1.6313 | 0.051418 | *TSSK6* |
| ENSG00000121068 | 17 | 59467257 | 59496827 | 88 | 15 | 40506 | 1.6312 | 0.05142 | *TBX2* |
| ENSG00000233198 | 9 | 140112018 | 140134090 | 55 | 15 | 54565 | 1.6308 | 0.051464 | *RNF224* |
| ENSG00000231852 | 6 | 31996042 | 32019447 | 67 | 25 | 40247 | 1.6303 | 0.051515 | *CYP21A2* |

| | | | | | | | | | |
|---|---|---|---|---|---|---|---|---|---|
| ENSG00000182749 | 1 | 26177701 | 26207744 | 49 | 10 | 36599 | 1.6301 | 0.051545 | *PAQR7* |
| ENSG00000181143 | 19 | 8949520 | 9102018 | 614 | 72 | 46732 | 1.63 | 0.051551 | *MUC16* |
| ENSG00000184349 | 5 | 106702590 | 107016596 | 811 | 142 | 45513 | 1.6299 | 0.051563 | *EFNA5* |
| ENSG00000105173 | 19 | 30292805 | 30325215 | 98 | 17 | 48258 | 1.6299 | 0.051563 | *CCNE1* |
| ENSG00000134321 | 2 | 6995937 | 7048370 | 83 | 19 | 49625 | 1.6298 | 0.051575 | *RSAD2* |
| ENSG00000266953 | 19 | 34877220 | 34910269 | 69 | 17 | 44190 | 1.6296 | 0.051596 | *RP11-618P17.4* |
| ENSG00000149636 | 20 | 35370194 | 35412221 | 46 | 24 | 47167 | 1.6294 | 0.051609 | *DSN1* |
| ENSG00000120675 | 13 | 43587339 | 43693045 | 292 | 25 | 39780 | 1.6291 | 0.051648 | *DNAJC15* |
| ENSG00000258388 | 6 | 32111622 | 32149755 | 106 | 22 | 48029 | 1.6288 | 0.051674 | *PPT2-EGFL8* |
| ENSG00000184203 | 3 | 195231221 | 195280209 | 179 | 23 | 45571 | 1.6287 | 0.051685 | *PPP1R2* |
| ENSG00000155858 | 5 | 157160703 | 157197717 | 111 | 12 | 51606 | 1.6283 | 0.051735 | *LSM11* |
| ENSG00000131495 | 5 | 140008325 | 140037370 | 77 | 10 | 53361 | 1.6281 | 0.051753 | *NDUFA2* |
| ENSG00000149532 | 11 | 61160121 | 61207503 | 52 | 9 | 38884 | 1.6271 | 0.05186 | *CPSF7* |
| ENSG00000105281 | 19 | 47268140 | 47301851 | 145 | 23 | 47883 | 1.6266 | 0.051914 | *SLC1A5* |
| ENSG00000166006 | 12 | 75423857 | 75613648 | 531 | 58 | 41676 | 1.6266 | 0.051916 | *KCNC2* |
| ENSG00000148704 | 10 | 118878032 | 118907812 | 41 | 10 | 39314 | 1.6265 | 0.051918 | *VAX1* |
| ENSG00000114098 | 3 | 137896109 | 138027231 | 132 | 38 | 49693 | 1.6265 | 0.051924 | *ARMC8* |
| ENSG00000063180 | 19 | 49131199 | 49159569 | 64 | 14 | 35608 | 1.6262 | 0.051956 | *CA11* |
| ENSG00000145741 | 5 | 72784233 | 72811460 | 45 | 15 | 42239 | 1.626 | 0.05198 | *BTF3* |
| ENSG00000237541 | 6 | 32699119 | 32724992 | 581 | 28 | 36865 | 1.6246 | 0.052124 | *HLA-DQA2* |
| ENSG00000127445 | 19 | 9935933 | 9970358 | 71 | 16 | 42500 | 1.624 | 0.052188 | *PIN1* |
| ENSG00000180539 | 9 | 139911916 | 139941234 | 76 | 15 | 53118 | 1.6238 | 0.052205 | *C9orf139* |
| ENSG00000136026 | 12 | 106621655 | 106708057 | 271 | 58 | 46166 | 1.6237 | 0.052225 | *CKAP4* |
| ENSG00000173486 | 11 | 63998475 | 64021604 | 54 | 11 | 44878 | 1.6235 | 0.052244 | *FKBP2* |
| ENSG00000136110 | 13 | 53267399 | 53323947 | 183 | 24 | 50005 | 1.6232 | 0.052274 | *LECT1* |
| ENSG00000268923 | 6 | 31963945 | 31984881 | 22 | 9 | 43717 | 1.623 | 0.052298 | *AL645922.1* |
| ENSG00000148343 | 9 | 131788900 | 131844361 | 113 | 20 | 31105 | 1.6227 | 0.052328 | *FAM73B* |
| ENSG00000261717 | 16 | 75436582 | 75508604 | 343 | 18 | 45840 | 1.6225 | 0.052347 | *RP11-77K12.1* |
| ENSG00000124194 | 20 | 42865887 | 42919013 | 151 | 16 | 30739 | 1.6217 | 0.052436 | *GDAP1L1* |
| ENSG00000135119 | 12 | 117166096 | 117301436 | 446 | 68 | 44757 | 1.6214 | 0.052463 | *RNFT2* |
| ENSG00000188807 | 1 | 9638932 | 9684935 | 65 | 13 | 43037 | 1.621 | 0.052506 | *TMEM201* |
| ENSG00000188573 | 5 | 167946121 | 167967639 | 96 | 11 | 45994 | 1.6209 | 0.05252 | *FBLL1* |
| ENSG00000137218 | 6 | 41727914 | 41764280 | 96 | 22 | 48889 | 1.6209 | 0.052521 | *FRS3* |
| ENSG00000179253 | 20 | 60282562 | 60304804 | 99 | 17 | 23857 | 1.6205 | 0.052561 | *RP11-429E11.3* |
| ENSG00000132185 | 1 | 161666762 | 161694142 | 120 | 29 | 41796 | 1.6203 | 0.052579 | *FCRLA* |
| ENSG00000251201 | 5 | 114904339 | 114971858 | 154 | 21 | 51111 | 1.6201 | 0.052603 | *TMED7-TICAM2* |

| | | | | | | | | | |
|---|---|---|---|---|---|---|---|---|---|
| ENSG00000243414 | 5 | 114904339 | 114971876 | 154 | 21 | 51111 | 1.6201 | 0.052603 | *TICAM2* |
| ENSG00000168539 | 11 | 62666151 | 62699279 | 75 | 22 | 47579 | 1.6192 | 0.0527 | *CHRM1* |
| ENSG00000205809 | 12 | 10569453 | 10604899 | 63 | 12 | 39735 | 1.6192 | 0.0527 | *KLRC2* |
| ENSG00000125995 | 20 | 34277194 | 34298906 | 58 | 11 | 46065 | 1.6191 | 0.05271 | *ROMO1* |
| ENSG00000177058 | 5 | 54911673 | 55079022 | 901 | 32 | 43569 | 1.619 | 0.052718 | *SLC38A9* |
| ENSG00000144524 | 2 | 232636381 | 232683963 | 117 | 16 | 54566 | 1.619 | 0.052727 | *COPS7B* |
| ENSG00000179813 | 13 | 43345686 | 43375683 | 102 | 14 | 51874 | 1.6188 | 0.052745 | *FAM216B* |
| ENSG00000149809 | 11 | 64869317 | 64893856 | 46 | 15 | 47326 | 1.6183 | 0.052794 | *TM7SF2* |
| ENSG00000084774 | 2 | 27430258 | 27476811 | 144 | 17 | 43154 | 1.618 | 0.052831 | *CAD* |
| ENSG00000065559 | 17 | 11914141 | 12057147 | 278 | 30 | 52129 | 1.6175 | 0.052885 | *MAP2K4* |
| ENSG00000269121 | 16 | 12052000 | 12072333 | 92 | 15 | 33336 | 1.6175 | 0.052887 | *AC007216.2* |
| ENSG00000048991 | 2 | 136279025 | 136492840 | 197 | 23 | 25716 | 1.6173 | 0.052902 | *R3HDM1* |
| ENSG00000011007 | 1 | 24059645 | 24098549 | 46 | 19 | 46056 | 1.617 | 0.052937 | *TCEB3* |
| ENSG00000243696 | 3 | 52837298 | 52879745 | 92 | 12 | 51866 | 1.6169 | 0.052945 | *RP5-966M1.6* |
| ENSG00000167536 | 17 | 27214799 | 27240089 | 26 | 9 | 46615 | 1.6166 | 0.052981 | *DHRS13* |
| ENSG00000172487 | 11 | 56117691 | 56138764 | 96 | 12 | 56092 | 1.6166 | 0.052987 | *OR8J1* |
| ENSG00000129636 | 16 | 47178298 | 47508060 | 377 | 36 | 43163 | 1.6159 | 0.053059 | *ITFG1* |
| ENSG00000185966 | 1 | 152528130 | 152549248 | 59 | 14 | 40834 | 1.6157 | 0.053084 | *LCE3E* |
| ENSG00000168803 | 15 | 43612872 | 43656096 | 79 | 11 | 38573 | 1.6156 | 0.053089 | *ADAL* |
| ENSG00000083097 | 6 | 83767385 | 83891069 | 208 | 22 | 49934 | 1.6143 | 0.053235 | *DOPEY1* |
| ENSG00000125457 | 17 | 73252309 | 73277308 | 81 | 14 | 34617 | 1.6142 | 0.053239 | *MIF4GD* |
| ENSG00000124406 | 4 | 42400390 | 42669122 | 935 | 83 | 50954 | 1.6141 | 0.053255 | *ATP8A1* |
| ENSG00000179213 | 19 | 51739603 | 51782584 | 131 | 18 | 26565 | 1.6139 | 0.053274 | *SIGLECL1* |
| ENSG00000138814 | 4 | 101934566 | 102279435 | 801 | 85 | 36578 | 1.6137 | 0.053299 | *PPP3CA* |
| ENSG00000130545 | 19 | 6453788 | 6477232 | 93 | 18 | 41923 | 1.6136 | 0.05331 | *CRB3* |
| ENSG00000135426 | 12 | 55331802 | 55388530 | 97 | 25 | 42264 | 1.6135 | 0.053321 | *TESPA1* |
| ENSG00000113272 | 5 | 157148205 | 157178456 | 102 | 17 | 48824 | 1.6133 | 0.053335 | *THG1L* |
| ENSG00000267988 | 7 | 45892684 | 45916045 | 93 | 18 | 47940 | 1.6126 | 0.053411 | *AC096582.1* |
| ENSG00000174891 | 3 | 157813644 | 158273519 | 1628 | 48 | 51600 | 1.6125 | 0.053422 | *RSRC1* |
| ENSG00000185022 | 22 | 38587889 | 38622518 | 115 | 16 | 38650 | 1.6125 | 0.053429 | *MAFF* |
| ENSG00000102699 | 13 | 24985064 | 25096948 | 705 | 36 | 50800 | 1.6121 | 0.053475 | *PARP4* |
| ENSG00000165084 | 8 | 69232957 | 69741257 | 1584 | 129 | 42162 | 1.6115 | 0.053536 | *C8orf34* |
| ENSG00000148204 | 9 | 126108449 | 126152603 | 149 | 31 | 46074 | 1.6115 | 0.053539 | *CRB2* |
| ENSG00000114209 | 3 | 167391086 | 167462727 | 113 | 9 | 31890 | 1.6113 | 0.053561 | *PDCD10* |
| ENSG00000109103 | 17 | 26863725 | 26889686 | 22 | 9 | 39028 | 1.6112 | 0.053566 | *UNC119* |
| ENSG00000144036 | 2 | 72393113 | 73063170 | 1182 | 39 | 35554 | 1.6111 | 0.053577 | *EXOC6B* |

| | | | | | | | | | |
|---|---|---|---|---|---|---|---|---|---|
| ENSG00000131771 | 17 | 37772993 | 37802879 | 76 | 14 | 31753 | 1.6106 | 0.053636 | *PPP1R1B* |
| ENSG00000124641 | 6 | 41863092 | 41898877 | 76 | 18 | 52901 | 1.61 | 0.053697 | *MED20* |
| ENSG00000138068 | 2 | 37384963 | 37433741 | 348 | 27 | 47069 | 1.6097 | 0.053732 | *SULT6B1* |
| ENSG00000117640 | 1 | 26135131 | 26169432 | 88 | 8 | 40823 | 1.6095 | 0.053758 | *MTFR1L* |
| ENSG00000120063 | 17 | 62996833 | 63062957 | 57 | 17 | 44445 | 1.6091 | 0.053796 | *GNA13* |
| ENSG00000197838 | 19 | 41584377 | 41612100 | 127 | 12 | 43386 | 1.6091 | 0.053801 | *CYP2A13* |
| ENSG00000124659 | 6 | 42702219 | 42724558 | 50 | 12 | 54206 | 1.6084 | 0.053872 | *TBCC* |
| ENSG00000179271 | 19 | 13054972 | 13078050 | 32 | 14 | 47100 | 1.6075 | 0.053969 | *GADD45GIP1* |
| ENSG00000105479 | 19 | 48789714 | 48835151 | 184 | 25 | 49468 | 1.6067 | 0.054062 | *CCDC114* |
| ENSG00000166166 | 14 | 103985521 | 104013410 | 81 | 14 | 51845 | 1.6061 | 0.054131 | *TRMT61A* |
| ENSG00000134146 | 15 | 35499546 | 35848394 | 815 | 73 | 36640 | 1.6059 | 0.054144 | *DPH6* |
| ENSG00000172489 | 11 | 56009676 | 56030698 | 66 | 9 | 56499 | 1.6059 | 0.054153 | *OR5T3* |
| ENSG00000087111 | 17 | 26870401 | 26908890 | 25 | 10 | 49597 | 1.6049 | 0.05426 | *PIGS* |
| ENSG00000132963 | 13 | 29223241 | 29263062 | 112 | 14 | 38661 | 1.6048 | 0.054268 | *POMP* |
| ENSG00000001617 | 3 | 50182478 | 50236508 | 93 | 17 | 38140 | 1.6046 | 0.054286 | *SEMA3F* |
| ENSG00000132874 | 18 | 42782960 | 43273072 | 1707 | 185 | 48594 | 1.6046 | 0.05429 | *SLC14A2* |
| ENSG00000114631 | 3 | 127338024 | 127401652 | 77 | 21 | 55165 | 1.604 | 0.05436 | *PODXL2* |
| ENSG00000142973 | 1 | 47213510 | 47295085 | 221 | 37 | 40030 | 1.6038 | 0.054376 | *CYP4B1* |
| ENSG00000011590 | 19 | 36185429 | 36217940 | 83 | 20 | 47858 | 1.6037 | 0.054395 | *ZBTB32* |
| ENSG00000107185 | 9 | 35739203 | 35768572 | 42 | 13 | 52627 | 1.6036 | 0.054403 | *RGP1* |
| ENSG00000174177 | 16 | 88762871 | 88791794 | 181 | 37 | 47263 | 1.6029 | 0.054473 | *CTU2* |
| ENSG00000132604 | 16 | 69379464 | 69452474 | 142 | 32 | 50833 | 1.6028 | 0.054489 | *TERF2* |
| ENSG00000055955 | 3 | 52836991 | 52875495 | 84 | 11 | 52379 | 1.6022 | 0.05456 | *ITIH4* |
| ENSG00000136158 | 13 | 80900111 | 80925086 | 49 | 17 | 39882 | 1.6019 | 0.054585 | *SPRY2* |
| ENSG00000131653 | 16 | 2195699 | 2238130 | 62 | 28 | 41508 | 1.6019 | 0.05459 | *TRAF7* |
| ENSG00000167515 | 16 | 88912628 | 88939094 | 138 | 25 | 41036 | 1.6018 | 0.054597 | *TRAPPC2L* |
| ENSG00000064300 | 17 | 47562655 | 47602379 | 118 | 38 | 43738 | 1.6016 | 0.054627 | *NGFR* |
| ENSG00000112640 | 6 | 42942237 | 42990080 | 160 | 25 | 37025 | 1.6015 | 0.054631 | *PPP2R5D* |
| ENSG00000173662 | 1 | 6605241 | 6649817 | 126 | 14 | 33752 | 1.601 | 0.054686 | *TAS1R1* |
| ENSG00000249992 | 3 | 45255958 | 45277770 | 96 | 16 | 38864 | 1.6006 | 0.054736 | *TMEM158* |
| ENSG00000197191 | 9 | 140109087 | 140130763 | 54 | 14 | 56206 | 1.6004 | 0.054754 | *C9orf169* |
| ENSG00000132837 | 5 | 78283438 | 78541861 | 750 | 65 | 36606 | 1.6002 | 0.054772 | *DMGDH* |
| ENSG00000167775 | 19 | 8357011 | 8383240 | 86 | 10 | 29559 | 1.5988 | 0.05493 | *CD320* |
| ENSG00000183571 | 15 | 99501459 | 99561024 | 274 | 42 | 39424 | 1.5987 | 0.054945 | *PGPEP1L* |
| ENSG00000256530 | 15 | 76020311 | 76041568 | 68 | 8 | 48786 | 1.5983 | 0.054991 | *AC019294.1* |
| ENSG00000134909 | 11 | 128824955 | 129159219 | 924 | 67 | 51162 | 1.5982 | 0.054999 | *ARHGAP32* |

| | | | | | | | | | |
|---|---|---|---|---|---|---|---|---|---|
| ENSG00000258713 | 20 | 2785633 | 2806479 | 44 | 15 | 40066 | 1.5982 | 0.055002 | *C20orf141* |
| ENSG00000125618 | 2 | 113963574 | 114046527 | 327 | 34 | 48881 | 1.598 | 0.055019 | *PAX8* |
| ENSG00000062650 | 10 | 88185013 | 88291572 | 276 | 34 | 43884 | 1.5979 | 0.055029 | *WAPAL* |
| ENSG00000162711 | 1 | 247569458 | 247622410 | 297 | 59 | 47626 | 1.5978 | 0.055048 | *NLRP3* |
| ENSG00000167766 | 19 | 53087313 | 53203749 | 667 | 78 | 49252 | 1.5975 | 0.055072 | *ZNF83* |
| ENSG00000179965 | 16 | 30408618 | 30450920 | 47 | 18 | 38150 | 1.5974 | 0.055084 | *ZNF771* |
| ENSG00000163141 | 1 | 150999046 | 151030076 | 78 | 20 | 50222 | 1.5969 | 0.055148 | *BNIPL* |
| ENSG00000104756 | 8 | 25275366 | 25325992 | 154 | 10 | 49559 | 1.5968 | 0.055153 | *KCTD9* |
| ENSG00000176884 | 9 | 140022842 | 140073207 | 115 | 31 | 38259 | 1.5967 | 0.055162 | *GRIN1* |
| ENSG00000162727 | 1 | 248298450 | 248319388 | 55 | 11 | 35648 | 1.5963 | 0.055212 | *OR2M5* |
| ENSG00000141012 | 16 | 88870142 | 88933378 | 404 | 45 | 48750 | 1.596 | 0.055242 | *GALNS* |
| ENSG00000259571 | 11 | 121976062 | 121996923 | 66 | 16 | 51243 | 1.596 | 0.055244 | *BLID* |
| ENSG00000170037 | 17 | 7825419 | 7863236 | 113 | 11 | 47270 | 1.5956 | 0.055293 | *CNTROB* |
| ENSG00000159167 | 8 | 23689428 | 23722320 | 102 | 17 | 49549 | 1.5936 | 0.055511 | *STC1* |
| ENSG00000106804 | 9 | 123704616 | 123822554 | 304 | 33 | 42340 | 1.593 | 0.055579 | *C5* |
| ENSG00000268556 | 2 | 105857759 | 105879601 | 43 | 11 | 37794 | 1.5923 | 0.055656 | *AC012360.1* |
| ENSG00000081177 | 14 | 69648228 | 69719075 | 160 | 23 | 34662 | 1.5922 | 0.05567 | *EXD2* |
| ENSG00000003137 | 2 | 72346367 | 72385167 | 90 | 22 | 38045 | 1.5922 | 0.055671 | *CYP26B1* |
| ENSG00000066117 | 12 | 50468755 | 50504495 | 51 | 14 | 37674 | 1.5921 | 0.055686 | *SMARCD1* |
| ENSG00000137965 | 1 | 79105481 | 79139763 | 82 | 16 | 52111 | 1.5919 | 0.055699 | *IFI44* |
| ENSG00000124568 | 6 | 25773125 | 25842287 | 235 | 26 | 36153 | 1.5913 | 0.055769 | *SLC17A1* |
| ENSG00000152256 | 2 | 173410101 | 173499823 | 323 | 28 | 49944 | 1.5912 | 0.055781 | *PDK1* |
| ENSG00000116032 | 19 | 990418 | 1019731 | 253 | 43 | 50662 | 1.5912 | 0.055785 | *GRIN3B* |
| ENSG00000134508 | 18 | 20704528 | 20850431 | 328 | 55 | 43021 | 1.5908 | 0.055832 | *CABLES1* |
| ENSG00000182578 | 5 | 149422854 | 149502935 | 257 | 59 | 41340 | 1.59 | 0.055917 | *CSF1R* |
| ENSG00000185532 | 10 | 52740945 | 54068110 | 4881 | 332 | 42836 | 1.5895 | 0.055976 | *PRKG1* |
| ENSG00000137474 | 11 | 76829310 | 76936284 | 431 | 61 | 41650 | 1.5893 | 0.055995 | *MYO7A* |
| ENSG00000146067 | 5 | 176936789 | 176991542 | 75 | 14 | 30601 | 1.5889 | 0.056038 | *FAM193B* |
| ENSG00000176986 | 10 | 75494120 | 75541919 | 75 | 12 | 42167 | 1.5888 | 0.056052 | *SEC24C* |
| ENSG00000188064 | 22 | 46306242 | 46383009 | 369 | 65 | 43809 | 1.5887 | 0.056065 | *WNT7B* |
| ENSG00000188389 | 2 | 242782033 | 242811060 | 78 | 15 | 50432 | 1.5882 | 0.05612 | *PDCD1* |
| ENSG00000086506 | 16 | 220452 | 241180 | 47 | 14 | 47897 | 1.5878 | 0.056165 | *HBQ1* |
| ENSG00000160216 | 21 | 45275067 | 45416417 | 390 | 50 | 45282 | 1.5865 | 0.056315 | *AGPAT3* |
| ENSG00000179262 | 19 | 13046669 | 13074456 | 40 | 15 | 47702 | 1.5861 | 0.05636 | *RAD23A* |
| ENSG00000180658 | 6 | 132011520 | 132032635 | 37 | 9 | 31631 | 1.586 | 0.05637 | *OR2A4* |
| ENSG00000198373 | 16 | 69786209 | 69985644 | 555 | 48 | 44580 | 1.5857 | 0.056402 | *WWP2* |

| | | | | | | | | | |
|---|---|---|---|---|---|---|---|---|---|
| ENSG00000114654 | 3 | 128710472 | 128769585 | 85 | 24 | 44547 | 1.5851 | 0.056469 | *EFCC1* |
| ENSG00000242372 | 20 | 33856714 | 33882788 | 71 | 10 | 44669 | 1.5849 | 0.056491 | *EIF6* |
| ENSG00000109606 | 4 | 24509064 | 24596173 | 251 | 36 | 45115 | 1.5847 | 0.056518 | *DHX15* |
| ENSG00000153832 | 2 | 230777018 | 230887825 | 194 | 28 | 48948 | 1.5839 | 0.056609 | *FBXO36* |
| ENSG00000176595 | 8 | 1912044 | 1965102 | 242 | 44 | 44056 | 1.5837 | 0.056627 | *KBTBD11* |
| ENSG00000184363 | 11 | 382614 | 414908 | 127 | 17 | 35475 | 1.5835 | 0.05665 | *PKP3* |
| ENSG00000006114 | 17 | 35864900 | 35979544 | 210 | 35 | 43335 | 1.5834 | 0.056665 | *SYNRG* |
| ENSG00000081692 | 1 | 227908126 | 227933112 | 52 | 14 | 50473 | 1.5831 | 0.056705 | *JMJD4* |
| ENSG00000187147 | 1 | 44860866 | 45127396 | 743 | 137 | 44073 | 1.5827 | 0.05674 | *RNF220* |
| ENSG00000170369 | 20 | 23794406 | 23817368 | 163 | 25 | 46940 | 1.5827 | 0.056741 | *CST2* |
| ENSG00000115523 | 2 | 85902298 | 85935977 | 140 | 15 | 45430 | 1.582 | 0.056822 | *GNLY* |
| ENSG00000149084 | 11 | 43567986 | 43888167 | 854 | 73 | 50336 | 1.582 | 0.056826 | *HSD17B12* |
| ENSG00000009830 | 14 | 77731299 | 77797227 | 235 | 40 | 50853 | 1.5819 | 0.056831 | *POMT2* |
| ENSG00000163636 | 3 | 63986225 | 64019658 | 122 | 25 | 46536 | 1.5815 | 0.05688 | *PSMD6* |
| ENSG00000198646 | 20 | 33274722 | 33423452 | 281 | 23 | 52359 | 1.5815 | 0.056884 | *NCOA6* |
| ENSG00000137776 | 15 | 59161244 | 59235852 | 106 | 17 | 34641 | 1.58 | 0.057049 | *SLTM* |
| ENSG00000162889 | 1 | 206848289 | 206917628 | 209 | 31 | 41646 | 1.5798 | 0.057075 | *MAPKAPK2* |
| ENSG00000140265 | 15 | 43640370 | 43673223 | 45 | 8 | 38148 | 1.5781 | 0.057276 | *ZSCAN29* |
| ENSG00000157741 | 7 | 138905102 | 139002982 | 159 | 22 | 39947 | 1.5776 | 0.057329 | *UBN2* |
| ENSG00000085733 | 11 | 70234510 | 70292690 | 118 | 16 | 50335 | 1.5772 | 0.057377 | *CTTN* |
| ENSG00000256632 | 12 | 133254635 | 133301498 | 155 | 22 | 49001 | 1.5771 | 0.05739 | *RP13-672B3.2* |
| ENSG00000134014 | 8 | 27937190 | 28058673 | 314 | 32 | 41586 | 1.5767 | 0.057428 | *ELP3* |
| ENSG00000115204 | 2 | 27522360 | 27558547 | 46 | 13 | 40710 | 1.5761 | 0.0575 | *MPV17* |
| ENSG00000136270 | 7 | 45129699 | 45161646 | 146 | 23 | 41901 | 1.5752 | 0.057604 | *TBRG4* |
| ENSG00000077152 | 1 | 202290785 | 202321108 | 44 | 10 | 41367 | 1.5752 | 0.057608 | *UBE2T* |
| ENSG00000232268 | 11 | 4605037 | 4626243 | 86 | 16 | 51975 | 1.5752 | 0.057608 | *OR52I1* |
| ENSG00000160460 | 19 | 40962148 | 41092370 | 344 | 63 | 44762 | 1.575 | 0.05763 | *SPTBN4* |
| ENSG00000132589 | 17 | 27196353 | 27234697 | 57 | 14 | 42637 | 1.5749 | 0.057634 | *FLOT2* |
| ENSG00000137642 | 11 | 121312912 | 121514402 | 447 | 80 | 40055 | 1.5749 | 0.057635 | *SORL1* |
| ENSG00000153714 | 9 | 12765020 | 12832130 | 211 | 51 | 45147 | 1.5747 | 0.057668 | *LURAP1L* |
| ENSG00000143363 | 1 | 150970896 | 151018189 | 140 | 32 | 46636 | 1.5745 | 0.057687 | *PRUNE* |
| ENSG00000117461 | 1 | 46495812 | 46652160 | 300 | 21 | 51720 | 1.5743 | 0.057709 | *PIK3R3* |
| ENSG00000188786 | 1 | 38265239 | 38335292 | 148 | 23 | 42664 | 1.5742 | 0.057716 | *MTF1* |
| ENSG00000154655 | 18 | 5944705 | 6425236 | 1341 | 155 | 43035 | 1.5741 | 0.057733 | *L3MBTL4* |
| ENSG00000075711 | 3 | 196759431 | 197036171 | 835 | 57 | 47278 | 1.5738 | 0.057767 | *DLG1* |
| ENSG00000146373 | 6 | 125273691 | 125423779 | 328 | 57 | 45281 | 1.573 | 0.057854 | *RNF217* |

| | | | | | | | | | |
|---|---|---|---|---|---|---|---|---|---|
| ENSG00000112425 | 6 | 145812719 | 146067160 | 847 | 34 | 53940 | 1.573 | 0.057862 | *EPM2A* |
| ENSG00000112175 | 6 | 55608443 | 55750362 | 334 | 62 | 50431 | 1.5726 | 0.057909 | *BMP5* |
| ENSG00000058272 | 12 | 80157343 | 80339240 | 239 | 21 | 24357 | 1.5724 | 0.05793 | *PPP1R12A* |
| ENSG00000164898 | 7 | 139014203 | 139041065 | 98 | 8 | 31173 | 1.5722 | 0.05795 | *C7orf55* |
| ENSG00000133574 | 7 | 150254365 | 150281041 | 153 | 16 | 45242 | 1.5722 | 0.05795 | *GIMAP4* |
| ENSG00000108342 | 17 | 38161614 | 38184066 | 90 | 10 | 47395 | 1.5722 | 0.057954 | *CSF3* |
| ENSG00000198218 | 3 | 49057140 | 49141796 | 96 | 21 | 41165 | 1.5719 | 0.057989 | *QRICH1* |
| ENSG00000142515 | 19 | 51348171 | 51374020 | 191 | 34 | 48892 | 1.5717 | 0.058016 | *KLK3* |
| ENSG00000214655 | 10 | 75535340 | 75571551 | 39 | 8 | 42042 | 1.5714 | 0.058048 | *ZSWIM8* |
| ENSG00000092201 | 14 | 21809631 | 21862425 | 128 | 17 | 40521 | 1.5711 | 0.058078 | *SUPT16H* |
| ENSG00000162976 | 2 | 11285324 | 11329000 | 187 | 29 | 44684 | 1.5709 | 0.058101 | *PQLC3* |
| ENSG00000131620 | 11 | 69914408 | 70045634 | 570 | 88 | 52781 | 1.5709 | 0.058101 | *ANO1* |
| ENSG00000137692 | 11 | 102922805 | 102972944 | 88 | 17 | 52104 | 1.5709 | 0.058102 | *DCUN1D5* |
| ENSG00000187848 | 12 | 133185366 | 133208972 | 105 | 14 | 55883 | 1.5708 | 0.058115 | *P2RX2* |
| ENSG00000149269 | 11 | 77022752 | 77195680 | 431 | 35 | 45048 | 1.5706 | 0.058138 | *PAK1* |
| ENSG00000178343 | 4 | 42389856 | 42414504 | 95 | 23 | 50629 | 1.5706 | 0.05814 | *SHISA3* |
| ENSG00000133710 | 5 | 147395246 | 147526852 | 653 | 38 | 33033 | 1.5706 | 0.05814 | *SPINK5* |
| ENSG00000205927 | 21 | 34388153 | 34411504 | 66 | 22 | 48978 | 1.57 | 0.058205 | *OLIG2* |
| ENSG00000023171 | 11 | 123386344 | 123508482 | 418 | 84 | 45993 | 1.57 | 0.058206 | *GRAMD1B* |
| ENSG00000204502 | 1 | 13349819 | 13379057 | 9 | 4 | 42657 | 1.5696 | 0.058256 | *PRAMEF5* |
| ENSG00000101194 | 20 | 61574052 | 61609949 | 146 | 36 | 46734 | 1.5695 | 0.05827 | *SLC17A9* |
| ENSG00000223474 | 1 | 26135212 | 26157288 | 57 | 7 | 36949 | 1.5692 | 0.058305 | *AL020996.1* |
| ENSG00000139567 | 12 | 52290692 | 52327145 | 133 | 29 | 48214 | 1.5691 | 0.058314 | *ACVRL1* |
| ENSG00000198824 | 13 | 115069988 | 115102796 | 78 | 11 | 46739 | 1.5689 | 0.058331 | *CHAMP1* |
| ENSG00000241404 | 6 | 32122360 | 32146058 | 71 | 19 | 47635 | 1.5688 | 0.058344 | *EGFL8* |
| ENSG00000272916 | 10 | 75546272 | 75581409 | 47 | 8 | 39248 | 1.5685 | 0.058383 | *RP11-574K11.31* |
| ENSG00000204614 | 6 | 30093885 | 30126512 | 315 | 20 | 46997 | 1.5666 | 0.058603 | *TRIM40* |
| ENSG00000137809 | 15 | 68584050 | 68734501 | 552 | 81 | 43368 | 1.5663 | 0.058644 | *ITGA11* |
| ENSG00000269169 | 19 | 989601 | 1009952 | 171 | 31 | 48812 | 1.5661 | 0.058657 | *AC004528.1* |
| ENSG00000011132 | 19 | 3740817 | 3771697 | 130 | 31 | 52432 | 1.5656 | 0.058723 | *APBA3* |
| ENSG00000150433 | 11 | 124956398 | 124991659 | 172 | 32 | 48905 | 1.5655 | 0.058732 | *TMEM218* |
| ENSG00000101444 | 20 | 32858074 | 32909608 | 73 | 9 | 33428 | 1.5655 | 0.058738 | *AHCY* |
| ENSG00000162302 | 11 | 64116620 | 64149687 | 93 | 10 | 38439 | 1.5654 | 0.058742 | *RPS6KA4* |
| ENSG00000244734 | 11 | 5236694 | 5260625 | 95 | 21 | 42843 | 1.5654 | 0.058748 | *HBB* |
| ENSG00000213760 | 6 | 31502239 | 31526204 | 165 | 31 | 48709 | 1.5644 | 0.058859 | *ATP6V1G2* |
| ENSG00000144802 | 3 | 101536835 | 101589866 | 85 | 17 | 46896 | 1.5644 | 0.058865 | *NFKBIZ* |

| | | | | | | | | | |
|---|---|---|---|---|---|---|---|---|---|
| ENSG00000171067 | 11 | 68018803 | 68049469 | 86 | 14 | 47409 | 1.5641 | 0.058899 | *C11orf24* |
| ENSG00000125998 | 20 | 33863534 | 33890204 | 65 | 10 | 42960 | 1.564 | 0.058903 | *FAM83C* |
| ENSG00000218537 | 22 | 24226613 | 24251117 | 154 | 23 | 44879 | 1.564 | 0.058912 | *AP000350.4* |
| ENSG00000177084 | 12 | 133190348 | 133273951 | 288 | 19 | 55124 | 1.5638 | 0.058933 | *POLE* |
| ENSG00000148408 | 9 | 140762241 | 141029076 | 527 | 61 | 43671 | 1.5632 | 0.059003 | *CACNA1B* |
| ENSG00000175279 | 1 | 10480159 | 10522210 | 130 | 15 | 40818 | 1.5627 | 0.059059 | *APITD1* |
| ENSG00000251503 | 1 | 10480159 | 10522210 | 130 | 15 | 40818 | 1.5627 | 0.059059 | *APITD1-CORT* |
| ENSG00000144028 | 2 | 96930074 | 96981297 | 59 | 13 | 36907 | 1.562 | 0.059144 | *SNRNP200* |
| ENSG00000161714 | 17 | 43176335 | 43220721 | 117 | 30 | 50487 | 1.5619 | 0.059151 | *PLCD3* |
| ENSG00000107960 | 10 | 105632300 | 105687963 | 161 | 17 | 44260 | 1.5619 | 0.05916 | *OBFC1* |
| ENSG00000197364 | 1 | 153399534 | 153422425 | 58 | 11 | 46784 | 1.5618 | 0.059171 | *S100A7L2* |
| ENSG00000162337 | 11 | 68070077 | 68226743 | 416 | 59 | 43055 | 1.5606 | 0.059305 | *LRP5* |
| ENSG00000174611 | 3 | 134311980 | 134380478 | 174 | 12 | 53698 | 1.5601 | 0.059371 | *KY* |
| ENSG00000171431 | 17 | 39022193 | 39051479 | 89 | 18 | 41011 | 1.5589 | 0.059508 | *KRT20* |
| ENSG00000133561 | 7 | 150312463 | 150339473 | 78 | 14 | 54375 | 1.5589 | 0.059509 | *GIMAP6* |
| ENSG00000166275 | 10 | 104603980 | 104634718 | 94 | 15 | 49343 | 1.5588 | 0.059521 | *C10orf32* |
| ENSG00000123545 | 6 | 97327189 | 97355757 | 113 | 17 | 53480 | 1.5583 | 0.059581 | *NDUFAF4* |
| ENSG00000168280 | 2 | 149622819 | 149893273 | 275 | 24 | 33681 | 1.5582 | 0.05959 | *KIF5C* |
| ENSG00000115738 | 2 | 8808975 | 8834583 | 63 | 18 | 38344 | 1.5582 | 0.059595 | *ID2* |
| ENSG00000064763 | 12 | 29292036 | 29503913 | 737 | 61 | 48860 | 1.5575 | 0.059672 | *FAR2* |
| ENSG00000166507 | 10 | 75551669 | 75581589 | 44 | 8 | 39007 | 1.5573 | 0.059703 | *NDST2* |
| ENSG00000176894 | 12 | 133254192 | 133307276 | 176 | 25 | 49176 | 1.5564 | 0.05981 | *PXMP2* |
| ENSG00000241690 | 20 | 2785614 | 2808712 | 50 | 18 | 41178 | 1.5563 | 0.059817 | *TMEM239* |
| ENSG00000110917 | 12 | 121114672 | 121149667 | 115 | 25 | 45658 | 1.5554 | 0.059925 | *MLEC* |
| ENSG00000174276 | 11 | 64873875 | 64895170 | 39 | 12 | 48203 | 1.5551 | 0.059958 | *ZNHIT2* |
| ENSG00000164023 | 4 | 108735719 | 108846203 | 249 | 51 | 45304 | 1.5548 | 0.059993 | *SGMS2* |
| ENSG00000066777 | 8 | 68075747 | 68265912 | 269 | 32 | 43704 | 1.5546 | 0.060021 | *ARFGEF1* |
| ENSG00000258973 | 14 | 24791516 | 24816588 | 109 | 18 | 33315 | 1.5538 | 0.060113 | *RP11-934B9.3* |
| ENSG00000008988 | 8 | 56969854 | 56997069 | 108 | 13 | 46849 | 1.5534 | 0.060163 | *RPS20* |
| ENSG00000069011 | 5 | 134353425 | 134380503 | 85 | 11 | 35323 | 1.553 | 0.06021 | *PITX1* |
| ENSG00000135596 | 6 | 109755265 | 109797171 | 112 | 23 | 37948 | 1.5524 | 0.060285 | *MICAL1* |
| ENSG00000185909 | 3 | 49199044 | 49223917 | 37 | 7 | 49970 | 1.5524 | 0.060286 | *KLHDC8B* |
| ENSG00000171612 | 1 | 9589541 | 9655237 | 107 | 16 | 44746 | 1.5523 | 0.060292 | *SLC25A33* |
| ENSG00000172531 | 11 | 67155654 | 67198654 | 65 | 16 | 45416 | 1.5522 | 0.060312 | *PPP1CA* |
| ENSG00000070610 | 9 | 35726863 | 35759983 | 47 | 16 | 54077 | 1.5521 | 0.060315 | *GBA2* |
| ENSG00000135047 | 9 | 90330434 | 90356308 | 97 | 14 | 34072 | 1.5514 | 0.060409 | *CTSL* |

| | | | | | | | | | |
|---|---|---|---|---|---|---|---|---|---|
| ENSG00000107872 | 10 | 104168946 | 104192893 | 33 | 11 | 51847 | 1.5512 | 0.060424 | *FBXL15* |
| ENSG00000163348 | 1 | 154919502 | 154946329 | 28 | 11 | 46987 | 1.5511 | 0.060435 | *PYGO2* |
| ENSG00000180304 | 15 | 64969772 | 65005480 | 64 | 15 | 38540 | 1.5511 | 0.060435 | *OAZ2* |
| ENSG00000173588 | 12 | 94690225 | 94863764 | 372 | 47 | 53856 | 1.5511 | 0.060443 | *CCDC41* |
| ENSG00000129988 | 20 | 36964759 | 37015665 | 219 | 37 | 44971 | 1.5508 | 0.060473 | *LBP* |
| ENSG00000117569 | 1 | 97177221 | 97299294 | 311 | 30 | 43984 | 1.55 | 0.060571 | *PTBP2* |
| ENSG00000165810 | 5 | 180457225 | 180498523 | 194 | 29 | 41539 | 1.5496 | 0.060616 | *BTNL9* |
| ENSG00000169609 | 15 | 83647193 | 83690393 | 112 | 26 | 49802 | 1.5493 | 0.060659 | *C15orf40* |
| ENSG00000242110 | 5 | 33976283 | 34018220 | 106 | 18 | 36877 | 1.549 | 0.060696 | *AMACR* |
| ENSG00000165494 | 11 | 82858030 | 82908493 | 107 | 16 | 40722 | 1.5487 | 0.060726 | *PCF11* |
| ENSG00000175634 | 11 | 67185931 | 67212872 | 40 | 11 | 40124 | 1.5484 | 0.060767 | *RPS6KB2* |
| ENSG00000103253 | 16 | 766936 | 795525 | 83 | 15 | 32476 | 1.5484 | 0.060768 | *HAGHL* |
| ENSG00000081248 | 1 | 200998642 | 201091694 | 322 | 64 | 35950 | 1.5483 | 0.060773 | *CACNA1S* |
| ENSG00000166927 | 11 | 60135955 | 60173424 | 102 | 13 | 54191 | 1.5483 | 0.060774 | *MS4A7* |
| ENSG00000125450 | 17 | 73191754 | 73241853 | 99 | 14 | 48177 | 1.5482 | 0.060793 | *NUP85* |
| ENSG00000177453 | 5 | 43182173 | 43290952 | 310 | 47 | 46752 | 1.548 | 0.060811 | *NIM1K* |
| ENSG00000242252 | 1 | 156201753 | 156223112 | 48 | 16 | 50592 | 1.5478 | 0.060841 | *BGLAP* |
| ENSG00000185670 | 11 | 62505791 | 62531660 | 44 | 8 | 43262 | 1.5471 | 0.060919 | *ZBTB3* |
| ENSG00000122966 | 12 | 120113595 | 120325095 | 598 | 65 | 46787 | 1.5471 | 0.06092 | *CIT* |
| ENSG00000108176 | 10 | 69546427 | 69607924 | 164 | 27 | 36833 | 1.5471 | 0.060923 | *DNAJC12* |
| ENSG00000185187 | 11 | 395716 | 427455 | 90 | 21 | 40091 | 1.5465 | 0.060992 | *SIGIRR* |
| ENSG00000144029 | 2 | 95742952 | 95825179 | 82 | 20 | 48915 | 1.5461 | 0.061041 | *MRPS5* |
| ENSG00000168802 | 16 | 69141913 | 69176487 | 64 | 20 | 54560 | 1.5458 | 0.061082 | *CHTF8* |
| ENSG00000184381 | 22 | 38497502 | 38611697 | 466 | 36 | 47936 | 1.5457 | 0.061086 | *PLA2G6* |
| ENSG00000164054 | 3 | 48499197 | 48552259 | 80 | 16 | 34983 | 1.5457 | 0.06109 | *SHISA5* |
| ENSG00000214021 | 3 | 9839770 | 9906822 | 212 | 31 | 40981 | 1.5456 | 0.061104 | *TTLL3* |
| ENSG00000185721 | 22 | 31785509 | 31934726 | 423 | 41 | 48625 | 1.5456 | 0.061104 | *DRG1* |
| ENSG00000064651 | 5 | 127409458 | 127535380 | 221 | 21 | 41421 | 1.5452 | 0.061153 | *SLC12A2* |
| ENSG00000169908 | 3 | 149076809 | 149105652 | 103 | 21 | 55286 | 1.5452 | 0.061154 | *TM4SF1* |
| ENSG00000167186 | 16 | 19068921 | 19101417 | 99 | 14 | 32862 | 1.545 | 0.061173 | *COQ7* |
| ENSG00000173511 | 11 | 63992010 | 64016259 | 59 | 14 | 45913 | 1.5442 | 0.061268 | *VEGFB* |
| ENSG00000168234 | 18 | 21562737 | 21725574 | 428 | 77 | 34257 | 1.5441 | 0.061279 | *TTC39C* |
| ENSG00000125414 | 17 | 10414465 | 10463274 | 122 | 12 | 37566 | 1.5438 | 0.061314 | *MYH2* |
| ENSG00000145740 | 5 | 68379473 | 68436896 | 142 | 17 | 45393 | 1.5435 | 0.061358 | *SLC30A5* |
| ENSG00000181552 | 14 | 21226586 | 21249107 | 176 | 26 | 33635 | 1.5434 | 0.061361 | *EDDM3B* |
| ENSG00000006611 | 11 | 17505442 | 17575963 | 262 | 47 | 40775 | 1.5431 | 0.061406 | *USH1C* |

| | | | | | | | | | |
|---|---|---|---|---|---|---|---|---|---|
| ENSG00000186207 | 1 | 152473320 | 152494653 | 65 | 10 | 36678 | 1.543 | 0.06141 | *LCE5A* |
| ENSG00000142224 | 1 | 206962215 | 207026324 | 185 | 31 | 48895 | 1.5428 | 0.061439 | *IL19* |
| ENSG00000165661 | 9 | 139088179 | 139147687 | 218 | 44 | 48291 | 1.5425 | 0.061477 | *QSOX2* |
| ENSG00000119927 | 10 | 113899624 | 113985135 | 222 | 35 | 44479 | 1.5424 | 0.061484 | *GPAM* |
| ENSG00000122406 | 1 | 93287582 | 93317481 | 59 | 8 | 46183 | 1.5423 | 0.061502 | *RPL5* |
| ENSG00000204323 | 17 | 73619514 | 73647484 | 50 | 16 | 47698 | 1.5413 | 0.061622 | *SMIM5* |
| ENSG00000166741 | 11 | 114118509 | 114194007 | 178 | 43 | 48284 | 1.541 | 0.061661 | *NNMT* |
| ENSG00000175514 | 11 | 67208772 | 67230200 | 36 | 11 | 36952 | 1.5407 | 0.06169 | *GPR152* |
| ENSG00000167562 | 19 | 53049075 | 53100427 | 293 | 44 | 44705 | 1.5398 | 0.061801 | *ZNF701* |
| ENSG00000164465 | 6 | 117764980 | 117901021 | 508 | 52 | 49506 | 1.539 | 0.061905 | *DCBLD1* |
| ENSG00000011465 | 12 | 91529025 | 91586900 | 69 | 16 | 36107 | 1.5382 | 0.061996 | *DCN* |
| ENSG00000198877 | 11 | 55530914 | 55551858 | 120 | 10 | 50163 | 1.5381 | 0.062011 | *OR5D13* |
| ENSG00000102893 | 16 | 47485034 | 47745434 | 244 | 25 | 42436 | 1.5377 | 0.062057 | *PHKB* |
| ENSG00000112659 | 6 | 43139913 | 43202325 | 156 | 26 | 42877 | 1.5375 | 0.062091 | *CUL9* |
| ENSG00000175920 | 4 | 3455033 | 3513200 | 401 | 55 | 48395 | 1.5373 | 0.062114 | *DOK7* |
| ENSG00000163888 | 3 | 183967001 | 183989251 | 54 | 11 | 53196 | 1.537 | 0.062147 | *CAMK2N2* |
| ENSG00000155100 | 8 | 92072424 | 92109323 | 61 | 9 | 47058 | 1.537 | 0.062152 | *OTUD6B* |
| ENSG00000142453 | 19 | 10972189 | 11043453 | 131 | 21 | 53952 | 1.5365 | 0.062212 | *CARM1* |
| ENSG00000180488 | 1 | 78235309 | 78354106 | 237 | 42 | 39001 | 1.5362 | 0.062248 | *FAM73A* |
| ENSG00000141314 | 17 | 30583195 | 30661680 | 242 | 36 | 40412 | 1.5362 | 0.062249 | *RHBDL3* |
| ENSG00000141560 | 17 | 80664559 | 80698204 | 193 | 22 | 44438 | 1.5357 | 0.062302 | *FN3KRP* |
| ENSG00000163357 | 1 | 154996300 | 155033406 | 62 | 17 | 41383 | 1.5357 | 0.062307 | *DCST1* |
| ENSG00000168671 | 5 | 36025119 | 36081460 | 198 | 31 | 39176 | 1.5354 | 0.06234 | *UGT3A2* |
| ENSG00000040487 | 1 | 19628820 | 19665794 | 182 | 27 | 52947 | 1.5353 | 0.062359 | *PQLC2* |
| ENSG00000244731 | 6 | 31939801 | 31980458 | 63 | 21 | 38049 | 1.5346 | 0.062435 | *C4A* |
| ENSG00000107175 | 9 | 35722332 | 35747001 | 33 | 14 | 52956 | 1.5345 | 0.062453 | *CREB3* |
| ENSG00000189067 | 16 | 11631853 | 11740237 | 503 | 74 | 45156 | 1.5342 | 0.062492 | *LITAF* |
| ENSG00000138684 | 4 | 123523783 | 123552224 | 85 | 15 | 42631 | 1.533 | 0.062639 | *IL21* |
| ENSG00000088298 | 20 | 33693167 | 33875928 | 489 | 48 | 43481 | 1.532 | 0.062756 | *EDEM2* |
| ENSG00000112149 | 6 | 14107872 | 14147149 | 123 | 27 | 47144 | 1.5319 | 0.062768 | *CD83* |
| ENSG00000183675 | 10 | 48726874 | 48837966 | 7 | 4 | 46511 | 1.5319 | 0.062779 | *PTPN20B* |
| ENSG00000096433 | 6 | 33578142 | 33674351 | 475 | 64 | 43246 | 1.5316 | 0.062815 | *ITPR3* |
| ENSG00000189143 | 7 | 73203872 | 73257014 | 137 | 11 | 28858 | 1.5315 | 0.062817 | *CLDN4* |
| ENSG00000255054 | 1 | 26127961 | 26160235 | 82 | 9 | 40458 | 1.5294 | 0.063081 | *RP1-317E23.6* |
| ENSG00000222009 | 1 | 45264154 | 45291257 | 59 | 10 | 51083 | 1.529 | 0.063129 | *BTBD19* |
| ENSG00000169696 | 17 | 79924683 | 79985282 | 137 | 25 | 40099 | 1.529 | 0.06313 | *ASPSCR1* |

| | | | | | | | | | |
|---|---|---|---|---|---|---|---|---|---|
| ENSG00000171790 | 1 | 41471269 | 41498909 | 88 | 16 | 48561 | 1.5288 | 0.063163 | *SLFNL1* |
| ENSG00000167930 | 16 | 274545 | 329942 | 190 | 38 | 39174 | 1.5284 | 0.063201 | *ITFG3* |
| ENSG00000135547 | 6 | 126058810 | 126092415 | 58 | 9 | 41460 | 1.5284 | 0.063203 | *HEY2* |
| ENSG00000154975 | 17 | 49697674 | 50247377 | 1486 | 132 | 44808 | 1.5281 | 0.063245 | *CA10* |
| ENSG00000243708 | 15 | 42110283 | 42150345 | 141 | 20 | 46487 | 1.528 | 0.063259 | *PLA2G4B* |
| ENSG00000168970 | 15 | 42110293 | 42150353 | 141 | 20 | 46487 | 1.528 | 0.063259 | *JMJD7-PLA2G4B* |
| ENSG00000151292 | 5 | 122837793 | 122962739 | 242 | 21 | 46691 | 1.5277 | 0.063296 | *CSNK1G3* |
| ENSG00000091436 | 2 | 173930163 | 174142738 | 726 | 78 | 43315 | 1.5276 | 0.06331 | *MLTK* |
| ENSG00000145386 | 4 | 122727599 | 122755087 | 68 | 12 | 43267 | 1.5275 | 0.06332 | *CCNA2* |
| ENSG00000105409 | 19 | 42460734 | 42511649 | 44 | 17 | 42949 | 1.5267 | 0.063414 | *ATP1A3* |
| ENSG00000124256 | 20 | 56168902 | 56205632 | 104 | 33 | 49250 | 1.5258 | 0.063527 | *ZBP1* |
| ENSG00000037965 | 12 | 54392832 | 54417570 | 30 | 10 | 51228 | 1.5258 | 0.06353 | *HOXC8* |
| ENSG00000133863 | 8 | 30679060 | 30758122 | 199 | 19 | 30816 | 1.5255 | 0.063568 | *TEX15* |
| ENSG00000145832 | 5 | 135160338 | 135234326 | 292 | 40 | 43658 | 1.5251 | 0.06362 | *SLC25A48* |
| ENSG00000185024 | 14 | 105665623 | 105791926 | 313 | 27 | 46391 | 1.5251 | 0.06362 | *BRF1* |
| ENSG00000166825 | 15 | 90318120 | 90368633 | 206 | 34 | 46214 | 1.525 | 0.063626 | *ANPEP* |
| ENSG00000177363 | 11 | 62443874 | 62467371 | 39 | 7 | 50599 | 1.5248 | 0.063655 | *LRRN4CL* |
| ENSG00000014138 | 11 | 65019233 | 65083060 | 89 | 18 | 44193 | 1.5246 | 0.063674 | *POLA2* |
| ENSG00000111142 | 12 | 95857296 | 95919615 | 266 | 29 | 47571 | 1.5244 | 0.06371 | *METAP2* |
| ENSG00000124593 | 6 | 41738087 | 41767879 | 71 | 17 | 47110 | 1.5241 | 0.063738 | *PRICKLE4* |
| ENSG00000244414 | 1 | 196778887 | 196811319 | 22 | 13 | 40922 | 1.5241 | 0.063742 | *CFHR1* |
| ENSG00000095574 | 10 | 124740322 | 124778333 | 141 | 19 | 40497 | 1.5241 | 0.063747 | *IKZF5* |
| ENSG00000127824 | 2 | 220104433 | 220152892 | 109 | 17 | 30283 | 1.5235 | 0.063815 | *TUBA4A* |
| ENSG00000182923 | 3 | 134194585 | 134303859 | 297 | 10 | 52797 | 1.5235 | 0.063815 | *CEP63* |
| ENSG00000198601 | 1 | 248333288 | 248354331 | 127 | 13 | 45058 | 1.5232 | 0.063858 | *OR2M2* |
| ENSG00000166471 | 11 | 9292201 | 9346327 | 184 | 20 | 43888 | 1.523 | 0.063881 | *TMEM41B* |
| ENSG00000115750 | 2 | 9973483 | 10084545 | 617 | 39 | 40261 | 1.5228 | 0.063909 | *TAF1B* |
| ENSG00000149260 | 11 | 76767979 | 76847201 | 248 | 54 | 44178 | 1.522 | 0.064009 | *CAPN5* |
| ENSG00000112715 | 6 | 43727921 | 43764224 | 96 | 28 | 41675 | 1.5214 | 0.064082 | *VEGFA* |
| ENSG00000226174 | 14 | 105854916 | 105926443 | 54 | 35 | 43925 | 1.5214 | 0.064084 | *TEX22* |
| ENSG00000254870 | 6 | 31487996 | 31524385 | 262 | 36 | 50632 | 1.5213 | 0.064091 | *ATP6V1G2-DDX39B* |
| ENSG00000255874 | 13 | 111511578 | 111532162 | 52 | 13 | 33638 | 1.5212 | 0.064101 | *LINC00346* |
| ENSG00000251493 | 5 | 72730654 | 72754352 | 29 | 7 | 52111 | 1.5205 | 0.064187 | *FOXD1* |
| ENSG00000111860 | 6 | 118771935 | 119041238 | 851 | 51 | 51719 | 1.5203 | 0.064221 | *CEP85L* |
| ENSG00000156127 | 14 | 75978768 | 76023358 | 130 | 24 | 47136 | 1.5196 | 0.064301 | *BATF* |
| ENSG00000175352 | 11 | 8992123 | 9035596 | 114 | 16 | 48116 | 1.5193 | 0.064339 | *NRIP3* |

| | | | | | | | | | |
|---|---|---|---|---|---|---|---|---|---|
| ENSG00000158901 | 20 | 44169792 | 44217965 | 226 | 8 | 26005 | 1.5192 | 0.064353 | *WFDC8* |
| ENSG00000235376 | 10 | 104995644 | 105017773 | 43 | 10 | 50222 | 1.5191 | 0.064364 | *RPEL1* |
| ENSG00000106803 | 9 | 101974346 | 102002897 | 86 | 9 | 55379 | 1.5191 | 0.064369 | *SEC61B* |
| ENSG00000253426 | 8 | 8999252 | 9035646 | 66 | 15 | 36097 | 1.5191 | 0.064374 | *RP11-10A14.4* |
| ENSG00000167106 | 9 | 130692858 | 130752792 | 143 | 16 | 35769 | 1.519 | 0.064386 | *FAM102A* |
| ENSG00000161011 | 5 | 179223388 | 179275078 | 212 | 43 | 42769 | 1.5189 | 0.064395 | *SQSTM1* |
| ENSG00000182512 | 14 | 95989840 | 96021061 | 142 | 27 | 45628 | 1.5189 | 0.064396 | *GLRX5* |
| ENSG00000196470 | 16 | 48380275 | 48492313 | 189 | 30 | 43711 | 1.5187 | 0.064416 | *SIAH1* |
| ENSG00000117676 | 1 | 26846252 | 26911521 | 143 | 22 | 37508 | 1.5178 | 0.064528 | *RPS6KA1* |
| ENSG00000140450 | 15 | 98452784 | 98527068 | 383 | 58 | 39179 | 1.5176 | 0.064553 | *ARRDC4* |
| ENSG00000134470 | 10 | 5980855 | 6030150 | 239 | 49 | 44966 | 1.5175 | 0.064575 | *IL15RA* |
| ENSG00000147437 | 8 | 25266776 | 25292170 | 53 | 9 | 44808 | 1.5172 | 0.064611 | *GNRH1* |
| ENSG00000114626 | 3 | 127381778 | 127409768 | 60 | 13 | 55482 | 1.5169 | 0.064648 | *ABTB1* |
| ENSG00000146587 | 7 | 5075452 | 5119119 | 177 | 24 | 46470 | 1.5168 | 0.064665 | *RBAK* |
| ENSG00000240972 | 22 | 24226191 | 24247414 | 118 | 21 | 44721 | 1.5166 | 0.064678 | *MIF* |
| ENSG00000115504 | 2 | 62890986 | 63283622 | 503 | 61 | 53104 | 1.5166 | 0.064688 | *EHBP1* |
| ENSG00000270024 | 8 | 67569831 | 67782707 | 362 | 28 | 38830 | 1.5166 | 0.064688 | *C8orf44-SGK3* |
| ENSG00000067057 | 10 | 3098525 | 3189904 | 720 | 91 | 46741 | 1.5164 | 0.064705 | *PFKP* |
| ENSG00000221836 | 7 | 143737461 | 143758473 | 95 | 10 | 47643 | 1.5156 | 0.06481 | *OR2A5* |
| ENSG00000115263 | 2 | 162989392 | 163018914 | 83 | 17 | 39381 | 1.5155 | 0.064821 | *GCG* |
| ENSG00000145348 | 4 | 106952756 | 107252652 | 771 | 41 | 52101 | 1.5154 | 0.064832 | *TBCK* |
| ENSG00000073792 | 3 | 185351527 | 185552844 | 391 | 47 | 45525 | 1.5152 | 0.06486 | *IGF2BP2* |
| ENSG00000160471 | 19 | 55850674 | 55876182 | 80 | 21 | 45903 | 1.5149 | 0.0649 | *COX6B2* |
| ENSG00000170275 | 3 | 33145471 | 33199265 | 143 | 25 | 41841 | 1.5149 | 0.064902 | *CRTAP* |
| ENSG00000197530 | 1 | 1540795 | 1575990 | 41 | 13 | 31822 | 1.5144 | 0.064957 | *MIB2* |
| ENSG00000187242 | 17 | 39007555 | 39033462 | 86 | 16 | 44065 | 1.5141 | 0.064996 | *KRT12* |
| ENSG00000203301 | 1 | 2133360 | 2155620 | 54 | 18 | 44549 | 1.5138 | 0.065038 | *AL590822.1* |
| ENSG00000112977 | 5 | 10669342 | 10771384 | 339 | 32 | 44554 | 1.5138 | 0.065038 | *DAP* |
| ENSG00000228768 | 17 | 29888161 | 29912678 | 84 | 28 | 53822 | 1.5137 | 0.065054 | *AC003101.1* |
| ENSG00000153989 | 6 | 117986665 | 118041803 | 151 | 30 | 47250 | 1.5136 | 0.065061 | *NUS1* |
| ENSG00000183963 | 22 | 31450091 | 31510743 | 228 | 31 | 45700 | 1.5135 | 0.065082 | *SMTN* |
| ENSG00000141543 | 17 | 78099013 | 78130982 | 135 | 21 | 46792 | 1.5133 | 0.065107 | *EIF4A3* |
| ENSG00000134253 | 1 | 117643682 | 117675209 | 91 | 17 | 55216 | 1.5129 | 0.065151 | *TRIM45* |
| ENSG00000204604 | 19 | 53331261 | 53370902 | 212 | 28 | 42472 | 1.5126 | 0.065197 | *ZNF468* |
| ENSG00000141933 | 19 | 497299 | 529654 | 137 | 39 | 42516 | 1.5125 | 0.065202 | *TPGS1* |
| ENSG00000167645 | 19 | 38785753 | 38817913 | 114 | 18 | 53405 | 1.5125 | 0.065203 | *YIF1B* |

| | | | | | | | | | |
|---|---|---|---|---|---|---|---|---|---|
| ENSG00000267812 | 21 | 37260353 | 37280727 | 27 | 9 | 47317 | 1.512 | 0.065262 | *FKSG68* |
| ENSG00000137634 | 11 | 114431313 | 114476484 | 131 | 23 | 40968 | 1.512 | 0.065267 | *NXPE4* |
| ENSG00000176749 | 17 | 30803637 | 30828274 | 41 | 11 | 38598 | 1.5112 | 0.065367 | *CDK5R1* |
| ENSG00000101204 | 20 | 61965420 | 62019753 | 344 | 64 | 44395 | 1.5109 | 0.065412 | *CHRNA4* |
| ENSG00000204852 | 12 | 111041832 | 111097235 | 114 | 13 | 48239 | 1.5097 | 0.065555 | *TCTN1* |
| ENSG00000167523 | 16 | 89714210 | 89747680 | 151 | 24 | 40746 | 1.5097 | 0.065557 | *SPATA33* |
| ENSG00000092847 | 1 | 36325409 | 36405211 | 64 | 18 | 50920 | 1.5097 | 0.06556 | *AGO1* |
| ENSG00000168925 | 16 | 75242898 | 75268822 | 177 | 25 | 45500 | 1.5095 | 0.065585 | *CTRB1* |
| ENSG00000131778 | 1 | 146704291 | 146777443 | 389 | 31 | 44426 | 1.5093 | 0.065608 | *CHD1L* |
| ENSG00000130935 | 17 | 65703949 | 65750647 | 140 | 15 | 42496 | 1.5088 | 0.065675 | *NOL11* |
| ENSG00000167778 | 12 | 53448388 | 53483204 | 76 | 18 | 42102 | 1.5087 | 0.065694 | *SPRYD3* |
| ENSG00000183765 | 22 | 29073731 | 29148410 | 211 | 37 | 47181 | 1.5086 | 0.065705 | *CHEK2* |
| ENSG00000175087 | 1 | 26427664 | 26462034 | 96 | 5 | 47876 | 1.5086 | 0.065706 | *PDIK1L* |
| ENSG00000120242 | 9 | 21399146 | 21420184 | 78 | 19 | 50996 | 1.5084 | 0.065723 | *IFNA8* |
| ENSG00000151012 | 4 | 139075251 | 139173503 | 285 | 37 | 43506 | 1.5083 | 0.06574 | *SLC7A11* |
| ENSG00000087266 | 4 | 2784750 | 2852825 | 275 | 44 | 42988 | 1.5082 | 0.065753 | *SH3BP2* |
| ENSG00000016864 | 3 | 52718505 | 52750048 | 64 | 8 | 52851 | 1.5081 | 0.065766 | *GLT8D1* |
| ENSG00000120659 | 13 | 43126872 | 43192149 | 217 | 28 | 54149 | 1.508 | 0.065782 | *TNFSF11* |
| ENSG00000012124 | 19 | 35800164 | 35848258 | 175 | 60 | 42524 | 1.5079 | 0.065792 | *CD22* |
| ENSG00000197774 | 16 | 1813208 | 1841709 | 103 | 20 | 46260 | 1.5074 | 0.065852 | *EME2* |
| ENSG00000167941 | 17 | 41821099 | 41846156 | 70 | 16 | 48963 | 1.5069 | 0.065924 | *SOST* |
| ENSG00000091428 | 2 | 173590002 | 173927621 | 1058 | 118 | 47821 | 1.506 | 0.06603 | *RAPGEF4* |
| ENSG00000196182 | 1 | 36795225 | 36861497 | 106 | 21 | 46881 | 1.5056 | 0.066084 | *STK40* |
| ENSG00000236980 | 3 | 49205065 | 49239291 | 54 | 8 | 46397 | 1.5056 | 0.066091 | *C3orf84* |
| ENSG00000162104 | 16 | 3993388 | 4176186 | 727 | 115 | 40932 | 1.5047 | 0.066203 | *ADCY9* |
| ENSG00000142235 | 19 | 48978528 | 49026446 | 107 | 24 | 41413 | 1.5042 | 0.066267 | *LMTK3* |
| ENSG00000233024 | 16 | 18441943 | 18483188 | 4 | 1 | 46364 | 1.5037 | 0.066328 | *NPIPA7* |
| ENSG00000119285 | 1 | 236702305 | 236777804 | 543 | 29 | 28718 | 1.5036 | 0.06634 | *HEATR1* |
| ENSG00000135269 | 7 | 115840547 | 115908837 | 278 | 20 | 47588 | 1.5032 | 0.066391 | *TES* |
| ENSG00000242173 | 16 | 308726 | 343003 | 141 | 39 | 38001 | 1.5032 | 0.066392 | *ARHGDIG* |
| ENSG00000258052 | 12 | 70185449 | 70259143 | 260 | 32 | 41750 | 1.5031 | 0.066412 | *AC025263.3* |
| ENSG00000259642 | 15 | 80205113 | 80227194 | 126 | 11 | 54665 | 1.5027 | 0.066462 | *C15orf37* |
| ENSG00000165566 | 13 | 25725822 | 25756426 | 68 | 21 | 44522 | 1.5027 | 0.066464 | *AMER2* |
| ENSG00000119421 | 9 | 124884745 | 124932098 | 174 | 25 | 35050 | 1.5019 | 0.066566 | *NDUFA8* |
| ENSG00000183662 | 3 | 68043359 | 68604776 | 2122 | 157 | 49589 | 1.5011 | 0.06666 | *FAM19A1* |
| ENSG00000111339 | 12 | 14968503 | 15006429 | 112 | 13 | 54592 | 1.5011 | 0.066668 | *ART4* |

| | | | | | | | | | |
|---|---|---|---|---|---|---|---|---|---|
| ENSG00000232125 | 2 | 207506345 | 207593120 | 359 | 46 | 46001 | 1.5008 | 0.066704 | *DYTN* |
| ENSG00000005513 | 16 | 1021808 | 1046979 | 142 | 30 | 42988 | 1.4998 | 0.066827 | *SOX8* |
| ENSG00000250173 | 5 | 969480 | 991415 | 40 | 14 | 45566 | 1.4994 | 0.066884 | *RP11-661C8.3* |
| ENSG00000257028 | 15 | 80205507 | 80227194 | 124 | 11 | 54627 | 1.4993 | 0.066899 | *C15ORF37* |
| ENSG00000130449 | 5 | 60618100 | 60851997 | 437 | 42 | 49096 | 1.4993 | 0.066904 | *ZSWIM6* |
| ENSG00000146521 | 6 | 168175217 | 168207539 | 174 | 24 | 40445 | 1.4991 | 0.066924 | *C6orf123* |
| ENSG00000127483 | 1 | 21059154 | 21123816 | 183 | 24 | 51082 | 1.4985 | 0.066997 | *HP1BP3* |
| ENSG00000108506 | 17 | 59932731 | 60015377 | 150 | 18 | 40981 | 1.4984 | 0.067016 | *INTS2* |
| ENSG00000118058 | 11 | 118297205 | 118407539 | 142 | 26 | 40907 | 1.4983 | 0.067029 | *KMT2A* |
| ENSG00000180884 | 19 | 35437258 | 35464953 | 110 | 16 | 51135 | 1.498 | 0.067067 | *ZNF792* |
| ENSG00000084734 | 2 | 27709709 | 27756554 | 105 | 17 | 48409 | 1.4977 | 0.067111 | *GCKR* |
| ENSG00000179071 | 11 | 85384893 | 85407320 | 50 | 14 | 49161 | 1.4973 | 0.067152 | *CCDC89* |
| ENSG00000075340 | 2 | 70824750 | 71005357 | 785 | 82 | 46381 | 1.4973 | 0.067162 | *ADD2* |
| ENSG00000116977 | 1 | 236671300 | 236726281 | 374 | 19 | 27029 | 1.4966 | 0.067249 | *LGALS8* |
| ENSG00000184887 | 14 | 105704827 | 105727430 | 73 | 7 | 52719 | 1.4966 | 0.067249 | *BTBD6* |
| ENSG00000105983 | 7 | 156451646 | 156695924 | 967 | 65 | 51955 | 1.4963 | 0.067287 | *LMBR1* |
| ENSG00000114902 | 3 | 52728971 | 52752182 | 51 | 8 | 53042 | 1.4963 | 0.067291 | *SPCS1* |
| ENSG00000108604 | 17 | 61899444 | 61930425 | 75 | 6 | 23538 | 1.4961 | 0.067319 | *SMARCD2* |
| ENSG00000179889 | 16 | 15058448 | 15243196 | 319 | 46 | 40991 | 1.496 | 0.067333 | *PDXDC1* |
| ENSG00000197604 | 10 | 72184585 | 72206314 | 49 | 11 | 44350 | 1.4959 | 0.067334 | *AC022532.1* |
| ENSG00000128309 | 22 | 37405676 | 37435863 | 113 | 24 | 45709 | 1.4958 | 0.067354 | *MPST* |
| ENSG00000256374 | 1 | 148191752 | 148212536 | 3 | 2 | 42528 | 1.4958 | 0.067355 | *PPIAL4D* |
| ENSG00000168412 | 4 | 187444809 | 187486721 | 177 | 29 | 41216 | 1.4957 | 0.067366 | *MTNR1A* |
| ENSG00000161179 | 22 | 21972378 | 21994353 | 46 | 10 | 30184 | 1.4953 | 0.067423 | *YDJC* |
| ENSG00000114480 | 3 | 81528850 | 81821312 | 575 | 53 | 40236 | 1.4945 | 0.067517 | *GBE1* |
| ENSG00000128872 | 15 | 52033758 | 52118565 | 263 | 18 | 53165 | 1.4945 | 0.067522 | *TMOD2* |
| ENSG00000147789 | 8 | 146042849 | 146082894 | 135 | 12 | 43141 | 1.4941 | 0.067576 | *ZNF7* |
| ENSG00000122203 | 5 | 175763064 | 175798971 | 98 | 15 | 52039 | 1.4939 | 0.0676 | *KIAA1191* |
| ENSG00000237765 | 4 | 15673285 | 15717188 | 125 | 17 | 53476 | 1.4938 | 0.067614 | *FAM200B* |
| ENSG00000153339 | 18 | 29399136 | 29543099 | 415 | 37 | 49991 | 1.493 | 0.067719 | *TRAPPC8* |
| ENSG00000169252 | 5 | 148196156 | 148218196 | 83 | 12 | 39538 | 1.4929 | 0.067727 | *ADRB2* |
| ENSG00000269242 | 19 | 12744645 | 12769211 | 48 | 12 | 32899 | 1.4926 | 0.067772 | *CTD-2192J16.22* |
| ENSG00000143458 | 1 | 151033054 | 151108018 | 189 | 26 | 41143 | 1.4926 | 0.067773 | *GABPB2* |
| ENSG00000141977 | 19 | 16262179 | 16294336 | 107 | 13 | 39646 | 1.4923 | 0.067817 | *CIB3* |
| ENSG00000157823 | 15 | 90363831 | 90447574 | 238 | 19 | 52232 | 1.4921 | 0.067831 | *AP3S2* |
| ENSG00000197622 | 1 | 151013447 | 151052801 | 94 | 20 | 45552 | 1.492 | 0.067853 | *CDC42SE1* |

| | | | | | | | | | |
|---|---|---|---|---|---|---|---|---|---|
| ENSG00000115207 | 2 | 27538716 | 27589868 | 67 | 15 | 42863 | 1.4919 | 0.067866 | *GTF3C2* |
| ENSG00000115290 | 2 | 165339322 | 165488358 | 279 | 43 | 44182 | 1.4918 | 0.067875 | *GRB14* |
| ENSG00000166822 | 16 | 75466952 | 75509395 | 193 | 16 | 42163 | 1.4918 | 0.067882 | *TMEM170A* |
| ENSG00000109061 | 17 | 10385624 | 10431860 | 74 | 10 | 30811 | 1.4913 | 0.067948 | *MYH1* |
| ENSG00000137166 | 6 | 41504164 | 41580122 | 281 | 40 | 47576 | 1.4907 | 0.068017 | *FOXP4* |
| ENSG00000205329 | 12 | 55715485 | 55736420 | 89 | 10 | 44067 | 1.4902 | 0.068083 | *OR6C3* |
| ENSG00000131051 | 20 | 34281531 | 34340234 | 197 | 13 | 45569 | 1.49 | 0.068106 | *RBM39* |
| ENSG00000135175 | 7 | 97603996 | 97630481 | 158 | 16 | 48320 | 1.4897 | 0.068157 | *OCM2* |
| ENSG00000130427 | 7 | 100308423 | 100331323 | 59 | 12 | 53842 | 1.4896 | 0.06816 | *EPO* |
| ENSG00000151690 | 2 | 191263081 | 191383931 | 351 | 32 | 46129 | 1.4896 | 0.068165 | *MFSD6* |
| ENSG00000213246 | 17 | 56412539 | 56440454 | 73 | 17 | 51718 | 1.4893 | 0.068201 | *SUPT4H1* |
| ENSG00000172482 | 2 | 241797896 | 241829919 | 161 | 17 | 52003 | 1.4884 | 0.068323 | *AGXT* |
| ENSG00000010256 | 3 | 48626435 | 48658409 | 31 | 13 | 43100 | 1.4882 | 0.06835 | *UQCRC1* |
| ENSG00000134255 | 1 | 111672249 | 111737724 | 139 | 23 | 43999 | 1.4877 | 0.068409 | *CEPT1* |
| ENSG00000180992 | 6 | 44071194 | 44105194 | 145 | 20 | 40952 | 1.4872 | 0.068482 | *MRPL14* |
| ENSG00000186119 | 11 | 55577086 | 55598073 | 83 | 12 | 51985 | 1.4869 | 0.068521 | *OR5D18* |
| ENSG00000136908 | 9 | 130687378 | 130710763 | 47 | 7 | 29004 | 1.4868 | 0.06854 | *DPM2* |
| ENSG00000108405 | 17 | 3789886 | 3829794 | 108 | 28 | 40983 | 1.4865 | 0.068578 | *P2RX1* |
| ENSG00000107485 | 10 | 8085567 | 8127161 | 166 | 30 | 49016 | 1.4864 | 0.068585 | *GATA3* |
| ENSG00000170322 | 11 | 129723531 | 129775485 | 117 | 19 | 49634 | 1.4863 | 0.068606 | *NFRKB* |
| ENSG00000176978 | 9 | 139994994 | 140019629 | 89 | 17 | 45413 | 1.4857 | 0.068675 | *DPP7* |
| ENSG00000153815 | 16 | 81468775 | 81755367 | 1261 | 184 | 45884 | 1.4853 | 0.068735 | *CMIP* |
| ENSG00000160339 | 9 | 137762654 | 137789924 | 145 | 26 | 53015 | 1.4851 | 0.068763 | *FCN2* |
| ENSG00000161847 | 19 | 10416888 | 10454316 | 151 | 26 | 52855 | 1.4848 | 0.068798 | *RAVER1* |
| ENSG00000115657 | 2 | 220064490 | 220093712 | 44 | 11 | 38836 | 1.4843 | 0.068868 | *ABCB6* |
| ENSG00000169704 | 3 | 128769610 | 128791249 | 18 | 6 | 32564 | 1.4837 | 0.068943 | *GP9* |
| ENSG00000125304 | 13 | 100143671 | 100226260 | 219 | 29 | 48379 | 1.4835 | 0.068973 | *TM9SF2* |
| ENSG00000163632 | 3 | 63795038 | 63844312 | 84 | 17 | 44583 | 1.4832 | 0.069013 | *C3orf49* |
| ENSG00000129055 | 3 | 134186548 | 134215558 | 77 | 7 | 50002 | 1.4827 | 0.069072 | *ANAPC13* |
| ENSG00000178776 | 5 | 147250289 | 147296101 | 212 | 28 | 40748 | 1.4827 | 0.069075 | *C5orf46* |
| ENSG00000180357 | 15 | 64742941 | 64988264 | 469 | 41 | 37982 | 1.4827 | 0.069082 | *ZNF609* |
| ENSG00000188242 | 5 | 456239 | 483213 | 153 | 17 | 30838 | 1.4826 | 0.069084 | *CTD-2228K2.5* |
| ENSG00000184857 | 16 | 8864241 | 8901505 | 226 | 38 | 46028 | 1.4823 | 0.069127 | *TMEM186* |
| ENSG00000094963 | 1 | 171144347 | 171191822 | 253 | 23 | 41024 | 1.4817 | 0.069212 | *FMO2* |
| ENSG00000213782 | 12 | 12956250 | 12992915 | 132 | 27 | 47195 | 1.4816 | 0.069229 | *DDX47* |
| ENSG00000205856 | 22 | 32534993 | 32565309 | 164 | 28 | 42206 | 1.4811 | 0.069291 | *C22orf42* |

| | | | | | | | | | |
|---|---|---|---|---|---|---|---|---|---|
| ENSG00000156234 | 4 | 78422907 | 78542988 | 229 | 51 | 47113 | 1.4809 | 0.06931 | *CXCL13* |
| ENSG00000163067 | 2 | 95821177 | 95860065 | 47 | 17 | 48758 | 1.4809 | 0.069312 | *ZNF2* |
| ENSG00000120324 | 5 | 140561942 | 140585215 | 96 | 14 | 43098 | 1.4802 | 0.069415 | *PCDHB10* |
| ENSG00000158792 | 16 | 89752751 | 89778113 | 75 | 16 | 37806 | 1.48 | 0.069431 | *SPATA2L* |
| ENSG00000112964 | 5 | 42413879 | 42731979 | 804 | 82 | 45553 | 1.4799 | 0.069455 | *GHR* |
| ENSG00000168610 | 17 | 40455342 | 40550586 | 205 | 21 | 45813 | 1.4792 | 0.069544 | *STAT3* |
| ENSG00000184517 | 16 | 75172390 | 75216134 | 191 | 23 | 40154 | 1.4787 | 0.069609 | *ZFP1* |
| ENSG00000090372 | 19 | 47212764 | 47260251 | 112 | 22 | 53120 | 1.4786 | 0.069626 | *STRN4* |
| ENSG00000136982 | 8 | 120836216 | 120878250 | 223 | 22 | 38089 | 1.4785 | 0.069642 | *DSCC1* |
| ENSG00000144736 | 3 | 72788428 | 72921065 | 225 | 18 | 45043 | 1.4775 | 0.069771 | *SHQ1* |
| ENSG00000163794 | 2 | 27520268 | 27541313 | 32 | 11 | 42155 | 1.4775 | 0.069777 | *UCN* |
| ENSG00000144063 | 2 | 110831447 | 110884143 | 50 | 16 | 28789 | 1.477 | 0.069837 | *MALL* |
| ENSG00000106399 | 7 | 7666149 | 7768238 | 387 | 42 | 50801 | 1.477 | 0.069844 | *RPA3* |
| ENSG00000184601 | 14 | 105036021 | 105066852 | 90 | 15 | 36193 | 1.4768 | 0.069864 | *C14orf180* |
| ENSG00000076344 | 16 | 308300 | 335980 | 97 | 29 | 35112 | 1.4768 | 0.06987 | *RGS11* |
| ENSG00000186010 | 19 | 19616545 | 19654285 | 94 | 10 | 46032 | 1.4767 | 0.06988 | *NDUFA13* |
| ENSG00000213921 | 19 | 40257235 | 40286775 | 102 | 18 | 46877 | 1.4765 | 0.069906 | *LEUTX* |
| ENSG00000146674 | 7 | 45941949 | 45971473 | 63 | 16 | 54011 | 1.4763 | 0.069929 | *IGFBP3* |
| ENSG00000198563 | 6 | 31487996 | 31520225 | 232 | 32 | 51683 | 1.4759 | 0.069988 | *DDX39B* |
| ENSG00000188822 | 1 | 24187016 | 24295549 | 293 | 20 | 45589 | 1.4757 | 0.070015 | *CNR2* |
| ENSG00000100218 | 22 | 23391593 | 23497208 | 232 | 26 | 50162 | 1.4753 | 0.070062 | *RTDR1* |
| ENSG00000171291 | 19 | 11949541 | 12004565 | 173 | 24 | 39519 | 1.4752 | 0.070074 | *ZNF439* |
| ENSG00000173826 | 17 | 61590695 | 61636338 | 80 | 17 | 51765 | 1.475 | 0.070109 | *KCNH6* |
| ENSG00000204511 | 6 | 31486494 | 31508009 | 169 | 22 | 53751 | 1.4747 | 0.070151 | *MCCD1* |
| ENSG00000255046 | 8 | 11645437 | 11670077 | 174 | 30 | 52756 | 1.4744 | 0.070188 | *RP11-297N6.4* |
| ENSG00000116161 | 1 | 174958300 | 174990851 | 63 | 16 | 53941 | 1.4737 | 0.070278 | *CACYBP* |
| ENSG00000162687 | 1 | 196184909 | 196588355 | 773 | 49 | 46370 | 1.4735 | 0.070307 | *KCNT2* |
| ENSG00000149781 | 11 | 63964150 | 64001354 | 89 | 20 | 48948 | 1.4732 | 0.070342 | *FERMT3* |
| ENSG00000175354 | 18 | 12775477 | 12939642 | 525 | 66 | 40881 | 1.473 | 0.07037 | *PTPN2* |
| ENSG00000159377 | 1 | 151362010 | 151384420 | 26 | 12 | 42572 | 1.473 | 0.070372 | *PSMB4* |
| ENSG00000053900 | 4 | 25368835 | 25430120 | 234 | 23 | 38497 | 1.473 | 0.070382 | *ANAPC4* |
| ENSG00000204084 | 1 | 38316369 | 38422729 | 270 | 30 | 40368 | 1.4727 | 0.07041 | *INPP5B* |
| ENSG00000117305 | 1 | 24118375 | 24175110 | 83 | 8 | 51617 | 1.4724 | 0.070458 | *HMGCL* |
| ENSG00000272573 | 3 | 52857130 | 52879235 | 56 | 7 | 52552 | 1.4714 | 0.070588 | *MUSTN1* |
| ENSG00000140280 | 15 | 52005208 | 52053782 | 162 | 17 | 54041 | 1.4711 | 0.070638 | *LYSMD2* |
| ENSG00000197446 | 19 | 41610337 | 41644271 | 181 | 24 | 45771 | 1.471 | 0.070642 | *CYP2F1* |

| | | | | | | | | | |
|---|---|---|---|---|---|---|---|---|---|
| ENSG00000169554 | 2 | 145131648 | 145292147 | 255 | 61 | 45478 | 1.471 | 0.07065 | *ZEB2* |
| ENSG00000178772 | 3 | 194050494 | 194082057 | 162 | 26 | 42562 | 1.4709 | 0.07066 | *CPN2* |
| ENSG00000162004 | 16 | 762582 | 786954 | 88 | 14 | 29024 | 1.4708 | 0.070668 | *CCDC78* |
| ENSG00000008838 | 17 | 38165350 | 38227468 | 162 | 18 | 49245 | 1.4708 | 0.07067 | *MED24* |
| ENSG00000187715 | 3 | 127624075 | 127716514 | 300 | 35 | 45682 | 1.4708 | 0.070674 | *KBTBD12* |
| ENSG00000183395 | 12 | 102580237 | 102601623 | 36 | 7 | 51529 | 1.4708 | 0.070677 | *PMCH* |
| ENSG00000140057 | 14 | 96848448 | 96965764 | 295 | 52 | 48832 | 1.4707 | 0.070692 | *AK7* |
| ENSG00000166441 | 11 | 8693958 | 8746306 | 154 | 35 | 44719 | 1.4703 | 0.070741 | *RPL27A* |
| ENSG00000172663 | 11 | 67221824 | 67246743 | 42 | 15 | 43046 | 1.4702 | 0.070748 | *TMEM134* |
| ENSG00000198848 | 16 | 55826763 | 55877249 | 98 | 25 | 48272 | 1.4699 | 0.0708 | *CES1* |
| ENSG00000112874 | 5 | 102874556 | 102908494 | 95 | 14 | 52882 | 1.4694 | 0.070856 | *NUDT12* |
| ENSG00000174332 | 1 | 53961910 | 54209877 | 652 | 70 | 41629 | 1.4694 | 0.070867 | *GLIS1* |
| ENSG00000159650 | 3 | 126190124 | 126246616 | 225 | 48 | 45804 | 1.4686 | 0.070972 | *UROC1* |
| ENSG00000183682 | 1 | 39947318 | 40001607 | 144 | 26 | 36446 | 1.468 | 0.071047 | *BMP8A* |
| ENSG00000198326 | 20 | 2785614 | 2810930 | 54 | 19 | 41497 | 1.4678 | 0.071084 | *TMEM239* |
| ENSG00000019549 | 8 | 49820249 | 49844299 | 33 | 13 | 45229 | 1.4677 | 0.071094 | *SNAI2* |
| ENSG00000080709 | 5 | 113686642 | 113842337 | 406 | 59 | 43267 | 1.4672 | 0.071162 | *KCNN2* |
| ENSG00000181001 | 11 | 5799084 | 5820046 | 70 | 7 | 45398 | 1.467 | 0.071195 | *OR52N1* |
| ENSG00000128266 | 22 | 23402540 | 23477224 | 168 | 19 | 50836 | 1.4668 | 0.071211 | *GNAZ* |
| ENSG00000113303 | 5 | 180316077 | 180387906 | 209 | 16 | 44709 | 1.4665 | 0.071253 | *BTNL8* |
| ENSG00000127528 | 19 | 16425628 | 16448685 | 95 | 20 | 50545 | 1.4661 | 0.071315 | *KLF2* |
| ENSG00000269693 | 19 | 12626534 | 12701789 | 159 | 37 | 35886 | 1.466 | 0.071319 | *CTD-2192J16.20* |
| ENSG00000145241 | 4 | 68327521 | 68421324 | 336 | 23 | 49924 | 1.466 | 0.071327 | *CENPC* |
| ENSG00000007174 | 17 | 11491748 | 11883065 | 1098 | 140 | 46613 | 1.466 | 0.07133 | *DNAH9* |
| ENSG00000135097 | 12 | 120769133 | 120816983 | 114 | 14 | 50263 | 1.4659 | 0.071344 | *MSI1* |
| ENSG00000213190 | 1 | 151020234 | 151050970 | 65 | 16 | 44274 | 1.4657 | 0.071363 | *MLLT11* |
| ENSG00000120322 | 5 | 140547371 | 140570081 | 87 | 13 | 42708 | 1.4655 | 0.071386 | *PCDHB8* |
| ENSG00000168071 | 11 | 64097695 | 64135006 | 81 | 12 | 35499 | 1.4646 | 0.071516 | *CCDC88B* |
| ENSG00000188536 | 16 | 212846 | 233709 | 38 | 13 | 44898 | 1.4645 | 0.07153 | *HBA2* |
| ENSG00000177191 | 19 | 41921264 | 41944635 | 59 | 5 | 53728 | 1.4644 | 0.071539 | *B3GNT8* |
| ENSG00000120279 | 6 | 153009030 | 153055702 | 206 | 19 | 33752 | 1.4639 | 0.071617 | *MYCT1* |
| ENSG00000101413 | 20 | 36651948 | 36730768 | 153 | 15 | 37995 | 1.4636 | 0.071658 | *RPRD1B* |
| ENSG00000170677 | 18 | 67946137 | 68007436 | 250 | 40 | 47460 | 1.4632 | 0.071708 | *SOCS6* |
| ENSG00000180953 | 15 | 80181182 | 80226044 | 248 | 23 | 53502 | 1.4631 | 0.071717 | *ST20* |
| ENSG00000221986 | 1 | 109824987 | 109859663 | 90 | 22 | 42710 | 1.4621 | 0.071852 | *MYBPHL* |
| ENSG00000145833 | 5 | 134084469 | 134200823 | 147 | 36 | 40808 | 1.4619 | 0.071879 | *DDX46* |

| | | | | | | | | | |
|---|---|---|---|---|---|---|---|---|---|
| ENSG00000174963 | 3 | 147093833 | 147134647 | 85 | 18 | 53551 | 1.4612 | 0.071985 | *ZIC4* |
| ENSG00000104946 | 19 | 50370682 | 50402005 | 70 | 14 | 42886 | 1.4602 | 0.072111 | *TBC1D17* |
| ENSG00000075884 | 2 | 143838931 | 144535921 | 1822 | 169 | 47514 | 1.4602 | 0.072118 | *ARHGAP15* |
| ENSG00000205047 | 16 | 87721749 | 87749290 | 125 | 29 | 46835 | 1.4601 | 0.07213 | *FLJ00104* |
| ENSG00000064201 | 11 | 2313227 | 2349430 | 150 | 22 | 46123 | 1.4598 | 0.072169 | *TSPAN32* |
| ENSG00000105605 | 19 | 54402589 | 54457195 | 75 | 33 | 36232 | 1.4598 | 0.072178 | *CACNG7* |
| ENSG00000117114 | 1 | 81761845 | 82468120 | 2559 | 270 | 48165 | 1.4596 | 0.072198 | *LPHN2* |
| ENSG00000008197 | 6 | 50671541 | 50750701 | 152 | 19 | 46645 | 1.4595 | 0.07221 | *TFAP2D* |
| ENSG00000172053 | 3 | 49123365 | 49152553 | 31 | 9 | 40918 | 1.4591 | 0.07227 | *QARS* |
| ENSG00000169224 | 1 | 247660360 | 247750992 | 320 | 53 | 48080 | 1.4591 | 0.072272 | *GCSAML* |
| ENSG00000116106 | 2 | 222272747 | 222448922 | 491 | 67 | 46097 | 1.4588 | 0.072312 | *EPHA4* |
| ENSG00000206172 | 16 | 216679 | 237521 | 46 | 14 | 47982 | 1.4583 | 0.072379 | *HBA1* |
| ENSG00000268927 | 16 | 70685570 | 70709739 | 84 | 26 | 42877 | 1.4582 | 0.07239 | *FLJ00418* |
| ENSG00000142945 | 1 | 45195490 | 45243439 | 136 | 18 | 42406 | 1.4581 | 0.072401 | *KIF2C* |
| ENSG00000170558 | 18 | 25520930 | 25767410 | 602 | 73 | 47189 | 1.4581 | 0.072406 | *CDH2* |
| ENSG00000004838 | 3 | 50368541 | 50394283 | 17 | 6 | 54038 | 1.458 | 0.072421 | *ZMYND10* |
| ENSG00000077092 | 3 | 25205823 | 25649423 | 1931 | 185 | 49327 | 1.4577 | 0.072466 | *RARB* |
| ENSG00000081377 | 9 | 99242523 | 99392112 | 191 | 29 | 42166 | 1.4576 | 0.072481 | *CDC14B* |
| ENSG00000175084 | 2 | 220273099 | 220301461 | 67 | 14 | 36253 | 1.4575 | 0.072483 | *DES* |
| ENSG00000059758 | 12 | 96662039 | 96804338 | 389 | 42 | 47921 | 1.4574 | 0.072498 | *CDK17* |
| ENSG00000197343 | 7 | 99146029 | 99184076 | 47 | 13 | 39520 | 1.4574 | 0.072506 | *ZNF655* |
| ENSG00000139350 | 12 | 97291001 | 97357129 | 161 | 16 | 54811 | 1.4574 | 0.072507 | *NEDD1* |
| ENSG00000221949 | 12 | 62985531 | 63007214 | 61 | 15 | 55761 | 1.4573 | 0.072515 | *C12orf61* |
| ENSG00000250021 | 15 | 90367540 | 90466114 | 260 | 22 | 51152 | 1.4573 | 0.072519 | *C15orf38-AP3S2* |
| ENSG00000197536 | 5 | 131736328 | 131821736 | 271 | 32 | 37922 | 1.4566 | 0.072609 | *C5orf56* |
| ENSG00000197119 | 14 | 100747448 | 100782884 | 87 | 25 | 49806 | 1.4562 | 0.072667 | *SLC25A29* |
| ENSG00000167910 | 8 | 59392737 | 59422795 | 82 | 16 | 40325 | 1.4562 | 0.07267 | *CYP7A1* |
| ENSG00000170043 | 17 | 7823663 | 7845441 | 59 | 10 | 44661 | 1.4559 | 0.072705 | *TRAPPC1* |
| ENSG00000151062 | 12 | 1891123 | 2038002 | 548 | 85 | 44496 | 1.4558 | 0.072725 | *CACNA2D4* |
| ENSG00000112273 | 6 | 22559678 | 22581892 | 94 | 16 | 49244 | 1.4557 | 0.072735 | *HDGFL1* |
| ENSG00000169174 | 1 | 55495221 | 55540525 | 156 | 37 | 44747 | 1.4555 | 0.072768 | *PCSK9* |
| ENSG00000176809 | 17 | 62840430 | 62925598 | 26 | 17 | 36301 | 1.4555 | 0.072771 | *LRRC37A3* |
| ENSG00000160111 | 19 | 16993758 | 17147625 | 490 | 97 | 44527 | 1.4547 | 0.072874 | *CPAMD8* |
| ENSG00000156574 | 10 | 72182071 | 72217707 | 75 | 16 | 43089 | 1.4546 | 0.072887 | *NODAL* |
| ENSG00000025293 | 20 | 34349896 | 34548303 | 564 | 29 | 43314 | 1.4545 | 0.072897 | *PHF20* |
| ENSG00000141668 | 18 | 70193915 | 70315756 | 317 | 45 | 37377 | 1.4544 | 0.072916 | *CBLN2* |

| | | | | | | | | | |
|---|---|---|---|---|---|---|---|---|---|
| ENSG00000132274 | 11 | 5700919 | 5768319 | 430 | 52 | 49391 | 1.4543 | 0.072932 | *TRIM22* |
| ENSG00000130204 | 19 | 45383826 | 45416946 | 120 | 28 | 47470 | 1.4539 | 0.072986 | *TOMM40* |
| ENSG00000226180 | 16 | 87717144 | 87739753 | 103 | 23 | 50067 | 1.4526 | 0.073171 | *AC010536.1* |
| ENSG00000154768 | 17 | 34077916 | 34102098 | 87 | 11 | 50020 | 1.4514 | 0.073339 | *C17orf50* |
| ENSG00000131732 | 5 | 80587409 | 80619116 | 98 | 15 | 50244 | 1.4512 | 0.073358 | *ZCCHC9* |
| ENSG00000185156 | 17 | 8690436 | 8712667 | 82 | 14 | 43913 | 1.4512 | 0.073368 | *MFSD6L* |
| ENSG00000139797 | 13 | 98818039 | 98839519 | 55 | 15 | 42325 | 1.4508 | 0.073412 | *RNF113B* |
| ENSG00000239839 | 8 | 6863391 | 6885823 | 58 | 20 | 47495 | 1.4508 | 0.073417 | *DEFA3* |
| ENSG00000187118 | 3 | 28273075 | 28376633 | 223 | 25 | 44881 | 1.4508 | 0.073421 | *CMC1* |
| ENSG00000137948 | 1 | 92404928 | 92489983 | 285 | 18 | 39426 | 1.45 | 0.073527 | *BRDT* |
| ENSG00000154099 | 16 | 84168865 | 84222373 | 266 | 47 | 51086 | 1.4495 | 0.073596 | *DNAAF1* |
| ENSG00000170801 | 4 | 8261492 | 8318838 | 273 | 46 | 43985 | 1.4489 | 0.073685 | *HTRA3* |
| ENSG00000215372 | 8 | 7203039 | 7253080 | 22 | 7 | 41453 | 1.4485 | 0.073736 | *ZNF705G* |
| ENSG00000006016 | 19 | 18673030 | 18728551 | 203 | 27 | 36461 | 1.4485 | 0.073739 | *CRLF1* |
| ENSG00000160058 | 1 | 32820704 | 32870332 | 33 | 20 | 55099 | 1.4485 | 0.073746 | *BSDC1* |
| ENSG00000172955 | 4 | 100113795 | 100150694 | 101 | 18 | 40369 | 1.4482 | 0.073785 | *ADH6* |
| ENSG00000180447 | 9 | 89549279 | 89572104 | 59 | 13 | 43797 | 1.4481 | 0.073801 | *GAS1* |
| ENSG00000154719 | 21 | 26947968 | 26989829 | 164 | 13 | 26331 | 1.4479 | 0.073829 | *MRPL39* |
| ENSG00000108179 | 10 | 81097225 | 81125093 | 70 | 14 | 42688 | 1.4478 | 0.073835 | *PPIF* |
| ENSG00000135052 | 9 | 88631061 | 88725088 | 285 | 25 | 30384 | 1.4476 | 0.07386 | *GOLM1* |
| ENSG00000125945 | 1 | 23675941 | 23705935 | 79 | 14 | 38687 | 1.4475 | 0.073885 | *ZNF436* |
| ENSG00000146414 | 6 | 146175381 | 146295559 | 222 | 15 | 55975 | 1.4472 | 0.073917 | *SHPRH* |
| ENSG00000087191 | 17 | 61894512 | 61919379 | 50 | 5 | 22829 | 1.447 | 0.073949 | *PSMC5* |
| ENSG00000149806 | 11 | 64878100 | 64899945 | 37 | 12 | 45281 | 1.4469 | 0.073962 | *FAU* |
| ENSG00000096996 | 19 | 18159805 | 18219754 | 282 | 42 | 47688 | 1.4468 | 0.07397 | *IL12RB1* |
| ENSG00000198890 | 1 | 107589267 | 107611916 | 33 | 7 | 55133 | 1.4467 | 0.073989 | *PRMT6* |
| ENSG00000156687 | 8 | 35082975 | 35664068 | 1184 | 85 | 43279 | 1.4464 | 0.074031 | *UNC5D* |
| ENSG00000121390 | 13 | 20238896 | 20367142 | 289 | 28 | 40887 | 1.4464 | 0.074032 | *PSPC1* |
| ENSG00000115652 | 2 | 106699759 | 106820795 | 361 | 43 | 49823 | 1.4463 | 0.07404 | *UXS1* |
| ENSG00000170619 | 8 | 146056427 | 146089121 | 110 | 13 | 43585 | 1.4461 | 0.074076 | *COMMD5* |
| ENSG00000198520 | 1 | 45130364 | 45201263 | 159 | 43 | 45137 | 1.4451 | 0.074212 | *C1orf228* |
| ENSG00000105750 | 19 | 21096028 | 21143503 | 332 | 12 | 54259 | 1.4451 | 0.074222 | *ZNF85* |
| ENSG00000181789 | 3 | 128958449 | 129006614 | 101 | 11 | 41285 | 1.4449 | 0.074243 | *COPG1* |
| ENSG00000119922 | 10 | 91051712 | 91079033 | 53 | 16 | 38313 | 1.4449 | 0.074245 | *IFIT2* |
| ENSG00000187239 | 9 | 132639466 | 132815473 | 694 | 69 | 49620 | 1.4449 | 0.074249 | *FNBP1* |
| ENSG00000170929 | 19 | 9193855 | 9214889 | 103 | 14 | 50288 | 1.4443 | 0.07433 | *OR1M1* |

| | | | | | | | | | |
|---|---|---|---|---|---|---|---|---|---|
| ENSG00000117308 | 1 | 24112089 | 24137271 | 21 | 5 | 54602 | 1.4442 | 0.074339 | *GALE* |
| ENSG00000170890 | 12 | 120749914 | 120775592 | 65 | 9 | 46387 | 1.4442 | 0.074346 | *PLA2G1B* |
| ENSG00000183742 | 7 | 20164278 | 20267027 | 298 | 36 | 37636 | 1.4436 | 0.074431 | *MACC1* |
| ENSG00000118971 | 12 | 4372938 | 4424516 | 93 | 37 | 45186 | 1.4431 | 0.074497 | *CCND2* |
| ENSG00000168175 | 14 | 55508349 | 55546910 | 83 | 15 | 52055 | 1.4429 | 0.074528 | *MAPK1IP1L* |
| ENSG00000181908 | 11 | 64206546 | 64229126 | 49 | 15 | 44823 | 1.4425 | 0.074577 | *AP003774.4* |
| ENSG00000143952 | 2 | 64109280 | 64256206 | 493 | 48 | 40142 | 1.4424 | 0.074589 | *VPS54* |
| ENSG00000073417 | 15 | 85513671 | 85692376 | 641 | 47 | 38215 | 1.4424 | 0.074601 | *PDE8A* |
| ENSG00000153233 | 12 | 71021853 | 71324623 | 1065 | 125 | 41946 | 1.4423 | 0.074613 | *PTPRR* |
| ENSG00000144935 | 3 | 142432916 | 142536730 | 230 | 23 | 42345 | 1.442 | 0.074656 | *TRPC1* |
| ENSG00000172009 | 19 | 2775458 | 2823599 | 108 | 16 | 33898 | 1.4419 | 0.074659 | *THOP1* |
| ENSG00000059915 | 10 | 104152376 | 104191296 | 61 | 18 | 48456 | 1.4412 | 0.074759 | *PSD* |
| ENSG00000118017 | 3 | 137832560 | 137861229 | 55 | 9 | 44573 | 1.4412 | 0.074766 | *A4GNT* |
| ENSG00000255103 | 1 | 39866151 | 39892154 | 46 | 12 | 47698 | 1.4405 | 0.074863 | *KIAA0754* |
| ENSG00000105447 | 19 | 48939030 | 48970279 | 92 | 16 | 29304 | 1.4402 | 0.074905 | *GRWD1* |
| ENSG00000177479 | 3 | 48946254 | 49033815 | 110 | 18 | 38479 | 1.4402 | 0.074912 | *ARIH2* |
| ENSG00000167633 | 19 | 55225969 | 55388448 | 419 | 47 | 37172 | 1.4396 | 0.074988 | *KIR3DL1* |
| ENSG00000081014 | 15 | 51190869 | 51308097 | 327 | 23 | 54552 | 1.4393 | 0.075035 | *AP4E1* |
| ENSG00000185347 | 14 | 105946192 | 105975912 | 54 | 7 | 29757 | 1.4391 | 0.075066 | *C14orf80* |
| ENSG00000145050 | 3 | 51412478 | 51436828 | 18 | 8 | 50862 | 1.439 | 0.075074 | *MANF* |
| ENSG00000168509 | 1 | 145403095 | 145427545 | 20 | 9 | 44165 | 1.439 | 0.075076 | *HFE2* |
| ENSG00000156973 | 2 | 232587135 | 232660982 | 211 | 20 | 53105 | 1.4389 | 0.075084 | *PDE6D* |
| ENSG00000147592 | 8 | 71537553 | 71591409 | 189 | 21 | 34507 | 1.4388 | 0.075109 | *LACTB2* |
| ENSG00000164749 | 8 | 76310149 | 76489078 | 787 | 64 | 42684 | 1.4387 | 0.075115 | *HNF4G* |
| ENSG00000100890 | 14 | 35581052 | 35753271 | 622 | 52 | 48914 | 1.4385 | 0.075146 | *KIAA0391* |
| ENSG00000177370 | 17 | 890357 | 916911 | 70 | 15 | 44675 | 1.4381 | 0.0752 | *TIMM22* |
| ENSG00000139865 | 14 | 38055052 | 38520647 | 1177 | 108 | 45994 | 1.4378 | 0.075251 | *TTC6* |
| ENSG00000140795 | 16 | 46730891 | 46834319 | 95 | 18 | 37343 | 1.4374 | 0.075305 | *MYLK3* |
| ENSG00000122735 | 9 | 34447412 | 34530982 | 161 | 22 | 51590 | 1.4368 | 0.075388 | *DNAI1* |
| ENSG00000182557 | 17 | 4326983 | 4401503 | 408 | 64 | 50990 | 1.4367 | 0.0754 | *SPNS3* |
| ENSG00000131871 | 15 | 101801022 | 101827705 | 84 | 17 | 34197 | 1.4366 | 0.075414 | *VIMP* |
| ENSG00000178150 | 19 | 48665575 | 48800865 | 672 | 81 | 46674 | 1.4365 | 0.07543 | *ZNF114* |
| ENSG00000174156 | 6 | 52751437 | 52784483 | 115 | 15 | 33955 | 1.4364 | 0.07544 | *GSTA3* |
| ENSG00000114904 | 3 | 52734800 | 52814965 | 179 | 16 | 48780 | 1.4364 | 0.075444 | *NEK4* |
| ENSG00000224867 | 1 | 220593286 | 220618023 | 92 | 13 | 46976 | 1.4362 | 0.075472 | *AC096644.1* |
| ENSG00000213937 | 16 | 3052457 | 3074506 | 63 | 18 | 50253 | 1.4356 | 0.075564 | *CLDN9* |

| | | | | | | | | | |
|---|---|---|---|---|---|---|---|---|---|
| ENSG00000136319 | 14 | 20714717 | 20784153 | 245 | 42 | 49642 | 1.4355 | 0.075566 | *TTC5* |
| ENSG00000126218 | 13 | 113767128 | 113813843 | 82 | 26 | 39054 | 1.4355 | 0.075568 | *F10* |
| ENSG00000137501 | 11 | 85395267 | 85532184 | 358 | 35 | 49204 | 1.4351 | 0.07563 | *SYTL2* |
| ENSG00000004766 | 7 | 92851653 | 92998338 | 255 | 45 | 41898 | 1.4348 | 0.075674 | *CCDC132* |
| ENSG00000086504 | 16 | 407384 | 430527 | 159 | 23 | 39896 | 1.4343 | 0.075739 | *MRPL28* |
| ENSG00000266302 | 17 | 16583575 | 16717767 | 125 | 18 | 33851 | 1.4335 | 0.075853 | *RP11-219A15.1* |
| ENSG00000099840 | 19 | 2086380 | 2109592 | 67 | 16 | 48251 | 1.4335 | 0.075863 | *IZUMO4* |
| ENSG00000130254 | 19 | 5577010 | 5634057 | 150 | 23 | 49702 | 1.4334 | 0.075878 | *SAFB2* |
| ENSG00000265818 | 6 | 8005959 | 8112763 | 473 | 43 | 44944 | 1.4332 | 0.075897 | *EEF1E1-BLOC1S5* |
| ENSG00000124541 | 6 | 42979383 | 43011894 | 85 | 16 | 36950 | 1.4332 | 0.075907 | *RRP36* |
| ENSG00000241233 | 11 | 71239071 | 71260253 | 99 | 8 | 49504 | 1.4322 | 0.07605 | *KRTAP5-8* |
| ENSG00000166816 | 16 | 75135758 | 75160669 | 86 | 20 | 42715 | 1.4317 | 0.07612 | *LDHD* |
| ENSG00000144045 | 2 | 74735258 | 74763463 | 16 | 8 | 51888 | 1.4315 | 0.076143 | *DQX1* |
| ENSG00000136114 | 13 | 52941305 | 52990629 | 153 | 14 | 51465 | 1.4311 | 0.076194 | *THSD1* |
| ENSG00000122257 | 16 | 24539014 | 24594184 | 188 | 18 | 51551 | 1.4309 | 0.076233 | *RBBP6* |
| ENSG00000114268 | 3 | 48545117 | 48609448 | 80 | 19 | 39496 | 1.4308 | 0.076237 | *PFKFB4* |
| ENSG00000153774 | 16 | 75317596 | 75477383 | 637 | 23 | 48165 | 1.4308 | 0.076249 | *CFDP1* |
| ENSG00000183463 | 13 | 28541851 | 28572791 | 88 | 16 | 41612 | 1.4304 | 0.076302 | *URAD* |
| ENSG00000178980 | 19 | 48271829 | 48297943 | 86 | 22 | 48169 | 1.4302 | 0.07633 | *SEPW1* |
| ENSG00000234719 | 16 | 12011230 | 12080500 | 279 | 38 | 33734 | 1.43 | 0.076366 | *RP11-166B2.1* |
| ENSG00000166928 | 11 | 60136003 | 60195161 | 175 | 21 | 51440 | 1.4294 | 0.076445 | *MS4A14* |
| ENSG00000269531 | 12 | 80188742 | 80212321 | 31 | 9 | 26176 | 1.4294 | 0.076449 | *AC073569.1* |
| ENSG00000153234 | 2 | 157170944 | 157208860 | 49 | 11 | 35949 | 1.429 | 0.076497 | *NR4A2* |
| ENSG00000161013 | 5 | 179214597 | 179243952 | 114 | 28 | 46564 | 1.429 | 0.076508 | *MGAT4B* |
| ENSG00000156990 | 3 | 9869533 | 9896286 | 79 | 16 | 37313 | 1.4288 | 0.076527 | *RPUSD3* |
| ENSG00000170160 | 17 | 16582851 | 16717767 | 126 | 18 | 33684 | 1.4288 | 0.076532 | *CCDC144A* |
| ENSG00000185000 | 8 | 145529954 | 145560573 | 42 | 12 | 43708 | 1.4286 | 0.076554 | *DGAT1* |
| ENSG00000155561 | 7 | 135232667 | 135343505 | 560 | 35 | 50797 | 1.4286 | 0.076558 | *NUP205* |
| ENSG00000131773 | 8 | 136459700 | 136678965 | 558 | 50 | 45138 | 1.4282 | 0.076621 | *KHDRBS3* |
| ENSG00000077150 | 10 | 104143867 | 104172281 | 57 | 16 | 46497 | 1.4281 | 0.076628 | *NFKB2* |
| ENSG00000174942 | 11 | 56174734 | 56195708 | 109 | 14 | 46822 | 1.4281 | 0.076633 | *OR5R1* |
| ENSG00000144331 | 2 | 180296709 | 180736232 | 1768 | 176 | 50356 | 1.428 | 0.076647 | *ZNF385B* |
| ENSG00000268301 | 12 | 11177087 | 11197275 | 93 | 11 | 32240 | 1.4276 | 0.0767 | *AC018630.1* |
| ENSG00000165458 | 11 | 71924745 | 71960149 | 72 | 16 | 47486 | 1.4276 | 0.076704 | *INPPL1* |
| ENSG00000169682 | 16 | 28975542 | 29005869 | 53 | 17 | 41004 | 1.4272 | 0.076756 | *SPNS1* |
| ENSG00000103160 | 16 | 84145886 | 84188797 | 150 | 32 | 52805 | 1.4272 | 0.076765 | *HSDL1* |

| | | | | | | | | | |
|---|---|---|---|---|---|---|---|---|---|
| ENSG00000198258 | 19 | 9928568 | 9950791 | 52 | 16 | 36983 | 1.4267 | 0.076827 | *UBL5* |
| ENSG00000178206 | 2 | 132034330 | 132060127 | 47 | 11 | 36887 | 1.4266 | 0.07685 | *CYP4F31P* |
| ENSG00000178999 | 17 | 8098056 | 8123918 | 90 | 18 | 47224 | 1.4263 | 0.076897 | *AURKB* |
| ENSG00000181371 | 11 | 56247825 | 56268871 | 88 | 13 | 51812 | 1.426 | 0.076937 | *OR5M8* |
| ENSG00000181016 | 7 | 112110908 | 112141117 | 107 | 20 | 43707 | 1.4259 | 0.076943 | *LSMEM1* |
| ENSG00000183098 | 13 | 93869095 | 95069655 | 3705 | 323 | 46159 | 1.4257 | 0.076983 | *GPC6* |
| ENSG00000171132 | 2 | 45868484 | 46425129 | 2132 | 243 | 46996 | 1.4252 | 0.077054 | *PRKCE* |
| ENSG00000129226 | 17 | 7472785 | 7495429 | 50 | 12 | 46227 | 1.4251 | 0.07706 | *CD68* |
| ENSG00000006125 | 17 | 33895065 | 34063436 | 568 | 44 | 54703 | 1.4251 | 0.077063 | *AP2B1* |
| ENSG00000150471 | 4 | 62056976 | 62954053 | 2274 | 211 | 41931 | 1.4247 | 0.077125 | *LPHN3* |
| ENSG00000267824 | 16 | 20489024 | 20510981 | 36 | 9 | 33586 | 1.424 | 0.077218 | *AC137056.1* |
| ENSG00000213676 | 6 | 32055953 | 32106030 | 136 | 38 | 36961 | 1.424 | 0.077225 | *ATF6B* |
| ENSG00000170185 | 4 | 144096070 | 144154983 | 138 | 12 | 55910 | 1.424 | 0.077229 | *USP38* |
| ENSG00000139266 | 12 | 58138881 | 58164190 | 28 | 11 | 42066 | 1.4239 | 0.077239 | *Mar-09* |
| ENSG00000196284 | 6 | 44767054 | 45355690 | 2250 | 56 | 48445 | 1.4237 | 0.07727 | *SUPT3H* |
| ENSG00000068028 | 3 | 50357219 | 50388411 | 19 | 8 | 52022 | 1.4236 | 0.077275 | *RASSF1* |
| ENSG00000186115 | 19 | 15978833 | 16018930 | 250 | 22 | 54478 | 1.4235 | 0.077298 | *CYP4F2* |
| ENSG00000044459 | 9 | 17124980 | 17513921 | 1465 | 52 | 34415 | 1.4228 | 0.077398 | *CNTLN* |
| ENSG00000122034 | 13 | 27988681 | 28019958 | 149 | 20 | 40172 | 1.4227 | 0.077409 | *GTF3A* |
| ENSG00000197548 | 3 | 11303995 | 11609139 | 803 | 54 | 45398 | 1.4225 | 0.077447 | *ATG7* |
| ENSG00000109065 | 17 | 72756686 | 72782506 | 81 | 10 | 41383 | 1.4224 | 0.077456 | *NAT9* |
| ENSG00000105662 | 19 | 18784487 | 18903004 | 336 | 52 | 48250 | 1.4223 | 0.07747 | *CRTC1* |
| ENSG00000184661 | 8 | 25306513 | 25375436 | 198 | 15 | 47811 | 1.4222 | 0.077478 | *CDCA2* |
| ENSG00000162188 | 11 | 62465130 | 62486673 | 43 | 9 | 54484 | 1.4222 | 0.077489 | *GNG3* |
| ENSG00000099899 | 22 | 20089389 | 20114915 | 92 | 18 | 51399 | 1.4221 | 0.077497 | *TRMT2A* |
| ENSG00000204287 | 6 | 32397619 | 32422823 | 486 | 26 | 42534 | 1.4218 | 0.077538 | *HLA-DRA* |
| ENSG00000077157 | 1 | 202307827 | 202571834 | 387 | 23 | 41763 | 1.4217 | 0.077551 | *PPP1R12B* |
| ENSG00000135446 | 12 | 58131510 | 58159796 | 30 | 15 | 41571 | 1.4214 | 0.077593 | *CDK4* |
| ENSG00000117906 | 15 | 77213960 | 77252601 | 89 | 23 | 42736 | 1.4213 | 0.07761 | *RCN2* |
| ENSG00000183103 | 19 | 42331148 | 42358760 | 44 | 7 | 38520 | 1.4211 | 0.077643 | *LYPD4* |
| ENSG00000172340 | 3 | 67400884 | 67715038 | 1343 | 125 | 46310 | 1.4207 | 0.077705 | *SUCLG2* |
| ENSG00000100365 | 22 | 37247030 | 37284057 | 137 | 26 | 38209 | 1.4206 | 0.077718 | *NCF4* |
| ENSG00000172830 | 11 | 67060919 | 67090078 | 41 | 13 | 34664 | 1.4202 | 0.07778 | *SSH3* |
| ENSG00000205629 | 16 | 25113050 | 25199552 | 186 | 16 | 53262 | 1.42 | 0.077802 | *LCMT1* |
| ENSG00000082438 | 2 | 165500134 | 165710189 | 684 | 58 | 43720 | 1.4199 | 0.077819 | *COBLL1* |
| ENSG00000164120 | 4 | 175401328 | 175454305 | 208 | 38 | 50097 | 1.4197 | 0.077846 | *HPGD* |

| | | | | | | | | | |
|---|---|---|---|---|---|---|---|---|---|
| ENSG00000258941 | 14 | 39693106 | 39830397 | 418 | 34 | 44464 | 1.4194 | 0.077893 | *RP11-407N17.3* |
| ENSG00000069248 | 1 | 229567045 | 229654103 | 197 | 14 | 46859 | 1.4191 | 0.077933 | *NUP133* |
| ENSG00000196248 | 11 | 123837368 | 123858488 | 87 | 14 | 52649 | 1.4184 | 0.07803 | *OR10S1* |
| ENSG00000143545 | 1 | 153944127 | 153968834 | 42 | 14 | 52170 | 1.4184 | 0.078031 | *RAB13* |
| ENSG00000106080 | 7 | 30040203 | 30076300 | 79 | 11 | 55619 | 1.4184 | 0.078042 | *FKBP14* |
| ENSG00000179165 | 6 | 36348328 | 36420666 | 257 | 22 | 36395 | 1.4176 | 0.07816 | *PXT1* |
| ENSG00000104938 | 19 | 7818035 | 7844491 | 124 | 20 | 44407 | 1.4175 | 0.078165 | *CLEC4M* |
| ENSG00000025796 | 6 | 108178960 | 108289393 | 226 | 21 | 48555 | 1.4175 | 0.078171 | *SEC63* |
| ENSG00000154438 | 7 | 116993276 | 117078177 | 117 | 20 | 36959 | 1.4175 | 0.078172 | *ASZ1* |
| ENSG00000177465 | 14 | 74048410 | 74073200 | 123 | 23 | 37807 | 1.4174 | 0.078186 | *ACOT4* |
| ENSG00000205143 | 9 | 34611376 | 34638104 | 49 | 12 | 51145 | 1.4172 | 0.078212 | *ARID3C* |
| ENSG00000253958 | 8 | 8549448 | 8571616 | 143 | 6 | 55196 | 1.4169 | 0.078253 | *CLDN23* |
| ENSG00000012061 | 19 | 45900591 | 45992086 | 287 | 48 | 45439 | 1.4167 | 0.078285 | *ERCC1* |
| ENSG00000168936 | 4 | 1707679 | 1733085 | 133 | 16 | 46363 | 1.4163 | 0.078342 | *TMEM129* |
| ENSG00000149792 | 11 | 64879655 | 64904843 | 41 | 13 | 45375 | 1.4163 | 0.078347 | *MRPL49* |
| ENSG00000183317 | 1 | 38169552 | 38240805 | 289 | 35 | 50471 | 1.4156 | 0.07844 | *EPHA10* |
| ENSG00000140470 | 15 | 100501794 | 100892210 | 2129 | 228 | 41948 | 1.4156 | 0.078453 | *ADAMTS17* |
| ENSG00000185885 | 11 | 303506 | 325272 | 71 | 22 | 36653 | 1.4154 | 0.078477 | *IFITM1* |
| ENSG00000106236 | 7 | 98236609 | 98269180 | 68 | 19 | 40661 | 1.4151 | 0.078519 | *NPTX2* |
| ENSG00000144278 | 2 | 154718426 | 155320361 | 1988 | 135 | 45245 | 1.4145 | 0.078604 | *GALNT13* |
| ENSG00000075651 | 3 | 171308195 | 171538740 | 732 | 45 | 42053 | 1.4145 | 0.078605 | *PLD1* |
| ENSG00000067225 | 15 | 72481370 | 72534164 | 95 | 21 | 47123 | 1.4143 | 0.07863 | *PKM* |
| ENSG00000111737 | 12 | 120522899 | 120565306 | 105 | 23 | 36432 | 1.4142 | 0.078658 | *RAB35* |
| ENSG00000166159 | 12 | 1919433 | 1955918 | 121 | 39 | 49548 | 1.4141 | 0.07866 | *LRTM2* |
| ENSG00000267303 | 19 | 10416147 | 10441354 | 102 | 19 | 51222 | 1.4139 | 0.078702 | *CTD-2369P2.12* |
| ENSG00000138658 | 4 | 113450492 | 113568151 | 265 | 28 | 48059 | 1.4128 | 0.078856 | *C4orf21* |
| ENSG00000132781 | 1 | 45784835 | 45816142 | 44 | 13 | 48211 | 1.4128 | 0.078858 | *MUTYH* |
| ENSG00000162408 | 1 | 6571407 | 6624595 | 195 | 9 | 36916 | 1.4128 | 0.078859 | *NOL9* |
| ENSG00000105668 | 19 | 36147715 | 36179367 | 139 | 17 | 47149 | 1.4127 | 0.078873 | *UPK1A* |
| ENSG00000171793 | 1 | 41435007 | 41488235 | 169 | 30 | 50603 | 1.4126 | 0.07888 | *CTPS1* |
| ENSG00000139725 | 12 | 122205664 | 122251309 | 124 | 18 | 36668 | 1.4125 | 0.078896 | *RHOF* |
| ENSG00000169885 | 1 | 1836266 | 1858735 | 56 | 10 | 36943 | 1.4123 | 0.078928 | *CALML6* |
| ENSG00000158296 | 20 | 45176463 | 45314714 | 366 | 81 | 43917 | 1.4122 | 0.078952 | *SLC13A3* |
| ENSG00000172890 | 11 | 71154155 | 71245153 | 244 | 21 | 23568 | 1.4121 | 0.078963 | *NADSYN1* |
| ENSG00000187595 | 17 | 40167594 | 40227383 | 115 | 18 | 38592 | 1.4121 | 0.078965 | *ZNF385C* |
| ENSG00000135763 | 1 | 229751981 | 229805946 | 111 | 13 | 52579 | 1.412 | 0.078982 | *URB2* |

| | | | | | | | | | |
|---|---|---|---|---|---|---|---|---|---|
| ENSG00000186364 | 1 | 145576115 | 145599439 | 25 | 10 | 43915 | 1.4119 | 0.078984 | *NUDT17* |
| ENSG00000132773 | 1 | 45795342 | 45819647 | 43 | 11 | 47059 | 1.4117 | 0.079026 | *TOE1* |
| ENSG00000268635 | 11 | 77174416 | 77198074 | 62 | 15 | 45464 | 1.4114 | 0.079057 | *DKFZP434E1119* |
| ENSG00000162222 | 11 | 62485541 | 62517765 | 69 | 10 | 51077 | 1.4113 | 0.07908 | *TTC9C* |
| ENSG00000160396 | 19 | 40875178 | 40906094 | 69 | 21 | 49196 | 1.4112 | 0.079088 | *HIPK4* |
| ENSG00000257341 | 14 | 105943536 | 105968604 | 25 | 10 | 28693 | 1.4112 | 0.079089 | *CRIP1* |
| ENSG00000121270 | 16 | 48190821 | 48291479 | 285 | 16 | 33813 | 1.4111 | 0.079113 | *ABCC11* |
| ENSG00000122679 | 7 | 45187390 | 45235901 | 332 | 22 | 30836 | 1.4107 | 0.079173 | *RAMP3* |
| ENSG00000104774 | 19 | 12747325 | 12787556 | 70 | 19 | 35266 | 1.4106 | 0.079175 | *MAN2B1* |
| ENSG00000258674 | 19 | 19617036 | 19656885 | 93 | 10 | 46860 | 1.4101 | 0.079253 | *CTC-260F20.3* |
| ENSG00000129465 | 14 | 24795227 | 24819251 | 105 | 17 | 34141 | 1.4101 | 0.07926 | *RIPK3* |
| ENSG00000165171 | 7 | 73238920 | 73266865 | 77 | 4 | 21791 | 1.41 | 0.079265 | *WBSCR27* |
| ENSG00000186792 | 3 | 50320262 | 50346899 | 26 | 10 | 48910 | 1.4099 | 0.079291 | *HYAL3* |
| ENSG00000077097 | 3 | 25629475 | 25716398 | 193 | 18 | 48890 | 1.4098 | 0.079297 | *TOP2B* |
| ENSG00000163346 | 1 | 154906552 | 154938599 | 55 | 14 | 39213 | 1.4094 | 0.079353 | *PBXIP1* |
| ENSG00000168421 | 4 | 40182673 | 40258587 | 259 | 59 | 43613 | 1.4092 | 0.079381 | *RHOH* |
| ENSG00000185028 | 5 | 181626 | 205468 | 122 | 21 | 33437 | 1.4092 | 0.079394 | *LRRC14B* |
| ENSG00000093217 | 3 | 38378251 | 38472839 | 335 | 33 | 33090 | 1.4091 | 0.079406 | *XYLB* |
| ENSG00000122180 | 1 | 203042260 | 203065164 | 58 | 19 | 46091 | 1.4088 | 0.079441 | *MYOG* |
| ENSG00000212928 | 14 | 57661888 | 57683291 | 43 | 5 | 26092 | 1.4088 | 0.079445 | *AL391152.1* |
| ENSG00000197603 | 5 | 37096330 | 37259530 | 225 | 24 | 46575 | 1.4087 | 0.079455 | *C5orf42* |
| ENSG00000134824 | 11 | 61550452 | 61644826 | 215 | 29 | 42649 | 1.4081 | 0.079551 | *FADS2* |
| ENSG00000186603 | 1 | 45782545 | 45804347 | 24 | 11 | 53468 | 1.4078 | 0.079591 | *HPDL* |
| ENSG00000182771 | 10 | 87349312 | 88136250 | 2565 | 246 | 43196 | 1.4071 | 0.079694 | *GRID1* |
| ENSG00000198429 | 19 | 11988599 | 12035144 | 137 | 21 | 36267 | 1.4071 | 0.079701 | *ZNF69* |
| ENSG00000135631 | 2 | 73290510 | 73393849 | 335 | 31 | 37618 | 1.4066 | 0.079769 | *RAB11FIP5* |
| ENSG00000143851 | 1 | 202106141 | 202140716 | 122 | 35 | 48591 | 1.4064 | 0.079797 | *PTPN7* |
| ENSG00000243477 | 3 | 50323833 | 50346852 | 25 | 10 | 48561 | 1.4051 | 0.079997 | *NAT6* |
| ENSG00000139990 | 14 | 69507598 | 69629867 | 230 | 27 | 41266 | 1.4049 | 0.080027 | *DCAF5* |
| ENSG00000089123 | 20 | 13236709 | 13629587 | 1198 | 87 | 45878 | 1.4048 | 0.080039 | *TASP1* |
| ENSG00000178385 | 2 | 208683027 | 208900284 | 436 | 56 | 45012 | 1.4045 | 0.080081 | *PLEKHM3* |
| ENSG00000130703 | 20 | 60803580 | 60881268 | 238 | 26 | 46083 | 1.4043 | 0.080111 | *OSBPL2* |
| ENSG00000250565 | 2 | 46707889 | 46779696 | 247 | 38 | 42695 | 1.4038 | 0.080195 | *ATP6V1E2* |
| ENSG00000135913 | 2 | 219304974 | 219443084 | 286 | 19 | 46222 | 1.4037 | 0.080197 | *USP37* |
| ENSG00000185201 | 11 | 297631 | 325272 | 92 | 30 | 35365 | 1.4036 | 0.080215 | *IFITM2* |
| ENSG00000181074 | 11 | 5765923 | 5786959 | 163 | 13 | 50830 | 1.4036 | 0.080218 | *OR52N4* |

| | | | | | | | | | |
|---|---|---|---|---|---|---|---|---|---|
| ENSG00000146776 | 7 | 105235514 | 105527050 | 857 | 163 | 44240 | 1.4033 | 0.080264 | *ATXN7L1* |
| ENSG00000153086 | 2 | 135586117 | 135669604 | 102 | 14 | 30679 | 1.4032 | 0.080277 | *ACMSD* |
| ENSG00000204310 | 6 | 32125989 | 32155873 | 98 | 22 | 46276 | 1.4028 | 0.080345 | *AGPAT1* |
| ENSG00000189320 | 7 | 135403096 | 135443594 | 149 | 29 | 49519 | 1.4025 | 0.080389 | *FAM180A* |
| ENSG00000162585 | 1 | 2105903 | 2154159 | 111 | 30 | 45531 | 1.4024 | 0.080394 | *C1orf86* |
| ENSG00000079459 | 8 | 11643082 | 11706818 | 439 | 55 | 48493 | 1.4013 | 0.080565 | *FDFT1* |
| ENSG00000089692 | 12 | 6871678 | 6897621 | 47 | 19 | 46433 | 1.4012 | 0.080574 | *LAG3* |
| ENSG00000100604 | 14 | 93379425 | 93411638 | 131 | 27 | 54494 | 1.4009 | 0.080618 | *CHGA* |
| ENSG00000139624 | 12 | 50513575 | 50571288 | 114 | 11 | 31880 | 1.4008 | 0.080641 | *CERS5* |
| ENSG00000152977 | 3 | 147101209 | 147238080 | 336 | 31 | 54971 | 1.4007 | 0.080659 | *ZIC1* |
| ENSG00000103653 | 15 | 75064398 | 75105539 | 86 | 16 | 28890 | 1.3998 | 0.080783 | *CSK* |
| ENSG00000151917 | 6 | 56809773 | 56902140 | 202 | 16 | 49017 | 1.3994 | 0.080844 | *BEND6* |
| ENSG00000124657 | 6 | 27915019 | 27935960 | 33 | 13 | 40230 | 1.3994 | 0.08085 | *OR2B6* |
| ENSG00000086159 | 12 | 50350977 | 50380922 | 44 | 16 | 31028 | 1.3993 | 0.080857 | *AQP6* |
| ENSG00000153560 | 3 | 33419828 | 33492863 | 255 | 22 | 40329 | 1.3992 | 0.08087 | *UBP1* |
| ENSG00000205883 | 8 | 11829830 | 11852099 | 92 | 20 | 42381 | 1.3991 | 0.080886 | *DEFB135* |
| ENSG00000173110 | 1 | 161484036 | 161506681 | 71 | 16 | 41600 | 1.3991 | 0.080898 | *HSPA6* |
| ENSG00000205086 | 2 | 42152508 | 42191406 | 120 | 24 | 46651 | 1.399 | 0.080908 | *C2orf91* |
| ENSG00000164093 | 4 | 111528579 | 111573279 | 80 | 15 | 53319 | 1.3989 | 0.080919 | *PITX2* |
| ENSG00000171530 | 5 | 76976991 | 77174604 | 781 | 64 | 41547 | 1.3989 | 0.080926 | *TBCA* |
| ENSG00000010803 | 1 | 41482872 | 41717826 | 706 | 43 | 50446 | 1.3988 | 0.080943 | *SCMH1* |
| ENSG00000152785 | 4 | 81942119 | 81988685 | 114 | 19 | 35821 | 1.3987 | 0.080946 | *BMP3* |
| ENSG00000204498 | 6 | 31504647 | 31536606 | 205 | 35 | 49227 | 1.3986 | 0.080969 | *NFKBIL1* |
| ENSG00000197454 | 1 | 248175250 | 248196188 | 53 | 9 | 42083 | 1.3984 | 0.080999 | *OR2L5* |
| ENSG00000011405 | 11 | 17089277 | 17239530 | 299 | 29 | 43959 | 1.3982 | 0.081022 | *PIK3C2A* |
| ENSG00000250151 | 3 | 9824766 | 9887085 | 195 | 23 | 41393 | 1.3982 | 0.081033 | *ARPC4-TTLL3* |
| ENSG00000164512 | 5 | 55385507 | 55539186 | 518 | 70 | 38343 | 1.3981 | 0.081042 | *ANKRD55* |
| ENSG00000198242 | 17 | 27036411 | 27061377 | 43 | 9 | 46621 | 1.3975 | 0.081135 | *RPL23A* |
| ENSG00000170946 | 11 | 31381387 | 31463396 | 137 | 17 | 47723 | 1.3971 | 0.081194 | *DNAJC24* |
| ENSG00000104731 | 16 | 87720091 | 87809598 | 465 | 51 | 47505 | 1.3966 | 0.081268 | *KLHDC4* |
| ENSG00000127603 | 1 | 39536988 | 39962849 | 937 | 70 | 45203 | 1.3952 | 0.081485 | *MACF1* |
| ENSG00000196569 | 6 | 129194342 | 129847714 | 1979 | 148 | 42580 | 1.3951 | 0.081485 | *LAMA2* |
| ENSG00000184454 | 1 | 24872602 | 24945819 | 358 | 51 | 48406 | 1.395 | 0.081505 | *NCMAP* |
| ENSG00000108312 | 17 | 42272401 | 42308994 | 78 | 16 | 42642 | 1.3946 | 0.081563 | *UBTF* |
| ENSG00000182393 | 19 | 39776965 | 39799313 | 81 | 9 | 32809 | 1.3946 | 0.081567 | *IFNL1* |
| ENSG00000272333 | 19 | 36198921 | 36239779 | 104 | 20 | 50924 | 1.3946 | 0.081575 | *KMT2B* |

| | | | | | | | | | |
|---|---|---|---|---|---|---|---|---|---|
| ENSG00000163938 | 3 | 52705172 | 52738508 | 69 | 8 | 50841 | 1.3945 | 0.081584 | *GNL3* |
| ENSG00000163155 | 1 | 151122224 | 151148424 | 30 | 13 | 38550 | 1.394 | 0.081658 | *LYSMD1* |
| ENSG00000188686 | 8 | 145480549 | 145502470 | 4 | 2 | 50616 | 1.3939 | 0.081675 | *SCXA* |
| ENSG00000180881 | 12 | 75659759 | 75794708 | 283 | 33 | 49109 | 1.3936 | 0.081717 | *CAPS2* |
| ENSG00000155886 | 9 | 19497450 | 19796926 | 959 | 124 | 42952 | 1.3936 | 0.081724 | *SLC24A2* |
| ENSG00000188624 | 19 | 46613328 | 46637931 | 57 | 15 | 50671 | 1.3935 | 0.081741 | *IGFL3* |
| ENSG00000171877 | 15 | 44152962 | 44497450 | 462 | 53 | 33384 | 1.3933 | 0.081763 | *FRMD5* |
| ENSG00000089057 | 20 | 4823002 | 5000939 | 519 | 91 | 44385 | 1.3927 | 0.081858 | *SLC23A2* |
| ENSG00000138604 | 15 | 69442923 | 69574556 | 261 | 23 | 40878 | 1.3923 | 0.081913 | *GLCE* |
| ENSG00000166483 | 11 | 9585228 | 9625004 | 113 | 22 | 43804 | 1.3923 | 0.081915 | *WEE1* |
| ENSG00000173137 | 8 | 145586790 | 145628457 | 65 | 11 | 34916 | 1.392 | 0.081955 | *ADCK5* |
| ENSG00000136536 | 2 | 160559000 | 160637538 | 356 | 28 | 48070 | 1.3918 | 0.081993 | *Mar-07* |
| ENSG00000100038 | 22 | 22301397 | 22347213 | 142 | 19 | 49636 | 1.3917 | 0.082002 | *TOP3B* |
| ENSG00000013810 | 4 | 1713227 | 1756898 | 247 | 22 | 44315 | 1.3911 | 0.082098 | *TACC3* |
| ENSG00000166946 | 15 | 43467316 | 43497396 | 30 | 12 | 48081 | 1.3908 | 0.082137 | *CCNDBP1* |
| ENSG00000186790 | 1 | 47871744 | 47893723 | 51 | 10 | 37702 | 1.3908 | 0.082149 | *FOXE3* |
| ENSG00000137821 | 15 | 71135578 | 71352414 | 294 | 34 | 47652 | 1.3902 | 0.082234 | *LRRC49* |
| ENSG00000149761 | 11 | 63983413 | 64008158 | 62 | 17 | 48193 | 1.3901 | 0.082246 | *NUDT22* |
| ENSG00000125255 | 13 | 103686350 | 103729196 | 240 | 39 | 53055 | 1.3901 | 0.082256 | *SLC10A2* |
| ENSG00000137204 | 6 | 43253432 | 43283276 | 60 | 14 | 39503 | 1.3898 | 0.08229 | *SLC22A7* |
| ENSG00000124215 | 20 | 58523471 | 58619066 | 251 | 44 | 44793 | 1.3898 | 0.082297 | *CDH26* |
| ENSG00000221947 | 8 | 71571600 | 71712606 | 562 | 25 | 33273 | 1.3896 | 0.082329 | *XKR9* |
| ENSG00000163684 | 3 | 58281974 | 58320422 | 196 | 15 | 33736 | 1.3889 | 0.082435 | *RPP14* |
| ENSG00000115137 | 2 | 25156505 | 25204963 | 116 | 18 | 42855 | 1.3888 | 0.082444 | *DNAJC27* |
| ENSG00000164758 | 8 | 118522952 | 118562501 | 128 | 23 | 54502 | 1.3886 | 0.082474 | *MED30* |
| ENSG00000150527 | 14 | 39724488 | 39866156 | 421 | 33 | 43290 | 1.3886 | 0.082474 | *CTAGE5* |
| ENSG00000089154 | 12 | 120555007 | 120642513 | 132 | 14 | 28726 | 1.3884 | 0.082513 | *GCN1L1* |
| ENSG00000087274 | 4 | 2835584 | 2941803 | 203 | 29 | 47993 | 1.3882 | 0.082532 | *ADD1* |
| ENSG00000135127 | 12 | 120417673 | 120542298 | 279 | 26 | 33307 | 1.388 | 0.082572 | *CCDC64* |
| ENSG00000109381 | 4 | 139939266 | 140108372 | 355 | 46 | 50244 | 1.3878 | 0.082595 | *ELF2* |
| ENSG00000103260 | 16 | 755115 | 779655 | 78 | 14 | 28222 | 1.3876 | 0.082631 | *METRN* |
| ENSG00000115604 | 2 | 102917989 | 103025218 | 471 | 30 | 44620 | 1.3876 | 0.082633 | *IL18R1* |
| ENSG00000259159 | 11 | 119199652 | 119227368 | 75 | 8 | 50582 | 1.3875 | 0.082651 | *MFRP* |
| ENSG00000223953 | 11 | 119199652 | 119227383 | 75 | 8 | 50582 | 1.3875 | 0.082651 | *C1QTNF5* |
| ENSG00000235718 | 11 | 119199652 | 119227383 | 75 | 8 | 50582 | 1.3875 | 0.082651 | *MFRP* |
| ENSG00000105664 | 19 | 18883583 | 18912123 | 60 | 22 | 48007 | 1.3872 | 0.08269 | *COMP* |

| | | | | | | | | | |
|---|---|---|---|---|---|---|---|---|---|
| ENSG00000229183 | 11 | 60979688 | 61028929 | 21 | 11 | 37037 | 1.3869 | 0.082731 | *PGA4* |
| ENSG00000013725 | 11 | 60729115 | 60797849 | 275 | 50 | 42270 | 1.3868 | 0.082758 | *CD6* |
| ENSG00000104969 | 19 | 2744712 | 2793369 | 128 | 13 | 32008 | 1.3867 | 0.08276 | *SGTA* |
| ENSG00000177432 | 4 | 89607066 | 89629386 | 61 | 14 | 37885 | 1.3864 | 0.082805 | *NAP1L5* |
| ENSG00000215114 | 8 | 59313823 | 59374060 | 188 | 24 | 55514 | 1.3864 | 0.082811 | *UBXN2B* |
| ENSG00000180053 | 8 | 23549964 | 23574111 | 106 | 17 | 46662 | 1.386 | 0.082871 | *NKX2-6* |
| ENSG00000250067 | 19 | 19617036 | 19658390 | 101 | 12 | 45936 | 1.386 | 0.082872 | *YJEFN3* |
| ENSG00000182004 | 1 | 203820731 | 203849678 | 122 | 10 | 56112 | 1.386 | 0.082881 | *SNRPE* |
| ENSG00000114812 | 3 | 42520791 | 42589059 | 202 | 45 | 36194 | 1.3854 | 0.082971 | *VIPR1* |
| ENSG00000166862 | 22 | 36949968 | 37109603 | 482 | 94 | 45557 | 1.3853 | 0.082976 | *CACNG2* |
| ENSG00000164733 | 8 | 11690033 | 11736957 | 268 | 34 | 52111 | 1.3853 | 0.082977 | *CTSB* |
| ENSG00000256188 | 12 | 11275557 | 11297243 | 114 | 8 | 27817 | 1.3852 | 0.08299 | *TAS2R30* |
| ENSG00000197681 | 17 | 36327711 | 36368166 | 10 | 2 | 42859 | 1.3849 | 0.083048 | *TBC1D3* |
| ENSG00000108688 | 17 | 32587240 | 32609261 | 76 | 13 | 53359 | 1.3845 | 0.083102 | *CCL7* |
| ENSG00000058056 | 3 | 179360543 | 179517189 | 424 | 67 | 42631 | 1.3836 | 0.083235 | *USP13* |
| ENSG00000010610 | 12 | 6886024 | 6939974 | 164 | 35 | 40607 | 1.3834 | 0.083269 | *CD4* |
| ENSG00000155097 | 8 | 104023291 | 104095279 | 228 | 33 | 47872 | 1.3833 | 0.08329 | *ATP6V1C1* |
| ENSG00000169991 | 1 | 19220775 | 19293180 | 284 | 47 | 43577 | 1.3822 | 0.083455 | *IFFO2* |
| ENSG00000214212 | 19 | 10937251 | 10990466 | 82 | 15 | 54657 | 1.3818 | 0.083518 | *C19orf38* |
| ENSG00000117791 | 1 | 220911567 | 220968150 | 173 | 19 | 30983 | 1.3815 | 0.083562 | *Mar-02* |
| ENSG00000109113 | 17 | 27031299 | 27055447 | 38 | 8 | 48701 | 1.3812 | 0.083605 | *RAB34* |
| ENSG00000182053 | 11 | 49040504 | 49069579 | 130 | 11 | 22744 | 1.3811 | 0.08363 | *TRIM49B* |
| ENSG00000076554 | 8 | 80860571 | 81153467 | 849 | 70 | 43202 | 1.381 | 0.083638 | *TPD52* |
| ENSG00000157168 | 8 | 31486902 | 32632548 | 3860 | 259 | 45780 | 1.3807 | 0.083681 | *NRG1* |
| ENSG00000011454 | 9 | 125693112 | 125877145 | 258 | 37 | 46973 | 1.3807 | 0.083682 | *RABGAP1* |
| ENSG00000148935 | 11 | 22637188 | 22844601 | 477 | 63 | 40525 | 1.3804 | 0.083728 | *GAS2* |
| ENSG00000115041 | 2 | 95953052 | 96061825 | 139 | 16 | 51282 | 1.3802 | 0.083767 | *KCNIP3* |
| ENSG00000131864 | 19 | 57620506 | 57653294 | 99 | 13 | 46665 | 1.3798 | 0.083825 | *USP29* |
| ENSG00000168131 | 6 | 27868963 | 27890174 | 60 | 17 | 42833 | 1.3794 | 0.083888 | *OR2B2* |
| ENSG00000248485 | 1 | 161218517 | 161265240 | 204 | 27 | 47697 | 1.3791 | 0.083929 | *PCP4L1* |
| ENSG00000177239 | 9 | 139971379 | 140013635 | 123 | 22 | 46262 | 1.3786 | 0.084008 | *MAN1B1* |
| ENSG00000168661 | 19 | 35407807 | 35446074 | 139 | 11 | 37587 | 1.3784 | 0.084048 | *ZNF30* |
| ENSG00000172586 | 10 | 75531805 | 75553410 | 21 | 4 | 45023 | 1.3782 | 0.084078 | *CHCHD1* |
| ENSG00000161526 | 17 | 73653196 | 73714142 | 101 | 21 | 44680 | 1.378 | 0.084104 | *SAP30BP* |
| ENSG00000086015 | 1 | 46242659 | 46511796 | 609 | 22 | 55214 | 1.3779 | 0.084112 | *MAST2* |
| ENSG00000160229 | 19 | 20949110 | 21001922 | 488 | 19 | 55137 | 1.3775 | 0.084183 | *ZNF66* |

| | | | | | | | | | |
|---|---|---|---|---|---|---|---|---|---|
| ENSG00000091136 | 7 | 107554244 | 107653700 | 252 | 49 | 44880 | 1.3774 | 0.084198 | *LAMB1* |
| ENSG00000163206 | 1 | 152840793 | 152867523 | 19 | 9 | 38017 | 1.377 | 0.084257 | *SMCP* |
| ENSG00000188629 | 19 | 9425021 | 9503293 | 250 | 18 | 45434 | 1.3768 | 0.084288 | *ZNF177* |
| ENSG00000160193 | 21 | 44253204 | 44309678 | 322 | 28 | 43327 | 1.3765 | 0.084341 | *WDR4* |
| ENSG00000115525 | 2 | 86056267 | 86126137 | 246 | 22 | 53520 | 1.3764 | 0.084351 | *ST3GAL5* |
| ENSG00000147606 | 8 | 92211722 | 92420378 | 507 | 62 | 48738 | 1.3762 | 0.084375 | *SLC26A7* |
| ENSG00000239264 | 6 | 7871483 | 8036646 | 727 | 85 | 42552 | 1.3761 | 0.084392 | *TXNDC5* |
| ENSG00000149743 | 11 | 63981271 | 64003726 | 48 | 15 | 50449 | 1.3757 | 0.084457 | *TRPT1* |
| ENSG00000270011 | 19 | 9424850 | 9503293 | 251 | 18 | 45482 | 1.3756 | 0.084475 | *ZNF559-ZNF177* |
| ENSG00000101448 | 20 | 44155625 | 44186391 | 105 | 8 | 33397 | 1.3754 | 0.0845 | *EPPIN* |
| ENSG00000197077 | 22 | 25338697 | 25603415 | 779 | 97 | 46110 | 1.3753 | 0.084522 | *KIAA1671* |
| ENSG00000174013 | 3 | 196285482 | 196325930 | 121 | 27 | 43704 | 1.3752 | 0.084535 | *FBXO45* |
| ENSG00000187889 | 1 | 57174477 | 57295369 | 418 | 83 | 44467 | 1.3751 | 0.084548 | *C1orf168* |
| ENSG00000178741 | 15 | 75202132 | 75240509 | 81 | 12 | 31999 | 1.3747 | 0.084614 | *COX5A* |
| ENSG00000164684 | 8 | 81530686 | 81797016 | 432 | 41 | 42961 | 1.3745 | 0.084637 | *ZNF704* |
| ENSG00000196510 | 12 | 110800705 | 110851535 | 161 | 19 | 43063 | 1.3735 | 0.084796 | *ANAPC7* |
| ENSG00000196263 | 19 | 57009212 | 57051590 | 180 | 16 | 55407 | 1.3727 | 0.084924 | *ZNF471* |
| ENSG00000158321 | 7 | 69053905 | 70268054 | 2837 | 191 | 49980 | 1.3723 | 0.084992 | *AUTS2* |
| ENSG00000015676 | 7 | 44408720 | 44540479 | 302 | 15 | 54883 | 1.3721 | 0.085016 | *NUDCD3* |
| ENSG00000197125 | 11 | 124300022 | 124321054 | 69 | 16 | 48411 | 1.372 | 0.085025 | *OR8B8* |
| ENSG00000165752 | 10 | 134010996 | 134155351 | 697 | 98 | 42070 | 1.3716 | 0.085101 | *STK32C* |
| ENSG00000124380 | 2 | 70110692 | 70142707 | 62 | 8 | 32254 | 1.3715 | 0.085106 | *SNRNP27* |
| ENSG00000179218 | 19 | 13039392 | 13065303 | 38 | 13 | 50372 | 1.3715 | 0.085109 | *CALR* |
| ENSG00000168439 | 11 | 63942744 | 63982015 | 105 | 18 | 45915 | 1.3711 | 0.085179 | *STIP1* |
| ENSG00000242616 | 9 | 114413615 | 114442526 | 140 | 19 | 36466 | 1.3707 | 0.085241 | *GNG10* |
| ENSG00000143147 | 1 | 168043997 | 168116821 | 171 | 23 | 49464 | 1.3702 | 0.085306 | *GPR161* |
| ENSG00000144119 | 2 | 119903819 | 119926465 | 76 | 15 | 43689 | 1.3699 | 0.085361 | *C1QL2* |
| ENSG00000137494 | 11 | 82894781 | 82981736 | 219 | 27 | 49867 | 1.3699 | 0.085362 | *ANKRD42* |
| ENSG00000204480 | 1 | 13684889 | 13708405 | 7 | 4 | 25554 | 1.3691 | 0.085481 | *PRAMEF19* |
| ENSG00000162813 | 1 | 220220824 | 220273804 | 137 | 28 | 49280 | 1.369 | 0.085496 | *BPNT1* |
| ENSG00000140400 | 15 | 75638133 | 75670971 | 34 | 11 | 37469 | 1.3688 | 0.085527 | *MAN2C1* |
| ENSG00000144635 | 3 | 32557463 | 32622366 | 180 | 32 | 51559 | 1.3687 | 0.085547 | *DYNC1LI1* |
| ENSG00000116984 | 1 | 236948610 | 237077281 | 363 | 28 | 42217 | 1.3684 | 0.085598 | *MTR* |
| ENSG00000088448 | 13 | 111520887 | 111577416 | 207 | 23 | 38130 | 1.3682 | 0.085625 | *ANKRD10* |
| ENSG00000022567 | 8 | 142207265 | 142328404 | 464 | 70 | 51180 | 1.3682 | 0.085628 | *SLC45A4* |
| ENSG00000198961 | 5 | 108660410 | 108755695 | 395 | 25 | 41821 | 1.368 | 0.085648 | *PJA2* |

| | | | | | | | | | |
|---|---|---|---|---|---|---|---|---|---|
| ENSG00000164867 | 7 | 150678083 | 150721676 | 95 | 27 | 46304 | 1.3679 | 0.085664 | *NOS3* |
| ENSG00000155229 | 10 | 99208081 | 99268551 | 111 | 18 | 50013 | 1.3674 | 0.08575 | *MMS19* |
| ENSG00000187857 | 12 | 55748895 | 55769833 | 85 | 7 | 39439 | 1.3672 | 0.085775 | *OR6C75* |
| ENSG00000115297 | 2 | 74730590 | 74754274 | 12 | 6 | 57454 | 1.3666 | 0.085877 | *TLX2* |
| ENSG00000100170 | 22 | 32429019 | 32519016 | 273 | 17 | 50353 | 1.3666 | 0.085879 | *SLC5A1* |
| ENSG00000172594 | 6 | 123100315 | 123140865 | 120 | 32 | 38612 | 1.3663 | 0.085922 | *SMPDL3A* |
| ENSG00000180979 | 15 | 42824720 | 42851000 | 46 | 12 | 47302 | 1.3662 | 0.085935 | *LRRC57* |
| ENSG00000083845 | 19 | 58887767 | 58916173 | 85 | 16 | 49236 | 1.3658 | 0.085994 | *RPS5* |
| ENSG00000105549 | 19 | 351750 | 386013 | 148 | 48 | 37674 | 1.3656 | 0.08603 | *THEG* |
| ENSG00000176920 | 19 | 49189228 | 49219207 | 101 | 14 | 27243 | 1.3653 | 0.086087 | *FUT2* |
| ENSG00000048649 | 11 | 77361041 | 77542063 | 444 | 23 | 36350 | 1.3649 | 0.086142 | *RSF1* |
| ENSG00000249109 | 5 | 177199198 | 177231397 | 16 | 6 | 33259 | 1.3649 | 0.086143 | *RP11-1026M7.2* |
| ENSG00000182580 | 3 | 184269572 | 184310197 | 157 | 26 | 52974 | 1.3647 | 0.086172 | *EPHB3* |
| ENSG00000138688 | 4 | 123063488 | 123293913 | 391 | 44 | 45878 | 1.3646 | 0.086185 | *KIAA1109* |
| ENSG00000164751 | 8 | 77882494 | 77923280 | 119 | 22 | 44768 | 1.3646 | 0.086187 | *PEX2* |
| ENSG00000197953 | 3 | 151441704 | 151489127 | 182 | 23 | 53983 | 1.3645 | 0.0862 | *AADACL2* |
| ENSG00000164197 | 5 | 63451671 | 63678696 | 382 | 31 | 52530 | 1.3639 | 0.086304 | *RNF180* |
| ENSG00000066654 | 16 | 20734986 | 20763406 | 80 | 11 | 36217 | 1.3637 | 0.086336 | *THUMPD1* |
| ENSG00000174173 | 3 | 101270706 | 101295290 | 68 | 14 | 48487 | 1.3636 | 0.086348 | *TRMT10C* |
| ENSG00000072134 | 17 | 19108928 | 19250028 | 85 | 18 | 47731 | 1.3634 | 0.086372 | *EPN2* |
| ENSG00000120251 | 4 | 158115334 | 158297227 | 338 | 39 | 49777 | 1.3634 | 0.086377 | *GRIA2* |
| ENSG00000115839 | 2 | 135799835 | 135943964 | 221 | 18 | 27243 | 1.3633 | 0.086392 | *RAB3GAP1* |
| ENSG00000140285 | 15 | 49705293 | 49790972 | 306 | 29 | 38484 | 1.3627 | 0.086484 | *FGF7* |
| ENSG00000154889 | 18 | 11872621 | 11919222 | 229 | 44 | 35890 | 1.3626 | 0.086507 | *MPPE1* |
| ENSG00000115307 | 2 | 74743772 | 74767066 | 14 | 8 | 51102 | 1.3626 | 0.08651 | *AUP1* |
| ENSG00000146670 | 11 | 64823772 | 64861636 | 97 | 23 | 43079 | 1.3624 | 0.086543 | *CDCA5* |
| ENSG00000166206 | 15 | 26778693 | 27194686 | 1493 | 149 | 41046 | 1.3623 | 0.086559 | *GABRB3* |
| ENSG00000162571 | 1 | 1099264 | 1143315 | 220 | 31 | 37152 | 1.3621 | 0.086579 | *TTLL10* |
| ENSG00000173875 | 19 | 12711732 | 12752735 | 120 | 15 | 32163 | 1.362 | 0.086596 | *ZNF791* |
| ENSG00000105676 | 19 | 19134384 | 19180563 | 93 | 12 | 56498 | 1.3618 | 0.086623 | *ARMC6* |
| ENSG00000120333 | 1 | 174969925 | 175002561 | 70 | 18 | 53001 | 1.3617 | 0.086648 | *MRPS14* |
| ENSG00000114127 | 3 | 142015449 | 142176904 | 276 | 30 | 51038 | 1.3616 | 0.086657 | *XRN1* |
| ENSG00000237412 | 2 | 233375173 | 233400422 | 90 | 21 | 54716 | 1.3615 | 0.086674 | *PRSS56* |
| ENSG00000214736 | 6 | 41745400 | 41767636 | 57 | 14 | 45434 | 1.3613 | 0.086711 | *TOMM6* |
| ENSG00000122133 | 9 | 138443602 | 138468801 | 52 | 18 | 45728 | 1.3612 | 0.086722 | *PAEP* |
| ENSG00000118898 | 16 | 4922508 | 5020742 | 466 | 70 | 43080 | 1.3611 | 0.086739 | *PPL* |

| | | | | | | | | | |
|---|---|---|---|---|---|---|---|---|---|
| ENSG00000204334 | 2 | 171630296 | 171665481 | 122 | 12 | 49501 | 1.3609 | 0.086769 | *ERICH2* |
| ENSG00000253379 | 8 | 72305675 | 72514260 | 515 | 71 | 42890 | 1.3607 | 0.086809 | *RP11-1102P16.1* |
| ENSG00000125637 | 2 | 113904902 | 113976973 | 202 | 48 | 41348 | 1.3607 | 0.086811 | *PSD4* |
| ENSG00000145335 | 4 | 90635250 | 90769466 | 451 | 36 | 35847 | 1.3604 | 0.086852 | *SNCA* |
| ENSG00000126246 | 19 | 36220058 | 36243354 | 69 | 14 | 51099 | 1.3595 | 0.086994 | *IGFLR1* |
| ENSG00000204919 | 13 | 57705052 | 57728073 | 21 | 8 | 41768 | 1.3594 | 0.087004 | *PRR20A* |
| ENSG00000267880 | 19 | 51367552 | 51388040 | 101 | 20 | 44357 | 1.3594 | 0.087014 | *AC037199.1* |
| ENSG00000137814 | 15 | 42831008 | 42872192 | 75 | 14 | 47506 | 1.3591 | 0.087052 | *HAUS2* |
| ENSG00000179988 | 10 | 124703897 | 124767029 | 199 | 24 | 50461 | 1.3591 | 0.087059 | *PSTK* |
| ENSG00000112137 | 6 | 12707893 | 13298645 | 1585 | 131 | 41294 | 1.3589 | 0.087093 | *PHACTR1* |
| ENSG00000110013 | 11 | 124493009 | 124575603 | 125 | 22 | 46299 | 1.3585 | 0.087153 | *SIAE* |
| ENSG00000025156 | 6 | 122710691 | 122764264 | 128 | 15 | 55788 | 1.3581 | 0.087217 | *HSF2* |
| ENSG00000214753 | 11 | 62470102 | 62504821 | 70 | 13 | 53908 | 1.3581 | 0.087224 | *HNRNPUL2* |
| ENSG00000268328 | 2 | 70119482 | 70139841 | 40 | 6 | 32153 | 1.3575 | 0.08731 | *AC019206.1* |
| ENSG00000042062 | 20 | 49192645 | 49318065 | 437 | 62 | 39143 | 1.3574 | 0.08733 | *FAM65C* |
| ENSG00000117984 | 11 | 1763982 | 1795222 | 100 | 16 | 47554 | 1.3572 | 0.087355 | *CTSD* |
| ENSG00000170631 | 8 | 146145744 | 146186274 | 126 | 22 | 42344 | 1.3571 | 0.08738 | *ZNF16* |
| ENSG00000144118 | 2 | 120987640 | 121062289 | 250 | 17 | 46705 | 1.3571 | 0.087382 | *RALB* |
| ENSG00000269591 | 13 | 20258547 | 20279240 | 46 | 10 | 34401 | 1.3569 | 0.08741 | *AL354808.2* |
| ENSG00000258790 | 14 | 35581755 | 35796680 | 786 | 59 | 48357 | 1.3568 | 0.087415 | *KIAA0391* |
| ENSG00000166689 | 11 | 16789842 | 17045990 | 904 | 96 | 42061 | 1.3567 | 0.087434 | *PLEKHA7* |
| ENSG00000182050 | 12 | 86362516 | 87242681 | 2623 | 102 | 48223 | 1.3564 | 0.087484 | *MGAT4C* |
| ENSG00000158955 | 17 | 44900567 | 44974096 | 183 | 29 | 33235 | 1.3563 | 0.087504 | *WNT9B* |
| ENSG00000073111 | 3 | 127307066 | 127351276 | 51 | 20 | 54903 | 1.356 | 0.087552 | *MCM2* |
| ENSG00000196331 | 6 | 27851203 | 27871669 | 51 | 11 | 38778 | 1.3557 | 0.087601 | *HIST1H2BO* |
| ENSG00000196544 | 17 | 8081652 | 8103564 | 66 | 15 | 46999 | 1.3556 | 0.087609 | *C17orf59* |
| ENSG00000145945 | 6 | 3839620 | 3861551 | 84 | 12 | 36494 | 1.3555 | 0.087636 | *FAM50B* |
| ENSG00000176742 | 11 | 5210965 | 5231930 | 133 | 19 | 39088 | 1.3553 | 0.087656 | *OR51V1* |
| ENSG00000170522 | 4 | 110957002 | 111130355 | 675 | 110 | 45055 | 1.3553 | 0.087664 | *ELOVL6* |
| ENSG00000146828 | 7 | 100414442 | 100474631 | 160 | 28 | 46698 | 1.3549 | 0.087727 | *SLC12A9* |
| ENSG00000249139 | 20 | 44155625 | 44186065 | 100 | 8 | 34414 | 1.3545 | 0.087782 | *EPPIN-WFDC6* |
| ENSG00000151576 | 3 | 113714680 | 113817269 | 196 | 30 | 53794 | 1.3537 | 0.087917 | *QTRTD1* |
| ENSG00000197859 | 9 | 136387286 | 136450641 | 109 | 33 | 45872 | 1.3537 | 0.087919 | *ADAMTSL2* |
| ENSG00000203691 | 1 | 225590404 | 225612040 | 63 | 12 | 44029 | 1.3536 | 0.08794 | *AC092811.1* |
| ENSG00000135776 | 1 | 229642329 | 229704442 | 123 | 16 | 51060 | 1.3534 | 0.087969 | *ABCB10* |
| ENSG00000126822 | 14 | 65160820 | 65223610 | 142 | 22 | 52989 | 1.3531 | 0.088017 | *PLEKHG3* |

| | | | | | | | | | |
|---|---|---|---|---|---|---|---|---|---|
| ENSG00000176092 | 1 | 26638350 | 26690621 | 189 | 29 | 45716 | 1.353 | 0.088021 | *AIM1L* |
| ENSG00000145040 | 3 | 48589160 | 48611206 | 32 | 12 | 45931 | 1.3529 | 0.088046 | *UCN2* |
| ENSG00000159409 | 1 | 151664880 | 151699290 | 55 | 11 | 38523 | 1.3528 | 0.088065 | *CELF3* |
| ENSG00000119689 | 14 | 75338594 | 75380448 | 120 | 15 | 41628 | 1.3527 | 0.088079 | *DLST* |
| ENSG00000176531 | 19 | 43969106 | 44019087 | 155 | 25 | 51087 | 1.3526 | 0.088096 | *PHLDB3* |
| ENSG00000148795 | 10 | 104580288 | 104607290 | 76 | 14 | 46425 | 1.3517 | 0.088237 | *CYP17A1* |
| ENSG00000121797 | 3 | 46438654 | 46464488 | 121 | 14 | 26825 | 1.3515 | 0.088265 | *CCRL2* |
| ENSG00000162267 | 3 | 52818784 | 52853025 | 80 | 13 | 51124 | 1.3515 | 0.088268 | *ITIH3* |
| ENSG00000172016 | 2 | 79374132 | 79396879 | 90 | 14 | 53328 | 1.3514 | 0.088285 | *REG3A* |
| ENSG00000185324 | 16 | 89737145 | 89772772 | 146 | 22 | 40017 | 1.3509 | 0.088356 | *CDK10* |
| ENSG00000108576 | 17 | 28511337 | 28573020 | 113 | 25 | 38146 | 1.3507 | 0.08839 | *SLC6A4* |
| ENSG00000267748 | 19 | 38769791 | 38805550 | 114 | 18 | 54022 | 1.3507 | 0.088402 | *CTB-102L5.4* |
| ENSG00000014123 | 6 | 96959471 | 97013152 | 91 | 16 | 50553 | 1.3506 | 0.088408 | *UFL1* |
| ENSG00000185483 | 1 | 64229693 | 64657181 | 1031 | 97 | 41962 | 1.3502 | 0.088481 | *ROR1* |
| ENSG00000167333 | 11 | 4609902 | 4639489 | 98 | 15 | 52671 | 1.3495 | 0.088588 | *TRIM68* |
| ENSG00000131914 | 1 | 26727269 | 26766213 | 98 | 18 | 33162 | 1.3493 | 0.088623 | *LIN28A* |
| ENSG00000006715 | 7 | 38752563 | 38981994 | 831 | 61 | 38519 | 1.3493 | 0.088625 | *VPS41* |
| ENSG00000088386 | 13 | 99326055 | 99414908 | 336 | 73 | 38665 | 1.3492 | 0.088642 | *SLC15A1* |
| ENSG00000229292 | 19 | 56270507 | 56294545 | 58 | 18 | 42588 | 1.349 | 0.088662 | *RFPL4AL1* |
| ENSG00000196110 | 19 | 9394951 | 9430514 | 101 | 18 | 40796 | 1.3489 | 0.088683 | *ZNF699* |
| ENSG00000120054 | 10 | 101791950 | 101851634 | 175 | 28 | 32692 | 1.3488 | 0.088697 | *CPN1* |
| ENSG00000165388 | 10 | 48345024 | 48383866 | 51 | 18 | 47066 | 1.3487 | 0.088712 | *ZNF488* |
| ENSG00000160323 | 9 | 136269478 | 136334508 | 262 | 31 | 47839 | 1.3487 | 0.088713 | *ADAMTS13* |
| ENSG00000182183 | 1 | 53089016 | 53145355 | 153 | 32 | 44291 | 1.3486 | 0.088726 | *FAM159A* |
| ENSG00000163874 | 1 | 37930153 | 37959980 | 73 | 17 | 42289 | 1.3486 | 0.088735 | *ZC3H12A* |
| ENSG00000140398 | 15 | 75629296 | 75657592 | 42 | 15 | 41566 | 1.3485 | 0.088745 | *NEIL1* |
| ENSG00000268996 | 9 | 139970807 | 139991121 | 37 | 13 | 42511 | 1.3483 | 0.088783 | *AL807752.1* |
| ENSG00000198939 | 5 | 178312895 | 178370213 | 268 | 26 | 53630 | 1.3483 | 0.088788 | *ZFP2* |
| ENSG00000092853 | 1 | 36175819 | 36245568 | 60 | 15 | 43689 | 1.3482 | 0.0888 | *CLSPN* |
| ENSG00000182308 | 4 | 41973713 | 41998476 | 107 | 14 | 42491 | 1.3482 | 0.088802 | *DCAF4L1* |
| ENSG00000135245 | 7 | 128085903 | 128108472 | 89 | 22 | 50616 | 1.348 | 0.088829 | *HILPDA* |
| ENSG00000136488 | 17 | 61962275 | 61984021 | 77 | 14 | 27487 | 1.3475 | 0.088912 | *CSH1* |
| ENSG00000122678 | 7 | 44101846 | 44132139 | 73 | 8 | 53041 | 1.3472 | 0.088957 | *POLM* |
| ENSG00000133315 | 11 | 63756030 | 63943578 | 436 | 74 | 40391 | 1.3472 | 0.088959 | *MACROD1* |
| ENSG00000109851 | 11 | 20167701 | 20192159 | 90 | 31 | 51079 | 1.3471 | 0.088978 | *DBX1* |
| ENSG00000188186 | 7 | 99736530 | 99763567 | 41 | 8 | 50713 | 1.3466 | 0.08905 | *LAMTOR4* |

| | | | | | | | | | |
|---|---|---|---|---|---|---|---|---|---|
| ENSG00000232810 | 6 | 31533344 | 31556113 | 91 | 24 | 48766 | 1.3466 | 0.089051 | *TNF* |
| ENSG00000133020 | 17 | 10283639 | 10335267 | 89 | 11 | 29617 | 1.3466 | 0.089054 | *MYH8* |
| ENSG00000041515 | 13 | 109238500 | 109870355 | 2756 | 252 | 45314 | 1.3465 | 0.089068 | *MYO16* |
| ENSG00000157379 | 14 | 24749804 | 24779039 | 97 | 15 | 36300 | 1.3465 | 0.089073 | *DHRS1* |
| ENSG00000117682 | 1 | 26748773 | 26807785 | 116 | 20 | 41242 | 1.3462 | 0.089113 | *DHDDS* |
| ENSG00000164265 | 5 | 147240245 | 147271754 | 209 | 25 | 44510 | 1.3462 | 0.089118 | *SCGB3A2* |
| ENSG00000108691 | 17 | 32572304 | 32594222 | 57 | 8 | 55369 | 1.3462 | 0.089127 | *CCL2* |
| ENSG00000122786 | 7 | 134419003 | 134665479 | 746 | 94 | 43585 | 1.3461 | 0.089139 | *CALD1* |
| ENSG00000205330 | 12 | 55704346 | 55725408 | 72 | 12 | 45539 | 1.3459 | 0.089163 | *OR6C1* |
| ENSG00000163832 | 3 | 47527130 | 47565251 | 80 | 17 | 42844 | 1.3458 | 0.089191 | *ELP6* |
| ENSG00000181035 | 19 | 19164808 | 19233697 | 159 | 20 | 55693 | 1.3455 | 0.089237 | *SLC25A42* |
| ENSG00000120915 | 8 | 27338296 | 27413081 | 231 | 28 | 42763 | 1.345 | 0.089306 | *EPHX2* |
| ENSG00000166575 | 11 | 86738886 | 87044800 | 1764 | 71 | 45122 | 1.345 | 0.089306 | *TMEM135* |
| ENSG00000103005 | 16 | 58023450 | 58065522 | 95 | 15 | 54058 | 1.3449 | 0.089336 | *USB1* |
| ENSG00000108091 | 10 | 61538521 | 61676414 | 553 | 42 | 45204 | 1.3448 | 0.089346 | *CCDC6* |
| ENSG00000163946 | 3 | 56644161 | 56727265 | 372 | 28 | 38532 | 1.3448 | 0.089352 | *FAM208A* |
| ENSG00000120211 | 9 | 5221419 | 5245304 | 50 | 8 | 47018 | 1.3442 | 0.089441 | *INSL4* |
| ENSG00000065057 | 16 | 2079816 | 2107867 | 59 | 21 | 40462 | 1.3441 | 0.089455 | *NTHL1* |
| ENSG00000137193 | 6 | 37127979 | 37153202 | 56 | 14 | 49499 | 1.344 | 0.089478 | *PIM1* |
| ENSG00000198879 | 10 | 7190586 | 7463450 | 1091 | 117 | 48818 | 1.344 | 0.089479 | *SFMBT2* |
| ENSG00000204659 | 5 | 179095584 | 179117975 | 69 | 12 | 47281 | 1.3437 | 0.089519 | *CBY3* |
| ENSG00000084676 | 2 | 24704783 | 25003571 | 762 | 58 | 50339 | 1.3437 | 0.089531 | *NCOA1* |
| ENSG00000254901 | 19 | 19277712 | 19313400 | 130 | 15 | 51121 | 1.3431 | 0.089619 | *MEF2BNB* |
| ENSG00000205978 | 14 | 24857992 | 24898494 | 114 | 37 | 45481 | 1.3425 | 0.089722 | *NYNRIN* |
| ENSG00000172967 | 22 | 17254302 | 17312589 | 160 | 19 | 48694 | 1.3425 | 0.089722 | *XKR3* |
| ENSG00000110011 | 11 | 63987750 | 64011758 | 56 | 16 | 46906 | 1.3423 | 0.089744 | *DNAJC4* |
| ENSG00000136960 | 8 | 120559326 | 120695693 | 369 | 57 | 44629 | 1.3423 | 0.089756 | *ENPP2* |
| ENSG00000156510 | 10 | 70970059 | 71037315 | 266 | 51 | 40587 | 1.3422 | 0.089768 | *HKDC1* |
| ENSG00000141068 | 17 | 25773670 | 25963461 | 422 | 51 | 50985 | 1.3421 | 0.089778 | *KSR1* |
| ENSG00000180104 | 5 | 433273 | 482052 | 258 | 23 | 30442 | 1.3418 | 0.089824 | *EXOC3* |
| ENSG00000198515 | 4 | 47927994 | 48028689 | 385 | 30 | 37374 | 1.3417 | 0.089849 | *CNGA1* |
| ENSG00000164877 | 7 | 1458101 | 1509138 | 179 | 24 | 42183 | 1.3416 | 0.089859 | *MICALL2* |
| ENSG00000254598 | 11 | 11363489 | 11384904 | 58 | 18 | 42160 | 1.3411 | 0.089937 | *CSNK2A3* |
| ENSG00000136100 | 13 | 52976737 | 53034763 | 149 | 14 | 51955 | 1.341 | 0.089968 | *VPS36* |
| ENSG00000120471 | 11 | 128794626 | 128823040 | 106 | 28 | 43784 | 1.3408 | 0.089995 | *TP53AIP1* |
| ENSG00000007372 | 11 | 31796340 | 31849509 | 84 | 15 | 48348 | 1.3406 | 0.09002 | *PAX6* |

| | | | | | | | | | |
|---|---|---|---|---|---|---|---|---|---|
| ENSG00000006015 | 19 | 18689495 | 18713146 | 80 | 15 | 37904 | 1.3406 | 0.090022 | *C19orf60* |
| ENSG00000196150 | 8 | 146066632 | 146137553 | 157 | 21 | 43378 | 1.3406 | 0.090029 | *ZNF250* |
| ENSG00000119596 | 14 | 75220069 | 75332244 | 222 | 18 | 40596 | 1.3405 | 0.090037 | *YLPM1* |
| ENSG00000113048 | 5 | 71505236 | 71626473 | 191 | 28 | 39448 | 1.3402 | 0.090091 | *MRPS27* |
| ENSG00000185681 | 9 | 124912190 | 124972367 | 160 | 35 | 36837 | 1.3402 | 0.090095 | *MORN5* |
| ENSG00000105695 | 19 | 35773028 | 35814707 | 101 | 29 | 47518 | 1.3401 | 0.090106 | *MAG* |
| ENSG00000183484 | 14 | 105505728 | 105541782 | 84 | 10 | 26911 | 1.3378 | 0.090479 | *GPR132* |
| ENSG00000183597 | 22 | 19994537 | 20063449 | 263 | 42 | 39875 | 1.3377 | 0.090502 | *TANGO2* |
| ENSG00000150201 | 10 | 43857090 | 43881783 | 86 | 20 | 32854 | 1.3371 | 0.090589 | *FXYD4* |
| ENSG00000113263 | 5 | 156559944 | 156692201 | 515 | 60 | 48848 | 1.3371 | 0.09059 | *ITK* |
| ENSG00000005189 | 16 | 20807751 | 20870987 | 97 | 19 | 40734 | 1.337 | 0.090615 | *AC004381.6* |
| ENSG00000157554 | 21 | 39741949 | 40043704 | 1102 | 147 | 43587 | 1.3367 | 0.090664 | *ERG* |
| ENSG00000139970 | 14 | 60052694 | 60347684 | 667 | 63 | 47241 | 1.3366 | 0.090674 | *RTN1* |
| ENSG00000144021 | 2 | 96921870 | 96949087 | 34 | 10 | 40089 | 1.3364 | 0.09071 | *CIAO1* |
| ENSG00000145649 | 5 | 54388476 | 54416080 | 51 | 10 | 47369 | 1.3359 | 0.090785 | *GZMA* |
| ENSG00000119946 | 10 | 101078856 | 101164087 | 215 | 33 | 38164 | 1.3356 | 0.090844 | *CNNM1* |
| ENSG00000255275 | 1 | 19165767 | 19257615 | 464 | 89 | 47198 | 1.3354 | 0.090878 | *RP13-279N23.2* |
| ENSG00000205246 | 19 | 23935807 | 24020937 | 187 | 27 | 47628 | 1.3353 | 0.090895 | *RPSAP58* |
| ENSG00000137824 | 15 | 41018082 | 41058049 | 87 | 14 | 51884 | 1.3349 | 0.090946 | *RMDN3* |
| ENSG00000172987 | 10 | 100208875 | 101005619 | 2283 | 104 | 38864 | 1.3347 | 0.090982 | *HPSE2* |
| ENSG00000144580 | 2 | 219423303 | 219471803 | 124 | 11 | 45423 | 1.3345 | 0.091025 | *RQCD1* |
| ENSG00000182993 | 12 | 14946506 | 15069520 | 346 | 22 | 52602 | 1.3342 | 0.091062 | *C12orf60* |
| ENSG00000112214 | 6 | 97000424 | 97074512 | 239 | 23 | 53067 | 1.3341 | 0.091082 | *FHL5* |
| ENSG00000066422 | 3 | 101357733 | 101406339 | 105 | 13 | 54233 | 1.334 | 0.091102 | *ZBTB11* |
| ENSG00000144460 | 2 | 226255364 | 226528734 | 544 | 59 | 43060 | 1.3338 | 0.091127 | *NYAP2* |
| ENSG00000151322 | 14 | 33394139 | 34283382 | 2971 | 366 | 45497 | 1.3337 | 0.091144 | *NPAS3* |
| ENSG00000178821 | 1 | 1839029 | 1860712 | 54 | 9 | 37002 | 1.3335 | 0.091189 | *TMEM52* |
| ENSG00000164283 | 5 | 54263692 | 54328499 | 229 | 29 | 39103 | 1.3334 | 0.091204 | *ESM1* |
| ENSG00000113732 | 5 | 172400760 | 172472448 | 198 | 37 | 38976 | 1.333 | 0.091261 | *ATP6V0E1* |
| ENSG00000134245 | 1 | 112999163 | 113082787 | 222 | 48 | 45757 | 1.333 | 0.09127 | *WNT2B* |
| ENSG00000106333 | 7 | 100189800 | 100215798 | 38 | 9 | 49861 | 1.3329 | 0.091284 | *PCOLCE* |
| ENSG00000255374 | 12 | 11233886 | 11254912 | 144 | 11 | 38281 | 1.3326 | 0.091339 | *TAS2R43* |
| ENSG00000008256 | 7 | 6191407 | 6322275 | 344 | 45 | 39452 | 1.3324 | 0.091358 | *CYTH3* |
| ENSG00000143537 | 1 | 155013042 | 155045252 | 46 | 16 | 47155 | 1.3323 | 0.091383 | *ADAM15* |
| ENSG00000127561 | 16 | 2029661 | 2054276 | 129 | 15 | 46143 | 1.3323 | 0.091388 | *SYNGR3* |
| ENSG00000160606 | 17 | 27041366 | 27064953 | 39 | 9 | 44727 | 1.3315 | 0.091517 | *TLCD1* |

| | | | | | | | | | |
|---|---|---|---|---|---|---|---|---|---|
| ENSG00000149798 | 11 | 65072289 | 65099900 | 46 | 14 | 48185 | 1.3312 | 0.091569 | *CDC42EP2* |
| ENSG00000114771 | 3 | 151521825 | 151556276 | 177 | 18 | 20955 | 1.3311 | 0.09157 | *AADAC* |
| ENSG00000167701 | 8 | 145718356 | 145742557 | 38 | 10 | 43627 | 1.3307 | 0.091647 | *GPT* |
| ENSG00000188394 | 9 | 125786806 | 125807975 | 16 | 9 | 54697 | 1.3306 | 0.091664 | *GPR21* |
| ENSG00000135100 | 12 | 121406346 | 121450315 | 180 | 28 | 50766 | 1.3303 | 0.091712 | *HNF1A* |
| ENSG00000125533 | 20 | 61627331 | 61648387 | 67 | 24 | 48282 | 1.3302 | 0.091729 | *BHLHE23* |
| ENSG00000110328 | 11 | 11282423 | 11653552 | 1761 | 246 | 44256 | 1.3295 | 0.091847 | *GALNT18* |
| ENSG00000166170 | 14 | 104012881 | 104039168 | 61 | 10 | 51295 | 1.3293 | 0.091874 | *BAG5* |
| ENSG00000175274 | 11 | 44897454 | 44982840 | 266 | 58 | 40739 | 1.3291 | 0.0919 | *TP53I11* |
| ENSG00000085382 | 6 | 105165968 | 105317794 | 420 | 29 | 43351 | 1.3291 | 0.09191 | *HACE1* |
| ENSG00000103254 | 16 | 760581 | 782601 | 75 | 13 | 28829 | 1.3288 | 0.091951 | *FAM173A* |
| ENSG00000178722 | 5 | 60923535 | 61057590 | 457 | 59 | 46853 | 1.3287 | 0.091973 | *C5orf64* |
| ENSG00000142789 | 1 | 22318149 | 22349032 | 154 | 26 | 28090 | 1.3287 | 0.091979 | *CELA3A* |
| ENSG00000256436 | 12 | 11172986 | 11194006 | 100 | 12 | 34120 | 1.3286 | 0.091992 | *TAS2R31* |
| ENSG00000118707 | 20 | 35191891 | 35232353 | 48 | 21 | 43813 | 1.328 | 0.09209 | *TGIF2* |
| ENSG00000206422 | 18 | 7221123 | 7242045 | 52 | 12 | 40745 | 1.3279 | 0.092098 | *LRRC30* |
| ENSG00000070366 | 17 | 1953133 | 2217065 | 755 | 43 | 38711 | 1.3276 | 0.092162 | *SMG6* |
| ENSG00000185475 | 11 | 62544887 | 62567877 | 30 | 8 | 37944 | 1.3275 | 0.092172 | *TMEM179B* |
| ENSG00000152795 | 4 | 83333717 | 83361294 | 88 | 14 | 45850 | 1.3272 | 0.092229 | *HNRNPDL* |
| ENSG00000116132 | 1 | 170621869 | 170718560 | 201 | 37 | 48226 | 1.3269 | 0.092279 | *PRRX1* |
| ENSG00000164344 | 4 | 187120133 | 187189625 | 315 | 34 | 45452 | 1.3268 | 0.092285 | *KLKB1* |
| ENSG00000124942 | 11 | 62191016 | 62333707 | 315 | 55 | 41780 | 1.3265 | 0.092343 | *AHNAK* |
| ENSG00000115661 | 2 | 220100177 | 220125059 | 52 | 11 | 34756 | 1.3259 | 0.092445 | *STK16* |
| ENSG00000259040 | 6 | 7871755 | 8074597 | 872 | 89 | 43083 | 1.3258 | 0.092445 | *BLOC1S5-TXNDC5* |
| ENSG00000203443 | 3 | 172024218 | 172049513 | 62 | 14 | 47336 | 1.3256 | 0.092479 | *AC092964.1* |
| ENSG00000100938 | 14 | 24691628 | 24718448 | 31 | 11 | 41023 | 1.3248 | 0.092618 | *GMPR2* |
| ENSG00000103742 | 15 | 65663802 | 65725410 | 152 | 32 | 42116 | 1.3238 | 0.092778 | *IGDCC4* |
| ENSG00000136274 | 7 | 45110037 | 45138513 | 123 | 18 | 45824 | 1.3234 | 0.092846 | *NACAD* |
| ENSG00000076662 | 19 | 10434452 | 10460499 | 100 | 21 | 54686 | 1.3234 | 0.092856 | *ICAM3* |
| ENSG00000096384 | 6 | 44204824 | 44231620 | 97 | 24 | 51526 | 1.3234 | 0.092859 | *HSP90AB1* |
| ENSG00000160783 | 1 | 156172784 | 156222874 | 133 | 19 | 45706 | 1.3233 | 0.092875 | *PMF1* |
| ENSG00000185104 | 1 | 50895150 | 51435935 | 826 | 59 | 50883 | 1.3228 | 0.092959 | *FAF1* |
| ENSG00000206199 | 3 | 149468892 | 149696172 | 587 | 58 | 51057 | 1.3225 | 0.093002 | *ANKUB1* |
| ENSG00000129925 | 16 | 410773 | 447113 | 227 | 33 | 35093 | 1.3216 | 0.093144 | *TMEM8A* |
| ENSG00000140650 | 16 | 8872680 | 8953188 | 528 | 49 | 50207 | 1.3214 | 0.093186 | *PMM2* |
| ENSG00000159212 | 21 | 36031688 | 36100525 | 160 | 21 | 50636 | 1.3211 | 0.093239 | *CLIC6* |

| | | | | | | | | | |
|---|---|---|---|---|---|---|---|---|---|
| ENSG00000154027 | 1 | 77737736 | 78035651 | 1207 | 122 | 40262 | 1.321 | 0.093246 | *AK5* |
| ENSG00000169509 | 1 | 152476978 | 152498486 | 65 | 9 | 34658 | 1.321 | 0.093254 | *CRCT1* |
| ENSG00000107614 | 10 | 17174253 | 17254053 | 321 | 18 | 51653 | 1.3206 | 0.093314 | *TRDMT1* |
| ENSG00000214314 | 7 | 99030552 | 99050747 | 40 | 8 | 33865 | 1.32 | 0.093424 | *AC073063.1* |
| ENSG00000088899 | 20 | 3133263 | 3164192 | 100 | 27 | 50221 | 1.3197 | 0.093474 | *LZTS3* |
| ENSG00000173401 | 12 | 75718419 | 75774340 | 113 | 16 | 49504 | 1.3196 | 0.093487 | *GLIPR1L1* |
| ENSG00000108510 | 17 | 60009966 | 60152643 | 196 | 23 | 40609 | 1.3194 | 0.093518 | *MED13* |
| ENSG00000162664 | 1 | 90450671 | 90511090 | 100 | 31 | 44077 | 1.3194 | 0.093519 | *ZNF326* |
| ENSG00000086205 | 11 | 49158187 | 49240222 | 326 | 27 | 41286 | 1.3194 | 0.093525 | *FOLH1* |
| ENSG00000114270 | 3 | 48591506 | 48642700 | 56 | 18 | 46904 | 1.3192 | 0.093556 | *COL7A1* |
| ENSG00000196968 | 10 | 75522049 | 75550009 | 37 | 6 | 44384 | 1.3191 | 0.093561 | *FUT11* |
| ENSG00000055118 | 7 | 150632049 | 150685403 | 134 | 24 | 45719 | 1.3189 | 0.093604 | *KCNH2* |
| ENSG00000267120 | 19 | 36220153 | 36246333 | 72 | 14 | 50133 | 1.3189 | 0.093604 | *AD000671.6* |
| ENSG00000152672 | 2 | 71025775 | 71057732 | 179 | 12 | 48277 | 1.318 | 0.093749 | *CLEC4F* |
| ENSG00000083857 | 4 | 187498937 | 187657876 | 545 | 91 | 43222 | 1.3177 | 0.093802 | *FAT1* |
| ENSG00000137275 | 6 | 3054225 | 3125421 | 131 | 41 | 40382 | 1.3175 | 0.093833 | *RIPK1* |
| ENSG00000134222 | 1 | 109812178 | 109835808 | 88 | 21 | 43533 | 1.3173 | 0.093867 | *PSRC1* |
| ENSG00000179057 | 11 | 18715852 | 18757777 | 147 | 30 | 48918 | 1.3169 | 0.093943 | *IGSF22* |
| ENSG00000179477 | 17 | 7965954 | 8001021 | 111 | 31 | 47960 | 1.3167 | 0.093966 | *ALOX12B* |
| ENSG00000176402 | 7 | 99510892 | 99537243 | 64 | 13 | 41942 | 1.3165 | 0.094009 | *GJC3* |
| ENSG00000100526 | 14 | 54853567 | 54896936 | 128 | 20 | 38246 | 1.3161 | 0.094077 | *CDKN3* |
| ENSG00000149357 | 11 | 71786941 | 71824433 | 82 | 6 | 24254 | 1.316 | 0.09408 | *LAMTOR1* |
| ENSG00000164256 | 5 | 23497264 | 23538706 | 81 | 19 | 38301 | 1.3157 | 0.094131 | *PRDM9* |
| ENSG00000109111 | 17 | 26979109 | 27039697 | 63 | 14 | 51730 | 1.3156 | 0.094162 | *SUPT6H* |
| ENSG00000173213 | 18 | 37225 | 83545 | 107 | 15 | 41947 | 1.3155 | 0.094172 | *RP11-683L23.1* |
| ENSG00000106268 | 7 | 2271857 | 2300781 | 141 | 27 | 47618 | 1.3151 | 0.09424 | *NUDT1* |
| ENSG00000260238 | 1 | 156172773 | 156223123 | 135 | 20 | 45876 | 1.3151 | 0.094246 | *PMF1-BGLAP* |
| ENSG00000171722 | 1 | 162332993 | 162356660 | 91 | 28 | 45320 | 1.3148 | 0.094293 | *C1orf111* |
| ENSG00000106852 | 9 | 124954856 | 125001905 | 77 | 30 | 47457 | 1.3148 | 0.094293 | *LHX6* |
| ENSG00000160752 | 1 | 155268539 | 155300457 | 30 | 6 | 38734 | 1.3142 | 0.094392 | *FDPS* |
| ENSG00000204513 | 1 | 12874468 | 12901264 | 34 | 24 | 36732 | 1.3141 | 0.094402 | *PRAMEF11* |
| ENSG00000077942 | 22 | 45888118 | 46007015 | 474 | 73 | 43628 | 1.3136 | 0.09449 | *FBLN1* |
| ENSG00000246366 | 8 | 71510812 | 71585514 | 247 | 25 | 36989 | 1.3135 | 0.094501 | *RP11-382J12.1* |
| ENSG00000090621 | 1 | 40016488 | 40052462 | 109 | 20 | 41701 | 1.3135 | 0.094512 | *PABPC4* |
| ENSG00000182156 | 17 | 77694681 | 77726021 | 124 | 19 | 45349 | 1.3134 | 0.094524 | *ENPP7* |
| ENSG00000227507 | 6 | 31538302 | 31560299 | 94 | 27 | 45844 | 1.3133 | 0.09454 | *LTB* |

| | | | | | | | | | |
|---|---|---|---|---|---|---|---|---|---|
| ENSG00000221910 | 7 | 143622259 | 143643372 | 82 | 8 | 28475 | 1.3132 | 0.094557 | *OR2F2* |
| ENSG00000166960 | 18 | 30507366 | 31031065 | 1186 | 38 | 39753 | 1.313 | 0.094589 | *CCDC178* |
| ENSG00000187522 | 10 | 14870163 | 14923740 | 88 | 19 | 35920 | 1.3128 | 0.094618 | *HSPA14* |
| ENSG00000222038 | 2 | 131359054 | 131425664 | 25 | 11 | 44310 | 1.3122 | 0.094719 | *POTEJ* |
| ENSG00000116688 | 1 | 12030238 | 12083571 | 161 | 19 | 46250 | 1.3121 | 0.094743 | *MFN2* |
| ENSG00000091651 | 16 | 46713555 | 46742306 | 22 | 6 | 30980 | 1.3121 | 0.094747 | *ORC6* |
| ENSG00000051341 | 3 | 121140278 | 121274853 | 335 | 26 | 48673 | 1.3116 | 0.094833 | *POLQ* |
| ENSG00000167103 | 9 | 130673158 | 130703076 | 64 | 12 | 35001 | 1.3114 | 0.094867 | *PIP5KL1* |
| ENSG00000135924 | 2 | 220133989 | 220161622 | 64 | 16 | 35093 | 1.3107 | 0.094979 | *DNAJB2* |
| ENSG00000268484 | 15 | 96894487 | 96914900 | 49 | 16 | 44917 | 1.3107 | 0.094986 | *AC087477.1* |
| ENSG00000106125 | 7 | 30801033 | 30942002 | 559 | 65 | 37915 | 1.3105 | 0.095006 | *FAM188B* |
| ENSG00000156239 | 21 | 30234513 | 30267693 | 178 | 16 | 41576 | 1.3103 | 0.095043 | *N6AMT1* |
| ENSG00000179002 | 1 | 19156093 | 19196176 | 254 | 42 | 51006 | 1.3095 | 0.095178 | *TAS1R2* |
| ENSG00000155085 | 6 | 109804059 | 110022420 | 793 | 33 | 31622 | 1.3092 | 0.095235 | *AK9* |
| ENSG00000198846 | 8 | 59707977 | 60041767 | 905 | 121 | 46098 | 1.309 | 0.095259 | *TOX* |
| ENSG00000196678 | 16 | 20781515 | 20921671 | 244 | 26 | 40695 | 1.3088 | 0.095296 | *ERI2* |
| ENSG00000185651 | 22 | 21893736 | 21988323 | 131 | 12 | 31385 | 1.3088 | 0.095296 | *UBE2L3* |
| ENSG00000169508 | 13 | 99936784 | 99969659 | 85 | 19 | 41383 | 1.3087 | 0.095322 | *GPR183* |
| ENSG00000134970 | 5 | 114939205 | 114978689 | 114 | 13 | 53913 | 1.3082 | 0.095399 | *TMED7* |
| ENSG00000159459 | 15 | 43225095 | 43408311 | 228 | 29 | 35505 | 1.3081 | 0.095416 | *UBR1* |
| ENSG00000162878 | 2 | 42265160 | 42295668 | 140 | 15 | 45003 | 1.308 | 0.095434 | *PKDCC* |
| ENSG00000197046 | 18 | 43395477 | 43434045 | 87 | 17 | 29728 | 1.3079 | 0.095461 | *SIGLEC15* |
| ENSG00000162413 | 1 | 6640784 | 6684667 | 88 | 17 | 55404 | 1.3073 | 0.095555 | *KLHL21* |
| ENSG00000106993 | 9 | 4669559 | 4718398 | 191 | 16 | 28805 | 1.3066 | 0.09567 | *CDC37L1* |
| ENSG00000165996 | 10 | 17621958 | 17669376 | 230 | 29 | 52733 | 1.3066 | 0.095675 | *PTPLA* |
| ENSG00000149823 | 11 | 64846796 | 64889332 | 71 | 16 | 45456 | 1.3064 | 0.095708 | *VPS51* |
| ENSG00000185615 | 16 | 323152 | 347215 | 109 | 26 | 40715 | 1.3063 | 0.095729 | *PDIA2* |
| ENSG00000167992 | 11 | 61015762 | 61072896 | 42 | 18 | 40896 | 1.3059 | 0.095786 | *VWCE* |
| ENSG00000196405 | 14 | 100427786 | 100620573 | 609 | 65 | 43038 | 1.3058 | 0.09581 | *EVL* |
| ENSG00000187416 | 7 | 103959104 | 104559001 | 2013 | 197 | 46280 | 1.3055 | 0.095857 | *LHFPL3* |
| ENSG00000115226 | 2 | 27704750 | 27728112 | 40 | 7 | 43500 | 1.3055 | 0.095859 | *FNDC4* |
| ENSG00000196616 | 4 | 100216121 | 100252558 | 120 | 23 | 38462 | 1.3052 | 0.095908 | *ADH1B* |
| ENSG00000160087 | 1 | 1179289 | 1219265 | 136 | 24 | 44186 | 1.3051 | 0.095927 | *UBE2J2* |
| ENSG00000103023 | 16 | 58303901 | 58338951 | 199 | 13 | 51846 | 1.3051 | 0.095932 | *PRSS54* |
| ENSG00000170448 | 4 | 47839257 | 47926653 | 253 | 20 | 46877 | 1.305 | 0.095946 | *NFXL1* |
| ENSG00000234857 | 11 | 62447747 | 62504856 | 112 | 13 | 52754 | 1.305 | 0.095946 | *HNRNPUL2-BSCL2* |

| | | | | | | | | | |
|---|---|---|---|---|---|---|---|---|---|
| ENSG00000148634 | 10 | 69671665 | 69845105 | 377 | 37 | 34559 | 1.3049 | 0.095963 | *HERC4* |
| ENSG00000179837 | 3 | 51418731 | 51445330 | 19 | 9 | 55920 | 1.3048 | 0.095978 | *RBM15B* |
| ENSG00000124159 | 20 | 43912085 | 43947169 | 122 | 15 | 44335 | 1.3048 | 0.095988 | *MATN4* |
| ENSG00000163606 | 3 | 112630056 | 112703969 | 377 | 34 | 46339 | 1.3047 | 0.095996 | *CD200R1* |
| ENSG00000261115 | 7 | 140764032 | 141190180 | 1060 | 126 | 42287 | 1.3046 | 0.096013 | *TMEM178B* |
| ENSG00000165272 | 9 | 33431152 | 33457609 | 101 | 24 | 51049 | 1.3046 | 0.09602 | *AQP3* |
| ENSG00000162551 | 1 | 21825858 | 21914905 | 431 | 61 | 45757 | 1.3044 | 0.096051 | *ALPL* |
| ENSG00000151812 | 14 | 58020640 | 58458912 | 1936 | 179 | 47415 | 1.3044 | 0.096052 | *SLC35F4* |
| ENSG00000173548 | 15 | 75930247 | 75964642 | 60 | 16 | 42052 | 1.3037 | 0.096173 | *SNX33* |
| ENSG00000137392 | 6 | 35752759 | 35775088 | 65 | 19 | 32590 | 1.3032 | 0.096248 | *CLPS* |
| ENSG00000058453 | 1 | 17056768 | 17309474 | 179 | 34 | 39976 | 1.3028 | 0.096329 | *CROCC* |
| ENSG00000196502 | 16 | 28606903 | 28644946 | 66 | 18 | 37769 | 1.3027 | 0.096333 | *SULT1A1* |
| ENSG00000153162 | 6 | 7717030 | 7891655 | 658 | 96 | 44441 | 1.3025 | 0.096369 | *BMP6* |
| ENSG00000104205 | 8 | 67614653 | 67784257 | 307 | 21 | 37606 | 1.3022 | 0.096424 | *SGK3* |
| ENSG00000152475 | 19 | 58868985 | 58902427 | 117 | 16 | 50413 | 1.302 | 0.096459 | *ZNF837* |
| ENSG00000146083 | 5 | 175943698 | 175975026 | 80 | 17 | 38382 | 1.3019 | 0.096481 | *RNF44* |
| ENSG00000196652 | 7 | 99092274 | 99142323 | 57 | 16 | 40731 | 1.3016 | 0.09653 | *ZKSCAN5* |
| ENSG00000175544 | 11 | 67209877 | 67236699 | 45 | 14 | 39071 | 1.301 | 0.096623 | *CABP4* |
| ENSG00000197563 | 18 | 59700800 | 59864351 | 691 | 38 | 39511 | 1.3009 | 0.09665 | *PIGN* |
| ENSG00000182986 | 19 | 53357043 | 53410946 | 242 | 33 | 50748 | 1.3007 | 0.096678 | *ZNF320* |
| ENSG00000138180 | 10 | 95246389 | 95298849 | 227 | 30 | 35432 | 1.3006 | 0.096703 | *CEP55* |
| ENSG00000187514 | 2 | 232561605 | 232588251 | 88 | 14 | 46628 | 1.3003 | 0.096746 | *PTMA* |
| ENSG00000175463 | 11 | 67161386 | 67187560 | 43 | 12 | 45784 | 1.2993 | 0.096926 | *TBC1D10C* |
| ENSG00000179292 | 11 | 66049341 | 66074135 | 39 | 15 | 44330 | 1.2987 | 0.097018 | *TMEM151A* |
| ENSG00000164300 | 5 | 79397050 | 79561898 | 704 | 107 | 47030 | 1.2986 | 0.097041 | *SERINC5* |
| ENSG00000118689 | 6 | 108871038 | 109015977 | 250 | 30 | 36718 | 1.298 | 0.097143 | *FOXO3* |
| ENSG00000077522 | 1 | 236839754 | 236937931 | 401 | 60 | 43033 | 1.2977 | 0.097195 | *ACTN2* |
| ENSG00000163939 | 3 | 52569368 | 52729933 | 401 | 21 | 49113 | 1.2977 | 0.0972 | *PBRM1* |
| ENSG00000068383 | 10 | 134341324 | 134606979 | 913 | 71 | 53690 | 1.2977 | 0.0972 | *INPP5A* |
| ENSG00000124224 | 20 | 56796188 | 56894495 | 320 | 30 | 44267 | 1.2972 | 0.097279 | *PPP4R1L* |
| ENSG00000100433 | 14 | 88639113 | 88803251 | 678 | 99 | 43781 | 1.297 | 0.097317 | *KCNK10* |
| ENSG00000065150 | 13 | 98595912 | 98686551 | 153 | 26 | 33570 | 1.2969 | 0.097326 | *IPO5* |
| ENSG00000185803 | 8 | 145567795 | 145594932 | 45 | 12 | 47355 | 1.2965 | 0.097394 | *SLC52A2* |
| ENSG00000167748 | 19 | 51312404 | 51337043 | 91 | 18 | 45828 | 1.2961 | 0.097476 | *KLK1* |
| ENSG00000130294 | 2 | 241643181 | 241769725 | 386 | 66 | 43712 | 1.2952 | 0.097617 | *KIF1A* |
| ENSG00000134339 | 11 | 18250770 | 18280190 | 76 | 24 | 35151 | 1.2948 | 0.097695 | *SAA2* |

| | | | | | | | | | |
|---|---|---|---|---|---|---|---|---|---|
| ENSG00000163347 | 3 | 190013490 | 190050264 | 210 | 34 | 52473 | 1.2948 | 0.097698 | *CLDN1* |
| ENSG00000119522 | 9 | 126131933 | 126702431 | 1558 | 107 | 41278 | 1.2947 | 0.097707 | *DENND1A* |
| ENSG00000137822 | 15 | 43651419 | 43709293 | 69 | 13 | 41850 | 1.2944 | 0.09777 | *TUBGCP4* |
| ENSG00000141664 | 18 | 60180240 | 60264942 | 234 | 30 | 39221 | 1.294 | 0.097825 | *ZCCHC2* |
| ENSG00000138435 | 2 | 175602320 | 175639200 | 97 | 15 | 30607 | 1.2937 | 0.097877 | *CHRNA1* |
| ENSG00000145526 | 5 | 19463060 | 20585982 | 3314 | 207 | 42231 | 1.2937 | 0.097888 | *CDH18* |
| ENSG00000165457 | 11 | 71917645 | 71942994 | 65 | 14 | 48489 | 1.2934 | 0.097936 | *FOLR2* |
| ENSG00000215472 | 18 | 46998051 | 47027956 | 134 | 14 | 54124 | 1.2932 | 0.097976 | *RPL17-C18orf32* |
| ENSG00000106086 | 7 | 30057020 | 30180096 | 335 | 21 | 54471 | 1.293 | 0.098009 | *PLEKHA8* |
| ENSG00000153993 | 7 | 84614869 | 84826171 | 360 | 49 | 45917 | 1.2929 | 0.098026 | *SEMA3D* |
| ENSG00000152223 | 18 | 43417574 | 43557240 | 347 | 34 | 33299 | 1.2926 | 0.098072 | *EPG5* |
| ENSG00000182824 | 22 | 18711427 | 18755407 | 12 | 7 | 42602 | 1.2926 | 0.098078 | *AC008132.1* |
| ENSG00000140943 | 16 | 84077368 | 84160511 | 426 | 57 | 49409 | 1.2923 | 0.098131 | *MBTPS1* |
| ENSG00000160691 | 1 | 154924774 | 154956871 | 32 | 10 | 44113 | 1.2923 | 0.098134 | *SHC1* |
| ENSG00000115866 | 2 | 136654247 | 136753670 | 78 | 6 | 19825 | 1.2922 | 0.098136 | *DARS* |
| ENSG00000149418 | 11 | 130019457 | 130090271 | 222 | 47 | 48756 | 1.2918 | 0.098205 | *ST14* |
| ENSG00000116991 | 1 | 232523711 | 232707304 | 506 | 60 | 44929 | 1.2917 | 0.098233 | *SIPA1L2* |
| ENSG00000243566 | 7 | 76129745 | 76658340 | 1474 | 136 | 37204 | 1.2915 | 0.098265 | *UPK3B* |
| ENSG00000089335 | 19 | 35158544 | 35187302 | 137 | 18 | 54552 | 1.2911 | 0.098328 | *ZNF302* |
| ENSG00000069329 | 16 | 46680054 | 46733430 | 41 | 9 | 35992 | 1.291 | 0.098355 | *VPS35* |
| ENSG00000163435 | 1 | 201967073 | 201996316 | 97 | 18 | 38930 | 1.2908 | 0.098387 | *ELF3* |
| ENSG00000221909 | 7 | 99133931 | 99166159 | 39 | 13 | 40077 | 1.2908 | 0.09839 | *FAM200A* |
| ENSG00000168000 | 11 | 62447747 | 62487317 | 74 | 11 | 52322 | 1.2907 | 0.098412 | *BSCL2* |
| ENSG00000116830 | 1 | 117592925 | 117660075 | 216 | 34 | 40028 | 1.2906 | 0.098424 | *TTF2* |
| ENSG00000122986 | 12 | 111055646 | 111152755 | 217 | 22 | 46348 | 1.2906 | 0.098424 | *HVCN1* |
| ENSG00000188167 | 3 | 33121913 | 33148293 | 68 | 13 | 35961 | 1.2903 | 0.098468 | *TMPPE* |
| ENSG00000185624 | 17 | 79791035 | 79828570 | 104 | 12 | 55133 | 1.2896 | 0.098587 | *P4HB* |
| ENSG00000112658 | 6 | 43129037 | 43159243 | 88 | 16 | 40470 | 1.2896 | 0.098594 | *SRF* |
| ENSG00000035403 | 10 | 75747872 | 75889918 | 276 | 40 | 41707 | 1.2896 | 0.098602 | *VCL* |
| ENSG00000049089 | 1 | 40756159 | 40793488 | 131 | 19 | 34014 | 1.2892 | 0.098663 | *COL9A2* |
| ENSG00000089163 | 12 | 120730119 | 120761052 | 96 | 17 | 47965 | 1.2892 | 0.098664 | *SIRT4* |
| ENSG00000176476 | 16 | 28555236 | 28613111 | 165 | 18 | 40682 | 1.289 | 0.098698 | *CCDC101* |
| ENSG00000069849 | 3 | 141584966 | 141655356 | 283 | 28 | 52694 | 1.2882 | 0.098832 | *ATP1B3* |
| ENSG00000158545 | 16 | 88626789 | 88708374 | 347 | 44 | 52418 | 1.2882 | 0.098837 | *ZC3H18* |
| ENSG00000269120 | 13 | 41101138 | 41121323 | 71 | 17 | 37717 | 1.2882 | 0.098845 | *AL133318.1* |
| ENSG00000183709 | 19 | 39749154 | 39770732 | 61 | 12 | 35484 | 1.2875 | 0.098959 | *IFNL2* |

| | | | | | | | | | |
|---|---|---|---|---|---|---|---|---|---|
| ENSG00000122420 | 1 | 78759568 | 79015434 | 787 | 94 | 46704 | 1.2875 | 0.098968 | *PTGFR* |
| ENSG00000175806 | 8 | 9901778 | 10296401 | 1747 | 152 | 42227 | 1.2874 | 0.09897 | *MSRA* |
| ENSG00000116251 | 1 | 6231329 | 6279449 | 132 | 20 | 30978 | 1.2872 | 0.099012 | *RPL22* |
| ENSG00000152332 | 1 | 162457041 | 162509419 | 152 | 11 | 52251 | 1.2871 | 0.09903 | *UHMK1* |
| ENSG00000100968 | 14 | 24824879 | 24858810 | 120 | 18 | 39227 | 1.2863 | 0.099163 | *NFATC4* |
| ENSG00000075415 | 12 | 98977369 | 99005946 | 84 | 17 | 53934 | 1.2863 | 0.099176 | *SLC25A3* |
| ENSG00000122194 | 6 | 161113270 | 161184347 | 226 | 30 | 39083 | 1.2858 | 0.099254 | *PLG* |
| ENSG00000141582 | 17 | 77796955 | 77823228 | 81 | 23 | 45216 | 1.2856 | 0.099291 | *CBX4* |
| ENSG00000170044 | 3 | 101808088 | 102208685 | 1061 | 71 | 45594 | 1.2854 | 0.099326 | *ZPLD1* |
| ENSG00000133106 | 13 | 43450524 | 43576407 | 393 | 71 | 48047 | 1.2852 | 0.099358 | *EPSTI1* |
| ENSG00000183747 | 16 | 20452783 | 20508991 | 135 | 13 | 31018 | 1.2849 | 0.09942 | *ACSM2A* |
| ENSG00000269339 | 12 | 51609860 | 51630085 | 54 | 10 | 39087 | 1.2849 | 0.099421 | *AC139768.1* |
| ENSG00000101310 | 20 | 18478137 | 18552059 | 302 | 22 | 39961 | 1.2848 | 0.099431 | *SEC23B* |
| ENSG00000169193 | 7 | 23626998 | 23694327 | 116 | 18 | 47610 | 1.2847 | 0.099454 | *CCDC126* |
| ENSG00000151500 | 11 | 134108173 | 134133264 | 71 | 16 | 48870 | 1.2845 | 0.099487 | *THYN1* |
| ENSG00000114378 | 3 | 50327320 | 50359812 | 35 | 15 | 46525 | 1.2842 | 0.099528 | *HYAL1* |
| ENSG00000132763 | 1 | 45955725 | 45986739 | 76 | 8 | 56451 | 1.2833 | 0.099698 | *MMACHC* |
| ENSG00000233224 | 6 | 27850477 | 27870963 | 55 | 12 | 39322 | 1.2831 | 0.099733 | *HIST1H2AM* |
| ENSG00000123737 | 4 | 122712472 | 122748176 | 86 | 14 | 40065 | 1.2828 | 0.099773 | *EXOSC9* |
| ENSG00000158106 | 8 | 144441057 | 144476390 | 112 | 24 | 45870 | 1.2824 | 0.099844 | *RHPN1* |
| ENSG00000162144 | 11 | 61106217 | 61139771 | 15 | 9 | 42439 | 1.2823 | 0.099866 | *CYB561A3* |
| ENSG00000152422 | 5 | 82363317 | 82659606 | 754 | 43 | 39914 | 1.2821 | 0.099909 | *XRCC4* |
| ENSG00000126217 | 13 | 113538692 | 113764053 | 908 | 139 | 48127 | 1.2819 | 0.099939 | *MCF2L* |
| ENSG00000076382 | 17 | 26894588 | 26936297 | 34 | 18 | 51781 | 1.2818 | 0.099952 | *SPAG5* |
| ENSG00000176153 | 14 | 65395870 | 65419531 | 99 | 18 | 38922 | 1.2818 | 0.099964 | *GPX2* |
| ENSG00000265681 | 18 | 47004851 | 47028906 | 111 | 13 | 54383 | 1.2815 | 0.10001 | *RPL17* |
| ENSG00000154342 | 1 | 228184752 | 228258961 | 153 | 21 | 46776 | 1.2811 | 0.10008 | *WNT3A* |
| ENSG00000173818 | 17 | 78378965 | 78421886 | 184 | 35 | 49721 | 1.2811 | 0.10008 | *ENDOV* |
| ENSG00000253276 | 7 | 106287211 | 106311442 | 85 | 26 | 40702 | 1.2806 | 0.10016 | *CCDC71L* |
| ENSG00000182674 | 8 | 73439626 | 73860584 | 1151 | 102 | 42265 | 1.2805 | 0.10018 | *KCNB2* |
| ENSG00000095059 | 19 | 12776531 | 12802716 | 31 | 15 | 45923 | 1.2805 | 0.10019 | *DHPS* |
| ENSG00000163719 | 3 | 9681117 | 9754077 | 135 | 24 | 43905 | 1.2797 | 0.10032 | *MTMR14* |
| ENSG00000109047 | 17 | 9789637 | 9818938 | 149 | 19 | 34772 | 1.2795 | 0.10036 | *RCVRN* |
| ENSG00000177294 | 17 | 6669542 | 6710420 | 178 | 31 | 49831 | 1.2794 | 0.10037 | *FBXO39* |
| ENSG00000100647 | 14 | 70068313 | 70191859 | 201 | 35 | 47377 | 1.2794 | 0.10038 | *KIAA0247* |
| ENSG00000189306 | 22 | 42895974 | 42925808 | 97 | 11 | 44364 | 1.2792 | 0.10041 | *RRP7A* |

| | | | | | | | | | |
|---|---|---|---|---|---|---|---|---|---|
| ENSG00000100523 | 14 | 53500686 | 53630000 | 281 | 24 | 35645 | 1.2791 | 0.10042 | *DDHD1* |
| ENSG00000267964 | 10 | 75521935 | 75542378 | 31 | 6 | 44700 | 1.2791 | 0.10044 | *AC022400.2* |
| ENSG00000028116 | 2 | 58124786 | 58397055 | 619 | 60 | 47285 | 1.279 | 0.10045 | *VRK2* |
| ENSG00000173542 | 4 | 71758043 | 71898166 | 218 | 41 | 34428 | 1.2789 | 0.10047 | *MOB1B* |
| ENSG00000167858 | 17 | 6682452 | 6745080 | 292 | 40 | 52968 | 1.2789 | 0.10047 | *TEKT1* |
| ENSG00000108294 | 17 | 36898989 | 36930484 | 90 | 23 | 44327 | 1.2789 | 0.10047 | *PSMB3* |
| ENSG00000104140 | 15 | 41154412 | 41176487 | 57 | 6 | 35878 | 1.2788 | 0.10048 | *RHOV* |
| ENSG00000106605 | 7 | 43788279 | 43856939 | 160 | 19 | 47170 | 1.2787 | 0.1005 | *BLVRA* |
| ENSG00000115234 | 2 | 27583389 | 27609995 | 47 | 11 | 41894 | 1.2786 | 0.10052 | *SNX17* |
| ENSG00000136108 | 13 | 53019564 | 53060763 | 118 | 12 | 47689 | 1.2784 | 0.10056 | *CKAP2* |
| ENSG00000177683 | 7 | 108184987 | 108220194 | 85 | 13 | 36191 | 1.2777 | 0.10067 | *THAP5* |
| ENSG00000152061 | 1 | 174118548 | 174974445 | 2145 | 49 | 43675 | 1.2777 | 0.10068 | *RABGAP1L* |
| ENSG00000118655 | 1 | 114437763 | 114466708 | 56 | 14 | 41242 | 1.2776 | 0.1007 | *DCLRE1B* |
| ENSG00000074047 | 2 | 121483199 | 121760229 | 704 | 126 | 44506 | 1.2774 | 0.10072 | *GLI2* |
| ENSG00000148288 | 9 | 136018340 | 136049332 | 98 | 25 | 41413 | 1.2774 | 0.10073 | *GBGT1* |
| ENSG00000172572 | 12 | 20512179 | 20847315 | 1279 | 132 | 45074 | 1.2774 | 0.10073 | *PDE3A* |
| ENSG00000175166 | 3 | 184006497 | 184036842 | 66 | 16 | 43749 | 1.2773 | 0.10074 | *PSMD2* |
| ENSG00000134215 | 1 | 108103782 | 108517766 | 1551 | 146 | 48355 | 1.2772 | 0.10077 | *VAV3* |
| ENSG00000105341 | 19 | 41927223 | 41956622 | 71 | 10 | 47442 | 1.277 | 0.1008 | *ATP5SL* |
| ENSG00000038274 | 5 | 162920120 | 162956342 | 115 | 21 | 44302 | 1.2769 | 0.10082 | *MAT2B* |
| ENSG00000187801 | 1 | 40905774 | 40939390 | 137 | 31 | 46983 | 1.2768 | 0.10084 | *ZFP69B* |
| ENSG00000162980 | 2 | 152635498 | 152695006 | 148 | 22 | 40804 | 1.2766 | 0.10087 | *ARL5A* |
| ENSG00000204103 | 20 | 39304488 | 39327880 | 42 | 8 | 47033 | 1.2762 | 0.10094 | *MAFB* |
| ENSG00000111897 | 6 | 122754499 | 122802967 | 135 | 16 | 56419 | 1.276 | 0.10098 | *SERINC1* |
| ENSG00000132139 | 17 | 34061530 | 34089897 | 98 | 17 | 47387 | 1.2752 | 0.10113 | *GAS2L2* |
| ENSG00000170892 | 19 | 54683789 | 54707585 | 84 | 17 | 40091 | 1.2751 | 0.10113 | *TSEN34* |
| ENSG00000022556 | 19 | 55454498 | 55522510 | 382 | 53 | 45338 | 1.2752 | 0.10113 | *NLRP2* |
| ENSG00000154328 | 8 | 11617148 | 11654855 | 235 | 37 | 50363 | 1.275 | 0.10116 | *NEIL2* |
| ENSG00000138801 | 4 | 108501433 | 108651608 | 467 | 43 | 49204 | 1.2749 | 0.10117 | *PAPSS1* |
| ENSG00000122687 | 7 | 2263866 | 2291840 | 128 | 25 | 49166 | 1.2748 | 0.1012 | *FTSJ2* |
| ENSG00000168273 | 3 | 52558029 | 52623253 | 169 | 16 | 48101 | 1.2746 | 0.10122 | *SMIM4* |
| ENSG00000197372 | 19 | 23698433 | 23880022 | 556 | 46 | 47211 | 1.2747 | 0.10122 | *ZNF675* |
| ENSG00000126550 | 4 | 70906119 | 70934562 | 108 | 19 | 49192 | 1.2742 | 0.1013 | *HTN1* |
| ENSG00000100105 | 22 | 31711790 | 31752218 | 88 | 22 | 47353 | 1.2739 | 0.10135 | *PATZ1* |
| ENSG00000132824 | 20 | 43114862 | 43160750 | 87 | 13 | 44746 | 1.2736 | 0.1014 | *SERINC3* |
| ENSG00000169567 | 5 | 130484720 | 130517428 | 90 | 10 | 24813 | 1.2735 | 0.10142 | *HINT1* |

| | | | | | | | | | |
|---|---|---|---|---|---|---|---|---|---|
| ENSG00000113645 | 5 | 167708656 | 167909308 | 689 | 117 | 43404 | 1.2735 | 0.10142 | *WWC1* |
| ENSG00000103066 | 16 | 68269207 | 68304961 | 82 | 14 | 51928 | 1.2735 | 0.10142 | *PLA2G15* |
| ENSG00000172354 | 7 | 100261154 | 100286797 | 68 | 16 | 53265 | 1.2733 | 0.10146 | *GNB2* |
| ENSG00000172551 | 12 | 55214303 | 55262177 | 252 | 23 | 39767 | 1.2732 | 0.10148 | *MUCL1* |
| ENSG00000171867 | 20 | 4656882 | 4692236 | 115 | 27 | 46085 | 1.2732 | 0.10148 | *PRNP* |
| ENSG00000185133 | 22 | 31508717 | 31540682 | 95 | 26 | 42672 | 1.273 | 0.10151 | *INPP5J* |
| ENSG00000140107 | 14 | 100779674 | 100806715 | 63 | 16 | 45222 | 1.2728 | 0.10155 | *SLC25A47* |
| ENSG00000150551 | 2 | 133392426 | 133439152 | 89 | 9 | 36482 | 1.2724 | 0.10162 | *LYPD1* |
| ENSG00000268024 | 17 | 32571900 | 32592306 | 51 | 9 | 55121 | 1.2717 | 0.10175 | *AC005549.3* |
| ENSG00000175764 | 9 | 124574207 | 124865885 | 1130 | 95 | 43169 | 1.2714 | 0.10179 | *TTLL11* |
| ENSG00000139116 | 12 | 39677030 | 39847192 | 307 | 31 | 36750 | 1.2713 | 0.10181 | *KIF21A* |
| ENSG00000172073 | 2 | 88814169 | 88839102 | 111 | 12 | 50379 | 1.2713 | 0.10182 | *TEX37* |
| ENSG00000213658 | 16 | 28986147 | 29012104 | 47 | 20 | 40200 | 1.2709 | 0.10188 | *LAT* |
| ENSG00000180667 | 1 | 207207194 | 207236325 | 40 | 9 | 54167 | 1.2706 | 0.10193 | *YOD1* |
| ENSG00000142279 | 19 | 34961874 | 35007258 | 133 | 31 | 33819 | 1.2705 | 0.10195 | *WTIP* |
| ENSG00000135655 | 12 | 62644119 | 62821211 | 476 | 35 | 55486 | 1.2702 | 0.102 | *USP15* |
| ENSG00000174992 | 16 | 29779561 | 29803096 | 15 | 9 | 45706 | 1.2701 | 0.10203 | *ZG16* |
| ENSG00000119535 | 1 | 36921644 | 36958879 | 116 | 26 | 49138 | 1.2699 | 0.10207 | *CSF3R* |
| ENSG00000268424 | 5 | 101559696 | 101581279 | 93 | 16 | 41670 | 1.2698 | 0.10207 | *AC008948.1* |
| ENSG00000112414 | 6 | 142612991 | 142777403 | 274 | 35 | 41979 | 1.2698 | 0.10208 | *GPR126* |
| ENSG00000163884 | 3 | 126051478 | 126086285 | 72 | 15 | 54466 | 1.2695 | 0.10214 | *KLF15* |
| ENSG00000204335 | 2 | 171561861 | 171584588 | 68 | 11 | 47942 | 1.2694 | 0.10216 | *SP5* |
| ENSG00000198502 | 6 | 32475120 | 32508064 | 59 | 13 | 35958 | 1.2693 | 0.10217 | *HLA-DRB5* |
| ENSG00000164961 | 8 | 126026502 | 126114082 | 274 | 24 | 30195 | 1.2691 | 0.10221 | *KIAA0196* |
| ENSG00000168268 | 3 | 52548386 | 52579070 | 73 | 13 | 45420 | 1.269 | 0.10223 | *NT5DC2* |
| ENSG00000133398 | 5 | 6361994 | 6388707 | 100 | 16 | 57017 | 1.2689 | 0.10224 | *MED10* |
| ENSG00000167815 | 19 | 12897634 | 12922694 | 80 | 11 | 33358 | 1.2687 | 0.10228 | *PRDX2* |
| ENSG00000139974 | 14 | 61437832 | 61560451 | 154 | 30 | 49408 | 1.2684 | 0.10232 | *SLC38A6* |
| ENSG00000110237 | 11 | 73009334 | 73090136 | 195 | 19 | 38403 | 1.2683 | 0.10234 | *ARHGEF17* |
| ENSG00000182979 | 14 | 105876159 | 105947066 | 61 | 33 | 42177 | 1.2679 | 0.10241 | *MTA1* |
| ENSG00000259120 | 17 | 73632323 | 73654057 | 44 | 12 | 44677 | 1.268 | 0.10241 | *SMIM6* |
| ENSG00000165923 | 11 | 47671143 | 47746941 | 142 | 18 | 39349 | 1.2677 | 0.10246 | *AGBL2* |
| ENSG00000151876 | 5 | 41915356 | 41951845 | 57 | 10 | 44238 | 1.2674 | 0.10251 | *FBXO4* |
| ENSG00000095713 | 10 | 99614757 | 99800585 | 336 | 60 | 49946 | 1.2671 | 0.10257 | *CRTAC1* |
| ENSG00000243244 | 2 | 48746522 | 48836025 | 416 | 49 | 48600 | 1.2669 | 0.1026 | *STON1* |
| ENSG00000153015 | 5 | 64054757 | 64324590 | 598 | 35 | 53354 | 1.2667 | 0.10263 | *CWC27* |

| | | | | | | | | | |
|---|---|---|---|---|---|---|---|---|---|
| ENSG00000138381 | 2 | 190516111 | 190545557 | 69 | 13 | 44481 | 1.2667 | 0.10264 | *ASNSD1* |
| ENSG00000111666 | 12 | 102080725 | 102147918 | 228 | 32 | 45875 | 1.2664 | 0.10268 | *CHPT1* |
| ENSG00000240403 | 19 | 55351898 | 55388662 | 181 | 16 | 43918 | 1.2663 | 0.1027 | *KIR3DL2* |
| ENSG00000136244 | 7 | 22755503 | 22781621 | 100 | 19 | 35944 | 1.2661 | 0.10273 | *IL6* |
| ENSG00000110711 | 11 | 67240512 | 67268574 | 35 | 12 | 41728 | 1.2661 | 0.10273 | *AIP* |
| ENSG00000135537 | 6 | 108606098 | 108857999 | 478 | 53 | 41193 | 1.2658 | 0.10278 | *LACE1* |
| ENSG00000166123 | 16 | 46908290 | 46975209 | 80 | 18 | 41993 | 1.2659 | 0.10278 | *GPT2* |
| ENSG00000179029 | 17 | 8066555 | 8089717 | 62 | 14 | 50001 | 1.2655 | 0.10285 | *TMEM107* |
| ENSG00000197594 | 6 | 132119156 | 132226295 | 349 | 60 | 40148 | 1.2654 | 0.10287 | *ENPP1* |
| ENSG00000065000 | 19 | 2090988 | 2174464 | 374 | 46 | 44493 | 1.2653 | 0.10289 | *AP3D1* |
| ENSG00000173692 | 2 | 231911578 | 232047541 | 253 | 30 | 38894 | 1.2652 | 0.10291 | *PSMD1* |
| ENSG00000180318 | 12 | 85663885 | 85705562 | 81 | 14 | 35997 | 1.2651 | 0.10291 | *ALX1* |
| ENSG00000127527 | 19 | 16456050 | 16592896 | 319 | 42 | 46399 | 1.2649 | 0.10296 | *EPS15L1* |
| ENSG00000086232 | 7 | 6051881 | 6108861 | 264 | 32 | 46368 | 1.2647 | 0.10299 | *EIF2AK1* |
| ENSG00000008382 | 19 | 4333524 | 4370083 | 122 | 25 | 51192 | 1.2647 | 0.10299 | *MPND* |
| ENSG00000188428 | 6 | 8003800 | 8074647 | 354 | 26 | 43478 | 1.2645 | 0.10303 | *BLOC1S5* |
| ENSG00000162298 | 11 | 64879252 | 64912004 | 58 | 17 | 45599 | 1.2643 | 0.10306 | *SYVN1* |
| ENSG00000155792 | 8 | 120875957 | 121073152 | 931 | 57 | 39430 | 1.2642 | 0.10309 | *DEPTOR* |
| ENSG00000104973 | 19 | 50311539 | 50352073 | 94 | 20 | 42781 | 1.2638 | 0.10316 | *MED25* |
| ENSG00000146830 | 7 | 100267130 | 100297071 | 64 | 15 | 55845 | 1.2637 | 0.10317 | *GIGYF1* |
| ENSG00000184154 | 11 | 71781382 | 71831828 | 117 | 7 | 23571 | 1.2636 | 0.10319 | *LRTOMT* |
| ENSG00000091583 | 17 | 64198151 | 64262643 | 292 | 37 | 42298 | 1.2626 | 0.10336 | *APOH* |
| ENSG00000170011 | 3 | 39840405 | 40311812 | 1641 | 126 | 40166 | 1.2625 | 0.10338 | *MYRIP* |
| ENSG00000071539 | 5 | 882758 | 929472 | 138 | 15 | 38364 | 1.2622 | 0.10344 | *TRIP13* |
| ENSG00000026297 | 6 | 167332992 | 167380679 | 188 | 30 | 35018 | 1.2616 | 0.10354 | *RNASET2* |
| ENSG00000078804 | 20 | 33282094 | 33311243 | 57 | 8 | 49157 | 1.2616 | 0.10355 | *TP53INP2* |
| ENSG00000243646 | 21 | 34628663 | 34679539 | 165 | 25 | 46918 | 1.2612 | 0.10361 | *IL10RB* |
| ENSG00000125944 | 1 | 23620264 | 23680829 | 75 | 16 | 41772 | 1.2609 | 0.10367 | *HNRNPR* |
| ENSG00000163273 | 2 | 232776530 | 232801113 | 63 | 24 | 47283 | 1.2609 | 0.10367 | *NPPC* |
| ENSG00000155329 | 5 | 132322677 | 132372296 | 145 | 11 | 40804 | 1.2607 | 0.10371 | *ZCCHC10* |
| ENSG00000132017 | 19 | 14053304 | 14082254 | 28 | 13 | 43958 | 1.2605 | 0.10375 | *DCAF15* |
| ENSG00000179163 | 1 | 24161567 | 24204784 | 151 | 8 | 49970 | 1.2604 | 0.10376 | *FUCA1* |
| ENSG00000125485 | 9 | 135458384 | 135555788 | 237 | 32 | 51950 | 1.2601 | 0.10382 | *DDX31* |
| ENSG00000233812 | 15 | 32684108 | 32705493 | 11 | 7 | 40266 | 1.2597 | 0.10388 | *AC139426.2* |
| ENSG00000115540 | 2 | 198370295 | 198428423 | 87 | 15 | 53628 | 1.2597 | 0.1039 | *MOB4* |
| ENSG00000175315 | 11 | 65769312 | 65790976 | 75 | 14 | 42853 | 1.2592 | 0.10398 | *CST6* |

| | | | | | | | | | |
|---|---|---|---|---|---|---|---|---|---|
| ENSG00000142619 | 1 | 17565593 | 17620728 | 256 | 37 | 55875 | 1.2591 | 0.10399 | *PADI3* |
| ENSG00000143816 | 1 | 228096357 | 228145599 | 149 | 22 | 48677 | 1.259 | 0.10401 | *WNT9A* |
| ENSG00000178467 | 3 | 49017319 | 49054587 | 30 | 9 | 37843 | 1.2589 | 0.10404 | *P4HTM* |
| ENSG00000119392 | 9 | 131256979 | 131314567 | 94 | 13 | 34833 | 1.2586 | 0.10408 | *GLE1* |
| ENSG00000182141 | 19 | 21463962 | 21522227 | 196 | 16 | 42116 | 1.2579 | 0.10422 | *ZNF708* |
| ENSG00000188818 | 5 | 700471 | 861101 | 179 | 42 | 40693 | 1.2575 | 0.10428 | *ZDHHC11* |
| ENSG00000177576 | 18 | 46998028 | 47023622 | 112 | 13 | 53678 | 1.2574 | 0.1043 | *C18orf32* |
| ENSG00000153132 | 4 | 141299609 | 141359122 | 168 | 17 | 49651 | 1.2573 | 0.10432 | *CLGN* |
| ENSG00000214513 | 2 | 73419386 | 73449641 | 92 | 13 | 41870 | 1.2569 | 0.10439 | *NOTO* |
| ENSG00000050748 | 5 | 179650143 | 179729099 | 310 | 50 | 51515 | 1.2568 | 0.10442 | *MAPK9* |
| ENSG00000182545 | 14 | 20963696 | 20989328 | 59 | 19 | 41182 | 1.2568 | 0.10442 | *RNASE10* |
| ENSG00000154556 | 4 | 186496598 | 186887806 | 1815 | 225 | 47394 | 1.2567 | 0.10443 | *SORBS2* |
| ENSG00000172336 | 7 | 100293676 | 100315118 | 64 | 12 | 56823 | 1.2567 | 0.10443 | *POP7* |
| ENSG00000240247 | 8 | 6844288 | 6885778 | 65 | 23 | 47109 | 1.2567 | 0.10444 | *DEFA1B* |
| ENSG00000167202 | 15 | 78266378 | 78380066 | 531 | 52 | 40262 | 1.2566 | 0.10444 | *TBC1D2B* |
| ENSG00000242498 | 15 | 90433159 | 90466188 | 100 | 12 | 52478 | 1.2564 | 0.10448 | *C15orf38* |
| ENSG00000112208 | 6 | 57027124 | 57064631 | 51 | 9 | 51500 | 1.2563 | 0.1045 | *BAG2* |
| ENSG00000206190 | 15 | 25912420 | 26120317 | 965 | 146 | 43201 | 1.2563 | 0.10451 | *ATP10A* |
| ENSG00000121895 | 4 | 38958365 | 39044542 | 393 | 56 | 48541 | 1.2562 | 0.10452 | *TMEM156* |
| ENSG00000175970 | 12 | 121138238 | 121171443 | 122 | 21 | 47069 | 1.2561 | 0.10454 | *UNC119B* |
| ENSG00000180061 | 19 | 55814169 | 55855416 | 158 | 24 | 49184 | 1.2561 | 0.10454 | *TMEM150B* |
| ENSG00000124120 | 20 | 43094526 | 43133244 | 68 | 12 | 45499 | 1.2561 | 0.10454 | *TTPAL* |
| ENSG00000064607 | 19 | 19091697 | 19154832 | 142 | 17 | 55007 | 1.256 | 0.10456 | *SUGP2* |
| ENSG00000172014 | 9 | 69371812 | 69435560 | 6 | 3 | 42650 | 1.2558 | 0.1046 | *ANKRD20A4* |
| ENSG00000257230 | 12 | 59907059 | 59927965 | 81 | 16 | 48214 | 1.2555 | 0.10466 | *RP11-272B17.2* |
| ENSG00000103479 | 16 | 53457889 | 53535561 | 147 | 12 | 44728 | 1.2553 | 0.10468 | *RBL2* |
| ENSG00000169410 | 15 | 75749462 | 75881630 | 219 | 23 | 33884 | 1.2553 | 0.10469 | *PTPN9* |
| ENSG00000130226 | 7 | 153574182 | 154695995 | 4030 | 372 | 43173 | 1.2552 | 0.1047 | *DPP6* |
| ENSG00000254550 | 11 | 76803886 | 76824377 | 56 | 13 | 48760 | 1.2552 | 0.1047 | *OMP* |
| ENSG00000111674 | 12 | 7012909 | 7042861 | 54 | 8 | 36748 | 1.2551 | 0.10473 | *ENO2* |
| ENSG00000221933 | 7 | 143761275 | 143782288 | 71 | 10 | 46068 | 1.2549 | 0.10475 | *OR2A25* |
| ENSG00000137040 | 9 | 6001043 | 6025618 | 49 | 11 | 42484 | 1.2549 | 0.10475 | *RANBP6* |
| ENSG00000163154 | 1 | 151119105 | 151142225 | 29 | 11 | 39451 | 1.2546 | 0.10481 | *TNFAIP8L2* |
| ENSG00000165898 | 14 | 74950423 | 74973809 | 65 | 9 | 54722 | 1.2538 | 0.10496 | *ISCA2* |
| ENSG00000148734 | 10 | 71997085 | 72053432 | 233 | 46 | 43194 | 1.2537 | 0.10497 | *NPFFR1* |
| ENSG00000125730 | 19 | 6667715 | 6740573 | 298 | 50 | 49025 | 1.2537 | 0.10497 | *C3* |

| | | | | | | | | | |
|---|---|---|---|---|---|---|---|---|---|
| ENSG00000187024 | 9 | 130445257 | 130497152 | 74 | 12 | 45843 | 1.2537 | 0.10498 | *PTRH1* |
| ENSG00000163497 | 2 | 219835809 | 219860379 | 27 | 8 | 27586 | 1.2534 | 0.10504 | *FEV* |
| ENSG00000152056 | 2 | 224606403 | 224712744 | 417 | 38 | 43340 | 1.2529 | 0.10513 | *AP1S3* |
| ENSG00000122863 | 10 | 73714123 | 73783322 | 237 | 29 | 40355 | 1.2528 | 0.10513 | *CHST3* |
| ENSG00000104951 | 19 | 50382911 | 50442796 | 189 | 39 | 42618 | 1.2527 | 0.10515 | *IL4I1* |
| ENSG00000142025 | 19 | 42338806 | 42366401 | 51 | 7 | 40136 | 1.2523 | 0.10523 | *DMRTC2* |
| ENSG00000137709 | 11 | 120097349 | 120200653 | 221 | 54 | 43259 | 1.2523 | 0.10524 | *POU2F3* |
| ENSG00000117523 | 1 | 171444651 | 171572650 | 363 | 27 | 48936 | 1.252 | 0.10528 | *PRRC2C* |
| ENSG00000160209 | 21 | 45128975 | 45192188 | 216 | 44 | 45896 | 1.2516 | 0.10536 | *PDXK* |
| ENSG00000140839 | 16 | 74432529 | 74465368 | 42 | 16 | 47791 | 1.2515 | 0.10537 | *CLEC18B* |
| ENSG00000203733 | 6 | 142444227 | 142466288 | 43 | 8 | 47487 | 1.2514 | 0.1054 | *GJE1* |
| ENSG00000114423 | 3 | 105364305 | 105598396 | 720 | 56 | 52045 | 1.2511 | 0.10544 | *CBLB* |
| ENSG00000141504 | 17 | 7519552 | 7541194 | 56 | 16 | 44267 | 1.2508 | 0.10551 | *SAT2* |
| ENSG00000128802 | 10 | 48403092 | 48426853 | 37 | 14 | 45966 | 1.2504 | 0.10557 | *GDF2* |
| ENSG00000165055 | 7 | 128106783 | 128156656 | 219 | 31 | 53859 | 1.2502 | 0.10561 | *METTL2B* |
| ENSG00000139679 | 13 | 48953707 | 49028840 | 93 | 15 | 35472 | 1.2502 | 0.10561 | *LPAR6* |
| ENSG00000139977 | 14 | 57847262 | 57892635 | 49 | 13 | 34332 | 1.2502 | 0.10561 | *NAA30* |
| ENSG00000138152 | 10 | 124020821 | 124107677 | 353 | 42 | 43305 | 1.25 | 0.10565 | *BTBD16* |
| ENSG00000034971 | 1 | 171594557 | 171631823 | 158 | 26 | 46422 | 1.2498 | 0.10569 | *MYOC* |
| ENSG00000100593 | 14 | 77930740 | 77975210 | 155 | 19 | 40815 | 1.2497 | 0.1057 | *ISM2* |
| ENSG00000177519 | 2 | 154323852 | 154345322 | 40 | 9 | 53996 | 1.2497 | 0.10571 | *RPRM* |
| ENSG00000225663 | 17 | 79770287 | 79801178 | 89 | 13 | 53768 | 1.2492 | 0.10579 | *FAM195B* |
| ENSG00000106330 | 7 | 100199725 | 100223007 | 43 | 8 | 53910 | 1.2491 | 0.10581 | *MOSPD3* |
| ENSG00000167985 | 11 | 61187514 | 61225001 | 55 | 8 | 35523 | 1.2491 | 0.10581 | *SDHAF2* |
| ENSG00000182670 | 21 | 38435526 | 38585413 | 584 | 26 | 54513 | 1.249 | 0.10583 | *TTC3* |
| ENSG00000103245 | 16 | 769753 | 801329 | 80 | 17 | 37441 | 1.2489 | 0.10585 | *NARFL* |
| ENSG00000139842 | 13 | 113852552 | 113929399 | 166 | 13 | 28847 | 1.2487 | 0.10588 | *CUL4A* |
| ENSG00000143595 | 1 | 154283566 | 154307801 | 80 | 13 | 46257 | 1.2483 | 0.10596 | *AQP10* |
| ENSG00000064655 | 20 | 45513263 | 45827492 | 975 | 141 | 41741 | 1.2483 | 0.10596 | *EYA2* |
| ENSG00000087365 | 11 | 65808200 | 65846779 | 68 | 13 | 45374 | 1.2478 | 0.10605 | *SF3B2* |
| ENSG00000108587 | 17 | 28794380 | 28864610 | 117 | 14 | 43905 | 1.2478 | 0.10605 | *GOSR1* |
| ENSG00000131808 | 11 | 30242563 | 30266808 | 55 | 6 | 49759 | 1.2476 | 0.10609 | *FSHB* |
| ENSG00000164089 | 4 | 109653196 | 109694210 | 62 | 15 | 34488 | 1.2475 | 0.10611 | *ETNPPL* |
| ENSG00000164168 | 4 | 148528534 | 148603195 | 48 | 10 | 34188 | 1.2474 | 0.10613 | *TMEM184C* |
| ENSG00000008130 | 1 | 1672671 | 1721896 | 93 | 19 | 37651 | 1.2471 | 0.10619 | *NADK* |
| ENSG00000182676 | 17 | 79781368 | 79802926 | 62 | 8 | 54434 | 1.2466 | 0.10627 | *PPP1R27* |

| | | | | | | | | | |
|---|---|---|---|---|---|---|---|---|---|
| ENSG00000100241 | 22 | 50873429 | 50923454 | 183 | 36 | 45896 | 1.2461 | 0.10636 | *SBF1* |
| ENSG00000138641 | 4 | 89432199 | 89639693 | 491 | 53 | 38176 | 1.2457 | 0.10643 | *HERC3* |
| ENSG00000103404 | 16 | 23062727 | 23170591 | 296 | 21 | 55087 | 1.2455 | 0.10647 | *USP31* |
| ENSG00000106609 | 7 | 66376212 | 66433538 | 162 | 17 | 51219 | 1.2452 | 0.10653 | *TMEM248* |
| ENSG00000164654 | 7 | 7596503 | 7658560 | 261 | 31 | 46315 | 1.245 | 0.10657 | *MIOS* |
| ENSG00000157343 | 6 | 35694809 | 35726856 | 65 | 19 | 46037 | 1.2448 | 0.1066 | *ARMC12* |
| ENSG00000117228 | 1 | 89508002 | 89541043 | 111 | 15 | 52307 | 1.2443 | 0.1067 | *GBP1* |
| ENSG00000050555 | 9 | 133874469 | 133979860 | 386 | 59 | 40325 | 1.2441 | 0.10673 | *LAMC3* |
| ENSG00000154914 | 17 | 9538015 | 9643008 | 274 | 42 | 33016 | 1.2441 | 0.10673 | *USP43* |
| ENSG00000119408 | 9 | 127009885 | 127125586 | 307 | 40 | 48935 | 1.2436 | 0.10682 | *NEK6* |
| ENSG00000137860 | 15 | 45534428 | 45578149 | 164 | 10 | 19777 | 1.2435 | 0.10683 | *SLC28A2* |
| ENSG00000197880 | 1 | 23897985 | 23977058 | 170 | 41 | 46051 | 1.2431 | 0.10692 | *MDS2* |
| ENSG00000169715 | 16 | 56649387 | 56671024 | 109 | 28 | 43305 | 1.2431 | 0.10692 | *MT1E* |
| ENSG00000143457 | 1 | 150608701 | 150679630 | 227 | 14 | 52034 | 1.2429 | 0.10695 | *GOLPH3L* |
| ENSG00000231672 | 2 | 218138742 | 218631316 | 1507 | 136 | 38982 | 1.2429 | 0.10695 | *DIRC3* |
| ENSG00000108469 | 17 | 73612925 | 73673269 | 137 | 21 | 48624 | 1.2428 | 0.10698 | *RECQL5* |
| ENSG00000113387 | 5 | 32521739 | 32614185 | 248 | 39 | 36150 | 1.2422 | 0.10708 | *SUB1* |
| ENSG00000178761 | 15 | 75182328 | 75209462 | 51 | 10 | 33142 | 1.2421 | 0.1071 | *FAM219B* |
| ENSG00000166068 | 15 | 38534527 | 38659450 | 420 | 22 | 33312 | 1.2412 | 0.10726 | *SPRED1* |
| ENSG00000114742 | 3 | 39083489 | 39148155 | 198 | 26 | 40608 | 1.2408 | 0.10734 | *WDR48* |
| ENSG00000074071 | 16 | 1811891 | 1833156 | 67 | 15 | 45339 | 1.2407 | 0.10736 | *MRPS34* |
| ENSG00000259332 | 15 | 80127537 | 80226096 | 398 | 36 | 53680 | 1.2405 | 0.10739 | *ST20-MTHFS* |
| ENSG00000164690 | 7 | 155582680 | 155614967 | 73 | 29 | 46125 | 1.2394 | 0.1076 | *SHH* |
| ENSG00000170085 | 5 | 175655365 | 175782994 | 347 | 28 | 50856 | 1.2393 | 0.10761 | *SIMC1* |
| ENSG00000109471 | 4 | 123362625 | 123387880 | 58 | 13 | 41991 | 1.2392 | 0.10763 | *IL2* |
| ENSG00000154721 | 21 | 27001584 | 27099874 | 316 | 41 | 37757 | 1.2387 | 0.10772 | *JAM2* |
| ENSG00000149596 | 20 | 42730335 | 42826218 | 405 | 68 | 51159 | 1.2385 | 0.10776 | *JPH2* |
| ENSG00000197165 | 16 | 28593264 | 28618430 | 61 | 10 | 38445 | 1.2384 | 0.10779 | *SULT1A2* |
| ENSG00000104760 | 8 | 17711889 | 17777874 | 456 | 36 | 28837 | 1.2377 | 0.10791 | *FGL1* |
| ENSG00000160917 | 7 | 99026545 | 99064994 | 74 | 12 | 34334 | 1.2372 | 0.10801 | *CPSF4* |
| ENSG00000169762 | 4 | 16152128 | 16239033 | 314 | 34 | 48452 | 1.2369 | 0.10806 | *TAPT1* |
| ENSG00000163637 | 3 | 64069543 | 64441152 | 1181 | 138 | 42227 | 1.2368 | 0.10808 | *PRICKLE2* |
| ENSG00000171126 | 2 | 42659157 | 42731237 | 202 | 36 | 45325 | 1.2366 | 0.10811 | *KCNG3* |
| ENSG00000255537 | 11 | 125355110 | 125379424 | 51 | 9 | 55101 | 1.2362 | 0.1082 | *AP000708.1* |
| ENSG00000124209 | 20 | 56874752 | 56952563 | 271 | 34 | 47432 | 1.2358 | 0.10827 | *RAB22A* |
| ENSG00000162290 | 3 | 53307447 | 53391654 | 153 | 22 | 48591 | 1.2357 | 0.10829 | *DCP1A* |

| | | | | | | | | | |
|---|---|---|---|---|---|---|---|---|---|
| ENSG00000251369 | 19 | 58036625 | 58081231 | 62 | 14 | 56828 | 1.2355 | 0.10832 | *ZNF550* |
| ENSG00000115602 | 2 | 102917962 | 102978497 | 289 | 21 | 47980 | 1.2355 | 0.10833 | *IL1RL1* |
| ENSG00000106992 | 9 | 130618759 | 130650022 | 81 | 26 | 40383 | 1.2354 | 0.10833 | *AK1* |
| ENSG00000105583 | 19 | 12768885 | 12792170 | 39 | 16 | 43643 | 1.2354 | 0.10835 | *WDR83OS* |
| ENSG00000243896 | 7 | 143945700 | 143966815 | 5 | 2 | 43844 | 1.2352 | 0.10838 | *OR2A7* |
| ENSG00000244693 | 7 | 143953794 | 143976381 | 5 | 2 | 43844 | 1.2352 | 0.10838 | *CTAGE8* |
| ENSG00000140199 | 15 | 34515460 | 34640261 | 589 | 42 | 42252 | 1.2351 | 0.10839 | *SLC12A6* |
| ENSG00000120645 | 12 | 165931 | 297626 | 504 | 97 | 41926 | 1.2345 | 0.1085 | *IQSEC3* |
| ENSG00000221883 | 3 | 48945221 | 48966818 | 36 | 9 | 40832 | 1.2344 | 0.10852 | *ARIH2OS* |
| ENSG00000169375 | 15 | 75651720 | 75758183 | 131 | 18 | 34298 | 1.2342 | 0.10857 | *SIN3A* |
| ENSG00000177283 | 10 | 35917177 | 35940362 | 51 | 11 | 45119 | 1.2341 | 0.10858 | *FZD8* |
| ENSG00000177693 | 15 | 102452245 | 102473298 | 5 | 1 | 43527 | 1.2338 | 0.10863 | *OR4F4* |
| ENSG00000268626 | 17 | 72336906 | 72358271 | 76 | 15 | 45291 | 1.2339 | 0.10863 | *AC103809.2* |
| ENSG00000167637 | 19 | 44321444 | 44363307 | 199 | 17 | 53680 | 1.2336 | 0.10868 | *ZNF283* |
| ENSG00000163156 | 1 | 151119140 | 151152773 | 38 | 15 | 41303 | 1.2335 | 0.1087 | *SCNM1* |
| ENSG00000165724 | 9 | 140466531 | 140494942 | 59 | 10 | 47750 | 1.2335 | 0.1087 | *ZMYND19* |
| ENSG00000204520 | 6 | 31361356 | 31393092 | 540 | 42 | 31041 | 1.2332 | 0.10874 | *MICA* |
| ENSG00000083635 | 13 | 45503384 | 45573618 | 116 | 24 | 36964 | 1.2329 | 0.10881 | *NUFIP1* |
| ENSG00000112312 | 6 | 24765159 | 24796327 | 92 | 25 | 44997 | 1.2322 | 0.10894 | *GMNN* |
| ENSG00000183542 | 12 | 10549983 | 10572356 | 54 | 10 | 37846 | 1.2318 | 0.10901 | *KLRC4* |
| ENSG00000061987 | 12 | 62850597 | 63001363 | 459 | 38 | 54452 | 1.2318 | 0.10901 | *MON2* |
| ENSG00000092295 | 14 | 24708320 | 24743638 | 75 | 20 | 38567 | 1.2317 | 0.10903 | *TGM1* |
| ENSG00000112333 | 6 | 108477262 | 108520013 | 80 | 22 | 42326 | 1.2313 | 0.1091 | *NR2E1* |
| ENSG00000185988 | 19 | 1514073 | 1545455 | 164 | 26 | 33488 | 1.2312 | 0.10912 | *PLK5* |
| ENSG00000073712 | 14 | 53313986 | 53429153 | 282 | 34 | 44718 | 1.2311 | 0.10914 | *FERMT2* |
| ENSG00000172061 | 3 | 194065976 | 194100472 | 237 | 34 | 41509 | 1.2307 | 0.10922 | *LRRC15* |
| ENSG00000007866 | 6 | 35431374 | 35474853 | 141 | 31 | 38741 | 1.2305 | 0.10925 | *TEAD3* |
| ENSG00000110911 | 12 | 51363184 | 51432349 | 217 | 18 | 36029 | 1.2305 | 0.10926 | *SLC11A2* |
| ENSG00000059728 | 2 | 70114820 | 70180077 | 142 | 13 | 30183 | 1.2304 | 0.10927 | *MXD1* |
| ENSG00000131730 | 5 | 80519104 | 80572216 | 146 | 18 | 51462 | 1.2301 | 0.10933 | *CKMT2* |
| ENSG00000196821 | 6 | 34545065 | 34674636 | 323 | 26 | 37158 | 1.2298 | 0.10938 | *C6orf106* |
| ENSG00000109736 | 4 | 2922288 | 2946586 | 61 | 16 | 45023 | 1.2296 | 0.10942 | *MFSD10* |
| ENSG00000177030 | 11 | 634233 | 716715 | 376 | 40 | 46007 | 1.2296 | 0.10942 | *DEAF1* |
| ENSG00000183578 | 15 | 51338795 | 51407473 | 152 | 20 | 54540 | 1.2293 | 0.10948 | *TNFAIP8L3* |
| ENSG00000125037 | 3 | 9994221 | 10062800 | 213 | 11 | 42983 | 1.2285 | 0.10963 | *EMC3* |
| ENSG00000136444 | 17 | 48546161 | 48573336 | 65 | 13 | 35278 | 1.2285 | 0.10963 | *RSAD1* |

| | | | | | | | | | |
|---|---|---|---|---|---|---|---|---|---|
| ENSG00000112308 | 6 | 24695089 | 24731064 | 126 | 34 | 48744 | 1.2285 | 0.10964 | *C6orf62* |
| ENSG00000146826 | 7 | 99742043 | 99766338 | 37 | 8 | 49161 | 1.2279 | 0.10974 | *C7orf43* |
| ENSG00000164100 | 4 | 118944773 | 119189803 | 864 | 53 | 35015 | 1.2278 | 0.10976 | *NDST3* |
| ENSG00000184478 | 11 | 5958570 | 5979591 | 72 | 16 | 54343 | 1.2277 | 0.10979 | *OR56A3* |
| ENSG00000155265 | 10 | 99599996 | 99641337 | 97 | 20 | 40050 | 1.2269 | 0.10994 | *GOLGA7B* |
| ENSG00000247077 | 12 | 133277405 | 133309228 | 134 | 26 | 47497 | 1.2264 | 0.11002 | *PGAM5* |
| ENSG00000178078 | 19 | 4314040 | 4352783 | 141 | 27 | 50307 | 1.2263 | 0.11004 | *STAP2* |
| ENSG00000075223 | 7 | 80361854 | 80561675 | 503 | 87 | 46497 | 1.2262 | 0.11007 | *SEMA3C* |
| ENSG00000173705 | 3 | 33181537 | 33270707 | 328 | 39 | 48017 | 1.2258 | 0.11014 | *SUSD5* |
| ENSG00000030419 | 2 | 213854429 | 214027151 | 501 | 48 | 39143 | 1.2255 | 0.11019 | *IKZF2* |
| ENSG00000150627 | 4 | 176976985 | 177113978 | 555 | 43 | 53435 | 1.2255 | 0.11019 | *WDR17* |
| ENSG00000134376 | 1 | 197160592 | 197457585 | 489 | 60 | 44219 | 1.2254 | 0.11021 | *CRB1* |
| ENSG00000066032 | 2 | 79402357 | 80885905 | 5011 | 403 | 42777 | 1.2253 | 0.11023 | *CTNNA2* |
| ENSG00000169758 | 15 | 76342178 | 76531462 | 515 | 73 | 40850 | 1.2249 | 0.1103 | *C15orf27* |
| ENSG00000162621 | 1 | 74925562 | 74988298 | 107 | 20 | 33587 | 1.2249 | 0.11031 | *LRRC53* |
| ENSG00000178802 | 15 | 75172346 | 75201798 | 51 | 10 | 33142 | 1.2249 | 0.11031 | *MPI* |
| ENSG00000167074 | 22 | 41753337 | 41805330 | 111 | 10 | 48229 | 1.2249 | 0.11031 | *TEF* |
| ENSG00000164287 | 5 | 54398799 | 54479005 | 197 | 20 | 54118 | 1.2248 | 0.11033 | *CDC20B* |
| ENSG00000185436 | 1 | 24470647 | 24524449 | 198 | 33 | 44977 | 1.2247 | 0.11034 | *IFNLR1* |
| ENSG00000147439 | 8 | 22467931 | 22536661 | 209 | 24 | 47308 | 1.2247 | 0.11035 | *BIN3* |
| ENSG00000188321 | 19 | 9424448 | 9471838 | 152 | 15 | 44967 | 1.2243 | 0.11043 | *ZNF559* |
| ENSG00000186377 | 1 | 47417036 | 47526423 | 249 | 26 | 36687 | 1.2242 | 0.11044 | *CYP4X1* |
| ENSG00000002330 | 11 | 64027302 | 64062176 | 50 | 20 | 47717 | 1.224 | 0.11048 | *BAD* |
| ENSG00000119772 | 2 | 25445845 | 25575459 | 263 | 57 | 50199 | 1.2239 | 0.11049 | *DNMT3A* |
| ENSG00000070540 | 17 | 66407089 | 66463654 | 146 | 27 | 51538 | 1.2233 | 0.11061 | *WIPI1* |
| ENSG00000147869 | 9 | 14709722 | 14732715 | 120 | 27 | 44066 | 1.223 | 0.11066 | *CER1* |
| ENSG00000204613 | 6 | 30109722 | 30138711 | 254 | 21 | 45937 | 1.2227 | 0.11072 | *TRIM10* |
| ENSG00000197892 | 8 | 28914796 | 29130641 | 450 | 55 | 46543 | 1.2227 | 0.11073 | *KIF13B* |
| ENSG00000155906 | 6 | 151715989 | 151783259 | 268 | 30 | 38675 | 1.222 | 0.11085 | *RMND1* |
| ENSG00000128284 | 22 | 36526372 | 36572225 | 158 | 30 | 37730 | 1.2213 | 0.11098 | *APOL3* |
| ENSG00000197747 | 1 | 151945391 | 151976866 | 109 | 10 | 47696 | 1.2213 | 0.11099 | *S100A10* |
| ENSG00000171298 | 17 | 78065355 | 78103678 | 259 | 27 | 47333 | 1.2213 | 0.11099 | *GAA* |
| ENSG00000183318 | 17 | 8646424 | 8671877 | 81 | 19 | 46700 | 1.2212 | 0.111 | *SPDYE4* |
| ENSG00000214239 | 6 | 159037471 | 159059322 | 99 | 14 | 52622 | 1.2211 | 0.11102 | *AL591025.1* |
| ENSG00000188883 | 7 | 139127436 | 139178458 | 208 | 35 | 40380 | 1.2204 | 0.11115 | *KLRG2* |
| ENSG00000198538 | 19 | 53290662 | 53370853 | 373 | 58 | 38851 | 1.2203 | 0.11117 | *ZNF28* |

| | | | | | | | | | |
|---|---|---|---|---|---|---|---|---|---|
| ENSG00000105329 | 19 | 41797492 | 41869816 | 152 | 25 | 52120 | 1.2196 | 0.11131 | *TGFB1* |
| ENSG00000163762 | 3 | 149026285 | 149062201 | 110 | 26 | 51913 | 1.2183 | 0.11155 | *TM4SF18* |
| ENSG00000130307 | 19 | 17349985 | 17403595 | 219 | 31 | 44277 | 1.2182 | 0.11156 | *USHBP1* |
| ENSG00000157593 | 6 | 44211833 | 44235291 | 77 | 23 | 54387 | 1.218 | 0.11162 | *SLC35B2* |
| ENSG00000145934 | 5 | 166701804 | 167701162 | 2518 | 293 | 42196 | 1.2178 | 0.11164 | *TENM2* |
| ENSG00000114853 | 3 | 42685176 | 42719072 | 81 | 21 | 47279 | 1.2175 | 0.1117 | *ZBTB47* |
| ENSG00000250733 | 8 | 140933416 | 140956190 | 84 | 16 | 49381 | 1.2175 | 0.1117 | *C8orf17* |
| ENSG00000249773 | 7 | 55944970 | 56032932 | 283 | 35 | 53038 | 1.2174 | 0.11172 | *MRPS17* |
| ENSG00000172731 | 10 | 72048729 | 72152382 | 430 | 63 | 47887 | 1.2173 | 0.11174 | *LRRC20* |
| ENSG00000185480 | 12 | 102503956 | 102601298 | 197 | 20 | 54031 | 1.2172 | 0.11176 | *PARPBP* |
| ENSG00000052802 | 4 | 166238775 | 166274312 | 145 | 26 | 41865 | 1.217 | 0.1118 | *MSMO1* |
| ENSG00000065154 | 10 | 126075872 | 126117545 | 139 | 18 | 55146 | 1.2166 | 0.11188 | *OAT* |
| ENSG00000160908 | 7 | 99074142 | 99107947 | 45 | 16 | 45352 | 1.2164 | 0.11192 | *ZNF394* |
| ENSG00000145949 | 6 | 2653863 | 2761200 | 529 | 67 | 52137 | 1.2162 | 0.11195 | *MYLK4* |
| ENSG00000132313 | 2 | 86416478 | 86450917 | 116 | 14 | 39024 | 1.2159 | 0.11201 | *MRPL35* |
| ENSG00000206503 | 6 | 29899037 | 29923661 | 807 | 46 | 43495 | 1.2159 | 0.11201 | *HLA-A* |
| ENSG00000119699 | 14 | 76414442 | 76459334 | 75 | 18 | 46859 | 1.2156 | 0.11208 | *TGFB3* |
| ENSG00000163686 | 3 | 58213233 | 58291420 | 341 | 26 | 35640 | 1.2153 | 0.11212 | *ABHD6* |
| ENSG00000184986 | 14 | 105982940 | 106006539 | 24 | 5 | 40119 | 1.2151 | 0.11216 | *TMEM121* |
| ENSG00000161960 | 17 | 7466024 | 7492323 | 63 | 13 | 46441 | 1.2148 | 0.11221 | *EIF4A1* |
| ENSG00000179873 | 19 | 56286770 | 56358166 | 403 | 76 | 37885 | 1.2149 | 0.11221 | *NLRP11* |
| ENSG00000104415 | 8 | 134193282 | 134252587 | 177 | 46 | 46441 | 1.2146 | 0.11226 | *WISP1* |
| ENSG00000169756 | 2 | 109140857 | 109313702 | 597 | 38 | 35095 | 1.2146 | 0.11227 | *LIMS1* |
| ENSG00000080815 | 14 | 73593126 | 73700399 | 254 | 36 | 51088 | 1.2145 | 0.11228 | *PSEN1* |
| ENSG00000129255 | 17 | 7476847 | 7506107 | 67 | 14 | 44640 | 1.2144 | 0.1123 | *MPDU1* |
| ENSG00000131055 | 20 | 30215691 | 30242809 | 78 | 8 | 33095 | 1.2144 | 0.1123 | *COX4I2* |
| ENSG00000198488 | 11 | 76735385 | 76763096 | 91 | 19 | 47667 | 1.214 | 0.11237 | *B3GNT6* |
| ENSG00000205784 | 19 | 4880449 | 4912879 | 110 | 22 | 41134 | 1.2137 | 0.11244 | *ARRDC5* |
| ENSG00000051180 | 15 | 40976972 | 41034354 | 130 | 13 | 51644 | 1.2132 | 0.11252 | *RAD51* |
| ENSG00000108551 | 17 | 17387751 | 17409709 | 46 | 12 | 31105 | 1.2131 | 0.11254 | *RASD1* |
| ENSG00000156374 | 10 | 105052553 | 105120891 | 119 | 18 | 44545 | 1.213 | 0.11256 | *PCGF6* |
| ENSG00000157103 | 3 | 11024410 | 11090933 | 218 | 54 | 47226 | 1.2128 | 0.1126 | *SLC6A1* |
| ENSG00000130508 | 2 | 1625659 | 1758624 | 496 | 79 | 45808 | 1.2125 | 0.11265 | *PXDN* |
| ENSG00000174562 | 19 | 51318545 | 51350469 | 134 | 34 | 47361 | 1.2124 | 0.11268 | *KLK15* |
| ENSG00000121058 | 17 | 55005548 | 55048415 | 105 | 26 | 50298 | 1.2123 | 0.11269 | *COIL* |
| ENSG00000182687 | 17 | 74060875 | 74083622 | 38 | 7 | 39076 | 1.2122 | 0.11272 | *GALR2* |

| | | | | | | | | | |
|---|---|---|---|---|---|---|---|---|---|
| ENSG00000272104 | 3 | 50378323 | 50415354 | 23 | 8 | 54265 | 1.212 | 0.11276 | *XXcos-LUCA11.5* |
| ENSG00000197471 | 16 | 29664300 | 29692187 | 48 | 11 | 31698 | 1.2119 | 0.11277 | *SPN* |
| ENSG00000101266 | 20 | 449116 | 534465 | 193 | 33 | 43634 | 1.2116 | 0.11284 | *CSNK2A1* |
| ENSG00000231145 | 2 | 190632909 | 190653157 | 55 | 12 | 44359 | 1.2115 | 0.11285 | *AC013468.1* |
| ENSG00000198944 | 5 | 132139033 | 132162488 | 32 | 8 | 44208 | 1.2115 | 0.11285 | *SOWAHA* |
| ENSG00000105610 | 19 | 12985237 | 13007995 | 51 | 11 | 43033 | 1.2115 | 0.11285 | *KLF1* |
| ENSG00000132702 | 1 | 156579086 | 156605517 | 39 | 8 | 26890 | 1.211 | 0.11294 | *HAPLN2* |
| ENSG00000121057 | 17 | 55152453 | 55208710 | 225 | 42 | 46126 | 1.2108 | 0.11299 | *AKAP1* |
| ENSG00000264424 | 17 | 10336607 | 10382876 | 86 | 11 | 29850 | 1.2107 | 0.113 | *MYH4* |
| ENSG00000106266 | 7 | 2281405 | 2403953 | 436 | 63 | 42661 | 1.2104 | 0.11305 | *SNX8* |
| ENSG00000162430 | 1 | 26116667 | 26154713 | 107 | 13 | 46394 | 1.2103 | 0.11308 | *SEPN1* |
| ENSG00000164081 | 3 | 51686709 | 51748339 | 55 | 24 | 52631 | 1.2101 | 0.11313 | *TEX264* |
| ENSG00000154217 | 17 | 65363575 | 65703372 | 935 | 193 | 48301 | 1.2099 | 0.11316 | *PITPNC1* |
| ENSG00000269407 | 20 | 23320015 | 23344066 | 82 | 13 | 37609 | 1.2099 | 0.11316 | *AL096677.1* |
| ENSG00000166925 | 7 | 100050982 | 100086902 | 64 | 12 | 52244 | 1.2094 | 0.11326 | *TSC22D4* |
| ENSG00000176428 | 7 | 73072155 | 73096442 | 89 | 10 | 50976 | 1.2092 | 0.11329 | *VPS37D* |
| ENSG00000143420 | 1 | 150563327 | 150612088 | 129 | 21 | 47039 | 1.2088 | 0.11337 | *ENSA* |
| ENSG00000064199 | 11 | 124533694 | 124577414 | 60 | 15 | 44318 | 1.2087 | 0.11338 | *SPA17* |
| ENSG00000161265 | 19 | 36223365 | 36246346 | 64 | 13 | 49404 | 1.2087 | 0.11338 | *U2AF1L4* |
| ENSG00000186509 | 11 | 57781353 | 57959088 | 669 | 50 | 42990 | 1.2086 | 0.11341 | *OR9Q1* |
| ENSG00000129534 | 14 | 45662393 | 45732743 | 134 | 19 | 44307 | 1.2086 | 0.11341 | *MIS18BP1* |
| ENSG00000113583 | 5 | 133281201 | 133314478 | 60 | 17 | 55794 | 1.2085 | 0.11342 | *C5orf15* |
| ENSG00000154309 | 1 | 222978406 | 223189337 | 721 | 70 | 50383 | 1.2081 | 0.1135 | *DISP1* |
| ENSG00000109920 | 11 | 47728072 | 47798995 | 153 | 20 | 38496 | 1.2077 | 0.11357 | *FNBP4* |
| ENSG00000033867 | 3 | 27404214 | 27535911 | 497 | 40 | 52004 | 1.2077 | 0.11358 | *SLC4A7* |
| ENSG00000090971 | 19 | 55986371 | 56008935 | 95 | 23 | 50002 | 1.2075 | 0.11363 | *NAT14* |
| ENSG00000149571 | 11 | 126283254 | 126883355 | 2178 | 189 | 47414 | 1.2074 | 0.11364 | *KIRREL3* |
| ENSG00000163293 | 4 | 47906159 | 48052188 | 517 | 39 | 39705 | 1.2073 | 0.11366 | *NIPAL1* |
| ENSG00000172262 | 5 | 43055278 | 43202123 | 536 | 42 | 46870 | 1.2073 | 0.11367 | *ZNF131* |
| ENSG00000131115 | 19 | 44701700 | 44751421 | 86 | 14 | 38730 | 1.2069 | 0.11373 | *ZNF227* |
| ENSG00000081059 | 5 | 133440402 | 133497556 | 122 | 17 | 38942 | 1.2068 | 0.11376 | *TCF7* |
| ENSG00000034677 | 8 | 101259288 | 101358446 | 222 | 21 | 51504 | 1.2063 | 0.11385 | *RNF19A* |
| ENSG00000104537 | 8 | 124683034 | 124759647 | 212 | 40 | 47153 | 1.2061 | 0.1139 | *ANXA13* |
| ENSG00000108733 | 17 | 33891814 | 33915882 | 87 | 16 | 54780 | 1.206 | 0.11392 | *PEX12* |
| ENSG00000112041 | 6 | 35455651 | 35490715 | 118 | 23 | 47457 | 1.2057 | 0.11396 | *TULP1* |
| ENSG00000176871 | 12 | 118460712 | 118510235 | 169 | 32 | 43190 | 1.2057 | 0.11396 | *WSB2* |

| | | | | | | | | | |
|---|---|---|---|---|---|---|---|---|---|
| ENSG00000126267 | 19 | 36129125 | 36159763 | 92 | 17 | 42749 | 1.2055 | 0.114 | *COX6B1* |
| ENSG00000197296 | 20 | 42921478 | 42949809 | 66 | 9 | 28941 | 1.2055 | 0.114 | *FITM2* |
| ENSG00000028839 | 6 | 134263308 | 134321570 | 133 | 19 | 36190 | 1.205 | 0.1141 | *TBPL1* |
| ENSG00000204308 | 6 | 32136131 | 32161930 | 76 | 20 | 45287 | 1.2049 | 0.11412 | *RNF5* |
| ENSG00000114933 | 2 | 206848445 | 206961027 | 306 | 31 | 37261 | 1.2039 | 0.11431 | *INO80D* |
| ENSG00000196776 | 3 | 107752145 | 107819872 | 108 | 26 | 51894 | 1.2035 | 0.11439 | *CD47* |
| ENSG00000163521 | 2 | 220091328 | 220120200 | 67 | 14 | 38533 | 1.2032 | 0.11444 | *GLB1L* |
| ENSG00000185689 | 6 | 4069440 | 4141185 | 241 | 27 | 44618 | 1.2025 | 0.11458 | *C6orf201* |
| ENSG00000172301 | 17 | 30168883 | 30196356 | 70 | 15 | 47541 | 1.2025 | 0.11459 | *COPRS* |
| ENSG00000196427 | 1 | 108755963 | 108796689 | 42 | 8 | 48730 | 1.2021 | 0.11466 | *NBPF4* |
| ENSG00000079393 | 10 | 76844192 | 76878979 | 110 | 19 | 39691 | 1.2021 | 0.11467 | *DUSP13* |
| ENSG00000204572 | 11 | 71266609 | 71289980 | 154 | 22 | 46901 | 1.2021 | 0.11467 | *KRTAP5-10* |
| ENSG00000136238 | 7 | 6404154 | 6453608 | 225 | 32 | 42687 | 1.202 | 0.11468 | *RAC1* |
| ENSG00000137947 | 1 | 89308615 | 89367627 | 189 | 24 | 35982 | 1.202 | 0.11469 | *GTF2B* |
| ENSG00000069275 | 1 | 205671947 | 205729404 | 161 | 28 | 39737 | 1.2019 | 0.1147 | *NUCKS1* |
| ENSG00000168310 | 4 | 185298867 | 185405734 | 378 | 97 | 47809 | 1.2018 | 0.11471 | *IRF2* |
| ENSG00000169221 | 16 | 30358423 | 30391585 | 42 | 11 | 31295 | 1.2017 | 0.11473 | *TBC1D10B* |
| ENSG00000137497 | 11 | 71703910 | 71801739 | 229 | 13 | 29796 | 1.2011 | 0.11485 | *NUMA1* |
| ENSG00000121281 | 16 | 50270048 | 50362046 | 233 | 48 | 45278 | 1.2007 | 0.11493 | *ADCY7* |
| ENSG00000152377 | 5 | 136300987 | 136944068 | 1935 | 176 | 45635 | 1.2003 | 0.11501 | *SPOCK1* |
| ENSG00000224689 | 19 | 9790600 | 9821452 | 141 | 13 | 40860 | 1.2003 | 0.11502 | *ZNF812* |
| ENSG00000169964 | 3 | 44893361 | 44917162 | 47 | 15 | 37128 | 1.1999 | 0.11509 | *TMEM42* |
| ENSG00000018699 | 2 | 32843099 | 33056118 | 881 | 91 | 50529 | 1.1998 | 0.11511 | *TTC27* |
| ENSG00000212124 | 12 | 11164218 | 11185219 | 109 | 16 | 36446 | 1.1996 | 0.11515 | *TAS2R19* |
| ENSG00000103024 | 16 | 1810287 | 1831731 | 65 | 15 | 45420 | 1.1995 | 0.11517 | *NME3* |
| ENSG00000117450 | 1 | 45966708 | 45998719 | 88 | 9 | 56778 | 1.1993 | 0.1152 | *PRDX1* |
| ENSG00000174132 | 5 | 99861009 | 99932445 | 178 | 17 | 36836 | 1.199 | 0.11526 | *FAM174A* |
| ENSG00000143376 | 1 | 151574541 | 151681567 | 185 | 25 | 41287 | 1.199 | 0.11527 | *SNX27* |
| ENSG00000163462 | 1 | 155135873 | 155167447 | 48 | 10 | 48990 | 1.1988 | 0.1153 | *TRIM46* |
| ENSG00000163220 | 1 | 153320330 | 153343503 | 86 | 8 | 49202 | 1.1987 | 0.11533 | *S100A9* |
| ENSG00000083817 | 19 | 58072934 | 58100295 | 50 | 7 | 57279 | 1.1986 | 0.11535 | *ZNF416* |
| ENSG00000165474 | 13 | 20751609 | 20777037 | 93 | 18 | 37635 | 1.1985 | 0.11536 | *GJB2* |
| ENSG00000130876 | 19 | 33689570 | 33726756 | 121 | 31 | 33605 | 1.1984 | 0.11538 | *SLC7A10* |
| ENSG00000269422 | 16 | 88884366 | 88904545 | 187 | 17 | 53332 | 1.1984 | 0.11539 | *AC092384.1* |
| ENSG00000249160 | 5 | 10495108 | 10532196 | 139 | 28 | 44508 | 1.1983 | 0.1154 | *RP11-1C1.5* |
| ENSG00000121903 | 1 | 33928246 | 33972107 | 105 | 11 | 48734 | 1.1977 | 0.11552 | *ZSCAN20* |

| | | | | | | | | | |
|---|---|---|---|---|---|---|---|---|---|
| ENSG00000168743 | 4 | 106805932 | 106935184 | 466 | 42 | 34889 | 1.1975 | 0.11556 | *NPNT* |
| ENSG00000172403 | 4 | 119799996 | 119992402 | 716 | 106 | 52436 | 1.1974 | 0.11557 | *SYNPO2* |
| ENSG00000104960 | 19 | 50343992 | 50374001 | 81 | 19 | 49883 | 1.1973 | 0.1156 | *PTOV1* |
| ENSG00000175170 | 20 | 25734102 | 25858861 | 198 | 22 | 36465 | 1.1973 | 0.1156 | *FAM182B* |
| ENSG00000143552 | 1 | 153955161 | 154137592 | 312 | 30 | 49829 | 1.1971 | 0.11564 | *NUP210L* |
| ENSG00000058668 | 1 | 203585689 | 203723209 | 255 | 55 | 38561 | 1.1965 | 0.11575 | *ATP2B4* |
| ENSG00000134910 | 11 | 125451607 | 125505110 | 204 | 16 | 40852 | 1.1961 | 0.11583 | *STT3A* |
| ENSG00000188175 | 7 | 92807899 | 92865837 | 110 | 27 | 45661 | 1.1959 | 0.11586 | *HEPACAM2* |
| ENSG00000250264 | 6 | 32771544 | 32816599 | 468 | 53 | 43549 | 1.1958 | 0.11588 | *TAP2* |
| ENSG00000175497 | 2 | 115189876 | 116613328 | 3859 | 263 | 46817 | 1.1954 | 0.11596 | *DPP10* |
| ENSG00000133884 | 11 | 65091225 | 65130720 | 65 | 17 | 41885 | 1.195 | 0.11604 | *DPF2* |
| ENSG00000212127 | 12 | 11080005 | 11334172 | 1425 | 36 | 35345 | 1.1948 | 0.11607 | *TAS2R14* |
| ENSG00000229615 | 2 | 87079023 | 87102827 | 51 | 7 | 40835 | 1.1947 | 0.1161 | *AC111200.1* |
| ENSG00000099203 | 19 | 10933114 | 10956994 | 32 | 9 | 52754 | 1.1947 | 0.1161 | *TMED1* |
| ENSG00000037897 | 12 | 58152254 | 58176576 | 31 | 5 | 37437 | 1.1947 | 0.11611 | *METTL1* |
| ENSG00000143387 | 1 | 150758684 | 150790799 | 55 | 12 | 48426 | 1.1946 | 0.11612 | *CTSK* |
| ENSG00000115221 | 2 | 160946177 | 161138399 | 621 | 63 | 45494 | 1.1946 | 0.11612 | *ITGB6* |
| ENSG00000092330 | 14 | 24698849 | 24721880 | 27 | 11 | 44736 | 1.1946 | 0.11612 | *TINF2* |
| ENSG00000167104 | 20 | 31609454 | 31641853 | 98 | 18 | 47949 | 1.1946 | 0.11612 | *BPIFB6* |
| ENSG00000260286 | 6 | 24787549 | 24809117 | 84 | 19 | 38613 | 1.1941 | 0.11621 | *C6orf229* |
| ENSG00000170365 | 4 | 146392346 | 146489231 | 164 | 24 | 33118 | 1.194 | 0.11625 | *SMAD1* |
| ENSG00000142494 | 17 | 19388698 | 19492347 | 218 | 52 | 44201 | 1.1938 | 0.11628 | *SLC47A1* |
| ENSG00000187164 | 10 | 118633742 | 118896097 | 351 | 36 | 36189 | 1.1935 | 0.11634 | *KIAA1598* |
| ENSG00000115607 | 2 | 103025149 | 103079025 | 248 | 19 | 38028 | 1.1935 | 0.11635 | *IL18RAP* |
| ENSG00000167178 | 15 | 74382652 | 74440881 | 39 | 18 | 41286 | 1.1933 | 0.11637 | *ISLR2* |
| ENSG00000101255 | 20 | 351261 | 388203 | 178 | 44 | 37793 | 1.1931 | 0.11641 | *TRIB3* |
| ENSG00000170903 | 11 | 105856350 | 105903130 | 93 | 16 | 54992 | 1.1931 | 0.11642 | *MSANTD4* |
| ENSG00000254959 | 7 | 30781753 | 30941696 | 630 | 75 | 37977 | 1.1928 | 0.11648 | *INMT-FAM188B* |
| ENSG00000184465 | 6 | 169847307 | 170112159 | 708 | 61 | 36290 | 1.1926 | 0.11651 | *WDR27* |
| ENSG00000170871 | 4 | 6773102 | 6895897 | 221 | 39 | 43895 | 1.1923 | 0.11656 | *KIAA0232* |
| ENSG00000179046 | 4 | 189002427 | 189040757 | 264 | 42 | 35500 | 1.1923 | 0.11657 | *TRIML2* |
| ENSG00000171861 | 17 | 675513 | 705749 | 112 | 25 | 53544 | 1.1923 | 0.11657 | *RNMTL1* |
| ENSG00000105483 | 19 | 48674027 | 48769203 | 454 | 53 | 47370 | 1.1919 | 0.11664 | *CARD8* |
| ENSG00000163218 | 1 | 153292596 | 153331316 | 191 | 13 | 40529 | 1.1914 | 0.11674 | *PGLYRP4* |
| ENSG00000158717 | 16 | 88752903 | 88782829 | 159 | 33 | 48087 | 1.1914 | 0.11675 | *RNF166* |
| ENSG00000108797 | 17 | 40824631 | 40861832 | 43 | 11 | 51177 | 1.1914 | 0.11675 | *CNTNAP1* |

| | | | | | | | | | |
|---|---|---|---|---|---|---|---|---|---|
| ENSG00000213121 | 6 | 153542455 | 153678623 | 538 | 62 | 49900 | 1.1912 | 0.11678 | *AL590867.1* |
| ENSG00000265944 | 18 | 6501415 | 6600652 | 365 | 70 | 36397 | 1.1911 | 0.11682 | *C18orf64* |
| ENSG00000262039 | 17 | 47438102 | 47564350 | 382 | 76 | 44444 | 1.1909 | 0.11685 | *RP11-81K2.1* |
| ENSG00000163898 | 3 | 185214050 | 185280401 | 220 | 20 | 34302 | 1.1907 | 0.11688 | *LIPH* |
| ENSG00000065534 | 3 | 123318896 | 123613178 | 497 | 63 | 43342 | 1.1907 | 0.11689 | *MYLK* |
| ENSG00000068615 | 2 | 86431116 | 86575206 | 519 | 41 | 38594 | 1.1903 | 0.11696 | *REEP1* |
| ENSG00000241852 | 8 | 22447114 | 22471663 | 60 | 7 | 54606 | 1.1903 | 0.11696 | *C8orf58* |
| ENSG00000121749 | 12 | 72223487 | 72330629 | 283 | 30 | 45667 | 1.1902 | 0.11698 | *TBC1D15* |
| ENSG00000162407 | 1 | 56950419 | 57120974 | 639 | 70 | 38963 | 1.1897 | 0.11709 | *PPAP2B* |
| ENSG00000079112 | 8 | 95129399 | 95239531 | 362 | 54 | 43694 | 1.1896 | 0.1171 | *CDH17* |
| ENSG00000184227 | 14 | 73993818 | 74020498 | 26 | 13 | 47460 | 1.1893 | 0.11715 | *ACOT1* |
| ENSG00000146618 | 7 | 19174405 | 19195044 | 51 | 14 | 48257 | 1.1891 | 0.1172 | *FERD3L* |
| ENSG00000198807 | 14 | 37116773 | 37158920 | 161 | 27 | 46939 | 1.1891 | 0.1172 | *PAX9* |
| ENSG00000089327 | 19 | 35635633 | 35670786 | 141 | 26 | 48751 | 1.189 | 0.11722 | *FXYD5* |
| ENSG00000182912 | 21 | 45927099 | 45955835 | 43 | 20 | 46915 | 1.1889 | 0.11724 | *C21orf90* |
| ENSG00000103018 | 16 | 69448428 | 69510169 | 209 | 26 | 51923 | 1.1888 | 0.11726 | *CYB5B* |
| ENSG00000116774 | 1 | 114512063 | 114534876 | 88 | 16 | 41089 | 1.1888 | 0.11727 | *OLFML3* |
| ENSG00000010165 | 1 | 171740788 | 171793163 | 199 | 12 | 52120 | 1.1886 | 0.11729 | *METTL13* |
| ENSG00000121988 | 2 | 135884486 | 136298806 | 627 | 26 | 20361 | 1.1886 | 0.11729 | *ZRANB3* |
| ENSG00000136237 | 7 | 22147856 | 22406763 | 868 | 109 | 49537 | 1.1886 | 0.1173 | *RAPGEF5* |
| ENSG00000163354 | 1 | 154980996 | 155016257 | 68 | 14 | 40530 | 1.1879 | 0.11744 | *DCST2* |
| ENSG00000156831 | 8 | 126093921 | 126389362 | 750 | 84 | 40011 | 1.1879 | 0.11744 | *NSMCE2* |
| ENSG00000175893 | 9 | 14601069 | 14703469 | 476 | 56 | 50245 | 1.1878 | 0.11746 | *ZDHHC21* |
| ENSG00000179294 | 17 | 36817961 | 36841187 | 45 | 11 | 41298 | 1.1873 | 0.11755 | *C17orf96* |
| ENSG00000178562 | 2 | 204561198 | 204613635 | 134 | 18 | 50302 | 1.1868 | 0.11764 | *CD28* |
| ENSG00000187144 | 1 | 16703612 | 16773919 | 207 | 35 | 46958 | 1.1867 | 0.11767 | *SPATA21* |
| ENSG00000163617 | 3 | 113672984 | 113785460 | 189 | 33 | 53458 | 1.1867 | 0.11768 | *KIAA1407* |
| ENSG00000143515 | 1 | 154288029 | 154333783 | 145 | 18 | 44884 | 1.1865 | 0.11771 | *ATP8B2* |
| ENSG00000145824 | 5 | 134896373 | 134924969 | 50 | 11 | 49208 | 1.1865 | 0.11772 | *CXCL14* |
| ENSG00000135914 | 2 | 231962944 | 231999832 | 57 | 15 | 39219 | 1.1861 | 0.1178 | *HTR2B* |
| ENSG00000163633 | 4 | 87787358 | 87867354 | 229 | 27 | 47236 | 1.1855 | 0.11791 | *C4orf36* |
| ENSG00000166924 | 7 | 100071550 | 100102422 | 58 | 8 | 55131 | 1.1855 | 0.11792 | *NYAP1* |
| ENSG00000176046 | 16 | 28538606 | 28560495 | 55 | 11 | 43952 | 1.1854 | 0.11793 | *NUPR1* |
| ENSG00000173264 | 11 | 64027534 | 64066972 | 50 | 19 | 47714 | 1.1853 | 0.11794 | *GPR137* |
| ENSG00000262730 | 17 | 7819249 | 7843940 | 63 | 13 | 44258 | 1.1848 | 0.11804 | *RP11-1099M24.7* |
| ENSG00000129295 | 8 | 133574320 | 133697838 | 236 | 31 | 37768 | 1.184 | 0.11821 | *LRRC6* |

| | | | | | | | | | |
|---|---|---|---|---|---|---|---|---|---|
| ENSG00000019485 | 11 | 45105564 | 45257734 | 353 | 47 | 38143 | 1.1837 | 0.11827 | *PRDM11* |
| ENSG00000108651 | 17 | 30177923 | 30238784 | 186 | 19 | 46411 | 1.1836 | 0.11829 | *UTP6* |
| ENSG00000124019 | 2 | 225233415 | 225276802 | 169 | 20 | 52144 | 1.1835 | 0.1183 | *FAM124B* |
| ENSG00000107281 | 9 | 139923922 | 139950655 | 84 | 18 | 52154 | 1.1833 | 0.11835 | *NPDC1* |
| ENSG00000226321 | 2 | 241848202 | 241942728 | 256 | 39 | 50490 | 1.1832 | 0.11836 | *AC104809.3* |
| ENSG00000144040 | 2 | 73159165 | 73312747 | 297 | 31 | 41888 | 1.1831 | 0.11839 | *SFXN5* |
| ENSG00000168259 | 17 | 40118451 | 40183394 | 96 | 17 | 45444 | 1.1828 | 0.11844 | *DNAJC7* |
| ENSG00000039987 | 19 | 12852516 | 12879272 | 28 | 17 | 43917 | 1.1826 | 0.11848 | *BEST2* |
| ENSG00000185559 | 14 | 101182042 | 101211539 | 91 | 26 | 44868 | 1.1826 | 0.11849 | *DLK1* |
| ENSG00000037749 | 5 | 153408466 | 153610038 | 604 | 53 | 46089 | 1.1825 | 0.1185 | *MFAP3* |
| ENSG00000142657 | 1 | 10448649 | 10490201 | 130 | 21 | 51378 | 1.1824 | 0.11852 | *PGD* |
| ENSG00000151687 | 2 | 190529016 | 190635919 | 223 | 28 | 48472 | 1.1823 | 0.11854 | *ANKAR* |
| ENSG00000159259 | 21 | 37747676 | 37801313 | 179 | 22 | 41215 | 1.1823 | 0.11854 | *CHAF1B* |
| ENSG00000138101 | 2 | 25590067 | 25906503 | 566 | 77 | 45017 | 1.1819 | 0.11863 | *DTNB* |
| ENSG00000181781 | 19 | 453346 | 484983 | 150 | 29 | 39553 | 1.1818 | 0.11864 | *ODF3L2* |
| ENSG00000170561 | 5 | 2735959 | 2762969 | 73 | 19 | 42764 | 1.1815 | 0.1187 | *IRX2* |
| ENSG00000204673 | 19 | 50362295 | 50391716 | 65 | 12 | 44459 | 1.1815 | 0.1187 | *AKT1S1* |
| ENSG00000124116 | 20 | 44366583 | 44430571 | 304 | 22 | 39136 | 1.1814 | 0.11871 | *WFDC3* |
| ENSG00000177954 | 1 | 153953235 | 153974626 | 38 | 16 | 52818 | 1.1814 | 0.11872 | *RPS27* |
| ENSG00000168159 | 1 | 228664762 | 228693467 | 74 | 15 | 44439 | 1.1814 | 0.11872 | *RNF187* |
| ENSG00000160870 | 7 | 99292660 | 99342819 | 94 | 7 | 27954 | 1.1811 | 0.11878 | *CYP3A7* |
| ENSG00000029363 | 6 | 136568001 | 136620989 | 41 | 19 | 51095 | 1.1811 | 0.11879 | *BCLAF1* |
| ENSG00000164451 | 6 | 116840174 | 116890031 | 136 | 16 | 41118 | 1.1807 | 0.11885 | *FAM26D* |
| ENSG00000147862 | 9 | 14071842 | 14408982 | 777 | 158 | 41566 | 1.1806 | 0.11888 | *NFIB* |
| ENSG00000179941 | 12 | 76728254 | 76752222 | 46 | 13 | 37405 | 1.1806 | 0.11888 | *BBS10* |
| ENSG00000196408 | 16 | 2018918 | 2044193 | 136 | 18 | 49563 | 1.1806 | 0.11888 | *NOXO1* |
| ENSG00000160117 | 19 | 17382454 | 17408455 | 125 | 15 | 43703 | 1.1805 | 0.11889 | *ANKLE1* |
| ENSG00000157778 | 7 | 1596966 | 1620641 | 77 | 15 | 49000 | 1.1804 | 0.11892 | *PSMG3* |
| ENSG00000212126 | 12 | 11128512 | 11149511 | 105 | 9 | 27191 | 1.1801 | 0.11897 | *TAS2R50* |
| ENSG00000243789 | 15 | 42110283 | 42139779 | 95 | 13 | 44849 | 1.18 | 0.119 | *JMJD7* |
| ENSG00000104695 | 8 | 30621973 | 30681830 | 169 | 19 | 36789 | 1.1797 | 0.11907 | *PPP2CB* |
| ENSG00000232263 | 21 | 31651463 | 31671832 | 84 | 16 | 39255 | 1.1796 | 0.11908 | *KRTAP25-1* |
| ENSG00000091039 | 12 | 76735577 | 76963589 | 483 | 34 | 38479 | 1.1796 | 0.11909 | *OSBPL8* |
| ENSG00000269095 | 19 | 17336749 | 17376257 | 162 | 35 | 46149 | 1.1796 | 0.11909 | *AC010646.3* |
| ENSG00000120860 | 12 | 102396705 | 102465927 | 149 | 24 | 53421 | 1.1795 | 0.11911 | *CCDC53* |
| ENSG00000198931 | 16 | 88865747 | 88888352 | 172 | 26 | 47572 | 1.1793 | 0.11914 | *APRT* |

| | | | | | | | | | |
|---|---|---|---|---|---|---|---|---|---|
| ENSG00000206536 | 3 | 98099510 | 98120475 | 82 | 11 | 47698 | 1.1791 | 0.11917 | *OR5K3* |
| ENSG00000198832 | 22 | 31490758 | 31526055 | 118 | 21 | 43094 | 1.1789 | 0.11921 | *SELM* |
| ENSG00000121361 | 12 | 21907889 | 21938515 | 23 | 12 | 55196 | 1.1783 | 0.11934 | *KCNJ8* |
| ENSG00000143013 | 1 | 87784151 | 87824606 | 102 | 31 | 40223 | 1.1782 | 0.11936 | *LMO4* |
| ENSG00000173227 | 11 | 66764249 | 66828334 | 81 | 32 | 46838 | 1.1782 | 0.11936 | *SYT12* |
| ENSG00000159917 | 19 | 44722882 | 44819199 | 189 | 19 | 35156 | 1.1781 | 0.11938 | *ZNF235* |
| ENSG00000186431 | 19 | 55375704 | 55411838 | 326 | 40 | 47378 | 1.1777 | 0.11946 | *FCAR* |
| ENSG00000066697 | 9 | 103179438 | 103223511 | 134 | 11 | 49204 | 1.1775 | 0.1195 | *MSANTD3* |
| ENSG00000115423 | 2 | 84733579 | 85056713 | 909 | 60 | 44323 | 1.1773 | 0.11954 | *DNAH6* |
| ENSG00000174914 | 11 | 56457864 | 56478781 | 72 | 13 | 46476 | 1.177 | 0.1196 | *OR9G1* |
| ENSG00000269047 | 16 | 1021252 | 1041663 | 121 | 23 | 42250 | 1.1769 | 0.11962 | *AC009041.2* |
| ENSG00000111424 | 12 | 48225320 | 48346831 | 375 | 61 | 44061 | 1.1767 | 0.11966 | *VDR* |
| ENSG00000136487 | 17 | 61947578 | 61969295 | 84 | 9 | 20188 | 1.1756 | 0.11987 | *GH2* |
| ENSG00000107331 | 9 | 139891686 | 139933367 | 120 | 23 | 54417 | 1.1754 | 0.11993 | *ABCA2* |
| ENSG00000126778 | 14 | 61100133 | 61134977 | 48 | 12 | 39552 | 1.1752 | 0.11996 | *SIX1* |
| ENSG00000160953 | 19 | 1275890 | 1388430 | 270 | 68 | 44276 | 1.1751 | 0.11998 | *MUM1* |
| ENSG00000151014 | 4 | 139926943 | 139976906 | 94 | 24 | 47055 | 1.1751 | 0.11999 | *CCRN4L* |
| ENSG00000156795 | 8 | 124418965 | 124489470 | 363 | 14 | 46986 | 1.1749 | 0.12001 | *WDYHV1* |
| ENSG00000055957 | 3 | 52801603 | 52836078 | 82 | 10 | 50619 | 1.1746 | 0.12007 | *ITIH1* |
| ENSG00000186665 | 17 | 65977217 | 65999765 | 35 | 8 | 33872 | 1.1745 | 0.12009 | *C17orf58* |
| ENSG00000182950 | 15 | 76006318 | 76030029 | 48 | 8 | 47828 | 1.1745 | 0.1201 | *ODF3L1* |
| ENSG00000182255 | 11 | 30021288 | 30048570 | 84 | 15 | 51217 | 1.1744 | 0.12012 | *KCNA4* |
| ENSG00000269307 | 19 | 17368227 | 17403775 | 149 | 20 | 43978 | 1.1743 | 0.12013 | *CTD-2278I10.6* |
| ENSG00000169676 | 4 | 9773258 | 9795632 | 61 | 12 | 36142 | 1.1743 | 0.12014 | *DRD5* |
| ENSG00000054983 | 14 | 88294164 | 88470009 | 890 | 62 | 43970 | 1.1743 | 0.12014 | *GALC* |
| ENSG00000154736 | 21 | 28280231 | 28348832 | 245 | 32 | 43797 | 1.1741 | 0.12018 | *ADAMTS5* |
| ENSG00000178988 | 4 | 6699428 | 6721607 | 45 | 17 | 42106 | 1.1738 | 0.12024 | *MRFAP1L1* |
| ENSG00000180096 | 16 | 30379454 | 30417312 | 65 | 15 | 41326 | 1.1736 | 0.12029 | *Sep-01* |
| ENSG00000164221 | 5 | 114592885 | 114642528 | 107 | 22 | 45321 | 1.1733 | 0.12033 | *CCDC112* |
| ENSG00000125257 | 13 | 95662083 | 95963687 | 1318 | 146 | 39230 | 1.1733 | 0.12033 | *ABCC4* |
| ENSG00000205022 | 16 | 88918034 | 88943068 | 106 | 18 | 39621 | 1.1732 | 0.12036 | *PABPN1L* |
| ENSG00000141198 | 17 | 52966748 | 53049310 | 306 | 31 | 51369 | 1.1732 | 0.12036 | *TOM1L1* |
| ENSG00000140022 | 14 | 81717000 | 81912809 | 581 | 68 | 44801 | 1.1731 | 0.12037 | *STON2* |
| ENSG00000124212 | 20 | 48110411 | 48194683 | 178 | 46 | 45132 | 1.173 | 0.1204 | *PTGIS* |
| ENSG00000146070 | 6 | 46661938 | 46713430 | 102 | 19 | 34716 | 1.1728 | 0.12043 | *PLA2G7* |
| ENSG00000006747 | 7 | 12600203 | 12703228 | 449 | 77 | 49751 | 1.1727 | 0.12045 | *SCIN* |

| | | | | | | | | | |
|---|---|---|---|---|---|---|---|---|---|
| ENSG00000175567 | 11 | 73675712 | 73704352 | 90 | 14 | 43336 | 1.1723 | 0.12054 | *UCP2* |
| ENSG00000163993 | 4 | 6684796 | 6708897 | 83 | 24 | 37627 | 1.1717 | 0.12066 | *S100P* |
| ENSG00000152484 | 13 | 27630293 | 27756033 | 422 | 51 | 44461 | 1.1717 | 0.12066 | *USP12* |
| ENSG00000025434 | 11 | 47259851 | 47300396 | 91 | 14 | 41296 | 1.1711 | 0.12078 | *NR1H3* |
| ENSG00000134545 | 12 | 10584863 | 10617284 | 85 | 15 | 42754 | 1.1708 | 0.12084 | *KLRC1* |
| ENSG00000255622 | 5 | 140525577 | 140548639 | 82 | 14 | 42780 | 1.1706 | 0.12088 | *PCDHB17* |
| ENSG00000249884 | 2 | 86722791 | 86958245 | 556 | 58 | 42022 | 1.1702 | 0.12096 | *RNF103-CHMP3* |
| ENSG00000028310 | 5 | 840406 | 902939 | 159 | 19 | 41952 | 1.1702 | 0.12097 | *BRD9* |
| ENSG00000186051 | 9 | 108414738 | 108435367 | 64 | 13 | 37934 | 1.1702 | 0.12097 | *TAL2* |
| ENSG00000138835 | 9 | 116197011 | 116370018 | 505 | 26 | 55539 | 1.17 | 0.12099 | *RGS3* |
| ENSG00000164077 | 3 | 49936302 | 49977606 | 64 | 11 | 39782 | 1.1697 | 0.12106 | *MON1A* |
| ENSG00000129292 | 8 | 133777618 | 133871052 | 205 | 18 | 50023 | 1.1697 | 0.12107 | *PHF20L1* |
| ENSG00000129946 | 19 | 406583 | 470996 | 291 | 57 | 39259 | 1.1696 | 0.12109 | *SHC2* |
| ENSG00000115561 | 2 | 86720554 | 86958245 | 566 | 59 | 41903 | 1.1693 | 0.12114 | *CHMP3* |
| ENSG00000128699 | 2 | 190625049 | 190659097 | 90 | 15 | 46368 | 1.1692 | 0.12117 | *ORMDL1* |
| ENSG00000213934 | 11 | 5259313 | 5281122 | 68 | 10 | 37277 | 1.1689 | 0.12122 | *HBG1* |
| ENSG00000112033 | 6 | 35300335 | 35405968 | 309 | 36 | 28252 | 1.1688 | 0.12123 | *PPARD* |
| ENSG00000196357 | 19 | 36663188 | 36747159 | 286 | 26 | 52993 | 1.1688 | 0.12123 | *ZNF565* |
| ENSG00000067365 | 16 | 8705540 | 8750081 | 164 | 18 | 49211 | 1.1687 | 0.12126 | *METTL22* |
| ENSG00000128881 | 15 | 43020932 | 43223007 | 277 | 36 | 36291 | 1.1685 | 0.12129 | *TTBK2* |
| ENSG00000106327 | 7 | 100208039 | 100250402 | 80 | 12 | 55015 | 1.168 | 0.12139 | *TFR2* |
| ENSG00000092068 | 14 | 23584504 | 23662883 | 208 | 59 | 42184 | 1.168 | 0.1214 | *SLC7A8* |
| ENSG00000233828 | 5 | 86502423 | 86544822 | 64 | 17 | 45391 | 1.1678 | 0.12144 | *AC008394.1* |
| ENSG00000123700 | 17 | 68154814 | 68186189 | 59 | 13 | 50625 | 1.1676 | 0.12149 | *KCNJ2* |
| ENSG00000116793 | 1 | 114229453 | 114312111 | 162 | 19 | 48476 | 1.1674 | 0.12152 | *PHTF1* |
| ENSG00000196119 | 11 | 124429893 | 124451037 | 85 | 16 | 55013 | 1.1669 | 0.12163 | *OR8A1* |
| ENSG00000180376 | 3 | 56581189 | 56665846 | 402 | 34 | 42638 | 1.1668 | 0.12164 | *CCDC66* |
| ENSG00000167112 | 9 | 131061382 | 131095021 | 83 | 15 | 35683 | 1.1668 | 0.12165 | *TRUB2* |
| ENSG00000149262 | 11 | 77579766 | 77715724 | 417 | 32 | 52271 | 1.1666 | 0.12169 | *INTS4* |
| ENSG00000179934 | 3 | 39361197 | 39385171 | 78 | 17 | 40356 | 1.1662 | 0.12177 | *CCR8* |
| ENSG00000203993 | 9 | 140499784 | 140523358 | 51 | 10 | 47594 | 1.1661 | 0.12178 | *C9orf37* |
| ENSG00000090659 | 19 | 7794879 | 7822464 | 113 | 25 | 40303 | 1.1661 | 0.12178 | *CD209* |
| ENSG00000164743 | 8 | 13414352 | 13435796 | 135 | 26 | 53391 | 1.1661 | 0.12179 | *C8orf48* |
| ENSG00000184076 | 22 | 30153358 | 30176402 | 75 | 16 | 38883 | 1.1661 | 0.1218 | *UQCR10* |
| ENSG00000196329 | 7 | 150409341 | 150457121 | 166 | 24 | 49169 | 1.1656 | 0.12188 | *GIMAP5* |
| ENSG00000159251 | 15 | 35070297 | 35098340 | 112 | 40 | 47942 | 1.1651 | 0.122 | *ACTC1* |

| | | | | | | | | | |
|---|---|---|---|---|---|---|---|---|---|
| ENSG00000126561 | 17 | 40429565 | 40473961 | 85 | 13 | 38221 | 1.1651 | 0.122 | *STAT5A* |
| ENSG00000179615 | 12 | 55958199 | 55979128 | 82 | 11 | 36861 | 1.1647 | 0.12206 | *OR2AP1* |
| ENSG00000080298 | 9 | 3208297 | 3536004 | 790 | 72 | 37628 | 1.1644 | 0.12213 | *RFX3* |
| ENSG00000163075 | 2 | 120292008 | 120429827 | 269 | 32 | 47697 | 1.1642 | 0.12217 | *PCDP1* |
| ENSG00000132823 | 20 | 42815136 | 42849431 | 131 | 26 | 32990 | 1.1641 | 0.12219 | *OSER1* |
| ENSG00000114391 | 3 | 101389935 | 101415626 | 61 | 11 | 55633 | 1.164 | 0.12221 | *RPL24* |
| ENSG00000149476 | 11 | 61090682 | 61130767 | 21 | 13 | 44584 | 1.164 | 0.12221 | *DAK* |
| ENSG00000030066 | 11 | 47789639 | 47880107 | 188 | 19 | 35747 | 1.1639 | 0.12224 | *NUP160* |
| ENSG00000073169 | 22 | 50629408 | 50666045 | 153 | 30 | 44395 | 1.1636 | 0.1223 | *SELO* |
| ENSG00000008513 | 8 | 134457091 | 134594183 | 552 | 119 | 44694 | 1.1631 | 0.1224 | *ST3GAL1* |
| ENSG00000263812 | 18 | 74230612 | 74332925 | 509 | 86 | 42325 | 1.1629 | 0.12243 | *LINC00908* |
| ENSG00000256100 | 11 | 63732092 | 63765818 | 62 | 8 | 27111 | 1.1628 | 0.12245 | *AP000721.4* |
| ENSG00000167754 | 19 | 51436559 | 51466349 | 128 | 24 | 35264 | 1.1628 | 0.12245 | *KLK5* |
| ENSG00000144644 | 3 | 30757692 | 30946257 | 677 | 71 | 41285 | 1.1628 | 0.12246 | *GADL1* |
| ENSG00000166801 | 11 | 58900221 | 58932512 | 93 | 7 | 55266 | 1.1627 | 0.12247 | *FAM111A* |
| ENSG00000134817 | 11 | 56991051 | 57014709 | 66 | 17 | 52380 | 1.1626 | 0.12249 | *APLNR* |
| ENSG00000071894 | 8 | 145608444 | 145644753 | 67 | 14 | 35695 | 1.1622 | 0.12257 | *CPSF1* |
| ENSG00000198556 | 7 | 99060464 | 99111273 | 66 | 17 | 40482 | 1.1622 | 0.12258 | *ZNF789* |
| ENSG00000148824 | 10 | 135197598 | 135244811 | 206 | 29 | 42193 | 1.1622 | 0.12258 | *MTG1* |
| ENSG00000169019 | 4 | 47442885 | 47475736 | 100 | 15 | 41608 | 1.162 | 0.12263 | *COMMD8* |
| ENSG00000181935 | 11 | 55329604 | 55350536 | 181 | 17 | 50905 | 1.1619 | 0.12265 | *OR4C16* |
| ENSG00000138107 | 10 | 104228986 | 104272482 | 146 | 10 | 50060 | 1.1618 | 0.12266 | *ACTR1A* |
| ENSG00000188690 | 10 | 127467146 | 127521817 | 126 | 9 | 55738 | 1.1618 | 0.12266 | *UROS* |
| ENSG00000036257 | 2 | 225324867 | 225460110 | 404 | 24 | 55556 | 1.1614 | 0.12273 | *CUL3* |
| ENSG00000134109 | 3 | 5219331 | 5271642 | 140 | 28 | 34714 | 1.1614 | 0.12274 | *EDEM1* |
| ENSG00000204472 | 6 | 31572961 | 31594798 | 90 | 18 | 36825 | 1.1613 | 0.12276 | *AIF1* |
| ENSG00000152457 | 10 | 14929358 | 15006431 | 217 | 46 | 46021 | 1.1612 | 0.12279 | *DCLRE1C* |
| ENSG00000227782 | 17 | 15933505 | 15953994 | 49 | 8 | 47326 | 1.1605 | 0.12293 | *AC002553.1* |
| ENSG00000125166 | 16 | 58731035 | 58778261 | 191 | 23 | 45095 | 1.1603 | 0.12296 | *GOT2* |
| ENSG00000065268 | 19 | 973174 | 1004569 | 213 | 28 | 46532 | 1.1603 | 0.12296 | *WDR18* |
| ENSG00000197153 | 6 | 27848093 | 27870884 | 60 | 13 | 40580 | 1.1601 | 0.123 | *HIST1H3J* |
| ENSG00000231861 | 3 | 98206448 | 98227496 | 51 | 16 | 35055 | 1.16 | 0.12302 | *OR5K2* |
| ENSG00000188163 | 9 | 140128036 | 140152222 | 72 | 15 | 46998 | 1.16 | 0.12303 | *FAM166A* |
| ENSG00000254834 | 11 | 56334212 | 56355263 | 76 | 10 | 50577 | 1.1596 | 0.12311 | *OR5M10* |
| ENSG00000129245 | 17 | 7484548 | 7528189 | 89 | 16 | 42176 | 1.1595 | 0.12312 | *FXR2* |
| ENSG00000133488 | 22 | 30874900 | 30911698 | 162 | 19 | 50879 | 1.1595 | 0.12313 | *SEC14L4* |

| | | | | | | | | | |
|---|---|---|---|---|---|---|---|---|---|
| ENSG00000187105 | 14 | 73935189 | 74035651 | 210 | 48 | 41320 | 1.159 | 0.12323 | *HEATR4* |
| ENSG00000079931 | 6 | 132607194 | 132732684 | 330 | 49 | 34374 | 1.1588 | 0.12326 | *MOXD1* |
| ENSG00000175826 | 17 | 7136910 | 7165810 | 38 | 6 | 56442 | 1.1588 | 0.12326 | *CTDNEP1* |
| ENSG00000074755 | 17 | 3897739 | 4056314 | 611 | 41 | 47774 | 1.1587 | 0.12328 | *ZZEF1* |
| ENSG00000224586 | 6 | 28483702 | 28512729 | 75 | 17 | 42684 | 1.1587 | 0.12329 | *GPX5* |
| ENSG00000258890 | 17 | 62492706 | 62548579 | 95 | 20 | 43587 | 1.1586 | 0.12331 | *CEP95* |
| ENSG00000112200 | 6 | 56941642 | 57045105 | 140 | 14 | 53871 | 1.1584 | 0.12334 | *ZNF451* |
| ENSG00000120159 | 9 | 26830683 | 26902802 | 198 | 32 | 45586 | 1.1584 | 0.12335 | *CAAP1* |
| ENSG00000177098 | 11 | 117994092 | 118033603 | 80 | 23 | 45980 | 1.1584 | 0.12335 | *SCN4B* |
| ENSG00000215621 | 17 | 79770293 | 79794533 | 68 | 10 | 54038 | 1.1583 | 0.12338 | *AC174470.1* |
| ENSG00000196092 | 9 | 36823272 | 37044103 | 860 | 123 | 48488 | 1.1578 | 0.12348 | *PAX5* |
| ENSG00000110700 | 11 | 17085936 | 17109334 | 52 | 11 | 42359 | 1.1577 | 0.12349 | *RPS13* |
| ENSG00000144824 | 3 | 111441344 | 111705364 | 1092 | 93 | 45424 | 1.1575 | 0.12354 | *PHLDB2* |
| ENSG00000267883 | 21 | 47173565 | 47200005 | 48 | 8 | 49788 | 1.1573 | 0.12357 | *PRED60* |
| ENSG00000187837 | 6 | 26045968 | 26066699 | 54 | 12 | 33475 | 1.1573 | 0.12358 | *HIST1H1C* |
| ENSG00000214102 | 7 | 141398153 | 141441071 | 71 | 9 | 51327 | 1.1566 | 0.12371 | *WEE2* |
| ENSG00000264194 | 17 | 10038831 | 10060277 | 110 | 30 | 45262 | 1.1564 | 0.12376 | *AC000003.2* |
| ENSG00000129450 | 19 | 51618165 | 51649908 | 45 | 22 | 34824 | 1.1563 | 0.12378 | *SIGLEC9* |
| ENSG00000226979 | 6 | 31529831 | 31552101 | 97 | 22 | 50690 | 1.1563 | 0.12379 | *LTA* |
| ENSG00000110200 | 11 | 71807424 | 71833826 | 65 | 7 | 29059 | 1.156 | 0.12384 | *ANAPC15* |
| ENSG00000145293 | 4 | 83341715 | 83392244 | 154 | 30 | 42963 | 1.1557 | 0.12391 | *ENOPH1* |
| ENSG00000196352 | 1 | 207484853 | 207544311 | 140 | 16 | 46318 | 1.155 | 0.12405 | *CD55* |
| ENSG00000143434 | 1 | 151094161 | 151129104 | 82 | 19 | 41493 | 1.1548 | 0.12408 | *SEMA6C* |
| ENSG00000188215 | 16 | 20856247 | 20921706 | 122 | 19 | 39396 | 1.1548 | 0.12408 | *DCUN1D3* |
| ENSG00000124208 | 20 | 48687661 | 48780174 | 235 | 24 | 48757 | 1.1545 | 0.12415 | *TMEM189-UBE2V1* |
| ENSG00000240849 | 20 | 48687663 | 48780335 | 235 | 24 | 48757 | 1.1545 | 0.12415 | *TMEM189* |
| ENSG00000172381 | 11 | 57788402 | 57809403 | 104 | 13 | 49505 | 1.1539 | 0.12426 | *OR6Q1* |
| ENSG00000141522 | 17 | 79815597 | 79839282 | 68 | 9 | 54664 | 1.1539 | 0.12426 | *ARHGDIA* |
| ENSG00000173621 | 11 | 66614118 | 66637946 | 28 | 11 | 49436 | 1.1539 | 0.12427 | *LRFN4* |
| ENSG00000198569 | 9 | 140115209 | 140141006 | 67 | 17 | 52866 | 1.1536 | 0.12433 | *SLC34A3* |
| ENSG00000270757 | 2 | 198355137 | 198425450 | 106 | 16 | 53866 | 1.1531 | 0.12444 | *HSPE1-MOB4* |
| ENSG00000138246 | 3 | 132126370 | 132267876 | 332 | 39 | 43548 | 1.1525 | 0.12455 | *DNAJC13* |
| ENSG00000169900 | 16 | 31217283 | 31238680 | 19 | 9 | 44139 | 1.1525 | 0.12455 | *PYDC1* |
| ENSG00000267706 | 19 | 55853669 | 55880884 | 84 | 22 | 44349 | 1.1524 | 0.12457 | *CTD-2105E13.6* |
| ENSG00000189362 | 2 | 191359068 | 191409448 | 126 | 19 | 45542 | 1.1522 | 0.12463 | *TMEM194B* |
| ENSG00000164114 | 4 | 156253810 | 156308122 | 234 | 20 | 32367 | 1.152 | 0.12466 | *MAP9* |

| | | | | | | | | | |
|---|---|---|---|---|---|---|---|---|---|
| ENSG00000171208 | 16 | 47101614 | 47187908 | 82 | 13 | 39696 | 1.1519 | 0.12469 | *NETO2* |
| ENSG00000188612 | 17 | 73153408 | 73189078 | 95 | 22 | 37186 | 1.1517 | 0.12472 | *SUMO2* |
| ENSG00000180305 | 20 | 44248165 | 44269835 | 83 | 8 | 44393 | 1.1517 | 0.12472 | *WFDC10A* |
| ENSG00000014824 | 4 | 41982489 | 42102474 | 477 | 18 | 30532 | 1.1516 | 0.12475 | *SLC30A9* |
| ENSG00000139998 | 14 | 65402532 | 65449494 | 139 | 28 | 43692 | 1.1515 | 0.12476 | *RAB15* |
| ENSG00000066056 | 1 | 43756664 | 43798779 | 61 | 12 | 36673 | 1.1509 | 0.12488 | *TIE1* |
| ENSG00000105369 | 19 | 42371190 | 42395439 | 19 | 4 | 44038 | 1.1509 | 0.12488 | *CD79A* |
| ENSG00000128534 | 7 | 117814086 | 117842878 | 149 | 11 | 51678 | 1.1508 | 0.12491 | *NAA38* |
| ENSG00000130158 | 19 | 11299971 | 11383157 | 243 | 33 | 39126 | 1.1505 | 0.12498 | *DOCK6* |
| ENSG00000140612 | 15 | 85202775 | 85269947 | 158 | 24 | 47163 | 1.1503 | 0.12501 | *SEC11A* |
| ENSG00000065029 | 6 | 35216686 | 35273762 | 140 | 17 | 30894 | 1.1501 | 0.12505 | *ZNF76* |
| ENSG00000156966 | 2 | 232250254 | 232275875 | 74 | 16 | 41669 | 1.15 | 0.12507 | *B3GNT7* |
| ENSG00000185821 | 12 | 55810038 | 55830976 | 56 | 9 | 52635 | 1.15 | 0.12507 | *OR6C76* |
| ENSG00000197780 | 1 | 109595108 | 109628624 | 136 | 24 | 50753 | 1.1497 | 0.12513 | *TAF13* |
| ENSG00000095739 | 10 | 28956271 | 28981868 | 68 | 17 | 52536 | 1.1496 | 0.12515 | *BAMBI* |
| ENSG00000225119 | 10 | 38707074 | 38746820 | 136 | 12 | 45879 | 1.1495 | 0.12518 | *LINC00999* |
| ENSG00000204020 | 10 | 90511163 | 90547999 | 163 | 23 | 47154 | 1.1495 | 0.12518 | *LIPN* |
| ENSG00000096264 | 6 | 41293393 | 41328625 | 183 | 24 | 39464 | 1.1491 | 0.12526 | *NCR2* |
| ENSG00000185267 | 10 | 14851249 | 14890574 | 97 | 21 | 38512 | 1.1489 | 0.12529 | *CDNF* |
| ENSG00000149635 | 20 | 45159585 | 45189213 | 88 | 22 | 45299 | 1.1488 | 0.12533 | *OCSTAMP* |
| ENSG00000256591 | 11 | 61186692 | 61263294 | 126 | 17 | 37654 | 1.1487 | 0.12534 | *RP11-286N22.8* |
| ENSG00000176797 | 8 | 7728726 | 7750186 | 3 | 1 | 43121 | 1.1474 | 0.1256 | *DEFB103A* |
| ENSG00000109062 | 17 | 72734791 | 72775492 | 96 | 18 | 44705 | 1.1474 | 0.1256 | *SLC9A3R1* |
| ENSG00000205358 | 16 | 56693726 | 56715041 | 113 | 32 | 41668 | 1.1473 | 0.12562 | *MT1H* |
| ENSG00000197406 | 14 | 102017688 | 102039789 | 46 | 14 | 45363 | 1.1472 | 0.12565 | *DIO3* |
| ENSG00000111206 | 12 | 2956847 | 2996206 | 89 | 16 | 34500 | 1.147 | 0.12569 | *FOXM1* |
| ENSG00000142606 | 1 | 2512078 | 2574481 | 229 | 16 | 32388 | 1.1465 | 0.12578 | *MMEL1* |
| ENSG00000138760 | 4 | 77069890 | 77145046 | 298 | 24 | 33167 | 1.1461 | 0.12588 | *SCARB2* |
| ENSG00000130513 | 19 | 18475541 | 18509986 | 117 | 18 | 47683 | 1.1459 | 0.12592 | *GDF15* |
| ENSG00000124107 | 20 | 43870880 | 43893205 | 85 | 9 | 40427 | 1.1458 | 0.12594 | *SLPI* |
| ENSG00000137720 | 11 | 111739659 | 111766699 | 62 | 9 | 43326 | 1.1455 | 0.126 | *C11orf1* |
| ENSG00000198467 | 9 | 35671989 | 35701017 | 80 | 20 | 50415 | 1.1455 | 0.12601 | *TPM2* |
| ENSG00000176340 | 11 | 63732079 | 63754015 | 44 | 7 | 28242 | 1.1455 | 0.12601 | *COX8A* |
| ENSG00000100626 | 14 | 69715994 | 69831183 | 435 | 72 | 43301 | 1.1453 | 0.12605 | *GALNT16* |
| ENSG00000132334 | 10 | 129695325 | 129894119 | 871 | 167 | 52316 | 1.1452 | 0.12607 | *PTPRE* |
| ENSG00000105248 | 19 | 4237076 | 4279087 | 129 | 11 | 32694 | 1.1449 | 0.12612 | *CCDC94* |

| | | | | | | | | | |
|---|---|---|---|---|---|---|---|---|---|
| ENSG00000106635 | 7 | 72940686 | 72982332 | 85 | 13 | 52817 | 1.1449 | 0.12613 | *BCL7B* |
| ENSG00000176909 | 19 | 49205999 | 49232978 | 96 | 9 | 25243 | 1.1449 | 0.12613 | *MAMSTR* |
| ENSG00000163635 | 3 | 63840233 | 63999138 | 351 | 44 | 45583 | 1.1444 | 0.12623 | *ATXN7* |
| ENSG00000059769 | 9 | 114383632 | 114442508 | 254 | 35 | 40438 | 1.1444 | 0.12623 | *DNAJC25* |
| ENSG00000244115 | 9 | 114383666 | 114442509 | 254 | 35 | 40438 | 1.1444 | 0.12623 | *DNAJC25-GNG10* |
| ENSG00000269197 | 8 | 70840403 | 70860635 | 91 | 14 | 53841 | 1.1443 | 0.12624 | *AC090574.1* |
| ENSG00000151348 | 11 | 44107099 | 44276979 | 360 | 37 | 52164 | 1.1444 | 0.12624 | *EXT2* |
| ENSG00000234409 | 22 | 20125950 | 20148399 | 86 | 15 | 48940 | 1.1442 | 0.12627 | *AC006547.14* |
| ENSG00000106341 | 7 | 31716329 | 31758069 | 133 | 29 | 47634 | 1.144 | 0.12631 | *PPP1R17* |
| ENSG00000164219 | 5 | 114536527 | 114608569 | 168 | 28 | 38270 | 1.144 | 0.12632 | *PGGT1B* |
| ENSG00000136286 | 7 | 44992261 | 45028697 | 99 | 18 | 47021 | 1.1439 | 0.12633 | *MYO1G* |
| ENSG00000203791 | 10 | 126426718 | 126490439 | 142 | 22 | 47048 | 1.1439 | 0.12633 | *METTL10* |
| ENSG00000167770 | 11 | 63743325 | 63779283 | 74 | 12 | 30357 | 1.1439 | 0.12634 | *OTUB1* |
| ENSG00000159216 | 21 | 36150098 | 37386965 | 3991 | 312 | 45528 | 1.1439 | 0.12634 | *RUNX1* |
| ENSG00000198721 | 6 | 4105923 | 4145831 | 127 | 15 | 42249 | 1.1438 | 0.12636 | *ECI2* |
| ENSG00000089157 | 12 | 120624489 | 120649038 | 43 | 12 | 31125 | 1.1437 | 0.12638 | *RPLP0* |
| ENSG00000161010 | 5 | 179251436 | 179299173 | 183 | 26 | 39871 | 1.1436 | 0.1264 | *C5orf45* |
| ENSG00000079215 | 5 | 36596457 | 36698436 | 276 | 56 | 40399 | 1.1435 | 0.12641 | *SLC1A3* |
| ENSG00000204859 | 1 | 6630061 | 6659340 | 63 | 20 | 50298 | 1.1434 | 0.12644 | *ZBTB48* |
| ENSG00000114744 | 3 | 149446257 | 149480286 | 93 | 14 | 47372 | 1.1434 | 0.12644 | *COMMD2* |
| ENSG00000142089 | 11 | 309669 | 337537 | 83 | 22 | 39075 | 1.1432 | 0.12648 | *IFITM3* |
| ENSG00000198720 | 17 | 27906787 | 27951779 | 69 | 22 | 41965 | 1.1432 | 0.12648 | *ANKRD13B* |
| ENSG00000133247 | 19 | 55841221 | 55869488 | 91 | 20 | 47114 | 1.1429 | 0.12654 | *SUV420H2* |
| ENSG00000169126 | 10 | 28054115 | 28297977 | 638 | 50 | 43832 | 1.1426 | 0.1266 | *ARMC4* |
| ENSG00000080822 | 3 | 98206756 | 98251910 | 106 | 22 | 39493 | 1.1424 | 0.12663 | *CLDND1* |
| ENSG00000112335 | 6 | 108522426 | 108592464 | 137 | 22 | 40267 | 1.1425 | 0.12663 | *SNX3* |
| ENSG00000168026 | 3 | 39139152 | 39190394 | 178 | 24 | 34833 | 1.1424 | 0.12665 | *TTC21A* |
| ENSG00000081154 | 3 | 101282939 | 101323281 | 117 | 16 | 50030 | 1.1423 | 0.12666 | *PCNP* |
| ENSG00000114650 | 3 | 47445203 | 47528616 | 101 | 18 | 47457 | 1.1417 | 0.12679 | *SCAP* |
| ENSG00000113494 | 5 | 35038861 | 35240794 | 711 | 78 | 37287 | 1.1416 | 0.12681 | *PRLR* |
| ENSG00000256713 | 11 | 60998514 | 61028920 | 20 | 11 | 36756 | 1.1415 | 0.12684 | *PGA5* |
| ENSG00000130287 | 19 | 19312782 | 19373042 | 153 | 26 | 38877 | 1.1408 | 0.12699 | *NCAN* |
| ENSG00000112706 | 6 | 76620832 | 76792395 | 387 | 63 | 49436 | 1.1405 | 0.12705 | *IMPG1* |
| ENSG00000105287 | 19 | 47167532 | 47230384 | 155 | 24 | 47768 | 1.1405 | 0.12705 | *PRKD2* |
| ENSG00000175198 | 13 | 100731269 | 101192686 | 1055 | 74 | 38788 | 1.1403 | 0.12708 | *PCCA* |
| ENSG00000205085 | 7 | 128302342 | 128336929 | 184 | 15 | 54917 | 1.1402 | 0.1271 | *FAM71F2* |

| | | | | | | | | | |
|---|---|---|---|---|---|---|---|---|---|
| ENSG00000239887 | 1 | 162326698 | 162366600 | 146 | 36 | 45959 | 1.1398 | 0.12719 | *C1orf226* |
| ENSG00000106462 | 7 | 148494475 | 148591413 | 231 | 32 | 52774 | 1.1397 | 0.1272 | *EZH2* |
| ENSG00000067798 | 12 | 78214685 | 78616790 | 1490 | 156 | 48803 | 1.1396 | 0.12723 | *NAV3* |
| ENSG00000003393 | 2 | 202555277 | 202655912 | 356 | 32 | 38012 | 1.1395 | 0.12724 | *ALS2* |
| ENSG00000079337 | 12 | 48118455 | 48174823 | 206 | 34 | 50057 | 1.1395 | 0.12725 | *RAPGEF3* |
| ENSG00000215883 | 1 | 54628009 | 54675709 | 110 | 35 | 47744 | 1.1394 | 0.12726 | *CYB5RL* |
| ENSG00000205155 | 19 | 36226015 | 36247911 | 58 | 13 | 49548 | 1.1393 | 0.12729 | *PSENEN* |
| ENSG00000141519 | 17 | 78000435 | 78084412 | 450 | 48 | 45259 | 1.1392 | 0.1273 | *CCDC40* |
| ENSG00000089693 | 12 | 6847170 | 6886641 | 145 | 21 | 39759 | 1.1387 | 0.12741 | *MLF2* |
| ENSG00000222040 | 2 | 96768707 | 96791984 | 33 | 7 | 50152 | 1.1383 | 0.12751 | *ADRA2B* |
| ENSG00000166106 | 11 | 130308869 | 130356532 | 160 | 40 | 47039 | 1.1383 | 0.12751 | *ADAMTS15* |
| ENSG00000169564 | 2 | 70304585 | 70326332 | 32 | 7 | 33767 | 1.138 | 0.12755 | *PCBP1* |
| ENSG00000176715 | 16 | 89144783 | 89232254 | 511 | 40 | 36393 | 1.138 | 0.12756 | *ACSF3* |
| ENSG00000213218 | 17 | 61939372 | 61961126 | 96 | 6 | 21009 | 1.138 | 0.12756 | *CSH2* |
| ENSG00000056586 | 9 | 125596835 | 125677620 | 155 | 27 | 43385 | 1.1379 | 0.12758 | *RC3H2* |
| ENSG00000203783 | 1 | 153180060 | 153201793 | 83 | 8 | 42375 | 1.1379 | 0.12759 | *PRR9* |
| ENSG00000160972 | 8 | 145693352 | 145737504 | 64 | 13 | 44243 | 1.1378 | 0.1276 | *PPP1R16A* |
| ENSG00000101193 | 20 | 61559471 | 61586996 | 96 | 20 | 47072 | 1.1376 | 0.12765 | *GID8* |
| ENSG00000143995 | 2 | 66650584 | 66811001 | 399 | 72 | 40351 | 1.1372 | 0.12773 | *MEIS1* |
| ENSG00000171385 | 1 | 112303284 | 112541777 | 774 | 137 | 43857 | 1.1368 | 0.1278 | *KCND3* |
| ENSG00000189253 | 11 | 89592450 | 89619185 | 2 | 1 | 43587 | 1.1367 | 0.12783 | *TRIM64B* |
| ENSG00000141052 | 17 | 12559207 | 12682266 | 405 | 77 | 40357 | 1.1367 | 0.12783 | *MYOCD* |
| ENSG00000136630 | 1 | 221041699 | 221068401 | 80 | 17 | 49363 | 1.1366 | 0.12785 | *HLX* |
| ENSG00000179142 | 8 | 143981975 | 144009259 | 116 | 13 | 32324 | 1.1365 | 0.12788 | *CYP11B2* |
| ENSG00000053501 | 19 | 17316155 | 17340638 | 134 | 20 | 39424 | 1.1361 | 0.12796 | *USE1* |
| ENSG00000091073 | 7 | 76080993 | 76145312 | 122 | 15 | 30143 | 1.136 | 0.12798 | *DTX2* |
| ENSG00000139687 | 13 | 48867887 | 49066122 | 252 | 29 | 35828 | 1.1359 | 0.12801 | *RB1* |
| ENSG00000115520 | 2 | 198308147 | 198350032 | 52 | 9 | 53099 | 1.1352 | 0.12814 | *COQ10B* |
| ENSG00000203864 | 1 | 117226734 | 117259225 | 59 | 16 | 43555 | 1.1351 | 0.12816 | *C1orf137* |
| ENSG00000174915 | 11 | 438268 | 501393 | 247 | 27 | 52505 | 1.1351 | 0.12818 | *PTDSS2* |
| ENSG00000255038 | 11 | 65824748 | 65847090 | 39 | 11 | 49379 | 1.135 | 0.12819 | *RP11-1167A19.2* |
| ENSG00000006652 | 7 | 112053023 | 112131072 | 305 | 42 | 39556 | 1.1349 | 0.12821 | *IFRD1* |
| ENSG00000143036 | 1 | 95275898 | 95370802 | 395 | 66 | 50508 | 1.1344 | 0.12832 | *SLC44A3* |
| ENSG00000253206 | 8 | 50438945 | 50476973 | 79 | 7 | 40350 | 1.1341 | 0.12837 | *RP11-738G5.2* |
| ENSG00000154380 | 1 | 225664537 | 225850844 | 493 | 31 | 52982 | 1.1339 | 0.12842 | *ENAH* |
| ENSG00000269205 | 16 | 2021633 | 2042918 | 126 | 16 | 50008 | 1.1337 | 0.12846 | *AC005606.1* |

| | | | | | | | | | |
|---|---|---|---|---|---|---|---|---|---|
| ENSG00000127554 | 16 | 2024208 | 2047750 | 124 | 14 | 49798 | 1.1335 | 0.1285 | *GFER* |
| ENSG00000091317 | 3 | 32512804 | 32554900 | 110 | 23 | 53158 | 1.1333 | 0.12855 | *CMTM6* |
| ENSG00000120725 | 5 | 138272409 | 138639246 | 565 | 47 | 41966 | 1.133 | 0.1286 | *SIL1* |
| ENSG00000198736 | 16 | 1978211 | 2003327 | 88 | 27 | 34347 | 1.133 | 0.12861 | *MSRB1* |
| ENSG00000258292 | 12 | 96186875 | 96227085 | 152 | 23 | 44015 | 1.1329 | 0.12863 | *RP11-536G4.1* |
| ENSG00000254536 | 10 | 135194338 | 135243999 | 227 | 33 | 41709 | 1.1326 | 0.12869 | *RP11-108K14.8* |
| ENSG00000170291 | 17 | 7144735 | 7173259 | 47 | 7 | 55411 | 1.1325 | 0.12871 | *ELP5* |
| ENSG00000177455 | 16 | 28933260 | 28960667 | 31 | 12 | 36711 | 1.1323 | 0.12876 | *CD19* |
| ENSG00000132688 | 1 | 156628555 | 156657189 | 76 | 14 | 46797 | 1.1322 | 0.12877 | *NES* |
| ENSG00000170279 | 7 | 148277657 | 148322952 | 152 | 39 | 42983 | 1.1317 | 0.12888 | *C7orf33* |
| ENSG00000186960 | 14 | 29231910 | 29292493 | 74 | 29 | 50500 | 1.1316 | 0.1289 | *C14orf23* |
| ENSG00000137076 | 9 | 35686945 | 35742392 | 92 | 20 | 50717 | 1.1314 | 0.12895 | *TLN1* |
| ENSG00000054356 | 2 | 220144345 | 220184370 | 111 | 22 | 38919 | 1.1312 | 0.12898 | *PTPRN* |
| ENSG00000168487 | 8 | 22012249 | 22079839 | 205 | 48 | 48267 | 1.1311 | 0.12901 | *BMP1* |
| ENSG00000145863 | 5 | 160964069 | 161139599 | 449 | 45 | 43886 | 1.1308 | 0.12906 | *GABRA6* |
| ENSG00000117425 | 1 | 45275516 | 45318735 | 104 | 13 | 50366 | 1.1305 | 0.12912 | *PTCH2* |
| ENSG00000068650 | 13 | 113334643 | 113551482 | 696 | 85 | 40783 | 1.1304 | 0.12915 | *ATP11A* |
| ENSG00000254706 | 1 | 162325197 | 162361905 | 137 | 36 | 46338 | 1.1301 | 0.12921 | *RP11-565P22.6* |
| ENSG00000101335 | 20 | 35159887 | 35188228 | 31 | 14 | 46978 | 1.13 | 0.12923 | *MYL9* |
| ENSG00000062370 | 19 | 44820708 | 44881377 | 230 | 27 | 39803 | 1.1297 | 0.12931 | *ZNF112* |
| ENSG00000198570 | 1 | 211639864 | 211676259 | 141 | 28 | 47716 | 1.1294 | 0.12936 | *RD3* |
| ENSG00000189157 | 4 | 77125193 | 77214933 | 436 | 38 | 43446 | 1.1293 | 0.12939 | *FAM47E* |
| ENSG00000125841 | 20 | 317426 | 350304 | 134 | 25 | 41343 | 1.129 | 0.12944 | *NRSN2* |
| ENSG00000024526 | 1 | 68929835 | 68972904 | 141 | 18 | 44518 | 1.1288 | 0.1295 | *DEPDC1* |
| ENSG00000181693 | 11 | 56047529 | 56068566 | 78 | 15 | 55668 | 1.1287 | 0.12952 | *OR8H1* |
| ENSG00000143479 | 1 | 206798881 | 206867764 | 189 | 30 | 42345 | 1.1285 | 0.12955 | *DYRK3* |
| ENSG00000020426 | 14 | 61191460 | 61446671 | 337 | 42 | 40354 | 1.1282 | 0.12961 | *MNAT1* |
| ENSG00000241468 | 7 | 99036098 | 99073954 | 67 | 10 | 31446 | 1.1281 | 0.12964 | *ATP5J2* |
| ENSG00000171189 | 21 | 30899254 | 31322351 | 1356 | 114 | 39992 | 1.1281 | 0.12965 | *GRIK1* |
| ENSG00000122035 | 13 | 27834464 | 27857827 | 51 | 14 | 39135 | 1.1278 | 0.1297 | *RASL11A* |
| ENSG00000163950 | 4 | 1684527 | 1724282 | 147 | 21 | 44734 | 1.1274 | 0.12979 | *SLBP* |
| ENSG00000121067 | 17 | 47666246 | 47765596 | 128 | 33 | 52671 | 1.1273 | 0.1298 | *SPOP* |
| ENSG00000005436 | 2 | 75869126 | 75948115 | 250 | 28 | 43994 | 1.1272 | 0.12982 | *GCFC2* |
| ENSG00000244482 | 19 | 54710737 | 54756649 | 126 | 31 | 38745 | 1.1272 | 0.12982 | *LILRA6* |
| ENSG00000063978 | 4 | 2453947 | 2637047 | 473 | 61 | 47809 | 1.1269 | 0.12989 | *RNF4* |
| ENSG00000182325 | 8 | 145569091 | 145593036 | 39 | 10 | 47423 | 1.1264 | 0.12999 | *FBXL6* |

| | | | | | | | | | |
|---|---|---|---|---|---|---|---|---|---|
| ENSG00000164610 | 7 | 33124409 | 33159013 | 111 | 16 | 52987 | 1.1264 | 0.13001 | *RP9* |
| ENSG00000163931 | 3 | 53248723 | 53300068 | 226 | 23 | 43794 | 1.1259 | 0.13011 | *TKT* |
| ENSG00000164631 | 7 | 6718064 | 6756554 | 149 | 21 | 35124 | 1.1258 | 0.13012 | *ZNF12* |
| ENSG00000115524 | 2 | 198244508 | 198309815 | 88 | 15 | 52087 | 1.1257 | 0.13014 | *SF3B1* |
| ENSG00000124227 | 20 | 56783551 | 56813709 | 69 | 21 | 47126 | 1.1257 | 0.13014 | *ANKRD60* |
| ENSG00000243725 | 1 | 55171495 | 55218330 | 228 | 10 | 29714 | 1.1256 | 0.13016 | *TTC4* |
| ENSG00000096968 | 9 | 4975033 | 5138183 | 468 | 30 | 45747 | 1.1256 | 0.13017 | *JAK2* |
| ENSG00000126583 | 19 | 54372444 | 54420906 | 90 | 28 | 34198 | 1.1256 | 0.13017 | *PRKCG* |
| ENSG00000143443 | 1 | 151010216 | 151034462 | 48 | 14 | 49501 | 1.1255 | 0.13019 | *C1orf56* |
| ENSG00000114383 | 3 | 50347458 | 50375682 | 25 | 12 | 47483 | 1.1254 | 0.13022 | *TUSC2* |
| ENSG00000163349 | 1 | 114461814 | 114530426 | 162 | 29 | 46601 | 1.1251 | 0.13027 | *HIPK1* |
| ENSG00000129625 | 5 | 112202084 | 112268236 | 250 | 26 | 41078 | 1.1251 | 0.13027 | *REEP5* |
| ENSG00000186275 | 1 | 146363546 | 146477744 | 8 | 6 | 42738 | 1.125 | 0.1303 | *NBPF12* |
| ENSG00000128989 | 15 | 52829242 | 52872080 | 119 | 14 | 46019 | 1.1249 | 0.13031 | *ARPP19* |
| ENSG00000164284 | 5 | 148714993 | 148744146 | 55 | 10 | 45557 | 1.1247 | 0.13036 | *GRPEL2* |
| ENSG00000135549 | 6 | 122783076 | 123057518 | 791 | 74 | 45878 | 1.1246 | 0.13039 | *PKIB* |
| ENSG00000084234 | 11 | 129929732 | 130024699 | 319 | 29 | 38499 | 1.1246 | 0.13039 | *APLP2* |
| ENSG00000167118 | 9 | 131123598 | 131163015 | 79 | 17 | 40088 | 1.1245 | 0.1304 | *URM1* |
| ENSG00000151611 | 4 | 146529415 | 146591187 | 157 | 25 | 45513 | 1.1243 | 0.13044 | *MMAA* |
| ENSG00000162892 | 1 | 207060788 | 207087484 | 69 | 10 | 34234 | 1.1241 | 0.13049 | *IL24* |
| ENSG00000169871 | 7 | 100718720 | 100745017 | 117 | 20 | 42478 | 1.1241 | 0.13049 | *TRIM56* |
| ENSG00000107949 | 10 | 127502115 | 127552264 | 85 | 13 | 52949 | 1.1236 | 0.1306 | *BCCIP* |
| ENSG00000123124 | 8 | 87344967 | 87500649 | 488 | 35 | 46541 | 1.1234 | 0.13064 | *WWP1* |
| ENSG00000064218 | 9 | 966964 | 1001731 | 127 | 23 | 49049 | 1.1234 | 0.13064 | *DMRT3* |
| ENSG00000137252 | 6 | 55029050 | 55157418 | 473 | 47 | 49105 | 1.1233 | 0.13065 | *HCRTR2* |
| ENSG00000114670 | 3 | 130735694 | 131079309 | 817 | 62 | 41448 | 1.1233 | 0.13066 | *NEK11* |
| ENSG00000180035 | 16 | 30379427 | 30421429 | 67 | 15 | 41136 | 1.123 | 0.13071 | *ZNF48* |
| ENSG00000255154 | 3 | 58293341 | 58313847 | 96 | 12 | 35063 | 1.1228 | 0.13075 | *RPP14* |
| ENSG00000105393 | 19 | 17368159 | 17402058 | 144 | 19 | 43518 | 1.1228 | 0.13075 | *BABAM1* |
| ENSG00000144290 | 2 | 162270843 | 162851792 | 1371 | 55 | 50203 | 1.1228 | 0.13076 | *SLC4A10* |
| ENSG00000204525 | 6 | 31226526 | 31249907 | 807 | 49 | 43279 | 1.1224 | 0.13085 | *HLA-C* |
| ENSG00000182261 | 11 | 7970971 | 7996973 | 81 | 18 | 51601 | 1.1223 | 0.13086 | *NLRP10* |
| ENSG00000185442 | 15 | 93150673 | 93363114 | 948 | 103 | 37551 | 1.1221 | 0.1309 | *FAM174B* |
| ENSG00000163539 | 3 | 33527737 | 33769848 | 450 | 38 | 52135 | 1.122 | 0.13093 | *CLASP2* |
| ENSG00000136535 | 2 | 162262605 | 162292381 | 55 | 15 | 50768 | 1.1211 | 0.13112 | *TBR1* |
| ENSG00000173320 | 4 | 184764584 | 184954679 | 556 | 110 | 45215 | 1.1211 | 0.13113 | *STOX2* |

| | | | | | | | | | |
|---|---|---|---|---|---|---|---|---|---|
| ENSG00000229974 | 7 | 134822797 | 134842916 | 59 | 5 | 55356 | 1.1211 | 0.13113 | *AC083862.1* |
| ENSG00000101191 | 20 | 61499090 | 61579304 | 307 | 41 | 44657 | 1.121 | 0.13114 | *DIDO1* |
| ENSG00000258289 | 14 | 65371079 | 65421309 | 211 | 20 | 32500 | 1.1209 | 0.13116 | *CHURC1* |
| ENSG00000185808 | 21 | 38421470 | 38455470 | 156 | 20 | 54645 | 1.1208 | 0.13119 | *PIGP* |
| ENSG00000117151 | 1 | 85005289 | 85050163 | 131 | 18 | 36355 | 1.1206 | 0.13122 | *CTBS* |
| ENSG00000105699 | 19 | 35729233 | 35768867 | 131 | 20 | 50604 | 1.1205 | 0.13126 | *LSR* |
| ENSG00000138942 | 22 | 31546168 | 31613005 | 176 | 26 | 41314 | 1.1203 | 0.1313 | *RNF185* |
| ENSG00000196456 | 7 | 150055879 | 150119558 | 214 | 28 | 45027 | 1.12 | 0.13136 | *ZNF775* |
| ENSG00000144218 | 2 | 100152323 | 100769201 | 1513 | 123 | 48744 | 1.1193 | 0.1315 | *AFF3* |
| ENSG00000248919 | 7 | 99007372 | 99073820 | 122 | 19 | 35346 | 1.1191 | 0.13154 | *ATP5J2-PTCD1* |
| ENSG00000164871 | 8 | 7290206 | 7331196 | 2 | 1 | 42757 | 1.1192 | 0.13154 | *SPAG11B* |
| ENSG00000015592 | 8 | 27082840 | 27125937 | 160 | 25 | 44908 | 1.1191 | 0.13155 | *STMN4* |
| ENSG00000166734 | 15 | 44570927 | 44717956 | 235 | 15 | 34379 | 1.119 | 0.13156 | *CASC4* |
| ENSG00000169764 | 2 | 64058074 | 64128696 | 180 | 29 | 42357 | 1.1189 | 0.13159 | *UGP2* |
| ENSG00000243667 | 2 | 68340068 | 68394692 | 115 | 18 | 43395 | 1.1188 | 0.13161 | *WDR92* |
| ENSG00000113924 | 3 | 120337020 | 120411418 | 289 | 50 | 50689 | 1.1183 | 0.13171 | *HGD* |
| ENSG00000112494 | 6 | 167674657 | 167739507 | 410 | 52 | 46438 | 1.118 | 0.13179 | *UNC93A* |
| ENSG00000173456 | 11 | 119195237 | 119218023 | 45 | 13 | 46507 | 1.1178 | 0.13182 | *RNF26* |
| ENSG00000127022 | 5 | 179095629 | 179167926 | 204 | 27 | 46133 | 1.1177 | 0.13186 | *CANX* |
| ENSG00000160683 | 11 | 118744475 | 118778508 | 66 | 25 | 48269 | 1.1176 | 0.13186 | *CXCR5* |
| ENSG00000182463 | 20 | 51578946 | 52121869 | 1623 | 226 | 44438 | 1.1176 | 0.13186 | *TSHZ2* |
| ENSG00000212643 | 5 | 112217313 | 112238791 | 85 | 11 | 38953 | 1.1174 | 0.1319 | *ZRSR1* |
| ENSG00000103591 | 15 | 67483371 | 67557533 | 168 | 15 | 52332 | 1.1173 | 0.13192 | *AAGAB* |
| ENSG00000188199 | 10 | 81452983 | 81484437 | 34 | 7 | 35992 | 1.1171 | 0.13197 | *NUTM2B* |
| ENSG00000171848 | 2 | 10252455 | 10281545 | 58 | 13 | 44527 | 1.1168 | 0.13203 | *RRM2* |
| ENSG00000197653 | 12 | 124237042 | 124430753 | 560 | 73 | 45822 | 1.1167 | 0.13207 | *DNAH10* |
| ENSG00000172466 | 18 | 32902176 | 32935446 | 53 | 14 | 35550 | 1.1165 | 0.1321 | *ZNF24* |
| ENSG00000268864 | 19 | 54092882 | 54116751 | 79 | 19 | 44713 | 1.1164 | 0.13212 | *CTB-167G5.5* |
| ENSG00000106603 | 7 | 43638055 | 43779316 | 470 | 24 | 40590 | 1.1164 | 0.13213 | *COA1* |
| ENSG00000107742 | 10 | 73808793 | 73858790 | 142 | 20 | 35090 | 1.1162 | 0.13218 | *SPOCK2* |
| ENSG00000177548 | 16 | 28905742 | 28957847 | 68 | 17 | 38281 | 1.116 | 0.13221 | *RABEP2* |
| ENSG00000116221 | 1 | 54639714 | 54701137 | 155 | 34 | 49164 | 1.1155 | 0.13232 | *MRPL37* |
| ENSG00000234438 | 15 | 65359154 | 65382276 | 48 | 15 | 53358 | 1.1151 | 0.13241 | *KBTBD13* |
| ENSG00000117090 | 1 | 160567890 | 160627085 | 204 | 42 | 42188 | 1.1148 | 0.13247 | *SLAMF1* |
| ENSG00000164941 | 8 | 95815539 | 95903974 | 314 | 27 | 55254 | 1.1147 | 0.13248 | *INTS8* |
| ENSG00000111863 | 6 | 11702287 | 11817279 | 367 | 60 | 38490 | 1.1141 | 0.13262 | *ADTRP* |

| | | | | | | | | | |
|---|---|---|---|---|---|---|---|---|---|
| ENSG00000167525 | 17 | 27020215 | 27048872 | 32 | 9 | 51612 | 1.1139 | 0.13266 | *PROCA1* |
| ENSG00000152128 | 2 | 135203330 | 135486570 | 768 | 89 | 38523 | 1.1135 | 0.13274 | *TMEM163* |
| ENSG00000134575 | 11 | 47250853 | 47280457 | 65 | 12 | 43935 | 1.1135 | 0.13274 | *ACP2* |
| ENSG00000104870 | 19 | 50000073 | 50039590 | 117 | 18 | 40034 | 1.1135 | 0.13275 | *FCGRT* |
| ENSG00000137675 | 11 | 102552218 | 102586537 | 185 | 26 | 44606 | 1.1134 | 0.13277 | *MMP27* |
| ENSG00000198925 | 2 | 220064494 | 220104439 | 70 | 14 | 41957 | 1.1132 | 0.13281 | *ATG9A* |
| ENSG00000087258 | 16 | 56215302 | 56401356 | 566 | 75 | 46270 | 1.1131 | 0.13283 | *GNAO1* |
| ENSG00000258817 | 11 | 49963943 | 49984971 | 98 | 14 | 42889 | 1.1124 | 0.13299 | *OR4C13* |
| ENSG00000204577 | 19 | 54710147 | 54756602 | 127 | 31 | 38894 | 1.1122 | 0.13303 | *LILRB3* |
| ENSG00000166046 | 12 | 106685707 | 106750793 | 186 | 25 | 48817 | 1.1121 | 0.13305 | *TCP11L2* |
| ENSG00000221946 | 19 | 35624154 | 35655204 | 76 | 24 | 50746 | 1.112 | 0.13307 | *FXYD7* |
| ENSG00000129810 | 3 | 20192085 | 20237784 | 182 | 23 | 42689 | 1.1119 | 0.13309 | *SGOL1* |
| ENSG00000134716 | 1 | 60348980 | 60402462 | 129 | 19 | 44992 | 1.1117 | 0.13313 | *CYP2J2* |
| ENSG00000272647 | 7 | 99146291 | 99215433 | 91 | 19 | 39608 | 1.1114 | 0.1332 | *GS1-259H13.10* |
| ENSG00000167513 | 16 | 88859621 | 88885666 | 149 | 26 | 46472 | 1.1114 | 0.1332 | *CDT1* |
| ENSG00000162975 | 2 | 11042063 | 11064350 | 82 | 24 | 42602 | 1.1113 | 0.13321 | *KCNF1* |
| ENSG00000115548 | 2 | 86657770 | 86729839 | 200 | 27 | 44685 | 1.1112 | 0.13324 | *KDM3A* |
| ENSG00000143878 | 2 | 20636835 | 20659200 | 53 | 18 | 54446 | 1.1112 | 0.13325 | *RHOB* |
| ENSG00000136935 | 9 | 127630646 | 127720771 | 151 | 17 | 54136 | 1.1111 | 0.13326 | *GOLGA1* |
| ENSG00000204486 | 1 | 13506066 | 13536943 | 4 | 1 | 42634 | 1.1111 | 0.13327 | *PRAMEF21* |
| ENSG00000241563 | 1 | 10499776 | 10521959 | 46 | 8 | 30972 | 1.1108 | 0.13332 | *CORT* |
| ENSG00000063438 | 5 | 294291 | 448406 | 598 | 66 | 38478 | 1.1106 | 0.13338 | *AHRR* |
| ENSG00000123609 | 2 | 152116979 | 152156571 | 171 | 15 | 42120 | 1.1104 | 0.13341 | *NMI* |
| ENSG00000122862 | 10 | 70837862 | 70874567 | 121 | 17 | 31622 | 1.1103 | 0.13343 | *SRGN* |
| ENSG00000124802 | 6 | 8063593 | 8112811 | 179 | 25 | 47960 | 1.11 | 0.13351 | *EEF1E1* |
| ENSG00000177535 | 1 | 247604255 | 247625308 | 202 | 27 | 48893 | 1.1099 | 0.13352 | *OR2B11* |
| ENSG00000173221 | 5 | 95077023 | 95168709 | 255 | 48 | 51190 | 1.1093 | 0.13366 | *GLRX* |
| ENSG00000105849 | 7 | 19725085 | 19758710 | 149 | 17 | 37186 | 1.1093 | 0.13366 | *TWISTNB* |
| ENSG00000107593 | 10 | 102037903 | 102100243 | 157 | 32 | 38333 | 1.1093 | 0.13366 | *PKD2L1* |
| ENSG00000197355 | 9 | 139961953 | 139988991 | 60 | 16 | 44585 | 1.1087 | 0.13377 | *UAP1L1* |
| ENSG00000109572 | 4 | 170523784 | 170654824 | 362 | 34 | 47185 | 1.1087 | 0.13378 | *CLCN3* |
| ENSG00000129559 | 14 | 24676058 | 24711660 | 49 | 14 | 40954 | 1.1081 | 0.1339 | *NEDD8* |
| ENSG00000198208 | 14 | 75360657 | 75400099 | 106 | 16 | 45410 | 1.108 | 0.13394 | *RPS6KL1* |
| ENSG00000124193 | 20 | 42076568 | 42102245 | 92 | 24 | 40920 | 1.1074 | 0.13405 | *SRSF6* |
| ENSG00000106246 | 7 | 99004362 | 99073787 | 125 | 19 | 35517 | 1.1072 | 0.1341 | *PTCD1* |
| ENSG00000230610 | 1 | 38282232 | 38303886 | 53 | 12 | 42250 | 1.1068 | 0.13419 | *AL929472.1* |

| | | | | | | | | | |
|---|---|---|---|---|---|---|---|---|---|
| ENSG00000255526 | 14 | 24673161 | 24711558 | 58 | 14 | 41967 | 1.1062 | 0.13432 | *NEDD8-MDP1* |
| ENSG00000273088 | 1 | 155131885 | 155169748 | 63 | 12 | 47497 | 1.1059 | 0.13439 | *RP11-201K10.3* |
| ENSG00000131738 | 17 | 39509746 | 39536052 | 55 | 14 | 41882 | 1.1056 | 0.13446 | *KRT33B* |
| ENSG00000104388 | 8 | 61419416 | 61546186 | 328 | 21 | 47046 | 1.1054 | 0.1345 | *RAB2A* |
| ENSG00000181626 | 18 | 12083852 | 12139748 | 205 | 14 | 38461 | 1.1052 | 0.13453 | *ANKRD62* |
| ENSG00000269155 | 6 | 168186210 | 168208927 | 121 | 17 | 40506 | 1.1052 | 0.13454 | *AL009178.1* |
| ENSG00000107959 | 10 | 3169920 | 3225003 | 352 | 30 | 48997 | 1.1047 | 0.13465 | *PITRM1* |
| ENSG00000119655 | 14 | 74932895 | 74970880 | 105 | 9 | 54110 | 1.1043 | 0.13473 | *NPC2* |
| ENSG00000172399 | 4 | 120046939 | 120118944 | 273 | 29 | 44289 | 1.1041 | 0.13477 | *MYOZ2* |
| ENSG00000137752 | 11 | 104886170 | 104982158 | 350 | 30 | 46615 | 1.1041 | 0.13477 | *CASP1* |
| ENSG00000263264 | 19 | 7403848 | 7458271 | 155 | 27 | 40332 | 1.1038 | 0.13484 | *CTB-133G6.1* |
| ENSG00000144445 | 2 | 210876147 | 211046107 | 155 | 30 | 46985 | 1.1035 | 0.1349 | *KANSL1L* |
| ENSG00000197147 | 1 | 89980395 | 90073423 | 263 | 25 | 39510 | 1.1032 | 0.13497 | *LRRC8B* |
| ENSG00000213920 | 14 | 24673143 | 24695276 | 41 | 10 | 42111 | 1.1031 | 0.13499 | *MDP1* |
| ENSG00000154415 | 7 | 113506832 | 113725975 | 438 | 23 | 38894 | 1.103 | 0.135 | *PPP1R3A* |
| ENSG00000116691 | 1 | 12069523 | 12102102 | 78 | 18 | 44335 | 1.103 | 0.13502 | *MIIP* |
| ENSG00000182272 | 11 | 359796 | 392116 | 161 | 29 | 38703 | 1.1028 | 0.13505 | *B4GALNT4* |
| ENSG00000147654 | 8 | 110541940 | 110588225 | 120 | 24 | 43267 | 1.1026 | 0.1351 | *EBAG9* |
| ENSG00000186118 | 1 | 47124527 | 47149266 | 50 | 7 | 37346 | 1.1026 | 0.13511 | *TEX38* |
| ENSG00000162843 | 1 | 241805580 | 241975435 | 702 | 84 | 50455 | 1.1025 | 0.13512 | *WDR64* |
| ENSG00000128655 | 2 | 178477980 | 178983066 | 2085 | 94 | 44881 | 1.1022 | 0.13519 | *PDE11A* |
| ENSG00000180875 | 1 | 240642873 | 240785449 | 446 | 105 | 45100 | 1.1019 | 0.13525 | *GREM2* |
| ENSG00000157470 | 15 | 59654892 | 59825748 | 569 | 77 | 44521 | 1.1019 | 0.13525 | *FAM81A* |
| ENSG00000135248 | 7 | 128339115 | 128381797 | 168 | 32 | 51572 | 1.1019 | 0.13526 | *FAM71F1* |
| ENSG00000165821 | 14 | 21979232 | 22015350 | 97 | 23 | 46060 | 1.1019 | 0.13526 | *SALL2* |
| ENSG00000167700 | 8 | 145724457 | 145746596 | 32 | 7 | 40840 | 1.101 | 0.13544 | *MFSD3* |
| ENSG00000205882 | 8 | 11840692 | 11863821 | 77 | 15 | 43370 | 1.1009 | 0.13547 | *DEFB134* |
| ENSG00000150990 | 12 | 125421371 | 125483668 | 363 | 47 | 44081 | 1.1008 | 0.1355 | *DHX37* |
| ENSG00000114554 | 3 | 126697437 | 126766235 | 258 | 32 | 43588 | 1.1006 | 0.13553 | *PLXNA1* |
| ENSG00000181649 | 11 | 2939503 | 2960685 | 55 | 17 | 39214 | 1.1005 | 0.13556 | *PHLDA2* |
| ENSG00000151150 | 10 | 61776056 | 62503248 | 2563 | 184 | 48915 | 1.1002 | 0.13562 | *ANK3* |
| ENSG00000145920 | 5 | 175213313 | 175321023 | 328 | 54 | 47079 | 1.1 | 0.13566 | *CPLX2* |
| ENSG00000138594 | 15 | 52111825 | 52249492 | 363 | 18 | 52172 | 1.0998 | 0.1357 | *TMOD3* |
| ENSG00000111641 | 12 | 6656029 | 6687857 | 52 | 14 | 40662 | 1.0998 | 0.13571 | *NOP2* |
| ENSG00000143603 | 1 | 154659931 | 154852756 | 570 | 125 | 46408 | 1.0997 | 0.13573 | *KCNN3* |
| ENSG00000171467 | 6 | 43264872 | 43347216 | 142 | 20 | 43671 | 1.0989 | 0.13591 | *ZNF318* |

| | | | | | | | | | |
|---|---|---|---|---|---|---|---|---|---|
| ENSG00000174990 | 16 | 87911625 | 87980135 | 371 | 56 | 40591 | 1.0985 | 0.13598 | *CA5A* |
| ENSG00000213402 | 11 | 67192981 | 67215538 | 39 | 8 | 36227 | 1.0985 | 0.13599 | *PTPRCAP* |
| ENSG00000149658 | 20 | 61816781 | 61857586 | 128 | 18 | 50856 | 1.0983 | 0.13605 | *YTHDF1* |
| ENSG00000095464 | 10 | 95362345 | 95435767 | 258 | 47 | 44427 | 1.0979 | 0.13612 | *PDE6C* |
| ENSG00000257315 | 1 | 203755437 | 203779686 | 80 | 8 | 35277 | 1.0974 | 0.13623 | *ZBED6* |
| ENSG00000012822 | 12 | 54094903 | 54131529 | 103 | 23 | 46929 | 1.0972 | 0.13627 | *CALCOCO1* |
| ENSG00000146197 | 6 | 35172190 | 35230856 | 114 | 18 | 34660 | 1.097 | 0.13632 | *SCUBE3* |
| ENSG00000217455 | 7 | 3169868 | 3224287 | 351 | 49 | 48822 | 1.097 | 0.13632 | *AC091801.1* |
| ENSG00000184451 | 17 | 40820907 | 40845935 | 37 | 10 | 51436 | 1.0968 | 0.13636 | *CCR10* |
| ENSG00000160957 | 8 | 145726667 | 145753229 | 39 | 8 | 40331 | 1.0964 | 0.13644 | *RECQL4* |
| ENSG00000108854 | 17 | 62528413 | 62668186 | 285 | 24 | 39324 | 1.0962 | 0.1365 | *SMURF2* |
| ENSG00000104435 | 8 | 80513049 | 80588410 | 259 | 38 | 46512 | 1.0961 | 0.13652 | *STMN2* |
| ENSG00000096088 | 6 | 41694449 | 41731847 | 111 | 35 | 49045 | 1.0959 | 0.13656 | *PGC* |
| ENSG00000188986 | 9 | 140139625 | 140177998 | 108 | 11 | 37352 | 1.0959 | 0.13656 | *NELFB* |
| ENSG00000187266 | 19 | 11478236 | 11505018 | 42 | 10 | 43669 | 1.0959 | 0.13657 | *EPOR* |
| ENSG00000272325 | 6 | 34237456 | 34370451 | 317 | 18 | 47152 | 1.0956 | 0.13663 | *NUDT3* |
| ENSG00000196104 | 4 | 167644535 | 168165947 | 1954 | 153 | 48737 | 1.0955 | 0.13666 | *SPOCK3* |
| ENSG00000221890 | 22 | 39204457 | 39249987 | 159 | 24 | 48182 | 1.0955 | 0.13666 | *NPTXR* |
| ENSG00000262526 | 17 | 7133746 | 7157954 | 29 | 6 | 55644 | 1.0953 | 0.13669 | *CTD-2545G14.7* |
| ENSG00000126432 | 11 | 64075560 | 64099283 | 35 | 11 | 42164 | 1.0953 | 0.1367 | *PRDX5* |
| ENSG00000183654 | 5 | 16057248 | 16190871 | 268 | 45 | 42964 | 1.0952 | 0.13672 | *Mar-11* |
| ENSG00000110080 | 11 | 126215535 | 126320239 | 301 | 59 | 45865 | 1.0948 | 0.1368 | *ST3GAL4* |
| ENSG00000154122 | 5 | 14694910 | 14881887 | 530 | 78 | 46521 | 1.0947 | 0.13682 | *ANKH* |
| ENSG00000268155 | 9 | 5067163 | 5094580 | 88 | 11 | 50321 | 1.0947 | 0.13682 | *AL161450.1* |
| ENSG00000125888 | 20 | 17664317 | 17726516 | 224 | 35 | 51253 | 1.0947 | 0.13682 | *BANF2* |
| ENSG00000248235 | 8 | 22436787 | 22469597 | 86 | 14 | 54469 | 1.0946 | 0.13684 | *AC037459.4* |
| ENSG00000187758 | 4 | 100187524 | 100222185 | 107 | 18 | 38286 | 1.0945 | 0.13687 | *ADH1A* |
| ENSG00000101144 | 20 | 55733804 | 55851685 | 415 | 50 | 45178 | 1.0945 | 0.13688 | *BMP7* |
| ENSG00000186130 | 9 | 125660335 | 125685609 | 55 | 18 | 42185 | 1.0944 | 0.13689 | *ZBTB6* |
| ENSG00000156398 | 10 | 104464295 | 104513249 | 116 | 19 | 43315 | 1.094 | 0.13699 | *SFXN2* |
| ENSG00000147421 | 8 | 28737911 | 28932281 | 270 | 32 | 43676 | 1.0938 | 0.13702 | *HMBOX1* |
| ENSG00000145002 | 8 | 12272913 | 12303915 | 4 | 2 | 42704 | 1.0938 | 0.13703 | *FAM86B2* |
| ENSG00000112837 | 6 | 85387069 | 85484237 | 279 | 31 | 45400 | 1.0934 | 0.13711 | *TBX18* |
| ENSG00000171448 | 9 | 125667845 | 125703779 | 71 | 20 | 43573 | 1.0933 | 0.13713 | *ZBTB26* |
| ENSG00000136811 | 9 | 131207465 | 131273571 | 104 | 11 | 33790 | 1.0932 | 0.13715 | *ODF2* |
| ENSG00000013374 | 7 | 151028785 | 151085535 | 221 | 45 | 45355 | 1.0932 | 0.13716 | *NUB1* |

| | | | | | | | | | |
|---|---|---|---|---|---|---|---|---|---|
| ENSG00000236882 | 5 | 95177936 | 95205837 | 94 | 22 | 43555 | 1.0931 | 0.13717 | *C5orf27* |
| ENSG00000124795 | 6 | 18214099 | 18275054 | 144 | 22 | 35089 | 1.0931 | 0.13717 | *DEK* |
| ENSG00000123983 | 2 | 223715652 | 223819357 | 318 | 31 | 45939 | 1.093 | 0.1372 | *ACSL3* |
| ENSG00000106367 | 7 | 100787678 | 100814877 | 90 | 17 | 37421 | 1.0927 | 0.13727 | *AP1S1* |
| ENSG00000171130 | 7 | 149560057 | 149587784 | 132 | 13 | 42561 | 1.0926 | 0.13728 | *ATP6V0E2* |
| ENSG00000049883 | 5 | 71606194 | 71666052 | 67 | 13 | 36485 | 1.092 | 0.13741 | *PTCD2* |
| ENSG00000270466 | 16 | 30379755 | 30403863 | 41 | 9 | 41703 | 1.0919 | 0.13743 | *Sep-01* |
| ENSG00000167986 | 11 | 61056923 | 61120068 | 34 | 18 | 41611 | 1.0914 | 0.13754 | *DDB1* |
| ENSG00000169393 | 19 | 48487908 | 48538410 | 322 | 36 | 51143 | 1.0913 | 0.13756 | *ELSPBP1* |
| ENSG00000197070 | 9 | 140490106 | 140519812 | 68 | 11 | 47705 | 1.091 | 0.13764 | *ARRDC1* |
| ENSG00000105372 | 19 | 42353988 | 42386994 | 54 | 6 | 42939 | 1.0909 | 0.13765 | *RPS19* |
| ENSG00000126351 | 17 | 38204543 | 38260120 | 114 | 27 | 44968 | 1.0909 | 0.13766 | *THRA* |
| ENSG00000175550 | 11 | 65676728 | 65699032 | 43 | 12 | 39535 | 1.0907 | 0.13769 | *DRAP1* |
| ENSG00000174943 | 16 | 29906333 | 29948356 | 69 | 11 | 41991 | 1.0903 | 0.1378 | *KCTD13* |
| ENSG00000180209 | 16 | 30372255 | 30399312 | 42 | 9 | 41098 | 1.0902 | 0.13781 | *MYLPF* |
| ENSG00000151715 | 11 | 129675714 | 129739898 | 221 | 26 | 49456 | 1.0901 | 0.13782 | *TMEM45B* |
| ENSG00000165794 | 14 | 21457414 | 21480030 | 63 | 22 | 44730 | 1.0902 | 0.13782 | *SLC39A2* |
| ENSG00000175505 | 11 | 67121639 | 67151648 | 36 | 10 | 40032 | 1.09 | 0.13786 | *CLCF1* |
| ENSG00000172724 | 9 | 34679564 | 34701274 | 64 | 15 | 52874 | 1.0899 | 0.13789 | *CCL19* |
| ENSG00000216937 | 10 | 32725068 | 32873492 | 631 | 51 | 42733 | 1.0898 | 0.13791 | *CCDC7* |
| ENSG00000148730 | 10 | 72154135 | 72198374 | 94 | 15 | 44583 | 1.0896 | 0.13795 | *EIF4EBP2* |
| ENSG00000158941 | 8 | 22452145 | 22489027 | 131 | 13 | 46254 | 1.0893 | 0.138 | *CCAR2* |
| ENSG00000169570 | 5 | 118163017 | 118334240 | 391 | 49 | 43548 | 1.0891 | 0.13805 | *DTWD2* |
| ENSG00000130222 | 9 | 92209928 | 92231470 | 76 | 14 | 47310 | 1.0891 | 0.13805 | *GADD45G* |
| ENSG00000167588 | 12 | 50487602 | 50515102 | 52 | 10 | 38642 | 1.089 | 0.13807 | *GPD1* |
| ENSG00000183508 | 1 | 118138556 | 118180994 | 156 | 17 | 47364 | 1.0888 | 0.13812 | *FAM46C* |
| ENSG00000204252 | 6 | 32961955 | 32987389 | 236 | 39 | 40988 | 1.0888 | 0.13812 | *HLA-DOA* |
| ENSG00000159556 | 15 | 76619065 | 76644817 | 65 | 17 | 39177 | 1.0883 | 0.13823 | *ISL2* |
| ENSG00000147697 | 8 | 130750442 | 130809134 | 218 | 30 | 40495 | 1.0883 | 0.13824 | *GSDMC* |
| ENSG00000141076 | 16 | 69155194 | 69275033 | 277 | 39 | 54745 | 1.0882 | 0.13826 | *CIRH1A* |
| ENSG00000146021 | 5 | 136943189 | 137081779 | 315 | 35 | 49837 | 1.0881 | 0.13828 | *KLHL3* |
| ENSG00000120162 | 9 | 27315207 | 27539779 | 829 | 117 | 44613 | 1.0875 | 0.13842 | *MOB3B* |
| ENSG00000111321 | 12 | 6474211 | 6510733 | 103 | 28 | 39601 | 1.087 | 0.13852 | *LTBR* |
| ENSG00000152242 | 18 | 43743519 | 43856949 | 222 | 18 | 44300 | 1.087 | 0.13852 | *C18orf25* |
| ENSG00000214300 | 7 | 99895325 | 99929819 | 38 | 9 | 52447 | 1.0869 | 0.13853 | *SPDYE3* |
| ENSG00000163960 | 3 | 196064533 | 196169345 | 249 | 32 | 49367 | 1.0869 | 0.13854 | *UBXN7* |

| | | | | | | | | | |
|---|---|---|---|---|---|---|---|---|---|
| ENSG00000134028 | 8 | 24231798 | 24273526 | 61 | 13 | 46693 | 1.0868 | 0.13855 | *ADAMDEC1* |
| ENSG00000181761 | 11 | 55879849 | 55900787 | 163 | 7 | 56184 | 1.0866 | 0.1386 | *OR8H3* |
| ENSG00000139343 | 12 | 96242706 | 96307606 | 275 | 35 | 48585 | 1.0866 | 0.13861 | *SNRPF* |
| ENSG00000124733 | 6 | 42969832 | 42991706 | 61 | 13 | 37171 | 1.0865 | 0.13862 | *MEA1* |
| ENSG00000056487 | 22 | 45267042 | 45415880 | 595 | 85 | 41557 | 1.0865 | 0.13862 | *PHF21B* |
| ENSG00000204767 | 5 | 169281268 | 169417744 | 393 | 75 | 41147 | 1.0865 | 0.13863 | *FAM196B* |
| ENSG00000244462 | 20 | 34226847 | 34262878 | 99 | 12 | 35147 | 1.0863 | 0.13867 | *RBM12* |
| ENSG00000084731 | 2 | 26139471 | 26215618 | 207 | 36 | 44798 | 1.0862 | 0.13869 | *KIF3C* |
| ENSG00000236624 | 1 | 45949538 | 45975751 | 49 | 7 | 55250 | 1.0862 | 0.1387 | *CCDC163P* |
| ENSG00000117400 | 1 | 43793478 | 43828443 | 46 | 10 | 39219 | 1.086 | 0.13874 | *MPL* |
| ENSG00000119777 | 2 | 27245778 | 27274563 | 32 | 9 | 41292 | 1.086 | 0.13874 | *TMEM214* |
| ENSG00000204574 | 6 | 30529153 | 30574956 | 110 | 13 | 33912 | 1.086 | 0.13874 | *ABCF1* |
| ENSG00000155428 | 7 | 72420016 | 72449997 | 5 | 3 | 36248 | 1.0854 | 0.13888 | *TRIM74* |
| ENSG00000112186 | 6 | 17383447 | 17568023 | 499 | 76 | 45390 | 1.0853 | 0.13889 | *CAP2* |
| ENSG00000258740 | 14 | 75695319 | 75745986 | 130 | 30 | 38410 | 1.0853 | 0.13889 | *RP11-293M10.1* |
| ENSG00000108861 | 17 | 41833489 | 41866356 | 99 | 13 | 50586 | 1.0853 | 0.1389 | *DUSP3* |
| ENSG00000160862 | 7 | 99554343 | 99583780 | 67 | 12 | 49017 | 1.0851 | 0.13893 | *AZGP1* |
| ENSG00000171903 | 19 | 16013177 | 16055677 | 211 | 19 | 47959 | 1.0849 | 0.13899 | *CYP4F11* |
| ENSG00000166130 | 12 | 98997183 | 99048891 | 149 | 24 | 52889 | 1.0848 | 0.139 | *IKBIP* |
| ENSG00000100453 | 14 | 25090160 | 25113473 | 79 | 16 | 55267 | 1.0846 | 0.13904 | *GZMB* |
| ENSG00000105701 | 19 | 18632561 | 18664887 | 61 | 21 | 38045 | 1.0844 | 0.1391 | *FKBP8* |
| ENSG00000116337 | 1 | 110148726 | 110184673 | 84 | 23 | 41799 | 1.0842 | 0.13914 | *AMPD2* |
| ENSG00000177946 | 16 | 90026206 | 90048942 | 111 | 22 | 32678 | 1.084 | 0.13917 | *CENPBD1* |
| ENSG00000103550 | 16 | 19704902 | 19739557 | 76 | 9 | 41130 | 1.0838 | 0.13922 | *KNOP1* |
| ENSG00000114446 | 3 | 107869659 | 107951417 | 393 | 9 | 50729 | 1.0838 | 0.13923 | *IFT57* |
| ENSG00000198715 | 1 | 156249880 | 156275463 | 58 | 8 | 54597 | 1.0835 | 0.13929 | *C1orf85* |
| ENSG00000144231 | 2 | 128593840 | 128625731 | 102 | 16 | 42376 | 1.0831 | 0.13939 | *POLR2D* |
| ENSG00000172818 | 11 | 65544493 | 65574690 | 87 | 16 | 34258 | 1.0828 | 0.13944 | *OVOL1* |
| ENSG00000272781 | 14 | 47301134 | 48153999 | 2924 | 175 | 46716 | 1.0829 | 0.13944 | *MDGA2* |
| ENSG00000189292 | 2 | 269558 | 298851 | 62 | 11 | 50657 | 1.0824 | 0.13954 | *FAM150B* |
| ENSG00000127540 | 19 | 1587171 | 1615480 | 42 | 12 | 56013 | 1.0822 | 0.13958 | *UQCR11* |
| ENSG00000268791 | 4 | 6683782 | 6704189 | 74 | 21 | 36835 | 1.0818 | 0.13967 | *AC093323.1* |
| ENSG00000075151 | 1 | 21122963 | 21513377 | 941 | 53 | 49813 | 1.0818 | 0.13968 | *EIF4G3* |
| ENSG00000184486 | 6 | 99272580 | 99296660 | 62 | 12 | 54081 | 1.0816 | 0.13971 | *POU3F2* |
| ENSG00000162897 | 1 | 207121310 | 207153970 | 73 | 12 | 45145 | 1.0816 | 0.13972 | *FCAMR* |
| ENSG00000103248 | 16 | 86553782 | 86598841 | 180 | 31 | 50225 | 1.0816 | 0.13973 | *MTHFSD* |

| | | | | | | | | | |
|---|---|---|---|---|---|---|---|---|---|
| ENSG00000123154 | 19 | 12767614 | 12796646 | 47 | 17 | 41265 | 1.0815 | 0.13974 | *WDR83* |
| ENSG00000100625 | 14 | 61166246 | 61201066 | 42 | 8 | 47741 | 1.0813 | 0.13978 | *SIX4* |
| ENSG00000203485 | 14 | 105145943 | 105195942 | 210 | 38 | 46559 | 1.0808 | 0.13988 | *INF2* |
| ENSG00000086288 | 7 | 37878199 | 37950003 | 419 | 50 | 44135 | 1.0805 | 0.13995 | *NME8* |
| ENSG00000160703 | 11 | 119027277 | 119064725 | 49 | 13 | 35527 | 1.0805 | 0.13996 | *NLRX1* |
| ENSG00000105383 | 19 | 51718320 | 51757115 | 94 | 19 | 35068 | 1.0801 | 0.14006 | *CD33* |
| ENSG00000118690 | 6 | 109159619 | 109305186 | 432 | 50 | 45793 | 1.0799 | 0.1401 | *ARMC2* |
| ENSG00000070019 | 12 | 14755576 | 14859519 | 178 | 29 | 42853 | 1.0798 | 0.14012 | *GUCY2C* |
| ENSG00000260259 | 16 | 89739028 | 89762977 | 114 | 16 | 41815 | 1.0792 | 0.14024 | *RP11-368I7.4* |
| ENSG00000173679 | 9 | 125413845 | 125434927 | 45 | 16 | 43108 | 1.0792 | 0.14025 | *OR1L1* |
| ENSG00000170074 | 5 | 177124982 | 177220399 | 98 | 16 | 39328 | 1.0792 | 0.14026 | *FAM153A* |
| ENSG00000178665 | 7 | 55945169 | 56019918 | 241 | 31 | 52934 | 1.0786 | 0.14039 | *ZNF713* |
| ENSG00000248835 | 1 | 54625333 | 54647997 | 62 | 18 | 50759 | 1.0784 | 0.14043 | *AL357673.1* |
| ENSG00000272897 | 20 | 34210262 | 34272466 | 185 | 16 | 34674 | 1.0775 | 0.14062 | *RP1-309K20.6* |
| ENSG00000116260 | 1 | 180113969 | 180183165 | 289 | 38 | 39935 | 1.0773 | 0.14067 | *QSOX1* |
| ENSG00000124134 | 20 | 43710951 | 43739753 | 75 | 18 | 41691 | 1.0773 | 0.14067 | *KCNS1* |
| ENSG00000134516 | 5 | 169054251 | 169520386 | 1627 | 179 | 40630 | 1.0773 | 0.14068 | *DOCK2* |
| ENSG00000145908 | 5 | 150263954 | 150294545 | 108 | 10 | 32150 | 1.0772 | 0.1407 | *ZNF300* |
| ENSG00000165953 | 14 | 94943611 | 94994181 | 201 | 43 | 47415 | 1.0768 | 0.14079 | *SERPINA12* |
| ENSG00000081307 | 3 | 132363290 | 132406941 | 59 | 14 | 37629 | 1.0767 | 0.14081 | *UBA5* |
| ENSG00000167419 | 17 | 56285909 | 56355879 | 125 | 27 | 48259 | 1.0763 | 0.14089 | *LPO* |
| ENSG00000268781 | 4 | 3501523 | 3521678 | 151 | 20 | 54720 | 1.076 | 0.14097 | *AL590235.1* |
| ENSG00000172236 | 16 | 1280697 | 1302555 | 89 | 17 | 45968 | 1.0758 | 0.14101 | *TPSAB1* |
| ENSG00000163472 | 1 | 156242726 | 156272976 | 73 | 8 | 54609 | 1.0757 | 0.14103 | *TMEM79* |
| ENSG00000132002 | 19 | 14615582 | 14650582 | 113 | 19 | 47130 | 1.0756 | 0.14104 | *DNAJB1* |
| ENSG00000134070 | 3 | 10196549 | 10295427 | 315 | 49 | 36132 | 1.0756 | 0.14105 | *IRAK2* |
| ENSG00000134668 | 1 | 32246023 | 32291652 | 93 | 19 | 45063 | 1.0755 | 0.14107 | *SPOCD1* |
| ENSG00000138385 | 2 | 170638443 | 170678574 | 174 | 24 | 48670 | 1.0755 | 0.14107 | *SSB* |
| ENSG00000182087 | 19 | 999647 | 1031117 | 275 | 45 | 50772 | 1.0753 | 0.14112 | *TMEM259* |
| ENSG00000151615 | 4 | 147550045 | 147573626 | 73 | 10 | 32564 | 1.0748 | 0.14123 | *POU4F2* |
| ENSG00000176230 | 14 | 20575566 | 20596597 | 65 | 14 | 53025 | 1.0747 | 0.14124 | *OR4K17* |
| ENSG00000203797 | 6 | 110702974 | 110746765 | 195 | 36 | 44639 | 1.0747 | 0.14125 | *DDO* |
| ENSG00000135436 | 12 | 49966668 | 50009422 | 73 | 13 | 39273 | 1.0744 | 0.14131 | *FAM186B* |
| ENSG00000204179 | 10 | 46539955 | 46651045 | 2 | 1 | 42531 | 1.0742 | 0.14136 | *PTPN20A* |
| ENSG00000139915 | 14 | 47298826 | 48154157 | 2929 | 174 | 46719 | 1.0742 | 0.14137 | *MDGA2* |
| ENSG00000179954 | 19 | 55989771 | 56040465 | 259 | 45 | 44952 | 1.0739 | 0.14143 | *SSC5D* |

| | | | | | | | | | |
|---|---|---|---|---|---|---|---|---|---|
| ENSG00000170049 | 17 | 7815177 | 7843121 | 74 | 15 | 46024 | 1.0738 | 0.14145 | *KCNAB3* |
| ENSG00000164543 | 7 | 43612357 | 43676385 | 215 | 17 | 41712 | 1.0736 | 0.14149 | *STK17A* |
| ENSG00000204956 | 5 | 140700252 | 140902546 | 558 | 65 | 43995 | 1.0735 | 0.14152 | *PCDHGA1* |
| ENSG00000146963 | 7 | 139015105 | 139118198 | 371 | 13 | 30301 | 1.0733 | 0.14157 | *C7orf55-LUC7L2* |
| ENSG00000049656 | 5 | 1307859 | 1355214 | 155 | 21 | 40299 | 1.073 | 0.14164 | *CLPTM1L* |
| ENSG00000005187 | 16 | 20611565 | 20818903 | 573 | 43 | 34845 | 1.073 | 0.14164 | *ACSM3* |
| ENSG00000011009 | 1 | 24107460 | 24132029 | 20 | 8 | 49508 | 1.0729 | 0.14166 | *LYPLA2* |
| ENSG00000139351 | 12 | 102112426 | 102143250 | 68 | 17 | 46012 | 1.0729 | 0.14166 | *SYCP3* |
| ENSG00000214160 | 3 | 183950089 | 183977336 | 55 | 12 | 53168 | 1.0727 | 0.14171 | *ALG3* |
| ENSG00000170088 | 4 | 165985574 | 166139701 | 601 | 49 | 32181 | 1.0727 | 0.14171 | *TMEM192* |
| ENSG00000140511 | 15 | 89410519 | 89448857 | 158 | 25 | 48072 | 1.0726 | 0.14172 | *HAPLN3* |
| ENSG00000152818 | 6 | 144596837 | 145184170 | 1290 | 127 | 40971 | 1.0726 | 0.14174 | *UTRN* |
| ENSG00000262302 | 17 | 7140148 | 7175408 | 55 | 9 | 55688 | 1.0725 | 0.14176 | *RP1-4G17.5* |
| ENSG00000214078 | 20 | 34203953 | 34262878 | 162 | 15 | 34538 | 1.0723 | 0.14179 | *CPNE1* |
| ENSG00000105989 | 7 | 116906685 | 116973343 | 171 | 35 | 43677 | 1.0722 | 0.14182 | *WNT2* |
| ENSG00000005108 | 7 | 11399984 | 11881824 | 2067 | 186 | 47450 | 1.072 | 0.14186 | *THSD7A* |
| ENSG00000169957 | 16 | 30525325 | 30548142 | 19 | 10 | 39885 | 1.072 | 0.14186 | *ZNF768* |
| ENSG00000196549 | 3 | 154731913 | 154911497 | 452 | 69 | 42633 | 1.0717 | 0.14193 | *MME* |
| ENSG00000189030 | 1 | 156258415 | 156279428 | 49 | 8 | 55577 | 1.0714 | 0.14198 | *VHLL* |
| ENSG00000164056 | 4 | 124307950 | 124334910 | 83 | 22 | 42151 | 1.0709 | 0.1421 | *SPRY1* |
| ENSG00000100429 | 22 | 50673612 | 50699834 | 122 | 18 | 37469 | 1.0704 | 0.14222 | *HDAC10* |
| ENSG00000100239 | 22 | 50771733 | 50893514 | 469 | 50 | 43540 | 1.0698 | 0.14234 | *PPP6R2* |
| ENSG00000172538 | 10 | 50329199 | 50352053 | 140 | 19 | 43112 | 1.0698 | 0.14236 | *FAM170B* |
| ENSG00000116120 | 2 | 223425255 | 223531056 | 305 | 35 | 43483 | 1.0696 | 0.14241 | *FARSB* |
| ENSG00000183269 | 11 | 5867904 | 5888964 | 139 | 10 | 48208 | 1.0695 | 0.14242 | *OR52E8* |
| ENSG00000188026 | 12 | 123945925 | 124028265 | 254 | 35 | 44399 | 1.0694 | 0.14245 | *RILPL1* |
| ENSG00000180113 | 6 | 46645612 | 46682056 | 43 | 14 | 43526 | 1.0692 | 0.14248 | *TDRD6* |
| ENSG00000197442 | 6 | 136868185 | 137123656 | 522 | 67 | 41340 | 1.0693 | 0.14248 | *MAP3K5* |
| ENSG00000089818 | 12 | 8224807 | 8260367 | 60 | 18 | 36667 | 1.0692 | 0.14248 | *NECAP1* |
| ENSG00000196151 | 2 | 160082304 | 160153310 | 332 | 27 | 25314 | 1.069 | 0.14253 | *WDSUB1* |
| ENSG00000113761 | 5 | 176439697 | 176518190 | 240 | 26 | 41298 | 1.069 | 0.14254 | *ZNF346* |
| ENSG00000221990 | 5 | 431645 | 453258 | 110 | 14 | 29903 | 1.0686 | 0.14263 | *C5orf55* |
| ENSG00000204991 | 16 | 89874587 | 89947727 | 292 | 31 | 42833 | 1.0686 | 0.14263 | *SPIRE2* |
| ENSG00000140030 | 14 | 88461468 | 88488419 | 97 | 15 | 48572 | 1.0685 | 0.14264 | *GPR65* |
| ENSG00000065518 | 3 | 120305156 | 120331347 | 124 | 16 | 48661 | 1.0682 | 0.14271 | *NDUFB4* |
| ENSG00000171989 | 15 | 59489042 | 59510705 | 109 | 16 | 38110 | 1.068 | 0.14276 | *LDHAL6B* |

| | | | | | | | | | |
|---|---|---|---|---|---|---|---|---|---|
| ENSG00000204290 | 6 | 32351740 | 32384905 | 516 | 29 | 48773 | 1.0675 | 0.14288 | *BTNL2* |
| ENSG00000140526 | 15 | 89620690 | 89755591 | 341 | 49 | 50498 | 1.0675 | 0.14288 | *ABHD2* |
| ENSG00000179862 | 1 | 41316729 | 41338018 | 95 | 25 | 42230 | 1.0674 | 0.14289 | *CITED4* |
| ENSG00000034713 | 16 | 75590249 | 75621779 | 50 | 9 | 35563 | 1.0674 | 0.14289 | *GABARAPL2* |
| ENSG00000180043 | 19 | 55856276 | 55884628 | 85 | 22 | 43473 | 1.0674 | 0.14289 | *FAM71E2* |
| ENSG00000129472 | 14 | 21917179 | 21955132 | 113 | 16 | 40296 | 1.0671 | 0.14296 | *RAB2B* |
| ENSG00000131899 | 17 | 18118901 | 18158189 | 93 | 14 | 51478 | 1.0671 | 0.14296 | *LLGL1* |
| ENSG00000143761 | 1 | 228260361 | 228296912 | 51 | 15 | 50328 | 1.0667 | 0.14305 | *ARF1* |
| ENSG00000151502 | 11 | 134084539 | 134127686 | 89 | 24 | 48832 | 1.0663 | 0.14313 | *VPS26B* |
| ENSG00000133216 | 1 | 23027332 | 23251818 | 469 | 84 | 37081 | 1.0663 | 0.14315 | *EPHB2* |
| ENSG00000010626 | 12 | 6972733 | 7033407 | 140 | 10 | 30554 | 1.0662 | 0.14317 | *LRRC23* |
| ENSG00000159289 | 15 | 74352198 | 74384891 | 22 | 13 | 46545 | 1.066 | 0.14321 | *GOLGA6A* |
| ENSG00000173805 | 17 | 39863994 | 39900896 | 146 | 16 | 38809 | 1.0658 | 0.14326 | *HAP1* |
| ENSG00000082196 | 5 | 34009553 | 34053937 | 129 | 16 | 32693 | 1.0657 | 0.14328 | *C1QTNF3* |
| ENSG00000172428 | 2 | 241055980 | 241086224 | 202 | 23 | 51086 | 1.0656 | 0.1433 | *MYEOV2* |
| ENSG00000151224 | 10 | 82021576 | 82059440 | 144 | 24 | 43458 | 1.0649 | 0.14345 | *MAT1A* |
| ENSG00000167635 | 19 | 36695504 | 36739676 | 149 | 14 | 53867 | 1.0649 | 0.14346 | *ZNF146* |
| ENSG00000115091 | 2 | 114637537 | 114730173 | 313 | 29 | 35652 | 1.0647 | 0.14351 | *ACTR3* |
| ENSG00000099994 | 22 | 24567227 | 24595078 | 73 | 18 | 46349 | 1.0645 | 0.14354 | *SUSD2* |
| ENSG00000111704 | 12 | 7930390 | 7958655 | 199 | 19 | 38762 | 1.0645 | 0.14355 | *NANOG* |
| ENSG00000173113 | 11 | 64073932 | 64095556 | 31 | 10 | 41683 | 1.0645 | 0.14356 | *TRMT112* |
| ENSG00000154319 | 8 | 11268972 | 11342224 | 446 | 60 | 45532 | 1.0643 | 0.14359 | *FAM167A* |
| ENSG00000105131 | 19 | 15327730 | 15354246 | 29 | 16 | 47707 | 1.0642 | 0.14362 | *EPHX3* |
| ENSG00000160602 | 17 | 27042915 | 27080473 | 65 | 15 | 45076 | 1.064 | 0.14366 | *NEK8* |
| ENSG00000181982 | 4 | 24797739 | 24991826 | 619 | 81 | 42732 | 1.0635 | 0.14378 | *CCDC149* |
| ENSG00000269955 | 7 | 139016106 | 139117345 | 369 | 13 | 30155 | 1.0635 | 0.14378 | *LUC7L2* |
| ENSG00000255221 | 11 | 104953196 | 104982158 | 97 | 10 | 50115 | 1.0633 | 0.14382 | *CARD17* |
| ENSG00000123243 | 10 | 7591232 | 7718961 | 526 | 97 | 48355 | 1.0633 | 0.14383 | *ITIH5* |
| ENSG00000181192 | 10 | 12100971 | 12175224 | 259 | 30 | 37089 | 1.0633 | 0.14383 | *DHTKD1* |
| ENSG00000205669 | 14 | 74067649 | 74096592 | 125 | 20 | 40263 | 1.063 | 0.14388 | *ACOT6* |
| ENSG00000187600 | 2 | 46646329 | 46721564 | 231 | 25 | 47112 | 1.063 | 0.14389 | *TMEM247* |
| ENSG00000160781 | 1 | 156203206 | 156227881 | 52 | 17 | 53615 | 1.0622 | 0.14407 | *PAQR6* |
| ENSG00000108582 | 17 | 28695923 | 28807007 | 178 | 15 | 42279 | 1.062 | 0.14411 | *CPD* |
| ENSG00000255302 | 15 | 49160083 | 49182380 | 60 | 14 | 36461 | 1.0617 | 0.14418 | *EID1* |
| ENSG00000079950 | 6 | 132757006 | 132844337 | 289 | 46 | 40504 | 1.0614 | 0.14425 | *STX7* |
| ENSG00000146215 | 6 | 43257448 | 43286535 | 58 | 12 | 40463 | 1.0605 | 0.14445 | *CRIP3* |

| | | | | | | | | | |
|---|---|---|---|---|---|---|---|---|---|
| ENSG00000142347 | 19 | 8575674 | 8652461 | 238 | 42 | 41378 | 1.0605 | 0.14446 | *MYO1F* |
| ENSG00000164967 | 9 | 34600483 | 34622101 | 42 | 11 | 49768 | 1.0604 | 0.14448 | *RPP25L* |
| ENSG00000036672 | 11 | 119215925 | 119262436 | 113 | 14 | 53307 | 1.0604 | 0.14448 | *USP2* |
| ENSG00000099337 | 19 | 38800484 | 38829660 | 112 | 16 | 51725 | 1.0604 | 0.14448 | *KCNK6* |
| ENSG00000162572 | 1 | 1205816 | 1237409 | 100 | 24 | 41177 | 1.0603 | 0.14449 | *SCNN1D* |
| ENSG00000131979 | 14 | 55298726 | 55379570 | 274 | 28 | 43771 | 1.0603 | 0.1445 | *GCH1* |
| ENSG00000204315 | 6 | 32086484 | 32108068 | 56 | 20 | 40848 | 1.0601 | 0.14456 | *FKBPL* |
| ENSG00000211448 | 14 | 80653873 | 80864100 | 466 | 60 | 40537 | 1.0601 | 0.14456 | *DIO2* |
| ENSG00000128590 | 7 | 108200012 | 108225294 | 75 | 14 | 40131 | 1.0599 | 0.14459 | *DNAJB9* |
| ENSG00000167807 | 19 | 10406103 | 10436685 | 109 | 22 | 45485 | 1.0598 | 0.14463 | *CTD-2369P2.10* |
| ENSG00000267673 | 19 | 10406103 | 10436691 | 109 | 22 | 45485 | 1.0598 | 0.14463 | *FDX1L* |
| ENSG00000198336 | 17 | 45267812 | 45311045 | 106 | 26 | 51842 | 1.0595 | 0.14468 | *MYL4* |
| ENSG00000139610 | 12 | 51712227 | 51750463 | 113 | 21 | 45371 | 1.0594 | 0.14471 | *CELA1* |
| ENSG00000175387 | 18 | 45347922 | 45467515 | 270 | 19 | 48088 | 1.0593 | 0.14472 | *SMAD2* |
| ENSG00000171492 | 1 | 90276573 | 90412170 | 218 | 52 | 50416 | 1.0593 | 0.14473 | *LRRC8D* |
| ENSG00000168228 | 4 | 25304407 | 25382005 | 258 | 31 | 38423 | 1.0591 | 0.14477 | *ZCCHC4* |
| ENSG00000182329 | 2 | 202927978 | 203072585 | 395 | 49 | 49943 | 1.0591 | 0.14478 | *AC079354.1* |
| ENSG0000080618 | 13 | 46617321 | 46689211 | 235 | 17 | 52207 | 1.0589 | 0.14482 | *CPB2* |
| ENSG00000204305 | 6 | 32138745 | 32162101 | 70 | 18 | 45499 | 1.0587 | 0.14487 | *AGER* |
| ENSG00000068489 | 17 | 57222860 | 57292066 | 127 | 20 | 40374 | 1.0587 | 0.14487 | *PRR11* |
| ENSG00000180353 | 3 | 121340246 | 121389774 | 144 | 18 | 48536 | 1.0583 | 0.14496 | *HCLS1* |
| ENSG00000143797 | 2 | 8982820 | 9153942 | 320 | 34 | 47839 | 1.0582 | 0.14498 | *MBOAT2* |
| ENSG00000169006 | 2 | 11788304 | 11820290 | 119 | 31 | 47530 | 1.0578 | 0.14507 | *NTSR2* |
| ENSG00000149212 | 11 | 94888704 | 94975705 | 124 | 20 | 52206 | 1.0577 | 0.14509 | *SESN3* |
| ENSG00000079462 | 19 | 42791185 | 42817698 | 22 | 4 | 50681 | 1.0575 | 0.14513 | *PAFAH1B3* |
| ENSG00000162378 | 1 | 53182126 | 53303014 | 291 | 25 | 31202 | 1.0575 | 0.14514 | *ZYG11B* |
| ENSG00000145198 | 3 | 183938217 | 183970117 | 77 | 13 | 51627 | 1.0573 | 0.14518 | *VWA5B2* |
| ENSG00000166855 | 15 | 65430557 | 65487680 | 57 | 15 | 39686 | 1.057 | 0.14525 | *CLPX* |
| ENSG00000255837 | 12 | 11139094 | 11160474 | 128 | 6 | 23481 | 1.0569 | 0.14527 | *TAS2R20* |
| ENSG00000139323 | 12 | 89803495 | 89929801 | 356 | 25 | 55483 | 1.0569 | 0.14529 | *POC1B* |
| ENSG00000143365 | 1 | 151768547 | 151814348 | 103 | 28 | 48732 | 1.0567 | 0.14532 | *RORC* |
| ENSG00000121578 | 3 | 118920579 | 118969950 | 214 | 30 | 50421 | 1.0567 | 0.14533 | *B4GALT4* |
| ENSG00000160408 | 9 | 130637600 | 130677687 | 83 | 32 | 44942 | 1.0566 | 0.14534 | *ST6GALNAC6* |
| ENSG00000111331 | 12 | 113366157 | 113421054 | 193 | 20 | 39966 | 1.0566 | 0.14534 | *OAS3* |
| ENSG00000170835 | 9 | 135927365 | 135957248 | 62 | 17 | 37566 | 1.0566 | 0.14535 | *CEL* |
| ENSG00000123472 | 1 | 47088409 | 47149539 | 124 | 11 | 38846 | 1.0566 | 0.14536 | *ATPAF1* |

| | | | | | | | | | |
|---|---|---|---|---|---|---|---|---|---|
| ENSG00000174059 | 1 | 208047594 | 208094747 | 175 | 31 | 36352 | 1.0565 | 0.14536 | *CD34* |
| ENSG00000159915 | 19 | 44744318 | 44789470 | 98 | 11 | 34067 | 1.0565 | 0.14537 | *ZNF233* |
| ENSG00000092531 | 15 | 42773431 | 42847547 | 123 | 18 | 40543 | 1.0563 | 0.14542 | *SNAP23* |
| ENSG00000178460 | 8 | 67772984 | 67844283 | 101 | 15 | 44193 | 1.0559 | 0.1455 | *MCMDC2* |
| ENSG00000171987 | 11 | 4582653 | 4609050 | 180 | 40 | 42067 | 1.0559 | 0.14551 | *C11orf40* |
| ENSG00000179055 | 9 | 107446660 | 107467766 | 85 | 14 | 37714 | 1.0557 | 0.14555 | *OR13D1* |
| ENSG00000197768 | 9 | 140135713 | 140157934 | 76 | 10 | 39654 | 1.0555 | 0.1456 | *C9orf173* |
| ENSG00000118193 | 1 | 200510628 | 200599862 | 373 | 39 | 46111 | 1.0553 | 0.14565 | *KIF14* |
| ENSG00000250641 | 6 | 31664681 | 31695695 | 76 | 20 | 28979 | 1.0548 | 0.14576 | *MEGT1* |
| ENSG00000144837 | 3 | 119306689 | 119358658 | 197 | 26 | 41509 | 1.0546 | 0.14581 | *PLA1A* |
| ENSG00000121905 | 1 | 33341595 | 33374042 | 42 | 13 | 54458 | 1.0544 | 0.14585 | *HPCA* |
| ENSG00000175573 | 11 | 65674279 | 65696588 | 42 | 12 | 38061 | 1.0544 | 0.14585 | *C11orf68* |
| ENSG00000236311 | 10 | 102839078 | 102900883 | 199 | 30 | 51281 | 1.0542 | 0.1459 | *TLX1NB* |
| ENSG00000174628 | 16 | 19717778 | 19878907 | 314 | 30 | 40712 | 1.0539 | 0.14595 | *IQCK* |
| ENSG00000179152 | 3 | 44369611 | 44460943 | 207 | 19 | 44638 | 1.0537 | 0.14601 | *TCAIM* |
| ENSG00000161807 | 19 | 9215274 | 9236439 | 137 | 11 | 36954 | 1.0536 | 0.14602 | *OR7G1* |
| ENSG00000172725 | 11 | 67195519 | 67221292 | 44 | 10 | 36934 | 1.0533 | 0.14611 | *CORO1B* |
| ENSG00000197256 | 19 | 11264943 | 11318467 | 218 | 30 | 49465 | 1.0531 | 0.14614 | *KANK2* |
| ENSG00000163956 | 4 | 3498103 | 3544286 | 270 | 31 | 54519 | 1.0531 | 0.14615 | *LRPAP1* |
| ENSG00000065325 | 17 | 9715523 | 9805419 | 367 | 34 | 28512 | 1.0525 | 0.14628 | *GLP2R* |
| ENSG00000010327 | 3 | 52519354 | 52568511 | 91 | 19 | 49060 | 1.0525 | 0.14629 | *STAB1* |
| ENSG00000185798 | 3 | 196271056 | 196305545 | 85 | 13 | 48966 | 1.0524 | 0.14631 | *WDR53* |
| ENSG00000167978 | 16 | 2792330 | 2832539 | 54 | 17 | 41946 | 1.0524 | 0.14632 | *SRRM2* |
| ENSG00000159200 | 21 | 35875440 | 35997441 | 567 | 42 | 38683 | 1.0521 | 0.14637 | *RCAN1* |
| ENSG00000137767 | 15 | 45913346 | 45993492 | 374 | 50 | 44844 | 1.052 | 0.1464 | *SQRDL* |
| ENSG00000131459 | 5 | 179717690 | 179790387 | 380 | 49 | 49763 | 1.0517 | 0.14648 | *GFPT2* |
| ENSG00000128849 | 15 | 57658165 | 57852925 | 875 | 123 | 42903 | 1.0513 | 0.14655 | *CGNL1* |
| ENSG00000213853 | 16 | 10612279 | 10684555 | 349 | 73 | 40107 | 1.0512 | 0.14659 | *EMP2* |
| ENSG00000164708 | 7 | 44092326 | 44115186 | 69 | 10 | 51637 | 1.0509 | 0.14665 | *PGAM2* |
| ENSG00000176771 | 2 | 133419374 | 134336034 | 2489 | 272 | 46083 | 1.0509 | 0.14666 | *NCKAP5* |
| ENSG00000149600 | 20 | 31280493 | 31341803 | 212 | 15 | 47715 | 1.0503 | 0.14678 | *COMMD7* |
| ENSG00000164294 | 5 | 54445946 | 54472899 | 67 | 8 | 56444 | 1.0503 | 0.14679 | *GPX8* |
| ENSG00000120913 | 8 | 22425792 | 22465538 | 94 | 19 | 50232 | 1.0501 | 0.14682 | *PDLIM2* |
| ENSG00000069869 | 15 | 56109120 | 56295944 | 848 | 50 | 52998 | 1.0501 | 0.14683 | *NEDD4* |
| ENSG00000141433 | 18 | 894944 | 922173 | 109 | 31 | 45932 | 1.0501 | 0.14685 | *ADCYAP1* |
| ENSG00000126107 | 1 | 45458212 | 45487001 | 80 | 12 | 49512 | 1.05 | 0.14686 | *HECTD3* |

| | | | | | | | | | |
|---|---|---|---|---|---|---|---|---|---|
| ENSG00000123240 | 10 | 13131449 | 13190291 | 189 | 41 | 45637 | 1.05 | 0.14686 | *OPTN* |
| ENSG00000166165 | 14 | 103975996 | 103999448 | 61 | 11 | 44599 | 1.05 | 0.14686 | *CKB* |
| ENSG00000091164 | 18 | 54254439 | 54328831 | 167 | 21 | 42962 | 1.05 | 0.14686 | *TXNL1* |
| ENSG00000181767 | 11 | 55862519 | 55883457 | 179 | 7 | 55570 | 1.0499 | 0.14688 | *OR8H2* |
| ENSG00000064666 | 19 | 1016298 | 1049068 | 226 | 48 | 49311 | 1.0496 | 0.14694 | *CNN2* |
| ENSG00000095066 | 19 | 12863817 | 12993554 | 324 | 37 | 38283 | 1.0492 | 0.14704 | *HOOK2* |
| ENSG00000143862 | 1 | 202092532 | 202123869 | 89 | 28 | 43440 | 1.0491 | 0.14706 | *ARL8A* |
| ENSG00000180347 | 7 | 31543704 | 31708334 | 603 | 55 | 47672 | 1.0491 | 0.14706 | *CCDC129* |
| ENSG00000179761 | 17 | 27267531 | 27394234 | 234 | 49 | 47384 | 1.049 | 0.14709 | *PIPOX* |
| ENSG00000120262 | 6 | 151805165 | 151952328 | 638 | 76 | 42195 | 1.0486 | 0.14718 | *CCDC170* |
| ENSG00000196670 | 5 | 180264611 | 180298286 | 148 | 17 | 42194 | 1.0485 | 0.14721 | *ZFP62* |
| ENSG00000111642 | 12 | 6669249 | 6726642 | 94 | 11 | 36925 | 1.0485 | 0.14722 | *CHD4* |
| ENSG00000140694 | 16 | 14519558 | 14736585 | 388 | 39 | 51661 | 1.0482 | 0.14727 | *PARN* |
| ENSG00000133114 | 13 | 45553687 | 45617746 | 169 | 22 | 32612 | 1.0481 | 0.14731 | *GPALPP1* |
| ENSG00000141150 | 17 | 34048668 | 34080540 | 94 | 25 | 48343 | 1.048 | 0.14732 | *RASL10B* |
| ENSG00000114395 | 3 | 50378126 | 50405891 | 16 | 8 | 53035 | 1.0476 | 0.1474 | *CYB561D2* |
| ENSG00000059122 | 16 | 2951938 | 3011209 | 357 | 51 | 46602 | 1.0474 | 0.14745 | *FLYWCH1* |
| ENSG00000112039 | 6 | 35410138 | 35444880 | 121 | 20 | 29091 | 1.0474 | 0.14746 | *FANCE* |
| ENSG00000169714 | 3 | 128878327 | 128912765 | 41 | 16 | 45927 | 1.047 | 0.14754 | *CNBP* |
| ENSG00000264324 | 2 | 74428731 | 74600504 | 274 | 45 | 31294 | 1.0467 | 0.14762 | *RP11-287D1.3* |
| ENSG00000123836 | 1 | 207212801 | 207264369 | 83 | 15 | 47234 | 1.0464 | 0.14769 | *PFKFB2* |
| ENSG00000204822 | 2 | 74689085 | 74710449 | 17 | 11 | 50515 | 1.0456 | 0.14788 | *MRPL53* |
| ENSG00000158457 | 7 | 128774712 | 128818671 | 143 | 22 | 54063 | 1.0455 | 0.14789 | *TSPAN33* |
| ENSG00000145476 | 4 | 187102674 | 187144610 | 212 | 25 | 45991 | 1.0454 | 0.14791 | *CYP4V2* |
| ENSG00000137513 | 11 | 78137007 | 78295919 | 393 | 19 | 33698 | 1.045 | 0.14801 | *NARS2* |
| ENSG00000181885 | 17 | 7153222 | 7177302 | 49 | 9 | 54288 | 1.045 | 0.14802 | *CLDN7* |
| ENSG00000146143 | 6 | 57169603 | 57523375 | 693 | 76 | 39586 | 1.0444 | 0.14816 | *PRIM2* |
| ENSG00000155850 | 5 | 149330300 | 149383018 | 96 | 19 | 45879 | 1.0443 | 0.14817 | *SLC26A2* |
| ENSG00000113140 | 5 | 151030657 | 151076726 | 111 | 21 | 36185 | 1.0442 | 0.14819 | *SPARC* |
| ENSG00000050405 | 12 | 50559571 | 50687329 | 187 | 14 | 37787 | 1.0442 | 0.1482 | *LIMA1* |
| ENSG00000100079 | 22 | 37956255 | 37988623 | 115 | 24 | 35921 | 1.0439 | 0.14826 | *LGALS2* |
| ENSG00000174939 | 16 | 29901696 | 29941185 | 68 | 13 | 44988 | 1.0438 | 0.14828 | *ASPHD1* |
| ENSG00000118491 | 6 | 144175573 | 144269483 | 243 | 38 | 47777 | 1.0437 | 0.14832 | *ZC2HC1B* |
| ENSG00000173976 | 19 | 3759087 | 3782233 | 81 | 23 | 45705 | 1.0436 | 0.14833 | *RAX2* |
| ENSG00000162522 | 1 | 33197486 | 33250571 | 86 | 21 | 54024 | 1.0435 | 0.14835 | *KIAA1522* |
| ENSG00000171936 | 19 | 15842203 | 15863153 | 90 | 9 | 47880 | 1.0435 | 0.14835 | *OR10H3* |

| | | | | | | | | | |
|---|---|---|---|---|---|---|---|---|---|
| ENSG00000198704 | 6 | 28461073 | 28505992 | 136 | 22 | 36866 | 1.0433 | 0.14839 | *GPX6* |
| ENSG00000251357 | 22 | 24194375 | 24247410 | 226 | 41 | 45471 | 1.0431 | 0.14846 | *AP000350.10* |
| ENSG00000151577 | 3 | 113837499 | 113928254 | 283 | 45 | 44283 | 1.0427 | 0.14853 | *DRD3* |
| ENSG00000171044 | 8 | 10743555 | 11068875 | 1231 | 63 | 39709 | 1.0425 | 0.14858 | *XKR6* |
| ENSG00000184743 | 11 | 63381559 | 63449393 | 110 | 15 | 48146 | 1.0424 | 0.14861 | *ATL3* |
| ENSG00000132122 | 1 | 48751044 | 48947845 | 403 | 51 | 47561 | 1.0422 | 0.14865 | *SPATA6* |
| ENSG00000164113 | 4 | 123290121 | 123360957 | 146 | 25 | 41784 | 1.0421 | 0.14867 | *ADAD1* |
| ENSG00000050820 | 16 | 75252928 | 75311951 | 284 | 33 | 47661 | 1.0422 | 0.14867 | *BCAR1* |
| ENSG00000198399 | 2 | 24415733 | 24593583 | 516 | 25 | 51917 | 1.042 | 0.14872 | *ITSN2* |
| ENSG00000164402 | 5 | 132076509 | 132152933 | 65 | 25 | 43161 | 1.0419 | 0.14873 | *Sep-08* |
| ENSG00000092931 | 17 | 74721947 | 74787531 | 45 | 20 | 44662 | 1.0418 | 0.14875 | *MFSD11* |
| ENSG00000205693 | 12 | 27905671 | 27934209 | 117 | 8 | 41443 | 1.0416 | 0.14881 | *MANSC4* |
| ENSG00000188676 | 8 | 39782133 | 39883910 | 556 | 56 | 49033 | 1.0412 | 0.1489 | *IDO2* |
| ENSG00000117507 | 1 | 171096879 | 171140707 | 210 | 27 | 47697 | 1.0411 | 0.14891 | *FMO6P* |
| ENSG00000132661 | 20 | 23321373 | 23345414 | 83 | 13 | 37376 | 1.0411 | 0.14892 | *NXT1* |
| ENSG00000106785 | 9 | 100821557 | 100891494 | 143 | 24 | 39283 | 1.0405 | 0.14905 | *TRIM14* |
| ENSG00000273154 | 20 | 62330216 | 62380456 | 161 | 13 | 39475 | 1.0405 | 0.14905 | *RP4-583P15.15* |
| ENSG00000169217 | 16 | 30352087 | 30376682 | 33 | 10 | 24643 | 1.0405 | 0.14906 | *CD2BP2* |
| ENSG00000203795 | 10 | 124660217 | 124682627 | 59 | 9 | 54120 | 1.0402 | 0.14912 | *FAM24A* |
| ENSG00000188033 | 19 | 12678775 | 12760912 | 196 | 19 | 29092 | 1.04 | 0.14916 | *ZNF490* |
| ENSG00000159217 | 17 | 47064774 | 47143012 | 161 | 24 | 53007 | 1.0399 | 0.14919 | *IGF2BP1* |
| ENSG00000213339 | 19 | 10802106 | 10834113 | 84 | 14 | 53974 | 1.0399 | 0.1492 | *QTRT1* |
| ENSG00000151572 | 12 | 101101304 | 101532419 | 1451 | 163 | 51247 | 1.0398 | 0.14921 | *ANO4* |
| ENSG00000203863 | 6 | 89664246 | 89686685 | 50 | 17 | 51114 | 1.0391 | 0.14937 | *AL079342.1* |
| ENSG00000076513 | 12 | 110426991 | 110487568 | 131 | 20 | 39282 | 1.0391 | 0.14938 | *ANKRD13A* |
| ENSG00000056050 | 4 | 170640616 | 170689104 | 122 | 31 | 44890 | 1.0389 | 0.14942 | *C4orf27* |
| ENSG00000177511 | 18 | 55008044 | 55048962 | 82 | 13 | 34625 | 1.0389 | 0.14942 | *ST8SIA3* |
| ENSG00000204464 | 1 | 15480840 | 15507813 | 123 | 18 | 48334 | 1.0388 | 0.14944 | *C1orf195* |
| ENSG00000127152 | 14 | 99625624 | 99747861 | 271 | 54 | 36722 | 1.0388 | 0.14944 | *BCL11B* |
| ENSG00000162300 | 11 | 64841695 | 64865870 | 49 | 12 | 40934 | 1.0388 | 0.14945 | *ZFPL1* |
| ENSG00000255561 | 11 | 111734780 | 111761967 | 65 | 10 | 41636 | 1.0387 | 0.14947 | *FDXACB1* |
| ENSG00000197093 | 7 | 99746867 | 99776373 | 39 | 8 | 48611 | 1.0386 | 0.14949 | *GAL3ST4* |
| ENSG00000115155 | 2 | 26670071 | 26791566 | 379 | 65 | 37579 | 1.0386 | 0.1495 | *OTOF* |
| ENSG00000197114 | 20 | 62328817 | 62377494 | 146 | 11 | 37998 | 1.0386 | 0.1495 | *ZGPAT* |
| ENSG00000163515 | 3 | 108452271 | 108486231 | 110 | 15 | 42763 | 1.0386 | 0.14951 | *RETNLB* |
| ENSG00000072609 | 12 | 133388773 | 133542890 | 484 | 43 | 45990 | 1.0385 | 0.14953 | *CHFR* |

| | | | | | | | | | |
|---|---|---|---|---|---|---|---|---|---|
| ENSG00000153404 | 5 | 130373 | 200085 | 287 | 26 | 28426 | 1.0383 | 0.14957 | *PLEKHG4B* |
| ENSG00000100578 | 14 | 58884103 | 59025216 | 165 | 39 | 51553 | 1.0383 | 0.14957 | *KIAA0586* |
| ENSG00000125895 | 20 | 1151205 | 1176059 | 52 | 25 | 47729 | 1.0381 | 0.14962 | *TMEM74B* |
| ENSG00000178449 | 12 | 50495762 | 50524240 | 61 | 8 | 38218 | 1.0374 | 0.14977 | *COX14* |
| ENSG00000197647 | 19 | 12115547 | 12156556 | 76 | 11 | 37271 | 1.0374 | 0.14978 | *ZNF433* |
| ENSG00000214929 | 9 | 84593687 | 84620171 | 34 | 13 | 45290 | 1.0372 | 0.14981 | *SPATA31D1* |
| ENSG00000113649 | 5 | 145816874 | 145901524 | 217 | 20 | 38002 | 1.0372 | 0.14982 | *TCERG1* |
| ENSG00000132664 | 20 | 18437771 | 18475287 | 122 | 24 | 44412 | 1.0372 | 0.14983 | *POLR3F* |
| ENSG00000011295 | 17 | 15892694 | 15958329 | 167 | 17 | 42133 | 1.0367 | 0.14994 | *TTC19* |
| ENSG00000089351 | 19 | 35475688 | 35527375 | 176 | 23 | 44970 | 1.0366 | 0.14997 | *GRAMD1A* |
| ENSG00000105669 | 19 | 19000323 | 19040206 | 102 | 20 | 40225 | 1.0365 | 0.14999 | *COPE* |
| ENSG00000014641 | 2 | 63805743 | 63844331 | 67 | 11 | 39381 | 1.0364 | 0.15001 | *MDH1* |
| ENSG00000111049 | 12 | 81100691 | 81123452 | 39 | 10 | 36480 | 1.0355 | 0.15021 | *MYF5* |
| ENSG00000157800 | 7 | 139983493 | 140114233 | 574 | 83 | 48485 | 1.0354 | 0.15023 | *SLC37A3* |
| ENSG00000136521 | 3 | 179312478 | 179355435 | 81 | 13 | 47037 | 1.0352 | 0.15028 | *NDUFB5* |
| ENSG00000100575 | 14 | 58865212 | 58904332 | 32 | 10 | 45808 | 1.0353 | 0.15028 | *TIMM9* |
| ENSG00000184674 | 22 | 24366133 | 24394680 | 4 | 2 | 42982 | 1.0351 | 0.15032 | *GSTT1* |
| ENSG00000183166 | 7 | 71234476 | 71922136 | 2223 | 142 | 37382 | 1.0349 | 0.15035 | *CALN1* |
| ENSG00000107669 | 10 | 123489939 | 123698316 | 804 | 67 | 39994 | 1.0348 | 0.15037 | *ATE1* |
| ENSG00000198033 | 13 | 19737910 | 19765992 | 240 | 27 | 48087 | 1.0348 | 0.15037 | *TUBA3C* |
| ENSG00000156172 | 8 | 96247147 | 96291429 | 238 | 30 | 56308 | 1.0345 | 0.15044 | *C8orf37* |
| ENSG00000183653 | 21 | 30958360 | 31013067 | 198 | 29 | 41365 | 1.0344 | 0.15048 | *GRIK1-AS2* |
| ENSG00000070367 | 14 | 57660518 | 57745726 | 117 | 17 | 34363 | 1.0342 | 0.15053 | *EXOC5* |
| ENSG00000196917 | 12 | 123094824 | 123225390 | 373 | 42 | 39679 | 1.0338 | 0.1506 | *HCAR1* |
| ENSG00000142694 | 1 | 36777632 | 36799755 | 40 | 12 | 40294 | 1.0338 | 0.15062 | *EVA1B* |
| ENSG00000186298 | 12 | 111147485 | 111190744 | 112 | 16 | 37740 | 1.0337 | 0.15064 | *PPP1CC* |
| ENSG00000070886 | 1 | 22880057 | 22940087 | 262 | 45 | 48738 | 1.0336 | 0.15065 | *EPHA8* |
| ENSG00000135407 | 12 | 58181159 | 58222487 | 52 | 12 | 38099 | 1.0331 | 0.15079 | *AVIL* |
| ENSG00000141026 | 17 | 17370300 | 17406540 | 82 | 16 | 28778 | 1.0329 | 0.15084 | *MED9* |
| ENSG00000163344 | 1 | 154887210 | 154919467 | 67 | 12 | 32137 | 1.0328 | 0.15085 | *PMVK* |
| ENSG00000267976 | 21 | 37848165 | 37870058 | 104 | 20 | 40444 | 1.0328 | 0.15086 | *AP000695.1* |
| ENSG00000112592 | 6 | 170853390 | 170891958 | 121 | 13 | 54150 | 1.0327 | 0.15087 | *TBP* |
| ENSG00000058063 | 3 | 182501288 | 182649423 | 337 | 22 | 44184 | 1.0327 | 0.15088 | *ATP11B* |
| ENSG00000137106 | 9 | 37412663 | 37446987 | 82 | 19 | 46037 | 1.0326 | 0.1509 | *GRHPR* |
| ENSG00000151773 | 13 | 44388045 | 44463827 | 336 | 34 | 40234 | 1.0326 | 0.1509 | *CCDC122* |
| ENSG00000132570 | 5 | 134230596 | 134353649 | 203 | 37 | 41624 | 1.0322 | 0.15099 | *PCBD2* |

| | | | | | | | | | |
|---|---|---|---|---|---|---|---|---|---|
| ENSG00000267531 | 19 | 55841254 | 55866561 | 83 | 18 | 47724 | 1.032 | 0.15103 | *AC020922.1* |
| ENSG00000198610 | 10 | 5227425 | 5270912 | 308 | 22 | 42428 | 1.0316 | 0.15112 | *AKR1C4* |
| ENSG00000197415 | 3 | 156967531 | 157261408 | 909 | 82 | 47941 | 1.0314 | 0.15117 | *VEPH1* |
| ENSG00000125144 | 16 | 56690643 | 56711977 | 113 | 32 | 42854 | 1.0312 | 0.15122 | *MT1G* |
| ENSG00000125386 | 4 | 2616988 | 2744292 | 381 | 38 | 45291 | 1.031 | 0.15128 | *FAM193A* |
| ENSG00000140876 | 16 | 77746411 | 77786157 | 318 | 36 | 43806 | 1.0309 | 0.15129 | *NUDT7* |
| ENSG00000104918 | 19 | 7723930 | 7745334 | 61 | 26 | 45658 | 1.0308 | 0.15131 | *RETN* |
| ENSG00000105655 | 19 | 18535198 | 18559111 | 44 | 7 | 37927 | 1.0307 | 0.15135 | *ISYNA1* |
| ENSG00000269686 | 2 | 133864577 | 133886625 | 94 | 12 | 30216 | 1.0306 | 0.15137 | *AC011755.1* |
| ENSG00000157303 | 9 | 95810989 | 95857420 | 158 | 24 | 43087 | 1.0305 | 0.15138 | *SUSD3* |
| ENSG00000125954 | 14 | 65371203 | 65538521 | 494 | 64 | 40658 | 1.0305 | 0.15139 | *CHURC1-FNTB* |
| ENSG00000184898 | 2 | 152094454 | 152128393 | 81 | 12 | 30251 | 1.0301 | 0.15149 | *RBM43* |
| ENSG00000120798 | 12 | 95404005 | 95477479 | 221 | 32 | 51023 | 1.0299 | 0.15153 | *NR2C1* |
| ENSG00000184394 | 14 | 20601895 | 20622821 | 62 | 13 | 53453 | 1.0298 | 0.15155 | *OR4N5* |
| ENSG00000114349 | 3 | 50219045 | 50243949 | 40 | 11 | 31795 | 1.0297 | 0.15157 | *GNAT1* |
| ENSG00000113722 | 5 | 149536358 | 149574120 | 112 | 22 | 48619 | 1.0297 | 0.15157 | *CDX1* |
| ENSG00000039560 | 5 | 34646342 | 34842732 | 699 | 111 | 46336 | 1.0296 | 0.1516 | *RAI14* |
| ENSG00000030304 | 9 | 113421051 | 113573859 | 503 | 74 | 45228 | 1.0293 | 0.15166 | *MUSK* |
| ENSG00000124222 | 20 | 57216328 | 57264582 | 109 | 24 | 47482 | 1.0291 | 0.15171 | *STX16* |
| ENSG00000134318 | 2 | 11309887 | 11498456 | 563 | 35 | 41434 | 1.0291 | 0.15173 | *ROCK2* |
| ENSG00000101311 | 20 | 6045492 | 6114191 | 329 | 41 | 46195 | 1.029 | 0.15173 | *FERMT1* |
| ENSG00000133318 | 11 | 63438918 | 63537363 | 134 | 17 | 50280 | 1.029 | 0.15175 | *RTN3* |
| ENSG00000178796 | 1 | 151672909 | 151712281 | 81 | 17 | 42203 | 1.0283 | 0.1519 | *RIIAD1* |
| ENSG00000162894 | 1 | 207067731 | 207106592 | 78 | 16 | 35202 | 1.0283 | 0.15192 | *FAIM3* |
| ENSG00000071655 | 19 | 1566639 | 1602882 | 50 | 16 | 51011 | 1.028 | 0.15198 | *MBD3* |
| ENSG00000175728 | 11 | 130532851 | 130597247 | 113 | 29 | 41301 | 1.0279 | 0.152 | *C11orf44* |
| ENSG00000151952 | 12 | 129546270 | 130398211 | 3425 | 383 | 46300 | 1.0275 | 0.15209 | *TMEM132D* |
| ENSG00000182871 | 21 | 46815052 | 46943634 | 658 | 84 | 43650 | 1.0275 | 0.15209 | *COL18A1* |
| ENSG00000183828 | 14 | 105629275 | 105657660 | 67 | 8 | 53737 | 1.0272 | 0.15215 | *NUDT14* |
| ENSG00000204420 | 6 | 31676371 | 31704491 | 60 | 17 | 28230 | 1.0272 | 0.15216 | *C6orf25* |
| ENSG00000111846 | 6 | 10482456 | 10639601 | 605 | 81 | 42200 | 1.0272 | 0.15217 | *GCNT2* |
| ENSG00000119599 | 14 | 73383040 | 73436411 | 238 | 33 | 49705 | 1.0271 | 0.15218 | *DCAF4* |
| ENSG00000105974 | 7 | 116154839 | 116211233 | 173 | 23 | 51003 | 1.0271 | 0.15219 | *CAV1* |
| ENSG00000272214 | 12 | 121397641 | 121420095 | 86 | 18 | 52976 | 1.0271 | 0.15219 | *AC079602.1* |
| ENSG00000182566 | 19 | 7783843 | 7808792 | 105 | 20 | 38805 | 1.0265 | 0.15232 | *CLEC4G* |
| ENSG00000166073 | 15 | 40081233 | 40223093 | 334 | 56 | 41398 | 1.0265 | 0.15233 | *GPR176* |

| | | | | | | | | | |
|---|---|---|---|---|---|---|---|---|---|
| ENSG00000184014 | 11 | 9150372 | 9296937 | 360 | 34 | 46552 | 1.0257 | 0.15251 | *DENND5A* |
| ENSG00000257365 | 14 | 65371203 | 65539368 | 496 | 64 | 40636 | 1.0257 | 0.15251 | *FNTB* |
| ENSG00000108309 | 17 | 42375781 | 42406039 | 64 | 14 | 32782 | 1.0257 | 0.15251 | *RUNDC3A* |
| ENSG00000126247 | 19 | 36620477 | 36651255 | 104 | 29 | 49445 | 1.0256 | 0.15254 | *CAPNS1* |
| ENSG00000128313 | 22 | 36103919 | 36135530 | 147 | 18 | 41433 | 1.0255 | 0.15257 | *APOL5* |
| ENSG00000129048 | 3 | 132306081 | 132347811 | 90 | 17 | 41500 | 1.0254 | 0.15258 | *ACKR4* |
| ENSG00000188223 | 19 | 36226579 | 36255420 | 77 | 15 | 47479 | 1.0252 | 0.15264 | *AC002398.9* |
| ENSG00000152102 | 2 | 131795449 | 131861033 | 94 | 11 | 39082 | 1.0251 | 0.15265 | *FAM168B* |
| ENSG00000162877 | 1 | 205787150 | 205829260 | 124 | 30 | 35675 | 1.0249 | 0.1527 | *PM20D1* |
| ENSG00000121933 | 1 | 112015970 | 112116584 | 308 | 57 | 41594 | 1.0246 | 0.15277 | *ADORA3* |
| ENSG00000146476 | 6 | 151763422 | 151801236 | 142 | 21 | 39898 | 1.0245 | 0.1528 | *C6orf211* |
| ENSG00000138002 | 2 | 27657238 | 27722656 | 105 | 11 | 38554 | 1.0244 | 0.15282 | *IFT172* |
| ENSG00000244005 | 20 | 34245977 | 34297281 | 148 | 15 | 35554 | 1.0244 | 0.15282 | *NFS1* |
| ENSG00000172901 | 5 | 115288151 | 115373316 | 444 | 68 | 47455 | 1.0243 | 0.15285 | *AQPEP* |
| ENSG00000206557 | 3 | 32849510 | 32949318 | 258 | 45 | 46731 | 1.0241 | 0.1529 | *TRIM71* |
| ENSG00000112511 | 6 | 33368176 | 33394230 | 56 | 15 | 50525 | 1.0239 | 0.15293 | *PHF1* |
| ENSG00000196376 | 6 | 118218689 | 118648839 | 1336 | 125 | 47847 | 1.0237 | 0.15299 | *SLC35F1* |
| ENSG00000160194 | 21 | 44289754 | 44343414 | 277 | 21 | 42436 | 1.0237 | 0.153 | *NDUFV3* |
| ENSG00000103460 | 16 | 52461917 | 52591714 | 308 | 56 | 44750 | 1.0235 | 0.15303 | *TOX3* |
| ENSG00000175482 | 11 | 67108248 | 67134443 | 17 | 7 | 41105 | 1.0234 | 0.15306 | *POLD4* |
| ENSG00000116604 | 1 | 156423519 | 156480620 | 110 | 20 | 39968 | 1.0228 | 0.1532 | *MEF2D* |
| ENSG00000163803 | 2 | 28670012 | 28876654 | 827 | 117 | 46273 | 1.0226 | 0.15326 | *PLB1* |
| ENSG00000156222 | 15 | 85417885 | 85528876 | 476 | 81 | 46303 | 1.0225 | 0.15328 | *SLC28A1* |
| ENSG00000103222 | 16 | 16033434 | 16246931 | 836 | 114 | 44022 | 1.0224 | 0.15329 | *ABCC1* |
| ENSG00000198482 | 19 | 53020905 | 53077717 | 267 | 29 | 37980 | 1.0224 | 0.15329 | *ZNF808* |
| ENSG00000166477 | 15 | 52220222 | 52274003 | 131 | 10 | 49266 | 1.0222 | 0.15333 | *LEO1* |
| ENSG00000119333 | 9 | 131385940 | 131429066 | 83 | 14 | 37722 | 1.0222 | 0.15334 | *WDR34* |
| ENSG00000117139 | 1 | 202686526 | 202788598 | 305 | 27 | 41064 | 1.0219 | 0.15341 | *KDM5B* |
| ENSG00000008710 | 16 | 2128711 | 2195899 | 182 | 29 | 36680 | 1.0218 | 0.15344 | *PKD1* |
| ENSG00000166971 | 16 | 53514952 | 53548323 | 78 | 15 | 44647 | 1.0218 | 0.15344 | *AKTIP* |
| ENSG00000148680 | 10 | 92490578 | 92627671 | 402 | 33 | 38188 | 1.0215 | 0.1535 | *HTR7* |
| ENSG00000115211 | 2 | 27577219 | 27603353 | 44 | 11 | 42864 | 1.0215 | 0.15351 | *EIF2B4* |
| ENSG00000081853 | 5 | 140708539 | 140902546 | 527 | 60 | 43924 | 1.0213 | 0.15356 | *PCDHGA2* |
| ENSG00000214891 | 11 | 49065266 | 49090664 | 107 | 15 | 32718 | 1.0212 | 0.15359 | *TRIM64C* |
| ENSG00000267281 | 12 | 53890472 | 54029763 | 274 | 28 | 45246 | 1.0211 | 0.15361 | *RP11-793H13.10* |
| ENSG00000170653 | 12 | 53891640 | 54030199 | 274 | 28 | 45246 | 1.0211 | 0.15361 | *ATF7* |

| | | | | | | | | | |
|---|---|---|---|---|---|---|---|---|---|
| ENSG00000144554 | 3 | 10058098 | 10153614 | 317 | 12 | 39659 | 1.021 | 0.15363 | *FANCD2* |
| ENSG00000174093 | 17 | 36327717 | 36431858 | 68 | 16 | 41066 | 1.0209 | 0.15365 | *RP11-1407O15.2* |
| ENSG00000104679 | 8 | 23117633 | 23163792 | 187 | 35 | 46839 | 1.0208 | 0.15367 | *R3HCC1* |
| ENSG00000187612 | 11 | 55671126 | 55692058 | 189 | 8 | 56705 | 1.0206 | 0.15372 | *OR5W2* |
| ENSG00000183751 | 16 | 2012038 | 2042934 | 181 | 24 | 50175 | 1.0206 | 0.15373 | *TBL3* |
| ENSG00000159450 | 1 | 152068793 | 152096556 | 21 | 8 | 33537 | 1.0205 | 0.15374 | *TCHH* |
| ENSG00000213066 | 6 | 167402670 | 167476201 | 442 | 19 | 43074 | 1.0201 | 0.15383 | *FGFR1OP* |
| ENSG00000111215 | 12 | 10967559 | 11334212 | 1736 | 43 | 34624 | 1.0201 | 0.15385 | *PRR4* |
| ENSG00000143061 | 1 | 117107031 | 117220375 | 155 | 43 | 43859 | 1.0194 | 0.15402 | *IGSF3* |
| ENSG00000269605 | 20 | 43224346 | 43258126 | 84 | 17 | 47054 | 1.0193 | 0.15402 | *Z97053.1* |
| ENSG00000198542 | 13 | 102094966 | 102385456 | 993 | 109 | 44164 | 1.0192 | 0.15406 | *ITGBL1* |
| ENSG00000124818 | 6 | 47739718 | 47810516 | 253 | 26 | 54151 | 1.0191 | 0.15407 | *OPN5* |
| ENSG00000167113 | 9 | 131074815 | 131106351 | 63 | 13 | 33163 | 1.0191 | 0.15409 | *COQ4* |
| ENSG00000116473 | 1 | 112074840 | 112269313 | 639 | 70 | 50224 | 1.0188 | 0.15416 | *RAP1A* |
| ENSG00000187726 | 11 | 73651364 | 73691411 | 138 | 25 | 43124 | 1.0188 | 0.15416 | *DNAJB13* |
| ENSG00000115361 | 2 | 211042663 | 211100215 | 59 | 14 | 43465 | 1.0185 | 0.15422 | *ACADL* |
| ENSG00000168036 | 3 | 41226328 | 41311587 | 148 | 20 | 39698 | 1.0184 | 0.15424 | *CTNNB1* |
| ENSG00000177731 | 17 | 18138150 | 18172230 | 66 | 15 | 54617 | 1.0182 | 0.15429 | *FLII* |
| ENSG00000154252 | 2 | 242706240 | 242753623 | 150 | 29 | 39621 | 1.0179 | 0.15437 | *GAL3ST2* |
| ENSG00000012779 | 10 | 45859661 | 45951561 | 325 | 38 | 51013 | 1.0175 | 0.15446 | *ALOX5* |
| ENSG00000041988 | 1 | 6674926 | 6705646 | 80 | 14 | 54929 | 1.0175 | 0.15447 | *THAP3* |
| ENSG00000146833 | 7 | 99464581 | 99527223 | 167 | 16 | 40547 | 1.0173 | 0.1545 | *TRIM4* |
| ENSG00000108799 | 17 | 40842293 | 40907071 | 62 | 10 | 46690 | 1.0173 | 0.15451 | *EZH1* |
| ENSG00000130684 | 20 | 25644851 | 25687477 | 103 | 13 | 25754 | 1.0173 | 0.15451 | *ZNF337* |
| ENSG00000104889 | 19 | 12907394 | 12934452 | 114 | 10 | 32473 | 1.017 | 0.15457 | *RNASEH2A* |
| ENSG00000173153 | 11 | 64063044 | 64094215 | 37 | 11 | 43070 | 1.0166 | 0.15468 | *ESRRA* |
| ENSG00000139053 | 12 | 15115956 | 15144799 | 134 | 15 | 42894 | 1.0162 | 0.15476 | *PDE6H* |
| ENSG00000152705 | 5 | 134293596 | 134357392 | 115 | 19 | 41344 | 1.0162 | 0.15478 | *CATSPER3* |
| ENSG00000105991 | 7 | 27122612 | 27145615 | 48 | 17 | 48319 | 1.0161 | 0.15478 | *HOXA1* |
| ENSG00000188229 | 9 | 140125665 | 140148159 | 68 | 17 | 48504 | 1.0161 | 0.1548 | *TUBB4B* |
| ENSG00000136279 | 7 | 44074239 | 44119055 | 147 | 14 | 51918 | 1.0159 | 0.15484 | *DBNL* |
| ENSG00000250644 | 11 | 1758897 | 1790281 | 105 | 17 | 43870 | 1.0158 | 0.15487 | *RP11-295K3.1* |
| ENSG00000240510 | 1 | 151672909 | 151712070 | 79 | 16 | 41827 | 1.0156 | 0.1549 | *AL589765.1* |
| ENSG00000175305 | 8 | 95881998 | 95918906 | 130 | 20 | 53836 | 1.0154 | 0.15495 | *CCNE2* |
| ENSG00000146005 | 5 | 139165406 | 139234051 | 121 | 22 | 47917 | 1.0154 | 0.15497 | *PSD2* |
| ENSG00000105514 | 19 | 11422722 | 11466946 | 111 | 20 | 33787 | 1.0153 | 0.15497 | *RAB3D* |

| | | | | | | | | | |
|---|---|---|---|---|---|---|---|---|---|
| ENSG00000170296 | 17 | 7133333 | 7156089 | 26 | 7 | 55930 | 1.0151 | 0.15502 | *GABARAP* |
| ENSG00000187260 | 7 | 151062995 | 151117813 | 179 | 44 | 43778 | 1.015 | 0.15505 | *WDR86* |
| ENSG00000062524 | 15 | 41785836 | 41816085 | 84 | 20 | 50737 | 1.0145 | 0.15518 | *LTK* |
| ENSG00000112339 | 6 | 135271516 | 135434194 | 406 | 24 | 49637 | 1.0139 | 0.15532 | *HBS1L* |
| ENSG00000034239 | 8 | 49613348 | 49657870 | 95 | 27 | 51552 | 1.0137 | 0.15536 | *EFCAB1* |
| ENSG00000114867 | 3 | 184022283 | 184063146 | 75 | 20 | 50175 | 1.0136 | 0.15538 | *EIF4G1* |
| ENSG00000128654 | 2 | 177124123 | 177212753 | 188 | 22 | 50733 | 1.0136 | 0.15539 | *MTX2* |
| ENSG00000125458 | 17 | 73116320 | 73137890 | 44 | 11 | 36065 | 1.0135 | 0.1554 | *NT5C* |
| ENSG00000257103 | 19 | 34653409 | 34730420 | 240 | 20 | 51428 | 1.0135 | 0.15542 | *LSM14A* |
| ENSG00000167881 | 17 | 74025184 | 74078734 | 104 | 14 | 36958 | 1.0134 | 0.15544 | *SRP68* |
| ENSG00000144712 | 3 | 12827971 | 12923415 | 363 | 45 | 39363 | 1.0133 | 0.15546 | *CAND2* |
| ENSG00000104852 | 19 | 49578676 | 49621862 | 140 | 28 | 52895 | 1.0132 | 0.15549 | *SNRNP70* |
| ENSG00000184916 | 14 | 105597318 | 105645161 | 125 | 16 | 49651 | 1.013 | 0.15552 | *JAG2* |
| ENSG00000168913 | 9 | 34511038 | 34533039 | 34 | 11 | 53994 | 1.0127 | 0.1556 | *ENHO* |
| ENSG00000184185 | 17 | 21269509 | 21333179 | 88 | 31 | 42451 | 1.0125 | 0.15564 | *KCNJ12* |
| ENSG00000160685 | 1 | 154965127 | 155000998 | 58 | 16 | 44903 | 1.0122 | 0.15571 | *ZBTB7B* |
| ENSG00000072071 | 19 | 14250750 | 14326999 | 182 | 36 | 46171 | 1.0119 | 0.1558 | *LPHN1* |
| ENSG00000125814 | 20 | 23345159 | 23412125 | 249 | 20 | 42107 | 1.0118 | 0.15583 | *NAPB* |
| ENSG00000188322 | 16 | 28293840 | 28345170 | 107 | 27 | 40489 | 1.0116 | 0.15587 | *SBK1* |
| ENSG00000204296 | 6 | 32246303 | 32349684 | 982 | 54 | 36672 | 1.0115 | 0.15588 | *C6orf10* |
| ENSG00000011083 | 5 | 149559520 | 149612351 | 207 | 39 | 44039 | 1.011 | 0.15601 | *SLC6A7* |
| ENSG00000102445 | 13 | 46906139 | 47022325 | 292 | 23 | 43807 | 1.0109 | 0.15602 | *KIAA0226L* |
| ENSG00000142230 | 19 | 47606531 | 47723886 | 142 | 30 | 47238 | 1.0109 | 0.15603 | *SAE1* |
| ENSG00000164508 | 6 | 25716291 | 25736790 | 98 | 10 | 31578 | 1.0107 | 0.15607 | *HIST1H2AA* |
| ENSG00000146047 | 6 | 25717137 | 25737573 | 98 | 10 | 31578 | 1.0107 | 0.15607 | *HIST1H2BA* |
| ENSG00000167962 | 16 | 2037655 | 2069824 | 132 | 16 | 44723 | 1.0107 | 0.15608 | *ZNF598* |
| ENSG00000181896 | 19 | 19769605 | 19804318 | 125 | 23 | 52392 | 1.0107 | 0.15608 | *ZNF101* |
| ENSG00000171053 | 11 | 125606188 | 125629762 | 137 | 16 | 54341 | 1.0106 | 0.1561 | *PATE1* |
| ENSG00000125122 | 16 | 67231042 | 67270951 | 54 | 14 | 38884 | 1.0104 | 0.15614 | *LRRC29* |
| ENSG00000138376 | 2 | 215580370 | 215684428 | 583 | 61 | 53096 | 1.0104 | 0.15615 | *BARD1* |
| ENSG00000177764 | 20 | 267737 | 290965 | 74 | 20 | 43576 | 1.0103 | 0.15617 | *ZCCHC3* |
| ENSG00000183624 | 3 | 128987671 | 129035029 | 98 | 9 | 41120 | 1.0102 | 0.15619 | *HMCES* |
| ENSG00000160959 | 8 | 145733376 | 145760557 | 41 | 12 | 40430 | 1.0102 | 0.1562 | *LRRC14* |
| ENSG00000165730 | 10 | 70577298 | 70665188 | 285 | 43 | 49424 | 1.0101 | 0.15623 | *STOX1* |
| ENSG00000269676 | 12 | 133039334 | 133060726 | 84 | 25 | 46334 | 1.0101 | 0.15623 | *MUC8* |
| ENSG00000140092 | 14 | 92325756 | 92424331 | 309 | 64 | 50058 | 1.01 | 0.15624 | *FBLN5* |

| | | | | | | | | | |
|---|---|---|---|---|---|---|---|---|---|
| ENSG00000165449 | 10 | 61400523 | 61505760 | 317 | 55 | 45122 | 1.01 | 0.15626 | *SLC16A9* |
| ENSG00000112799 | 6 | 6578341 | 6665216 | 443 | 84 | 47894 | 1.0095 | 0.15636 | *LY86* |
| ENSG00000104522 | 8 | 144684788 | 144710218 | 72 | 18 | 37904 | 1.0094 | 0.15639 | *TSTA3* |
| ENSG00000047346 | 15 | 52863514 | 53012014 | 324 | 41 | 40639 | 1.0094 | 0.15639 | *FAM214A* |
| ENSG00000196936 | 1 | 248102160 | 248123098 | 65 | 8 | 45701 | 1.0092 | 0.15644 | *OR2L8* |
| ENSG00000006059 | 17 | 39492344 | 39517064 | 49 | 12 | 43001 | 1.009 | 0.15648 | *KRT33A* |
| ENSG00000180787 | 17 | 4971543 | 5009669 | 112 | 21 | 35444 | 1.009 | 0.15649 | *ZFP3* |
| ENSG00000183346 | 10 | 63412719 | 63536524 | 481 | 44 | 37022 | 1.0086 | 0.15657 | *C10orf107* |
| ENSG00000103175 | 16 | 84318252 | 84373450 | 324 | 80 | 49278 | 1.0086 | 0.15657 | *WFDC1* |
| ENSG00000135480 | 12 | 52616304 | 52655970 | 129 | 29 | 47587 | 1.0084 | 0.15664 | *KRT7* |
| ENSG00000144908 | 3 | 125812412 | 125926837 | 557 | 61 | 47315 | 1.0082 | 0.15668 | *ALDH1L1* |
| ENSG00000165496 | 14 | 47110222 | 47131028 | 54 | 12 | 49977 | 1.0082 | 0.15668 | *RPL10L* |
| ENSG00000131477 | 17 | 40900465 | 40925059 | 25 | 10 | 42018 | 1.0081 | 0.1567 | *RAMP2* |
| ENSG00000166188 | 16 | 58018572 | 58044357 | 77 | 13 | 51900 | 1.008 | 0.15673 | *ZNF319* |
| ENSG00000167123 | 9 | 131164030 | 131209626 | 123 | 13 | 28722 | 1.0078 | 0.15679 | *CERCAM* |
| ENSG00000163468 | 1 | 156268759 | 156347664 | 200 | 20 | 53777 | 1.0076 | 0.15682 | *CCT3* |
| ENSG00000108960 | 17 | 53459974 | 53509353 | 100 | 20 | 38761 | 1.0075 | 0.15684 | *MMD* |
| ENSG00000151247 | 4 | 99782835 | 99861788 | 271 | 28 | 38086 | 1.0075 | 0.15685 | *EIF4E* |
| ENSG00000196860 | 14 | 58852634 | 58885419 | 34 | 13 | 48782 | 1.0072 | 0.15691 | *TOMM20L* |
| ENSG00000103067 | 16 | 68253014 | 68282005 | 47 | 9 | 45641 | 1.0072 | 0.15693 | *ESRP2* |
| ENSG00000107862 | 10 | 103995289 | 104152656 | 282 | 31 | 50717 | 1.007 | 0.15696 | *GBF1* |
| ENSG00000268059 | 6 | 13605728 | 13631126 | 47 | 10 | 53260 | 1.0069 | 0.15698 | *AL441883.1* |
| ENSG00000139209 | 12 | 47148546 | 47236191 | 295 | 46 | 36366 | 1.0066 | 0.15705 | *SLC38A4* |
| ENSG00000177150 | 18 | 13653346 | 13736662 | 319 | 31 | 48590 | 1.0066 | 0.15706 | *FAM210A* |
| ENSG00000241043 | 9 | 32556787 | 32578619 | 52 | 11 | 50817 | 1.0062 | 0.15715 | *GVQW1* |
| ENSG00000173611 | 9 | 127694887 | 127915785 | 444 | 37 | 53902 | 1.0061 | 0.15718 | *SCAI* |
| ENSG00000176105 | 18 | 711588 | 822547 | 424 | 35 | 41793 | 1.0061 | 0.15719 | *YES1* |
| ENSG00000135521 | 6 | 144154481 | 144194949 | 108 | 17 | 53146 | 1.0059 | 0.15722 | *LTV1* |
| ENSG00000138617 | 15 | 65516798 | 65602956 | 268 | 26 | 44052 | 1.0057 | 0.15727 | *PARP16* |
| ENSG00000213654 | 6 | 32148543 | 32173300 | 81 | 24 | 43847 | 1.0057 | 0.15728 | *GPSM3* |
| ENSG00000186891 | 1 | 1128888 | 1152071 | 101 | 17 | 37909 | 1.0056 | 0.1573 | *TNFRSF18* |
| ENSG00000237102 | 16 | 67234251 | 67270930 | 47 | 14 | 41778 | 1.0055 | 0.15734 | *AC040160.1* |
| ENSG00000116883 | 1 | 36779335 | 36804822 | 46 | 14 | 40266 | 1.0054 | 0.15736 | *RP11-268J15.5* |
| ENSG00000230062 | 6 | 46704654 | 46737243 | 84 | 18 | 33491 | 1.005 | 0.15744 | *ANKRD66* |
| ENSG00000196584 | 7 | 152331864 | 152383250 | 225 | 30 | 41139 | 1.005 | 0.15745 | *XRCC2* |
| ENSG00000167526 | 16 | 89617065 | 89640950 | 68 | 10 | 53396 | 1.0049 | 0.15748 | *RPL13* |

| | | | | | | | | | |
|---|---|---|---|---|---|---|---|---|---|
| ENSG00000249049 | 4 | 92230185 | 92256457 | 107 | 10 | 50954 | 1.0048 | 0.15749 | *RP11-763F8.1* |
| ENSG00000100450 | 14 | 25065686 | 25088905 | 94 | 18 | 55264 | 1.0048 | 0.15751 | *GZMH* |
| ENSG00000167524 | 17 | 26924982 | 26951218 | 24 | 11 | 52496 | 1.0047 | 0.15753 | *SGK494* |
| ENSG00000079313 | 19 | 1805248 | 1858452 | 158 | 19 | 37372 | 1.0045 | 0.15758 | *REXO1* |
| ENSG00000250424 | 7 | 30883010 | 30973427 | 294 | 54 | 36896 | 1.0042 | 0.15764 | *AQP1* |
| ENSG00000144891 | 3 | 148405571 | 148470795 | 212 | 32 | 38214 | 1.0039 | 0.15772 | *AGTR1* |
| ENSG00000158022 | 1 | 26367795 | 26404927 | 105 | 15 | 38513 | 1.0033 | 0.15787 | *TRIM63* |
| ENSG00000129566 | 14 | 20823826 | 20891588 | 263 | 49 | 48567 | 1.0028 | 0.15798 | *TEP1* |
| ENSG00000117859 | 1 | 52032851 | 52264889 | 456 | 53 | 39509 | 1.0027 | 0.15801 | *OSBPL9* |
| ENSG00000187862 | 1 | 156539519 | 156566562 | 99 | 12 | 24520 | 1.0026 | 0.15802 | *TTC24* |
| ENSG00000164172 | 5 | 52381509 | 52415893 | 158 | 14 | 56175 | 1.0026 | 0.15803 | *MOCS2* |
| ENSG00000134987 | 5 | 110417414 | 110476200 | 226 | 30 | 41181 | 1.0026 | 0.15803 | *WDR36* |
| ENSG00000270188 | 1 | 238097024 | 238118575 | 116 | 13 | 46594 | 1.0026 | 0.15804 | *MTRNR2L11* |
| ENSG00000233502 | 2 | 10213908 | 10236228 | 103 | 21 | 49603 | 1.0025 | 0.15804 | *AC104794.4* |
| ENSG00000138777 | 4 | 106280234 | 106405238 | 525 | 38 | 44312 | 1.0025 | 0.15805 | *PPA2* |
| ENSG00000142599 | 1 | 8402457 | 8887702 | 875 | 64 | 45641 | 1.0024 | 0.15808 | *RERE* |
| ENSG00000181222 | 17 | 7377685 | 7427933 | 239 | 18 | 47210 | 1.0023 | 0.15811 | *POLR2A* |
| ENSG00000188738 | 2 | 186593355 | 186708017 | 316 | 14 | 54678 | 1.0022 | 0.15812 | *FSIP2* |
| ENSG00000225828 | 1 | 32816871 | 32839879 | 13 | 8 | 53934 | 1.0022 | 0.15813 | *FAM229A* |
| ENSG00000066230 | 5 | 463425 | 534447 | 369 | 56 | 37848 | 1.0021 | 0.15814 | *SLC9A3* |
| ENSG00000167755 | 19 | 51451887 | 51482929 | 87 | 23 | 43172 | 1.0019 | 0.15821 | *KLK6* |
| ENSG00000065833 | 6 | 83910108 | 84150797 | 646 | 40 | 44972 | 1.0017 | 0.15825 | *ME1* |
| ENSG00000213563 | 8 | 145741117 | 145764516 | 31 | 10 | 43555 | 1.0016 | 0.15826 | *C8orf82* |
| ENSG00000123427 | 12 | 58155275 | 58186324 | 41 | 5 | 32613 | 1.0016 | 0.15828 | *METTL21B* |
| ENSG00000100410 | 22 | 41845721 | 41874729 | 50 | 8 | 50213 | 1.0012 | 0.15836 | *PHF5A* |
| ENSG00000162227 | 11 | 62528775 | 62564814 | 58 | 13 | 35494 | 1.0012 | 0.15837 | *TAF6L* |
| ENSG00000204217 | 2 | 203231659 | 203442474 | 427 | 37 | 51945 | 1.0008 | 0.15845 | *BMPR2* |
| ENSG00000110881 | 12 | 50441331 | 50487394 | 79 | 16 | 35136 | 1.0006 | 0.1585 | *ASIC1* |
| ENSG00000054938 | 11 | 74397474 | 74452430 | 142 | 35 | 42675 | 1.0002 | 0.1586 | *CHRDL2* |
| ENSG00000161558 | 19 | 48825613 | 48877494 | 189 | 28 | 54607 | 1.0001 | 0.15862 | *TMEM143* |
| ENSG00000175182 | 3 | 184043714 | 184074063 | 83 | 22 | 52349 | 1.0001 | 0.15863 | *FAM131A* |
| ENSG00000128694 | 2 | 190601386 | 190637953 | 79 | 16 | 47969 | 0.99997 | 0.15866 | *OSGEPL1* |
| ENSG00000145388 | 4 | 119596523 | 119646588 | 122 | 15 | 37266 | 0.99999 | 0.15866 | *METTL14* |
| ENSG00000165105 | 9 | 85584500 | 85688092 | 191 | 19 | 50863 | 0.99985 | 0.15869 | *RASEF* |
| ENSG00000188886 | 2 | 96779589 | 96814175 | 53 | 11 | 49566 | 0.9998 | 0.1587 | *ASTL* |
| ENSG00000184361 | 17 | 43321760 | 43349479 | 66 | 14 | 43445 | 0.9998 | 0.1587 | *SPATA32* |

| | | | | | | | | | |
|---|---|---|---|---|---|---|---|---|---|
| ENSG00000186854 | 2 | 85038774 | 85144132 | 368 | 39 | 42536 | 0.99973 | 0.15872 | *TRABD2A* |
| ENSG00000153922 | 5 | 98180908 | 98272240 | 207 | 24 | 43447 | 0.99959 | 0.15875 | *CHD1* |
| ENSG00000135723 | 16 | 67253290 | 67291561 | 40 | 17 | 49347 | 0.99953 | 0.15877 | *FHOD1* |
| ENSG00000079805 | 19 | 10818755 | 10954164 | 350 | 40 | 44153 | 0.99946 | 0.15879 | *DNM2* |
| ENSG00000188283 | 19 | 37698828 | 37744828 | 77 | 16 | 43055 | 0.99946 | 0.15879 | *ZNF383* |
| ENSG00000050327 | 7 | 144042381 | 144087725 | 26 | 6 | 52696 | 0.99932 | 0.15882 | *ARHGEF5* |
| ENSG00000168993 | 4 | 768745 | 829986 | 319 | 40 | 49389 | 0.99913 | 0.15887 | *CPLX1* |
| ENSG00000163466 | 2 | 219071817 | 219129079 | 123 | 16 | 44371 | 0.99903 | 0.15889 | *ARPC2* |
| ENSG00000086289 | 7 | 37713446 | 38001543 | 1182 | 111 | 45226 | 0.99897 | 0.1589 | *EPDR1* |
| ENSG00000142507 | 17 | 4689439 | 4711790 | 60 | 19 | 40017 | 0.99877 | 0.15895 | *PSMB6* |
| ENSG00000120457 | 11 | 128751251 | 128800930 | 189 | 37 | 46710 | 0.99852 | 0.15901 | *KCNJ5* |
| ENSG00000146592 | 7 | 28328940 | 28875511 | 1842 | 256 | 48966 | 0.99848 | 0.15902 | *CREB5* |
| ENSG00000122550 | 7 | 23135353 | 23227533 | 296 | 18 | 36749 | 0.99828 | 0.15907 | *KLHL7* |
| ENSG00000196169 | 17 | 72312349 | 72361959 | 176 | 37 | 42809 | 0.99822 | 0.15909 | *KIF19* |
| ENSG00000154144 | 11 | 124482732 | 124515287 | 55 | 20 | 45874 | 0.99808 | 0.15912 | *TBRG1* |
| ENSG00000135801 | 1 | 229718858 | 229771794 | 113 | 16 | 54120 | 0.99784 | 0.15918 | *TAF5L* |
| ENSG00000103021 | 16 | 58255061 | 58327740 | 358 | 26 | 48552 | 0.99762 | 0.15923 | *CCDC113* |
| ENSG00000116701 | 1 | 183514698 | 183570011 | 162 | 27 | 45521 | 0.99742 | 0.15928 | *NCF2* |
| ENSG00000160201 | 21 | 44503066 | 44537697 | 59 | 12 | 43471 | 0.99735 | 0.1593 | *U2AF1* |
| ENSG00000212122 | 5 | 112758251 | 112780728 | 106 | 13 | 33790 | 0.99723 | 0.15933 | *TSSK1B* |
| ENSG00000259171 | 14 | 21142746 | 21178735 | 158 | 33 | 49797 | 0.99711 | 0.15935 | *AL163636.6* |
| ENSG00000185386 | 22 | 50692142 | 50719196 | 124 | 26 | 48654 | 0.99714 | 0.15935 | *MAPK11* |
| ENSG00000257594 | 12 | 89903185 | 89930039 | 95 | 11 | 54610 | 0.99697 | 0.15939 | *GALNT4* |
| ENSG00000259075 | 12 | 89903185 | 89930039 | 95 | 11 | 54610 | 0.99697 | 0.15939 | *POC1B-GALNT4* |
| ENSG00000114353 | 3 | 50253724 | 50306787 | 58 | 18 | 45225 | 0.9965 | 0.1595 | *GNAI2* |
| ENSG00000167964 | 16 | 2180804 | 2214166 | 56 | 16 | 37758 | 0.99623 | 0.15957 | *RAB26* |
| ENSG00000073350 | 17 | 73511161 | 73581289 | 409 | 49 | 38099 | 0.99616 | 0.15959 | *LLGL2* |
| ENSG00000177238 | 16 | 31215342 | 31246510 | 25 | 10 | 43105 | 0.99596 | 0.15963 | *TRIM72* |
| ENSG00000107807 | 10 | 102879257 | 102907545 | 89 | 24 | 49343 | 0.99578 | 0.15968 | *TLX1* |
| ENSG00000180613 | 4 | 54955690 | 54978672 | 42 | 8 | 41144 | 0.99574 | 0.15969 | *GSX2* |
| ENSG00000147604 | 8 | 74192506 | 74218024 | 87 | 13 | 53374 | 0.99553 | 0.15974 | *RPL7* |
| ENSG00000091972 | 3 | 112041194 | 112091659 | 176 | 23 | 44211 | 0.99546 | 0.15976 | *CD200* |
| ENSG00000175455 | 3 | 123606152 | 123690564 | 133 | 20 | 41449 | 0.99539 | 0.15977 | *CCDC14* |
| ENSG00000214193 | 1 | 36761988 | 36800484 | 66 | 14 | 41402 | 0.99501 | 0.15987 | *SH3D21* |
| ENSG00000126860 | 17 | 29634578 | 29658902 | 39 | 4 | 25678 | 0.9946 | 0.15997 | *EVI2A* |
| ENSG00000125247 | 13 | 101246181 | 101337347 | 285 | 26 | 47772 | 0.9944 | 0.16001 | *TMTC4* |

| | | | | | | | | | |
|---|---|---|---|---|---|---|---|---|---|
| ENSG00000153558 | 3 | 33308517 | 33455154 | 389 | 34 | 41979 | 0.99391 | 0.16013 | *FBXL2* |
| ENSG00000135632 | 2 | 73431350 | 73464365 | 89 | 16 | 42835 | 0.9937 | 0.16018 | *SMYD5* |
| ENSG00000124702 | 6 | 42971951 | 42999036 | 68 | 13 | 34647 | 0.99373 | 0.16018 | *KLHDC3* |
| ENSG00000181903 | 11 | 55422614 | 55443671 | 18 | 8 | 44334 | 0.99362 | 0.1602 | *OR4C6* |
| ENSG00000188130 | 22 | 50673879 | 50710254 | 155 | 25 | 40711 | 0.99364 | 0.1602 | *MAPK12* |
| ENSG00000146700 | 7 | 76008651 | 76049012 | 186 | 26 | 47504 | 0.99349 | 0.16024 | *SRCRB4D* |
| ENSG00000114302 | 3 | 48772030 | 48895279 | 120 | 21 | 39401 | 0.99292 | 0.16037 | *PRKAR2A* |
| ENSG00000170396 | 2 | 185453093 | 185814219 | 878 | 63 | 43517 | 0.99271 | 0.16043 | *ZNF804A* |
| ENSG00000172071 | 2 | 88846259 | 88937094 | 146 | 24 | 40860 | 0.99247 | 0.16048 | *EIF2AK3* |
| ENSG00000197233 | 9 | 125263081 | 125284022 | 57 | 21 | 42104 | 0.9925 | 0.16048 | *OR1J2* |
| ENSG00000134690 | 1 | 38148090 | 38185391 | 128 | 14 | 46741 | 0.99233 | 0.16052 | *CDCA8* |
| ENSG00000180205 | 20 | 44226578 | 44269907 | 169 | 16 | 47625 | 0.99227 | 0.16053 | *WFDC9* |
| ENSG00000118985 | 5 | 95210802 | 95307775 | 270 | 20 | 29903 | 0.9922 | 0.16055 | *ELL2* |
| ENSG00000146856 | 7 | 134661259 | 134842715 | 480 | 21 | 52072 | 0.99217 | 0.16056 | *AGBL3* |
| ENSG00000185313 | 3 | 38728293 | 38845501 | 429 | 59 | 45799 | 0.99181 | 0.16064 | *SCN10A* |
| ENSG00000181019 | 16 | 69730899 | 69770854 | 109 | 21 | 48674 | 0.9915 | 0.16072 | *NQO1* |
| ENSG00000116096 | 2 | 73104489 | 73129287 | 25 | 6 | 36737 | 0.99112 | 0.16081 | *SPR* |
| ENSG00000177885 | 17 | 73304157 | 73411790 | 306 | 19 | 28949 | 0.99068 | 0.16092 | *GRB2* |
| ENSG00000177352 | 3 | 49189968 | 49213754 | 32 | 6 | 47812 | 0.99058 | 0.16094 | *CCDC71* |
| ENSG00000164118 | 4 | 175194828 | 175264531 | 159 | 32 | 43032 | 0.99025 | 0.16103 | *CEP44* |
| ENSG00000231360 | 1 | 144329738 | 144531058 | 5 | 3 | 33454 | 0.99004 | 0.16108 | *AL592284.1* |
| ENSG00000172318 | 2 | 168665182 | 168740551 | 220 | 40 | 46018 | 0.98993 | 0.1611 | *B3GALT1* |
| ENSG00000100814 | 14 | 20769527 | 20811471 | 147 | 20 | 45200 | 0.98986 | 0.16112 | *CCNB1IP1* |
| ENSG00000146918 | 7 | 158414003 | 158507520 | 319 | 26 | 48658 | 0.9898 | 0.16113 | *NCAPG2* |
| ENSG00000257921 | 12 | 58156811 | 58190829 | 49 | 5 | 32111 | 0.98969 | 0.16116 | *RP11-571M6.15* |
| ENSG00000132681 | 1 | 160111360 | 160166767 | 225 | 55 | 45940 | 0.98941 | 0.16123 | *ATP1A4* |
| ENSG00000173432 | 11 | 18277721 | 18301524 | 95 | 16 | 33907 | 0.98939 | 0.16123 | *SAA1* |
| ENSG00000240891 | 3 | 111383523 | 111575294 | 714 | 81 | 42877 | 0.98931 | 0.16125 | *PLCXD2* |
| ENSG00000079257 | 3 | 158353611 | 158400482 | 168 | 10 | 56776 | 0.9891 | 0.16131 | *LXN* |
| ENSG00000116035 | 2 | 71117720 | 71170576 | 193 | 38 | 48745 | 0.98888 | 0.16136 | *VAX2* |
| ENSG00000173757 | 17 | 40341186 | 40438725 | 120 | 15 | 48634 | 0.98818 | 0.16153 | *STAT5B* |
| ENSG00000163449 | 2 | 216936589 | 216977506 | 91 | 16 | 43800 | 0.98808 | 0.16156 | *TMEM169* |
| ENSG00000206053 | 16 | 1718257 | 1762281 | 75 | 11 | 40487 | 0.98787 | 0.16161 | *HN1L* |
| ENSG00000177721 | 5 | 43029335 | 43053272 | 55 | 13 | 34192 | 0.98783 | 0.16162 | *ANXA2R* |
| ENSG00000175489 | 19 | 18491954 | 18518427 | 98 | 19 | 47313 | 0.98752 | 0.16169 | *LRRC25* |
| ENSG00000178878 | 12 | 12868851 | 12992909 | 350 | 88 | 46458 | 0.98732 | 0.16174 | *APOLD1* |

| | | | | | | | | | |
|---|---|---|---|---|---|---|---|---|---|
| ENSG00000141431 | 18 | 31148579 | 31341156 | 509 | 55 | 46012 | 0.98725 | 0.16176 | *ASXL3* |
| ENSG00000180210 | 11 | 46730730 | 46771056 | 92 | 15 | 43134 | 0.98707 | 0.1618 | *F2* |
| ENSG00000135363 | 11 | 33870122 | 33923836 | 156 | 37 | 50323 | 0.98684 | 0.16186 | *LMO2* |
| ENSG00000240583 | 7 | 30883010 | 30975131 | 296 | 54 | 36735 | 0.98676 | 0.16188 | *AQP1* |
| ENSG00000167182 | 17 | 45963516 | 46016323 | 133 | 19 | 50881 | 0.98663 | 0.16191 | *SP2* |
| ENSG00000108622 | 17 | 62069954 | 62107994 | 56 | 13 | 50409 | 0.98625 | 0.16201 | *ICAM2* |
| ENSG00000164176 | 5 | 83226373 | 83690611 | 1087 | 138 | 49001 | 0.98618 | 0.16202 | *EDIL3* |
| ENSG00000204469 | 6 | 31578497 | 31615548 | 161 | 26 | 41711 | 0.9857 | 0.16214 | *PRRC2A* |
| ENSG00000165929 | 14 | 92236095 | 92343880 | 248 | 36 | 47120 | 0.98543 | 0.16221 | *TC2N* |
| ENSG00000204397 | 11 | 104902053 | 104982158 | 268 | 27 | 44534 | 0.98509 | 0.16229 | *CARD16* |
| ENSG00000100949 | 14 | 24724744 | 24750945 | 60 | 15 | 38817 | 0.98489 | 0.16234 | *RABGGTA* |
| ENSG00000129159 | 11 | 17746359 | 17814602 | 167 | 38 | 47205 | 0.98441 | 0.16246 | *KCNC1* |
| ENSG00000140488 | 15 | 72567068 | 72622470 | 87 | 16 | 50364 | 0.98411 | 0.16253 | *CELF6* |
| ENSG00000174792 | 4 | 76471258 | 76501095 | 112 | 20 | 45438 | 0.98408 | 0.16254 | *C4orf26* |
| ENSG00000139718 | 12 | 122232086 | 122280562 | 79 | 13 | 35674 | 0.984 | 0.16256 | *SETD1B* |
| ENSG00000152580 | 3 | 151133172 | 151186497 | 196 | 31 | 38832 | 0.98356 | 0.16267 | *IGSF10* |
| ENSG00000269590 | 19 | 12765642 | 12790211 | 40 | 16 | 39556 | 0.98352 | 0.16267 | *CTD-2192J16.24* |
| ENSG00000004487 | 1 | 23335941 | 23420182 | 129 | 15 | 33192 | 0.98337 | 0.16271 | *KDM1A* |
| ENSG00000262601 | 16 | 85194882 | 85228969 | 146 | 33 | 46231 | 0.98274 | 0.16287 | *CTC-786C10.1* |
| ENSG00000213214 | 7 | 143873176 | 144001230 | 19 | 9 | 42154 | 0.98254 | 0.16292 | *ARHGEF35* |
| ENSG00000204283 | 17 | 75865109 | 75888659 | 86 | 15 | 50139 | 0.98254 | 0.16292 | *FLJ45079* |
| ENSG00000008441 | 19 | 13096422 | 13219610 | 225 | 36 | 44962 | 0.9823 | 0.16297 | *NFIX* |
| ENSG00000156453 | 5 | 141222938 | 141268811 | 88 | 31 | 45160 | 0.98226 | 0.16299 | *PCDH1* |
| ENSG00000171307 | 10 | 99195927 | 99227127 | 59 | 14 | 49220 | 0.98181 | 0.1631 | *ZDHHC16* |
| ENSG00000258365 | 12 | 94661534 | 94686620 | 118 | 22 | 54109 | 0.98181 | 0.1631 | *RP11-1105G2.3* |
| ENSG00000169618 | 2 | 68860721 | 68892708 | 112 | 26 | 35714 | 0.98156 | 0.16316 | *PROKR1* |
| ENSG00000138395 | 2 | 202645184 | 202770273 | 338 | 55 | 38198 | 0.98109 | 0.16327 | *CDK15* |
| ENSG00000182230 | 5 | 175477692 | 175569261 | 115 | 23 | 34957 | 0.981 | 0.1633 | *FAM153B* |
| ENSG00000185303 | 10 | 81305608 | 81330153 | 121 | 20 | 53250 | 0.98036 | 0.16345 | *SFTPA2* |
| ENSG00000142149 | 21 | 33235628 | 33426946 | 730 | 122 | 45994 | 0.98024 | 0.16348 | *HUNK* |
| ENSG00000119608 | 14 | 75309736 | 75340537 | 72 | 13 | 37686 | 0.9801 | 0.16352 | *PROX2* |
| ENSG00000214967 | 16 | 16462912 | 16497811 | 3 | 2 | 42947 | 0.97997 | 0.16355 | *NPIPA7* |
| ENSG00000165209 | 9 | 125861779 | 126040855 | 181 | 44 | 47801 | 0.97992 | 0.16356 | *STRBP* |
| ENSG00000163362 | 1 | 200850176 | 200894863 | 159 | 40 | 42545 | 0.97987 | 0.16358 | *C1orf106* |
| ENSG00000250741 | 2 | 18726811 | 18780830 | 115 | 26 | 40022 | 0.97934 | 0.16371 | *NT5C1B-RDH14* |
| ENSG00000185013 | 2 | 18727050 | 18780838 | 115 | 26 | 40022 | 0.97934 | 0.16371 | *NT5C1B* |

| | | | | | | | | | |
|---|---|---|---|---|---|---|---|---|---|
| ENSG00000085871 | 4 | 140576922 | 140671899 | 351 | 64 | 44387 | 0.97934 | 0.16371 | *MGST2* |
| ENSG00000100271 | 22 | 43425522 | 43495434 | 274 | 42 | 33970 | 0.97875 | 0.16385 | *TTLL1* |
| ENSG00000174937 | 11 | 56226964 | 56248014 | 83 | 14 | 52025 | 0.97847 | 0.16392 | *OR5M3* |
| ENSG00000268898 | 2 | 40478147 | 40500117 | 107 | 21 | 42200 | 0.97828 | 0.16397 | *AC007377.1* |
| ENSG00000172493 | 4 | 87846154 | 88072206 | 618 | 62 | 46235 | 0.97802 | 0.16403 | *AFF1* |
| ENSG00000130749 | 19 | 47557444 | 47627009 | 131 | 23 | 37539 | 0.97802 | 0.16403 | *ZC3H4* |
| ENSG00000136824 | 9 | 106846541 | 106913698 | 174 | 15 | 50725 | 0.9779 | 0.16406 | *SMC2* |
| ENSG00000124493 | 6 | 33979628 | 34133399 | 693 | 86 | 33963 | 0.97787 | 0.16407 | *GRM4* |
| ENSG00000185332 | 17 | 79275074 | 79314474 | 158 | 25 | 45182 | 0.9778 | 0.16409 | *TMEM105* |
| ENSG00000163833 | 3 | 121301966 | 121359139 | 190 | 34 | 45977 | 0.97754 | 0.16415 | *FBXO40* |
| ENSG00000185926 | 11 | 51505282 | 51526211 | 93 | 12 | 44331 | 0.97742 | 0.16418 | *OR4C46* |
| ENSG00000011376 | 3 | 45419998 | 45600913 | 400 | 44 | 42198 | 0.97731 | 0.16421 | *LARS2* |
| ENSG00000180697 | 3 | 126235842 | 126287808 | 212 | 31 | 42209 | 0.97731 | 0.16421 | *C3orf22* |
| ENSG00000065618 | 10 | 105781044 | 105855760 | 145 | 28 | 42596 | 0.97724 | 0.16422 | *COL17A1* |
| ENSG00000188372 | 7 | 76016835 | 76081388 | 249 | 35 | 44660 | 0.9771 | 0.16426 | *ZP3* |
| ENSG00000081019 | 1 | 114294454 | 114365098 | 127 | 20 | 44520 | 0.97693 | 0.1643 | *RSBN1* |
| ENSG00000198838 | 15 | 33593163 | 34168303 | 2486 | 256 | 43840 | 0.97673 | 0.16435 | *RYR3* |
| ENSG00000205176 | 8 | 86558695 | 86585726 | 3 | 1 | 43326 | 0.97664 | 0.16437 | *REXO1L1P* |
| ENSG00000176209 | 8 | 42386298 | 42419603 | 110 | 14 | 35343 | 0.97509 | 0.16476 | *SMIM19* |
| ENSG00000205111 | 2 | 39392787 | 39466729 | 174 | 21 | 39036 | 0.97483 | 0.16482 | *CDKL4* |
| ENSG00000137496 | 11 | 71699587 | 71726761 | 74 | 10 | 35897 | 0.97472 | 0.16485 | *IL18BP* |
| ENSG00000169683 | 17 | 79971178 | 79999027 | 87 | 14 | 36365 | 0.97466 | 0.16486 | *LRRC45* |
| ENSG00000213002 | 8 | 71475681 | 71496671 | 48 | 11 | 39763 | 0.9742 | 0.16498 | *AC120194.1* |
| ENSG00000148655 | 10 | 77350998 | 78329925 | 2129 | 216 | 38467 | 0.97403 | 0.16502 | *C10orf11* |
| ENSG00000176165 | 14 | 29225050 | 29248870 | 27 | 16 | 50954 | 0.97401 | 0.16503 | *FOXG1* |
| ENSG00000168701 | 16 | 67251006 | 67273181 | 19 | 8 | 52806 | 0.974 | 0.16503 | *TMEM208* |
| ENSG00000067715 | 12 | 79247773 | 79855788 | 1618 | 127 | 47293 | 0.97352 | 0.16515 | *SYT1* |
| ENSG00000105810 | 7 | 92224235 | 92475908 | 315 | 53 | 40672 | 0.97346 | 0.16516 | *CDK6* |
| ENSG00000204424 | 6 | 31664640 | 31695581 | 75 | 19 | 28596 | 0.97293 | 0.1653 | *LY6G6F* |
| ENSG00000159674 | 4 | 1150720 | 1212750 | 174 | 36 | 36309 | 0.97283 | 0.16532 | *SPON2* |
| ENSG00000129194 | 17 | 7481496 | 7503488 | 43 | 11 | 44756 | 0.97277 | 0.16533 | *SOX15* |
| ENSG00000119396 | 9 | 123930415 | 123995292 | 144 | 24 | 40753 | 0.97276 | 0.16534 | *RAB14* |
| ENSG00000099904 | 22 | 20106979 | 20145530 | 155 | 18 | 48832 | 0.9724 | 0.16543 | *ZDHHC8* |
| ENSG00000141505 | 17 | 7066750 | 7092883 | 78 | 25 | 44985 | 0.9719 | 0.16555 | *ASGR1* |
| ENSG00000213722 | 6 | 31684815 | 31708394 | 49 | 16 | 30966 | 0.97187 | 0.16556 | *DDAH2* |
| ENSG00000101421 | 20 | 32389110 | 32452172 | 83 | 14 | 35315 | 0.97179 | 0.16558 | *CHMP4B* |

| | | | | | | | | | |
|---|---|---|---|---|---|---|---|---|---|
| ENSG00000214097 | 3 | 196223748 | 196252237 | 86 | 18 | 33517 | 0.97152 | 0.16564 | *SMCO1* |
| ENSG00000165338 | 10 | 93160096 | 93284586 | 143 | 23 | 30918 | 0.97152 | 0.16564 | *HECTD2* |
| ENSG00000079616 | 16 | 29792040 | 29826706 | 24 | 14 | 47527 | 0.97094 | 0.16579 | *KIF22* |
| ENSG00000115457 | 2 | 217487551 | 217539159 | 122 | 28 | 43639 | 0.97066 | 0.16586 | *IGFBP2* |
| ENSG00000180917 | 16 | 71305292 | 71333618 | 97 | 15 | 43655 | 0.97056 | 0.16588 | *CMTR2* |
| ENSG00000174282 | 17 | 7352685 | 7397582 | 167 | 17 | 49122 | 0.97058 | 0.16588 | *ZBTB4* |
| ENSG00000206044 | 12 | 2948397 | 2976213 | 74 | 15 | 32188 | 0.9705 | 0.1659 | *AC005841.1* |
| ENSG00000140497 | 15 | 75126071 | 75175706 | 82 | 16 | 36514 | 0.97043 | 0.16592 | *SCAMP2* |
| ENSG00000103257 | 16 | 87853629 | 87913094 | 452 | 51 | 33889 | 0.97028 | 0.16595 | *SLC7A5* |
| ENSG00000136160 | 13 | 78459616 | 78503903 | 125 | 18 | 42220 | 0.97023 | 0.16596 | *EDNRB* |
| ENSG00000179111 | 17 | 8013908 | 8037410 | 67 | 19 | 46741 | 0.97013 | 0.16599 | *HES7* |
| ENSG00000129911 | 19 | 1842399 | 1873567 | 91 | 13 | 30929 | 0.96979 | 0.16608 | *KLF16* |
| ENSG00000168389 | 1 | 40410802 | 40445638 | 66 | 21 | 39083 | 0.96974 | 0.16609 | *MFSD2A* |
| ENSG00000179314 | 17 | 5665554 | 6037747 | 1394 | 196 | 41480 | 0.9697 | 0.1661 | *WSCD1* |
| ENSG00000139714 | 12 | 122079024 | 122120537 | 154 | 23 | 35683 | 0.96946 | 0.16616 | *MORN3* |
| ENSG00000124900 | 11 | 55640773 | 55669286 | 149 | 10 | 50557 | 0.96913 | 0.16624 | *TRIM51* |
| ENSG00000198856 | 4 | 109561740 | 109598976 | 105 | 5 | 51130 | 0.96909 | 0.16625 | *OSTC* |
| ENSG00000123104 | 12 | 26480342 | 26996131 | 1808 | 170 | 41470 | 0.96856 | 0.16638 | *ITPR2* |
| ENSG00000115306 | 2 | 54673422 | 54906812 | 798 | 69 | 46121 | 0.96846 | 0.16641 | *SPTBN1* |
| ENSG00000005020 | 7 | 26696681 | 27044858 | 980 | 44 | 50982 | 0.96781 | 0.16657 | *SKAP2* |
| ENSG00000100030 | 22 | 22098789 | 22231970 | 378 | 37 | 53388 | 0.96779 | 0.16657 | *MAPK1* |
| ENSG00000259224 | 17 | 7374721 | 7396383 | 103 | 11 | 50742 | 0.96772 | 0.16659 | *SLC35G6* |
| ENSG00000162891 | 1 | 207028699 | 207052568 | 39 | 13 | 40659 | 0.96727 | 0.1667 | *IL20* |
| ENSG00000069188 | 17 | 71320523 | 71650228 | 1063 | 176 | 46513 | 0.9664 | 0.16692 | *SDK2* |
| ENSG00000136280 | 7 | 45029074 | 45126068 | 378 | 29 | 47239 | 0.96629 | 0.16695 | *CCM2* |
| ENSG00000116641 | 1 | 62910399 | 63163969 | 406 | 17 | 38496 | 0.96622 | 0.16697 | *DOCK7* |
| ENSG00000168556 | 4 | 184416147 | 184442249 | 69 | 17 | 48234 | 0.96605 | 0.16701 | *ING2* |
| ENSG00000198453 | 19 | 37397231 | 37499602 | 523 | 23 | 51758 | 0.96602 | 0.16702 | *ZNF568* |
| ENSG00000117152 | 1 | 163028565 | 163056592 | 85 | 20 | 52037 | 0.96593 | 0.16704 | *RGS4* |
| ENSG00000138411 | 2 | 197049094 | 197468416 | 1261 | 96 | 37636 | 0.96576 | 0.16708 | *HECW2* |
| ENSG00000011332 | 19 | 38691646 | 38730354 | 102 | 22 | 43925 | 0.96557 | 0.16713 | *DPF1* |
| ENSG00000100403 | 22 | 41687526 | 41766151 | 132 | 17 | 43951 | 0.96552 | 0.16714 | *ZC3H7B* |
| ENSG00000151276 | 3 | 65329200 | 66034509 | 2530 | 235 | 43122 | 0.96547 | 0.16715 | *MAGI1* |
| ENSG00000198108 | 5 | 129230165 | 129532327 | 549 | 41 | 50582 | 0.96534 | 0.16719 | *CHSY3* |
| ENSG00000267963 | 2 | 241043907 | 241074304 | 193 | 23 | 46155 | 0.96502 | 0.16727 | *AC013469.1* |
| ENSG00000175121 | 20 | 43728093 | 43753813 | 94 | 14 | 37644 | 0.96487 | 0.16731 | *WFDC5* |

| | | | | | | | | | |
|---|---|---|---|---|---|---|---|---|---|
| ENSG00000105982 | 7 | 156422975 | 156479824 | 159 | 26 | 38902 | 0.96475 | 0.16733 | *RNF32* |
| ENSG00000181904 | 5 | 134171370 | 134205427 | 38 | 13 | 48770 | 0.96445 | 0.16741 | *C5orf24* |
| ENSG00000196415 | 19 | 830963 | 858175 | 95 | 31 | 43987 | 0.96427 | 0.16745 | *PRTN3* |
| ENSG00000143740 | 1 | 227906240 | 227978927 | 194 | 30 | 44218 | 0.96393 | 0.16754 | *SNAP47* |
| ENSG00000107014 | 9 | 5289868 | 5314969 | 79 | 17 | 45439 | 0.96388 | 0.16755 | *RLN2* |
| ENSG00000140464 | 15 | 74277014 | 74350153 | 198 | 24 | 48625 | 0.96383 | 0.16757 | *PML* |
| ENSG00000117266 | 1 | 205463723 | 205511921 | 223 | 47 | 42653 | 0.96357 | 0.16763 | *CDK18* |
| ENSG00000172590 | 14 | 23289088 | 23314246 | 93 | 24 | 50263 | 0.96358 | 0.16763 | *MRPL52* |
| ENSG00000130312 | 19 | 17393418 | 17427652 | 157 | 19 | 43227 | 0.96323 | 0.16772 | *MRPL34* |
| ENSG00000138686 | 4 | 122735595 | 122801652 | 135 | 21 | 42442 | 0.96292 | 0.16779 | *BBS7* |
| ENSG00000203909 | 6 | 74052785 | 74074018 | 86 | 18 | 54015 | 0.96253 | 0.16789 | *DPPA5* |
| ENSG00000110066 | 11 | 67912330 | 67991295 | 181 | 22 | 52863 | 0.96245 | 0.16791 | *SUV420H1* |
| ENSG00000254466 | 11 | 59234846 | 59255869 | 63 | 13 | 32642 | 0.96225 | 0.16796 | *OR4D10* |
| ENSG00000166272 | 10 | 104493727 | 104586021 | 284 | 31 | 39028 | 0.96221 | 0.16797 | *WBP1L* |
| ENSG00000148399 | 9 | 140439356 | 140483387 | 84 | 18 | 47764 | 0.96194 | 0.16804 | *DPH7* |
| ENSG00000106483 | 7 | 37935543 | 38075297 | 656 | 88 | 48879 | 0.96149 | 0.16815 | *SFRP4* |
| ENSG00000168077 | 8 | 27481385 | 27544293 | 230 | 19 | 48743 | 0.96147 | 0.16816 | *SCARA3* |
| ENSG00000258818 | 14 | 21142259 | 21178761 | 162 | 34 | 49957 | 0.96133 | 0.16819 | *RNASE4* |
| ENSG00000224838 | 5 | 59716565 | 59736939 | 95 | 14 | 40196 | 0.96129 | 0.1682 | *FKSG52* |
| ENSG00000073060 | 12 | 125251402 | 125377214 | 344 | 87 | 44900 | 0.96061 | 0.16837 | *SCARB1* |
| ENSG00000134686 | 1 | 33779224 | 33906653 | 432 | 35 | 46021 | 0.96042 | 0.16842 | *PHC2* |
| ENSG00000127249 | 3 | 193109866 | 193320900 | 605 | 78 | 52323 | 0.96041 | 0.16842 | *ATP13A4* |
| ENSG00000168003 | 11 | 62613518 | 62666352 | 84 | 19 | 40056 | 0.96036 | 0.16844 | *SLC3A2* |
| ENSG00000137713 | 11 | 111587632 | 111647151 | 131 | 13 | 32031 | 0.96037 | 0.16844 | *PPP2R1B* |
| ENSG00000116539 | 1 | 155295059 | 155542598 | 232 | 37 | 40692 | 0.95993 | 0.16855 | *ASH1L* |
| ENSG00000269881 | 16 | 289546 | 329942 | 140 | 28 | 35827 | 0.95981 | 0.16857 | *ITFG3* |
| ENSG00000186716 | 22 | 23511891 | 23670224 | 549 | 71 | 47343 | 0.95958 | 0.16863 | *BCR* |
| ENSG00000133812 | 11 | 9790214 | 10325754 | 2017 | 67 | 45737 | 0.95941 | 0.16868 | *SBF2* |
| ENSG00000163131 | 1 | 150692672 | 150748433 | 135 | 13 | 51354 | 0.95928 | 0.16871 | *CTSS* |
| ENSG00000072401 | 10 | 60084735 | 60140513 | 129 | 20 | 37779 | 0.9592 | 0.16873 | *UBE2D1* |
| ENSG00000168453 | 8 | 21961928 | 22000897 | 122 | 21 | 52504 | 0.95904 | 0.16877 | *HR* |
| ENSG00000204304 | 6 | 32142512 | 32167963 | 78 | 22 | 44474 | 0.95888 | 0.16881 | *PBX2* |
| ENSG00000146859 | 7 | 134822824 | 134860967 | 120 | 9 | 53678 | 0.95877 | 0.16884 | *TMEM140* |
| ENSG00000152402 | 11 | 106534738 | 106899250 | 1256 | 100 | 51214 | 0.95868 | 0.16886 | *GUCY1A2* |
| ENSG00000231256 | 17 | 41847803 | 41874980 | 75 | 9 | 47365 | 0.95862 | 0.16887 | *C17orf105* |
| ENSG00000256655 | 8 | 128949126 | 128970591 | 67 | 17 | 44533 | 0.95851 | 0.1689 | *TMEM75* |

| | | | | | | | | | |
|---|---|---|---|---|---|---|---|---|---|
| ENSG00000115896 | 2 | 198659426 | 199447305 | 1721 | 137 | 48864 | 0.95839 | 0.16893 | *PLCL1* |
| ENSG00000173930 | 5 | 101559690 | 101642253 | 390 | 40 | 46272 | 0.95834 | 0.16895 | *SLCO4C1* |
| ENSG00000268358 | 5 | 111745280 | 111766310 | 29 | 14 | 45561 | 0.95772 | 0.1691 | *EPB41L4A-AS2* |
| ENSG00000146039 | 6 | 25744927 | 25791419 | 188 | 23 | 34289 | 0.95774 | 0.1691 | *SLC17A4* |
| ENSG00000254245 | 5 | 140713601 | 140902546 | 515 | 58 | 43659 | 0.95751 | 0.16916 | *PCDHGA3* |
| ENSG00000170027 | 7 | 75946116 | 75998348 | 137 | 21 | 42549 | 0.95696 | 0.16929 | *YWHAG* |
| ENSG00000105711 | 19 | 35511588 | 35541352 | 67 | 14 | 51509 | 0.95698 | 0.16929 | *SCN1B* |
| ENSG00000167383 | 19 | 44911685 | 44962766 | 266 | 37 | 42141 | 0.95694 | 0.1693 | *ZNF229* |
| ENSG00000003756 | 3 | 50116341 | 50166454 | 80 | 12 | 38690 | 0.95668 | 0.16936 | *RBM5* |
| ENSG00000162078 | 16 | 2870170 | 2898967 | 71 | 14 | 49762 | 0.95664 | 0.16937 | *ZG16B* |
| ENSG00000162989 | 2 | 155544811 | 155724863 | 623 | 86 | 44983 | 0.95657 | 0.16939 | *KCNJ3* |
| ENSG00000267796 | 19 | 36229262 | 36255420 | 72 | 15 | 47392 | 0.95657 | 0.16939 | *LIN37* |
| ENSG00000064042 | 4 | 41351624 | 41712061 | 1010 | 87 | 37577 | 0.9563 | 0.16946 | *LIMCH1* |
| ENSG00000146411 | 6 | 134299835 | 134383774 | 186 | 42 | 42627 | 0.95622 | 0.16948 | *SLC2A12* |
| ENSG00000268317 | 16 | 56652971 | 56676150 | 128 | 23 | 40909 | 0.95612 | 0.16951 | *AC026461.1* |
| ENSG00000255468 | 11 | 66105421 | 66142275 | 69 | 9 | 35838 | 0.95601 | 0.16953 | *RP11-867G23.8* |
| ENSG00000133422 | 22 | 31311117 | 31374284 | 99 | 15 | 43923 | 0.9558 | 0.16959 | *MORC2* |
| ENSG00000186446 | 3 | 44761088 | 44788575 | 52 | 10 | 46764 | 0.9557 | 0.16961 | *ZNF501* |
| ENSG00000231389 | 6 | 33022346 | 33058552 | 555 | 26 | 24520 | 0.95566 | 0.16962 | *HLA-DPA1* |
| ENSG00000168143 | 6 | 54701569 | 54816820 | 458 | 42 | 41679 | 0.95563 | 0.16963 | *FAM83B* |
| ENSG00000173727 | 11 | 65212728 | 65244028 | 103 | 11 | 48970 | 0.95556 | 0.16965 | *AP000769.1* |
| ENSG00000154262 | 17 | 67064843 | 67148029 | 169 | 20 | 38790 | 0.95497 | 0.1698 | *ABCA6* |
| ENSG00000129007 | 15 | 68473043 | 68508417 | 71 | 12 | 35558 | 0.95468 | 0.16987 | *CALML4* |
| ENSG00000069956 | 15 | 52234303 | 52368462 | 288 | 36 | 38925 | 0.95454 | 0.16991 | *MAPK6* |
| ENSG00000159363 | 1 | 17302453 | 17348423 | 121 | 20 | 41227 | 0.95404 | 0.17003 | *ATP13A2* |
| ENSG00000139410 | 12 | 113850042 | 113886081 | 97 | 10 | 41846 | 0.95406 | 0.17003 | *SDSL* |
| ENSG00000255552 | 6 | 31669548 | 31691842 | 55 | 16 | 27179 | 0.95396 | 0.17005 | *LY6G6E* |
| ENSG00000154124 | 5 | 14654773 | 14709820 | 110 | 20 | 45329 | 0.95393 | 0.17006 | *FAM105B* |
| ENSG00000142621 | 1 | 15563768 | 15736779 | 574 | 90 | 42816 | 0.95383 | 0.17008 | *FHAD1* |
| ENSG00000145975 | 6 | 4039668 | 4097578 | 215 | 29 | 48614 | 0.95375 | 0.1701 | *FAM217A* |
| ENSG00000189332 | 11 | 18220685 | 18246109 | 110 | 9 | 40167 | 0.95379 | 0.1701 | *RP11-113D6.10* |
| ENSG00000167720 | 17 | 2196677 | 2238554 | 95 | 16 | 38140 | 0.9537 | 0.17012 | *SRR* |
| ENSG00000213341 | 10 | 101938055 | 101999376 | 149 | 17 | 33364 | 0.95358 | 0.17015 | *CHUK* |
| ENSG00000039650 | 19 | 50354461 | 50381166 | 78 | 17 | 46556 | 0.95357 | 0.17015 | *PNKP* |
| ENSG00000149136 | 11 | 57083459 | 57113351 | 75 | 14 | 52470 | 0.95353 | 0.17016 | *SSRP1* |
| ENSG00000214479 | 6 | 83772692 | 83792854 | 41 | 7 | 52756 | 0.95346 | 0.17018 | *AL139333.1* |

| | | | | | | | | | |
|---|---|---|---|---|---|---|---|---|---|
| ENSG00000164483 | 6 | 130455460 | 130696570 | 1200 | 73 | 45690 | 0.95307 | 0.17028 | *SAMD3* |
| ENSG00000081803 | 7 | 121948481 | 122536813 | 1662 | 137 | 49436 | 0.95306 | 0.17028 | *CADPS2* |
| ENSG00000154001 | 14 | 63828075 | 64020092 | 555 | 50 | 36571 | 0.95303 | 0.17029 | *PPP2R5E* |
| ENSG00000124615 | 6 | 39857354 | 39912290 | 206 | 36 | 45826 | 0.95261 | 0.1704 | *MOCS1* |
| ENSG00000106305 | 7 | 6038876 | 6073465 | 179 | 27 | 50366 | 0.95258 | 0.1704 | *AIMP2* |
| ENSG00000150048 | 12 | 10212153 | 10274226 | 125 | 19 | 29948 | 0.95256 | 0.17041 | *CLEC1A* |
| ENSG00000139437 | 12 | 110328069 | 110431646 | 261 | 26 | 37947 | 0.95209 | 0.17053 | *TCHP* |
| ENSG00000100078 | 22 | 31520795 | 31546593 | 66 | 20 | 39027 | 0.95201 | 0.17055 | *PLA2G3* |
| ENSG00000119514 | 9 | 101559981 | 101622363 | 173 | 33 | 47387 | 0.95189 | 0.17058 | *GALNT12* |
| ENSG00000101670 | 18 | 47077069 | 47129272 | 144 | 35 | 50241 | 0.95138 | 0.17071 | *LIPG* |
| ENSG00000100100 | 22 | 31667579 | 31698520 | 80 | 15 | 36251 | 0.95122 | 0.17075 | *PIK3IP1* |
| ENSG00000166033 | 10 | 124211041 | 124284424 | 316 | 43 | 45182 | 0.95117 | 0.17076 | *HTRA1* |
| ENSG00000166926 | 11 | 60092304 | 60174069 | 304 | 29 | 53273 | 0.95072 | 0.17087 | *MS4A6E* |
| ENSG00000100253 | 22 | 50915213 | 50939077 | 74 | 35 | 47852 | 0.95067 | 0.17089 | *MIOX* |
| ENSG00000104044 | 15 | 27990021 | 28354504 | 891 | 88 | 37333 | 0.95038 | 0.17096 | *OCA2* |
| ENSG00000138092 | 2 | 25006005 | 25055245 | 203 | 26 | 44889 | 0.95031 | 0.17098 | *CENPO* |
| ENSG00000145685 | 5 | 77771038 | 78075844 | 741 | 90 | 41600 | 0.95017 | 0.17101 | *LHFPL2* |
| ENSG0000053108 | 5 | 132522147 | 132958255 | 1320 | 202 | 48513 | 0.94994 | 0.17107 | *FSTL4* |
| ENSG00000197757 | 12 | 54374408 | 54434607 | 113 | 24 | 50109 | 0.94951 | 0.17118 | *HOXC6* |
| ENSG00000198185 | 20 | 45119701 | 45152198 | 72 | 14 | 49573 | 0.94946 | 0.17119 | *ZNF334* |
| ENSG00000123739 | 4 | 110621145 | 110661233 | 81 | 16 | 50652 | 0.94944 | 0.1712 | *PLA2G12A* |
| ENSG00000188687 | 2 | 74433369 | 74580541 | 239 | 42 | 31370 | 0.9494 | 0.17121 | *SLC4A5* |
| ENSG00000099954 | 22 | 17830837 | 18047850 | 915 | 137 | 36610 | 0.9494 | 0.17121 | *CECR2* |
| ENSG00000179918 | 16 | 30444952 | 30467502 | 29 | 14 | 40133 | 0.94928 | 0.17124 | *SEPHS2* |
| ENSG00000134086 | 3 | 10172692 | 10203904 | 124 | 17 | 39683 | 0.94923 | 0.17125 | *VHL* |
| ENSG00000162438 | 1 | 15754935 | 15785737 | 84 | 19 | 49482 | 0.94902 | 0.17131 | *CTRC* |
| ENSG00000149231 | 11 | 96075933 | 96133087 | 142 | 22 | 54686 | 0.94894 | 0.17133 | *CCDC82* |
| ENSG00000011143 | 17 | 56272803 | 56306966 | 60 | 15 | 44746 | 0.94875 | 0.17137 | *MKS1* |
| ENSG00000269223 | 20 | 62464779 | 62485273 | 88 | 13 | 25653 | 0.94859 | 0.17141 | *AL158091.1* |
| ENSG00000143140 | 1 | 147218332 | 147255484 | 152 | 25 | 50727 | 0.94857 | 0.17142 | *GJA5* |
| ENSG00000215346 | 8 | 10973980 | 10997745 | 106 | 18 | 37658 | 0.94826 | 0.1715 | *AF131215.5* |
| ENSG00000185880 | 15 | 45011186 | 45070027 | 225 | 23 | 36313 | 0.94814 | 0.17153 | *TRIM69* |
| ENSG00000130023 | 6 | 170141718 | 170191680 | 170 | 17 | 37528 | 0.94807 | 0.17155 | *ERMARD* |
| ENSG00000145362 | 4 | 113729265 | 114314896 | 1847 | 214 | 44913 | 0.94803 | 0.17156 | *ANK2* |
| ENSG00000149177 | 11 | 47992113 | 48199670 | 431 | 45 | 39470 | 0.94801 | 0.17156 | *PTPRJ* |
| ENSG00000272869 | 5 | 112186885 | 112238776 | 209 | 22 | 39694 | 0.94758 | 0.17167 | *CTC-487M23.8* |

| | | | | | | | | | |
|---|---|---|---|---|---|---|---|---|---|
| ENSG00000126088 | 1 | 45467819 | 45491247 | 55 | 9 | 51106 | 0.94746 | 0.1717 | *UROD* |
| ENSG00000204508 | 1 | 13025543 | 13048381 | 3 | 1 | 42590 | 0.94736 | 0.17173 | *PRAMEF22* |
| ENSG00000124507 | 6 | 34423916 | 34513006 | 243 | 43 | 38128 | 0.94726 | 0.17175 | *PACSIN1* |
| ENSG00000205683 | 14 | 73076004 | 73370809 | 991 | 138 | 44280 | 0.94729 | 0.17175 | *DPF3* |
| ENSG00000112079 | 6 | 36451669 | 36525247 | 238 | 19 | 29987 | 0.9468 | 0.17187 | *STK38* |
| ENSG00000108840 | 17 | 42144114 | 42211070 | 130 | 21 | 34874 | 0.94667 | 0.1719 | *HDAC5* |
| ENSG00000159527 | 1 | 153260338 | 153293194 | 83 | 18 | 40414 | 0.94626 | 0.17201 | *PGLYRP3* |
| ENSG00000121380 | 12 | 12192778 | 12374018 | 587 | 90 | 48444 | 0.94612 | 0.17204 | *BCL2L14* |
| ENSG00000182500 | 12 | 122054455 | 122090583 | 92 | 17 | 41102 | 0.94602 | 0.17207 | *ORAI1* |
| ENSG00000158786 | 1 | 20455819 | 20486879 | 67 | 27 | 48713 | 0.94598 | 0.17208 | *PLA2G2F* |
| ENSG00000120057 | 10 | 99516508 | 99541709 | 74 | 10 | 31257 | 0.94585 | 0.17211 | *SFRP5* |
| ENSG00000124664 | 6 | 34495579 | 34534110 | 117 | 19 | 35432 | 0.94553 | 0.17219 | *SPDEF* |
| ENSG00000168461 | 18 | 9698162 | 9872548 | 763 | 152 | 44663 | 0.94526 | 0.17226 | *RAB31* |
| ENSG00000172940 | 3 | 38297303 | 38329806 | 58 | 16 | 38110 | 0.94519 | 0.17228 | *SLC22A13* |
| ENSG00000254415 | 19 | 52135806 | 52160054 | 76 | 16 | 45607 | 0.94495 | 0.17234 | *SIGLEC14* |
| ENSG00000159197 | 21 | 35726323 | 35753688 | 65 | 14 | 40429 | 0.94493 | 0.17235 | *KCNE2* |
| ENSG00000061273 | 12 | 48166505 | 48236915 | 203 | 44 | 52018 | 0.94468 | 0.17241 | *HDAC7* |
| ENSG00000187080 | 1 | 248118535 | 248139641 | 104 | 21 | 38734 | 0.94457 | 0.17244 | *OR2AK2* |
| ENSG00000142552 | 19 | 50020875 | 50060219 | 167 | 10 | 38817 | 0.94429 | 0.17251 | *RCN3* |
| ENSG00000162086 | 16 | 3345406 | 3378852 | 59 | 15 | 37960 | 0.94409 | 0.17256 | *ZNF75A* |
| ENSG00000272968 | 7 | 5013349 | 5122852 | 500 | 40 | 48353 | 0.94406 | 0.17257 | *RBAK-RBAKDN* |
| ENSG00000185420 | 1 | 245902642 | 246680614 | 3502 | 183 | 33456 | 0.94389 | 0.17261 | *SMYD3* |
| ENSG00000168884 | 4 | 2733375 | 2768103 | 159 | 28 | 45205 | 0.9438 | 0.17263 | *TNIP2* |
| ENSG00000119650 | 14 | 76358479 | 76560928 | 512 | 61 | 49225 | 0.94381 | 0.17263 | *IFT43* |
| ENSG00000008283 | 17 | 61499665 | 61533739 | 51 | 15 | 47851 | 0.9436 | 0.17269 | *CYB561* |
| ENSG00000150244 | 11 | 55019658 | 55048595 | 165 | 19 | 48590 | 0.94356 | 0.1727 | *TRIM48* |
| ENSG00000065883 | 7 | 39979636 | 40146733 | 360 | 36 | 33068 | 0.94333 | 0.17276 | *CDK13* |
| ENSG00000172716 | 17 | 33667324 | 33710720 | 150 | 30 | 34929 | 0.94329 | 0.17277 | *SLFN11* |
| ENSG00000134020 | 8 | 22560769 | 22867513 | 1064 | 171 | 47292 | 0.94302 | 0.17283 | *PEBP4* |
| ENSG00000070756 | 8 | 101688044 | 101745037 | 136 | 16 | 46863 | 0.94285 | 0.17288 | *PABPC1* |
| ENSG00000172824 | 16 | 67012492 | 67053661 | 42 | 14 | 46802 | 0.94282 | 0.17289 | *CES4A* |
| ENSG00000107984 | 10 | 54064056 | 54087802 | 59 | 12 | 32579 | 0.94267 | 0.17292 | *DKK1* |
| ENSG00000160113 | 19 | 17332692 | 17366749 | 169 | 31 | 44244 | 0.94258 | 0.17295 | *NR2F6* |
| ENSG00000065911 | 2 | 74415689 | 74454692 | 87 | 15 | 29609 | 0.94244 | 0.17298 | *MTHFD2* |
| ENSG00000153208 | 2 | 112646056 | 112797138 | 588 | 38 | 52837 | 0.94242 | 0.17299 | *MERTK* |
| ENSG00000011426 | 7 | 36419415 | 36503400 | 311 | 41 | 52446 | 0.94221 | 0.17304 | *ANLN* |

| | | | | | | | | | |
|---|---|---|---|---|---|---|---|---|---|
| ENSG00000152670 | 5 | 55023845 | 55122985 | 297 | 31 | 41348 | 0.94212 | 0.17307 | *DDX4* |
| ENSG00000205710 | 17 | 4792713 | 4816227 | 95 | 16 | 41032 | 0.94183 | 0.17314 | *C17orf107* |
| ENSG00000158104 | 12 | 122267433 | 122311502 | 120 | 17 | 42181 | 0.9416 | 0.1732 | *HPD* |
| ENSG00000139055 | 12 | 15056969 | 15102016 | 117 | 12 | 46212 | 0.94145 | 0.17324 | *ERP27* |
| ENSG00000178966 | 9 | 86585626 | 86628985 | 79 | 11 | 55599 | 0.94122 | 0.1733 | *RMI1* |
| ENSG00000254402 | 8 | 145737761 | 145762416 | 34 | 11 | 40866 | 0.94075 | 0.17342 | *LRRC24* |
| ENSG00000263620 | 17 | 8044134 | 8076293 | 78 | 15 | 45124 | 0.94065 | 0.17344 | *RP11-599B13.6* |
| ENSG00000183389 | 11 | 6013216 | 6034424 | 106 | 16 | 53330 | 0.94011 | 0.17358 | *OR56A4* |
| ENSG00000164307 | 5 | 96086521 | 96153803 | 368 | 35 | 46923 | 0.94002 | 0.1736 | *ERAP1* |
| ENSG00000116885 | 1 | 36871428 | 36926086 | 283 | 19 | 55561 | 0.93999 | 0.17361 | *OSCP1* |
| ENSG00000163026 | 2 | 24242210 | 24282445 | 106 | 20 | 50159 | 0.93982 | 0.17365 | *C2orf44* |
| ENSG00000118762 | 4 | 88918820 | 89008929 | 249 | 34 | 32740 | 0.93985 | 0.17365 | *PKD2* |
| ENSG00000111335 | 12 | 113406200 | 113459528 | 190 | 27 | 42890 | 0.93968 | 0.17369 | *OAS2* |
| ENSG00000143889 | 2 | 38779120 | 38840728 | 185 | 24 | 42976 | 0.93962 | 0.17371 | *HNRNPLL* |
| ENSG00000178075 | 3 | 113537029 | 113676021 | 318 | 56 | 48580 | 0.93956 | 0.17372 | *GRAMD1C* |
| ENSG00000149735 | 11 | 64691943 | 64713360 | 43 | 9 | 42374 | 0.93927 | 0.1738 | *GPHA2* |
| ENSG00000116678 | 1 | 65876248 | 66117242 | 756 | 54 | 33121 | 0.93887 | 0.1739 | *LEPR* |
| ENSG00000106665 | 7 | 73693805 | 73830273 | 274 | 40 | 36693 | 0.93886 | 0.1739 | *CLIP2* |
| ENSG00000112514 | 6 | 33374219 | 33396094 | 41 | 13 | 50090 | 0.93863 | 0.17396 | *CUTA* |
| ENSG00000095485 | 10 | 101982055 | 102037437 | 160 | 15 | 33682 | 0.93801 | 0.17412 | *CWF19L1* |
| ENSG00000153563 | 2 | 87001729 | 87045519 | 116 | 17 | 49655 | 0.93765 | 0.17421 | *CD8A* |
| ENSG00000150873 | 2 | 11263179 | 11296916 | 123 | 21 | 45248 | 0.93759 | 0.17423 | *C2orf50* |
| ENSG00000170681 | 9 | 103330361 | 103360188 | 112 | 16 | 46117 | 0.93751 | 0.17425 | *MURC* |
| ENSG00000205808 | 9 | 4652298 | 4675256 | 145 | 18 | 26016 | 0.93737 | 0.17428 | *PPAPDC2* |
| ENSG00000108370 | 17 | 63123549 | 63233821 | 332 | 42 | 33861 | 0.93717 | 0.17434 | *RGS9* |
| ENSG00000182732 | 14 | 72389156 | 73040654 | 2741 | 233 | 48196 | 0.93713 | 0.17435 | *RGS6* |
| ENSG00000267889 | 2 | 203094292 | 203114435 | 47 | 11 | 52634 | 0.93699 | 0.17438 | *AC079354.2* |
| ENSG00000198901 | 15 | 91499270 | 91548859 | 169 | 22 | 34411 | 0.93701 | 0.17438 | *PRC1* |
| ENSG00000178764 | 8 | 123783633 | 123996750 | 541 | 132 | 47972 | 0.9368 | 0.17443 | *ZHX2* |
| ENSG00000188707 | 7 | 150016938 | 150039808 | 63 | 6 | 51268 | 0.93663 | 0.17448 | *ZBED6CL* |
| ENSG00000183617 | 19 | 3752662 | 3778573 | 93 | 24 | 49500 | 0.9366 | 0.17448 | *MRPL54* |
| ENSG00000113249 | 5 | 156446424 | 156496130 | 216 | 33 | 45285 | 0.93638 | 0.17454 | *HAVCR1* |
| ENSG00000205209 | 19 | 35074234 | 35176604 | 512 | 47 | 50372 | 0.93636 | 0.17454 | *SCGB2B2* |
| ENSG00000168116 | 6 | 56901347 | 56930023 | 40 | 8 | 51469 | 0.93625 | 0.17457 | *KIAA1586* |
| ENSG00000149507 | 11 | 59797748 | 59825517 | 73 | 18 | 51816 | 0.93627 | 0.17457 | *OOSP2* |
| ENSG00000159720 | 16 | 67461917 | 67525140 | 79 | 20 | 46121 | 0.93622 | 0.17458 | *ATP6V0D1* |

| | | | | | | | | | |
|---|---|---|---|---|---|---|---|---|---|
| ENSG00000214274 | 14 | 21142336 | 21177130 | 156 | 32 | 50246 | 0.93595 | 0.17465 | *ANG* |
| ENSG00000135378 | 11 | 32841489 | 32889669 | 175 | 20 | 55416 | 0.93559 | 0.17474 | *PRRG4* |
| ENSG00000197386 | 4 | 3066408 | 3255676 | 480 | 40 | 39029 | 0.93518 | 0.17485 | *HTT* |
| ENSG00000165802 | 9 | 140332022 | 140363786 | 41 | 20 | 47019 | 0.93509 | 0.17487 | *NSMF* |
| ENSG00000100462 | 14 | 23379720 | 23408794 | 102 | 10 | 43576 | 0.93511 | 0.17487 | *PRMT5* |
| ENSG00000205937 | 16 | 2293117 | 2328413 | 67 | 13 | 45827 | 0.93501 | 0.17489 | *RNPS1* |
| ENSG00000166173 | 15 | 71113863 | 71156498 | 92 | 15 | 47859 | 0.93455 | 0.17501 | *LARP6* |
| ENSG00000162373 | 1 | 49183195 | 49252641 | 124 | 24 | 31909 | 0.93447 | 0.17503 | *BEND5* |
| ENSG00000048140 | 5 | 176064388 | 176096058 | 84 | 18 | 31565 | 0.93438 | 0.17505 | *TSPAN17* |
| ENSG00000171459 | 9 | 125502019 | 125523062 | 27 | 10 | 43171 | 0.93423 | 0.17509 | *OR1L6* |
| ENSG00000111671 | 12 | 6970099 | 7008522 | 81 | 8 | 28738 | 0.93422 | 0.1751 | *SPSB2* |
| ENSG00000178773 | 16 | 89632176 | 89673654 | 168 | 37 | 48215 | 0.93394 | 0.17517 | *CPNE7* |
| ENSG00000270033 | 17 | 60339848 | 60370276 | 119 | 18 | 42911 | 0.93384 | 0.17519 | *RP11-51L5.7* |
| ENSG00000198919 | 3 | 108298529 | 108423693 | 185 | 20 | 47192 | 0.93373 | 0.17522 | *DZIP3* |
| ENSG00000197965 | 1 | 167680429 | 167771156 | 304 | 43 | 51056 | 0.9331 | 0.17538 | *MPZL1* |
| ENSG00000215504 | 11 | 85553600 | 85575986 | 79 | 9 | 54060 | 0.93311 | 0.17538 | *AP000974.1* |
| ENSG00000115386 | 2 | 79337488 | 79360545 | 91 | 20 | 47568 | 0.93302 | 0.1754 | *REG1A* |
| ENSG00000140853 | 16 | 57013397 | 57127443 | 414 | 111 | 47534 | 0.93305 | 0.1754 | *NLRC5* |
| ENSG00000226761 | 12 | 11203964 | 11224893 | 71 | 14 | 37710 | 0.93292 | 0.17543 | *TAS2R46* |
| ENSG00000273398 | 2 | 68348370 | 68498362 | 345 | 23 | 51165 | 0.93277 | 0.17547 | *RP11-474G23.1* |
| ENSG00000163705 | 3 | 10113001 | 10159915 | 162 | 9 | 41309 | 0.93276 | 0.17547 | *FANCD2OS* |
| ENSG00000148358 | 9 | 132805705 | 132912448 | 445 | 53 | 48557 | 0.93268 | 0.17549 | *GPR107* |
| ENSG00000151838 | 14 | 59961257 | 60053549 | 421 | 26 | 43467 | 0.93236 | 0.17558 | *CCDC175* |
| ENSG00000163467 | 1 | 156297105 | 156326786 | 64 | 10 | 53896 | 0.93214 | 0.17563 | *TSACC* |
| ENSG00000110075 | 11 | 68218186 | 68392802 | 452 | 31 | 49517 | 0.93203 | 0.17566 | *PPP6R3* |
| ENSG00000127666 | 19 | 4805944 | 4841716 | 155 | 24 | 42834 | 0.93174 | 0.17574 | *TICAM1* |
| ENSG00000166828 | 16 | 23184036 | 23238204 | 242 | 20 | 54485 | 0.93168 | 0.17575 | *SCNN1G* |
| ENSG00000161281 | 19 | 36631824 | 36653771 | 94 | 24 | 47523 | 0.93148 | 0.1758 | *COX7A1* |
| ENSG00000188655 | 14 | 21014252 | 21039090 | 73 | 21 | 38010 | 0.93142 | 0.17582 | *RNASE9* |
| ENSG00000137161 | 6 | 42886938 | 42917025 | 85 | 19 | 43460 | 0.93138 | 0.17583 | *CNPY3* |
| ENSG00000064687 | 19 | 1030102 | 1075571 | 230 | 42 | 45367 | 0.93124 | 0.17586 | *ABCA7* |
| ENSG00000175877 | 7 | 73265489 | 73290223 | 57 | 9 | 31033 | 0.93113 | 0.17589 | *WBSCR28* |
| ENSG00000105819 | 7 | 102927869 | 102979958 | 90 | 22 | 50103 | 0.93071 | 0.176 | *PMPCB* |
| ENSG00000100325 | 22 | 30174597 | 30244271 | 196 | 27 | 39768 | 0.93065 | 0.17602 | *ASCC2* |
| ENSG00000111224 | 12 | 3890213 | 3992608 | 350 | 41 | 40718 | 0.93055 | 0.17604 | *PARP11* |
| ENSG00000067606 | 1 | 1971909 | 2126834 | 481 | 55 | 43439 | 0.93025 | 0.17612 | *PRKCZ* |

| | | | | | | | | | |
|---|---|---|---|---|---|---|---|---|---|
| ENSG00000174669 | 11 | 66119992 | 66149685 | 60 | 8 | 37149 | 0.9298 | 0.17624 | *SLC29A2* |
| ENSG00000173389 | 3 | 51918892 | 51947351 | 51 | 11 | 42247 | 0.92972 | 0.17626 | *IQCF1* |
| ENSG00000179284 | 19 | 13065973 | 13095567 | 44 | 20 | 39443 | 0.92933 | 0.17636 | *DAND5* |
| ENSG00000162763 | 1 | 165503244 | 165543198 | 194 | 22 | 37580 | 0.92886 | 0.17648 | *LRRC52* |
| ENSG00000079785 | 2 | 15721302 | 15781235 | 215 | 18 | 37523 | 0.92874 | 0.17651 | *DDX1* |
| ENSG00000110906 | 12 | 109876461 | 109925349 | 221 | 24 | 50471 | 0.92872 | 0.17652 | *KCTD10* |
| ENSG00000204291 | 9 | 101695461 | 101843069 | 414 | 60 | 41953 | 0.92841 | 0.1766 | *COL15A1* |
| ENSG00000117289 | 1 | 145428469 | 145452635 | 15 | 9 | 44098 | 0.92807 | 0.17669 | *TXNIP* |
| ENSG00000105996 | 7 | 27129721 | 27152430 | 51 | 13 | 41929 | 0.92802 | 0.1767 | *HOXA2* |
| ENSG00000178919 | 9 | 100605536 | 100628986 | 103 | 8 | 36509 | 0.92788 | 0.17674 | *FOXE1* |
| ENSG00000060971 | 3 | 38134620 | 38188733 | 100 | 24 | 42116 | 0.92716 | 0.17692 | *ACAA1* |
| ENSG00000167173 | 15 | 75477984 | 75514510 | 41 | 16 | 40737 | 0.927 | 0.17696 | *C15orf39* |
| ENSG00000213625 | 1 | 65876270 | 65911690 | 81 | 19 | 38786 | 0.92691 | 0.17699 | *LEPROT* |
| ENSG00000124104 | 20 | 44452449 | 44481914 | 88 | 19 | 44143 | 0.9266 | 0.17707 | *SNX21* |
| ENSG00000184566 | 11 | 33892189 | 33913338 | 65 | 18 | 53660 | 0.92635 | 0.17713 | *AC132216.1* |
| ENSG00000169062 | 13 | 115037059 | 115081283 | 127 | 24 | 35401 | 0.92611 | 0.17719 | *UPF3A* |
| ENSG00000114648 | 3 | 47314407 | 47398306 | 96 | 20 | 46317 | 0.92607 | 0.1772 | *KLHL18* |
| ENSG00000181518 | 11 | 123767109 | 123788183 | 95 | 15 | 55602 | 0.92555 | 0.17734 | *OR8D4* |
| ENSG00000105971 | 7 | 115917434 | 116158595 | 770 | 70 | 52350 | 0.92545 | 0.17737 | *CAV2* |
| ENSG00000106868 | 9 | 114793065 | 114947688 | 438 | 69 | 48373 | 0.92505 | 0.17747 | *SUSD1* |
| ENSG00000182902 | 22 | 18033139 | 18083760 | 141 | 37 | 39379 | 0.92477 | 0.17754 | *SLC25A18* |
| ENSG00000268170 | 7 | 142909163 | 142929360 | 22 | 7 | 44162 | 0.92472 | 0.17756 | *AC073342.1* |
| ENSG00000257987 | 12 | 49111218 | 49169569 | 129 | 23 | 41842 | 0.92364 | 0.17784 | *LINC00935* |
| ENSG00000136527 | 3 | 185623694 | 185665924 | 93 | 16 | 38547 | 0.92342 | 0.17789 | *TRA2B* |
| ENSG00000206052 | 18 | 67058291 | 67526323 | 1866 | 224 | 48429 | 0.92343 | 0.17789 | *DOK6* |
| ENSG00000072840 | 4 | 5702924 | 5840772 | 780 | 124 | 48611 | 0.9234 | 0.1779 | *EVC* |
| ENSG00000173175 | 3 | 122991143 | 123178605 | 417 | 47 | 40695 | 0.92324 | 0.17794 | *ADCY5* |
| ENSG00000122783 | 7 | 134767115 | 134865547 | 307 | 15 | 52668 | 0.92289 | 0.17803 | *C7orf49* |
| ENSG00000242515 | 2 | 234535100 | 234691951 | 584 | 56 | 46382 | 0.92279 | 0.17806 | *UGT1A10* |
| ENSG00000061656 | 20 | 34193814 | 34218971 | 68 | 12 | 35173 | 0.92259 | 0.17811 | *SPAG4* |
| ENSG00000125787 | 20 | 3014268 | 3036393 | 55 | 12 | 47671 | 0.92252 | 0.17813 | *GNRH2* |
| ENSG00000005471 | 7 | 87021013 | 87119751 | 267 | 25 | 35969 | 0.92235 | 0.17817 | *ABCB4* |
| ENSG00000172081 | 19 | 2061037 | 2106672 | 121 | 34 | 49232 | 0.92228 | 0.17819 | *MOB3A* |
| ENSG00000189159 | 17 | 73121343 | 73174376 | 122 | 22 | 38187 | 0.92227 | 0.1782 | *HN1* |
| ENSG00000197106 | 1 | 110683108 | 110754824 | 160 | 23 | 31091 | 0.92207 | 0.17825 | *SLC6A17* |
| ENSG00000111145 | 12 | 96578160 | 96673613 | 282 | 55 | 49359 | 0.92194 | 0.17828 | *ELK3* |

| | | | | | | | | | |
|---|---|---|---|---|---|---|---|---|---|
| ENSG00000167085 | 17 | 47471414 | 47502246 | 92 | 24 | 39778 | 0.92188 | 0.1783 | *PHB* |
| ENSG00000173825 | 11 | 65112238 | 65135084 | 32 | 10 | 39255 | 0.92179 | 0.17832 | *TIGD3* |
| ENSG00000122877 | 10 | 64561756 | 64689660 | 439 | 45 | 42671 | 0.9215 | 0.1784 | *EGR2* |
| ENSG00000196072 | 10 | 102023713 | 102056469 | 95 | 14 | 38810 | 0.92122 | 0.17847 | *BLOC1S2* |
| ENSG00000204421 | 6 | 31676425 | 31699622 | 52 | 14 | 27143 | 0.92081 | 0.17857 | *LY6G6C* |
| ENSG00000269415 | 3 | 59947215 | 59967583 | 125 | 22 | 41403 | 0.92068 | 0.17861 | *NPCDR1* |
| ENSG00000114388 | 3 | 50374761 | 50398522 | 14 | 6 | 53339 | 0.92063 | 0.17862 | *NPRL2* |
| ENSG00000168734 | 20 | 43150426 | 43262888 | 243 | 29 | 45115 | 0.92036 | 0.17869 | *PKIG* |
| ENSG00000181788 | 3 | 150448914 | 150491264 | 54 | 15 | 33218 | 0.92025 | 0.17872 | *SIAH2* |
| ENSG00000108262 | 17 | 27890487 | 27931072 | 71 | 19 | 42855 | 0.92027 | 0.17872 | *GIT1* |
| ENSG00000163634 | 3 | 63809546 | 63859579 | 84 | 19 | 52686 | 0.92001 | 0.17878 | *THOC7* |
| ENSG00000204021 | 10 | 90474301 | 90522543 | 246 | 14 | 42965 | 0.91988 | 0.17882 | *LIPK* |
| ENSG00000106526 | 7 | 149931005 | 150030814 | 289 | 33 | 32079 | 0.91966 | 0.17888 | *ACTR3C* |
| ENSG00000182957 | 13 | 24543944 | 24891212 | 1588 | 217 | 43415 | 0.91939 | 0.17895 | *SPATA13* |
| ENSG00000133641 | 12 | 88417623 | 88453937 | 39 | 12 | 48856 | 0.91909 | 0.17902 | *C12orf29* |
| ENSG00000143641 | 1 | 230183536 | 230427870 | 954 | 132 | 41457 | 0.91903 | 0.17904 | *GALNT2* |
| ENSG00000112078 | 6 | 36400544 | 36468920 | 185 | 18 | 35520 | 0.91895 | 0.17906 | *KCTD20* |
| ENSG00000116922 | 1 | 38137242 | 38167914 | 90 | 14 | 45249 | 0.9189 | 0.17907 | *C1orf109* |
| ENSG00000132554 | 8 | 100963164 | 101153496 | 379 | 49 | 44231 | 0.91856 | 0.17916 | *RGS22* |
| ENSG00000184209 | 12 | 123932188 | 123967701 | 67 | 15 | 47185 | 0.91858 | 0.17916 | *SNRNP35* |
| ENSG00000164651 | 7 | 20813906 | 20836505 | 28 | 10 | 53253 | 0.91846 | 0.17919 | *SP8* |
| ENSG00000088256 | 19 | 3084408 | 3134002 | 208 | 37 | 43619 | 0.91847 | 0.17919 | *GNA11* |
| ENSG00000142065 | 19 | 36817162 | 36880101 | 171 | 17 | 54398 | 0.91844 | 0.17919 | *ZFP14* |
| ENSG00000114805 | 3 | 155083369 | 155472856 | 1224 | 75 | 33994 | 0.91839 | 0.17921 | *PLCH1* |
| ENSG00000178226 | 16 | 31140246 | 31171415 | 41 | 15 | 31968 | 0.91806 | 0.17929 | *PRSS36* |
| ENSG00000163904 | 3 | 185290284 | 185361339 | 215 | 15 | 43004 | 0.91805 | 0.1793 | *SENP2* |
| ENSG00000168404 | 16 | 74695753 | 74744858 | 208 | 40 | 48233 | 0.91803 | 0.1793 | *MLKL* |
| ENSG00000164292 | 5 | 95039226 | 95170087 | 357 | 60 | 49166 | 0.91789 | 0.17934 | *RHOBTB3* |
| ENSG00000185955 | 7 | 100044238 | 100071894 | 41 | 10 | 50797 | 0.91778 | 0.17937 | *C7orf61* |
| ENSG00000183230 | 10 | 67662276 | 69465927 | 7353 | 329 | 40226 | 0.91756 | 0.17943 | *CTNNA3* |
| ENSG00000273025 | 15 | 72549087 | 72622287 | 138 | 24 | 44925 | 0.91698 | 0.17958 | *CELF6* |
| ENSG00000167634 | 19 | 55424877 | 55487680 | 316 | 57 | 46043 | 0.91688 | 0.1796 | *NLRP7* |
| ENSG00000168002 | 11 | 62519016 | 62544182 | 43 | 10 | 35591 | 0.91657 | 0.17968 | *POLR2G* |
| ENSG00000143194 | 1 | 166948346 | 167001451 | 150 | 19 | 42603 | 0.91646 | 0.17971 | *MAEL* |
| ENSG00000163098 | 19 | 53782856 | 53804875 | 141 | 29 | 43300 | 0.91637 | 0.17974 | *BIRC8* |
| ENSG00000163006 | 2 | 109393213 | 109511933 | 276 | 23 | 43010 | 0.91593 | 0.17985 | *CCDC138* |

| | | | | | | | | | |
|---|---|---|---|---|---|---|---|---|---|
| ENSG00000136051 | 12 | 105491102 | 105572912 | 264 | 27 | 49045 | 0.91555 | 0.17995 | *KIAA1033* |
| ENSG00000123297 | 12 | 58166372 | 58211854 | 68 | 9 | 32920 | 0.91537 | 0.18 | *TSFM* |
| ENSG00000129214 | 17 | 7507382 | 7546700 | 122 | 17 | 35293 | 0.91524 | 0.18003 | *SHBG* |
| ENSG00000170537 | 16 | 18985256 | 19085264 | 303 | 27 | 37200 | 0.91457 | 0.18021 | *TMC7* |
| ENSG00000196843 | 2 | 97192480 | 97228375 | 60 | 20 | 34425 | 0.91452 | 0.18022 | *ARID5A* |
| ENSG00000186474 | 19 | 51522348 | 51548486 | 48 | 12 | 43454 | 0.91453 | 0.18022 | *KLK12* |
| ENSG00000135931 | 2 | 232053260 | 232249548 | 447 | 56 | 41532 | 0.91429 | 0.18028 | *ARMC9* |
| ENSG00000204576 | 6 | 30514663 | 30541500 | 47 | 10 | 48757 | 0.91362 | 0.18046 | *PRR3* |
| ENSG00000115694 | 2 | 242422089 | 242459145 | 108 | 20 | 53031 | 0.91346 | 0.1805 | *STK25* |
| ENSG00000008018 | 6 | 170834205 | 170872429 | 121 | 14 | 52565 | 0.91321 | 0.18057 | *PSMB1* |
| ENSG00000221954 | 11 | 49993009 | 50014071 | 78 | 13 | 41239 | 0.91314 | 0.18058 | *OR4C12* |
| ENSG00000131558 | 7 | 132927829 | 133761342 | 1653 | 82 | 46598 | 0.91295 | 0.18063 | *EXOC4* |
| ENSG00000174944 | 3 | 150919905 | 151006255 | 265 | 36 | 50520 | 0.91261 | 0.18072 | *P2RY14* |
| ENSG00000071994 | 6 | 170874383 | 170903780 | 81 | 10 | 55228 | 0.91246 | 0.18076 | *PDCD2* |
| ENSG00000170100 | 16 | 89274118 | 89305363 | 171 | 15 | 24919 | 0.91246 | 0.18076 | *ZNF778* |
| ENSG00000089050 | 20 | 18457184 | 18487887 | 118 | 19 | 40196 | 0.91227 | 0.18081 | *RBBP9* |
| ENSG00000198113 | 9 | 140162201 | 140187093 | 46 | 20 | 42741 | 0.91226 | 0.18082 | *TOR4A* |
| ENSG00000231213 | 3 | 146284342 | 146334003 | 338 | 28 | 56432 | 0.91209 | 0.18086 | *PLSCR5* |
| ENSG00000179636 | 14 | 21482268 | 21514435 | 109 | 39 | 45707 | 0.91205 | 0.18087 | *TPPP2* |
| ENSG00000137802 | 15 | 42056632 | 42130053 | 149 | 21 | 45775 | 0.91188 | 0.18092 | *MAPKBP1* |
| ENSG00000204428 | 6 | 31634461 | 31661817 | 64 | 16 | 36848 | 0.91167 | 0.18097 | *LY6G5C* |
| ENSG00000164694 | 6 | 159580429 | 159703141 | 480 | 31 | 34724 | 0.91168 | 0.18097 | *FNDC1* |
| ENSG00000198130 | 2 | 191044461 | 191218919 | 544 | 25 | 49067 | 0.91149 | 0.18102 | *HIBCH* |
| ENSG00000077080 | 7 | 100230720 | 100264084 | 58 | 15 | 52974 | 0.91144 | 0.18103 | *ACTL6B* |
| ENSG00000164167 | 4 | 147086837 | 147131152 | 152 | 16 | 36512 | 0.91138 | 0.18105 | *LSM6* |
| ENSG00000179403 | 1 | 1360241 | 1388262 | 48 | 15 | 41284 | 0.91134 | 0.18106 | *VWA1* |
| ENSG00000268193 | 19 | 19007822 | 19055206 | 103 | 23 | 40055 | 0.91134 | 0.18106 | *AC002985.3* |
| ENSG00000173947 | 1 | 111878910 | 111905635 | 157 | 25 | 47015 | 0.91116 | 0.18111 | *PIFO* |
| ENSG00000082996 | 3 | 149520495 | 149689926 | 405 | 39 | 51397 | 0.91081 | 0.1812 | *RNF13* |
| ENSG00000244122 | 2 | 234580584 | 234691945 | 472 | 42 | 45188 | 0.91027 | 0.18134 | *UGT1A7* |
| ENSG00000254221 | 5 | 140719828 | 140902546 | 506 | 57 | 43498 | 0.91002 | 0.18141 | *PCDHGB1* |
| ENSG00000198586 | 2 | 171837333 | 172097824 | 578 | 35 | 31602 | 0.90988 | 0.18144 | *TLK1* |
| ENSG00000265118 | 17 | 29622231 | 29655849 | 57 | 6 | 28454 | 0.90989 | 0.18144 | *CTD-2370N5.3* |
| ENSG00000133392 | 16 | 15787029 | 15960890 | 726 | 98 | 46489 | 0.90897 | 0.18168 | *MYH11* |
| ENSG00000166135 | 10 | 102278829 | 102329755 | 139 | 8 | 55643 | 0.90888 | 0.18171 | *HIF1AN* |
| ENSG00000173214 | 6 | 111570551 | 111602370 | 87 | 18 | 44504 | 0.90847 | 0.18181 | *KIAA1919* |

| | | | | | | | | | |
|---|---|---|---|---|---|---|---|---|---|
| ENSG00000152782 | 10 | 91332745 | 91415215 | 350 | 40 | 37357 | 0.9084 | 0.18183 | *PANK1* |
| ENSG00000177143 | 18 | 570367 | 591524 | 111 | 25 | 51870 | 0.90832 | 0.18185 | *CETN1* |
| ENSG00000110844 | 12 | 49952001 | 50048449 | 144 | 20 | 40438 | 0.90803 | 0.18193 | *PRPF40B* |
| ENSG00000127418 | 4 | 993724 | 1030685 | 161 | 31 | 49913 | 0.90789 | 0.18197 | *FGFRL1* |
| ENSG00000168484 | 8 | 22004426 | 22031992 | 79 | 26 | 50981 | 0.9079 | 0.18197 | *SFTPC* |
| ENSG00000174456 | 12 | 110455872 | 110521491 | 182 | 29 | 41467 | 0.90782 | 0.18199 | *C12orf76* |
| ENSG00000178235 | 13 | 84441344 | 84466528 | 41 | 10 | 50404 | 0.90765 | 0.18203 | *SLITRK1* |
| ENSG00000123500 | 6 | 116430086 | 116489910 | 147 | 25 | 45578 | 0.90761 | 0.18204 | *COL10A1* |
| ENSG00000155660 | 7 | 148690154 | 148735733 | 167 | 20 | 35561 | 0.90739 | 0.1821 | *PDIA4* |
| ENSG00000140545 | 15 | 89431916 | 89466642 | 160 | 26 | 48117 | 0.90732 | 0.18212 | *MFGE8* |
| ENSG00000052795 | 4 | 159680290 | 159839201 | 261 | 23 | 49461 | 0.90674 | 0.18227 | *FNIP2* |
| ENSG00000130669 | 19 | 39606410 | 39683456 | 254 | 34 | 53239 | 0.90668 | 0.18229 | *PAK4* |
| ENSG00000052344 | 16 | 31132756 | 31157083 | 35 | 10 | 32004 | 0.90649 | 0.18234 | *PRSS8* |
| ENSG00000165646 | 10 | 118990604 | 119048941 | 165 | 29 | 37508 | 0.90643 | 0.18235 | *SLC18A2* |
| ENSG00000171219 | 11 | 64580859 | 64622041 | 88 | 19 | 35377 | 0.90637 | 0.18237 | *CDC42BPG* |
| ENSG00000188334 | 19 | 48461303 | 48505427 | 195 | 30 | 45819 | 0.90626 | 0.1824 | *BSPH1* |
| ENSG00000168569 | 11 | 62529101 | 62569493 | 62 | 13 | 35056 | 0.90622 | 0.18241 | *TMEM223* |
| ENSG00000153230 | 1 | 247891917 | 247912861 | 58 | 9 | 43546 | 0.90617 | 0.18242 | *OR14K1* |
| ENSG00000169435 | 4 | 74427267 | 74496348 | 189 | 18 | 51836 | 0.90608 | 0.18245 | *RASSF6* |
| ENSG00000132855 | 1 | 63053158 | 63081830 | 28 | 6 | 39404 | 0.90604 | 0.18246 | *ANGPTL3* |
| ENSG00000179397 | 1 | 244607679 | 244814479 | 693 | 49 | 34401 | 0.90592 | 0.18249 | *C1orf101* |
| ENSG00000165113 | 9 | 86344336 | 86454431 | 271 | 15 | 52189 | 0.9059 | 0.18249 | *GKAP1* |
| ENSG00000168209 | 10 | 74023678 | 74045794 | 51 | 9 | 26385 | 0.90593 | 0.18249 | *DDIT4* |
| ENSG00000150261 | 11 | 56103421 | 56124507 | 84 | 11 | 54846 | 0.90581 | 0.18252 | *OR8K1* |
| ENSG00000175445 | 8 | 19749228 | 19834769 | 369 | 39 | 40551 | 0.90557 | 0.18258 | *LPL* |
| ENSG00000119953 | 10 | 112040488 | 112074709 | 93 | 17 | 39568 | 0.90523 | 0.18267 | *SMNDC1* |
| ENSG00000100918 | 14 | 24631062 | 24659463 | 57 | 13 | 47027 | 0.90496 | 0.18274 | *REC8* |
| ENSG00000101150 | 20 | 62486596 | 62532898 | 127 | 19 | 34472 | 0.90497 | 0.18274 | *TPD52L2* |
| ENSG00000175604 | 16 | 12170603 | 12194159 | 111 | 15 | 49106 | 0.90481 | 0.18278 | *RP11-276H1.3* |
| ENSG00000156171 | 1 | 111649955 | 111692838 | 102 | 11 | 32779 | 0.90474 | 0.1828 | *DRAM2* |
| ENSG00000198830 | 1 | 26788941 | 26812463 | 53 | 13 | 37612 | 0.9046 | 0.18284 | *HMGN2* |
| ENSG00000167434 | 17 | 58217297 | 58258260 | 71 | 21 | 37291 | 0.90459 | 0.18284 | *CA4* |
| ENSG00000080511 | 19 | 10113925 | 10142955 | 166 | 35 | 51679 | 0.9045 | 0.18286 | *RDH8* |
| ENSG00000162777 | 1 | 111719796 | 111757157 | 89 | 28 | 50831 | 0.90446 | 0.18288 | *DENND2D* |
| ENSG00000218336 | 4 | 183055140 | 183734177 | 2700 | 311 | 47042 | 0.90442 | 0.18289 | *TENM3* |
| ENSG00000188385 | 10 | 133908175 | 134008313 | 627 | 79 | 47135 | 0.9042 | 0.18294 | *JAKMIP3* |

| | | | | | | | | | |
|---|---|---|---|---|---|---|---|---|---|
| ENSG00000163630 | 3 | 63203991 | 63612597 | 1723 | 179 | 47648 | 0.90408 | 0.18298 | *SYNPR* |
| ENSG00000106460 | 7 | 12240867 | 12292993 | 322 | 20 | 36175 | 0.90399 | 0.183 | *TMEM106B* |
| ENSG00000112159 | 6 | 90342218 | 90539442 | 493 | 46 | 53783 | 0.90378 | 0.18306 | *MDN1* |
| ENSG00000116406 | 1 | 184649365 | 184734047 | 194 | 23 | 36331 | 0.90363 | 0.1831 | *EDEM3* |
| ENSG00000099204 | 10 | 116180872 | 116454762 | 848 | 107 | 39586 | 0.90362 | 0.1831 | *ABLIM1* |
| ENSG00000174807 | 11 | 66071958 | 66094515 | 52 | 14 | 38196 | 0.90305 | 0.18325 | *CD248* |
| ENSG00000125285 | 13 | 95351886 | 95374389 | 42 | 12 | 51594 | 0.90302 | 0.18326 | *SOX21* |
| ENSG00000204711 | 9 | 72425709 | 72531148 | 320 | 44 | 51914 | 0.90275 | 0.18333 | *C9orf135* |
| ENSG00000146678 | 7 | 45917956 | 45943267 | 73 | 15 | 50939 | 0.90271 | 0.18334 | *IGFBP1* |
| ENSG00000172116 | 2 | 87032462 | 87099047 | 188 | 25 | 48098 | 0.9025 | 0.1834 | *CD8B* |
| ENSG00000266733 | 17 | 28874130 | 28900511 | 44 | 12 | 50397 | 0.90219 | 0.18348 | *TBC1D29* |
| ENSG00000160678 | 1 | 153590402 | 153614513 | 61 | 9 | 50412 | 0.90208 | 0.18351 | *S100A1* |
| ENSG00000149115 | 11 | 57057112 | 57102426 | 131 | 17 | 51251 | 0.90172 | 0.1836 | *TNKS1BP1* |
| ENSG00000088367 | 20 | 34669426 | 34830721 | 180 | 26 | 37009 | 0.90168 | 0.18361 | *EPB41L1* |
| ENSG00000099949 | 22 | 21323751 | 21363327 | 142 | 35 | 44603 | 0.9016 | 0.18363 | *LZTR1* |
| ENSG00000263711 | 18 | 70808476 | 70941715 | 381 | 45 | 34000 | 0.90145 | 0.18367 | *RP11-169F17.1* |
| ENSG00000124098 | 20 | 54923971 | 54953719 | 91 | 14 | 37726 | 0.90147 | 0.18367 | *FAM210B* |
| ENSG00000117505 | 1 | 93801445 | 93845055 | 103 | 13 | 39280 | 0.90133 | 0.18371 | *DR1* |
| ENSG00000172640 | 12 | 48586081 | 48607170 | 147 | 10 | 51464 | 0.90121 | 0.18374 | *OR10AD1* |
| ENSG00000168148 | 1 | 228602546 | 228623026 | 48 | 11 | 42651 | 0.90114 | 0.18376 | *HIST3H3* |
| ENSG00000135241 | 7 | 108100866 | 108220110 | 358 | 35 | 41970 | 0.901 | 0.18379 | *PNPLA8* |
| ENSG00000009844 | 6 | 142458367 | 142555826 | 206 | 17 | 39203 | 0.9007 | 0.18387 | *VTA1* |
| ENSG00000255093 | 11 | 111312064 | 111338876 | 74 | 19 | 50335 | 0.90065 | 0.18389 | *RP11-794P6.2* |
| ENSG00000119048 | 5 | 133696870 | 133737683 | 95 | 8 | 37867 | 0.90052 | 0.18392 | *UBE2B* |
| ENSG00000110719 | 11 | 67796483 | 67828362 | 75 | 22 | 52245 | 0.90046 | 0.18394 | *TCIRG1* |
| ENSG00000213047 | 1 | 197463878 | 197754826 | 424 | 34 | 51468 | 0.90023 | 0.184 | *DENND1B* |
| ENSG00000068971 | 11 | 64675025 | 64711945 | 78 | 13 | 42149 | 0.90009 | 0.18404 | *PPP2R5B* |
| ENSG00000182348 | 7 | 88378682 | 88976346 | 2326 | 158 | 44505 | 0.89978 | 0.18412 | *ZNF804B* |
| ENSG00000197483 | 19 | 55977699 | 56005854 | 118 | 30 | 47222 | 0.89975 | 0.18413 | *ZNF628* |
| ENSG00000140506 | 15 | 75095057 | 75128099 | 73 | 13 | 30200 | 0.89951 | 0.18419 | *LMAN1L* |
| ENSG00000111229 | 12 | 110862630 | 110898227 | 94 | 13 | 35820 | 0.89938 | 0.18423 | *ARPC3* |
| ENSG00000105821 | 7 | 102942921 | 102995320 | 92 | 23 | 49250 | 0.899 | 0.18433 | *DNAJC2* |
| ENSG00000234224 | 7 | 123660973 | 123683523 | 72 | 9 | 46072 | 0.89851 | 0.18446 | *TMEM229A* |
| ENSG00000180806 | 12 | 54378679 | 54407121 | 52 | 15 | 47846 | 0.89822 | 0.18453 | *HOXC9* |
| ENSG00000164530 | 6 | 36912209 | 36942613 | 133 | 21 | 50506 | 0.89764 | 0.18469 | *PI16* |
| ENSG00000105426 | 19 | 5148506 | 5350814 | 797 | 148 | 47341 | 0.8976 | 0.1847 | *PTPRS* |

| | | | | | | | | | |
|---|---|---|---|---|---|---|---|---|---|
| ENSG00000187905 | 22 | 21390249 | 21428457 | 131 | 39 | 46593 | 0.89757 | 0.18471 | *AC002472.13* |
| ENSG00000123146 | 19 | 14481313 | 14529537 | 85 | 35 | 44865 | 0.89748 | 0.18473 | *CD97* |
| ENSG00000055332 | 2 | 37316353 | 37394208 | 306 | 31 | 50666 | 0.89745 | 0.18474 | *EIF2AK2* |
| ENSG00000165264 | 9 | 32542997 | 32583160 | 89 | 17 | 50815 | 0.89716 | 0.18482 | *NDUFB6* |
| ENSG00000163808 | 3 | 44793209 | 44924868 | 190 | 27 | 40672 | 0.89704 | 0.18485 | *KIF15* |
| ENSG00000078487 | 7 | 99988449 | 100036615 | 79 | 10 | 51962 | 0.897 | 0.18486 | *ZCWPW1* |
| ENSG00000108556 | 17 | 4791069 | 4816369 | 99 | 17 | 39900 | 0.8968 | 0.18491 | *CHRNE* |
| ENSG00000173821 | 17 | 78224665 | 78382586 | 558 | 86 | 41850 | 0.8965 | 0.18499 | *RNF213* |
| ENSG00000167528 | 12 | 48723791 | 48755197 | 78 | 15 | 41261 | 0.89646 | 0.185 | *ZNF641* |
| ENSG00000131097 | 17 | 42913721 | 42937848 | 76 | 15 | 49293 | 0.89629 | 0.18505 | *HIGD1B* |
| ENSG00000146250 | 6 | 84212194 | 84245423 | 113 | 22 | 44899 | 0.89585 | 0.18517 | *PRSS35* |
| ENSG00000166136 | 10 | 102257203 | 102299757 | 100 | 5 | 56685 | 0.89558 | 0.18524 | *NDUFB8* |
| ENSG00000100228 | 22 | 23477513 | 23516537 | 132 | 14 | 52441 | 0.89554 | 0.18525 | *RAB36* |
| ENSG00000135045 | 9 | 77551497 | 77577802 | 62 | 14 | 41818 | 0.89551 | 0.18526 | *C9orf40* |
| ENSG00000163113 | 1 | 149899705 | 149992625 | 139 | 18 | 47354 | 0.89525 | 0.18533 | *OTUD7B* |
| ENSG00000213024 | 19 | 50400082 | 50443020 | 149 | 29 | 42082 | 0.89523 | 0.18533 | *NUP62* |
| ENSG00000197465 | 4 | 144782020 | 144836716 | 165 | 16 | 46608 | 0.89485 | 0.18543 | *GYPE* |
| ENSG00000017260 | 3 | 130559439 | 130745556 | 367 | 43 | 45450 | 0.8946 | 0.1855 | *ATP2C1* |
| ENSG00000221882 | 17 | 3171193 | 3192268 | 86 | 6 | 21342 | 0.89453 | 0.18552 | *OR3A2* |
| ENSG00000143442 | 1 | 151365200 | 151441941 | 86 | 13 | 50023 | 0.89437 | 0.18556 | *POGZ* |
| ENSG00000188368 | 19 | 42796250 | 42824973 | 31 | 6 | 50766 | 0.89423 | 0.1856 | *PRR19* |
| ENSG00000148814 | 10 | 134135614 | 134205010 | 307 | 32 | 44396 | 0.89401 | 0.18566 | *LRRC27* |
| ENSG00000255524 | 16 | 28638975 | 28680003 | 37 | 10 | 40343 | 0.89395 | 0.18567 | *NPIPB8* |
| ENSG00000090615 | 12 | 133335495 | 133415444 | 352 | 45 | 43767 | 0.89372 | 0.18574 | *GOLGA3* |
| ENSG00000029534 | 8 | 41500739 | 41764280 | 856 | 134 | 46207 | 0.89364 | 0.18576 | *ANK1* |
| ENSG00000171729 | 1 | 15469028 | 15556976 | 538 | 54 | 51304 | 0.89348 | 0.1858 | *TMEM51* |
| ENSG00000115946 | 2 | 68374976 | 68413370 | 92 | 13 | 50218 | 0.89348 | 0.1858 | *PNO1* |
| ENSG00000157240 | 7 | 90883783 | 90908123 | 50 | 10 | 52919 | 0.89349 | 0.1858 | *FZD1* |
| ENSG00000174775 | 11 | 522242 | 547287 | 148 | 20 | 52374 | 0.89303 | 0.18592 | *HRAS* |
| ENSG00000204611 | 19 | 52606344 | 52653175 | 160 | 23 | 42163 | 0.89287 | 0.18596 | *ZNF616* |
| ENSG00000165678 | 10 | 85889196 | 85923001 | 103 | 18 | 30793 | 0.89271 | 0.18601 | *GHITM* |
| ENSG00000269554 | 1 | 2111237 | 2133179 | 44 | 16 | 44572 | 0.89232 | 0.18611 | *AL590822.2* |
| ENSG00000148965 | 11 | 18242896 | 18268440 | 66 | 12 | 31820 | 0.8919 | 0.18622 | *SAA4* |
| ENSG00000174514 | 1 | 205528013 | 205582046 | 193 | 39 | 37762 | 0.89178 | 0.18626 | *MFSD4* |
| ENSG00000139324 | 12 | 88526073 | 88603664 | 105 | 18 | 43218 | 0.89167 | 0.18629 | *TMTC3* |
| ENSG00000144843 | 3 | 119288115 | 119318792 | 116 | 12 | 29518 | 0.89156 | 0.18631 | *ADPRH* |

| | | | | | | | | | |
|---|---|---|---|---|---|---|---|---|---|
| ENSG00000125878 | 20 | 574441 | 601042 | 100 | 30 | 43995 | 0.89153 | 0.18632 | *TCF15* |
| ENSG00000203867 | 10 | 112394155 | 112609227 | 779 | 105 | 43730 | 0.89141 | 0.18635 | *RBM20* |
| ENSG00000101773 | 18 | 20368224 | 20616451 | 613 | 39 | 31174 | 0.89088 | 0.1865 | *RBBP8* |
| ENSG00000128973 | 15 | 68489330 | 68559549 | 161 | 15 | 31511 | 0.89061 | 0.18657 | *CLN6* |
| ENSG00000184307 | 3 | 113656748 | 113694248 | 73 | 22 | 51447 | 0.89018 | 0.18669 | *ZDHHC23* |
| ENSG00000121388 | 13 | 19746173 | 19771496 | 233 | 25 | 50024 | 0.89015 | 0.18669 | *RP11-408E5.4* |
| ENSG00000153822 | 17 | 68039570 | 68141749 | 364 | 30 | 46920 | 0.88974 | 0.1868 | *KCNJ16* |
| ENSG00000116726 | 1 | 12824984 | 12848049 | 62 | 12 | 46929 | 0.88963 | 0.18683 | *PRAMEF12* |
| ENSG00000203943 | 1 | 84754049 | 84865640 | 264 | 45 | 42567 | 0.88919 | 0.18695 | *SAMD13* |
| ENSG00000182504 | 3 | 101432769 | 101499406 | 143 | 17 | 51486 | 0.88906 | 0.18698 | *CEP97* |
| ENSG00000198420 | 7 | 143538468 | 143609291 | 66 | 16 | 46019 | 0.88887 | 0.18704 | *FAM115A* |
| ENSG00000143367 | 1 | 151502781 | 151566059 | 104 | 22 | 38875 | 0.88868 | 0.18709 | *TUFT1* |
| ENSG00000118432 | 6 | 88839583 | 88886078 | 124 | 25 | 44398 | 0.88863 | 0.1871 | *CNR1* |
| ENSG00000101246 | 20 | 62319996 | 62349377 | 88 | 11 | 37044 | 0.88863 | 0.1871 | *ARFRP1* |
| ENSG00000185862 | 17 | 29620784 | 29651130 | 48 | 6 | 29485 | 0.88861 | 0.18711 | *EVI2B* |
| ENSG00000165406 | 10 | 45940035 | 46100354 | 599 | 24 | 44060 | 0.88847 | 0.18714 | *Mar-08* |
| ENSG00000255339 | 10 | 102255385 | 102299638 | 102 | 6 | 56693 | 0.88848 | 0.18714 | *NDUFB8* |
| ENSG00000160712 | 1 | 154367669 | 154451926 | 228 | 36 | 43856 | 0.88846 | 0.18715 | *IL6R* |
| ENSG00000167531 | 12 | 48951467 | 48973849 | 61 | 17 | 38580 | 0.88827 | 0.1872 | *LALBA* |
| ENSG00000110172 | 11 | 89924328 | 89966532 | 183 | 16 | 53528 | 0.88809 | 0.18725 | *CHORDC1* |
| ENSG00000111077 | 12 | 53430753 | 53468156 | 106 | 24 | 35713 | 0.88793 | 0.18729 | *TENC1* |
| ENSG00000091127 | 7 | 105070108 | 105172714 | 435 | 37 | 34762 | 0.88774 | 0.18734 | *PUS7* |
| ENSG00000154734 | 21 | 28198066 | 28227728 | 97 | 23 | 48215 | 0.88774 | 0.18734 | *ADAMTS1* |
| ENSG00000100385 | 22 | 37511878 | 37581094 | 217 | 61 | 48245 | 0.88773 | 0.18734 | *IL2RB* |
| ENSG00000164815 | 7 | 103756788 | 103858495 | 405 | 25 | 31388 | 0.8876 | 0.18738 | *ORC5* |
| ENSG00000151135 | 12 | 107339497 | 107382556 | 124 | 23 | 41711 | 0.88748 | 0.18741 | *C12orf23* |
| ENSG00000131094 | 17 | 43027061 | 43055439 | 104 | 24 | 48697 | 0.88739 | 0.18743 | *C1QL1* |
| ENSG00000131951 | 14 | 60376431 | 60540277 | 346 | 20 | 30686 | 0.88736 | 0.18744 | *LRRC9* |
| ENSG00000214871 | 7 | 23200760 | 23244503 | 148 | 10 | 39604 | 0.88702 | 0.18753 | *AC005082.1* |
| ENSG00000015475 | 22 | 18206906 | 18267536 | 245 | 53 | 40393 | 0.88703 | 0.18753 | *BID* |
| ENSG00000215915 | 1 | 1375069 | 1415538 | 53 | 18 | 45324 | 0.88683 | 0.18758 | *ATAD3C* |
| ENSG00000113361 | 5 | 31183857 | 31339253 | 390 | 72 | 45446 | 0.88686 | 0.18758 | *CDH6* |
| ENSG00000187545 | 1 | 12942727 | 12968101 | 11 | 7 | 35743 | 0.88679 | 0.1876 | *PRAMEF10* |
| ENSG00000175946 | 8 | 124647767 | 124675190 | 125 | 20 | 43582 | 0.88678 | 0.1876 | *KLHL38* |
| ENSG00000198176 | 13 | 114229013 | 114305785 | 141 | 18 | 42685 | 0.88646 | 0.18768 | *TFDP1* |
| ENSG00000104903 | 19 | 13199847 | 13223975 | 43 | 16 | 47706 | 0.88614 | 0.18777 | *LYL1* |

| | | | | | | | | | |
|---|---|---|---|---|---|---|---|---|---|
| ENSG00000169203 | 16 | 29485010 | 29527320 | 2 | 1 | 28126 | 0.8861 | 0.18778 | *RP11-231C14.4* |
| ENSG00000269363 | 2 | 232561517 | 232581621 | 68 | 10 | 45266 | 0.886 | 0.18781 | *MGC4771* |
| ENSG00000221938 | 7 | 143816151 | 143837162 | 91 | 5 | 43824 | 0.88596 | 0.18782 | *OR2A14* |
| ENSG00000112379 | 6 | 138473058 | 138675800 | 628 | 74 | 39552 | 0.88578 | 0.18787 | *KIAA1244* |
| ENSG00000006837 | 5 | 133531305 | 133716738 | 395 | 21 | 40553 | 0.8855 | 0.18794 | *CDKL3* |
| ENSG00000146223 | 6 | 42837356 | 42867663 | 98 | 15 | 40466 | 0.88535 | 0.18798 | *RPL7L1* |
| ENSG00000214706 | 3 | 50315163 | 50340349 | 23 | 9 | 46125 | 0.88531 | 0.188 | *IFRD2* |
| ENSG00000048052 | 7 | 18116572 | 19052039 | 2593 | 278 | 43694 | 0.8852 | 0.18802 | *HDAC9* |
| ENSG00000144724 | 3 | 61537243 | 62293288 | 2566 | 270 | 45163 | 0.88508 | 0.18806 | *PTPRG* |
| ENSG00000085552 | 1 | 159886829 | 159925386 | 110 | 29 | 41933 | 0.88497 | 0.18809 | *IGSF9* |
| ENSG00000171631 | 11 | 72965550 | 73019662 | 123 | 18 | 41819 | 0.88464 | 0.18818 | *P2RY6* |
| ENSG00000165119 | 9 | 86572998 | 86605569 | 63 | 12 | 52529 | 0.88448 | 0.18822 | *HNRNPK* |
| ENSG00000220201 | 19 | 10405479 | 10430556 | 88 | 19 | 43883 | 0.88438 | 0.18824 | *ZGLP1* |
| ENSG00000123992 | 2 | 220228268 | 220274744 | 95 | 22 | 42377 | 0.88436 | 0.18825 | *DNPEP* |
| ENSG00000137831 | 15 | 70936893 | 71065932 | 232 | 27 | 36247 | 0.8843 | 0.18827 | *UACA* |
| ENSG00000185177 | 7 | 57177321 | 57217571 | 161 | 12 | 35966 | 0.88416 | 0.1883 | *ZNF479* |
| ENSG00000221823 | 2 | 68395989 | 68493369 | 227 | 17 | 53570 | 0.88379 | 0.1884 | *PPP3R1* |
| ENSG00000145506 | 5 | 998944 | 1049058 | 235 | 33 | 45518 | 0.88358 | 0.18846 | *NKD2* |
| ENSG00000172382 | 16 | 2752419 | 2780552 | 21 | 13 | 42708 | 0.88354 | 0.18847 | *PRSS27* |
| ENSG00000196826 | 19 | 12562373 | 12672288 | 360 | 30 | 46075 | 0.88354 | 0.18847 | *ZNF709* |
| ENSG00000267059 | 19 | 1568338 | 1615444 | 72 | 19 | 52502 | 0.88351 | 0.18848 | *UQCR11* |
| ENSG00000071082 | 2 | 101608177 | 101650494 | 125 | 16 | 48934 | 0.88309 | 0.1886 | *RPL31* |
| ENSG00000035115 | 2 | 207730 | 276398 | 160 | 17 | 51323 | 0.88234 | 0.1888 | *SH3YL1* |
| ENSG00000183783 | 4 | 44165926 | 44460824 | 835 | 96 | 52921 | 0.88188 | 0.18892 | *KCTD8* |
| ENSG00000268211 | 9 | 131052367 | 131074212 | 65 | 13 | 34051 | 0.8819 | 0.18892 | *AL359091.2* |
| ENSG00000260007 | 15 | 68476139 | 68532056 | 135 | 14 | 34654 | 0.88126 | 0.18909 | *RP11-315D16.2* |
| ENSG00000160401 | 9 | 130459268 | 130488281 | 30 | 8 | 44153 | 0.88105 | 0.18915 | *C9orf117* |
| ENSG00000248710 | 3 | 159935241 | 160177617 | 469 | 52 | 48636 | 0.88059 | 0.18927 | *RP11-432B6.3* |
| ENSG00000156502 | 10 | 70929988 | 70978855 | 167 | 24 | 34147 | 0.88061 | 0.18927 | *SUPV3L1* |
| ENSG00000213578 | 15 | 75108888 | 75134141 | 48 | 12 | 36683 | 0.88034 | 0.18934 | *CPLX3* |
| ENSG00000100911 | 14 | 24602574 | 24626779 | 41 | 17 | 43547 | 0.88025 | 0.18936 | *PSME2* |
| ENSG00000143409 | 1 | 150959025 | 150990851 | 87 | 19 | 41828 | 0.88022 | 0.18937 | *FAM63A* |
| ENSG00000174791 | 11 | 66087713 | 66114311 | 59 | 12 | 35049 | 0.88013 | 0.18939 | *RIN1* |
| ENSG00000183793 | 16 | 15447516 | 15484904 | 45 | 13 | 38263 | 0.88011 | 0.1894 | *NPIPA5* |
| ENSG00000255587 | 6 | 36673256 | 36709870 | 148 | 24 | 44298 | 0.88008 | 0.18941 | *RAB44* |
| ENSG00000206559 | 3 | 28380637 | 28589613 | 365 | 32 | 38154 | 0.88002 | 0.18943 | *ZCWPW2* |

| | | | | | | | | | |
|---|---|---|---|---|---|---|---|---|---|
| ENSG00000152086 | 2 | 130939318 | 130966034 | 70 | 7 | 50076 | 0.87943 | 0.18958 | *TUBA3E* |
| ENSG00000117143 | 1 | 162521323 | 162579627 | 113 | 20 | 50805 | 0.87931 | 0.18962 | *UAP1* |
| ENSG00000082516 | 5 | 154256976 | 154327769 | 202 | 21 | 34516 | 0.87925 | 0.18963 | *GEMIN5* |
| ENSG00000141298 | 17 | 27942956 | 28267294 | 485 | 31 | 40846 | 0.87923 | 0.18964 | *SSH2* |
| ENSG00000168827 | 3 | 158352067 | 158420364 | 251 | 14 | 56370 | 0.8792 | 0.18965 | *GFM1* |
| ENSG00000204351 | 6 | 31916857 | 31947532 | 108 | 27 | 39694 | 0.87919 | 0.18965 | *SKIV2L* |
| ENSG00000213185 | 10 | 124598594 | 124649157 | 84 | 19 | 55764 | 0.8791 | 0.18967 | *FAM24B* |
| ENSG00000136243 | 7 | 23211446 | 23250630 | 137 | 9 | 39888 | 0.87898 | 0.18971 | *NUPL2* |
| ENSG00000169247 | 5 | 148293202 | 148452726 | 527 | 59 | 49127 | 0.87892 | 0.18972 | *SH3TC2* |
| ENSG00000176515 | 6 | 1070164 | 1115181 | 160 | 28 | 39724 | 0.87878 | 0.18976 | *AL033381.1* |
| ENSG00000031823 | 19 | 5906150 | 5988153 | 176 | 33 | 52869 | 0.87863 | 0.1898 | *RANBP3* |
| ENSG00000180974 | 11 | 5895501 | 5916527 | 117 | 10 | 56352 | 0.87838 | 0.18987 | *OR52E4* |
| ENSG00000154096 | 11 | 119278090 | 119305695 | 75 | 10 | 50183 | 0.87833 | 0.18988 | *THY1* |
| ENSG00000143171 | 1 | 165360159 | 165424433 | 281 | 45 | 42241 | 0.87816 | 0.18993 | *RXRG* |
| ENSG00000103047 | 16 | 68867507 | 69129083 | 624 | 63 | 47901 | 0.87813 | 0.18994 | *TANGO6* |
| ENSG00000169814 | 3 | 15632848 | 15697329 | 156 | 25 | 39075 | 0.87809 | 0.18995 | *BTD* |
| ENSG00000137672 | 11 | 101312295 | 101753293 | 2008 | 97 | 45436 | 0.878 | 0.18997 | *TRPC6* |
| ENSG00000147416 | 8 | 20044878 | 20094330 | 108 | 24 | 51396 | 0.87772 | 0.19005 | *ATP6V1B2* |
| ENSG00000135220 | 4 | 69784181 | 69827509 | 155 | 14 | 34307 | 0.87762 | 0.19007 | *UGT2A3* |
| ENSG00000155975 | 8 | 17094080 | 17169936 | 271 | 37 | 38232 | 0.87738 | 0.19014 | *VPS37A* |
| ENSG00000104369 | 8 | 75136935 | 75243563 | 272 | 37 | 49958 | 0.8773 | 0.19016 | *JPH1* |
| ENSG00000142511 | 19 | 51263721 | 51284989 | 65 | 26 | 39469 | 0.87728 | 0.19017 | *GPR32* |
| ENSG00000181817 | 1 | 36846839 | 36873493 | 110 | 10 | 53013 | 0.87717 | 0.1902 | *LSM10* |
| ENSG00000204344 | 6 | 31928868 | 31960598 | 80 | 22 | 39538 | 0.87709 | 0.19022 | *STK19* |
| ENSG00000006740 | 17 | 12682856 | 12904960 | 936 | 116 | 42332 | 0.87679 | 0.1903 | *ARHGAP44* |
| ENSG00000160679 | 1 | 153596525 | 153628782 | 75 | 9 | 50968 | 0.87674 | 0.19032 | *CHTOP* |
| ENSG00000145309 | 4 | 71190683 | 71212833 | 154 | 12 | 49325 | 0.87649 | 0.19038 | *CABS1* |
| ENSG00000204169 | 10 | 51454162 | 51496327 | 25 | 8 | 44003 | 0.8765 | 0.19038 | *AGAP7* |
| ENSG00000165816 | 10 | 115989018 | 116061272 | 212 | 28 | 48467 | 0.87647 | 0.19039 | *VWA2* |
| ENSG00000137500 | 11 | 82960139 | 83007450 | 145 | 16 | 49137 | 0.87647 | 0.19039 | *CCDC90B* |
| ENSG00000158887 | 1 | 161264525 | 161289762 | 94 | 19 | 35111 | 0.87637 | 0.19041 | *MPZ* |
| ENSG00000144339 | 2 | 192803769 | 193070435 | 560 | 97 | 43770 | 0.8764 | 0.19041 | *TMEFF2* |
| ENSG00000233863 | 8 | 35639365 | 35663384 | 13 | 8 | 46075 | 0.87638 | 0.19041 | *AC012215.1* |
| ENSG00000143452 | 1 | 150660536 | 150703364 | 130 | 8 | 51955 | 0.87634 | 0.19042 | *HORMAD1* |
| ENSG00000162739 | 1 | 160444820 | 160503052 | 223 | 33 | 32011 | 0.87624 | 0.19045 | *SLAMF6* |
| ENSG00000135604 | 6 | 144461663 | 144519507 | 113 | 27 | 37074 | 0.87625 | 0.19045 | *STX11* |

| | | | | | | | | | |
|---|---|---|---|---|---|---|---|---|---|
| ENSG00000127399 | 7 | 150009728 | 150045239 | 104 | 10 | 50280 | 0.87626 | 0.19045 | *LRRC61* |
| ENSG00000133619 | 7 | 149401872 | 149441664 | 117 | 19 | 40784 | 0.87607 | 0.1905 | *KRBA1* |
| ENSG00000109466 | 4 | 166118770 | 166254308 | 395 | 40 | 35202 | 0.87596 | 0.19053 | *KLHL2* |
| ENSG00000196228 | 2 | 108853651 | 108891807 | 143 | 20 | 33979 | 0.87589 | 0.19055 | *SULT1C3* |
| ENSG00000179564 | 3 | 50306458 | 50335545 | 22 | 8 | 47539 | 0.87562 | 0.19062 | *LSMEM2* |
| ENSG00000100249 | 22 | 29444660 | 29467832 | 83 | 22 | 41783 | 0.87533 | 0.1907 | *C22orf31* |
| ENSG00000145423 | 4 | 154691744 | 154720272 | 74 | 12 | 48321 | 0.87515 | 0.19075 | *SFRP2* |
| ENSG00000153113 | 5 | 95850971 | 96125299 | 1097 | 111 | 46027 | 0.87508 | 0.19076 | *CAST* |
| ENSG00000198283 | 11 | 58264648 | 58285578 | 27 | 9 | 51072 | 0.87503 | 0.19078 | *OR5B21* |
| ENSG00000162009 | 16 | 1112756 | 1141454 | 165 | 32 | 41020 | 0.87471 | 0.19086 | *SSTR5* |
| ENSG00000241123 | 21 | 45989332 | 46010481 | 92 | 12 | 32500 | 0.87458 | 0.1909 | *KRTAP10-5* |
| ENSG00000163516 | 2 | 220084479 | 220111391 | 66 | 13 | 41123 | 0.87412 | 0.19103 | *ANKZF1* |
| ENSG00000254087 | 8 | 56782372 | 56933940 | 556 | 59 | 50250 | 0.87409 | 0.19103 | *LYN* |
| ENSG00000117877 | 19 | 45899467 | 45924024 | 67 | 14 | 43123 | 0.87397 | 0.19107 | *CD3EAP* |
| ENSG00000198755 | 6 | 35426185 | 35448562 | 76 | 13 | 28244 | 0.87369 | 0.19114 | *RPL10A* |
| ENSG00000114745 | 3 | 39128150 | 39159854 | 87 | 20 | 36614 | 0.87341 | 0.19122 | *GORASP1* |
| ENSG00000187066 | 11 | 64842451 | 64866847 | 46 | 12 | 41775 | 0.87341 | 0.19122 | *TMEM262* |
| ENSG00000121454 | 1 | 180189421 | 180259380 | 274 | 56 | 44611 | 0.87339 | 0.19123 | *LHX4* |
| ENSG00000089250 | 12 | 117635947 | 117899975 | 767 | 118 | 43940 | 0.87338 | 0.19123 | *NOS1* |
| ENSG00000159387 | 16 | 55347672 | 55374672 | 112 | 15 | 50808 | 0.87311 | 0.1913 | *IRX6* |
| ENSG00000058404 | 7 | 44246749 | 44384176 | 413 | 55 | 49728 | 0.87304 | 0.19132 | *CAMK2B* |
| ENSG00000159658 | 1 | 47114366 | 47194824 | 181 | 14 | 37644 | 0.87266 | 0.19142 | *EFCAB14* |
| ENSG00000145337 | 4 | 89432136 | 89454964 | 21 | 10 | 37799 | 0.87258 | 0.19144 | *PYURF* |
| ENSG00000151655 | 10 | 7735232 | 7801483 | 304 | 48 | 43254 | 0.87245 | 0.19148 | *ITIH2* |
| ENSG00000104863 | 19 | 49607581 | 49631717 | 86 | 17 | 52012 | 0.87236 | 0.1915 | *LIN7B* |
| ENSG00000204952 | 17 | 37082685 | 37133655 | 66 | 17 | 51344 | 0.87228 | 0.19153 | *FBXO47* |
| ENSG00000100106 | 22 | 38083011 | 38182563 | 294 | 34 | 43928 | 0.8721 | 0.19158 | *TRIOBP* |
| ENSG00000124171 | 20 | 49338081 | 49383332 | 143 | 22 | 48083 | 0.87205 | 0.19159 | *PARD6B* |
| ENSG00000130813 | 19 | 10186798 | 10213928 | 44 | 16 | 46186 | 0.87201 | 0.1916 | *C19orf66* |
| ENSG00000118523 | 6 | 132259316 | 132282513 | 54 | 16 | 43520 | 0.87199 | 0.19161 | *CTGF* |
| ENSG00000102897 | 16 | 20901190 | 20946328 | 81 | 15 | 37787 | 0.87157 | 0.19172 | *LYRM1* |
| ENSG00000254505 | 14 | 24668789 | 24693075 | 52 | 11 | 42761 | 0.87104 | 0.19187 | *CHMP4A* |
| ENSG00000099901 | 22 | 20093461 | 20124878 | 120 | 16 | 49082 | 0.87103 | 0.19187 | *RANBP1* |
| ENSG00000159182 | 17 | 46789084 | 46809884 | 45 | 12 | 46292 | 0.871 | 0.19188 | *PRAC1* |
| ENSG00000173928 | 19 | 11475361 | 11497627 | 32 | 9 | 43522 | 0.87098 | 0.19188 | *SWSAP1* |
| ENSG00000105011 | 19 | 14220321 | 14257768 | 63 | 10 | 48469 | 0.87088 | 0.19191 | *ASF1B* |

| | | | | | | | | | |
|---|---|---|---|---|---|---|---|---|---|
| ENSG00000142544 | 19 | 51590863 | 51621627 | 85 | 22 | 45631 | 0.87039 | 0.19204 | *CTU1* |
| ENSG00000115286 | 19 | 1373526 | 1405583 | 98 | 24 | 36204 | 0.87013 | 0.19211 | *NDUFS7* |
| ENSG00000162929 | 2 | 61283006 | 61401960 | 306 | 28 | 47274 | 0.86985 | 0.19219 | *KIAA1841* |
| ENSG00000175548 | 12 | 38700380 | 38727784 | 99 | 12 | 36756 | 0.8698 | 0.19221 | *ALG10B* |
| ENSG00000189068 | 19 | 54534079 | 54577207 | 196 | 37 | 48902 | 0.86975 | 0.19222 | *VSTM1* |
| ENSG00000152700 | 5 | 133926834 | 133994961 | 99 | 21 | 40580 | 0.86958 | 0.19226 | *SAR1B* |
| ENSG00000149503 | 11 | 61881445 | 61930635 | 203 | 16 | 33469 | 0.86942 | 0.19231 | *INCENP* |
| ENSG00000198967 | 1 | 158566229 | 158587170 | 75 | 11 | 51180 | 0.86882 | 0.19247 | *OR10Z1* |
| ENSG00000100461 | 14 | 23359854 | 23398393 | 112 | 14 | 46396 | 0.86878 | 0.19248 | *RBM23* |
| ENSG00000048471 | 16 | 12060594 | 12678146 | 3113 | 226 | 38181 | 0.86878 | 0.19248 | *SNX29* |
| ENSG00000096080 | 6 | 43629040 | 43665528 | 116 | 16 | 30813 | 0.86848 | 0.19257 | *MRPS18A* |
| ENSG00000214814 | 8 | 124854227 | 125142302 | 701 | 79 | 51664 | 0.86845 | 0.19257 | *FER1L6* |
| ENSG00000172179 | 6 | 22277480 | 22307730 | 67 | 20 | 42247 | 0.86843 | 0.19258 | *PRL* |
| ENSG00000173581 | 19 | 56142428 | 56174527 | 94 | 20 | 41081 | 0.86845 | 0.19258 | *CCDC106* |
| ENSG00000154678 | 7 | 31780793 | 32348941 | 2356 | 224 | 46379 | 0.86837 | 0.19259 | *PDE1C* |
| ENSG00000113430 | 5 | 1867541 | 1897350 | 161 | 32 | 52776 | 0.86834 | 0.1926 | *IRX4* |
| ENSG00000117643 | 1 | 25933959 | 26122698 | 329 | 49 | 41931 | 0.86833 | 0.19261 | *MAN1C1* |
| ENSG00000160145 | 3 | 123788870 | 124455172 | 2217 | 181 | 47262 | 0.86813 | 0.19266 | *KALRN* |
| ENSG00000158161 | 1 | 28286855 | 28425207 | 379 | 41 | 45982 | 0.86805 | 0.19268 | *EYA3* |
| ENSG00000103932 | 15 | 41799374 | 41846467 | 154 | 19 | 45195 | 0.86738 | 0.19287 | *RPAP1* |
| ENSG00000163157 | 1 | 151132462 | 151158760 | 22 | 11 | 47812 | 0.86733 | 0.19288 | *TMOD4* |
| ENSG00000023191 | 11 | 484512 | 517300 | 189 | 21 | 54619 | 0.86703 | 0.19296 | *RNH1* |
| ENSG00000182175 | 15 | 93576636 | 93642433 | 266 | 58 | 44630 | 0.86696 | 0.19298 | *RGMA* |
| ENSG00000105929 | 7 | 138381040 | 138494305 | 487 | 96 | 49267 | 0.86694 | 0.19299 | *ATP6V0A4* |
| ENSG00000187689 | 4 | 71374257 | 71408459 | 222 | 19 | 37082 | 0.86671 | 0.19305 | *AMTN* |
| ENSG00000113555 | 5 | 141313150 | 141359304 | 114 | 27 | 46042 | 0.86628 | 0.19317 | *PCDH12* |
| ENSG00000119004 | 2 | 204093663 | 204173009 | 172 | 19 | 32408 | 0.8662 | 0.19319 | *CYP20A1* |
| ENSG00000131373 | 3 | 15592211 | 15653338 | 137 | 30 | 43404 | 0.86618 | 0.1932 | *HACL1* |
| ENSG00000137770 | 15 | 44709432 | 44831236 | 239 | 20 | 35737 | 0.86591 | 0.19327 | *CTDSPL2* |
| ENSG00000169397 | 14 | 21349558 | 21370507 | 39 | 10 | 44888 | 0.8658 | 0.1933 | *RNASE3* |
| ENSG00000253953 | 5 | 140757452 | 140902546 | 413 | 50 | 43816 | 0.86558 | 0.19336 | *PCDHGB4* |
| ENSG00000203896 | 20 | 62356815 | 62380456 | 86 | 11 | 39612 | 0.86552 | 0.19338 | *LIME1* |
| ENSG00000103194 | 16 | 84723584 | 84823528 | 427 | 56 | 40886 | 0.86546 | 0.19339 | *USP10* |
| ENSG00000065328 | 10 | 13193554 | 13263104 | 307 | 60 | 49959 | 0.86539 | 0.19341 | *MCM10* |
| ENSG00000006625 | 7 | 30526237 | 30601095 | 212 | 31 | 46022 | 0.86511 | 0.19349 | *GGCT* |
| ENSG00000153292 | 6 | 46955440 | 47020099 | 173 | 36 | 43059 | 0.86494 | 0.19354 | *GPR110* |

| | | | | | | | | | |
|---|---|---|---|---|---|---|---|---|---|
| ENSG00000255974 | 19 | 41339443 | 41366352 | 106 | 26 | 38804 | 0.86493 | 0.19354 | *CYP2A6* |
| ENSG00000101596 | 18 | 2645737 | 2815015 | 655 | 45 | 47886 | 0.8649 | 0.19355 | *SMCHD1* |
| ENSG00000138696 | 4 | 95669119 | 96089599 | 1542 | 107 | 46639 | 0.86467 | 0.19361 | *BMPR1B* |
| ENSG00000157212 | 7 | 154725397 | 154804794 | 182 | 16 | 51361 | 0.8645 | 0.19366 | *PAXIP1* |
| ENSG00000120616 | 10 | 32546679 | 32677726 | 403 | 41 | 47197 | 0.86447 | 0.19366 | *EPC1* |
| ENSG00000145428 | 4 | 154621277 | 154691387 | 258 | 21 | 36094 | 0.86435 | 0.1937 | *RNF175* |
| ENSG00000119969 | 10 | 96295547 | 96383662 | 199 | 18 | 53024 | 0.86432 | 0.19371 | *HELLS* |
| ENSG00000135637 | 2 | 74689113 | 74720535 | 22 | 14 | 50812 | 0.86408 | 0.19377 | *CCDC142* |
| ENSG00000109171 | 4 | 48333339 | 48438229 | 348 | 28 | 49981 | 0.86407 | 0.19377 | *SLAIN2* |
| ENSG00000122585 | 7 | 24313782 | 24341484 | 130 | 20 | 55508 | 0.86383 | 0.19384 | *NPY* |
| ENSG00000157870 | 1 | 2507930 | 2532908 | 106 | 12 | 35161 | 0.86379 | 0.19385 | *FAM213B* |
| ENSG00000166164 | 16 | 50337398 | 50412845 | 191 | 22 | 45161 | 0.86352 | 0.19393 | *BRD7* |
| ENSG00000130045 | 9 | 91140016 | 91209189 | 87 | 32 | 43431 | 0.86346 | 0.19394 | *NXNL2* |
| ENSG00000272772 | 5 | 133492861 | 133571762 | 204 | 13 | 50262 | 0.86334 | 0.19397 | *CTD-2410N18.5* |
| ENSG00000184345 | 3 | 51885645 | 51907440 | 71 | 8 | 34673 | 0.86323 | 0.19401 | *IQCF2* |
| ENSG00000141027 | 17 | 15922471 | 16131499 | 448 | 27 | 43217 | 0.86259 | 0.19418 | *NCOR1* |
| ENSG00000118492 | 6 | 146910101 | 147146598 | 689 | 97 | 49621 | 0.86236 | 0.19424 | *ADGB* |
| ENSG00000008196 | 6 | 50776436 | 50825326 | 135 | 22 | 51468 | 0.86234 | 0.19425 | *TFAP2B* |
| ENSG00000171944 | 11 | 5142922 | 5163872 | 54 | 15 | 51346 | 0.86234 | 0.19425 | *OR52A5* |
| ENSG00000131203 | 8 | 39749794 | 39795963 | 143 | 19 | 48738 | 0.86221 | 0.19429 | *IDO1* |
| ENSG00000268759 | 15 | 32764822 | 32790287 | 1 | 1 | 42858 | 0.86216 | 0.1943 | *AC135983.2* |
| ENSG00000105404 | 19 | 42450833 | 42473542 | 30 | 7 | 45302 | 0.86211 | 0.19431 | *RABAC1* |
| ENSG00000139436 | 12 | 110357607 | 110444194 | 196 | 23 | 37722 | 0.86206 | 0.19433 | *GIT2* |
| ENSG00000169598 | 1 | 3763845 | 3811993 | 273 | 41 | 45142 | 0.86137 | 0.19452 | *DFFB* |
| ENSG00000174607 | 4 | 115509611 | 115609380 | 232 | 33 | 38124 | 0.86127 | 0.19455 | *UGT8* |
| ENSG00000109472 | 4 | 166272346 | 166429472 | 776 | 45 | 51122 | 0.86113 | 0.19458 | *CPE* |
| ENSG00000043093 | 3 | 182645862 | 182713741 | 108 | 16 | 31075 | 0.86096 | 0.19463 | *DCUN1D1* |
| ENSG00000243284 | 1 | 159814106 | 159842447 | 90 | 21 | 42086 | 0.8609 | 0.19465 | *VSIG8* |
| ENSG00000152784 | 4 | 81095033 | 81135483 | 140 | 22 | 31709 | 0.86087 | 0.19465 | *PRDM8* |
| ENSG00000065613 | 10 | 105716959 | 105798991 | 283 | 22 | 49328 | 0.8605 | 0.19476 | *SLK* |
| ENSG00000187231 | 2 | 179956419 | 180139517 | 541 | 46 | 41691 | 0.86034 | 0.1948 | *SESTD1* |
| ENSG00000055044 | 2 | 203120439 | 203178389 | 126 | 16 | 42098 | 0.8603 | 0.19481 | *NOP58* |
| ENSG00000175766 | 5 | 176047683 | 176083642 | 69 | 16 | 32628 | 0.85997 | 0.1949 | *EIF4E1B* |
| ENSG00000255245 | 11 | 117680878 | 117757382 | 235 | 61 | 47091 | 0.85957 | 0.19501 | *FXYD6-FXYD2* |
| ENSG00000080819 | 3 | 98229976 | 98322567 | 271 | 49 | 45550 | 0.85905 | 0.19516 | *CPOX* |
| ENSG00000138336 | 10 | 70310413 | 70464239 | 589 | 46 | 43537 | 0.85889 | 0.1952 | *TET1* |

| | | | | | | | | | |
|---|---|---|---|---|---|---|---|---|---|
| ENSG00000072310 | 17 | 17703713 | 17750325 | 74 | 9 | 27020 | 0.85881 | 0.19522 | *SREBF1* |
| ENSG00000184886 | 17 | 34880847 | 34905159 | 55 | 9 | 30380 | 0.85877 | 0.19523 | *PIGW* |
| ENSG00000101557 | 18 | 148383 | 224629 | 184 | 25 | 53436 | 0.85873 | 0.19525 | *USP14* |
| ENSG00000196605 | 19 | 9852669 | 9913856 | 142 | 15 | 36113 | 0.85859 | 0.19528 | *ZNF846* |
| ENSG00000133477 | 22 | 40380953 | 40449533 | 110 | 21 | 47737 | 0.85836 | 0.19535 | *FAM83F* |
| ENSG00000173557 | 2 | 26775450 | 26812400 | 96 | 23 | 43943 | 0.85818 | 0.1954 | *C2orf70* |
| ENSG00000177354 | 10 | 50497187 | 50545537 | 160 | 33 | 49003 | 0.8581 | 0.19542 | *C10orf71* |
| ENSG00000163961 | 3 | 196185654 | 196240639 | 185 | 27 | 37175 | 0.85774 | 0.19552 | *RNF168* |
| ENSG00000063515 | 22 | 19126089 | 19147796 | 77 | 18 | 43587 | 0.85772 | 0.19552 | *GSC2* |
| ENSG00000100034 | 22 | 22263793 | 22317209 | 172 | 21 | 50370 | 0.85774 | 0.19552 | *PPM1F* |
| ENSG00000165462 | 11 | 71940121 | 71966708 | 48 | 12 | 40775 | 0.85771 | 0.19553 | *PHOX2A* |
| ENSG00000179008 | 14 | 60853187 | 60992261 | 332 | 19 | 27791 | 0.85732 | 0.19564 | *C14orf39* |
| ENSG00000197891 | 11 | 64348113 | 64379820 | 74 | 10 | 31359 | 0.85718 | 0.19567 | *SLC22A12* |
| ENSG00000101265 | 20 | 4750669 | 4814291 | 331 | 64 | 48479 | 0.8567 | 0.19581 | *RASSF2* |
| ENSG00000082701 | 3 | 119530170 | 119823264 | 536 | 46 | 42213 | 0.85662 | 0.19583 | *GSK3B* |
| ENSG00000161692 | 17 | 42775976 | 42839632 | 127 | 24 | 54871 | 0.85602 | 0.19599 | *DBF4B* |
| ENSG00000177275 | 1 | 248087071 | 248108057 | 58 | 11 | 36260 | 0.85586 | 0.19604 | *OR2AJ1* |
| ENSG00000010932 | 1 | 171207638 | 171265117 | 185 | 31 | 39420 | 0.85578 | 0.19606 | *FMO1* |
| ENSG00000163900 | 3 | 185184284 | 185226845 | 58 | 9 | 48372 | 0.85568 | 0.19609 | *TMEM41A* |
| ENSG00000117036 | 1 | 157080983 | 157118266 | 65 | 17 | 32317 | 0.85563 | 0.1961 | *ETV3* |
| ENSG00000165495 | 11 | 125024583 | 125313285 | 868 | 138 | 44977 | 0.85564 | 0.1961 | *PKNOX2* |
| ENSG00000187918 | 11 | 5464638 | 5485707 | 142 | 22 | 39322 | 0.85534 | 0.19618 | *OR51I2* |
| ENSG00000100316 | 22 | 39698887 | 39726394 | 124 | 19 | 35286 | 0.85517 | 0.19623 | *RPL3* |
| ENSG00000165879 | 10 | 99069022 | 99091672 | 97 | 12 | 50755 | 0.85505 | 0.19626 | *FRAT1* |
| ENSG00000150526 | 14 | 39689435 | 39732859 | 133 | 20 | 46437 | 0.85502 | 0.19627 | *MIA2* |
| ENSG00000205363 | 15 | 74022141 | 74055088 | 119 | 21 | 43787 | 0.85454 | 0.1964 | *C15orf59* |
| ENSG00000171201 | 4 | 71225810 | 71265961 | 240 | 27 | 44016 | 0.85445 | 0.19643 | *SMR3B* |
| ENSG00000151962 | 4 | 155692365 | 155759965 | 144 | 16 | 38922 | 0.85437 | 0.19645 | *RBM46* |
| ENSG00000186529 | 19 | 15741707 | 15783635 | 345 | 28 | 47886 | 0.85438 | 0.19645 | *CYP4F3* |
| ENSG00000141456 | 17 | 4564679 | 4617632 | 181 | 20 | 37071 | 0.85422 | 0.19649 | *PELP1* |
| ENSG00000186141 | 1 | 145582605 | 145621025 | 34 | 10 | 43922 | 0.85412 | 0.19652 | *POLR3C* |
| ENSG00000132517 | 17 | 4925895 | 4965304 | 155 | 23 | 39326 | 0.85401 | 0.19655 | *SLC52A1* |
| ENSG00000163728 | 3 | 180309918 | 180345616 | 53 | 18 | 45188 | 0.85388 | 0.19659 | *TTC14* |
| ENSG00000113558 | 5 | 133474633 | 133522729 | 94 | 13 | 43785 | 0.85366 | 0.19665 | *SKP1* |
| ENSG00000151914 | 6 | 56312785 | 56829426 | 1151 | 71 | 46770 | 0.85366 | 0.19665 | *DST* |
| ENSG00000141682 | 18 | 57557180 | 57581538 | 67 | 20 | 47455 | 0.85343 | 0.19671 | *PMAIP1* |

| | | | | | | | | | |
|---|---|---|---|---|---|---|---|---|---|
| ENSG00000004468 | 4 | 15769898 | 15864853 | 311 | 29 | 34325 | 0.85338 | 0.19672 | *CD38* |
| ENSG00000101457 | 20 | 44410576 | 44450066 | 142 | 18 | 47585 | 0.8534 | 0.19672 | *DNTTIP1* |
| ENSG00000122565 | 7 | 26230782 | 26262976 | 69 | 10 | 50679 | 0.85244 | 0.19699 | *CBX3* |
| ENSG00000112183 | 6 | 17271577 | 17304106 | 96 | 23 | 53279 | 0.85235 | 0.19701 | *RBM24* |
| ENSG00000144560 | 3 | 11587544 | 11772220 | 684 | 95 | 39702 | 0.85228 | 0.19703 | *VGLL4* |
| ENSG00000204427 | 6 | 31644726 | 31681221 | 84 | 17 | 31735 | 0.85188 | 0.19714 | *ABHD16A* |
| ENSG00000104312 | 8 | 90759975 | 90813291 | 150 | 16 | 31922 | 0.85189 | 0.19714 | *RIPK2* |
| ENSG00000154162 | 5 | 21740782 | 22863731 | 3349 | 220 | 45201 | 0.85161 | 0.19722 | *CDH12* |
| ENSG00000138172 | 10 | 105196543 | 105222660 | 48 | 18 | 50522 | 0.8516 | 0.19722 | *CALHM2* |
| ENSG00000067704 | 1 | 220257444 | 220331380 | 185 | 26 | 53342 | 0.85157 | 0.19723 | *IARS2* |
| ENSG00000177462 | 1 | 248074320 | 248095258 | 84 | 13 | 28577 | 0.85153 | 0.19724 | *OR2T8* |
| ENSG00000143486 | 1 | 206734620 | 206795904 | 208 | 36 | 53149 | 0.85146 | 0.19726 | *EIF2D* |
| ENSG00000253767 | 5 | 140762381 | 140902546 | 401 | 49 | 43547 | 0.85131 | 0.1973 | *PCDHGA8* |
| ENSG00000166558 | 16 | 84033272 | 84086241 | 486 | 71 | 42075 | 0.85126 | 0.19731 | *SLC38A8* |
| ENSG00000101017 | 20 | 44736911 | 44768502 | 106 | 23 | 50675 | 0.85125 | 0.19732 | *CD40* |
| ENSG00000087206 | 5 | 176322006 | 176459634 | 514 | 23 | 45458 | 0.85096 | 0.1974 | *UIMC1* |
| ENSG00000134183 | 1 | 110135889 | 110165679 | 64 | 15 | 40116 | 0.85077 | 0.19745 | *GNAT2* |
| ENSG00000111725 | 12 | 120095558 | 120129435 | 59 | 9 | 52153 | 0.8502 | 0.19761 | *PRKAB1* |
| ENSG00000161888 | 19 | 11232196 | 11276484 | 247 | 32 | 45328 | 0.85014 | 0.19762 | *SPC24* |
| ENSG00000198774 | 12 | 86188331 | 86240348 | 229 | 23 | 52321 | 0.85006 | 0.19765 | *RASSF9* |
| ENSG00000168298 | 6 | 26146559 | 26167343 | 25 | 10 | 40967 | 0.84992 | 0.19769 | *HIST1H1E* |
| ENSG00000198625 | 1 | 204475511 | 204552871 | 189 | 15 | 32499 | 0.84964 | 0.19776 | *MDM4* |
| ENSG00000183864 | 22 | 41819496 | 41853027 | 70 | 6 | 51545 | 0.84955 | 0.19779 | *TOB2* |
| ENSG00000130997 | 4 | 2063645 | 2253848 | 498 | 19 | 27951 | 0.84952 | 0.1978 | *POLN* |
| ENSG00000179051 | 1 | 17723256 | 17776220 | 180 | 21 | 54909 | 0.84901 | 0.19794 | *RCC2* |
| ENSG00000167165 | 2 | 234590253 | 234691946 | 420 | 39 | 44226 | 0.84901 | 0.19794 | *UGT1A6* |
| ENSG00000160469 | 19 | 55783440 | 55833901 | 167 | 26 | 52383 | 0.84878 | 0.198 | *BRSK1* |
| ENSG00000179364 | 14 | 105756900 | 105874484 | 112 | 34 | 47362 | 0.84852 | 0.19807 | *PACS2* |
| ENSG00000102879 | 16 | 30184148 | 30210397 | 27 | 9 | 38004 | 0.84834 | 0.19812 | *CORO1A* |
| ENSG00000213088 | 1 | 159163097 | 159186290 | 82 | 21 | 37340 | 0.84793 | 0.19824 | *DARC* |
| ENSG00000116030 | 2 | 203060903 | 203113331 | 129 | 18 | 53242 | 0.84741 | 0.19838 | *SUMO1* |
| ENSG00000171772 | 10 | 135357404 | 135392876 | 163 | 5 | 49614 | 0.84721 | 0.19844 | *SYCE1* |
| ENSG00000141179 | 17 | 53818340 | 53930191 | 334 | 28 | 43582 | 0.84718 | 0.19845 | *PCTP* |
| ENSG00000143418 | 1 | 150923059 | 150957479 | 77 | 13 | 52190 | 0.84714 | 0.19846 | *CERS2* |
| ENSG00000162594 | 1 | 67622083 | 67735662 | 342 | 45 | 51757 | 0.84709 | 0.19847 | *IL23R* |
| ENSG00000175376 | 11 | 65754016 | 65779647 | 100 | 12 | 41621 | 0.8471 | 0.19847 | *EIF1AD* |

| | | | | | | | | | |
|---|---|---|---|---|---|---|---|---|---|
| ENSG00000125812 | 20 | 23332787 | 23363700 | 117 | 14 | 40746 | 0.84682 | 0.19855 | *GZF1* |
| ENSG00000203772 | 10 | 135224170 | 135392916 | 572 | 52 | 41499 | 0.84675 | 0.19857 | *SPRN* |
| ENSG00000118298 | 1 | 150219554 | 150247478 | 35 | 9 | 46634 | 0.84658 | 0.19862 | *CA14* |
| ENSG00000154485 | 10 | 127445022 | 127474390 | 47 | 14 | 52791 | 0.84655 | 0.19862 | *MMP21* |
| ENSG00000172828 | 16 | 66985140 | 67019051 | 50 | 21 | 48607 | 0.84657 | 0.19862 | *CES3* |
| ENSG00000138136 | 10 | 102976733 | 102999551 | 42 | 12 | 43684 | 0.84629 | 0.19869 | *LBX1* |
| ENSG00000172005 | 2 | 95681422 | 95729737 | 58 | 18 | 49417 | 0.84596 | 0.19879 | *MAL* |
| ENSG00000132321 | 2 | 237222794 | 237426185 | 477 | 51 | 49677 | 0.8458 | 0.19883 | *IQCA1* |
| ENSG00000169208 | 14 | 22027934 | 22048875 | 128 | 26 | 43317 | 0.84582 | 0.19883 | *OR10G3* |
| ENSG00000158806 | 8 | 21871636 | 21904408 | 83 | 26 | 44131 | 0.84572 | 0.19886 | *NPM2* |
| ENSG00000221840 | 11 | 51401378 | 51422448 | 95 | 11 | 45440 | 0.84534 | 0.19896 | *OR4A5* |
| ENSG00000163536 | 3 | 167443031 | 167553356 | 379 | 34 | 43812 | 0.84475 | 0.19913 | *SERPINI1* |
| ENSG00000160563 | 9 | 134725494 | 134965295 | 373 | 47 | 45967 | 0.8441 | 0.19931 | *MED27* |
| ENSG00000065457 | 16 | 75620879 | 75667198 | 109 | 19 | 37005 | 0.84396 | 0.19935 | *ADAT1* |
| ENSG00000171094 | 2 | 29405640 | 30154432 | 2757 | 231 | 43080 | 0.84369 | 0.19942 | *ALK* |
| ENSG00000112699 | 6 | 1614041 | 2255926 | 1771 | 203 | 45481 | 0.84352 | 0.19947 | *GMDS* |
| ENSG00000136235 | 7 | 23265586 | 23324727 | 98 | 16 | 40396 | 0.84353 | 0.19947 | *GPNMB* |
| ENSG00000170638 | 22 | 50614344 | 50648027 | 141 | 24 | 46018 | 0.84347 | 0.19948 | *TRABD* |
| ENSG00000153207 | 1 | 246992400 | 247105280 | 305 | 38 | 42347 | 0.8432 | 0.19956 | *AHCTF1* |
| ENSG00000204590 | 6 | 30499154 | 30534951 | 66 | 14 | 50044 | 0.84293 | 0.19964 | *GNL1* |
| ENSG00000137825 | 15 | 41775591 | 41805747 | 73 | 20 | 49246 | 0.84287 | 0.19965 | *ITPKA* |
| ENSG00000111906 | 6 | 125531108 | 125633282 | 340 | 34 | 43543 | 0.84255 | 0.19974 | *HDDC2* |
| ENSG00000118113 | 11 | 102572526 | 102607781 | 194 | 30 | 41816 | 0.8422 | 0.19984 | *MMP8* |
| ENSG00000043355 | 13 | 100624026 | 100649018 | 45 | 17 | 36665 | 0.84211 | 0.19986 | *ZIC2* |
| ENSG00000170006 | 4 | 153529784 | 153611317 | 243 | 50 | 47200 | 0.84205 | 0.19988 | *TMEM154* |
| ENSG00000188817 | 3 | 63628344 | 63674696 | 211 | 25 | 48859 | 0.84181 | 0.19995 | *SNTN* |
| ENSG00000150076 | 10 | 32822297 | 33181802 | 1550 | 60 | 42153 | 0.84144 | 0.20005 | *C10ORF68* |
| ENSG00000198691 | 1 | 94448393 | 94596688 | 584 | 100 | 43267 | 0.84138 | 0.20007 | *ABCA4* |
| ENSG00000171649 | 19 | 58079824 | 58115145 | 66 | 9 | 56400 | 0.84129 | 0.20009 | *ZIK1* |
| ENSG00000183605 | 10 | 120890279 | 120935179 | 144 | 21 | 45912 | 0.84121 | 0.20011 | *SFXN4* |
| ENSG00000251247 | 19 | 37331263 | 37413931 | 246 | 27 | 35576 | 0.84102 | 0.20017 | *ZNF345* |
| ENSG00000267173 | 19 | 44823848 | 44915774 | 344 | 49 | 38076 | 0.84077 | 0.20024 | *CTC-512J12.6* |
| ENSG00000156395 | 10 | 106390859 | 107034993 | 2086 | 113 | 46478 | 0.8407 | 0.20026 | *SORCS3* |
| ENSG00000177674 | 1 | 11786141 | 11824859 | 95 | 24 | 43122 | 0.84066 | 0.20027 | *AGTRAP* |
| ENSG00000134072 | 3 | 9789026 | 9821676 | 68 | 18 | 50185 | 0.84064 | 0.20028 | *CAMK1* |
| ENSG00000071189 | 7 | 17820385 | 17990124 | 419 | 30 | 39418 | 0.84055 | 0.2003 | *SNX13* |

| | | | | | | | | | |
|---|---|---|---|---|---|---|---|---|---|
| ENSG00000149527 | 1 | 2347419 | 2446969 | 356 | 74 | 44900 | 0.84048 | 0.20032 | *PLCH2* |
| ENSG00000223865 | 6 | 33033703 | 33064978 | 503 | 20 | 25755 | 0.84022 | 0.20039 | *HLA-DPB1* |
| ENSG00000198346 | 19 | 53960989 | 54016950 | 369 | 68 | 48080 | 0.84021 | 0.20039 | *ZNF813* |
| ENSG00000042813 | 7 | 49880017 | 50170925 | 867 | 41 | 50519 | 0.84015 | 0.20041 | *ZPBP* |
| ENSG00000269282 | 12 | 48749919 | 48771738 | 42 | 8 | 45879 | 0.84007 | 0.20043 | *AC024257.1* |
| ENSG00000135956 | 2 | 96904254 | 96941732 | 46 | 16 | 46560 | 0.84002 | 0.20045 | *TMEM127* |
| ENSG00000145721 | 5 | 96417574 | 96488576 | 151 | 29 | 45181 | 0.83994 | 0.20047 | *LIX1* |
| ENSG00000132622 | 20 | 3703314 | 3743758 | 132 | 41 | 45775 | 0.83993 | 0.20047 | *HSPA12B* |
| ENSG00000205649 | 4 | 70884130 | 70912255 | 104 | 17 | 49431 | 0.83984 | 0.2005 | *HTN3* |
| ENSG00000173039 | 11 | 65411067 | 65440565 | 56 | 12 | 43645 | 0.83983 | 0.2005 | *RELA* |
| ENSG00000113441 | 5 | 96261098 | 96383219 | 388 | 26 | 49121 | 0.83978 | 0.20052 | *LNPEP* |
| ENSG00000186652 | 11 | 57144267 | 57168130 | 46 | 10 | 51659 | 0.83956 | 0.20058 | *PRG2* |
| ENSG00000066813 | 16 | 20537548 | 20597749 | 159 | 21 | 26564 | 0.83955 | 0.20058 | *ACSM2B* |
| ENSG00000132199 | 18 | 660324 | 722676 | 322 | 27 | 40215 | 0.83936 | 0.20063 | *ENOSF1* |
| ENSG00000106348 | 7 | 128022331 | 128060306 | 127 | 25 | 51275 | 0.83929 | 0.20065 | *IMPDH1* |
| ENSG00000196504 | 2 | 153498107 | 153584511 | 244 | 25 | 49087 | 0.83918 | 0.20068 | *PRPF40A* |
| ENSG00000219016 | 22 | 48924712 | 48953199 | 114 | 29 | 45098 | 0.83908 | 0.20071 | *CTA-299D3.8* |
| ENSG00000115241 | 2 | 27594061 | 27642554 | 79 | 9 | 35220 | 0.83899 | 0.20074 | *PPM1G* |
| ENSG00000113389 | 5 | 32679176 | 32801819 | 368 | 50 | 52539 | 0.83899 | 0.20074 | *NPR3* |
| ENSG00000179094 | 17 | 8033790 | 8069824 | 111 | 19 | 47783 | 0.83899 | 0.20074 | *PER1* |
| ENSG00000101639 | 18 | 12981361 | 13135051 | 512 | 40 | 38875 | 0.83892 | 0.20076 | *CEP192* |
| ENSG00000165805 | 12 | 88363816 | 88437814 | 140 | 18 | 39327 | 0.8388 | 0.20079 | *C12orf50* |
| ENSG00000174970 | 11 | 55724975 | 55745990 | 195 | 7 | 54631 | 0.83829 | 0.20093 | *OR10AG1* |
| ENSG00000181827 | 15 | 56369478 | 56545483 | 552 | 27 | 39496 | 0.83721 | 0.20124 | *RFX7* |
| ENSG00000140284 | 15 | 50464393 | 50538592 | 338 | 40 | 42386 | 0.83705 | 0.20128 | *SLC27A2* |
| ENSG00000241119 | 2 | 234570499 | 234691946 | 507 | 46 | 45650 | 0.83671 | 0.20138 | *UGT1A9* |
| ENSG00000184226 | 13 | 66866967 | 67814468 | 2515 | 228 | 49804 | 0.83666 | 0.20139 | *PCDH9* |
| ENSG00000161267 | 3 | 197226654 | 197310194 | 376 | 54 | 45284 | 0.83646 | 0.20145 | *BDH1* |
| ENSG00000138294 | 10 | 51539498 | 51572517 | 52 | 14 | 45599 | 0.83618 | 0.20153 | *MSMB* |
| ENSG00000184840 | 5 | 177009159 | 177033125 | 90 | 17 | 51184 | 0.83603 | 0.20157 | *TMED9* |
| ENSG00000136161 | 13 | 49053095 | 49117369 | 79 | 15 | 33871 | 0.83604 | 0.20157 | *RCBTB2* |
| ENSG00000095627 | 10 | 115929029 | 116002063 | 215 | 18 | 53241 | 0.836 | 0.20158 | *TDRD1* |
| ENSG00000128165 | 22 | 50909985 | 50934869 | 75 | 29 | 45951 | 0.836 | 0.20158 | *ADM2* |
| ENSG00000095380 | 9 | 100809021 | 100855357 | 106 | 13 | 34824 | 0.8359 | 0.2016 | *NANS* |
| ENSG00000198171 | 20 | 3160996 | 3195331 | 176 | 29 | 49292 | 0.83572 | 0.20166 | *DDRGK1* |
| ENSG00000196967 | 19 | 37587636 | 37673643 | 254 | 21 | 47241 | 0.83551 | 0.20171 | *ZNF585A* |

| | | | | | | | | | |
|---|---|---|---|---|---|---|---|---|---|
| ENSG00000140937 | 16 | 64967656 | 65170015 | 655 | 65 | 44281 | 0.8353 | 0.20178 | *CDH11* |
| ENSG00000139197 | 12 | 7331281 | 7381170 | 189 | 25 | 40035 | 0.83513 | 0.20182 | *PEX5* |
| ENSG00000137936 | 1 | 94017347 | 94322706 | 1045 | 66 | 34568 | 0.83508 | 0.20184 | *BCAR3* |
| ENSG00000147872 | 9 | 19098373 | 19159288 | 228 | 41 | 45129 | 0.8349 | 0.20189 | *PLIN2* |
| ENSG00000072571 | 5 | 162877209 | 162928947 | 130 | 22 | 46695 | 0.83485 | 0.2019 | *HMMR* |
| ENSG00000253537 | 5 | 140752467 | 140902546 | 432 | 51 | 43877 | 0.83475 | 0.20193 | *PCDHGA7* |
| ENSG00000184584 | 5 | 138845119 | 138872520 | 46 | 15 | 39077 | 0.83467 | 0.20195 | *TMEM173* |
| ENSG00000127831 | 2 | 219273815 | 219328018 | 83 | 14 | 41168 | 0.83465 | 0.20196 | *VIL1* |
| ENSG00000107581 | 10 | 120784356 | 120850316 | 104 | 14 | 29393 | 0.83442 | 0.20202 | *EIF3A* |
| ENSG00000137992 | 1 | 100642475 | 100725390 | 201 | 19 | 29853 | 0.8344 | 0.20203 | *DBT* |
| ENSG00000113575 | 5 | 133520025 | 133571833 | 147 | 8 | 52591 | 0.83424 | 0.20207 | *PPP2CA* |
| ENSG00000037637 | 1 | 16563334 | 16688949 | 318 | 21 | 43435 | 0.83409 | 0.20211 | *FBXO42* |
| ENSG00000153574 | 2 | 88981162 | 89060452 | 205 | 13 | 27300 | 0.83391 | 0.20217 | *RPIA* |
| ENSG00000136834 | 9 | 125229220 | 125250235 | 90 | 18 | 48791 | 0.83333 | 0.20233 | *OR1J1* |
| ENSG00000125899 | 20 | 10998408 | 11020014 | 44 | 18 | 46265 | 0.83274 | 0.20249 | *C20orf187* |
| ENSG00000176204 | 2 | 76964845 | 77830445 | 3388 | 232 | 41485 | 0.83259 | 0.20254 | *LRRTM4* |
| ENSG00000185634 | 15 | 49105932 | 49265641 | 429 | 75 | 43838 | 0.83252 | 0.20256 | *SHC4* |
| ENSG00000116983 | 1 | 40134320 | 40167361 | 148 | 28 | 47896 | 0.83243 | 0.20258 | *HPCAL4* |
| ENSG00000244355 | 6 | 31673133 | 31695581 | 52 | 15 | 27144 | 0.83237 | 0.2026 | *LY6G6D* |
| ENSG00000255472 | 14 | 21490977 | 21512054 | 90 | 31 | 46560 | 0.8323 | 0.20262 | *RP11-998D10.1* |
| ENSG00000206150 | 14 | 21490979 | 21512944 | 90 | 31 | 46560 | 0.8323 | 0.20262 | *RNASE13* |
| ENSG00000273294 | 5 | 33977279 | 34134633 | 341 | 48 | 38795 | 0.83222 | 0.20264 | *RP11-1084J3.4* |
| ENSG00000205327 | 12 | 55876147 | 55897100 | 106 | 17 | 41579 | 0.83207 | 0.20269 | *OR6C68* |
| ENSG00000220205 | 17 | 8052467 | 8076864 | 65 | 12 | 46389 | 0.83207 | 0.20269 | *VAMP2* |
| ENSG00000151929 | 10 | 121400882 | 121447331 | 177 | 29 | 43525 | 0.83179 | 0.20276 | *BAG3* |
| ENSG00000144395 | 2 | 197494278 | 197638214 | 332 | 31 | 44795 | 0.83165 | 0.2028 | *CCDC150* |
| ENSG00000177663 | 22 | 17555844 | 17606583 | 151 | 38 | 44920 | 0.83164 | 0.20281 | *IL17RA* |
| ENSG00000117586 | 1 | 173142873 | 173186452 | 130 | 20 | 39978 | 0.83149 | 0.20285 | *TNFSF4* |
| ENSG00000095203 | 9 | 111924255 | 112093244 | 466 | 86 | 44173 | 0.83146 | 0.20286 | *EPB41L4B* |
| ENSG00000197776 | 14 | 50149823 | 50229870 | 103 | 21 | 45914 | 0.83112 | 0.20295 | *KLHDC1* |
| ENSG00000135472 | 12 | 50250679 | 50308000 | 222 | 44 | 47690 | 0.83109 | 0.20296 | *FAIM2* |
| ENSG00000187959 | 17 | 71234588 | 71268491 | 112 | 13 | 45302 | 0.8308 | 0.20304 | *CPSF4L* |
| ENSG00000013561 | 5 | 141327893 | 141379856 | 204 | 21 | 46945 | 0.83061 | 0.2031 | *RNF14* |
| ENSG00000107159 | 9 | 35663853 | 35691156 | 74 | 19 | 49443 | 0.83036 | 0.20317 | *CA9* |
| ENSG00000172938 | 11 | 68737490 | 68758455 | 67 | 15 | 44492 | 0.83033 | 0.20318 | *MRGPRD* |
| ENSG00000121716 | 7 | 99923737 | 99975356 | 103 | 10 | 52755 | 0.83006 | 0.20325 | *PILRB* |

| | | | | | | | | | |
|---|---|---|---|---|---|---|---|---|---|
| ENSG00000129244 | 17 | 7539945 | 7571086 | 141 | 11 | 28871 | 0.82953 | 0.2034 | *ATP1B2* |
| ENSG00000145391 | 4 | 140407095 | 140537853 | 375 | 76 | 49574 | 0.82894 | 0.20357 | *SETD7* |
| ENSG00000075826 | 10 | 102236399 | 102299628 | 135 | 8 | 56351 | 0.82895 | 0.20357 | *SEC31B* |
| ENSG00000142207 | 21 | 33673329 | 33775335 | 265 | 21 | 34592 | 0.82851 | 0.20369 | *URB1* |
| ENSG00000169727 | 17 | 79998567 | 80025346 | 83 | 19 | 38042 | 0.82833 | 0.20374 | *GPS1* |
| ENSG00000215217 | 5 | 7820491 | 7861603 | 133 | 23 | 44222 | 0.82826 | 0.20376 | *C5orf49* |
| ENSG00000185246 | 14 | 45543302 | 45595485 | 93 | 11 | 33203 | 0.82819 | 0.20378 | *PRPF39* |
| ENSG00000138944 | 22 | 44629547 | 44718731 | 372 | 85 | 44962 | 0.8278 | 0.20389 | *KIAA1644* |
| ENSG00000164122 | 4 | 177124824 | 177208722 | 355 | 41 | 48842 | 0.82747 | 0.20399 | *ASB5* |
| ENSG00000245317 | 5 | 179276013 | 179298283 | 71 | 14 | 38576 | 0.8272 | 0.20406 | *CTC-241N9.1* |
| ENSG00000253731 | 5 | 140743651 | 140902546 | 448 | 53 | 43870 | 0.82705 | 0.2041 | *PCDHGA6* |
| ENSG00000115806 | 2 | 171774974 | 171833639 | 135 | 15 | 26526 | 0.82696 | 0.20413 | *GORASP2* |
| ENSG00000161513 | 17 | 72848619 | 72879156 | 71 | 9 | 35557 | 0.82696 | 0.20413 | *FDXR* |
| ENSG00000033100 | 7 | 150919575 | 150945908 | 73 | 13 | 56916 | 0.82693 | 0.20414 | *CHPF2* |
| ENSG00000126353 | 17 | 38700021 | 38731724 | 67 | 24 | 44688 | 0.8267 | 0.2042 | *CCR7* |
| ENSG00000183309 | 8 | 144708183 | 144745900 | 96 | 22 | 36514 | 0.8266 | 0.20423 | *ZNF623* |
| ENSG00000139289 | 12 | 76409227 | 76437712 | 54 | 18 | 38743 | 0.8264 | 0.20429 | *PHLDA1* |
| ENSG00000144406 | 2 | 210626717 | 210874024 | 530 | 52 | 33300 | 0.82629 | 0.20432 | *UNC80* |
| ENSG00000125648 | 19 | 6426090 | 6475214 | 191 | 32 | 44322 | 0.8262 | 0.20435 | *SLC25A23* |
| ENSG00000157764 | 7 | 140409127 | 140634564 | 431 | 27 | 32638 | 0.82615 | 0.20436 | *BRAF* |
| ENSG00000068885 | 3 | 159964774 | 160127668 | 274 | 37 | 49544 | 0.82586 | 0.20444 | *IFT80* |
| ENSG00000165115 | 9 | 86441613 | 86546342 | 254 | 19 | 50655 | 0.82586 | 0.20444 | *KIF27* |
| ENSG00000132535 | 17 | 7083209 | 7133021 | 88 | 25 | 46990 | 0.8257 | 0.20449 | *DLG4* |
| ENSG00000111907 | 6 | 125430195 | 125595553 | 541 | 72 | 42454 | 0.82556 | 0.20453 | *TPD52L1* |
| ENSG00000101331 | 20 | 30588245 | 30629984 | 84 | 15 | 42904 | 0.8254 | 0.20457 | *CCM2L* |
| ENSG00000225556 | 1 | 151800295 | 151823033 | 63 | 17 | 43777 | 0.82458 | 0.20481 | *C2CD4D* |
| ENSG00000152520 | 13 | 28702643 | 28879475 | 360 | 34 | 40557 | 0.82453 | 0.20482 | *PAN3* |
| ENSG00000127481 | 1 | 19391000 | 19546770 | 350 | 34 | 54271 | 0.8245 | 0.20483 | *UBR4* |
| ENSG00000137100 | 9 | 34603548 | 34630515 | 51 | 12 | 49676 | 0.82425 | 0.2049 | *DCTN3* |
| ENSG00000119541 | 18 | 61046423 | 61099716 | 111 | 16 | 38855 | 0.82426 | 0.2049 | *VPS4B* |
| ENSG00000143727 | 2 | 254140 | 288283 | 73 | 10 | 51874 | 0.82417 | 0.20492 | *ACP1* |
| ENSG00000120942 | 1 | 11323263 | 11366106 | 99 | 15 | 33575 | 0.82377 | 0.20504 | *UBIAD1* |
| ENSG00000136732 | 2 | 127403509 | 127464246 | 278 | 46 | 46891 | 0.82377 | 0.20504 | *GYPC* |
| ENSG00000181007 | 19 | 36864022 | 36919558 | 156 | 15 | 51206 | 0.82354 | 0.2051 | *ZFP82* |
| ENSG00000205301 | 4 | 141353416 | 141429531 | 255 | 25 | 51827 | 0.8235 | 0.20511 | *RP11-542P2.1* |
| ENSG00000198598 | 12 | 132302938 | 132346328 | 176 | 43 | 40114 | 0.82331 | 0.20516 | *MMP17* |

| | | | | | | | | | |
|---|---|---|---|---|---|---|---|---|---|
| ENSG00000118960 | 2 | 20750208 | 20860849 | 482 | 74 | 48104 | 0.82321 | 0.20519 | *HS1BP3* |
| ENSG00000183307 | 22 | 17587189 | 17612257 | 109 | 25 | 42849 | 0.82323 | 0.20519 | *CECR6* |
| ENSG00000197182 | 22 | 46439749 | 46519808 | 232 | 53 | 44092 | 0.82251 | 0.20539 | *FLJ27365* |
| ENSG00000186777 | 4 | 254464 | 309110 | 225 | 23 | 52559 | 0.82242 | 0.20542 | *ZNF732* |
| ENSG00000137942 | 1 | 93903688 | 94030218 | 182 | 36 | 42004 | 0.82214 | 0.2055 | *FNBP1L* |
| ENSG00000267909 | 14 | 70028216 | 70050339 | 48 | 10 | 44589 | 0.82178 | 0.2056 | *CCDC177* |
| ENSG00000090382 | 12 | 69732121 | 69758014 | 108 | 17 | 39395 | 0.82162 | 0.20565 | *LYZ* |
| ENSG00000095110 | 11 | 114382437 | 114440617 | 187 | 11 | 31758 | 0.82159 | 0.20566 | *NXPE1* |
| ENSG00000128510 | 7 | 129922974 | 129974020 | 162 | 29 | 53062 | 0.82147 | 0.20569 | *CPA4* |
| ENSG00000240053 | 6 | 31627944 | 31651553 | 84 | 20 | 38995 | 0.82142 | 0.2057 | *LY6G5B* |
| ENSG00000169604 | 2 | 69230310 | 69486459 | 639 | 107 | 41338 | 0.82137 | 0.20572 | *ANTXR1* |
| ENSG00000188219 | 2 | 131965882 | 132032965 | 33 | 11 | 39658 | 0.82131 | 0.20573 | *POTEE* |
| ENSG00000106689 | 9 | 126753949 | 126805580 | 143 | 28 | 43923 | 0.82117 | 0.20578 | *LHX2* |
| ENSG00000115317 | 2 | 74746504 | 74770472 | 16 | 9 | 40808 | 0.8211 | 0.2058 | *HTRA2* |
| ENSG00000196517 | 1 | 44447172 | 44507139 | 115 | 32 | 39202 | 0.82092 | 0.20585 | *SLC6A9* |
| ENSG00000186496 | 18 | 32936661 | 32967301 | 63 | 14 | 37434 | 0.82091 | 0.20585 | *ZNF396* |
| ENSG00000183474 | 5 | 68846035 | 68900550 | 8 | 5 | 38855 | 0.82037 | 0.206 | *GTF2H2C* |
| ENSG00000187778 | 12 | 49940327 | 49971936 | 60 | 8 | 35040 | 0.82039 | 0.206 | *MCRS1* |
| ENSG00000181929 | 12 | 49386057 | 49422980 | 47 | 10 | 50989 | 0.82019 | 0.20605 | *PRKAG1* |
| ENSG00000179626 | 12 | 55934983 | 55956028 | 49 | 13 | 48637 | 0.82013 | 0.20607 | *OR6C4* |
| ENSG00000186895 | 11 | 69614992 | 69643792 | 130 | 18 | 52516 | 0.81995 | 0.20612 | *FGF3* |
| ENSG00000083444 | 1 | 11984262 | 12045595 | 174 | 24 | 43261 | 0.81982 | 0.20616 | *PLOD1* |
| ENSG00000152527 | 2 | 43854412 | 44005126 | 655 | 58 | 46705 | 0.81982 | 0.20616 | *PLEKHH2* |
| ENSG00000241635 | 2 | 234516291 | 234691956 | 655 | 64 | 46566 | 0.81964 | 0.20621 | *UGT1A8* |
| ENSG00000185888 | 1 | 227993394 | 228044171 | 199 | 34 | 42197 | 0.81957 | 0.20623 | *PRSS38* |
| ENSG00000268964 | 19 | 53537991 | 53564329 | 60 | 10 | 51202 | 0.81927 | 0.20632 | *ERVV-2* |
| ENSG00000174015 | 13 | 46266446 | 46298694 | 98 | 20 | 43599 | 0.81915 | 0.20635 | *SPERT* |
| ENSG00000204657 | 6 | 29545683 | 29566745 | 70 | 17 | 32787 | 0.81903 | 0.20639 | *OR2H2* |
| ENSG00000155368 | 2 | 120114497 | 120140126 | 128 | 14 | 53761 | 0.81874 | 0.20647 | *DBI* |
| ENSG00000142676 | 1 | 24008269 | 24032915 | 58 | 11 | 23370 | 0.81862 | 0.2065 | *RPL11* |
| ENSG00000149516 | 11 | 59814060 | 59848601 | 127 | 15 | 46074 | 0.81862 | 0.2065 | *MS4A3* |
| ENSG00000132950 | 13 | 20387622 | 20447776 | 212 | 11 | 45619 | 0.81862 | 0.2065 | *ZMYM5* |
| ENSG00000171988 | 10 | 64916981 | 65235722 | 789 | 50 | 37781 | 0.81826 | 0.2066 | *JMJD1C* |
| ENSG00000198797 | 1 | 177130633 | 177261558 | 287 | 39 | 44066 | 0.81819 | 0.20663 | *BRINP2* |
| ENSG00000162148 | 11 | 61238592 | 61268403 | 86 | 14 | 35504 | 0.81802 | 0.20667 | *PPP1R32* |
| ENSG00000187800 | 1 | 156853490 | 156896226 | 117 | 15 | 46361 | 0.81742 | 0.20684 | *PEAR1* |

| | | | | | | | | | |
|---|---|---|---|---|---|---|---|---|---|
| ENSG00000148690 | 10 | 95417640 | 95472329 | 210 | 18 | 38959 | 0.81733 | 0.20687 | *FRA10AC1* |
| ENSG00000136327 | 14 | 37039784 | 37061812 | 79 | 9 | 39447 | 0.81734 | 0.20687 | *NKX2-8* |
| ENSG00000204899 | 13 | 73272495 | 73311825 | 81 | 19 | 48014 | 0.81718 | 0.20691 | *MZT1* |
| ENSG00000180370 | 3 | 196456728 | 196569518 | 470 | 61 | 41117 | 0.8171 | 0.20694 | *PAK2* |
| ENSG00000158467 | 7 | 128854864 | 129080052 | 504 | 44 | 44635 | 0.81703 | 0.20696 | *AHCYL2* |
| ENSG00000120526 | 8 | 110243148 | 110356614 | 298 | 23 | 47828 | 0.81677 | 0.20703 | *NUDCD1* |
| ENSG00000262628 | 17 | 2955963 | 2976901 | 13 | 7 | 31230 | 0.81669 | 0.20705 | *OR1D5* |
| ENSG00000130413 | 11 | 8403418 | 8625836 | 841 | 54 | 47607 | 0.81657 | 0.20709 | *STK33* |
| ENSG00000175206 | 1 | 11895766 | 11918402 | 111 | 19 | 48214 | 0.81646 | 0.20712 | *NPPA* |
| ENSG00000074416 | 3 | 127397909 | 127552051 | 454 | 86 | 49466 | 0.81644 | 0.20712 | *MGLL* |
| ENSG00000167721 | 17 | 2215797 | 2250801 | 67 | 19 | 50247 | 0.81629 | 0.20717 | *TSR1* |
| ENSG00000162494 | 1 | 13791445 | 13850543 | 263 | 61 | 44737 | 0.81612 | 0.20722 | *LRRC38* |
| ENSG00000070501 | 8 | 42185972 | 42239326 | 105 | 14 | 37329 | 0.81608 | 0.20723 | *POLB* |
| ENSG00000198466 | 19 | 58351225 | 58386480 | 132 | 20 | 34551 | 0.81602 | 0.20724 | *ZNF587* |
| ENSG00000166349 | 11 | 36522259 | 36624706 | 242 | 33 | 48934 | 0.81589 | 0.20728 | *RAG1* |
| ENSG00000137055 | 9 | 26894081 | 26957461 | 223 | 23 | 46383 | 0.81563 | 0.20735 | *PLAA* |
| ENSG00000111652 | 12 | 6822907 | 6851041 | 146 | 23 | 47335 | 0.81537 | 0.20743 | *COPS7A* |
| ENSG00000168807 | 16 | 69211032 | 69352955 | 339 | 34 | 55376 | 0.81528 | 0.20746 | *SNTB2* |
| ENSG00000160785 | 1 | 156153880 | 156192587 | 104 | 11 | 36976 | 0.81481 | 0.20759 | *SLC25A44* |
| ENSG00000124440 | 19 | 46790303 | 46856690 | 307 | 42 | 49951 | 0.81473 | 0.20761 | *HIF3A* |
| ENSG00000157538 | 21 | 38585721 | 38650262 | 177 | 30 | 53862 | 0.81465 | 0.20764 | *DSCR3* |
| ENSG00000144827 | 3 | 111687857 | 111722210 | 178 | 19 | 53501 | 0.8146 | 0.20765 | *ABHD10* |
| ENSG00000204348 | 6 | 31927587 | 31950069 | 85 | 23 | 39553 | 0.81457 | 0.20766 | *DXO* |
| ENSG00000127220 | 19 | 17392940 | 17431045 | 186 | 21 | 43945 | 0.81452 | 0.20767 | *ABHD8* |
| ENSG00000153037 | 5 | 112186919 | 112215485 | 117 | 20 | 39928 | 0.81427 | 0.20775 | *SRP19* |
| ENSG00000007171 | 17 | 26073792 | 26137525 | 218 | 35 | 40542 | 0.81422 | 0.20776 | *NOS2* |
| ENSG00000029364 | 14 | 69854732 | 69939105 | 217 | 25 | 54559 | 0.81389 | 0.20785 | *SLC39A9* |
| ENSG00000259752 | 15 | 47415693 | 47436320 | 54 | 11 | 53274 | 0.81344 | 0.20798 | *FKSG62* |
| ENSG00000177103 | 11 | 117288489 | 117698240 | 1604 | 210 | 44942 | 0.81317 | 0.20806 | *DSCAML1* |
| ENSG00000011422 | 19 | 44140247 | 44184699 | 149 | 32 | 47354 | 0.81299 | 0.20811 | *PLAUR* |
| ENSG00000100150 | 22 | 32139944 | 32313012 | 529 | 45 | 51072 | 0.81286 | 0.20815 | *DEPDC5* |
| ENSG00000262576 | 5 | 140724768 | 140902546 | 492 | 55 | 43373 | 0.81278 | 0.20817 | *PCDHGA4* |
| ENSG00000112297 | 6 | 106949730 | 107028326 | 424 | 39 | 45370 | 0.81263 | 0.20821 | *AIM1* |
| ENSG00000121807 | 3 | 46385225 | 46412419 | 87 | 12 | 37525 | 0.81258 | 0.20823 | *CCR2* |
| ENSG00000172915 | 13 | 35506424 | 36257159 | 2252 | 130 | 49486 | 0.81239 | 0.20828 | *NBEA* |
| ENSG00000177374 | 17 | 1947448 | 1972981 | 68 | 18 | 47634 | 0.8121 | 0.20837 | *HIC1* |

| | | | | | | | | | |
|---|---|---|---|---|---|---|---|---|---|
| ENSG00000182611 | 6 | 27772112 | 27792607 | 51 | 11 | 41443 | 0.81202 | 0.20839 | *HIST1H2AJ* |
| ENSG00000196730 | 9 | 90102143 | 90333548 | 896 | 110 | 43469 | 0.812 | 0.2084 | *DAPK1* |
| ENSG00000253910 | 5 | 140729703 | 140902546 | 483 | 55 | 43573 | 0.81188 | 0.20843 | *PCDHGB2* |
| ENSG00000181785 | 11 | 55787895 | 55808869 | 209 | 8 | 54143 | 0.81179 | 0.20846 | *OR5AS1* |
| ENSG00000157827 | 2 | 153181751 | 153516348 | 1097 | 108 | 44083 | 0.81166 | 0.20849 | *FMNL2* |
| ENSG00000115468 | 2 | 233460767 | 233557491 | 311 | 51 | 47198 | 0.81169 | 0.20849 | *EFHD1* |
| ENSG00000134698 | 1 | 36263773 | 36333491 | 77 | 17 | 44228 | 0.8116 | 0.20851 | *AGO4* |
| ENSG00000198624 | 5 | 150550613 | 150613706 | 287 | 56 | 47262 | 0.81146 | 0.20855 | *CCDC69* |
| ENSG00000100479 | 14 | 50100273 | 50165140 | 152 | 14 | 42205 | 0.81135 | 0.20858 | *POLE2* |
| ENSG00000171169 | 9 | 130813512 | 130840485 | 53 | 7 | 31646 | 0.81127 | 0.2086 | *NAIF1* |
| ENSG00000197701 | 4 | 43179 | 98099 | 95 | 15 | 43856 | 0.8111 | 0.20866 | *ZNF595* |
| ENSG00000132872 | 18 | 40837843 | 40867615 | 55 | 15 | 46482 | 0.81097 | 0.20869 | *SYT4* |
| ENSG00000115274 | 2 | 74672150 | 74698011 | 8 | 5 | 44542 | 0.81077 | 0.20875 | *INO80B* |
| ENSG00000204435 | 6 | 31623013 | 31648120 | 88 | 20 | 36804 | 0.81062 | 0.20879 | *CSNK2B* |
| ENSG00000172201 | 6 | 19827617 | 19850915 | 84 | 23 | 47523 | 0.81046 | 0.20884 | *ID4* |
| ENSG00000079387 | 12 | 48426681 | 48510091 | 263 | 24 | 42978 | 0.81035 | 0.20887 | *SENP1* |
| ENSG00000175356 | 11 | 9031071 | 9169661 | 398 | 67 | 48732 | 0.80953 | 0.2091 | *SCUBE2* |
| ENSG00000067167 | 8 | 71475677 | 71530622 | 133 | 19 | 42773 | 0.80934 | 0.20916 | *TRAM1* |
| ENSG00000213139 | 3 | 186246230 | 186274491 | 37 | 11 | 37845 | 0.80923 | 0.20919 | *CRYGS* |
| ENSG00000119397 | 9 | 123827141 | 123949888 | 287 | 24 | 38517 | 0.80923 | 0.20919 | *CNTRL* |
| ENSG00000148291 | 9 | 136213428 | 136238045 | 72 | 10 | 50599 | 0.80879 | 0.20932 | *SURF2* |
| ENSG00000225362 | 15 | 71392583 | 71417833 | 50 | 9 | 30637 | 0.80878 | 0.20932 | *CT62* |
| ENSG00000204463 | 6 | 31596805 | 31630482 | 135 | 25 | 42524 | 0.80872 | 0.20934 | *BAG6* |
| ENSG00000197713 | 2 | 210857289 | 210896300 | 46 | 15 | 42673 | 0.80799 | 0.20955 | *RPE* |
| ENSG00000261934 | 5 | 140772520 | 140902546 | 367 | 46 | 43108 | 0.80791 | 0.20957 | *PCDHGA9* |
| ENSG00000178537 | 3 | 48884369 | 48946426 | 67 | 12 | 35860 | 0.8075 | 0.20969 | *SLC25A20* |
| ENSG00000134717 | 1 | 52511797 | 52566388 | 84 | 17 | 47519 | 0.80736 | 0.20973 | *BTF3L4* |
| ENSG00000112812 | 6 | 27205480 | 27234403 | 86 | 16 | 40738 | 0.80681 | 0.20989 | *PRSS16* |
| ENSG00000171241 | 16 | 46604466 | 46665538 | 50 | 27 | 45561 | 0.80677 | 0.2099 | *SHCBP1* |
| ENSG00000198718 | 14 | 45421411 | 45553634 | 271 | 21 | 37021 | 0.80673 | 0.20991 | *FAM179B* |
| ENSG00000172377 | 11 | 57875972 | 57896916 | 65 | 10 | 39696 | 0.80648 | 0.20998 | *OR9I1* |
| ENSG00000135341 | 6 | 91213292 | 91306764 | 315 | 28 | 55170 | 0.80644 | 0.20999 | *MAP3K7* |
| ENSG00000169507 | 2 | 165742696 | 165822035 | 265 | 28 | 43913 | 0.80625 | 0.21005 | *SLC38A11* |
| ENSG00000262209 | 5 | 140739831 | 140902546 | 456 | 53 | 43621 | 0.8062 | 0.21006 | *PCDHGB3* |
| ENSG00000184619 | 17 | 8261955 | 8290029 | 81 | 14 | 54007 | 0.8062 | 0.21006 | *KRBA2* |
| ENSG00000118515 | 6 | 134480384 | 134649250 | 528 | 93 | 46677 | 0.80618 | 0.21007 | *SGK1* |

| | | | | | | | | | |
|---|---|---|---|---|---|---|---|---|---|
| ENSG00000185219 | 3 | 44471262 | 44529162 | 92 | 17 | 39319 | 0.80615 | 0.21008 | *ZNF445* |
| ENSG00000136044 | 12 | 105557074 | 105640016 | 301 | 33 | 50365 | 0.80609 | 0.2101 | *APPL2* |
| ENSG00000204569 | 6 | 30558177 | 30596389 | 113 | 8 | 24389 | 0.80594 | 0.21014 | *PPP1R10* |
| ENSG00000226364 | 17 | 50929481 | 50986948 | 323 | 27 | 46082 | 0.80595 | 0.21014 | *AC102948.2* |
| ENSG00000184682 | 11 | 1900375 | 1922084 | 73 | 20 | 42646 | 0.80564 | 0.21023 | *C11orf89* |
| ENSG00000131398 | 19 | 50805194 | 50846772 | 97 | 31 | 46736 | 0.80547 | 0.21027 | *KCNC3* |
| ENSG00000149133 | 11 | 55751157 | 55772101 | 182 | 9 | 51957 | 0.80503 | 0.2104 | *OR5F1* |
| ENSG00000124391 | 16 | 88694999 | 88716881 | 103 | 34 | 44374 | 0.80485 | 0.21045 | *IL17C* |
| ENSG00000253250 | 8 | 91960865 | 92007485 | 85 | 14 | 38541 | 0.80483 | 0.21046 | *C8orf88* |
| ENSG00000083844 | 19 | 57692868 | 57734724 | 132 | 20 | 49381 | 0.8043 | 0.21061 | *ZNF264* |
| ENSG00000089847 | 19 | 4173351 | 4234811 | 210 | 41 | 44360 | 0.80415 | 0.21065 | *ANKRD24* |
| ENSG00000166126 | 14 | 103378993 | 103409933 | 88 | 29 | 45748 | 0.80406 | 0.21068 | *AMN* |
| ENSG00000136371 | 15 | 80115927 | 80199721 | 301 | 35 | 52877 | 0.80405 | 0.21068 | *MTHFS* |
| ENSG00000213719 | 6 | 31688358 | 31717540 | 69 | 21 | 34637 | 0.80399 | 0.2107 | *CLIC1* |
| ENSG00000170616 | 8 | 145544228 | 145569943 | 37 | 13 | 44668 | 0.80391 | 0.21072 | *SCRT1* |
| ENSG00000103174 | 16 | 5064845 | 5094142 | 100 | 21 | 45233 | 0.80392 | 0.21072 | *NAGPA* |
| ENSG00000079841 | 6 | 72586406 | 73122845 | 1566 | 84 | 47314 | 0.80387 | 0.21074 | *RIMS1* |
| ENSG00000105671 | 19 | 19020484 | 19049436 | 64 | 18 | 39521 | 0.8036 | 0.21081 | *DDX49* |
| ENSG00000179930 | 1 | 182013705 | 182040847 | 74 | 9 | 37633 | 0.80343 | 0.21086 | *ZNF648* |
| ENSG00000144426 | 2 | 203869602 | 204101101 | 381 | 36 | 38096 | 0.80345 | 0.21086 | *NBEAL1* |
| ENSG00000240303 | 3 | 132266982 | 132389567 | 240 | 32 | 38930 | 0.80328 | 0.21091 | *ACAD11* |
| ENSG00000061337 | 8 | 20093676 | 20171474 | 309 | 68 | 46812 | 0.80306 | 0.21097 | *LZTS1* |
| ENSG00000167825 | 11 | 55692932 | 55713876 | 156 | 7 | 56296 | 0.80305 | 0.21097 | *OR5I1* |
| ENSG00000173040 | 4 | 5534499 | 5721275 | 777 | 103 | 42262 | 0.80297 | 0.211 | *EVC2* |
| ENSG00000092200 | 14 | 21746098 | 21829460 | 267 | 41 | 48447 | 0.80292 | 0.21101 | *RPGRIP1* |
| ENSG00000116273 | 1 | 6663745 | 6694093 | 63 | 11 | 56282 | 0.80288 | 0.21102 | *PHF13* |
| ENSG00000180066 | 10 | 134247831 | 134272912 | 67 | 18 | 39779 | 0.80285 | 0.21103 | *C10orf91* |
| ENSG00000245680 | 19 | 37662481 | 37719055 | 95 | 17 | 47063 | 0.80282 | 0.21104 | *ZNF585B* |
| ENSG00000169071 | 9 | 94315373 | 94722444 | 1669 | 150 | 43818 | 0.80251 | 0.21113 | *ROR2* |
| ENSG00000179603 | 7 | 126068652 | 126903348 | 2479 | 144 | 48392 | 0.80243 | 0.21115 | *GRM8* |
| ENSG00000163382 | 1 | 156551554 | 156574091 | 101 | 7 | 21417 | 0.80222 | 0.21121 | *APOA1BP* |
| ENSG00000101746 | 18 | 31421064 | 31814916 | 939 | 104 | 53192 | 0.80221 | 0.21122 | *NOL4* |
| ENSG00000204347 | 17 | 72342555 | 72368085 | 67 | 15 | 45950 | 0.8021 | 0.21125 | *BTBD17* |
| ENSG00000134317 | 2 | 10075341 | 10152411 | 416 | 47 | 46779 | 0.80198 | 0.21128 | *GRHL1* |
| ENSG00000164045 | 3 | 48188636 | 48239892 | 83 | 13 | 46026 | 0.802 | 0.21128 | *CDC25A* |
| ENSG00000213064 | 1 | 168185176 | 168232263 | 101 | 14 | 37280 | 0.80193 | 0.2113 | *SFT2D2* |

| | | | | | | | | | |
|---|---|---|---|---|---|---|---|---|---|
| ENSG00000168427 | 2 | 239037363 | 239071588 | 134 | 34 | 49182 | 0.8019 | 0.21131 | *KLHL30* |
| ENSG00000082805 | 12 | 1089675 | 1615099 | 2053 | 103 | 42616 | 0.80161 | 0.21139 | *ERC1* |
| ENSG00000138615 | 15 | 65478337 | 65513826 | 53 | 9 | 39923 | 0.80149 | 0.21142 | *CILP* |
| ENSG00000118514 | 6 | 135228528 | 135281260 | 112 | 16 | 37879 | 0.8013 | 0.21148 | *ALDH8A1* |
| ENSG00000076864 | 1 | 21912708 | 22005801 | 397 | 82 | 49524 | 0.80089 | 0.2116 | *RAP1GAP* |
| ENSG00000178997 | 15 | 41464923 | 41532941 | 293 | 17 | 29223 | 0.80073 | 0.21164 | *EXD1* |
| ENSG00000031081 | 3 | 119003220 | 119149561 | 574 | 90 | 46167 | 0.8006 | 0.21168 | *ARHGAP31* |
| ENSG00000143158 | 1 | 167875967 | 167916278 | 72 | 11 | 41592 | 0.80029 | 0.21177 | *MPC2* |
| ENSG00000156802 | 8 | 124322090 | 124438590 | 209 | 36 | 41880 | 0.80031 | 0.21177 | *ATAD2* |
| ENSG00000253305 | 5 | 140777770 | 140902546 | 350 | 44 | 43292 | 0.80016 | 0.21181 | *PCDHGB6* |
| ENSG00000171222 | 20 | 34531539 | 34557394 | 64 | 10 | 30813 | 0.79987 | 0.21189 | *SCAND1* |
| ENSG00000168216 | 6 | 70375694 | 70517003 | 464 | 21 | 53643 | 0.79968 | 0.21195 | *LMBRD1* |
| ENSG00000135093 | 12 | 109450894 | 109535831 | 159 | 34 | 47420 | 0.79929 | 0.21206 | *USP30* |
| ENSG00000106682 | 7 | 73578575 | 73621431 | 63 | 13 | 47198 | 0.79919 | 0.21209 | *EIF4H* |
| ENSG00000197262 | 17 | 34629793 | 34651846 | 1 | 1 | 42834 | 0.79916 | 0.2121 | *CCL4L2* |
| ENSG00000033327 | 11 | 77916343 | 78139394 | 464 | 16 | 24966 | 0.79903 | 0.21214 | *GAB2* |
| ENSG00000197943 | 16 | 81762702 | 82001899 | 1532 | 187 | 46963 | 0.79891 | 0.21217 | *PLCG2* |
| ENSG00000173744 | 2 | 228326868 | 228435930 | 429 | 36 | 55404 | 0.79881 | 0.2122 | *AGFG1* |
| ENSG00000221937 | 7 | 142909130 | 142930162 | 24 | 7 | 45268 | 0.79869 | 0.21224 | *TAS2R40* |
| ENSG00000254873 | 11 | 118257810 | 118282610 | 49 | 11 | 41086 | 0.79843 | 0.21231 | *RP11-770J1.5* |
| ENSG00000185823 | 15 | 24910541 | 24938593 | 168 | 20 | 50801 | 0.79827 | 0.21236 | *NPAP1* |
| ENSG00000152034 | 6 | 100357786 | 100452123 | 277 | 37 | 51363 | 0.79789 | 0.21247 | *MCHR2* |
| ENSG00000237693 | 5 | 150216085 | 150290298 | 287 | 12 | 28092 | 0.79774 | 0.21251 | *IRGM* |
| ENSG00000088727 | 3 | 47259516 | 47334941 | 111 | 18 | 41498 | 0.79765 | 0.21254 | *KIF9* |
| ENSG00000153823 | 2 | 229705242 | 230146001 | 1557 | 171 | 48280 | 0.79741 | 0.21261 | *PID1* |
| ENSG00000269679 | 18 | 77265812 | 77286057 | 118 | 18 | 44775 | 0.7974 | 0.21261 | *AC018445.1* |
| ENSG00000166228 | 10 | 72632037 | 72658541 | 95 | 23 | 50426 | 0.79703 | 0.21272 | *PCBD1* |
| ENSG00000137960 | 1 | 78435226 | 78614133 | 523 | 49 | 46971 | 0.79694 | 0.21274 | *GIPC2* |
| ENSG00000183313 | 11 | 5997122 | 6018215 | 76 | 21 | 53884 | 0.79693 | 0.21275 | *OR52L1* |
| ENSG00000176895 | 11 | 4918600 | 4939538 | 75 | 17 | 40480 | 0.79689 | 0.21276 | *OR51A7* |
| ENSG00000167772 | 19 | 8418173 | 8449257 | 88 | 16 | 53513 | 0.79687 | 0.21276 | *ANGPTL4* |
| ENSG00000181938 | 16 | 58318984 | 58450048 | 516 | 67 | 44309 | 0.79682 | 0.21278 | *GINS3* |
| ENSG00000183773 | 22 | 21309396 | 21345649 | 92 | 28 | 41093 | 0.79676 | 0.21279 | *AIFM3* |
| ENSG00000196199 | 13 | 20197788 | 20257599 | 139 | 16 | 47363 | 0.79674 | 0.2128 | *MPHOSPH8* |
| ENSG00000171466 | 19 | 9749330 | 9795776 | 123 | 14 | 36033 | 0.79673 | 0.21281 | *ZNF562* |
| ENSG00000165949 | 14 | 94561182 | 94593033 | 123 | 24 | 34941 | 0.79648 | 0.21288 | *IFI27* |

| | | | | | | | | | |
|---|---|---|---|---|---|---|---|---|---|
| ENSG00000250361 | 4 | 144907257 | 145071844 | 611 | 52 | 47130 | 0.79631 | 0.21293 | *GYPB* |
| ENSG00000204262 | 2 | 189886622 | 190054605 | 518 | 32 | 43377 | 0.79609 | 0.21299 | *COL5A2* |
| ENSG00000225921 | 6 | 13605559 | 13642971 | 71 | 15 | 53157 | 0.79593 | 0.21304 | *NOL7* |
| ENSG00000261732 | 16 | 1672260 | 1746716 | 144 | 17 | 38830 | 0.79589 | 0.21305 | *LA16c-431H6.6* |
| ENSG00000254709 | 22 | 23219960 | 23248287 | 29 | 13 | 39677 | 0.79563 | 0.21312 | *IGLL5* |
| ENSG00000167619 | 19 | 42807477 | 42839214 | 33 | 7 | 51210 | 0.79561 | 0.21313 | *TMEM145* |
| ENSG00000155621 | 9 | 74516426 | 74610970 | 170 | 32 | 42691 | 0.79516 | 0.21326 | *C9orf85* |
| ENSG00000022976 | 14 | 102773714 | 102819044 | 123 | 24 | 39514 | 0.79473 | 0.21338 | *ZNF839* |
| ENSG00000140807 | 16 | 50572241 | 50680647 | 164 | 26 | 37791 | 0.79457 | 0.21343 | *NKD1* |
| ENSG00000101473 | 20 | 44460360 | 44496045 | 127 | 24 | 45573 | 0.79455 | 0.21344 | *ACOT8* |
| ENSG00000253485 | 5 | 140733898 | 140902546 | 470 | 54 | 43470 | 0.79436 | 0.21349 | *PCDHGA5* |
| ENSG00000063244 | 19 | 56155512 | 56196081 | 123 | 20 | 39923 | 0.79406 | 0.21358 | *U2AF2* |
| ENSG00000111665 | 12 | 6943957 | 6971230 | 41 | 13 | 42041 | 0.79398 | 0.2136 | *CDCA3* |
| ENSG00000188038 | 16 | 67908708 | 67932758 | 39 | 10 | 50793 | 0.79394 | 0.21361 | *NRN1L* |
| ENSG00000196409 | 9 | 40750700 | 40846415 | 54 | 7 | 43543 | 0.79386 | 0.21364 | *ZNF658* |
| ENSG00000250510 | 12 | 6920711 | 6949136 | 105 | 29 | 36258 | 0.79377 | 0.21366 | *GPR162* |
| ENSG00000242689 | 11 | 58380146 | 58403198 | 50 | 10 | 53092 | 0.7937 | 0.21369 | *CNTF* |
| ENSG00000162825 | 1 | 144136808 | 144234481 | 2 | 1 | 42552 | 0.79321 | 0.21383 | *NBPF8* |
| ENSG00000116685 | 1 | 11969648 | 11996485 | 105 | 19 | 53175 | 0.7931 | 0.21386 | *KIAA2013* |
| ENSG00000115364 | 2 | 75863909 | 75927977 | 210 | 28 | 45970 | 0.79308 | 0.21387 | *MRPL19* |
| ENSG00000166716 | 15 | 85281866 | 85359659 | 205 | 27 | 45805 | 0.79306 | 0.21387 | *ZNF592* |
| ENSG00000047849 | 3 | 47882182 | 48140769 | 326 | 25 | 40513 | 0.79297 | 0.2139 | *MAP4* |
| ENSG00000253873 | 5 | 140790762 | 140901835 | 312 | 41 | 43454 | 0.79277 | 0.21396 | *PCDHGA11* |
| ENSG00000186185 | 17 | 42992077 | 43035082 | 149 | 31 | 50586 | 0.79247 | 0.21404 | *KIF18B* |
| ENSG00000140993 | 16 | 3338832 | 3365729 | 41 | 10 | 38696 | 0.79243 | 0.21405 | *TIGD7* |
| ENSG00000255073 | 11 | 58336645 | 58402112 | 136 | 20 | 52790 | 0.7922 | 0.21412 | *ZFP91-CNTF* |
| ENSG00000138646 | 4 | 89368268 | 89437314 | 62 | 24 | 50892 | 0.79204 | 0.21417 | *HERC5* |
| ENSG00000179840 | 1 | 9702668 | 9724644 | 54 | 8 | 28882 | 0.79195 | 0.2142 | *C1orf200* |
| ENSG00000126878 | 9 | 133961863 | 134008539 | 157 | 28 | 40451 | 0.79191 | 0.21421 | *AIF1L* |
| ENSG00000140474 | 15 | 75118457 | 75145687 | 44 | 10 | 35895 | 0.79178 | 0.21424 | *ULK3* |
| ENSG00000105672 | 19 | 36122647 | 36145773 | 54 | 15 | 37994 | 0.79178 | 0.21425 | *ETV2* |
| ENSG00000079246 | 2 | 216962187 | 217081026 | 299 | 36 | 41944 | 0.79156 | 0.21431 | *XRCC5* |
| ENSG00000076706 | 11 | 119169241 | 119202231 | 40 | 16 | 42969 | 0.79155 | 0.21431 | *MCAM* |
| ENSG00000108654 | 17 | 62485734 | 62514317 | 57 | 12 | 36701 | 0.79117 | 0.21442 | *DDX5* |
| ENSG00000080986 | 18 | 2561510 | 2626634 | 218 | 29 | 53545 | 0.79101 | 0.21447 | *NDC80* |
| ENSG00000184022 | 1 | 248746052 | 248767100 | 7 | 4 | 43234 | 0.79095 | 0.21449 | *OR2T10* |

| | | | | | | | | | |
|---|---|---|---|---|---|---|---|---|---|
| ENSG00000156017 | 9 | 77585936 | 77653339 | 183 | 27 | 40871 | 0.79081 | 0.21453 | *C9orf41* |
| ENSG00000090104 | 1 | 192534857 | 192559161 | 85 | 14 | 36707 | 0.79043 | 0.21464 | *RGS1* |
| ENSG00000169813 | 10 | 43871065 | 43914614 | 173 | 22 | 32968 | 0.79042 | 0.21464 | *HNRNPF* |
| ENSG00000205765 | 5 | 41894290 | 41931738 | 57 | 12 | 43973 | 0.78997 | 0.21477 | *C5orf51* |
| ENSG00000136573 | 8 | 11341510 | 11432113 | 404 | 45 | 35790 | 0.78995 | 0.21478 | *BLK* |
| ENSG00000138346 | 10 | 70163821 | 70241879 | 329 | 28 | 29325 | 0.78984 | 0.21481 | *DNA2* |
| ENSG00000170145 | 11 | 111463115 | 111611577 | 359 | 23 | 27737 | 0.78969 | 0.21485 | *SIK2* |
| ENSG00000125454 | 17 | 73259073 | 73295591 | 125 | 19 | 33200 | 0.78968 | 0.21486 | *SLC25A19* |
| ENSG00000268706 | 18 | 61592042 | 61619601 | 80 | 7 | 38218 | 0.78966 | 0.21486 | *AC009802.1* |
| ENSG00000104490 | 8 | 102688771 | 103147135 | 1556 | 151 | 42903 | 0.78957 | 0.21489 | *NCALD* |
| ENSG00000206560 | 3 | 15698743 | 15911278 | 427 | 29 | 42001 | 0.78954 | 0.2149 | *ANKRD28* |
| ENSG00000267845 | 9 | 139209089 | 139229640 | 58 | 13 | 49102 | 0.78949 | 0.21491 | *DKFZP434A062* |
| ENSG00000126603 | 16 | 4354762 | 4399598 | 100 | 31 | 41394 | 0.78922 | 0.21499 | *GLIS2* |
| ENSG00000103569 | 15 | 58420368 | 58488110 | 236 | 43 | 47321 | 0.78908 | 0.21503 | *AQP9* |
| ENSG00000172154 | 11 | 55850753 | 55871762 | 187 | 8 | 54427 | 0.78903 | 0.21505 | *OR8I2* |
| ENSG00000172188 | 11 | 55360830 | 55381874 | 41 | 5 | 49331 | 0.78891 | 0.21508 | *OR4C11* |
| ENSG00000137522 | 11 | 71629747 | 71718643 | 248 | 15 | 39033 | 0.78893 | 0.21508 | *RNF121* |
| ENSG00000244476 | 6 | 11092722 | 11121965 | 137 | 13 | 52540 | 0.78889 | 0.21509 | *ERVFRD-1* |
| ENSG00000138587 | 15 | 56703742 | 56767335 | 160 | 24 | 48637 | 0.78879 | 0.21512 | *MNS1* |
| ENSG00000163814 | 3 | 45113770 | 45197914 | 321 | 72 | 48515 | 0.78868 | 0.21515 | *CDCP1* |
| ENSG00000197852 | 1 | 112213252 | 112308446 | 375 | 51 | 49967 | 0.7886 | 0.21517 | *FAM212B* |
| ENSG00000103035 | 16 | 74320673 | 74350186 | 99 | 17 | 50410 | 0.78856 | 0.21519 | *PSMD7* |
| ENSG00000228157 | 17 | 18986287 | 19010150 | 1 | 1 | 43266 | 0.78851 | 0.2152 | *AC007952.5* |
| ENSG00000165807 | 14 | 65006620 | 65066098 | 194 | 28 | 51619 | 0.78843 | 0.21522 | *PPP1R36* |
| ENSG00000125991 | 20 | 34119770 | 34155405 | 93 | 11 | 41686 | 0.78845 | 0.21522 | *ERGIC3* |
| ENSG00000103227 | 16 | 893634 | 1041318 | 861 | 74 | 35388 | 0.78838 | 0.21524 | *LMF1* |
| ENSG00000138794 | 4 | 110599785 | 110634739 | 78 | 11 | 52724 | 0.78827 | 0.21527 | *CASP6* |
| ENSG00000100902 | 14 | 35737839 | 35796699 | 217 | 24 | 44914 | 0.78818 | 0.2153 | *PSMA6* |
| ENSG00000057757 | 1 | 24094895 | 24124722 | 27 | 10 | 50609 | 0.78799 | 0.21535 | *PITHD1* |
| ENSG00000185716 | 16 | 22008959 | 22108855 | 112 | 22 | 44836 | 0.78778 | 0.21541 | *C16orf52* |
| ENSG00000101542 | 18 | 58990815 | 59233006 | 687 | 164 | 46150 | 0.7877 | 0.21544 | *CDH20* |
| ENSG00000125901 | 20 | 3016591 | 3038900 | 65 | 13 | 48367 | 0.78761 | 0.21546 | *MRPS26* |
| ENSG00000184786 | 6 | 170130210 | 170161655 | 88 | 15 | 33495 | 0.78759 | 0.21547 | *TCTE3* |
| ENSG00000183734 | 11 | 2279725 | 2302182 | 109 | 19 | 37485 | 0.78718 | 0.21559 | *ASCL2* |
| ENSG00000180919 | 11 | 6118914 | 6140065 | 98 | 6 | 21621 | 0.7871 | 0.21561 | *OR56B4* |
| ENSG00000171425 | 19 | 56136825 | 56166988 | 97 | 21 | 43583 | 0.78688 | 0.21568 | *ZNF581* |

| | | | | | | | | | |
|---|---|---|---|---|---|---|---|---|---|
| ENSG00000122481 | 1 | 95689711 | 95722781 | 126 | 24 | 47013 | 0.78673 | 0.21572 | *RWDD3* |
| ENSG00000088833 | 20 | 1412807 | 1464487 | 137 | 22 | 46472 | 0.78672 | 0.21572 | *NSFL1C* |
| ENSG00000139908 | 14 | 24664903 | 24687568 | 61 | 11 | 41987 | 0.78655 | 0.21577 | *TSSK4* |
| ENSG00000164989 | 9 | 15542895 | 16071661 | 1847 | 52 | 44630 | 0.78647 | 0.2158 | *CCDC171* |
| ENSG00000263020 | 6 | 31623879 | 31651323 | 91 | 21 | 37492 | 0.78636 | 0.21583 | *CSNK2B-LY6G5B-1181* |
| ENSG00000186193 | 9 | 139946581 | 139975040 | 79 | 17 | 47530 | 0.78623 | 0.21587 | *SAPCD2* |
| ENSG00000050130 | 14 | 59941161 | 59982128 | 187 | 16 | 40721 | 0.78612 | 0.2159 | *JKAMP* |
| ENSG00000183889 | 16 | 16401301 | 16454447 | 5 | 3 | 36983 | 0.7861 | 0.2159 | *NPIPA7* |
| ENSG00000104499 | 8 | 143905663 | 144007922 | 443 | 30 | 31645 | 0.78606 | 0.21592 | *GML* |
| ENSG00000173566 | 8 | 21954383 | 21976932 | 88 | 12 | 53692 | 0.78595 | 0.21595 | *NUDT18* |
| ENSG00000186628 | 15 | 83414114 | 83484822 | 179 | 25 | 44262 | 0.78576 | 0.216 | *FSD2* |
| ENSG00000149972 | 11 | 98881683 | 100239616 | 6192 | 390 | 45679 | 0.78571 | 0.21602 | *CNTN5* |
| ENSG00000186660 | 11 | 58336584 | 58398515 | 129 | 20 | 52888 | 0.78558 | 0.21606 | *ZFP91* |
| ENSG00000273047 | 20 | 62359623 | 62381751 | 88 | 13 | 40119 | 0.78553 | 0.21607 | *RP4-583P15.14* |
| ENSG00000225932 | 7 | 143870559 | 143893146 | 17 | 5 | 44454 | 0.78547 | 0.21609 | *CTAGE4* |
| ENSG00000254122 | 5 | 140787427 | 140902546 | 320 | 42 | 43345 | 0.78532 | 0.21613 | *PCDHGB7* |
| ENSG00000106688 | 9 | 4480444 | 4597469 | 441 | 73 | 45846 | 0.78526 | 0.21615 | *SLC1A1* |
| ENSG00000004776 | 19 | 36235469 | 36258980 | 58 | 14 | 45381 | 0.78507 | 0.21621 | *HSPB6* |
| ENSG00000124003 | 2 | 223526505 | 223584649 | 203 | 39 | 34414 | 0.78495 | 0.21624 | *MOGAT1* |
| ENSG00000162039 | 16 | 1873984 | 1944295 | 541 | 12 | 52610 | 0.78475 | 0.2163 | *MEIOB* |
| ENSG00000232941 | 8 | 52720143 | 52740969 | 59 | 11 | 33148 | 0.7847 | 0.21631 | *AC090186.1* |
| ENSG00000188404 | 1 | 169649808 | 169690839 | 190 | 25 | 46819 | 0.78464 | 0.21633 | *SELL* |
| ENSG00000134308 | 2 | 9714101 | 9781143 | 140 | 21 | 40654 | 0.78423 | 0.21645 | *YWHAQ* |
| ENSG00000005073 | 7 | 27211129 | 27234842 | 41 | 18 | 48133 | 0.78421 | 0.21646 | *HOXA11* |
| ENSG00000167549 | 17 | 27931774 | 27959925 | 35 | 12 | 44214 | 0.78391 | 0.21655 | *CORO6* |
| ENSG00000078269 | 6 | 158392888 | 158530208 | 497 | 59 | 50479 | 0.78382 | 0.21657 | *SYNJ2* |
| ENSG00000253159 | 5 | 140800185 | 140902546 | 302 | 40 | 43429 | 0.7837 | 0.21661 | *PCDHGA12* |
| ENSG00000099804 | 19 | 521714 | 552097 | 193 | 43 | 41354 | 0.78369 | 0.21661 | *CDC34* |
| ENSG00000253846 | 5 | 140782743 | 140902546 | 338 | 44 | 43310 | 0.78368 | 0.21662 | *PCDHGA10* |
| ENSG00000102921 | 16 | 48562637 | 48664059 | 217 | 11 | 38917 | 0.78345 | 0.21668 | *N4BP1* |
| ENSG00000186231 | 6 | 97362605 | 97598630 | 798 | 84 | 44951 | 0.78334 | 0.21671 | *KLHL32* |
| ENSG00000115507 | 2 | 63267192 | 63294971 | 22 | 10 | 52610 | 0.78331 | 0.21672 | *OTX1* |
| ENSG00000198554 | 14 | 55395668 | 55503823 | 289 | 28 | 42934 | 0.78328 | 0.21673 | *WDHD1* |
| ENSG00000127989 | 7 | 91311323 | 91520034 | 417 | 30 | 44933 | 0.78325 | 0.21674 | *MTERF* |
| ENSG00000172922 | 11 | 65472367 | 65498418 | 95 | 16 | 38039 | 0.78322 | 0.21675 | *RNASEH2C* |
| ENSG00000131386 | 3 | 16206156 | 16283499 | 280 | 52 | 46343 | 0.78306 | 0.2168 | *GALNT15* |

| | | | | | | | | | |
|---|---|---|---|---|---|---|---|---|---|
| ENSG00000137757 | 11 | 104854962 | 104903895 | 194 | 22 | 51428 | 0.7829 | 0.21684 | *CASP5* |
| ENSG00000185008 | 3 | 75945846 | 77709115 | 5774 | 417 | 46219 | 0.78246 | 0.21697 | *ROBO2* |
| ENSG00000111667 | 12 | 6951292 | 6985796 | 36 | 8 | 43632 | 0.78248 | 0.21697 | *USP5* |
| ENSG00000267360 | 19 | 37573921 | 37711414 | 356 | 29 | 47235 | 0.78243 | 0.21698 | *CTC-454I21.3* |
| ENSG00000136840 | 9 | 130660165 | 130689317 | 60 | 25 | 41157 | 0.78198 | 0.21711 | *ST6GALNAC4* |
| ENSG00000134827 | 11 | 59610273 | 59644048 | 36 | 15 | 51283 | 0.78199 | 0.21711 | *TCN1* |
| ENSG00000181610 | 17 | 55906842 | 55937417 | 96 | 20 | 48579 | 0.78185 | 0.21715 | *MRPS23* |
| ENSG00000183287 | 18 | 57088172 | 57374612 | 1194 | 147 | 43768 | 0.78144 | 0.21727 | *CCBE1* |
| ENSG00000162415 | 1 | 45472071 | 45781881 | 707 | 50 | 45175 | 0.78126 | 0.21732 | *ZSWIM5* |
| ENSG00000130635 | 9 | 137523620 | 137746686 | 1052 | 133 | 44319 | 0.78117 | 0.21735 | *COL5A1* |
| ENSG00000189433 | 1 | 35215342 | 35239325 | 95 | 18 | 51433 | 0.78109 | 0.21738 | *GJB4* |
| ENSG00000075420 | 3 | 171747418 | 172129455 | 1069 | 134 | 45089 | 0.78106 | 0.21738 | *FNDC3B* |
| ENSG00000171055 | 2 | 36768570 | 36883230 | 433 | 59 | 46953 | 0.78081 | 0.21746 | *FEZ2* |
| ENSG00000149483 | 11 | 61119473 | 61146981 | 8 | 4 | 38968 | 0.78076 | 0.21747 | *TMEM138* |
| ENSG00000139155 | 12 | 20838289 | 20916320 | 364 | 35 | 38386 | 0.78072 | 0.21748 | *SLCO1C1* |
| ENSG00000099849 | 11 | 550404 | 574021 | 145 | 23 | 48360 | 0.78062 | 0.21751 | *RASSF7* |
| ENSG00000258247 | 12 | 95760984 | 95784672 | 120 | 18 | 41128 | 0.78037 | 0.21759 | *RP11-167N24.6* |
| ENSG00000269358 | 1 | 224205760 | 224226371 | 20 | 6 | 48056 | 0.78033 | 0.2176 | *AC138393.1* |
| ENSG00000198471 | 3 | 187406047 | 187430345 | 118 | 17 | 39736 | 0.78025 | 0.21762 | *RTP2* |
| ENSG00000101745 | 18 | 9126226 | 9295983 | 536 | 42 | 45901 | 0.78024 | 0.21762 | *ANKRD12* |
| ENSG00000266118 | 2 | 24337236 | 24423305 | 329 | 32 | 47975 | 0.78016 | 0.21765 | *RP11-507M3.1* |
| ENSG00000126062 | 3 | 50382180 | 50407041 | 14 | 6 | 53400 | 0.78016 | 0.21765 | *TMEM115* |
| ENSG00000120868 | 12 | 99028919 | 99139204 | 265 | 30 | 53233 | 0.78009 | 0.21767 | *APAF1* |
| ENSG00000172020 | 3 | 115332171 | 115450337 | 331 | 75 | 47885 | 0.77996 | 0.21771 | *GAP43* |
| ENSG00000204568 | 6 | 30575486 | 30604172 | 81 | 9 | 27194 | 0.77994 | 0.21771 | *MRPS18B* |
| ENSG00000263809 | 17 | 8261956 | 8296505 | 106 | 18 | 54137 | 0.77997 | 0.21771 | *RP11-849F2.7* |
| ENSG00000100412 | 22 | 41855129 | 41934993 | 117 | 18 | 48846 | 0.77951 | 0.21784 | *ACO2* |
| ENSG00000147676 | 8 | 120167273 | 120267913 | 344 | 43 | 39929 | 0.77948 | 0.21785 | *MAL2* |
| ENSG00000118596 | 12 | 59979848 | 60186395 | 614 | 46 | 43600 | 0.77918 | 0.21794 | *SLC16A7* |
| ENSG00000109181 | 4 | 69671711 | 69706914 | 147 | 19 | 44440 | 0.77891 | 0.21802 | *UGT2B10* |
| ENSG00000113716 | 5 | 149369884 | 149442386 | 191 | 21 | 37118 | 0.77867 | 0.21809 | *HMGXB3* |
| ENSG00000085514 | 7 | 99955153 | 100007719 | 80 | 10 | 51048 | 0.77848 | 0.21814 | *PILRA* |
| ENSG00000133027 | 17 | 17398877 | 17505022 | 296 | 34 | 33720 | 0.77843 | 0.21816 | *PEMT* |
| ENSG00000215454 | 21 | 45983606 | 46004987 | 82 | 12 | 35474 | 0.77835 | 0.21818 | *KRTAP10-4* |
| ENSG00000258529 | 11 | 111647010 | 111760149 | 244 | 22 | 39224 | 0.77783 | 0.21833 | *ALG9* |
| ENSG00000121073 | 17 | 47768305 | 47796376 | 45 | 14 | 48886 | 0.77779 | 0.21835 | *SLC35B1* |

| | | | | | | | | | |
|---|---|---|---|---|---|---|---|---|---|
| ENSG00000161970 | 17 | 8270838 | 8296531 | 84 | 14 | 55454 | 0.77736 | 0.21847 | *RPL26* |
| ENSG00000269855 | 19 | 58897457 | 58918407 | 72 | 13 | 46259 | 0.77657 | 0.2187 | *AC012313.1* |
| ENSG00000135392 | 12 | 56204744 | 56234608 | 39 | 16 | 43646 | 0.77656 | 0.21871 | *DNAJC14* |
| ENSG00000160539 | 9 | 134155081 | 134194649 | 133 | 17 | 51154 | 0.77629 | 0.21879 | *PPAPDC3* |
| ENSG00000184047 | 12 | 122682210 | 122722081 | 73 | 14 | 38140 | 0.77595 | 0.21889 | *DIABLO* |
| ENSG00000138175 | 10 | 104423488 | 104484164 | 117 | 21 | 41475 | 0.77578 | 0.21894 | *ARL3* |
| ENSG00000066294 | 1 | 160500885 | 160559306 | 245 | 29 | 31255 | 0.7757 | 0.21896 | *CD84* |
| ENSG00000160325 | 9 | 136315089 | 136345970 | 96 | 20 | 47831 | 0.77543 | 0.21904 | *CACFD1* |
| ENSG00000185274 | 7 | 70587155 | 71188585 | 2225 | 206 | 43933 | 0.77523 | 0.2191 | *WBSCR17* |
| ENSG00000217075 | 2 | 36748948 | 36789411 | 120 | 22 | 53188 | 0.77509 | 0.21914 | *AC007401.2* |
| ENSG00000115275 | 2 | 74678184 | 74702537 | 11 | 6 | 44081 | 0.77509 | 0.21914 | *MOGS* |
| ENSG00000180767 | 3 | 126233126 | 126272134 | 161 | 26 | 42038 | 0.7751 | 0.21914 | *CHST13* |
| ENSG00000220032 | 18 | 74197477 | 74220045 | 49 | 14 | 49861 | 0.77496 | 0.21918 | *RP11-17M16.1* |
| ENSG00000026950 | 6 | 26392465 | 26425444 | 153 | 10 | 50101 | 0.77485 | 0.21921 | *BTN3A1* |
| ENSG00000204422 | 6 | 31644739 | 31691849 | 109 | 22 | 30321 | 0.77481 | 0.21923 | *XXbac-BPG32J3.20* |
| ENSG00000174744 | 11 | 66094804 | 66122596 | 61 | 11 | 37318 | 0.77434 | 0.21937 | *BRMS1* |
| ENSG00000158552 | 2 | 220049959 | 220084373 | 52 | 15 | 31658 | 0.77407 | 0.21944 | *ZFAND2B* |
| ENSG00000153944 | 17 | 55323212 | 55772046 | 1362 | 220 | 47072 | 0.77408 | 0.21944 | *MSI2* |
| ENSG00000134283 | 12 | 42622249 | 42863517 | 704 | 39 | 48446 | 0.77369 | 0.21956 | *PPHLN1* |
| ENSG00000018236 | 12 | 41076244 | 41476220 | 1498 | 101 | 49641 | 0.77357 | 0.21959 | *CNTN1* |
| ENSG00000184056 | 15 | 91531646 | 91575833 | 147 | 24 | 39675 | 0.77354 | 0.2196 | *VPS33B* |
| ENSG00000173077 | 9 | 117894097 | 118174923 | 1099 | 107 | 47296 | 0.77345 | 0.21963 | *Dec-01* |
| ENSG00000099330 | 19 | 17327013 | 17350028 | 128 | 19 | 42896 | 0.77306 | 0.21974 | *OCEL1* |
| ENSG00000129562 | 14 | 23023805 | 23068175 | 286 | 21 | 34290 | 0.77296 | 0.21977 | *DAD1* |
| ENSG00000146063 | 5 | 180639499 | 180672809 | 124 | 25 | 35370 | 0.77251 | 0.21991 | *TRIM41* |
| ENSG00000145777 | 5 | 110395760 | 110423722 | 58 | 17 | 41190 | 0.77233 | 0.21996 | *TSLP* |
| ENSG00000143369 | 1 | 150470538 | 150496265 | 66 | 14 | 37090 | 0.77212 | 0.22002 | *ECM1* |
| ENSG00000164500 | 7 | 50125632 | 50209426 | 332 | 22 | 51013 | 0.77203 | 0.22005 | *C7orf72* |
| ENSG00000148948 | 11 | 40125753 | 41491323 | 4219 | 288 | 49548 | 0.77194 | 0.22008 | *LRRC4C* |
| ENSG00000116514 | 1 | 33392046 | 33440286 | 108 | 14 | 49857 | 0.77183 | 0.22011 | *RNF19B* |
| ENSG00000116954 | 1 | 39293870 | 39335495 | 108 | 20 | 40898 | 0.77156 | 0.22019 | *RRAGC* |
| ENSG00000163817 | 3 | 45786942 | 45848027 | 143 | 47 | 45961 | 0.77145 | 0.22022 | *SLC6A20* |
| ENSG00000256073 | 21 | 33755439 | 33776254 | 53 | 7 | 36176 | 0.77138 | 0.22024 | *C21orf119* |
| ENSG00000166147 | 15 | 48690503 | 48948046 | 536 | 35 | 40041 | 0.77053 | 0.22049 | *FBN1* |
| ENSG00000132485 | 1 | 71518974 | 71556980 | 102 | 19 | 52912 | 0.77051 | 0.2205 | *ZRANB2* |
| ENSG00000183840 | 2 | 133164147 | 133414132 | 750 | 101 | 40931 | 0.77045 | 0.22052 | *GPR39* |

| | | | | | | | | | |
|---|---|---|---|---|---|---|---|---|---|
| ENSG00000138375 | 2 | 217267137 | 217357776 | 365 | 36 | 40350 | 0.77044 | 0.22052 | *SMARCAL1* |
| ENSG00000164266 | 5 | 147194131 | 147221349 | 71 | 19 | 46180 | 0.77039 | 0.22053 | *SPINK1* |
| ENSG00000058091 | 7 | 90085738 | 90849905 | 2704 | 150 | 49820 | 0.7704 | 0.22053 | *CDK14* |
| ENSG00000212123 | 19 | 5772971 | 5794776 | 97 | 15 | 32967 | 0.77036 | 0.22054 | *PRR22* |
| ENSG00000196562 | 20 | 46275092 | 46425360 | 660 | 145 | 47192 | 0.77034 | 0.22055 | *SULF2* |
| ENSG00000136267 | 7 | 14174674 | 15024402 | 3017 | 231 | 41408 | 0.77021 | 0.22059 | *DGKB* |
| ENSG00000139168 | 12 | 42695880 | 42729920 | 93 | 12 | 50487 | 0.76958 | 0.22077 | *ZCRB1* |
| ENSG00000196734 | 1 | 152774447 | 152795585 | 47 | 15 | 50148 | 0.76955 | 0.22078 | *LCE1B* |
| ENSG00000232833 | 9 | 67774944 | 67796624 | 1 | 1 | 43041 | 0.76949 | 0.2208 | *FAM27E3* |
| ENSG00000196242 | 1 | 247683434 | 247707141 | 97 | 18 | 51505 | 0.76946 | 0.22081 | *OR2C3* |
| ENSG00000211460 | 2 | 122484679 | 122535429 | 88 | 15 | 36513 | 0.76944 | 0.22081 | *TSN* |
| ENSG00000100897 | 14 | 24573404 | 24604451 | 64 | 11 | 31260 | 0.7692 | 0.22089 | *DCAF11* |
| ENSG00000021776 | 15 | 35137732 | 35272040 | 362 | 32 | 40208 | 0.76913 | 0.22091 | *AQR* |
| ENSG00000148225 | 9 | 116065502 | 116112578 | 177 | 19 | 38342 | 0.76909 | 0.22092 | *WDR31* |
| ENSG00000270800 | 6 | 34246547 | 34403825 | 342 | 20 | 44814 | 0.76889 | 0.22098 | *RPS10-NUDT3* |
| ENSG00000174197 | 15 | 41903422 | 42072141 | 372 | 35 | 36643 | 0.76874 | 0.22102 | *MGA* |
| ENSG00000214941 | 17 | 15869874 | 15913031 | 135 | 15 | 42091 | 0.76866 | 0.22105 | *ZSWIM7* |
| ENSG00000132970 | 13 | 27121840 | 27273085 | 557 | 50 | 52147 | 0.76863 | 0.22106 | *WASF3* |
| ENSG00000186340 | 6 | 169605875 | 169664139 | 326 | 58 | 51278 | 0.7686 | 0.22107 | *THBS2* |
| ENSG00000213015 | 19 | 56136382 | 56164835 | 91 | 20 | 44465 | 0.76859 | 0.22107 | *ZNF580* |
| ENSG00000025039 | 6 | 90064355 | 90131989 | 175 | 36 | 49651 | 0.76853 | 0.22109 | *RRAGD* |
| ENSG00000261272 | 6 | 30968251 | 31013179 | 676 | 59 | 48384 | 0.76837 | 0.22113 | *MUC22* |
| ENSG00000119707 | 14 | 73515144 | 73598122 | 189 | 33 | 50988 | 0.7683 | 0.22115 | *RBM25* |
| ENSG00000215018 | 7 | 7385834 | 7585484 | 728 | 64 | 44050 | 0.76827 | 0.22116 | *COL28A1* |
| ENSG00000204930 | 9 | 35806388 | 35838744 | 58 | 12 | 40905 | 0.76826 | 0.22117 | *FAM221B* |
| ENSG00000183470 | 7 | 132323553 | 132423528 | 257 | 40 | 44128 | 0.76818 | 0.22119 | *AC009365.3* |
| ENSG00000134121 | 3 | 228279 | 461090 | 958 | 113 | 38301 | 0.76768 | 0.22134 | *CHL1* |
| ENSG00000149380 | 11 | 73936846 | 74032702 | 242 | 25 | 46041 | 0.76732 | 0.22145 | *P4HA3* |
| ENSG00000181790 | 8 | 143520791 | 143636370 | 301 | 67 | 43671 | 0.76712 | 0.2215 | *BAI1* |
| ENSG00000268111 | 20 | 44085910 | 44117452 | 119 | 20 | 33771 | 0.76692 | 0.22156 | *AL031663.1* |
| ENSG00000165983 | 10 | 16468942 | 16565736 | 419 | 44 | 36911 | 0.76626 | 0.22176 | *PTER* |
| ENSG00000145982 | 6 | 5251277 | 5781813 | 1940 | 248 | 44616 | 0.76604 | 0.22182 | *FARS2* |
| ENSG00000269549 | 20 | 44064071 | 44087220 | 59 | 14 | 33834 | 0.76606 | 0.22182 | *AL031663.2* |
| ENSG00000103490 | 16 | 31202806 | 31224771 | 20 | 8 | 40197 | 0.76604 | 0.22183 | *PYCARD* |
| ENSG00000261359 | 16 | 31203206 | 31224773 | 20 | 8 | 40197 | 0.76604 | 0.22183 | *C16orf98* |
| ENSG00000204681 | 6 | 29513406 | 29611753 | 380 | 43 | 44496 | 0.76597 | 0.22185 | *GABBR1* |

| | | | | | | | | | |
|---|---|---|---|---|---|---|---|---|---|
| ENSG00000171680 | 1 | 6516152 | 6590121 | 195 | 32 | 38911 | 0.76591 | 0.22187 | *PLEKHG5* |
| ENSG00000101654 | 18 | 13716659 | 13774557 | 190 | 25 | 47125 | 0.76546 | 0.222 | *RNMT* |
| ENSG00000168490 | 8 | 22067222 | 22099854 | 91 | 22 | 45961 | 0.76531 | 0.22204 | *PHYHIP* |
| ENSG00000264813 | 17 | 61552184 | 61609209 | 140 | 21 | 48146 | 0.76516 | 0.22209 | *ACE* |
| ENSG00000269048 | 6 | 166935369 | 166957709 | 107 | 13 | 54039 | 0.76498 | 0.22214 | *Z98049.1* |
| ENSG00000148483 | 10 | 17784251 | 17852867 | 37 | 12 | 16080 | 0.76482 | 0.22219 | *TMEM236* |
| ENSG00000259371 | 14 | 24580560 | 24611038 | 58 | 9 | 33103 | 0.7648 | 0.2222 | *RP11-468E2.6* |
| ENSG00000108001 | 10 | 131623547 | 131772105 | 343 | 105 | 44554 | 0.76474 | 0.22221 | *EBF3* |
| ENSG00000257040 | 14 | 72446191 | 72468326 | 122 | 15 | 52297 | 0.76468 | 0.22223 | *AC005477.1* |
| ENSG00000256618 | 17 | 22012437 | 22033991 | 107 | 20 | 52437 | 0.76466 | 0.22224 | *MTRNR2L1* |
| ENSG00000162444 | 1 | 10047264 | 10086078 | 82 | 14 | 38258 | 0.76458 | 0.22226 | *RBP7* |
| ENSG00000172156 | 17 | 32602687 | 32625353 | 78 | 13 | 53837 | 0.76453 | 0.22228 | *CCL11* |
| ENSG00000144115 | 2 | 88459835 | 88496146 | 115 | 25 | 43713 | 0.76392 | 0.22246 | *THNSL2* |
| ENSG00000106013 | 7 | 117844727 | 117892785 | 187 | 16 | 45148 | 0.76379 | 0.2225 | *ANKRD7* |
| ENSG00000212734 | 17 | 6544971 | 6566791 | 100 | 13 | 44443 | 0.76376 | 0.2225 | *C17orf100* |
| ENSG00000133121 | 13 | 33667272 | 33934767 | 848 | 113 | 48201 | 0.76373 | 0.22251 | *STARD13* |
| ENSG00000113282 | 5 | 157202751 | 157296183 | 284 | 22 | 42507 | 0.76373 | 0.22252 | *CLINT1* |
| ENSG00000260914 | 16 | 69323585 | 69364614 | 105 | 16 | 54566 | 0.76316 | 0.22268 | *RP11-343C2.11* |
| ENSG00000133059 | 1 | 205101632 | 205190727 | 232 | 18 | 30446 | 0.76304 | 0.22272 | *DSTYK* |
| ENSG00000122557 | 7 | 35662269 | 35745181 | 216 | 21 | 29245 | 0.76304 | 0.22272 | *HERPUD2* |
| ENSG00000050426 | 12 | 51431745 | 51464207 | 113 | 17 | 34389 | 0.76274 | 0.22281 | *LETMD1* |
| ENSG00000162951 | 2 | 80505483 | 80541874 | 79 | 25 | 38135 | 0.76261 | 0.22285 | *LRRTM1* |
| ENSG00000176900 | 11 | 4893049 | 4914113 | 43 | 9 | 32351 | 0.76229 | 0.22294 | *OR51T1* |
| ENSG00000134640 | 11 | 92692886 | 92728232 | 106 | 17 | 43542 | 0.76216 | 0.22298 | *MTNR1B* |
| ENSG00000178127 | 18 | 9092628 | 9144343 | 134 | 23 | 49077 | 0.76205 | 0.22302 | *NDUFV2* |
| ENSG00000081985 | 1 | 67763047 | 67872583 | 323 | 33 | 38642 | 0.76187 | 0.22307 | *IL12RB2* |
| ENSG00000268503 | 17 | 7507264 | 7527427 | 39 | 10 | 38871 | 0.76186 | 0.22307 | *AC007421.1* |
| ENSG00000177489 | 1 | 247741662 | 247762615 | 93 | 17 | 50473 | 0.7616 | 0.22315 | *OR2G2* |
| ENSG00000072415 | 14 | 67697826 | 67812536 | 255 | 34 | 45509 | 0.76152 | 0.22317 | *MPP5* |
| ENSG00000198937 | 6 | 37440696 | 37477698 | 90 | 21 | 44258 | 0.76149 | 0.22318 | *CCDC167* |
| ENSG00000125999 | 20 | 31851286 | 31907684 | 160 | 26 | 46410 | 0.76141 | 0.22321 | *BPIFB1* |
| ENSG00000078124 | 11 | 76561911 | 76747841 | 572 | 26 | 41694 | 0.7613 | 0.22324 | *ACER3* |
| ENSG00000111880 | 6 | 89309985 | 89683348 | 864 | 43 | 45333 | 0.76126 | 0.22325 | *RNGTT* |
| ENSG00000112701 | 6 | 76301225 | 76437997 | 387 | 34 | 38753 | 0.76119 | 0.22327 | *SENP6* |
| ENSG00000131781 | 1 | 146636930 | 146724700 | 488 | 25 | 47381 | 0.76118 | 0.22328 | *FMO5* |
| ENSG00000132323 | 2 | 239069042 | 239122370 | 262 | 23 | 48819 | 0.76106 | 0.22331 | *ILKAP* |

| | | | | | | | | | |
|---|---|---|---|---|---|---|---|---|---|
| ENSG00000182197 | 8 | 118796729 | 119134092 | 916 | 110 | 42983 | 0.76055 | 0.22346 | *EXT1* |
| ENSG00000183260 | 20 | 62482566 | 62504341 | 59 | 12 | 36918 | 0.76044 | 0.2235 | *ABHD16B* |
| ENSG00000010810 | 6 | 111971535 | 112204655 | 713 | 71 | 34667 | 0.7603 | 0.22354 | *FYN* |
| ENSG00000204843 | 2 | 74578281 | 74629214 | 62 | 9 | 28740 | 0.76023 | 0.22356 | *DCTN1* |
| ENSG00000233538 | 2 | 232306906 | 232327864 | 65 | 13 | 43872 | 0.76016 | 0.22358 | *AC017104.2* |
| ENSG00000154978 | 7 | 55493749 | 55650681 | 793 | 23 | 52491 | 0.76011 | 0.2236 | *VOPP1* |
| ENSG00000182774 | 15 | 83195504 | 83219315 | 13 | 4 | 47204 | 0.76009 | 0.2236 | *RPS17L* |
| ENSG00000103740 | 15 | 78449810 | 78548030 | 355 | 62 | 45927 | 0.76006 | 0.22361 | *ACSBG1* |
| ENSG00000130702 | 20 | 60873011 | 60952368 | 379 | 46 | 44131 | 0.76005 | 0.22361 | *LAMA5* |
| ENSG00000133627 | 7 | 152446834 | 152562463 | 349 | 41 | 42159 | 0.76003 | 0.22362 | *ACTR3B* |
| ENSG00000065243 | 1 | 89139905 | 89311938 | 523 | 26 | 27351 | 0.7599 | 0.22366 | *PKN2* |
| ENSG00000157224 | 7 | 90003035 | 90152716 | 488 | 60 | 48230 | 0.75984 | 0.22368 | *CLDN12* |
| ENSG00000171403 | 17 | 39712096 | 39738310 | 61 | 9 | 47502 | 0.75913 | 0.22389 | *KRT9* |
| ENSG00000183570 | 21 | 47053608 | 47372368 | 848 | 52 | 40114 | 0.75899 | 0.22393 | *PCBP3* |
| ENSG00000103197 | 16 | 2087466 | 2148716 | 156 | 40 | 41946 | 0.75894 | 0.22395 | *TSC2* |
| ENSG00000013392 | 6 | 83893098 | 83918651 | 63 | 15 | 39824 | 0.7588 | 0.22399 | *RWDD2A* |
| ENSG00000130649 | 10 | 135323910 | 135384724 | 261 | 14 | 42211 | 0.75871 | 0.22401 | *CYP2E1* |
| ENSG00000204370 | 11 | 111947497 | 112000353 | 77 | 25 | 35028 | 0.75854 | 0.22406 | *SDHD* |
| ENSG00000131153 | 16 | 85699804 | 85733679 | 103 | 17 | 45628 | 0.75799 | 0.22423 | *GINS2* |
| ENSG00000162944 | 2 | 198422948 | 198550769 | 263 | 23 | 52790 | 0.75775 | 0.2243 | *RFTN2* |
| ENSG00000221821 | 6 | 42848005 | 42868554 | 75 | 12 | 41006 | 0.75757 | 0.22435 | *C6orf226* |
| ENSG00000184163 | 1 | 1167826 | 1192102 | 92 | 18 | 42523 | 0.75753 | 0.22436 | *FAM132A* |
| ENSG00000116981 | 1 | 40114793 | 40147710 | 97 | 22 | 50200 | 0.75739 | 0.22441 | *NT5C1A* |
| ENSG00000072182 | 2 | 220368892 | 220413494 | 98 | 20 | 45707 | 0.75739 | 0.22441 | *ASIC4* |
| ENSG00000164816 | 8 | 6902831 | 6924256 | 123 | 19 | 51562 | 0.75736 | 0.22442 | *DEFA5* |
| ENSG00000176383 | 12 | 122678090 | 122703499 | 51 | 13 | 39379 | 0.7571 | 0.2245 | *B3GNT4* |
| ENSG00000198952 | 1 | 156209015 | 156262620 | 125 | 13 | 55799 | 0.75701 | 0.22452 | *SMG5* |
| ENSG00000074211 | 4 | 6312305 | 6575327 | 1038 | 141 | 44633 | 0.75683 | 0.22458 | *PPP2R2C* |
| ENSG00000183762 | 22 | 29459066 | 29574321 | 414 | 47 | 45255 | 0.75646 | 0.22469 | *KREMEN1* |
| ENSG00000174876 | 1 | 104220037 | 104249302 | 1 | 1 | 43098 | 0.75642 | 0.2247 | *AMY1B* |
| ENSG00000219073 | 1 | 22293514 | 22335135 | 303 | 35 | 32455 | 0.75633 | 0.22473 | *CELA3B* |
| ENSG00000168582 | 2 | 209015464 | 209038300 | 125 | 21 | 37294 | 0.75613 | 0.22479 | *CRYGA* |
| ENSG00000169750 | 17 | 79979500 | 80002080 | 74 | 11 | 32710 | 0.7559 | 0.22486 | *RAC3* |
| ENSG00000273167 | 13 | 24543839 | 24905169 | 1642 | 217 | 43385 | 0.75581 | 0.22488 | *RP11-307N16.6* |
| ENSG00000164574 | 5 | 153560290 | 153810544 | 949 | 87 | 45894 | 0.7556 | 0.22494 | *GALNT10* |
| ENSG00000148290 | 9 | 136208610 | 136233552 | 67 | 10 | 50703 | 0.75547 | 0.22498 | *SURF1* |

| | | | | | | | | | |
|---|---|---|---|---|---|---|---|---|---|
| ENSG00000206177 | 16 | 193891 | 226767 | 89 | 21 | 39648 | 0.75511 | 0.22509 | *HBM* |
| ENSG00000116898 | 1 | 36911319 | 36940038 | 114 | 13 | 52449 | 0.75495 | 0.22514 | *MRPS15* |
| ENSG00000269514 | 12 | 48582170 | 48605814 | 156 | 13 | 51522 | 0.75489 | 0.22516 | *DKFZP779L1853* |
| ENSG00000188735 | 12 | 122140658 | 122230907 | 286 | 41 | 36624 | 0.75469 | 0.22522 | *TMEM120B* |
| ENSG00000206077 | 5 | 700475 | 777067 | 72 | 30 | 37385 | 0.7545 | 0.22528 | *ZDHHC11B* |
| ENSG00000155111 | 6 | 110921181 | 111147161 | 540 | 44 | 44040 | 0.7543 | 0.22533 | *CDK19* |
| ENSG00000181958 | 11 | 55125360 | 55146394 | 118 | 17 | 44340 | 0.75425 | 0.22535 | *OR4A15* |
| ENSG00000110619 | 11 | 3012152 | 3088843 | 256 | 24 | 42679 | 0.75421 | 0.22536 | *CARS* |
| ENSG00000169692 | 9 | 139557595 | 139591875 | 90 | 24 | 49392 | 0.75414 | 0.22538 | *AGPAT2* |
| ENSG00000154277 | 4 | 41248430 | 41280472 | 117 | 19 | 48636 | 0.75408 | 0.2254 | *UCHL1* |
| ENSG00000204128 | 2 | 231892205 | 231924434 | 96 | 20 | 41130 | 0.75403 | 0.22542 | *C2orf72* |
| ENSG00000170382 | 1 | 204576298 | 204664861 | 234 | 35 | 40156 | 0.75391 | 0.22545 | *LRRN2* |
| ENSG00000106392 | 7 | 7186565 | 7298282 | 644 | 51 | 42064 | 0.75386 | 0.22547 | *C1GALT1* |
| ENSG00000144381 | 2 | 198341305 | 198391461 | 65 | 12 | 54986 | 0.7538 | 0.22548 | *HSPD1* |
| ENSG00000128606 | 7 | 102543438 | 102595396 | 89 | 15 | 45097 | 0.75343 | 0.2256 | *LRRC17* |
| ENSG00000169105 | 15 | 40753160 | 40775353 | 36 | 9 | 39641 | 0.75343 | 0.2256 | *CHST14* |
| ENSG00000198353 | 12 | 54400715 | 54459813 | 103 | 21 | 52693 | 0.75332 | 0.22563 | *HOXC4* |
| ENSG0000084710 | 2 | 25254999 | 25388243 | 315 | 41 | 38890 | 0.75327 | 0.22564 | *EFR3B* |
| ENSG00000110975 | 12 | 33517173 | 33602754 | 287 | 27 | 37599 | 0.75305 | 0.22571 | *SYT10* |
| ENSG00000164117 | 4 | 175147809 | 175215415 | 173 | 28 | 44238 | 0.75291 | 0.22575 | *FBXO8* |
| ENSG00000183283 | 12 | 51622076 | 51675146 | 139 | 27 | 40035 | 0.75277 | 0.22579 | *DAZAP2* |
| ENSG00000235711 | 15 | 79565146 | 79600580 | 71 | 14 | 50914 | 0.75266 | 0.22583 | *ANKRD34C* |
| ENSG00000114841 | 3 | 52340335 | 52444507 | 147 | 17 | 50601 | 0.75257 | 0.22585 | *DNAH1* |
| ENSG00000162849 | 1 | 245308287 | 245882733 | 2333 | 286 | 45119 | 0.75225 | 0.22595 | *KIF26B* |
| ENSG00000188316 | 10 | 118599023 | 118681299 | 114 | 25 | 47089 | 0.75194 | 0.22604 | *ENO4* |
| ENSG00000189280 | 1 | 35210648 | 35234113 | 89 | 21 | 47292 | 0.7519 | 0.22606 | *GJB5* |
| ENSG00000131013 | 6 | 149815869 | 149877174 | 33 | 12 | 35630 | 0.75175 | 0.2261 | *PPIL4* |
| ENSG00000188610 | 1 | 120827756 | 120865681 | 5 | 3 | 42933 | 0.7517 | 0.22612 | *FAM72B* |
| ENSG00000104549 | 8 | 126000739 | 126044525 | 164 | 19 | 34539 | 0.75166 | 0.22613 | *SQLE* |
| ENSG00000146872 | 17 | 60526019 | 60702842 | 243 | 28 | 48616 | 0.75147 | 0.22619 | *TLK2* |
| ENSG00000204442 | 13 | 107810883 | 108529083 | 2667 | 312 | 45936 | 0.75139 | 0.22621 | *FAM155A* |
| ENSG00000186844 | 1 | 152789949 | 152810573 | 20 | 9 | 47870 | 0.75109 | 0.2263 | *LCE1A* |
| ENSG00000120594 | 10 | 20095168 | 20588785 | 2305 | 162 | 43287 | 0.75059 | 0.22645 | *PLXDC2* |
| ENSG00000152503 | 5 | 114450459 | 114526243 | 186 | 35 | 40551 | 0.75052 | 0.22647 | *TRIM36* |
| ENSG0000078018 | 2 | 210278782 | 210608842 | 890 | 71 | 33551 | 0.75033 | 0.22653 | *MAP2* |
| ENSG00000214954 | 8 | 92104060 | 92241464 | 411 | 27 | 46536 | 0.74985 | 0.22667 | *LRRC69* |

| | | | | | | | | | |
|---|---|---|---|---|---|---|---|---|---|
| ENSG00000150510 | 13 | 51786503 | 51868377 | 228 | 43 | 41389 | 0.74979 | 0.22669 | *FAM124A* |
| ENSG00000186867 | 4 | 122240467 | 122312214 | 420 | 44 | 34572 | 0.74962 | 0.22674 | *QRFPR* |
| ENSG00000136839 | 9 | 107369529 | 107390485 | 119 | 9 | 31744 | 0.74953 | 0.22677 | *OR13C9* |
| ENSG00000164932 | 8 | 104373743 | 104405225 | 126 | 16 | 54601 | 0.74909 | 0.2269 | *CTHRC1* |
| ENSG00000136738 | 10 | 17676124 | 17767913 | 469 | 38 | 50714 | 0.74898 | 0.22694 | *STAM* |
| ENSG00000267979 | 3 | 172009438 | 172029686 | 54 | 12 | 47154 | 0.74868 | 0.22702 | *AC092964.2* |
| ENSG00000269259 | 10 | 102872536 | 102893624 | 82 | 14 | 48251 | 0.74867 | 0.22703 | *HUG1* |
| ENSG00000182541 | 22 | 31598225 | 31686066 | 192 | 27 | 37587 | 0.74867 | 0.22703 | *LIMK2* |
| ENSG00000132003 | 19 | 13896274 | 13953044 | 125 | 16 | 35824 | 0.74828 | 0.22715 | *ZSWIM4* |
| ENSG00000100285 | 22 | 29866219 | 29897379 | 86 | 18 | 45302 | 0.748 | 0.22723 | *NEFH* |
| ENSG00000181803 | 14 | 21098855 | 21119850 | 138 | 13 | 54159 | 0.74787 | 0.22727 | *OR6S1* |
| ENSG00000112499 | 6 | 160582093 | 160708670 | 530 | 58 | 43594 | 0.74767 | 0.22733 | *SLC22A2* |
| ENSG00000130748 | 19 | 47539165 | 47561888 | 79 | 17 | 40933 | 0.74766 | 0.22733 | *TMEM160* |
| ENSG00000131069 | 20 | 33449949 | 33525769 | 135 | 13 | 45884 | 0.7475 | 0.22738 | *ACSS2* |
| ENSG00000170743 | 11 | 7250009 | 7500273 | 1042 | 130 | 46112 | 0.74727 | 0.22745 | *SYT9* |
| ENSG00000085511 | 6 | 161402759 | 161548417 | 440 | 51 | 43005 | 0.74707 | 0.22751 | *MAP3K4* |
| ENSG00000099875 | 19 | 2027470 | 2061243 | 106 | 31 | 50007 | 0.74708 | 0.22751 | *MKNK2* |
| ENSG00000100926 | 14 | 24648349 | 24692679 | 99 | 21 | 40708 | 0.74672 | 0.22762 | *TM9SF1* |
| ENSG00000254692 | 14 | 24648363 | 24692679 | 99 | 21 | 40708 | 0.74672 | 0.22762 | *TM9SF1* |
| ENSG00000148019 | 9 | 80840978 | 80904606 | 161 | 25 | 52545 | 0.74642 | 0.22771 | *CEP78* |
| ENSG00000070831 | 1 | 22369120 | 22429437 | 190 | 26 | 51064 | 0.74614 | 0.22779 | *CDC42* |
| ENSG00000268000 | 19 | 49993781 | 50014614 | 57 | 10 | 33720 | 0.7461 | 0.2278 | *hsa-mir-150* |
| ENSG00000121900 | 1 | 33350195 | 33377039 | 32 | 9 | 55770 | 0.74595 | 0.22785 | *TMEM54* |
| ENSG00000140105 | 14 | 100790125 | 100853142 | 202 | 16 | 39031 | 0.74552 | 0.22798 | *WARS* |
| ENSG00000103485 | 16 | 29664600 | 29720020 | 91 | 20 | 37640 | 0.74507 | 0.22812 | *QPRT* |
| ENSG00000089041 | 12 | 121560622 | 121633876 | 394 | 41 | 48088 | 0.74468 | 0.22823 | *P2RX7* |
| ENSG00000095261 | 9 | 123567774 | 123615262 | 88 | 10 | 51276 | 0.74428 | 0.22835 | *PSMD5* |
| ENSG00000167595 | 19 | 36239044 | 36271930 | 106 | 19 | 41281 | 0.74412 | 0.2284 | *C19orf55* |
| ENSG00000135838 | 1 | 182748428 | 182809519 | 127 | 16 | 32195 | 0.74403 | 0.22843 | *NPL* |
| ENSG00000165659 | 13 | 72002098 | 72451330 | 630 | 123 | 38429 | 0.744 | 0.22844 | *DACH1* |
| ENSG00000125089 | 4 | 8173799 | 8253530 | 300 | 64 | 40894 | 0.74389 | 0.22847 | *SH3TC1* |
| ENSG00000174502 | 1 | 205872176 | 205922588 | 202 | 37 | 36300 | 0.74377 | 0.22851 | *SLC26A9* |
| ENSG00000069702 | 1 | 92135902 | 92381892 | 840 | 106 | 46086 | 0.74357 | 0.22857 | *TGFBR3* |
| ENSG00000125823 | 20 | 23410322 | 23435567 | 80 | 15 | 35888 | 0.74336 | 0.22863 | *CSTL1* |
| ENSG00000183580 | 5 | 15490305 | 15949900 | 1118 | 86 | 51392 | 0.74292 | 0.22877 | *FBXL7* |
| ENSG00000155256 | 10 | 99486878 | 99530664 | 156 | 19 | 42845 | 0.74288 | 0.22878 | *ZFYVE27* |

| | | | | | | | | | |
|---|---|---|---|---|---|---|---|---|---|
| ENSG00000172687 | 19 | 21531732 | 21572104 | 213 | 10 | 46329 | 0.74287 | 0.22878 | *ZNF738* |
| ENSG00000166260 | 17 | 53019263 | 53056146 | 148 | 15 | 53859 | 0.74285 | 0.22879 | *COX11* |
| ENSG00000135617 | 2 | 73445134 | 73470366 | 62 | 15 | 42924 | 0.74265 | 0.22885 | *PRADC1* |
| ENSG00000249016 | 5 | 54242063 | 54263674 | 105 | 15 | 42476 | 0.74264 | 0.22885 | *RP11-45H22.3* |
| ENSG00000113810 | 3 | 160107062 | 160162750 | 117 | 19 | 50989 | 0.7424 | 0.22892 | *SMC4* |
| ENSG00000089060 | 12 | 113726564 | 113807298 | 238 | 31 | 51730 | 0.7424 | 0.22892 | *SLC8B1* |
| ENSG00000255994 | 14 | 70027483 | 70048032 | 40 | 9 | 44603 | 0.74233 | 0.22895 | *CCDC177* |
| ENSG00000131183 | 5 | 176796236 | 176835849 | 62 | 13 | 44363 | 0.74204 | 0.22903 | *SLC34A1* |
| ENSG00000186832 | 17 | 39756030 | 39782151 | 68 | 8 | 45747 | 0.74191 | 0.22907 | *KRT16* |
| ENSG00000176438 | 14 | 95873831 | 95952173 | 413 | 88 | 44222 | 0.74176 | 0.22912 | *SYNE3* |
| ENSG00000100031 | 22 | 24969718 | 25034972 | 127 | 29 | 40931 | 0.74174 | 0.22912 | *GGT1* |
| ENSG00000104885 | 19 | 2154148 | 2242577 | 314 | 44 | 44710 | 0.74129 | 0.22926 | *DOT1L* |
| ENSG00000158636 | 11 | 76145967 | 76274069 | 299 | 33 | 37982 | 0.74119 | 0.22929 | *C11orf30* |
| ENSG00000018625 | 1 | 160075549 | 160123381 | 179 | 32 | 54466 | 0.74074 | 0.22942 | *ATP1A2* |
| ENSG00000187238 | 1 | 152576287 | 152596574 | 64 | 6 | 49762 | 0.74072 | 0.22943 | *LCE3B* |
| ENSG00000119778 | 2 | 23961534 | 24159984 | 688 | 40 | 55097 | 0.74061 | 0.22947 | *ATAD2B* |
| ENSG00000134061 | 5 | 66468103 | 66502627 | 90 | 22 | 35022 | 0.7405 | 0.2295 | *CD180* |
| ENSG00000204149 | 10 | 51738078 | 51780259 | 95 | 20 | 31117 | 0.74018 | 0.2296 | *AGAP6* |
| ENSG00000102572 | 13 | 99092455 | 99240194 | 516 | 53 | 46653 | 0.74006 | 0.22963 | *STK24* |
| ENSG00000124160 | 20 | 44679624 | 44728591 | 136 | 17 | 50188 | 0.74008 | 0.22963 | *NCOA5* |
| ENSG00000120253 | 6 | 150035451 | 150080801 | 99 | 8 | 45716 | 0.74004 | 0.22964 | *NUP43* |
| ENSG00000196653 | 3 | 44744135 | 44775323 | 54 | 12 | 43048 | 0.7398 | 0.22971 | *ZNF502* |
| ENSG00000146376 | 6 | 129887277 | 130041370 | 581 | 84 | 51305 | 0.73931 | 0.22986 | *ARHGAP18* |
| ENSG00000039600 | 5 | 157042687 | 157108488 | 212 | 24 | 46831 | 0.73928 | 0.22987 | *SOX30* |
| ENSG00000110583 | 11 | 63696431 | 63734800 | 86 | 12 | 29000 | 0.73928 | 0.22987 | *NAA40* |
| ENSG00000181939 | 11 | 55311783 | 55332895 | 163 | 12 | 50351 | 0.73921 | 0.22989 | *OR4C15* |
| ENSG00000129910 | 16 | 89228175 | 89271900 | 173 | 23 | 44829 | 0.7392 | 0.22989 | *CDH15* |
| ENSG00000204510 | 1 | 12966450 | 12990569 | 3 | 2 | 32765 | 0.73918 | 0.2299 | *PRAMEF7* |
| ENSG00000148735 | 10 | 115501213 | 115553188 | 210 | 20 | 51795 | 0.73914 | 0.22991 | *PLEKHS1* |
| ENSG00000167566 | 12 | 50174929 | 50232533 | 178 | 25 | 41424 | 0.739 | 0.22995 | *NCKAP5L* |
| ENSG00000259207 | 17 | 45321212 | 45431658 | 417 | 42 | 41595 | 0.73899 | 0.22996 | *ITGB3* |
| ENSG00000259753 | 17 | 45321263 | 45431658 | 417 | 42 | 41595 | 0.73899 | 0.22996 | *ITGB3* |
| ENSG00000234278 | 13 | 57731331 | 57754352 | 26 | 8 | 43490 | 0.73867 | 0.23005 | *PRR20E* |
| ENSG00000163584 | 3 | 170572664 | 170598272 | 71 | 16 | 51152 | 0.73867 | 0.23006 | *RPL22L1* |
| ENSG00000142623 | 1 | 17521621 | 17582500 | 256 | 60 | 50801 | 0.73835 | 0.23015 | *PADI1* |
| ENSG00000188011 | 2 | 242801752 | 242825975 | 81 | 12 | 45713 | 0.73835 | 0.23015 | *CXXC11* |

| | | | | | | | | | |
|---|---|---|---|---|---|---|---|---|---|
| ENSG00000167323 | 11 | 3865757 | 4124439 | 711 | 59 | 38929 | 0.73832 | 0.23016 | *STIM1* |
| ENSG00000119638 | 14 | 75538822 | 75604047 | 140 | 11 | 44959 | 0.73806 | 0.23024 | *NEK9* |
| ENSG00000182263 | 2 | 164439906 | 164602522 | 337 | 64 | 45045 | 0.73796 | 0.23027 | *FIGN* |
| ENSG00000105327 | 19 | 47714081 | 47746023 | 57 | 19 | 46823 | 0.73788 | 0.23029 | *BBC3* |
| ENSG00000155508 | 5 | 154227113 | 154266353 | 92 | 11 | 32278 | 0.73765 | 0.23036 | *CNOT8* |
| ENSG00000186191 | 20 | 31657450 | 31709557 | 210 | 27 | 50418 | 0.73757 | 0.23039 | *BPIFB4* |
| ENSG00000255501 | 11 | 104998448 | 105020453 | 105 | 9 | 53113 | 0.73698 | 0.23057 | *CARD18* |
| ENSG00000143815 | 1 | 225579204 | 225626627 | 140 | 24 | 44435 | 0.73693 | 0.23058 | *LBR* |
| ENSG00000154710 | 7 | 66137151 | 66286451 | 568 | 34 | 52318 | 0.73687 | 0.2306 | *RABGEF1* |
| ENSG00000183172 | 22 | 42465695 | 42490288 | 62 | 9 | 48420 | 0.73675 | 0.23064 | *SMDT1* |
| ENSG00000268632 | 15 | 42063346 | 42086229 | 46 | 14 | 40715 | 0.73664 | 0.23067 | *AC073657.1* |
| ENSG00000064309 | 11 | 125815691 | 125943230 | 493 | 70 | 53549 | 0.73656 | 0.2307 | *CDON* |
| ENSG00000197938 | 3 | 97991732 | 98012676 | 119 | 13 | 28271 | 0.7364 | 0.23074 | *OR5H2* |
| ENSG00000104812 | 19 | 49461382 | 49506567 | 92 | 20 | 50422 | 0.7364 | 0.23074 | *GYS1* |
| ENSG00000213799 | 19 | 53827002 | 53868122 | 205 | 43 | 45091 | 0.73634 | 0.23076 | *ZNF845* |
| ENSG00000139914 | 14 | 24590484 | 24612058 | 42 | 7 | 30533 | 0.73615 | 0.23082 | *FITM1* |
| ENSG00000137124 | 9 | 38382661 | 38408658 | 121 | 39 | 48598 | 0.73589 | 0.2309 | *ALDH1B1* |
| ENSG00000128564 | 7 | 100795790 | 100818874 | 89 | 17 | 37943 | 0.73558 | 0.23099 | *VGF* |
| ENSG00000086848 | 11 | 111642919 | 111752305 | 232 | 21 | 38578 | 0.73552 | 0.23101 | *ALG9* |
| ENSG00000075188 | 12 | 102457967 | 102523902 | 138 | 20 | 55470 | 0.73518 | 0.23112 | *NUP37* |
| ENSG00000166589 | 16 | 66932025 | 66962887 | 51 | 12 | 31391 | 0.73502 | 0.23116 | *CDH16* |
| ENSG00000135094 | 12 | 113820250 | 113874106 | 138 | 16 | 44695 | 0.73467 | 0.23127 | *SDS* |
| ENSG00000115170 | 2 | 158582958 | 158742374 | 261 | 58 | 41600 | 0.73455 | 0.23131 | *ACVR1* |
| ENSG00000133069 | 1 | 205187304 | 205252471 | 195 | 23 | 32728 | 0.73447 | 0.23133 | *TMCC2* |
| ENSG00000100101 | 22 | 38067680 | 38180137 | 329 | 40 | 44796 | 0.73446 | 0.23133 | *NOL12* |
| ENSG00000169488 | 14 | 20433603 | 20454818 | 47 | 12 | 44188 | 0.73439 | 0.23136 | *OR4K15* |
| ENSG00000162714 | 1 | 247450714 | 247505148 | 154 | 21 | 43878 | 0.7341 | 0.23144 | *ZNF496* |
| ENSG00000221916 | 19 | 49611655 | 49632397 | 79 | 14 | 50439 | 0.73401 | 0.23147 | *C19orf73* |
| ENSG00000166145 | 15 | 41126216 | 41160405 | 92 | 15 | 29341 | 0.73351 | 0.23162 | *SPINT1* |
| ENSG00000172000 | 19 | 2857333 | 2888515 | 111 | 18 | 39971 | 0.73314 | 0.23174 | *ZNF556* |
| ENSG00000108107 | 19 | 55886713 | 55924612 | 184 | 35 | 38455 | 0.7331 | 0.23175 | *RPL28* |
| ENSG00000093167 | 3 | 37084117 | 37235180 | 323 | 22 | 38083 | 0.73301 | 0.23178 | *LRRFIP2* |
| ENSG00000166405 | 11 | 8117597 | 8200602 | 342 | 35 | 46142 | 0.73296 | 0.23179 | *RIC3* |
| ENSG00000150403 | 13 | 114135310 | 114214542 | 493 | 22 | 30910 | 0.73278 | 0.23185 | *TMCO3* |
| ENSG00000174498 | 15 | 65609465 | 65680378 | 187 | 39 | 50561 | 0.73275 | 0.23185 | *IGDCC3* |
| ENSG00000131050 | 20 | 31739574 | 31779218 | 59 | 14 | 35231 | 0.7326 | 0.2319 | *BPIFA2* |

| | | | | | | | | | |
|---|---|---|---|---|---|---|---|---|---|
| ENSG00000197959 | 1 | 171800621 | 172397606 | 1571 | 124 | 44922 | 0.73259 | 0.23191 | *DNM3* |
| ENSG00000110042 | 11 | 58928903 | 58986060 | 159 | 21 | 49290 | 0.73237 | 0.23197 | *DTX4* |
| ENSG00000155592 | 16 | 25237322 | 25279252 | 141 | 17 | 46482 | 0.73224 | 0.23201 | *ZKSCAN2* |
| ENSG00000226650 | 5 | 154383260 | 154407685 | 102 | 19 | 39683 | 0.73205 | 0.23207 | *KIF4B* |
| ENSG00000184389 | 1 | 33762367 | 33796699 | 102 | 16 | 45496 | 0.73197 | 0.23209 | *A3GALT2* |
| ENSG00000133019 | 1 | 239539865 | 240088750 | 1520 | 176 | 46854 | 0.73182 | 0.23214 | *CHRM3* |
| ENSG00000006704 | 7 | 73858120 | 74026931 | 348 | 62 | 45972 | 0.73165 | 0.23219 | *GTF2IRD1* |
| ENSG00000239332 | 2 | 47033836 | 47096146 | 272 | 51 | 45356 | 0.73102 | 0.23238 | *LINC01119* |
| ENSG00000107623 | 10 | 48415815 | 48448976 | 57 | 20 | 47181 | 0.73097 | 0.2324 | *GDF10* |
| ENSG00000144815 | 3 | 101488046 | 101557073 | 125 | 16 | 53336 | 0.73093 | 0.23241 | *NXPE3* |
| ENSG00000161973 | 17 | 8623252 | 8658537 | 140 | 22 | 48150 | 0.73093 | 0.23241 | *CCDC42* |
| ENSG00000144134 | 2 | 114374806 | 114410973 | 78 | 14 | 32068 | 0.73091 | 0.23242 | *RABL2A* |
| ENSG00000189171 | 1 | 153581263 | 153616568 | 82 | 11 | 46226 | 0.73084 | 0.23244 | *S100A13* |
| ENSG00000230989 | 16 | 83831448 | 83863342 | 157 | 31 | 51853 | 0.7308 | 0.23245 | *HSBP1* |
| ENSG00000029639 | 6 | 155568643 | 155645627 | 368 | 34 | 48538 | 0.73049 | 0.23255 | *TFB1M* |
| ENSG00000267596 | 17 | 34313476 | 34339084 | 117 | 15 | 43656 | 0.73048 | 0.23255 | *CCL15* |
| ENSG00000123080 | 1 | 51416417 | 51450305 | 47 | 13 | 50166 | 0.7304 | 0.23257 | *CDKN2C* |
| ENSG00000118507 | 6 | 131446806 | 131614675 | 427 | 49 | 50588 | 0.73039 | 0.23258 | *AKAP7* |
| ENSG00000180644 | 10 | 72347104 | 72372531 | 81 | 20 | 48641 | 0.73037 | 0.23258 | *PRF1* |
| ENSG00000089022 | 12 | 112269782 | 112344343 | 157 | 7 | 55083 | 0.73037 | 0.23258 | *MAPKAPK5* |
| ENSG00000125798 | 20 | 22551643 | 22576093 | 33 | 7 | 31412 | 0.73021 | 0.23263 | *FOXA2* |
| ENSG00000141219 | 17 | 71218372 | 71255091 | 154 | 8 | 49871 | 0.73018 | 0.23264 | *C17orf80* |
| ENSG00000197363 | 8 | 146014261 | 146046554 | 116 | 18 | 34739 | 0.73006 | 0.23268 | *ZNF517* |
| ENSG00000107186 | 9 | 13095703 | 13289589 | 334 | 50 | 37335 | 0.72985 | 0.23274 | *MPDZ* |
| ENSG00000172889 | 9 | 139543308 | 139577130 | 100 | 24 | 51471 | 0.72916 | 0.23295 | *EGFL7* |
| ENSG00000169045 | 5 | 179031179 | 179071785 | 171 | 27 | 43331 | 0.72903 | 0.23299 | *HNRNPH1* |
| ENSG00000174871 | 11 | 66035661 | 66062772 | 49 | 12 | 52115 | 0.72851 | 0.23315 | *CNIH2* |
| ENSG00000171209 | 4 | 71098305 | 71127145 | 188 | 12 | 37358 | 0.72828 | 0.23322 | *CSN3* |
| ENSG00000115053 | 2 | 232308242 | 232358352 | 183 | 27 | 48349 | 0.72825 | 0.23323 | *NCL* |
| ENSG00000133895 | 11 | 64560982 | 64588766 | 27 | 12 | 46254 | 0.72784 | 0.23335 | *MEN1* |
| ENSG00000123353 | 12 | 56201703 | 56225663 | 32 | 10 | 38007 | 0.72759 | 0.23343 | *ORMDL2* |
| ENSG00000120075 | 17 | 46658619 | 46681323 | 60 | 15 | 43644 | 0.72761 | 0.23343 | *HOXB5* |
| ENSG00000181562 | 14 | 21204051 | 21226539 | 146 | 29 | 52551 | 0.72755 | 0.23345 | *EDDM3A* |
| ENSG00000176915 | 12 | 133292254 | 133348474 | 246 | 44 | 45918 | 0.72741 | 0.23349 | *ANKLE2* |
| ENSG00000170777 | 9 | 6318349 | 6341900 | 57 | 15 | 44137 | 0.72737 | 0.2335 | *TPD52L3* |
| ENSG00000079335 | 1 | 100800584 | 100995833 | 363 | 35 | 34577 | 0.72727 | 0.23353 | *CDC14A* |

| | | | | | | | | | |
|---|---|---|---|---|---|---|---|---|---|
| ENSG00000100341 | 22 | 44265558 | 44297893 | 150 | 24 | 43002 | 0.72721 | 0.23355 | *PNPLA5* |
| ENSG00000214128 | 7 | 138472695 | 138532846 | 183 | 32 | 52030 | 0.7271 | 0.23358 | *TMEM213* |
| ENSG00000179674 | 3 | 160384948 | 160406233 | 73 | 13 | 46998 | 0.72692 | 0.23364 | *ARL14* |
| ENSG00000198707 | 12 | 88432793 | 88545993 | 126 | 24 | 50784 | 0.72683 | 0.23366 | *CEP290* |
| ENSG00000203784 | 1 | 153165919 | 153187596 | 65 | 14 | 42525 | 0.72679 | 0.23368 | *LELP1* |
| ENSG00000255072 | 4 | 89432724 | 89452940 | 19 | 10 | 38107 | 0.72676 | 0.23369 | *PIGY* |
| ENSG00000160818 | 1 | 156554279 | 156581288 | 108 | 8 | 22082 | 0.72665 | 0.23372 | *GPATCH4* |
| ENSG00000197013 | 19 | 21669484 | 21749072 | 291 | 17 | 54469 | 0.72664 | 0.23372 | *ZNF429* |
| ENSG00000141946 | 19 | 57635464 | 57666570 | 138 | 17 | 36255 | 0.72665 | 0.23372 | *ZIM3* |
| ENSG00000146648 | 7 | 55076714 | 55334313 | 846 | 132 | 44481 | 0.72659 | 0.23374 | *EGFR* |
| ENSG00000139684 | 13 | 47335391 | 47381367 | 96 | 15 | 35938 | 0.72655 | 0.23375 | *ESD* |
| ENSG00000182168 | 4 | 96073655 | 96480357 | 1485 | 180 | 41883 | 0.72647 | 0.23378 | *UNC5C* |
| ENSG00000135926 | 2 | 219128915 | 219167309 | 88 | 8 | 47291 | 0.72638 | 0.2338 | *TMBIM1* |
| ENSG00000135750 | 1 | 233739750 | 233818258 | 286 | 69 | 47745 | 0.72625 | 0.23384 | *KCNK1* |
| ENSG00000154222 | 1 | 52801395 | 52841865 | 39 | 15 | 48942 | 0.72582 | 0.23397 | *CC2D1B* |
| ENSG00000162669 | 1 | 91716323 | 91880426 | 432 | 38 | 49784 | 0.72578 | 0.23399 | *HFM1* |
| ENSG00000172977 | 11 | 65469467 | 65497075 | 101 | 17 | 38307 | 0.72534 | 0.23412 | *KAT5* |
| ENSG00000143786 | 1 | 224612362 | 224938251 | 915 | 100 | 45277 | 0.72522 | 0.23416 | *CNIH3* |
| ENSG00000136297 | 7 | 4935620 | 5008844 | 282 | 39 | 43591 | 0.72519 | 0.23417 | *MMD2* |
| ENSG00000164604 | 7 | 112708386 | 112737833 | 91 | 19 | 47514 | 0.72496 | 0.23424 | *GPR85* |
| ENSG00000214511 | 12 | 51337705 | 51374289 | 180 | 17 | 43504 | 0.72448 | 0.23439 | *HIGD1C* |
| ENSG00000196628 | 18 | 52879562 | 53342018 | 859 | 107 | 46166 | 0.72446 | 0.23439 | *TCF4* |
| ENSG00000171497 | 4 | 159620286 | 159654548 | 50 | 10 | 50767 | 0.72424 | 0.23446 | *PPID* |
| ENSG00000204099 | 2 | 242739920 | 242768739 | 102 | 22 | 47154 | 0.72398 | 0.23454 | *NEU4* |
| ENSG00000164615 | 5 | 134064191 | 134097847 | 40 | 17 | 42527 | 0.72373 | 0.23462 | *CAMLG* |
| ENSG00000242852 | 19 | 12561998 | 12634668 | 278 | 17 | 47943 | 0.7227 | 0.23493 | *ZNF709* |
| ENSG00000066629 | 14 | 100194030 | 100418397 | 696 | 91 | 41273 | 0.72241 | 0.23502 | *EML1* |
| ENSG00000177613 | 10 | 53445247 | 53469355 | 89 | 12 | 43965 | 0.72217 | 0.2351 | *CSTF2T* |
| ENSG00000126746 | 12 | 6765643 | 6808738 | 35 | 17 | 50653 | 0.72216 | 0.2351 | *ZNF384* |
| ENSG00000183354 | 9 | 5871596 | 6017901 | 340 | 35 | 42645 | 0.722 | 0.23515 | *KIAA2026* |
| ENSG00000167693 | 17 | 692553 | 893010 | 1121 | 170 | 41176 | 0.72114 | 0.23541 | *NXN* |
| ENSG00000164039 | 4 | 103990592 | 104031040 | 84 | 13 | 52555 | 0.72104 | 0.23544 | *BDH2* |
| ENSG00000078328 | 16 | 6059095 | 7773340 | 10865 | 562 | 41895 | 0.72089 | 0.23549 | *RBFOX1* |
| ENSG00000124103 | 20 | 55082243 | 55111198 | 61 | 13 | 34295 | 0.7209 | 0.23549 | *FAM209A* |
| ENSG00000122952 | 10 | 58106989 | 58131036 | 33 | 10 | 39580 | 0.72048 | 0.23561 | *ZWINT* |
| ENSG00000009413 | 6 | 111610234 | 111814918 | 523 | 46 | 43556 | 0.72027 | 0.23568 | *REV3L* |

| | | | | | | | | | |
|---|---|---|---|---|---|---|---|---|---|
| ENSG00000130956 | 9 | 99202483 | 99263618 | 140 | 12 | 35503 | 0.72026 | 0.23568 | *HABP4* |
| ENSG00000099937 | 22 | 21118167 | 21152008 | 108 | 13 | 53548 | 0.71998 | 0.23577 | *SERPIND1* |
| ENSG00000161328 | 11 | 527527 | 564916 | 209 | 30 | 50362 | 0.7198 | 0.23583 | *LRRC56* |
| ENSG00000122691 | 7 | 19050614 | 19167295 | 262 | 48 | 35031 | 0.71968 | 0.23586 | *TWIST1* |
| ENSG00000102755 | 13 | 28864489 | 29079265 | 613 | 84 | 43302 | 0.71955 | 0.2359 | *FLT1* |
| ENSG00000160813 | 7 | 100022905 | 100044188 | 40 | 6 | 54236 | 0.71939 | 0.23595 | *PPP1R35* |
| ENSG00000215912 | 1 | 2557415 | 2728286 | 368 | 31 | 41321 | 0.71907 | 0.23605 | *TTC34* |
| ENSG00000175398 | 12 | 56020644 | 56041638 | 79 | 13 | 45671 | 0.71901 | 0.23607 | *OR10P1* |
| ENSG00000125520 | 20 | 62361214 | 62384858 | 94 | 13 | 40571 | 0.71895 | 0.23609 | *SLC2A4RG* |
| ENSG00000127995 | 7 | 94128531 | 94196331 | 157 | 19 | 41478 | 0.71845 | 0.23624 | *CASD1* |
| ENSG00000081870 | 1 | 54372247 | 54421602 | 111 | 13 | 34924 | 0.71828 | 0.23629 | *HSPB11* |
| ENSG00000138303 | 10 | 73846278 | 73986892 | 191 | 31 | 38342 | 0.71818 | 0.23632 | *ASCC1* |
| ENSG00000117472 | 1 | 46630745 | 46661630 | 49 | 12 | 45528 | 0.71814 | 0.23633 | *TSPAN1* |
| ENSG00000124939 | 11 | 61966140 | 61991408 | 60 | 15 | 34591 | 0.71815 | 0.23633 | *SCGB2A1* |
| ENSG00000132749 | 11 | 68464908 | 68529032 | 128 | 13 | 52948 | 0.71783 | 0.23643 | *MTL5* |
| ENSG00000152082 | 2 | 130929310 | 130958302 | 70 | 7 | 49913 | 0.71762 | 0.2365 | *MZT2B* |
| ENSG00000111664 | 12 | 6939118 | 6966557 | 58 | 16 | 43622 | 0.71762 | 0.2365 | *GNB3* |
| ENSG00000196218 | 19 | 38914339 | 39088204 | 608 | 72 | 41145 | 0.71741 | 0.23656 | *RYR1* |
| ENSG00000109846 | 11 | 111769289 | 111804446 | 74 | 13 | 38710 | 0.71739 | 0.23657 | *CRYAB* |
| ENSG00000118873 | 1 | 220311635 | 220455796 | 327 | 35 | 46749 | 0.71734 | 0.23658 | *RAB3GAP2* |
| ENSG00000198250 | 10 | 47647581 | 47711443 | 81 | 25 | 39022 | 0.71733 | 0.23659 | *ANTXRL* |
| ENSG00000186399 | 15 | 30685943 | 30716463 | 1 | 1 | 42772 | 0.71728 | 0.2366 | *GOLGA8R* |
| ENSG00000183431 | 1 | 38412647 | 38466593 | 167 | 20 | 48366 | 0.71703 | 0.23668 | *SF3A3* |
| ENSG00000167792 | 11 | 67364323 | 67390006 | 71 | 10 | 39218 | 0.71694 | 0.23671 | *NDUFV1* |
| ENSG00000146049 | 6 | 24347131 | 24368512 | 70 | 14 | 38738 | 0.71675 | 0.23677 | *KAAG1* |
| ENSG00000180530 | 21 | 16323556 | 16447321 | 288 | 45 | 40768 | 0.7167 | 0.23678 | *NRIP1* |
| ENSG00000130733 | 19 | 11023444 | 11049357 | 58 | 12 | 56262 | 0.7166 | 0.23681 | *YIPF2* |
| ENSG00000147905 | 9 | 37110536 | 37368146 | 389 | 35 | 48032 | 0.71649 | 0.23685 | *ZCCHC7* |
| ENSG00000161040 | 7 | 102443308 | 102725286 | 498 | 38 | 46542 | 0.71645 | 0.23686 | *FBXL13* |
| ENSG00000170439 | 12 | 56065330 | 56088395 | 55 | 12 | 39766 | 0.71628 | 0.23691 | *METTL7B* |
| ENSG00000268537 | 3 | 46917699 | 46940171 | 46 | 13 | 41995 | 0.71625 | 0.23692 | *AC109583.1* |
| ENSG00000128011 | 19 | 39787208 | 39815976 | 54 | 15 | 45102 | 0.71609 | 0.23697 | *LRFN1* |
| ENSG00000156206 | 15 | 81289374 | 81451516 | 437 | 56 | 39011 | 0.71597 | 0.23701 | *C15orf26* |
| ENSG00000130363 | 6 | 159383903 | 159431219 | 272 | 11 | 42151 | 0.71593 | 0.23702 | *RSPH3* |
| ENSG00000179178 | 1 | 43725665 | 43749673 | 64 | 17 | 45398 | 0.71548 | 0.23716 | *TMEM125* |
| ENSG00000147912 | 9 | 37500889 | 37598871 | 320 | 42 | 47499 | 0.7154 | 0.23718 | *FBXO10* |

| | | | | | | | | | |
|---|---|---|---|---|---|---|---|---|---|
| ENSG00000159423 | 1 | 19187926 | 19239275 | 252 | 46 | 46890 | 0.71538 | 0.23719 | *ALDH4A1* |
| ENSG00000122718 | 9 | 35947105 | 35968151 | 112 | 16 | 51718 | 0.71538 | 0.23719 | *OR2S2* |
| ENSG00000138606 | 15 | 45449412 | 45503373 | 110 | 18 | 39993 | 0.71528 | 0.23722 | *SHF* |
| ENSG00000141127 | 17 | 18733398 | 18844581 | 363 | 23 | 53220 | 0.71495 | 0.23732 | *PRPSAP2* |
| ENSG00000163807 | 3 | 44769153 | 44813154 | 71 | 12 | 49709 | 0.71484 | 0.23735 | *KIAA1143* |
| ENSG00000183688 | 17 | 279769 | 305730 | 66 | 13 | 33533 | 0.71456 | 0.23744 | *FAM101B* |
| ENSG00000111912 | 6 | 126092307 | 126262266 | 340 | 45 | 44313 | 0.71447 | 0.23747 | *NCOA7* |
| ENSG00000150676 | 11 | 85556144 | 85641064 | 304 | 28 | 52666 | 0.71445 | 0.23747 | *CCDC83* |
| ENSG00000081138 | 18 | 63407488 | 63558638 | 514 | 43 | 35164 | 0.71429 | 0.23752 | *CDH7* |
| ENSG00000273049 | 12 | 54369629 | 54438672 | 134 | 29 | 48817 | 0.71419 | 0.23755 | *RP11-834C11.12* |
| ENSG00000075391 | 1 | 178052864 | 178458644 | 739 | 54 | 45915 | 0.714 | 0.23761 | *RASAL2* |
| ENSG00000204438 | 6 | 31619006 | 31644060 | 90 | 22 | 37436 | 0.71375 | 0.23769 | *GPANK1* |
| ENSG00000104142 | 15 | 41176628 | 41206173 | 55 | 10 | 33609 | 0.71375 | 0.23769 | *VPS18* |
| ENSG00000170866 | 19 | 54789854 | 54819952 | 127 | 16 | 28374 | 0.7134 | 0.2378 | *LILRA3* |
| ENSG00000198844 | 17 | 8203559 | 8235829 | 108 | 24 | 50098 | 0.71337 | 0.23781 | *ARHGEF15* |
| ENSG00000213316 | 5 | 179210981 | 179233648 | 75 | 20 | 46433 | 0.71312 | 0.23788 | *LTC4S* |
| ENSG00000176890 | 18 | 647604 | 683578 | 147 | 15 | 36274 | 0.71313 | 0.23788 | *TYMS* |
| ENSG00000217930 | 16 | 4371550 | 4415608 | 123 | 29 | 42761 | 0.7129 | 0.23795 | *PAM16* |
| ENSG00000176219 | 14 | 20681791 | 20702964 | 188 | 22 | 45442 | 0.71285 | 0.23797 | *OR11H6* |
| ENSG00000168140 | 16 | 4411849 | 4443529 | 94 | 19 | 40223 | 0.71273 | 0.23801 | *VASN* |
| ENSG00000131788 | 1 | 145565233 | 145596546 | 45 | 9 | 44166 | 0.71268 | 0.23802 | *PIAS3* |
| ENSG00000184436 | 22 | 21343393 | 21366485 | 112 | 20 | 44326 | 0.71254 | 0.23807 | *THAP7* |
| ENSG00000130382 | 19 | 6202966 | 6289959 | 363 | 39 | 36111 | 0.71243 | 0.2381 | *MLLT1* |
| ENSG00000266258 | 18 | 67992675 | 68029695 | 148 | 19 | 39199 | 0.71232 | 0.23813 | *RP11-41O4.1* |
| ENSG00000144136 | 2 | 113393434 | 113431404 | 108 | 11 | 30716 | 0.71223 | 0.23816 | *SLC20A1* |
| ENSG00000144227 | 2 | 139418342 | 139547918 | 343 | 60 | 48738 | 0.71167 | 0.23833 | *NXPH2* |
| ENSG00000118308 | 12 | 25163936 | 25271268 | 463 | 77 | 47257 | 0.71161 | 0.23835 | *LRMP* |
| ENSG00000158859 | 1 | 161144098 | 161178846 | 71 | 20 | 54192 | 0.71151 | 0.23838 | *ADAMTS4* |
| ENSG00000228804 | 3 | 187410101 | 187461637 | 202 | 28 | 39932 | 0.71147 | 0.2384 | *RP11-211G3.3* |
| ENSG00000124216 | 20 | 48589536 | 48615423 | 114 | 14 | 33057 | 0.71133 | 0.23844 | *SNAI1* |
| ENSG00000184925 | 9 | 139834003 | 139859949 | 107 | 15 | 47773 | 0.71131 | 0.23845 | *LCN12* |
| ENSG00000124787 | 6 | 4984966 | 5014297 | 181 | 18 | 52890 | 0.71082 | 0.2386 | *RPP40* |
| ENSG00000249222 | 22 | 43025809 | 43046607 | 63 | 10 | 48263 | 0.71065 | 0.23865 | *ATP5L2* |
| ENSG00000116729 | 1 | 68554142 | 68708803 | 656 | 111 | 47574 | 0.71054 | 0.23869 | *WLS* |
| ENSG00000110987 | 12 | 122447328 | 122509948 | 177 | 30 | 44295 | 0.71029 | 0.23876 | *BCL7A* |
| ENSG00000125492 | 9 | 135447572 | 135475653 | 60 | 21 | 43855 | 0.7096 | 0.23898 | *BARHL1* |

| | | | | | | | | | |
|---|---|---|---|---|---|---|---|---|---|
| ENSG00000168243 | 1 | 235700987 | 235824054 | 400 | 59 | 43376 | 0.70954 | 0.23899 | *GNG4* |
| ENSG00000056558 | 9 | 123654671 | 123701451 | 152 | 15 | 47254 | 0.70944 | 0.23902 | *TRAF1* |
| ENSG00000183186 | 19 | 395438 | 419139 | 125 | 42 | 40208 | 0.7093 | 0.23907 | *C2CD4C* |
| ENSG00000137038 | 9 | 7786490 | 7898380 | 422 | 66 | 50564 | 0.70926 | 0.23908 | *TMEM261* |
| ENSG00000180626 | 17 | 5072831 | 5105178 | 65 | 13 | 36781 | 0.70827 | 0.23939 | *ZNF594* |
| ENSG00000221962 | 3 | 152047484 | 152068779 | 44 | 11 | 51537 | 0.70815 | 0.23942 | *TMEM14E* |
| ENSG00000058600 | 16 | 22298730 | 22356424 | 85 | 24 | 41470 | 0.70771 | 0.23956 | *POLR3E* |
| ENSG00000223501 | 6 | 33208049 | 33249824 | 129 | 16 | 39594 | 0.70696 | 0.2398 | *VPS52* |
| ENSG00000267952 | 19 | 7561938 | 7594508 | 94 | 18 | 52309 | 0.70669 | 0.23988 | *CTD-2207O23.12* |
| ENSG00000143105 | 1 | 111049839 | 111071797 | 75 | 21 | 43382 | 0.70655 | 0.23992 | *KCNA10* |
| ENSG00000187815 | 1 | 40932887 | 40972015 | 132 | 21 | 43271 | 0.70635 | 0.23999 | *ZFP69* |
| ENSG00000172613 | 11 | 67149176 | 67175881 | 38 | 9 | 44275 | 0.70626 | 0.24001 | *RAD9A* |
| ENSG00000135365 | 11 | 45940871 | 46152985 | 342 | 31 | 32425 | 0.70621 | 0.24003 | *PHF21A* |
| ENSG00000256525 | 17 | 62463902 | 62503154 | 87 | 10 | 32657 | 0.70605 | 0.24008 | *POLG2* |
| ENSG00000166167 | 10 | 103103820 | 103327078 | 528 | 26 | 32020 | 0.70603 | 0.24009 | *BTRC* |
| ENSG00000083642 | 13 | 33150564 | 33362157 | 506 | 23 | 42725 | 0.7059 | 0.24013 | *PDS5B* |
| ENSG00000089876 | 10 | 127514906 | 127595005 | 119 | 19 | 53332 | 0.70577 | 0.24016 | *DHX32* |
| ENSG00000091129 | 7 | 107778068 | 108107161 | 1315 | 93 | 43244 | 0.70557 | 0.24023 | *NRCAM* |
| ENSG00000197238 | 6 | 27781884 | 27802257 | 57 | 14 | 39025 | 0.70545 | 0.24026 | *HIST1H4J* |
| ENSG00000074201 | 11 | 77215981 | 77358850 | 339 | 30 | 46728 | 0.70537 | 0.24029 | *CLNS1A* |
| ENSG00000117016 | 1 | 41076351 | 41141329 | 187 | 30 | 46925 | 0.70535 | 0.2403 | *RIMS3* |
| ENSG00000156096 | 4 | 70335883 | 70401732 | 386 | 17 | 50905 | 0.70534 | 0.2403 | *UGT2B4* |
| ENSG00000074696 | 15 | 65812756 | 65880687 | 164 | 13 | 45539 | 0.705 | 0.2404 | *PTPLAD1* |
| ENSG00000105088 | 19 | 9954394 | 10057228 | 276 | 42 | 44035 | 0.70473 | 0.24049 | *OLFM2* |
| ENSG00000162595 | 1 | 68501645 | 68527314 | 34 | 5 | 27814 | 0.7046 | 0.24053 | *DIRAS3* |
| ENSG00000136522 | 3 | 179296073 | 179332442 | 71 | 7 | 51046 | 0.70449 | 0.24056 | *MRPL47* |
| ENSG00000023608 | 14 | 62219075 | 62273146 | 186 | 16 | 46049 | 0.7045 | 0.24056 | *SNAPC1* |
| ENSG00000151923 | 10 | 121324199 | 121366541 | 97 | 17 | 49248 | 0.70446 | 0.24057 | *TIAL1* |
| ENSG00000163909 | 1 | 40079825 | 40115617 | 101 | 30 | 44098 | 0.70445 | 0.24058 | *HEYL* |
| ENSG00000123643 | 5 | 150806607 | 150881942 | 254 | 33 | 41753 | 0.70439 | 0.24059 | *SLC36A1* |
| ENSG00000196374 | 6 | 27772822 | 27793267 | 51 | 11 | 42573 | 0.70438 | 0.2406 | *HIST1H2BM* |
| ENSG00000088926 | 4 | 187177099 | 187220835 | 170 | 35 | 47646 | 0.70422 | 0.24065 | *F11* |
| ENSG00000051596 | 5 | 175334876 | 175471683 | 48 | 21 | 30077 | 0.7042 | 0.24065 | *THOC3* |
| ENSG00000022277 | 20 | 55033647 | 55103943 | 199 | 15 | 32480 | 0.70413 | 0.24068 | *RTFDC1* |
| ENSG00000147475 | 8 | 37584117 | 37626619 | 30 | 17 | 55936 | 0.70384 | 0.24076 | *ERLIN2* |
| ENSG00000256029 | 1 | 159817690 | 159852843 | 109 | 20 | 44251 | 0.70375 | 0.24079 | *RP11-190A12.7* |

| | | | | | | | | | |
|---|---|---|---|---|---|---|---|---|---|
| ENSG00000054793 | 20 | 50203053 | 50395173 | 949 | 73 | 48279 | 0.70357 | 0.24085 | *ATP9A* |
| ENSG00000099991 | 22 | 24397642 | 24584596 | 351 | 44 | 48097 | 0.70359 | 0.24085 | *CABIN1* |
| ENSG00000179902 | 1 | 109638573 | 109666479 | 116 | 20 | 39597 | 0.70355 | 0.24086 | *C1orf194* |
| ENSG00000214597 | 8 | 145565878 | 145588574 | 39 | 12 | 47397 | 0.70336 | 0.24092 | *TMEM249* |
| ENSG00000113805 | 3 | 74301719 | 74580291 | 777 | 115 | 41649 | 0.70333 | 0.24093 | *CNTN3* |
| ENSG00000163694 | 4 | 40415272 | 40642892 | 724 | 146 | 43649 | 0.70318 | 0.24097 | *RBM47* |
| ENSG00000130511 | 19 | 18519674 | 18555372 | 70 | 12 | 38951 | 0.70316 | 0.24098 | *SSBP4* |
| ENSG00000181009 | 11 | 5788864 | 5809897 | 35 | 7 | 43625 | 0.70304 | 0.24102 | *OR52N5* |
| ENSG00000162482 | 1 | 19599052 | 19625744 | 130 | 9 | 55519 | 0.70289 | 0.24106 | *AKR7A3* |
| ENSG00000187772 | 6 | 105394923 | 105541207 | 242 | 42 | 47720 | 0.70288 | 0.24107 | *LIN28B* |
| ENSG00000162231 | 11 | 62549595 | 62583774 | 52 | 9 | 36412 | 0.70283 | 0.24108 | *NXF1* |
| ENSG00000174851 | 11 | 66042051 | 66066641 | 37 | 10 | 49189 | 0.70283 | 0.24108 | *YIF1A* |
| ENSG00000172115 | 7 | 25149710 | 25174980 | 65 | 13 | 43890 | 0.70276 | 0.2411 | *CYCS* |
| ENSG00000159399 | 2 | 75051108 | 75130486 | 243 | 47 | 46870 | 0.7023 | 0.24125 | *HK2* |
| ENSG00000267157 | 19 | 5774016 | 5795630 | 100 | 15 | 32644 | 0.70219 | 0.24128 | *CTB-54O9.9* |
| ENSG00000130711 | 9 | 133529981 | 133568368 | 122 | 23 | 35454 | 0.70195 | 0.24136 | *PRDM12* |
| ENSG00000137251 | 6 | 54162657 | 54264950 | 329 | 39 | 38638 | 0.70191 | 0.24137 | *TINAG* |
| ENSG00000130584 | 20 | 62365019 | 62472597 | 527 | 33 | 43696 | 0.70181 | 0.2414 | *ZBTB46* |
| ENSG00000118680 | 18 | 3251907 | 3288282 | 143 | 17 | 40337 | 0.70178 | 0.24141 | *MYL12B* |
| ENSG00000164078 | 3 | 49914435 | 49951299 | 49 | 9 | 37897 | 0.70144 | 0.24151 | *MST1R* |
| ENSG00000150783 | 11 | 112028095 | 112053282 | 77 | 14 | 43763 | 0.70144 | 0.24151 | *TEX12* |
| ENSG00000256222 | 20 | 55923496 | 55944878 | 99 | 9 | 47716 | 0.70147 | 0.24151 | *MTRNR2L3* |
| ENSG00000148120 | 9 | 97478983 | 97859441 | 903 | 64 | 40340 | 0.70131 | 0.24155 | *C9orf3* |
| ENSG00000132329 | 2 | 238757536 | 238830756 | 282 | 69 | 43714 | 0.70127 | 0.24157 | *RAMP1* |
| ENSG00000186400 | 1 | 158538709 | 158559689 | 72 | 16 | 49427 | 0.70123 | 0.24158 | *OR10X1* |
| ENSG00000185869 | 19 | 37369026 | 37417193 | 170 | 18 | 34631 | 0.7011 | 0.24162 | *ZNF829* |
| ENSG00000205531 | 11 | 2955667 | 3023607 | 273 | 28 | 40659 | 0.70099 | 0.24166 | *NAP1L4* |
| ENSG00000134278 | 18 | 12436511 | 12668133 | 787 | 58 | 40790 | 0.70091 | 0.24168 | *SPIRE1* |
| ENSG00000164841 | 8 | 109609079 | 109809844 | 686 | 69 | 47081 | 0.70056 | 0.24179 | *TMEM74* |
| ENSG00000109814 | 4 | 39490375 | 39539931 | 222 | 29 | 37313 | 0.70048 | 0.24181 | *UGDH* |
| ENSG00000169427 | 8 | 140603081 | 140725299 | 326 | 60 | 42511 | 0.70047 | 0.24182 | *KCNK9* |
| ENSG00000170271 | 5 | 154188051 | 154248812 | 102 | 16 | 41116 | 0.70042 | 0.24183 | *FAXDC2* |
| ENSG00000141401 | 18 | 11971024 | 12040876 | 404 | 51 | 32766 | 0.70019 | 0.24191 | *IMPA2* |
| ENSG00000157368 | 16 | 70603798 | 70704585 | 365 | 39 | 41303 | 0.70008 | 0.24194 | *IL34* |
| ENSG00000203871 | 6 | 88096840 | 88119467 | 118 | 7 | 51082 | 0.69965 | 0.24207 | *C6orf164* |
| ENSG00000180891 | 17 | 55928604 | 56042684 | 310 | 63 | 44961 | 0.69967 | 0.24207 | *CUEDC1* |

| | | | | | | | | | |
|---|---|---|---|---|---|---|---|---|---|
| ENSG00000237435 | 1 | 96393457 | 96498436 | 354 | 36 | 54121 | 0.69949 | 0.24212 | *RP11-147C23.1* |
| ENSG00000108239 | 10 | 96152261 | 96305687 | 289 | 25 | 51800 | 0.6995 | 0.24212 | *TBC1D12* |
| ENSG00000175602 | 11 | 65647875 | 65669105 | 50 | 12 | 38525 | 0.69926 | 0.2422 | *CCDC85B* |
| ENSG00000114547 | 3 | 125677987 | 125712297 | 109 | 17 | 47481 | 0.69905 | 0.24226 | *ROPN1B* |
| ENSG00000158113 | 12 | 122642285 | 122698018 | 163 | 24 | 45007 | 0.69889 | 0.24231 | *LRRC43* |
| ENSG00000126861 | 17 | 29589031 | 29634557 | 67 | 8 | 30764 | 0.6988 | 0.24234 | *OMG* |
| ENSG00000113088 | 5 | 54310081 | 54340398 | 80 | 14 | 33920 | 0.69875 | 0.24235 | *GZMK* |
| ENSG00000118418 | 6 | 79900962 | 79954406 | 123 | 18 | 45957 | 0.6985 | 0.24243 | *HMGN3* |
| ENSG00000156928 | 7 | 23328358 | 23361348 | 83 | 15 | 31458 | 0.69808 | 0.24256 | *MALSU1* |
| ENSG00000148426 | 10 | 11855338 | 11924276 | 254 | 38 | 52392 | 0.6981 | 0.24256 | *PROSER2* |
| ENSG00000213928 | 14 | 24620262 | 24645774 | 51 | 13 | 49763 | 0.69808 | 0.24256 | *IRF9* |
| ENSG00000135624 | 2 | 73450548 | 73490149 | 112 | 19 | 36156 | 0.69798 | 0.2426 | *CCT7* |
| ENSG00000133328 | 11 | 63310242 | 63340855 | 55 | 16 | 40774 | 0.69795 | 0.2426 | *HRASLS2* |
| ENSG00000161016 | 8 | 146005150 | 146027972 | 89 | 14 | 33710 | 0.69788 | 0.24263 | *RPL8* |
| ENSG00000271698 | 8 | 145566888 | 145589269 | 37 | 11 | 46917 | 0.6974 | 0.24278 | *GS1-393G12.13* |
| ENSG00000175334 | 11 | 65759550 | 65781620 | 83 | 11 | 40768 | 0.6974 | 0.24278 | *BANF1* |
| ENSG00000198265 | 17 | 65056554 | 65252105 | 414 | 38 | 39199 | 0.69738 | 0.24278 | *HELZ* |
| ENSG00000007923 | 1 | 6684228 | 6771984 | 216 | 24 | 54575 | 0.69722 | 0.24283 | *DNAJC11* |
| ENSG00000112276 | 6 | 105534697 | 105595049 | 204 | 21 | 33607 | 0.69713 | 0.24286 | *BVES* |
| ENSG00000188580 | 6 | 124115286 | 125156803 | 3197 | 299 | 45439 | 0.69693 | 0.24292 | *NKAIN2* |
| ENSG00000161574 | 17 | 34300692 | 34338660 | 169 | 26 | 43636 | 0.69688 | 0.24294 | *CCL15-CCL14* |
| ENSG00000157483 | 15 | 59417113 | 59675099 | 846 | 121 | 35661 | 0.69686 | 0.24295 | *MYO1E* |
| ENSG00000167548 | 12 | 49402758 | 49463557 | 63 | 11 | 49910 | 0.69679 | 0.24297 | *KMT2D* |
| ENSG00000133056 | 1 | 204381756 | 204473852 | 228 | 31 | 36136 | 0.69643 | 0.24308 | *PIK3C2B* |
| ENSG00000206013 | 11 | 288200 | 309526 | 77 | 24 | 33177 | 0.69612 | 0.24318 | *IFITM5* |
| ENSG00000126705 | 1 | 27850546 | 27940942 | 94 | 25 | 42385 | 0.69595 | 0.24323 | *AHDC1* |
| ENSG00000149548 | 11 | 124814017 | 124921385 | 401 | 28 | 37554 | 0.69594 | 0.24323 | *CCDC15* |
| ENSG00000105352 | 19 | 42115344 | 42143442 | 99 | 21 | 44495 | 0.69587 | 0.24325 | *CEACAM4* |
| ENSG00000171401 | 17 | 39647233 | 39671957 | 69 | 23 | 48484 | 0.69585 | 0.24326 | *KRT13* |
| ENSG00000015153 | 12 | 42540906 | 42642151 | 231 | 27 | 45589 | 0.69574 | 0.2433 | *YAF2* |
| ENSG00000175066 | 3 | 141872414 | 141954449 | 232 | 34 | 48458 | 0.69567 | 0.24332 | *GK5* |
| ENSG00000126500 | 11 | 63860660 | 63896645 | 114 | 21 | 38778 | 0.69562 | 0.24333 | *FLRT1* |
| ENSG00000016402 | 6 | 137311108 | 137376298 | 168 | 32 | 38959 | 0.69554 | 0.24336 | *IL20RA* |
| ENSG00000221858 | 7 | 143782141 | 143803186 | 52 | 6 | 49341 | 0.69536 | 0.24341 | *OR2A12* |
| ENSG00000189337 | 1 | 14915200 | 15454539 | 2023 | 270 | 44991 | 0.69515 | 0.24348 | *KAZN* |
| ENSG00000096070 | 6 | 36154521 | 36210567 | 77 | 16 | 49365 | 0.69511 | 0.24349 | *BRPF3* |

| | | | | | | | | | |
|---|---|---|---|---|---|---|---|---|---|
| ENSG00000180878 | 11 | 6216796 | 6242362 | 62 | 13 | 32128 | 0.69504 | 0.24352 | *C11orf42* |
| ENSG00000158373 | 6 | 26148349 | 26181577 | 52 | 14 | 42464 | 0.69499 | 0.24353 | *HIST1H2BD* |
| ENSG00000139926 | 14 | 51945818 | 52207445 | 977 | 153 | 50412 | 0.69496 | 0.24354 | *FRMD6* |
| ENSG00000143164 | 1 | 167895021 | 168055081 | 254 | 24 | 43301 | 0.6949 | 0.24356 | *DCAF6* |
| ENSG00000174527 | 12 | 109775708 | 109903328 | 651 | 57 | 48690 | 0.69491 | 0.24356 | *MYO1H* |
| ENSG00000197283 | 6 | 33377847 | 33431466 | 108 | 23 | 49495 | 0.69477 | 0.2436 | *SYNGAP1* |
| ENSG00000101230 | 20 | 13192418 | 13291298 | 434 | 72 | 51237 | 0.69462 | 0.24365 | *ISM1* |
| ENSG00000170584 | 5 | 162863532 | 162897146 | 71 | 14 | 41813 | 0.69449 | 0.24369 | *NUDCD2* |
| ENSG00000165891 | 12 | 77405027 | 77469360 | 229 | 53 | 43528 | 0.69444 | 0.2437 | *E2F7* |
| ENSG00000187166 | 12 | 48712763 | 48734062 | 73 | 12 | 46844 | 0.69441 | 0.24371 | *H1FNT* |
| ENSG00000152133 | 2 | 37301594 | 37336387 | 92 | 15 | 43971 | 0.69361 | 0.24396 | *GPATCH11* |
| ENSG00000186919 | 17 | 74058042 | 74089747 | 69 | 9 | 42780 | 0.69362 | 0.24396 | *ZACN* |
| ENSG00000105617 | 19 | 54648899 | 54673620 | 70 | 20 | 41362 | 0.69363 | 0.24396 | *LENG1* |
| ENSG00000136492 | 17 | 59748627 | 59950882 | 372 | 35 | 48945 | 0.69354 | 0.24398 | *BRIP1* |
| ENSG00000241978 | 9 | 112532769 | 112944792 | 1444 | 173 | 46449 | 0.69351 | 0.244 | *AKAP2* |
| ENSG00000109445 | 4 | 142132041 | 142165851 | 96 | 9 | 45181 | 0.69294 | 0.24417 | *ZNF330* |
| ENSG00000183691 | 17 | 54661060 | 54682951 | 43 | 12 | 40463 | 0.69294 | 0.24417 | *NOG* |
| ENSG00000242550 | 18 | 61554408 | 61613345 | 232 | 9 | 44545 | 0.69294 | 0.24417 | *SERPINB10* |
| ENSG00000197632 | 18 | 61528926 | 61581124 | 172 | 7 | 47136 | 0.69292 | 0.24418 | *SERPINB2* |
| ENSG00000173482 | 18 | 7556780 | 8416859 | 2711 | 275 | 44790 | 0.69283 | 0.24421 | *PTPRM* |
| ENSG00000065357 | 12 | 56311103 | 56357811 | 52 | 14 | 53798 | 0.69273 | 0.24424 | *DGKA* |
| ENSG00000269563 | 2 | 47789601 | 47811998 | 79 | 24 | 50653 | 0.69262 | 0.24427 | *AC138655.1* |
| ENSG00000197045 | 14 | 54931202 | 54965914 | 62 | 11 | 45605 | 0.69256 | 0.24429 | *GMFB* |
| ENSG00000175928 | 3 | 3831121 | 3899387 | 210 | 32 | 29676 | 0.69254 | 0.2443 | *LRRN1* |
| ENSG00000085465 | 1 | 111946936 | 111980399 | 122 | 16 | 37113 | 0.69225 | 0.24439 | *OVGP1* |
| ENSG00000134242 | 1 | 114346433 | 114424381 | 154 | 20 | 39946 | 0.69215 | 0.24442 | *PTPN22* |
| ENSG00000158711 | 1 | 205567071 | 205611090 | 139 | 25 | 41651 | 0.69203 | 0.24446 | *ELK4* |
| ENSG00000102805 | 13 | 77554795 | 77586652 | 109 | 18 | 43551 | 0.69188 | 0.24451 | *CLN5* |
| ENSG00000114026 | 3 | 9781628 | 9839903 | 131 | 24 | 50207 | 0.69163 | 0.24458 | *OGG1* |
| ENSG00000184374 | 8 | 119997691 | 120128821 | 689 | 45 | 44965 | 0.69157 | 0.2446 | *COLEC10* |
| ENSG00000177156 | 11 | 737329 | 775024 | 90 | 16 | 35241 | 0.69155 | 0.24461 | *TALDO1* |
| ENSG00000228278 | 9 | 117082149 | 117105532 | 48 | 17 | 46059 | 0.69151 | 0.24462 | *ORM2* |
| ENSG00000167613 | 19 | 54855362 | 54892165 | 159 | 35 | 42529 | 0.69152 | 0.24462 | *LAIR1* |
| ENSG00000163554 | 1 | 158570496 | 158666488 | 273 | 33 | 45648 | 0.69125 | 0.2447 | *SPTA1* |
| ENSG00000213494 | 17 | 34300327 | 34339100 | 171 | 26 | 43539 | 0.69107 | 0.24476 | *CCL14* |
| ENSG00000110195 | 11 | 71890602 | 71917345 | 79 | 17 | 39158 | 0.69092 | 0.24481 | *FOLR1* |

| | | | | | | | | | |
|---|---|---|---|---|---|---|---|---|---|
| ENSG00000160062 | 1 | 32995028 | 33081540 | 233 | 16 | 51689 | 0.69078 | 0.24485 | *ZBTB8A* |
| ENSG00000227868 | 1 | 23327327 | 23352343 | 41 | 7 | 34932 | 0.69069 | 0.24488 | *C1orf234* |
| ENSG00000095906 | 16 | 1822902 | 1849192 | 151 | 18 | 36002 | 0.6907 | 0.24488 | *NUBP2* |
| ENSG00000101470 | 20 | 44441853 | 44472384 | 95 | 19 | 46683 | 0.6907 | 0.24488 | *TNNC2* |
| ENSG00000173262 | 12 | 7955108 | 8053744 | 439 | 67 | 44855 | 0.69052 | 0.24493 | *SLC2A14* |
| ENSG00000157654 | 9 | 112532589 | 112944792 | 1445 | 173 | 46457 | 0.69043 | 0.24496 | *PALM2-AKAP2* |
| ENSG00000104267 | 8 | 86366081 | 86403722 | 68 | 15 | 51855 | 0.69036 | 0.24498 | *CA2* |
| ENSG00000087586 | 20 | 54934445 | 54977393 | 101 | 29 | 43034 | 0.6903 | 0.245 | *AURKA* |
| ENSG00000157601 | 21 | 42782231 | 42841141 | 259 | 30 | 31589 | 0.69019 | 0.24504 | *MX1* |
| ENSG00000128383 | 22 | 39338746 | 39369188 | 72 | 18 | 46425 | 0.6899 | 0.24513 | *APOBEC3A* |
| ENSG00000133935 | 14 | 76106134 | 76137532 | 67 | 17 | 43384 | 0.68985 | 0.24514 | *C14orf1* |
| ENSG00000241553 | 3 | 9824179 | 9859410 | 92 | 18 | 48344 | 0.68955 | 0.24524 | *ARPC4* |
| ENSG00000103647 | 15 | 68861308 | 69030145 | 545 | 68 | 39514 | 0.68938 | 0.24529 | *CORO2B* |
| ENSG00000173988 | 13 | 46776083 | 46861501 | 309 | 39 | 38360 | 0.68893 | 0.24543 | *LRRC63* |
| ENSG00000182831 | 16 | 9175505 | 9225497 | 230 | 27 | 39771 | 0.68889 | 0.24545 | *C16orf72* |
| ENSG00000130592 | 11 | 1864200 | 1923497 | 241 | 42 | 47584 | 0.6887 | 0.24551 | *LSP1* |
| ENSG00000186666 | 12 | 50221573 | 50246912 | 62 | 14 | 51061 | 0.6883 | 0.24563 | *BCDIN3D* |
| ENSG00000168918 | 2 | 233914677 | 234126549 | 739 | 95 | 37765 | 0.68824 | 0.24565 | *INPP5D* |
| ENSG00000170854 | 3 | 97650662 | 97701301 | 74 | 17 | 53667 | 0.68825 | 0.24565 | *MINA* |
| ENSG00000171217 | 6 | 155575147 | 155607682 | 174 | 17 | 50622 | 0.68822 | 0.24566 | *CLDN20* |
| ENSG00000185127 | 6 | 170092233 | 170116401 | 52 | 12 | 35921 | 0.688 | 0.24573 | *C6orf120* |
| ENSG00000127946 | 7 | 75152621 | 75378280 | 838 | 93 | 48842 | 0.6878 | 0.24579 | *HIP1* |
| ENSG00000267772 | 17 | 58631908 | 58673984 | 38 | 14 | 33792 | 0.68773 | 0.24581 | *RP11-15E18.4* |
| ENSG00000152217 | 18 | 42250138 | 42658475 | 867 | 142 | 51599 | 0.68765 | 0.24584 | *SETBP1* |
| ENSG00000187145 | 1 | 150256289 | 150291414 | 128 | 13 | 33773 | 0.68756 | 0.24587 | *MRPS21* |
| ENSG00000122176 | 1 | 203299756 | 203330617 | 91 | 19 | 41764 | 0.68753 | 0.24587 | *FMOD* |
| ENSG00000184863 | 7 | 155427145 | 155584179 | 492 | 27 | 45380 | 0.68712 | 0.246 | *RBM33* |
| ENSG00000223496 | 16 | 70274134 | 70295833 | 21 | 12 | 35571 | 0.68709 | 0.24601 | *EXOSC6* |
| ENSG00000078699 | 20 | 32067881 | 32247842 | 165 | 33 | 49421 | 0.68696 | 0.24605 | *CBFA2T2* |
| ENSG00000146834 | 7 | 100016413 | 100041741 | 45 | 6 | 53342 | 0.68695 | 0.24606 | *MEPCE* |
| ENSG00000158122 | 9 | 99391859 | 99427585 | 27 | 16 | 43574 | 0.6869 | 0.24607 | *AAED1* |
| ENSG00000171984 | 20 | 5721039 | 5854558 | 590 | 98 | 49283 | 0.68663 | 0.24616 | *C20orf196* |
| ENSG00000157191 | 1 | 16757167 | 16796573 | 120 | 21 | 46183 | 0.68658 | 0.24617 | *NECAP2* |
| ENSG00000204444 | 6 | 31610193 | 31635987 | 100 | 22 | 41559 | 0.68647 | 0.24621 | *APOM* |
| ENSG00000080573 | 19 | 10060237 | 10131147 | 423 | 69 | 47305 | 0.68639 | 0.24623 | *COL5A3* |
| ENSG00000087116 | 5 | 178527852 | 178782431 | 983 | 175 | 47225 | 0.68633 | 0.24625 | *ADAMTS2* |

| | | | | | | | | | |
|---|---|---|---|---|---|---|---|---|---|
| ENSG00000176783 | 5 | 178967559 | 179047027 | 289 | 31 | 50594 | 0.68601 | 0.24635 | *RUFY1* |
| ENSG00000187244 | 19 | 45302328 | 45334673 | 131 | 31 | 47952 | 0.68599 | 0.24636 | *BCAM* |
| ENSG00000154640 | 21 | 18955971 | 18995265 | 78 | 19 | 45741 | 0.68581 | 0.24642 | *BTG3* |
| ENSG00000130024 | 6 | 170094001 | 170134151 | 88 | 17 | 34812 | 0.68578 | 0.24643 | *PHF10* |
| ENSG00000197223 | 2 | 68258262 | 68348080 | 137 | 29 | 42747 | 0.68574 | 0.24644 | *C1D* |
| ENSG00000132026 | 19 | 12926292 | 12956242 | 79 | 12 | 41044 | 0.68536 | 0.24656 | *RTBDN* |
| ENSG00000186417 | 15 | 51623826 | 51710210 | 354 | 47 | 37096 | 0.68533 | 0.24657 | *GLDN* |
| ENSG00000166704 | 19 | 58478421 | 58524717 | 179 | 16 | 51856 | 0.68523 | 0.2466 | *ZNF606* |
| ENSG00000137103 | 9 | 35804448 | 35864844 | 107 | 18 | 43407 | 0.68499 | 0.24668 | *TMEM8B* |
| ENSG00000196177 | 10 | 124758495 | 124827827 | 271 | 37 | 45380 | 0.68484 | 0.24672 | *ACADSB* |
| ENSG00000157107 | 5 | 72241808 | 72396349 | 301 | 19 | 49849 | 0.6848 | 0.24673 | *FCHO2* |
| ENSG00000021762 | 11 | 3098346 | 3197969 | 509 | 91 | 40784 | 0.68481 | 0.24673 | *OSBPL5* |
| ENSG00000113971 | 3 | 132266986 | 132451303 | 373 | 45 | 38585 | 0.68447 | 0.24684 | *NPHP3* |
| ENSG00000256966 | 9 | 37502544 | 37602466 | 326 | 43 | 47183 | 0.68443 | 0.24685 | *RP11-613M10.8* |
| ENSG00000171148 | 3 | 9811544 | 9844695 | 78 | 13 | 49714 | 0.68422 | 0.24692 | *TADA3* |
| ENSG00000254979 | 11 | 57144838 | 57201532 | 116 | 25 | 51388 | 0.6842 | 0.24693 | *RP11-872D17.8* |
| ENSG00000133816 | 11 | 12105543 | 12295334 | 726 | 140 | 45055 | 0.68416 | 0.24694 | *MICAL2* |
| ENSG00000141994 | 19 | 5774843 | 5801249 | 121 | 18 | 33035 | 0.68414 | 0.24694 | *DUS3L* |
| ENSG00000130175 | 19 | 11536109 | 11571783 | 94 | 22 | 37425 | 0.6841 | 0.24696 | *PRKCSH* |
| ENSG00000161057 | 7 | 102974701 | 103019842 | 85 | 18 | 49820 | 0.68404 | 0.24698 | *PSMC2* |
| ENSG00000171033 | 8 | 79418374 | 79527502 | 286 | 29 | 50813 | 0.68389 | 0.24702 | *PKIA* |
| ENSG00000013306 | 17 | 42386993 | 42412238 | 55 | 12 | 30387 | 0.68378 | 0.24706 | *SLC25A39* |
| ENSG00000185792 | 19 | 56209798 | 56259768 | 218 | 52 | 48340 | 0.6837 | 0.24708 | *NLRP9* |
| ENSG00000111445 | 12 | 118441393 | 118480935 | 124 | 26 | 49160 | 0.68368 | 0.24709 | *RFC5* |
| ENSG00000171503 | 4 | 159583277 | 159640775 | 64 | 19 | 47218 | 0.68354 | 0.24713 | *ETFDH* |
| ENSG00000268085 | 8 | 23402287 | 23430651 | 156 | 24 | 45856 | 0.68352 | 0.24714 | *AC051642.1* |
| ENSG00000168612 | 20 | 44499866 | 44523905 | 74 | 10 | 44220 | 0.68347 | 0.24716 | *ZSWIM1* |
| ENSG00000166152 | 16 | 49397734 | 49443342 | 134 | 17 | 40914 | 0.68342 | 0.24717 | *C16orf78* |
| ENSG00000196900 | 5 | 125957414 | 125981979 | 99 | 24 | 37597 | 0.68335 | 0.24719 | *C5orf48* |
| ENSG00000154930 | 20 | 24976868 | 25049616 | 204 | 36 | 51518 | 0.68318 | 0.24725 | *ACSS1* |
| ENSG00000124602 | 6 | 40984772 | 41016928 | 78 | 8 | 53929 | 0.68305 | 0.24729 | *UNC5CL* |
| ENSG00000269570 | 11 | 58368589 | 58388759 | 55 | 13 | 50861 | 0.68262 | 0.24742 | *AP001350.1* |
| ENSG00000185561 | 17 | 1601064 | 1623732 | 59 | 16 | 38013 | 0.68246 | 0.24747 | *TLCD2* |
| ENSG00000184293 | 12 | 9858456 | 9895895 | 194 | 18 | 47464 | 0.6824 | 0.24749 | *CLECL1* |
| ENSG00000101417 | 20 | 32284512 | 32318125 | 75 | 16 | 47132 | 0.68231 | 0.24752 | *PXMP4* |
| ENSG00000196586 | 6 | 76448909 | 76639254 | 413 | 38 | 41415 | 0.68229 | 0.24753 | *MYO6* |

| | | | | | | | | | |
|---|---|---|---|---|---|---|---|---|---|
| ENSG00000135968 | 2 | 109055017 | 109135871 | 383 | 26 | 43170 | 0.68222 | 0.24755 | *GCC2* |
| ENSG00000115268 | 19 | 1428358 | 1450583 | 41 | 12 | 49392 | 0.68219 | 0.24756 | *RPS15* |
| ENSG00000093000 | 22 | 45549722 | 45593896 | 173 | 20 | 44574 | 0.68218 | 0.24756 | *NUP50* |
| ENSG00000229544 | 10 | 126125592 | 126148753 | 76 | 14 | 49046 | 0.68214 | 0.24758 | *NKX1-2* |
| ENSG00000219435 | 11 | 64057863 | 64082242 | 21 | 8 | 51150 | 0.68214 | 0.24758 | *TEX40* |
| ENSG00000101367 | 20 | 31397699 | 31448211 | 181 | 29 | 45219 | 0.6821 | 0.24759 | *MAPRE1* |
| ENSG00000165948 | 14 | 94537628 | 94580192 | 195 | 29 | 39808 | 0.68145 | 0.24779 | *IFI27L1* |
| ENSG00000131389 | 3 | 14434076 | 14540857 | 302 | 72 | 45792 | 0.68141 | 0.24781 | *SLC6A6* |
| ENSG00000141696 | 17 | 39948199 | 39978856 | 76 | 15 | 50779 | 0.68133 | 0.24783 | *LEPREL4* |
| ENSG00000180346 | 4 | 90023968 | 90046050 | 117 | 17 | 31172 | 0.68127 | 0.24785 | *TIGD2* |
| ENSG00000153684 | 15 | 28613767 | 28647170 | 3 | 2 | 42660 | 0.68109 | 0.24791 | *GOLGA8F* |
| ENSG00000093183 | 3 | 42579461 | 42652572 | 216 | 22 | 36255 | 0.68102 | 0.24793 | *SEC22C* |
| ENSG00000204439 | 6 | 31616075 | 31638549 | 82 | 20 | 38369 | 0.68095 | 0.24795 | *C6orf47* |
| ENSG00000143314 | 1 | 156697095 | 156721382 | 54 | 14 | 38629 | 0.6807 | 0.24803 | *MRPL24* |
| ENSG00000170906 | 19 | 54596036 | 54622564 | 106 | 23 | 48992 | 0.68051 | 0.24809 | *NDUFA3* |
| ENSG00000130177 | 13 | 114990362 | 115048198 | 176 | 19 | 29923 | 0.68047 | 0.2481 | *CDC16* |
| ENSG00000167325 | 11 | 4105937 | 4170106 | 244 | 17 | 30993 | 0.68032 | 0.24815 | *RRM1* |
| ENSG00000264278 | 18 | 74496688 | 74544251 | 215 | 34 | 48551 | 0.68022 | 0.24818 | *RP11-162A12.2* |
| ENSG00000162896 | 1 | 207091863 | 207129811 | 57 | 13 | 33557 | 0.67954 | 0.2484 | *PIGR* |
| ENSG00000172732 | 11 | 65614597 | 65645124 | 61 | 8 | 28816 | 0.67951 | 0.24841 | *MUS81* |
| ENSG00000089048 | 20 | 13684969 | 13775532 | 295 | 36 | 49230 | 0.67924 | 0.24849 | *ESF1* |
| ENSG00000204393 | 20 | 31165281 | 31206695 | 81 | 22 | 37824 | 0.67913 | 0.24853 | *RP11-410N8.4* |
| ENSG00000130522 | 19 | 18380563 | 18402432 | 106 | 13 | 52965 | 0.67892 | 0.24859 | *JUND* |
| ENSG00000174485 | 15 | 65940384 | 66094631 | 340 | 22 | 33534 | 0.67872 | 0.24866 | *DENND4A* |
| ENSG00000129028 | 15 | 71163681 | 71195124 | 44 | 12 | 52704 | 0.6787 | 0.24866 | *THAP10* |
| ENSG00000182450 | 11 | 64048774 | 64077503 | 28 | 12 | 49030 | 0.67862 | 0.24869 | *KCNK4* |
| ENSG00000167749 | 19 | 51399608 | 51423994 | 89 | 17 | 32828 | 0.67862 | 0.24869 | *KLK4* |
| ENSG00000110514 | 11 | 47280712 | 47361582 | 183 | 20 | 38695 | 0.67848 | 0.24873 | *MADD* |
| ENSG00000173960 | 2 | 24140155 | 24237779 | 382 | 30 | 55620 | 0.67832 | 0.24878 | *UBXN2A* |
| ENSG00000005469 | 7 | 86964997 | 87039111 | 205 | 23 | 35722 | 0.67784 | 0.24894 | *CROT* |
| ENSG00000157322 | 16 | 69974810 | 70008141 | 45 | 12 | 39841 | 0.67771 | 0.24898 | *CLEC18A* |
| ENSG00000186522 | 2 | 110290559 | 110381783 | 264 | 29 | 43990 | 0.67732 | 0.2491 | *Sep-10* |
| ENSG00000143067 | 1 | 120152045 | 120200396 | 148 | 22 | 49353 | 0.67708 | 0.24918 | *ZNF697* |
| ENSG00000138442 | 2 | 203729505 | 203889521 | 374 | 23 | 46519 | 0.67707 | 0.24918 | *WDR12* |
| ENSG00000116455 | 1 | 111972512 | 112001998 | 73 | 13 | 42286 | 0.67702 | 0.2492 | *WDR77* |
| ENSG00000144152 | 2 | 112885962 | 112955791 | 221 | 42 | 49328 | 0.677 | 0.2492 | *FBLN7* |

| | | | | | | | | | |
|---|---|---|---|---|---|---|---|---|---|
| ENSG00000143198 | 1 | 165590098 | 165641033 | 224 | 28 | 40511 | 0.67695 | 0.24922 | *MGST3* |
| ENSG00000170175 | 17 | 7338380 | 7371026 | 78 | 14 | 45198 | 0.67658 | 0.24934 | *CHRNB1* |
| ENSG00000110243 | 11 | 116650083 | 116673136 | 65 | 17 | 44344 | 0.67651 | 0.24936 | *APOA5* |
| ENSG00000005812 | 13 | 77556740 | 77611330 | 191 | 22 | 45358 | 0.67652 | 0.24936 | *FBXL3* |
| ENSG00000131149 | 16 | 85635015 | 85719810 | 217 | 58 | 47501 | 0.6765 | 0.24936 | *GSE1* |
| ENSG00000130517 | 19 | 18441397 | 18490760 | 150 | 32 | 46630 | 0.67645 | 0.24938 | *PGPEP1* |
| ENSG00000186453 | 2 | 24387938 | 24433718 | 179 | 20 | 43636 | 0.67642 | 0.24939 | *FAM228A* |
| ENSG00000180708 | 1 | 158379718 | 158400656 | 84 | 10 | 31104 | 0.67622 | 0.24945 | *OR10K2* |
| ENSG00000004897 | 17 | 45185069 | 45276788 | 240 | 31 | 51684 | 0.67584 | 0.24957 | *CDC27* |
| ENSG00000184428 | 8 | 144376554 | 144452149 | 285 | 27 | 37083 | 0.67576 | 0.2496 | *TOP1MT* |
| ENSG00000158669 | 8 | 41424706 | 41492520 | 241 | 22 | 50429 | 0.67572 | 0.24961 | *AGPAT6* |
| ENSG00000198924 | 10 | 115584488 | 115624142 | 47 | 23 | 45082 | 0.6756 | 0.24965 | *DCLRE1A* |
| ENSG00000054118 | 1 | 36680017 | 36780958 | 143 | 21 | 44794 | 0.67545 | 0.24969 | *THRAP3* |
| ENSG00000124588 | 6 | 2978221 | 3029996 | 378 | 50 | 49361 | 0.67545 | 0.24969 | *NQO2* |
| ENSG00000187790 | 14 | 45595143 | 45680093 | 146 | 21 | 37217 | 0.67523 | 0.24976 | *FANCM* |
| ENSG00000185269 | 17 | 79900388 | 79929716 | 93 | 10 | 41554 | 0.67522 | 0.24977 | *NOTUM* |
| ENSG00000167384 | 19 | 44969854 | 45014576 | 233 | 28 | 44507 | 0.67496 | 0.24985 | *ZNF180* |
| ENSG00000121053 | 17 | 56260098 | 56292535 | 77 | 21 | 40654 | 0.67479 | 0.24991 | *EPX* |
| ENSG00000057593 | 13 | 113750105 | 113784995 | 92 | 18 | 41594 | 0.67428 | 0.25007 | *F7* |
| ENSG00000115009 | 2 | 228668558 | 228692272 | 44 | 19 | 42364 | 0.67424 | 0.25008 | *CCL20* |
| ENSG00000011021 | 1 | 11856207 | 11913201 | 329 | 23 | 52118 | 0.67414 | 0.25011 | *CLCN6* |
| ENSG00000166532 | 12 | 8824196 | 8945691 | 160 | 28 | 47445 | 0.6738 | 0.25022 | *RIMKLB* |
| ENSG00000184012 | 21 | 42826478 | 42913043 | 395 | 58 | 46458 | 0.67368 | 0.25026 | *TMPRSS2* |
| ENSG00000148248 | 9 | 136218325 | 136252970 | 90 | 13 | 51491 | 0.67364 | 0.25027 | *SURF4* |
| ENSG00000165606 | 10 | 50562237 | 50613497 | 116 | 18 | 50277 | 0.67364 | 0.25027 | *DRGX* |
| ENSG00000124827 | 6 | 10863456 | 10892174 | 80 | 19 | 45551 | 0.67362 | 0.25028 | *GCM2* |
| ENSG00000184983 | 22 | 42471529 | 42496959 | 72 | 9 | 48271 | 0.67342 | 0.25034 | *NDUFA6* |
| ENSG00000090539 | 3 | 184087861 | 184118524 | 77 | 25 | 46691 | 0.6733 | 0.25038 | *CHRD* |
| ENSG00000173848 | 10 | 5444514 | 5510426 | 184 | 17 | 52001 | 0.6731 | 0.25044 | *NET1* |
| ENSG00000166450 | 15 | 55893744 | 56045288 | 406 | 45 | 32718 | 0.6731 | 0.25044 | *PRTG* |
| ENSG00000169733 | 17 | 79995778 | 80019707 | 78 | 17 | 38195 | 0.673 | 0.25047 | *RFNG* |
| ENSG00000179456 | 1 | 244204585 | 244230778 | 40 | 17 | 49905 | 0.67275 | 0.25055 | *ZBTB18* |
| ENSG00000151893 | 10 | 120423679 | 120524761 | 356 | 36 | 45690 | 0.67268 | 0.25058 | *CACUL1* |
| ENSG00000117262 | 1 | 145754411 | 145837103 | 11 | 4 | 43194 | 0.67229 | 0.2507 | *GPR89A* |
| ENSG00000180938 | 8 | 125975540 | 126001631 | 89 | 6 | 50747 | 0.67223 | 0.25072 | *ZNF572* |
| ENSG00000141480 | 17 | 4603784 | 4634794 | 120 | 18 | 42081 | 0.67207 | 0.25077 | *ARRB2* |

| | | | | | | | | | |
|---|---|---|---|---|---|---|---|---|---|
| ENSG00000079134 | 18 | 204520 | 278050 | 164 | 20 | 54847 | 0.67192 | 0.25082 | *THOC1* |
| ENSG00000178752 | 2 | 239057623 | 239087541 | 124 | 25 | 49163 | 0.67185 | 0.25084 | *FAM132B* |
| ENSG00000149295 | 11 | 113270318 | 113356413 | 231 | 32 | 39725 | 0.67178 | 0.25086 | *DRD2* |
| ENSG00000178026 | 22 | 24971588 | 24999175 | 75 | 15 | 42163 | 0.67176 | 0.25087 | *FAM211B* |
| ENSG00000138111 | 10 | 104211149 | 104246802 | 130 | 15 | 50556 | 0.67165 | 0.2509 | *TMEM180* |
| ENSG00000152904 | 1 | 235480665 | 235517847 | 107 | 13 | 41236 | 0.67157 | 0.25093 | *GGPS1* |
| ENSG00000136939 | 9 | 125476269 | 125497204 | 66 | 8 | 43211 | 0.67134 | 0.251 | *OR1L4* |
| ENSG00000011566 | 2 | 39466407 | 39674453 | 510 | 42 | 42827 | 0.67127 | 0.25102 | *MAP4K3* |
| ENSG00000196937 | 7 | 120978905 | 121046418 | 158 | 14 | 31250 | 0.67127 | 0.25103 | *FAM3C* |
| ENSG00000068001 | 3 | 50345221 | 50370337 | 20 | 10 | 47222 | 0.67105 | 0.2511 | *HYAL2* |
| ENSG00000177830 | 11 | 857357 | 925058 | 162 | 23 | 36761 | 0.67081 | 0.25117 | *CHID1* |
| ENSG00000112584 | 6 | 170589791 | 170726153 | 378 | 35 | 48789 | 0.67055 | 0.25125 | *FAM120B* |
| ENSG00000136518 | 3 | 179270668 | 179316196 | 77 | 8 | 51255 | 0.67039 | 0.25131 | *ACTL6A* |
| ENSG00000004777 | 19 | 36255434 | 36289724 | 109 | 17 | 44265 | 0.67032 | 0.25133 | *ARHGAP33* |
| ENSG00000196712 | 17 | 29411945 | 29719134 | 447 | 20 | 30749 | 0.67015 | 0.25138 | *NF1* |
| ENSG00000080224 | 3 | 96523425 | 97481304 | 1366 | 114 | 39880 | 0.67009 | 0.2514 | *EPHA6* |
| ENSG00000117394 | 1 | 43381052 | 43434530 | 207 | 28 | 46209 | 0.66975 | 0.25151 | *SLC2A1* |
| ENSG00000269496 | 3 | 172351483 | 172371740 | 45 | 12 | 53077 | 0.66975 | 0.25151 | *AC007919.2* |
| ENSG00000099250 | 10 | 33456420 | 33635190 | 740 | 84 | 46947 | 0.6697 | 0.25153 | *NRP1* |
| ENSG00000124232 | 20 | 43925491 | 43955803 | 109 | 20 | 43052 | 0.66939 | 0.25162 | *RBPJL* |
| ENSG00000078401 | 6 | 12280596 | 12307427 | 84 | 22 | 42951 | 0.66928 | 0.25166 | *EDN1* |
| ENSG00000168476 | 8 | 21985533 | 22009464 | 53 | 15 | 52504 | 0.66927 | 0.25166 | *REEP4* |
| ENSG00000171847 | 12 | 8363856 | 8390214 | 34 | 16 | 38209 | 0.66927 | 0.25166 | *FAM90A1* |
| ENSG00000075292 | 2 | 71493691 | 71672199 | 566 | 46 | 40781 | 0.66919 | 0.25169 | *ZNF638* |
| ENSG00000253293 | 7 | 27200210 | 27229880 | 52 | 21 | 45200 | 0.66907 | 0.25172 | *HOXA10* |
| ENSG00000249861 | 19 | 40136534 | 40161287 | 64 | 9 | 49895 | 0.66904 | 0.25173 | *LGALS16* |
| ENSG00000136848 | 9 | 124319336 | 124557809 | 590 | 115 | 40697 | 0.66903 | 0.25174 | *DAB2IP* |
| ENSG00000188732 | 7 | 23709749 | 23752868 | 148 | 23 | 44616 | 0.6688 | 0.25181 | *FAM221A* |
| ENSG00000178473 | 10 | 5396972 | 5426169 | 129 | 19 | 35840 | 0.66866 | 0.25186 | *UCN3* |
| ENSG00000101146 | 20 | 55916066 | 55964267 | 207 | 14 | 46646 | 0.66852 | 0.2519 | *RAE1* |
| ENSG00000105852 | 7 | 94979256 | 95035680 | 119 | 17 | 49782 | 0.6685 | 0.25191 | *PON3* |
| ENSG00000196796 | 16 | 29039923 | 29074047 | 14 | 6 | 39430 | 0.66849 | 0.25191 | *CTB-134H23.2* |
| ENSG00000023892 | 6 | 35255595 | 35299548 | 116 | 17 | 29084 | 0.66832 | 0.25197 | *DEF6* |
| ENSG00000178623 | 2 | 241534848 | 241580676 | 219 | 45 | 50134 | 0.66828 | 0.25198 | *GPR35* |
| ENSG00000121440 | 3 | 73421584 | 73684091 | 716 | 117 | 47178 | 0.66822 | 0.252 | *PDZRN3* |
| ENSG00000140443 | 15 | 99182200 | 99517759 | 1067 | 137 | 41726 | 0.66809 | 0.25204 | *IGF1R* |

| | | | | | | | | | |
|---|---|---|---|---|---|---|---|---|---|
| ENSG00000203963 | 1 | 67547848 | 67707536 | 558 | 58 | 47597 | 0.66803 | 0.25206 | *C1orf141* |
| ENSG00000197579 | 9 | 32530542 | 32562551 | 62 | 12 | 52106 | 0.66798 | 0.25207 | *TOPORS* |
| ENSG00000145247 | 4 | 48877036 | 48918954 | 87 | 15 | 51363 | 0.66784 | 0.25212 | *OCIAD2* |
| ENSG00000178084 | 3 | 183760835 | 183788459 | 101 | 21 | 45947 | 0.6676 | 0.25219 | *HTR3C* |
| ENSG00000160049 | 1 | 10506579 | 10542583 | 67 | 14 | 32954 | 0.66739 | 0.25226 | *DFFA* |
| ENSG00000140481 | 15 | 74499613 | 74638813 | 391 | 58 | 31501 | 0.66701 | 0.25238 | *CCDC33* |
| ENSG00000268858 | 20 | 62482879 | 62503217 | 55 | 12 | 37083 | 0.66686 | 0.25243 | *C20ORF135* |
| ENSG00000205639 | 2 | 24222951 | 24296191 | 232 | 33 | 51123 | 0.66681 | 0.25245 | *MFSD2B* |
| ENSG00000205212 | 17 | 20729760 | 20809453 | 215 | 36 | 43132 | 0.6667 | 0.25248 | *CCDC144NL* |
| ENSG00000126773 | 14 | 60548629 | 60645851 | 252 | 10 | 25411 | 0.66639 | 0.25258 | *PCNXL4* |
| ENSG00000138430 | 2 | 174927175 | 175123426 | 550 | 37 | 43725 | 0.66617 | 0.25265 | *OLA1* |
| ENSG00000121039 | 8 | 74196847 | 74247516 | 153 | 22 | 52587 | 0.66598 | 0.25271 | *RDH10* |
| ENSG00000169181 | 16 | 27788850 | 28084830 | 1036 | 151 | 48600 | 0.66562 | 0.25283 | *GSG1L* |
| ENSG00000251349 | 9 | 103194553 | 103349918 | 431 | 33 | 46268 | 0.66522 | 0.25295 | *MSANTD3-TMEFF1* |
| ENSG00000241697 | 9 | 103194560 | 103349918 | 431 | 33 | 46268 | 0.66522 | 0.25295 | *TMEFF1* |
| ENSG00000110395 | 11 | 119066752 | 119188859 | 181 | 22 | 39574 | 0.66522 | 0.25296 | *CBL* |
| ENSG00000196355 | 4 | 38618029 | 38676430 | 159 | 26 | 45133 | 0.66507 | 0.253 | *AC021860.1* |
| ENSG00000155816 | 1 | 240167648 | 240648489 | 1957 | 181 | 48272 | 0.66469 | 0.25312 | *FMN2* |
| ENSG00000181856 | 17 | 7174986 | 7201576 | 50 | 12 | 39954 | 0.6647 | 0.25312 | *SLC2A4* |
| ENSG00000104327 | 8 | 91060836 | 91117703 | 159 | 21 | 41777 | 0.66466 | 0.25314 | *CALB1* |
| ENSG00000107242 | 9 | 71310575 | 71634092 | 1021 | 90 | 40733 | 0.66465 | 0.25314 | *PIP5K1B* |
| ENSG00000214188 | 7 | 116583953 | 116748860 | 212 | 44 | 48614 | 0.6646 | 0.25315 | *ST7-OT4* |
| ENSG00000136143 | 13 | 48500622 | 48622125 | 564 | 35 | 40409 | 0.66455 | 0.25317 | *SUCLA2* |
| ENSG00000262246 | 16 | 4394543 | 4485706 | 256 | 37 | 43693 | 0.66449 | 0.25319 | *CORO7* |
| ENSG00000157150 | 3 | 12184551 | 12210851 | 80 | 16 | 45307 | 0.6641 | 0.25331 | *TIMP4* |
| ENSG00000172935 | 11 | 68761863 | 68790877 | 63 | 16 | 49108 | 0.66409 | 0.25332 | *MRGPRF* |
| ENSG00000118922 | 13 | 74250226 | 74718394 | 1515 | 174 | 44277 | 0.66405 | 0.25333 | *KLF12* |
| ENSG00000227051 | 14 | 96495661 | 96570417 | 300 | 41 | 52401 | 0.66338 | 0.25354 | *C14orf132* |
| ENSG00000244242 | 11 | 1743640 | 1781821 | 144 | 17 | 40103 | 0.66333 | 0.25356 | *IFITM10* |
| ENSG00000138834 | 16 | 1746184 | 1830318 | 211 | 32 | 39808 | 0.66304 | 0.25365 | *MAPK8IP3* |
| ENSG00000105619 | 19 | 54600320 | 54629055 | 110 | 20 | 50683 | 0.66285 | 0.25371 | *TFPT* |
| ENSG00000162526 | 1 | 32807122 | 32839913 | 20 | 13 | 52200 | 0.66247 | 0.25384 | *TSSK3* |
| ENSG00000185101 | 11 | 407933 | 452011 | 127 | 25 | 42741 | 0.66229 | 0.25389 | *ANO9* |
| ENSG00000253896 | 8 | 12601 | 39775 | 1 | 1 | 43670 | 0.66227 | 0.2539 | *AC144568.2* |
| ENSG00000204388 | 6 | 31785512 | 31808031 | 77 | 28 | 41131 | 0.66218 | 0.25393 | *HSPA1B* |
| ENSG00000184434 | 9 | 26983134 | 27015691 | 80 | 21 | 45605 | 0.66217 | 0.25393 | *LRRC19* |

| | | | | | | | | | |
|---|---|---|---|---|---|---|---|---|---|
| ENSG00000156671 | 10 | 76849344 | 76951881 | 267 | 27 | 41745 | 0.66201 | 0.25398 | *SAMD8* |
| ENSG00000166833 | 11 | 19362271 | 20153144 | 2356 | 274 | 43121 | 0.66198 | 0.25399 | *NAV2* |
| ENSG00000162746 | 1 | 161681353 | 161707933 | 128 | 22 | 40953 | 0.66172 | 0.25407 | *FCRLB* |
| ENSG00000163430 | 3 | 120101140 | 120180100 | 261 | 35 | 37806 | 0.66167 | 0.25409 | *FSTL1* |
| ENSG00000113273 | 5 | 78063032 | 78291910 | 894 | 90 | 42259 | 0.66166 | 0.25409 | *ARSB* |
| ENSG00000025423 | 12 | 57135945 | 57191574 | 117 | 19 | 37481 | 0.66128 | 0.25422 | *HSD17B6* |
| ENSG00000184206 | 15 | 84894525 | 84924120 | 56 | 17 | 43634 | 0.66128 | 0.25422 | *GOLGA6L4* |
| ENSG00000144681 | 3 | 36411836 | 36599499 | 847 | 68 | 53581 | 0.66122 | 0.25423 | *STAC* |
| ENSG00000197128 | 19 | 57968031 | 57998938 | 145 | 10 | 51027 | 0.66124 | 0.25423 | *ZNF772* |
| ENSG00000162607 | 1 | 62891968 | 62927475 | 84 | 11 | 40711 | 0.66117 | 0.25425 | *USP1* |
| ENSG00000109625 | 4 | 8584387 | 8631488 | 274 | 47 | 42035 | 0.66112 | 0.25427 | *CPZ* |
| ENSG00000243335 | 7 | 66083868 | 66286446 | 777 | 40 | 52435 | 0.66105 | 0.25429 | *KCTD7* |
| ENSG00000198758 | 1 | 110282702 | 110316649 | 90 | 20 | 46509 | 0.66083 | 0.25436 | *EPS8L3* |
| ENSG00000255223 | 11 | 56299746 | 56320757 | 116 | 13 | 39313 | 0.66072 | 0.2544 | *OR5M11* |
| ENSG00000016082 | 5 | 50668921 | 50700564 | 65 | 13 | 48860 | 0.66055 | 0.25445 | *ISL1* |
| ENSG00000268218 | 16 | 89324512 | 89349452 | 105 | 20 | 39058 | 0.6605 | 0.25447 | *AC137932.1* |
| ENSG00000135185 | 7 | 86815478 | 86859903 | 147 | 16 | 54057 | 0.66039 | 0.2545 | *TMEM243* |
| ENSG00000147601 | 8 | 73911099 | 73970357 | 131 | 15 | 36164 | 0.66023 | 0.25455 | *TERF1* |
| ENSG00000151575 | 15 | 56526207 | 56748195 | 654 | 43 | 47422 | 0.66024 | 0.25455 | *TEX9* |
| ENSG00000205593 | 22 | 50737459 | 50775489 | 160 | 21 | 42751 | 0.66022 | 0.25456 | *DENND6B* |
| ENSG00000231031 | 2 | 15820906 | 15894463 | 278 | 44 | 42216 | 0.66015 | 0.25458 | *AC008271.1* |
| ENSG00000213588 | 6 | 33412356 | 33435325 | 47 | 12 | 50318 | 0.66011 | 0.25459 | *ZBTB9* |
| ENSG00000156983 | 3 | 9763413 | 9799702 | 76 | 25 | 50365 | 0.66006 | 0.25461 | *BRPF1* |
| ENSG00000170473 | 12 | 56285197 | 56336402 | 71 | 12 | 53320 | 0.65969 | 0.25473 | *WIBG* |
| ENSG00000162924 | 2 | 61098656 | 61168745 | 128 | 26 | 41355 | 0.65948 | 0.25479 | *REL* |
| ENSG00000130844 | 19 | 54014235 | 54093523 | 497 | 56 | 47491 | 0.65948 | 0.25479 | *ZNF331* |
| ENSG00000141469 | 18 | 43294092 | 43342485 | 229 | 22 | 42564 | 0.65942 | 0.25481 | *SLC14A1* |
| ENSG00000164649 | 7 | 21930518 | 21995702 | 251 | 46 | 45413 | 0.65932 | 0.25485 | *CDCA7L* |
| ENSG00000105141 | 19 | 15150195 | 15179104 | 148 | 19 | 46061 | 0.65853 | 0.2551 | *CASP14* |
| ENSG00000107902 | 10 | 126140403 | 126316457 | 1000 | 129 | 44935 | 0.65833 | 0.25516 | *LHPP* |
| ENSG00000236287 | 11 | 10823621 | 10890343 | 192 | 29 | 44678 | 0.65816 | 0.25522 | *ZBED5* |
| ENSG00000104412 | 8 | 109445830 | 109509145 | 225 | 16 | 49726 | 0.65809 | 0.25524 | *EMC2* |
| ENSG00000085721 | 16 | 15143879 | 15198174 | 140 | 24 | 37635 | 0.65807 | 0.25525 | *RRN3* |
| ENSG00000130255 | 19 | 5664958 | 5701887 | 109 | 24 | 50117 | 0.65807 | 0.25525 | *RPL36* |
| ENSG00000117010 | 1 | 40987233 | 41023841 | 161 | 16 | 36310 | 0.658 | 0.25527 | *ZNF684* |
| ENSG00000223601 | 10 | 22487743 | 22508950 | 89 | 10 | 28094 | 0.65797 | 0.25528 | *EBLN1* |

| | | | | | | | | | |
|---|---|---|---|---|---|---|---|---|---|
| ENSG00000181778 | 9 | 71141496 | 71165783 | 46 | 12 | 39288 | 0.6579 | 0.2553 | *TMEM252* |
| ENSG00000205359 | 5 | 101697486 | 101844720 | 607 | 29 | 53407 | 0.65788 | 0.25531 | *SLCO6A1* |
| ENSG00000214194 | 7 | 112746773 | 112768668 | 57 | 9 | 36277 | 0.6575 | 0.25543 | *LINC00998* |
| ENSG00000162923 | 1 | 224562845 | 224634735 | 134 | 11 | 36201 | 0.65691 | 0.25562 | *WDR26* |
| ENSG00000172927 | 11 | 69051605 | 69192494 | 260 | 46 | 40900 | 0.65665 | 0.2557 | *MYEOV* |
| ENSG00000183292 | 2 | 131318402 | 131351734 | 2 | 1 | 43541 | 0.65649 | 0.25575 | *AC140481.2* |
| ENSG00000174827 | 1 | 145716918 | 145774074 | 22 | 11 | 43941 | 0.65643 | 0.25577 | *PDZK1* |
| ENSG00000100532 | 14 | 54966530 | 55015567 | 59 | 12 | 40024 | 0.65584 | 0.25596 | *CGRRF1* |
| ENSG00000118894 | 16 | 5124305 | 5157809 | 149 | 33 | 48985 | 0.65586 | 0.25596 | *FAM86A* |
| ENSG00000259529 | 14 | 24610427 | 24646611 | 68 | 19 | 47966 | 0.65578 | 0.25598 | *RP11-468E2.4* |
| ENSG00000149452 | 11 | 62746626 | 62793311 | 96 | 28 | 48090 | 0.6556 | 0.25604 | *SLC22A8* |
| ENSG00000142233 | 19 | 49154664 | 49186338 | 61 | 15 | 42758 | 0.65549 | 0.25608 | *NTN5* |
| ENSG00000116299 | 1 | 109646301 | 109759401 | 422 | 59 | 39834 | 0.65516 | 0.25618 | *KIAA1324* |
| ENSG00000180185 | 16 | 1866968 | 1900208 | 314 | 11 | 53790 | 0.65489 | 0.25627 | *FAHD1* |
| ENSG00000137343 | 6 | 30584619 | 30624600 | 91 | 17 | 31044 | 0.65486 | 0.25628 | *ATAT1* |
| ENSG00000127074 | 1 | 192595275 | 192639390 | 118 | 18 | 35975 | 0.6548 | 0.2563 | *RGS13* |
| ENSG00000166105 | 11 | 134134139 | 134199458 | 188 | 35 | 45504 | 0.65473 | 0.25632 | *GLB1L3* |
| ENSG00000135124 | 12 | 121637660 | 121681909 | 185 | 23 | 54087 | 0.65466 | 0.25634 | *P2RX4* |
| ENSG00000142698 | 1 | 34622484 | 34694732 | 231 | 40 | 43357 | 0.65465 | 0.25635 | *C1orf94* |
| ENSG00000178395 | 1 | 223556715 | 223578812 | 53 | 7 | 50069 | 0.6546 | 0.25636 | *C1orf65* |
| ENSG00000136478 | 17 | 62214587 | 62350661 | 379 | 32 | 41869 | 0.6546 | 0.25636 | *TEX2* |
| ENSG00000103710 | 15 | 65335679 | 65379028 | 98 | 17 | 54086 | 0.65447 | 0.2564 | *RASL12* |
| ENSG00000070761 | 16 | 58137496 | 58173354 | 138 | 13 | 45715 | 0.65432 | 0.25645 | *C16orf80* |
| ENSG00000123119 | 8 | 91793778 | 91981636 | 402 | 28 | 30341 | 0.65423 | 0.25648 | *NECAB1* |
| ENSG00000226397 | 12 | 25136358 | 25160453 | 94 | 24 | 51511 | 0.65412 | 0.25652 | *C12orf77* |
| ENSG00000131061 | 20 | 32309463 | 32390075 | 121 | 23 | 38364 | 0.65413 | 0.25652 | *ZNF341* |
| ENSG00000147588 | 8 | 82342561 | 82369758 | 88 | 10 | 32673 | 0.65399 | 0.25656 | *PMP2* |
| ENSG00000205758 | 21 | 34951647 | 35026232 | 116 | 28 | 53637 | 0.65398 | 0.25656 | *CRYZL1* |
| ENSG00000123607 | 2 | 166703985 | 166820353 | 283 | 33 | 49038 | 0.65391 | 0.25658 | *TTC21B* |
| ENSG00000183826 | 6 | 38126227 | 38617924 | 1290 | 129 | 44392 | 0.65389 | 0.25659 | *BTBD9* |
| ENSG00000109787 | 4 | 38655817 | 38712663 | 181 | 39 | 45188 | 0.65378 | 0.25663 | *KLF3* |
| ENSG00000008516 | 16 | 3086682 | 3120727 | 127 | 23 | 52159 | 0.65372 | 0.25665 | *MMP25* |
| ENSG00000268192 | 11 | 119017590 | 119037930 | 37 | 7 | 28526 | 0.65347 | 0.25673 | *AP002956.1* |
| ENSG00000117448 | 1 | 46006215 | 46045721 | 97 | 10 | 53588 | 0.6533 | 0.25678 | *AKR1A1* |
| ENSG00000140718 | 16 | 53727875 | 54165853 | 1431 | 176 | 42474 | 0.65317 | 0.25682 | *FTO* |
| ENSG00000128422 | 17 | 39765689 | 39791094 | 42 | 14 | 44733 | 0.65319 | 0.25682 | *KRT17* |

| | | | | | | | | | |
|---|---|---|---|---|---|---|---|---|---|
| ENSG00000182606 | 3 | 42045294 | 42277381 | 719 | 76 | 49298 | 0.65308 | 0.25685 | *TRAK1* |
| ENSG00000172803 | 11 | 65591112 | 65634367 | 94 | 15 | 33877 | 0.652 | 0.2572 | *SNX32* |
| ENSG00000101441 | 20 | 23656277 | 23679677 | 92 | 21 | 45479 | 0.6519 | 0.25723 | *CST4* |
| ENSG00000204954 | 12 | 104333980 | 104369486 | 191 | 25 | 40437 | 0.65179 | 0.25727 | *C12orf73* |
| ENSG00000128487 | 17 | 19902657 | 20232339 | 842 | 42 | 41348 | 0.65172 | 0.25729 | *SPECC1* |
| ENSG00000128159 | 22 | 50646118 | 50693421 | 207 | 31 | 39357 | 0.65139 | 0.2574 | *TUBGCP6* |
| ENSG00000167114 | 9 | 131092925 | 131133749 | 87 | 18 | 37413 | 0.65125 | 0.25744 | *SLC27A4* |
| ENSG00000178201 | 19 | 57956542 | 57977854 | 111 | 11 | 51878 | 0.65125 | 0.25744 | *VN1R1* |
| ENSG00000124449 | 19 | 44210162 | 44234173 | 84 | 15 | 44559 | 0.65124 | 0.25745 | *IRGC* |
| ENSG00000080644 | 15 | 78875394 | 78923637 | 153 | 18 | 44642 | 0.65117 | 0.25747 | *CHRNA3* |
| ENSG00000139973 | 14 | 62443803 | 62578431 | 604 | 83 | 45060 | 0.65108 | 0.2575 | *SYT16* |
| ENSG00000269753 | 1 | 2248581 | 2269042 | 99 | 26 | 44449 | 0.65097 | 0.25753 | *AL589739.1* |
| ENSG00000241106 | 6 | 32770540 | 32794825 | 303 | 27 | 38782 | 0.65071 | 0.25762 | *HLA-DOB* |
| ENSG00000196353 | 3 | 131242399 | 132014254 | 3355 | 199 | 47846 | 0.65068 | 0.25763 | *CPNE4* |
| ENSG00000087494 | 12 | 28101017 | 28135638 | 122 | 21 | 50047 | 0.65055 | 0.25767 | *PTHLH* |
| ENSG00000215853 | 1 | 152116071 | 152141704 | 34 | 9 | 30419 | 0.65051 | 0.25768 | *RPTN* |
| ENSG00000102794 | 13 | 77512632 | 77542777 | 82 | 24 | 44056 | 0.65041 | 0.25771 | *IRG1* |
| ENSG00000249624 | 21 | 34609079 | 34665517 | 184 | 26 | 49596 | 0.65032 | 0.25774 | *AP000295.9* |
| ENSG00000204978 | 19 | 41939063 | 41960670 | 47 | 10 | 42536 | 0.65023 | 0.25777 | *C19orf69* |
| ENSG00000115598 | 2 | 102793433 | 102866462 | 313 | 24 | 47410 | 0.64937 | 0.25805 | *IL1RL2* |
| ENSG00000163138 | 4 | 20687905 | 20764530 | 318 | 28 | 55082 | 0.64929 | 0.25808 | *PACRGL* |
| ENSG00000117360 | 1 | 150283925 | 150335671 | 154 | 17 | 37850 | 0.64909 | 0.25814 | *PRPF3* |
| ENSG00000108590 | 17 | 6536635 | 6564954 | 127 | 16 | 43727 | 0.64872 | 0.25826 | *MED31* |
| ENSG00000158856 | 8 | 21896506 | 21950038 | 216 | 35 | 44220 | 0.64867 | 0.25828 | *DMTN* |
| ENSG00000197416 | 8 | 82427216 | 82453613 | 74 | 15 | 34876 | 0.64859 | 0.2583 | *FABP12* |
| ENSG00000196576 | 22 | 50703408 | 50756056 | 224 | 47 | 45526 | 0.64831 | 0.25839 | *PLXNB2* |
| ENSG00000121671 | 11 | 45858669 | 45914798 | 128 | 23 | 46762 | 0.64821 | 0.25842 | *CRY2* |
| ENSG00000143248 | 1 | 163070911 | 163301577 | 999 | 96 | 50835 | 0.6482 | 0.25843 | *RGS5* |
| ENSG00000168874 | 2 | 85968467 | 86025189 | 149 | 29 | 39795 | 0.6477 | 0.25859 | *ATOH8* |
| ENSG00000255582 | 14 | 22091992 | 22113096 | 114 | 21 | 50261 | 0.6476 | 0.25862 | *OR10G2* |
| ENSG00000087086 | 19 | 49458558 | 49480135 | 54 | 13 | 49459 | 0.64745 | 0.25867 | *FTL* |
| ENSG00000150907 | 13 | 41119804 | 41250734 | 275 | 22 | 36235 | 0.64722 | 0.25875 | *FOXO1* |
| ENSG00000175040 | 3 | 142828173 | 142851800 | 59 | 9 | 46291 | 0.64715 | 0.25877 | *CHST2* |
| ENSG00000102908 | 16 | 69588997 | 69748569 | 398 | 32 | 46469 | 0.64711 | 0.25878 | *NFAT5* |
| ENSG00000197050 | 19 | 37488759 | 37631216 | 483 | 24 | 51364 | 0.64683 | 0.25887 | *ZNF420* |
| ENSG00000180424 | 20 | 30018322 | 30048060 | 87 | 11 | 42225 | 0.64664 | 0.25893 | *DEFB123* |

| | | | | | | | | | |
|---|---|---|---|---|---|---|---|---|---|
| ENSG00000179085 | 1 | 155102367 | 155123071 | 36 | 10 | 48074 | 0.64656 | 0.25896 | *DPM3* |
| ENSG00000243660 | 10 | 43922282 | 44001517 | 315 | 28 | 35870 | 0.64641 | 0.25901 | *ZNF487* |
| ENSG00000014216 | 11 | 64938037 | 64989477 | 122 | 19 | 40930 | 0.64631 | 0.25904 | *CAPN1* |
| ENSG00000137726 | 11 | 117697693 | 117758201 | 168 | 41 | 47721 | 0.64623 | 0.25906 | *FXYD6* |
| ENSG00000159640 | 17 | 61544422 | 61609205 | 153 | 24 | 47821 | 0.646 | 0.25914 | *ACE* |
| ENSG00000164764 | 8 | 73966775 | 74046323 | 250 | 30 | 43631 | 0.6458 | 0.2592 | *SBSPON* |
| ENSG00000184608 | 8 | 11215911 | 11306167 | 456 | 48 | 36920 | 0.64578 | 0.25921 | *C8orf12* |
| ENSG00000125629 | 2 | 118836028 | 118878573 | 143 | 33 | 50868 | 0.64571 | 0.25923 | *INSIG2* |
| ENSG00000182346 | 13 | 106108216 | 106153383 | 176 | 17 | 49781 | 0.64562 | 0.25926 | *DAOA* |
| ENSG00000133193 | 17 | 71193492 | 71242892 | 234 | 11 | 50978 | 0.64541 | 0.25933 | *FAM104A* |
| ENSG00000119685 | 14 | 76089968 | 76431421 | 723 | 61 | 47058 | 0.64527 | 0.25938 | *TTLL5* |
| ENSG00000064547 | 19 | 19724477 | 19749739 | 39 | 10 | 49327 | 0.6452 | 0.2594 | *LPAR2* |
| ENSG00000197587 | 1 | 46962668 | 46989898 | 82 | 20 | 40859 | 0.64508 | 0.25944 | *DMBX1* |
| ENSG00000139971 | 14 | 58456453 | 58774857 | 884 | 99 | 48081 | 0.64506 | 0.25945 | *C14orf37* |
| ENSG00000138639 | 4 | 86386267 | 86933823 | 1660 | 120 | 36880 | 0.64486 | 0.25951 | *ARHGAP24* |
| ENSG00000198010 | 8 | 1439532 | 1666642 | 1073 | 87 | 48469 | 0.64484 | 0.25951 | *DLGAP2* |
| ENSG00000131067 | 20 | 33422523 | 33470663 | 93 | 15 | 49059 | 0.64467 | 0.25957 | *GGT7* |
| ENSG00000130958 | 9 | 99072988 | 99155992 | 332 | 28 | 48798 | 0.64455 | 0.25961 | *SLC35D2* |
| ENSG00000109576 | 4 | 170971373 | 171022850 | 152 | 26 | 45890 | 0.64422 | 0.25972 | *AADAT* |
| ENSG00000176222 | 19 | 44366515 | 44415537 | 110 | 10 | 51133 | 0.64402 | 0.25978 | *ZNF404* |
| ENSG00000135643 | 12 | 70750056 | 70838072 | 354 | 52 | 45558 | 0.64391 | 0.25982 | *KCNMB4* |
| ENSG00000135116 | 12 | 117283949 | 117329246 | 108 | 21 | 41044 | 0.64357 | 0.25993 | *HRK* |
| ENSG00000133321 | 11 | 63294281 | 63323934 | 48 | 16 | 42541 | 0.64334 | 0.26 | *RARRES3* |
| ENSG00000241128 | 1 | 247876401 | 247897345 | 53 | 9 | 44967 | 0.64311 | 0.26008 | *OR14A2* |
| ENSG00000135916 | 2 | 231719354 | 231753963 | 159 | 40 | 44831 | 0.643 | 0.26011 | *ITM2C* |
| ENSG00000146409 | 6 | 133080509 | 133129701 | 207 | 23 | 40237 | 0.64265 | 0.26023 | *SLC18B1* |
| ENSG00000164241 | 5 | 126368250 | 126419184 | 138 | 13 | 28511 | 0.64229 | 0.26034 | *C5orf63* |
| ENSG00000131096 | 17 | 42020106 | 42091837 | 167 | 32 | 44080 | 0.6423 | 0.26034 | *PYY* |
| ENSG00000185958 | 12 | 50710013 | 50800405 | 123 | 10 | 31211 | 0.64223 | 0.26036 | *FAM186A* |
| ENSG00000160973 | 8 | 145688795 | 145711718 | 35 | 9 | 43355 | 0.64221 | 0.26037 | *FOXH1* |
| ENSG00000064995 | 6 | 34835555 | 34865866 | 86 | 13 | 30692 | 0.64215 | 0.26039 | *TAF11* |
| ENSG00000145782 | 5 | 115153893 | 115187555 | 66 | 18 | 46985 | 0.642 | 0.26044 | *ATG12* |
| ENSG00000197658 | 11 | 62837412 | 62921693 | 452 | 26 | 39374 | 0.64171 | 0.26053 | *SLC22A24* |
| ENSG00000104419 | 8 | 134239414 | 134324265 | 316 | 59 | 47810 | 0.64169 | 0.26054 | *NDRG1* |
| ENSG00000133243 | 19 | 1975447 | 2044880 | 287 | 53 | 45097 | 0.64157 | 0.26057 | *BTBD2* |
| ENSG00000072195 | 2 | 220289568 | 220373009 | 162 | 30 | 42688 | 0.64148 | 0.2606 | *SPEG* |

| | | | | | | | | | |
|---|---|---|---|---|---|---|---|---|---|
| ENSG00000269832 | 14 | 50281993 | 50303258 | 17 | 7 | 54742 | 0.64127 | 0.26067 | *AL627171.2* |
| ENSG00000085415 | 18 | 12937132 | 12997535 | 185 | 25 | 35624 | 0.64091 | 0.26079 | *SEH1L* |
| ENSG00000122859 | 10 | 71321454 | 71342994 | 47 | 18 | 45319 | 0.64078 | 0.26083 | *NEUROG3* |
| ENSG00000136144 | 13 | 50096082 | 50169719 | 420 | 30 | 29008 | 0.64077 | 0.26084 | *RCBTB1* |
| ENSG00000164251 | 5 | 76104758 | 76141140 | 98 | 16 | 37158 | 0.64074 | 0.26085 | *F2RL1* |
| ENSG00000113621 | 5 | 134199493 | 134247215 | 72 | 19 | 42121 | 0.64066 | 0.26087 | *TXNDC15* |
| ENSG00000113552 | 5 | 141361314 | 141402606 | 171 | 18 | 45767 | 0.64043 | 0.26095 | *GNPDA1* |
| ENSG00000122705 | 9 | 36180853 | 36314778 | 436 | 46 | 45036 | 0.64039 | 0.26096 | *CLTA* |
| ENSG00000014914 | 1 | 149890543 | 149918791 | 48 | 7 | 51992 | 0.64019 | 0.26102 | *MTMR11* |
| ENSG00000179454 | 14 | 45383522 | 45521525 | 306 | 22 | 34571 | 0.64013 | 0.26104 | *KLHL28* |
| ENSG00000105656 | 19 | 18543473 | 18642937 | 250 | 18 | 33696 | 0.64013 | 0.26104 | *ELL* |
| ENSG00000085563 | 7 | 87123175 | 87352611 | 525 | 64 | 47304 | 0.64011 | 0.26105 | *ABCB1* |
| ENSG00000100632 | 14 | 69836848 | 69875344 | 115 | 19 | 55084 | 0.63996 | 0.2611 | *ERH* |
| ENSG00000188729 | 3 | 190907030 | 190993404 | 342 | 43 | 45074 | 0.63988 | 0.26113 | *OSTN* |
| ENSG00000160075 | 1 | 1467053 | 1520249 | 177 | 23 | 38744 | 0.63896 | 0.26142 | *SSU72* |
| ENSG00000187193 | 16 | 56706336 | 56728108 | 98 | 22 | 42269 | 0.63872 | 0.2615 | *MT1X* |
| ENSG00000143368 | 1 | 149885209 | 149910236 | 46 | 6 | 50806 | 0.63825 | 0.26166 | *SF3B4* |
| ENSG00000185129 | 5 | 139477362 | 139506321 | 23 | 7 | 53075 | 0.63808 | 0.26171 | *PURA* |
| ENSG00000014164 | 8 | 144509825 | 144633623 | 452 | 51 | 52170 | 0.63809 | 0.26171 | *ZC3H3* |
| ENSG00000215066 | 9 | 6457618 | 6480375 | 68 | 11 | 41566 | 0.63807 | 0.26171 | *C9orf38* |
| ENSG00000151704 | 11 | 128696210 | 128747268 | 173 | 33 | 48643 | 0.63808 | 0.26171 | *KCNJ1* |
| ENSG00000180176 | 11 | 2175159 | 2203107 | 88 | 16 | 42100 | 0.63793 | 0.26176 | *TH* |
| ENSG00000214106 | 7 | 154710208 | 154751618 | 81 | 10 | 55766 | 0.63785 | 0.26179 | *PAXIP1-AS2* |
| ENSG00000131095 | 17 | 42972376 | 43004305 | 122 | 22 | 52683 | 0.63783 | 0.26179 | *GFAP* |
| ENSG00000175564 | 11 | 73701326 | 73730480 | 61 | 10 | 45288 | 0.63781 | 0.2618 | *UCP3* |
| ENSG00000176893 | 11 | 4925900 | 4946922 | 94 | 20 | 47236 | 0.63779 | 0.26181 | *OR51G2* |
| ENSG00000168070 | 11 | 64694989 | 64749557 | 152 | 15 | 43252 | 0.63776 | 0.26181 | *C11orf85* |
| ENSG00000173614 | 1 | 9993486 | 10055559 | 132 | 22 | 36762 | 0.63753 | 0.26189 | *NMNAT1* |
| ENSG00000173838 | 17 | 60768675 | 60895705 | 351 | 41 | 44056 | 0.63724 | 0.26198 | *Mar-10* |
| ENSG00000164659 | 7 | 86496222 | 86699015 | 538 | 28 | 51413 | 0.63708 | 0.26204 | *KIAA1324L* |
| ENSG00000146013 | 5 | 137578068 | 137620360 | 102 | 17 | 51266 | 0.63704 | 0.26205 | *GFRA3* |
| ENSG00000260836 | 15 | 83198783 | 83234682 | 48 | 12 | 44298 | 0.63658 | 0.2622 | *RP11-152F13.10* |
| ENSG00000267885 | 1 | 33429268 | 33449642 | 46 | 8 | 40096 | 0.63619 | 0.26233 | *FKSG48* |
| ENSG00000143374 | 1 | 150449887 | 150490078 | 132 | 15 | 35675 | 0.63616 | 0.26234 | *TARS2* |
| ENSG00000174946 | 3 | 150905619 | 150930988 | 81 | 16 | 33169 | 0.63611 | 0.26235 | *GPR171* |
| ENSG00000167550 | 12 | 49448468 | 49473808 | 24 | 6 | 47407 | 0.636 | 0.26239 | *RHEBL1* |

| | | | | | | | | | |
|---|---|---|---|---|---|---|---|---|---|
| ENSG00000196268 | 19 | 21569921 | 21620375 | 192 | 18 | 44621 | 0.63566 | 0.2625 | *ZNF493* |
| ENSG00000168703 | 20 | 43742066 | 43763106 | 77 | 11 | 46980 | 0.63567 | 0.2625 | *WFDC12* |
| ENSG00000176919 | 9 | 139829698 | 139851426 | 93 | 15 | 47884 | 0.63552 | 0.26254 | *C8G* |
| ENSG00000183072 | 5 | 172649112 | 172672360 | 55 | 16 | 37463 | 0.63552 | 0.26255 | *NKX2-5* |
| ENSG00000067221 | 15 | 74265547 | 74296963 | 89 | 12 | 41719 | 0.63546 | 0.26256 | *STOML1* |
| ENSG00000266956 | 17 | 34300692 | 34327306 | 112 | 23 | 44995 | 0.63534 | 0.2626 | *CTB-186H2.3* |
| ENSG00000100246 | 22 | 39164513 | 39200203 | 80 | 19 | 40182 | 0.63513 | 0.26267 | *DNAL4* |
| ENSG00000204406 | 2 | 148768580 | 149285805 | 1317 | 93 | 49417 | 0.63497 | 0.26272 | *MBD5* |
| ENSG00000140950 | 16 | 84501681 | 84597639 | 692 | 95 | 36583 | 0.63475 | 0.2628 | *TLDC1* |
| ENSG00000132170 | 3 | 12318867 | 12485855 | 429 | 46 | 44168 | 0.63468 | 0.26282 | *PPARG* |
| ENSG00000103426 | 16 | 4380252 | 4480495 | 280 | 43 | 43026 | 0.63458 | 0.26285 | *CORO7-PAM16* |
| ENSG00000132205 | 18 | 2837028 | 2925991 | 231 | 72 | 39295 | 0.63459 | 0.26285 | *EMILIN2* |
| ENSG00000101443 | 20 | 44088346 | 44120172 | 123 | 21 | 34289 | 0.63443 | 0.2629 | *WFDC2* |
| ENSG00000087237 | 16 | 56985762 | 57027757 | 173 | 38 | 50227 | 0.63432 | 0.26293 | *CETP* |
| ENSG00000163072 | 2 | 169633049 | 169732024 | 307 | 48 | 45727 | 0.6342 | 0.26297 | *NOSTRIN* |
| ENSG00000127125 | 1 | 42911788 | 42949056 | 107 | 10 | 42205 | 0.63407 | 0.26302 | *PPCS* |
| ENSG00000113504 | 5 | 1040499 | 1122150 | 354 | 68 | 46578 | 0.63407 | 0.26302 | *SLC12A7* |
| ENSG00000151233 | 12 | 42465647 | 42548681 | 236 | 28 | 54073 | 0.63397 | 0.26305 | *GXYLT1* |
| ENSG00000114859 | 3 | 184053973 | 184089439 | 94 | 23 | 47985 | 0.63373 | 0.26313 | *CLCN2* |
| ENSG00000110046 | 11 | 64652007 | 64694722 | 87 | 13 | 39345 | 0.63372 | 0.26313 | *ATG2A* |
| ENSG00000147854 | 9 | 6403151 | 6517054 | 332 | 31 | 41056 | 0.63353 | 0.26319 | *UHRF2* |
| ENSG00000116016 | 2 | 46510806 | 46623836 | 378 | 67 | 39977 | 0.63345 | 0.26322 | *EPAS1* |
| ENSG00000039523 | 16 | 67542321 | 67590691 | 72 | 19 | 45147 | 0.63333 | 0.26326 | *FAM65A* |
| ENSG00000126216 | 13 | 113129325 | 113252481 | 353 | 27 | 47553 | 0.63326 | 0.26328 | *TUBGCP3* |
| ENSG00000169840 | 13 | 28356780 | 28378905 | 74 | 18 | 48137 | 0.63307 | 0.26334 | *GSX1* |
| ENSG00000181577 | 6 | 43958317 | 43983695 | 148 | 21 | 29326 | 0.63283 | 0.26342 | *C6orf223* |
| ENSG00000215041 | 17 | 7208947 | 7242712 | 84 | 15 | 48067 | 0.6328 | 0.26343 | *NEURL4* |
| ENSG00000116459 | 1 | 111981486 | 112015395 | 83 | 17 | 47168 | 0.63278 | 0.26344 | *ATP5F1* |
| ENSG00000178917 | 3 | 44530462 | 44562128 | 39 | 10 | 39280 | 0.6326 | 0.2635 | *ZNF852* |
| ENSG00000175485 | 11 | 6210376 | 6231489 | 57 | 14 | 29744 | 0.63259 | 0.2635 | *OR52W1* |
| ENSG00000158019 | 2 | 28102808 | 28571768 | 1085 | 81 | 40362 | 0.63244 | 0.26355 | *BRE* |
| ENSG00000010361 | 19 | 50300126 | 50330633 | 98 | 17 | 43191 | 0.63235 | 0.26358 | *FUZ* |
| ENSG00000198663 | 6 | 36829646 | 36906740 | 271 | 26 | 50875 | 0.63233 | 0.26359 | *C6orf89* |
| ENSG00000177096 | 22 | 42460255 | 42485445 | 56 | 9 | 49845 | 0.63228 | 0.2636 | *FAM109B* |
| ENSG00000198754 | 1 | 40225195 | 40247020 | 71 | 17 | 48566 | 0.63211 | 0.26366 | *OXCT2* |
| ENSG00000163191 | 1 | 151994982 | 152030383 | 58 | 13 | 35324 | 0.63202 | 0.26369 | *S100A11* |

| | | | | | | | | | |
|---|---|---|---|---|---|---|---|---|---|
| ENSG00000183715 | 11 | 132274871 | 133412414 | 3939 | 386 | 42379 | 0.63197 | 0.2637 | *OPCML* |
| ENSG00000249115 | 19 | 36093646 | 36126251 | 69 | 15 | 44808 | 0.63182 | 0.26375 | *HAUS5* |
| ENSG00000197380 | 19 | 47140869 | 47174395 | 85 | 27 | 44621 | 0.63149 | 0.26386 | *DACT3* |
| ENSG00000172789 | 12 | 54416637 | 54439142 | 51 | 9 | 53703 | 0.63128 | 0.26393 | *HOXC5* |
| ENSG00000157093 | 3 | 42428570 | 42462092 | 66 | 19 | 45390 | 0.63124 | 0.26394 | *LYZL4* |
| ENSG00000121316 | 12 | 14646595 | 14731283 | 210 | 36 | 42357 | 0.6311 | 0.26399 | *PLBD1* |
| ENSG00000122025 | 13 | 28567411 | 28684729 | 438 | 53 | 42272 | 0.63102 | 0.26401 | *FLT3* |
| ENSG00000228259 | 5 | 179068298 | 179089445 | 67 | 20 | 45150 | 0.63093 | 0.26404 | *AC136604.1* |
| ENSG00000205133 | 8 | 93885758 | 94039901 | 353 | 47 | 39282 | 0.63087 | 0.26406 | *TRIQK* |
| ENSG00000254553 | 1 | 32920658 | 33076393 | 375 | 23 | 53361 | 0.63076 | 0.2641 | *RP1-27O5.3* |
| ENSG00000258804 | 14 | 87362122 | 87399099 | 131 | 13 | 48566 | 0.63047 | 0.26419 | *RP11-322L20.1* |
| ENSG00000148572 | 10 | 64883050 | 64924783 | 113 | 16 | 32167 | 0.63018 | 0.26429 | *NRBF2* |
| ENSG00000134375 | 1 | 201914619 | 201949789 | 125 | 23 | 52727 | 0.63015 | 0.2643 | *TIMM17A* |
| ENSG00000125975 | 20 | 34101014 | 34127481 | 63 | 12 | 38286 | 0.62999 | 0.26435 | *C20orf173* |
| ENSG00000118263 | 2 | 207928861 | 208041991 | 345 | 60 | 48151 | 0.62997 | 0.26436 | *KLF7* |
| ENSG00000176393 | 1 | 201941500 | 201985275 | 222 | 27 | 48442 | 0.62952 | 0.26451 | *RNPEP* |
| ENSG00000204960 | 7 | 155139680 | 155170629 | 212 | 28 | 40073 | 0.62926 | 0.26459 | *BLACE* |
| ENSG00000129315 | 12 | 49072247 | 49120681 | 126 | 17 | 46562 | 0.62912 | 0.26464 | *CCNT1* |
| ENSG00000138439 | 2 | 203489901 | 203644480 | 249 | 36 | 45324 | 0.6289 | 0.26471 | *FAM117B* |
| ENSG00000150938 | 2 | 36573069 | 36788278 | 612 | 91 | 44617 | 0.62865 | 0.26479 | *CRIM1* |
| ENSG00000204642 | 6 | 29680552 | 29716305 | 389 | 29 | 43787 | 0.62833 | 0.26489 | *HLA-F* |
| ENSG00000006194 | 16 | 3303743 | 3361401 | 106 | 21 | 41043 | 0.62822 | 0.26493 | *ZNF263* |
| ENSG00000007306 | 19 | 42167235 | 42220895 | 177 | 22 | 42951 | 0.62807 | 0.26498 | *CEACAM7* |
| ENSG00000125968 | 20 | 30183086 | 30204318 | 69 | 4 | 24315 | 0.62777 | 0.26508 | *ID1* |
| ENSG00000113300 | 5 | 179911412 | 180015405 | 509 | 34 | 48843 | 0.62769 | 0.2651 | *CNOT6* |
| ENSG00000180537 | 6 | 13914677 | 13990533 | 178 | 20 | 39157 | 0.62765 | 0.26512 | *RNF182* |
| ENSG00000105063 | 19 | 55731148 | 55780363 | 147 | 29 | 53089 | 0.62755 | 0.26515 | *PPP6R1* |
| ENSG00000182909 | 19 | 54962981 | 54984894 | 81 | 16 | 43461 | 0.62747 | 0.26517 | *LENG9* |
| ENSG00000197885 | 3 | 23923151 | 23998082 | 280 | 25 | 45671 | 0.62746 | 0.26518 | *NKIRAS1* |
| ENSG00000170545 | 12 | 51628342 | 51674674 | 122 | 25 | 39567 | 0.6274 | 0.2652 | *SMAGP* |
| ENSG00000196497 | 14 | 24639425 | 24668170 | 70 | 16 | 41295 | 0.62737 | 0.26521 | *IPO4* |
| ENSG00000021826 | 2 | 211332406 | 211553831 | 632 | 70 | 48984 | 0.62704 | 0.26532 | *CPS1* |
| ENSG00000109208 | 4 | 71216478 | 71242875 | 149 | 20 | 40097 | 0.62691 | 0.26536 | *SMR3A* |
| ENSG00000127452 | 19 | 9910943 | 9948492 | 74 | 17 | 34458 | 0.62687 | 0.26537 | *FBXL12* |
| ENSG00000132819 | 20 | 55956463 | 55994389 | 119 | 28 | 44186 | 0.62684 | 0.26538 | *RBM38* |
| ENSG00000153071 | 5 | 39361780 | 39472402 | 387 | 58 | 52631 | 0.62672 | 0.26542 | *DAB2* |

| | | | | | | | | | |
|---|---|---|---|---|---|---|---|---|---|
| ENSG00000131626 | 11 | 70106806 | 70240509 | 324 | 16 | 37488 | 0.62648 | 0.2655 | *PPFIA1* |
| ENSG00000173230 | 3 | 121372046 | 121478602 | 240 | 25 | 53503 | 0.62637 | 0.26554 | *GOLGB1* |
| ENSG00000078674 | 8 | 17770349 | 17895478 | 617 | 46 | 33218 | 0.62632 | 0.26555 | *PCM1* |
| ENSG00000203666 | 1 | 245123007 | 245300466 | 530 | 52 | 47412 | 0.62624 | 0.26558 | *EFCAB2* |
| ENSG00000054796 | 20 | 55894815 | 55929050 | 114 | 9 | 47047 | 0.62621 | 0.26559 | *SPO11* |
| ENSG00000154035 | 17 | 21132183 | 21166722 | 106 | 28 | 46249 | 0.626 | 0.26566 | *C17orf103* |
| ENSG00000257184 | 7 | 27193154 | 27229632 | 70 | 26 | 42573 | 0.62544 | 0.26584 | *RP1-170O19.20* |
| ENSG00000204710 | 11 | 64927625 | 64950715 | 42 | 15 | 44987 | 0.62487 | 0.26603 | *SPDYC* |
| ENSG00000260170 | 15 | 45869549 | 45978512 | 342 | 48 | 45131 | 0.62487 | 0.26603 | *RP11-96O20.4* |
| ENSG00000173950 | 3 | 194779008 | 195001896 | 895 | 93 | 40047 | 0.62476 | 0.26606 | *XXYLT1* |
| ENSG00000128594 | 7 | 127657124 | 127682160 | 47 | 12 | 44306 | 0.62435 | 0.2662 | *LRRC4* |
| ENSG00000257727 | 12 | 56693626 | 56720120 | 45 | 8 | 44672 | 0.62417 | 0.26626 | *CNPY2* |
| ENSG00000181322 | 3 | 137970279 | 138058728 | 92 | 30 | 50953 | 0.62386 | 0.26636 | *NME9* |
| ENSG00000153048 | 16 | 8936799 | 8972866 | 239 | 41 | 50641 | 0.62378 | 0.26639 | *CARHSP1* |
| ENSG00000162814 | 1 | 217794666 | 218055038 | 872 | 74 | 46321 | 0.62344 | 0.2665 | *SPATA17* |
| ENSG00000183347 | 1 | 89819617 | 89862020 | 141 | 21 | 50664 | 0.62341 | 0.26651 | *GBP6* |
| ENSG00000126790 | 14 | 59917081 | 59961148 | 158 | 18 | 43877 | 0.62317 | 0.26659 | *L3HYPDH* |
| ENSG00000129317 | 12 | 44112410 | 44162620 | 154 | 18 | 41072 | 0.62284 | 0.26669 | *PUS7L* |
| ENSG00000259522 | 14 | 24639601 | 24669622 | 73 | 17 | 41337 | 0.62279 | 0.26671 | *RP11-468E2.2* |
| ENSG00000172939 | 3 | 38196580 | 38306979 | 168 | 27 | 47306 | 0.62276 | 0.26672 | *OXSR1* |
| ENSG00000184271 | 12 | 51570719 | 51621477 | 142 | 31 | 42763 | 0.62272 | 0.26673 | *POU6F1* |
| ENSG00000188694 | 21 | 31643629 | 31665276 | 85 | 13 | 43680 | 0.62249 | 0.26681 | *KRTAP24-1* |
| ENSG00000152495 | 5 | 110549351 | 110840584 | 858 | 88 | 47827 | 0.62246 | 0.26682 | *CAMK4* |
| ENSG00000110092 | 11 | 69445855 | 69479242 | 90 | 18 | 37512 | 0.62215 | 0.26692 | *CCND1* |
| ENSG00000183117 | 8 | 2782875 | 4862494 | 17820 | 791 | 46936 | 0.62191 | 0.267 | *CSMD1* |
| ENSG00000257127 | 12 | 92805307 | 92834778 | 95 | 20 | 40646 | 0.62192 | 0.267 | *CLLU1* |
| ENSG00000143401 | 1 | 150180717 | 150218504 | 72 | 11 | 39673 | 0.62154 | 0.26712 | *ANP32E* |
| ENSG00000189430 | 19 | 55407508 | 55437508 | 214 | 40 | 40698 | 0.62131 | 0.2672 | *NCR1* |
| ENSG00000159899 | 9 | 35782151 | 35819729 | 80 | 12 | 36581 | 0.62115 | 0.26725 | *NPR2* |
| ENSG00000119782 | 2 | 24262571 | 24296551 | 108 | 14 | 47430 | 0.62113 | 0.26726 | *FKBP1B* |
| ENSG00000188162 | 11 | 17558920 | 17678697 | 333 | 54 | 45709 | 0.62112 | 0.26726 | *OTOG* |
| ENSG00000203722 | 6 | 150228014 | 150254257 | 75 | 13 | 36202 | 0.62104 | 0.26729 | *RAET1G* |
| ENSG00000135090 | 12 | 118577606 | 118820750 | 489 | 44 | 50289 | 0.621 | 0.2673 | *TAOK3* |
| ENSG00000119703 | 14 | 75520873 | 75555126 | 70 | 8 | 44762 | 0.62095 | 0.26732 | *ZC2HC1C* |
| ENSG00000116698 | 1 | 183431351 | 183577381 | 289 | 44 | 49451 | 0.62065 | 0.26742 | *SMG7* |
| ENSG00000146410 | 6 | 136542162 | 136581473 | 22 | 14 | 49495 | 0.62048 | 0.26747 | *MTFR2* |

| | | | | | | | | | |
|---|---|---|---|---|---|---|---|---|---|
| ENSG00000148660 | 10 | 75562259 | 75644343 | 114 | 16 | 41845 | 0.62029 | 0.26753 | *CAMK2G* |
| ENSG00000184697 | 16 | 3054713 | 3080072 | 61 | 17 | 52402 | 0.62 | 0.26763 | *CLDN6* |
| ENSG00000266076 | 17 | 63524220 | 63647151 | 345 | 53 | 35975 | 0.61985 | 0.26768 | *CTD-2535L24.2* |
| ENSG00000147679 | 8 | 117768742 | 117871702 | 264 | 43 | 31214 | 0.61948 | 0.2678 | *UTP23* |
| ENSG00000135333 | 6 | 93939738 | 94139265 | 557 | 81 | 45619 | 0.61915 | 0.26791 | *EPHA7* |
| ENSG00000119965 | 10 | 124680419 | 124723919 | 96 | 20 | 53359 | 0.61913 | 0.26792 | *C10orf88* |
| ENSG00000065054 | 16 | 2065357 | 2099027 | 86 | 20 | 41742 | 0.61907 | 0.26794 | *SLC9A3R2* |
| ENSG00000010017 | 6 | 13611730 | 13721796 | 205 | 31 | 53955 | 0.61902 | 0.26795 | *RANBP9* |
| ENSG00000169385 | 14 | 21413611 | 21434595 | 50 | 12 | 53024 | 0.61902 | 0.26795 | *RNASE2* |
| ENSG00000174684 | 11 | 66102843 | 66125163 | 43 | 10 | 37197 | 0.61874 | 0.26804 | *B3GNT1* |
| ENSG00000137098 | 9 | 35798042 | 35822269 | 45 | 11 | 32741 | 0.6187 | 0.26806 | *SPAG8* |
| ENSG00000156136 | 4 | 71848255 | 71906631 | 82 | 29 | 41216 | 0.61848 | 0.26813 | *DCK* |
| ENSG00000261210 | 16 | 19287061 | 19343467 | 212 | 31 | 38431 | 0.61826 | 0.2682 | *CLEC19A* |
| ENSG00000163320 | 3 | 88091094 | 88209035 | 292 | 12 | 30392 | 0.61763 | 0.26841 | *CGGBP1* |
| ENSG00000221869 | 8 | 48639471 | 48661648 | 15 | 10 | 46431 | 0.61709 | 0.26859 | *CEBPD* |
| ENSG00000214940 | 16 | 18401799 | 18451131 | 1 | 1 | 42718 | 0.61675 | 0.2687 | *NPIPA8* |
| ENSG00000060339 | 10 | 70470769 | 70562134 | 229 | 25 | 49060 | 0.6167 | 0.26872 | *CCAR1* |
| ENSG00000143643 | 1 | 231031989 | 231124621 | 374 | 30 | 48081 | 0.616 | 0.26895 | *TTC13* |
| ENSG00000179695 | 12 | 55835998 | 55856936 | 73 | 10 | 57013 | 0.616 | 0.26895 | *OR6C2* |
| ENSG00000148498 | 10 | 34388488 | 35114253 | 1858 | 115 | 39240 | 0.61597 | 0.26896 | *PARD3* |
| ENSG00000177558 | 19 | 35705703 | 35729632 | 131 | 21 | 45329 | 0.61585 | 0.269 | *FAM187B* |
| ENSG00000148339 | 9 | 130820480 | 130881524 | 169 | 8 | 32092 | 0.61577 | 0.26902 | *SLC25A25* |
| ENSG00000268788 | 4 | 76861883 | 76888526 | 79 | 15 | 47784 | 0.61567 | 0.26905 | *AC110615.1* |
| ENSG00000188641 | 1 | 97533299 | 98396605 | 2516 | 234 | 50888 | 0.6154 | 0.26915 | *DPYD* |
| ENSG00000126251 | 19 | 35851831 | 35873855 | 59 | 12 | 49517 | 0.61536 | 0.26916 | *GPR42* |
| ENSG00000176532 | 7 | 29593427 | 29616911 | 88 | 21 | 48272 | 0.61515 | 0.26923 | *PRR15* |
| ENSG00000189051 | 17 | 8284022 | 8311144 | 72 | 19 | 54681 | 0.61512 | 0.26924 | *RNF222* |
| ENSG00000248369 | 4 | 133346957 | 133378390 | 85 | 13 | 49924 | 0.61504 | 0.26926 | *RP11-62N21.1* |
| ENSG00000127993 | 7 | 92148087 | 92177319 | 60 | 9 | 52732 | 0.61507 | 0.26926 | *RBM48* |
| ENSG00000268614 | 19 | 7588481 | 7614945 | 97 | 23 | 50671 | 0.61503 | 0.26927 | *CTD-2207O23.10* |
| ENSG00000095539 | 10 | 102719275 | 102755628 | 66 | 10 | 34517 | 0.61472 | 0.26937 | *SEMA4G* |
| ENSG00000182372 | 8 | 1693944 | 1744738 | 204 | 17 | 54642 | 0.61439 | 0.26948 | *CLN8* |
| ENSG00000142185 | 21 | 45760046 | 45872964 | 327 | 40 | 47325 | 0.61437 | 0.26949 | *TRPM2* |
| ENSG00000164463 | 5 | 172473355 | 172576291 | 305 | 28 | 36683 | 0.61422 | 0.26953 | *CREBRF* |
| ENSG00000140750 | 16 | 24920706 | 25036987 | 234 | 26 | 45162 | 0.6141 | 0.26957 | *ARHGAP17* |
| ENSG00000111669 | 12 | 6966283 | 6990112 | 32 | 6 | 41097 | 0.61394 | 0.26963 | *TPI1* |

| | | | | | | | | | |
|---|---|---|---|---|---|---|---|---|---|
| ENSG00000136715 | 2 | 128688791 | 128795694 | 227 | 23 | 36185 | 0.61386 | 0.26965 | *SAP130* |
| ENSG00000268179 | 1 | 851264 | 876445 | 67 | 16 | 37667 | 0.61332 | 0.26983 | *AL645608.1* |
| ENSG00000140287 | 15 | 50524144 | 50568223 | 167 | 28 | 42263 | 0.61313 | 0.2699 | *HDC* |
| ENSG00000269049 | 16 | 14554842 | 14575054 | 29 | 12 | 49011 | 0.61306 | 0.26992 | *AC092291.2* |
| ENSG00000157227 | 14 | 23295766 | 23328236 | 110 | 30 | 50266 | 0.61299 | 0.26994 | *MMP14* |
| ENSG00000124225 | 20 | 56213448 | 56296592 | 270 | 63 | 39508 | 0.61299 | 0.26994 | *PMEPA1* |
| ENSG00000110931 | 12 | 121665497 | 121746111 | 260 | 46 | 47933 | 0.61281 | 0.27 | *CAMKK2* |
| ENSG00000164169 | 4 | 148548936 | 148615381 | 46 | 12 | 35470 | 0.6123 | 0.27017 | *PRMT10* |
| ENSG00000104957 | 19 | 13832574 | 13884110 | 149 | 20 | 50181 | 0.61203 | 0.27026 | *CCDC130* |
| ENSG00000242220 | 21 | 33938862 | 33969683 | 113 | 18 | 48898 | 0.61201 | 0.27027 | *TCP10L* |
| ENSG00000143971 | 2 | 67614451 | 67647677 | 108 | 11 | 55808 | 0.61149 | 0.27044 | *ETAA1* |
| ENSG00000107077 | 9 | 6710863 | 7185648 | 2452 | 201 | 42682 | 0.61149 | 0.27044 | *KDM4C* |
| ENSG00000107518 | 10 | 116843124 | 117718503 | 2130 | 80 | 46005 | 0.61111 | 0.27056 | *ATRNL1* |
| ENSG00000149346 | 20 | 10405951 | 10627477 | 745 | 65 | 46283 | 0.61099 | 0.2706 | *SLX4IP* |
| ENSG00000064933 | 2 | 190639107 | 190752355 | 279 | 34 | 41164 | 0.61097 | 0.27061 | *PMS1* |
| ENSG00000111203 | 12 | 2911788 | 2978957 | 157 | 19 | 28991 | 0.61077 | 0.27068 | *ITFG2* |
| ENSG00000159761 | 16 | 67690719 | 67712661 | 28 | 10 | 44884 | 0.61066 | 0.27071 | *C16orf86* |
| ENSG00000104442 | 8 | 66504694 | 66556442 | 201 | 11 | 54140 | 0.61053 | 0.27076 | *ARMC1* |
| ENSG00000141867 | 19 | 15337647 | 15453356 | 147 | 27 | 52537 | 0.61013 | 0.27089 | *BRD4* |
| ENSG00000124143 | 20 | 37220577 | 37289678 | 235 | 39 | 40518 | 0.61005 | 0.27091 | *ARHGAP40* |
| ENSG00000137200 | 6 | 37390995 | 37460603 | 122 | 26 | 42388 | 0.60988 | 0.27097 | *CMTR1* |
| ENSG00000087995 | 17 | 60491228 | 60537454 | 47 | 17 | 50257 | 0.60988 | 0.27097 | *METTL2A* |
| ENSG00000105429 | 19 | 42819761 | 42892921 | 116 | 10 | 46499 | 0.60981 | 0.27099 | *MEGF8* |
| ENSG00000127743 | 5 | 148740887 | 148793765 | 159 | 30 | 51438 | 0.60966 | 0.27104 | *IL17B* |
| ENSG00000183091 | 2 | 152331850 | 152601001 | 534 | 66 | 39538 | 0.6096 | 0.27106 | *NEB* |
| ENSG00000170265 | 7 | 148882577 | 148933339 | 170 | 24 | 39787 | 0.60958 | 0.27107 | *ZNF282* |
| ENSG00000167615 | 19 | 54950065 | 54983217 | 171 | 24 | 43558 | 0.60949 | 0.2711 | *LENG8* |
| ENSG00000146469 | 6 | 153061933 | 153090900 | 59 | 16 | 36954 | 0.60929 | 0.27116 | *VIP* |
| ENSG00000168787 | 6 | 29354416 | 29375448 | 132 | 9 | 39293 | 0.60892 | 0.27129 | *OR12D2* |
| ENSG00000139211 | 12 | 47459490 | 47483734 | 58 | 17 | 48279 | 0.60882 | 0.27132 | *AMIGO2* |
| ENSG00000147649 | 8 | 98646407 | 98750998 | 225 | 26 | 41360 | 0.60856 | 0.27141 | *MTDH* |
| ENSG00000114956 | 2 | 74143953 | 74196088 | 154 | 17 | 41960 | 0.60819 | 0.27153 | *DGUOK* |
| ENSG00000177380 | 19 | 49612646 | 49664283 | 194 | 30 | 48276 | 0.60795 | 0.27161 | *PPFIA3* |
| ENSG00000187627 | 2 | 87125076 | 87251104 | 28 | 12 | 25451 | 0.60787 | 0.27164 | *RGPD1* |
| ENSG00000112531 | 6 | 163825032 | 164009628 | 433 | 40 | 44154 | 0.60786 | 0.27164 | *QKI* |
| ENSG00000106829 | 9 | 82176688 | 82351658 | 399 | 50 | 46621 | 0.60784 | 0.27165 | *TLE4* |

| | | | | | | | | | |
|---|---|---|---|---|---|---|---|---|---|
| ENSG00000184110 | 16 | 28689879 | 28757051 | 14 | 9 | 39739 | 0.60779 | 0.27166 | *EIF3C* |
| ENSG00000116001 | 2 | 70426576 | 70485792 | 137 | 11 | 48548 | 0.60778 | 0.27167 | *TIA1* |
| ENSG00000171444 | 5 | 112347796 | 112834527 | 2121 | 115 | 40599 | 0.60772 | 0.27169 | *MCC* |
| ENSG00000137177 | 6 | 17749414 | 17997854 | 822 | 68 | 49403 | 0.60767 | 0.2717 | *KIF13A* |
| ENSG00000206344 | 6 | 31155537 | 31181745 | 507 | 38 | 47822 | 0.60744 | 0.27178 | *HCG27* |
| ENSG00000017373 | 17 | 36676251 | 36772183 | 218 | 40 | 42278 | 0.60744 | 0.27178 | *SRCIN1* |
| ENSG00000067066 | 2 | 231270657 | 231418805 | 553 | 66 | 44241 | 0.60733 | 0.27182 | *SP100* |
| ENSG00000091622 | 17 | 6344584 | 6469814 | 440 | 72 | 41189 | 0.60732 | 0.27182 | *PITPNM3* |
| ENSG00000101003 | 20 | 25378363 | 25443264 | 217 | 15 | 41525 | 0.6063 | 0.27216 | *GINS1* |
| ENSG00000074527 | 12 | 96041583 | 96194930 | 643 | 84 | 45773 | 0.60626 | 0.27217 | *NTN4* |
| ENSG00000101464 | 20 | 33138346 | 33274910 | 225 | 18 | 49074 | 0.60625 | 0.27217 | *PIGU* |
| ENSG00000171560 | 4 | 155494278 | 155521918 | 42 | 10 | 49548 | 0.60619 | 0.27219 | *FGA* |
| ENSG00000125900 | 20 | 1504897 | 1549489 | 156 | 23 | 43283 | 0.60614 | 0.27221 | *SIRPD* |
| ENSG00000163479 | 1 | 155968839 | 156000750 | 71 | 16 | 35686 | 0.60594 | 0.27228 | *SSR2* |
| ENSG00000214681 | 3 | 51897737 | 51919600 | 59 | 9 | 41203 | 0.60591 | 0.27229 | *IQCF5* |
| ENSG00000100557 | 14 | 57926019 | 57970585 | 116 | 15 | 39443 | 0.60575 | 0.27234 | *C14orf105* |
| ENSG00000027697 | 6 | 137508621 | 137550586 | 68 | 21 | 44635 | 0.60479 | 0.27266 | *IFNGR1* |
| ENSG00000186222 | 4 | 6707842 | 6729387 | 56 | 17 | 42586 | 0.60477 | 0.27267 | *BLOC1S4* |
| ENSG00000107882 | 10 | 104253744 | 104403292 | 386 | 25 | 46225 | 0.60466 | 0.2727 | *SUFU* |
| ENSG00000155833 | 9 | 105747593 | 105790770 | 187 | 20 | 43971 | 0.60459 | 0.27273 | *CYLC2* |
| ENSG00000132470 | 17 | 73707408 | 73763899 | 119 | 33 | 44537 | 0.60436 | 0.2728 | *ITGB4* |
| ENSG00000119915 | 10 | 103976085 | 103999346 | 41 | 9 | 48932 | 0.60419 | 0.27286 | *ELOVL3* |
| ENSG00000268750 | 19 | 58321087 | 58380309 | 211 | 29 | 41824 | 0.60416 | 0.27287 | *CTD-2583A14.10* |
| ENSG00000269343 | 19 | 58321099 | 58380018 | 211 | 29 | 41824 | 0.60416 | 0.27287 | *ZNF587B* |
| ENSG00000230178 | 5 | 180784269 | 180805263 | 1 | 1 | 42734 | 0.60407 | 0.2729 | *OR4F3* |
| ENSG00000082014 | 7 | 150925850 | 150984982 | 163 | 30 | 52068 | 0.60389 | 0.27296 | *SMARCD3* |
| ENSG00000124226 | 20 | 48542948 | 48580429 | 160 | 18 | 36207 | 0.60383 | 0.27298 | *RNF114* |
| ENSG00000007944 | 6 | 16119356 | 16158479 | 79 | 20 | 46272 | 0.60338 | 0.27313 | *MYLIP* |
| ENSG00000127325 | 12 | 70027140 | 70103256 | 355 | 36 | 46348 | 0.6032 | 0.27319 | *BEST3* |
| ENSG00000092140 | 14 | 31018329 | 31099269 | 202 | 33 | 46351 | 0.60316 | 0.2732 | *G2E3* |
| ENSG00000159214 | 1 | 44447031 | 44472200 | 46 | 13 | 34103 | 0.60311 | 0.27322 | *CCDC24* |
| ENSG00000271786 | 10 | 18921405 | 18954123 | 134 | 17 | 55503 | 0.60232 | 0.27348 | *RP11-139J15.7* |
| ENSG00000163702 | 3 | 9948758 | 9985314 | 105 | 21 | 32977 | 0.60206 | 0.27357 | *IL17RC* |
| ENSG00000121742 | 13 | 20786110 | 20816534 | 111 | 21 | 49911 | 0.60194 | 0.27361 | *GJB6* |
| ENSG00000254656 | 14 | 101336992 | 101361184 | 73 | 21 | 41958 | 0.60185 | 0.27364 | *RTL1* |
| ENSG00000163171 | 2 | 37859032 | 37975611 | 465 | 63 | 36984 | 0.6018 | 0.27365 | *CDC42EP3* |

| | | | | | | | | | |
|---|---|---|---|---|---|---|---|---|---|
| ENSG00000164096 | 4 | 120208207 | 120235600 | 102 | 16 | 56100 | 0.60173 | 0.27368 | *C4orf3* |
| ENSG00000165424 | 10 | 81132081 | 81215383 | 364 | 67 | 46253 | 0.60154 | 0.27374 | *ZCCHC24* |
| ENSG00000162032 | 16 | 1816713 | 1853701 | 227 | 19 | 33768 | 0.60141 | 0.27378 | *SPSB3* |
| ENSG00000004534 | 3 | 49967440 | 50147478 | 352 | 24 | 38150 | 0.60134 | 0.27381 | *RBM6* |
| ENSG00000160007 | 19 | 47411933 | 47518334 | 170 | 28 | 53067 | 0.60112 | 0.27388 | *ARHGAP35* |
| ENSG00000116478 | 1 | 32747687 | 32809236 | 44 | 27 | 52119 | 0.60089 | 0.27396 | *HDAC1* |
| ENSG00000179796 | 3 | 26654297 | 26762267 | 298 | 44 | 39255 | 0.60083 | 0.27398 | *LRRC3B* |
| ENSG00000177628 | 1 | 155194243 | 155224490 | 65 | 6 | 26727 | 0.60046 | 0.2741 | *GBA* |
| ENSG00000196365 | 19 | 5681845 | 5730583 | 162 | 28 | 49977 | 0.60039 | 0.27412 | *LONP1* |
| ENSG00000087245 | 16 | 55413612 | 55550603 | 703 | 84 | 45702 | 0.60031 | 0.27415 | *MMP2* |
| ENSG00000168214 | 4 | 26155077 | 26446541 | 649 | 87 | 47578 | 0.60004 | 0.27424 | *RBPJ* |
| ENSG00000080503 | 9 | 2005342 | 2203624 | 778 | 141 | 41439 | 0.59988 | 0.27429 | *SMARCA2* |
| ENSG00000270394 | 4 | 117210016 | 117231520 | 59 | 11 | 33151 | 0.59976 | 0.27433 | *MTRNR2L13* |
| ENSG00000180574 | 12 | 10648201 | 10685734 | 155 | 13 | 53668 | 0.59953 | 0.27441 | *EIF2S3L* |
| ENSG00000248385 | 19 | 54563149 | 54594634 | 137 | 19 | 53207 | 0.59954 | 0.27441 | *TARM1* |
| ENSG00000198398 | 3 | 190136444 | 190177665 | 159 | 29 | 48271 | 0.5995 | 0.27442 | *TMEM207* |
| ENSG00000142534 | 19 | 49989622 | 50012946 | 67 | 9 | 31294 | 0.59937 | 0.27446 | *RPS11* |
| ENSG00000158882 | 1 | 161185793 | 161210408 | 82 | 20 | 50148 | 0.59889 | 0.27462 | *TOMM40L* |
| ENSG00000060558 | 19 | 3126191 | 3173767 | 275 | 50 | 47685 | 0.5989 | 0.27462 | *GNA15* |
| ENSG00000197665 | 17 | 19049040 | 19072424 | 2 | 1 | 43316 | 0.59877 | 0.27466 | *AC007952.6* |
| ENSG00000119314 | 9 | 114970715 | 115105947 | 450 | 21 | 41170 | 0.5987 | 0.27469 | *PTBP3* |
| ENSG00000141756 | 17 | 39958932 | 39989465 | 90 | 12 | 52049 | 0.59861 | 0.27472 | *FKBP10* |
| ENSG00000151694 | 2 | 9618615 | 9705921 | 208 | 21 | 42680 | 0.59852 | 0.27475 | *ADAM17* |
| ENSG00000241370 | 6 | 30302908 | 30324661 | 76 | 21 | 38283 | 0.5985 | 0.27475 | *RPP21* |
| ENSG00000254585 | 15 | 23878691 | 23901175 | 76 | 20 | 37352 | 0.59842 | 0.27478 | *MAGEL2* |
| ENSG00000108849 | 17 | 42008172 | 42029836 | 60 | 13 | 39323 | 0.59836 | 0.2748 | *PPY* |
| ENSG00000012660 | 6 | 53122196 | 53223947 | 325 | 19 | 39170 | 0.598 | 0.27492 | *ELOVL5* |
| ENSG00000110717 | 11 | 67788084 | 67814111 | 70 | 12 | 51627 | 0.59796 | 0.27493 | *NDUFS8* |
| ENSG00000033178 | 4 | 68468370 | 68576897 | 405 | 23 | 53436 | 0.59773 | 0.27501 | *UBA6* |
| ENSG00000145965 | 6 | 3972909 | 3994372 | 83 | 14 | 47570 | 0.59764 | 0.27504 | *C6ORF50* |
| ENSG00000184828 | 18 | 45543044 | 45947123 | 1569 | 236 | 47396 | 0.59754 | 0.27507 | *ZBTB7C* |
| ENSG00000140093 | 14 | 94739650 | 94769608 | 80 | 16 | 36856 | 0.5974 | 0.27512 | *SERPINA10* |
| ENSG00000182319 | 8 | 8165258 | 8254008 | 369 | 53 | 42833 | 0.59722 | 0.27518 | *SGK223* |
| ENSG00000204524 | 19 | 57741973 | 57776503 | 141 | 16 | 50807 | 0.59706 | 0.27523 | *ZNF805* |
| ENSG00000215475 | 13 | 46344405 | 46435871 | 320 | 51 | 43325 | 0.59685 | 0.2753 | *SIAH3* |
| ENSG00000236032 | 3 | 97858170 | 97879249 | 102 | 6 | 51358 | 0.59679 | 0.27533 | *OR5H14* |

| | | | | | | | | | |
|---|---|---|---|---|---|---|---|---|---|
| ENSG00000064490 | 19 | 19293008 | 19322678 | 77 | 19 | 48584 | 0.59657 | 0.2754 | *RFXANK* |
| ENSG00000116580 | 1 | 155709508 | 155839191 | 210 | 23 | 42482 | 0.59653 | 0.27541 | *GON4L* |
| ENSG00000171798 | 10 | 134963951 | 135049916 | 436 | 49 | 42826 | 0.5965 | 0.27542 | *KNDC1* |
| ENSG00000213186 | 3 | 160140233 | 160213561 | 169 | 26 | 51244 | 0.59645 | 0.27544 | *TRIM59* |
| ENSG00000204352 | 9 | 96070481 | 96118696 | 205 | 31 | 39236 | 0.59645 | 0.27544 | *C9orf129* |
| ENSG00000109680 | 4 | 26568059 | 26766973 | 260 | 21 | 42673 | 0.59639 | 0.27546 | *TBC1D19* |
| ENSG00000168496 | 11 | 61550109 | 61574716 | 29 | 5 | 30431 | 0.59606 | 0.27557 | *FEN1* |
| ENSG00000214866 | 2 | 3741453 | 3846122 | 439 | 65 | 48463 | 0.59602 | 0.27558 | *DCDC2C* |
| ENSG00000149651 | 20 | 44340990 | 44364469 | 119 | 7 | 20253 | 0.59603 | 0.27558 | *SPINT4* |
| ENSG00000269360 | 15 | 93739295 | 93761277 | 108 | 17 | 53743 | 0.59563 | 0.27571 | *AC112693.2* |
| ENSG00000174796 | 4 | 76429156 | 76485683 | 170 | 24 | 47186 | 0.59557 | 0.27573 | *THAP6* |
| ENSG00000134363 | 5 | 52766239 | 52792964 | 97 | 18 | 47532 | 0.59553 | 0.27575 | *FST* |
| ENSG00000198734 | 1 | 169473404 | 169565826 | 397 | 41 | 47934 | 0.59548 | 0.27576 | *F5* |
| ENSG00000174325 | 2 | 189588882 | 189664831 | 287 | 22 | 52829 | 0.59529 | 0.27582 | *DIRC1* |
| ENSG00000156269 | 4 | 80136848 | 80257204 | 263 | 27 | 42687 | 0.59506 | 0.2759 | *NAA11* |
| ENSG00000092098 | 14 | 24605892 | 24639870 | 57 | 21 | 47163 | 0.59428 | 0.27616 | *RNF31* |
| ENSG00000074181 | 19 | 15260444 | 15321792 | 190 | 18 | 40436 | 0.5943 | 0.27616 | *NOTCH3* |
| ENSG00000198835 | 1 | 228327553 | 228357527 | 42 | 13 | 45470 | 0.59421 | 0.27619 | *GJC2* |
| ENSG00000143847 | 1 | 202985626 | 203057868 | 229 | 40 | 51535 | 0.59389 | 0.27629 | *PPFIA4* |
| ENSG00000138640 | 4 | 89637106 | 90042549 | 1462 | 99 | 44473 | 0.59388 | 0.2763 | *FAM13A* |
| ENSG00000177398 | 21 | 43473068 | 43573563 | 592 | 94 | 47534 | 0.59382 | 0.27632 | *UMODL1* |
| ENSG00000161509 | 17 | 72828162 | 72867627 | 92 | 8 | 42005 | 0.59371 | 0.27635 | *GRIN2C* |
| ENSG00000141905 | 19 | 3349561 | 3479215 | 469 | 90 | 40944 | 0.5936 | 0.27639 | *NFIC* |
| ENSG00000136698 | 2 | 131340339 | 131367123 | 1 | 1 | 43671 | 0.59327 | 0.2765 | *CFC1* |
| ENSG00000142856 | 1 | 63896441 | 64069392 | 487 | 30 | 46308 | 0.5931 | 0.27656 | *ITGB3BP* |
| ENSG00000258839 | 16 | 89968527 | 89997385 | 117 | 19 | 32705 | 0.59308 | 0.27656 | *MC1R* |
| ENSG00000110318 | 11 | 101775746 | 101881789 | 288 | 27 | 39857 | 0.593 | 0.27659 | *KIAA1377* |
| ENSG00000152430 | 2 | 198581603 | 198661486 | 129 | 16 | 55096 | 0.59297 | 0.2766 | *BOLL* |
| ENSG00000186350 | 9 | 137198944 | 137342431 | 486 | 59 | 45827 | 0.59291 | 0.27662 | *RXRA* |
| ENSG00000204183 | 20 | 34010827 | 34033248 | 70 | 10 | 29693 | 0.59278 | 0.27666 | *GDF5OS* |
| ENSG00000186684 | 2 | 127931696 | 127987654 | 191 | 26 | 50618 | 0.59272 | 0.27668 | *CYP27C1* |
| ENSG00000130164 | 19 | 11190038 | 11254492 | 287 | 37 | 44086 | 0.59253 | 0.27675 | *LDLR* |
| ENSG00000095917 | 16 | 1296060 | 1318532 | 115 | 19 | 34192 | 0.59215 | 0.27687 | *TPSD1* |
| ENSG00000088035 | 1 | 63823261 | 63914233 | 240 | 28 | 50071 | 0.59214 | 0.27688 | *ALG6* |
| ENSG00000177494 | 3 | 111301747 | 111324290 | 54 | 14 | 46436 | 0.59208 | 0.2769 | *ZBED2* |
| ENSG00000176732 | 2 | 24328241 | 24356347 | 109 | 9 | 54347 | 0.59193 | 0.27695 | *PFN4* |

| | | | | | | | | | |
|---|---|---|---|---|---|---|---|---|---|
| ENSG00000164438 | 5 | 170726288 | 170749138 | 53 | 14 | 50087 | 0.59171 | 0.27702 | *TLX3* |
| ENSG00000167733 | 19 | 5670615 | 5698533 | 79 | 17 | 51338 | 0.59172 | 0.27702 | *HSD11B1L* |
| ENSG00000184939 | 16 | 68553993 | 68619975 | 279 | 13 | 52640 | 0.59161 | 0.27705 | *ZFP90* |
| ENSG00000130173 | 19 | 11338178 | 11362619 | 49 | 9 | 30804 | 0.59155 | 0.27707 | *C19orf80* |
| ENSG00000115641 | 2 | 105964169 | 106064970 | 351 | 70 | 47901 | 0.59154 | 0.27708 | *FHL2* |
| ENSG00000181234 | 12 | 128741948 | 129202460 | 1506 | 211 | 43029 | 0.59149 | 0.2771 | *TMEM132C* |
| ENSG00000137504 | 11 | 85360752 | 85403951 | 99 | 19 | 47023 | 0.59143 | 0.27711 | *CREBZF* |
| ENSG00000106245 | 7 | 98996264 | 99027239 | 46 | 14 | 41765 | 0.59118 | 0.2772 | *BUD31* |
| ENSG00000107859 | 10 | 103979943 | 104011231 | 58 | 10 | 50979 | 0.59117 | 0.2772 | *PITX3* |
| ENSG00000100299 | 22 | 51051182 | 51076607 | 119 | 29 | 42404 | 0.59092 | 0.27729 | *ARSA* |
| ENSG00000108424 | 17 | 45716842 | 45772871 | 107 | 16 | 49247 | 0.59073 | 0.27735 | *KPNB1* |
| ENSG00000141194 | 17 | 56222494 | 56243517 | 61 | 11 | 41731 | 0.59052 | 0.27742 | *OR4D1* |
| ENSG00000102547 | 13 | 49872786 | 50028262 | 459 | 58 | 44624 | 0.59029 | 0.2775 | *CAB39L* |
| ENSG00000213780 | 6 | 30865961 | 30891883 | 103 | 15 | 40797 | 0.59026 | 0.27751 | *GTF2H4* |
| ENSG00000181449 | 3 | 181419714 | 181442221 | 20 | 7 | 43063 | 0.59019 | 0.27753 | *SOX2* |
| ENSG00000182898 | 1 | 152046620 | 152071540 | 23 | 12 | 44541 | 0.59011 | 0.27756 | *TCHHL1* |
| ENSG00000167977 | 16 | 2722476 | 2769031 | 92 | 22 | 43710 | 0.58998 | 0.2776 | *KCTD5* |
| ENSG00000187747 | 11 | 5592107 | 5613114 | 160 | 16 | 36887 | 0.58944 | 0.27778 | *OR52B6* |
| ENSG00000229453 | 3 | 48338336 | 48379831 | 111 | 5 | 25904 | 0.58935 | 0.27781 | *SPINK8* |
| ENSG00000115128 | 2 | 24280454 | 24309313 | 104 | 12 | 49853 | 0.58933 | 0.27782 | *SF3B14* |
| ENSG00000100263 | 22 | 29645841 | 29674198 | 63 | 22 | 50489 | 0.58929 | 0.27783 | *RHBDD3* |
| ENSG00000144354 | 2 | 174209548 | 174243725 | 141 | 24 | 47733 | 0.58908 | 0.27791 | *CDCA7* |
| ENSG00000239779 | 2 | 74675456 | 74698018 | 7 | 5 | 42735 | 0.58843 | 0.27812 | *WBP1* |
| ENSG00000135473 | 12 | 56700007 | 56737837 | 55 | 9 | 44358 | 0.58842 | 0.27812 | *PAN2* |
| ENSG00000269846 | 20 | 35714188 | 35735581 | 32 | 12 | 51504 | 0.58826 | 0.27818 | *RBL1* |
| ENSG00000103510 | 16 | 31117075 | 31152714 | 56 | 8 | 23906 | 0.58816 | 0.27821 | *KAT8* |
| ENSG00000101446 | 20 | 44131101 | 44154264 | 57 | 12 | 41033 | 0.58804 | 0.27825 | *SPINT3* |
| ENSG00000171711 | 8 | 7742151 | 7764233 | 7 | 2 | 42939 | 0.588 | 0.27827 | *DEFB4A* |
| ENSG00000166226 | 12 | 69969114 | 70005350 | 113 | 12 | 46540 | 0.58769 | 0.27837 | *CCT2* |
| ENSG00000163053 | 2 | 230889698 | 230943715 | 199 | 29 | 49253 | 0.58764 | 0.27839 | *SLC16A14* |
| ENSG00000184224 | 11 | 67360351 | 67384177 | 44 | 8 | 46305 | 0.58763 | 0.27839 | *C11orf72* |
| ENSG00000182533 | 3 | 8765486 | 8893492 | 605 | 80 | 37607 | 0.5876 | 0.2784 | *CAV3* |
| ENSG00000181619 | 14 | 59885740 | 59942060 | 243 | 24 | 42968 | 0.58759 | 0.2784 | *GPR135* |
| ENSG00000169495 | 8 | 38821683 | 38856181 | 92 | 9 | 53916 | 0.58697 | 0.27861 | *HTRA4* |
| ENSG00000256566 | 20 | 2437858 | 2499553 | 131 | 27 | 45263 | 0.58698 | 0.27861 | *RP4-734P14.4* |
| ENSG00000161647 | 17 | 41868167 | 41920538 | 99 | 13 | 46005 | 0.58676 | 0.27868 | *MPP3* |

| | | | | | | | | | |
|---|---|---|---|---|---|---|---|---|---|
| ENSG00000105967 | 7 | 115565202 | 115809950 | 681 | 61 | 46375 | 0.58662 | 0.27873 | *TFEC* |
| ENSG00000064652 | 5 | 122169134 | 122375049 | 738 | 38 | 35432 | 0.58645 | 0.27879 | *SNX24* |
| ENSG00000095383 | 9 | 100951311 | 101027915 | 354 | 40 | 46256 | 0.58633 | 0.27883 | *TBC1D2* |
| ENSG00000176912 | 18 | 631320 | 668340 | 148 | 25 | 40332 | 0.58628 | 0.27884 | *C18orf56* |
| ENSG00000253304 | 1 | 29435940 | 29460447 | 69 | 11 | 34339 | 0.58627 | 0.27885 | *TMEM200B* |
| ENSG00000106346 | 7 | 6134515 | 6211195 | 237 | 29 | 42602 | 0.58618 | 0.27888 | *USP42* |
| ENSG00000131848 | 19 | 56722681 | 56889752 | 603 | 113 | 40925 | 0.58545 | 0.27912 | *ZSCAN5A* |
| ENSG00000120694 | 13 | 31700762 | 31746525 | 101 | 14 | 43143 | 0.58495 | 0.27929 | *HSPH1* |
| ENSG00000105726 | 19 | 19746007 | 19784502 | 114 | 22 | 54437 | 0.58492 | 0.2793 | *ATP13A1* |
| ENSG00000092010 | 14 | 24595367 | 24618176 | 44 | 11 | 35101 | 0.58473 | 0.27936 | *PSME1* |
| ENSG00000170909 | 19 | 54587933 | 54616000 | 146 | 21 | 48794 | 0.58454 | 0.27943 | *OSCAR* |
| ENSG00000124074 | 16 | 67686848 | 67711168 | 28 | 11 | 43270 | 0.58415 | 0.27956 | *ENKD1* |
| ENSG00000168014 | 11 | 73713763 | 73892255 | 375 | 35 | 42276 | 0.58401 | 0.27961 | *C2CD3* |
| ENSG00000176834 | 12 | 118491398 | 118583831 | 330 | 67 | 46635 | 0.5839 | 0.27964 | *VSIG10* |
| ENSG00000159023 | 1 | 29203603 | 29456553 | 584 | 47 | 33846 | 0.58386 | 0.27966 | *EPB41* |
| ENSG00000261459 | 16 | 30527244 | 30556173 | 32 | 14 | 38620 | 0.58373 | 0.2797 | *ZNF747* |
| ENSG00000008323 | 12 | 6409602 | 6447672 | 83 | 22 | 40177 | 0.58356 | 0.27976 | *PLEKHG6* |
| ENSG00000164088 | 3 | 52269841 | 52294613 | 38 | 8 | 45979 | 0.58338 | 0.27982 | *PPM1M* |
| ENSG00000174226 | 8 | 101575116 | 101685643 | 505 | 58 | 43002 | 0.58339 | 0.27982 | *SNX31* |
| ENSG00000186493 | 5 | 2742245 | 2765508 | 53 | 15 | 40838 | 0.58331 | 0.27984 | *C5orf38* |
| ENSG00000164220 | 5 | 75901307 | 75929259 | 149 | 24 | 48465 | 0.58327 | 0.27986 | *F2RL2* |
| ENSG00000182472 | 19 | 39210827 | 39270544 | 278 | 27 | 49392 | 0.58301 | 0.27994 | *CAPN12* |
| ENSG00000143799 | 1 | 226538392 | 226605780 | 226 | 17 | 43026 | 0.58286 | 0.27999 | *PARP1* |
| ENSG00000268602 | 11 | 107572998 | 107605135 | 85 | 16 | 43090 | 0.58286 | 0.28 | *AP002353.1* |
| ENSG00000105364 | 19 | 10352577 | 10380721 | 62 | 18 | 53981 | 0.58277 | 0.28003 | *MRPL4* |
| ENSG00000165972 | 12 | 96250826 | 96346752 | 405 | 50 | 46187 | 0.58268 | 0.28006 | *CCDC38* |
| ENSG00000101247 | 20 | 13755596 | 13809067 | 176 | 31 | 47504 | 0.5826 | 0.28008 | *NDUFAF5* |
| ENSG00000140931 | 16 | 66627777 | 66657795 | 70 | 13 | 40348 | 0.58201 | 0.28028 | *CMTM3* |
| ENSG00000241360 | 22 | 38044734 | 38072941 | 108 | 17 | 47926 | 0.58165 | 0.2804 | *PDXP* |
| ENSG00000132631 | 20 | 18784370 | 18805033 | 83 | 23 | 52140 | 0.58157 | 0.28043 | *SCP2D1* |
| ENSG00000179059 | 4 | 188906925 | 188936204 | 143 | 21 | 26442 | 0.58121 | 0.28055 | *ZFP42* |
| ENSG00000173610 | 4 | 70444135 | 70528965 | 415 | 38 | 52517 | 0.58112 | 0.28058 | *UGT2A1* |
| ENSG00000270386 | 4 | 70444912 | 70528967 | 415 | 38 | 52517 | 0.58112 | 0.28058 | *UGT2A1* |
| ENSG00000160949 | 8 | 145644165 | 145679827 | 94 | 11 | 49657 | 0.58097 | 0.28063 | *TONSL* |
| ENSG00000088205 | 2 | 118562226 | 118599955 | 130 | 20 | 51812 | 0.58089 | 0.28066 | *DDX18* |
| ENSG00000137486 | 11 | 74965226 | 75072873 | 251 | 58 | 45382 | 0.58087 | 0.28067 | *ARRB1* |

| | | | | | | | | | |
|---|---|---|---|---|---|---|---|---|---|
| ENSG00000244405 | 3 | 185754097 | 185838107 | 137 | 23 | 45005 | 0.58063 | 0.28074 | *ETV5* |
| ENSG00000141161 | 17 | 33464836 | 33526364 | 168 | 29 | 46473 | 0.58044 | 0.28081 | *UNC45B* |
| ENSG00000169413 | 14 | 21239210 | 21260626 | 149 | 25 | 36404 | 0.5801 | 0.28092 | *RNASE6* |
| ENSG00000097007 | 9 | 133579333 | 133773062 | 601 | 66 | 34204 | 0.57984 | 0.28101 | *ABL1* |
| ENSG00000108298 | 17 | 37346536 | 37370980 | 53 | 14 | 49150 | 0.5797 | 0.28106 | *RPL19* |
| ENSG00000174100 | 17 | 36442989 | 36489101 | 87 | 16 | 34564 | 0.57963 | 0.28108 | *MRPL45* |
| ENSG00000138161 | 10 | 124581665 | 124649146 | 132 | 26 | 54967 | 0.57957 | 0.2811 | *CUZD1* |
| ENSG00000103507 | 16 | 31107428 | 31134110 | 42 | 5 | 19078 | 0.57934 | 0.28118 | *BCKDK* |
| ENSG00000089159 | 12 | 120638250 | 120713574 | 120 | 21 | 40408 | 0.57929 | 0.2812 | *PXN* |
| ENSG00000112053 | 6 | 35901291 | 36002645 | 287 | 37 | 45679 | 0.57922 | 0.28122 | *SLC26A8* |
| ENSG00000168958 | 2 | 228179867 | 228232550 | 224 | 24 | 39109 | 0.57913 | 0.28125 | *MFF* |
| ENSG00000113739 | 5 | 172731716 | 172766506 | 62 | 25 | 40762 | 0.57896 | 0.28131 | *STC2* |
| ENSG00000177000 | 1 | 11835780 | 11876977 | 253 | 19 | 52420 | 0.57857 | 0.28144 | *MTHFR* |
| ENSG00000100056 | 22 | 19107792 | 19142197 | 114 | 25 | 48869 | 0.57852 | 0.28146 | *DGCR14* |
| ENSG00000269624 | 1 | 63063437 | 63085249 | 23 | 6 | 41275 | 0.57807 | 0.28161 | *AL138847.1* |
| ENSG00000107819 | 10 | 102780991 | 102810998 | 61 | 24 | 43837 | 0.57797 | 0.28164 | *SFXN3* |
| ENSG00000167283 | 11 | 118261869 | 118312211 | 98 | 17 | 40506 | 0.57787 | 0.28168 | *ATP5L* |
| ENSG00000156886 | 16 | 31394633 | 31447826 | 95 | 23 | 51840 | 0.57781 | 0.2817 | *ITGAD* |
| ENSG00000196159 | 4 | 126227554 | 126424087 | 591 | 71 | 49367 | 0.57776 | 0.28171 | *FAT4* |
| ENSG00000165660 | 10 | 126480354 | 126535239 | 117 | 20 | 45997 | 0.57718 | 0.28191 | *FAM175B* |
| ENSG00000155090 | 8 | 103651007 | 103678130 | 99 | 20 | 47134 | 0.5771 | 0.28194 | *KLF10* |
| ENSG00000078142 | 18 | 39525171 | 39677794 | 291 | 27 | 45732 | 0.57691 | 0.282 | *PIK3C3* |
| ENSG00000197084 | 1 | 152767311 | 152789107 | 59 | 13 | 51422 | 0.5765 | 0.28214 | *LCE1C* |
| ENSG00000215749 | 15 | 82920438 | 82949159 | 2 | 1 | 43294 | 0.5762 | 0.28224 | *GOLGA6L18* |
| ENSG00000134313 | 2 | 8855408 | 8987760 | 252 | 37 | 46353 | 0.57617 | 0.28225 | *KIDINS220* |
| ENSG00000179751 | 19 | 39683471 | 39704906 | 52 | 5 | 54675 | 0.57611 | 0.28227 | *SYCN* |
| ENSG00000171815 | 5 | 140420979 | 140443512 | 37 | 13 | 48293 | 0.57597 | 0.28232 | *PCDHB1* |
| ENSG00000143436 | 1 | 151722119 | 151746040 | 69 | 11 | 51252 | 0.57595 | 0.28233 | *MRPL9* |
| ENSG00000119640 | 14 | 75509924 | 75546186 | 78 | 10 | 45425 | 0.57584 | 0.28236 | *ACYP1* |
| ENSG00000256097 | 3 | 52087076 | 52107567 | 23 | 7 | 31009 | 0.57578 | 0.28238 | *LINC00696* |
| ENSG00000268146 | 1 | 244217632 | 244241211 | 55 | 17 | 52336 | 0.57574 | 0.28239 | *AL590483.1* |
| ENSG00000119616 | 14 | 75169847 | 75213394 | 64 | 11 | 48391 | 0.5757 | 0.28241 | *FCF1* |
| ENSG00000055950 | 10 | 102719215 | 102757272 | 72 | 11 | 33940 | 0.57525 | 0.28256 | *MRPL43* |
| ENSG00000110925 | 12 | 51444990 | 51487447 | 118 | 17 | 34632 | 0.57522 | 0.28257 | *CSRNP2* |
| ENSG00000151468 | 10 | 12928627 | 13151652 | 1071 | 140 | 44290 | 0.57504 | 0.28263 | *CCDC3* |
| ENSG00000122026 | 13 | 27815446 | 27840828 | 70 | 16 | 44634 | 0.57479 | 0.28272 | *RPL21* |

| | | | | | | | | | |
|---|---|---|---|---|---|---|---|---|---|
| ENSG00000205730 | 16 | 19115254 | 19142946 | 44 | 7 | 31233 | 0.57443 | 0.28284 | *ITPRIPL2* |
| ENSG00000167614 | 19 | 54916393 | 54958080 | 200 | 36 | 43638 | 0.57439 | 0.28285 | *TTYH1* |
| ENSG00000145882 | 5 | 148727570 | 148759216 | 68 | 12 | 49188 | 0.57413 | 0.28294 | *PCYOX1L* |
| ENSG00000139547 | 12 | 57335219 | 57363158 | 77 | 9 | 34106 | 0.57394 | 0.28301 | *RDH16* |
| ENSG00000043591 | 10 | 115793806 | 115816667 | 74 | 8 | 53832 | 0.57383 | 0.28304 | *ADRB1* |
| ENSG00000108599 | 17 | 19797615 | 19891656 | 175 | 16 | 53073 | 0.57375 | 0.28307 | *AKAP10* |
| ENSG00000143793 | 1 | 228278427 | 228303112 | 37 | 14 | 50715 | 0.57353 | 0.28314 | *C1orf35* |
| ENSG00000129646 | 17 | 74260130 | 74313761 | 171 | 20 | 34042 | 0.57332 | 0.28321 | *QRICH2* |
| ENSG00000071282 | 3 | 8533393 | 8619805 | 331 | 36 | 39450 | 0.5733 | 0.28322 | *LMCD1* |
| ENSG00000088280 | 1 | 23745056 | 23821061 | 207 | 22 | 42900 | 0.57304 | 0.28331 | *ASAP3* |
| ENSG00000174137 | 4 | 1609642 | 1696029 | 320 | 33 | 46395 | 0.57292 | 0.28335 | *FAM53A* |
| ENSG00000167671 | 19 | 4434996 | 4467819 | 118 | 20 | 51674 | 0.57287 | 0.28337 | *UBXN6* |
| ENSG00000139344 | 12 | 96327071 | 96372370 | 190 | 20 | 49162 | 0.57276 | 0.2834 | *AMDHD1* |
| ENSG00000072062 | 19 | 14192500 | 14238896 | 68 | 15 | 49712 | 0.57269 | 0.28343 | *PRKACA* |
| ENSG00000104687 | 8 | 30525583 | 30595443 | 201 | 28 | 35948 | 0.57212 | 0.28362 | *GSR* |
| ENSG00000170276 | 11 | 111772966 | 111799574 | 59 | 11 | 36989 | 0.57205 | 0.28364 | *HSPB2* |
| ENSG00000127585 | 16 | 732500 | 765829 | 77 | 15 | 29920 | 0.57178 | 0.28373 | *FBXL16* |
| ENSG00000181023 | 11 | 5747681 | 5768768 | 113 | 15 | 47690 | 0.57174 | 0.28375 | *OR56B1* |
| ENSG00000196099 | 11 | 123666043 | 123687095 | 82 | 24 | 45576 | 0.57161 | 0.28379 | *OR6M1* |
| ENSG00000155506 | 5 | 154082462 | 154207167 | 262 | 39 | 42726 | 0.57143 | 0.28386 | *LARP1* |
| ENSG00000110031 | 11 | 58284344 | 58355693 | 140 | 20 | 50877 | 0.57141 | 0.28386 | *LPXN* |
| ENSG00000184261 | 2 | 47733720 | 47808078 | 207 | 53 | 44385 | 0.57116 | 0.28395 | *KCNK12* |
| ENSG00000138795 | 4 | 108958701 | 109100112 | 303 | 32 | 37843 | 0.57113 | 0.28396 | *LEF1* |
| ENSG00000163106 | 4 | 95209686 | 95274027 | 171 | 16 | 51620 | 0.57103 | 0.28399 | *HPGDS* |
| ENSG00000148541 | 10 | 60995890 | 61132939 | 348 | 47 | 45025 | 0.57095 | 0.28402 | *FAM13C* |
| ENSG00000110852 | 12 | 9995583 | 10032735 | 162 | 29 | 45164 | 0.57087 | 0.28404 | *CLEC2B* |
| ENSG00000143190 | 1 | 167180066 | 167406582 | 470 | 57 | 45275 | 0.5708 | 0.28407 | *POU2F1* |
| ENSG00000171864 | 20 | 4692556 | 4719106 | 138 | 28 | 42278 | 0.57044 | 0.28419 | *PRND* |
| ENSG00000109991 | 11 | 57095848 | 57148073 | 138 | 21 | 42077 | 0.57039 | 0.28421 | *P2RX3* |
| ENSG00000091664 | 11 | 22349643 | 22411049 | 191 | 42 | 48825 | 0.57034 | 0.28422 | *SLC17A6* |
| ENSG00000186469 | 14 | 52282913 | 52456060 | 659 | 99 | 39265 | 0.57028 | 0.28424 | *GNG2* |
| ENSG00000135740 | 16 | 67261586 | 67316093 | 60 | 23 | 48451 | 0.57025 | 0.28425 | *SLC9A5* |
| ENSG00000198836 | 3 | 193300933 | 193425612 | 387 | 26 | 54353 | 0.57021 | 0.28427 | *OPA1* |
| ENSG00000132801 | 20 | 44476256 | 44517761 | 153 | 17 | 45856 | 0.57002 | 0.28433 | *ZSWIM3* |
| ENSG00000187944 | 2 | 197659726 | 197685000 | 49 | 12 | 44076 | 0.56966 | 0.28445 | *C2orf66* |
| ENSG00000163060 | 2 | 95527178 | 95552574 | 53 | 10 | 43199 | 0.5696 | 0.28447 | *TEKT4* |

| | | | | | | | | | |
|---|---|---|---|---|---|---|---|---|---|
| ENSG00000105854 | 7 | 95024175 | 95074510 | 181 | 23 | 49543 | 0.56953 | 0.2845 | *PON2* |
| ENSG00000070444 | 17 | 2277354 | 2314412 | 117 | 19 | 37393 | 0.56946 | 0.28452 | *MNT* |
| ENSG00000115216 | 2 | 27640657 | 27675126 | 57 | 6 | 34065 | 0.56942 | 0.28453 | *NRBP1* |
| ENSG00000137075 | 9 | 36326393 | 36497545 | 309 | 43 | 51202 | 0.56941 | 0.28454 | *RNF38* |
| ENSG00000186090 | 3 | 183739332 | 183767157 | 119 | 22 | 44457 | 0.56919 | 0.28462 | *HTR3D* |
| ENSG00000203805 | 10 | 122206466 | 122359367 | 614 | 95 | 45239 | 0.56881 | 0.28474 | *PPAPDC1A* |
| ENSG00000101452 | 20 | 37580942 | 37678366 | 199 | 16 | 55482 | 0.56876 | 0.28476 | *DHX35* |
| ENSG00000168040 | 11 | 70039269 | 70063496 | 81 | 22 | 51168 | 0.5687 | 0.28478 | *FADD* |
| ENSG00000147316 | 8 | 6254113 | 6511144 | 1431 | 141 | 41177 | 0.56834 | 0.2849 | *MCPH1* |
| ENSG00000143774 | 1 | 228317663 | 228346685 | 36 | 11 | 47979 | 0.56827 | 0.28492 | *GUK1* |
| ENSG00000180008 | 14 | 55483948 | 55526206 | 91 | 13 | 54321 | 0.56822 | 0.28494 | *SOCS4* |
| ENSG00000259803 | 16 | 89252406 | 89278072 | 109 | 16 | 46353 | 0.56801 | 0.28502 | *SLC22A31* |
| ENSG00000183439 | 4 | 165865598 | 165908820 | 135 | 15 | 43145 | 0.56796 | 0.28503 | *TRIM61* |
| ENSG00000106538 | 7 | 150025408 | 150048763 | 80 | 11 | 49107 | 0.56733 | 0.28524 | *RARRES2* |
| ENSG00000176845 | 17 | 81027567 | 81062864 | 138 | 40 | 46704 | 0.56728 | 0.28526 | *METRNL* |
| ENSG00000187672 | 3 | 55532336 | 56512391 | 3287 | 230 | 47521 | 0.5671 | 0.28532 | *ERC2* |
| ENSG00000143178 | 1 | 168240278 | 168293664 | 141 | 29 | 40213 | 0.56706 | 0.28534 | *TBX19* |
| ENSG00000174586 | 19 | 58855723 | 58884214 | 75 | 12 | 38940 | 0.56702 | 0.28535 | *ZNF497* |
| ENSG00000167139 | 15 | 74155949 | 74191555 | 101 | 20 | 39113 | 0.56697 | 0.28537 | *TBC1D21* |
| ENSG00000121774 | 1 | 32469430 | 32536451 | 56 | 25 | 45710 | 0.56667 | 0.28547 | *KHDRBS1* |
| ENSG00000165757 | 10 | 30291729 | 30414423 | 317 | 58 | 41244 | 0.5665 | 0.28553 | *KIAA1462* |
| ENSG00000159256 | 21 | 37682487 | 37768446 | 314 | 22 | 46974 | 0.56638 | 0.28557 | *MORC3* |
| ENSG00000166548 | 16 | 66531906 | 66596447 | 128 | 10 | 23807 | 0.5663 | 0.28559 | *TK2* |
| ENSG00000205328 | 12 | 55784214 | 55805289 | 70 | 7 | 34343 | 0.56619 | 0.28563 | *OR6C65* |
| ENSG00000158050 | 2 | 96798905 | 96821179 | 30 | 10 | 47117 | 0.56601 | 0.28569 | *DUSP2* |
| ENSG00000205281 | 15 | 82625130 | 82651706 | 1 | 1 | 43440 | 0.56599 | 0.2857 | *GOLGA6L10* |
| ENSG00000229086 | 21 | 34527776 | 34552541 | 99 | 23 | 53745 | 0.56597 | 0.28571 | *C21orf54* |
| ENSG00000119866 | 2 | 60668302 | 60790702 | 213 | 48 | 42495 | 0.56581 | 0.28576 | *BCL11A* |
| ENSG00000173926 | 5 | 126193406 | 126376500 | 520 | 24 | 28785 | 0.56557 | 0.28584 | *Mar-03* |
| ENSG00000059588 | 1 | 234517059 | 234624849 | 511 | 66 | 44237 | 0.56547 | 0.28588 | *TARBP1* |
| ENSG00000163125 | 1 | 150325567 | 150459042 | 462 | 25 | 40541 | 0.56535 | 0.28592 | *RPRD2* |
| ENSG00000130429 | 7 | 98961872 | 99002424 | 50 | 11 | 48226 | 0.56523 | 0.28596 | *ARPC1B* |
| ENSG00000268163 | 19 | 57939338 | 57998904 | 236 | 19 | 50575 | 0.56523 | 0.28596 | *AC004076.9* |
| ENSG00000072422 | 10 | 62619196 | 62771198 | 442 | 58 | 45371 | 0.56505 | 0.28602 | *RHOBTB1* |
| ENSG00000273079 | 12 | 13683165 | 14143053 | 1601 | 170 | 47124 | 0.56506 | 0.28602 | *GRIN2B* |
| ENSG00000267849 | 6 | 44390472 | 44410720 | 42 | 15 | 36003 | 0.56497 | 0.28605 | *AL133262.1* |

| | | | | | | | | | |
|---|---|---|---|---|---|---|---|---|---|
| ENSG00000269650 | 20 | 34746461 | 34768180 | 38 | 8 | 32534 | 0.5646 | 0.28617 | *AL121895.1* |
| ENSG00000188582 | 3 | 142658006 | 142692178 | 119 | 24 | 35963 | 0.56447 | 0.28622 | *PAQR9* |
| ENSG00000076201 | 3 | 47412501 | 47464931 | 65 | 11 | 43698 | 0.56417 | 0.28632 | *PTPN23* |
| ENSG00000113749 | 5 | 175075033 | 175123245 | 87 | 20 | 38484 | 0.56418 | 0.28632 | *HRH2* |
| ENSG00000183036 | 21 | 41229243 | 41311322 | 369 | 78 | 51364 | 0.56391 | 0.28641 | *PCP4* |
| ENSG00000140104 | 14 | 105442112 | 105486819 | 174 | 16 | 36754 | 0.56375 | 0.28646 | *C14orf79* |
| ENSG00000124091 | 20 | 55056548 | 55110981 | 137 | 17 | 34917 | 0.56352 | 0.28654 | *GCNT7* |
| ENSG00000088832 | 20 | 1339622 | 1383806 | 154 | 25 | 47242 | 0.56345 | 0.28657 | *FKBP1A* |
| ENSG00000157450 | 15 | 59147374 | 59399618 | 601 | 53 | 43057 | 0.56334 | 0.2866 | *RNF111* |
| ENSG00000082258 | 2 | 135665805 | 135726912 | 107 | 13 | 33616 | 0.5632 | 0.28665 | *CCNT2* |
| ENSG00000113448 | 5 | 58254865 | 59827947 | 4424 | 284 | 44481 | 0.56309 | 0.28669 | *PDE4D* |
| ENSG00000173207 | 1 | 154937129 | 154961725 | 30 | 7 | 38805 | 0.56306 | 0.2867 | *CKS1B* |
| ENSG00000171227 | 2 | 120177477 | 120206096 | 112 | 20 | 47940 | 0.56306 | 0.2867 | *TMEM37* |
| ENSG00000145220 | 4 | 4259428 | 4301896 | 224 | 33 | 38699 | 0.56304 | 0.2867 | *LYAR* |
| ENSG00000068781 | 2 | 48747064 | 49013654 | 1154 | 117 | 47587 | 0.56296 | 0.28673 | *STON1-GTF2A1L* |
| ENSG00000204851 | 19 | 46980163 | 47009755 | 155 | 26 | 48780 | 0.56287 | 0.28676 | *PNMAL2* |
| ENSG00000122852 | 10 | 81360695 | 81385196 | 113 | 19 | 46017 | 0.56213 | 0.28701 | *SFTPA1* |
| ENSG00000119411 | 9 | 116101821 | 116143513 | 265 | 38 | 39655 | 0.56211 | 0.28702 | *BSPRY* |
| ENSG00000117154 | 1 | 18424240 | 18714977 | 1103 | 172 | 47789 | 0.56196 | 0.28707 | *IGSF21* |
| ENSG00000118257 | 2 | 206536714 | 206672857 | 328 | 81 | 45186 | 0.56188 | 0.2871 | *NRP2* |
| ENSG00000171469 | 19 | 9705356 | 9742075 | 89 | 13 | 36098 | 0.56179 | 0.28713 | *ZNF561* |
| ENSG00000161021 | 5 | 179149851 | 179233512 | 247 | 31 | 48664 | 0.56151 | 0.28722 | *MAML1* |
| ENSG00000114331 | 3 | 194985465 | 195173807 | 618 | 27 | 24191 | 0.56077 | 0.28748 | *ACAP2* |
| ENSG00000157873 | 1 | 2477078 | 2506821 | 89 | 12 | 45182 | 0.56064 | 0.28752 | *TNFRSF14* |
| ENSG00000149575 | 11 | 118022666 | 118057388 | 98 | 17 | 40481 | 0.56034 | 0.28762 | *SCN2B* |
| ENSG00000119402 | 9 | 123504256 | 123565690 | 103 | 14 | 52408 | 0.56017 | 0.28768 | *FBXW2* |
| ENSG00000122958 | 10 | 70873268 | 70942617 | 195 | 19 | 31874 | 0.55989 | 0.28778 | *VPS26A* |
| ENSG00000170802 | 2 | 48531776 | 48616433 | 287 | 27 | 49672 | 0.55983 | 0.2878 | *FOXN2* |
| ENSG00000168619 | 8 | 39432008 | 39597583 | 342 | 36 | 43434 | 0.55974 | 0.28783 | *ADAM18* |
| ENSG00000228273 | 7 | 100537257 | 100560424 | 76 | 16 | 41708 | 0.55968 | 0.28785 | *MUC3A* |
| ENSG00000125459 | 1 | 155569979 | 155728153 | 188 | 30 | 41192 | 0.55939 | 0.28795 | *MSTO1* |
| ENSG00000106105 | 7 | 30624297 | 30683649 | 165 | 24 | 38731 | 0.55931 | 0.28797 | *GARS* |
| ENSG00000147596 | 8 | 70953886 | 70993928 | 145 | 23 | 40577 | 0.55934 | 0.28797 | *PRDM14* |
| ENSG00000174748 | 3 | 23948036 | 23975183 | 81 | 14 | 49367 | 0.55924 | 0.288 | *RPL15* |
| ENSG00000112212 | 6 | 41000293 | 41022076 | 65 | 8 | 53932 | 0.55873 | 0.28817 | *TSPO2* |
| ENSG00000109685 | 4 | 1863151 | 1993934 | 124 | 50 | 49382 | 0.55862 | 0.28821 | *WHSC1* |

| | | | | | | | | | |
|---|---|---|---|---|---|---|---|---|---|
| ENSG00000267710 | 19 | 56774138 | 56831819 | 177 | 30 | 37314 | 0.55851 | 0.28825 | *AC006116.20* |
| ENSG00000144893 | 3 | 150793484 | 151164860 | 1160 | 94 | 42053 | 0.55832 | 0.28831 | *MED12L* |
| ENSG00000147642 | 8 | 110576207 | 110714020 | 358 | 36 | 45169 | 0.5582 | 0.28835 | *SYBU* |
| ENSG00000158528 | 7 | 94526514 | 94935727 | 803 | 78 | 44612 | 0.55814 | 0.28837 | *PPP1R9A* |
| ENSG00000186300 | 19 | 2831433 | 2870482 | 168 | 19 | 43056 | 0.55794 | 0.28844 | *ZNF555* |
| ENSG00000180090 | 17 | 3184929 | 3205876 | 107 | 9 | 19836 | 0.5579 | 0.28846 | *OR3A1* |
| ENSG00000167702 | 8 | 145681426 | 145709585 | 54 | 13 | 42515 | 0.55787 | 0.28847 | *KIFC2* |
| ENSG00000132915 | 5 | 149227519 | 149334356 | 302 | 57 | 45461 | 0.55779 | 0.28849 | *PDE6A* |
| ENSG00000134802 | 11 | 57164427 | 57205053 | 84 | 21 | 51677 | 0.55781 | 0.28849 | *SLC43A3* |
| ENSG00000257355 | 19 | 12118703 | 12173782 | 113 | 13 | 32066 | 0.5575 | 0.28859 | *CTD-2006C1.10* |
| ENSG00000101138 | 20 | 54957427 | 54989518 | 89 | 27 | 37682 | 0.55744 | 0.28861 | *CSTF1* |
| ENSG00000172650 | 10 | 75424033 | 75467639 | 35 | 8 | 35147 | 0.55742 | 0.28862 | *AGAP5* |
| ENSG00000243710 | 1 | 43627820 | 43730029 | 407 | 40 | 35751 | 0.55738 | 0.28863 | *WDR65* |
| ENSG00000239305 | 2 | 86820516 | 86860989 | 128 | 20 | 40795 | 0.55723 | 0.28868 | *RNF103* |
| ENSG00000053254 | 14 | 89581215 | 90095493 | 1532 | 228 | 45511 | 0.55711 | 0.28873 | *FOXN3* |
| ENSG00000244754 | 13 | 32996554 | 33122970 | 218 | 27 | 33475 | 0.55676 | 0.28884 | *N4BP2L2* |
| ENSG00000107938 | 10 | 127398084 | 127462712 | 82 | 17 | 45364 | 0.55656 | 0.28891 | *EDRF1* |
| ENSG00000047249 | 8 | 54618117 | 54766118 | 175 | 15 | 29458 | 0.55654 | 0.28892 | *ATP6V1H* |
| ENSG00000174370 | 11 | 128759460 | 128785930 | 92 | 25 | 48619 | 0.55646 | 0.28895 | *C11orf45* |
| ENSG00000167705 | 17 | 1539444 | 1563371 | 61 | 14 | 46359 | 0.55596 | 0.28912 | *RILP* |
| ENSG00000182481 | 17 | 66021635 | 66052958 | 100 | 10 | 31614 | 0.55596 | 0.28912 | *KPNA2* |
| ENSG00000141385 | 18 | 12318943 | 12387313 | 183 | 19 | 38969 | 0.55588 | 0.28915 | *AFG3L2* |
| ENSG00000156150 | 1 | 110592616 | 110623322 | 53 | 6 | 48878 | 0.55585 | 0.28916 | *ALX3* |
| ENSG00000173786 | 17 | 40108759 | 40139749 | 56 | 14 | 45172 | 0.55511 | 0.28941 | *CNP* |
| ENSG00000256053 | 14 | 104019299 | 104083860 | 195 | 18 | 49020 | 0.55448 | 0.28962 | *APOPT1* |
| ENSG00000132693 | 1 | 159672079 | 159694379 | 65 | 14 | 42755 | 0.55422 | 0.28971 | *CRP* |
| ENSG00000134030 | 18 | 46055417 | 46399588 | 1197 | 148 | 43149 | 0.55408 | 0.28976 | *CTIF* |
| ENSG00000271303 | 20 | 617259 | 644014 | 84 | 27 | 41637 | 0.55389 | 0.28983 | *SRXN1* |
| ENSG00000229729 | 3 | 88098424 | 88198123 | 256 | 11 | 30067 | 0.55374 | 0.28988 | *RP11-159G9.5* |
| ENSG00000255423 | 3 | 73100810 | 73122488 | 75 | 10 | 46279 | 0.55368 | 0.2899 | *EBLN2* |
| ENSG00000113108 | 5 | 139927853 | 139983337 | 96 | 15 | 52047 | 0.55365 | 0.28991 | *APBB3* |
| ENSG00000085999 | 1 | 46703360 | 46754145 | 51 | 15 | 33747 | 0.55333 | 0.29002 | *RAD54L* |
| ENSG00000139780 | 13 | 103328094 | 103356854 | 141 | 28 | 49461 | 0.55318 | 0.29007 | *METTL21C* |
| ENSG00000215644 | 17 | 79752008 | 79781889 | 56 | 11 | 52155 | 0.55279 | 0.2902 | *GCGR* |
| ENSG00000132613 | 16 | 70685107 | 70729969 | 153 | 35 | 41974 | 0.55271 | 0.29023 | *MTSS1L* |
| ENSG00000132434 | 7 | 55423141 | 55511435 | 268 | 21 | 52878 | 0.55259 | 0.29027 | *LANCL2* |

| | | | | | | | | | |
|---|---|---|---|---|---|---|---|---|---|
| ENSG00000167011 | 7 | 100803774 | 100833557 | 97 | 18 | 42647 | 0.55255 | 0.29029 | *NAT16* |
| ENSG00000101004 | 20 | 25423341 | 25576153 | 423 | 35 | 37800 | 0.55253 | 0.29029 | *NINL* |
| ENSG00000256061 | 15 | 55692723 | 55810432 | 443 | 43 | 40266 | 0.55247 | 0.29031 | *DYX1C1* |
| ENSG00000148308 | 9 | 135896076 | 135943890 | 127 | 22 | 47329 | 0.55236 | 0.29035 | *GTF3C5* |
| ENSG00000144674 | 3 | 37274668 | 37418242 | 315 | 23 | 41427 | 0.5522 | 0.2904 | *GOLGA4* |
| ENSG00000100351 | 22 | 40287086 | 40379725 | 118 | 30 | 34381 | 0.55216 | 0.29042 | *GRAP2* |
| ENSG00000186980 | 21 | 31710717 | 31730924 | 100 | 15 | 39185 | 0.55207 | 0.29045 | *KRTAP23-1* |
| ENSG00000109971 | 11 | 122918197 | 122943938 | 123 | 16 | 33991 | 0.55202 | 0.29047 | *HSPA8* |
| ENSG00000112290 | 6 | 110411022 | 110511207 | 300 | 21 | 34686 | 0.55184 | 0.29053 | *WASF1* |
| ENSG00000101460 | 20 | 33124658 | 33158149 | 56 | 11 | 46320 | 0.55168 | 0.29058 | *MAP1LC3A* |
| ENSG00000205300 | 20 | 4040799 | 4065812 | 210 | 26 | 34815 | 0.55131 | 0.29071 | *RP11-352D3.2* |
| ENSG00000100614 | 14 | 60702470 | 60775805 | 159 | 15 | 32324 | 0.55115 | 0.29076 | *PPM1A* |
| ENSG00000164091 | 3 | 52278437 | 52332036 | 95 | 14 | 44127 | 0.55113 | 0.29077 | *WDR82* |
| ENSG00000145244 | 4 | 47586015 | 47850123 | 757 | 60 | 49566 | 0.55114 | 0.29077 | *CORIN* |
| ENSG00000119950 | 10 | 111957363 | 112057123 | 286 | 27 | 40675 | 0.55107 | 0.29079 | *MXI1* |
| ENSG00000136383 | 15 | 85349911 | 85426713 | 231 | 40 | 48166 | 0.55097 | 0.29083 | *ALPK3* |
| ENSG00000138380 | 2 | 203766937 | 203861786 | 209 | 14 | 47468 | 0.55088 | 0.29086 | *CARF* |
| ENSG00000111670 | 12 | 102129275 | 102234716 | 268 | 36 | 48286 | 0.55073 | 0.29091 | *GNPTAB* |
| ENSG00000099282 | 10 | 71201229 | 71277425 | 364 | 29 | 52834 | 0.55059 | 0.29096 | *TSPAN15* |
| ENSG00000186998 | 22 | 29591840 | 29665586 | 239 | 42 | 43446 | 0.55052 | 0.29098 | *EMID1* |
| ENSG00000183640 | 21 | 32175016 | 32195570 | 78 | 10 | 47369 | 0.55044 | 0.29101 | *KRTAP8-1* |
| ENSG00000205971 | 7 | 1722446 | 1743961 | 52 | 10 | 51107 | 0.55025 | 0.29107 | *AC074389.6* |
| ENSG00000204610 | 6 | 30120993 | 30150473 | 168 | 20 | 43438 | 0.55018 | 0.2911 | *TRIM15* |
| ENSG00000100997 | 20 | 25265379 | 25381619 | 350 | 17 | 44235 | 0.55006 | 0.29114 | *ABHD12* |
| ENSG00000171552 | 20 | 30242255 | 30321792 | 123 | 13 | 35573 | 0.55006 | 0.29114 | *BCL2L1* |
| ENSG00000134259 | 1 | 115818539 | 115890857 | 262 | 57 | 40758 | 0.55002 | 0.29115 | *NGF* |
| ENSG00000055070 | 1 | 16669070 | 16734640 | 196 | 22 | 46800 | 0.54993 | 0.29118 | *SZRD1* |
| ENSG00000170312 | 10 | 62528089 | 62564610 | 136 | 17 | 54954 | 0.54984 | 0.29121 | *CDK1* |
| ENSG00000182352 | 17 | 72570818 | 72600348 | 126 | 15 | 42478 | 0.54981 | 0.29122 | *C17orf77* |
| ENSG00000169116 | 4 | 75848305 | 75985325 | 258 | 42 | 45121 | 0.54978 | 0.29123 | *PARM1* |
| ENSG00000135164 | 7 | 86771677 | 86835653 | 230 | 16 | 55873 | 0.54979 | 0.29123 | *DMTF1* |
| ENSG00000178338 | 5 | 178276954 | 178325123 | 343 | 16 | 48788 | 0.54978 | 0.29124 | *ZNF354B* |
| ENSG00000149054 | 11 | 6937635 | 7015863 | 325 | 38 | 47816 | 0.54972 | 0.29126 | *ZNF215* |
| ENSG00000127837 | 2 | 219118850 | 219144980 | 52 | 7 | 43845 | 0.54946 | 0.29135 | *AAMP* |
| ENSG00000196660 | 1 | 219848769 | 220141989 | 1106 | 85 | 41648 | 0.54943 | 0.29136 | *SLC30A10* |
| ENSG00000163596 | 2 | 203630690 | 203746708 | 266 | 21 | 49512 | 0.54927 | 0.29141 | *ICA1L* |

| | | | | | | | | | |
|---|---|---|---|---|---|---|---|---|---|
| ENSG00000166289 | 19 | 30145963 | 30176364 | 77 | 16 | 33396 | 0.54904 | 0.29149 | *PLEKHF1* |
| ENSG00000164035 | 4 | 101306498 | 101811283 | 1869 | 118 | 48671 | 0.54881 | 0.29157 | *EMCN* |
| ENSG00000198795 | 18 | 22631890 | 22942154 | 862 | 140 | 46988 | 0.54874 | 0.29159 | *ZNF521* |
| ENSG00000158863 | 8 | 21936670 | 21972409 | 148 | 25 | 50186 | 0.54857 | 0.29165 | *FAM160B2* |
| ENSG00000164970 | 9 | 34388182 | 34468568 | 214 | 29 | 47002 | 0.54836 | 0.29172 | *FAM219A* |
| ENSG00000271092 | 1 | 95573479 | 95722781 | 527 | 59 | 44465 | 0.5483 | 0.29174 | *TMEM56-RWDD3* |
| ENSG00000109066 | 17 | 72762622 | 72845918 | 225 | 28 | 47314 | 0.54829 | 0.29175 | *TMEM104* |
| ENSG00000059378 | 7 | 139713544 | 139773521 | 240 | 37 | 40551 | 0.54819 | 0.29178 | *PARP12* |
| ENSG00000114854 | 3 | 52475118 | 52498086 | 50 | 10 | 51947 | 0.54786 | 0.29189 | *TNNC1* |
| ENSG00000080200 | 3 | 97585819 | 97673810 | 133 | 13 | 45845 | 0.54788 | 0.29189 | *CRYBG3* |
| ENSG00000269074 | 2 | 11593927 | 11616274 | 103 | 11 | 53781 | 0.54775 | 0.29193 | *AC099344.1* |
| ENSG00000100077 | 22 | 25950816 | 26135261 | 501 | 39 | 51627 | 0.54764 | 0.29197 | *ADRBK2* |
| ENSG00000159921 | 9 | 36204438 | 36287053 | 245 | 27 | 45849 | 0.54753 | 0.29201 | *GNE* |
| ENSG00000198933 | 17 | 45761447 | 45799416 | 77 | 14 | 47043 | 0.54745 | 0.29203 | *TBKBP1* |
| ENSG00000104324 | 8 | 97647455 | 98171882 | 920 | 107 | 40896 | 0.54738 | 0.29206 | *CPQ* |
| ENSG00000106025 | 7 | 120417376 | 120508456 | 163 | 20 | 37575 | 0.54735 | 0.29207 | *TSPAN12* |
| ENSG00000157992 | 2 | 27655233 | 27679348 | 33 | 5 | 34577 | 0.54726 | 0.2921 | *KRTCAP3* |
| ENSG00000178734 | 13 | 76435187 | 76467947 | 159 | 16 | 46459 | 0.54723 | 0.29211 | *C13orf45* |
| ENSG00000005381 | 17 | 56337217 | 56368296 | 64 | 14 | 52855 | 0.54715 | 0.29214 | *MPO* |
| ENSG00000243444 | 9 | 112393068 | 112723755 | 1372 | 136 | 49535 | 0.54685 | 0.29224 | *PALM2* |
| ENSG00000064601 | 20 | 44508783 | 44537459 | 69 | 16 | 37699 | 0.54669 | 0.29229 | *CTSA* |
| ENSG00000253787 | 5 | 67475704 | 67504235 | 99 | 25 | 52257 | 0.54647 | 0.29237 | *RP11-404L6.2* |
| ENSG00000163933 | 3 | 53112499 | 53174478 | 164 | 13 | 44178 | 0.54638 | 0.2924 | *RFT1* |
| ENSG00000150401 | 13 | 114100134 | 114155267 | 201 | 23 | 35601 | 0.54638 | 0.2924 | *DCUN1D2* |
| ENSG00000197822 | 5 | 68778119 | 68863931 | 71 | 17 | 47820 | 0.5462 | 0.29246 | *OCLN* |
| ENSG00000104497 | 8 | 82701816 | 82765101 | 300 | 19 | 54050 | 0.54609 | 0.2925 | *SNX16* |
| ENSG00000152455 | 10 | 14910819 | 14956314 | 91 | 24 | 37113 | 0.54608 | 0.29251 | *SUV39H2* |
| ENSG00000155846 | 5 | 149099861 | 149244585 | 502 | 68 | 49966 | 0.54591 | 0.29257 | *PPARGC1B* |
| ENSG00000177409 | 7 | 92749368 | 92787682 | 76 | 21 | 43654 | 0.54558 | 0.29268 | *SAMD9L* |
| ENSG00000143499 | 1 | 214444445 | 214520474 | 266 | 34 | 44289 | 0.54545 | 0.29272 | *SMYD2* |
| ENSG00000251322 | 22 | 51102843 | 51181726 | 211 | 49 | 42740 | 0.54546 | 0.29272 | *SHANK3* |
| ENSG00000134780 | 11 | 61437905 | 61524473 | 194 | 28 | 48081 | 0.54526 | 0.29279 | *DAGLA* |
| ENSG00000149639 | 20 | 35395845 | 35502089 | 168 | 33 | 41525 | 0.5451 | 0.29284 | *SOGA1* |
| ENSG00000092345 | 3 | 16618299 | 16721813 | 531 | 71 | 48715 | 0.5447 | 0.29298 | *DAZL* |
| ENSG00000143257 | 1 | 161189456 | 161218092 | 105 | 18 | 51500 | 0.54467 | 0.29299 | *NR1I3* |
| ENSG00000269529 | 12 | 50680489 | 50701718 | 37 | 5 | 38111 | 0.54442 | 0.29308 | *AC140061.12* |

| | | | | | | | | | |
|---|---|---|---|---|---|---|---|---|---|
| ENSG00000137168 | 6 | 36812603 | 36852800 | 134 | 19 | 54953 | 0.54438 | 0.29309 | *PPIL1* |
| ENSG00000170426 | 12 | 57306938 | 57338189 | 95 | 15 | 40103 | 0.5443 | 0.29312 | *SDR9C7* |
| ENSG00000153531 | 13 | 114066260 | 114117839 | 210 | 49 | 45150 | 0.54375 | 0.29331 | *ADPRHL1* |
| ENSG00000186009 | 13 | 114293119 | 114322501 | 53 | 17 | 41990 | 0.5433 | 0.29346 | *ATP4B* |
| ENSG00000236939 | 8 | 104135191 | 104163703 | 137 | 23 | 56368 | 0.54323 | 0.29348 | *C8orf56* |
| ENSG00000171133 | 9 | 114079763 | 114100800 | 43 | 9 | 49023 | 0.54317 | 0.29351 | *OR2K2* |
| ENSG00000196155 | 16 | 67301413 | 67333402 | 40 | 15 | 45911 | 0.54258 | 0.29371 | *PLEKHG4* |
| ENSG00000183562 | 11 | 3001093 | 3022563 | 87 | 15 | 38568 | 0.54253 | 0.29373 | *AC131971.1* |
| ENSG00000132405 | 4 | 6900969 | 7044845 | 577 | 47 | 51470 | 0.54239 | 0.29378 | *TBC1D14* |
| ENSG00000140675 | 16 | 31484323 | 31512181 | 20 | 9 | 43920 | 0.5422 | 0.29384 | *SLC5A2* |
| ENSG00000150722 | 2 | 182808968 | 183006125 | 491 | 63 | 48541 | 0.54216 | 0.29385 | *PPP1R1C* |
| ENSG00000204764 | 5 | 170278874 | 170737019 | 978 | 39 | 39778 | 0.54199 | 0.29391 | *RANBP17* |
| ENSG00000057294 | 12 | 32933679 | 33059774 | 376 | 57 | 40379 | 0.54159 | 0.29405 | *PKP2* |
| ENSG00000176635 | 22 | 30466163 | 30583064 | 255 | 31 | 40988 | 0.54158 | 0.29405 | *HORMAD2* |
| ENSG00000113456 | 5 | 34895369 | 34929094 | 71 | 15 | 55017 | 0.54158 | 0.29406 | *RAD1* |
| ENSG00000165526 | 11 | 126061993 | 126091587 | 97 | 19 | 42179 | 0.54155 | 0.29406 | *RPUSD4* |
| ENSG00000160439 | 19 | 55540476 | 55592659 | 349 | 50 | 51652 | 0.54138 | 0.29412 | *RDH13* |
| ENSG00000233280 | 3 | 97530884 | 97604132 | 101 | 13 | 32879 | 0.54126 | 0.29416 | *CRYBG3* |
| ENSG00000214357 | 5 | 172058269 | 172128543 | 221 | 42 | 46519 | 0.54125 | 0.29417 | *NEURL1B* |
| ENSG00000151967 | 3 | 159547650 | 159625149 | 127 | 27 | 47094 | 0.54121 | 0.29418 | *SCHIP1* |
| ENSG00000140505 | 15 | 75031185 | 75058543 | 75 | 9 | 25225 | 0.54107 | 0.29423 | *CYP1A2* |
| ENSG00000099338 | 19 | 38816415 | 38871589 | 209 | 26 | 46802 | 0.54097 | 0.29426 | *CATSPERG* |
| ENSG00000157426 | 4 | 57194456 | 57263666 | 301 | 28 | 49063 | 0.54079 | 0.29433 | *AASDH* |
| ENSG00000163564 | 1 | 158890586 | 158956844 | 184 | 32 | 47306 | 0.54056 | 0.29441 | *PYHIN1* |
| ENSG00000107147 | 9 | 138584031 | 138694992 | 478 | 78 | 47753 | 0.54042 | 0.29445 | *KCNT1* |
| ENSG00000221837 | 21 | 46037040 | 46058294 | 99 | 15 | 38727 | 0.54015 | 0.29455 | *KRTAP10-9* |
| ENSG00000170604 | 19 | 46376866 | 46399376 | 85 | 19 | 46744 | 0.54011 | 0.29456 | *IRF2BP1* |
| ENSG00000147874 | 9 | 19043141 | 19113117 | 332 | 29 | 44204 | 0.54002 | 0.29459 | *HAUS6* |
| ENSG00000133065 | 1 | 205748221 | 205792876 | 135 | 23 | 32859 | 0.53994 | 0.29462 | *SLC41A1* |
| ENSG00000168438 | 6 | 110491344 | 110585478 | 296 | 32 | 42162 | 0.53993 | 0.29462 | *CDC40* |
| ENSG00000204410 | 6 | 31697725 | 31742622 | 120 | 28 | 34544 | 0.53984 | 0.29465 | *MSH5* |
| ENSG00000255152 | 6 | 31697797 | 31742628 | 120 | 28 | 34544 | 0.53984 | 0.29465 | *MSH5-SAPCD1* |
| ENSG00000162881 | 2 | 42979642 | 43001401 | 67 | 15 | 37153 | 0.53969 | 0.29471 | *OXER1* |
| ENSG00000223931 | 16 | 32053311 | 32073601 | 4 | 2 | 20358 | 0.53969 | 0.29471 | *AC142381.1* |
| ENSG00000228008 | 3 | 49931278 | 49964370 | 45 | 9 | 39823 | 0.53957 | 0.29475 | *CTD-2330K9.3* |
| ENSG00000162949 | 2 | 30935637 | 31053408 | 462 | 76 | 47705 | 0.53955 | 0.29476 | *CAPN13* |

| | | | | | | | | | |
|---|---|---|---|---|---|---|---|---|---|
| ENSG00000184924 | 2 | 25002603 | 25026251 | 86 | 17 | 47449 | 0.53933 | 0.29483 | *PTRHD1* |
| ENSG00000184232 | 11 | 120071475 | 120111041 | 125 | 24 | 48802 | 0.53929 | 0.29484 | *OAF* |
| ENSG00000124789 | 6 | 17605269 | 17716656 | 325 | 43 | 49348 | 0.53906 | 0.29492 | *NUP153* |
| ENSG00000145075 | 3 | 180310646 | 180598793 | 731 | 57 | 35988 | 0.53895 | 0.29496 | *CCDC39* |
| ENSG00000132383 | 17 | 1722996 | 1813376 | 280 | 33 | 36621 | 0.53858 | 0.29509 | *RPA1* |
| ENSG00000156471 | 8 | 97263943 | 97359223 | 217 | 32 | 39852 | 0.53849 | 0.29512 | *PTDSS1* |
| ENSG00000166743 | 16 | 20624559 | 20720212 | 279 | 27 | 33440 | 0.5385 | 0.29512 | *ACSM1* |
| ENSG00000164112 | 4 | 122670088 | 122696582 | 122 | 16 | 39606 | 0.53842 | 0.29514 | *TMEM155* |
| ENSG00000105246 | 19 | 4219495 | 4247528 | 88 | 17 | 29935 | 0.53811 | 0.29525 | *EBI3* |
| ENSG00000168924 | 4 | 1803206 | 1867974 | 64 | 33 | 42356 | 0.53778 | 0.29537 | *LETM1* |
| ENSG00000122779 | 7 | 138135079 | 138284738 | 332 | 35 | 48400 | 0.53776 | 0.29537 | *TRIM24* |
| ENSG00000125779 | 20 | 3859486 | 3917605 | 169 | 21 | 47672 | 0.53762 | 0.29542 | *PANK2* |
| ENSG00000235478 | 22 | 17592476 | 17622994 | 151 | 28 | 43084 | 0.53752 | 0.29546 | *AC006946.15* |
| ENSG00000197930 | 14 | 53096634 | 53172618 | 177 | 26 | 53138 | 0.53734 | 0.29552 | *ERO1L* |
| ENSG00000160190 | 21 | 43906118 | 44011550 | 452 | 42 | 36133 | 0.53721 | 0.29556 | *SLC37A1* |
| ENSG00000220008 | 19 | 2279774 | 2318156 | 127 | 30 | 34553 | 0.537 | 0.29563 | *LINGO3* |
| ENSG00000114204 | 3 | 167149577 | 167206792 | 203 | 22 | 39016 | 0.53695 | 0.29565 | *SERPINI2* |
| ENSG00000164556 | 7 | 38714946 | 38736637 | 96 | 23 | 42868 | 0.5368 | 0.2957 | *FAM183B* |
| ENSG00000171611 | 6 | 42873727 | 42903573 | 76 | 12 | 38294 | 0.5365 | 0.29581 | *PTCRA* |
| ENSG00000151689 | 2 | 191198196 | 191246391 | 148 | 15 | 35767 | 0.5364 | 0.29584 | *INPP1* |
| ENSG00000204193 | 9 | 113055801 | 113110127 | 238 | 43 | 38785 | 0.53641 | 0.29584 | *TXNDC8* |
| ENSG00000255529 | 15 | 57874231 | 58084960 | 800 | 107 | 46424 | 0.53607 | 0.29595 | *POLR2M* |
| ENSG00000233932 | 15 | 48473736 | 48505953 | 1 | 1 | 12906 | 0.53594 | 0.296 | *CTXN2* |
| ENSG00000172500 | 11 | 65641212 | 65666010 | 60 | 11 | 36509 | 0.5358 | 0.29605 | *FIBP* |
| ENSG00000119280 | 1 | 230962865 | 231015335 | 164 | 27 | 45521 | 0.53575 | 0.29607 | *C1orf198* |
| ENSG00000173208 | 12 | 39933835 | 40023553 | 205 | 32 | 42515 | 0.53491 | 0.29636 | *ABCD2* |
| ENSG00000154263 | 17 | 67133355 | 67250987 | 303 | 27 | 42202 | 0.5349 | 0.29636 | *ABCA10* |
| ENSG00000151365 | 11 | 77764907 | 77789397 | 116 | 13 | 53657 | 0.53471 | 0.29643 | *THRSP* |
| ENSG00000164291 | 5 | 94880778 | 94950768 | 125 | 15 | 26550 | 0.53458 | 0.29647 | *ARSK* |
| ENSG00000105612 | 19 | 12976025 | 13002282 | 54 | 10 | 42706 | 0.53451 | 0.29649 | *DNASE2* |
| ENSG00000205502 | 15 | 62445734 | 62467482 | 53 | 12 | 50915 | 0.5344 | 0.29653 | *C2CD4B* |
| ENSG00000013288 | 4 | 6566902 | 6635089 | 219 | 40 | 45335 | 0.53424 | 0.29659 | *MAN2B2* |
| ENSG00000148180 | 9 | 123960072 | 124105121 | 406 | 42 | 38517 | 0.53397 | 0.29668 | *GSN* |
| ENSG00000183908 | 11 | 56939221 | 56969191 | 96 | 15 | 49675 | 0.53398 | 0.29668 | *LRRC55* |
| ENSG00000163359 | 2 | 238222646 | 238333018 | 324 | 60 | 45291 | 0.53393 | 0.29669 | *COL6A3* |
| ENSG00000169436 | 8 | 139590478 | 139936249 | 1315 | 154 | 42688 | 0.53395 | 0.29669 | *COL22A1* |

| | | | | | | | | | |
|---|---|---|---|---|---|---|---|---|---|
| ENSG00000185982 | 20 | 158494 | 180322 | 113 | 15 | 43394 | 0.5339 | 0.2967 | *DEFB128* |
| ENSG00000188991 | 12 | 16331419 | 16440619 | 495 | 29 | 47467 | 0.53387 | 0.29672 | *SLC15A5* |
| ENSG00000166329 | 17 | 55811842 | 55832673 | 93 | 16 | 53173 | 0.5338 | 0.29674 | *CCDC182* |
| ENSG00000138670 | 4 | 82337547 | 82975397 | 2356 | 165 | 43685 | 0.53378 | 0.29675 | *RASGEF1B* |
| ENSG00000198590 | 3 | 37417760 | 37486988 | 158 | 25 | 43650 | 0.53373 | 0.29676 | *C3orf35* |
| ENSG00000198742 | 7 | 98615061 | 98751723 | 229 | 48 | 50227 | 0.53342 | 0.29687 | *SMURF1* |
| ENSG00000169976 | 6 | 144406018 | 144426754 | 69 | 18 | 42229 | 0.53336 | 0.29689 | *SF3B5* |
| ENSG00000204897 | 17 | 38894273 | 38921584 | 80 | 17 | 48527 | 0.53336 | 0.29689 | *KRT25* |
| ENSG00000187987 | 6 | 28389707 | 28421279 | 119 | 15 | 49441 | 0.53324 | 0.29693 | *ZSCAN23* |
| ENSG00000253831 | 1 | 157051836 | 157079600 | 126 | 19 | 42085 | 0.53318 | 0.29695 | *ETV3L* |
| ENSG00000099290 | 10 | 51817648 | 51903269 | 74 | 9 | 36875 | 0.53226 | 0.29727 | *FAM21A* |
| ENSG00000073584 | 17 | 38771214 | 38814760 | 95 | 17 | 51611 | 0.53224 | 0.29728 | *SMARCE1* |
| ENSG00000267985 | 7 | 72449478 | 72474996 | 5 | 3 | 37143 | 0.53218 | 0.2973 | *AC005488.1* |
| ENSG00000187954 | 8 | 145664965 | 145701060 | 70 | 20 | 45313 | 0.53202 | 0.29736 | *CYHR1* |
| ENSG00000088038 | 19 | 54631444 | 54669419 | 114 | 28 | 44407 | 0.53184 | 0.29742 | *CNOT3* |
| ENSG00000162521 | 1 | 33106743 | 33161812 | 144 | 12 | 37655 | 0.53173 | 0.29746 | *RBBP4* |
| ENSG00000010818 | 6 | 143062604 | 143276338 | 588 | 87 | 50390 | 0.53156 | 0.29752 | *HIVEP2* |
| ENSG00000197445 | 16 | 73150536 | 73188346 | 120 | 21 | 40793 | 0.53153 | 0.29753 | *C16orf47* |
| ENSG00000129484 | 14 | 20801741 | 20836064 | 126 | 15 | 42046 | 0.53142 | 0.29756 | *PARP2* |
| ENSG00000141338 | 17 | 66853433 | 66961533 | 270 | 32 | 44775 | 0.53118 | 0.29765 | *ABCA8* |
| ENSG00000198246 | 10 | 73069015 | 73133142 | 237 | 37 | 38204 | 0.53115 | 0.29766 | *SLC29A3* |
| ENSG00000007545 | 16 | 1652326 | 1737909 | 185 | 22 | 39022 | 0.531 | 0.29771 | *CRAMP1L* |
| ENSG00000116852 | 1 | 200928520 | 201002828 | 204 | 25 | 34552 | 0.53088 | 0.29775 | *KIF21B* |
| ENSG00000186432 | 3 | 160202783 | 160293376 | 196 | 22 | 48916 | 0.53085 | 0.29776 | *KPNA4* |
| ENSG00000121236 | 11 | 5607339 | 5644188 | 210 | 47 | 48338 | 0.53076 | 0.29779 | *TRIM6* |
| ENSG00000109424 | 4 | 141470588 | 141499959 | 93 | 15 | 36773 | 0.53071 | 0.29781 | *UCP1* |
| ENSG00000168916 | 5 | 123962608 | 124094500 | 417 | 70 | 48027 | 0.53068 | 0.29782 | *ZNF608* |
| ENSG00000240505 | 17 | 16822849 | 16885432 | 296 | 40 | 43131 | 0.53004 | 0.29804 | *TNFRSF13B* |
| ENSG00000104365 | 8 | 42118820 | 42199973 | 131 | 19 | 41261 | 0.52931 | 0.29829 | *IKBKB* |
| ENSG00000214686 | 3 | 51802580 | 51823009 | 19 | 7 | 36185 | 0.529 | 0.2984 | *IQCF6* |
| ENSG00000139496 | 13 | 25865662 | 25933938 | 65 | 25 | 43506 | 0.52868 | 0.29851 | *NUPL1* |
| ENSG00000111186 | 12 | 1629057 | 1766409 | 593 | 138 | 43120 | 0.52848 | 0.29858 | *WNT5B* |
| ENSG00000099769 | 16 | 1830414 | 1854972 | 194 | 15 | 31713 | 0.52839 | 0.29861 | *IGFALS* |
| ENSG00000130589 | 20 | 62179439 | 62215592 | 133 | 32 | 43395 | 0.528 | 0.29875 | *HELZ2* |
| ENSG00000236334 | 1 | 143757144 | 143777881 | 2 | 1 | 42709 | 0.52787 | 0.29879 | *PPIAL4G* |
| ENSG00000182870 | 12 | 132670924 | 132915935 | 590 | 99 | 48107 | 0.52784 | 0.29881 | *GALNT9* |

| | | | | | | | | | |
|---|---|---|---|---|---|---|---|---|---|
| ENSG00000177947 | 11 | 186738 | 210261 | 66 | 17 | 42592 | 0.52757 | 0.2989 | *ODF3* |
| ENSG00000112473 | 6 | 33158222 | 33182216 | 56 | 13 | 48567 | 0.52723 | 0.29902 | *SLC39A7* |
| ENSG00000141279 | 17 | 45590308 | 45710642 | 261 | 22 | 47218 | 0.52695 | 0.29911 | *NPEPPS* |
| ENSG00000203778 | 6 | 112398802 | 112433993 | 112 | 6 | 55481 | 0.52642 | 0.2993 | *FAM229B* |
| ENSG00000168930 | 11 | 89520823 | 89551743 | 16 | 7 | 41047 | 0.52638 | 0.29931 | *TRIM49* |
| ENSG00000084754 | 2 | 26403504 | 26477594 | 146 | 8 | 52431 | 0.52617 | 0.29939 | *HADHA* |
| ENSG00000105146 | 19 | 57732377 | 57756916 | 108 | 15 | 53796 | 0.52603 | 0.29943 | *AURKC* |
| ENSG00000074317 | 5 | 176037085 | 176067530 | 48 | 13 | 36013 | 0.52582 | 0.29951 | *SNCB* |
| ENSG00000198677 | 5 | 94789599 | 94900711 | 246 | 14 | 26329 | 0.52515 | 0.29974 | *TTC37* |
| ENSG00000228727 | 6 | 31720576 | 31742628 | 61 | 18 | 34299 | 0.52504 | 0.29978 | *SAPCD1* |
| ENSG00000188425 | 19 | 46406475 | 46428036 | 118 | 21 | 52418 | 0.52502 | 0.29978 | *NANOS2* |
| ENSG00000104881 | 19 | 45872892 | 45919607 | 123 | 26 | 42598 | 0.52491 | 0.29982 | *PPP1R13L* |
| ENSG00000114982 | 2 | 97248907 | 97318524 | 70 | 17 | 27413 | 0.52487 | 0.29984 | *KANSL3* |
| ENSG00000134954 | 11 | 128318656 | 128467453 | 369 | 74 | 46902 | 0.52456 | 0.29994 | *ETS1* |
| ENSG00000117411 | 1 | 44434615 | 44466840 | 55 | 14 | 34727 | 0.52423 | 0.30006 | *B4GALT2* |
| ENSG00000122376 | 10 | 88843918 | 88961225 | 347 | 29 | 42439 | 0.52424 | 0.30006 | *FAM35A* |
| ENSG00000102786 | 13 | 51918213 | 52038400 | 246 | 26 | 51228 | 0.52423 | 0.30006 | *INTS6* |
| ENSG00000196396 | 20 | 49116891 | 49211299 | 258 | 33 | 55039 | 0.52398 | 0.30015 | *PTPN1* |
| ENSG00000148300 | 9 | 136261186 | 136293164 | 152 | 13 | 53099 | 0.52374 | 0.30023 | *REXO4* |
| ENSG00000137133 | 9 | 35802957 | 35825351 | 40 | 12 | 37444 | 0.52347 | 0.30032 | *HINT2* |
| ENSG00000109654 | 4 | 154063494 | 154270472 | 613 | 85 | 44322 | 0.52345 | 0.30033 | *TRIM2* |
| ENSG00000123342 | 12 | 56219217 | 56246750 | 28 | 15 | 45075 | 0.52339 | 0.30035 | *MMP19* |
| ENSG00000176597 | 3 | 182961032 | 183026292 | 184 | 14 | 41791 | 0.52329 | 0.30039 | *B3GNT5* |
| ENSG00000146425 | 6 | 159047506 | 159075771 | 88 | 13 | 52271 | 0.52329 | 0.30039 | *DYNLT1* |
| ENSG00000137054 | 9 | 37475932 | 37513694 | 197 | 14 | 52104 | 0.52329 | 0.30039 | *POLR1E* |
| ENSG00000258223 | 7 | 141941963 | 141967878 | 150 | 11 | 30462 | 0.5229 | 0.30052 | *PRSS58* |
| ENSG00000182698 | 2 | 220182131 | 220207899 | 102 | 12 | 25877 | 0.52285 | 0.30054 | *RESP18* |
| ENSG00000205835 | 3 | 190560666 | 190620218 | 205 | 36 | 40305 | 0.52286 | 0.30054 | *GMNC* |
| ENSG00000171617 | 5 | 73913234 | 73947249 | 91 | 14 | 49002 | 0.52251 | 0.30066 | *ENC1* |
| ENSG00000107105 | 9 | 23680102 | 23836335 | 486 | 76 | 49578 | 0.5225 | 0.30066 | *ELAVL2* |
| ENSG00000159261 | 21 | 37822919 | 37958867 | 576 | 59 | 35951 | 0.52215 | 0.30078 | *CLDN14* |
| ENSG00000241973 | 22 | 21051979 | 21223705 | 560 | 28 | 53527 | 0.52194 | 0.30086 | *PI4KA* |
| ENSG00000120708 | 5 | 135354584 | 135409507 | 142 | 17 | 44386 | 0.52173 | 0.30093 | *TGFBI* |
| ENSG00000186160 | 1 | 47523160 | 47593991 | 229 | 9 | 23310 | 0.52151 | 0.30101 | *CYP4Z1* |
| ENSG00000221818 | 8 | 25689246 | 25912913 | 742 | 112 | 46411 | 0.52115 | 0.30113 | *EBF2* |
| ENSG00000162399 | 1 | 55454606 | 55486556 | 95 | 16 | 39682 | 0.52104 | 0.30117 | *BSND* |

| | | | | | | | | | |
|---|---|---|---|---|---|---|---|---|---|
| ENSG00000167699 | 17 | 650337 | 695581 | 159 | 25 | 50153 | 0.52063 | 0.30131 | *GLOD4* |
| ENSG00000256812 | 16 | 55590584 | 55611599 | 51 | 8 | 37793 | 0.52049 | 0.30136 | *CAPNS2* |
| ENSG00000113360 | 5 | 31390604 | 31542303 | 438 | 58 | 38984 | 0.52046 | 0.30137 | *DROSHA* |
| ENSG00000100889 | 14 | 24553262 | 24589807 | 64 | 19 | 38931 | 0.52046 | 0.30137 | *PCK2* |
| ENSG00000170099 | 14 | 94760585 | 94799731 | 149 | 26 | 42123 | 0.52007 | 0.30151 | *SERPINA6* |
| ENSG00000269103 | 22 | 21348030 | 21370736 | 117 | 20 | 43435 | 0.52005 | 0.30151 | *AC002472.1* |
| ENSG00000196993 | 16 | 28753108 | 28794128 | 3 | 2 | 43308 | 0.51994 | 0.30155 | *NPIPB9* |
| ENSG00000268798 | 19 | 1430838 | 1451937 | 47 | 12 | 51580 | 0.51991 | 0.30156 | *AC027307.3* |
| ENSG00000109101 | 17 | 26823261 | 26875914 | 113 | 22 | 47091 | 0.51989 | 0.30157 | *FOXN1* |
| ENSG00000167799 | 11 | 67385409 | 67407401 | 105 | 14 | 34196 | 0.5198 | 0.3016 | *NUDT8* |
| ENSG00000088881 | 20 | 2663524 | 2750753 | 219 | 41 | 41154 | 0.51939 | 0.30174 | *EBF4* |
| ENSG00000011260 | 17 | 49327889 | 49385297 | 167 | 21 | 42518 | 0.51934 | 0.30176 | *UTP18* |
| ENSG00000134709 | 1 | 60270458 | 60352050 | 189 | 22 | 43168 | 0.51933 | 0.30177 | *HOOK1* |
| ENSG00000163541 | 2 | 84640647 | 84697169 | 150 | 15 | 31001 | 0.51927 | 0.30179 | *SUCLG1* |
| ENSG00000177679 | 7 | 75821216 | 75926605 | 192 | 30 | 43231 | 0.51906 | 0.30186 | *SRRM3* |
| ENSG00000072657 | 12 | 72471046 | 73069422 | 1414 | 120 | 38215 | 0.51896 | 0.30189 | *TRHDE* |
| ENSG00000100242 | 22 | 39120730 | 39200148 | 163 | 34 | 42178 | 0.51879 | 0.30195 | *SUN2* |
| ENSG00000163795 | 2 | 27590098 | 27613657 | 40 | 9 | 37148 | 0.51877 | 0.30196 | *ZNF513* |
| ENSG00000171954 | 19 | 15609304 | 15673128 | 240 | 50 | 49071 | 0.51872 | 0.30198 | *CYP4F22* |
| ENSG00000068654 | 2 | 86237339 | 86343278 | 248 | 27 | 33772 | 0.51864 | 0.30201 | *POLR1A* |
| ENSG00000182742 | 17 | 46642875 | 46667473 | 67 | 16 | 40217 | 0.51835 | 0.30211 | *HOXB4* |
| ENSG00000131747 | 17 | 38534768 | 38584202 | 101 | 16 | 54822 | 0.5183 | 0.30212 | *TOP2A* |
| ENSG00000140367 | 15 | 76125622 | 76203419 | 171 | 24 | 39618 | 0.51829 | 0.30213 | *UBE2Q2* |
| ENSG00000235645 | 10 | 71434655 | 71462208 | 144 | 19 | 54281 | 0.51824 | 0.30214 | *RP11-242G20.1* |
| ENSG00000261086 | 15 | 83199620 | 83241124 | 67 | 14 | 46044 | 0.51819 | 0.30216 | *RP11-379H8.1* |
| ENSG00000100055 | 22 | 37668068 | 37721382 | 168 | 34 | 43116 | 0.51777 | 0.30231 | *CYTH4* |
| ENSG00000118420 | 6 | 83592117 | 83785560 | 580 | 46 | 49504 | 0.51771 | 0.30233 | *UBE3D* |
| ENSG00000205857 | 12 | 7907812 | 7936717 | 47 | 19 | 49365 | 0.51749 | 0.30241 | *NANOGNB* |
| ENSG00000130529 | 19 | 49650998 | 49725093 | 250 | 37 | 48062 | 0.51746 | 0.30242 | *TRPM4* |
| ENSG00000164087 | 3 | 52099269 | 52198706 | 137 | 20 | 35106 | 0.51736 | 0.30245 | *POC1A* |
| ENSG00000095209 | 9 | 108446825 | 108548893 | 364 | 16 | 22734 | 0.51687 | 0.30262 | *TMEM38B* |
| ENSG00000164002 | 1 | 40964413 | 40992228 | 110 | 16 | 41601 | 0.51685 | 0.30263 | *EXO5* |
| ENSG00000153266 | 3 | 62345356 | 62369999 | 40 | 11 | 48717 | 0.51674 | 0.30267 | *FEZF2* |
| ENSG00000127507 | 19 | 14833205 | 14899353 | 372 | 45 | 44012 | 0.51667 | 0.30269 | *EMR2* |
| ENSG00000132466 | 4 | 73929093 | 74134515 | 204 | 36 | 48215 | 0.51657 | 0.30273 | *ANKRD17* |
| ENSG00000144730 | 3 | 57114010 | 57214334 | 271 | 33 | 42738 | 0.51636 | 0.3028 | *IL17RD* |

| | | | | | | | | | |
|---|---|---|---|---|---|---|---|---|---|
| ENSG00000008324 | 3 | 42613332 | 42646606 | 91 | 13 | 35861 | 0.5162 | 0.30286 | *SS18L2* |
| ENSG00000165030 | 9 | 94161327 | 94196144 | 101 | 23 | 43625 | 0.51616 | 0.30287 | *NFIL3* |
| ENSG00000198674 | 11 | 123854870 | 123875868 | 85 | 15 | 49745 | 0.51613 | 0.30288 | *OR10G6* |
| ENSG00000196141 | 2 | 201160604 | 201356986 | 507 | 66 | 42100 | 0.51595 | 0.30294 | *SPATS2L* |
| ENSG00000146072 | 6 | 47189268 | 47287641 | 229 | 54 | 45135 | 0.51592 | 0.30296 | *TNFRSF21* |
| ENSG00000119632 | 14 | 94584116 | 94606590 | 63 | 10 | 31097 | 0.51592 | 0.30296 | *IFI27L2* |
| ENSG00000074935 | 6 | 112381980 | 112418732 | 79 | 7 | 55877 | 0.51586 | 0.30297 | *TUBE1* |
| ENSG00000149633 | 20 | 36828890 | 36899174 | 197 | 33 | 45914 | 0.51579 | 0.303 | *KIAA1755* |
| ENSG00000143947 | 2 | 55449039 | 55472989 | 81 | 18 | 41607 | 0.5157 | 0.30303 | *RPS27A* |
| ENSG00000256861 | 12 | 122682326 | 122760970 | 165 | 21 | 39950 | 0.51566 | 0.30305 | *RP11-512M8.5* |
| ENSG00000153317 | 8 | 131054353 | 131465906 | 1481 | 71 | 55063 | 0.51543 | 0.30313 | *ASAP1* |
| ENSG00000115541 | 2 | 198354718 | 198378181 | 33 | 7 | 55126 | 0.51538 | 0.30314 | *HSPE1* |
| ENSG00000163463 | 1 | 155131884 | 155155951 | 44 | 9 | 47094 | 0.5153 | 0.30317 | *KRTCAP2* |
| ENSG00000260861 | 20 | 1510790 | 1610655 | 223 | 32 | 46489 | 0.51509 | 0.30325 | *RP4-576H24.4* |
| ENSG00000160282 | 21 | 47546176 | 47585481 | 237 | 51 | 42591 | 0.51499 | 0.30328 | *FTCD* |
| ENSG00000197566 | 17 | 16514051 | 16567170 | 189 | 14 | 46402 | 0.51425 | 0.30354 | *ZNF624* |
| ENSG00000132300 | 2 | 86323305 | 86379280 | 151 | 18 | 31373 | 0.51422 | 0.30355 | *PTCD3* |
| ENSG00000174705 | 5 | 171742185 | 171891527 | 584 | 53 | 36705 | 0.51398 | 0.30363 | *SH3PXD2B* |
| ENSG00000159147 | 21 | 34921848 | 34971014 | 60 | 12 | 51311 | 0.51393 | 0.30365 | *DONSON* |
| ENSG00000106031 | 7 | 27223122 | 27249725 | 58 | 15 | 43191 | 0.5137 | 0.30373 | *HOXA13* |
| ENSG00000258053 | 12 | 71431182 | 71522027 | 291 | 22 | 35560 | 0.51367 | 0.30374 | *CTD-2021H9.3* |
| ENSG00000185666 | 22 | 32898539 | 33464358 | 1923 | 205 | 45490 | 0.51351 | 0.3038 | *SYN3* |
| ENSG00000165238 | 9 | 95937198 | 96092854 | 493 | 60 | 40642 | 0.51322 | 0.3039 | *WNK2* |
| ENSG00000119682 | 14 | 75110140 | 75189818 | 108 | 11 | 40768 | 0.51314 | 0.30393 | *AREL1* |
| ENSG00000099812 | 19 | 741126 | 774319 | 100 | 40 | 42994 | 0.51311 | 0.30394 | *MISP* |
| ENSG00000196683 | 7 | 22842251 | 22872470 | 161 | 19 | 41143 | 0.51294 | 0.304 | *TOMM7* |
| ENSG00000262484 | 19 | 39389620 | 39412798 | 71 | 15 | 42824 | 0.51288 | 0.30402 | *CCER2* |
| ENSG00000122482 | 1 | 91370859 | 91497829 | 246 | 21 | 55526 | 0.51267 | 0.30409 | *ZNF644* |
| ENSG00000164123 | 4 | 159804286 | 159969912 | 525 | 43 | 51603 | 0.51234 | 0.3042 | *C4orf45* |
| ENSG00000162068 | 16 | 2511500 | 2534146 | 20 | 13 | 46606 | 0.51236 | 0.3042 | *NTN3* |
| ENSG00000258256 | 8 | 86124675 | 86167946 | 76 | 10 | 53780 | 0.51227 | 0.30423 | *RP11-219B4.5* |
| ENSG00000251246 | 1 | 155026224 | 155069283 | 78 | 18 | 45910 | 0.51223 | 0.30424 | *EFNA3* |
| ENSG00000143590 | 1 | 155026224 | 155070014 | 78 | 18 | 45910 | 0.51223 | 0.30424 | *EFNA3* |
| ENSG00000188089 | 15 | 42263780 | 42353388 | 416 | 56 | 46621 | 0.51214 | 0.30428 | *PLA2G4E* |
| ENSG00000101493 | 18 | 74059644 | 74217146 | 564 | 88 | 44431 | 0.51207 | 0.3043 | *ZNF516* |
| ENSG00000124067 | 16 | 67967377 | 68013504 | 62 | 13 | 35340 | 0.51205 | 0.30431 | *SLC12A4* |

| | | | | | | | | | |
|---|---|---|---|---|---|---|---|---|---|
| ENSG00000120875 | 8 | 29180581 | 29218185 | 103 | 18 | 34475 | 0.512 | 0.30433 | *DUSP4* |
| ENSG00000168237 | 3 | 52311105 | 52339272 | 52 | 11 | 45017 | 0.51187 | 0.30437 | *GLYCTK* |
| ENSG00000196867 | 19 | 57040317 | 57078169 | 161 | 21 | 45504 | 0.51178 | 0.3044 | *ZFP28* |
| ENSG00000119042 | 2 | 200124223 | 200345989 | 318 | 39 | 33209 | 0.5117 | 0.30443 | *SATB2* |
| ENSG00000180611 | 3 | 192504604 | 192645950 | 489 | 57 | 42840 | 0.51166 | 0.30444 | *MB21D2* |
| ENSG00000157077 | 1 | 52598046 | 52822358 | 282 | 39 | 45803 | 0.51164 | 0.30445 | *ZFYVE9* |
| ENSG00000167970 | 16 | 2293738 | 2322343 | 59 | 11 | 46511 | 0.51148 | 0.30451 | *AC009065.1* |
| ENSG00000167447 | 17 | 57276761 | 57302608 | 59 | 10 | 37942 | 0.51143 | 0.30452 | *SMG8* |
| ENSG00000254470 | 11 | 65533364 | 65558273 | 58 | 11 | 37677 | 0.51124 | 0.30459 | *AP5B1* |
| ENSG00000178882 | 12 | 124447788 | 124810570 | 1220 | 150 | 45667 | 0.51111 | 0.30464 | *FAM101A* |
| ENSG00000215527 | 18 | 12648042 | 12673470 | 91 | 12 | 40932 | 0.511 | 0.30467 | *AP005482.1* |
| ENSG00000118600 | 12 | 64163583 | 64213338 | 89 | 23 | 53607 | 0.511 | 0.30468 | *TMEM5* |
| ENSG00000198951 | 22 | 42444358 | 42476846 | 65 | 10 | 49530 | 0.5109 | 0.30471 | *NAGA* |
| ENSG00000220702 | 22 | 44751431 | 44772164 | 132 | 33 | 50348 | 0.51087 | 0.30472 | *RP1-32I10.10* |
| ENSG00000254469 | 11 | 71566555 | 71649700 | 384 | 18 | 37401 | 0.51073 | 0.30477 | *RP11-849H4.2* |
| ENSG00000033011 | 16 | 5073703 | 5147380 | 275 | 59 | 49645 | 0.51058 | 0.30482 | *ALG1* |
| ENSG00000172197 | 6 | 20090935 | 20222670 | 415 | 83 | 44625 | 0.51054 | 0.30484 | *MBOAT1* |
| ENSG00000156049 | 9 | 80027995 | 80273223 | 825 | 148 | 44733 | 0.51054 | 0.30484 | *GNA14* |
| ENSG00000197696 | 15 | 85188360 | 85211794 | 69 | 15 | 41170 | 0.51047 | 0.30486 | *NMB* |
| ENSG00000142687 | 1 | 35889091 | 36033551 | 106 | 43 | 48429 | 0.51036 | 0.3049 | *KIAA0319L* |
| ENSG00000054267 | 1 | 235284949 | 235501534 | 594 | 27 | 43119 | 0.5103 | 0.30492 | *ARID4B* |
| ENSG00000133265 | 19 | 55763599 | 55801749 | 99 | 20 | 52869 | 0.51029 | 0.30492 | *HSPBP1* |
| ENSG00000088247 | 19 | 6403359 | 6434805 | 111 | 16 | 49715 | 0.51024 | 0.30494 | *KHSRP* |
| ENSG00000110944 | 12 | 56722663 | 56744193 | 40 | 7 | 45411 | 0.51004 | 0.30501 | *IL23A* |
| ENSG00000177169 | 12 | 132369196 | 132417712 | 117 | 16 | 29156 | 0.51004 | 0.30501 | *ULK1* |
| ENSG00000108511 | 17 | 46661639 | 46692354 | 82 | 19 | 44281 | 0.51002 | 0.30502 | *HOXB6* |
| ENSG00000110076 | 11 | 64363646 | 64500660 | 218 | 25 | 36731 | 0.50946 | 0.30521 | *NRXN2* |
| ENSG00000165118 | 9 | 86543226 | 86581901 | 82 | 15 | 50560 | 0.50907 | 0.30535 | *C9orf64* |
| ENSG00000150275 | 10 | 55552531 | 57397702 | 7132 | 330 | 41492 | 0.50898 | 0.30538 | *PCDH15* |
| ENSG00000170323 | 8 | 82380654 | 82405498 | 78 | 13 | 44656 | 0.5089 | 0.30541 | *FABP4* |
| ENSG00000169718 | 17 | 80005382 | 80033763 | 76 | 18 | 38080 | 0.50869 | 0.30549 | *DUS1L* |
| ENSG00000167791 | 11 | 67276383 | 67300899 | 36 | 14 | 41817 | 0.50865 | 0.3055 | *CABP2* |
| ENSG00000170348 | 14 | 75588173 | 75653334 | 154 | 13 | 47824 | 0.50858 | 0.30552 | *TMED10* |
| ENSG00000140678 | 16 | 31356455 | 31404318 | 96 | 24 | 43675 | 0.50846 | 0.30557 | *ITGAX* |
| ENSG00000271723 | 1 | 55097459 | 55217981 | 499 | 49 | 41453 | 0.50805 | 0.30571 | *MROH7-TTC4* |
| ENSG00000101307 | 20 | 1534167 | 1610707 | 145 | 26 | 47768 | 0.50788 | 0.30577 | *SIRPB1* |

| | | | | | | | | | |
|---|---|---|---|---|---|---|---|---|---|
| ENSG00000159723 | 16 | 67506474 | 67527716 | 27 | 9 | 41706 | 0.50778 | 0.3058 | *AGRP* |
| ENSG00000100908 | 14 | 24598174 | 24620797 | 43 | 13 | 37680 | 0.50773 | 0.30582 | *EMC9* |
| ENSG00000162688 | 1 | 100305640 | 100399579 | 380 | 40 | 49995 | 0.50764 | 0.30585 | *AGL* |
| ENSG00000116652 | 1 | 64004588 | 64025405 | 55 | 12 | 50270 | 0.50761 | 0.30586 | *DLEU2L* |
| ENSG00000169955 | 16 | 30527244 | 30556668 | 33 | 15 | 39186 | 0.50745 | 0.30592 | *ZNF747* |
| ENSG00000115365 | 2 | 211285973 | 211352376 | 186 | 18 | 48047 | 0.50731 | 0.30597 | *LANCL1* |
| ENSG00000187210 | 9 | 79024752 | 79132332 | 468 | 55 | 48775 | 0.5071 | 0.30604 | *GCNT1* |
| ENSG00000093144 | 6 | 127599855 | 127674754 | 153 | 16 | 49241 | 0.50705 | 0.30606 | *ECHDC1* |
| ENSG00000269367 | 12 | 42614050 | 42638075 | 58 | 11 | 47020 | 0.50689 | 0.30612 | *AC020629.1* |
| ENSG00000184276 | 11 | 71534246 | 71558756 | 107 | 12 | 44299 | 0.50667 | 0.30619 | *DEFB108B* |
| ENSG00000134463 | 10 | 11774365 | 11816069 | 129 | 16 | 48750 | 0.50653 | 0.30624 | *ECHDC3* |
| ENSG00000088387 | 13 | 99435741 | 99748879 | 1184 | 64 | 46625 | 0.50655 | 0.30624 | *DOCK9* |
| ENSG00000248167 | 6 | 30287359 | 30324631 | 139 | 27 | 39683 | 0.50645 | 0.30627 | *TRIM39-RPP21* |
| ENSG00000132128 | 1 | 46716868 | 46779280 | 78 | 20 | 35860 | 0.50607 | 0.30641 | *LRRC41* |
| ENSG00000108846 | 17 | 48702138 | 48779613 | 191 | 54 | 45922 | 0.50599 | 0.30643 | *ABCC3* |
| ENSG00000125875 | 20 | 406124 | 453197 | 129 | 24 | 46133 | 0.50601 | 0.30643 | *TBC1D20* |
| ENSG00000214841 | 7 | 34675709 | 34709772 | 203 | 27 | 38772 | 0.50574 | 0.30652 | *AC005493.1* |
| ENSG0000087448 | 12 | 27922953 | 27965973 | 167 | 24 | 34871 | 0.50575 | 0.30652 | *KLHL42* |
| ENSG00000113916 | 3 | 187429165 | 187473515 | 145 | 32 | 43965 | 0.50558 | 0.30658 | *BCL6* |
| ENSG00000213160 | 2 | 170540998 | 170643499 | 558 | 59 | 52699 | 0.5055 | 0.3066 | *KLHL23* |
| ENSG00000142632 | 1 | 16514349 | 16549104 | 146 | 10 | 49224 | 0.50517 | 0.30672 | *ARHGEF19* |
| ENSG00000009335 | 7 | 156921607 | 157072066 | 550 | 40 | 43463 | 0.50516 | 0.30672 | *UBE3C* |
| ENSG00000114166 | 3 | 20071515 | 20205896 | 577 | 66 | 46098 | 0.50504 | 0.30677 | *KAT2B* |
| ENSG00000261190 | 16 | 52029399 | 52122386 | 258 | 41 | 51971 | 0.50496 | 0.30679 | *C16orf97* |
| ENSG00000204514 | 19 | 58350099 | 58410442 | 187 | 26 | 37903 | 0.50488 | 0.30682 | *ZNF814* |
| ENSG00000156194 | 4 | 76771020 | 76833724 | 160 | 23 | 36487 | 0.50478 | 0.30686 | *PPEF2* |
| ENSG00000166598 | 12 | 104313885 | 104357423 | 231 | 36 | 42184 | 0.50469 | 0.30689 | *HSP90B1* |
| ENSG00000032219 | 14 | 58755103 | 58850605 | 169 | 19 | 52203 | 0.50465 | 0.3069 | *ARID4A* |
| ENSG00000122674 | 7 | 5928356 | 5975605 | 118 | 30 | 40593 | 0.50453 | 0.30694 | *CCZ1* |
| ENSG00000154359 | 8 | 12569403 | 12623582 | 177 | 24 | 44892 | 0.50453 | 0.30694 | *LONRF1* |
| ENSG00000117395 | 1 | 43619846 | 43746607 | 492 | 48 | 36370 | 0.50443 | 0.30698 | *EBNA1BP2* |
| ENSG00000167608 | 19 | 54653846 | 54686944 | 97 | 28 | 44263 | 0.50422 | 0.30705 | *TMC4* |
| ENSG00000138468 | 3 | 101033049 | 101242085 | 670 | 22 | 53779 | 0.50413 | 0.30708 | *SENP7* |
| ENSG00000171595 | 17 | 72260386 | 72321023 | 245 | 36 | 38903 | 0.50404 | 0.30712 | *DNAI2* |
| ENSG00000106410 | 7 | 144084333 | 144117320 | 104 | 11 | 40090 | 0.5039 | 0.30716 | *NOBOX* |
| ENSG00000112062 | 6 | 35985488 | 36089013 | 286 | 26 | 42756 | 0.50386 | 0.30718 | *MAPK14* |

| | | | | | | | | | |
|---|---|---|---|---|---|---|---|---|---|
| ENSG00000249751 | 5 | 138774245 | 138852328 | 30 | 11 | 40528 | 0.50376 | 0.30722 | *ECSCR* |
| ENSG00000178732 | 3 | 194105550 | 194129995 | 96 | 12 | 26583 | 0.50273 | 0.30758 | *GP5* |
| ENSG00000134160 | 15 | 31283264 | 31463476 | 679 | 105 | 46623 | 0.50235 | 0.30771 | *TRPM1* |
| ENSG00000159069 | 9 | 139824887 | 139849148 | 108 | 17 | 49481 | 0.50193 | 0.30786 | *FBXW5* |
| ENSG00000163982 | 4 | 4180530 | 4238616 | 302 | 38 | 44113 | 0.50166 | 0.30795 | *OTOP1* |
| ENSG00000078725 | 9 | 121905736 | 122141745 | 579 | 85 | 44334 | 0.50168 | 0.30795 | *BRINP1* |
| ENSG00000178363 | 10 | 5556924 | 5578225 | 71 | 23 | 51570 | 0.50166 | 0.30795 | *CALML3* |
| ENSG00000136999 | 8 | 120418546 | 120446593 | 143 | 10 | 45539 | 0.50113 | 0.30814 | *NOV* |
| ENSG00000104472 | 8 | 141511397 | 141537236 | 73 | 20 | 51942 | 0.50088 | 0.30823 | *CHRAC1* |
| ENSG00000230657 | 12 | 11450017 | 11473369 | 71 | 17 | 46227 | 0.50081 | 0.30825 | *PRB4* |
| ENSG00000077800 | 7 | 72732167 | 72782634 | 113 | 12 | 51567 | 0.50077 | 0.30827 | *FKBP6* |
| ENSG00000170162 | 6 | 117576721 | 117604728 | 78 | 16 | 34089 | 0.50062 | 0.30832 | *VGLL2* |
| ENSG00000096401 | 6 | 44345262 | 44428163 | 172 | 39 | 40525 | 0.50056 | 0.30834 | *CDC5L* |
| ENSG00000126243 | 19 | 36416260 | 36446669 | 85 | 14 | 33672 | 0.50056 | 0.30834 | *LRFN3* |
| ENSG00000105251 | 19 | 4268598 | 4300721 | 98 | 25 | 45658 | 0.50052 | 0.30835 | *SHD* |
| ENSG00000143179 | 1 | 165786768 | 165890855 | 207 | 36 | 44036 | 0.50012 | 0.3085 | *UCK2* |
| ENSG00000198298 | 10 | 44091855 | 44123351 | 129 | 25 | 48874 | 0.50012 | 0.3085 | *ZNF485* |
| ENSG00000132612 | 16 | 69335259 | 69368949 | 92 | 17 | 53697 | 0.49998 | 0.30854 | *VPS4A* |
| ENSG00000131374 | 3 | 17188654 | 18496309 | 3280 | 115 | 41953 | 0.49977 | 0.30862 | *TBC1D5* |
| ENSG00000162517 | 1 | 32085463 | 32120497 | 72 | 8 | 32080 | 0.49972 | 0.30864 | *PEF1* |
| ENSG00000203872 | 6 | 88044567 | 88085181 | 138 | 15 | 51487 | 0.49928 | 0.30879 | *C6orf163* |
| ENSG00000196923 | 5 | 176900395 | 176934607 | 44 | 20 | 35586 | 0.49926 | 0.3088 | *PDLIM7* |
| ENSG00000198162 | 1 | 117900071 | 118081494 | 581 | 41 | 53986 | 0.49912 | 0.30885 | *MAN1A2* |
| ENSG00000233438 | 2 | 27750253 | 27800011 | 117 | 15 | 51929 | 0.49906 | 0.30887 | *AC109829.1* |
| ENSG00000008438 | 19 | 46512411 | 46536323 | 79 | 13 | 55348 | 0.49853 | 0.30906 | *PGLYRP1* |
| ENSG00000204950 | 11 | 61266272 | 61288482 | 62 | 11 | 28452 | 0.49828 | 0.30914 | *LRRC10B* |
| ENSG00000197851 | 7 | 96100938 | 96142835 | 108 | 22 | 48495 | 0.49822 | 0.30916 | *C7orf76* |
| ENSG00000266202 | 17 | 26115832 | 26230391 | 236 | 28 | 41828 | 0.49766 | 0.30936 | *RP1-66C13.4* |
| ENSG00000250038 | 4 | 28353901 | 28414486 | 269 | 23 | 48264 | 0.49742 | 0.30945 | *RP11-180C1.1* |
| ENSG00000110660 | 11 | 107651717 | 107809019 | 725 | 68 | 43529 | 0.49721 | 0.30952 | *SLC35F2* |
| ENSG00000272297 | 4 | 187337700 | 187486464 | 786 | 112 | 42563 | 0.49718 | 0.30953 | *RP11-215A19.2* |
| ENSG00000160948 | 8 | 145639000 | 145663931 | 75 | 9 | 49969 | 0.49719 | 0.30953 | *VPS28* |
| ENSG00000159788 | 4 | 3284755 | 3451640 | 469 | 76 | 42380 | 0.49703 | 0.30958 | *RGS12* |
| ENSG00000171819 | 1 | 11239398 | 11266038 | 49 | 6 | 26000 | 0.49692 | 0.30962 | *ANGPTL7* |
| ENSG00000168067 | 11 | 64546290 | 64580713 | 35 | 9 | 43468 | 0.49686 | 0.30964 | *MAP4K2* |
| ENSG00000038002 | 4 | 178341924 | 178373657 | 92 | 23 | 47045 | 0.49668 | 0.30971 | *AGA* |

| | | | | | | | | | |
|---|---|---|---|---|---|---|---|---|---|
| ENSG00000117298 | 1 | 21533740 | 21681997 | 454 | 71 | 43383 | 0.49661 | 0.30973 | *ECE1* |
| ENSG00000217825 | 7 | 154978273 | 155000165 | 165 | 19 | 36875 | 0.4966 | 0.30974 | *AC099552.4* |
| ENSG00000136541 | 2 | 158165137 | 158194225 | 41 | 10 | 31015 | 0.4963 | 0.30984 | *ERMN* |
| ENSG00000082556 | 8 | 54128284 | 54174257 | 235 | 25 | 42252 | 0.4963 | 0.30984 | *OPRK1* |
| ENSG00000110057 | 11 | 67748575 | 67782452 | 78 | 17 | 51107 | 0.49624 | 0.30986 | *UNC93B1* |
| ENSG00000109943 | 11 | 122699208 | 122753347 | 200 | 39 | 46961 | 0.49625 | 0.30986 | *CRTAM* |
| ENSG00000168903 | 5 | 180405845 | 180443727 | 55 | 11 | 36251 | 0.49614 | 0.3099 | *BTNL3* |
| ENSG00000153982 | 17 | 57287828 | 57363328 | 156 | 15 | 33981 | 0.49612 | 0.3099 | *GDPD1* |
| ENSG00000215612 | 4 | 8837802 | 8883543 | 31 | 18 | 29761 | 0.49598 | 0.30995 | *HMX1* |
| ENSG00000169016 | 2 | 11574501 | 11616297 | 199 | 14 | 52896 | 0.49596 | 0.30996 | *E2F6* |
| ENSG00000245888 | 16 | 27269526 | 27311813 | 88 | 16 | 43654 | 0.49576 | 0.31003 | *CTD-3203P2.2* |
| ENSG00000108819 | 17 | 48201104 | 48237991 | 60 | 15 | 48624 | 0.49575 | 0.31004 | *PPP1R9B* |
| ENSG00000130595 | 11 | 1930792 | 1969936 | 117 | 32 | 45689 | 0.49569 | 0.31006 | *TNNT3* |
| ENSG00000148606 | 10 | 79724907 | 79799303 | 137 | 22 | 39475 | 0.49559 | 0.31009 | *POLR3A* |
| ENSG00000105202 | 19 | 40315098 | 40347054 | 112 | 20 | 55498 | 0.4954 | 0.31016 | *FBL* |
| ENSG00000113460 | 5 | 34905481 | 34936101 | 58 | 12 | 53898 | 0.49525 | 0.31021 | *BRIX1* |
| ENSG00000149716 | 11 | 69457844 | 69500184 | 84 | 18 | 35883 | 0.49513 | 0.31025 | *ORAOV1* |
| ENSG00000268265 | 21 | 34937431 | 34957645 | 24 | 7 | 53026 | 0.4945 | 0.31048 | *AP000304.1* |
| ENSG00000121691 | 11 | 34450472 | 34503609 | 173 | 22 | 37169 | 0.49437 | 0.31052 | *CAT* |
| ENSG00000126785 | 14 | 63660832 | 63769937 | 312 | 40 | 41601 | 0.49423 | 0.31057 | *RHOJ* |
| ENSG00000243509 | 20 | 62318021 | 62340037 | 65 | 11 | 36600 | 0.49412 | 0.31061 | *TNFRSF6B* |
| ENSG00000108244 | 17 | 39068948 | 39103886 | 184 | 28 | 36685 | 0.49411 | 0.31062 | *KRT23* |
| ENSG00000198805 | 14 | 20927113 | 20955253 | 88 | 18 | 41535 | 0.49402 | 0.31065 | *PNP* |
| ENSG00000173801 | 17 | 39765692 | 39953183 | 592 | 71 | 42495 | 0.49359 | 0.3108 | *JUP* |
| ENSG00000205863 | 13 | 24455238 | 24486794 | 123 | 18 | 47096 | 0.49349 | 0.31083 | *C1QTNF9B* |
| ENSG00000142920 | 1 | 33536705 | 33596131 | 101 | 14 | 48561 | 0.49345 | 0.31085 | *ADC* |
| ENSG00000196090 | 20 | 40691392 | 41828610 | 3744 | 332 | 43273 | 0.49344 | 0.31085 | *PTPRT* |
| ENSG00000244255 | 6 | 31885475 | 31929825 | 132 | 24 | 45429 | 0.49331 | 0.3109 | *CFB* |
| ENSG00000243649 | 6 | 31885475 | 31929861 | 132 | 24 | 45429 | 0.49331 | 0.3109 | *CFB* |
| ENSG00000138802 | 4 | 110344928 | 110472052 | 277 | 21 | 48300 | 0.4932 | 0.31094 | *SEC24B* |
| ENSG00000015171 | 10 | 170405 | 310577 | 275 | 28 | 40345 | 0.49298 | 0.31101 | *ZMYND11* |
| ENSG00000147896 | 9 | 27514312 | 27536496 | 109 | 13 | 42199 | 0.49289 | 0.31104 | *IFNK* |
| ENSG00000175105 | 3 | 88178254 | 88203815 | 61 | 6 | 30971 | 0.49281 | 0.31108 | *ZNF654* |
| ENSG00000155324 | 5 | 125685824 | 125842186 | 427 | 92 | 46705 | 0.49211 | 0.31132 | *GRAMD3* |
| ENSG00000117602 | 1 | 24819387 | 24877530 | 201 | 29 | 39523 | 0.49178 | 0.31144 | *RCAN3* |
| ENSG00000143858 | 1 | 202549724 | 202689545 | 351 | 49 | 38290 | 0.49166 | 0.31148 | *SYT2* |

| | | | | | | | | | |
|---|---|---|---|---|---|---|---|---|---|
| ENSG00000005339 | 16 | 3765055 | 3940727 | 223 | 53 | 36971 | 0.4916 | 0.3115 | *CREBBP* |
| ENSG00000163785 | 3 | 133774147 | 133979689 | 533 | 46 | 36158 | 0.49148 | 0.31154 | *RYK* |
| ENSG00000196684 | 19 | 16234838 | 16279386 | 136 | 24 | 48478 | 0.49136 | 0.31159 | *HSH2D* |
| ENSG00000149485 | 11 | 61557099 | 61606790 | 82 | 9 | 33995 | 0.49126 | 0.31162 | *FADS1* |
| ENSG00000255872 | 9 | 37578410 | 38078684 | 1612 | 118 | 50001 | 0.49115 | 0.31166 | *RP11-613M10.9* |
| ENSG00000079101 | 18 | 586988 | 660334 | 251 | 54 | 44740 | 0.49112 | 0.31167 | *CLUL1* |
| ENSG00000171551 | 2 | 233334537 | 233362538 | 118 | 28 | 50302 | 0.49074 | 0.3118 | *ECEL1* |
| ENSG00000146242 | 6 | 83062923 | 83090545 | 65 | 11 | 53546 | 0.49076 | 0.3118 | *TPBG* |
| ENSG00000176083 | 1 | 26678125 | 26711013 | 122 | 20 | 38501 | 0.49069 | 0.31182 | *ZNF683* |
| ENSG00000184588 | 1 | 66248197 | 66850259 | 1583 | 163 | 45117 | 0.49066 | 0.31183 | *PDE4B* |
| ENSG00000105402 | 19 | 47980894 | 48028497 | 70 | 12 | 42001 | 0.49067 | 0.31183 | *NAPA* |
| ENSG00000118322 | 5 | 159980127 | 160289221 | 1066 | 113 | 48303 | 0.49052 | 0.31188 | *ATP10B* |
| ENSG00000261794 | 15 | 30886329 | 30916764 | 18 | 8 | 38175 | 0.49023 | 0.31198 | *GOLGA8H* |
| ENSG00000212710 | 18 | 19983564 | 20007878 | 44 | 17 | 38363 | 0.49015 | 0.31201 | *CTAGE1* |
| ENSG00000185760 | 6 | 73321520 | 73918574 | 1691 | 182 | 43057 | 0.49008 | 0.31204 | *KCNQ5* |
| ENSG00000249852 | 7 | 217554 | 239557 | 96 | 22 | 48100 | 0.4898 | 0.31214 | *AC145676.2* |
| ENSG00000196639 | 3 | 11168779 | 11315243 | 475 | 71 | 38859 | 0.48953 | 0.31223 | *HRH1* |
| ENSG00000141380 | 18 | 23586578 | 23681181 | 217 | 20 | 36756 | 0.48932 | 0.31231 | *SS18* |
| ENSG00000186440 | 1 | 158522441 | 158543394 | 73 | 10 | 50475 | 0.48927 | 0.31232 | *OR6P1* |
| ENSG00000215277 | 14 | 23644525 | 23752686 | 413 | 53 | 45096 | 0.48929 | 0.31232 | *C14orf164* |
| ENSG00000170425 | 17 | 15838231 | 15889060 | 118 | 21 | 39779 | 0.48925 | 0.31233 | *ADORA2B* |
| ENSG00000166535 | 12 | 8965068 | 9049597 | 286 | 50 | 46113 | 0.4892 | 0.31235 | *A2ML1* |
| ENSG00000166436 | 11 | 8623584 | 8703413 | 235 | 29 | 52653 | 0.48917 | 0.31236 | *TRIM66* |
| ENSG00000198003 | 19 | 11521272 | 11556603 | 64 | 22 | 41251 | 0.489 | 0.31242 | *CCDC151* |
| ENSG00000143437 | 1 | 150772181 | 150859244 | 166 | 22 | 49480 | 0.4886 | 0.31256 | *ARNT* |
| ENSG00000085433 | 1 | 109502836 | 109594850 | 294 | 37 | 54597 | 0.48857 | 0.31257 | *WDR47* |
| ENSG00000176014 | 18 | 12297668 | 12354319 | 206 | 15 | 40094 | 0.48855 | 0.31258 | *TUBB6* |
| ENSG00000100744 | 14 | 96819814 | 96863625 | 60 | 14 | 46348 | 0.48843 | 0.31262 | *GSKIP* |
| ENSG00000156575 | 11 | 57134242 | 57158623 | 47 | 11 | 40318 | 0.48839 | 0.31264 | *PRG3* |
| ENSG00000095002 | 2 | 47620108 | 47799450 | 575 | 66 | 40835 | 0.48834 | 0.31265 | *MSH2* |
| ENSG00000121764 | 1 | 32073287 | 32108119 | 80 | 7 | 31630 | 0.48819 | 0.31271 | *HCRTR1* |
| ENSG00000163009 | 2 | 10271509 | 10361851 | 348 | 48 | 40199 | 0.48813 | 0.31273 | *C2orf48* |
| ENSG00000256407 | 1 | 54596897 | 54675746 | 237 | 53 | 47190 | 0.48794 | 0.3128 | *RP11-446E24.4* |
| ENSG00000244165 | 19 | 10212214 | 10236048 | 72 | 19 | 51645 | 0.48786 | 0.31282 | *P2RY11* |
| ENSG00000188112 | 6 | 42058856 | 42120357 | 262 | 51 | 48750 | 0.48782 | 0.31284 | *C6orf132* |
| ENSG00000071626 | 19 | 1397568 | 1445719 | 79 | 20 | 43509 | 0.48747 | 0.31296 | *DAZAP1* |

| | | | | | | | | | |
|---|---|---|---|---|---|---|---|---|---|
| ENSG00000134326 | 2 | 6970701 | 7016766 | 47 | 14 | 47118 | 0.4874 | 0.31299 | *CMPK2* |
| ENSG00000142627 | 1 | 16440832 | 16492582 | 106 | 25 | 38924 | 0.48696 | 0.31314 | *EPHA2* |
| ENSG00000087088 | 19 | 49448072 | 49475055 | 76 | 18 | 46511 | 0.48695 | 0.31315 | *BAX* |
| ENSG00000187764 | 9 | 91965702 | 92123045 | 625 | 92 | 45784 | 0.48684 | 0.31319 | *SEMA4D* |
| ENSG00000177464 | 19 | 46083022 | 46115466 | 70 | 21 | 38750 | 0.48675 | 0.31322 | *GPR4* |
| ENSG00000141720 | 17 | 36911942 | 36966379 | 133 | 26 | 38672 | 0.48653 | 0.31329 | *PIP4K2B* |
| ENSG00000173451 | 12 | 72046789 | 72084419 | 48 | 9 | 49441 | 0.48629 | 0.31338 | *THAP2* |
| ENSG00000164038 | 4 | 103931025 | 104016986 | 155 | 28 | 52026 | 0.4859 | 0.31352 | *SLC9B2* |
| ENSG00000137714 | 11 | 110290607 | 110345605 | 201 | 17 | 46249 | 0.48578 | 0.31356 | *FDX1* |
| ENSG00000033050 | 7 | 150894923 | 150934316 | 129 | 22 | 52812 | 0.4855 | 0.31366 | *ABCF2* |
| ENSG00000005156 | 17 | 33297513 | 33342083 | 42 | 13 | 39097 | 0.48546 | 0.31368 | *LIG3* |
| ENSG00000168824 | 4 | 4339867 | 4430785 | 391 | 61 | 42094 | 0.48542 | 0.31369 | *NSG1* |
| ENSG00000268457 | 20 | 30299001 | 30319134 | 25 | 5 | 30680 | 0.4854 | 0.31369 | *AL160175.1* |
| ENSG00000254999 | 3 | 10147276 | 10178874 | 170 | 10 | 48675 | 0.48534 | 0.31372 | *BRK1* |
| ENSG00000087510 | 20 | 55194358 | 55224339 | 76 | 17 | 49919 | 0.48531 | 0.31373 | *TFAP2C* |
| ENSG00000133424 | 22 | 33548212 | 34328829 | 2977 | 302 | 47741 | 0.48513 | 0.31379 | *LARGE* |
| ENSG00000152463 | 10 | 15064226 | 15125851 | 307 | 39 | 48081 | 0.485 | 0.31384 | *OLAH* |
| ENSG00000168038 | 3 | 41278090 | 42013922 | 2504 | 94 | 39815 | 0.48493 | 0.31386 | *ULK4* |
| ENSG00000108984 | 17 | 67400839 | 67549472 | 361 | 60 | 44400 | 0.4849 | 0.31387 | *MAP2K6* |
| ENSG00000109911 | 11 | 31521297 | 31815546 | 452 | 26 | 35045 | 0.48471 | 0.31394 | *ELP4* |
| ENSG00000196378 | 8 | 145988499 | 146022730 | 111 | 13 | 30027 | 0.48469 | 0.31395 | *ZNF34* |
| ENSG00000117868 | 7 | 158513686 | 158632944 | 492 | 30 | 34826 | 0.48448 | 0.31402 | *ESYT2* |
| ENSG00000144357 | 2 | 170673968 | 170950641 | 1191 | 47 | 43437 | 0.48445 | 0.31403 | *UBR3* |
| ENSG00000107618 | 10 | 48371487 | 48400991 | 46 | 18 | 48533 | 0.4841 | 0.31416 | *RBP3* |
| ENSG00000169313 | 3 | 151045168 | 151112600 | 253 | 36 | 44568 | 0.48337 | 0.31442 | *P2RY12* |
| ENSG00000198822 | 7 | 86263230 | 86504200 | 531 | 63 | 42570 | 0.48334 | 0.31443 | *GRM3* |
| ENSG00000157110 | 8 | 30231944 | 30439778 | 440 | 39 | 49229 | 0.48333 | 0.31443 | *RBPMS* |
| ENSG00000120937 | 1 | 11907521 | 11928988 | 84 | 17 | 47418 | 0.48299 | 0.31455 | *NPPB* |
| ENSG00000135346 | 6 | 87785216 | 87814824 | 103 | 14 | 44903 | 0.48288 | 0.31459 | *CGA* |
| ENSG00000103316 | 16 | 21240195 | 21324404 | 164 | 25 | 42075 | 0.48258 | 0.3147 | *CRYM* |
| ENSG00000069998 | 22 | 17608401 | 17656177 | 249 | 38 | 44664 | 0.48253 | 0.31471 | *CECR5* |
| ENSG00000173166 | 2 | 204249068 | 204410133 | 272 | 33 | 33591 | 0.48251 | 0.31472 | *RAPH1* |
| ENSG00000269237 | 19 | 21577961 | 21684068 | 388 | 26 | 50279 | 0.4825 | 0.31472 | *CTD-2561J22.3* |
| ENSG00000157184 | 1 | 53652101 | 53689869 | 72 | 14 | 34006 | 0.48229 | 0.3148 | *CPT2* |
| ENSG00000187838 | 17 | 7283046 | 7317416 | 94 | 28 | 47681 | 0.48222 | 0.31482 | *TMEM256-PLSCR3* |
| ENSG00000262481 | 17 | 7283053 | 7317416 | 94 | 28 | 47681 | 0.48222 | 0.31482 | *C17orf61-PLSCR3* |

| | | | | | | | | | |
|---|---|---|---|---|---|---|---|---|---|
| ENSG00000187498 | 13 | 110791318 | 110969496 | 867 | 139 | 52005 | 0.48194 | 0.31493 | *COL4A1* |
| ENSG00000117600 | 1 | 99719509 | 99785146 | 218 | 31 | 41753 | 0.48176 | 0.31499 | *LPPR4* |
| ENSG00000166595 | 16 | 66955959 | 66978326 | 26 | 9 | 34742 | 0.48171 | 0.31501 | *FAM96B* |
| ENSG00000004455 | 1 | 33463585 | 33556597 | 166 | 19 | 51746 | 0.48132 | 0.31514 | *AK2* |
| ENSG00000128815 | 10 | 49882921 | 50201001 | 1168 | 118 | 40963 | 0.48091 | 0.31529 | *WDFY4* |
| ENSG00000106244 | 7 | 98979671 | 99016452 | 45 | 13 | 46633 | 0.48085 | 0.31531 | *PDAP1* |
| ENSG00000182070 | 11 | 5162239 | 5217612 | 343 | 26 | 40657 | 0.48084 | 0.31532 | *OR52A1* |
| ENSG00000174136 | 5 | 98094354 | 98144347 | 130 | 24 | 43125 | 0.48076 | 0.31534 | *RGMB* |
| ENSG00000128965 | 15 | 41235160 | 41258710 | 85 | 13 | 38734 | 0.48063 | 0.31539 | *CHAC1* |
| ENSG00000178602 | 2 | 241068446 | 241093979 | 161 | 27 | 49852 | 0.48059 | 0.3154 | *OTOS* |
| ENSG00000122566 | 7 | 26219547 | 26251149 | 84 | 11 | 52130 | 0.48042 | 0.31547 | *HNRNPA2B1* |
| ENSG00000181274 | 10 | 99082255 | 99104458 | 64 | 10 | 48399 | 0.48006 | 0.31559 | *FRAT2* |
| ENSG00000137225 | 6 | 44116548 | 44162139 | 151 | 40 | 45454 | 0.48003 | 0.3156 | *CAPN11* |
| ENSG00000006607 | 2 | 242285658 | 242444256 | 473 | 48 | 54497 | 0.47978 | 0.31569 | *FARP2* |
| ENSG00000205581 | 21 | 40704241 | 40731573 | 105 | 13 | 53105 | 0.47956 | 0.31577 | *HMGN1* |
| ENSG00000100983 | 20 | 33506236 | 33553620 | 78 | 11 | 50835 | 0.47953 | 0.31578 | *GSS* |
| ENSG00000108883 | 17 | 42917311 | 42987030 | 238 | 25 | 52712 | 0.47948 | 0.3158 | *EFTUD2* |
| ENSG00000051128 | 19 | 19030010 | 19062070 | 47 | 15 | 38790 | 0.47944 | 0.31581 | *HOMER3* |
| ENSG00000182973 | 3 | 32716637 | 32825367 | 510 | 35 | 49532 | 0.47908 | 0.31594 | *CNOT10* |
| ENSG00000183023 | 2 | 40314410 | 40848193 | 2136 | 165 | 39891 | 0.47886 | 0.31602 | *SLC8A1* |
| ENSG00000138448 | 2 | 187444792 | 187555628 | 222 | 27 | 51465 | 0.47876 | 0.31605 | *ITGAV* |
| ENSG00000133226 | 1 | 24948207 | 25009758 | 92 | 18 | 48322 | 0.47873 | 0.31607 | *SRRM1* |
| ENSG00000214447 | 17 | 42967135 | 42992758 | 98 | 15 | 53384 | 0.47873 | 0.31607 | *FAM187A* |
| ENSG00000203684 | 1 | 228341787 | 228363213 | 28 | 9 | 46317 | 0.47863 | 0.3161 | *IBA57-AS1* |
| ENSG00000103966 | 15 | 42180950 | 42274776 | 281 | 44 | 45946 | 0.47863 | 0.3161 | *EHD4* |
| ENSG00000130811 | 19 | 10215693 | 10240596 | 69 | 17 | 50980 | 0.47862 | 0.31611 | *EIF3G* |
| ENSG00000002587 | 4 | 11384774 | 11441389 | 211 | 57 | 47894 | 0.47839 | 0.31619 | *HS3ST1* |
| ENSG00000222017 | 2 | 198547830 | 198649547 | 183 | 18 | 53862 | 0.47831 | 0.31622 | *AC011997.1* |
| ENSG00000205090 | 1 | 1460554 | 1485833 | 59 | 19 | 42753 | 0.47817 | 0.31626 | *TMEM240* |
| ENSG00000038358 | 16 | 67896926 | 67928406 | 43 | 12 | 48595 | 0.47812 | 0.31628 | *EDC4* |
| ENSG00000183607 | 2 | 69162364 | 69190102 | 57 | 14 | 41737 | 0.47765 | 0.31645 | *GKN2* |
| ENSG00000117533 | 1 | 171659300 | 171721387 | 262 | 19 | 47774 | 0.47753 | 0.31649 | *VAMP4* |
| ENSG00000137834 | 15 | 66984566 | 67084338 | 307 | 50 | 53515 | 0.47726 | 0.31659 | *SMAD6* |
| ENSG00000136699 | 2 | 130898981 | 130950323 | 125 | 10 | 48689 | 0.47711 | 0.31664 | *SMPD4* |
| ENSG00000135829 | 1 | 182798504 | 182866886 | 109 | 15 | 35206 | 0.47696 | 0.31669 | *DHX9* |
| ENSG00000172831 | 16 | 66958347 | 66988999 | 36 | 11 | 38602 | 0.47695 | 0.3167 | *CES2* |

| | | | | | | | | | |
|---|---|---|---|---|---|---|---|---|---|
| ENSG00000054179 | 9 | 139932550 | 139958497 | 70 | 15 | 51658 | 0.47689 | 0.31672 | *ENTPD2* |
| ENSG00000100612 | 14 | 60600838 | 60646574 | 128 | 7 | 26111 | 0.47671 | 0.31678 | *DHRS7* |
| ENSG00000112877 | 5 | 602387 | 677283 | 324 | 36 | 48437 | 0.47656 | 0.31684 | *CEP72* |
| ENSG00000148943 | 11 | 27506123 | 27538320 | 52 | 10 | 35649 | 0.47629 | 0.31693 | *LIN7C* |
| ENSG00000265303 | 17 | 57264927 | 57302587 | 83 | 12 | 35004 | 0.4763 | 0.31693 | *CTD-2510F5.6* |
| ENSG00000166225 | 12 | 69854129 | 69983562 | 318 | 25 | 45516 | 0.4762 | 0.31696 | *FRS2* |
| ENSG00000198842 | 1 | 167053282 | 167108402 | 213 | 28 | 48681 | 0.47612 | 0.31699 | *DUSP27* |
| ENSG00000171295 | 19 | 11915099 | 11956016 | 131 | 8 | 50508 | 0.47601 | 0.31703 | *ZNF440* |
| ENSG00000168546 | 8 | 21537915 | 21679869 | 308 | 86 | 43505 | 0.47495 | 0.31741 | *GFRA2* |
| ENSG00000120160 | 9 | 27274656 | 27307137 | 139 | 21 | 40374 | 0.4749 | 0.31743 | *EQTN* |
| ENSG00000109063 | 17 | 10521843 | 10570626 | 170 | 17 | 33915 | 0.47446 | 0.31759 | *MYH3* |
| ENSG00000172331 | 7 | 134321560 | 134374565 | 218 | 15 | 36159 | 0.47428 | 0.31765 | *BPGM* |
| ENSG00000196482 | 1 | 216666588 | 217321097 | 2383 | 241 | 46090 | 0.47426 | 0.31766 | *ESRRG* |
| ENSG00000140986 | 16 | 1983975 | 2017607 | 163 | 29 | 46109 | 0.4741 | 0.31771 | *RPL3L* |
| ENSG00000259511 | 15 | 84831242 | 84860986 | 19 | 6 | 39860 | 0.47371 | 0.31785 | *UBE2Q2L* |
| ENSG00000120727 | 5 | 138667276 | 138715406 | 55 | 14 | 41147 | 0.47367 | 0.31787 | *PAIP2* |
| ENSG00000196109 | 19 | 22351893 | 22389753 | 142 | 11 | 44211 | 0.47355 | 0.31791 | *ZNF676* |
| ENSG00000158480 | 20 | 48509928 | 48542080 | 105 | 13 | 38249 | 0.47344 | 0.31795 | *SPATA2* |
| ENSG00000143412 | 1 | 150944493 | 150978110 | 84 | 18 | 41997 | 0.47334 | 0.31799 | *ANXA9* |
| ENSG00000174898 | 19 | 5710688 | 5788745 | 252 | 54 | 44577 | 0.47333 | 0.31799 | *CATSPERD* |
| ENSG00000185033 | 15 | 90693836 | 90782911 | 345 | 41 | 39152 | 0.47301 | 0.3181 | *SEMA4B* |
| ENSG00000111196 | 12 | 10748612 | 10776222 | 101 | 12 | 36612 | 0.473 | 0.31811 | *MAGOHB* |
| ENSG00000032389 | 2 | 3182696 | 3391653 | 817 | 63 | 44178 | 0.4729 | 0.31814 | *TSSC1* |
| ENSG00000110446 | 11 | 60694556 | 60730002 | 119 | 20 | 41865 | 0.4728 | 0.31818 | *SLC15A3* |
| ENSG00000163513 | 3 | 30637994 | 30745634 | 341 | 61 | 49922 | 0.47263 | 0.31824 | *TGFBR2* |
| ENSG00000132429 | 6 | 105596155 | 105637870 | 114 | 17 | 36240 | 0.47248 | 0.31829 | *POPDC3* |
| ENSG00000139719 | 12 | 122704111 | 122761068 | 119 | 16 | 40048 | 0.47212 | 0.31842 | *VPS33A* |
| ENSG00000228919 | 4 | 7930728 | 7952022 | 86 | 12 | 44718 | 0.47211 | 0.31843 | *AC097381.1* |
| ENSG00000113569 | 5 | 37278239 | 37381283 | 193 | 21 | 47256 | 0.47186 | 0.31851 | *NUP155* |
| ENSG00000182866 | 1 | 32706840 | 32761766 | 73 | 26 | 39641 | 0.47182 | 0.31853 | *LCK* |
| ENSG00000151790 | 4 | 156765890 | 156851558 | 230 | 36 | 47160 | 0.47182 | 0.31853 | *TDO2* |
| ENSG00000197870 | 12 | 11408857 | 11432739 | 53 | 9 | 32155 | 0.47173 | 0.31856 | *PRB3* |
| ENSG00000161583 | 17 | 34735936 | 34818104 | 25 | 7 | 23686 | 0.47172 | 0.31856 | *TBC1D3G* |
| ENSG00000084693 | 2 | 27255232 | 27303490 | 64 | 12 | 40389 | 0.47132 | 0.31871 | *AGBL5* |
| ENSG00000164761 | 8 | 119925796 | 119974439 | 180 | 25 | 47117 | 0.47108 | 0.31879 | *TNFRSF11B* |
| ENSG00000163395 | 1 | 201149953 | 201208080 | 251 | 46 | 42564 | 0.47102 | 0.31881 | *IGFN1* |

| | | | | | | | | | |
|---|---|---|---|---|---|---|---|---|---|
| ENSG00000089220 | 12 | 118563663 | 118593389 | 75 | 18 | 49508 | 0.47098 | 0.31883 | *PEBP1* |
| ENSG00000197914 | 6 | 27788952 | 27809305 | 49 | 14 | 40196 | 0.47093 | 0.31885 | *HIST1H4K* |
| ENSG00000243480 | 1 | 104149999 | 104178402 | 9 | 6 | 47354 | 0.47057 | 0.31897 | *AMY2A* |
| ENSG00000122512 | 7 | 6002870 | 6058756 | 208 | 30 | 48586 | 0.47058 | 0.31897 | *PMS2* |
| ENSG00000180340 | 17 | 42624925 | 42646907 | 57 | 17 | 48413 | 0.47059 | 0.31897 | *FZD2* |
| ENSG00000113838 | 3 | 186253862 | 186298332 | 113 | 12 | 31027 | 0.47037 | 0.31905 | *TBCCD1* |
| ENSG00000137819 | 15 | 69581286 | 69710119 | 307 | 37 | 31975 | 0.47013 | 0.31913 | *PAQR5* |
| ENSG00000219438 | 22 | 48875272 | 49256724 | 2428 | 251 | 45034 | 0.46992 | 0.31921 | *FAM19A5* |
| ENSG00000134440 | 18 | 55257888 | 55299445 | 148 | 23 | 50929 | 0.46988 | 0.31922 | *NARS* |
| ENSG00000103241 | 16 | 86534133 | 86558076 | 113 | 30 | 47255 | 0.46972 | 0.31928 | *FOXF1* |
| ENSG00000151718 | 4 | 184010446 | 184251930 | 510 | 70 | 51440 | 0.46966 | 0.3193 | *WWC2* |
| ENSG00000131143 | 16 | 85822239 | 85850650 | 80 | 14 | 32800 | 0.46952 | 0.31935 | *COX4I1* |
| ENSG00000116819 | 1 | 36028971 | 36070929 | 28 | 16 | 37687 | 0.46944 | 0.31938 | *TFAP2E* |
| ENSG00000136653 | 1 | 206670879 | 206772616 | 267 | 46 | 52481 | 0.46904 | 0.31952 | *RASSF5* |
| ENSG00000100075 | 22 | 19153095 | 19176343 | 88 | 17 | 42792 | 0.46904 | 0.31952 | *SLC25A1* |
| ENSG00000188004 | 1 | 159794264 | 159835137 | 119 | 35 | 41041 | 0.46895 | 0.31955 | *C1orf204* |
| ENSG00000106211 | 7 | 75921861 | 75943612 | 80 | 17 | 40810 | 0.46893 | 0.31956 | *HSPB1* |
| ENSG00000125898 | 20 | 804358 | 848106 | 204 | 50 | 45962 | 0.4688 | 0.3196 | *FAM110A* |
| ENSG00000102683 | 13 | 23745091 | 23909304 | 1179 | 110 | 49332 | 0.46846 | 0.31973 | *SGCG* |
| ENSG00000143321 | 1 | 156701899 | 156746717 | 117 | 24 | 36152 | 0.46809 | 0.31986 | *HDGF* |
| ENSG00000148156 | 9 | 111606871 | 111629239 | 124 | 20 | 51735 | 0.46788 | 0.31994 | *ACTL7B* |
| ENSG00000016490 | 1 | 86924051 | 86975972 | 184 | 40 | 51328 | 0.4676 | 0.32004 | *CLCA1* |
| ENSG00000077809 | 7 | 74061994 | 74185026 | 168 | 27 | 45596 | 0.46689 | 0.32029 | *GTF2I* |
| ENSG00000171557 | 4 | 155515286 | 155544119 | 41 | 9 | 48629 | 0.46682 | 0.32031 | *FGG* |
| ENSG00000177300 | 4 | 184229220 | 184251927 | 60 | 14 | 45443 | 0.46639 | 0.32047 | *CLDN22* |
| ENSG00000267937 | 21 | 38878740 | 38898973 | 57 | 13 | 53677 | 0.46633 | 0.32049 | *AP001421.1* |
| ENSG00000166455 | 16 | 81077102 | 81120872 | 186 | 9 | 46933 | 0.4663 | 0.3205 | *C16orf46* |
| ENSG00000075413 | 14 | 103841729 | 103980168 | 379 | 21 | 38478 | 0.4659 | 0.32065 | *MARK3* |
| ENSG00000237190 | 5 | 133727778 | 133757589 | 43 | 7 | 40384 | 0.4658 | 0.32068 | *CDKN2AIPNL* |
| ENSG00000196350 | 19 | 22459210 | 22509978 | 108 | 19 | 44578 | 0.46566 | 0.32073 | *ZNF729* |
| ENSG00000110777 | 11 | 111212977 | 111336355 | 497 | 54 | 40211 | 0.46562 | 0.32074 | *POU2AF1* |
| ENSG00000162598 | 1 | 60442941 | 60549442 | 276 | 52 | 55017 | 0.46538 | 0.32083 | *C1orf87* |
| ENSG00000186103 | 3 | 121279551 | 121315481 | 111 | 14 | 46866 | 0.46531 | 0.32086 | *ARGFX* |
| ENSG00000133250 | 19 | 8565462 | 8589048 | 117 | 8 | 44383 | 0.46524 | 0.32088 | *ZNF414* |
| ENSG00000159208 | 1 | 150244953 | 150269505 | 62 | 10 | 32774 | 0.46508 | 0.32094 | *C1orf51* |
| ENSG00000113734 | 5 | 172561445 | 172601390 | 126 | 20 | 42096 | 0.46502 | 0.32096 | *BNIP1* |

| | | | | | | | | | |
|---|---|---|---|---|---|---|---|---|---|
| ENSG00000197753 | 6 | 35763070 | 35811651 | 152 | 14 | 44705 | 0.46468 | 0.32108 | *LHFPL5* |
| ENSG00000087589 | 20 | 54977168 | 55044396 | 217 | 34 | 34649 | 0.46451 | 0.32114 | *CASS4* |
| ENSG00000168952 | 14 | 25268862 | 25529503 | 908 | 109 | 44545 | 0.46436 | 0.3212 | *STXBP6* |
| ENSG00000168615 | 8 | 38844388 | 38972663 | 292 | 27 | 49694 | 0.46403 | 0.32131 | *ADAM9* |
| ENSG00000136950 | 9 | 127614409 | 127650003 | 72 | 12 | 51807 | 0.46385 | 0.32138 | *ARPC5L* |
| ENSG00000234188 | 6 | 116432320 | 116453093 | 57 | 16 | 45293 | 0.46369 | 0.32144 | *AL121963.1* |
| ENSG00000143549 | 1 | 154117784 | 154177124 | 90 | 16 | 45023 | 0.46354 | 0.32149 | *TPM3* |
| ENSG00000076067 | 12 | 56905713 | 56994745 | 214 | 26 | 43023 | 0.46348 | 0.32151 | *RBMS2* |
| ENSG00000243073 | 1 | 12929045 | 12956025 | 20 | 13 | 34217 | 0.46319 | 0.32161 | *PRAMEF4* |
| ENSG00000170775 | 7 | 124376051 | 124415681 | 98 | 15 | 49296 | 0.46313 | 0.32164 | *GPR37* |
| ENSG00000187753 | 9 | 88825180 | 88884572 | 136 | 24 | 43097 | 0.46311 | 0.32164 | *C9orf153* |
| ENSG00000138041 | 2 | 55764428 | 55856015 | 302 | 33 | 40455 | 0.46308 | 0.32165 | *SMEK2* |
| ENSG00000083896 | 4 | 69166105 | 69225807 | 112 | 19 | 52627 | 0.46289 | 0.32172 | *YTHDC1* |
| ENSG00000147804 | 8 | 145625126 | 145652279 | 54 | 19 | 44850 | 0.46265 | 0.32181 | *SLC39A4* |
| ENSG00000197121 | 2 | 197687728 | 197802520 | 227 | 17 | 34904 | 0.46244 | 0.32188 | *PGAP1* |
| ENSG00000173597 | 4 | 70576880 | 70663679 | 315 | 33 | 45493 | 0.46236 | 0.32191 | *SULT1B1* |
| ENSG00000168878 | 2 | 85874437 | 85905864 | 59 | 20 | 42962 | 0.46223 | 0.32196 | *SFTPB* |
| ENSG00000164879 | 8 | 86275665 | 86371269 | 201 | 26 | 50553 | 0.46198 | 0.32205 | *CA3* |
| ENSG00000125462 | 1 | 156364042 | 156410493 | 67 | 28 | 41262 | 0.4617 | 0.32215 | *C1orf61* |
| ENSG00000243364 | 1 | 155026207 | 155052029 | 43 | 11 | 46155 | 0.46163 | 0.32218 | *EFNA4* |
| ENSG00000226524 | 6 | 88022323 | 88119459 | 418 | 31 | 48382 | 0.46157 | 0.3222 | *RP1-102H19.8* |
| ENSG00000270249 | 7 | 102171948 | 102242891 | 5 | 3 | 43125 | 0.46145 | 0.32224 | *RP11-514P8.7* |
| ENSG00000183207 | 19 | 49486705 | 49529252 | 129 | 14 | 51612 | 0.46145 | 0.32224 | *RUVBL2* |
| ENSG00000213953 | 2 | 187351840 | 187375393 | 56 | 9 | 54383 | 0.46142 | 0.32225 | *AC018867.1* |
| ENSG00000139734 | 13 | 60229717 | 60748121 | 1484 | 72 | 49523 | 0.46116 | 0.32234 | *DIAPH3* |
| ENSG00000125651 | 19 | 6369580 | 6403992 | 131 | 17 | 45518 | 0.46114 | 0.32235 | *GTF2F1* |
| ENSG00000165202 | 9 | 125366948 | 125388026 | 109 | 19 | 47983 | 0.4611 | 0.32236 | *OR1Q1* |
| ENSG00000189139 | 14 | 44963545 | 44986482 | 127 | 15 | 45276 | 0.4611 | 0.32236 | *FSCB* |
| ENSG00000074266 | 11 | 85945586 | 85999855 | 144 | 21 | 51064 | 0.46101 | 0.32239 | *EED* |
| ENSG00000166793 | 11 | 57402560 | 57427417 | 35 | 8 | 43262 | 0.46099 | 0.3224 | *YPEL4* |
| ENSG00000228208 | 1 | 218673438 | 218709320 | 144 | 10 | 25858 | 0.46078 | 0.32248 | *C1orf143* |
| ENSG00000215717 | 1 | 109622425 | 109649556 | 101 | 22 | 45913 | 0.46041 | 0.32261 | *TMEM167B* |
| ENSG00000151729 | 4 | 186054395 | 186081536 | 81 | 15 | 39033 | 0.46022 | 0.32268 | *SLC25A4* |
| ENSG00000198853 | 9 | 35480124 | 35571895 | 166 | 26 | 46566 | 0.45988 | 0.3228 | *RUSC2* |
| ENSG00000187688 | 17 | 16308856 | 16350317 | 117 | 18 | 46022 | 0.45976 | 0.32284 | *TRPV2* |
| ENSG00000239382 | 19 | 36490022 | 36515141 | 36 | 11 | 35888 | 0.45969 | 0.32287 | *ALKBH6* |

| | | | | | | | | | |
|---|---|---|---|---|---|---|---|---|---|
| ENSG00000121406 | 19 | 58028693 | 58078910 | 110 | 14 | 55663 | 0.45967 | 0.32288 | *ZNF549* |
| ENSG00000119471 | 9 | 115132217 | 115244690 | 428 | 30 | 53567 | 0.45962 | 0.32289 | *HSDL2* |
| ENSG00000174899 | 3 | 157251035 | 157405538 | 526 | 60 | 44339 | 0.45958 | 0.32291 | *C3orf55* |
| ENSG00000166171 | 10 | 103320317 | 103379425 | 117 | 15 | 46498 | 0.45958 | 0.32291 | *DPCD* |
| ENSG00000069812 | 1 | 6462478 | 6494730 | 133 | 17 | 39780 | 0.45934 | 0.323 | *HES2* |
| ENSG00000116191 | 1 | 178684300 | 178899238 | 500 | 43 | 43622 | 0.45926 | 0.32302 | *RALGPS2* |
| ENSG00000132906 | 1 | 15807327 | 15863029 | 234 | 14 | 50307 | 0.45921 | 0.32304 | *CASP9* |
| ENSG00000143776 | 1 | 227167566 | 227516175 | 2013 | 45 | 51423 | 0.45913 | 0.32307 | *CDC42BPA* |
| ENSG00000167131 | 17 | 42966510 | 42992758 | 100 | 15 | 53467 | 0.4591 | 0.32308 | *CCDC103* |
| ENSG00000086062 | 9 | 33094080 | 33177354 | 342 | 41 | 49144 | 0.45884 | 0.32318 | *B4GALT1* |
| ENSG00000169136 | 19 | 50421959 | 50447192 | 84 | 19 | 39891 | 0.45876 | 0.3232 | *ATF5* |
| ENSG00000170500 | 2 | 100879753 | 100949195 | 285 | 24 | 37397 | 0.45872 | 0.32322 | *LONRF2* |
| ENSG00000001561 | 6 | 46087730 | 46124436 | 131 | 11 | 36963 | 0.45842 | 0.32333 | *ENPP4* |
| ENSG00000158966 | 1 | 64926428 | 65168741 | 553 | 101 | 37156 | 0.45831 | 0.32336 | *CACHD1* |
| ENSG00000040633 | 17 | 7128347 | 7153041 | 29 | 8 | 55008 | 0.4583 | 0.32337 | *PHF23* |
| ENSG00000133739 | 8 | 86009382 | 86068311 | 116 | 19 | 45337 | 0.45818 | 0.32341 | *LRRCC1* |
| ENSG00000162384 | 1 | 53669771 | 53696289 | 56 | 12 | 38133 | 0.45791 | 0.32351 | *C1orf123* |
| ENSG00000196778 | 11 | 4500109 | 4521138 | 73 | 13 | 48248 | 0.45785 | 0.32353 | *OR52K1* |
| ENSG00000203883 | 20 | 62669076 | 62690994 | 58 | 16 | 45153 | 0.45751 | 0.32365 | *SOX18* |
| ENSG00000204396 | 6 | 31723367 | 31755108 | 78 | 23 | 32358 | 0.45742 | 0.32369 | *VWA7* |
| ENSG00000104361 | 8 | 99192061 | 99316621 | 313 | 54 | 45391 | 0.45714 | 0.32379 | *NIPAL2* |
| ENSG00000139178 | 12 | 7232183 | 7271869 | 91 | 18 | 48176 | 0.45711 | 0.3238 | *C1RL* |
| ENSG00000046889 | 8 | 68854353 | 69159265 | 1100 | 108 | 47125 | 0.457 | 0.32384 | *PREX2* |
| ENSG00000167747 | 19 | 51290961 | 51318186 | 119 | 17 | 48251 | 0.45662 | 0.32397 | *C19orf48* |
| ENSG00000144771 | 3 | 54942264 | 55011115 | 206 | 26 | 39731 | 0.45659 | 0.32398 | *LRTM1* |
| ENSG00000222011 | 7 | 102379418 | 102459672 | 218 | 34 | 39623 | 0.45627 | 0.3241 | *FAM185A* |
| ENSG00000129128 | 4 | 177231115 | 177263396 | 189 | 27 | 47624 | 0.45618 | 0.32413 | *SPCS3* |
| ENSG00000088002 | 19 | 49045332 | 49112682 | 332 | 49 | 42889 | 0.45608 | 0.32417 | *SULT2B1* |
| ENSG00000129250 | 17 | 4891243 | 4941696 | 154 | 28 | 43309 | 0.45603 | 0.32419 | *KIF1C* |
| ENSG00000256715 | 21 | 14731931 | 14755386 | 134 | 19 | 33997 | 0.45567 | 0.32431 | *AL050302.1* |
| ENSG00000120899 | 8 | 27158999 | 27326903 | 530 | 44 | 46612 | 0.45555 | 0.32436 | *PTK2B* |
| ENSG00000171873 | 20 | 4191329 | 4239721 | 167 | 35 | 46702 | 0.45528 | 0.32445 | *ADRA1D* |
| ENSG00000122733 | 9 | 34947484 | 34992541 | 89 | 23 | 41828 | 0.45524 | 0.32447 | *KIAA1045* |
| ENSG00000213828 | 2 | 240073493 | 240094053 | 84 | 11 | 56253 | 0.45493 | 0.32458 | *AC017028.1* |
| ENSG00000248511 | 4 | 93208402 | 93235329 | 37 | 12 | 44942 | 0.45476 | 0.32464 | *RP11-9B6.1* |
| ENSG00000168994 | 6 | 3712848 | 3762260 | 274 | 29 | 36006 | 0.45476 | 0.32464 | *PXDC1* |

| | | | | | | | | | |
|---|---|---|---|---|---|---|---|---|---|
| ENSG00000206432 | 18 | 5872071 | 5905954 | 107 | 26 | 36593 | 0.45476 | 0.32464 | *TMEM200C* |
| ENSG00000101447 | 20 | 37544955 | 37591703 | 117 | 12 | 54744 | 0.45461 | 0.32469 | *FAM83D* |
| ENSG00000219626 | 2 | 24289396 | 24402509 | 442 | 29 | 50369 | 0.45444 | 0.32476 | *FAM228B* |
| ENSG00000131019 | 6 | 150374286 | 150400231 | 116 | 18 | 46196 | 0.45436 | 0.32478 | *ULBP3* |
| ENSG00000179331 | 11 | 107789229 | 107844208 | 192 | 32 | 44182 | 0.4543 | 0.32481 | *RAB39A* |
| ENSG00000154764 | 3 | 13847755 | 13931618 | 300 | 56 | 34314 | 0.45418 | 0.32485 | *WNT7A* |
| ENSG00000143622 | 1 | 155857599 | 155891195 | 71 | 11 | 34751 | 0.4538 | 0.32499 | *RIT1* |
| ENSG00000132952 | 13 | 31181830 | 31243686 | 89 | 13 | 40199 | 0.4538 | 0.32499 | *USPL1* |
| ENSG00000197540 | 19 | 534034 | 559919 | 181 | 38 | 40792 | 0.45372 | 0.32501 | *GZMM* |
| ENSG00000130300 | 19 | 17452257 | 17498159 | 192 | 45 | 42665 | 0.45357 | 0.32507 | *PLVAP* |
| ENSG00000121904 | 1 | 33969609 | 34641443 | 1950 | 228 | 42458 | 0.45354 | 0.32508 | *CSMD2* |
| ENSG00000166266 | 11 | 107869459 | 107988503 | 280 | 31 | 49858 | 0.45352 | 0.32509 | *CUL5* |
| ENSG00000166133 | 15 | 40851499 | 40876659 | 89 | 9 | 33719 | 0.45341 | 0.32513 | *RPUSD2* |
| ENSG00000153214 | 2 | 112802800 | 112886895 | 279 | 41 | 48756 | 0.45333 | 0.32516 | *TMEM87B* |
| ENSG00000218416 | 2 | 241378732 | 241406131 | 129 | 35 | 48666 | 0.45333 | 0.32516 | *AC110619.2* |
| ENSG00000127838 | 2 | 219125115 | 219221516 | 260 | 25 | 46862 | 0.45313 | 0.32523 | *PNKD* |
| ENSG00000169905 | 1 | 179799102 | 179856938 | 136 | 20 | 49362 | 0.45303 | 0.32526 | *TOR1AIP2* |
| ENSG00000072756 | 3 | 3158600 | 3202563 | 195 | 33 | 36012 | 0.45291 | 0.32531 | *TRNT1* |
| ENSG00000010030 | 6 | 36312419 | 36366164 | 202 | 41 | 37338 | 0.45275 | 0.32536 | *ETV7* |
| ENSG00000166510 | 18 | 52558740 | 52636739 | 211 | 25 | 47706 | 0.45259 | 0.32542 | *CCDC68* |
| ENSG00000197353 | 8 | 143821568 | 143843952 | 57 | 11 | 46440 | 0.45252 | 0.32545 | *LYPD2* |
| ENSG00000060069 | 18 | 77429801 | 77524510 | 399 | 38 | 45020 | 0.45251 | 0.32545 | *CTDP1* |
| ENSG00000137817 | 15 | 72523522 | 72575340 | 122 | 19 | 39566 | 0.45226 | 0.32554 | *PARP6* |
| ENSG00000163995 | 4 | 7957039 | 8170559 | 827 | 176 | 44608 | 0.45221 | 0.32556 | *ABLIM2* |
| ENSG00000197345 | 11 | 68648744 | 68681303 | 87 | 14 | 50305 | 0.45205 | 0.32562 | *MRPL21* |
| ENSG00000082074 | 5 | 39095338 | 39284630 | 509 | 67 | 42512 | 0.45197 | 0.32565 | *FYB* |
| ENSG00000110344 | 11 | 118220300 | 118279926 | 116 | 14 | 36377 | 0.45189 | 0.32567 | *UBE4A* |
| ENSG00000181847 | 3 | 113985760 | 114039135 | 167 | 32 | 46545 | 0.45168 | 0.32575 | *TIGIT* |
| ENSG00000146205 | 2 | 242117924 | 242174792 | 193 | 47 | 37984 | 0.45152 | 0.32581 | *ANO7* |
| ENSG00000235173 | 8 | 145182672 | 145205746 | 8 | 2 | 40245 | 0.4512 | 0.32592 | *FAM203A* |
| ENSG00000137601 | 4 | 170304426 | 170543780 | 626 | 35 | 45256 | 0.45072 | 0.32609 | *NEK1* |
| ENSG00000170290 | 11 | 107568104 | 107600419 | 87 | 18 | 44857 | 0.45068 | 0.32611 | *SLN* |
| ENSG00000163050 | 1 | 227075237 | 227185246 | 334 | 57 | 43378 | 0.45062 | 0.32613 | *ADCK3* |
| ENSG00000144034 | 2 | 73946231 | 73974527 | 62 | 5 | 42181 | 0.45048 | 0.32618 | *TPRKB* |
| ENSG00000110921 | 12 | 110001060 | 110045067 | 159 | 20 | 54684 | 0.45049 | 0.32618 | *MVK* |
| ENSG00000268797 | 19 | 41297202 | 41414187 | 460 | 78 | 44883 | 0.45032 | 0.32624 | *CTC-490E21.12* |

| | | | | | | | | | |
|---|---|---|---|---|---|---|---|---|---|
| ENSG00000068793 | 15 | 22882005 | 23016016 | 371 | 90 | 45381 | 0.45022 | 0.32627 | *CYFIP1* |
| ENSG00000271605 | 17 | 62451569 | 62474760 | 23 | 5 | 35565 | 0.45015 | 0.3263 | *MILR1* |
| ENSG00000188981 | 4 | 3236096 | 3283465 | 170 | 19 | 40270 | 0.45007 | 0.32633 | *MSANTD1* |
| ENSG00000145864 | 5 | 160705436 | 160986050 | 792 | 91 | 47157 | 0.44998 | 0.32636 | *GABRB2* |
| ENSG00000143870 | 2 | 10913517 | 10988103 | 543 | 37 | 38713 | 0.44989 | 0.3264 | *PDIA6* |
| ENSG00000145819 | 5 | 142139949 | 142618576 | 1080 | 134 | 35712 | 0.44977 | 0.32644 | *ARHGAP26* |
| ENSG00000160633 | 19 | 5613046 | 5678489 | 141 | 24 | 48147 | 0.44967 | 0.32647 | *SAFB* |
| ENSG00000155269 | 4 | 8550452 | 8631486 | 399 | 76 | 41430 | 0.44964 | 0.32648 | *GPR78* |
| ENSG00000168303 | 7 | 40155622 | 40184258 | 38 | 10 | 35234 | 0.44948 | 0.32654 | *MPLKIP* |
| ENSG00000104848 | 19 | 49560675 | 49586198 | 102 | 16 | 47293 | 0.44945 | 0.32655 | *KCNA7* |
| ENSG00000175216 | 11 | 46754598 | 46877847 | 222 | 30 | 34589 | 0.44925 | 0.32662 | *CKAP5* |
| ENSG00000076242 | 3 | 37024823 | 37117380 | 193 | 17 | 40140 | 0.44923 | 0.32663 | *MLH1* |
| ENSG00000139187 | 12 | 9092640 | 9173356 | 246 | 19 | 46175 | 0.4492 | 0.32664 | *KLRG1* |
| ENSG00000106123 | 7 | 142542792 | 142578847 | 111 | 16 | 29389 | 0.44912 | 0.32667 | *EPHB6* |
| ENSG00000180869 | 1 | 85083913 | 85110703 | 81 | 19 | 39653 | 0.44908 | 0.32669 | *C1orf180* |
| ENSG00000268885 | 5 | 657759 | 678839 | 109 | 21 | 46319 | 0.44903 | 0.32671 | *AC026740.1* |
| ENSG00000171777 | 19 | 38889695 | 38926945 | 149 | 28 | 44645 | 0.44885 | 0.32677 | *RASGRP4* |
| ENSG00000114686 | 3 | 131171056 | 131231827 | 133 | 20 | 35278 | 0.44883 | 0.32678 | *MRPL3* |
| ENSG00000205186 | 8 | 82360576 | 82383814 | 68 | 10 | 33925 | 0.44881 | 0.32678 | *FABP9* |
| ENSG00000120832 | 12 | 107361069 | 107390944 | 85 | 22 | 41772 | 0.44873 | 0.32681 | *MTERFD3* |
| ENSG00000196535 | 17 | 27390528 | 27517430 | 301 | 33 | 35456 | 0.44864 | 0.32685 | *MYO18A* |
| ENSG00000187658 | 5 | 157088561 | 157117151 | 107 | 18 | 43225 | 0.44854 | 0.32688 | *C5orf52* |
| ENSG00000134444 | 18 | 59844491 | 59984355 | 488 | 36 | 50780 | 0.44817 | 0.32702 | *KIAA1468* |
| ENSG00000139537 | 12 | 49287893 | 49335623 | 152 | 10 | 51687 | 0.44802 | 0.32707 | *CCDC65* |
| ENSG00000138964 | 22 | 44558836 | 44625413 | 331 | 54 | 47937 | 0.44788 | 0.32712 | *PARVG* |
| ENSG00000162520 | 1 | 33135507 | 33179197 | 104 | 11 | 37728 | 0.44778 | 0.32716 | *SYNC* |
| ENSG00000105856 | 7 | 106799406 | 106852974 | 120 | 11 | 53869 | 0.44765 | 0.3272 | *HBP1* |
| ENSG00000177508 | 16 | 54307216 | 54330675 | 59 | 15 | 47034 | 0.44755 | 0.32724 | *IRX3* |
| ENSG00000197863 | 19 | 37298330 | 37351689 | 94 | 17 | 42893 | 0.44735 | 0.32731 | *ZNF790* |
| ENSG00000177025 | 19 | 58459805 | 58495902 | 133 | 13 | 45928 | 0.44708 | 0.32741 | *C19orf18* |
| ENSG00000129521 | 14 | 34383437 | 34941980 | 2139 | 255 | 44898 | 0.44693 | 0.32746 | *EGLN3* |
| ENSG00000149328 | 11 | 134191768 | 134258235 | 200 | 60 | 49030 | 0.44681 | 0.32751 | *GLB1L2* |
| ENSG00000105497 | 19 | 52064551 | 52102991 | 187 | 35 | 38608 | 0.4467 | 0.32755 | *ZNF175* |
| ENSG00000132000 | 19 | 14032000 | 14074204 | 56 | 19 | 43944 | 0.44659 | 0.32759 | *PODNL1* |
| ENSG00000267368 | 7 | 102267472 | 102293238 | 1 | 1 | 42811 | 0.44655 | 0.3276 | *UPK3BL* |
| ENSG00000166965 | 15 | 91488100 | 91516349 | 101 | 20 | 38270 | 0.44651 | 0.32761 | *RCCD1* |

| | | | | | | | | | |
|---|---|---|---|---|---|---|---|---|---|
| ENSG00000152092 | 1 | 176816438 | 177144109 | 833 | 102 | 42175 | 0.44649 | 0.32762 | *ASTN1* |
| ENSG00000124831 | 2 | 238526219 | 238732325 | 809 | 89 | 39899 | 0.44545 | 0.328 | *LRRFIP1* |
| ENSG00000269028 | 3 | 96325981 | 96347000 | 80 | 12 | 38245 | 0.44542 | 0.32801 | *MTRNR2L12* |
| ENSG00000153130 | 4 | 141168440 | 141316880 | 416 | 55 | 41414 | 0.44487 | 0.32821 | *SCOC* |
| ENSG00000162377 | 1 | 53142508 | 53174038 | 85 | 12 | 44111 | 0.44461 | 0.3283 | *COA7* |
| ENSG00000117155 | 1 | 85099390 | 85166486 | 193 | 24 | 45773 | 0.44461 | 0.3283 | *SSX2IP* |
| ENSG00000069696 | 11 | 627293 | 650706 | 109 | 21 | 47472 | 0.44447 | 0.32835 | *DRD4* |
| ENSG00000121075 | 17 | 59519765 | 59572471 | 105 | 22 | 52616 | 0.44444 | 0.32836 | *TBX4* |
| ENSG00000163159 | 1 | 151132463 | 151177797 | 49 | 15 | 50001 | 0.44433 | 0.3284 | *VPS72* |
| ENSG00000185664 | 12 | 56337889 | 56377101 | 56 | 19 | 46294 | 0.44434 | 0.3284 | *PMEL* |
| ENSG00000152443 | 19 | 58248164 | 58279527 | 70 | 12 | 39461 | 0.44422 | 0.32844 | *ZNF776* |
| ENSG00000088726 | 3 | 12765024 | 12820956 | 190 | 31 | 36007 | 0.44404 | 0.32851 | *TMEM40* |
| ENSG00000163755 | 3 | 148837371 | 148901519 | 235 | 36 | 48910 | 0.444 | 0.32852 | *HPS3* |
| ENSG00000181555 | 3 | 47047919 | 47215457 | 254 | 23 | 43124 | 0.44392 | 0.32855 | *SETD2* |
| ENSG00000169418 | 1 | 153641113 | 153676468 | 72 | 15 | 38889 | 0.44369 | 0.32863 | *NPR1* |
| ENSG00000204228 | 6 | 33162419 | 33184608 | 48 | 13 | 51024 | 0.44344 | 0.32872 | *HSD17B8* |
| ENSG00000105223 | 19 | 40844363 | 40896346 | 146 | 32 | 43151 | 0.44331 | 0.32877 | *PLD3* |
| ENSG00000215910 | 1 | 11811844 | 11859642 | 230 | 22 | 49143 | 0.44325 | 0.32879 | *C1orf167* |
| ENSG00000172425 | 11 | 118388187 | 118411912 | 47 | 6 | 26800 | 0.44318 | 0.32882 | *TTC36* |
| ENSG00000214376 | 11 | 93541398 | 93593697 | 192 | 26 | 28854 | 0.443 | 0.32888 | *VSTM5* |
| ENSG00000177119 | 12 | 45599770 | 45844187 | 628 | 45 | 28473 | 0.44294 | 0.32891 | *ANO6* |
| ENSG00000129535 | 14 | 24539316 | 24594223 | 91 | 25 | 38514 | 0.44247 | 0.32908 | *NRL* |
| ENSG00000197497 | 19 | 53652466 | 53706639 | 229 | 36 | 38072 | 0.44242 | 0.32909 | *ZNF665* |
| ENSG00000134207 | 1 | 114621913 | 114706541 | 322 | 76 | 50186 | 0.44224 | 0.32916 | *SYT6* |
| ENSG00000146707 | 7 | 76229303 | 76266578 | 106 | 10 | 40693 | 0.44224 | 0.32916 | *POMZP3* |
| ENSG00000107201 | 9 | 32445300 | 32536322 | 260 | 30 | 54698 | 0.44211 | 0.3292 | *DDX58* |
| ENSG00000010319 | 3 | 52457069 | 52489101 | 40 | 11 | 52104 | 0.4417 | 0.32935 | *SEMA3G* |
| ENSG00000046604 | 18 | 29068006 | 29138971 | 248 | 30 | 42336 | 0.44158 | 0.3294 | *DSG2* |
| ENSG00000096092 | 6 | 52525907 | 52561386 | 170 | 22 | 48372 | 0.44145 | 0.32944 | *TMEM14A* |
| ENSG00000159184 | 17 | 46792125 | 46816540 | 50 | 15 | 47175 | 0.44144 | 0.32945 | *HOXB13* |
| ENSG00000188747 | 9 | 140307802 | 140338858 | 71 | 12 | 52072 | 0.4414 | 0.32946 | *NOXA1* |
| ENSG00000080845 | 20 | 34884258 | 35167040 | 317 | 64 | 42728 | 0.4413 | 0.3295 | *DLGAP4* |
| ENSG00000138271 | 3 | 151001891 | 151044740 | 161 | 23 | 54326 | 0.44126 | 0.32951 | *GPR87* |
| ENSG00000163932 | 3 | 53180025 | 53236733 | 124 | 24 | 40930 | 0.44109 | 0.32957 | *PRKCD* |
| ENSG00000170266 | 3 | 33028100 | 33148722 | 406 | 56 | 45429 | 0.44093 | 0.32963 | *GLB1* |
| ENSG00000092445 | 15 | 41839873 | 41881536 | 104 | 17 | 46883 | 0.44063 | 0.32974 | *TYRO3* |

| | | | | | | | | | |
|---|---|---|---|---|---|---|---|---|---|
| ENSG00000176658 | 17 | 30809540 | 31214195 | 899 | 96 | 46831 | 0.4405 | 0.32979 | *MYO1D* |
| ENSG00000102524 | 13 | 108893588 | 108970832 | 223 | 51 | 48775 | 0.44041 | 0.32982 | *TNFSF13B* |
| ENSG00000101574 | 18 | 2527524 | 2581508 | 209 | 25 | 42478 | 0.44027 | 0.32987 | *METTL4* |
| ENSG00000138669 | 4 | 81999837 | 82146218 | 327 | 38 | 39086 | 0.44021 | 0.32989 | *PRKG2* |
| ENSG00000163040 | 2 | 132275248 | 132301239 | 86 | 12 | 30440 | 0.43984 | 0.33003 | *CCDC74A* |
| ENSG00000103121 | 16 | 80999698 | 81063875 | 358 | 20 | 35954 | 0.43973 | 0.33007 | *CMC2* |
| ENSG00000154917 | 3 | 133533083 | 133624680 | 304 | 61 | 42788 | 0.4396 | 0.33011 | *RAB6B* |
| ENSG00000138036 | 2 | 43991178 | 44047149 | 227 | 20 | 36657 | 0.43957 | 0.33012 | *DYNC2LI1* |
| ENSG00000162599 | 1 | 61320931 | 61938465 | 1682 | 251 | 46350 | 0.43937 | 0.3302 | *NFIA* |
| ENSG00000159625 | 16 | 57718705 | 57775717 | 175 | 35 | 35718 | 0.43895 | 0.33035 | *CCDC135* |
| ENSG00000125449 | 17 | 73096047 | 73136360 | 124 | 23 | 39638 | 0.43894 | 0.33035 | *ARMC7* |
| ENSG00000169220 | 5 | 176774838 | 176809602 | 68 | 13 | 43500 | 0.43889 | 0.33037 | *RGS14* |
| ENSG00000080839 | 20 | 35614752 | 35734398 | 191 | 28 | 42134 | 0.43887 | 0.33038 | *RBL1* |
| ENSG00000189042 | 19 | 37168514 | 37228603 | 151 | 15 | 53883 | 0.43874 | 0.33042 | *ZNF567* |
| ENSG00000134574 | 11 | 47226493 | 47270767 | 82 | 17 | 43380 | 0.4383 | 0.33058 | *DDB2* |
| ENSG00000100121 | 22 | 22978780 | 23000368 | 35 | 13 | 33237 | 0.43771 | 0.3308 | *GGTLC2* |
| ENSG00000215356 | 8 | 7773859 | 7822271 | 2 | 1 | 42688 | 0.43725 | 0.33096 | *ZNF705B* |
| ENSG00000183520 | 1 | 38464930 | 38500496 | 59 | 24 | 46677 | 0.43714 | 0.331 | *UTP11L* |
| ENSG00000198730 | 11 | 10762534 | 10811290 | 130 | 31 | 42580 | 0.43703 | 0.33104 | *CTR9* |
| ENSG00000187566 | 6 | 18110718 | 18132851 | 96 | 12 | 54726 | 0.43698 | 0.33106 | *NHLRC1* |
| ENSG00000138778 | 4 | 104016963 | 104129566 | 258 | 19 | 53930 | 0.43694 | 0.33108 | *CENPE* |
| ENSG00000182333 | 10 | 90414198 | 90448571 | 97 | 20 | 43792 | 0.43675 | 0.33115 | *LIPF* |
| ENSG00000180745 | 10 | 129666105 | 129701211 | 128 | 39 | 50458 | 0.43663 | 0.33119 | *CLRN3* |
| ENSG00000145632 | 5 | 57739809 | 57766087 | 137 | 19 | 52019 | 0.43661 | 0.3312 | *PLK2* |
| ENSG00000124205 | 20 | 57865482 | 57911047 | 97 | 35 | 43659 | 0.43654 | 0.33122 | *EDN3* |
| ENSG00000126549 | 4 | 70851647 | 70878287 | 59 | 15 | 50356 | 0.43645 | 0.33126 | *STATH* |
| ENSG00000132746 | 11 | 67419633 | 67458671 | 103 | 16 | 34408 | 0.43638 | 0.33128 | *ALDH3B2* |
| ENSG00000221887 | 18 | 61606535 | 61659008 | 179 | 29 | 43366 | 0.43639 | 0.33128 | *HMSD* |
| ENSG00000140254 | 15 | 45399569 | 45432136 | 48 | 21 | 41879 | 0.43631 | 0.33131 | *DUOXA1* |
| ENSG00000140044 | 14 | 75884419 | 75950814 | 189 | 37 | 43108 | 0.43627 | 0.33132 | *JDP2* |
| ENSG00000118260 | 2 | 208384461 | 208478155 | 166 | 15 | 49179 | 0.43587 | 0.33147 | *CREB1* |
| ENSG00000152104 | 1 | 214512039 | 214735792 | 909 | 108 | 47716 | 0.43583 | 0.33148 | *PTPN14* |
| ENSG00000056972 | 6 | 111867657 | 111937481 | 218 | 32 | 38000 | 0.43581 | 0.33149 | *TRAF3IP2* |
| ENSG00000100083 | 22 | 37994481 | 38039571 | 118 | 18 | 40313 | 0.43578 | 0.3315 | *GGA1* |
| ENSG00000163618 | 3 | 62374022 | 62871054 | 2016 | 195 | 46366 | 0.43572 | 0.33152 | *CADPS* |
| ENSG00000187997 | 17 | 76132434 | 76172258 | 188 | 43 | 42558 | 0.43573 | 0.33152 | *C17orf99* |

| | | | | | | | | | |
|---|---|---|---|---|---|---|---|---|---|
| ENSG00000138363 | 2 | 216166540 | 216224487 | 205 | 24 | 43474 | 0.43545 | 0.33162 | *ATIC* |
| ENSG00000105205 | 19 | 40211890 | 40238668 | 138 | 12 | 47426 | 0.43538 | 0.33164 | *CLC* |
| ENSG00000162994 | 2 | 55391927 | 55469699 | 232 | 29 | 46193 | 0.43522 | 0.3317 | *CLHC1* |
| ENSG00000128271 | 22 | 24803847 | 24848328 | 122 | 24 | 40936 | 0.43515 | 0.33173 | *ADORA2A* |
| ENSG00000239810 | 1 | 13042394 | 13062998 | 4 | 2 | 42884 | 0.435 | 0.33178 | *WI2-3308P17.2* |
| ENSG00000183251 | 11 | 5312244 | 5333226 | 106 | 23 | 43029 | 0.43499 | 0.33179 | *OR51B4* |
| ENSG00000132510 | 17 | 7733222 | 7768114 | 79 | 20 | 40735 | 0.43479 | 0.33186 | *KDM6B* |
| ENSG00000109163 | 4 | 68595046 | 68630078 | 126 | 17 | 48817 | 0.43467 | 0.3319 | *GNRHR* |
| ENSG00000129173 | 11 | 19235610 | 19273167 | 72 | 18 | 42286 | 0.43459 | 0.33193 | *E2F8* |
| ENSG00000169851 | 4 | 30712037 | 31158422 | 1405 | 113 | 43581 | 0.43454 | 0.33195 | *PCDH7* |
| ENSG00000107672 | 10 | 123706603 | 123744732 | 164 | 15 | 49473 | 0.43451 | 0.33196 | *NSMCE4A* |
| ENSG00000100478 | 14 | 31484312 | 31572818 | 234 | 41 | 48090 | 0.43448 | 0.33197 | *AP4S1* |
| ENSG00000197993 | 7 | 142628201 | 142669768 | 116 | 17 | 29254 | 0.43439 | 0.332 | *KEL* |
| ENSG00000012504 | 12 | 100857486 | 100968191 | 185 | 40 | 38948 | 0.4342 | 0.33207 | *NR1H4* |
| ENSG00000165283 | 9 | 35089888 | 35113154 | 44 | 10 | 42745 | 0.43397 | 0.33216 | *STOML2* |
| ENSG00000008735 | 22 | 51029114 | 51062409 | 159 | 28 | 40816 | 0.43376 | 0.33223 | *MAPK8IP2* |
| ENSG00000120215 | 9 | 5880802 | 5920606 | 90 | 17 | 42379 | 0.43358 | 0.3323 | *MLANA* |
| ENSG00000123200 | 13 | 46518600 | 46636894 | 288 | 18 | 54512 | 0.43313 | 0.33246 | *ZC3H13* |
| ENSG00000233412 | 3 | 97877544 | 97898485 | 156 | 5 | 48509 | 0.43273 | 0.33261 | *OR5H15* |
| ENSG00000106302 | 7 | 123459037 | 123527532 | 141 | 26 | 52758 | 0.43272 | 0.33261 | *HYAL4* |
| ENSG00000161610 | 17 | 40326078 | 40347470 | 15 | 9 | 47514 | 0.43258 | 0.33266 | *HCRT* |
| ENSG00000135423 | 12 | 56854736 | 56892198 | 71 | 16 | 44436 | 0.43238 | 0.33273 | *GLS2* |
| ENSG00000227152 | 1 | 248594508 | 248615434 | 17 | 10 | 37429 | 0.43234 | 0.33275 | *OR2T7* |
| ENSG00000130762 | 1 | 3360990 | 3407677 | 153 | 23 | 44205 | 0.4323 | 0.33276 | *ARHGEF16* |
| ENSG00000142484 | 17 | 4665179 | 4696506 | 109 | 22 | 42645 | 0.43232 | 0.33276 | *TM4SF5* |
| ENSG00000115594 | 2 | 102671004 | 102806334 | 445 | 58 | 39910 | 0.43204 | 0.33286 | *IL1R1* |
| ENSG00000181541 | 4 | 151493077 | 151515843 | 27 | 12 | 44252 | 0.43175 | 0.33296 | *MAB21L2* |
| ENSG00000187546 | 7 | 15229943 | 15611640 | 1441 | 139 | 45792 | 0.4317 | 0.33298 | *AGMO* |
| ENSG00000004779 | 16 | 23582323 | 23617677 | 130 | 16 | 49612 | 0.43121 | 0.33316 | *NDUFAB1* |
| ENSG00000146151 | 6 | 55289167 | 55454012 | 840 | 62 | 51679 | 0.43108 | 0.33321 | *HMGCLL1* |
| ENSG00000063169 | 19 | 48101453 | 48216533 | 382 | 45 | 51216 | 0.431 | 0.33323 | *GLTSCR1* |
| ENSG00000184979 | 22 | 18622666 | 18670164 | 157 | 28 | 41941 | 0.43088 | 0.33328 | *USP18* |
| ENSG00000144191 | 2 | 98952618 | 99025064 | 245 | 31 | 41811 | 0.43076 | 0.33332 | *CNGA3* |
| ENSG00000244067 | 6 | 52604897 | 52638367 | 122 | 15 | 32669 | 0.43072 | 0.33334 | *GSTA2* |
| ENSG00000167800 | 11 | 67388774 | 67417031 | 114 | 20 | 36299 | 0.43067 | 0.33335 | *TBX10* |
| ENSG00000151148 | 12 | 109905207 | 109984507 | 264 | 26 | 50116 | 0.43054 | 0.3334 | *UBE3B* |

| | | | | | | | | | |
|---|---|---|---|---|---|---|---|---|---|
| ENSG00000158008 | 1 | 26336375 | 26372955 | 73 | 13 | 39435 | 0.43041 | 0.33345 | *EXTL1* |
| ENSG00000258539 | 10 | 126295649 | 126490296 | 465 | 57 | 48524 | 0.43032 | 0.33348 | *RP11-12J10.3* |
| ENSG00000214226 | 17 | 54859274 | 54926134 | 201 | 16 | 52398 | 0.43027 | 0.3335 | *C17orf67* |
| ENSG00000213398 | 16 | 67963653 | 67988034 | 24 | 7 | 35955 | 0.43019 | 0.33353 | *LCAT* |
| ENSG00000047188 | 5 | 112839380 | 112940982 | 478 | 30 | 36566 | 0.43006 | 0.33357 | *YTHDC2* |
| ENSG00000144591 | 2 | 220353589 | 220381710 | 60 | 10 | 47184 | 0.43001 | 0.33359 | *GMPPA* |
| ENSG00000106927 | 9 | 116812407 | 116850752 | 158 | 30 | 49772 | 0.42998 | 0.3336 | *AMBP* |
| ENSG00000153347 | 5 | 94717048 | 94796158 | 275 | 38 | 39949 | 0.42997 | 0.33361 | *FAM81B* |
| ENSG00000169612 | 15 | 83644959 | 83669809 | 62 | 15 | 48164 | 0.42983 | 0.33366 | *FAM103A1* |
| ENSG00000218819 | 2 | 21336789 | 21376144 | 78 | 14 | 42794 | 0.42975 | 0.33369 | *TDRD15* |
| ENSG00000100554 | 14 | 67751088 | 67836982 | 187 | 32 | 44455 | 0.42971 | 0.3337 | *ATP6V1D* |
| ENSG00000179021 | 3 | 88188893 | 88227879 | 90 | 7 | 31929 | 0.42961 | 0.33374 | *C3orf38* |
| ENSG00000053918 | 11 | 2455914 | 2880339 | 1238 | 217 | 43435 | 0.42949 | 0.33378 | *KCNQ1* |
| ENSG00000160505 | 19 | 56337944 | 56403220 | 310 | 68 | 43174 | 0.42913 | 0.33391 | *NLRP4* |
| ENSG00000155099 | 8 | 91996024 | 92063292 | 140 | 21 | 40335 | 0.42906 | 0.33394 | *TMEM55A* |
| ENSG00000146378 | 6 | 132928161 | 132955414 | 143 | 26 | 44273 | 0.42904 | 0.33395 | *TAAR2* |
| ENSG00000081320 | 2 | 196988290 | 197051227 | 230 | 19 | 47001 | 0.42882 | 0.33403 | *STK17B* |
| ENSG00000255292 | 11 | 111947627 | 112074528 | 285 | 40 | 40840 | 0.42862 | 0.3341 | *SDHD* |
| ENSG00000198169 | 8 | 145936298 | 145991802 | 105 | 21 | 34979 | 0.42845 | 0.33416 | *ZNF251* |
| ENSG00000106536 | 7 | 39007598 | 39542694 | 1765 | 139 | 48764 | 0.42816 | 0.33427 | *POU6F2* |
| ENSG00000129474 | 14 | 23430383 | 23461851 | 57 | 11 | 52205 | 0.42812 | 0.33428 | *AJUBA* |
| ENSG00000176231 | 19 | 16049818 | 16070768 | 121 | 6 | 36781 | 0.42806 | 0.33431 | *OR10H4* |
| ENSG00000117362 | 1 | 150227804 | 150251980 | 31 | 11 | 40144 | 0.42796 | 0.33434 | *APH1A* |
| ENSG00000173862 | 7 | 33755593 | 33778068 | 56 | 17 | 30230 | 0.42755 | 0.33449 | *RP11-89N17.1* |
| ENSG00000164037 | 4 | 103796205 | 103950896 | 369 | 13 | 46453 | 0.42751 | 0.3345 | *SLC9B1* |
| ENSG00000215428 | 9 | 133264029 | 133292639 | 135 | 37 | 41361 | 0.42752 | 0.3345 | *AL354898.1* |
| ENSG00000198855 | 12 | 108898962 | 108929614 | 90 | 17 | 48670 | 0.42751 | 0.3345 | *FICD* |
| ENSG00000157765 | 4 | 25646923 | 25690370 | 102 | 30 | 39556 | 0.42733 | 0.33457 | *SLC34A2* |
| ENSG00000149634 | 20 | 44505128 | 44526274 | 59 | 10 | 39539 | 0.42731 | 0.33458 | *SPATA25* |
| ENSG00000141384 | 18 | 23795900 | 23981649 | 555 | 50 | 39861 | 0.42719 | 0.33462 | *TAF4B* |
| ENSG00000156219 | 4 | 76922337 | 77043955 | 780 | 31 | 46620 | 0.42701 | 0.33469 | *ART3* |
| ENSG00000103111 | 16 | 77214732 | 77246302 | 175 | 18 | 39029 | 0.42696 | 0.3347 | *MON1B* |
| ENSG00000166140 | 15 | 41089284 | 41116767 | 85 | 10 | 40397 | 0.42657 | 0.33485 | *ZFYVE19* |
| ENSG00000132153 | 3 | 47834399 | 47901685 | 77 | 14 | 38468 | 0.42629 | 0.33495 | *DHX30* |
| ENSG00000268396 | 2 | 239182329 | 239202498 | 55 | 13 | 51513 | 0.4258 | 0.33513 | *AC012485.1* |
| ENSG00000130311 | 19 | 17410327 | 17444110 | 145 | 17 | 45850 | 0.4257 | 0.33516 | *DDA1* |

| | | | | | | | | | |
|---|---|---|---|---|---|---|---|---|---|
| ENSG00000133466 | 22 | 37566207 | 37605425 | 149 | 21 | 44790 | 0.42558 | 0.33521 | *C1QTNF6* |
| ENSG00000152684 | 5 | 52073774 | 52109880 | 147 | 16 | 50910 | 0.4254 | 0.33527 | *PELO* |
| ENSG00000079739 | 1 | 64048947 | 64135916 | 279 | 53 | 47223 | 0.42539 | 0.33528 | *PGM1* |
| ENSG00000103423 | 16 | 4465806 | 4516776 | 132 | 21 | 46327 | 0.42525 | 0.33533 | *DNAJA3* |
| ENSG00000145757 | 5 | 94977885 | 95044415 | 212 | 26 | 48261 | 0.42502 | 0.33541 | *SPATA9* |
| ENSG00000214946 | 17 | 15625561 | 15659476 | 80 | 15 | 41303 | 0.42496 | 0.33543 | *TBC1D26* |
| ENSG00000173726 | 1 | 235262651 | 235302251 | 88 | 11 | 50029 | 0.42471 | 0.33552 | *TOMM20* |
| ENSG00000120889 | 8 | 22867646 | 22936692 | 218 | 42 | 43338 | 0.42463 | 0.33555 | *TNFRSF10B* |
| ENSG00000204387 | 6 | 31792385 | 31817541 | 91 | 30 | 38740 | 0.4241 | 0.33574 | *C6orf48* |
| ENSG00000120949 | 1 | 12113434 | 12214264 | 254 | 57 | 46637 | 0.42404 | 0.33577 | *TNFRSF8* |
| ENSG00000221866 | 7 | 131798091 | 132343447 | 1712 | 194 | 41506 | 0.42398 | 0.33579 | *PLXNA4* |
| ENSG00000177427 | 17 | 18153848 | 18179866 | 59 | 12 | 55649 | 0.42337 | 0.33601 | *MIEF2* |
| ENSG00000136731 | 2 | 128838774 | 128963251 | 368 | 25 | 41147 | 0.42318 | 0.33608 | *UGGT1* |
| ENSG00000140945 | 16 | 82650408 | 83840204 | 7153 | 496 | 47460 | 0.42304 | 0.33613 | *CDH13* |
| ENSG00000168411 | 16 | 74645292 | 74710779 | 283 | 26 | 46867 | 0.423 | 0.33615 | *RFWD3* |
| ENSG00000167191 | 16 | 19858616 | 19907489 | 130 | 23 | 40186 | 0.42293 | 0.33617 | *GPRC5B* |
| ENSG00000197586 | 20 | 25166329 | 25217365 | 167 | 23 | 37816 | 0.42294 | 0.33617 | *ENTPD6* |
| ENSG00000137135 | 9 | 35648872 | 35685863 | 125 | 18 | 44225 | 0.42289 | 0.33619 | *ARHGEF39* |
| ENSG00000116183 | 1 | 176422307 | 176824735 | 961 | 87 | 48014 | 0.42281 | 0.33622 | *PAPPA2* |
| ENSG00000169271 | 5 | 53741445 | 53762207 | 107 | 18 | 53166 | 0.42246 | 0.33635 | *HSPB3* |
| ENSG00000081189 | 5 | 88003975 | 88209922 | 434 | 46 | 42627 | 0.42152 | 0.33669 | *MEF2C* |
| ENSG00000181873 | 1 | 228343516 | 228379958 | 65 | 12 | 50370 | 0.42149 | 0.3367 | *IBA57* |
| ENSG00000135355 | 6 | 90594188 | 90615839 | 76 | 19 | 53779 | 0.42128 | 0.33678 | *GJA10* |
| ENSG00000109667 | 4 | 9762777 | 10066560 | 1354 | 68 | 32939 | 0.42124 | 0.33679 | *SLC2A9* |
| ENSG00000269526 | 19 | 53507174 | 53529375 | 39 | 10 | 35131 | 0.42104 | 0.33686 | *ERVV-1* |
| ENSG00000176040 | 3 | 111743690 | 111810116 | 287 | 37 | 46331 | 0.4208 | 0.33695 | *TMPRSS7* |
| ENSG00000005844 | 16 | 30473979 | 30544506 | 108 | 30 | 37656 | 0.42042 | 0.33709 | *ITGAL* |
| ENSG00000240224 | 2 | 234611638 | 234691945 | 335 | 32 | 42077 | 0.42032 | 0.33713 | *UGT1A5* |
| ENSG00000204227 | 6 | 33166272 | 33190499 | 54 | 15 | 52498 | 0.42031 | 0.33713 | *RING1* |
| ENSG00000138472 | 3 | 108616639 | 108682742 | 277 | 38 | 51180 | 0.42017 | 0.33718 | *GUCA1C* |
| ENSG00000132541 | 8 | 99104572 | 99139469 | 141 | 25 | 54261 | 0.41988 | 0.33729 | *HRSP12* |
| ENSG00000214562 | 10 | 89107425 | 89140452 | 30 | 12 | 42123 | 0.41983 | 0.33731 | *NUTM2D* |
| ENSG00000257962 | 8 | 86148524 | 86172077 | 44 | 8 | 54211 | 0.41978 | 0.33732 | *RP11-219B4.6* |
| ENSG00000150347 | 10 | 63651059 | 63866703 | 562 | 112 | 51122 | 0.41947 | 0.33744 | *ARID5B* |
| ENSG00000264116 | 18 | 73108967 | 73140995 | 100 | 17 | 41191 | 0.4194 | 0.33746 | *RP11-321M21.3* |
| ENSG00000188779 | 15 | 68102042 | 68136899 | 120 | 16 | 38743 | 0.41921 | 0.33753 | *SKOR1* |

| | | | | | | | | | |
|---|---|---|---|---|---|---|---|---|---|
| ENSG00000144785 | 12 | 56669700 | 56719843 | 96 | 12 | 44469 | 0.4191 | 0.33757 | *RP11-977G19.10* |
| ENSG00000131791 | 1 | 146616685 | 146654129 | 158 | 13 | 39238 | 0.41894 | 0.33763 | *PRKAB2* |
| ENSG00000206026 | 18 | 73111431 | 73149658 | 122 | 16 | 40217 | 0.41827 | 0.33788 | *SMIM21* |
| ENSG00000157916 | 1 | 2313267 | 2346883 | 99 | 26 | 41380 | 0.41795 | 0.33799 | *RER1* |
| ENSG00000125870 | 20 | 16700606 | 16732421 | 126 | 32 | 46753 | 0.41768 | 0.33809 | *SNRPB2* |
| ENSG00000128833 | 15 | 52474519 | 52597995 | 251 | 49 | 40089 | 0.41754 | 0.33814 | *MYO5C* |
| ENSG00000143514 | 1 | 223957601 | 224043674 | 209 | 22 | 38558 | 0.41742 | 0.33818 | *TP53BP2* |
| ENSG00000071677 | 2 | 238465217 | 238485818 | 126 | 18 | 41209 | 0.41722 | 0.33826 | *PRLH* |
| ENSG00000173209 | 2 | 61394553 | 61428338 | 88 | 15 | 48459 | 0.417 | 0.33834 | *AHSA2* |
| ENSG00000149646 | 20 | 34546512 | 34628622 | 139 | 21 | 28078 | 0.41683 | 0.3384 | *CNBD2* |
| ENSG00000078295 | 5 | 7386321 | 7840194 | 1322 | 133 | 45253 | 0.41666 | 0.33846 | *ADCY2* |
| ENSG00000204850 | 19 | 46987765 | 47009755 | 106 | 22 | 50377 | 0.41668 | 0.33846 | *AC011484.1* |
| ENSG00000136709 | 2 | 128448596 | 128578761 | 396 | 46 | 44738 | 0.41613 | 0.33866 | *WDR33* |
| ENSG00000127080 | 9 | 95365466 | 95442547 | 233 | 21 | 42262 | 0.41544 | 0.33891 | *IPPK* |
| ENSG00000184954 | 12 | 55852984 | 55873922 | 101 | 11 | 55541 | 0.41526 | 0.33898 | *OR6C70* |
| ENSG00000124596 | 6 | 40991366 | 41075526 | 203 | 20 | 50522 | 0.41509 | 0.33904 | *OARD1* |
| ENSG00000135845 | 1 | 172329329 | 172423230 | 255 | 33 | 39980 | 0.415 | 0.33907 | *PIGC* |
| ENSG00000082175 | 11 | 100890355 | 101011255 | 372 | 35 | 45716 | 0.415 | 0.33907 | *PGR* |
| ENSG00000162069 | 16 | 3067683 | 3096927 | 85 | 12 | 54131 | 0.41488 | 0.33912 | *CCDC64B* |
| ENSG00000116990 | 1 | 40351098 | 40377928 | 71 | 21 | 49475 | 0.41483 | 0.33913 | *MYCL* |
| ENSG00000121966 | 2 | 136861919 | 136885735 | 37 | 9 | 37326 | 0.41474 | 0.33917 | *CXCR4* |
| ENSG00000159648 | 16 | 58000339 | 58032020 | 96 | 16 | 45644 | 0.41449 | 0.33926 | *TEPP* |
| ENSG00000162735 | 1 | 160236602 | 160266138 | 49 | 10 | 37540 | 0.41436 | 0.3393 | *PEX19* |
| ENSG00000160789 | 1 | 156042364 | 156119880 | 196 | 23 | 27397 | 0.41421 | 0.33936 | *LMNA* |
| ENSG00000163507 | 3 | 108258716 | 108318491 | 123 | 17 | 48827 | 0.41395 | 0.33946 | *KIAA1524* |
| ENSG00000114656 | 3 | 128618709 | 128731533 | 372 | 38 | 36738 | 0.41391 | 0.33947 | *KIAA1257* |
| ENSG00000140350 | 15 | 69060874 | 69123236 | 132 | 24 | 31340 | 0.41388 | 0.33948 | *ANP32A* |
| ENSG00000176946 | 2 | 242513820 | 242586864 | 233 | 19 | 45482 | 0.41382 | 0.3395 | *THAP4* |
| ENSG00000187994 | 19 | 39348470 | 39378919 | 45 | 13 | 37949 | 0.4136 | 0.33958 | *RINL* |
| ENSG00000237649 | 6 | 33349313 | 33387701 | 103 | 20 | 50925 | 0.41358 | 0.33959 | *KIFC1* |
| ENSG00000183340 | 11 | 96113153 | 96250738 | 374 | 42 | 53660 | 0.41347 | 0.33963 | *JRKL* |
| ENSG00000179583 | 16 | 10961055 | 11036079 | 185 | 45 | 44853 | 0.41345 | 0.33964 | *CIITA* |
| ENSG00000185522 | 11 | 544855 | 570779 | 158 | 28 | 48264 | 0.41311 | 0.33976 | *C11orf35* |
| ENSG00000141968 | 19 | 6762725 | 6867377 | 403 | 64 | 43455 | 0.4131 | 0.33977 | *VAV1* |
| ENSG00000186198 | 15 | 65327708 | 65355734 | 61 | 12 | 51663 | 0.41307 | 0.33978 | *SLC51B* |
| ENSG00000118946 | 13 | 58195944 | 58313445 | 287 | 45 | 49560 | 0.41278 | 0.33988 | *PCDH17* |

| | | | | | | | | | |
|---|---|---|---|---|---|---|---|---|---|
| ENSG00000171960 | 1 | 43114096 | 43152429 | 81 | 16 | 52805 | 0.41263 | 0.33994 | *PPIH* |
| ENSG00000108821 | 17 | 48250650 | 48288993 | 95 | 31 | 42084 | 0.41249 | 0.33999 | *COL1A1* |
| ENSG00000169490 | 8 | 38836327 | 38864343 | 77 | 8 | 53822 | 0.41241 | 0.34002 | *TM2D2* |
| ENSG00000245848 | 19 | 33780840 | 33803470 | 70 | 11 | 51957 | 0.41228 | 0.34007 | *CEBPA* |
| ENSG00000178645 | 10 | 50877697 | 50928307 | 182 | 22 | 38867 | 0.41219 | 0.3401 | *C10orf53* |
| ENSG00000269897 | 10 | 22595317 | 22628471 | 22 | 12 | 43093 | 0.41217 | 0.34011 | *COMMD3-BMI1* |
| ENSG00000124196 | 20 | 42344804 | 42365638 | 50 | 18 | 53775 | 0.41198 | 0.34018 | *GTSF1L* |
| ENSG00000131409 | 19 | 51010149 | 51081302 | 168 | 45 | 43273 | 0.41166 | 0.34029 | *LRRC4B* |
| ENSG00000138162 | 10 | 123738689 | 124024060 | 1234 | 156 | 43245 | 0.41144 | 0.34038 | *TACC2* |
| ENSG00000011600 | 19 | 36385303 | 36409197 | 65 | 11 | 31666 | 0.4112 | 0.34046 | *TYROBP* |
| ENSG00000155890 | 3 | 140386881 | 140429992 | 159 | 21 | 44848 | 0.41116 | 0.34048 | *TRIM42* |
| ENSG00000178928 | 19 | 48294500 | 48332308 | 210 | 40 | 47959 | 0.41077 | 0.34062 | *TPRX1* |
| ENSG00000166685 | 17 | 71179129 | 71214646 | 144 | 14 | 47420 | 0.41061 | 0.34068 | *COG1* |
| ENSG00000172733 | 8 | 30843321 | 30901231 | 121 | 23 | 35748 | 0.41046 | 0.34074 | *PURG* |
| ENSG00000160799 | 3 | 46953216 | 47033500 | 125 | 17 | 46756 | 0.41041 | 0.34075 | *CCDC12* |
| ENSG00000152467 | 19 | 58535400 | 58575999 | 92 | 18 | 47089 | 0.41037 | 0.34077 | *ZSCAN1* |
| ENSG00000036054 | 3 | 99969844 | 100054095 | 164 | 16 | 46911 | 0.4102 | 0.34083 | *TBC1D23* |
| ENSG00000144619 | 3 | 2130497 | 3109645 | 4546 | 426 | 45084 | 0.41018 | 0.34084 | *CNTN4* |
| ENSG00000070915 | 16 | 56889119 | 56959762 | 316 | 50 | 46295 | 0.41018 | 0.34084 | *SLC12A3* |
| ENSG00000244020 | 1 | 237157403 | 237177718 | 56 | 17 | 42527 | 0.41013 | 0.34085 | *MT1HL1* |
| ENSG00000166261 | 11 | 123584885 | 123622383 | 132 | 12 | 52157 | 0.41011 | 0.34086 | *ZNF202* |
| ENSG00000179476 | 14 | 45356498 | 45386460 | 72 | 11 | 30674 | 0.41005 | 0.34089 | *C14orf28* |
| ENSG00000178199 | 6 | 149758794 | 149816197 | 188 | 13 | 45790 | 0.40986 | 0.34096 | *ZC3H12D* |
| ENSG00000101198 | 20 | 61862136 | 61914046 | 302 | 46 | 48119 | 0.40978 | 0.34098 | *NKAIN4* |
| ENSG00000225968 | 7 | 1717755 | 1797590 | 195 | 40 | 49580 | 0.40976 | 0.34099 | *ELFN1* |
| ENSG00000197601 | 11 | 13680217 | 13763893 | 212 | 29 | 54693 | 0.40954 | 0.34107 | *FAR1* |
| ENSG00000182327 | 17 | 4682254 | 4703685 | 64 | 15 | 40892 | 0.40938 | 0.34113 | *GLTPD2* |
| ENSG00000183655 | 15 | 86292554 | 86348261 | 248 | 42 | 46688 | 0.40913 | 0.34122 | *KLHL25* |
| ENSG00000212768 | 15 | 35260552 | 35282268 | 60 | 13 | 37679 | 0.40906 | 0.34125 | *AC114546.1* |
| ENSG00000099797 | 19 | 14617897 | 14686792 | 211 | 40 | 49708 | 0.40904 | 0.34126 | *TECR* |
| ENSG00000173080 | 1 | 155901480 | 155922625 | 26 | 6 | 27860 | 0.40883 | 0.34133 | *RXFP4* |
| ENSG00000074660 | 17 | 1527152 | 1559041 | 86 | 22 | 49724 | 0.40877 | 0.34135 | *SCARF1* |
| ENSG00000083828 | 19 | 58271023 | 58341307 | 207 | 25 | 42446 | 0.40865 | 0.3414 | *ZNF586* |
| ENSG00000081760 | 12 | 125539925 | 125637873 | 285 | 50 | 49600 | 0.40861 | 0.34141 | *AACS* |
| ENSG00000086300 | 7 | 26321541 | 26423949 | 343 | 46 | 36379 | 0.4084 | 0.34149 | *SNX10* |
| ENSG00000127337 | 12 | 69743483 | 69794576 | 222 | 20 | 45813 | 0.40831 | 0.34152 | *YEATS4* |

| | | | | | | | | | |
|---|---|---|---|---|---|---|---|---|---|
| ENSG00000137691 | 11 | 101908174 | 101965291 | 151 | 14 | 40296 | 0.40829 | 0.34153 | *C11orf70* |
| ENSG00000197595 | 13 | 113291358 | 113348811 | 201 | 35 | 49720 | 0.40828 | 0.34153 | *C13orf35* |
| ENSG00000126264 | 19 | 36383382 | 36405279 | 61 | 10 | 30528 | 0.4082 | 0.34156 | *HCST* |
| ENSG00000188833 | 9 | 140318816 | 140346268 | 64 | 13 | 51053 | 0.40812 | 0.34159 | *ENTPD8* |
| ENSG00000255639 | 12 | 4756800 | 4863747 | 344 | 40 | 40401 | 0.40777 | 0.34172 | *RP11-234B24.6* |
| ENSG00000163001 | 2 | 55736740 | 55783015 | 139 | 26 | 44073 | 0.40727 | 0.3419 | *CCDC104* |
| ENSG00000135226 | 4 | 70136217 | 70170768 | 25 | 8 | 43668 | 0.40682 | 0.34207 | *UGT2B28* |
| ENSG00000172350 | 11 | 119009722 | 119043360 | 65 | 11 | 27728 | 0.40666 | 0.34213 | *ABCG4* |
| ENSG00000100567 | 14 | 58701549 | 58748730 | 88 | 16 | 51292 | 0.4064 | 0.34222 | *PSMA3* |
| ENSG00000153707 | 9 | 8304246 | 10622723 | 11142 | 609 | 43721 | 0.40615 | 0.34231 | *PTPRD* |
| ENSG00000117226 | 1 | 89462349 | 89498577 | 85 | 12 | 29465 | 0.40615 | 0.34232 | *GBP3* |
| ENSG00000078399 | 7 | 27192054 | 27220117 | 51 | 17 | 43114 | 0.40613 | 0.34232 | *HOXA9* |
| ENSG00000119403 | 9 | 123607977 | 123649606 | 98 | 15 | 48129 | 0.40587 | 0.34242 | *PHF19* |
| ENSG00000113851 | 3 | 3180676 | 3231394 | 212 | 30 | 36438 | 0.40568 | 0.34249 | *CRBN* |
| ENSG00000178498 | 12 | 57988405 | 58013587 | 43 | 16 | 43527 | 0.40568 | 0.34249 | *DTX3* |
| ENSG00000164237 | 5 | 10265987 | 10318138 | 189 | 27 | 24898 | 0.40553 | 0.34255 | *CMBL* |
| ENSG00000173250 | 5 | 145882666 | 145905753 | 71 | 15 | 41612 | 0.40544 | 0.34258 | *GPR151* |
| ENSG00000187446 | 15 | 41513037 | 41584043 | 213 | 20 | 33332 | 0.40525 | 0.34265 | *CHP1* |
| ENSG00000128789 | 18 | 12648737 | 12735739 | 280 | 23 | 38779 | 0.40503 | 0.34273 | *PSMG2* |
| ENSG00000117528 | 1 | 94873933 | 94994222 | 158 | 23 | 40988 | 0.40483 | 0.3428 | *ABCD3* |
| ENSG00000116679 | 1 | 185255520 | 185296461 | 53 | 14 | 49314 | 0.4042 | 0.34303 | *IVNS1ABP* |
| ENSG00000267802 | 14 | 52426014 | 52446247 | 55 | 8 | 31045 | 0.40399 | 0.34311 | *AL358333.1* |
| ENSG00000101190 | 20 | 61462467 | 61503115 | 207 | 27 | 42877 | 0.40366 | 0.34323 | *TCFL5* |
| ENSG00000160360 | 9 | 139211932 | 139264057 | 202 | 38 | 51016 | 0.40347 | 0.3433 | *GPSM1* |
| ENSG00000197857 | 19 | 12325501 | 12415702 | 238 | 16 | 43808 | 0.4033 | 0.34336 | *ZNF44* |
| ENSG00000163930 | 3 | 52425029 | 52454366 | 28 | 8 | 55014 | 0.40323 | 0.34339 | *BAP1* |
| ENSG00000131584 | 1 | 1217756 | 1254989 | 91 | 25 | 37877 | 0.4032 | 0.3434 | *ACAP3* |
| ENSG00000161055 | 5 | 180007103 | 180028540 | 77 | 21 | 48926 | 0.40317 | 0.34341 | *SCGB3A1* |
| ENSG00000143502 | 1 | 223384161 | 223547544 | 416 | 44 | 42122 | 0.40314 | 0.34342 | *SUSD4* |
| ENSG00000175854 | 9 | 131027658 | 131061269 | 77 | 13 | 35335 | 0.40309 | 0.34344 | *SWI5* |
| ENSG00000182795 | 1 | 207181866 | 207216101 | 61 | 9 | 51477 | 0.4029 | 0.34351 | *C1orf116* |
| ENSG00000175311 | 16 | 21234986 | 21273750 | 77 | 27 | 43439 | 0.40279 | 0.34355 | *ANKS4B* |
| ENSG00000108018 | 10 | 108323421 | 108934292 | 1798 | 191 | 41010 | 0.40252 | 0.34365 | *SORCS1* |
| ENSG00000106780 | 9 | 123353091 | 123486748 | 185 | 15 | 42181 | 0.40222 | 0.34376 | *MEGF9* |
| ENSG00000135773 | 1 | 230873130 | 230947749 | 355 | 55 | 38282 | 0.40213 | 0.34379 | *CAPN9* |
| ENSG00000196171 | 1 | 158659369 | 158680486 | 52 | 10 | 40084 | 0.40212 | 0.3438 | *OR6K2* |

| | | | | | | | | | |
|---|---|---|---|---|---|---|---|---|---|
| ENSG00000170456 | 12 | 31525157 | 31754031 | 983 | 56 | 47468 | 0.40203 | 0.34383 | *DENND5B* |
| ENSG00000116584 | 1 | 155906630 | 155986861 | 128 | 21 | 33133 | 0.40172 | 0.34395 | *ARHGEF2* |
| ENSG00000177225 | 11 | 757220 | 787488 | 86 | 17 | 37670 | 0.40151 | 0.34402 | *PDDC1* |
| ENSG00000106554 | 7 | 132459629 | 132776848 | 660 | 49 | 48585 | 0.40137 | 0.34407 | *CHCHD3* |
| ENSG00000116157 | 1 | 53058044 | 53084723 | 79 | 13 | 51043 | 0.40126 | 0.34411 | *GPX7* |
| ENSG00000142864 | 1 | 67863493 | 67906098 | 68 | 13 | 37660 | 0.40124 | 0.34412 | *SERBP1* |
| ENSG00000250254 | 4 | 37952056 | 37972746 | 90 | 12 | 41229 | 0.40102 | 0.3442 | *PTTG2* |
| ENSG00000139531 | 12 | 56380964 | 56410425 | 49 | 13 | 41674 | 0.40103 | 0.3442 | *SUOX* |
| ENSG00000121274 | 16 | 50176829 | 50279221 | 231 | 30 | 41822 | 0.4008 | 0.34428 | *PAPD5* |
| ENSG00000163958 | 3 | 195914320 | 195948308 | 197 | 34 | 51546 | 0.40064 | 0.34434 | *ZDHHC19* |
| ENSG00000143393 | 1 | 151254273 | 151310191 | 67 | 14 | 49100 | 0.40039 | 0.34443 | *PI4KB* |
| ENSG00000196407 | 1 | 151809739 | 151836173 | 81 | 18 | 37406 | 0.4004 | 0.34443 | *THEM5* |
| ENSG00000124788 | 6 | 16289343 | 16771722 | 1555 | 238 | 44791 | 0.40035 | 0.34445 | *ATXN1* |
| ENSG00000129204 | 17 | 5009733 | 5088329 | 191 | 26 | 39778 | 0.40033 | 0.34446 | *USP6* |
| ENSG00000185775 | 9 | 43614507 | 43640730 | 5 | 2 | 42759 | 0.40012 | 0.34454 | *SPATA31A6* |
| ENSG00000267981 | 11 | 129926836 | 129954012 | 121 | 13 | 41020 | 0.40005 | 0.34456 | *AP003041.2* |
| ENSG00000099864 | 19 | 698953 | 758329 | 279 | 65 | 45313 | 0.39994 | 0.3446 | *PALM* |
| ENSG00000269071 | 5 | 138842108 | 138862506 | 26 | 7 | 36318 | 0.3999 | 0.34462 | *AC138517.1* |
| ENSG00000170779 | 14 | 105465910 | 105497485 | 138 | 11 | 32754 | 0.39985 | 0.34463 | *CDCA4* |
| ENSG00000136378 | 15 | 79041545 | 79113773 | 302 | 31 | 42803 | 0.39944 | 0.34478 | *ADAMTS7* |
| ENSG00000204345 | 17 | 72565504 | 72598422 | 146 | 17 | 40541 | 0.39944 | 0.34479 | *CD300LD* |
| ENSG00000144057 | 2 | 107408056 | 107513564 | 368 | 49 | 42294 | 0.39935 | 0.34482 | *ST6GAL2* |
| ENSG00000145192 | 3 | 186320712 | 186349107 | 103 | 17 | 47885 | 0.39935 | 0.34482 | *AHSG* |
| ENSG00000268925 | 10 | 135331026 | 135352661 | 78 | 16 | 32919 | 0.39922 | 0.34486 | *AL161645.2* |
| ENSG00000119943 | 10 | 100133322 | 100184941 | 223 | 15 | 30147 | 0.39907 | 0.34492 | *PYROXD2* |
| ENSG00000161981 | 16 | 93010 | 117669 | 75 | 12 | 47363 | 0.39904 | 0.34493 | *SNRNP25* |
| ENSG00000154016 | 17 | 18913986 | 18960950 | 28 | 14 | 34037 | 0.39882 | 0.34501 | *GRAP* |
| ENSG00000165655 | 10 | 77147588 | 77171664 | 24 | 14 | 45082 | 0.39874 | 0.34504 | *ZNF503* |
| ENSG00000131142 | 19 | 8107651 | 8137534 | 147 | 28 | 48703 | 0.39874 | 0.34504 | *CCL25* |
| ENSG00000189275 | 10 | 133594734 | 133632535 | 136 | 17 | 55267 | 0.39859 | 0.3451 | *AL450307.1* |
| ENSG00000184502 | 17 | 39858578 | 39882221 | 126 | 13 | 34956 | 0.39856 | 0.34511 | *GAST* |
| ENSG00000083123 | 6 | 80806364 | 81065987 | 845 | 44 | 37246 | 0.39852 | 0.34512 | *BCKDHB* |
| ENSG00000107854 | 10 | 93548069 | 93635033 | 235 | 20 | 45741 | 0.39853 | 0.34512 | *TNKS2* |
| ENSG00000231500 | 6 | 33229787 | 33254287 | 87 | 14 | 33427 | 0.39838 | 0.34518 | *RPS18* |
| ENSG00000168016 | 3 | 36858311 | 36996548 | 328 | 35 | 38318 | 0.39759 | 0.34547 | *TRANK1* |
| ENSG00000134955 | 11 | 124922963 | 124969131 | 223 | 42 | 47094 | 0.39752 | 0.34549 | *SLC37A2* |

| | | | | | | | | | |
|---|---|---|---|---|---|---|---|---|---|
| ENSG00000243207 | 19 | 10206899 | 10235456 | 79 | 22 | 50835 | 0.39753 | 0.34549 | *PPAN-P2RY11* |
| ENSG00000167920 | 17 | 38965358 | 39002522 | 118 | 20 | 41748 | 0.3974 | 0.34554 | *TMEM99* |
| ENSG00000138642 | 4 | 89289891 | 89374263 | 102 | 35 | 42642 | 0.39727 | 0.34558 | *HERC6* |
| ENSG00000164296 | 5 | 149362681 | 149390730 | 62 | 10 | 41163 | 0.39727 | 0.34559 | *TIGD6* |
| ENSG00000134262 | 1 | 114427370 | 114457823 | 54 | 10 | 35018 | 0.39713 | 0.34563 | *AP4B1* |
| ENSG00000141449 | 18 | 18812203 | 19115378 | 375 | 41 | 36032 | 0.39687 | 0.34573 | *GREB1L* |
| ENSG00000185015 | 8 | 86122816 | 86206302 | 139 | 15 | 52884 | 0.39669 | 0.3458 | *CA13* |
| ENSG00000094804 | 17 | 38433885 | 38469171 | 39 | 8 | 45833 | 0.3966 | 0.34583 | *CDC6* |
| ENSG00000214670 | 14 | 75148879 | 75189800 | 61 | 7 | 41966 | 0.39655 | 0.34585 | *AC007956.1* |
| ENSG00000139793 | 13 | 97863688 | 98056374 | 466 | 65 | 42271 | 0.39651 | 0.34587 | *MBNL2* |
| ENSG00000161996 | 16 | 689311 | 727833 | 88 | 15 | 31049 | 0.39641 | 0.3459 | *WDR90* |
| ENSG00000118503 | 6 | 138178351 | 138214449 | 79 | 14 | 38482 | 0.39608 | 0.34602 | *TNFAIP3* |
| ENSG00000172936 | 3 | 38169969 | 38194513 | 36 | 8 | 41476 | 0.39602 | 0.34604 | *MYD88* |
| ENSG00000085276 | 3 | 168791287 | 169391406 | 1948 | 217 | 44877 | 0.39597 | 0.34606 | *MECOM* |
| ENSG00000092421 | 5 | 115769312 | 115920630 | 560 | 110 | 45965 | 0.39593 | 0.34608 | *SEMA6A* |
| ENSG00000183798 | 20 | 39978606 | 40005467 | 57 | 13 | 47152 | 0.39584 | 0.34611 | *EMILIN3* |
| ENSG00000174738 | 3 | 23976751 | 24032109 | 211 | 23 | 45141 | 0.39577 | 0.34614 | *NR1D2* |
| ENSG00000153291 | 6 | 46610678 | 46655930 | 69 | 15 | 49163 | 0.3955 | 0.34624 | *SLC25A27* |
| ENSG00000170373 | 20 | 23718190 | 23741905 | 178 | 21 | 46066 | 0.39549 | 0.34624 | *CST1* |
| ENSG00000075290 | 10 | 102212798 | 102253501 | 83 | 5 | 54630 | 0.39533 | 0.3463 | *WNT8B* |
| ENSG00000114503 | 3 | 196652273 | 196679468 | 90 | 16 | 34663 | 0.39529 | 0.34631 | *NCBP2* |
| ENSG00000146755 | 7 | 72716535 | 72752085 | 50 | 15 | 46714 | 0.39513 | 0.34637 | *TRIM50* |
| ENSG00000197448 | 7 | 142931186 | 142977947 | 28 | 14 | 54252 | 0.39509 | 0.34639 | *GSTK1* |
| ENSG00000112304 | 6 | 24657263 | 24715293 | 205 | 30 | 46419 | 0.39507 | 0.3464 | *ACOT13* |
| ENSG00000163600 | 2 | 204791471 | 204836300 | 185 | 29 | 39995 | 0.39494 | 0.34644 | *ICOS* |
| ENSG00000103855 | 15 | 73966307 | 74016859 | 165 | 25 | 42526 | 0.39494 | 0.34645 | *CD276* |
| ENSG00000104371 | 8 | 42221586 | 42244750 | 49 | 12 | 35442 | 0.39472 | 0.34652 | *DKK4* |
| ENSG00000164930 | 8 | 104300661 | 104355094 | 207 | 21 | 53233 | 0.39469 | 0.34653 | *FZD6* |
| ENSG00000112851 | 5 | 65212303 | 65388377 | 240 | 26 | 35505 | 0.39429 | 0.34668 | *ERBB2IP* |
| ENSG00000100413 | 22 | 41911808 | 41950610 | 84 | 9 | 49203 | 0.39423 | 0.34671 | *POLR3H* |
| ENSG00000215906 | 1 | 23269536 | 23301831 | 105 | 19 | 40040 | 0.39407 | 0.34677 | *LACTBL1* |
| ENSG00000120256 | 6 | 150129934 | 150196199 | 135 | 13 | 36431 | 0.39388 | 0.34683 | *LRP11* |
| ENSG00000182963 | 17 | 42865816 | 42918184 | 139 | 24 | 50058 | 0.39375 | 0.34688 | *GJC1* |
| ENSG00000042781 | 1 | 215786236 | 216606738 | 2514 | 248 | 49192 | 0.39369 | 0.34691 | *USH2A* |
| ENSG00000161544 | 17 | 74513438 | 74557257 | 126 | 25 | 40687 | 0.39366 | 0.34691 | *CYGB* |
| ENSG00000091010 | 5 | 145708587 | 145730083 | 61 | 11 | 45251 | 0.39336 | 0.34703 | *POU4F3* |

| | | | | | | | | | |
|---|---|---|---|---|---|---|---|---|---|
| ENSG00000205213 | 11 | 27377508 | 27504322 | 277 | 48 | 41762 | 0.39327 | 0.34706 | *LGR4* |
| ENSG00000050628 | 1 | 71308036 | 71523491 | 683 | 75 | 49510 | 0.39317 | 0.3471 | *PTGER3* |
| ENSG00000051009 | 11 | 6222565 | 6265941 | 99 | 16 | 33826 | 0.39301 | 0.34715 | *FAM160A2* |
| ENSG00000188511 | 22 | 49798176 | 50061190 | 1103 | 131 | 40642 | 0.39294 | 0.34718 | *C22orf34* |
| ENSG00000144161 | 2 | 112959102 | 113022713 | 103 | 16 | 50317 | 0.39287 | 0.34721 | *ZC3H8* |
| ENSG00000179627 | 14 | 105256933 | 105281049 | 52 | 16 | 34980 | 0.39259 | 0.34731 | *ZBTB42* |
| ENSG00000217128 | 5 | 130967407 | 131142710 | 277 | 21 | 34697 | 0.39248 | 0.34735 | *FNIP1* |
| ENSG00000204661 | 5 | 179058545 | 179082047 | 76 | 21 | 44364 | 0.39245 | 0.34736 | *C5orf60* |
| ENSG00000127774 | 17 | 3562109 | 3582962 | 101 | 15 | 51777 | 0.39194 | 0.34755 | *EMC6* |
| ENSG00000269858 | 19 | 41294901 | 41324338 | 121 | 24 | 46828 | 0.39193 | 0.34755 | *EGLN2* |
| ENSG00000121940 | 1 | 109462130 | 109516111 | 231 | 34 | 46182 | 0.39188 | 0.34757 | *CLCC1* |
| ENSG00000105613 | 19 | 12934765 | 12995765 | 145 | 16 | 40260 | 0.39181 | 0.3476 | *MAST1* |
| ENSG00000065413 | 2 | 197821741 | 198185897 | 1117 | 98 | 43248 | 0.39147 | 0.34772 | *ANKRD44* |
| ENSG00000106077 | 7 | 73140424 | 73163197 | 57 | 13 | 38238 | 0.3912 | 0.34783 | *ABHD11* |
| ENSG00000154611 | 18 | 23703816 | 23783319 | 192 | 18 | 34342 | 0.39109 | 0.34786 | *PSMA8* |
| ENSG00000158987 | 5 | 130749614 | 130980929 | 377 | 23 | 36582 | 0.39093 | 0.34792 | *RAPGEF6* |
| ENSG00000158270 | 18 | 309361 | 510722 | 987 | 146 | 49243 | 0.39089 | 0.34794 | *COLEC12* |
| ENSG00000187091 | 3 | 38038987 | 38081253 | 139 | 26 | 49031 | 0.39087 | 0.34795 | *PLCD1* |
| ENSG00000144455 | 3 | 3732498 | 4518965 | 3341 | 205 | 34390 | 0.39084 | 0.34796 | *SUMF1* |
| ENSG00000269408 | 9 | 138140512 | 138161275 | 78 | 32 | 40554 | 0.39083 | 0.34796 | *AL390778.1* |
| ENSG00000143303 | 1 | 156688234 | 156716752 | 57 | 16 | 41427 | 0.39063 | 0.34803 | *RRNAD1* |
| ENSG00000110934 | 12 | 51664822 | 51728452 | 237 | 26 | 45895 | 0.39035 | 0.34814 | *BIN2* |
| ENSG00000180386 | 17 | 39421911 | 39442949 | 38 | 5 | 26681 | 0.39032 | 0.34815 | *KRTAP9-7* |
| ENSG00000178694 | 3 | 93771760 | 93857389 | 82 | 25 | 41655 | 0.39026 | 0.34817 | *NSUN3* |
| ENSG00000118292 | 1 | 150230600 | 150263327 | 49 | 12 | 36182 | 0.39021 | 0.34819 | *C1orf54* |
| ENSG00000185774 | 4 | 20720239 | 21960422 | 5071 | 315 | 46118 | 0.39009 | 0.34824 | *KCNIP4* |
| ENSG00000008405 | 12 | 107375142 | 107497607 | 340 | 29 | 42568 | 0.39005 | 0.34825 | *CRY1* |
| ENSG00000099940 | 22 | 21203271 | 21255506 | 159 | 14 | 53637 | 0.39005 | 0.34825 | *SNAP29* |
| ENSG00000169299 | 4 | 37818255 | 37874558 | 254 | 25 | 30118 | 0.38973 | 0.34837 | *PGM2* |
| ENSG00000119718 | 14 | 75459614 | 75486292 | 52 | 6 | 46893 | 0.38952 | 0.34845 | *EIF2B2* |
| ENSG00000160213 | 21 | 45182393 | 45206326 | 92 | 23 | 42539 | 0.38951 | 0.34845 | *CSTB* |
| ENSG00000172543 | 11 | 65637280 | 65661212 | 51 | 9 | 33858 | 0.38911 | 0.3486 | *CTSW* |
| ENSG00000102890 | 16 | 67223014 | 67247932 | 45 | 9 | 32892 | 0.38893 | 0.34866 | *ELMO3* |
| ENSG00000118162 | 19 | 47968401 | 47997525 | 39 | 10 | 43194 | 0.38888 | 0.34868 | *KPTN* |
| ENSG00000101333 | 20 | 9039410 | 9471889 | 1012 | 114 | 45994 | 0.38866 | 0.34876 | *PLCB4* |
| ENSG00000163645 | 3 | 150367672 | 150431758 | 212 | 41 | 40285 | 0.38835 | 0.34888 | *FAM194A* |

| | | | | | | | | | |
|---|---|---|---|---|---|---|---|---|---|
| ENSG00000143398 | 1 | 151160425 | 151232012 | 107 | 15 | 48703 | 0.38823 | 0.34892 | *PIP5K1A* |
| ENSG00000138029 | 2 | 26456038 | 26523336 | 125 | 11 | 51278 | 0.38823 | 0.34892 | *HADHB* |
| ENSG00000135976 | 2 | 97769233 | 97940258 | 86 | 25 | 38564 | 0.38807 | 0.34898 | *ANKRD36* |
| ENSG00000150991 | 12 | 125386150 | 125411914 | 61 | 15 | 43787 | 0.38806 | 0.34899 | *UBC* |
| ENSG00000120055 | 10 | 104199594 | 104221300 | 55 | 19 | 50031 | 0.38795 | 0.34903 | *C10orf95* |
| ENSG00000221977 | 14 | 22123297 | 22144238 | 112 | 17 | 50434 | 0.38776 | 0.3491 | *OR4E2* |
| ENSG00000150269 | 11 | 56219945 | 56240877 | 74 | 13 | 50394 | 0.38747 | 0.3492 | *OR5M9* |
| ENSG00000187961 | 1 | 885967 | 911095 | 93 | 19 | 38736 | 0.38731 | 0.34926 | *KLHL17* |
| ENSG00000181004 | 4 | 123643857 | 123676098 | 81 | 10 | 40054 | 0.38723 | 0.34929 | *BBS12* |
| ENSG00000114923 | 2 | 220482049 | 220516702 | 49 | 15 | 45087 | 0.38709 | 0.34935 | *SLC4A3* |
| ENSG00000146122 | 6 | 39750142 | 39882648 | 446 | 72 | 43362 | 0.38703 | 0.34937 | *DAAM2* |
| ENSG00000185668 | 1 | 38499523 | 38522450 | 57 | 10 | 51988 | 0.38699 | 0.34938 | *POU3F1* |
| ENSG00000110245 | 11 | 116690422 | 116713788 | 105 | 20 | 30879 | 0.38686 | 0.34943 | *APOC3* |
| ENSG00000087303 | 14 | 52461521 | 52545712 | 437 | 47 | 43828 | 0.38687 | 0.34943 | *NID2* |
| ENSG00000166278 | 6 | 31855562 | 31923449 | 208 | 39 | 41143 | 0.38679 | 0.34946 | *C2* |
| ENSG00000181381 | 4 | 169267886 | 169468937 | 892 | 99 | 44803 | 0.38668 | 0.34949 | *DDX60L* |
| ENSG00000205364 | 16 | 56656145 | 56677898 | 118 | 19 | 40410 | 0.38644 | 0.34958 | *MT1M* |
| ENSG00000268171 | 4 | 57266682 | 57287131 | 63 | 10 | 51832 | 0.38629 | 0.34964 | *AC068620.1* |
| ENSG00000132464 | 4 | 71484461 | 71562533 | 112 | 24 | 36398 | 0.38628 | 0.34964 | *ENAM* |
| ENSG00000205853 | 22 | 32745893 | 32788039 | 230 | 34 | 41016 | 0.38627 | 0.34965 | *RFPL3S* |
| ENSG00000137628 | 4 | 169127444 | 169249958 | 383 | 37 | 49742 | 0.38622 | 0.34967 | *DDX60* |
| ENSG00000166579 | 17 | 8306449 | 8403729 | 192 | 26 | 53677 | 0.386 | 0.34975 | *NDEL1* |
| ENSG00000243543 | 20 | 44152835 | 44178134 | 50 | 8 | 47993 | 0.38599 | 0.34975 | *WFDC6* |
| ENSG00000165502 | 14 | 50075237 | 50097403 | 80 | 9 | 41246 | 0.38598 | 0.34976 | *RPL36AL* |
| ENSG00000041982 | 9 | 117772806 | 117890536 | 397 | 57 | 53174 | 0.3859 | 0.34979 | *TNC* |
| ENSG00000198355 | 22 | 50344161 | 50367728 | 59 | 21 | 43604 | 0.38583 | 0.34981 | *PIM3* |
| ENSG00000174405 | 13 | 108849787 | 108880716 | 95 | 18 | 50351 | 0.3858 | 0.34982 | *LIG4* |
| ENSG00000160991 | 7 | 102063553 | 102107268 | 125 | 26 | 48799 | 0.38577 | 0.34983 | *ORAI2* |
| ENSG00000185088 | 15 | 63408071 | 63460220 | 189 | 23 | 37168 | 0.38574 | 0.34985 | *RPS27L* |
| ENSG00000184302 | 14 | 60965669 | 60989568 | 51 | 8 | 27574 | 0.38562 | 0.34989 | *SIX6* |
| ENSG00000136122 | 13 | 73292061 | 73340336 | 113 | 26 | 50395 | 0.3856 | 0.3499 | *BORA* |
| ENSG00000173641 | 1 | 16330523 | 16356089 | 151 | 17 | 55159 | 0.38554 | 0.34992 | *HSPB7* |
| ENSG00000066185 | 1 | 42886000 | 42931938 | 149 | 14 | 47598 | 0.38517 | 0.35005 | *ZMYND12* |
| ENSG00000166897 | 22 | 37754000 | 37833505 | 325 | 49 | 44905 | 0.38517 | 0.35006 | *ELFN2* |
| ENSG00000146232 | 6 | 44215903 | 44243500 | 71 | 21 | 51892 | 0.38498 | 0.35012 | *NFKBIE* |
| ENSG00000136159 | 13 | 48601703 | 48631358 | 101 | 21 | 46528 | 0.38494 | 0.35014 | *NUDT15* |

| | | | | | | | | | |
|---|---|---|---|---|---|---|---|---|---|
| ENSG00000114796 | 3 | 183343356 | 183412265 | 265 | 7 | 49817 | 0.38486 | 0.35017 | *KLHL24* |
| ENSG00000152591 | 4 | 88519681 | 88548062 | 84 | 14 | 44812 | 0.38484 | 0.35018 | *DSPP* |
| ENSG00000147471 | 8 | 37610111 | 37647283 | 23 | 13 | 55979 | 0.38455 | 0.35029 | *PROSC* |
| ENSG00000180398 | 2 | 47119009 | 47178994 | 289 | 35 | 37579 | 0.38449 | 0.35031 | *MCFD2* |
| ENSG00000180532 | 19 | 58170303 | 58200520 | 88 | 10 | 50500 | 0.38442 | 0.35033 | *ZSCAN4* |
| ENSG00000018408 | 3 | 149225022 | 149464501 | 838 | 110 | 47769 | 0.38437 | 0.35035 | *WWTR1* |
| ENSG00000205126 | 11 | 44059531 | 44091527 | 112 | 19 | 41075 | 0.38399 | 0.35049 | *ACCSL* |
| ENSG00000131015 | 6 | 150253136 | 150280371 | 42 | 16 | 38831 | 0.38367 | 0.35061 | *ULBP2* |
| ENSG00000128573 | 7 | 113716382 | 114343827 | 1103 | 102 | 38199 | 0.38339 | 0.35071 | *FOXP2* |
| ENSG00000172757 | 11 | 65580493 | 65639497 | 138 | 16 | 29806 | 0.38341 | 0.35071 | *CFL1* |
| ENSG00000137411 | 6 | 30866019 | 30904236 | 176 | 18 | 41711 | 0.38326 | 0.35076 | *VARS2* |
| ENSG00000132821 | 20 | 36521499 | 36583752 | 185 | 36 | 44994 | 0.38263 | 0.351 | *VSTM2L* |
| ENSG00000084674 | 2 | 21214301 | 21276945 | 120 | 30 | 34144 | 0.38248 | 0.35105 | *APOB* |
| ENSG00000129235 | 17 | 6534078 | 6557861 | 104 | 14 | 42641 | 0.38214 | 0.35118 | *TXNDC17* |
| ENSG00000185737 | 10 | 83625070 | 84756935 | 3540 | 268 | 42616 | 0.38193 | 0.35126 | *NRG3* |
| ENSG00000095319 | 9 | 131699978 | 131779375 | 90 | 10 | 42332 | 0.38163 | 0.35137 | *NUP188* |
| ENSG00000162873 | 1 | 205295220 | 205336218 | 207 | 28 | 52234 | 0.38144 | 0.35144 | *KLHDC8A* |
| ENSG00000127415 | 4 | 970785 | 1008316 | 142 | 33 | 51758 | 0.3807 | 0.35171 | *IDUA* |
| ENSG00000253633 | 8 | 103530968 | 103560896 | 188 | 28 | 50691 | 0.38072 | 0.35171 | *KB-1980E6.3* |
| ENSG00000170191 | 20 | 25583571 | 25614811 | 97 | 10 | 28989 | 0.38038 | 0.35183 | *NANP* |
| ENSG00000204544 | 6 | 30941495 | 30967680 | 173 | 16 | 50826 | 0.38034 | 0.35185 | *MUC21* |
| ENSG00000268674 | 1 | 16855561 | 16876070 | 33 | 7 | 44119 | 0.38018 | 0.3519 | *FAM231B* |
| ENSG00000184922 | 17 | 43288811 | 43334687 | 111 | 23 | 37148 | 0.38012 | 0.35193 | *FMNL1* |
| ENSG00000206043 | 18 | 71973048 | 72036422 | 268 | 22 | 48427 | 0.37998 | 0.35198 | *C18orf63* |
| ENSG00000272162 | 6 | 10738027 | 10940656 | 524 | 90 | 48652 | 0.37973 | 0.35207 | *RP11-637O19.3* |
| ENSG00000099256 | 10 | 25127536 | 25251533 | 459 | 63 | 42384 | 0.37947 | 0.35217 | *PRTFDC1* |
| ENSG00000111911 | 6 | 126267927 | 126311387 | 61 | 5 | 53591 | 0.37945 | 0.35218 | *HINT3* |
| ENSG00000240038 | 1 | 104086437 | 104132156 | 68 | 22 | 49797 | 0.37928 | 0.35224 | *AMY2B* |
| ENSG00000196313 | 7 | 72339936 | 72431979 | 72 | 36 | 43998 | 0.37919 | 0.35227 | *POM121* |
| ENSG00000114770 | 3 | 183627722 | 183745803 | 351 | 32 | 51882 | 0.37918 | 0.35228 | *ABCC5* |
| ENSG00000108231 | 10 | 95507566 | 95567916 | 170 | 39 | 44078 | 0.37908 | 0.35231 | *LGI1* |
| ENSG00000198873 | 10 | 120957101 | 121225131 | 978 | 128 | 43395 | 0.37894 | 0.35237 | *GRK5* |
| ENSG00000184385 | 21 | 43512244 | 43538644 | 168 | 23 | 49850 | 0.37887 | 0.35239 | *C21orf128* |
| ENSG00000164893 | 8 | 87216281 | 87343375 | 392 | 40 | 35462 | 0.37879 | 0.35242 | *SLC7A13* |
| ENSG00000005379 | 17 | 56368592 | 56416152 | 148 | 26 | 53965 | 0.37871 | 0.35245 | *BZRAP1* |
| ENSG00000158816 | 1 | 20607412 | 20691387 | 336 | 43 | 45193 | 0.37846 | 0.35254 | *VWA5B1* |

| | | | | | | | | | |
|---|---|---|---|---|---|---|---|---|---|
| ENSG00000178568 | 2 | 212230446 | 213413565 | 4795 | 280 | 42765 | 0.37832 | 0.3526 | *ERBB4* |
| ENSG00000173473 | 3 | 47616762 | 47833596 | 285 | 22 | 37673 | 0.37817 | 0.35265 | *SMARCC1* |
| ENSG00000138135 | 10 | 90955694 | 90977071 | 36 | 12 | 44655 | 0.37814 | 0.35266 | *CH25H* |
| ENSG00000224531 | 6 | 11084266 | 11148964 | 268 | 30 | 50483 | 0.3779 | 0.35275 | *SMIM13* |
| ENSG00000243716 | 16 | 22480442 | 22557842 | 8 | 5 | 43377 | 0.3779 | 0.35275 | *NPIPB5* |
| ENSG00000183684 | 17 | 79835713 | 79859462 | 29 | 12 | 52082 | 0.37783 | 0.35278 | *ALYREF* |
| ENSG00000148925 | 11 | 13399548 | 13494844 | 289 | 22 | 44513 | 0.37777 | 0.3528 | *BTBD10* |
| ENSG00000203942 | 10 | 99339450 | 99360690 | 55 | 12 | 48865 | 0.37762 | 0.35285 | *C10orf62* |
| ENSG00000128951 | 15 | 48613208 | 48645570 | 51 | 8 | 44886 | 0.37754 | 0.35289 | *DUT* |
| ENSG00000168564 | 4 | 184355744 | 184379351 | 142 | 12 | 30816 | 0.37744 | 0.35292 | *CDKN2AIP* |
| ENSG00000090530 | 3 | 189664517 | 189850226 | 768 | 72 | 47368 | 0.37732 | 0.35297 | *LEPREL1* |
| ENSG00000164107 | 4 | 174436120 | 174461380 | 54 | 13 | 53891 | 0.37732 | 0.35297 | *HAND2* |
| ENSG00000135905 | 2 | 225619807 | 225917162 | 855 | 81 | 43635 | 0.37716 | 0.35303 | *DOCK10* |
| ENSG00000270999 | 2 | 90448201 | 90468671 | 1 | 1 | 43032 | 0.37696 | 0.3531 | *CH17-132F21.1* |
| ENSG00000151366 | 11 | 77769350 | 77801265 | 131 | 14 | 53649 | 0.37694 | 0.35311 | *NDUFC2* |
| ENSG00000133983 | 14 | 70781798 | 70836448 | 203 | 24 | 45217 | 0.37663 | 0.35322 | *COX16* |
| ENSG00000203908 | 6 | 74062400 | 74083894 | 67 | 18 | 54904 | 0.37646 | 0.35329 | *KHDC3L* |
| ENSG00000155714 | 16 | 21985186 | 22022433 | 39 | 15 | 44825 | 0.37638 | 0.35332 | *PDZD9* |
| ENSG00000160801 | 3 | 46909236 | 46955287 | 93 | 23 | 44045 | 0.37632 | 0.35334 | *PTH1R* |
| ENSG00000141873 | 19 | 2722202 | 2750150 | 109 | 19 | 37650 | 0.37631 | 0.35334 | *SLC39A3* |
| ENSG00000166262 | 15 | 49609159 | 49923128 | 1107 | 51 | 44127 | 0.37606 | 0.35344 | *FAM227B* |
| ENSG00000078579 | 8 | 16839678 | 16869690 | 222 | 35 | 45036 | 0.37602 | 0.35345 | *FGF20* |
| ENSG00000177463 | 3 | 14979091 | 15105107 | 295 | 33 | 53630 | 0.37588 | 0.3535 | *NR2C2* |
| ENSG00000163638 | 3 | 64491333 | 64683676 | 491 | 77 | 34371 | 0.3758 | 0.35353 | *ADAMTS9* |
| ENSG00000134287 | 12 | 49319506 | 49361334 | 146 | 12 | 48159 | 0.37542 | 0.35367 | *ARF3* |
| ENSG00000124257 | 20 | 44507264 | 44529926 | 60 | 12 | 36849 | 0.37536 | 0.3537 | *NEURL2* |
| ENSG00000107554 | 10 | 101625334 | 101779676 | 560 | 42 | 48599 | 0.37524 | 0.35374 | *DNMBP* |
| ENSG00000155438 | 2 | 122474521 | 122504499 | 55 | 8 | 26419 | 0.37508 | 0.3538 | *NIFK* |
| ENSG00000131746 | 17 | 38622080 | 38667849 | 129 | 30 | 45558 | 0.37506 | 0.35381 | *TNS4* |
| ENSG00000166948 | 20 | 2351554 | 2423399 | 314 | 44 | 38204 | 0.37494 | 0.35385 | *TGM6* |
| ENSG00000232237 | 1 | 201073081 | 201106312 | 141 | 26 | 42768 | 0.37482 | 0.3539 | *ASCL5* |
| ENSG00000203837 | 10 | 118177379 | 118247469 | 197 | 26 | 38777 | 0.37454 | 0.354 | *PNLIPRP3* |
| ENSG00000253313 | 1 | 43737554 | 43761288 | 55 | 18 | 43060 | 0.37442 | 0.35404 | *C1orf210* |
| ENSG00000089127 | 12 | 113334582 | 113379990 | 165 | 7 | 25937 | 0.37438 | 0.35406 | *OAS1* |
| ENSG00000170464 | 5 | 138734279 | 138790180 | 63 | 9 | 30399 | 0.37437 | 0.35407 | *DNAJC18* |
| ENSG00000111344 | 12 | 113526624 | 113584044 | 248 | 24 | 50987 | 0.37436 | 0.35407 | *RASAL1* |

| | | | | | | | | | |
|---|---|---|---|---|---|---|---|---|---|
| ENSG00000104432 | 8 | 79577978 | 79727758 | 397 | 37 | 48553 | 0.37429 | 0.3541 | *IL7* |
| ENSG00000124766 | 6 | 21583972 | 21608847 | 28 | 13 | 43444 | 0.37405 | 0.35418 | *SOX4* |
| ENSG00000182187 | 2 | 208997297 | 209020892 | 106 | 16 | 45346 | 0.37348 | 0.35439 | *CRYGB* |
| ENSG00000138443 | 2 | 204182942 | 204322446 | 241 | 32 | 33074 | 0.37343 | 0.35441 | *ABI2* |
| ENSG00000109511 | 4 | 169003666 | 169118841 | 329 | 39 | 46844 | 0.37341 | 0.35442 | *ANXA10* |
| ENSG00000103496 | 16 | 31034210 | 31064296 | 42 | 6 | 22216 | 0.37336 | 0.35444 | *STX4* |
| ENSG00000171634 | 17 | 65811640 | 65990494 | 346 | 21 | 32274 | 0.37336 | 0.35444 | *BPTF* |
| ENSG00000257955 | 12 | 48403554 | 48429165 | 107 | 18 | 41355 | 0.37333 | 0.35445 | *RP1-228P16.5* |
| ENSG00000172638 | 11 | 65623912 | 65651063 | 65 | 10 | 29885 | 0.3733 | 0.35446 | *EFEMP2* |
| ENSG00000165181 | 9 | 114438453 | 114567288 | 545 | 35 | 42625 | 0.3731 | 0.35454 | *C9orf84* |
| ENSG00000180329 | 17 | 42740437 | 42777147 | 63 | 18 | 48484 | 0.37308 | 0.35455 | *CCDC43* |
| ENSG00000204104 | 2 | 239219082 | 239319541 | 322 | 28 | 42143 | 0.37303 | 0.35456 | *TRAF3IP1* |
| ENSG00000153956 | 7 | 81565760 | 82083114 | 1780 | 198 | 47908 | 0.37292 | 0.3546 | *CACNA2D1* |
| ENSG00000198711 | 1 | 54693710 | 54714736 | 58 | 14 | 52378 | 0.3729 | 0.35461 | *SSBP3-AS1* |
| ENSG00000248540 | 15 | 74408256 | 74431619 | 17 | 9 | 43406 | 0.37283 | 0.35464 | *RP11-247C2.2* |
| ENSG00000138798 | 4 | 110824040 | 110943422 | 245 | 27 | 41803 | 0.37278 | 0.35465 | *EGF* |
| ENSG00000157060 | 1 | 182859000 | 182932660 | 135 | 22 | 33944 | 0.37254 | 0.35475 | *SHCBP1L* |
| ENSG00000120903 | 8 | 27307279 | 27347400 | 153 | 25 | 46214 | 0.37236 | 0.35481 | *CHRNA2* |
| ENSG00000112759 | 6 | 44177242 | 44211888 | 101 | 27 | 50878 | 0.3723 | 0.35484 | *SLC29A1* |
| ENSG00000137857 | 15 | 45412131 | 45467774 | 125 | 23 | 34331 | 0.37203 | 0.35494 | *DUOX1* |
| ENSG00000176979 | 4 | 165943105 | 165972896 | 86 | 16 | 38093 | 0.3719 | 0.35498 | *TRIM60* |
| ENSG00000130653 | 9 | 140344404 | 140454986 | 210 | 67 | 44295 | 0.37163 | 0.35508 | *PNPLA7* |
| ENSG00000241685 | 7 | 98913521 | 98995787 | 123 | 26 | 39856 | 0.37159 | 0.3551 | *ARPC1A* |
| ENSG00000140836 | 16 | 72806784 | 73103597 | 831 | 163 | 46491 | 0.37144 | 0.35516 | *ZFHX3* |
| ENSG00000167601 | 19 | 41715108 | 41777671 | 170 | 31 | 47449 | 0.37139 | 0.35517 | *AXL* |
| ENSG00000267001 | 19 | 2707767 | 2750046 | 167 | 26 | 38338 | 0.37135 | 0.35519 | *AC006538.4* |
| ENSG00000152284 | 2 | 85350533 | 85547511 | 737 | 103 | 47784 | 0.37121 | 0.35524 | *TCF7L1* |
| ENSG00000130701 | 20 | 60975293 | 61012589 | 168 | 35 | 45455 | 0.37088 | 0.35536 | *RBBP8NL* |
| ENSG00000198483 | 1 | 145539230 | 145578526 | 61 | 11 | 44260 | 0.3708 | 0.35539 | *ANKRD35* |
| ENSG00000133687 | 12 | 29643773 | 29947692 | 1276 | 135 | 48288 | 0.37077 | 0.3554 | *TMTC1* |
| ENSG00000198914 | 2 | 105461969 | 105486929 | 40 | 13 | 51220 | 0.37075 | 0.35541 | *POU3F3* |
| ENSG00000185128 | 17 | 36273971 | 36304915 | 4 | 2 | 35724 | 0.37054 | 0.35549 | *TBC1D3F* |
| ENSG00000215014 | 1 | 1500355 | 1521373 | 97 | 13 | 40305 | 0.37046 | 0.35552 | *AL645728.1* |
| ENSG00000100243 | 22 | 43003846 | 43055574 | 135 | 21 | 45171 | 0.37037 | 0.35555 | *CYB5R3* |
| ENSG00000204366 | 6 | 31857384 | 31879769 | 73 | 25 | 36374 | 0.37034 | 0.35556 | *ZBTB12* |
| ENSG00000181588 | 19 | 1544668 | 1578057 | 62 | 30 | 41819 | 0.37033 | 0.35557 | *MEX3D* |

| | | | | | | | | | |
|---|---|---|---|---|---|---|---|---|---|
| ENSG00000128595 | 7 | 128369346 | 128421861 | 150 | 18 | 36608 | 0.37022 | 0.35561 | *CALU* |
| ENSG00000122299 | 16 | 11834442 | 11901123 | 212 | 31 | 38992 | 0.36994 | 0.35571 | *ZC3H7A* |
| ENSG00000076604 | 17 | 27061002 | 27087974 | 44 | 9 | 46527 | 0.36994 | 0.35571 | *TRAF4* |
| ENSG00000100994 | 20 | 25218705 | 25288650 | 265 | 19 | 39973 | 0.36981 | 0.35576 | *PYGB* |
| ENSG00000135452 | 12 | 58121796 | 58153994 | 44 | 17 | 41515 | 0.36971 | 0.3558 | *TSPAN31* |
| ENSG00000104043 | 15 | 50140435 | 50485014 | 1193 | 111 | 44669 | 0.36958 | 0.35585 | *ATP8B4* |
| ENSG00000187555 | 16 | 8975951 | 9068371 | 297 | 37 | 45102 | 0.36948 | 0.35588 | *USP7* |
| ENSG00000198464 | 19 | 52790430 | 52839175 | 246 | 33 | 44082 | 0.36949 | 0.35588 | *ZNF480* |
| ENSG00000176371 | 15 | 85134217 | 85181027 | 110 | 19 | 41493 | 0.3694 | 0.35591 | *ZSCAN2* |
| ENSG00000116675 | 1 | 65703902 | 65891552 | 524 | 57 | 39957 | 0.36917 | 0.356 | *DNAJC6* |
| ENSG00000137404 | 6 | 30645824 | 30669197 | 100 | 18 | 36850 | 0.36916 | 0.356 | *NRM* |
| ENSG00000145012 | 3 | 187861072 | 188618460 | 2267 | 261 | 44225 | 0.36909 | 0.35603 | *LPP* |
| ENSG00000143771 | 1 | 224534552 | 224577161 | 98 | 17 | 37831 | 0.36905 | 0.35605 | *CNIH4* |
| ENSG00000273217 | 5 | 130751584 | 131142676 | 635 | 31 | 35834 | 0.36894 | 0.35609 | *CTC-432M15.3* |
| ENSG00000183971 | 16 | 2049927 | 2080756 | 115 | 20 | 42444 | 0.36871 | 0.35617 | *NPW* |
| ENSG00000017427 | 12 | 102779645 | 102884423 | 181 | 30 | 51974 | 0.3686 | 0.35621 | *IGF1* |
| ENSG00000101306 | 20 | 30397111 | 30432492 | 87 | 10 | 40444 | 0.36862 | 0.35621 | *MYLK2* |
| ENSG00000181092 | 3 | 186550463 | 186586252 | 116 | 29 | 49421 | 0.36851 | 0.35625 | *ADIPOQ* |
| ENSG00000204599 | 6 | 30284256 | 30321506 | 150 | 26 | 41178 | 0.36803 | 0.35642 | *TRIM39* |
| ENSG00000152969 | 4 | 6017926 | 6212318 | 656 | 127 | 44043 | 0.368 | 0.35644 | *JAKMIP1* |
| ENSG00000104626 | 8 | 8849657 | 8984256 | 619 | 58 | 42381 | 0.36798 | 0.35644 | *ERI1* |
| ENSG00000174450 | 15 | 23674645 | 23702381 | 81 | 9 | 44853 | 0.36762 | 0.35658 | *GOLGA6L2* |
| ENSG00000179922 | 19 | 56122108 | 56145967 | 64 | 18 | 49485 | 0.36743 | 0.35665 | *ZNF784* |
| ENSG00000141096 | 16 | 67999566 | 68024732 | 42 | 11 | 38593 | 0.36737 | 0.35667 | *DPEP3* |
| ENSG00000205867 | 11 | 1608409 | 1629524 | 87 | 15 | 53655 | 0.36707 | 0.35678 | *KRTAP5-2* |
| ENSG00000148341 | 9 | 131759315 | 131800582 | 79 | 15 | 36443 | 0.36704 | 0.3568 | *SH3GLB2* |
| ENSG00000116218 | 1 | 179509674 | 179555087 | 192 | 25 | 44451 | 0.36688 | 0.35685 | *NPHS2* |
| ENSG00000151692 | 2 | 7047523 | 7218417 | 586 | 69 | 42679 | 0.36678 | 0.35689 | *RNF144A* |
| ENSG00000183914 | 17 | 7610672 | 7747062 | 334 | 57 | 35218 | 0.36665 | 0.35694 | *DNAH2* |
| ENSG00000137875 | 15 | 52391460 | 52414972 | 55 | 13 | 46267 | 0.36659 | 0.35696 | *BCL2L10* |
| ENSG00000161791 | 12 | 50021724 | 50111948 | 156 | 16 | 34641 | 0.36652 | 0.35699 | *FMNL3* |
| ENSG00000136867 | 9 | 115903222 | 115936417 | 160 | 27 | 46509 | 0.36649 | 0.357 | *SLC31A2* |
| ENSG00000148303 | 9 | 136205069 | 136228281 | 66 | 12 | 52411 | 0.36642 | 0.35702 | *RPL7A* |
| ENSG00000268960 | 18 | 45768672 | 45789208 | 105 | 22 | 41469 | 0.3664 | 0.35703 | *AC091150.1* |
| ENSG00000272822 | 12 | 49287286 | 49361148 | 250 | 14 | 49916 | 0.36604 | 0.35717 | *RP11-302B13.5* |
| ENSG00000166197 | 10 | 103901933 | 103933627 | 54 | 17 | 35465 | 0.36598 | 0.35719 | *NOLC1* |

| | | | | | | | | | |
|---|---|---|---|---|---|---|---|---|---|
| ENSG00000161091 | 19 | 3528259 | 3584288 | 169 | 43 | 37602 | 0.36592 | 0.35721 | *MFSD12* |
| ENSG00000229937 | 7 | 18056405 | 18077486 | 60 | 13 | 39250 | 0.36587 | 0.35723 | *PRPS1L1* |
| ENSG00000136152 | 13 | 46029060 | 46120765 | 291 | 23 | 42086 | 0.36562 | 0.35733 | *COG3* |
| ENSG00000105520 | 19 | 11456062 | 11486374 | 51 | 16 | 41398 | 0.36558 | 0.35734 | *DKFZP761J1410* |
| ENSG00000100346 | 22 | 39956758 | 40095742 | 324 | 44 | 40895 | 0.36557 | 0.35734 | *CACNA1I* |
| ENSG00000176571 | 8 | 87868670 | 88637447 | 2536 | 147 | 48963 | 0.36528 | 0.35745 | *CNBD1* |
| ENSG00000189060 | 22 | 38191114 | 38213442 | 64 | 10 | 50923 | 0.36511 | 0.35752 | *H1F0* |
| ENSG00000104064 | 15 | 50559389 | 50657605 | 297 | 23 | 33944 | 0.36498 | 0.35756 | *GABPB1* |
| ENSG00000107815 | 10 | 102737124 | 102764158 | 59 | 8 | 30225 | 0.3648 | 0.35763 | *C10orf2* |
| ENSG00000147459 | 8 | 25032238 | 25285598 | 740 | 95 | 38582 | 0.36465 | 0.35769 | *DOCK5* |
| ENSG00000186047 | 13 | 51275144 | 51428075 | 450 | 56 | 43431 | 0.36457 | 0.35772 | *DLEU7* |
| ENSG00000175592 | 11 | 65649520 | 65678044 | 55 | 14 | 36188 | 0.3645 | 0.35774 | *FOSL1* |
| ENSG00000187806 | 15 | 72680614 | 72710370 | 59 | 9 | 50996 | 0.36444 | 0.35776 | *TMEM202* |
| ENSG00000172840 | 16 | 66902492 | 66939657 | 77 | 16 | 36071 | 0.36441 | 0.35778 | *PDP2* |
| ENSG00000091542 | 17 | 18076392 | 18123268 | 112 | 13 | 42524 | 0.36408 | 0.3579 | *ALKBH5* |
| ENSG00000069424 | 1 | 6041526 | 6171253 | 395 | 83 | 49520 | 0.36401 | 0.35793 | *KCNAB2* |
| ENSG00000110442 | 11 | 36285051 | 36320999 | 188 | 21 | 47291 | 0.36377 | 0.35801 | *COMMD9* |
| ENSG00000168056 | 11 | 65296276 | 65336401 | 62 | 17 | 51199 | 0.36379 | 0.35801 | *LTBP3* |
| ENSG00000035141 | 2 | 70513107 | 70539222 | 53 | 8 | 44549 | 0.36376 | 0.35802 | *FAM136A* |
| ENSG00000011258 | 17 | 49244786 | 49347524 | 292 | 25 | 44550 | 0.36371 | 0.35804 | *MBTD1* |
| ENSG00000105357 | 19 | 50681443 | 50823802 | 674 | 112 | 46097 | 0.3637 | 0.35804 | *MYH14* |
| ENSG00000198728 | 10 | 103857317 | 103890210 | 66 | 15 | 45413 | 0.36348 | 0.35812 | *LDB1* |
| ENSG00000026036 | 20 | 62280653 | 62340037 | 241 | 27 | 37307 | 0.36312 | 0.35826 | *RTEL1-TNFRSF6B* |
| ENSG00000185467 | 7 | 98761197 | 98815129 | 257 | 17 | 51208 | 0.36253 | 0.35848 | *KPNA7* |
| ENSG00000135766 | 1 | 231489497 | 231570790 | 356 | 11 | 33053 | 0.36245 | 0.35851 | *EGLN1* |
| ENSG00000185722 | 17 | 4057201 | 4177274 | 259 | 26 | 38065 | 0.36241 | 0.35852 | *ANKFY1* |
| ENSG00000130656 | 16 | 192686 | 214502 | 67 | 16 | 39347 | 0.36237 | 0.35854 | *HBZ* |
| ENSG00000257767 | 12 | 112181694 | 112239222 | 39 | 8 | 50849 | 0.36234 | 0.35855 | *RP11-162P23.2* |
| ENSG00000108061 | 10 | 112669301 | 112783425 | 225 | 31 | 46595 | 0.36209 | 0.35864 | *SHOC2* |
| ENSG00000182809 | 14 | 105929299 | 105956499 | 28 | 15 | 35551 | 0.36196 | 0.35869 | *CRIP2* |
| ENSG00000214050 | 8 | 28195804 | 28357835 | 278 | 62 | 41572 | 0.36163 | 0.35881 | *FBXO16* |
| ENSG00000042753 | 19 | 47331393 | 47364249 | 52 | 24 | 44931 | 0.36154 | 0.35885 | *AP2S1* |
| ENSG00000115129 | 2 | 24290303 | 24318731 | 114 | 10 | 54092 | 0.36147 | 0.35887 | *TP53I3* |
| ENSG00000137878 | 15 | 57874106 | 58016943 | 575 | 67 | 46190 | 0.36136 | 0.35892 | *GCOM1* |
| ENSG00000148158 | 9 | 115503118 | 115653951 | 505 | 41 | 50253 | 0.36119 | 0.35898 | *SNX30* |
| ENSG00000187135 | 19 | 30007406 | 30065386 | 283 | 45 | 40341 | 0.3611 | 0.35901 | *VSTM2B* |

| | | | | | | | | | |
|---|---|---|---|---|---|---|---|---|---|
| ENSG00000134569 | 11 | 46868419 | 46950193 | 147 | 21 | 33147 | 0.36096 | 0.35907 | *LRP4* |
| ENSG00000261514 | 17 | 42005731 | 42026327 | 61 | 12 | 35830 | 0.36081 | 0.35912 | *RP11-527L4.2* |
| ENSG00000070018 | 12 | 12258959 | 12429946 | 433 | 42 | 49868 | 0.36024 | 0.35933 | *LRP6* |
| ENSG00000264668 | 8 | 144319280 | 144368573 | 178 | 15 | 34908 | 0.36007 | 0.3594 | *ZFP41* |
| ENSG00000188647 | 9 | 72314438 | 72384875 | 147 | 20 | 42452 | 0.35998 | 0.35943 | *PTAR1* |
| ENSG00000267106 | 19 | 9721967 | 9766572 | 86 | 12 | 34809 | 0.35961 | 0.35957 | *C19orf82* |
| ENSG00000185658 | 21 | 40546102 | 40703485 | 537 | 31 | 47342 | 0.35955 | 0.35959 | *BRWD1* |
| ENSG00000198074 | 7 | 134202344 | 134236160 | 166 | 17 | 45915 | 0.35924 | 0.35971 | *AKR1B10* |
| ENSG00000125505 | 19 | 54667107 | 54703733 | 103 | 24 | 46323 | 0.35898 | 0.3598 | *MBOAT7* |
| ENSG00000153140 | 5 | 89678078 | 89715603 | 39 | 12 | 51380 | 0.35883 | 0.35986 | *CETN3* |
| ENSG00000172139 | 3 | 111849734 | 112023105 | 862 | 34 | 54005 | 0.35863 | 0.35993 | *SLC9C1* |
| ENSG00000179776 | 16 | 66390533 | 66448686 | 185 | 35 | 50611 | 0.35854 | 0.35997 | *CDH5* |
| ENSG00000133808 | 11 | 12287627 | 12390691 | 503 | 62 | 38949 | 0.35844 | 0.36001 | *MICALCL* |
| ENSG00000151789 | 3 | 21449915 | 22424812 | 4979 | 373 | 46264 | 0.35835 | 0.36004 | *ZNF385D* |
| ENSG00000186766 | 10 | 129525499 | 129549450 | 154 | 17 | 32174 | 0.35789 | 0.36021 | *FOXI2* |
| ENSG00000101294 | 20 | 30092231 | 30167370 | 173 | 9 | 26541 | 0.35769 | 0.36029 | *HM13* |
| ENSG00000104825 | 19 | 39380340 | 39409533 | 77 | 16 | 39469 | 0.35727 | 0.36044 | *NFKBIB* |
| ENSG00000134285 | 12 | 49305301 | 49330257 | 90 | 8 | 52445 | 0.35716 | 0.36049 | *FKBP11* |
| ENSG00000255098 | 8 | 11860545 | 11883043 | 87 | 17 | 37448 | 0.35707 | 0.36052 | *RP11-481A20.11* |
| ENSG00000143627 | 1 | 155249630 | 155281225 | 52 | 7 | 31893 | 0.35705 | 0.36053 | *PKLR* |
| ENSG00000185262 | 17 | 74251283 | 74277375 | 83 | 16 | 46124 | 0.35693 | 0.36057 | *UBALD2* |
| ENSG00000105325 | 19 | 3496271 | 3548328 | 132 | 23 | 34330 | 0.35641 | 0.36077 | *FZR1* |
| ENSG00000150867 | 10 | 22813778 | 23013484 | 696 | 69 | 46217 | 0.35622 | 0.36084 | *PIP4K2A* |
| ENSG00000163013 | 2 | 73471810 | 73521559 | 93 | 23 | 33717 | 0.35614 | 0.36087 | *FBXO41* |
| ENSG00000258465 | 1 | 160177254 | 160264920 | 131 | 18 | 36785 | 0.35612 | 0.36088 | *DCAF8* |
| ENSG00000179468 | 7 | 142713245 | 142734282 | 44 | 8 | 48424 | 0.35608 | 0.36089 | *OR9A2* |
| ENSG00000205057 | 12 | 92803870 | 92831924 | 86 | 17 | 41288 | 0.35607 | 0.36089 | *CLLU1OS* |
| ENSG00000163508 | 3 | 27747440 | 27774206 | 69 | 16 | 47315 | 0.35593 | 0.36095 | *EOMES* |
| ENSG00000197375 | 5 | 131695444 | 131741306 | 145 | 15 | 32900 | 0.35572 | 0.36102 | *SLC22A5* |
| ENSG00000172006 | 19 | 2809872 | 2845771 | 135 | 20 | 41362 | 0.3556 | 0.36107 | *ZNF554* |
| ENSG00000182511 | 15 | 91416925 | 91449006 | 89 | 17 | 49416 | 0.35559 | 0.36108 | *FES* |
| ENSG00000221996 | 11 | 4378493 | 4399616 | 57 | 14 | 45921 | 0.35555 | 0.36109 | *OR52B4* |
| ENSG00000108641 | 17 | 19230867 | 19291495 | 49 | 12 | 47412 | 0.35552 | 0.3611 | *B9D1* |
| ENSG00000167236 | 17 | 34330096 | 34355005 | 107 | 17 | 34897 | 0.35523 | 0.36121 | *CCL23* |
| ENSG00000105618 | 19 | 54608837 | 54645140 | 130 | 29 | 50641 | 0.35514 | 0.36124 | *PRPF31* |
| ENSG00000100319 | 22 | 30116945 | 30173000 | 201 | 25 | 41718 | 0.35487 | 0.36135 | *ZMAT5* |

| | | | | | | | | | |
|---|---|---|---|---|---|---|---|---|---|
| ENSG00000204592 | 6 | 30447244 | 30471982 | 88 | 18 | 45944 | 0.35472 | 0.3614 | *HLA-E* |
| ENSG00000149292 | 11 | 113175251 | 113264266 | 202 | 28 | 39684 | 0.35418 | 0.3616 | *TTC12* |
| ENSG00000226757 | 4 | 122675740 | 122697962 | 97 | 16 | 37935 | 0.35411 | 0.36163 | *AC079341.1* |
| ENSG00000171570 | 19 | 41274147 | 41324103 | 190 | 30 | 46108 | 0.35406 | 0.36165 | *RAB4B-EGLN2* |
| ENSG00000025772 | 20 | 43560771 | 43599127 | 70 | 18 | 48092 | 0.35403 | 0.36166 | *TOMM34* |
| ENSG00000197728 | 12 | 56425637 | 56448116 | 36 | 10 | 44449 | 0.35386 | 0.36172 | *RPS26* |
| ENSG00000172366 | 16 | 681813 | 708474 | 62 | 13 | 33076 | 0.35374 | 0.36177 | *FAM195A* |
| ENSG00000196387 | 12 | 133646424 | 133694130 | 40 | 8 | 46129 | 0.35357 | 0.36183 | *ZNF140* |
| ENSG00000132716 | 1 | 160175505 | 160264920 | 132 | 18 | 36942 | 0.35328 | 0.36194 | *DCAF8* |
| ENSG00000197961 | 19 | 9661029 | 9705209 | 153 | 6 | 25055 | 0.35323 | 0.36196 | *ZNF121* |
| ENSG00000168297 | 3 | 58308607 | 58421748 | 502 | 32 | 36236 | 0.35307 | 0.36202 | *PXK* |
| ENSG00000185551 | 15 | 96859167 | 96893492 | 43 | 21 | 43728 | 0.35303 | 0.36203 | *NR2F2* |
| ENSG00000124164 | 20 | 56954178 | 57036157 | 263 | 27 | 49144 | 0.35292 | 0.36207 | *VAPB* |
| ENSG00000171914 | 15 | 62672725 | 63146830 | 1428 | 185 | 44183 | 0.35268 | 0.36216 | *TLN2* |
| ENSG00000163046 | 2 | 132895164 | 133025542 | 208 | 26 | 23863 | 0.35262 | 0.36219 | *ANKRD30BL* |
| ENSG00000205078 | 16 | 77223294 | 77257112 | 210 | 24 | 42423 | 0.35235 | 0.36229 | *SYCE1L* |
| ENSG00000267757 | 19 | 46134752 | 46156098 | 75 | 14 | 53225 | 0.35221 | 0.36234 | *C19orf83* |
| ENSG00000071564 | 19 | 1599291 | 1662604 | 120 | 32 | 48590 | 0.35192 | 0.36245 | *TCF3* |
| ENSG00000122592 | 7 | 27183335 | 27207555 | 62 | 14 | 35155 | 0.35185 | 0.36248 | *HOXA7* |
| ENSG00000242384 | 17 | 34571086 | 34818061 | 30 | 10 | 26886 | 0.35171 | 0.36253 | *TBC1D3H* |
| ENSG00000254445 | 11 | 111773460 | 111807595 | 62 | 12 | 35819 | 0.3515 | 0.36261 | *HSPB2-C11orf52* |
| ENSG00000118137 | 11 | 116696467 | 116718666 | 88 | 17 | 28220 | 0.35149 | 0.36261 | *APOA1* |
| ENSG00000198028 | 19 | 9567183 | 9619283 | 188 | 10 | 37467 | 0.35145 | 0.36262 | *ZNF560* |
| ENSG00000186579 | 8 | 7672694 | 7696575 | 2 | 1 | 42594 | 0.35125 | 0.3627 | *DEFB106A* |
| ENSG00000176782 | 8 | 7683993 | 7708764 | 2 | 1 | 42594 | 0.35125 | 0.3627 | *DEFB104A* |
| ENSG00000138326 | 10 | 79783518 | 79826570 | 72 | 18 | 37653 | 0.3511 | 0.36276 | *RPS24* |
| ENSG00000137975 | 1 | 86879769 | 86932241 | 266 | 27 | 56061 | 0.35107 | 0.36277 | *CLCA2* |
| ENSG00000181195 | 8 | 57339233 | 57369293 | 134 | 16 | 49361 | 0.35084 | 0.36286 | *PENK* |
| ENSG00000119147 | 2 | 106669702 | 106704615 | 130 | 31 | 52350 | 0.35063 | 0.36293 | *C2orf40* |
| ENSG00000171928 | 17 | 18674308 | 18720027 | 119 | 15 | 49055 | 0.35058 | 0.36295 | *TVP23B* |
| ENSG00000172155 | 1 | 152759227 | 152780657 | 56 | 11 | 50962 | 0.3504 | 0.36302 | *LCE1D* |
| ENSG00000139890 | 14 | 23342374 | 23366895 | 50 | 12 | 54179 | 0.35032 | 0.36305 | *REM2* |
| ENSG00000117862 | 1 | 52475803 | 52531843 | 108 | 16 | 49430 | 0.35025 | 0.36307 | *TXNDC12* |
| ENSG00000135966 | 2 | 105870871 | 105956491 | 278 | 34 | 43743 | 0.35026 | 0.36307 | *TGFBRAP1* |
| ENSG00000178996 | 5 | 53803589 | 53852415 | 231 | 32 | 47991 | 0.35025 | 0.36308 | *SNX18* |
| ENSG00000170581 | 12 | 56725381 | 56763939 | 54 | 8 | 47762 | 0.35021 | 0.36309 | *STAT2* |

| | | | | | | | | | |
|---|---|---|---|---|---|---|---|---|---|
| ENSG00000018280 | 2 | 219236752 | 219271617 | 72 | 25 | 47262 | 0.35017 | 0.36311 | *SLC11A1* |
| ENSG00000197937 | 19 | 53617325 | 53672328 | 216 | 33 | 37508 | 0.35004 | 0.36315 | *ZNF347* |
| ENSG00000104755 | 8 | 39591254 | 39705808 | 275 | 33 | 49349 | 0.35003 | 0.36316 | *ADAM2* |
| ENSG00000124459 | 19 | 44406781 | 44449430 | 146 | 15 | 53397 | 0.35002 | 0.36316 | *ZNF45* |
| ENSG00000166682 | 11 | 113548272 | 113587095 | 168 | 21 | 44745 | 0.34983 | 0.36323 | *TMPRSS5* |
| ENSG00000187323 | 18 | 49856542 | 51067784 | 5236 | 273 | 41078 | 0.34964 | 0.3633 | *DCC* |
| ENSG00000117560 | 1 | 172618154 | 172646014 | 64 | 11 | 45306 | 0.34959 | 0.36332 | *FASLG* |
| ENSG00000166143 | 15 | 41097650 | 41130907 | 94 | 11 | 40250 | 0.34956 | 0.36333 | *PPP1R14D* |
| ENSG00000014257 | 3 | 132026211 | 132097142 | 293 | 29 | 53125 | 0.34954 | 0.36334 | *ACPP* |
| ENSG00000140988 | 16 | 2002053 | 2024861 | 140 | 16 | 51153 | 0.34941 | 0.36339 | *RPS2* |
| ENSG00000253110 | 5 | 171190656 | 171211743 | 82 | 15 | 51610 | 0.34934 | 0.36342 | *CTB-78H18.1* |
| ENSG00000101292 | 20 | 5272317 | 5307378 | 274 | 16 | 52817 | 0.34934 | 0.36342 | *PROKR2* |
| ENSG00000269308 | 1 | 808043 | 829983 | 21 | 11 | 28935 | 0.34927 | 0.36344 | *AL645608.2* |
| ENSG00000109944 | 11 | 122743391 | 122840506 | 348 | 53 | 49754 | 0.34921 | 0.36347 | *C11orf63* |
| ENSG00000170142 | 3 | 23837394 | 23942807 | 265 | 36 | 47019 | 0.34916 | 0.36349 | *UBE2E1* |
| ENSG00000100336 | 22 | 36575172 | 36610886 | 217 | 25 | 35610 | 0.34911 | 0.3635 | *APOL4* |
| ENSG00000083782 | 12 | 91347456 | 91408803 | 254 | 17 | 31127 | 0.349 | 0.36354 | *EPYC* |
| ENSG00000135018 | 9 | 86264878 | 86333118 | 214 | 17 | 36924 | 0.34895 | 0.36356 | *UBQLN1* |
| ENSG00000124780 | 6 | 39256777 | 39292329 | 165 | 32 | 48491 | 0.34887 | 0.36359 | *KCNK17* |
| ENSG00000143110 | 1 | 112006414 | 112031134 | 73 | 22 | 48857 | 0.34874 | 0.36364 | *C1orf162* |
| ENSG00000073803 | 3 | 184990729 | 185216885 | 606 | 52 | 39059 | 0.34875 | 0.36364 | *MAP3K13* |
| ENSG00000073921 | 11 | 85658727 | 85790924 | 446 | 34 | 53066 | 0.34855 | 0.36371 | *PICALM* |
| ENSG00000132780 | 1 | 46039518 | 46094566 | 113 | 9 | 53582 | 0.34846 | 0.36375 | *NASP* |
| ENSG00000130402 | 19 | 39128289 | 39232223 | 346 | 26 | 49201 | 0.34839 | 0.36377 | *ACTN4* |
| ENSG00000096872 | 9 | 26937037 | 27072928 | 301 | 42 | 47866 | 0.34831 | 0.3638 | *IFT74* |
| ENSG00000085760 | 2 | 55453731 | 55506483 | 181 | 28 | 38906 | 0.34808 | 0.36389 | *MTIF2* |
| ENSG00000186335 | 5 | 150684539 | 150737151 | 210 | 30 | 37565 | 0.34806 | 0.3639 | *SLC36A2* |
| ENSG00000011485 | 19 | 46840251 | 46906238 | 193 | 34 | 45147 | 0.34801 | 0.36392 | *PPP5C* |
| ENSG00000168724 | 5 | 34919698 | 34969069 | 91 | 16 | 50951 | 0.34782 | 0.36399 | *DNAJC21* |
| ENSG00000185962 | 1 | 152585310 | 152605579 | 109 | 8 | 50412 | 0.34776 | 0.36401 | *LCE3A* |
| ENSG00000076351 | 17 | 26711661 | 26744215 | 62 | 12 | 42287 | 0.34772 | 0.36402 | *SLC46A1* |
| ENSG00000176182 | 19 | 46383278 | 46415862 | 154 | 26 | 49456 | 0.34752 | 0.3641 | *MYPOP* |
| ENSG00000152954 | 6 | 24116350 | 24165128 | 195 | 40 | 38912 | 0.34736 | 0.36416 | *NRSN1* |
| ENSG00000157111 | 5 | 72406119 | 72437644 | 81 | 18 | 46551 | 0.3471 | 0.36426 | *TMEM171* |
| ENSG00000089486 | 16 | 4550676 | 4598829 | 118 | 14 | 35975 | 0.34694 | 0.36432 | *CDIP1* |
| ENSG00000215455 | 21 | 45948864 | 45970078 | 45 | 14 | 44572 | 0.34688 | 0.36434 | *KRTAP10-1* |

| | | | | | | | | | |
|---|---|---|---|---|---|---|---|---|---|
| ENSG00000184752 | 12 | 95280831 | 95407546 | 717 | 80 | 52751 | 0.34675 | 0.36439 | *NDUFA12* |
| ENSG00000267906 | 7 | 16818866 | 16839246 | 117 | 26 | 46968 | 0.34667 | 0.36442 | *AC073333.1* |
| ENSG00000169876 | 7 | 100653353 | 100712020 | 177 | 31 | 45285 | 0.34667 | 0.36442 | *MUC17* |
| ENSG00000163389 | 3 | 119177785 | 119223555 | 140 | 21 | 47269 | 0.3465 | 0.36448 | *POGLUT1* |
| ENSG00000100767 | 14 | 73694205 | 73751348 | 163 | 34 | 46993 | 0.3465 | 0.36448 | *PAPLN* |
| ENSG00000151892 | 10 | 117806444 | 118042979 | 724 | 110 | 48593 | 0.34643 | 0.36451 | *GFRA1* |
| ENSG00000234127 | 6 | 30142232 | 30191204 | 217 | 24 | 43990 | 0.34634 | 0.36454 | *TRIM26* |
| ENSG00000090470 | 15 | 65399717 | 65436174 | 39 | 14 | 49048 | 0.3463 | 0.36456 | *PDCD7* |
| ENSG00000006634 | 7 | 87495531 | 87548856 | 109 | 16 | 49628 | 0.34622 | 0.36459 | *DBF4* |
| ENSG00000130810 | 19 | 10206965 | 10235414 | 78 | 21 | 50774 | 0.34591 | 0.36471 | *PPAN* |
| ENSG00000185339 | 22 | 30992825 | 31033265 | 280 | 37 | 38749 | 0.34577 | 0.36476 | *TCN2* |
| ENSG00000114993 | 2 | 74642963 | 74679549 | 30 | 9 | 51028 | 0.34554 | 0.36484 | *RTKN* |
| ENSG00000255112 | 18 | 11841395 | 11864448 | 73 | 13 | 41101 | 0.34513 | 0.365 | *CHMP1B* |
| ENSG00000211450 | 11 | 57498825 | 57520986 | 25 | 7 | 45679 | 0.34509 | 0.36501 | *C11orf31* |
| ENSG00000269179 | 19 | 50402758 | 50471648 | 232 | 49 | 42469 | 0.34487 | 0.3651 | *CTC-326K19.6* |
| ENSG00000183888 | 1 | 16320731 | 16345302 | 177 | 20 | 55548 | 0.34469 | 0.36517 | *C1orf64* |
| ENSG00000196547 | 15 | 91435448 | 91475814 | 97 | 21 | 42051 | 0.34442 | 0.36526 | *MAN2A2* |
| ENSG00000167757 | 19 | 51515472 | 51541295 | 70 | 15 | 44673 | 0.34439 | 0.36528 | *KLK11* |
| ENSG00000150782 | 11 | 112003974 | 112044840 | 114 | 17 | 43704 | 0.34412 | 0.36538 | *IL18* |
| ENSG00000180245 | 4 | 110739150 | 110775760 | 93 | 10 | 33790 | 0.34407 | 0.3654 | *RRH* |
| ENSG00000164318 | 5 | 38248511 | 38475123 | 632 | 102 | 44561 | 0.34397 | 0.36543 | *EGFLAM* |
| ENSG00000141570 | 17 | 77755931 | 77785482 | 57 | 20 | 46780 | 0.34399 | 0.36543 | *CBX8* |
| ENSG00000085265 | 9 | 137791431 | 137819809 | 86 | 15 | 50894 | 0.34372 | 0.36553 | *FCN1* |
| ENSG00000178297 | 19 | 2379769 | 2436237 | 356 | 40 | 40941 | 0.34372 | 0.36553 | *TMPRSS9* |
| ENSG00000162441 | 1 | 9972173 | 10013465 | 93 | 17 | 38212 | 0.34351 | 0.36561 | *LZIC* |
| ENSG00000187957 | 2 | 230212345 | 230589274 | 1443 | 159 | 47190 | 0.34337 | 0.36566 | *DNER* |
| ENSG00000125965 | 20 | 34011145 | 34052568 | 99 | 15 | 29404 | 0.3429 | 0.36584 | *GDF5* |
| ENSG00000107560 | 10 | 119754427 | 119816114 | 99 | 18 | 49861 | 0.34269 | 0.36592 | *RAB11FIP2* |
| ENSG00000154040 | 18 | 21708942 | 21751567 | 108 | 19 | 32359 | 0.34267 | 0.36592 | *CABYR* |
| ENSG00000269000 | 2 | 107089728 | 107113848 | 83 | 17 | 51709 | 0.34261 | 0.36595 | *AC108868.1* |
| ENSG00000182253 | 15 | 99628420 | 99685798 | 271 | 38 | 50167 | 0.34254 | 0.36597 | *SYNM* |
| ENSG00000035720 | 4 | 68414446 | 68483055 | 265 | 37 | 38352 | 0.34232 | 0.36605 | *STAP1* |
| ENSG00000012232 | 8 | 28447986 | 28623116 | 394 | 62 | 46599 | 0.34233 | 0.36605 | *EXTL3* |
| ENSG00000196911 | 6 | 116992350 | 117073029 | 163 | 23 | 44984 | 0.34231 | 0.36606 | *KPNA5* |
| ENSG00000150681 | 1 | 192117587 | 192164945 | 89 | 16 | 53250 | 0.3422 | 0.3661 | *RGS18* |
| ENSG00000118407 | 6 | 75991575 | 76213454 | 383 | 31 | 51992 | 0.34199 | 0.36618 | *FILIP1* |

| | | | | | | | | | |
|---|---|---|---|---|---|---|---|---|---|
| ENSG00000114999 | 2 | 113229731 | 113309316 | 234 | 28 | 46651 | 0.34196 | 0.36619 | *TTL* |
| ENSG00000172867 | 12 | 53028342 | 53055948 | 92 | 18 | 49780 | 0.34187 | 0.36622 | *KRT2* |
| ENSG00000115474 | 2 | 233621174 | 233651278 | 89 | 14 | 49660 | 0.34143 | 0.36639 | *KCNJ13* |
| ENSG00000179010 | 4 | 6631818 | 6654472 | 76 | 17 | 47377 | 0.34137 | 0.36641 | *MRFAP1* |
| ENSG00000267314 | 19 | 5894852 | 5924718 | 87 | 18 | 52636 | 0.3413 | 0.36644 | *AC104532.2* |
| ENSG00000173349 | 2 | 128448597 | 128471385 | 80 | 18 | 40258 | 0.34128 | 0.36645 | *SFT2D3* |
| ENSG00000159433 | 15 | 42857857 | 43023179 | 244 | 28 | 28341 | 0.34108 | 0.36652 | *STARD9* |
| ENSG00000169169 | 19 | 50184155 | 50226988 | 107 | 24 | 41057 | 0.34105 | 0.36653 | *CPT1C* |
| ENSG00000188155 | 21 | 46001149 | 46022386 | 93 | 11 | 28608 | 0.34089 | 0.36659 | *KRTAP10-6* |
| ENSG00000128581 | 7 | 100946975 | 100975104 | 134 | 15 | 52705 | 0.3408 | 0.36663 | *RABL5* |
| ENSG00000029153 | 12 | 27475787 | 27586241 | 376 | 60 | 50790 | 0.34073 | 0.36665 | *ARNTL2* |
| ENSG00000175283 | 9 | 131697809 | 131719898 | 28 | 5 | 50142 | 0.34038 | 0.36678 | *DOLK* |
| ENSG00000165626 | 10 | 13470484 | 13580974 | 337 | 39 | 47328 | 0.34041 | 0.36678 | *BEND7* |
| ENSG00000145384 | 4 | 120228405 | 120253545 | 193 | 11 | 54842 | 0.34034 | 0.3668 | *FABP2* |
| ENSG00000148297 | 9 | 136195160 | 136224986 | 85 | 14 | 50269 | 0.34031 | 0.36681 | *MED22* |
| ENSG00000101276 | 20 | 730724 | 759131 | 155 | 37 | 48071 | 0.34006 | 0.36691 | *SLC52A3* |
| ENSG00000121207 | 4 | 155538097 | 155684270 | 274 | 25 | 29734 | 0.33995 | 0.36695 | *LRAT* |
| ENSG00000165861 | 14 | 73426159 | 73503920 | 365 | 33 | 52851 | 0.33986 | 0.36698 | *ZFYVE1* |
| ENSG00000105221 | 19 | 40726224 | 40801443 | 173 | 24 | 38111 | 0.33967 | 0.36705 | *AKT2* |
| ENSG00000146433 | 6 | 158947468 | 159066460 | 455 | 42 | 49638 | 0.33965 | 0.36706 | *TMEM181* |
| ENSG00000229387 | 11 | 22636860 | 22657369 | 31 | 10 | 36765 | 0.33963 | 0.36707 | *AC103801.2* |
| ENSG00000141424 | 18 | 33678495 | 33719348 | 132 | 18 | 51208 | 0.33952 | 0.36711 | *SLC39A6* |
| ENSG00000230000 | 7 | 62799239 | 62822151 | 10 | 2 | 47180 | 0.33944 | 0.36714 | *AC006455.1* |
| ENSG00000133606 | 7 | 140142840 | 140189369 | 165 | 14 | 30503 | 0.33927 | 0.3672 | *MKRN1* |
| ENSG00000174697 | 7 | 127871337 | 127907681 | 98 | 19 | 43182 | 0.33924 | 0.36721 | *LEP* |
| ENSG00000149313 | 11 | 105936228 | 105979437 | 69 | 13 | 43054 | 0.3392 | 0.36723 | *AASDHPPT* |
| ENSG00000172717 | 14 | 67646110 | 67705267 | 154 | 26 | 44904 | 0.33918 | 0.36724 | *FAM71D* |
| ENSG00000011523 | 2 | 65273500 | 65324138 | 173 | 26 | 51581 | 0.33902 | 0.3673 | *CEP68* |
| ENSG00000130299 | 19 | 17435729 | 17463544 | 105 | 16 | 45859 | 0.33884 | 0.36736 | *GTPBP3* |
| ENSG00000112972 | 5 | 43279497 | 43323614 | 120 | 8 | 44609 | 0.33873 | 0.36741 | *HMGCS1* |
| ENSG00000172113 | 3 | 48324754 | 48353175 | 73 | 5 | 34272 | 0.33835 | 0.36755 | *NME6* |
| ENSG00000181638 | 8 | 144318991 | 144354875 | 118 | 11 | 34203 | 0.33835 | 0.36755 | *ZFP41* |
| ENSG00000198576 | 8 | 143682405 | 143706833 | 84 | 15 | 39900 | 0.33815 | 0.36763 | *ARC* |
| ENSG00000143669 | 1 | 235814341 | 236056940 | 356 | 29 | 38216 | 0.33807 | 0.36765 | *LYST* |
| ENSG00000069966 | 15 | 52403117 | 52493566 | 262 | 29 | 50039 | 0.33795 | 0.3677 | *GNB5* |
| ENSG00000257115 | 14 | 19367522 | 19388606 | 3 | 1 | 43467 | 0.33788 | 0.36773 | *OR11H12* |

| | | | | | | | | | |
|---|---|---|---|---|---|---|---|---|---|
| ENSG00000180383 | 20 | 30043309 | 30074560 | 67 | 15 | 39612 | 0.33784 | 0.36774 | *DEFB124* |
| ENSG00000100092 | 22 | 38020661 | 38072939 | 179 | 27 | 44411 | 0.3378 | 0.36776 | *SH3BP1* |
| ENSG00000011243 | 19 | 15480859 | 15539952 | 105 | 11 | 51874 | 0.33745 | 0.36789 | *AKAP8L* |
| ENSG00000005059 | 4 | 110471361 | 110619874 | 438 | 41 | 48735 | 0.33728 | 0.36795 | *CCDC109B* |
| ENSG00000113520 | 5 | 131999678 | 132028368 | 46 | 10 | 31797 | 0.3373 | 0.36795 | *IL4* |
| ENSG00000183086 | 7 | 74369083 | 74448803 | 18 | 10 | 34420 | 0.33721 | 0.36798 | *GATSL1* |
| ENSG00000143632 | 1 | 229556992 | 229579845 | 62 | 16 | 37331 | 0.33713 | 0.36801 | *ACTA1* |
| ENSG00000116005 | 2 | 70474518 | 70518323 | 102 | 8 | 48827 | 0.33706 | 0.36804 | *PCYOX1* |
| ENSG00000112818 | 6 | 46751127 | 46817515 | 176 | 20 | 50991 | 0.33675 | 0.36815 | *MEP1A* |
| ENSG00000197177 | 10 | 134874433 | 134955179 | 409 | 68 | 44061 | 0.33675 | 0.36815 | *GPR123* |
| ENSG00000189091 | 16 | 70547691 | 70618820 | 118 | 14 | 36929 | 0.33675 | 0.36815 | *SF3B3* |
| ENSG00000144659 | 3 | 39414839 | 39448842 | 148 | 15 | 52146 | 0.3367 | 0.36817 | *SLC25A38* |
| ENSG00000177302 | 17 | 18164742 | 18228321 | 173 | 17 | 48422 | 0.33661 | 0.3682 | *TOP3A* |
| ENSG00000137078 | 9 | 35639292 | 35660937 | 100 | 17 | 43895 | 0.33622 | 0.36835 | *SIT1* |
| ENSG00000090674 | 19 | 7577512 | 7608895 | 73 | 22 | 53233 | 0.33612 | 0.36839 | *MCOLN1* |
| ENSG00000178301 | 11 | 77290436 | 77331400 | 104 | 11 | 44382 | 0.33604 | 0.36842 | *AQP11* |
| ENSG00000072110 | 14 | 69330860 | 69456157 | 493 | 85 | 47603 | 0.33602 | 0.36843 | *ACTN1* |
| ENSG00000111615 | 12 | 75880684 | 75915416 | 100 | 12 | 52235 | 0.33593 | 0.36846 | *KRR1* |
| ENSG00000132305 | 2 | 86361055 | 86432893 | 239 | 16 | 43128 | 0.33591 | 0.36847 | *IMMT* |
| ENSG00000143498 | 1 | 222721244 | 222773275 | 140 | 20 | 45309 | 0.33549 | 0.36863 | *TAF1A* |
| ENSG00000111850 | 6 | 88022301 | 88062043 | 195 | 16 | 47449 | 0.33513 | 0.36876 | *SMIM8* |
| ENSG00000157890 | 15 | 66177417 | 66556085 | 1263 | 171 | 45738 | 0.33472 | 0.36892 | *MEGF11* |
| ENSG00000139697 | 12 | 123763656 | 123859390 | 405 | 23 | 52225 | 0.33469 | 0.36893 | *SBNO1* |
| ENSG00000115020 | 2 | 209120991 | 209233475 | 365 | 26 | 32780 | 0.33448 | 0.36901 | *PIKFYVE* |
| ENSG00000141293 | 17 | 46200802 | 46517637 | 825 | 57 | 44693 | 0.33445 | 0.36902 | *SKAP1* |
| ENSG00000179921 | 2 | 219114219 | 219138582 | 44 | 8 | 42626 | 0.33415 | 0.36913 | *GPBAR1* |
| ENSG00000137880 | 15 | 41046218 | 41069906 | 48 | 16 | 43817 | 0.33415 | 0.36913 | *GCHFR* |
| ENSG00000168676 | 16 | 67313331 | 67370666 | 65 | 21 | 46197 | 0.33416 | 0.36913 | *KCTD19* |
| ENSG00000088325 | 20 | 30317074 | 30399608 | 159 | 16 | 36386 | 0.33406 | 0.36917 | *TPX2* |
| ENSG00000133030 | 17 | 16935859 | 17130993 | 524 | 48 | 39287 | 0.33377 | 0.36928 | *MPRIP* |
| ENSG00000261915 | 17 | 7205980 | 7232493 | 62 | 13 | 48604 | 0.33357 | 0.36935 | *RP11-542C16.2* |
| ENSG00000174453 | 2 | 215265789 | 215453683 | 591 | 60 | 44981 | 0.33353 | 0.36937 | *VWC2L* |
| ENSG00000095794 | 10 | 35405719 | 35511886 | 270 | 12 | 53263 | 0.33332 | 0.36945 | *CREM* |
| ENSG00000152556 | 12 | 48488922 | 48550187 | 173 | 22 | 46720 | 0.33325 | 0.36947 | *PFKM* |
| ENSG00000176731 | 8 | 86116311 | 86142650 | 38 | 7 | 51259 | 0.33322 | 0.36948 | *C8orf59* |
| ENSG00000181666 | 19 | 37793739 | 37870267 | 175 | 23 | 35474 | 0.33322 | 0.36948 | *HKR1* |

| | | | | | | | | | |
|---|---|---|---|---|---|---|---|---|---|
| ENSG00000267680 | 19 | 44588492 | 44622479 | 98 | 11 | 22398 | 0.33315 | 0.36951 | *ZNF224* |
| ENSG00000139668 | 13 | 52148644 | 52346171 | 289 | 39 | 51297 | 0.33306 | 0.36954 | *WDFY2* |
| ENSG00000105355 | 19 | 4828353 | 4877780 | 214 | 41 | 50683 | 0.33307 | 0.36954 | *PLIN3* |
| ENSG00000116703 | 1 | 186402698 | 186440254 | 76 | 10 | 46642 | 0.33272 | 0.36967 | *PDC* |
| ENSG00000115762 | 2 | 131852420 | 132121282 | 423 | 27 | 31148 | 0.33256 | 0.36973 | *PLEKHB2* |
| ENSG00000188897 | 16 | 11443702 | 11627444 | 1100 | 122 | 45118 | 0.33232 | 0.36982 | *CTD-3088G3.8* |
| ENSG00000175470 | 10 | 133737955 | 133783331 | 142 | 13 | 54092 | 0.33226 | 0.36985 | *PPP2R2D* |
| ENSG00000040933 | 2 | 99051317 | 99220853 | 293 | 17 | 54948 | 0.33218 | 0.36988 | *INPP4A* |
| ENSG00000105855 | 7 | 20360325 | 20465377 | 423 | 42 | 47277 | 0.33199 | 0.36995 | *ITGB8* |
| ENSG00000102452 | 13 | 101696130 | 102078843 | 1587 | 138 | 44796 | 0.332 | 0.36995 | *NALCN* |
| ENSG00000109743 | 4 | 15694573 | 15749936 | 183 | 33 | 43225 | 0.33168 | 0.37006 | *BST1* |
| ENSG00000117632 | 1 | 26200672 | 26243482 | 68 | 9 | 42591 | 0.33167 | 0.37007 | *STMN1* |
| ENSG00000020181 | 8 | 37631709 | 37712414 | 138 | 39 | 46276 | 0.33144 | 0.37016 | *GPR124* |
| ENSG00000244474 | 2 | 234617424 | 234691945 | 316 | 31 | 42066 | 0.33096 | 0.37034 | *UGT1A4* |
| ENSG00000186326 | 19 | 33156313 | 33179206 | 106 | 21 | 37736 | 0.33096 | 0.37034 | *RGS9BP* |
| ENSG00000203286 | 9 | 3442305 | 3479181 | 63 | 18 | 38359 | 0.33089 | 0.37036 | *AL365202.1* |
| ENSG00000100815 | 14 | 92422335 | 92517240 | 283 | 26 | 50434 | 0.3309 | 0.37036 | *TRIP11* |
| ENSG00000104131 | 15 | 44819255 | 44865227 | 77 | 12 | 35315 | 0.33046 | 0.37053 | *EIF3J* |
| ENSG00000102554 | 13 | 73619114 | 73661676 | 163 | 33 | 37119 | 0.33034 | 0.37057 | *KLF5* |
| ENSG00000121064 | 17 | 55045466 | 55094129 | 83 | 17 | 49646 | 0.33021 | 0.37062 | *SCPEP1* |
| ENSG00000168995 | 19 | 51635556 | 51666783 | 34 | 19 | 41934 | 0.32986 | 0.37075 | *SIGLEC7* |
| ENSG00000198046 | 19 | 56940694 | 56999434 | 155 | 20 | 46843 | 0.32983 | 0.37076 | *ZNF667* |
| ENSG00000104853 | 19 | 45447842 | 45506599 | 221 | 19 | 47793 | 0.32974 | 0.3708 | *CLPTM1* |
| ENSG00000171540 | 5 | 76914538 | 76945513 | 78 | 14 | 49693 | 0.32969 | 0.37082 | *OTP* |
| ENSG00000065989 | 19 | 10517449 | 10590305 | 170 | 26 | 50245 | 0.32966 | 0.37083 | *PDE4A* |
| ENSG00000163207 | 1 | 152871021 | 152894362 | 32 | 13 | 43863 | 0.32963 | 0.37084 | *IVL* |
| ENSG00000214113 | 6 | 5092827 | 5271172 | 719 | 68 | 40677 | 0.32934 | 0.37095 | *LYRM4* |
| ENSG00000187151 | 11 | 101751405 | 101797253 | 133 | 25 | 41443 | 0.32933 | 0.37095 | *ANGPTL5* |
| ENSG00000118849 | 3 | 158404681 | 158460485 | 210 | 22 | 52820 | 0.32914 | 0.37102 | *RARRES1* |
| ENSG00000115593 | 2 | 88357299 | 88422906 | 235 | 41 | 45749 | 0.32913 | 0.37103 | *SMYD1* |
| ENSG00000165410 | 14 | 35169593 | 35194029 | 105 | 17 | 31337 | 0.3291 | 0.37104 | *CFL2* |
| ENSG00000144589 | 2 | 220452582 | 220491173 | 40 | 23 | 47880 | 0.32884 | 0.37114 | *STK11IP* |
| ENSG00000139644 | 12 | 50091508 | 50168717 | 243 | 19 | 32709 | 0.32885 | 0.37114 | *TMBIM6* |
| ENSG00000100979 | 20 | 44517399 | 44550794 | 84 | 22 | 42086 | 0.32883 | 0.37114 | *PLTP* |
| ENSG00000197785 | 1 | 1437531 | 1480067 | 110 | 30 | 41531 | 0.32876 | 0.37117 | *ATAD3A* |
| ENSG00000203737 | 1 | 174407212 | 174428683 | 46 | 10 | 40165 | 0.32874 | 0.37117 | *GPR52* |

| | | | | | | | | | |
|---|---|---|---|---|---|---|---|---|---|
| ENSG00000079974 | 22 | 51195929 | 51232091 | 57 | 12 | 47090 | 0.32865 | 0.37121 | *RABL2B* |
| ENSG00000132130 | 17 | 35284084 | 35311917 | 56 | 14 | 31186 | 0.32856 | 0.37124 | *LHX1* |
| ENSG00000121957 | 1 | 109407972 | 109487167 | 255 | 31 | 44890 | 0.32851 | 0.37126 | *GPSM2* |
| ENSG00000181652 | 7 | 150699297 | 150731586 | 88 | 29 | 51216 | 0.32846 | 0.37128 | *ATG9B* |
| ENSG00000121297 | 19 | 31755851 | 31850453 | 219 | 52 | 42256 | 0.32781 | 0.37153 | *TSHZ3* |
| ENSG00000165506 | 14 | 50081892 | 50111948 | 92 | 9 | 36305 | 0.32761 | 0.3716 | *DNAAF2* |
| ENSG00000126001 | 20 | 34032985 | 34109804 | 144 | 13 | 31490 | 0.32717 | 0.37177 | *CEP250* |
| ENSG00000135249 | 7 | 105162532 | 105218124 | 202 | 13 | 26655 | 0.32693 | 0.37186 | *RINT1* |
| ENSG00000160796 | 3 | 47011173 | 47061193 | 83 | 15 | 44570 | 0.32677 | 0.37192 | *NBEAL2* |
| ENSG00000102900 | 16 | 56754017 | 56888797 | 350 | 41 | 52810 | 0.32654 | 0.37201 | *NUP93* |
| ENSG00000125813 | 20 | 21676297 | 21706620 | 43 | 18 | 43225 | 0.32655 | 0.37201 | *PAX1* |
| ENSG00000171357 | 1 | 46659006 | 46696933 | 53 | 13 | 46702 | 0.32641 | 0.37206 | *LURAP1* |
| ENSG00000148773 | 10 | 129884923 | 129934649 | 231 | 33 | 53948 | 0.32624 | 0.37212 | *MKI67* |
| ENSG00000132382 | 17 | 4432192 | 4468926 | 128 | 49 | 45116 | 0.32625 | 0.37212 | *MYBBP1A* |
| ENSG00000204398 | 13 | 112230548 | 112334955 | 415 | 55 | 45618 | 0.32603 | 0.3722 | *RP11-65D24.2* |
| ENSG00000131042 | 19 | 54767675 | 54795039 | 112 | 24 | 34893 | 0.32588 | 0.37226 | *LILRB2* |
| ENSG00000101134 | 20 | 53082136 | 53277710 | 552 | 60 | 38475 | 0.3258 | 0.37229 | *DOK5* |
| ENSG00000112787 | 12 | 133056137 | 133171774 | 565 | 66 | 47520 | 0.3256 | 0.37236 | *FBRSL1* |
| ENSG00000184530 | 6 | 127830600 | 127922962 | 383 | 34 | 32091 | 0.32539 | 0.37244 | *C6orf58* |
| ENSG00000198920 | 17 | 6471468 | 6554247 | 295 | 24 | 39289 | 0.32538 | 0.37245 | *KIAA0753* |
| ENSG00000141378 | 17 | 57741997 | 57794987 | 74 | 15 | 42523 | 0.32537 | 0.37245 | *PTRH2* |
| ENSG00000001167 | 6 | 41030684 | 41077715 | 104 | 16 | 48336 | 0.32535 | 0.37246 | *NFYA* |
| ENSG00000120693 | 13 | 37408968 | 37504902 | 239 | 43 | 39685 | 0.32535 | 0.37246 | *SMAD9* |
| ENSG00000151461 | 10 | 11952021 | 12095169 | 544 | 40 | 48956 | 0.32502 | 0.37258 | *UPF2* |
| ENSG00000187123 | 2 | 150176499 | 150340662 | 368 | 43 | 41082 | 0.32462 | 0.37273 | *LYPD6* |
| ENSG00000180902 | 2 | 242663994 | 242718231 | 211 | 36 | 37724 | 0.32461 | 0.37274 | *D2HGDH* |
| ENSG00000198087 | 6 | 47435525 | 47604999 | 486 | 24 | 52945 | 0.32458 | 0.37275 | *CD2AP* |
| ENSG00000138614 | 15 | 65861091 | 65913627 | 111 | 10 | 45732 | 0.32448 | 0.37279 | *VWA9* |
| ENSG00000154165 | 3 | 98240743 | 98261960 | 52 | 15 | 45822 | 0.32438 | 0.37283 | *GPR15* |
| ENSG00000171282 | 17 | 79363540 | 79443357 | 246 | 59 | 44314 | 0.32392 | 0.373 | *RP11-1055B8.7* |
| ENSG00000127184 | 5 | 85903721 | 85926779 | 27 | 14 | 43242 | 0.3238 | 0.37304 | *COX7C* |
| ENSG00000206107 | 21 | 31699331 | 31720012 | 108 | 14 | 34834 | 0.32378 | 0.37305 | *KRTAP27-1* |
| ENSG00000110203 | 11 | 71815915 | 71860936 | 170 | 22 | 38087 | 0.32365 | 0.3731 | *FOLR3* |
| ENSG00000162496 | 1 | 12617939 | 12687737 | 292 | 51 | 48757 | 0.32351 | 0.37316 | *DHRS3* |
| ENSG00000101624 | 18 | 12651832 | 12712776 | 200 | 18 | 38477 | 0.3234 | 0.3732 | *CEP76* |
| ENSG00000130347 | 6 | 107008903 | 107087373 | 348 | 24 | 43930 | 0.32327 | 0.37325 | *RTN4IP1* |

| | | | | | | | | | |
|---|---|---|---|---|---|---|---|---|---|
| ENSG00000113594 | 5 | 38465065 | 38618456 | 273 | 40 | 46032 | 0.32305 | 0.37333 | *LIFR* |
| ENSG00000156284 | 21 | 31576324 | 31598391 | 68 | 21 | 43258 | 0.32306 | 0.37333 | *CLDN8* |
| ENSG00000186146 | 4 | 9436260 | 9462240 | 66 | 16 | 45308 | 0.32296 | 0.37336 | *DEFB131* |
| ENSG00000165799 | 14 | 21500385 | 21522393 | 79 | 31 | 46243 | 0.32294 | 0.37337 | *RNASE7* |
| ENSG00000164346 | 5 | 74052817 | 74082737 | 70 | 11 | 43711 | 0.3228 | 0.37342 | *NSA2* |
| ENSG00000050438 | 12 | 51775101 | 51912980 | 345 | 32 | 52784 | 0.32273 | 0.37345 | *SLC4A8* |
| ENSG00000103512 | 16 | 14917538 | 15000017 | 49 | 28 | 35972 | 0.32267 | 0.37347 | *NOMO1* |
| ENSG00000133789 | 11 | 9675624 | 9784538 | 384 | 24 | 42483 | 0.3221 | 0.37369 | *SWAP70* |
| ENSG00000268941 | 20 | 57199828 | 57220772 | 69 | 13 | 36457 | 0.32199 | 0.37373 | *MGC4294* |
| ENSG00000162931 | 1 | 228585641 | 228614562 | 56 | 13 | 42105 | 0.32195 | 0.37374 | *TRIM17* |
| ENSG00000143630 | 1 | 155237374 | 155269639 | 54 | 6 | 30662 | 0.32194 | 0.37375 | *HCN3* |
| ENSG00000196693 | 10 | 43074555 | 43143992 | 192 | 23 | 36081 | 0.3218 | 0.3738 | *ZNF33B* |
| ENSG00000095370 | 9 | 130490596 | 130551020 | 149 | 26 | 37463 | 0.32176 | 0.37382 | *SH2D3C* |
| ENSG00000165312 | 10 | 23718198 | 23741308 | 34 | 10 | 38413 | 0.32172 | 0.37383 | *OTUD1* |
| ENSG00000056097 | 5 | 32344456 | 32454867 | 352 | 33 | 52150 | 0.32136 | 0.37397 | *ZFR* |
| ENSG00000005448 | 2 | 74638805 | 74662882 | 24 | 7 | 48795 | 0.32129 | 0.37399 | *WDR54* |
| ENSG00000171481 | 9 | 125427315 | 125448432 | 55 | 12 | 46638 | 0.32129 | 0.37399 | *OR1L3* |
| ENSG00000117399 | 1 | 43814626 | 43838874 | 36 | 10 | 45670 | 0.32126 | 0.37401 | *CDC20* |
| ENSG00000144035 | 2 | 73857959 | 73879520 | 90 | 8 | 45619 | 0.32117 | 0.37404 | *NAT8* |
| ENSG00000205208 | 4 | 159577831 | 159603407 | 19 | 7 | 40344 | 0.32103 | 0.37409 | *C4orf46* |
| ENSG00000164031 | 4 | 100807405 | 100877883 | 139 | 21 | 42259 | 0.32098 | 0.37411 | *DNAJB14* |
| ENSG00000186074 | 17 | 72680452 | 72719117 | 89 | 24 | 40965 | 0.32089 | 0.37415 | *CD300LF* |
| ENSG00000142186 | 11 | 65282548 | 65316175 | 44 | 14 | 54789 | 0.32082 | 0.37417 | *SCYL1* |
| ENSG00000257019 | 9 | 107356924 | 107377951 | 152 | 9 | 25884 | 0.32064 | 0.37424 | *OR13C2* |
| ENSG00000151320 | 14 | 32788479 | 33310567 | 1755 | 209 | 38782 | 0.32033 | 0.37436 | *AKAP6* |
| ENSG00000125810 | 20 | 23049986 | 23076977 | 55 | 11 | 48344 | 0.32031 | 0.37437 | *CD93* |
| ENSG00000074370 | 17 | 3817169 | 3877736 | 231 | 33 | 44205 | 0.32023 | 0.3744 | *ATP2A3* |
| ENSG00000084110 | 12 | 96356440 | 96400143 | 220 | 34 | 44509 | 0.3201 | 0.37445 | *HAL* |
| ENSG00000100234 | 22 | 33187687 | 33269030 | 214 | 38 | 47067 | 0.32009 | 0.37445 | *TIMP3* |
| ENSG00000164398 | 5 | 131132683 | 131357936 | 406 | 27 | 35465 | 0.31985 | 0.37454 | *ACSL6* |
| ENSG00000119547 | 18 | 55092917 | 55168529 | 226 | 35 | 37081 | 0.31975 | 0.37458 | *ONECUT2* |
| ENSG00000156162 | 8 | 95721931 | 95816064 | 324 | 39 | 45393 | 0.31959 | 0.37464 | *DPY19L4* |
| ENSG00000170989 | 1 | 101692444 | 101717074 | 48 | 17 | 44905 | 0.31957 | 0.37465 | *S1PR1* |
| ENSG00000188613 | 10 | 120779228 | 120803854 | 37 | 9 | 34959 | 0.319 | 0.37486 | *NANOS1* |
| ENSG00000061936 | 12 | 132185626 | 132294282 | 278 | 31 | 30434 | 0.31901 | 0.37486 | *SFSWAP* |
| ENSG00000104814 | 19 | 39068281 | 39119522 | 101 | 19 | 35551 | 0.319 | 0.37486 | *MAP4K1* |

| | | | | | | | | | |
|---|---|---|---|---|---|---|---|---|---|
| ENSG00000268509 | 3 | 5219227 | 5239982 | 39 | 14 | 38961 | 0.31892 | 0.37489 | *AC026202.1* |
| ENSG00000083457 | 17 | 3607922 | 3714537 | 408 | 55 | 40834 | 0.31872 | 0.37497 | *ITGAE* |
| ENSG00000150593 | 10 | 112621565 | 112669764 | 105 | 21 | 48441 | 0.31865 | 0.375 | *PDCD4* |
| ENSG00000007384 | 16 | 98058 | 136354 | 108 | 18 | 39126 | 0.3186 | 0.37502 | *RHBDF1* |
| ENSG00000139517 | 13 | 28110050 | 28204541 | 342 | 38 | 51674 | 0.31855 | 0.37503 | *LNX2* |
| ENSG00000183876 | 5 | 149665906 | 149728870 | 145 | 32 | 53148 | 0.31854 | 0.37504 | *ARSI* |
| ENSG00000178607 | 17 | 62106502 | 62218179 | 218 | 38 | 43687 | 0.31843 | 0.37508 | *ERN1* |
| ENSG00000183856 | 1 | 156485197 | 156552396 | 174 | 23 | 30044 | 0.31836 | 0.37511 | *IQGAP3* |
| ENSG00000149179 | 11 | 46948240 | 47195936 | 382 | 36 | 46952 | 0.31808 | 0.37521 | *C11orf49* |
| ENSG00000165280 | 9 | 35046061 | 35083246 | 70 | 13 | 43899 | 0.31807 | 0.37522 | *VCP* |
| ENSG00000124701 | 6 | 41011043 | 41042250 | 89 | 8 | 52321 | 0.31797 | 0.37525 | *APOBEC2* |
| ENSG00000129460 | 14 | 23928897 | 23989071 | 170 | 23 | 35054 | 0.31795 | 0.37526 | *NGDN* |
| ENSG00000108700 | 17 | 32636055 | 32658421 | 85 | 9 | 51867 | 0.31784 | 0.3753 | *CCL8* |
| ENSG00000198492 | 1 | 29053133 | 29106287 | 104 | 12 | 41891 | 0.31762 | 0.37539 | *YTHDF2* |
| ENSG00000125482 | 9 | 135241008 | 135292209 | 111 | 20 | 32308 | 0.31748 | 0.37544 | *TTF1* |
| ENSG00000143919 | 2 | 44579089 | 45009731 | 1146 | 61 | 44149 | 0.31738 | 0.37548 | *CAMKMT* |
| ENSG00000138613 | 15 | 63558217 | 63611325 | 141 | 31 | 41310 | 0.31713 | 0.37557 | *APH1B* |
| ENSG00000102466 | 13 | 102362134 | 103064124 | 2077 | 186 | 42209 | 0.31706 | 0.3756 | *FGF14* |
| ENSG00000205076 | 19 | 39251611 | 39274132 | 99 | 15 | 45173 | 0.31644 | 0.37583 | *LGALS7* |
| ENSG00000162783 | 1 | 181047638 | 181069977 | 55 | 13 | 50334 | 0.31644 | 0.37584 | *IER5* |
| ENSG00000135144 | 12 | 113484514 | 113545833 | 259 | 41 | 44776 | 0.31642 | 0.37584 | *DTX1* |
| ENSG00000204918 | 13 | 57711622 | 57734643 | 8 | 4 | 49992 | 0.31639 | 0.37585 | *PRR20B* |
| ENSG00000221931 | 11 | 123614196 | 123635253 | 77 | 12 | 40815 | 0.31638 | 0.37586 | *OR6X1* |
| ENSG00000185946 | 1 | 104058313 | 104107861 | 81 | 21 | 50939 | 0.31606 | 0.37598 | *RNPC3* |
| ENSG00000166908 | 12 | 57974957 | 58007198 | 64 | 16 | 37647 | 0.31604 | 0.37599 | *PIP4K2C* |
| ENSG00000143127 | 1 | 145514891 | 145553868 | 32 | 8 | 44256 | 0.31578 | 0.37609 | *ITGA10* |
| ENSG00000129696 | 8 | 33320904 | 33381119 | 230 | 17 | 49484 | 0.31551 | 0.37619 | *TTI2* |
| ENSG00000160886 | 8 | 143771529 | 143796545 | 78 | 9 | 46109 | 0.31546 | 0.37621 | *LY6K* |
| ENSG00000162779 | 1 | 179324855 | 179533870 | 1365 | 43 | 52929 | 0.31533 | 0.37626 | *AXDND1* |
| ENSG00000169184 | 22 | 28134265 | 28207486 | 180 | 67 | 46623 | 0.31532 | 0.37626 | *MN1* |
| ENSG00000088320 | 20 | 30053096 | 30082708 | 55 | 12 | 37746 | 0.31524 | 0.37629 | *REM1* |
| ENSG00000101197 | 20 | 61857235 | 61881859 | 144 | 22 | 52208 | 0.31515 | 0.37633 | *BIRC7* |
| ENSG00000258366 | 20 | 62279163 | 62338416 | 240 | 27 | 37353 | 0.31507 | 0.37635 | *RTEL1* |
| ENSG00000240184 | 5 | 140845580 | 140902542 | 159 | 29 | 46336 | 0.31494 | 0.3764 | *PCDHGC3* |
| ENSG00000157315 | 16 | 69367151 | 69395712 | 55 | 15 | 54315 | 0.31489 | 0.37642 | *TMED6* |
| ENSG00000120647 | 12 | 488439 | 561811 | 209 | 46 | 45795 | 0.31486 | 0.37643 | *CCDC77* |

| | | | | | | | | | |
|---|---|---|---|---|---|---|---|---|---|
| ENSG00000224153 | 3 | 184109325 | 184167436 | 255 | 43 | 45648 | 0.31473 | 0.37648 | *RP11-433C9.2* |
| ENSG00000119684 | 14 | 75470467 | 75528235 | 107 | 9 | 47496 | 0.31474 | 0.37648 | *MLH3* |
| ENSG00000134825 | 11 | 61525973 | 61570274 | 68 | 16 | 29862 | 0.31472 | 0.37649 | *TMEM258* |
| ENSG00000261857 | 19 | 41267553 | 41293395 | 88 | 15 | 43060 | 0.31453 | 0.37656 | *MIA* |
| ENSG00000165762 | 14 | 20334391 | 20355468 | 14 | 7 | 43473 | 0.31448 | 0.37658 | *OR4K2* |
| ENSG00000137269 | 6 | 53649295 | 53798919 | 444 | 33 | 51380 | 0.3142 | 0.37668 | *LRRC1* |
| ENSG00000143167 | 1 | 167012073 | 167069868 | 206 | 35 | 49417 | 0.31414 | 0.37671 | *GPA33* |
| ENSG00000127191 | 9 | 139766364 | 139831059 | 207 | 12 | 54492 | 0.31404 | 0.37675 | *TRAF2* |
| ENSG00000267908 | 19 | 56741773 | 56768863 | 160 | 37 | 46586 | 0.31402 | 0.37675 | *ZSCAN5D* |
| ENSG00000176697 | 11 | 27666440 | 27753605 | 168 | 25 | 41805 | 0.31368 | 0.37688 | *BDNF* |
| ENSG00000205021 | 17 | 34613842 | 34635731 | 3 | 2 | 42925 | 0.31359 | 0.37692 | *CCL3L1* |
| ENSG00000143033 | 1 | 93534792 | 93614638 | 148 | 18 | 39182 | 0.31331 | 0.37702 | *MTF2* |
| ENSG00000179388 | 8 | 22535172 | 22560815 | 41 | 14 | 47565 | 0.31332 | 0.37702 | *EGR3* |
| ENSG00000161036 | 7 | 102095376 | 102123615 | 104 | 15 | 40715 | 0.31318 | 0.37707 | *LRWD1* |
| ENSG00000172888 | 3 | 40556369 | 40626176 | 145 | 25 | 38746 | 0.31246 | 0.37735 | *ZNF621* |
| ENSG00000188505 | 19 | 39677601 | 39702524 | 65 | 7 | 54933 | 0.31209 | 0.37749 | *NCCRP1* |
| ENSG00000120533 | 8 | 110336553 | 110368182 | 79 | 16 | 54355 | 0.31196 | 0.37753 | *ENY2* |
| ENSG00000241058 | 10 | 18824490 | 18950551 | 581 | 35 | 53890 | 0.3118 | 0.3776 | *NSUN6* |
| ENSG00000149435 | 20 | 23955690 | 23979416 | 72 | 17 | 33221 | 0.31177 | 0.37761 | *GGTLC1* |
| ENSG00000121310 | 1 | 53351656 | 53402884 | 66 | 15 | 36400 | 0.31172 | 0.37763 | *ECHDC2* |
| ENSG00000100036 | 22 | 31021639 | 31075003 | 277 | 41 | 36729 | 0.31121 | 0.37782 | *SLC35E4* |
| ENSG00000181090 | 9 | 140503444 | 140774468 | 800 | 82 | 45072 | 0.31087 | 0.37795 | *EHMT1* |
| ENSG00000052749 | 10 | 99106115 | 99171127 | 277 | 13 | 53262 | 0.31074 | 0.378 | *RRP12* |
| ENSG00000198739 | 10 | 68675764 | 68869588 | 542 | 58 | 41918 | 0.31029 | 0.37817 | *LRRTM3* |
| ENSG00000158163 | 3 | 137770832 | 137844660 | 180 | 13 | 36501 | 0.31025 | 0.37818 | *DZIP1L* |
| ENSG00000167740 | 17 | 4036462 | 4100605 | 215 | 24 | 45718 | 0.31002 | 0.37827 | *CYB5D2* |
| ENSG00000205745 | 7 | 36108694 | 36135346 | 139 | 22 | 36270 | 0.31 | 0.37828 | *PP13004* |
| ENSG00000122644 | 7 | 12716481 | 12740559 | 135 | 26 | 43906 | 0.30994 | 0.3783 | *ARL4A* |
| ENSG00000147799 | 8 | 145744563 | 145921194 | 289 | 34 | 42281 | 0.30949 | 0.37847 | *ARHGAP39* |
| ENSG00000198816 | 19 | 7570178 | 7595912 | 71 | 17 | 52993 | 0.30941 | 0.3785 | *ZNF358* |
| ENSG00000008869 | 2 | 37185526 | 37321485 | 454 | 50 | 49562 | 0.30937 | 0.37852 | *HEATR5B* |
| ENSG00000082153 | 2 | 201665317 | 201698569 | 59 | 11 | 41258 | 0.30904 | 0.37864 | *BZW1* |
| ENSG00000240771 | 12 | 57993963 | 58023162 | 47 | 16 | 45657 | 0.30903 | 0.37865 | *ARHGEF25* |
| ENSG00000110497 | 11 | 46407964 | 46625675 | 228 | 31 | 48556 | 0.30895 | 0.37868 | *AMBRA1* |
| ENSG00000189050 | 17 | 58019601 | 58052122 | 57 | 12 | 36935 | 0.30883 | 0.37872 | *RNFT1* |
| ENSG00000268332 | 1 | 6090630 | 6111399 | 70 | 20 | 50813 | 0.30877 | 0.37875 | *AL035406.1* |

| | | | | | | | | | |
|---|---|---|---|---|---|---|---|---|---|
| ENSG00000007264 | 19 | 3767971 | 3812127 | 124 | 35 | 42602 | 0.3087 | 0.37877 | *MATK* |
| ENSG00000143653 | 1 | 246877349 | 246941439 | 276 | 27 | 42075 | 0.30869 | 0.37878 | *SCCPDH* |
| ENSG00000256574 | 10 | 45788102 | 45821056 | 165 | 14 | 39049 | 0.30854 | 0.37883 | *OR13A1* |
| ENSG00000115947 | 2 | 148677968 | 148789147 | 207 | 18 | 53562 | 0.30841 | 0.37889 | *ORC4* |
| ENSG00000143093 | 1 | 110564199 | 110627263 | 86 | 11 | 46894 | 0.30817 | 0.37898 | *STRIP1* |
| ENSG00000253729 | 8 | 48675669 | 48882743 | 180 | 45 | 45178 | 0.30801 | 0.37904 | *PRKDC* |
| ENSG00000161298 | 19 | 37085719 | 37129499 | 85 | 14 | 52765 | 0.30787 | 0.37909 | *ZNF382* |
| ENSG00000090534 | 3 | 184079723 | 184105932 | 65 | 19 | 46905 | 0.30784 | 0.3791 | *THPO* |
| ENSG00000105519 | 19 | 5901718 | 5925888 | 62 | 13 | 53118 | 0.30781 | 0.37911 | *CAPS* |
| ENSG00000115350 | 2 | 75175619 | 75207255 | 94 | 18 | 50007 | 0.30778 | 0.37912 | *POLE4* |
| ENSG00000198870 | 9 | 136233117 | 136281220 | 188 | 14 | 54607 | 0.30778 | 0.37912 | *C9orf96* |
| ENSG00000175322 | 18 | 14047456 | 14142489 | 257 | 29 | 50902 | 0.30777 | 0.37913 | *ZNF519* |
| ENSG00000007062 | 4 | 15954699 | 16096001 | 514 | 101 | 45758 | 0.30763 | 0.37918 | *PROM1* |
| ENSG00000121653 | 11 | 45897202 | 45938016 | 67 | 25 | 47477 | 0.30759 | 0.3792 | *MAPK8IP1* |
| ENSG00000132639 | 20 | 10189478 | 10298066 | 262 | 55 | 44805 | 0.30758 | 0.3792 | *SNAP25* |
| ENSG00000128274 | 22 | 43078127 | 43127304 | 274 | 33 | 41615 | 0.30755 | 0.37921 | *A4GALT* |
| ENSG00000139826 | 13 | 108860727 | 108896603 | 109 | 17 | 48304 | 0.30735 | 0.37929 | *ABHD13* |
| ENSG00000145911 | 5 | 177530444 | 177563088 | 96 | 16 | 52891 | 0.30714 | 0.37937 | *N4BP3* |
| ENSG00000255863 | 12 | 49305767 | 49343446 | 132 | 10 | 50188 | 0.30672 | 0.37953 | *AC073610.5* |
| ENSG00000139160 | 12 | 31790094 | 31836048 | 201 | 34 | 34942 | 0.30665 | 0.37955 | *METTL20* |
| ENSG00000114019 | 3 | 134064716 | 134104321 | 71 | 17 | 39288 | 0.30647 | 0.37962 | *AMOTL2* |
| ENSG00000143379 | 1 | 150888739 | 150947213 | 131 | 16 | 54066 | 0.30628 | 0.3797 | *SETDB1* |
| ENSG00000175115 | 11 | 65827834 | 66022218 | 377 | 32 | 49074 | 0.30609 | 0.37977 | *PACS1* |
| ENSG00000152078 | 1 | 95572894 | 95673163 | 339 | 44 | 43038 | 0.3059 | 0.37984 | *TMEM56* |
| ENSG00000121644 | 1 | 244806237 | 244882335 | 210 | 36 | 44330 | 0.30581 | 0.37987 | *DESI2* |
| ENSG00000084733 | 2 | 26246976 | 26370323 | 268 | 16 | 54048 | 0.30562 | 0.37995 | *RAB10* |
| ENSG00000183473 | 22 | 37590278 | 37618362 | 112 | 20 | 45677 | 0.30536 | 0.38004 | *SSTR3* |
| ENSG00000117500 | 1 | 93605299 | 93656285 | 116 | 17 | 46047 | 0.30529 | 0.38007 | *TMED5* |
| ENSG00000106003 | 7 | 2542163 | 2578811 | 121 | 31 | 41881 | 0.30525 | 0.38009 | *LFNG* |
| ENSG00000151376 | 11 | 86142150 | 86393678 | 1183 | 93 | 44351 | 0.30519 | 0.38011 | *ME3* |
| ENSG00000103534 | 16 | 19411818 | 19520435 | 499 | 50 | 37567 | 0.30512 | 0.38014 | *TMC5* |
| ENSG00000205858 | 7 | 16556505 | 16631193 | 279 | 47 | 49575 | 0.30455 | 0.38036 | *LRRC72* |
| ENSG00000001626 | 7 | 117095838 | 117366025 | 507 | 47 | 43855 | 0.30448 | 0.38038 | *CFTR* |
| ENSG00000107864 | 10 | 93796449 | 94060844 | 422 | 40 | 36882 | 0.30421 | 0.38049 | *CPEB3* |
| ENSG00000100439 | 14 | 23057146 | 23091265 | 111 | 27 | 40937 | 0.30399 | 0.38057 | *ABHD4* |
| ENSG00000107821 | 10 | 102811598 | 102837888 | 61 | 14 | 53716 | 0.30375 | 0.38066 | *KAZALD1* |

| | | | | | | | | | |
|---|---|---|---|---|---|---|---|---|---|
| ENSG00000103811 | 15 | 79203400 | 79251916 | 160 | 20 | 44618 | 0.30374 | 0.38066 | *CTSH* |
| ENSG00000102796 | 13 | 52332129 | 52388293 | 110 | 26 | 45211 | 0.30369 | 0.38068 | *DHRS12* |
| ENSG00000198580 | 7 | 6703376 | 6725991 | 102 | 19 | 39747 | 0.30359 | 0.38072 | *AC073343.1* |
| ENSG00000182759 | 8 | 144491352 | 144522576 | 165 | 30 | 42869 | 0.30355 | 0.38074 | *MAFA* |
| ENSG00000176925 | 11 | 4832551 | 4853686 | 101 | 10 | 46356 | 0.30353 | 0.38074 | *OR51F2* |
| ENSG00000163519 | 3 | 108531545 | 108583852 | 145 | 23 | 45926 | 0.30334 | 0.38082 | *TRAT1* |
| ENSG00000184575 | 12 | 64788130 | 64854907 | 120 | 21 | 44748 | 0.30323 | 0.38086 | *XPOT* |
| ENSG00000188313 | 3 | 146222967 | 146272651 | 223 | 26 | 54233 | 0.30316 | 0.38089 | *PLSCR1* |
| ENSG00000198001 | 12 | 44142747 | 44193346 | 129 | 18 | 42152 | 0.3031 | 0.38091 | *IRAK4* |
| ENSG00000214978 | 17 | 9684544 | 9735388 | 212 | 34 | 44388 | 0.30292 | 0.38098 | *RP11-477N12.3* |
| ENSG00000105696 | 19 | 18708240 | 18741849 | 124 | 23 | 41533 | 0.30272 | 0.38105 | *TMEM59L* |
| ENSG00000138622 | 15 | 73602200 | 73671605 | 212 | 40 | 44107 | 0.30244 | 0.38116 | *HCN4* |
| ENSG00000114638 | 3 | 118882364 | 118934000 | 225 | 45 | 48184 | 0.30235 | 0.38119 | *UPK1B* |
| ENSG00000137080 | 9 | 21155636 | 21176659 | 80 | 11 | 51891 | 0.30229 | 0.38122 | *IFNA21* |
| ENSG00000261652 | 15 | 55690746 | 55720962 | 134 | 14 | 33930 | 0.30196 | 0.38134 | *C15orf65* |
| ENSG00000005249 | 7 | 106675094 | 106812256 | 362 | 42 | 46050 | 0.30189 | 0.38137 | *PRKAR2B* |
| ENSG00000185758 | 4 | 184232917 | 184253579 | 57 | 13 | 45584 | 0.30186 | 0.38138 | *CLDN24* |
| ENSG00000112796 | 6 | 46116924 | 46148708 | 94 | 11 | 40086 | 0.30172 | 0.38143 | *ENPP5* |
| ENSG00000172457 | 11 | 56500303 | 56521287 | 89 | 16 | 50217 | 0.30171 | 0.38144 | *OR9G4* |
| ENSG00000186334 | 5 | 150646323 | 150693327 | 225 | 27 | 39760 | 0.30166 | 0.38145 | *SLC36A3* |
| ENSG00000130005 | 19 | 1387091 | 1411569 | 61 | 15 | 32619 | 0.30154 | 0.3815 | *GAMT* |
| ENSG00000132718 | 1 | 155819300 | 155864990 | 82 | 19 | 39855 | 0.30107 | 0.38168 | *SYT11* |
| ENSG00000197927 | 2 | 132469948 | 132534973 | 194 | 19 | 40595 | 0.30103 | 0.3817 | *C2orf27A* |
| ENSG00000169154 | 8 | 37781799 | 37807647 | 55 | 12 | 53325 | 0.30054 | 0.38188 | *GOT1L1* |
| ENSG00000132911 | 5 | 151761093 | 151822929 | 245 | 9 | 56196 | 0.30041 | 0.38193 | *NMUR2* |
| ENSG00000130383 | 19 | 5855837 | 5913798 | 285 | 41 | 52275 | 0.30041 | 0.38193 | *FUT5* |
| ENSG00000237787 | 3 | 153192284 | 153230486 | 151 | 22 | 36210 | 0.29996 | 0.38211 | *C3orf79* |
| ENSG00000250799 | 19 | 36280890 | 36314201 | 100 | 26 | 48103 | 0.2999 | 0.38213 | *PRODH2* |
| ENSG00000100360 | 22 | 37144246 | 37182300 | 81 | 13 | 45061 | 0.29957 | 0.38225 | *IFT27* |
| ENSG00000164116 | 4 | 156577863 | 156663501 | 307 | 44 | 46527 | 0.29936 | 0.38233 | *GUCY1A3* |
| ENSG00000085998 | 1 | 46644354 | 46695977 | 74 | 15 | 46333 | 0.29917 | 0.38241 | *POMGNT1* |
| ENSG00000062485 | 12 | 56655483 | 56704176 | 100 | 12 | 47067 | 0.29908 | 0.38244 | *CS* |
| ENSG00000116288 | 1 | 8004351 | 8055565 | 101 | 18 | 34578 | 0.299 | 0.38247 | *PARK7* |
| ENSG00000143195 | 1 | 166872443 | 166954719 | 169 | 25 | 54062 | 0.29889 | 0.38251 | *ILDR2* |
| ENSG00000176236 | 10 | 15127384 | 15149318 | 126 | 24 | 42166 | 0.2989 | 0.38251 | *C10orf111* |
| ENSG00000111726 | 12 | 22189108 | 22228608 | 87 | 15 | 37896 | 0.29877 | 0.38256 | *CMAS* |

| | | | | | | | | | |
|---|---|---|---|---|---|---|---|---|---|
| ENSG00000168826 | 4 | 4281924 | 4333513 | 231 | 22 | 42023 | 0.29872 | 0.38258 | *ZBTB49* |
| ENSG00000160993 | 7 | 102086685 | 102115323 | 113 | 18 | 42280 | 0.29872 | 0.38258 | *ALKBH4* |
| ENSG00000170390 | 4 | 150989426 | 151188609 | 660 | 67 | 46144 | 0.29868 | 0.38259 | *DCLK2* |
| ENSG00000197808 | 19 | 37118094 | 37167755 | 114 | 15 | 52791 | 0.29861 | 0.38262 | *ZNF461* |
| ENSG00000150093 | 10 | 33179247 | 33304720 | 464 | 56 | 37160 | 0.29854 | 0.38265 | *ITGB1* |
| ENSG00000203965 | 1 | 63979043 | 64048364 | 192 | 19 | 47639 | 0.29839 | 0.3827 | *EFCAB7* |
| ENSG00000134046 | 18 | 51669079 | 51761158 | 190 | 25 | 42819 | 0.29832 | 0.38273 | *MBD2* |
| ENSG00000124237 | 20 | 56715960 | 56746206 | 52 | 12 | 50756 | 0.29808 | 0.38282 | *C20orf85* |
| ENSG00000198812 | 12 | 69992351 | 70014942 | 66 | 10 | 51850 | 0.29801 | 0.38285 | *LRRC10* |
| ENSG00000172409 | 11 | 57406465 | 57439340 | 55 | 10 | 43281 | 0.29799 | 0.38286 | *CLP1* |
| ENSG00000101203 | 20 | 61914538 | 61976203 | 231 | 29 | 50823 | 0.29796 | 0.38287 | *COL20A1* |
| ENSG00000255633 | 6 | 62274008 | 62294534 | 170 | 12 | 48465 | 0.29781 | 0.38293 | *MTRNR2L9* |
| ENSG00000162738 | 1 | 160360376 | 160408468 | 133 | 24 | 42959 | 0.29777 | 0.38294 | *VANGL2* |
| ENSG00000205084 | 16 | 75562015 | 75600184 | 105 | 12 | 47113 | 0.29776 | 0.38294 | *TMEM231* |
| ENSG00000089639 | 19 | 19730285 | 19764476 | 68 | 19 | 51774 | 0.29773 | 0.38296 | *GMIP* |
| ENSG00000120093 | 17 | 46616232 | 46692274 | 202 | 34 | 41256 | 0.29761 | 0.383 | *HOXB3* |
| ENSG00000135250 | 7 | 104741151 | 105049755 | 630 | 37 | 35605 | 0.29744 | 0.38307 | *SRPK2* |
| ENSG00000162344 | 11 | 69503000 | 69529410 | 45 | 16 | 47206 | 0.29735 | 0.3831 | *FGF19* |
| ENSG00000136929 | 9 | 100679073 | 100717138 | 75 | 7 | 51683 | 0.2973 | 0.38312 | *HEMGN* |
| ENSG00000173627 | 1 | 183605541 | 183632451 | 149 | 5 | 23428 | 0.29715 | 0.38318 | *APOBEC4* |
| ENSG00000179088 | 12 | 103621369 | 103899749 | 713 | 66 | 40220 | 0.29713 | 0.38318 | *C12orf42* |
| ENSG00000141098 | 16 | 67698434 | 67763324 | 90 | 18 | 42459 | 0.297 | 0.38323 | *GFOD2* |
| ENSG00000116285 | 1 | 8054464 | 8096368 | 72 | 12 | 36028 | 0.29691 | 0.38327 | *ERRFI1* |
| ENSG00000130856 | 18 | 74524563 | 74692683 | 498 | 46 | 48941 | 0.29684 | 0.38329 | *ZNF236* |
| ENSG00000163520 | 3 | 13563824 | 13689922 | 516 | 74 | 46680 | 0.2968 | 0.38331 | *FBLN2* |
| ENSG00000126545 | 4 | 70786799 | 70822289 | 96 | 14 | 48196 | 0.2967 | 0.38335 | *CSN1S1* |
| ENSG00000169398 | 8 | 141657999 | 142022315 | 619 | 37 | 38455 | 0.29652 | 0.38341 | *PTK2* |
| ENSG00000131023 | 6 | 149969289 | 150049392 | 130 | 10 | 35049 | 0.29651 | 0.38342 | *LATS1* |
| ENSG00000076356 | 1 | 208185587 | 208427665 | 732 | 108 | 48615 | 0.29642 | 0.38346 | *PLXNA2* |
| ENSG00000083937 | 3 | 87266421 | 87314698 | 137 | 23 | 47030 | 0.29608 | 0.38359 | *CHMP2B* |
| ENSG00000104047 | 15 | 49903177 | 49947333 | 137 | 15 | 31921 | 0.29606 | 0.38359 | *DTWD1* |
| ENSG00000178105 | 11 | 108525752 | 108821657 | 623 | 42 | 48033 | 0.296 | 0.38361 | *DDX10* |
| ENSG00000115998 | 2 | 70367012 | 70485747 | 279 | 25 | 40312 | 0.29596 | 0.38363 | *C2orf42* |
| ENSG00000196326 | 10 | 5186335 | 5254449 | 386 | 31 | 37686 | 0.29597 | 0.38363 | *AKR1CL1* |
| ENSG00000118702 | 20 | 35869489 | 35900238 | 70 | 18 | 40787 | 0.29577 | 0.3837 | *GHRH* |
| ENSG00000177875 | 12 | 48567366 | 48589709 | 84 | 11 | 49756 | 0.29574 | 0.38371 | *C12orf68* |

| | | | | | | | | | |
|---|---|---|---|---|---|---|---|---|---|
| ENSG00000165215 | 7 | 73173328 | 73194600 | 60 | 10 | 42198 | 0.29548 | 0.38381 | *CLDN3* |
| ENSG00000165556 | 13 | 28526274 | 28555276 | 89 | 15 | 45421 | 0.29547 | 0.38382 | *CDX2* |
| ENSG00000138166 | 10 | 112247596 | 112281302 | 73 | 13 | 49072 | 0.2953 | 0.38388 | *DUSP5* |
| ENSG00000196743 | 5 | 150581711 | 150660001 | 278 | 48 | 40236 | 0.29497 | 0.38401 | *GM2A* |
| ENSG00000125148 | 16 | 56632111 | 56653409 | 97 | 25 | 43483 | 0.29493 | 0.38402 | *MT2A* |
| ENSG00000174500 | 3 | 111829688 | 111862152 | 95 | 19 | 49740 | 0.29474 | 0.38409 | *GCSAM* |
| ENSG00000198626 | 1 | 237195505 | 238007288 | 3107 | 281 | 44261 | 0.29469 | 0.38411 | *RYR2* |
| ENSG00000115677 | 2 | 242156679 | 242266476 | 308 | 35 | 47931 | 0.29428 | 0.38427 | *HDLBP* |
| ENSG00000152022 | 1 | 145467085 | 145511669 | 19 | 11 | 44144 | 0.29412 | 0.38433 | *LIX1L* |
| ENSG00000181392 | 19 | 36484209 | 36509695 | 42 | 12 | 40455 | 0.2941 | 0.38434 | *SYNE4* |
| ENSG00000227059 | 12 | 133784898 | 133822681 | 117 | 19 | 44274 | 0.29407 | 0.38435 | *ANHX* |
| ENSG00000133316 | 11 | 62589814 | 62619281 | 87 | 9 | 36828 | 0.29389 | 0.38442 | *WDR74* |
| ENSG00000205038 | 8 | 110364706 | 110552559 | 601 | 37 | 48931 | 0.29382 | 0.38445 | *PKHD1L1* |
| ENSG00000171045 | 8 | 143283441 | 143494601 | 898 | 82 | 46534 | 0.29376 | 0.38447 | *TSNARE1* |
| ENSG00000136352 | 14 | 36975602 | 37000354 | 56 | 16 | 39626 | 0.29376 | 0.38447 | *NKX2-1* |
| ENSG00000158006 | 1 | 26276258 | 26334648 | 153 | 13 | 33940 | 0.29367 | 0.38451 | *PAFAH2* |
| ENSG00000198445 | 22 | 17061667 | 17083700 | 56 | 17 | 40242 | 0.29356 | 0.38455 | *CCT8L2* |
| ENSG00000253626 | 10 | 81262357 | 81286188 | 99 | 16 | 46509 | 0.29334 | 0.38463 | *EIF5AL1* |
| ENSG00000069482 | 11 | 68441247 | 68468643 | 61 | 12 | 50513 | 0.29329 | 0.38465 | *GAL* |
| ENSG00000132359 | 17 | 2670350 | 2951033 | 1026 | 198 | 46832 | 0.2933 | 0.38465 | *RAP1GAP2* |
| ENSG00000106006 | 7 | 27175015 | 27200222 | 64 | 13 | 34262 | 0.29322 | 0.38468 | *HOXA6* |
| ENSG00000197956 | 1 | 153497075 | 153518720 | 43 | 7 | 36685 | 0.2931 | 0.38472 | *S100A6* |
| ENSG00000186409 | 1 | 42919001 | 43130335 | 712 | 37 | 46976 | 0.29304 | 0.38474 | *CCDC30* |
| ENSG00000097046 | 1 | 91956408 | 92001321 | 139 | 22 | 47089 | 0.29303 | 0.38475 | *CDC7* |
| ENSG00000177476 | 1 | 247758856 | 247779838 | 60 | 12 | 55074 | 0.29287 | 0.38481 | *OR2G3* |
| ENSG00000145779 | 5 | 118594387 | 118745383 | 553 | 63 | 37865 | 0.29286 | 0.38481 | *TNFAIP8* |
| ENSG00000049247 | 1 | 7893143 | 7923572 | 146 | 33 | 49028 | 0.2928 | 0.38484 | *UTS2* |
| ENSG00000265817 | 8 | 95380605 | 95459180 | 274 | 25 | 37710 | 0.29237 | 0.385 | *FSBP* |
| ENSG00000117543 | 1 | 101445179 | 101501644 | 85 | 11 | 51418 | 0.29212 | 0.3851 | *DPH5* |
| ENSG00000152683 | 2 | 32380933 | 32459448 | 237 | 27 | 52671 | 0.29205 | 0.38513 | *SLC30A6* |
| ENSG00000162981 | 2 | 14762810 | 14800933 | 64 | 13 | 42953 | 0.29174 | 0.38524 | *FAM84A* |
| ENSG00000149256 | 11 | 78353876 | 79161992 | 2362 | 270 | 43576 | 0.29136 | 0.38539 | *TENM4* |
| ENSG00000019505 | 11 | 45251852 | 45317870 | 187 | 23 | 35965 | 0.29134 | 0.3854 | *SYT13* |
| ENSG00000131379 | 3 | 14706606 | 14824541 | 343 | 57 | 44347 | 0.29129 | 0.38542 | *C3orf20* |
| ENSG00000181038 | 17 | 74712912 | 74740018 | 22 | 10 | 44890 | 0.29117 | 0.38546 | *METTL23* |
| ENSG00000187550 | 19 | 56031100 | 56058456 | 152 | 37 | 44236 | 0.29118 | 0.38546 | *SBK2* |

| | | | | | | | | | |
|---|---|---|---|---|---|---|---|---|---|
| ENSG00000166004 | 11 | 93384805 | 93473522 | 172 | 23 | 35770 | 0.29112 | 0.38548 | *KIAA1731* |
| ENSG00000172782 | 17 | 72863428 | 72899708 | 92 | 16 | 32103 | 0.29093 | 0.38555 | *FADS6* |
| ENSG00000187605 | 2 | 74219840 | 74345303 | 295 | 32 | 41674 | 0.29089 | 0.38557 | *TET3* |
| ENSG00000077063 | 7 | 117340705 | 117524193 | 417 | 73 | 45368 | 0.29078 | 0.38561 | *CTTNBP2* |
| ENSG00000148123 | 9 | 103780991 | 104097417 | 1157 | 109 | 46544 | 0.29076 | 0.38562 | *LPPR1* |
| ENSG00000133661 | 10 | 81687496 | 81752370 | 349 | 31 | 41350 | 0.29067 | 0.38565 | *SFTPD* |
| ENSG00000111275 | 12 | 112194691 | 112257782 | 57 | 12 | 51854 | 0.29064 | 0.38566 | *ALDH2* |
| ENSG00000155657 | 2 | 179380716 | 179705529 | 608 | 72 | 38089 | 0.29046 | 0.38573 | *TTN* |
| ENSG00000143512 | 1 | 222685602 | 222731445 | 159 | 19 | 40109 | 0.2903 | 0.38579 | *HHIPL2* |
| ENSG00000112983 | 5 | 137465455 | 137524675 | 78 | 12 | 54112 | 0.29029 | 0.3858 | *BRD8* |
| ENSG00000235631 | 7 | 122331718 | 122353021 | 56 | 10 | 53033 | 0.29014 | 0.38585 | *RNF148* |
| ENSG00000169884 | 12 | 49349123 | 49375546 | 61 | 9 | 44774 | 0.29002 | 0.3859 | *WNT10B* |
| ENSG00000168843 | 4 | 162295049 | 163095187 | 3136 | 220 | 44196 | 0.28995 | 0.38593 | *FSTL5* |
| ENSG00000145860 | 5 | 158574417 | 158647061 | 158 | 24 | 50655 | 0.28973 | 0.38601 | *RNF145* |
| ENSG00000163605 | 3 | 73035936 | 73128350 | 387 | 20 | 46391 | 0.28949 | 0.3861 | *PPP4R2* |
| ENSG00000003056 | 12 | 9082959 | 9112551 | 93 | 10 | 45504 | 0.28946 | 0.38611 | *M6PR* |
| ENSG00000137331 | 6 | 30700976 | 30722331 | 105 | 11 | 23560 | 0.2893 | 0.38618 | *IER3* |
| ENSG00000123374 | 12 | 56350553 | 56376568 | 44 | 15 | 45642 | 0.28927 | 0.38619 | *CDK2* |
| ENSG00000129657 | 17 | 75072798 | 75223179 | 636 | 90 | 49802 | 0.28891 | 0.38632 | *SEC14L1* |
| ENSG00000161798 | 12 | 50345653 | 50369464 | 40 | 13 | 28396 | 0.28872 | 0.3864 | *AQP5* |
| ENSG00000178381 | 7 | 1181707 | 1210395 | 259 | 19 | 46140 | 0.2885 | 0.38648 | *ZFAND2A* |
| ENSG00000147724 | 8 | 139132266 | 139519065 | 1406 | 153 | 38738 | 0.2884 | 0.38652 | *FAM135B* |
| ENSG00000170953 | 11 | 124402578 | 124423575 | 73 | 15 | 49791 | 0.28835 | 0.38654 | *OR8B12* |
| ENSG00000187951 | 15 | 30906697 | 31075196 | 402 | 31 | 38937 | 0.28832 | 0.38655 | *ARHGAP11B* |
| ENSG00000107018 | 9 | 5324969 | 5349873 | 83 | 12 | 37690 | 0.28827 | 0.38657 | *RLN1* |
| ENSG00000254732 | 11 | 57499635 | 57570715 | 108 | 17 | 46631 | 0.28811 | 0.38663 | *RP11-691N7.6* |
| ENSG00000166148 | 12 | 63529014 | 63554722 | 85 | 14 | 30887 | 0.28811 | 0.38663 | *AVPR1A* |
| ENSG00000168481 | 8 | 21994338 | 22024597 | 82 | 24 | 51131 | 0.28806 | 0.38665 | *LGI3* |
| ENSG00000272514 | 6 | 88107701 | 88184183 | 288 | 22 | 52414 | 0.28804 | 0.38666 | *C6ORF165* |
| ENSG00000175220 | 11 | 46688630 | 46732165 | 63 | 14 | 43818 | 0.288 | 0.38667 | *ARHGAP1* |
| ENSG00000197502 | 14 | 50290810 | 50321552 | 37 | 14 | 51598 | 0.28798 | 0.38668 | *AL627171.1* |
| ENSG00000117407 | 1 | 44388992 | 44412913 | 60 | 11 | 40490 | 0.28792 | 0.3867 | *ARTN* |
| ENSG00000256316 | 6 | 26240370 | 26260835 | 57 | 11 | 45343 | 0.28792 | 0.38671 | *HIST1H3F* |
| ENSG00000196961 | 19 | 50260225 | 50320370 | 182 | 32 | 49482 | 0.28786 | 0.38673 | *AP2A1* |
| ENSG00000112238 | 6 | 100044606 | 100073454 | 83 | 21 | 44099 | 0.2878 | 0.38675 | *PRDM13* |
| ENSG00000063015 | 17 | 27271919 | 27343458 | 105 | 20 | 50753 | 0.28748 | 0.38687 | *SEZ6* |

| | | | | | | | | | |
|---|---|---|---|---|---|---|---|---|---|
| ENSG00000123415 | 12 | 54548529 | 54592778 | 98 | 17 | 43716 | 0.28734 | 0.38693 | *SMUG1* |
| ENSG00000267740 | 19 | 5856182 | 5913798 | 284 | 41 | 52263 | 0.28717 | 0.38699 | *AC024592.12* |
| ENSG00000139146 | 12 | 31423518 | 31489992 | 69 | 21 | 41536 | 0.28699 | 0.38706 | *FAM60A* |
| ENSG00000216560 | 4 | 3568596 | 3602438 | 58 | 17 | 29015 | 0.28691 | 0.38709 | *LINC00955* |
| ENSG00000143217 | 1 | 161030785 | 161069389 | 97 | 26 | 49093 | 0.28675 | 0.38715 | *PVRL4* |
| ENSG00000268846 | 2 | 187357219 | 187377356 | 47 | 9 | 55983 | 0.28651 | 0.38724 | *AC018867.2* |
| ENSG00000162618 | 1 | 79345449 | 79482403 | 650 | 54 | 54092 | 0.28644 | 0.38727 | *ELTD1* |
| ENSG00000038295 | 4 | 166784410 | 167035047 | 640 | 109 | 43871 | 0.28617 | 0.38737 | *TLL1* |
| ENSG00000159239 | 2 | 74631304 | 74658718 | 35 | 6 | 49992 | 0.28609 | 0.3874 | *C2orf81* |
| ENSG00000107611 | 10 | 16855963 | 17181830 | 1312 | 135 | 43506 | 0.28607 | 0.38741 | *CUBN* |
| ENSG00000137868 | 15 | 74461807 | 74514608 | 137 | 19 | 32512 | 0.28586 | 0.38749 | *STRA6* |
| ENSG00000169242 | 1 | 155089936 | 155117333 | 57 | 13 | 39100 | 0.28532 | 0.3877 | *EFNA1* |
| ENSG00000077380 | 2 | 172533919 | 172614930 | 195 | 24 | 45423 | 0.28521 | 0.38774 | *DYNC1I2* |
| ENSG00000140835 | 16 | 71549136 | 71582649 | 71 | 18 | 38423 | 0.28506 | 0.3878 | *CHST4* |
| ENSG00000134389 | 1 | 196936667 | 196988804 | 107 | 17 | 35164 | 0.28479 | 0.3879 | *CFHR5* |
| ENSG00000197142 | 10 | 114123776 | 114198138 | 168 | 22 | 53307 | 0.28475 | 0.38792 | *ACSL5* |
| ENSG00000197818 | 20 | 48419250 | 48518779 | 354 | 43 | 48254 | 0.28451 | 0.38801 | *SLC9A8* |
| ENSG00000172771 | 3 | 129110164 | 129157494 | 127 | 20 | 40272 | 0.28437 | 0.38806 | *EFCAB12* |
| ENSG00000120029 | 10 | 103595356 | 103825950 | 346 | 43 | 51241 | 0.28426 | 0.38811 | *C10orf76* |
| ENSG00000160447 | 9 | 131454802 | 131493197 | 35 | 19 | 47889 | 0.28393 | 0.38823 | *PKN3* |
| ENSG00000151414 | 1 | 198116093 | 198301550 | 595 | 41 | 34768 | 0.28369 | 0.38832 | *NEK7* |
| ENSG00000133958 | 14 | 93789565 | 94184222 | 1011 | 57 | 49807 | 0.28365 | 0.38834 | *UNC79* |
| ENSG00000111540 | 12 | 56357697 | 56398490 | 79 | 15 | 43534 | 0.2836 | 0.38836 | *RAB5B* |
| ENSG00000167779 | 12 | 53481220 | 53506129 | 82 | 11 | 28836 | 0.28353 | 0.38838 | *IGFBP6* |
| ENSG00000117724 | 1 | 214766538 | 214847931 | 308 | 32 | 44803 | 0.28319 | 0.38851 | *CENPF* |
| ENSG00000006659 | 19 | 40184946 | 40210084 | 69 | 11 | 45854 | 0.28302 | 0.38858 | *LGALS14* |
| ENSG00000167578 | 19 | 41274121 | 41312847 | 146 | 25 | 45456 | 0.28303 | 0.38858 | *RAB4B* |
| ENSG00000255384 | 11 | 118293836 | 118315921 | 42 | 13 | 40303 | 0.28275 | 0.38869 | *RP11-770J1.4* |
| ENSG00000187017 | 1 | 6474848 | 6531430 | 188 | 25 | 37528 | 0.28268 | 0.38871 | *ESPN* |
| ENSG00000198478 | 6 | 80331000 | 80423372 | 253 | 36 | 49224 | 0.28264 | 0.38873 | *SH3BGRL2* |
| ENSG00000131446 | 5 | 180207541 | 180252652 | 228 | 19 | 45413 | 0.28261 | 0.38874 | *MGAT1* |
| ENSG00000188739 | 1 | 235284498 | 235334772 | 135 | 14 | 47596 | 0.28253 | 0.38877 | *RBM34* |
| ENSG00000187583 | 1 | 891877 | 921245 | 106 | 26 | 39592 | 0.28249 | 0.38878 | *PLEKHN1* |
| ENSG00000162910 | 1 | 228284380 | 228307013 | 31 | 11 | 49396 | 0.28234 | 0.38884 | *MRPL55* |
| ENSG00000013297 | 3 | 170126653 | 170588169 | 1374 | 117 | 44939 | 0.28236 | 0.38884 | *CLDN11* |
| ENSG00000115318 | 2 | 74749541 | 74792817 | 33 | 10 | 38557 | 0.28198 | 0.38898 | *LOXL3* |

| | | | | | | | | | |
|---|---|---|---|---|---|---|---|---|---|
| ENSG00000006534 | 11 | 67766048 | 67806744 | 103 | 18 | 51780 | 0.28185 | 0.38903 | *ALDH3B1* |
| ENSG00000178021 | 2 | 54470315 | 54493409 | 96 | 12 | 55447 | 0.28177 | 0.38906 | *TSPYL6* |
| ENSG00000187650 | 19 | 5894869 | 5920864 | 83 | 17 | 53480 | 0.28174 | 0.38907 | *VMAC* |
| ENSG00000251692 | 16 | 1525887 | 1548982 | 158 | 32 | 41004 | 0.28166 | 0.3891 | *PTX4* |
| ENSG00000113615 | 5 | 133974479 | 134073513 | 149 | 28 | 34758 | 0.28156 | 0.38914 | *SEC24A* |
| ENSG00000186088 | 7 | 76930068 | 77055717 | 455 | 48 | 41976 | 0.28146 | 0.38918 | *GSAP* |
| ENSG00000164600 | 7 | 31367075 | 31390508 | 44 | 13 | 39734 | 0.28129 | 0.38925 | *NEUROD6* |
| ENSG00000182816 | 21 | 31733709 | 31754557 | 76 | 17 | 44723 | 0.28125 | 0.38926 | *KRTAP13-2* |
| ENSG00000115760 | 2 | 32572096 | 32853966 | 680 | 61 | 53096 | 0.28121 | 0.38928 | *BIRC6* |
| ENSG00000168300 | 8 | 52720140 | 52821735 | 296 | 30 | 42118 | 0.28115 | 0.3893 | *PCMTD1* |
| ENSG00000198301 | 4 | 76852103 | 76922115 | 210 | 23 | 49610 | 0.28103 | 0.38934 | *SDAD1* |
| ENSG00000136449 | 17 | 48575745 | 48618862 | 145 | 23 | 43454 | 0.28102 | 0.38935 | *MYCBPAP* |
| ENSG00000042493 | 2 | 85611871 | 85655555 | 169 | 26 | 48765 | 0.28093 | 0.38938 | *CAPG* |
| ENSG00000155026 | 7 | 5955181 | 6020314 | 22 | 14 | 41140 | 0.28088 | 0.3894 | *RSPH10B* |
| ENSG00000137145 | 9 | 19220433 | 19384139 | 551 | 45 | 47460 | 0.28058 | 0.38952 | *DENND4C* |
| ENSG00000164048 | 3 | 48272590 | 48350743 | 154 | 12 | 41894 | 0.28055 | 0.38953 | *ZNF589* |
| ENSG00000235705 | 10 | 81256126 | 81280678 | 107 | 17 | 48530 | 0.2805 | 0.38955 | *AL133481.1* |
| ENSG00000091140 | 7 | 107521415 | 107582175 | 142 | 15 | 31594 | 0.28044 | 0.38957 | *DLD* |
| ENSG00000054803 | 20 | 54562496 | 54590528 | 41 | 14 | 41795 | 0.28036 | 0.3896 | *CBLN4* |
| ENSG00000106355 | 7 | 32514951 | 32544895 | 129 | 14 | 41952 | 0.28026 | 0.38964 | *LSM5* |
| ENSG00000247121 | 5 | 96139731 | 96281513 | 732 | 50 | 48532 | 0.28009 | 0.3897 | *CTD-2260A17.2* |
| ENSG00000071794 | 3 | 148737914 | 148814341 | 181 | 22 | 46984 | 0.28001 | 0.38973 | *HLTF* |
| ENSG00000122507 | 7 | 33158856 | 33655680 | 1703 | 115 | 48833 | 0.27995 | 0.38976 | *BBS9* |
| ENSG00000188888 | 17 | 36471413 | 36509730 | 71 | 20 | 42055 | 0.27981 | 0.38981 | *GPR179* |
| ENSG00000254614 | 11 | 64935077 | 64959305 | 52 | 14 | 44405 | 0.27975 | 0.38983 | *AP003068.23* |
| ENSG00000203811 | 1 | 149801110 | 149822765 | 15 | 5 | 42739 | 0.27966 | 0.38987 | *HIST2H3C* |
| ENSG00000183558 | 1 | 149803505 | 149824478 | 15 | 5 | 42739 | 0.27966 | 0.38987 | *HIST2H2AA3* |
| ENSG00000203812 | 1 | 149812643 | 149833191 | 15 | 5 | 42739 | 0.27966 | 0.38987 | *HIST2H2AA4* |
| ENSG00000203852 | 1 | 149814181 | 149835836 | 15 | 5 | 42739 | 0.27966 | 0.38987 | *HIST2H3A* |
| ENSG00000186073 | 15 | 36861812 | 37112449 | 769 | 97 | 51293 | 0.27965 | 0.38987 | *C15orf41* |
| ENSG00000180263 | 12 | 95460525 | 95621258 | 606 | 58 | 51052 | 0.2796 | 0.38989 | *FGD6* |
| ENSG00000145708 | 5 | 76238538 | 76286983 | 139 | 21 | 42533 | 0.27934 | 0.38999 | *CRHBP* |
| ENSG00000071537 | 14 | 81927893 | 82010205 | 149 | 37 | 44728 | 0.27904 | 0.39011 | *SEL1L* |
| ENSG00000109205 | 4 | 71052213 | 71080293 | 110 | 9 | 32738 | 0.27898 | 0.39013 | *ODAM* |
| ENSG00000271271 | 4 | 70444135 | 70515360 | 357 | 30 | 53141 | 0.27882 | 0.39019 | *UGT2A2* |
| ENSG00000169439 | 8 | 97495579 | 97634000 | 411 | 73 | 38797 | 0.27878 | 0.39021 | *SDC2* |

| | | | | | | | | | |
|---|---|---|---|---|---|---|---|---|---|
| ENSG00000140983 | 16 | 708086 | 734174 | 61 | 14 | 35309 | 0.27864 | 0.39026 | *RHOT2* |
| ENSG00000162592 | 1 | 3658962 | 3698209 | 167 | 31 | 46633 | 0.27838 | 0.39036 | *CCDC27* |
| ENSG00000165417 | 14 | 81631796 | 81697721 | 147 | 22 | 45266 | 0.27821 | 0.39043 | *GTF2A1* |
| ENSG00000038427 | 5 | 82757284 | 82888122 | 353 | 66 | 49371 | 0.27803 | 0.3905 | *VCAN* |
| ENSG00000198839 | 7 | 111836643 | 111993151 | 459 | 37 | 31720 | 0.27762 | 0.39065 | *ZNF277* |
| ENSG00000122965 | 12 | 114244543 | 114414176 | 747 | 68 | 44122 | 0.27761 | 0.39065 | *RBM19* |
| ENSG00000168955 | 2 | 228216872 | 228256711 | 188 | 37 | 52005 | 0.27729 | 0.39078 | *TM4SF20* |
| ENSG00000102854 | 16 | 800762 | 828865 | 145 | 34 | 38173 | 0.2772 | 0.39081 | *MSLN* |
| ENSG00000263155 | 15 | 57874139 | 57987562 | 497 | 54 | 46681 | 0.27621 | 0.39119 | *MYZAP* |
| ENSG00000146360 | 6 | 110289514 | 110311921 | 66 | 16 | 44983 | 0.27598 | 0.39128 | *GPR6* |
| ENSG00000175029 | 10 | 126666421 | 126859739 | 814 | 126 | 46047 | 0.27594 | 0.3913 | *CTBP2* |
| ENSG00000171234 | 4 | 69907081 | 69988705 | 455 | 18 | 29827 | 0.27578 | 0.39136 | *UGT2B7* |
| ENSG00000122218 | 1 | 160249063 | 160323190 | 141 | 26 | 37621 | 0.27569 | 0.39139 | *COPA* |
| ENSG00000111405 | 12 | 48093517 | 48129350 | 115 | 17 | 46978 | 0.27554 | 0.39145 | *ENDOU* |
| ENSG00000162959 | 2 | 32080129 | 32246299 | 410 | 35 | 54725 | 0.27545 | 0.39148 | *MEMO1* |
| ENSG00000125746 | 19 | 46100252 | 46158887 | 185 | 33 | 47782 | 0.27536 | 0.39152 | *EML2* |
| ENSG00000163788 | 3 | 43318004 | 43476256 | 228 | 38 | 47919 | 0.2752 | 0.39158 | *SNRK* |
| ENSG00000144452 | 2 | 215786266 | 216013151 | 576 | 77 | 38236 | 0.27492 | 0.39169 | *ABCA12* |
| ENSG00000176593 | 19 | 58503429 | 58532600 | 123 | 16 | 50203 | 0.27483 | 0.39172 | *CTD-2368P22.1* |
| ENSG00000186881 | 9 | 107256455 | 107277547 | 81 | 11 | 48396 | 0.27478 | 0.39174 | *OR13F1* |
| ENSG00000182217 | 1 | 149815607 | 149842714 | 11 | 4 | 46796 | 0.27477 | 0.39175 | *HIST2H4B* |
| ENSG00000132286 | 11 | 6492677 | 6515909 | 48 | 9 | 49579 | 0.27477 | 0.39175 | *TIMM10B* |
| ENSG00000137707 | 11 | 111328251 | 111393079 | 145 | 25 | 45540 | 0.27448 | 0.39186 | *BTG4* |
| ENSG00000142541 | 19 | 49980811 | 50005565 | 84 | 10 | 31293 | 0.27437 | 0.3919 | *RPL13A* |
| ENSG00000162734 | 1 | 160165127 | 160195166 | 38 | 13 | 41746 | 0.27432 | 0.39192 | *PEA15* |
| ENSG00000088179 | 2 | 120507207 | 120751394 | 489 | 35 | 51458 | 0.27424 | 0.39195 | *PTPN4* |
| ENSG00000108963 | 17 | 1923404 | 1956724 | 94 | 24 | 48746 | 0.27414 | 0.39199 | *DPH1* |
| ENSG00000137364 | 6 | 18118542 | 18165305 | 186 | 25 | 53388 | 0.274 | 0.39204 | *TPMT* |
| ENSG00000186132 | 2 | 120049801 | 120134404 | 340 | 40 | 52553 | 0.27387 | 0.39209 | *C2orf76* |
| ENSG00000107362 | 9 | 74467368 | 74535847 | 101 | 17 | 42871 | 0.27363 | 0.39218 | *ABHD17B* |
| ENSG00000183161 | 11 | 22634079 | 22657387 | 34 | 10 | 38558 | 0.27355 | 0.39222 | *FANCF* |
| ENSG00000113391 | 5 | 92943775 | 93457404 | 811 | 59 | 49863 | 0.27326 | 0.39233 | *FAM172A* |
| ENSG00000197275 | 8 | 95374188 | 95497337 | 481 | 34 | 37611 | 0.27288 | 0.39247 | *RAD54B* |
| ENSG00000243955 | 6 | 52646462 | 52678708 | 100 | 11 | 36570 | 0.27282 | 0.39249 | *GSTA1* |
| ENSG00000005483 | 7 | 104644626 | 104764808 | 170 | 22 | 35505 | 0.27268 | 0.39255 | *KMT2E* |
| ENSG00000204580 | 6 | 30834198 | 30877933 | 143 | 26 | 35807 | 0.27266 | 0.39256 | *DDR1* |

| | | | | | | | | | |
|---|---|---|---|---|---|---|---|---|---|
| ENSG00000135374 | 11 | 34490340 | 34545352 | 200 | 25 | 37875 | 0.27266 | 0.39256 | *ELF5* |
| ENSG00000175325 | 5 | 177409236 | 177433243 | 88 | 22 | 49474 | 0.2726 | 0.39258 | *PROP1* |
| ENSG00000084764 | 2 | 27183480 | 27260064 | 106 | 12 | 48358 | 0.27253 | 0.39261 | *MAPRE3* |
| ENSG00000223802 | 19 | 18969361 | 19017536 | 124 | 29 | 46717 | 0.272 | 0.39281 | *CERS1* |
| ENSG00000106304 | 7 | 123555286 | 123621468 | 160 | 24 | 47896 | 0.27189 | 0.39285 | *SPAM1* |
| ENSG00000172367 | 11 | 119046166 | 119070932 | 30 | 10 | 39107 | 0.27189 | 0.39285 | *PDZD3* |
| ENSG00000174358 | 5 | 1191710 | 1235232 | 206 | 48 | 46113 | 0.27183 | 0.39288 | *SLC6A19* |
| ENSG00000222000 | 2 | 98937852 | 98982468 | 187 | 19 | 38075 | 0.27127 | 0.39309 | *AC092675.3* |
| ENSG00000132670 | 20 | 2834830 | 3029722 | 364 | 35 | 37085 | 0.27127 | 0.39309 | *PTPRA* |
| ENSG00000145113 | 3 | 195463636 | 195549148 | 319 | 41 | 40986 | 0.27105 | 0.39318 | *MUC4* |
| ENSG00000009765 | 6 | 150680028 | 150737105 | 281 | 34 | 28168 | 0.27103 | 0.39318 | *IYD* |
| ENSG00000107731 | 10 | 72962327 | 73072621 | 396 | 76 | 43992 | 0.27098 | 0.3932 | *UNC5B* |
| ENSG00000188783 | 1 | 203434956 | 203470480 | 134 | 22 | 49849 | 0.27079 | 0.39327 | *PRELP* |
| ENSG00000125861 | 20 | 3629939 | 3654046 | 124 | 25 | 46379 | 0.27051 | 0.39338 | *GFRA4* |
| ENSG00000125743 | 19 | 46180712 | 46205827 | 60 | 13 | 54291 | 0.27029 | 0.39347 | *SNRPD2* |
| ENSG00000165091 | 9 | 75126717 | 75461267 | 1130 | 52 | 41389 | 0.27016 | 0.39352 | *TMC1* |
| ENSG00000116752 | 1 | 115100178 | 115134260 | 70 | 15 | 51926 | 0.2697 | 0.3937 | *BCAS2* |
| ENSG00000173908 | 17 | 38938455 | 38966211 | 97 | 23 | 45129 | 0.26963 | 0.39372 | *KRT28* |
| ENSG00000213085 | 1 | 159832154 | 159879953 | 144 | 21 | 41870 | 0.26962 | 0.39373 | *CCDC19* |
| ENSG00000198585 | 3 | 131090515 | 131117674 | 48 | 12 | 40169 | 0.26955 | 0.39375 | *NUDT16* |
| ENSG00000131759 | 17 | 38455444 | 38523094 | 101 | 14 | 48758 | 0.26941 | 0.39381 | *RARA* |
| ENSG00000137871 | 15 | 56912379 | 57220769 | 678 | 53 | 43175 | 0.26915 | 0.39391 | *ZNF280D* |
| ENSG00000136883 | 9 | 116838406 | 116871507 | 146 | 24 | 48423 | 0.26907 | 0.39394 | *KIF12* |
| ENSG00000183067 | 21 | 41107334 | 41184023 | 472 | 34 | 41659 | 0.26869 | 0.39408 | *IGSF5* |
| ENSG00000119686 | 14 | 76034960 | 76139557 | 243 | 53 | 38562 | 0.26857 | 0.39413 | *FLVCR2* |
| ENSG00000138771 | 4 | 77346253 | 77714406 | 1151 | 113 | 46577 | 0.26854 | 0.39414 | *SHROOM3* |
| ENSG00000063854 | 16 | 1835621 | 1887195 | 496 | 27 | 44703 | 0.26855 | 0.39414 | *HAGH* |
| ENSG00000166780 | 16 | 15518152 | 15728885 | 395 | 66 | 42510 | 0.26831 | 0.39423 | *C16orf45* |
| ENSG00000165813 | 10 | 115870621 | 115943979 | 161 | 19 | 53242 | 0.26823 | 0.39426 | *C10orf118* |
| ENSG00000131323 | 14 | 103233813 | 103387837 | 373 | 34 | 39155 | 0.26812 | 0.3943 | *TRAF3* |
| ENSG00000181135 | 8 | 144756622 | 144806068 | 146 | 35 | 36380 | 0.26807 | 0.39432 | *ZNF707* |
| ENSG00000090339 | 19 | 10371511 | 10407291 | 85 | 26 | 44540 | 0.26791 | 0.39439 | *ICAM1* |
| ENSG00000184305 | 4 | 91038686 | 92533064 | 5226 | 317 | 44443 | 0.26757 | 0.39452 | *CCSER1* |
| ENSG00000187094 | 3 | 42289317 | 42317699 | 67 | 17 | 41594 | 0.26747 | 0.39455 | *CCK* |
| ENSG00000115145 | 2 | 152963315 | 153042506 | 202 | 21 | 44141 | 0.26717 | 0.39467 | *STAM2* |
| ENSG00000115363 | 2 | 75686428 | 75806848 | 358 | 65 | 46596 | 0.26708 | 0.3947 | *EVA1A* |

| | | | | | | | | | |
|---|---|---|---|---|---|---|---|---|---|
| ENSG00000122729 | 9 | 32374618 | 32464767 | 261 | 47 | 52983 | 0.26706 | 0.39471 | *ACO1* |
| ENSG00000196260 | 6 | 30889130 | 30909952 | 110 | 15 | 42541 | 0.26692 | 0.39477 | *SFTA2* |
| ENSG00000258664 | 1 | 179823916 | 179844311 | 37 | 10 | 47948 | 0.26685 | 0.39479 | *IFRG15* |
| ENSG00000117322 | 1 | 207617575 | 207673240 | 163 | 28 | 47052 | 0.26679 | 0.39482 | *CR2* |
| ENSG00000066427 | 14 | 92514896 | 92582965 | 401 | 20 | 49415 | 0.26675 | 0.39483 | *ATXN3* |
| ENSG00000132740 | 11 | 68661310 | 68718070 | 190 | 19 | 49286 | 0.26621 | 0.39504 | *IGHMBP2* |
| ENSG00000082126 | 2 | 202499593 | 202573417 | 200 | 49 | 47558 | 0.26613 | 0.39507 | *MPP4* |
| ENSG00000175820 | 13 | 103371801 | 103399159 | 56 | 13 | 46565 | 0.26506 | 0.39548 | *CCDC168* |
| ENSG00000091640 | 17 | 4852521 | 4881167 | 59 | 17 | 43829 | 0.2648 | 0.39558 | *SPAG7* |
| ENSG00000081791 | 5 | 141293373 | 141331612 | 92 | 16 | 40825 | 0.26466 | 0.39564 | *KIAA0141* |
| ENSG00000125869 | 20 | 9485005 | 9521171 | 122 | 25 | 35563 | 0.2645 | 0.3957 | *LAMP5* |
| ENSG00000148843 | 10 | 105146405 | 105216049 | 111 | 21 | 47463 | 0.2643 | 0.39578 | *PDCD11* |
| ENSG00000140564 | 15 | 91401822 | 91436688 | 94 | 16 | 48368 | 0.26428 | 0.39578 | *FURIN* |
| ENSG00000179832 | 8 | 145192919 | 145326843 | 108 | 20 | 37973 | 0.26426 | 0.39579 | *MROH1* |
| ENSG00000140990 | 16 | 1999509 | 2021976 | 127 | 16 | 51457 | 0.26426 | 0.39579 | *NDUFB10* |
| ENSG00000103852 | 15 | 99666528 | 99801428 | 424 | 48 | 50805 | 0.26418 | 0.39582 | *TTC23* |
| ENSG00000154642 | 21 | 19151284 | 19201703 | 113 | 15 | 53098 | 0.26412 | 0.39584 | *C21orf91* |
| ENSG00000159592 | 1 | 46082976 | 46163785 | 205 | 9 | 55779 | 0.26407 | 0.39586 | *GPBP1L1* |
| ENSG00000248712 | 11 | 119050347 | 119077479 | 36 | 9 | 36846 | 0.26394 | 0.39591 | *CCDC153* |
| ENSG00000187714 | 10 | 50808347 | 50830765 | 60 | 21 | 37787 | 0.26392 | 0.39592 | *SLC18A3* |
| ENSG00000176945 | 3 | 195437753 | 195477994 | 26 | 14 | 41089 | 0.26384 | 0.39595 | *MUC20* |
| ENSG00000103202 | 16 | 436725 | 470367 | 136 | 19 | 33046 | 0.26377 | 0.39598 | *NME4* |
| ENSG00000215306 | 14 | 23276463 | 23301750 | 110 | 21 | 51176 | 0.26373 | 0.39599 | *AL135998.1* |
| ENSG00000164305 | 4 | 185538850 | 185580663 | 118 | 18 | 45516 | 0.26339 | 0.39612 | *CASP3* |
| ENSG00000131467 | 17 | 40966402 | 41005774 | 18 | 12 | 51620 | 0.26325 | 0.39618 | *PSME3* |
| ENSG00000175662 | 17 | 17736828 | 17885736 | 202 | 16 | 30040 | 0.2631 | 0.39624 | *TOM1L2* |
| ENSG00000102934 | 16 | 57280004 | 57328599 | 197 | 27 | 40571 | 0.26286 | 0.39633 | *PLLP* |
| ENSG00000140995 | 16 | 90004333 | 90044468 | 215 | 31 | 32202 | 0.26287 | 0.39633 | *DEF8* |
| ENSG00000125531 | 20 | 62174373 | 62198061 | 73 | 26 | 38209 | 0.26249 | 0.39647 | *C20orf195* |
| ENSG00000121210 | 4 | 154377498 | 154567863 | 532 | 86 | 42757 | 0.26244 | 0.39649 | *KIAA0922* |
| ENSG00000133706 | 5 | 145482601 | 145572223 | 388 | 10 | 55308 | 0.26243 | 0.39649 | *LARS* |
| ENSG00000135338 | 6 | 80184708 | 80257175 | 161 | 23 | 49665 | 0.2622 | 0.39658 | *LCA5* |
| ENSG00000129691 | 8 | 37952760 | 38011594 | 68 | 18 | 39217 | 0.26221 | 0.39658 | *ASH2L* |
| ENSG00000178567 | 3 | 37017357 | 37044795 | 47 | 8 | 38615 | 0.26209 | 0.39663 | *EPM2AIP1* |
| ENSG00000128040 | 4 | 57666026 | 57697908 | 137 | 25 | 32371 | 0.26198 | 0.39667 | *SPINK2* |
| ENSG00000170075 | 1 | 202081986 | 202112720 | 103 | 24 | 39124 | 0.26151 | 0.39685 | *GPR37L1* |

| | | | | | | | | | |
|---|---|---|---|---|---|---|---|---|---|
| ENSG00000164466 | 5 | 174894065 | 174966745 | 308 | 33 | 36190 | 0.26141 | 0.39689 | *SFXN1* |
| ENSG00000050767 | 5 | 177654619 | 178027556 | 1543 | 200 | 38253 | 0.26131 | 0.39693 | *COL23A1* |
| ENSG00000198851 | 11 | 118165260 | 118196890 | 104 | 13 | 55018 | 0.26108 | 0.39702 | *CD3E* |
| ENSG00000119698 | 14 | 94602465 | 94756072 | 402 | 32 | 36414 | 0.26094 | 0.39707 | *PPP4R4* |
| ENSG00000080854 | 11 | 133768459 | 133836880 | 199 | 41 | 46455 | 0.26092 | 0.39708 | *IGSF9B* |
| ENSG00000074771 | 6 | 155706504 | 155787037 | 240 | 61 | 46634 | 0.26071 | 0.39716 | *NOX3* |
| ENSG00000171446 | 17 | 38923060 | 38948786 | 91 | 21 | 47640 | 0.26054 | 0.39722 | *KRT27* |
| ENSG00000268975 | 19 | 41267553 | 41302590 | 130 | 21 | 44109 | 0.26055 | 0.39722 | *MIA-RAB4B* |
| ENSG00000177054 | 11 | 19128646 | 19207969 | 220 | 29 | 40212 | 0.26048 | 0.39725 | *ZDHHC13* |
| ENSG00000140682 | 16 | 31472906 | 31499281 | 33 | 11 | 46379 | 0.26031 | 0.39731 | *TGFB1I1* |
| ENSG00000175535 | 10 | 118295443 | 118337367 | 68 | 10 | 53959 | 0.26015 | 0.39737 | *PNLIP* |
| ENSG00000178015 | 5 | 94945782 | 94967846 | 41 | 17 | 42629 | 0.25953 | 0.39761 | *GPR150* |
| ENSG00000178852 | 17 | 45390656 | 45528678 | 480 | 27 | 37440 | 0.2593 | 0.3977 | *EFCAB13* |
| ENSG00000174886 | 19 | 5881287 | 5914017 | 160 | 24 | 53298 | 0.25932 | 0.3977 | *NDUFA11* |
| ENSG00000084207 | 11 | 67341066 | 67364131 | 94 | 13 | 48198 | 0.25928 | 0.39771 | *GSTP1* |
| ENSG00000114857 | 3 | 42632106 | 42700227 | 177 | 17 | 34349 | 0.25926 | 0.39772 | *NKTR* |
| ENSG00000173145 | 10 | 96065004 | 96132716 | 209 | 23 | 55224 | 0.2592 | 0.39774 | *NOC3L* |
| ENSG00000188393 | 12 | 10041272 | 10094980 | 194 | 44 | 41035 | 0.25916 | 0.39776 | *CLEC2A* |
| ENSG00000198561 | 11 | 57510715 | 57597018 | 117 | 18 | 47453 | 0.25913 | 0.39777 | *CTNND1* |
| ENSG00000160746 | 3 | 43386351 | 43743086 | 792 | 76 | 43846 | 0.25907 | 0.39779 | *ANO10* |
| ENSG00000136205 | 7 | 47304752 | 47632156 | 953 | 155 | 45895 | 0.25905 | 0.3978 | *TNS3* |
| ENSG00000255284 | 11 | 767578 | 794297 | 75 | 15 | 42287 | 0.25874 | 0.39792 | *AP006621.5* |
| ENSG00000186470 | 6 | 26355387 | 26388546 | 222 | 20 | 48217 | 0.25856 | 0.39799 | *BTN3A2* |
| ENSG00000149043 | 11 | 1838709 | 1868751 | 72 | 13 | 51990 | 0.25857 | 0.39799 | *SYT8* |
| ENSG00000269302 | 4 | 187101913 | 187122626 | 127 | 12 | 47022 | 0.25824 | 0.39811 | *AC110771.1* |
| ENSG00000239704 | 17 | 15329332 | 15416922 | 205 | 33 | 45692 | 0.25807 | 0.39818 | *CDRT4* |
| ENSG00000177873 | 3 | 40508604 | 40541727 | 76 | 11 | 46498 | 0.25799 | 0.39821 | *ZNF619* |
| ENSG00000157833 | 2 | 26385960 | 26422532 | 82 | 9 | 49877 | 0.25784 | 0.39827 | *GAREML* |
| ENSG00000164796 | 8 | 113225157 | 114459328 | 3434 | 131 | 44583 | 0.25771 | 0.39832 | *CSMD3* |
| ENSG00000168282 | 14 | 50077489 | 50100198 | 75 | 9 | 39557 | 0.25768 | 0.39833 | *MGAT2* |
| ENSG00000126233 | 8 | 143812362 | 143833829 | 62 | 10 | 55177 | 0.25743 | 0.39842 | *SLURP1* |
| ENSG00000185947 | 16 | 31875079 | 31938668 | 52 | 25 | 49635 | 0.25735 | 0.39846 | *ZNF267* |
| ENSG00000258064 | 12 | 72057984 | 72102748 | 73 | 14 | 43880 | 0.25727 | 0.39848 | *RP11-293I14.2* |
| ENSG00000116198 | 1 | 3718645 | 3783778 | 371 | 29 | 40435 | 0.25714 | 0.39853 | *CEP104* |
| ENSG00000186393 | 17 | 38912490 | 38938414 | 84 | 15 | 48876 | 0.25699 | 0.39859 | *KRT26* |
| ENSG00000268485 | 2 | 242157319 | 242178850 | 55 | 19 | 43486 | 0.25667 | 0.39872 | *AC104841.2* |

| | | | | | | | | | |
|---|---|---|---|---|---|---|---|---|---|
| ENSG00000197226 | 5 | 179279066 | 179344859 | 138 | 21 | 35742 | 0.25665 | 0.39872 | *TBC1D9B* |
| ENSG00000197459 | 6 | 26241879 | 26262303 | 53 | 12 | 44162 | 0.25663 | 0.39873 | *HIST1H2BH* |
| ENSG00000125657 | 19 | 6521010 | 6545931 | 76 | 23 | 42679 | 0.25655 | 0.39876 | *TNFSF9* |
| ENSG00000196428 | 3 | 150116122 | 150194218 | 241 | 28 | 50448 | 0.25651 | 0.39878 | *TSC22D2* |
| ENSG00000089280 | 16 | 31181431 | 31213127 | 25 | 10 | 42461 | 0.25638 | 0.39883 | *FUS* |
| ENSG00000110244 | 11 | 116681419 | 116704022 | 96 | 17 | 30738 | 0.25634 | 0.39884 | *APOA4* |
| ENSG00000170324 | 10 | 49354601 | 49492941 | 222 | 33 | 42021 | 0.25632 | 0.39885 | *FRMPD2* |
| ENSG00000168779 | 3 | 157804948 | 157834292 | 74 | 10 | 50023 | 0.25622 | 0.39889 | *SHOX2* |
| ENSG00000244623 | 7 | 99463610 | 99484680 | 52 | 10 | 38692 | 0.25608 | 0.39895 | *OR2AE1* |
| ENSG00000120686 | 13 | 38913986 | 38947140 | 37 | 10 | 41623 | 0.25601 | 0.39897 | *UFM1* |
| ENSG00000072864 | 16 | 15727124 | 15830210 | 284 | 42 | 42007 | 0.25592 | 0.39901 | *NDE1* |
| ENSG00000154429 | 1 | 229446758 | 229489041 | 110 | 15 | 54133 | 0.25585 | 0.39903 | *CCSAP* |
| ENSG00000109332 | 4 | 103705540 | 103800053 | 246 | 12 | 43115 | 0.25553 | 0.39916 | *UBE2D3* |
| ENSG00000138592 | 15 | 50706577 | 50803280 | 346 | 31 | 50781 | 0.25509 | 0.39933 | *USP8* |
| ENSG00000145416 | 4 | 164435450 | 165315202 | 3480 | 235 | 37548 | 0.25491 | 0.3994 | *Mar-01* |
| ENSG00000139428 | 12 | 109981542 | 110021679 | 170 | 20 | 51018 | 0.25482 | 0.39943 | *MMAB* |
| ENSG00000128203 | 22 | 26815239 | 26850981 | 108 | 23 | 45286 | 0.25456 | 0.39953 | *ASPHD2* |
| ENSG00000165507 | 10 | 45456429 | 45484258 | 96 | 23 | 47038 | 0.25449 | 0.39956 | *C10orf10* |
| ENSG00000165501 | 14 | 50055415 | 50091390 | 121 | 11 | 46669 | 0.25427 | 0.39964 | *LRR1* |
| ENSG00000211452 | 1 | 54346912 | 54386759 | 161 | 20 | 48995 | 0.25424 | 0.39966 | *DIO1* |
| ENSG00000171484 | 9 | 125380858 | 125401852 | 101 | 18 | 45044 | 0.25391 | 0.39978 | *OR1B1* |
| ENSG00000189334 | 1 | 153576731 | 153599462 | 43 | 9 | 41881 | 0.25389 | 0.39979 | *S100A14* |
| ENSG00000119979 | 10 | 120853598 | 120907496 | 173 | 14 | 45972 | 0.2538 | 0.39982 | *FAM45A* |
| ENSG00000147677 | 8 | 117644369 | 117789164 | 352 | 35 | 38605 | 0.25374 | 0.39985 | *EIF3H* |
| ENSG00000175193 | 3 | 183537173 | 183612721 | 253 | 28 | 51779 | 0.25363 | 0.39989 | *PARL* |
| ENSG00000204390 | 6 | 31767396 | 31793437 | 72 | 25 | 38264 | 0.25343 | 0.39997 | *HSPA1L* |
| ENSG00000105245 | 19 | 41162596 | 41206877 | 141 | 26 | 49386 | 0.25335 | 0.4 | *NUMBL* |
| ENSG00000088812 | 20 | 3441687 | 3641769 | 620 | 44 | 50732 | 0.25334 | 0.4 | *ATRN* |
| ENSG00000179630 | 13 | 44443420 | 44478068 | 161 | 20 | 44968 | 0.25325 | 0.40004 | *LACC1* |
| ENSG00000128039 | 4 | 56202276 | 56249263 | 146 | 23 | 49158 | 0.25311 | 0.40009 | *SRD5A3* |
| ENSG00000086570 | 5 | 150873654 | 150958505 | 287 | 40 | 49562 | 0.25305 | 0.40012 | *FAT2* |
| ENSG00000120341 | 1 | 177883091 | 177963438 | 325 | 52 | 45546 | 0.25295 | 0.40015 | *SEC16B* |
| ENSG00000188517 | 4 | 109721877 | 110233813 | 1779 | 123 | 33075 | 0.25279 | 0.40022 | *COL25A1* |
| ENSG00000126368 | 17 | 38239040 | 38266978 | 58 | 18 | 40796 | 0.25238 | 0.40037 | *NR1D1* |
| ENSG00000130528 | 19 | 49644455 | 49668681 | 99 | 20 | 46517 | 0.25222 | 0.40043 | *HRC* |
| ENSG00000186190 | 20 | 31633230 | 31671434 | 126 | 18 | 50591 | 0.25204 | 0.40051 | *BPIFB3* |

| | | | | | | | | | |
|---|---|---|---|---|---|---|---|---|---|
| ENSG00000091947 | 17 | 42078556 | 42111314 | 85 | 15 | 45430 | 0.25198 | 0.40053 | *TMEM101* |
| ENSG00000103091 | 16 | 74897468 | 75044071 | 505 | 93 | 44908 | 0.25191 | 0.40055 | *WDR59* |
| ENSG00000176022 | 1 | 1157629 | 1180421 | 132 | 9 | 47547 | 0.25151 | 0.40071 | *B3GALT6* |
| ENSG00000161653 | 17 | 42071914 | 42096431 | 66 | 15 | 44117 | 0.25107 | 0.40088 | *NAGS* |
| ENSG00000198841 | 1 | 52487775 | 52509488 | 51 | 11 | 50562 | 0.25091 | 0.40094 | *KTI12* |
| ENSG00000181323 | 17 | 7313679 | 7334951 | 43 | 19 | 40412 | 0.25055 | 0.40108 | *SPEM1* |
| ENSG00000166482 | 17 | 19276755 | 19300553 | 15 | 7 | 48280 | 0.25028 | 0.40119 | *MFAP4* |
| ENSG00000154025 | 17 | 18843658 | 18935908 | 167 | 53 | 39129 | 0.25008 | 0.40126 | *SLC5A10* |
| ENSG00000176490 | 19 | 2704565 | 2731416 | 124 | 20 | 40844 | 0.24994 | 0.40132 | *DIRAS1* |
| ENSG00000119414 | 9 | 127898852 | 127962218 | 173 | 15 | 53972 | 0.24987 | 0.40135 | *PPP6C* |
| ENSG00000138297 | 10 | 51582080 | 51633365 | 23 | 9 | 46062 | 0.24969 | 0.40141 | *TIMM23* |
| ENSG00000187676 | 13 | 31764073 | 31916413 | 431 | 52 | 41506 | 0.24961 | 0.40144 | *B3GALTL* |
| ENSG00000111981 | 6 | 150275143 | 150304846 | 67 | 13 | 29666 | 0.24954 | 0.40147 | *ULBP1* |
| ENSG00000203760 | 6 | 126651320 | 126680021 | 37 | 8 | 33366 | 0.24952 | 0.40148 | *CENPW* |
| ENSG00000166896 | 12 | 58325324 | 58361052 | 113 | 12 | 29817 | 0.24946 | 0.4015 | *XRCC6BP1* |
| ENSG00000151418 | 1 | 198482352 | 198520075 | 95 | 17 | 36496 | 0.24933 | 0.40155 | *ATP6V1G3* |
| ENSG00000231192 | 3 | 97841542 | 97862483 | 75 | 7 | 50102 | 0.2492 | 0.4016 | *OR5H1* |
| ENSG00000119231 | 3 | 196584727 | 196671585 | 254 | 24 | 34540 | 0.24902 | 0.40167 | *SENP5* |
| ENSG00000138071 | 2 | 65444887 | 65508387 | 196 | 20 | 42370 | 0.24875 | 0.40178 | *ACTR2* |
| ENSG00000173876 | 10 | 82828 | 130103 | 124 | 21 | 40294 | 0.2485 | 0.40187 | *TUBB8* |
| ENSG00000132467 | 4 | 71544196 | 71566267 | 33 | 11 | 41221 | 0.24838 | 0.40192 | *UTP3* |
| ENSG00000154473 | 10 | 124903793 | 124934886 | 63 | 10 | 46219 | 0.24834 | 0.40194 | *BUB3* |
| ENSG00000203910 | 1 | 92673497 | 92721370 | 85 | 13 | 27498 | 0.24795 | 0.40209 | *C1orf146* |
| ENSG00000068903 | 19 | 39359197 | 39400502 | 90 | 24 | 37495 | 0.24784 | 0.40213 | *SIRT2* |
| ENSG00000091106 | 2 | 32439522 | 32500923 | 155 | 20 | 53364 | 0.24759 | 0.40222 | *NLRC4* |
| ENSG00000145284 | 4 | 83540692 | 83730010 | 993 | 99 | 42618 | 0.24721 | 0.40237 | *SCD5* |
| ENSG00000166111 | 12 | 109294658 | 109469045 | 283 | 62 | 45204 | 0.24723 | 0.40237 | *SVOP* |
| ENSG00000183386 | 1 | 38452442 | 38481278 | 60 | 26 | 41403 | 0.2472 | 0.40238 | *FHL3* |
| ENSG00000119203 | 2 | 9553697 | 9623230 | 158 | 15 | 34474 | 0.24701 | 0.40245 | *CPSF3* |
| ENSG00000155926 | 8 | 134038973 | 134125298 | 384 | 36 | 48483 | 0.24683 | 0.40252 | *SLA* |
| ENSG00000099968 | 22 | 18101621 | 18223388 | 513 | 42 | 40187 | 0.24663 | 0.4026 | *BCL2L13* |
| ENSG00000198768 | 20 | 57024157 | 57100187 | 221 | 55 | 45731 | 0.24655 | 0.40263 | *APCDD1L* |
| ENSG00000112306 | 6 | 133125580 | 133148703 | 125 | 18 | 44376 | 0.24649 | 0.40265 | *RPS12* |
| ENSG00000154654 | 21 | 22360633 | 22925650 | 2012 | 175 | 47440 | 0.2462 | 0.40277 | *NCAM2* |
| ENSG00000097021 | 1 | 6314329 | 6464451 | 582 | 52 | 35056 | 0.24599 | 0.40284 | *ACOT7* |
| ENSG00000204683 | 10 | 21404692 | 21445488 | 94 | 16 | 40283 | 0.24601 | 0.40284 | *C10orf113* |

| | | | | | | | | | |
|---|---|---|---|---|---|---|---|---|---|
| ENSG00000004660 | 17 | 3753609 | 3808185 | 144 | 48 | 38408 | 0.24577 | 0.40293 | *CAMKK1* |
| ENSG00000185860 | 1 | 162784248 | 162848605 | 274 | 41 | 40835 | 0.24576 | 0.40294 | *C1orf110* |
| ENSG00000259003 | 14 | 23015534 | 23037939 | 145 | 22 | 36580 | 0.24559 | 0.403 | *AE000662.92* |
| ENSG00000066322 | 1 | 43819068 | 43843696 | 46 | 12 | 45358 | 0.24554 | 0.40302 | *ELOVL1* |
| ENSG00000048740 | 10 | 11037259 | 11388674 | 1274 | 146 | 41347 | 0.24507 | 0.4032 | *CELF2* |
| ENSG00000128609 | 7 | 123167051 | 123208309 | 109 | 16 | 54620 | 0.24504 | 0.40321 | *NDUFA5* |
| ENSG00000110108 | 11 | 60671346 | 60700915 | 71 | 11 | 39133 | 0.24505 | 0.40321 | *TMEM109* |
| ENSG00000170128 | 1 | 200832083 | 200853306 | 93 | 18 | 38279 | 0.24498 | 0.40324 | *GPR25* |
| ENSG00000126821 | 14 | 64140932 | 64204757 | 101 | 12 | 35451 | 0.24498 | 0.40324 | *SGPP1* |
| ENSG00000176087 | 5 | 139934041 | 139958688 | 36 | 11 | 48750 | 0.2449 | 0.40327 | *SLC35A4* |
| ENSG00000181965 | 5 | 134859991 | 134881639 | 54 | 17 | 42247 | 0.24487 | 0.40328 | *NEUROG1* |
| ENSG00000065183 | 1 | 118462343 | 118519165 | 119 | 14 | 31539 | 0.24464 | 0.40337 | *WDR3* |
| ENSG00000137547 | 8 | 55037770 | 55070461 | 31 | 12 | 41158 | 0.24421 | 0.40353 | *MRPL15* |
| ENSG00000204694 | 6 | 29383281 | 29434848 | 212 | 20 | 27079 | 0.24406 | 0.40359 | *OR11A1* |
| ENSG00000107738 | 10 | 73497316 | 73543255 | 191 | 40 | 50576 | 0.2437 | 0.40373 | *C10orf54* |
| ENSG00000213215 | 7 | 143647027 | 143668108 | 78 | 11 | 42633 | 0.24363 | 0.40376 | *OR2F1* |
| ENSG00000100191 | 22 | 32604465 | 32661328 | 247 | 36 | 45289 | 0.24353 | 0.4038 | *SLC5A4* |
| ENSG00000176984 | 11 | 120029685 | 120052387 | 58 | 15 | 39090 | 0.24339 | 0.40385 | *AP000679.2* |
| ENSG00000113205 | 5 | 140470234 | 140493406 | 70 | 7 | 52987 | 0.24335 | 0.40387 | *PCDHB3* |
| ENSG00000153930 | 17 | 54178319 | 54599020 | 1032 | 95 | 44240 | 0.2433 | 0.40389 | *ANKFN1* |
| ENSG00000124688 | 6 | 43587277 | 43618689 | 68 | 15 | 41899 | 0.24311 | 0.40396 | *MAD2L1BP* |
| ENSG00000140396 | 8 | 71011997 | 71326040 | 532 | 38 | 38718 | 0.24307 | 0.40397 | *NCOA2* |
| ENSG00000082515 | 5 | 154310630 | 154358971 | 124 | 17 | 44705 | 0.24265 | 0.40414 | *MRPL22* |
| ENSG00000151332 | 14 | 36757770 | 36799882 | 56 | 15 | 47749 | 0.2426 | 0.40416 | *MBIP* |
| ENSG00000255767 | 2 | 3569574 | 3605547 | 102 | 19 | 50961 | 0.24255 | 0.40418 | *RP13-512J5.1* |
| ENSG00000179409 | 17 | 637654 | 667239 | 112 | 22 | 47265 | 0.24238 | 0.40424 | *GEMIN4* |
| ENSG00000196458 | 12 | 133488019 | 133542892 | 75 | 18 | 42868 | 0.24237 | 0.40425 | *ZNF605* |
| ENSG00000221926 | 17 | 15521274 | 15597625 | 200 | 35 | 42578 | 0.24236 | 0.40425 | *TRIM16* |
| ENSG00000103502 | 16 | 29859678 | 29885057 | 26 | 6 | 48421 | 0.2423 | 0.40427 | *CDIPT* |
| ENSG00000168291 | 3 | 58403357 | 58429584 | 123 | 15 | 36855 | 0.24221 | 0.40431 | *PDHB* |
| ENSG00000171956 | 15 | 60286421 | 60363929 | 253 | 38 | 37113 | 0.24218 | 0.40432 | *FOXB1* |
| ENSG00000178700 | 3 | 93756680 | 93792233 | 31 | 12 | 42159 | 0.24214 | 0.40433 | *DHFRL1* |
| ENSG00000183695 | 11 | 19066002 | 19092228 | 137 | 18 | 48289 | 0.24212 | 0.40434 | *MRGPRX2* |
| ENSG00000213965 | 19 | 33172867 | 33214702 | 119 | 18 | 46303 | 0.24207 | 0.40436 | *NUDT19* |
| ENSG00000168772 | 4 | 105379469 | 105426058 | 61 | 14 | 39373 | 0.24205 | 0.40437 | *CXXC4* |
| ENSG00000144550 | 3 | 9735491 | 9781592 | 99 | 33 | 48376 | 0.24172 | 0.4045 | *CPNE9* |

| | | | | | | | | | |
|---|---|---|---|---|---|---|---|---|---|
| ENSG00000205060 | 7 | 133964084 | 134011803 | 136 | 7 | 46524 | 0.24173 | 0.4045 | *SLC35B4* |
| ENSG00000177984 | 9 | 139644086 | 139670707 | 128 | 10 | 39001 | 0.24169 | 0.40451 | *LCN15* |
| ENSG00000137409 | 6 | 36925917 | 36964074 | 158 | 25 | 44042 | 0.24152 | 0.40458 | *MTCH1* |
| ENSG00000159307 | 22 | 43583289 | 43749394 | 757 | 123 | 42832 | 0.24095 | 0.4048 | *SCUBE1* |
| ENSG00000178636 | 4 | 120103928 | 120143799 | 137 | 20 | 55728 | 0.24092 | 0.40481 | *RP11-455G16.1* |
| ENSG00000139304 | 12 | 80789774 | 81082802 | 839 | 77 | 33487 | 0.24073 | 0.40488 | *PTPRQ* |
| ENSG00000167617 | 19 | 54966210 | 54994411 | 124 | 30 | 44153 | 0.24074 | 0.40488 | *CDC42EP5* |
| ENSG00000189184 | 4 | 138430072 | 138463648 | 109 | 25 | 46720 | 0.24071 | 0.40489 | *PCDH18* |
| ENSG00000198793 | 1 | 11156592 | 11332564 | 385 | 19 | 24857 | 0.24066 | 0.40491 | *MTOR* |
| ENSG00000168887 | 2 | 85823777 | 85849189 | 43 | 10 | 34675 | 0.24062 | 0.40492 | *C2orf68* |
| ENSG00000041353 | 18 | 52375091 | 52572747 | 673 | 56 | 46424 | 0.24054 | 0.40495 | *RAB27B* |
| ENSG00000151553 | 10 | 116571503 | 116669591 | 315 | 24 | 45106 | 0.24032 | 0.40504 | *FAM160B1* |
| ENSG00000196505 | 1 | 118396107 | 118482253 | 112 | 20 | 35897 | 0.2403 | 0.40505 | *GDAP2* |
| ENSG00000159140 | 21 | 34904924 | 34959812 | 64 | 13 | 51942 | 0.24003 | 0.40515 | *SON* |
| ENSG00000170476 | 5 | 138713169 | 138735770 | 27 | 4 | 25614 | 0.23967 | 0.40529 | *MZB1* |
| ENSG00000143126 | 1 | 109782641 | 109828372 | 140 | 31 | 45815 | 0.23962 | 0.40531 | *CELSR2* |
| ENSG00000165553 | 14 | 77721826 | 77747655 | 88 | 19 | 41217 | 0.23958 | 0.40533 | *NGB* |
| ENSG00000183020 | 11 | 914894 | 1022239 | 366 | 18 | 31267 | 0.23944 | 0.40538 | *AP2A2* |
| ENSG00000100320 | 22 | 36124783 | 36434473 | 450 | 48 | 45019 | 0.23943 | 0.40539 | *RBFOX2* |
| ENSG00000155465 | 14 | 23232431 | 23309029 | 328 | 56 | 44260 | 0.23939 | 0.4054 | *SLC7A7* |
| ENSG00000268809 | 2 | 105872490 | 105892585 | 47 | 13 | 34072 | 0.23923 | 0.40546 | *AC012360.2* |
| ENSG00000171723 | 14 | 66964125 | 67658520 | 2338 | 42 | 43841 | 0.23899 | 0.40556 | *GPHN* |
| ENSG00000183598 | 1 | 149774826 | 149795236 | 36 | 5 | 50375 | 0.23894 | 0.40558 | *HIST2H3D* |
| ENSG00000170579 | 18 | 3486030 | 4465335 | 3394 | 351 | 44515 | 0.23865 | 0.40569 | *DLGAP1* |
| ENSG00000177800 | 1 | 229375383 | 229397557 | 65 | 12 | 54952 | 0.23851 | 0.40574 | *TMEM78* |
| ENSG00000175262 | 1 | 10996528 | 11052094 | 213 | 30 | 26343 | 0.2381 | 0.4059 | *C1orf127* |
| ENSG00000140987 | 16 | 3422085 | 3461065 | 102 | 16 | 50748 | 0.23808 | 0.40591 | *ZSCAN32* |
| ENSG00000123989 | 2 | 220393669 | 220418509 | 63 | 11 | 45757 | 0.23788 | 0.40599 | *CHPF* |
| ENSG00000180913 | 11 | 6139840 | 6160706 | 73 | 12 | 31752 | 0.23779 | 0.40602 | *OR56B3P* |
| ENSG00000126254 | 19 | 36109932 | 36138588 | 67 | 15 | 40807 | 0.23778 | 0.40603 | *RBM42* |
| ENSG00000178522 | 4 | 71447973 | 71483005 | 133 | 30 | 37588 | 0.23737 | 0.40619 | *AMBN* |
| ENSG00000145916 | 5 | 177547997 | 177587566 | 130 | 17 | 53574 | 0.23683 | 0.40639 | *RMND5B* |
| ENSG00000204671 | 12 | 122646577 | 122668746 | 72 | 10 | 49842 | 0.23672 | 0.40644 | *IL31* |
| ENSG00000092439 | 15 | 50834670 | 50989012 | 574 | 37 | 49716 | 0.23646 | 0.40654 | *TRPM7* |
| ENSG00000102678 | 13 | 22235522 | 22288637 | 126 | 26 | 49340 | 0.23632 | 0.40659 | *FGF9* |
| ENSG00000184564 | 13 | 86356925 | 86383623 | 148 | 13 | 44954 | 0.23625 | 0.40662 | *SLITRK6* |

| | | | | | | | | | |
|---|---|---|---|---|---|---|---|---|---|
| ENSG00000175077 | 3 | 186905274 | 186929253 | 74 | 16 | 42966 | 0.23616 | 0.40665 | *RTP1* |
| ENSG00000250210 | 8 | 132091 | 160563 | 20 | 7 | 36468 | 0.23614 | 0.40666 | *RP11-585F1.10* |
| ENSG00000120051 | 10 | 106103522 | 106224848 | 253 | 47 | 45907 | 0.23592 | 0.40675 | *CCDC147* |
| ENSG00000188133 | 9 | 32773497 | 32797397 | 79 | 12 | 42885 | 0.23588 | 0.40676 | *TMEM215* |
| ENSG00000148950 | 11 | 31443948 | 31541192 | 133 | 12 | 33993 | 0.2356 | 0.40687 | *IMMP1L* |
| ENSG00000134115 | 3 | 1124260 | 1455901 | 1731 | 186 | 42747 | 0.2353 | 0.40699 | *CNTN6* |
| ENSG00000122584 | 7 | 8463585 | 8802593 | 1607 | 119 | 45239 | 0.23508 | 0.40707 | *NXPH1* |
| ENSG00000159479 | 1 | 43839588 | 43865479 | 53 | 12 | 41310 | 0.23492 | 0.40713 | *MED8* |
| ENSG00000187678 | 5 | 141679992 | 141716020 | 104 | 19 | 46168 | 0.23491 | 0.40714 | *SPRY4* |
| ENSG00000172361 | 18 | 47743563 | 47802892 | 142 | 16 | 49705 | 0.23476 | 0.4072 | *CCDC11* |
| ENSG00000198056 | 12 | 57115380 | 57156157 | 73 | 12 | 44835 | 0.23469 | 0.40722 | *PRIM1* |
| ENSG00000204371 | 6 | 31837536 | 31875464 | 136 | 32 | 33187 | 0.23454 | 0.40728 | *EHMT2* |
| ENSG00000031003 | 5 | 137263649 | 137397650 | 330 | 24 | 52488 | 0.23438 | 0.40735 | *FAM13B* |
| ENSG00000187003 | 9 | 111614603 | 111636035 | 109 | 21 | 49309 | 0.23424 | 0.4074 | *ACTL7A* |
| ENSG00000074964 | 1 | 17856330 | 18034369 | 485 | 72 | 51117 | 0.23396 | 0.40751 | *ARHGEF10L* |
| ENSG00000119906 | 10 | 102662326 | 102734893 | 123 | 18 | 36033 | 0.23394 | 0.40752 | *FAM178A* |
| ENSG00000143772 | 1 | 226809391 | 226937024 | 267 | 48 | 47561 | 0.23389 | 0.40753 | *ITPKB* |
| ENSG00000168348 | 14 | 35993248 | 36016260 | 23 | 7 | 37148 | 0.23367 | 0.40762 | *INSM2* |
| ENSG00000159352 | 1 | 151217179 | 151249955 | 47 | 8 | 56010 | 0.23363 | 0.40763 | *PSMD4* |
| ENSG00000186943 | 9 | 107321449 | 107342411 | 57 | 9 | 31504 | 0.23365 | 0.40763 | *OR13C8* |
| ENSG00000063245 | 19 | 56176592 | 56231224 | 219 | 38 | 45477 | 0.23345 | 0.4077 | *EPN1* |
| ENSG00000005882 | 17 | 48162101 | 48199516 | 110 | 21 | 38670 | 0.23335 | 0.40775 | *PDK2* |
| ENSG00000185115 | 15 | 29550353 | 29572033 | 59 | 12 | 44029 | 0.23321 | 0.4078 | *NDNL2* |
| ENSG00000153012 | 4 | 24990469 | 25042501 | 236 | 45 | 37697 | 0.23278 | 0.40796 | *LGI2* |
| ENSG00000189283 | 3 | 59725036 | 61247133 | 6975 | 448 | 43684 | 0.23262 | 0.40803 | *FHIT* |
| ENSG00000119041 | 2 | 197617756 | 197674449 | 94 | 19 | 40932 | 0.2326 | 0.40804 | *GTF3C3* |
| ENSG00000123454 | 9 | 136491482 | 136534466 | 212 | 52 | 39177 | 0.23242 | 0.40811 | *DBH* |
| ENSG00000106976 | 9 | 130955658 | 131027527 | 176 | 31 | 38472 | 0.23228 | 0.40816 | *DNM1* |
| ENSG00000171853 | 2 | 3373446 | 3498865 | 527 | 65 | 44443 | 0.23226 | 0.40817 | *TRAPPC12* |
| ENSG00000204130 | 10 | 70090864 | 70177051 | 217 | 17 | 39853 | 0.23218 | 0.4082 | *RUFY2* |
| ENSG00000158195 | 1 | 27720730 | 27826669 | 169 | 21 | 37932 | 0.23216 | 0.40821 | *WASF2* |
| ENSG00000188770 | 1 | 203453271 | 203487992 | 142 | 24 | 48110 | 0.23192 | 0.4083 | *OPTC* |
| ENSG00000163751 | 3 | 148573043 | 148624983 | 233 | 17 | 36550 | 0.23172 | 0.40838 | *CPA3* |
| ENSG00000141510 | 17 | 7555097 | 7600856 | 138 | 31 | 34130 | 0.23165 | 0.40841 | *TP53* |
| ENSG00000183559 | 10 | 124447225 | 124469338 | 76 | 18 | 48122 | 0.2315 | 0.40846 | *C10orf120* |
| ENSG00000078140 | 4 | 39689664 | 39794412 | 350 | 28 | 44336 | 0.23146 | 0.40848 | *UBE2K* |

| | | | | | | | | | |
|---|---|---|---|---|---|---|---|---|---|
| ENSG00000183527 | 21 | 40536695 | 40565777 | 117 | 12 | 50585 | 0.23146 | 0.40848 | *PSMG1* |
| ENSG00000033170 | 14 | 65867310 | 66220839 | 973 | 75 | 42075 | 0.23135 | 0.40852 | *FUT8* |
| ENSG00000214140 | 17 | 74513668 | 74559660 | 138 | 25 | 39879 | 0.23122 | 0.40857 | *PRCD* |
| ENSG00000170613 | 5 | 156578857 | 156603275 | 76 | 13 | 50466 | 0.23115 | 0.4086 | *FAM71B* |
| ENSG00000005421 | 7 | 94916988 | 95035673 | 279 | 42 | 44706 | 0.2308 | 0.40873 | *PON1* |
| ENSG00000068831 | 11 | 64484383 | 64522928 | 51 | 8 | 34955 | 0.23081 | 0.40873 | *RASGRP2* |
| ENSG00000163754 | 3 | 148699128 | 148755419 | 129 | 21 | 50420 | 0.23073 | 0.40876 | *GYG1* |
| ENSG00000181104 | 5 | 76001868 | 76041606 | 107 | 19 | 45400 | 0.23063 | 0.4088 | *F2R* |
| ENSG00000196266 | 1 | 159273460 | 159294449 | 51 | 12 | 40648 | 0.23048 | 0.40886 | *OR10J3* |
| ENSG00000121851 | 1 | 145446236 | 145480388 | 10 | 7 | 44305 | 0.23032 | 0.40892 | *POLR3GL* |
| ENSG00000145414 | 4 | 164021225 | 164098073 | 181 | 24 | 51900 | 0.23011 | 0.409 | *NAF1* |
| ENSG00000088876 | 20 | 2452463 | 2515348 | 158 | 40 | 42407 | 0.23012 | 0.409 | *ZNF343* |
| ENSG00000153310 | 8 | 130841839 | 131039375 | 448 | 63 | 49004 | 0.23003 | 0.40903 | *FAM49B* |
| ENSG00000162927 | 2 | 61157357 | 61255394 | 186 | 35 | 42871 | 0.22979 | 0.40913 | *PUS10* |
| ENSG00000062194 | 5 | 56459775 | 56570506 | 243 | 36 | 46227 | 0.22979 | 0.40913 | *GPBP1* |
| ENSG00000167653 | 8 | 143741726 | 143774142 | 145 | 16 | 41119 | 0.22965 | 0.40918 | *PSCA* |
| ENSG00000196208 | 2 | 11664242 | 11792914 | 405 | 68 | 38997 | 0.22963 | 0.40919 | *GREB1* |
| ENSG00000133275 | 19 | 1931188 | 1991337 | 258 | 33 | 37679 | 0.22964 | 0.40919 | *CSNK1G2* |
| ENSG00000179639 | 1 | 159249504 | 159288014 | 102 | 20 | 42429 | 0.22936 | 0.40929 | *FCER1A* |
| ENSG00000119446 | 9 | 124989903 | 125037118 | 113 | 21 | 42432 | 0.22914 | 0.40938 | *RBM18* |
| ENSG00000007001 | 2 | 158723214 | 159002666 | 856 | 105 | 35362 | 0.22887 | 0.40949 | *UPP2* |
| ENSG00000172432 | 6 | 43563053 | 43606899 | 88 | 19 | 42316 | 0.22881 | 0.40951 | *GTPBP2* |
| ENSG00000186625 | 6 | 149906009 | 149980108 | 159 | 8 | 28577 | 0.22879 | 0.40952 | *KATNA1* |
| ENSG00000110851 | 12 | 108116643 | 108165049 | 154 | 13 | 29981 | 0.22866 | 0.40957 | *PRDM4* |
| ENSG00000063660 | 2 | 241365088 | 241417493 | 234 | 48 | 44714 | 0.22858 | 0.4096 | *GPC1* |
| ENSG00000227164 | 20 | 52095495 | 52116164 | 106 | 9 | 53017 | 0.22833 | 0.40969 | *AL354993.1* |
| ENSG00000070413 | 22 | 19013795 | 19119967 | 497 | 31 | 44438 | 0.22833 | 0.40969 | *DGCR2* |
| ENSG00000162456 | 1 | 47001316 | 47027199 | 68 | 15 | 42802 | 0.2282 | 0.40974 | *KNCN* |
| ENSG00000036530 | 14 | 100140641 | 100203638 | 178 | 30 | 49667 | 0.2282 | 0.40975 | *CYP46A1* |
| ENSG00000107443 | 10 | 97793151 | 97830627 | 78 | 18 | 45252 | 0.22798 | 0.40983 | *CCNJ* |
| ENSG00000138792 | 4 | 111276889 | 111496441 | 700 | 83 | 50549 | 0.22782 | 0.40989 | *ENPEP* |
| ENSG00000101327 | 20 | 1949403 | 1984732 | 62 | 16 | 33896 | 0.22762 | 0.40997 | *PDYN* |
| ENSG00000179933 | 14 | 23553974 | 23579665 | 98 | 18 | 48127 | 0.22734 | 0.41008 | *C14orf119* |
| ENSG00000165886 | 10 | 99248625 | 99340966 | 228 | 27 | 36186 | 0.22726 | 0.41011 | *UBTD1* |
| ENSG00000103942 | 15 | 83499838 | 83664661 | 424 | 51 | 48015 | 0.22688 | 0.41026 | *HOMER2* |
| ENSG00000170236 | 15 | 50782759 | 50848905 | 265 | 28 | 49200 | 0.22642 | 0.41044 | *USP50* |

| | | | | | | | | | |
|---|---|---|---|---|---|---|---|---|---|
| ENSG00000159388 | 1 | 203264619 | 203288730 | 56 | 18 | 45519 | 0.22628 | 0.41049 | *BTG2* |
| ENSG00000132603 | 16 | 69363333 | 69387014 | 57 | 13 | 54192 | 0.22608 | 0.41057 | *NIP7* |
| ENSG00000124089 | 20 | 54813788 | 54834871 | 94 | 19 | 38117 | 0.22607 | 0.41057 | *MC3R* |
| ENSG00000121381 | 12 | 10951693 | 10972767 | 36 | 8 | 38606 | 0.22596 | 0.41062 | *TAS2R9* |
| ENSG00000271949 | 1 | 90088699 | 90408809 | 646 | 65 | 52056 | 0.2259 | 0.41064 | *RP11-302M6.4* |
| ENSG00000128891 | 15 | 40810882 | 40867256 | 169 | 15 | 32642 | 0.22577 | 0.41069 | *C15orf57* |
| ENSG00000197723 | 17 | 40264756 | 40285371 | 38 | 4 | 25931 | 0.22573 | 0.41071 | *HSPB9* |
| ENSG00000268882 | 10 | 135182720 | 135203102 | 84 | 12 | 33217 | 0.22539 | 0.41084 | *AL360181.1* |
| ENSG00000185933 | 10 | 105203144 | 105228645 | 65 | 17 | 51682 | 0.22537 | 0.41085 | *CALHM1* |
| ENSG00000258484 | 15 | 69100560 | 69249150 | 257 | 40 | 36552 | 0.22532 | 0.41086 | *SPESP1* |
| ENSG00000134343 | 11 | 26200829 | 26694835 | 2959 | 160 | 48818 | 0.22525 | 0.41089 | *ANO3* |
| ENSG00000169894 | 7 | 100537187 | 100621118 | 124 | 29 | 43096 | 0.22518 | 0.41092 | *MUC3A* |
| ENSG00000179520 | 12 | 100740857 | 100825837 | 227 | 42 | 47736 | 0.22515 | 0.41093 | *SLC17A8* |
| ENSG00000196361 | 19 | 11552141 | 11601861 | 152 | 31 | 35311 | 0.22516 | 0.41093 | *ELAVL3* |
| ENSG00000159905 | 19 | 44445375 | 44481861 | 129 | 17 | 44088 | 0.22482 | 0.41106 | *ZNF221* |
| ENSG00000166169 | 10 | 103328639 | 103358027 | 54 | 7 | 45399 | 0.22479 | 0.41107 | *POLL* |
| ENSG00000204653 | 19 | 51004857 | 51027947 | 78 | 18 | 40801 | 0.22468 | 0.41111 | *ASPDH* |
| ENSG00000170540 | 16 | 18792991 | 18823000 | 60 | 11 | 47284 | 0.22441 | 0.41122 | *ARL6IP1* |
| ENSG00000164597 | 7 | 106832000 | 107214959 | 908 | 45 | 51452 | 0.22435 | 0.41124 | *COG5* |
| ENSG00000132518 | 17 | 7895912 | 7933657 | 107 | 24 | 47380 | 0.22431 | 0.41126 | *GUCY2D* |
| ENSG00000160218 | 21 | 45422200 | 45536433 | 230 | 18 | 29896 | 0.22423 | 0.41129 | *TRAPPC10* |
| ENSG00000181039 | 1 | 145460508 | 145485646 | 11 | 7 | 44229 | 0.22415 | 0.41132 | *ANKRD34A* |
| ENSG00000165097 | 6 | 18145560 | 18234084 | 338 | 44 | 50645 | 0.22383 | 0.41145 | *KDM1B* |
| ENSG00000100387 | 22 | 41337351 | 41379313 | 46 | 15 | 44112 | 0.22379 | 0.41146 | *RBX1* |
| ENSG00000143869 | 2 | 20856424 | 20883418 | 87 | 26 | 47411 | 0.22377 | 0.41147 | *GDF7* |
| ENSG00000138660 | 4 | 113142893 | 113201203 | 120 | 14 | 36322 | 0.22359 | 0.41154 | *AP1AR* |
| ENSG00000174917 | 19 | 5668432 | 5690907 | 53 | 14 | 49270 | 0.22326 | 0.41167 | *C19orf70* |
| ENSG00000164303 | 4 | 184999859 | 185152383 | 557 | 74 | 44193 | 0.22307 | 0.41174 | *ENPP6* |
| ENSG00000152214 | 18 | 40313192 | 40705657 | 989 | 109 | 43165 | 0.22291 | 0.4118 | *RIT2* |
| ENSG00000131375 | 3 | 15237659 | 15304425 | 88 | 19 | 35586 | 0.22287 | 0.41182 | *CAPN7* |
| ENSG00000090686 | 1 | 21994791 | 22120099 | 483 | 57 | 45991 | 0.2226 | 0.41192 | *USP48* |
| ENSG00000248050 | 8 | 110646344 | 110670313 | 62 | 16 | 43788 | 0.22242 | 0.41199 | *RP11-422N16.3* |
| ENSG00000198270 | 12 | 112359086 | 112460970 | 154 | 8 | 46911 | 0.22241 | 0.412 | *TMEM116* |
| ENSG00000116815 | 1 | 117047157 | 117123661 | 101 | 20 | 34948 | 0.22214 | 0.4121 | *CD58* |
| ENSG00000162888 | 1 | 206654449 | 206681061 | 87 | 17 | 44117 | 0.2219 | 0.4122 | *C1orf147* |
| ENSG00000185236 | 19 | 8444865 | 8479318 | 158 | 25 | 51587 | 0.2219 | 0.4122 | *RAB11B* |

| | | | | | | | | | |
|---|---|---|---|---|---|---|---|---|---|
| ENSG00000081026 | 1 | 113923371 | 114238545 | 737 | 52 | 41793 | 0.22162 | 0.4123 | *MAGI3* |
| ENSG00000198225 | 6 | 63911351 | 63932929 | 86 | 6 | 56123 | 0.22152 | 0.41234 | *FKBP1C* |
| ENSG00000128739 | 15 | 25058794 | 25233870 | 691 | 137 | 48194 | 0.22128 | 0.41244 | *SNRPN* |
| ENSG00000157911 | 1 | 2326236 | 2355236 | 96 | 23 | 45201 | 0.22121 | 0.41247 | *PEX10* |
| ENSG00000155974 | 12 | 66731211 | 67207966 | 1745 | 168 | 51861 | 0.22113 | 0.41249 | *GRIP1* |
| ENSG00000196711 | 8 | 53436597 | 53488067 | 125 | 16 | 38071 | 0.22089 | 0.41259 | *FAM150A* |
| ENSG00000171522 | 5 | 40669600 | 40703837 | 101 | 15 | 44473 | 0.22061 | 0.4127 | *PTGER4* |
| ENSG00000113742 | 5 | 173305283 | 173398979 | 249 | 25 | 37474 | 0.22042 | 0.41277 | *CPEB4* |
| ENSG00000180336 | 17 | 42723762 | 42777676 | 107 | 21 | 49833 | 0.22043 | 0.41277 | *C17orf104* |
| ENSG00000162704 | 1 | 183582401 | 183614892 | 214 | 9 | 24577 | 0.22041 | 0.41278 | *ARPC5* |
| ENSG00000163590 | 3 | 160463390 | 160806695 | 940 | 76 | 43183 | 0.22031 | 0.41281 | *PPM1L* |
| ENSG00000170215 | 9 | 67782938 | 67804111 | 1 | 1 | 42735 | 0.22009 | 0.4129 | *FAM27B* |
| ENSG00000135974 | 2 | 105943816 | 105975668 | 119 | 17 | 49832 | 0.21994 | 0.41296 | *C2orf49* |
| ENSG00000106537 | 7 | 16783160 | 16834161 | 268 | 52 | 49278 | 0.21993 | 0.41296 | *TSPAN13* |
| ENSG00000127914 | 7 | 91560181 | 91749987 | 387 | 12 | 52734 | 0.21994 | 0.41296 | *AKAP9* |
| ENSG00000147454 | 8 | 23376318 | 23442976 | 420 | 53 | 46629 | 0.21947 | 0.41314 | *SLC25A37* |
| ENSG00000103269 | 16 | 715666 | 738268 | 59 | 17 | 39797 | 0.21929 | 0.41321 | *RHBDL1* |
| ENSG00000204231 | 6 | 33151365 | 33178630 | 73 | 16 | 48716 | 0.21916 | 0.41326 | *RXRB* |
| ENSG00000143373 | 1 | 151244094 | 151274656 | 37 | 8 | 50403 | 0.21891 | 0.41336 | *ZNF687* |
| ENSG00000183729 | 8 | 53840991 | 53863677 | 122 | 17 | 44625 | 0.21885 | 0.41338 | *NPBWR1* |
| ENSG00000136643 | 1 | 213214589 | 213458116 | 348 | 35 | 50358 | 0.21863 | 0.41347 | *RPS6KC1* |
| ENSG00000186297 | 15 | 27101510 | 27204354 | 294 | 59 | 44400 | 0.21836 | 0.41357 | *GABRA5* |
| ENSG00000246223 | 14 | 98381947 | 98454461 | 283 | 38 | 42539 | 0.21835 | 0.41358 | *C14orf64* |
| ENSG00000196850 | 12 | 110959120 | 111031125 | 191 | 16 | 32127 | 0.21826 | 0.41361 | *PPTC7* |
| ENSG00000170262 | 21 | 33654124 | 33697095 | 104 | 24 | 42047 | 0.21811 | 0.41367 | *MRAP* |
| ENSG00000091536 | 17 | 18002020 | 18093116 | 184 | 26 | 36704 | 0.21787 | 0.41377 | *MYO15A* |
| ENSG00000144647 | 3 | 43110724 | 43157568 | 107 | 18 | 54586 | 0.21755 | 0.41389 | *POMGNT2* |
| ENSG00000182853 | 17 | 4678580 | 4699728 | 59 | 13 | 40277 | 0.21745 | 0.41393 | *VMO1* |
| ENSG00000096063 | 6 | 35790743 | 35899119 | 295 | 14 | 51377 | 0.21743 | 0.41394 | *SRPK1* |
| ENSG00000214688 | 10 | 73461458 | 73507581 | 104 | 21 | 44742 | 0.21735 | 0.41397 | *C10orf105* |
| ENSG00000269279 | 1 | 116905890 | 116936718 | 58 | 9 | 25750 | 0.21726 | 0.414 | *AL136376.1* |
| ENSG00000072444 | 10 | 47991603 | 48065018 | 3 | 2 | 42872 | 0.21714 | 0.41405 | *ASAH2C* |
| ENSG00000177879 | 5 | 115167178 | 115259778 | 190 | 31 | 49622 | 0.21705 | 0.41408 | *AP3S1* |
| ENSG00000260342 | 16 | 18789385 | 18822841 | 75 | 12 | 46377 | 0.21679 | 0.41419 | *RP11-1035H13.3* |
| ENSG00000129480 | 14 | 31905242 | 31936716 | 72 | 8 | 51752 | 0.21672 | 0.41421 | *DTD2* |
| ENSG00000031698 | 1 | 109746540 | 109790791 | 104 | 21 | 46255 | 0.21665 | 0.41424 | *SARS* |

| | | | | | | | | | |
|---|---|---|---|---|---|---|---|---|---|
| ENSG00000203780 | 10 | 127575108 | 127708161 | 383 | 29 | 51796 | 0.21661 | 0.41426 | *FANK1* |
| ENSG00000082146 | 2 | 202242581 | 202355569 | 191 | 17 | 39945 | 0.21654 | 0.41428 | *STRADB* |
| ENSG00000105738 | 19 | 38387868 | 38709012 | 919 | 95 | 42386 | 0.21654 | 0.41429 | *SIPA1L3* |
| ENSG00000213445 | 11 | 65395568 | 65428401 | 50 | 14 | 41584 | 0.21651 | 0.4143 | *SIPA1* |
| ENSG00000214306 | 11 | 107633129 | 107657344 | 88 | 17 | 53243 | 0.21627 | 0.41439 | *AP001024.2* |
| ENSG00000267318 | 17 | 58008269 | 58060462 | 90 | 16 | 38456 | 0.21625 | 0.4144 | *RP11-178C3.1* |
| ENSG00000205544 | 17 | 7296294 | 7317456 | 49 | 19 | 47901 | 0.21623 | 0.41441 | *TMEM256* |
| ENSG00000137766 | 15 | 54295101 | 54930806 | 2860 | 207 | 46251 | 0.21538 | 0.41474 | *UNC13C* |
| ENSG00000130283 | 19 | 18969361 | 19016905 | 122 | 28 | 46905 | 0.21526 | 0.41478 | *GDF1* |
| ENSG00000185669 | 16 | 88734090 | 88762901 | 90 | 20 | 52038 | 0.21508 | 0.41485 | *SNAI3* |
| ENSG00000122778 | 7 | 138506126 | 138676064 | 559 | 70 | 40241 | 0.21506 | 0.41486 | *KIAA1549* |
| ENSG00000203757 | 1 | 158676958 | 158697953 | 57 | 7 | 46150 | 0.21493 | 0.41491 | *OR6K3* |
| ENSG00000115808 | 2 | 37060783 | 37203615 | 385 | 40 | 43892 | 0.21492 | 0.41492 | *STRN* |
| ENSG00000087250 | 16 | 56612986 | 56635000 | 98 | 19 | 46354 | 0.21488 | 0.41493 | *MT3* |
| ENSG00000135069 | 9 | 80902059 | 80955009 | 231 | 40 | 43598 | 0.21478 | 0.41497 | *PSAT1* |
| ENSG00000174521 | 19 | 40711965 | 40734306 | 46 | 14 | 45625 | 0.2146 | 0.41504 | *TTC9B* |
| ENSG00000048540 | 12 | 16691307 | 16773528 | 98 | 34 | 46896 | 0.21455 | 0.41506 | *LMO3* |
| ENSG00000124203 | 20 | 57756075 | 57844168 | 273 | 48 | 38724 | 0.21451 | 0.41507 | *ZNF831* |
| ENSG00000196705 | 19 | 21314827 | 21383034 | 185 | 26 | 46586 | 0.21442 | 0.41511 | *ZNF431* |
| ENSG00000273266 | 12 | 54437661 | 54459814 | 40 | 14 | 52454 | 0.21421 | 0.41519 | *HOXC4* |
| ENSG00000185963 | 9 | 95463645 | 95537094 | 187 | 15 | 31416 | 0.21416 | 0.41521 | *BICD2* |
| ENSG00000254440 | 6 | 138527129 | 138549627 | 67 | 8 | 25906 | 0.21408 | 0.41524 | *PBOV1* |
| ENSG00000156011 | 8 | 18374811 | 18952240 | 3097 | 260 | 45247 | 0.21396 | 0.41529 | *PSD3* |
| ENSG00000182224 | 17 | 7751064 | 7775600 | 61 | 15 | 38384 | 0.21366 | 0.41541 | *CYB5D1* |
| ENSG00000140848 | 16 | 57116449 | 57191878 | 270 | 56 | 39863 | 0.21332 | 0.41554 | *CPNE2* |
| ENSG00000158985 | 5 | 130571186 | 130744140 | 257 | 25 | 33945 | 0.21317 | 0.4156 | *CDC42SE2* |
| ENSG00000170927 | 6 | 51470098 | 51962423 | 1803 | 106 | 43816 | 0.213 | 0.41567 | *PKHD1* |
| ENSG00000132386 | 17 | 1655253 | 1690868 | 151 | 24 | 45407 | 0.21294 | 0.41569 | *SERPINF1* |
| ENSG00000112761 | 6 | 112365275 | 112402171 | 82 | 13 | 53601 | 0.21269 | 0.41579 | *WISP3* |
| ENSG00000165125 | 7 | 142558956 | 142593507 | 105 | 12 | 23034 | 0.21264 | 0.4158 | *TRPV6* |
| ENSG00000162105 | 11 | 70303961 | 70973623 | 2053 | 231 | 44162 | 0.21266 | 0.4158 | *SHANK2* |
| ENSG00000132522 | 17 | 7204643 | 7228883 | 63 | 13 | 48671 | 0.21263 | 0.41581 | *GPS2* |
| ENSG00000160961 | 19 | 14790613 | 14854558 | 274 | 34 | 46497 | 0.21261 | 0.41582 | *ZNF333* |
| ENSG00000198042 | 8 | 33332268 | 33368778 | 133 | 12 | 49344 | 0.21248 | 0.41587 | *MAK16* |
| ENSG00000165521 | 14 | 89068775 | 89269096 | 417 | 35 | 36873 | 0.21242 | 0.41589 | *EML5* |
| ENSG00000153814 | 7 | 27860192 | 28230362 | 1047 | 131 | 46954 | 0.21228 | 0.41594 | *JAZF1* |

| | | | | | | | | | |
|---|---|---|---|---|---|---|---|---|---|
| ENSG00000174231 | 17 | 1543923 | 1598176 | 178 | 20 | 41119 | 0.21228 | 0.41594 | *PRPF8* |
| ENSG00000163535 | 2 | 201364731 | 201458505 | 244 | 29 | 54196 | 0.21221 | 0.41597 | *SGOL2* |
| ENSG00000162129 | 11 | 71993469 | 72155692 | 277 | 30 | 39537 | 0.21204 | 0.41604 | *CLPB* |
| ENSG00000166347 | 18 | 71910530 | 71969251 | 260 | 26 | 47903 | 0.21201 | 0.41605 | *CYB5A* |
| ENSG00000157404 | 4 | 55514085 | 55616881 | 279 | 44 | 43602 | 0.21183 | 0.41612 | *KIT* |
| ENSG00000197782 | 19 | 40565059 | 40606845 | 154 | 14 | 31890 | 0.21167 | 0.41618 | *ZNF780A* |
| ENSG00000168813 | 19 | 32826500 | 32888573 | 153 | 21 | 37169 | 0.21165 | 0.41619 | *ZNF507* |
| ENSG00000137955 | 1 | 76241879 | 76270764 | 109 | 18 | 37005 | 0.21164 | 0.4162 | *RABGGTB* |
| ENSG00000235750 | 1 | 175116123 | 175172135 | 197 | 26 | 43610 | 0.21156 | 0.41623 | *KIAA0040* |
| ENSG00000205220 | 16 | 67958405 | 67980990 | 16 | 5 | 31561 | 0.21148 | 0.41626 | *PSMB10* |
| ENSG00000136944 | 9 | 129366722 | 129473311 | 359 | 56 | 43899 | 0.21112 | 0.4164 | *LMX1B* |
| ENSG00000175215 | 12 | 58203710 | 58250522 | 63 | 10 | 34958 | 0.211 | 0.41644 | *CTDSP2* |
| ENSG00000188626 | 15 | 28937100 | 28993530 | 47 | 11 | 31404 | 0.21091 | 0.41648 | *GOLGA8M* |
| ENSG00000071909 | 2 | 171024655 | 171521681 | 1773 | 220 | 43991 | 0.21087 | 0.41649 | *MYO3B* |
| ENSG00000105707 | 19 | 35521410 | 35567475 | 115 | 23 | 49613 | 0.21088 | 0.41649 | *HPN* |
| ENSG00000103249 | 16 | 1484935 | 1535581 | 346 | 43 | 46703 | 0.21081 | 0.41652 | *CLCN7* |
| ENSG00000119820 | 2 | 32492979 | 32551663 | 141 | 29 | 44692 | 0.21057 | 0.41661 | *YIPF4* |
| ENSG00000129422 | 8 | 17491304 | 17668426 | 1146 | 122 | 39556 | 0.21057 | 0.41661 | *MTUS1* |
| ENSG00000183549 | 16 | 20410856 | 20462658 | 256 | 21 | 40345 | 0.21024 | 0.41674 | *ACSM5* |
| ENSG00000198077 | 19 | 41371344 | 41398657 | 117 | 20 | 47258 | 0.21011 | 0.41679 | *CYP2A7* |
| ENSG00000112530 | 6 | 163138164 | 163746524 | 2020 | 198 | 41699 | 0.2101 | 0.4168 | *PACRG* |
| ENSG00000167306 | 18 | 47339183 | 47731463 | 1615 | 124 | 47156 | 0.20983 | 0.4169 | *MYO5B* |
| ENSG00000105609 | 19 | 54744263 | 54771164 | 106 | 31 | 43208 | 0.20976 | 0.41693 | *LILRB5* |
| ENSG00000140459 | 15 | 74620100 | 74670081 | 117 | 18 | 37240 | 0.20952 | 0.41702 | *CYP11A1* |
| ENSG00000259132 | 14 | 23405479 | 23461467 | 110 | 14 | 41178 | 0.20936 | 0.41708 | *RP11-298I3.5* |
| ENSG00000177192 | 12 | 132403745 | 132438406 | 93 | 15 | 31258 | 0.20931 | 0.4171 | *PUS1* |
| ENSG00000165568 | 10 | 4818821 | 4900254 | 599 | 44 | 50042 | 0.20915 | 0.41716 | *AKR1E2* |
| ENSG00000010318 | 3 | 52434673 | 52467657 | 35 | 12 | 54564 | 0.20913 | 0.41717 | *PHF7* |
| ENSG00000145321 | 4 | 72597410 | 72679758 | 201 | 29 | 40659 | 0.20886 | 0.41728 | *GC* |
| ENSG00000162946 | 1 | 231752561 | 232187018 | 1344 | 185 | 49083 | 0.20879 | 0.41731 | *DISC1* |
| ENSG00000163882 | 3 | 184069506 | 184096384 | 67 | 18 | 44023 | 0.20853 | 0.41741 | *POLR2H* |
| ENSG00000076650 | 19 | 33561786 | 33631448 | 217 | 18 | 30259 | 0.20831 | 0.41749 | *GPATCH1* |
| ENSG00000128322 | 22 | 23905312 | 23932495 | 119 | 32 | 43938 | 0.2083 | 0.4175 | *IGLL1* |
| ENSG00000196591 | 6 | 114244192 | 114342472 | 273 | 38 | 54576 | 0.20819 | 0.41754 | *HDAC2* |
| ENSG00000162576 | 1 | 1278069 | 1307157 | 84 | 13 | 38099 | 0.20801 | 0.41761 | *MXRA8* |
| ENSG00000109861 | 11 | 88016760 | 88080955 | 241 | 32 | 41455 | 0.20802 | 0.41761 | *CTSC* |

| | | | | | | | | | |
|---|---|---|---|---|---|---|---|---|---|
| ENSG00000179715 | 12 | 47463386 | 47640443 | 567 | 81 | 40019 | 0.20778 | 0.4177 | *PCED1B* |
| ENSG00000166454 | 16 | 81059452 | 81090963 | 88 | 13 | 38187 | 0.20778 | 0.4177 | *ATMIN* |
| ENSG00000072210 | 17 | 19541449 | 19590911 | 155 | 15 | 52762 | 0.20773 | 0.41772 | *ALDH3A2* |
| ENSG00000134108 | 3 | 5153905 | 5232596 | 240 | 25 | 35086 | 0.20763 | 0.41776 | *ARL8B* |
| ENSG00000114850 | 3 | 156247929 | 156282973 | 112 | 16 | 54987 | 0.20763 | 0.41776 | *SSR3* |
| ENSG00000169246 | 16 | 21403558 | 21469888 | 1 | 1 | 42438 | 0.20752 | 0.4178 | *NPIPB3* |
| ENSG00000228594 | 1 | 1523392 | 1545476 | 32 | 10 | 43465 | 0.20748 | 0.41782 | *C1orf233* |
| ENSG00000157106 | 16 | 18806175 | 18947776 | 436 | 27 | 48374 | 0.20737 | 0.41786 | *SMG1* |
| ENSG00000118495 | 6 | 144251437 | 144395735 | 435 | 75 | 50870 | 0.20731 | 0.41788 | *PLAGL1* |
| ENSG00000157782 | 12 | 121068355 | 121115127 | 189 | 34 | 46247 | 0.20732 | 0.41788 | *CABP1* |
| ENSG00000165379 | 14 | 42066773 | 42383752 | 1224 | 82 | 43694 | 0.20732 | 0.41788 | *LRFN5* |
| ENSG00000166886 | 12 | 57472677 | 57499259 | 53 | 13 | 48353 | 0.2073 | 0.41789 | *NAB2* |
| ENSG00000131126 | 19 | 43895648 | 43932763 | 114 | 15 | 37934 | 0.2072 | 0.41793 | *TEX101* |
| ENSG00000144642 | 3 | 29312473 | 30061886 | 3091 | 285 | 45203 | 0.20714 | 0.41795 | *RBMS3* |
| ENSG00000248483 | 5 | 93060508 | 93087343 | 35 | 8 | 47825 | 0.20701 | 0.418 | *POU5F2* |
| ENSG00000143340 | 1 | 179702298 | 179795333 | 344 | 48 | 44783 | 0.20698 | 0.41801 | *FAM163A* |
| ENSG00000005102 | 17 | 41707756 | 41749322 | 121 | 22 | 39750 | 0.207 | 0.41801 | *MEOX1* |
| ENSG00000139372 | 12 | 104349582 | 104392652 | 203 | 29 | 45586 | 0.20623 | 0.4183 | *TDG* |
| ENSG00000162885 | 1 | 235603238 | 235677781 | 191 | 24 | 32581 | 0.20621 | 0.41831 | *B3GALNT2* |
| ENSG00000110367 | 11 | 118610034 | 118671858 | 165 | 18 | 54775 | 0.20614 | 0.41834 | *DDX6* |
| ENSG00000170469 | 5 | 138722252 | 138749777 | 30 | 6 | 27795 | 0.20608 | 0.41836 | *SPATA24* |
| ENSG00000206483 | 3 | 126280625 | 126337398 | 197 | 32 | 42243 | 0.2058 | 0.41847 | *TXNRD3NB* |
| ENSG00000134247 | 1 | 117442679 | 117542980 | 257 | 39 | 42302 | 0.20547 | 0.4186 | *PTGFRN* |
| ENSG00000154582 | 8 | 74841404 | 74894522 | 170 | 25 | 45053 | 0.20546 | 0.41861 | *TCEB1* |
| ENSG00000100350 | 22 | 36873237 | 36913148 | 140 | 22 | 38981 | 0.20533 | 0.41866 | *FOXRED2* |
| ENSG00000226124 | 2 | 200615267 | 200725896 | 253 | 50 | 45432 | 0.2053 | 0.41867 | *FTCDNL1* |
| ENSG00000250673 | 4 | 147135709 | 147160208 | 67 | 15 | 41894 | 0.20527 | 0.41868 | *RP11-6L6.2* |
| ENSG00000154080 | 18 | 24485595 | 24775281 | 938 | 85 | 47684 | 0.20528 | 0.41868 | *CHST9* |
| ENSG00000160883 | 5 | 176297870 | 176336333 | 94 | 25 | 44425 | 0.20504 | 0.41877 | *HK3* |
| ENSG00000139921 | 14 | 51696880 | 51732759 | 125 | 18 | 44692 | 0.20499 | 0.41879 | *TMX1* |
| ENSG00000091262 | 16 | 16232785 | 16327379 | 346 | 51 | 42228 | 0.20481 | 0.41886 | *ABCC6* |
| ENSG00000115232 | 2 | 182311929 | 182410914 | 329 | 40 | 49749 | 0.20476 | 0.41888 | *ITGA4* |
| ENSG00000136146 | 13 | 48617459 | 48679267 | 224 | 30 | 43775 | 0.20467 | 0.41891 | *MED4* |
| ENSG00000132109 | 11 | 4396127 | 4424926 | 156 | 16 | 49241 | 0.20458 | 0.41895 | *TRIM21* |
| ENSG00000133937 | 14 | 95224553 | 95246562 | 60 | 19 | 43302 | 0.20457 | 0.41895 | *GSC* |
| ENSG00000142675 | 1 | 26493894 | 26526377 | 93 | 16 | 42271 | 0.20439 | 0.41903 | *CNKSR1* |

| | | | | | | | | | |
|---|---|---|---|---|---|---|---|---|---|
| ENSG00000204006 | 1 | 51557906 | 51623752 | 103 | 17 | 40126 | 0.20431 | 0.41906 | *C1orf185* |
| ENSG00000186847 | 17 | 39728531 | 39753173 | 87 | 6 | 53020 | 0.20396 | 0.41919 | *KRT14* |
| ENSG00000149506 | 11 | 60625035 | 60653166 | 72 | 17 | 40638 | 0.20393 | 0.4192 | *ZP1* |
| ENSG00000143324 | 1 | 180591140 | 180869387 | 608 | 51 | 52158 | 0.20388 | 0.41922 | *XPR1* |
| ENSG00000219492 | 4 | 9375743 | 9400709 | 119 | 19 | 42858 | 0.20371 | 0.41929 | *RP11-1396O13.13* |
| ENSG00000124155 | 20 | 44034717 | 44064884 | 62 | 19 | 43957 | 0.20346 | 0.41939 | *PIGT* |
| ENSG00000204120 | 2 | 233552009 | 233735285 | 535 | 37 | 47363 | 0.20272 | 0.41968 | *GIGYF2* |
| ENSG00000066651 | 6 | 126297576 | 126370422 | 79 | 6 | 47812 | 0.20262 | 0.41972 | *TRMT11* |
| ENSG00000153551 | 3 | 32423163 | 32534559 | 481 | 66 | 52358 | 0.20249 | 0.41977 | *CMTM7* |
| ENSG00000166025 | 11 | 94429597 | 94619918 | 359 | 53 | 45337 | 0.20234 | 0.41982 | *AMOTL1* |
| ENSG00000122787 | 7 | 137677070 | 137812732 | 376 | 52 | 49313 | 0.20208 | 0.41993 | *AKR1D1* |
| ENSG00000215529 | 20 | 31436729 | 31559006 | 415 | 52 | 44551 | 0.20208 | 0.41993 | *EFCAB8* |
| ENSG00000158715 | 1 | 205616979 | 205659587 | 149 | 29 | 38101 | 0.20155 | 0.42013 | *SLC45A3* |
| ENSG00000237651 | 2 | 61362243 | 61401964 | 151 | 10 | 51030 | 0.2012 | 0.42027 | *C2orf74* |
| ENSG00000172339 | 1 | 95429963 | 95548501 | 403 | 42 | 49873 | 0.20083 | 0.42042 | *ALG14* |
| ENSG00000091181 | 3 | 3101233 | 3178297 | 351 | 95 | 45781 | 0.2008 | 0.42043 | *IL5RA* |
| ENSG00000166292 | 17 | 53786988 | 53819482 | 57 | 16 | 42343 | 0.20068 | 0.42048 | *TMEM100* |
| ENSG00000187474 | 19 | 52288416 | 52339442 | 235 | 32 | 51670 | 0.20066 | 0.42048 | *FPR3* |
| ENSG00000119318 | 9 | 110035418 | 110104475 | 280 | 29 | 50215 | 0.2006 | 0.42051 | *RAD23B* |
| ENSG00000104689 | 8 | 23037965 | 23092639 | 193 | 33 | 45134 | 0.20038 | 0.42059 | *TNFRSF10A* |
| ENSG00000174206 | 12 | 64570096 | 64626076 | 98 | 16 | 50374 | 0.19993 | 0.42077 | *C12orf66* |
| ENSG00000250571 | 8 | 144339603 | 144369101 | 105 | 13 | 36475 | 0.1999 | 0.42078 | *GLI4* |
| ENSG00000118156 | 19 | 48013942 | 48069113 | 75 | 13 | 36103 | 0.19983 | 0.42081 | *ZNF541* |
| ENSG00000185304 | 2 | 88045474 | 88295309 | 84 | 21 | 43507 | 0.19917 | 0.42107 | *RGPD2* |
| ENSG00000140905 | 16 | 81105566 | 81140008 | 173 | 11 | 46353 | 0.19909 | 0.42109 | *GCSH* |
| ENSG00000069206 | 8 | 24288443 | 24394483 | 345 | 35 | 47680 | 0.19906 | 0.42111 | *ADAM7* |
| ENSG00000162076 | 16 | 2923187 | 2959383 | 202 | 20 | 46008 | 0.19904 | 0.42111 | *FLYWCH2* |
| ENSG00000256294 | 19 | 44606334 | 44647027 | 114 | 16 | 29545 | 0.19907 | 0.42111 | *ZNF225* |
| ENSG00000197568 | 1 | 70810488 | 70861022 | 120 | 17 | 49398 | 0.19899 | 0.42113 | *HHLA3* |
| ENSG00000101188 | 20 | 61330189 | 61404123 | 295 | 50 | 38238 | 0.19885 | 0.42119 | *NTSR1* |
| ENSG00000258512 | 14 | 102186774 | 102208859 | 78 | 7 | 51626 | 0.19883 | 0.4212 | *RP11-796G6.2* |
| ENSG00000213977 | 17 | 3556196 | 3581976 | 121 | 19 | 48820 | 0.1988 | 0.42121 | *TAX1BP3* |
| ENSG00000178462 | 10 | 5425061 | 5456793 | 131 | 12 | 53467 | 0.19878 | 0.42122 | *TUBAL3* |
| ENSG00000165644 | 10 | 76983727 | 77005788 | 67 | 16 | 42268 | 0.19869 | 0.42125 | *COMTD1* |
| ENSG00000271741 | 1 | 35437134 | 35507342 | 55 | 10 | 31091 | 0.19867 | 0.42126 | *RP11-244H3.4* |
| ENSG00000100823 | 14 | 20913350 | 20935927 | 69 | 12 | 54616 | 0.19867 | 0.42126 | *APEX1* |

| | | | | | | | | | |
|---|---|---|---|---|---|---|---|---|---|
| ENSG00000174640 | 3 | 133641540 | 133781028 | 437 | 88 | 46919 | 0.19856 | 0.4213 | *SLCO2A1* |
| ENSG00000150628 | 4 | 177095789 | 177126822 | 114 | 22 | 55011 | 0.19842 | 0.42136 | *SPATA4* |
| ENSG00000102931 | 16 | 57269010 | 57297516 | 61 | 9 | 32448 | 0.1982 | 0.42144 | *ARL2BP* |
| ENSG00000140932 | 16 | 66603351 | 66632178 | 73 | 8 | 33581 | 0.19821 | 0.42144 | *CMTM2* |
| ENSG00000115461 | 2 | 217526828 | 217570248 | 112 | 19 | 39661 | 0.19804 | 0.42151 | *IGFBP5* |
| ENSG00000087302 | 14 | 52446193 | 52481420 | 98 | 19 | 45589 | 0.19802 | 0.42151 | *C14orf166* |
| ENSG00000125207 | 12 | 130812432 | 130867182 | 223 | 37 | 38918 | 0.19788 | 0.42157 | *PIWIL1* |
| ENSG00000171823 | 12 | 1665159 | 1713331 | 129 | 38 | 48956 | 0.19783 | 0.42159 | *FBXL14* |
| ENSG00000105289 | 19 | 3698107 | 3760811 | 233 | 45 | 50764 | 0.19774 | 0.42162 | *TJP3* |
| ENSG00000143842 | 1 | 204032243 | 204106863 | 208 | 55 | 50039 | 0.19703 | 0.4219 | *SOX13* |
| ENSG00000260916 | 15 | 55622230 | 55710708 | 363 | 29 | 33444 | 0.19678 | 0.422 | *CCPG1* |
| ENSG00000175197 | 12 | 57900371 | 57924300 | 32 | 5 | 47848 | 0.19658 | 0.42208 | *DDIT3* |
| ENSG00000174083 | 17 | 8696041 | 8780994 | 221 | 44 | 45369 | 0.19636 | 0.42217 | *PIK3R6* |
| ENSG00000169519 | 11 | 28119795 | 28365054 | 350 | 29 | 39422 | 0.19617 | 0.42224 | *METTL15* |
| ENSG00000181240 | 19 | 6416048 | 6443790 | 130 | 13 | 48604 | 0.19597 | 0.42232 | *SLC25A41* |
| ENSG00000075785 | 3 | 128434965 | 128543639 | 330 | 29 | 42170 | 0.19594 | 0.42233 | *RAB7A* |
| ENSG00000088756 | 18 | 6719717 | 6925715 | 598 | 84 | 38090 | 0.19571 | 0.42242 | *ARHGAP28* |
| ENSG00000150540 | 2 | 138711590 | 138783930 | 151 | 15 | 49183 | 0.19553 | 0.42249 | *HNMT* |
| ENSG00000116151 | 1 | 2242692 | 2333146 | 387 | 68 | 42808 | 0.19535 | 0.42256 | *MORN1* |
| ENSG00000173660 | 1 | 46759303 | 46792448 | 64 | 21 | 40570 | 0.19533 | 0.42257 | *UQCRH* |
| ENSG00000160214 | 21 | 45199394 | 45235174 | 186 | 17 | 32802 | 0.19513 | 0.42265 | *RRP1* |
| ENSG00000184357 | 6 | 27824570 | 27845359 | 60 | 13 | 34648 | 0.19491 | 0.42273 | *HIST1H1B* |
| ENSG00000173261 | 5 | 145453949 | 145493932 | 124 | 7 | 55183 | 0.19471 | 0.42281 | *PLAC8L1* |
| ENSG00000169223 | 5 | 176748563 | 176788853 | 74 | 13 | 38108 | 0.19463 | 0.42284 | *LMAN2* |
| ENSG00000132254 | 11 | 6486910 | 6512666 | 48 | 10 | 51778 | 0.19461 | 0.42285 | *ARFIP2* |
| ENSG00000135454 | 12 | 58007193 | 58037138 | 63 | 11 | 47091 | 0.19448 | 0.4229 | *B4GALNT1* |
| ENSG00000198917 | 9 | 131571930 | 131602100 | 105 | 12 | 38465 | 0.19444 | 0.42291 | *C9orf114* |
| ENSG00000163815 | 3 | 45033040 | 45087563 | 165 | 16 | 28078 | 0.19441 | 0.42293 | *CLEC3B* |
| ENSG00000100665 | 14 | 95017428 | 95046250 | 143 | 22 | 55806 | 0.19436 | 0.42295 | *SERPINA4* |
| ENSG00000256683 | 19 | 52457596 | 52500109 | 170 | 18 | 30842 | 0.19422 | 0.423 | *ZNF350* |
| ENSG00000198156 | 16 | 28343876 | 28384829 | 14 | 4 | 42756 | 0.19398 | 0.42309 | *NPIPB6* |
| ENSG00000163710 | 3 | 142524764 | 142618045 | 290 | 53 | 45563 | 0.19391 | 0.42312 | *PCOLCE2* |
| ENSG00000116882 | 1 | 119901402 | 119946753 | 137 | 18 | 36978 | 0.19382 | 0.42316 | *HAO2* |
| ENSG00000149196 | 11 | 86003253 | 86066969 | 276 | 22 | 47739 | 0.19372 | 0.4232 | *C11orf73* |
| ENSG00000186814 | 18 | 32821032 | 32880196 | 94 | 26 | 40316 | 0.19358 | 0.42325 | *ZSCAN30* |
| ENSG00000183495 | 12 | 132424508 | 132575005 | 270 | 30 | 36788 | 0.19337 | 0.42334 | *EP400* |

| | | | | | | | | | |
|---|---|---|---|---|---|---|---|---|---|
| ENSG00000167100 | 17 | 48177404 | 48217246 | 94 | 26 | 43193 | 0.19334 | 0.42334 | *SAMD14* |
| ENSG00000131779 | 1 | 145506252 | 145533730 | 10 | 6 | 44407 | 0.19308 | 0.42345 | *PEX11B* |
| ENSG00000188909 | 11 | 122838278 | 122862428 | 91 | 8 | 53435 | 0.19294 | 0.4235 | *BSX* |
| ENSG00000160094 | 1 | 33712146 | 33776320 | 140 | 24 | 51702 | 0.19288 | 0.42353 | *ZNF362* |
| ENSG00000176853 | 8 | 124770696 | 124837692 | 167 | 22 | 38563 | 0.19265 | 0.42361 | *FAM91A1* |
| ENSG00000132744 | 11 | 67400026 | 67428130 | 95 | 22 | 44153 | 0.19248 | 0.42368 | *ACY3* |
| ENSG00000187980 | 1 | 20477746 | 20513917 | 87 | 17 | 51590 | 0.19222 | 0.42379 | *PLA2G2C* |
| ENSG00000131242 | 17 | 29708642 | 29875236 | 512 | 90 | 47025 | 0.19209 | 0.42384 | *RAB11FIP4* |
| ENSG00000163092 | 2 | 167734997 | 168126263 | 1497 | 83 | 42165 | 0.19126 | 0.42416 | *XIRP2* |
| ENSG00000111961 | 6 | 148583440 | 148883186 | 1255 | 186 | 49794 | 0.19112 | 0.42421 | *SASH1* |
| ENSG00000204161 | 10 | 50352773 | 50406630 | 219 | 40 | 41314 | 0.19113 | 0.42421 | *C10orf128* |
| ENSG00000116717 | 1 | 68140744 | 68164021 | 88 | 14 | 46428 | 0.19103 | 0.42425 | *GADD45A* |
| ENSG00000176994 | 17 | 18208624 | 18236517 | 87 | 11 | 46727 | 0.19103 | 0.42425 | *SMCR8* |
| ENSG00000240857 | 2 | 18725989 | 18751946 | 55 | 21 | 42006 | 0.19098 | 0.42427 | *RDH14* |
| ENSG00000112685 | 6 | 475133 | 703117 | 835 | 94 | 39365 | 0.19082 | 0.42433 | *EXOC2* |
| ENSG00000144048 | 2 | 73979311 | 74017284 | 33 | 7 | 49363 | 0.19079 | 0.42434 | *DUSP11* |
| ENSG00000154451 | 1 | 89714633 | 89748544 | 57 | 11 | 44490 | 0.19078 | 0.42435 | *GBP5* |
| ENSG00000170782 | 11 | 6887856 | 6908850 | 109 | 11 | 54373 | 0.19074 | 0.42436 | *OR10A4* |
| ENSG00000159884 | 9 | 35648301 | 35671508 | 87 | 11 | 40498 | 0.19064 | 0.4244 | *CCDC107* |
| ENSG00000075426 | 2 | 28605315 | 28650179 | 98 | 20 | 44345 | 0.19013 | 0.4246 | *FOSL2* |
| ENSG00000196935 | 12 | 64228073 | 64551613 | 719 | 117 | 42101 | 0.19006 | 0.42463 | *SRGAP1* |
| ENSG00000154518 | 2 | 176030986 | 176059335 | 94 | 13 | 45694 | 0.18995 | 0.42467 | *ATP5G3* |
| ENSG00000100220 | 22 | 32773569 | 32818242 | 182 | 35 | 47091 | 0.18977 | 0.42474 | *RTCB* |
| ENSG00000165383 | 10 | 50107528 | 50153242 | 216 | 32 | 40911 | 0.18961 | 0.42481 | *LRRC18* |
| ENSG00000121634 | 1 | 147364946 | 147391393 | 88 | 21 | 50801 | 0.18955 | 0.42483 | *GJA8* |
| ENSG00000166401 | 18 | 61627159 | 61682278 | 178 | 39 | 46428 | 0.18943 | 0.42488 | *SERPINB8* |
| ENSG00000198327 | 6 | 26230561 | 26250976 | 67 | 9 | 49634 | 0.18938 | 0.4249 | *HIST1H4F* |
| ENSG00000214413 | 10 | 112648488 | 112689032 | 95 | 19 | 49528 | 0.18931 | 0.42493 | *BBIP1* |
| ENSG00000166189 | 10 | 103815147 | 103837792 | 35 | 12 | 53019 | 0.18926 | 0.42494 | *HPS6* |
| ENSG00000010322 | 3 | 52479134 | 52537087 | 109 | 10 | 53779 | 0.18898 | 0.42505 | *NISCH* |
| ENSG00000001630 | 7 | 91731465 | 91782266 | 112 | 7 | 51241 | 0.189 | 0.42505 | *CYP51A1* |
| ENSG00000155897 | 8 | 131782547 | 132064672 | 1084 | 96 | 45844 | 0.18877 | 0.42513 | *ADCY8* |
| ENSG00000105197 | 19 | 39961052 | 39994422 | 101 | 13 | 43583 | 0.1888 | 0.42513 | *TIMM50* |
| ENSG00000188056 | 6 | 41186062 | 41216620 | 187 | 16 | 36683 | 0.1887 | 0.42516 | *TREML4* |
| ENSG00000130598 | 11 | 1850219 | 1872910 | 34 | 17 | 47657 | 0.18867 | 0.42518 | *TNNI2* |
| ENSG00000172817 | 8 | 65490320 | 65721318 | 498 | 49 | 36293 | 0.18846 | 0.42526 | *CYP7B1* |

| | | | | | | | | | |
|---|---|---|---|---|---|---|---|---|---|
| ENSG00000123136 | 19 | 14509631 | 14540192 | 72 | 19 | 38510 | 0.18831 | 0.42532 | *DDX39A* |
| ENSG00000087338 | 2 | 70046774 | 70118528 | 135 | 16 | 43415 | 0.18829 | 0.42533 | *GMCL1* |
| ENSG00000198168 | 11 | 22825345 | 22861845 | 82 | 13 | 40406 | 0.18813 | 0.42539 | *SVIP* |
| ENSG00000141562 | 17 | 80406056 | 80458413 | 206 | 24 | 39012 | 0.18803 | 0.42543 | *NARF* |
| ENSG00000135519 | 12 | 49922940 | 49962091 | 82 | 12 | 34574 | 0.18796 | 0.42545 | *KCNH3* |
| ENSG00000168675 | 18 | 13207497 | 13662754 | 1304 | 182 | 43883 | 0.18796 | 0.42545 | *LDLRAD4* |
| ENSG00000139132 | 12 | 32542463 | 32808984 | 1186 | 95 | 49181 | 0.18727 | 0.42572 | *FGD4* |
| ENSG00000139083 | 12 | 11792788 | 12058336 | 683 | 144 | 43895 | 0.18712 | 0.42578 | *ETV6* |
| ENSG00000178719 | 8 | 145054226 | 145077583 | 53 | 6 | 50762 | 0.18693 | 0.42586 | *GRINA* |
| ENSG00000265629 | 2 | 14765215 | 14800933 | 62 | 13 | 42734 | 0.18691 | 0.42587 | *AC011897.1* |
| ENSG00000120963 | 8 | 102180106 | 102228421 | 97 | 14 | 30074 | 0.18644 | 0.42605 | *ZNF706* |
| ENSG00000262003 | 17 | 802872 | 824452 | 134 | 24 | 42940 | 0.18643 | 0.42606 | *RP11-676J12.7* |
| ENSG00000128059 | 4 | 57249528 | 57311781 | 202 | 22 | 49569 | 0.18631 | 0.4261 | *PPAT* |
| ENSG00000183668 | 19 | 43705943 | 43783682 | 236 | 36 | 31635 | 0.18627 | 0.42612 | *PSG9* |
| ENSG00000161654 | 17 | 42102003 | 42154987 | 93 | 17 | 44353 | 0.18615 | 0.42616 | *LSM12* |
| ENSG00000184672 | 8 | 85085022 | 85844079 | 2107 | 114 | 45548 | 0.18606 | 0.4262 | *RALYL* |
| ENSG00000239713 | 22 | 39426924 | 39493748 | 216 | 30 | 39291 | 0.18589 | 0.42626 | *APOBEC3G* |
| ENSG00000039319 | 5 | 79693832 | 79785169 | 252 | 15 | 30129 | 0.18584 | 0.42628 | *ZFYVE16* |
| ENSG00000142784 | 1 | 27551007 | 27645110 | 175 | 22 | 48401 | 0.18567 | 0.42635 | *WDTC1* |
| ENSG00000213988 | 19 | 20178803 | 20247885 | 200 | 28 | 31992 | 0.18564 | 0.42637 | *ZNF90* |
| ENSG00000156958 | 15 | 49437853 | 49670066 | 744 | 45 | 44359 | 0.18505 | 0.42659 | *GALK2* |
| ENSG00000152767 | 13 | 98784816 | 99112027 | 1291 | 182 | 43648 | 0.18502 | 0.42661 | *FARP1* |
| ENSG00000162642 | 1 | 85705639 | 85735355 | 107 | 23 | 55168 | 0.18473 | 0.42672 | *C1orf52* |
| ENSG00000161944 | 17 | 6994641 | 7029019 | 192 | 31 | 41585 | 0.18452 | 0.4268 | *ASGR2* |
| ENSG00000133028 | 17 | 10573654 | 10611692 | 75 | 8 | 38799 | 0.18444 | 0.42684 | *SCO1* |
| ENSG00000083067 | 9 | 73133979 | 74071820 | 3286 | 195 | 49767 | 0.18441 | 0.42685 | *TRPM3* |
| ENSG00000132677 | 1 | 156329003 | 156365011 | 100 | 24 | 44409 | 0.18424 | 0.42691 | *RHBG* |
| ENSG00000167670 | 19 | 4392659 | 4455015 | 198 | 21 | 51854 | 0.18416 | 0.42695 | *CHAF1A* |
| ENSG00000215012 | 22 | 19823661 | 19852419 | 90 | 18 | 38353 | 0.18407 | 0.42698 | *C22orf29* |
| ENSG00000121989 | 2 | 148592086 | 148698393 | 156 | 18 | 53496 | 0.18403 | 0.427 | *ACVR2A* |
| ENSG00000137074 | 9 | 32962604 | 33035166 | 347 | 14 | 53462 | 0.18386 | 0.42706 | *APTX* |
| ENSG00000163041 | 1 | 226239552 | 226269702 | 50 | 12 | 53482 | 0.18383 | 0.42707 | *H3F3A* |
| ENSG00000167210 | 18 | 44046935 | 44246996 | 698 | 111 | 48283 | 0.18384 | 0.42707 | *LOXHD1* |
| ENSG00000163867 | 1 | 35439523 | 35507569 | 52 | 9 | 30446 | 0.18379 | 0.42709 | *ZMYM6* |
| ENSG00000228856 | 4 | 9354855 | 9376447 | 60 | 7 | 41443 | 0.18377 | 0.4271 | *USP17L30* |
| ENSG00000224982 | 12 | 120021264 | 120089363 | 211 | 17 | 50194 | 0.18375 | 0.42711 | *TMEM233* |

| | | | | | | | | | |
|---|---|---|---|---|---|---|---|---|---|
| ENSG00000139219 | 12 | 48356748 | 48408269 | 207 | 31 | 45484 | 0.1836 | 0.42716 | *COL2A1* |
| ENSG00000079102 | 8 | 92957203 | 93125514 | 232 | 19 | 46958 | 0.18354 | 0.42719 | *RUNX1T1* |
| ENSG00000187240 | 11 | 102970160 | 103360591 | 2070 | 67 | 46685 | 0.18354 | 0.42719 | *DYNC2H1* |
| ENSG00000143977 | 2 | 70498494 | 70530903 | 70 | 8 | 46009 | 0.18341 | 0.42724 | *SNRPG* |
| ENSG00000137812 | 15 | 40876218 | 40966540 | 189 | 17 | 46931 | 0.18311 | 0.42735 | *CASC5* |
| ENSG00000048405 | 7 | 126976844 | 127081978 | 370 | 35 | 50564 | 0.18291 | 0.42743 | *ZNF800* |
| ENSG00000139291 | 12 | 72069867 | 72107836 | 59 | 13 | 40659 | 0.18242 | 0.42763 | *TMEM19* |
| ENSG00000100714 | 14 | 64844749 | 64936722 | 295 | 44 | 39362 | 0.18219 | 0.42772 | *MTHFD1* |
| ENSG00000070214 | 9 | 107996903 | 108211452 | 543 | 48 | 32540 | 0.182 | 0.42779 | *SLC44A1* |
| ENSG00000158525 | 7 | 129974630 | 130018571 | 197 | 27 | 55251 | 0.18184 | 0.42785 | *CPA5* |
| ENSG00000182257 | 22 | 46435358 | 46460024 | 84 | 17 | 39907 | 0.18174 | 0.42789 | *C22orf26* |
| ENSG00000100987 | 20 | 25041521 | 25072996 | 85 | 9 | 45561 | 0.18172 | 0.4279 | *VSX1* |
| ENSG00000021355 | 6 | 2822566 | 2852240 | 143 | 28 | 51727 | 0.18168 | 0.42791 | *SERPINB1* |
| ENSG00000117408 | 1 | 44402611 | 44443694 | 71 | 12 | 33048 | 0.18149 | 0.42799 | *IPO13* |
| ENSG00000168255 | 7 | 102168365 | 102223103 | 5 | 3 | 42887 | 0.18127 | 0.42808 | *POLR2J3* |
| ENSG00000269558 | 7 | 65102081 | 65123280 | 3 | 1 | 42945 | 0.18116 | 0.42812 | *AC104057.1* |
| ENSG00000123843 | 1 | 207252187 | 207283338 | 68 | 14 | 33867 | 0.18103 | 0.42817 | *C4BPB* |
| ENSG00000221914 | 8 | 26139007 | 26240196 | 261 | 43 | 50608 | 0.18096 | 0.4282 | *PPP2R2A* |
| ENSG00000228120 | 21 | 44569176 | 44592387 | 44 | 15 | 43878 | 0.1809 | 0.42822 | *AP001631.10* |
| ENSG00000101180 | 20 | 60780026 | 60805323 | 95 | 32 | 49167 | 0.18076 | 0.42828 | *HRH3* |
| ENSG00000185650 | 14 | 69244377 | 69273190 | 92 | 16 | 52083 | 0.18067 | 0.42831 | *ZFP36L1* |
| ENSG00000243709 | 1 | 226063982 | 226109082 | 168 | 25 | 41665 | 0.18058 | 0.42835 | *LEFTY1* |
| ENSG00000174038 | 9 | 35031092 | 35055988 | 48 | 12 | 43455 | 0.18029 | 0.42846 | *C9orf131* |
| ENSG00000213221 | 9 | 139243932 | 139268241 | 106 | 19 | 53116 | 0.18028 | 0.42847 | *DNLZ* |
| ENSG00000101098 | 20 | 43370449 | 43448979 | 163 | 34 | 45597 | 0.17993 | 0.4286 | *RIMS4* |
| ENSG00000066739 | 14 | 96737595 | 96840207 | 202 | 23 | 47352 | 0.17977 | 0.42867 | *ATG2B* |
| ENSG00000178425 | 6 | 116412012 | 116580660 | 448 | 46 | 39763 | 0.1797 | 0.42869 | *NT5DC1* |
| ENSG00000196570 | 5 | 176817108 | 176837637 | 24 | 9 | 46000 | 0.17939 | 0.42881 | *PFN3* |
| ENSG00000137764 | 15 | 67825047 | 68109461 | 723 | 55 | 41731 | 0.17928 | 0.42886 | *MAP2K5* |
| ENSG00000167554 | 19 | 52829498 | 52881031 | 225 | 35 | 48451 | 0.17928 | 0.42886 | *ZNF610* |
| ENSG00000171703 | 20 | 62671189 | 62713700 | 122 | 29 | 47627 | 0.17921 | 0.42889 | *TCEA2* |
| ENSG00000114790 | 3 | 153828792 | 153985616 | 451 | 39 | 48161 | 0.17918 | 0.4289 | *ARHGEF26* |
| ENSG00000015479 | 5 | 138599441 | 138677360 | 87 | 15 | 38638 | 0.17911 | 0.42893 | *MATR3* |
| ENSG00000108439 | 17 | 46008872 | 46035654 | 79 | 15 | 50957 | 0.17905 | 0.42895 | *PNPO* |
| ENSG00000132465 | 4 | 71511258 | 71557534 | 64 | 18 | 40236 | 0.17894 | 0.42899 | *IGJ* |
| ENSG00000167065 | 22 | 31038038 | 31073877 | 138 | 33 | 39865 | 0.17874 | 0.42907 | *DUSP18* |

| | | | | | | | | | |
|---|---|---|---|---|---|---|---|---|---|
| ENSG00000151151 | 10 | 59941278 | 60037694 | 405 | 17 | 31736 | 0.17858 | 0.42913 | *IPMK* |
| ENSG00000266964 | 19 | 35619712 | 35644013 | 50 | 13 | 53645 | 0.17824 | 0.42927 | *FXYD1* |
| ENSG00000105997 | 7 | 27135803 | 27202200 | 164 | 25 | 38199 | 0.17819 | 0.42929 | *HOXA3* |
| ENSG00000101019 | 20 | 33880369 | 34009944 | 249 | 17 | 30040 | 0.17779 | 0.42944 | *UQCC1* |
| ENSG00000128708 | 2 | 172768958 | 172858599 | 209 | 30 | 43066 | 0.17771 | 0.42947 | *HAT1* |
| ENSG00000168434 | 16 | 23389814 | 23474501 | 158 | 17 | 37237 | 0.1774 | 0.4296 | *COG7* |
| ENSG00000186510 | 1 | 16335370 | 16370545 | 238 | 27 | 53673 | 0.1769 | 0.4298 | *CLCNKA* |
| ENSG00000175449 | 5 | 94972458 | 95030477 | 155 | 23 | 48090 | 0.17678 | 0.42984 | *RFESD* |
| ENSG00000133742 | 8 | 86229837 | 86301243 | 156 | 27 | 50000 | 0.17672 | 0.42987 | *CA1* |
| ENSG00000157330 | 1 | 12796163 | 12833847 | 147 | 16 | 48687 | 0.17655 | 0.42993 | *C1orf158* |
| ENSG00000186575 | 22 | 29989545 | 30104587 | 280 | 33 | 45036 | 0.17638 | 0.43 | *NF2* |
| ENSG00000182010 | 10 | 63932794 | 64038466 | 319 | 23 | 48577 | 0.17595 | 0.43017 | *RTKN2* |
| ENSG00000177917 | 2 | 153564407 | 153627767 | 181 | 27 | 43627 | 0.17563 | 0.43029 | *ARL6IP6* |
| ENSG00000104059 | 15 | 29402457 | 29872927 | 1520 | 145 | 40864 | 0.17563 | 0.43029 | *FAM189A1* |
| ENSG00000186458 | 20 | 228377 | 251737 | 117 | 18 | 47929 | 0.17547 | 0.43035 | *DEFB132* |
| ENSG00000189376 | 8 | 124222196 | 124263638 | 20 | 14 | 47757 | 0.17537 | 0.43039 | *C8orf76* |
| ENSG00000129219 | 17 | 4700391 | 4736729 | 106 | 26 | 38494 | 0.17537 | 0.43039 | *PLD2* |
| ENSG00000138031 | 2 | 25032038 | 25152708 | 472 | 37 | 40973 | 0.17527 | 0.43043 | *ADCY3* |
| ENSG00000153790 | 7 | 25164316 | 25229975 | 284 | 23 | 38966 | 0.17518 | 0.43047 | *C7orf31* |
| ENSG00000225950 | 19 | 49549050 | 49578333 | 119 | 25 | 44758 | 0.17511 | 0.4305 | *NTF4* |
| ENSG00000107262 | 9 | 33237818 | 33274761 | 69 | 8 | 37038 | 0.17502 | 0.43053 | *BAG1* |
| ENSG00000175606 | 8 | 74874672 | 74905018 | 112 | 13 | 41669 | 0.17497 | 0.43055 | *TMEM70* |
| ENSG00000181915 | 10 | 64554516 | 64578238 | 48 | 10 | 31573 | 0.17489 | 0.43058 | *ADO* |
| ENSG00000161958 | 17 | 7331592 | 7358256 | 60 | 13 | 45245 | 0.17459 | 0.4307 | *FGF11* |
| ENSG00000006118 | 11 | 60681935 | 60714631 | 90 | 12 | 41491 | 0.17454 | 0.43072 | *TMEM132A* |
| ENSG00000091490 | 4 | 25739055 | 25875382 | 360 | 78 | 47046 | 0.17447 | 0.43075 | *SEL1L3* |
| ENSG00000113161 | 5 | 74622154 | 74667929 | 105 | 17 | 53323 | 0.17436 | 0.43079 | *HMGCR* |
| ENSG00000169972 | 1 | 1233947 | 1257057 | 42 | 14 | 36852 | 0.17349 | 0.43113 | *PUSL1* |
| ENSG00000172426 | 6 | 43602783 | 43650336 | 138 | 20 | 32264 | 0.17349 | 0.43113 | *RSPH9* |
| ENSG00000172243 | 12 | 10259376 | 10292857 | 122 | 17 | 27946 | 0.17349 | 0.43113 | *CLEC7A* |
| ENSG00000125912 | 19 | 3175561 | 3219573 | 167 | 21 | 48044 | 0.17349 | 0.43113 | *NCLN* |
| ENSG00000267697 | 7 | 135602022 | 135622198 | 73 | 14 | 39856 | 0.17323 | 0.43124 | *LUZP6* |
| ENSG00000117020 | 1 | 243641535 | 244024381 | 624 | 48 | 42625 | 0.17299 | 0.43133 | *AKT3* |
| ENSG00000255800 | 9 | 107350650 | 107371788 | 132 | 8 | 25449 | 0.17273 | 0.43143 | *OR13C5* |
| ENSG00000177951 | 11 | 157784 | 217428 | 95 | 20 | 45291 | 0.17229 | 0.4316 | *BET1L* |
| ENSG00000103365 | 16 | 23464863 | 23543316 | 203 | 19 | 44793 | 0.17222 | 0.43163 | *GGA2* |

| | | | | | | | | | |
|---|---|---|---|---|---|---|---|---|---|
| ENSG00000147894 | 9 | 27536544 | 27583864 | 217 | 32 | 50936 | 0.17217 | 0.43165 | *C9orf72* |
| ENSG00000180481 | 12 | 75774850 | 75836468 | 222 | 11 | 55244 | 0.17193 | 0.43175 | *GLIPR1L2* |
| ENSG00000171311 | 10 | 99185899 | 99215774 | 57 | 14 | 48417 | 0.17151 | 0.43191 | *EXOSC1* |
| ENSG00000186204 | 19 | 15773567 | 15817984 | 353 | 22 | 50375 | 0.17139 | 0.43196 | *CYP4F12* |
| ENSG00000122707 | 9 | 36026430 | 36134448 | 319 | 35 | 39805 | 0.17093 | 0.43214 | *RECK* |
| ENSG00000148396 | 9 | 139324549 | 139382141 | 204 | 29 | 43649 | 0.17091 | 0.43215 | *SEC16A* |
| ENSG00000074621 | 15 | 65893704 | 65963333 | 132 | 12 | 48664 | 0.17092 | 0.43215 | *SLC24A1* |
| ENSG00000197888 | 4 | 69392902 | 69444245 | 22 | 12 | 43072 | 0.17085 | 0.43217 | *UGT2B17* |
| ENSG00000162928 | 2 | 61234360 | 61289125 | 106 | 25 | 45386 | 0.17069 | 0.43223 | *PEX13* |
| ENSG00000117281 | 1 | 145685798 | 145725614 | 27 | 7 | 44158 | 0.17056 | 0.43229 | *CD160* |
| ENSG00000100258 | 22 | 50931376 | 50956135 | 100 | 27 | 49571 | 0.17045 | 0.43233 | *LMF2* |
| ENSG00000185838 | 22 | 19760747 | 19852462 | 258 | 45 | 42644 | 0.17037 | 0.43236 | *GNB1L* |
| ENSG00000006432 | 14 | 71179243 | 71286251 | 440 | 44 | 54675 | 0.17023 | 0.43241 | *MAP3K9* |
| ENSG00000128276 | 22 | 32740872 | 32767148 | 147 | 26 | 37395 | 0.17001 | 0.4325 | *RFPL3* |
| ENSG00000180628 | 10 | 92969908 | 93054088 | 199 | 13 | 27617 | 0.16995 | 0.43252 | *PCGF5* |
| ENSG00000187461 | 6 | 13459513 | 13496415 | 84 | 19 | 40776 | 0.16987 | 0.43256 | *AL583828.1* |
| ENSG00000087253 | 16 | 55532910 | 55630582 | 265 | 26 | 39237 | 0.16899 | 0.4329 | *LPCAT2* |
| ENSG00000203985 | 1 | 54464412 | 54493856 | 118 | 12 | 45932 | 0.1687 | 0.43301 | *LDLRAD1* |
| ENSG00000174938 | 16 | 29872480 | 29920868 | 57 | 14 | 44644 | 0.16869 | 0.43302 | *SEZ6L2* |
| ENSG00000112996 | 5 | 44799027 | 44830530 | 81 | 7 | 52614 | 0.16861 | 0.43305 | *MRPS30* |
| ENSG00000111271 | 12 | 112113857 | 112204903 | 60 | 17 | 51623 | 0.16843 | 0.43312 | *ACAD10* |
| ENSG00000035687 | 1 | 244561796 | 244625436 | 165 | 26 | 52419 | 0.16832 | 0.43317 | *ADSS* |
| ENSG00000184492 | 2 | 114246661 | 114268728 | 18 | 10 | 36052 | 0.1683 | 0.43317 | *FOXD4L1* |
| ENSG00000196224 | 11 | 1618795 | 1639693 | 120 | 17 | 55058 | 0.1683 | 0.43317 | *KRTAP5-3* |
| ENSG00000187796 | 9 | 139246355 | 139278133 | 137 | 19 | 54442 | 0.16827 | 0.43318 | *CARD9* |
| ENSG00000087128 | 4 | 69303167 | 69373322 | 251 | 22 | 44446 | 0.1681 | 0.43325 | *TMPRSS11E* |
| ENSG00000110841 | 12 | 27666364 | 27858497 | 643 | 86 | 48052 | 0.1678 | 0.43337 | *PPFIBP1* |
| ENSG00000204248 | 6 | 33120458 | 33170276 | 212 | 30 | 49049 | 0.1674 | 0.43353 | *COL11A2* |
| ENSG00000178934 | 19 | 39269851 | 39292389 | 96 | 12 | 35553 | 0.16736 | 0.43354 | *LGALS7B* |
| ENSG00000008282 | 7 | 105720949 | 105763022 | 142 | 26 | 45213 | 0.16711 | 0.43364 | *SYPL1* |
| ENSG00000268936 | 11 | 134845246 | 134866693 | 124 | 13 | 50608 | 0.16695 | 0.43371 | *AP003062.1* |
| ENSG00000198211 | 16 | 89975573 | 90012500 | 172 | 28 | 29731 | 0.16668 | 0.43381 | *TUBB3* |
| ENSG00000101412 | 20 | 32253489 | 32284210 | 42 | 16 | 47436 | 0.16667 | 0.43382 | *E2F1* |
| ENSG00000165244 | 9 | 99138223 | 99190611 | 156 | 13 | 47474 | 0.16639 | 0.43392 | *ZNF367* |
| ENSG00000137154 | 9 | 19365713 | 19390252 | 50 | 9 | 37582 | 0.16629 | 0.43396 | *RPS6* |
| ENSG00000103326 | 16 | 567717 | 614636 | 134 | 37 | 40926 | 0.16624 | 0.43398 | *CAPN15* |

| | | | | | | | | | |
|---|---|---|---|---|---|---|---|---|---|
| ENSG00000124731 | 6 | 41225664 | 41264457 | 122 | 26 | 41634 | 0.16617 | 0.43401 | *TREM1* |
| ENSG00000269058 | 19 | 16579868 | 16617003 | 154 | 17 | 47383 | 0.16584 | 0.43414 | *CALR3* |
| ENSG00000028137 | 1 | 12217060 | 12279285 | 196 | 29 | 50644 | 0.16581 | 0.43415 | *TNFRSF1B* |
| ENSG00000214491 | 22 | 30908786 | 30952669 | 173 | 21 | 53206 | 0.16579 | 0.43416 | *SEC14L6* |
| ENSG00000221989 | 7 | 143796607 | 143817657 | 87 | 6 | 26372 | 0.16531 | 0.43435 | *OR2A2* |
| ENSG00000174718 | 12 | 32102304 | 32156039 | 273 | 50 | 53469 | 0.16513 | 0.43442 | *KIAA1551* |
| ENSG00000083168 | 8 | 41776997 | 41919508 | 238 | 22 | 33438 | 0.16508 | 0.43444 | *KAT6A* |
| ENSG00000167552 | 12 | 49568579 | 49593107 | 42 | 7 | 35522 | 0.16504 | 0.43446 | *TUBA1A* |
| ENSG00000183760 | 19 | 39564553 | 39612133 | 202 | 24 | 53860 | 0.165 | 0.43447 | *PAPL* |
| ENSG00000187608 | 1 | 938803 | 959920 | 71 | 11 | 38603 | 0.1649 | 0.43451 | *ISG15* |
| ENSG00000128626 | 19 | 39411188 | 39433802 | 62 | 10 | 38070 | 0.16469 | 0.4346 | *MRPS12* |
| ENSG00000117091 | 1 | 160638536 | 160691641 | 202 | 44 | 43728 | 0.16459 | 0.43463 | *CD48* |
| ENSG00000177646 | 3 | 128588439 | 128644910 | 257 | 24 | 30716 | 0.16458 | 0.43464 | *ACAD9* |
| ENSG00000263563 | 17 | 21719601 | 21741762 | 72 | 10 | 47338 | 0.1645 | 0.43467 | *UBBP4* |
| ENSG00000154589 | 8 | 74893587 | 74951322 | 186 | 21 | 40163 | 0.16437 | 0.43472 | *LY96* |
| ENSG00000108055 | 10 | 112317449 | 112374394 | 216 | 19 | 40548 | 0.164 | 0.43487 | *SMC3* |
| ENSG00000231924 | 19 | 43360616 | 43393974 | 115 | 15 | 23710 | 0.16397 | 0.43488 | *PSG1* |
| ENSG00000163683 | 4 | 39537950 | 39650710 | 321 | 49 | 38341 | 0.16389 | 0.43491 | *SMIM14* |
| ENSG00000130713 | 9 | 133559108 | 133590248 | 89 | 10 | 31055 | 0.16362 | 0.43502 | *EXOSC2* |
| ENSG00000143882 | 2 | 10851775 | 10935236 | 309 | 43 | 42686 | 0.16354 | 0.43505 | *ATP6V1C2* |
| ENSG00000197580 | 11 | 112036190 | 112105422 | 189 | 25 | 43025 | 0.16331 | 0.43514 | *BCO2* |
| ENSG00000133105 | 13 | 32303674 | 32387009 | 292 | 29 | 46772 | 0.16325 | 0.43516 | *RXFP2* |
| ENSG00000164047 | 3 | 48254837 | 48276981 | 32 | 13 | 46955 | 0.16319 | 0.43518 | *CAMP* |
| ENSG00000076928 | 19 | 42377228 | 42444302 | 59 | 15 | 42691 | 0.1631 | 0.43522 | *ARHGEF1* |
| ENSG00000143355 | 1 | 197871037 | 197914608 | 110 | 23 | 45360 | 0.16277 | 0.43535 | *LHX9* |
| ENSG00000114544 | 3 | 125715198 | 125830404 | 476 | 32 | 48635 | 0.16272 | 0.43537 | *SLC41A3* |
| ENSG00000173281 | 8 | 8983765 | 9019084 | 118 | 18 | 36564 | 0.16268 | 0.43538 | *PPP1R3B* |
| ENSG00000214367 | 4 | 2219191 | 2253891 | 107 | 11 | 36606 | 0.16244 | 0.43548 | *HAUS3* |
| ENSG00000146085 | 6 | 49388073 | 49440904 | 119 | 14 | 55478 | 0.16241 | 0.43549 | *MUT* |
| ENSG00000128394 | 22 | 39426609 | 39459915 | 83 | 11 | 40713 | 0.16231 | 0.43553 | *APOBEC3F* |
| ENSG00000173585 | 3 | 45917996 | 45954667 | 97 | 17 | 35931 | 0.16216 | 0.43559 | *CCR9* |
| ENSG00000254852 | 16 | 14831923 | 14869270 | 9 | 6 | 37584 | 0.16214 | 0.4356 | *NPIPA2* |
| ENSG00000132155 | 3 | 12615100 | 12715725 | 321 | 25 | 34238 | 0.16204 | 0.43564 | *RAF1* |
| ENSG00000164946 | 9 | 14724664 | 14920993 | 856 | 154 | 47152 | 0.16197 | 0.43567 | *FREM1* |
| ENSG00000181350 | 17 | 16334891 | 16405467 | 247 | 30 | 48477 | 0.16192 | 0.43568 | *FAM211A* |
| ENSG00000092758 | 20 | 61437596 | 61482511 | 252 | 54 | 46519 | 0.16181 | 0.43573 | *COL9A3* |

| | | | | | | | | | |
|---|---|---|---|---|---|---|---|---|---|
| ENSG00000187642 | 1 | 900579 | 927497 | 97 | 21 | 36685 | 0.16175 | 0.43575 | *C1orf170* |
| ENSG00000127526 | 19 | 16650642 | 16693193 | 76 | 20 | 45817 | 0.16175 | 0.43575 | *SLC35E1* |
| ENSG00000125630 | 2 | 113289492 | 113344635 | 169 | 19 | 41913 | 0.16173 | 0.43576 | *POLR1B* |
| ENSG00000180083 | 20 | 44267202 | 44308909 | 137 | 12 | 46540 | 0.16165 | 0.43579 | *WFDC11* |
| ENSG00000188987 | 6 | 26178938 | 26199304 | 91 | 10 | 49506 | 0.16158 | 0.43582 | *HIST1H4D* |
| ENSG00000105397 | 19 | 10451209 | 10501352 | 151 | 24 | 54044 | 0.1613 | 0.43593 | *TYK2* |
| ENSG00000149564 | 11 | 124612026 | 124642186 | 63 | 15 | 39858 | 0.16127 | 0.43594 | *ESAM* |
| ENSG00000173193 | 3 | 122389465 | 122459687 | 258 | 34 | 48170 | 0.1612 | 0.43597 | *PARP14* |
| ENSG00000138759 | 4 | 78968724 | 79475423 | 2046 | 113 | 41724 | 0.16107 | 0.43602 | *FRAS1* |
| ENSG00000107951 | 10 | 30588730 | 30673377 | 267 | 38 | 42602 | 0.16107 | 0.43602 | *MTPAP* |
| ENSG00000160326 | 9 | 136326217 | 136354259 | 89 | 26 | 46904 | 0.16101 | 0.43604 | *SLC2A6* |
| ENSG00000143970 | 2 | 25946622 | 26111385 | 449 | 29 | 51206 | 0.16096 | 0.43606 | *ASXL2* |
| ENSG00000186153 | 16 | 78123310 | 79256564 | 7213 | 478 | 42541 | 0.16079 | 0.43613 | *WWOX* |
| ENSG00000105398 | 19 | 48363723 | 48399654 | 158 | 23 | 49583 | 0.16062 | 0.43619 | *SULT2A1* |
| ENSG00000228198 | 1 | 248356332 | 248377392 | 81 | 8 | 50476 | 0.16058 | 0.43621 | *OR2M3* |
| ENSG00000115295 | 2 | 29310571 | 29422509 | 297 | 42 | 42504 | 0.16039 | 0.43629 | *CLIP4* |
| ENSG00000171747 | 19 | 39282311 | 39314004 | 128 | 20 | 35766 | 0.16007 | 0.43641 | *LGALS4* |
| ENSG00000137203 | 6 | 10383419 | 10429892 | 109 | 27 | 38882 | 0.15986 | 0.4365 | *TFAP2A* |
| ENSG00000183513 | 2 | 99205773 | 99234978 | 57 | 6 | 55399 | 0.1598 | 0.43652 | *COA5* |
| ENSG00000168542 | 2 | 189829046 | 189887472 | 128 | 27 | 45854 | 0.15976 | 0.43654 | *COL3A1* |
| ENSG00000256797 | 12 | 10024088 | 10058432 | 134 | 15 | 37575 | 0.15976 | 0.43654 | *KLRF2* |
| ENSG00000246922 | 15 | 65375338 | 65417535 | 54 | 17 | 48845 | 0.15954 | 0.43662 | *UBAP1L* |
| ENSG00000117153 | 1 | 202850228 | 202907764 | 149 | 14 | 42838 | 0.15937 | 0.43669 | *KLHL12* |
| ENSG00000156232 | 15 | 83468380 | 83513611 | 132 | 24 | 48937 | 0.15921 | 0.43675 | *WHAMM* |
| ENSG00000181355 | 6 | 9586343 | 10221841 | 2320 | 193 | 47511 | 0.1589 | 0.43687 | *OFCC1* |
| ENSG00000159588 | 1 | 46075716 | 46099729 | 74 | 6 | 54886 | 0.15888 | 0.43688 | *CCDC17* |
| ENSG00000182631 | 5 | 33926491 | 33949023 | 49 | 13 | 23825 | 0.15883 | 0.4369 | *RXFP3* |
| ENSG00000189182 | 12 | 53073410 | 53107247 | 124 | 22 | 50848 | 0.15876 | 0.43693 | *KRT77* |
| ENSG00000268397 | 5 | 180672720 | 180694578 | 72 | 15 | 35240 | 0.15872 | 0.43694 | *AC008443.1* |
| ENSG00000030582 | 17 | 42412614 | 42440470 | 55 | 16 | 38368 | 0.15874 | 0.43694 | *GRN* |
| ENSG00000183423 | 4 | 110759358 | 110803471 | 104 | 14 | 45425 | 0.15871 | 0.43695 | *LRIT3* |
| ENSG00000216895 | 7 | 155393925 | 155447899 | 157 | 22 | 50951 | 0.15864 | 0.43698 | *AC009403.2* |
| ENSG00000075213 | 7 | 83575093 | 84132040 | 2020 | 131 | 43960 | 0.1584 | 0.43707 | *SEMA3A* |
| ENSG00000268977 | 16 | 79005647 | 79025739 | 118 | 35 | 45268 | 0.15839 | 0.43707 | *PIH1* |
| ENSG00000101546 | 18 | 77784358 | 77816397 | 148 | 16 | 44987 | 0.15819 | 0.43715 | *RBFA* |
| ENSG00000143079 | 1 | 112928803 | 113016078 | 216 | 15 | 32113 | 0.15798 | 0.43724 | *CTTNBP2NL* |

| | | | | | | | | | |
|---|---|---|---|---|---|---|---|---|---|
| ENSG00000272790 | 11 | 112031253 | 112081394 | 154 | 19 | 43486 | 0.1579 | 0.43727 | *AP002884.3* |
| ENSG00000123171 | 13 | 52426117 | 52450366 | 75 | 15 | 43581 | 0.15774 | 0.43733 | *CCDC70* |
| ENSG00000177234 | 10 | 122347721 | 122369629 | 93 | 18 | 49265 | 0.15765 | 0.43737 | *C10orf85* |
| ENSG00000108094 | 10 | 35287479 | 35389570 | 274 | 14 | 55140 | 0.15759 | 0.43739 | *CUL2* |
| ENSG00000166183 | 14 | 104542016 | 104589098 | 149 | 28 | 41273 | 0.15735 | 0.43748 | *ASPG* |
| ENSG00000108702 | 17 | 32677347 | 32700250 | 80 | 13 | 54756 | 0.15737 | 0.43748 | *CCL1* |
| ENSG00000198146 | 15 | 35260542 | 35290488 | 87 | 17 | 38408 | 0.15721 | 0.43754 | *ZNF770* |
| ENSG00000183977 | 3 | 20011453 | 20063822 | 195 | 24 | 44065 | 0.1571 | 0.43758 | *PP2D1* |
| ENSG00000011638 | 16 | 21159698 | 21201937 | 117 | 16 | 48604 | 0.15712 | 0.43758 | *TMEM159* |
| ENSG00000132975 | 13 | 27319341 | 27344922 | 100 | 16 | 52211 | 0.15653 | 0.43781 | *GPR12* |
| ENSG00000104835 | 19 | 39395906 | 39450495 | 168 | 23 | 40926 | 0.15625 | 0.43792 | *SARS2* |
| ENSG00000269547 | 19 | 39396044 | 39450495 | 168 | 23 | 40926 | 0.15625 | 0.43792 | *CTC-360G5.8* |
| ENSG00000133874 | 8 | 33395273 | 33434643 | 139 | 15 | 46966 | 0.15591 | 0.43805 | *RNF122* |
| ENSG00000119787 | 2 | 38512022 | 38614427 | 279 | 26 | 43801 | 0.1559 | 0.43806 | *ATL2* |
| ENSG00000144852 | 3 | 119489331 | 119547332 | 184 | 36 | 39872 | 0.15587 | 0.43807 | *NR1I2* |
| ENSG00000074603 | 15 | 65724801 | 65820042 | 261 | 24 | 42013 | 0.15579 | 0.4381 | *DPP8* |
| ENSG00000106009 | 7 | 2567511 | 2605361 | 141 | 22 | 40743 | 0.15566 | 0.43815 | *BRAT1* |
| ENSG00000100227 | 22 | 42969727 | 43020968 | 121 | 19 | 36326 | 0.15546 | 0.43823 | *POLDIP3* |
| ENSG00000113845 | 3 | 119207379 | 119253937 | 162 | 22 | 47381 | 0.15542 | 0.43825 | *TIMMDC1* |
| ENSG00000203880 | 20 | 62877094 | 62936855 | 117 | 21 | 44226 | 0.15535 | 0.43827 | *PCMTD2* |
| ENSG00000115993 | 2 | 202231930 | 202326302 | 180 | 16 | 41745 | 0.15516 | 0.43835 | *TRAK2* |
| ENSG00000121060 | 17 | 54955270 | 55001399 | 58 | 18 | 46302 | 0.15499 | 0.43841 | *TRIM25* |
| ENSG00000159596 | 1 | 46142886 | 46170115 | 60 | 4 | 54877 | 0.15487 | 0.43846 | *TMEM69* |
| ENSG00000183397 | 19 | 3529152 | 3554028 | 81 | 19 | 34765 | 0.15436 | 0.43866 | *C19orf71* |
| ENSG00000119714 | 14 | 91688876 | 91730269 | 115 | 19 | 42084 | 0.15381 | 0.43888 | *GPR68* |
| ENSG00000141527 | 17 | 78133791 | 78193130 | 238 | 46 | 46993 | 0.15379 | 0.43889 | *CARD14* |
| ENSG00000188050 | 7 | 122327766 | 122349210 | 62 | 12 | 52502 | 0.15355 | 0.43898 | *RNF133* |
| ENSG00000011198 | 3 | 43721605 | 43785863 | 200 | 20 | 37346 | 0.15312 | 0.43915 | *ABHD5* |
| ENSG00000107566 | 10 | 101899851 | 101958091 | 123 | 19 | 29932 | 0.1527 | 0.43932 | *ERLIN1* |
| ENSG00000156273 | 21 | 30556392 | 31013071 | 964 | 123 | 39866 | 0.1522 | 0.43951 | *BACH1* |
| ENSG00000066084 | 12 | 50888768 | 51152450 | 418 | 28 | 46615 | 0.15215 | 0.43954 | *DIP2B* |
| ENSG00000146955 | 7 | 140093843 | 140136050 | 170 | 38 | 43136 | 0.15173 | 0.4397 | *RAB19* |
| ENSG00000146276 | 6 | 89877220 | 89950997 | 394 | 49 | 52695 | 0.1516 | 0.43975 | *GABRR1* |
| ENSG00000214844 | 17 | 19031448 | 19051895 | 2 | 1 | 43509 | 0.15152 | 0.43978 | *AC007952.1* |
| ENSG00000178685 | 8 | 145041321 | 145096940 | 102 | 12 | 50024 | 0.15144 | 0.43982 | *PARP10* |
| ENSG00000187824 | 17 | 10592332 | 10643633 | 154 | 6 | 26635 | 0.15136 | 0.43985 | *TMEM220* |

| | | | | | | | | | |
|---|---|---|---|---|---|---|---|---|---|
| ENSG00000188958 | 3 | 190974957 | 191058325 | 365 | 41 | 45390 | 0.15117 | 0.43992 | *UTS2B* |
| ENSG00000100722 | 14 | 89019253 | 89089853 | 149 | 27 | 37627 | 0.15117 | 0.43992 | *ZC3H14* |
| ENSG00000072818 | 17 | 7229848 | 7264797 | 80 | 21 | 46005 | 0.15114 | 0.43993 | *ACAP1* |
| ENSG00000205250 | 16 | 67216072 | 67242821 | 48 | 9 | 32574 | 0.15099 | 0.43999 | *E2F4* |
| ENSG00000161940 | 17 | 6916339 | 6943609 | 55 | 9 | 45948 | 0.15087 | 0.44004 | *BCL6B* |
| ENSG00000120458 | 11 | 124626394 | 124680569 | 121 | 23 | 44904 | 0.15069 | 0.44011 | *MSANTD2* |
| ENSG00000204632 | 6 | 29784744 | 29808902 | 414 | 30 | 33329 | 0.15064 | 0.44013 | *HLA-G* |
| ENSG00000141748 | 17 | 37303147 | 37333737 | 128 | 23 | 48311 | 0.15065 | 0.44013 | *ARL5C* |
| ENSG00000167604 | 19 | 36368555 | 36403205 | 106 | 12 | 28111 | 0.15061 | 0.44014 | *NFKBID* |
| ENSG00000186226 | 1 | 152748690 | 152770902 | 55 | 10 | 37882 | 0.15054 | 0.44017 | *LCE1E* |
| ENSG00000232859 | 17 | 26195340 | 26231778 | 51 | 16 | 48891 | 0.15049 | 0.44019 | *LYRM9* |
| ENSG00000164949 | 8 | 95251481 | 95284578 | 113 | 16 | 47631 | 0.15024 | 0.44029 | *GEM* |
| ENSG00000129354 | 19 | 10673347 | 10707991 | 158 | 19 | 35539 | 0.15023 | 0.44029 | *AP1M2* |
| ENSG00000104611 | 8 | 19161128 | 19263729 | 356 | 61 | 48966 | 0.15006 | 0.44036 | *SH2D4A* |
| ENSG00000117614 | 1 | 25539170 | 25568993 | 22 | 14 | 52505 | 0.15 | 0.44038 | *SYF2* |
| ENSG00000197444 | 10 | 50932689 | 50980425 | 148 | 19 | 38735 | 0.14998 | 0.44039 | *OGDHL* |
| ENSG00000088836 | 20 | 3198063 | 3229836 | 127 | 27 | 46983 | 0.14997 | 0.4404 | *SLC4A11* |
| ENSG00000188827 | 16 | 3621182 | 3671599 | 157 | 12 | 49153 | 0.14969 | 0.44051 | *SLX4* |
| ENSG00000128253 | 22 | 32576422 | 32610718 | 185 | 16 | 36231 | 0.14966 | 0.44052 | *RFPL2* |
| ENSG00000103275 | 16 | 1345548 | 1387019 | 213 | 20 | 47765 | 0.14947 | 0.44059 | *UBE2I* |
| ENSG00000166257 | 11 | 123489895 | 123535952 | 209 | 29 | 52947 | 0.14942 | 0.44061 | *SCN3B* |
| ENSG00000145439 | 4 | 169774921 | 169941426 | 356 | 28 | 40112 | 0.14936 | 0.44063 | *CBR4* |
| ENSG00000094880 | 5 | 137513339 | 137559032 | 78 | 17 | 53438 | 0.14919 | 0.4407 | *CDC23* |
| ENSG00000165905 | 11 | 45933172 | 45960647 | 43 | 16 | 43451 | 0.14893 | 0.44081 | *GYLTL1B* |
| ENSG00000188938 | 9 | 96198776 | 96225874 | 111 | 19 | 50005 | 0.1489 | 0.44082 | *FAM120AOS* |
| ENSG00000266614 | 18 | 73961121 | 73982069 | 56 | 9 | 48300 | 0.14875 | 0.44087 | *RP11-94B19.4* |
| ENSG00000105427 | 19 | 42881171 | 42904439 | 50 | 5 | 41778 | 0.14867 | 0.44091 | *CNFN* |
| ENSG00000125347 | 5 | 131807301 | 131836490 | 104 | 8 | 51461 | 0.14865 | 0.44092 | *IRF1* |
| ENSG00000133678 | 10 | 81828402 | 81862313 | 167 | 24 | 50201 | 0.14863 | 0.44092 | *TMEM254* |
| ENSG00000082929 | 4 | 5516296 | 5539528 | 88 | 14 | 30937 | 0.14836 | 0.44103 | *C4orf6* |
| ENSG00000174428 | 7 | 74498364 | 74575623 | 24 | 13 | 35238 | 0.14829 | 0.44106 | *GTF2IRD2B* |
| ENSG00000113600 | 5 | 39274364 | 39434970 | 541 | 75 | 49136 | 0.14826 | 0.44107 | *C9* |
| ENSG00000179528 | 2 | 74714644 | 74740443 | 14 | 8 | 54141 | 0.14817 | 0.4411 | *LBX2* |
| ENSG00000145425 | 4 | 152010725 | 152035804 | 39 | 11 | 33471 | 0.14813 | 0.44112 | *RPS3A* |
| ENSG00000183134 | 11 | 60608413 | 60633444 | 117 | 25 | 43246 | 0.14806 | 0.44115 | *PTGDR2* |
| ENSG00000108509 | 17 | 4861287 | 4900960 | 117 | 22 | 41531 | 0.14789 | 0.44122 | *CAMTA2* |

| | | | | | | | | | |
|---|---|---|---|---|---|---|---|---|---|
| ENSG00000153446 | 16 | 5084123 | 5126111 | 144 | 37 | 51712 | 0.14776 | 0.44127 | *C16orf89* |
| ENSG00000154227 | 15 | 100930600 | 101095200 | 885 | 73 | 46521 | 0.14761 | 0.44133 | *CERS3* |
| ENSG00000186812 | 18 | 32810994 | 32857097 | 79 | 20 | 38231 | 0.14746 | 0.44138 | *ZNF397* |
| ENSG00000198670 | 6 | 160942515 | 161097407 | 323 | 37 | 44464 | 0.14726 | 0.44146 | *LPA* |
| ENSG00000268310 | 17 | 76200462 | 76230740 | 165 | 21 | 51843 | 0.14681 | 0.44164 | *AC087645.1* |
| ENSG00000174021 | 1 | 84954008 | 84982248 | 126 | 20 | 42076 | 0.14677 | 0.44166 | *GNG5* |
| ENSG00000134330 | 2 | 9603787 | 9646672 | 122 | 13 | 35281 | 0.14669 | 0.44169 | *IAH1* |
| ENSG00000185742 | 11 | 109282846 | 109309840 | 27 | 11 | 46093 | 0.14642 | 0.44179 | *C11orf87* |
| ENSG00000196975 | 2 | 69861557 | 70063596 | 469 | 55 | 44359 | 0.14633 | 0.44183 | *ANXA4* |
| ENSG00000059804 | 12 | 8061826 | 8098871 | 100 | 26 | 47591 | 0.14634 | 0.44183 | *SLC2A3* |
| ENSG00000163386 | 1 | 145279772 | 145380303 | 4 | 2 | 43158 | 0.14631 | 0.44184 | *NBPF10* |
| ENSG00000264058 | 17 | 38775049 | 38831393 | 133 | 17 | 51562 | 0.14629 | 0.44185 | *KRT222* |
| ENSG00000145555 | 5 | 16655395 | 16946372 | 1097 | 166 | 47986 | 0.14615 | 0.4419 | *MYO10* |
| ENSG00000047621 | 12 | 4586894 | 4657674 | 93 | 14 | 47815 | 0.14595 | 0.44198 | *C12orf4* |
| ENSG00000183628 | 22 | 18883541 | 18911751 | 77 | 24 | 44009 | 0.14581 | 0.44204 | *DGCR6* |
| ENSG00000152953 | 4 | 5043169 | 5512725 | 2283 | 184 | 36766 | 0.14574 | 0.44206 | *STK32B* |
| ENSG00000196388 | 17 | 4881425 | 4910905 | 98 | 21 | 41168 | 0.14573 | 0.44207 | *INCA1* |
| ENSG00000249581 | 4 | 17506788 | 17538727 | 128 | 21 | 52504 | 0.14566 | 0.4421 | *CLRN2* |
| ENSG00000081479 | 2 | 169973619 | 170229195 | 878 | 129 | 44395 | 0.14563 | 0.44211 | *LRP2* |
| ENSG00000196611 | 11 | 102650651 | 102678891 | 96 | 24 | 52927 | 0.1454 | 0.4422 | *MMP1* |
| ENSG00000170289 | 8 | 87556205 | 87765903 | 677 | 86 | 43387 | 0.14527 | 0.44225 | *CNGB3* |
| ENSG00000136059 | 3 | 38019550 | 38058679 | 132 | 17 | 50224 | 0.14525 | 0.44226 | *VILL* |
| ENSG00000273274 | 1 | 32920670 | 32972287 | 87 | 17 | 52871 | 0.14493 | 0.44238 | *ZBTB8B* |
| ENSG00000115590 | 2 | 102598306 | 102655006 | 198 | 19 | 36994 | 0.14464 | 0.4425 | *IL1R2* |
| ENSG00000184909 | 1 | 178501950 | 178527579 | 107 | 10 | 39839 | 0.1446 | 0.44251 | *C1ORF220* |
| ENSG00000138316 | 10 | 72422559 | 72532197 | 576 | 82 | 50731 | 0.14452 | 0.44255 | *ADAMTS14* |
| ENSG00000153157 | 6 | 10738027 | 10989553 | 707 | 107 | 45810 | 0.14402 | 0.44274 | *SYCP2L* |
| ENSG00000114698 | 3 | 145900126 | 145978966 | 360 | 31 | 39883 | 0.144 | 0.44275 | *PLSCR4* |
| ENSG00000182177 | 2 | 237092095 | 237183052 | 374 | 33 | 42810 | 0.14398 | 0.44276 | *ASB18* |
| ENSG00000110723 | 11 | 108366158 | 108474465 | 299 | 49 | 40489 | 0.14338 | 0.44299 | *EXPH5* |
| ENSG00000124356 | 2 | 74046086 | 74110786 | 194 | 28 | 39802 | 0.14332 | 0.44302 | *STAMBP* |
| ENSG00000152049 | 2 | 223906532 | 224073117 | 808 | 73 | 52324 | 0.14325 | 0.44305 | *KCNE4* |
| ENSG00000203859 | 1 | 119947554 | 119975658 | 81 | 13 | 28774 | 0.14311 | 0.4431 | *HSD3B2* |
| ENSG00000135222 | 4 | 70810974 | 70836725 | 96 | 13 | 50943 | 0.1431 | 0.4431 | *CSN2* |
| ENSG00000069493 | 12 | 9807565 | 9858413 | 280 | 27 | 44157 | 0.14298 | 0.44315 | *CLEC2D* |
| ENSG00000082397 | 18 | 5382383 | 5640699 | 837 | 123 | 43117 | 0.14277 | 0.44324 | *EPB41L3* |

| | | | | | | | | | |
|---|---|---|---|---|---|---|---|---|---|
| ENSG00000221995 | 17 | 27390537 | 27428537 | 92 | 12 | 34204 | 0.14256 | 0.44332 | *TIAF1* |
| ENSG00000138434 | 2 | 182746560 | 182805465 | 95 | 11 | 42141 | 0.14244 | 0.44337 | *SSFA2* |
| ENSG00000186230 | 19 | 57936697 | 57966853 | 93 | 22 | 49531 | 0.14226 | 0.44344 | *ZNF749* |
| ENSG00000187026 | 21 | 32109112 | 32129551 | 48 | 4 | 20790 | 0.14193 | 0.44357 | *KRTAP21-2* |
| ENSG00000100028 | 22 | 24941471 | 25015947 | 183 | 33 | 37972 | 0.14175 | 0.44364 | *SNRPD3* |
| ENSG00000189190 | 19 | 53257448 | 53300044 | 198 | 47 | 41299 | 0.14173 | 0.44365 | *ZNF600* |
| ENSG00000125378 | 14 | 54406454 | 54435479 | 80 | 22 | 43747 | 0.14166 | 0.44367 | *BMP4* |
| ENSG00000155760 | 2 | 202889310 | 202913160 | 70 | 12 | 47685 | 0.1415 | 0.44374 | *FZD7* |
| ENSG00000115325 | 2 | 74766153 | 74794681 | 25 | 7 | 36053 | 0.1413 | 0.44382 | *DOK1* |
| ENSG00000161929 | 17 | 5102256 | 5148155 | 136 | 25 | 41725 | 0.14127 | 0.44383 | *SCIMP* |
| ENSG00000143621 | 1 | 153624512 | 153653524 | 53 | 8 | 44408 | 0.14116 | 0.44387 | *ILF2* |
| ENSG00000146809 | 7 | 123197064 | 123287951 | 225 | 29 | 53827 | 0.14104 | 0.44392 | *ASB15* |
| ENSG00000101236 | 20 | 3897956 | 4006229 | 316 | 31 | 48563 | 0.14078 | 0.44402 | *RNF24* |
| ENSG00000182621 | 20 | 8102824 | 8959003 | 2950 | 288 | 47189 | 0.14075 | 0.44404 | *PLCB1* |
| ENSG00000188554 | 17 | 41312498 | 41373708 | 122 | 7 | 50938 | 0.14071 | 0.44405 | *NBR1* |
| ENSG00000148296 | 9 | 136187552 | 136213235 | 82 | 14 | 50470 | 0.14056 | 0.44411 | *SURF6* |
| ENSG00000174667 | 19 | 9314526 | 9335542 | 63 | 12 | 30334 | 0.14028 | 0.44422 | *OR7D4* |
| ENSG00000066136 | 1 | 41147320 | 41247275 | 308 | 28 | 41437 | 0.14019 | 0.44425 | *NFYC* |
| ENSG00000197409 | 6 | 26187068 | 26209521 | 125 | 12 | 50406 | 0.14019 | 0.44425 | *HIST1H3D* |
| ENSG00000140299 | 15 | 59941345 | 59991733 | 213 | 11 | 41773 | 0.14007 | 0.4443 | *BNIP2* |
| ENSG00000130520 | 19 | 18407040 | 18444084 | 98 | 20 | 47727 | 0.13999 | 0.44433 | *LSM4* |
| ENSG00000250127 | 5 | 57827394 | 57864070 | 120 | 19 | 36663 | 0.1399 | 0.44437 | *CTD-2117L12.1* |
| ENSG00000104524 | 8 | 144676083 | 144701943 | 107 | 20 | 36616 | 0.1399 | 0.44437 | *PYCRL* |
| ENSG00000170689 | 17 | 46688518 | 46713839 | 86 | 20 | 34550 | 0.13991 | 0.44437 | *HOXB9* |
| ENSG00000170458 | 5 | 140001313 | 140023286 | 71 | 9 | 53568 | 0.13981 | 0.4444 | *CD14* |
| ENSG00000184497 | 13 | 114452216 | 114524926 | 282 | 34 | 42419 | 0.13968 | 0.44446 | *TMEM255B* |
| ENSG00000156599 | 11 | 57425219 | 57478659 | 96 | 16 | 44532 | 0.13965 | 0.44447 | *ZDHHC5* |
| ENSG00000204488 | 1 | 13485254 | 13508260 | 1 | 1 | 44611 | 0.13957 | 0.4445 | *PRAMEF16* |
| ENSG00000135077 | 5 | 156502843 | 156579880 | 277 | 29 | 49959 | 0.13938 | 0.44458 | *HAVCR2* |
| ENSG00000203711 | 6 | 159280971 | 159341385 | 241 | 40 | 32923 | 0.13937 | 0.44458 | *C6orf99* |
| ENSG00000169992 | 17 | 7298193 | 7333179 | 78 | 31 | 45019 | 0.1392 | 0.44465 | *NLGN2* |
| ENSG00000144848 | 3 | 112241356 | 112290893 | 156 | 13 | 49036 | 0.13918 | 0.44466 | *ATG3* |
| ENSG00000229665 | 13 | 57718195 | 57741216 | 1 | 1 | 43388 | 0.13906 | 0.4447 | *PRR20C* |
| ENSG00000104826 | 19 | 49509237 | 49530338 | 100 | 12 | 50613 | 0.13861 | 0.44488 | *LHB* |
| ENSG00000269831 | 1 | 728532 | 749137 | 12 | 7 | 34598 | 0.13858 | 0.44489 | *AL669831.1* |
| ENSG00000144749 | 3 | 66419221 | 66561687 | 298 | 50 | 43765 | 0.13845 | 0.44494 | *LRIG1* |

| | | | | | | | | | |
|---|---|---|---|---|---|---|---|---|---|
| ENSG00000109670 | 4 | 153232410 | 153467253 | 523 | 22 | 50352 | 0.13801 | 0.44512 | *FBXW7* |
| ENSG00000198142 | 2 | 110361911 | 110386563 | 71 | 14 | 47686 | 0.13794 | 0.44515 | *SOWAHC* |
| ENSG00000171475 | 17 | 38365556 | 38450388 | 171 | 18 | 51039 | 0.13764 | 0.44526 | *WIPF2* |
| ENSG00000065548 | 2 | 187340883 | 187384090 | 101 | 13 | 52265 | 0.13762 | 0.44527 | *ZC3H15* |
| ENSG00000131795 | 1 | 145497598 | 145523536 | 9 | 5 | 43920 | 0.13756 | 0.44529 | *RBM8A* |
| ENSG00000256394 | 4 | 156740881 | 156797425 | 148 | 32 | 42653 | 0.13756 | 0.44529 | *ASIC5* |
| ENSG00000174292 | 17 | 7273853 | 7303093 | 95 | 23 | 48277 | 0.13754 | 0.4453 | *TNK1* |
| ENSG00000174145 | 4 | 37235842 | 37461087 | 788 | 118 | 48806 | 0.13753 | 0.44531 | *KIAA1239* |
| ENSG00000213057 | 1 | 178501887 | 178528024 | 109 | 10 | 39756 | 0.13698 | 0.44552 | *C1orf220* |
| ENSG00000182040 | 17 | 72902176 | 72929351 | 28 | 15 | 44094 | 0.137 | 0.44552 | *USH1G* |
| ENSG00000111605 | 12 | 69623317 | 69678138 | 137 | 17 | 47218 | 0.13677 | 0.44561 | *CPSF6* |
| ENSG00000141040 | 17 | 16444701 | 16482520 | 90 | 14 | 41692 | 0.13651 | 0.44571 | *ZNF287* |
| ENSG00000130590 | 20 | 62595466 | 62621361 | 79 | 12 | 45934 | 0.1364 | 0.44575 | *SAMD10* |
| ENSG00000170180 | 4 | 145020457 | 145071904 | 277 | 27 | 47403 | 0.13636 | 0.44577 | *GYPA* |
| ENSG00000197846 | 6 | 26189748 | 26210942 | 124 | 12 | 52140 | 0.13617 | 0.44584 | *HIST1H2BF* |
| ENSG00000178809 | 7 | 75014337 | 75050279 | 31 | 11 | 41532 | 0.13605 | 0.44589 | *TRIM73* |
| ENSG00000079263 | 2 | 231057826 | 231233762 | 616 | 67 | 45461 | 0.136 | 0.44591 | *SP140* |
| ENSG00000107719 | 10 | 72228577 | 72338205 | 454 | 67 | 43999 | 0.13598 | 0.44592 | *PALD1* |
| ENSG00000075891 | 10 | 102485360 | 102599698 | 270 | 40 | 46815 | 0.13589 | 0.44595 | *PAX2* |
| ENSG00000205436 | 14 | 103556481 | 103586896 | 83 | 16 | 47692 | 0.13588 | 0.44596 | *EXOC3L4* |
| ENSG00000103599 | 15 | 67537138 | 67804598 | 534 | 42 | 50442 | 0.1358 | 0.44599 | *IQCH* |
| ENSG00000106686 | 9 | 4543386 | 4676674 | 620 | 79 | 32651 | 0.13571 | 0.44603 | *SPATA6L* |
| ENSG00000169035 | 19 | 51469729 | 51497355 | 94 | 26 | 43527 | 0.13548 | 0.44612 | *KLK7* |
| ENSG00000197561 | 19 | 841014 | 866242 | 84 | 32 | 40302 | 0.13514 | 0.44625 | *ELANE* |
| ENSG00000157353 | 16 | 70478324 | 70524177 | 49 | 21 | 47655 | 0.13469 | 0.44643 | *FUK* |
| ENSG00000114784 | 3 | 40341175 | 40363915 | 55 | 5 | 52975 | 0.13452 | 0.44649 | *EIF1B* |
| ENSG00000213759 | 4 | 70055669 | 70090449 | 195 | 22 | 43854 | 0.13442 | 0.44654 | *UGT2B11* |
| ENSG00000108947 | 17 | 7598520 | 7624696 | 66 | 17 | 37521 | 0.13417 | 0.44663 | *EFNB3* |
| ENSG00000172728 | 8 | 33218342 | 33340940 | 417 | 37 | 45857 | 0.13402 | 0.44669 | *FUT10* |
| ENSG00000135457 | 12 | 51477446 | 51576926 | 307 | 29 | 43575 | 0.13381 | 0.44678 | *TFCP2* |
| ENSG00000162804 | 2 | 241928255 | 242044983 | 331 | 60 | 41766 | 0.13363 | 0.44685 | *SNED1* |
| ENSG00000269104 | 22 | 41675388 | 41695686 | 42 | 9 | 52661 | 0.13335 | 0.44696 | *AL035681.1* |
| ENSG00000159842 | 17 | 896758 | 1142315 | 1076 | 178 | 45979 | 0.13319 | 0.44702 | *ABR* |
| ENSG00000169738 | 17 | 79983012 | 80005608 | 77 | 10 | 29026 | 0.1332 | 0.44702 | *DCXR* |
| ENSG00000187634 | 1 | 850260 | 889955 | 123 | 27 | 41287 | 0.13226 | 0.44739 | *SAMD11* |
| ENSG00000020256 | 20 | 50658202 | 50830847 | 709 | 81 | 49244 | 0.13145 | 0.44771 | *ZFP64* |

| | | | | | | | | | |
|---|---|---|---|---|---|---|---|---|---|
| ENSG00000196866 | 6 | 26189079 | 26209471 | 120 | 12 | 51218 | 0.13144 | 0.44772 | *HIST1H2AD* |
| ENSG00000100116 | 22 | 38193912 | 38223183 | 80 | 12 | 49236 | 0.13142 | 0.44772 | *GCAT* |
| ENSG00000153107 | 2 | 112513848 | 112652267 | 397 | 34 | 48946 | 0.13137 | 0.44774 | *ANAPC1* |
| ENSG00000104907 | 19 | 13205716 | 13238381 | 72 | 26 | 47845 | 0.1313 | 0.44777 | *TRMT1* |
| ENSG00000105880 | 7 | 96639704 | 96664409 | 62 | 20 | 54059 | 0.13122 | 0.4478 | *DLX5* |
| ENSG00000120217 | 9 | 5440503 | 5480566 | 136 | 27 | 45684 | 0.13073 | 0.44799 | *CD274* |
| ENSG00000167850 | 17 | 72527247 | 72552282 | 119 | 17 | 45012 | 0.13075 | 0.44799 | *CD300C* |
| ENSG00000112139 | 6 | 37590284 | 37677082 | 400 | 76 | 37909 | 0.13006 | 0.44826 | *MDGA1* |
| ENSG00000176485 | 11 | 63330667 | 63394355 | 112 | 30 | 44115 | 0.13004 | 0.44827 | *PLA2G16* |
| ENSG00000196724 | 19 | 58423252 | 58456761 | 79 | 15 | 47684 | 0.13004 | 0.44827 | *ZNF418* |
| ENSG00000145014 | 3 | 194298402 | 194364418 | 394 | 46 | 40512 | 0.12991 | 0.44832 | *TMEM44* |
| ENSG00000109270 | 4 | 100789493 | 100825647 | 67 | 16 | 44299 | 0.12988 | 0.44833 | *LAMTOR3* |
| ENSG00000143553 | 1 | 153621130 | 153644306 | 39 | 6 | 47738 | 0.12968 | 0.44841 | *SNAPIN* |
| ENSG00000236027 | 11 | 125648006 | 125671495 | 84 | 11 | 51198 | 0.12966 | 0.44842 | *PATE3* |
| ENSG00000101558 | 18 | 9903999 | 9970018 | 224 | 37 | 52405 | 0.12959 | 0.44845 | *VAPA* |
| ENSG00000151650 | 10 | 135040908 | 135065433 | 105 | 21 | 46447 | 0.12941 | 0.44852 | *VENTX* |
| ENSG00000144713 | 3 | 12865984 | 12893087 | 93 | 11 | 33118 | 0.12866 | 0.44881 | *RPL32* |
| ENSG00000169067 | 5 | 56765846 | 56788636 | 71 | 14 | 49957 | 0.12863 | 0.44882 | *ACTBL2* |
| ENSG00000175581 | 11 | 73488361 | 73586178 | 368 | 27 | 48880 | 0.12857 | 0.44885 | *MRPL48* |
| ENSG00000128829 | 15 | 40216347 | 40337797 | 353 | 46 | 39188 | 0.12857 | 0.44885 | *EIF2AK4* |
| ENSG00000267140 | 18 | 33013835 | 33087904 | 160 | 29 | 53816 | 0.12853 | 0.44887 | *RP11-322E11.6* |
| ENSG00000240654 | 13 | 24871304 | 24906673 | 169 | 25 | 44486 | 0.12834 | 0.44894 | *C1QTNF9* |
| ENSG00000203668 | 1 | 241782155 | 241809232 | 99 | 21 | 53133 | 0.12831 | 0.44895 | *CHML* |
| ENSG00000044115 | 5 | 137936656 | 138280723 | 696 | 35 | 52438 | 0.128 | 0.44908 | *CTNNA1* |
| ENSG00000103769 | 15 | 66008392 | 66194329 | 393 | 35 | 38470 | 0.12793 | 0.4491 | *RAB11A* |
| ENSG00000179580 | 16 | 2006824 | 2028976 | 155 | 19 | 51756 | 0.12788 | 0.44912 | *RNF151* |
| ENSG00000182552 | 4 | 184550788 | 184590378 | 165 | 37 | 44712 | 0.12784 | 0.44914 | *RWDD4* |
| ENSG00000145358 | 4 | 101097027 | 101121939 | 123 | 24 | 54322 | 0.12768 | 0.4492 | *DDIT4L* |
| ENSG00000119681 | 14 | 74954873 | 75089306 | 338 | 50 | 39725 | 0.12755 | 0.44925 | *LTBP2* |
| ENSG00000196081 | 19 | 23394401 | 23443192 | 138 | 10 | 43902 | 0.12723 | 0.44938 | *ZNF724P* |
| ENSG00000167264 | 16 | 68011649 | 68123223 | 183 | 22 | 47493 | 0.1271 | 0.44943 | *DUS2* |
| ENSG00000119938 | 10 | 93378199 | 93402811 | 53 | 14 | 45702 | 0.12691 | 0.44951 | *PPP1R3C* |
| ENSG00000206203 | 22 | 19108321 | 19130134 | 68 | 12 | 52416 | 0.1269 | 0.44951 | *TSSK2* |
| ENSG00000232671 | 1 | 151242500 | 151264405 | 30 | 9 | 53014 | 0.1267 | 0.44959 | *RP11-126K1.2* |
| ENSG00000183780 | 1 | 234030679 | 234470262 | 1421 | 175 | 44543 | 0.12636 | 0.44973 | *SLC35F3* |
| ENSG00000144233 | 2 | 128609204 | 128653496 | 108 | 21 | 34290 | 0.12605 | 0.44985 | *AMMECR1L* |

| | | | | | | | | | |
|---|---|---|---|---|---|---|---|---|---|
| ENSG00000171596 | 2 | 232377871 | 232405206 | 124 | 13 | 55296 | 0.12602 | 0.44986 | *NMUR1* |
| ENSG00000155868 | 5 | 156554423 | 156596030 | 133 | 16 | 43625 | 0.12595 | 0.44989 | *MED7* |
| ENSG00000253117 | 8 | 133026467 | 133081627 | 205 | 30 | 50520 | 0.12593 | 0.44989 | *OC90* |
| ENSG00000171564 | 4 | 155474108 | 155502238 | 83 | 10 | 54200 | 0.12535 | 0.45012 | *FGB* |
| ENSG00000204560 | 6 | 30610896 | 30650814 | 112 | 14 | 27750 | 0.12524 | 0.45017 | *DHX16* |
| ENSG00000171396 | 17 | 39305910 | 39326983 | 33 | 8 | 36463 | 0.125 | 0.45026 | *KRTAP4-4* |
| ENSG00000163918 | 3 | 186497669 | 186534847 | 84 | 17 | 50547 | 0.1248 | 0.45034 | *RFC4* |
| ENSG00000121314 | 12 | 10948650 | 10969892 | 37 | 9 | 40259 | 0.12445 | 0.45048 | *TAS2R8* |
| ENSG00000121211 | 4 | 154255801 | 154346270 | 319 | 30 | 38878 | 0.12443 | 0.45049 | *MND1* |
| ENSG00000187783 | 10 | 45396648 | 45442450 | 130 | 22 | 50586 | 0.12411 | 0.45061 | *TMEM72* |
| ENSG00000158865 | 16 | 24847162 | 24932949 | 275 | 48 | 41621 | 0.12401 | 0.45065 | *SLC5A11* |
| ENSG00000146457 | 6 | 160136617 | 160187351 | 163 | 19 | 42374 | 0.1239 | 0.4507 | *WTAP* |
| ENSG00000099998 | 22 | 24605622 | 24651110 | 203 | 38 | 43172 | 0.12366 | 0.45079 | *GGT5* |
| ENSG00000167377 | 16 | 71471500 | 71506998 | 83 | 15 | 37942 | 0.12334 | 0.45092 | *ZNF23* |
| ENSG00000196437 | 19 | 37892057 | 37970177 | 163 | 25 | 37069 | 0.12333 | 0.45092 | *ZNF569* |
| ENSG00000167612 | 12 | 52271744 | 52295448 | 100 | 25 | 45701 | 0.1232 | 0.45097 | *ANKRD33* |
| ENSG00000083520 | 13 | 73319540 | 73366234 | 119 | 23 | 50622 | 0.12285 | 0.45111 | *DIS3* |
| ENSG00000103089 | 16 | 74736853 | 74818729 | 329 | 55 | 47281 | 0.1227 | 0.45117 | *FA2H* |
| ENSG00000205362 | 16 | 56662578 | 56683999 | 120 | 20 | 40957 | 0.12265 | 0.45119 | *MT1A* |
| ENSG00000143252 | 1 | 161274047 | 161342984 | 360 | 14 | 24989 | 0.12255 | 0.45123 | *SDHC* |
| ENSG00000168004 | 11 | 63218876 | 63268666 | 106 | 24 | 34600 | 0.12256 | 0.45123 | *HRASLS5* |
| ENSG00000112624 | 6 | 42739765 | 42846298 | 217 | 26 | 39653 | 0.12249 | 0.45126 | *GLTSCR1L* |
| ENSG00000256040 | 9 | 119150439 | 119172885 | 60 | 12 | 55810 | 0.12237 | 0.4513 | *PAPPA-AS1* |
| ENSG00000177981 | 12 | 48531571 | 48584996 | 148 | 20 | 50163 | 0.12236 | 0.45131 | *ASB8* |
| ENSG00000130560 | 9 | 138814815 | 138863226 | 227 | 30 | 49302 | 0.12222 | 0.45136 | *UBAC1* |
| ENSG00000163823 | 3 | 46233200 | 46259887 | 68 | 7 | 25447 | 0.12206 | 0.45143 | *CCR1* |
| ENSG00000136457 | 17 | 48531857 | 48556327 | 55 | 11 | 37856 | 0.12202 | 0.45144 | *CHAD* |
| ENSG00000163666 | 3 | 57221944 | 57270549 | 71 | 13 | 41949 | 0.12172 | 0.45156 | *HESX1* |
| ENSG00000197948 | 5 | 141008869 | 141040986 | 104 | 24 | 50919 | 0.12169 | 0.45157 | *FCHSD1* |
| ENSG00000125845 | 20 | 6738311 | 6770927 | 91 | 33 | 49584 | 0.12144 | 0.45167 | *BMP2* |
| ENSG00000015133 | 14 | 91727667 | 91894188 | 563 | 103 | 49163 | 0.12142 | 0.45168 | *CCDC88C* |
| ENSG00000124243 | 20 | 49401431 | 49503714 | 372 | 53 | 38371 | 0.12119 | 0.45177 | *BCAS4* |
| ENSG00000105993 | 7 | 157118075 | 157220133 | 447 | 49 | 42243 | 0.12111 | 0.4518 | *DNAJB6* |
| ENSG00000089685 | 17 | 76200267 | 76231717 | 170 | 22 | 51400 | 0.1211 | 0.45181 | *BIRC5* |
| ENSG00000189319 | 10 | 126297861 | 126442838 | 344 | 47 | 48150 | 0.121 | 0.45184 | *FAM53B* |
| ENSG00000156976 | 3 | 186490994 | 186517689 | 61 | 14 | 51831 | 0.12096 | 0.45186 | *EIF4A2* |

| | | | | | | | | | |
|---|---|---|---|---|---|---|---|---|---|
| ENSG00000119431 | 9 | 116125699 | 116149279 | 129 | 30 | 43330 | 0.12096 | 0.45186 | *HDHD3* |
| ENSG00000084070 | 1 | 40800522 | 40898998 | 280 | 51 | 47241 | 0.1209 | 0.45189 | *SMAP2* |
| ENSG00000239511 | 22 | 21467050 | 21492352 | 2 | 1 | 42797 | 0.12086 | 0.4519 | *POM121L7* |
| ENSG00000135999 | 2 | 149392009 | 149555130 | 403 | 24 | 38189 | 0.12082 | 0.45192 | *EPC2* |
| ENSG00000258588 | 11 | 5607955 | 5675628 | 337 | 64 | 43645 | 0.12078 | 0.45193 | *TRIM6-TRIM34* |
| ENSG00000170683 | 11 | 7950081 | 7971141 | 104 | 17 | 51182 | 0.12048 | 0.45205 | *OR10A3* |
| ENSG00000197245 | 1 | 26475511 | 26499119 | 74 | 17 | 40186 | 0.12031 | 0.45212 | *FAM110D* |
| ENSG00000148468 | 10 | 15243642 | 15423061 | 684 | 82 | 45951 | 0.11991 | 0.45228 | *FAM171A1* |
| ENSG00000204389 | 6 | 31773291 | 31795723 | 57 | 22 | 36800 | 0.11989 | 0.45229 | *HSPA1A* |
| ENSG00000225940 | 5 | 55797394 | 55912059 | 510 | 80 | 42333 | 0.11985 | 0.4523 | *AC022431.2* |
| ENSG00000183496 | 15 | 82324119 | 82348482 | 73 | 21 | 37647 | 0.11966 | 0.45238 | *MEX3B* |
| ENSG00000125249 | 13 | 98076476 | 98131382 | 94 | 15 | 46576 | 0.11929 | 0.45252 | *RAP2A* |
| ENSG00000166479 | 18 | 66330925 | 66392535 | 154 | 19 | 37141 | 0.11922 | 0.45255 | *TMX3* |
| ENSG00000164935 | 8 | 105341315 | 105378917 | 133 | 26 | 41458 | 0.11873 | 0.45275 | *DCSTAMP* |
| ENSG00000156711 | 6 | 36085586 | 36117842 | 62 | 16 | 43654 | 0.11865 | 0.45278 | *MAPK13* |
| ENSG00000206527 | 3 | 123199667 | 123314032 | 322 | 32 | 38566 | 0.11843 | 0.45286 | *PTPLB* |
| ENSG00000172795 | 5 | 112302399 | 112366667 | 217 | 29 | 47846 | 0.1182 | 0.45295 | *DCP2* |
| ENSG00000205002 | 8 | 117940438 | 117966726 | 54 | 17 | 49971 | 0.11818 | 0.45296 | *AARD* |
| ENSG00000092036 | 14 | 23405437 | 23436370 | 59 | 8 | 31906 | 0.11785 | 0.45309 | *HAUS4* |
| ENSG00000168621 | 5 | 37802779 | 37849788 | 157 | 31 | 45108 | 0.11776 | 0.45313 | *GDNF* |
| ENSG00000042304 | 2 | 228464806 | 228508036 | 376 | 38 | 46247 | 0.1177 | 0.45315 | *C2orf83* |
| ENSG00000085449 | 2 | 224710433 | 224820104 | 286 | 20 | 37488 | 0.11733 | 0.4533 | *WDFY1* |
| ENSG00000118972 | 12 | 4467393 | 4498894 | 88 | 17 | 44304 | 0.11732 | 0.4533 | *FGF23* |
| ENSG00000234602 | 5 | 54505442 | 54533143 | 91 | 10 | 56000 | 0.1171 | 0.45339 | *MCIDAS* |
| ENSG00000172819 | 12 | 53594354 | 53636764 | 85 | 26 | 34521 | 0.11685 | 0.45349 | *RARG* |
| ENSG00000151287 | 13 | 103408340 | 103436161 | 81 | 16 | 39070 | 0.11683 | 0.4535 | *TEX30* |
| ENSG00000090861 | 16 | 70276198 | 70333446 | 55 | 25 | 41532 | 0.1166 | 0.45359 | *AARS* |
| ENSG00000065882 | 4 | 37882708 | 38150796 | 951 | 137 | 46992 | 0.11657 | 0.4536 | *TBC1D1* |
| ENSG00000178342 | 18 | 77613668 | 77670184 | 207 | 27 | 37816 | 0.11654 | 0.45361 | *KCNG2* |
| ENSG00000121377 | 12 | 10944131 | 10965226 | 36 | 8 | 38129 | 0.11652 | 0.45362 | *TAS2R7* |
| ENSG00000100372 | 22 | 41155634 | 41225403 | 115 | 17 | 45414 | 0.11643 | 0.45366 | *SLC25A17* |
| ENSG00000186807 | 10 | 47736936 | 47773041 | 10 | 7 | 18783 | 0.11608 | 0.45379 | *ANXA8L2* |
| ENSG00000215033 | 10 | 47736962 | 47780871 | 10 | 7 | 18783 | 0.11608 | 0.45379 | *AL603965.1* |
| ENSG00000113638 | 5 | 40704577 | 40766077 | 135 | 16 | 49771 | 0.11598 | 0.45383 | *TTC33* |
| ENSG00000078618 | 1 | 52244863 | 52354477 | 245 | 37 | 44499 | 0.11512 | 0.45418 | *NRD1* |
| ENSG00000120068 | 17 | 46678739 | 46702478 | 77 | 19 | 36319 | 0.11509 | 0.45419 | *HOXB8* |

| | | | | | | | | | |
|---|---|---|---|---|---|---|---|---|---|
| ENSG00000166199 | 11 | 43892361 | 43951816 | 257 | 28 | 46542 | 0.115 | 0.45422 | *ALKBH3* |
| ENSG00000255307 | 11 | 6180560 | 6201638 | 72 | 16 | 24296 | 0.11489 | 0.45427 | *OR52B2* |
| ENSG00000187533 | 4 | 70989333 | 71052516 | 214 | 32 | 46176 | 0.11469 | 0.45435 | *C4orf40* |
| ENSG00000032444 | 19 | 7588890 | 7636650 | 259 | 33 | 46850 | 0.11468 | 0.45435 | *PNPLA6* |
| ENSG00000138069 | 2 | 65287835 | 65367240 | 278 | 30 | 53220 | 0.11456 | 0.4544 | *RAB1A* |
| ENSG00000163626 | 4 | 73911797 | 73945472 | 32 | 9 | 45941 | 0.11443 | 0.45445 | *COX18* |
| ENSG00000085117 | 11 | 44575977 | 44651913 | 316 | 58 | 48786 | 0.11438 | 0.45447 | *CD82* |
| ENSG00000081721 | 1 | 161709548 | 161737028 | 118 | 19 | 40593 | 0.1141 | 0.45458 | *DUSP12* |
| ENSG00000186648 | 14 | 24511206 | 24548937 | 81 | 33 | 46595 | 0.1141 | 0.45458 | *LRRC16B* |
| ENSG00000141084 | 16 | 67747005 | 67850555 | 140 | 23 | 39002 | 0.11384 | 0.45468 | *RANBP10* |
| ENSG00000159110 | 21 | 34592206 | 34647980 | 189 | 22 | 52997 | 0.11374 | 0.45472 | *IFNAR2* |
| ENSG00000170209 | 11 | 113248513 | 113281140 | 94 | 14 | 33691 | 0.11357 | 0.45479 | *ANKK1* |
| ENSG00000112357 | 6 | 137133717 | 137245075 | 303 | 28 | 51853 | 0.11346 | 0.45483 | *PEX7* |
| ENSG00000157570 | 11 | 44738015 | 44963972 | 683 | 119 | 40432 | 0.11346 | 0.45483 | *TSPAN18* |
| ENSG00000182827 | 1 | 226322380 | 226384431 | 164 | 22 | 54951 | 0.1132 | 0.45494 | *ACBD3* |
| ENSG00000116824 | 1 | 117287007 | 117321850 | 57 | 14 | 37148 | 0.11317 | 0.45495 | *CD2* |
| ENSG00000169926 | 15 | 31609058 | 31737868 | 354 | 59 | 48941 | 0.11308 | 0.45498 | *KLF13* |
| ENSG00000148187 | 9 | 125016882 | 125095743 | 89 | 24 | 48297 | 0.11304 | 0.455 | *MRRF* |
| ENSG00000101347 | 20 | 35508632 | 35590246 | 181 | 24 | 41613 | 0.11278 | 0.4551 | *SAMHD1* |
| ENSG00000183024 | 17 | 3019878 | 3040875 | 88 | 12 | 27312 | 0.11268 | 0.45514 | *OR1G1* |
| ENSG00000099866 | 19 | 479176 | 515347 | 163 | 35 | 34250 | 0.1126 | 0.45517 | *MADCAM1* |
| ENSG00000153029 | 1 | 180993067 | 181041074 | 175 | 33 | 44806 | 0.11242 | 0.45525 | *MR1* |
| ENSG00000099381 | 16 | 30958615 | 31006437 | 62 | 9 | 30688 | 0.11235 | 0.45527 | *SETD1A* |
| ENSG00000069667 | 15 | 60770483 | 61531518 | 2213 | 274 | 44370 | 0.11225 | 0.45531 | *RORA* |
| ENSG00000167136 | 9 | 131570753 | 131594956 | 95 | 11 | 39594 | 0.11186 | 0.45547 | *ENDOG* |
| ENSG00000106113 | 7 | 30682200 | 30749745 | 176 | 46 | 41924 | 0.11176 | 0.45551 | *CRHR2* |
| ENSG00000100296 | 22 | 29891868 | 29961205 | 199 | 20 | 47494 | 0.11173 | 0.45552 | *THOC5* |
| ENSG00000227268 | 10 | 89608918 | 89633194 | 41 | 11 | 53781 | 0.11135 | 0.45567 | *KLLN* |
| ENSG00000157350 | 16 | 70403338 | 70483140 | 95 | 34 | 47014 | 0.11116 | 0.45575 | *ST3GAL2* |
| ENSG00000125149 | 16 | 67133861 | 67192442 | 61 | 18 | 48133 | 0.11107 | 0.45578 | *C16orf70* |
| ENSG00000254106 | 5 | 109208883 | 109230452 | 80 | 16 | 39795 | 0.11101 | 0.4558 | *AC011366.3* |
| ENSG00000162236 | 11 | 62564369 | 62609560 | 110 | 11 | 35228 | 0.11059 | 0.45597 | *STX5* |
| ENSG00000136937 | 9 | 100385908 | 100446030 | 71 | 20 | 48527 | 0.11027 | 0.4561 | *NCBP1* |
| ENSG00000243627 | 21 | 35781013 | 35806264 | 33 | 12 | 44001 | 0.11007 | 0.45618 | *AP000322.53* |
| ENSG00000198752 | 14 | 103388716 | 103533799 | 380 | 58 | 42491 | 0.10989 | 0.45625 | *CDC42BPB* |
| ENSG00000111218 | 12 | 3480515 | 3713139 | 618 | 124 | 45922 | 0.10985 | 0.45627 | *PRMT8* |

| | | | | | | | | | |
|---|---|---|---|---|---|---|---|---|---|
| ENSG00000185989 | 13 | 114737194 | 114908086 | 747 | 65 | 38759 | 0.10966 | 0.45634 | *RASA3* |
| ENSG00000185924 | 17 | 1827971 | 1938639 | 444 | 72 | 47369 | 0.10945 | 0.45642 | *RTN4RL1* |
| ENSG00000204311 | 2 | 179306163 | 179336117 | 47 | 12 | 37589 | 0.10939 | 0.45645 | *DFNB59* |
| ENSG00000113073 | 5 | 139729787 | 139764728 | 44 | 18 | 53203 | 0.10936 | 0.45646 | *SLC4A9* |
| ENSG00000143751 | 1 | 226160403 | 226197032 | 54 | 14 | 45261 | 0.10926 | 0.4565 | *SDE2* |
| ENSG00000135048 | 9 | 74288282 | 74441606 | 533 | 43 | 47481 | 0.10902 | 0.45659 | *TMEM2* |
| ENSG00000148143 | 9 | 109615378 | 109785915 | 293 | 57 | 44446 | 0.10902 | 0.45659 | *ZNF462* |
| ENSG00000168267 | 10 | 23471256 | 23493181 | 33 | 11 | 47319 | 0.10879 | 0.45669 | *PTF1A* |
| ENSG00000134323 | 2 | 16070686 | 16097129 | 81 | 15 | 38524 | 0.10865 | 0.45674 | *MYCN* |
| ENSG00000197261 | 6 | 49508113 | 49539620 | 128 | 17 | 39700 | 0.10819 | 0.45692 | *C6orf141* |
| ENSG00000110148 | 11 | 6270966 | 6303357 | 118 | 24 | 38142 | 0.10814 | 0.45694 | *CCKBR* |
| ENSG00000128918 | 15 | 58235622 | 58800065 | 2162 | 245 | 43593 | 0.10812 | 0.45695 | *ALDH1A2* |
| ENSG00000164393 | 6 | 47614172 | 47675533 | 299 | 9 | 56039 | 0.10802 | 0.45699 | *GPR111* |
| ENSG00000163734 | 4 | 74892306 | 74914524 | 11 | 6 | 48056 | 0.10782 | 0.45707 | *CXCL3* |
| ENSG00000089902 | 14 | 103048998 | 103206913 | 483 | 37 | 47795 | 0.10782 | 0.45707 | *RCOR1* |
| ENSG00000164620 | 5 | 141006517 | 141030644 | 75 | 19 | 53400 | 0.10772 | 0.45711 | *RELL2* |
| ENSG00000148832 | 10 | 135182695 | 135215198 | 125 | 22 | 35461 | 0.10763 | 0.45714 | *PAOX* |
| ENSG00000235034 | 19 | 51142702 | 51172567 | 95 | 27 | 46388 | 0.1076 | 0.45716 | *C19orf81* |
| ENSG00000163082 | 2 | 223279236 | 223435667 | 464 | 75 | 46872 | 0.10749 | 0.4572 | *SGPP2* |
| ENSG00000137267 | 6 | 3143903 | 3167760 | 82 | 8 | 28347 | 0.10744 | 0.45722 | *TUBB2A* |
| ENSG00000226976 | 6 | 37002607 | 37023177 | 101 | 12 | 51988 | 0.10701 | 0.45739 | *COX6A1P2* |
| ENSG00000228567 | 19 | 53759929 | 53780972 | 106 | 18 | 53656 | 0.10683 | 0.45746 | *VN1R4* |
| ENSG00000180259 | 20 | 4701928 | 4731314 | 174 | 18 | 51771 | 0.10672 | 0.45751 | *PRNT* |
| ENSG00000267987 | 9 | 124093625 | 124120410 | 90 | 18 | 37118 | 0.10647 | 0.4576 | *AL161784.1* |
| ENSG00000188523 | 9 | 135275430 | 135458704 | 588 | 118 | 47264 | 0.10635 | 0.45765 | *C9orf171* |
| ENSG00000119688 | 14 | 74742126 | 74779759 | 100 | 17 | 38564 | 0.10627 | 0.45768 | *ABCD4* |
| ENSG00000145354 | 4 | 103780135 | 103820399 | 79 | 5 | 39228 | 0.10623 | 0.4577 | *CISD2* |
| ENSG00000100629 | 14 | 80933330 | 81435861 | 1781 | 127 | 43323 | 0.10609 | 0.45775 | *CEP128* |
| ENSG00000123159 | 19 | 14578572 | 14616944 | 175 | 31 | 43097 | 0.10596 | 0.45781 | *GIPC1* |
| ENSG00000136861 | 9 | 123141147 | 123352448 | 324 | 42 | 42813 | 0.10588 | 0.45784 | *CDK5RAP2* |
| ENSG00000236338 | 7 | 135601542 | 135623214 | 76 | 14 | 39384 | 0.10564 | 0.45793 | *AC015987.2* |
| ENSG00000145147 | 4 | 20244883 | 20632184 | 1131 | 168 | 49005 | 0.10559 | 0.45795 | *SLIT2* |
| ENSG00000227184 | 8 | 144929497 | 144962632 | 79 | 28 | 42518 | 0.10534 | 0.45805 | *EPPK1* |
| ENSG00000269117 | 20 | 31813091 | 31833387 | 55 | 5 | 38464 | 0.10517 | 0.45812 | *AL121901.1* |
| ENSG00000124935 | 11 | 61999682 | 62022280 | 61 | 14 | 38461 | 0.10487 | 0.45824 | *SCGB1D2* |
| ENSG00000144834 | 3 | 111707511 | 111742734 | 190 | 30 | 47685 | 0.10467 | 0.45832 | *TAGLN3* |

| | | | | | | | | | |
|---|---|---|---|---|---|---|---|---|---|
| ENSG00000165632 | 10 | 7850467 | 8068590 | 782 | 73 | 47462 | 0.10454 | 0.45837 | *TAF3* |
| ENSG00000100364 | 22 | 45576219 | 45646650 | 315 | 46 | 48511 | 0.10423 | 0.45849 | *KIAA0930* |
| ENSG00000164086 | 3 | 52072935 | 52100566 | 36 | 8 | 30560 | 0.10421 | 0.4585 | *DUSP7* |
| ENSG00000151503 | 11 | 134010014 | 134105348 | 237 | 36 | 48129 | 0.10383 | 0.45865 | *NCAPD3* |
| ENSG00000144535 | 2 | 232815955 | 233219060 | 1072 | 53 | 40465 | 0.1037 | 0.4587 | *DIS3L2* |
| ENSG00000079308 | 2 | 218654512 | 218877718 | 628 | 123 | 38238 | 0.10353 | 0.45877 | *TNS1* |
| ENSG00000185069 | 12 | 53151939 | 53181129 | 111 | 22 | 35632 | 0.10349 | 0.45879 | *KRT76* |
| ENSG00000225683 | 6 | 163576446 | 163622840 | 179 | 35 | 47393 | 0.10333 | 0.45885 | *AL078585.1* |
| ENSG00000109089 | 17 | 72973727 | 73011895 | 95 | 18 | 35849 | 0.10333 | 0.45885 | *CDR2L* |
| ENSG00000146112 | 6 | 30634166 | 30665672 | 131 | 19 | 31625 | 0.10307 | 0.45895 | *PPP1R18* |
| ENSG00000176024 | 19 | 52420400 | 52462012 | 162 | 18 | 32892 | 0.10301 | 0.45898 | *ZNF613* |
| ENSG00000196453 | 7 | 149118454 | 149168214 | 94 | 23 | 48039 | 0.10296 | 0.459 | *ZNF777* |
| ENSG00000186395 | 17 | 38964369 | 38988847 | 78 | 18 | 42973 | 0.10278 | 0.45907 | *KRT10* |
| ENSG00000170482 | 5 | 138692885 | 138730242 | 42 | 11 | 35904 | 0.10273 | 0.45909 | *SLC23A1* |
| ENSG00000132561 | 8 | 98871068 | 99058944 | 592 | 79 | 40587 | 0.10267 | 0.45911 | *MATN2* |
| ENSG00000172260 | 1 | 71851623 | 72758417 | 2039 | 156 | 42750 | 0.10266 | 0.45912 | *NEGR1* |
| ENSG00000189164 | 19 | 37851963 | 37893968 | 77 | 16 | 34751 | 0.10266 | 0.45912 | *ZNF527* |
| ENSG00000152804 | 10 | 94437945 | 94465403 | 62 | 13 | 53776 | 0.1023 | 0.45926 | *HHEX* |
| ENSG00000100628 | 14 | 94390499 | 94453137 | 235 | 48 | 43261 | 0.10183 | 0.45945 | *ASB2* |
| ENSG00000111596 | 12 | 70626774 | 70758773 | 412 | 32 | 52618 | 0.10179 | 0.45946 | *CNOT2* |
| ENSG00000188895 | 17 | 38268551 | 38303042 | 77 | 15 | 40797 | 0.10175 | 0.45948 | *MSL1* |
| ENSG00000154928 | 3 | 134306643 | 134989309 | 2236 | 189 | 42202 | 0.10167 | 0.45951 | *EPHB1* |
| ENSG00000176273 | 10 | 95643730 | 95725819 | 144 | 14 | 44576 | 0.10168 | 0.45951 | *SLC35G1* |
| ENSG00000092871 | 17 | 33323009 | 33426338 | 103 | 23 | 41366 | 0.10135 | 0.45964 | *RFFL* |
| ENSG00000013503 | 12 | 106741436 | 106913976 | 481 | 36 | 41926 | 0.10096 | 0.45979 | *POLR3B* |
| ENSG00000161905 | 17 | 4524197 | 4555589 | 85 | 27 | 38483 | 0.10095 | 0.45979 | *ALOX15* |
| ENSG00000175664 | 13 | 31496840 | 31559639 | 193 | 31 | 50380 | 0.10089 | 0.45982 | *TEX26* |
| ENSG00000183941 | 1 | 149794221 | 149821339 | 13 | 5 | 44906 | 0.10076 | 0.45987 | *HIST2H4A* |
| ENSG00000205821 | 17 | 2300279 | 2328731 | 75 | 20 | 38366 | 0.10077 | 0.45987 | *AC006435.1* |
| ENSG00000184140 | 15 | 102335902 | 102356876 | 53 | 10 | 41119 | 0.10027 | 0.46007 | *OR4F6* |
| ENSG00000106823 | 9 | 95246365 | 95308937 | 180 | 18 | 36563 | 0.10013 | 0.46012 | *ECM2* |
| ENSG00000203995 | 1 | 53298183 | 53370670 | 175 | 16 | 26585 | 0.10012 | 0.46013 | *ZYG11A* |
| ENSG00000139364 | 12 | 125661382 | 126156917 | 1634 | 223 | 47976 | 0.10007 | 0.46014 | *TMEM132B* |
| ENSG00000198959 | 20 | 36746863 | 36804980 | 165 | 17 | 32639 | 0.1 | 0.46017 | *TGM2* |
| ENSG00000161835 | 12 | 52390724 | 52419673 | 69 | 24 | 53169 | 0.09991 | 0.46021 | *GRASP* |
| ENSG00000109118 | 17 | 27222268 | 27288789 | 74 | 14 | 50119 | 0.099889 | 0.46022 | *PHF12* |

| | | | | | | | | | |
|---|---|---|---|---|---|---|---|---|---|
| ENSG00000197748 | 10 | 105879646 | 106002120 | 316 | 34 | 39630 | 0.099796 | 0.46025 | *WDR96* |
| ENSG00000116771 | 1 | 15888848 | 15921605 | 163 | 14 | 38017 | 0.099768 | 0.46026 | *AGMAT* |
| ENSG00000155087 | 8 | 103553800 | 103583245 | 164 | 28 | 52420 | 0.099612 | 0.46033 | *ODF1* |
| ENSG00000198821 | 1 | 167389877 | 167497847 | 378 | 84 | 43659 | 0.099483 | 0.46038 | *CD247* |
| ENSG00000198374 | 6 | 27823034 | 27843606 | 54 | 13 | 34273 | 0.099409 | 0.46041 | *HIST1H2AL* |
| ENSG00000145912 | 5 | 177566461 | 177590968 | 88 | 14 | 53218 | 0.099284 | 0.46046 | *NHP2* |
| ENSG00000065809 | 10 | 14550556 | 14826896 | 1184 | 152 | 49602 | 0.098791 | 0.46065 | *FAM107B* |
| ENSG00000099194 | 10 | 102096881 | 102134591 | 95 | 18 | 50210 | 0.0987 | 0.46069 | *SCD* |
| ENSG00000159337 | 15 | 42349207 | 42396752 | 223 | 21 | 53539 | 0.098671 | 0.4607 | *PLA2G4D* |
| ENSG00000206260 | 3 | 138712804 | 138735110 | 46 | 12 | 46946 | 0.098477 | 0.46078 | *PRR23A* |
| ENSG00000116127 | 2 | 73602886 | 73847920 | 537 | 28 | 33342 | 0.098276 | 0.46086 | *ALMS1* |
| ENSG00000176261 | 1 | 33055773 | 33126504 | 186 | 18 | 36959 | 0.098132 | 0.46091 | *ZBTB8OS* |
| ENSG00000197428 | 11 | 4650945 | 4672068 | 135 | 23 | 36751 | 0.098004 | 0.46096 | *OR51D1* |
| ENSG00000101216 | 20 | 62208955 | 62268394 | 231 | 29 | 41753 | 0.097788 | 0.46105 | *GMEB2* |
| ENSG00000168283 | 10 | 22600140 | 22630413 | 22 | 11 | 43181 | 0.097729 | 0.46107 | *BMI1* |
| ENSG00000116750 | 1 | 192971380 | 193039237 | 109 | 16 | 52333 | 0.097532 | 0.46115 | *UCHL5* |
| ENSG00000149308 | 11 | 108017942 | 108103369 | 144 | 16 | 51408 | 0.09743 | 0.46119 | *NPAT* |
| ENSG00000243729 | 6 | 29313007 | 29409744 | 440 | 47 | 38606 | 0.097259 | 0.46126 | *OR5V1* |
| ENSG00000154127 | 11 | 122516383 | 122695181 | 626 | 104 | 46261 | 0.096959 | 0.46138 | *UBASH3B* |
| ENSG00000133943 | 14 | 91516677 | 91701976 | 491 | 79 | 44609 | 0.096906 | 0.4614 | *C14orf159* |
| ENSG00000140416 | 15 | 63324831 | 63374114 | 198 | 28 | 47266 | 0.096911 | 0.4614 | *TPM1* |
| ENSG00000140688 | 16 | 31490792 | 31530630 | 64 | 9 | 35457 | 0.096889 | 0.46141 | *C16orf58* |
| ENSG00000172244 | 5 | 43476803 | 43525247 | 151 | 19 | 34758 | 0.096701 | 0.46148 | *C5orf34* |
| ENSG00000187695 | 3 | 128618717 | 128700173 | 312 | 31 | 35534 | 0.096633 | 0.46151 | *RP11-723O4.6* |
| ENSG00000028203 | 12 | 95601522 | 95706566 | 384 | 38 | 47001 | 0.096581 | 0.46153 | *VEZT* |
| ENSG00000255798 | 11 | 102542041 | 102564644 | 115 | 21 | 47957 | 0.096535 | 0.46155 | *RP11-817J15.3* |
| ENSG00000204501 | 1 | 13411176 | 13438191 | 1 | 1 | 42859 | 0.096404 | 0.4616 | *PRAMEF9* |
| ENSG00000129152 | 11 | 17731115 | 17753678 | 67 | 17 | 49776 | 0.096137 | 0.46171 | *MYOD1* |
| ENSG00000187391 | 7 | 77636393 | 79092890 | 5805 | 448 | 45183 | 0.096069 | 0.46173 | *MAGI2* |
| ENSG00000136045 | 12 | 108069509 | 108116944 | 118 | 16 | 39067 | 0.096011 | 0.46176 | *PWP1* |
| ENSG00000107020 | 9 | 5347973 | 5447878 | 379 | 49 | 40447 | 0.095858 | 0.46182 | *PLGRKT* |
| ENSG00000162694 | 1 | 101327943 | 101371554 | 53 | 13 | 44276 | 0.095812 | 0.46183 | *EXTL2* |
| ENSG00000099957 | 22 | 21354097 | 21393119 | 155 | 30 | 41988 | 0.095804 | 0.46184 | *P2RX6* |
| ENSG00000163811 | 2 | 29107509 | 29181088 | 267 | 16 | 44256 | 0.095259 | 0.46205 | *WDR43* |
| ENSG00000196531 | 12 | 57096212 | 57135412 | 79 | 14 | 49321 | 0.095221 | 0.46207 | *NACA* |
| ENSG00000163577 | 3 | 170596204 | 170636482 | 89 | 15 | 51128 | 0.094842 | 0.46222 | *EIF5A2* |

| | | | | | | | | | |
|---|---|---|---|---|---|---|---|---|---|
| ENSG00000185896 | 13 | 113941556 | 113987987 | 35 | 21 | 49983 | 0.09484 | 0.46222 | *LAMP1* |
| ENSG00000141101 | 16 | 69765770 | 69798843 | 80 | 18 | 44807 | 0.09466 | 0.46229 | *NOB1* |
| ENSG00000106004 | 7 | 27170671 | 27193287 | 55 | 12 | 35388 | 0.09451 | 0.46235 | *HOXA5* |
| ENSG00000106336 | 7 | 100171605 | 100208740 | 57 | 16 | 49683 | 0.094494 | 0.46236 | *FBXO24* |
| ENSG00000107164 | 9 | 133444352 | 133523739 | 210 | 34 | 42340 | 0.094431 | 0.46238 | *FUBP3* |
| ENSG00000136810 | 9 | 112996091 | 113028920 | 135 | 26 | 48486 | 0.094384 | 0.4624 | *TXN* |
| ENSG00000005243 | 17 | 46093533 | 46125392 | 105 | 20 | 46491 | 0.094305 | 0.46243 | *COPZ2* |
| ENSG00000167861 | 17 | 72936838 | 72979261 | 83 | 28 | 41072 | 0.094264 | 0.46245 | *HID1* |
| ENSG00000135218 | 7 | 79988891 | 80318593 | 990 | 107 | 37775 | 0.094221 | 0.46247 | *CD36* |
| ENSG00000198765 | 1 | 115387424 | 115547991 | 414 | 33 | 45902 | 0.094143 | 0.4625 | *SYCP1* |
| ENSG00000164744 | 7 | 48016745 | 48078716 | 167 | 16 | 49486 | 0.094104 | 0.46251 | *SUN3* |
| ENSG00000115282 | 2 | 74700200 | 74732013 | 21 | 14 | 51031 | 0.093973 | 0.46257 | *TTC31* |
| ENSG00000049249 | 1 | 7969907 | 8010926 | 57 | 16 | 38069 | 0.093734 | 0.46266 | *TNFRSF9* |
| ENSG00000008226 | 3 | 38070696 | 38175516 | 309 | 27 | 47682 | 0.093724 | 0.46266 | *DLEC1* |
| ENSG00000165997 | 10 | 18938334 | 18980568 | 146 | 21 | 54933 | 0.093672 | 0.46268 | *ARL5B* |
| ENSG00000204669 | 9 | 74656292 | 74697733 | 127 | 22 | 38808 | 0.093535 | 0.46274 | *C9orf57* |
| ENSG00000204983 | 7 | 142447319 | 142470923 | 42 | 9 | 33099 | 0.093436 | 0.46278 | *PRSS1* |
| ENSG00000182572 | 6 | 27829623 | 27850099 | 59 | 13 | 36254 | 0.093406 | 0.46279 | *HIST1H3I* |
| ENSG00000255835 | 1 | 226064396 | 226121978 | 196 | 32 | 43163 | 0.09324 | 0.46286 | *RP4-559A3.7* |
| ENSG00000105926 | 7 | 24602887 | 24743812 | 521 | 25 | 47705 | 0.093241 | 0.46286 | *MPP6* |
| ENSG00000163081 | 2 | 223152866 | 223179936 | 49 | 20 | 51164 | 0.093158 | 0.46289 | *CCDC140* |
| ENSG00000005175 | 12 | 48047070 | 48109844 | 146 | 21 | 50655 | 0.093125 | 0.4629 | *RPAP3* |
| ENSG00000132437 | 7 | 50516134 | 50643154 | 674 | 55 | 48565 | 0.092998 | 0.46295 | *DDC* |
| ENSG00000164253 | 5 | 76711795 | 76926436 | 908 | 78 | 38745 | 0.09299 | 0.46296 | *WDR41* |
| ENSG00000188846 | 3 | 40488783 | 40513861 | 77 | 10 | 43352 | 0.092827 | 0.46302 | *RPL14* |
| ENSG00000265590 | 21 | 33941132 | 33994591 | 181 | 24 | 48897 | 0.092752 | 0.46305 | *AP000275.65* |
| ENSG00000230185 | 9 | 115186096 | 115259484 | 267 | 15 | 54646 | 0.092509 | 0.46315 | *C9orf147* |
| ENSG00000171843 | 9 | 20331663 | 20632542 | 523 | 85 | 46857 | 0.092478 | 0.46316 | *MLLT3* |
| ENSG00000247626 | 2 | 198560087 | 198583113 | 59 | 8 | 52329 | 0.092261 | 0.46325 | *MARS2* |
| ENSG00000206538 | 3 | 86977119 | 87050269 | 102 | 21 | 48588 | 0.092171 | 0.46328 | *VGLL3* |
| ENSG00000163440 | 4 | 56412692 | 56468379 | 226 | 21 | 51924 | 0.091805 | 0.46343 | *PDCL2* |
| ENSG00000133111 | 13 | 37383361 | 37413241 | 37 | 12 | 40332 | 0.091707 | 0.46347 | *RFXAP* |
| ENSG00000227500 | 19 | 1895213 | 1936016 | 142 | 27 | 36825 | 0.09151 | 0.46354 | *SCAMP4* |
| ENSG00000180834 | 3 | 183523664 | 183553382 | 94 | 12 | 52416 | 0.091386 | 0.46359 | *MAP6D1* |
| ENSG00000089116 | 12 | 113889839 | 113920085 | 73 | 9 | 55486 | 0.091226 | 0.46366 | *LHX5* |
| ENSG00000258659 | 11 | 5630994 | 5675628 | 200 | 33 | 40862 | 0.091111 | 0.4637 | *TRIM34* |

| | | | | | | | | | |
|---|---|---|---|---|---|---|---|---|---|
| ENSG00000142449 | 19 | 8120286 | 8224730 | 572 | 90 | 41852 | 0.09099 | 0.46375 | *FBN3* |
| ENSG00000214216 | 3 | 158670717 | 158994096 | 761 | 83 | 42499 | 0.090917 | 0.46378 | *IQCJ* |
| ENSG00000161547 | 17 | 74720197 | 74743456 | 15 | 7 | 45319 | 0.090602 | 0.4639 | *SRSF2* |
| ENSG00000116138 | 1 | 15843308 | 15928874 | 366 | 21 | 43435 | 0.090182 | 0.46407 | *DNAJC16* |
| ENSG00000162695 | 1 | 101351632 | 101457309 | 181 | 21 | 53373 | 0.090123 | 0.46409 | *SLC30A7* |
| ENSG00000151470 | 4 | 130004472 | 130047795 | 155 | 16 | 39663 | 0.089606 | 0.4643 | *C4orf33* |
| ENSG00000135046 | 9 | 75756673 | 75795309 | 114 | 25 | 38754 | 0.089563 | 0.46432 | *ANXA1* |
| ENSG00000167874 | 17 | 7748383 | 7769417 | 50 | 14 | 37721 | 0.089478 | 0.46435 | *TMEM88* |
| ENSG00000125952 | 14 | 65462892 | 65579413 | 304 | 43 | 44535 | 0.089332 | 0.46441 | *MAX* |
| ENSG00000112981 | 5 | 137440866 | 137485132 | 80 | 17 | 53919 | 0.089187 | 0.46447 | *NME5* |
| ENSG00000205560 | 22 | 50997290 | 51027899 | 156 | 27 | 28960 | 0.088917 | 0.46457 | *CPT1B* |
| ENSG00000144566 | 3 | 19978571 | 20036667 | 244 | 27 | 41948 | 0.088805 | 0.46462 | *RAB5A* |
| ENSG00000156885 | 16 | 31429052 | 31449967 | 28 | 9 | 50384 | 0.088496 | 0.46474 | *COX6A2* |
| ENSG00000174844 | 3 | 57317727 | 57540071 | 915 | 55 | 40931 | 0.088299 | 0.46482 | *DNAH12* |
| ENSG00000143318 | 1 | 160150285 | 160181676 | 76 | 24 | 41973 | 0.088148 | 0.46488 | *CASQ1* |
| ENSG00000188760 | 2 | 220398385 | 220425317 | 84 | 13 | 45537 | 0.088011 | 0.46493 | *TMEM198* |
| ENSG00000127252 | 3 | 192948914 | 193005527 | 160 | 19 | 48018 | 0.087986 | 0.46494 | *HRASLS* |
| ENSG00000138035 | 2 | 55851400 | 55931045 | 278 | 33 | 39707 | 0.087977 | 0.46495 | *PNPT1* |
| ENSG00000133703 | 12 | 25347723 | 25413870 | 249 | 29 | 49238 | 0.08791 | 0.46497 | *KRAS* |
| ENSG00000125831 | 20 | 23421040 | 23443494 | 72 | 12 | 26418 | 0.087682 | 0.46506 | *CST11* |
| ENSG00000146477 | 6 | 160759300 | 160886014 | 311 | 34 | 51237 | 0.087504 | 0.46514 | *SLC22A3* |
| ENSG00000112992 | 5 | 43592794 | 43717507 | 370 | 23 | 45655 | 0.087454 | 0.46516 | *NNT* |
| ENSG00000149273 | 11 | 75100530 | 75143324 | 84 | 18 | 37759 | 0.087445 | 0.46516 | *RPS3* |
| ENSG00000107833 | 10 | 103531082 | 103553170 | 18 | 8 | 49074 | 0.087236 | 0.46524 | *NPM3* |
| ENSG00000158864 | 1 | 161156894 | 161194185 | 92 | 24 | 53663 | 0.086459 | 0.46555 | *NDUFS2* |
| ENSG00000110801 | 12 | 122316637 | 122366203 | 150 | 21 | 34753 | 0.086022 | 0.46572 | *PSMD9* |
| ENSG00000167842 | 17 | 5379605 | 5404134 | 115 | 11 | 29375 | 0.085813 | 0.46581 | *MIS12* |
| ENSG00000166670 | 11 | 102631234 | 102661359 | 119 | 25 | 54319 | 0.085725 | 0.46584 | *MMP10* |
| ENSG00000115289 | 2 | 74722170 | 74745707 | 11 | 5 | 57353 | 0.085608 | 0.46589 | *PCGF1* |
| ENSG00000214305 | 11 | 107640219 | 107662316 | 107 | 16 | 52424 | 0.085558 | 0.46591 | *AP001024.1* |
| ENSG00000198680 | 9 | 25666396 | 25688856 | 111 | 18 | 48585 | 0.085527 | 0.46592 | *TUSC1* |
| ENSG00000132356 | 5 | 40749481 | 40808476 | 136 | 19 | 50436 | 0.084597 | 0.46629 | *PRKAA1* |
| ENSG00000188522 | 17 | 18862102 | 18918117 | 103 | 30 | 36283 | 0.084363 | 0.46638 | *FAM83G* |
| ENSG00000109881 | 11 | 27342374 | 27395415 | 161 | 26 | 46214 | 0.084175 | 0.46646 | *CCDC34* |
| ENSG00000119801 | 2 | 30359807 | 30393399 | 53 | 15 | 45562 | 0.084069 | 0.4665 | *YPEL5* |
| ENSG00000070950 | 3 | 8807088 | 9015457 | 937 | 71 | 41298 | 0.084072 | 0.4665 | *RAD18* |

| | | | | | | | | | |
|---|---|---|---|---|---|---|---|---|---|
| ENSG00000198083 | 17 | 39401636 | 39422616 | 63 | 7 | 37075 | 0.084027 | 0.46652 | *KRTAP9-9* |
| ENSG00000005075 | 7 | 102103565 | 102129354 | 78 | 12 | 38754 | 0.083975 | 0.46654 | *POLR2J* |
| ENSG00000100288 | 22 | 51007378 | 51049884 | 214 | 36 | 32454 | 0.083966 | 0.46654 | *CHKB* |
| ENSG00000251606 | 5 | 96414779 | 96529354 | 310 | 56 | 45159 | 0.083432 | 0.46675 | *CTD-2215E18.1* |
| ENSG00000166863 | 12 | 57393784 | 57432667 | 82 | 11 | 53617 | 0.08336 | 0.46678 | *TAC3* |
| ENSG00000100600 | 14 | 93160152 | 93225047 | 173 | 43 | 42640 | 0.0833 | 0.46681 | *LGMN* |
| ENSG00000268194 | 2 | 121211911 | 121233697 | 76 | 20 | 38806 | 0.0829 | 0.46697 | *LINC01101* |
| ENSG00000170577 | 2 | 45222300 | 45246569 | 83 | 12 | 44723 | 0.082693 | 0.46705 | *SIX2* |
| ENSG00000123838 | 1 | 207267607 | 207328317 | 147 | 27 | 32696 | 0.082443 | 0.46715 | *C4BPA* |
| ENSG00000129167 | 11 | 18029111 | 18073973 | 89 | 11 | 41936 | 0.082042 | 0.46731 | *TPH1* |
| ENSG00000136928 | 9 | 101040391 | 101481479 | 1837 | 200 | 44712 | 0.081779 | 0.46741 | *GABBR2* |
| ENSG00000225614 | 16 | 88483879 | 88517165 | 104 | 31 | 45138 | 0.081726 | 0.46743 | *ZNF469* |
| ENSG00000135213 | 7 | 75036066 | 75125548 | 232 | 33 | 47005 | 0.081711 | 0.46744 | *POM121C* |
| ENSG00000140274 | 15 | 45396519 | 45420619 | 39 | 13 | 41836 | 0.081691 | 0.46745 | *DUOXA2* |
| ENSG00000171942 | 19 | 15828834 | 15849862 | 108 | 14 | 49713 | 0.081352 | 0.46758 | *OR10H2* |
| ENSG00000144468 | 2 | 227690297 | 227873931 | 412 | 27 | 41597 | 0.081306 | 0.4676 | *RHBDD1* |
| ENSG00000186174 | 11 | 118754584 | 118806317 | 127 | 31 | 44549 | 0.080874 | 0.46777 | *BCL9L* |
| ENSG00000148482 | 10 | 18230768 | 18342221 | 527 | 73 | 45611 | 0.080825 | 0.46779 | *SLC39A12* |
| ENSG00000180089 | 19 | 55728007 | 55751647 | 92 | 19 | 51986 | 0.080771 | 0.46781 | *TMEM86B* |
| ENSG00000269808 | 19 | 55728587 | 55751647 | 92 | 19 | 51986 | 0.080771 | 0.46781 | *AC010327.2* |
| ENSG00000268367 | 12 | 57800198 | 57820536 | 25 | 7 | 42498 | 0.080622 | 0.46787 | *AC126614.1* |
| ENSG00000171453 | 6 | 43467440 | 43507323 | 72 | 11 | 43558 | 0.080575 | 0.46789 | *POLR1C* |
| ENSG00000272047 | 6 | 158579384 | 158630376 | 151 | 20 | 45855 | 0.080458 | 0.46794 | *GTF2H5* |
| ENSG00000167680 | 19 | 4532600 | 4569820 | 108 | 28 | 46147 | 0.080215 | 0.46803 | *SEMA6B* |
| ENSG00000118777 | 4 | 89001416 | 89162474 | 525 | 65 | 42086 | 0.079939 | 0.46814 | *ABCG2* |
| ENSG00000267618 | 17 | 33328368 | 33458512 | 151 | 37 | 42514 | 0.079886 | 0.46816 | *RAD51L3-RFFL* |
| ENSG00000203818 | 1 | 149390131 | 149410542 | 1 | 1 | 42884 | 0.079544 | 0.4683 | *HIST2H3PS2* |
| ENSG00000100298 | 22 | 39483229 | 39510072 | 183 | 18 | 32177 | 0.079519 | 0.46831 | *APOBEC3H* |
| ENSG00000186020 | 19 | 37015676 | 37106178 | 234 | 27 | 53120 | 0.078978 | 0.46852 | *ZNF529* |
| ENSG00000162961 | 2 | 32082878 | 32274881 | 504 | 41 | 54536 | 0.07888 | 0.46856 | *DPY30* |
| ENSG00000145536 | 5 | 5130443 | 5330417 | 939 | 165 | 46456 | 0.078872 | 0.46857 | *ADAMTS16* |
| ENSG00000152583 | 4 | 88384487 | 88462213 | 299 | 28 | 36653 | 0.078767 | 0.46861 | *SPARCL1* |
| ENSG00000268387 | 1 | 161180160 | 161202296 | 62 | 17 | 50417 | 0.078673 | 0.46865 | *AL590714.1* |
| ENSG00000162419 | 1 | 28985244 | 29055865 | 119 | 13 | 34948 | 0.078611 | 0.46867 | *GMEB1* |
| ENSG00000175857 | 5 | 57777262 | 57802917 | 107 | 13 | 41705 | 0.078469 | 0.46873 | *GAPT* |
| ENSG00000251012 | 3 | 118856222 | 118916654 | 287 | 44 | 49106 | 0.078122 | 0.46887 | *RP11-484M3.5* |

| | | | | | | | | | |
|---|---|---|---|---|---|---|---|---|---|
| ENSG00000264472 | 18 | 66557018 | 66579492 | 91 | 11 | 45034 | 0.077982 | 0.46892 | *RP11-861L17.3* |
| ENSG00000171180 | 1 | 248392231 | 248413166 | 36 | 8 | 50022 | 0.077824 | 0.46898 | *OR2M4* |
| ENSG00000164011 | 1 | 43302280 | 43328148 | 44 | 10 | 52854 | 0.077816 | 0.46899 | *ZNF691* |
| ENSG00000115966 | 2 | 175926978 | 176043110 | 325 | 22 | 46467 | 0.077816 | 0.46899 | *ATF2* |
| ENSG00000116704 | 1 | 67455015 | 67529782 | 265 | 26 | 38306 | 0.07751 | 0.46911 | *SLC35D1* |
| ENSG00000196498 | 12 | 124798961 | 125062135 | 914 | 191 | 44072 | 0.077442 | 0.46914 | *NCOR2* |
| ENSG00000184166 | 17 | 2985352 | 3006290 | 62 | 12 | 37128 | 0.077212 | 0.46923 | *OR1D2* |
| ENSG00000135373 | 11 | 34632640 | 34692604 | 155 | 39 | 37853 | 0.077193 | 0.46924 | *EHF* |
| ENSG00000144199 | 2 | 97739320 | 97770619 | 20 | 11 | 29856 | 0.076893 | 0.46935 | *FAHD2B* |
| ENSG00000111801 | 6 | 26430700 | 26463643 | 110 | 18 | 39926 | 0.076778 | 0.4694 | *BTN3A3* |
| ENSG00000105127 | 19 | 15454196 | 15500609 | 84 | 12 | 53860 | 0.076789 | 0.4694 | *AKAP8* |
| ENSG00000175137 | 1 | 249094648 | 249130832 | 53 | 17 | 37975 | 0.076185 | 0.46964 | *SH3BP5L* |
| ENSG00000145451 | 4 | 175548065 | 175760465 | 911 | 113 | 49981 | 0.076087 | 0.46967 | *GLRA3* |
| ENSG00000185215 | 14 | 103579779 | 103613776 | 108 | 17 | 49913 | 0.076105 | 0.46967 | *TNFAIP2* |
| ENSG00000112167 | 6 | 39061840 | 39092965 | 89 | 23 | 39710 | 0.075981 | 0.46972 | *SAYSD1* |
| ENSG00000103429 | 16 | 14716672 | 14773093 | 62 | 20 | 55117 | 0.075584 | 0.46988 | *BFAR* |
| ENSG00000081277 | 1 | 201242580 | 201312121 | 292 | 31 | 34152 | 0.075543 | 0.46989 | *PKP1* |
| ENSG00000120664 | 13 | 36910568 | 36953738 | 145 | 20 | 54236 | 0.075366 | 0.46996 | *SPG20OS* |
| ENSG00000171368 | 5 | 650883 | 703510 | 215 | 34 | 44541 | 0.07532 | 0.46998 | *TPPP* |
| ENSG00000142798 | 1 | 22138738 | 22273790 | 475 | 70 | 47216 | 0.075229 | 0.47002 | *HSPG2* |
| ENSG00000162490 | 1 | 11741786 | 11795914 | 142 | 39 | 45892 | 0.074919 | 0.47014 | *DRAXIN* |
| ENSG00000198189 | 4 | 88247762 | 88322538 | 263 | 33 | 47489 | 0.074788 | 0.47019 | *HSD17B11* |
| ENSG00000143337 | 1 | 179841177 | 179904135 | 177 | 20 | 50060 | 0.074628 | 0.47026 | *TOR1AIP1* |
| ENSG00000150594 | 10 | 112826790 | 112850658 | 47 | 14 | 44940 | 0.074471 | 0.47032 | *ADRA2A* |
| ENSG00000198518 | 6 | 26194858 | 26216266 | 124 | 13 | 51552 | 0.074251 | 0.47041 | *HIST1H4E* |
| ENSG00000186187 | 16 | 75022928 | 75154892 | 311 | 40 | 37393 | 0.074015 | 0.4705 | *ZNRF1* |
| ENSG00000106617 | 7 | 151243197 | 151584210 | 1223 | 187 | 43728 | 0.073834 | 0.47057 | *PRKAG2* |
| ENSG00000072849 | 17 | 5364571 | 5400131 | 142 | 15 | 32445 | 0.073764 | 0.4706 | *DERL2* |
| ENSG00000197763 | 3 | 126280622 | 126383998 | 323 | 41 | 43795 | 0.073415 | 0.47074 | *TXNRD3* |
| ENSG00000155666 | 16 | 27204807 | 27243089 | 87 | 19 | 47691 | 0.073302 | 0.47078 | *KDM8* |
| ENSG00000170017 | 3 | 105075753 | 105305744 | 814 | 73 | 52314 | 0.073262 | 0.4708 | *ALCAM* |
| ENSG00000253719 | 12 | 74921551 | 74945223 | 101 | 12 | 35758 | 0.073243 | 0.47081 | *ATXN7L3B* |
| ENSG00000183955 | 12 | 123858320 | 123903905 | 152 | 22 | 44510 | 0.072989 | 0.47091 | *SETD8* |
| ENSG00000139620 | 12 | 49037184 | 49086021 | 131 | 18 | 45002 | 0.07269 | 0.47103 | *KANSL2* |
| ENSG00000197273 | 1 | 42618362 | 42640389 | 64 | 17 | 47710 | 0.072644 | 0.47104 | *GUCA2A* |
| ENSG00000130717 | 9 | 134389188 | 134416655 | 88 | 15 | 33368 | 0.072478 | 0.47111 | *UCK1* |

| | | | | | | | | | |
|---|---|---|---|---|---|---|---|---|---|
| ENSG00000183833 | 3 | 119411869 | 119495949 | 233 | 29 | 47700 | 0.072364 | 0.47116 | *MAATS1* |
| ENSG00000174839 | 3 | 57601184 | 57688816 | 239 | 26 | 45711 | 0.072242 | 0.4712 | *DENND6A* |
| ENSG00000165914 | 14 | 90996932 | 91292823 | 1087 | 123 | 46203 | 0.072221 | 0.47121 | *TTC7B* |
| ENSG00000106952 | 9 | 117646003 | 117702697 | 145 | 13 | 39267 | 0.072021 | 0.47129 | *TNFSF8* |
| ENSG00000158473 | 1 | 158139737 | 158164686 | 103 | 9 | 37050 | 0.071949 | 0.47132 | *CD1D* |
| ENSG00000091879 | 8 | 6347172 | 6430930 | 524 | 63 | 43939 | 0.07195 | 0.47132 | *ANGPT2* |
| ENSG00000129451 | 19 | 51505995 | 51533431 | 78 | 15 | 44908 | 0.071822 | 0.47137 | *KLK10* |
| ENSG00000171206 | 10 | 104394253 | 104428164 | 82 | 15 | 39414 | 0.071374 | 0.47155 | *TRIM8* |
| ENSG00000145703 | 5 | 75689074 | 76013957 | 1125 | 117 | 46208 | 0.071267 | 0.47159 | *IQGAP2* |
| ENSG00000155918 | 6 | 150329470 | 150356668 | 92 | 11 | 45534 | 0.071267 | 0.47159 | *RAET1L* |
| ENSG00000179598 | 17 | 17094309 | 17119629 | 82 | 20 | 46679 | 0.071256 | 0.4716 | *PLD6* |
| ENSG00000127314 | 12 | 68994619 | 69064372 | 159 | 28 | 39240 | 0.071007 | 0.4717 | *RAP1B* |
| ENSG00000179981 | 18 | 72912710 | 73011905 | 347 | 70 | 50724 | 0.070939 | 0.47172 | *TSHZ1* |
| ENSG00000173258 | 9 | 114277439 | 114350124 | 263 | 43 | 47871 | 0.070851 | 0.47176 | *ZNF483* |
| ENSG00000186197 | 1 | 236501562 | 236658214 | 722 | 74 | 39902 | 0.070772 | 0.47179 | *EDARADD* |
| ENSG00000164347 | 5 | 74007029 | 74073196 | 165 | 18 | 47573 | 0.070583 | 0.47186 | *GFM2* |
| ENSG00000148175 | 9 | 124091355 | 124142531 | 151 | 26 | 40426 | 0.070159 | 0.47203 | *STOM* |
| ENSG00000170190 | 17 | 73073822 | 73112257 | 181 | 27 | 40239 | 0.069801 | 0.47218 | *SLC16A5* |
| ENSG00000108021 | 10 | 5716801 | 5815703 | 307 | 40 | 52602 | 0.069431 | 0.47232 | *FAM208B* |
| ENSG00000158201 | 18 | 19220858 | 19294766 | 130 | 17 | 35567 | 0.069382 | 0.47234 | *ABHD3* |
| ENSG00000163162 | 2 | 101877681 | 101935163 | 120 | 24 | 50603 | 0.069362 | 0.47235 | *RNF149* |
| ENSG00000118004 | 2 | 3632426 | 3702048 | 263 | 42 | 45621 | 0.069334 | 0.47236 | *COLEC11* |
| ENSG00000129932 | 19 | 3480819 | 3510938 | 75 | 18 | 36969 | 0.069097 | 0.47246 | *DOHH* |
| ENSG00000183323 | 5 | 68566002 | 68638636 | 214 | 24 | 44706 | 0.069011 | 0.47249 | *CCDC125* |
| ENSG00000136813 | 9 | 114112972 | 114257025 | 403 | 34 | 39922 | 0.068931 | 0.47252 | *KIAA0368* |
| ENSG00000095139 | 11 | 118433105 | 118483748 | 103 | 11 | 31348 | 0.068945 | 0.47252 | *ARCN1* |
| ENSG00000104723 | 8 | 15264724 | 15634158 | 1962 | 153 | 45093 | 0.068909 | 0.47253 | *TUSC3* |
| ENSG00000148411 | 9 | 138888383 | 138997131 | 410 | 84 | 44707 | 0.068911 | 0.47253 | *NACC2* |
| ENSG00000185105 | 17 | 79887521 | 79915109 | 68 | 10 | 45371 | 0.068883 | 0.47254 | *MYADML2* |
| ENSG00000227124 | 3 | 75748794 | 75844734 | 155 | 15 | 42488 | 0.068856 | 0.47255 | *ZNF717* |
| ENSG00000143153 | 1 | 169064935 | 169111960 | 171 | 29 | 42210 | 0.068768 | 0.47259 | *ATP1B1* |
| ENSG00000136688 | 2 | 113720780 | 113753242 | 71 | 10 | 46724 | 0.068448 | 0.47271 | *IL36G* |
| ENSG00000214456 | 19 | 4512543 | 4545236 | 86 | 22 | 39646 | 0.068208 | 0.47281 | *PLIN5* |
| ENSG00000106070 | 7 | 50647760 | 50871159 | 818 | 73 | 46766 | 0.068135 | 0.47284 | *GRB10* |
| ENSG00000129158 | 11 | 17799595 | 18044709 | 484 | 58 | 46975 | 0.068094 | 0.47286 | *SERGEF* |
| ENSG00000188976 | 1 | 869584 | 904689 | 116 | 21 | 39693 | 0.068047 | 0.47287 | *NOC2L* |

| | | | | | | | | | |
|---|---|---|---|---|---|---|---|---|---|
| ENSG00000168032 | 3 | 40418647 | 40480110 | 133 | 10 | 38926 | 0.06789 | 0.47294 | *ENTPD3* |
| ENSG00000145817 | 5 | 143527723 | 143560278 | 110 | 10 | 50393 | 0.067877 | 0.47294 | *YIPF5* |
| ENSG00000112782 | 6 | 45858045 | 46058132 | 674 | 96 | 44577 | 0.067862 | 0.47295 | *CLIC5* |
| ENSG00000116128 | 1 | 147003182 | 147108017 | 327 | 66 | 46078 | 0.067803 | 0.47297 | *BCL9* |
| ENSG00000120729 | 5 | 137193480 | 137233540 | 44 | 14 | 42935 | 0.067807 | 0.47297 | *MYOT* |
| ENSG00000136997 | 8 | 128737680 | 128763674 | 70 | 25 | 39873 | 0.067758 | 0.47299 | *MYC* |
| ENSG00000025708 | 22 | 50954181 | 50978485 | 99 | 22 | 49186 | 0.067534 | 0.47308 | *TYMP* |
| ENSG00000151491 | 12 | 15763092 | 16045263 | 264 | 65 | 44310 | 0.06728 | 0.47318 | *EPS8* |
| ENSG00000145390 | 4 | 120123742 | 120226672 | 342 | 32 | 55377 | 0.067244 | 0.47319 | *USP53* |
| ENSG00000171124 | 19 | 5832899 | 5861485 | 92 | 30 | 38387 | 0.06688 | 0.47334 | *FUT3* |
| ENSG00000226777 | 14 | 106373838 | 106401825 | 19 | 9 | 45522 | 0.066646 | 0.47343 | *KIAA0125* |
| ENSG00000159335 | 12 | 6864682 | 6890116 | 48 | 12 | 35512 | 0.066614 | 0.47344 | *PTMS* |
| ENSG00000101199 | 20 | 61894137 | 61931142 | 141 | 17 | 49985 | 0.066636 | 0.47344 | *ARFGAP1* |
| ENSG00000154447 | 4 | 170005407 | 170202256 | 529 | 56 | 42685 | 0.066517 | 0.47348 | *SH3RF1* |
| ENSG00000135298 | 6 | 69335259 | 70109403 | 2411 | 139 | 48616 | 0.066533 | 0.47348 | *BAI3* |
| ENSG00000110628 | 11 | 2910951 | 2956476 | 159 | 44 | 45402 | 0.066512 | 0.47348 | *SLC22A18* |
| ENSG00000149443 | 20 | 18780372 | 18820797 | 135 | 31 | 52414 | 0.066508 | 0.47349 | *C20orf78* |
| ENSG00000204186 | 2 | 207129387 | 207189148 | 116 | 13 | 45380 | 0.066467 | 0.4735 | *ZDBF2* |
| ENSG00000169877 | 16 | 31529185 | 31550124 | 90 | 9 | 30615 | 0.066265 | 0.47358 | *AHSP* |
| ENSG00000159871 | 19 | 44290081 | 44341358 | 245 | 31 | 50239 | 0.066131 | 0.47364 | *LYPD5* |
| ENSG00000136531 | 2 | 166085912 | 166258818 | 500 | 48 | 54731 | 0.06594 | 0.47371 | *SCN2A* |
| ENSG00000129151 | 11 | 27052272 | 27159356 | 385 | 41 | 46644 | 0.065851 | 0.47375 | *BBOX1* |
| ENSG00000198604 | 14 | 35211937 | 35354853 | 501 | 41 | 49424 | 0.06583 | 0.47376 | *BAZ1A* |
| ENSG00000160349 | 9 | 138403284 | 138428378 | 90 | 23 | 46791 | 0.065782 | 0.47378 | *LCN1* |
| ENSG00000100065 | 22 | 37876400 | 37925549 | 181 | 26 | 41631 | 0.065544 | 0.47387 | *CARD10* |
| ENSG00000105371 | 19 | 10387643 | 10409198 | 68 | 14 | 39531 | 0.065531 | 0.47388 | *ICAM4* |
| ENSG00000168594 | 4 | 175740819 | 175909331 | 785 | 49 | 51818 | 0.06547 | 0.4739 | *ADAM29* |
| ENSG00000232434 | 9 | 139728867 | 139751797 | 35 | 13 | 50341 | 0.06541 | 0.47392 | *C9orf172* |
| ENSG00000071575 | 2 | 12847015 | 12892860 | 110 | 18 | 45196 | 0.06528 | 0.47398 | *TRIB2* |
| ENSG00000242612 | 16 | 441826 | 472487 | 110 | 15 | 36988 | 0.065204 | 0.47401 | *DECR2* |
| ENSG00000120656 | 1 | 28905835 | 28979597 | 112 | 20 | 46166 | 0.065171 | 0.47402 | *TAF12* |
| ENSG00000137207 | 6 | 43469565 | 43494728 | 45 | 9 | 46023 | 0.065136 | 0.47403 | *YIPF3* |
| ENSG00000164385 | 6 | 2612147 | 2644837 | 126 | 19 | 51087 | 0.065108 | 0.47404 | *C6orf195* |
| ENSG00000239961 | 19 | 54834456 | 54860421 | 110 | 39 | 47036 | 0.065059 | 0.47406 | *LILRA4* |
| ENSG00000137674 | 11 | 102437566 | 102506063 | 406 | 36 | 54177 | 0.064971 | 0.4741 | *MMP20* |
| ENSG00000170790 | 11 | 6880924 | 6901976 | 153 | 12 | 53786 | 0.064784 | 0.47417 | *OR10A2* |

| | | | | | | | | | |
|---|---|---|---|---|---|---|---|---|---|
| ENSG00000121621 | 11 | 28032167 | 28139855 | 160 | 21 | 37738 | 0.064725 | 0.4742 | *KIF18A* |
| ENSG00000183060 | 15 | 100245902 | 100283766 | 153 | 25 | 42248 | 0.064187 | 0.47441 | *LYSMD4* |
| ENSG00000173530 | 8 | 22983101 | 23031543 | 120 | 32 | 41173 | 0.064115 | 0.47444 | *TNFRSF10D* |
| ENSG00000143476 | 1 | 212198919 | 212290742 | 344 | 37 | 50228 | 0.064078 | 0.47445 | *DTL* |
| ENSG00000205323 | 12 | 56136247 | 56221540 | 92 | 22 | 40968 | 0.064064 | 0.47446 | *SARNP* |
| ENSG00000070047 | 11 | 566486 | 622222 | 243 | 25 | 45298 | 0.063576 | 0.47465 | *PHRF1* |
| ENSG00000139899 | 14 | 24885738 | 24910160 | 102 | 20 | 36979 | 0.063268 | 0.47478 | *CBLN3* |
| ENSG00000198750 | 7 | 74797499 | 74877509 | 6 | 4 | 43659 | 0.063162 | 0.47482 | *GATSL2* |
| ENSG00000166888 | 12 | 57479191 | 57535922 | 118 | 21 | 51929 | 0.063114 | 0.47484 | *STAT6* |
| ENSG00000185379 | 17 | 33416811 | 33458541 | 58 | 25 | 44822 | 0.06302 | 0.47488 | *RAD51D* |
| ENSG00000135722 | 16 | 67183834 | 67208473 | 17 | 10 | 46147 | 0.062887 | 0.47493 | *FBXL8* |
| ENSG00000144747 | 3 | 69058978 | 69111484 | 107 | 13 | 53352 | 0.062812 | 0.47496 | *TMF1* |
| ENSG00000152076 | 2 | 130886860 | 130912707 | 76 | 8 | 43450 | 0.062716 | 0.475 | *CCDC74B* |
| ENSG00000111245 | 12 | 111338623 | 111368526 | 116 | 19 | 47860 | 0.062708 | 0.475 | *MYL2* |
| ENSG00000124785 | 6 | 5988232 | 6017200 | 119 | 23 | 48175 | 0.062404 | 0.47512 | *NRN1* |
| ENSG00000007038 | 16 | 2857164 | 2886305 | 51 | 8 | 47336 | 0.062175 | 0.47521 | *PRSS21* |
| ENSG00000103152 | 16 | 117006 | 145852 | 75 | 14 | 31670 | 0.061868 | 0.47533 | *MPG* |
| ENSG00000153561 | 2 | 86937296 | 87015164 | 141 | 19 | 48256 | 0.061746 | 0.47538 | *RMND5A* |
| ENSG00000090520 | 3 | 186275192 | 186325061 | 147 | 14 | 34619 | 0.061747 | 0.47538 | *DNAJB11* |
| ENSG00000134419 | 16 | 18782617 | 18811705 | 77 | 12 | 48040 | 0.061665 | 0.47541 | *RPS15A* |
| ENSG00000092020 | 14 | 35544673 | 35601723 | 219 | 24 | 46413 | 0.061517 | 0.47547 | *PPP2R3C* |
| ENSG00000116062 | 2 | 47912669 | 48047240 | 417 | 56 | 41986 | 0.061357 | 0.47554 | *MSH6* |
| ENSG00000177076 | 9 | 19398925 | 19462018 | 181 | 24 | 46523 | 0.061265 | 0.47557 | *ACER2* |
| ENSG00000186184 | 13 | 28184903 | 28251548 | 267 | 46 | 48967 | 0.061212 | 0.4756 | *POLR1D* |
| ENSG00000166592 | 16 | 66945582 | 66969547 | 34 | 11 | 33174 | 0.061179 | 0.47561 | *RRAD* |
| ENSG00000127124 | 1 | 41962036 | 42511596 | 1471 | 134 | 48332 | 0.061156 | 0.47562 | *HIVEP3* |
| ENSG00000135900 | 2 | 224812121 | 224842431 | 74 | 11 | 41219 | 0.061161 | 0.47562 | *MRPL44* |
| ENSG00000105136 | 19 | 57989079 | 58016048 | 147 | 10 | 51259 | 0.061015 | 0.47567 | *ZNF419* |
| ENSG00000137274 | 6 | 3108608 | 3163812 | 107 | 14 | 29559 | 0.060987 | 0.47568 | *BPHL* |
| ENSG00000149050 | 11 | 7010549 | 7051599 | 157 | 15 | 49786 | 0.060628 | 0.47583 | *ZNF214* |
| ENSG00000141447 | 18 | 21732008 | 21987844 | 626 | 111 | 44302 | 0.060546 | 0.47586 | *OSBPL1A* |
| ENSG00000100796 | 14 | 91913955 | 91986898 | 213 | 15 | 50695 | 0.060521 | 0.47587 | *SMEK1* |
| ENSG00000075461 | 17 | 64951026 | 65039514 | 283 | 60 | 40732 | 0.060373 | 0.47593 | *CACNG4* |
| ENSG00000008517 | 16 | 3105298 | 3141908 | 127 | 29 | 46686 | 0.060337 | 0.47594 | *IL32* |
| ENSG00000086598 | 12 | 124059078 | 124093116 | 128 | 11 | 44298 | 0.060288 | 0.47596 | *TMED2* |
| ENSG00000175575 | 11 | 73577744 | 73648790 | 233 | 22 | 44964 | 0.060218 | 0.47599 | *PAAF1* |

| | | | | | | | | | |
|---|---|---|---|---|---|---|---|---|---|
| ENSG00000112096 | 6 | 160080089 | 160193561 | 381 | 34 | 46911 | 0.060196 | 0.476 | *SOD2* |
| ENSG00000172985 | 2 | 109735804 | 110272207 | 1300 | 119 | 42429 | 0.059976 | 0.47609 | *SH3RF3* |
| ENSG00000269510 | 9 | 35032390 | 35052824 | 39 | 12 | 42487 | 0.059913 | 0.47611 | *FLJ00273* |
| ENSG00000141639 | 18 | 48076448 | 48268194 | 496 | 74 | 51097 | 0.059914 | 0.47611 | *MAPK4* |
| ENSG00000198150 | 17 | 8251731 | 8273538 | 105 | 10 | 37694 | 0.059774 | 0.47617 | *AC135178.1* |
| ENSG00000167332 | 11 | 4691401 | 4729084 | 159 | 31 | 42382 | 0.059615 | 0.47623 | *OR51E2* |
| ENSG00000163545 | 1 | 205261187 | 205300883 | 142 | 23 | 49739 | 0.059589 | 0.47624 | *NUAK2* |
| ENSG00000139832 | 13 | 111165417 | 111224080 | 311 | 46 | 48887 | 0.059268 | 0.47637 | *RAB20* |
| ENSG00000054148 | 9 | 139733176 | 139755488 | 29 | 10 | 48886 | 0.059126 | 0.47643 | *PHPT1* |
| ENSG00000163297 | 4 | 80812303 | 81056608 | 558 | 51 | 42211 | 0.058932 | 0.4765 | *ANTXR2* |
| ENSG00000158169 | 9 | 97851336 | 98089991 | 289 | 37 | 48221 | 0.05886 | 0.47653 | *FANCC* |
| ENSG00000023287 | 8 | 53525016 | 53668403 | 208 | 34 | 42514 | 0.058753 | 0.47657 | *RB1CC1* |
| ENSG00000134201 | 1 | 110244864 | 110328050 | 276 | 46 | 38310 | 0.058715 | 0.47659 | *GSTM5* |
| ENSG00000170222 | 17 | 10590911 | 10624550 | 109 | 5 | 26557 | 0.058664 | 0.47661 | *ADPRM* |
| ENSG00000168631 | 6 | 30898749 | 30931998 | 120 | 25 | 40721 | 0.058547 | 0.47666 | *DPCR1* |
| ENSG00000185231 | 18 | 13872043 | 13925706 | 209 | 23 | 48831 | 0.058507 | 0.47667 | *MC2R* |
| ENSG00000181773 | 1 | 27709148 | 27732318 | 35 | 9 | 38363 | 0.05816 | 0.47681 | *GPR3* |
| ENSG00000196872 | 2 | 99400309 | 99562722 | 470 | 43 | 46529 | 0.058135 | 0.47682 | *KIAA1211L* |
| ENSG00000197901 | 11 | 62693857 | 62762455 | 153 | 22 | 47289 | 0.057872 | 0.47693 | *SLC22A6* |
| ENSG00000120265 | 6 | 150060579 | 150142556 | 199 | 9 | 30675 | 0.057465 | 0.47709 | *PCMT1* |
| ENSG00000174473 | 4 | 172723405 | 173972710 | 3171 | 182 | 42496 | 0.057349 | 0.47713 | *GALNTL6* |
| ENSG00000118096 | 11 | 118405243 | 118453685 | 128 | 7 | 25321 | 0.057262 | 0.47717 | *IFT46* |
| ENSG00000163581 | 3 | 170704137 | 170754539 | 126 | 11 | 30354 | 0.057063 | 0.47725 | *SLC2A2* |
| ENSG00000170549 | 5 | 3586168 | 3611517 | 97 | 24 | 47859 | 0.056655 | 0.47741 | *IRX1* |
| ENSG00000182359 | 11 | 105911825 | 105958492 | 93 | 18 | 49803 | 0.056198 | 0.47759 | *KBTBD3* |
| ENSG00000198929 | 1 | 162029564 | 162363321 | 1327 | 124 | 39416 | 0.055926 | 0.4777 | *NOS1AP* |
| ENSG00000188869 | 15 | 81613558 | 81676554 | 208 | 22 | 31297 | 0.055704 | 0.47779 | *TMC3* |
| ENSG00000167543 | 17 | 27883070 | 27910175 | 46 | 17 | 48009 | 0.055642 | 0.47781 | *TP53I13* |
| ENSG00000261833 | 16 | 76577314 | 76679520 | 335 | 61 | 50115 | 0.05554 | 0.47785 | *RP11-58C22.1* |
| ENSG00000094755 | 5 | 170180354 | 170251051 | 285 | 48 | 39900 | 0.055475 | 0.47788 | *GABRP* |
| ENSG00000131480 | 17 | 40986617 | 41012724 | 15 | 9 | 52729 | 0.055297 | 0.47795 | *AOC2* |
| ENSG00000132635 | 20 | 2805960 | 2831836 | 60 | 16 | 37601 | 0.055164 | 0.478 | *PCED1A* |
| ENSG00000134757 | 18 | 29017758 | 29068665 | 205 | 24 | 37212 | 0.054842 | 0.47813 | *DSG3* |
| ENSG00000130489 | 22 | 50951997 | 50974868 | 92 | 24 | 51791 | 0.054859 | 0.47813 | *SCO2* |
| ENSG00000169962 | 1 | 1256694 | 1280686 | 80 | 16 | 36842 | 0.054669 | 0.4782 | *TAS1R3* |
| ENSG00000256950 | 12 | 122316715 | 122369376 | 167 | 22 | 33798 | 0.054631 | 0.47822 | *RP11-87C12.2* |

| | | | | | | | | | |
|---|---|---|---|---|---|---|---|---|---|
| ENSG00000151948 | 12 | 129327972 | 129479509 | 716 | 120 | 43615 | 0.054331 | 0.47834 | *GLT1D1* |
| ENSG00000038532 | 16 | 11028345 | 11286046 | 793 | 62 | 46665 | 0.054096 | 0.47843 | *CLEC16A* |
| ENSG00000262664 | 17 | 1935327 | 1956724 | 64 | 19 | 50982 | 0.053984 | 0.47847 | *OVCA2* |
| ENSG00000148337 | 9 | 130918343 | 130976662 | 145 | 22 | 32582 | 0.053653 | 0.47861 | *CIZ1* |
| ENSG00000198183 | 20 | 31813801 | 31841115 | 62 | 6 | 38445 | 0.053568 | 0.47864 | *BPIFA1* |
| ENSG00000188176 | 17 | 4477294 | 4521614 | 197 | 44 | 40937 | 0.053517 | 0.47866 | *SMTNL2* |
| ENSG00000229924 | 4 | 9162135 | 9188453 | 6 | 3 | 42833 | 0.053251 | 0.47877 | *FAM90A26* |
| ENSG00000134324 | 2 | 11807721 | 11977535 | 590 | 92 | 43973 | 0.053159 | 0.4788 | *LPIN1* |
| ENSG00000196581 | 1 | 4704792 | 4862594 | 789 | 135 | 46187 | 0.052922 | 0.4789 | *AJAP1* |
| ENSG00000007933 | 1 | 171050018 | 171096959 | 210 | 27 | 43244 | 0.052826 | 0.47894 | *FMO3* |
| ENSG00000167460 | 19 | 16167831 | 16223813 | 177 | 32 | 48224 | 0.052675 | 0.479 | *TPM4* |
| ENSG00000130720 | 9 | 133767825 | 133824673 | 267 | 45 | 39347 | 0.052598 | 0.47903 | *FIBCD1* |
| ENSG00000265264 | 11 | 6492715 | 6540208 | 138 | 14 | 50332 | 0.052535 | 0.47905 | *TIMM10B* |
| ENSG00000130208 | 19 | 45407504 | 45432606 | 73 | 22 | 42055 | 0.05255 | 0.47905 | *APOC1* |
| ENSG00000072736 | 16 | 68108654 | 68273162 | 290 | 20 | 50652 | 0.052514 | 0.47906 | *NFATC3* |
| ENSG00000131368 | 3 | 15073967 | 15116842 | 107 | 19 | 54534 | 0.052453 | 0.47908 | *MRPS25* |
| ENSG00000259363 | 15 | 100337228 | 100427353 | 565 | 39 | 46600 | 0.051909 | 0.4793 | *CTD-2054N24.2* |
| ENSG00000204001 | 9 | 139638838 | 139662678 | 114 | 11 | 42439 | 0.051696 | 0.47939 | *LCN8* |
| ENSG00000197496 | 20 | 45328126 | 45374965 | 161 | 23 | 39466 | 0.051461 | 0.47948 | *SLC2A10* |
| ENSG00000160688 | 1 | 154945814 | 154975587 | 39 | 10 | 42210 | 0.05138 | 0.47951 | *FLAD1* |
| ENSG00000182645 | 10 | 118073940 | 118149541 | 232 | 28 | 34902 | 0.05113 | 0.47961 | *CCDC172* |
| ENSG00000111713 | 12 | 21679123 | 21767781 | 325 | 25 | 42491 | 0.050962 | 0.47968 | *GYS2* |
| ENSG00000163735 | 4 | 74851359 | 74874496 | 41 | 6 | 27964 | 0.050897 | 0.4797 | *CXCL5* |
| ENSG00000183718 | 5 | 180671417 | 180698119 | 97 | 17 | 36116 | 0.050889 | 0.47971 | *TRIM52* |
| ENSG00000021461 | 7 | 99415636 | 99473718 | 125 | 16 | 33814 | 0.050873 | 0.47971 | *CYP3A43* |
| ENSG00000172209 | 7 | 107100463 | 107126098 | 41 | 8 | 52921 | 0.050817 | 0.47974 | *GPR22* |
| ENSG00000064932 | 19 | 1097636 | 1184282 | 369 | 61 | 50353 | 0.050795 | 0.47974 | *SBNO2* |
| ENSG00000105392 | 19 | 48312703 | 48356587 | 323 | 34 | 45658 | 0.050778 | 0.47975 | *CRX* |
| ENSG00000148090 | 9 | 93966097 | 94134195 | 488 | 41 | 42933 | 0.050684 | 0.47979 | *AUH* |
| ENSG00000134871 | 13 | 110948159 | 111175374 | 1151 | 162 | 48555 | 0.050615 | 0.47982 | *COL4A2* |
| ENSG00000204876 | 7 | 155745326 | 155769037 | 87 | 29 | 44136 | 0.050464 | 0.47988 | *AC021218.2* |
| ENSG00000068137 | 17 | 40809932 | 40839048 | 44 | 9 | 50453 | 0.050341 | 0.47993 | *PLEKHH3* |
| ENSG00000196862 | 2 | 108433388 | 108517297 | 116 | 16 | 44940 | 0.050154 | 0.48 | *RGPD4* |
| ENSG00000110077 | 11 | 59929081 | 59962139 | 74 | 11 | 42343 | 0.050143 | 0.48 | *MS4A6A* |
| ENSG00000125844 | 20 | 17584323 | 17672940 | 216 | 40 | 39577 | 0.049924 | 0.48009 | *RRBP1* |
| ENSG00000101695 | 18 | 29588335 | 29663176 | 173 | 35 | 49218 | 0.049856 | 0.48012 | *RNF125* |

| | | | | | | | | | |
|---|---|---|---|---|---|---|---|---|---|
| ENSG00000158874 | 1 | 161182082 | 161203421 | 61 | 16 | 50515 | 0.049745 | 0.48016 | *APOA2* |
| ENSG00000104953 | 19 | 2967444 | 3005177 | 112 | 27 | 32649 | 0.049695 | 0.48018 | *TLE6* |
| ENSG00000259009 | 19 | 48327701 | 48374769 | 307 | 30 | 44825 | 0.049676 | 0.48019 | *TPRX2P* |
| ENSG00000135002 | 9 | 78990433 | 79019433 | 108 | 19 | 45953 | 0.049461 | 0.48028 | *RFK* |
| ENSG00000267882 | 20 | 45937246 | 45959467 | 26 | 10 | 43124 | 0.049456 | 0.48028 | *AL031666.2* |
| ENSG00000146006 | 5 | 138194612 | 138221057 | 43 | 5 | 52861 | 0.049375 | 0.48031 | *LRRTM2* |
| ENSG00000159792 | 16 | 67917175 | 67973581 | 70 | 15 | 47953 | 0.049187 | 0.48039 | *PSKH1* |
| ENSG00000169896 | 16 | 31261311 | 31354213 | 260 | 22 | 32014 | 0.049082 | 0.48043 | *ITGAM* |
| ENSG00000203814 | 1 | 149744245 | 149793928 | 56 | 9 | 43680 | 0.048925 | 0.48049 | *HIST2H2BF* |
| ENSG00000163798 | 2 | 27876338 | 27927840 | 36 | 13 | 41018 | 0.048924 | 0.48049 | *SLC4A1AP* |
| ENSG00000227345 | 10 | 51016325 | 51140715 | 203 | 20 | 44711 | 0.048738 | 0.48056 | *PARG* |
| ENSG00000006459 | 7 | 139774546 | 139886835 | 310 | 25 | 51898 | 0.048324 | 0.48073 | *KDM7A* |
| ENSG00000166451 | 16 | 81030103 | 81076719 | 179 | 13 | 39598 | 0.048265 | 0.48075 | *CENPN* |
| ENSG00000178935 | 19 | 58305209 | 58336281 | 94 | 15 | 47647 | 0.047929 | 0.48089 | *ZNF552* |
| ENSG00000181617 | 4 | 71081788 | 71110969 | 115 | 8 | 30920 | 0.046976 | 0.48127 | *FDCSP* |
| ENSG00000009709 | 1 | 18947500 | 19085360 | 437 | 93 | 46111 | 0.046747 | 0.48136 | *PAX7* |
| ENSG00000198558 | 6 | 27830926 | 27851289 | 62 | 14 | 37267 | 0.046693 | 0.48138 | *HIST1H4L* |
| ENSG00000164032 | 4 | 100859243 | 100881545 | 52 | 13 | 45315 | 0.04656 | 0.48143 | *H2AFZ* |
| ENSG00000151327 | 14 | 35504113 | 35592336 | 367 | 33 | 46736 | 0.046488 | 0.48146 | *FAM177A1* |
| ENSG00000174599 | 4 | 117994718 | 118016736 | 58 | 10 | 46341 | 0.046461 | 0.48147 | *TRAM1L1* |
| ENSG00000164327 | 5 | 38928021 | 39084510 | 385 | 27 | 52291 | 0.046337 | 0.48152 | *RICTOR* |
| ENSG00000171471 | 12 | 116987186 | 117024425 | 118 | 32 | 36792 | 0.045924 | 0.48169 | *MAP1LC3B2* |
| ENSG00000139330 | 12 | 91434268 | 91461760 | 136 | 22 | 34662 | 0.045832 | 0.48172 | *KERA* |
| ENSG00000132357 | 5 | 40831286 | 40870275 | 102 | 17 | 45846 | 0.045727 | 0.48176 | *CARD6* |
| ENSG00000244291 | 7 | 156421057 | 156443348 | 71 | 17 | 37627 | 0.045745 | 0.48176 | *C7orf13* |
| ENSG00000130772 | 1 | 28645513 | 28672476 | 47 | 11 | 50486 | 0.045691 | 0.48178 | *MED18* |
| ENSG00000171496 | 9 | 125319778 | 125340838 | 166 | 18 | 47360 | 0.045694 | 0.48178 | *OR1L8* |
| ENSG00000130731 | 16 | 674429 | 696358 | 54 | 11 | 50105 | 0.045667 | 0.48179 | *C16orf13* |
| ENSG00000167695 | 17 | 625652 | 656206 | 138 | 26 | 48686 | 0.045606 | 0.48181 | *FAM57A* |
| ENSG00000158553 | 6 | 27243682 | 27289949 | 175 | 22 | 49268 | 0.045595 | 0.48182 | *POM121L2* |
| ENSG00000179855 | 19 | 3575551 | 3603539 | 107 | 23 | 41221 | 0.045483 | 0.48186 | *GIPC3* |
| ENSG00000184545 | 11 | 1565274 | 1603150 | 130 | 12 | 54978 | 0.045445 | 0.48188 | *DUSP8* |
| ENSG00000231887 | 12 | 11023610 | 11046854 | 82 | 6 | 27289 | 0.045337 | 0.48192 | *PRH1* |
| ENSG00000258417 | 8 | 133026467 | 133107902 | 300 | 50 | 46190 | 0.045289 | 0.48194 | *OC90* |
| ENSG00000137492 | 11 | 76051000 | 76102015 | 113 | 23 | 51700 | 0.045241 | 0.48196 | *PRKRIR* |
| ENSG00000116711 | 1 | 186788085 | 186968113 | 610 | 69 | 43680 | 0.045174 | 0.48198 | *PLA2G4A* |

| | | | | | | | | | |
|---|---|---|---|---|---|---|---|---|---|
| ENSG00000180934 | 11 | 6037901 | 6058971 | 83 | 16 | 49046 | 0.045042 | 0.48204 | *OR56A1* |
| ENSG00000189366 | 3 | 125638118 | 125665882 | 149 | 11 | 29825 | 0.044991 | 0.48206 | *ALG1L* |
| ENSG00000151640 | 10 | 133990404 | 134029280 | 159 | 22 | 48924 | 0.044504 | 0.48225 | *DPYSL4* |
| ENSG00000266208 | 17 | 38506907 | 38530945 | 47 | 10 | 55283 | 0.044506 | 0.48225 | *CTD-2267D19.3* |
| ENSG00000183153 | 17 | 38507235 | 38530067 | 47 | 10 | 55283 | 0.044506 | 0.48225 | *GJD3* |
| ENSG00000146904 | 7 | 143077382 | 143115985 | 99 | 26 | 43911 | 0.044325 | 0.48232 | *EPHA1* |
| ENSG00000187005 | 21 | 32117439 | 32137746 | 42 | 4 | 21854 | 0.044257 | 0.48235 | *KRTAP21-1* |
| ENSG00000204394 | 6 | 31735295 | 31773730 | 95 | 29 | 35469 | 0.043949 | 0.48247 | *VARS* |
| ENSG00000156675 | 8 | 37706136 | 37766985 | 60 | 24 | 51793 | 0.043956 | 0.48247 | *RAB11FIP1* |
| ENSG00000174808 | 4 | 75659969 | 75729896 | 300 | 25 | 35996 | 0.043886 | 0.4825 | *BTC* |
| ENSG00000255823 | 11 | 10519434 | 10540723 | 95 | 20 | 38424 | 0.043874 | 0.4825 | *MTRNR2L8* |
| ENSG00000258227 | 7 | 141617157 | 141656807 | 93 | 15 | 41073 | 0.043811 | 0.48253 | *CLEC5A* |
| ENSG00000175224 | 11 | 46628826 | 46706368 | 76 | 14 | 51287 | 0.043734 | 0.48256 | *ATG13* |
| ENSG00000108774 | 17 | 40266994 | 40317035 | 110 | 12 | 32169 | 0.043729 | 0.48256 | *RAB5C* |
| ENSG00000152601 | 3 | 151951617 | 152193569 | 441 | 45 | 48791 | 0.043084 | 0.48282 | *MBNL1* |
| ENSG00000214109 | 19 | 13880884 | 13910972 | 130 | 14 | 25573 | 0.043048 | 0.48283 | *AC008686.1* |
| ENSG00000155754 | 2 | 202342148 | 202493901 | 490 | 30 | 45330 | 0.042876 | 0.4829 | *ALS2CR11* |
| ENSG00000137571 | 8 | 70569282 | 70757299 | 558 | 89 | 48952 | 0.042765 | 0.48294 | *SLCO5A1* |
| ENSG00000170367 | 20 | 23846572 | 23870387 | 75 | 15 | 43465 | 0.042745 | 0.48295 | *CST5* |
| ENSG00000163810 | 3 | 44906100 | 44966482 | 162 | 19 | 31126 | 0.042614 | 0.483 | *TGM4* |
| ENSG00000213380 | 16 | 69344043 | 69383570 | 109 | 19 | 54066 | 0.042562 | 0.48303 | *COG8* |
| ENSG00000163486 | 1 | 206506200 | 206647783 | 289 | 33 | 39809 | 0.042186 | 0.48318 | *SRGAP2* |
| ENSG00000117118 | 1 | 17335217 | 17390665 | 196 | 19 | 39134 | 0.041919 | 0.48328 | *SDHB* |
| ENSG00000106258 | 7 | 99235817 | 99287621 | 53 | 21 | 52728 | 0.041803 | 0.48333 | *CYP3A5* |
| ENSG00000158156 | 1 | 28275973 | 28304607 | 67 | 19 | 43552 | 0.041678 | 0.48338 | *XKR8* |
| ENSG00000135502 | 12 | 58003310 | 58029934 | 50 | 13 | 49085 | 0.041552 | 0.48343 | *SLC26A10* |
| ENSG00000142166 | 21 | 34686734 | 34742168 | 164 | 21 | 41396 | 0.041171 | 0.48358 | *IFNAR1* |
| ENSG00000119729 | 2 | 46758945 | 46820260 | 157 | 29 | 41194 | 0.041074 | 0.48362 | *RHOQ* |
| ENSG00000005007 | 19 | 18932747 | 18989045 | 118 | 21 | 50786 | 0.040981 | 0.48366 | *UPF1* |
| ENSG00000141698 | 17 | 39971335 | 40002523 | 103 | 8 | 52179 | 0.040915 | 0.48368 | *NT5C3B* |
| ENSG00000078177 | 4 | 40048446 | 40169872 | 328 | 39 | 48130 | 0.040794 | 0.48373 | *N4BP2* |
| ENSG00000203907 | 6 | 74068278 | 74114856 | 107 | 22 | 54479 | 0.040607 | 0.4838 | *OOEP* |
| ENSG00000253598 | 8 | 82595842 | 82618409 | 86 | 14 | 39368 | 0.040542 | 0.48383 | *SLC10A5* |
| ENSG00000132436 | 7 | 50501831 | 50528088 | 69 | 14 | 45896 | 0.040342 | 0.48391 | *FIGNL1* |
| ENSG00000107263 | 9 | 134442157 | 134625461 | 513 | 49 | 45063 | 0.040354 | 0.48391 | *RAPGEF1* |
| ENSG00000151632 | 10 | 5019967 | 5070223 | 309 | 45 | 41995 | 0.040235 | 0.48395 | *AKR1C2* |

| | | | | | | | | | |
|---|---|---|---|---|---|---|---|---|---|
| ENSG00000163029 | 2 | 17835079 | 17991509 | 362 | 35 | 38801 | 0.040053 | 0.48403 | *SMC6* |
| ENSG00000174417 | 8 | 110088850 | 110141813 | 167 | 22 | 46538 | 0.040029 | 0.48403 | *TRHR* |
| ENSG00000273452 | 17 | 12616199 | 12671542 | 135 | 36 | 40795 | 0.039884 | 0.48409 | *AC005358.1* |
| ENSG00000128617 | 7 | 128402545 | 128425844 | 74 | 11 | 36424 | 0.039864 | 0.4841 | *OPN1SW* |
| ENSG00000115616 | 2 | 103226166 | 103337777 | 285 | 46 | 40855 | 0.03985 | 0.48411 | *SLC9A2* |
| ENSG00000175938 | 16 | 30950387 | 30977782 | 20 | 6 | 30816 | 0.039847 | 0.48411 | *ORAI3* |
| ENSG00000134744 | 1 | 52863954 | 53029159 | 295 | 31 | 45348 | 0.039623 | 0.4842 | *ZCCHC11* |
| ENSG00000115464 | 2 | 61404598 | 61707904 | 802 | 50 | 47211 | 0.039486 | 0.48425 | *USP34* |
| ENSG00000166313 | 11 | 6406355 | 6450644 | 102 | 29 | 54005 | 0.039306 | 0.48432 | *APBB1* |
| ENSG00000256500 | 14 | 104019299 | 104162261 | 394 | 31 | 47037 | 0.039154 | 0.48438 | *RP11-73M18.2* |
| ENSG00000143811 | 1 | 226097578 | 226121978 | 55 | 18 | 53698 | 0.039056 | 0.48442 | *PYCR2* |
| ENSG00000165633 | 10 | 50212290 | 50333554 | 472 | 38 | 33542 | 0.039056 | 0.48442 | *VSTM4* |
| ENSG00000257860 | 12 | 103535620 | 103572087 | 132 | 22 | 47284 | 0.039066 | 0.48442 | *RP11-552I14.1* |
| ENSG00000158825 | 1 | 20905441 | 20955401 | 281 | 33 | 50404 | 0.039037 | 0.48443 | *CDA* |
| ENSG00000268327 | 15 | 59429899 | 59450054 | 60 | 16 | 44140 | 0.038914 | 0.48448 | *C15ORF31* |
| ENSG00000241837 | 21 | 35265757 | 35298284 | 120 | 19 | 54740 | 0.038837 | 0.48451 | *ATP5O* |
| ENSG00000255737 | 12 | 58110054 | 58132139 | 34 | 8 | 47791 | 0.038441 | 0.48467 | *AGAP2-AS1* |
| ENSG00000105887 | 7 | 135601509 | 135672101 | 286 | 33 | 45259 | 0.038133 | 0.48479 | *MTPN* |
| ENSG00000184967 | 12 | 132618993 | 132647013 | 100 | 16 | 35948 | 0.038142 | 0.48479 | *NOC4L* |
| ENSG00000126214 | 14 | 104018233 | 104177888 | 438 | 34 | 46914 | 0.037863 | 0.4849 | *KLC1* |
| ENSG00000152936 | 12 | 25552241 | 25811513 | 1066 | 101 | 44233 | 0.037723 | 0.48495 | *IFLTD1* |
| ENSG00000101000 | 20 | 33749876 | 33775165 | 63 | 8 | 36540 | 0.0376 | 0.485 | *PROCR* |
| ENSG00000182271 | 17 | 28633351 | 28671077 | 83 | 15 | 44618 | 0.03754 | 0.48503 | *TMIGD1* |
| ENSG00000234949 | 2 | 238489663 | 238514624 | 153 | 24 | 31328 | 0.037396 | 0.48508 | *AC104667.3* |
| ENSG00000123411 | 12 | 56391443 | 56442219 | 68 | 17 | 45301 | 0.037385 | 0.48509 | *IKZF4* |
| ENSG00000138193 | 10 | 95743746 | 96102580 | 811 | 101 | 41530 | 0.037301 | 0.48512 | *PLCE1* |
| ENSG00000196865 | 10 | 115604420 | 115686953 | 94 | 27 | 46921 | 0.037139 | 0.48519 | *NHLRC2* |
| ENSG00000215421 | 18 | 72255106 | 72787627 | 1137 | 88 | 46927 | 0.036975 | 0.48525 | *ZNF407* |
| ENSG00000154839 | 18 | 47891365 | 47930543 | 156 | 13 | 41020 | 0.036936 | 0.48527 | *SKA1* |
| ENSG00000186487 | 2 | 1782885 | 2345032 | 1897 | 183 | 46145 | 0.036753 | 0.48534 | *MYT1L* |
| ENSG00000189046 | 12 | 109515996 | 109541436 | 62 | 21 | 47415 | 0.036565 | 0.48542 | *ALKBH2* |
| ENSG00000154102 | 16 | 85713690 | 85794735 | 264 | 41 | 48922 | 0.036562 | 0.48542 | *C16orf74* |
| ENSG00000158014 | 1 | 26353743 | 26382624 | 67 | 10 | 31167 | 0.036519 | 0.48543 | *SLC30A2* |
| ENSG00000143353 | 1 | 219337186 | 219396207 | 117 | 15 | 50751 | 0.036449 | 0.48546 | *LYPLAL1* |
| ENSG00000100941 | 14 | 39634387 | 39662422 | 89 | 17 | 48003 | 0.036366 | 0.4855 | *PNN* |
| ENSG00000151726 | 4 | 185666749 | 185757972 | 329 | 43 | 46150 | 0.036287 | 0.48553 | *ACSL1* |

| | | | | | | | | | |
|---|---|---|---|---|---|---|---|---|---|
| ENSG00000079150 | 2 | 179318391 | 179353327 | 66 | 15 | 38241 | 0.03585 | 0.4857 | *FKBP7* |
| ENSG00000137463 | 4 | 140177317 | 140211492 | 97 | 16 | 51284 | 0.034953 | 0.48606 | *MGARP* |
| ENSG00000108406 | 17 | 57632886 | 57695706 | 58 | 12 | 32253 | 0.034936 | 0.48607 | *DHX40* |
| ENSG00000106479 | 7 | 149525456 | 149574568 | 121 | 21 | 45324 | 0.034534 | 0.48623 | *ZNF862* |
| ENSG00000186532 | 17 | 1672779 | 1743928 | 296 | 31 | 36238 | 0.0341 | 0.4864 | *SMYD4* |
| ENSG00000155729 | 2 | 201343675 | 201394507 | 144 | 36 | 48490 | 0.034052 | 0.48642 | *KCTD18* |
| ENSG00000105784 | 7 | 87246864 | 87471611 | 403 | 30 | 49918 | 0.034054 | 0.48642 | *RUNDC3B* |
| ENSG00000115459 | 2 | 85571517 | 85628875 | 174 | 30 | 49697 | 0.033995 | 0.48644 | *ELMOD3* |
| ENSG00000177992 | 9 | 90487741 | 90513814 | 102 | 27 | 33149 | 0.033973 | 0.48645 | *SPATA31E1* |
| ENSG00000172270 | 19 | 561297 | 593493 | 222 | 31 | 34532 | 0.033768 | 0.48653 | *BSG* |
| ENSG00000137101 | 9 | 35599530 | 35656807 | 179 | 36 | 48212 | 0.033706 | 0.48656 | *CD72* |
| ENSG00000164329 | 5 | 78897943 | 78992471 | 266 | 16 | 29265 | 0.033413 | 0.48667 | *PAPD4* |
| ENSG00000182117 | 15 | 34623917 | 34645378 | 113 | 12 | 49442 | 0.03321 | 0.48675 | *NOP10* |
| ENSG00000146192 | 6 | 36963422 | 37006846 | 203 | 34 | 48753 | 0.032989 | 0.48684 | *FGD2* |
| ENSG00000100330 | 22 | 30269144 | 30436855 | 451 | 43 | 48260 | 0.032537 | 0.48702 | *MTMR3* |
| ENSG00000196411 | 7 | 100390187 | 100435121 | 127 | 21 | 44916 | 0.032527 | 0.48703 | *EPHB4* |
| ENSG00000173714 | 17 | 48902011 | 48929709 | 84 | 12 | 36399 | 0.032507 | 0.48703 | *WFIKKN2* |
| ENSG00000175155 | 17 | 57399050 | 57489090 | 214 | 31 | 44919 | 0.032237 | 0.48714 | *YPEL2* |
| ENSG00000115267 | 2 | 163113589 | 163185213 | 98 | 16 | 39067 | 0.032082 | 0.4872 | *IFIH1* |
| ENSG00000179240 | 11 | 76082357 | 76135663 | 173 | 31 | 45839 | 0.032044 | 0.48722 | *RP11-111M22.2* |
| ENSG00000268466 | 14 | 58744751 | 58765865 | 44 | 6 | 52839 | 0.031919 | 0.48727 | *AL132989.1* |
| ENSG00000108423 | 17 | 57926851 | 57980304 | 128 | 16 | 38289 | 0.031844 | 0.4873 | *TUBD1* |
| ENSG00000103642 | 15 | 63403999 | 63444260 | 158 | 16 | 41243 | 0.031245 | 0.48754 | *LACTB* |
| ENSG00000112739 | 6 | 4011501 | 4075217 | 232 | 26 | 49335 | 0.03111 | 0.48759 | *PRPF4B* |
| ENSG00000085840 | 1 | 52828501 | 52880131 | 50 | 16 | 45444 | 0.030858 | 0.48769 | *ORC1* |
| ENSG00000064961 | 19 | 3562775 | 3589086 | 67 | 17 | 38161 | 0.030771 | 0.48773 | *HMG20B* |
| ENSG00000100142 | 22 | 38338614 | 38447922 | 126 | 17 | 44814 | 0.03076 | 0.48773 | *POLR2F* |
| ENSG00000163485 | 1 | 203049782 | 203146533 | 225 | 40 | 39989 | 0.030642 | 0.48778 | *ADORA1* |
| ENSG00000169981 | 3 | 44680219 | 44712283 | 33 | 10 | 42227 | 0.029855 | 0.48809 | *ZNF35* |
| ENSG00000157653 | 9 | 116162334 | 116201964 | 112 | 13 | 56773 | 0.02964 | 0.48818 | *C9orf43* |
| ENSG00000085832 | 1 | 51809935 | 51995000 | 667 | 42 | 53076 | 0.029572 | 0.4882 | *EPS15* |
| ENSG00000122642 | 7 | 32987017 | 33056543 | 251 | 43 | 41084 | 0.029452 | 0.48825 | *FKBP9* |
| ENSG00000204628 | 5 | 180653909 | 180685096 | 101 | 18 | 34680 | 0.029262 | 0.48833 | *GNB2L1* |
| ENSG00000158497 | 5 | 143181726 | 143210284 | 60 | 15 | 48059 | 0.028872 | 0.48848 | *HMHB1* |
| ENSG00000116039 | 2 | 71153012 | 71202536 | 180 | 43 | 44176 | 0.028718 | 0.48854 | *ATP6V1B1* |
| ENSG00000221843 | 2 | 27789389 | 27815588 | 41 | 9 | 55313 | 0.028681 | 0.48856 | *C2orf16* |

| | | | | | | | | | |
|---|---|---|---|---|---|---|---|---|---|
| ENSG00000196715 | 7 | 65328254 | 65434550 | 257 | 20 | 35127 | 0.028683 | 0.48856 | *VKORC1L1* |
| ENSG00000141759 | 18 | 77722867 | 77803949 | 292 | 23 | 42268 | 0.028677 | 0.48856 | *TXNL4A* |
| ENSG00000127922 | 7 | 96100938 | 96349203 | 605 | 56 | 40593 | 0.028617 | 0.48859 | *SHFM1* |
| ENSG00000105472 | 19 | 51216586 | 51238974 | 49 | 13 | 52624 | 0.028598 | 0.48859 | *CLEC11A* |
| ENSG00000163576 | 3 | 19910964 | 19998517 | 320 | 39 | 43117 | 0.028591 | 0.4886 | *EFHB* |
| ENSG00000188092 | 1 | 147390506 | 147475753 | 36 | 8 | 42269 | 0.028484 | 0.48864 | *GPR89B* |
| ENSG00000157014 | 3 | 10279707 | 10332902 | 152 | 24 | 36274 | 0.028478 | 0.48864 | *TATDN2* |
| ENSG00000144579 | 2 | 219252979 | 219280664 | 54 | 21 | 41825 | 0.028407 | 0.48867 | *CTDSP1* |
| ENSG00000168883 | 2 | 85819979 | 85886403 | 126 | 22 | 39703 | 0.028235 | 0.48874 | *USP39* |
| ENSG00000158435 | 2 | 101859264 | 101896778 | 71 | 17 | 53208 | 0.028224 | 0.48874 | *CNOT11* |
| ENSG00000188859 | 1 | 166016674 | 166146206 | 400 | 40 | 45609 | 0.02819 | 0.48876 | *FAM78B* |
| ENSG00000112852 | 5 | 140464227 | 140486962 | 86 | 8 | 54406 | 0.028013 | 0.48883 | *PCDHB2* |
| ENSG00000147647 | 8 | 105332552 | 105489281 | 507 | 77 | 41477 | 0.027924 | 0.48886 | *DPYS* |
| ENSG00000177082 | 15 | 85175999 | 85207574 | 86 | 15 | 39933 | 0.027901 | 0.48887 | *WDR73* |
| ENSG00000155093 | 7 | 157321750 | 158390480 | 4404 | 394 | 45266 | 0.027765 | 0.48892 | *PTPRN2* |
| ENSG00000172345 | 15 | 81591394 | 81626524 | 118 | 24 | 38008 | 0.027717 | 0.48894 | *STARD5* |
| ENSG00000122873 | 10 | 60018818 | 60059346 | 137 | 13 | 35901 | 0.027672 | 0.48896 | *CISD1* |
| ENSG00000119812 | 2 | 33798725 | 33834449 | 108 | 19 | 41446 | 0.027607 | 0.48899 | *FAM98A* |
| ENSG00000141428 | 18 | 33542046 | 33569241 | 59 | 9 | 52736 | 0.027609 | 0.48899 | *C18orf21* |
| ENSG00000157445 | 3 | 54146574 | 55118584 | 3210 | 331 | 49165 | 0.027509 | 0.48903 | *CACNA2D3* |
| ENSG00000103978 | 15 | 42492730 | 42575861 | 139 | 20 | 37773 | 0.027282 | 0.48912 | *TMEM87A* |
| ENSG00000232653 | 15 | 32875657 | 32906179 | 23 | 10 | 44040 | 0.027199 | 0.48915 | *GOLGA8N* |
| ENSG00000134186 | 1 | 109224945 | 109254425 | 21 | 13 | 51096 | 0.027104 | 0.48919 | *PRPF38B* |
| ENSG00000254685 | 1 | 74653896 | 74709770 | 152 | 12 | 27385 | 0.026989 | 0.48923 | *FPGT* |
| ENSG00000152779 | 10 | 91180051 | 91326398 | 417 | 77 | 44287 | 0.026948 | 0.48925 | *SLC16A12* |
| ENSG00000269175 | 1 | 101445761 | 101467653 | 29 | 7 | 55865 | 0.026823 | 0.4893 | *AC093157.1* |
| ENSG00000132326 | 2 | 239142679 | 239208743 | 220 | 29 | 48583 | 0.026596 | 0.48939 | *PER2* |
| ENSG00000268822 | 2 | 241614478 | 241634705 | 60 | 11 | 46663 | 0.026533 | 0.48942 | *AC011298.1* |
| ENSG00000142615 | 1 | 15773223 | 15808586 | 126 | 23 | 45692 | 0.026419 | 0.48946 | *CELA2A* |
| ENSG00000169684 | 15 | 78847862 | 78897611 | 128 | 17 | 47590 | 0.02631 | 0.48951 | *CHRNA5* |
| ENSG00000186818 | 19 | 55145340 | 55191810 | 240 | 36 | 41147 | 0.02612 | 0.48958 | *LILRB4* |
| ENSG00000127980 | 7 | 92106334 | 92167845 | 119 | 18 | 50768 | 0.025882 | 0.48968 | *PEX1* |
| ENSG00000221923 | 19 | 52863170 | 52899048 | 166 | 24 | 48021 | 0.025872 | 0.48968 | *ZNF880* |
| ENSG00000132259 | 11 | 6245995 | 6275659 | 63 | 15 | 41699 | 0.025746 | 0.48973 | *CNGA4* |
| ENSG00000175745 | 5 | 92909043 | 92940321 | 37 | 16 | 43400 | 0.025699 | 0.48975 | *NR2F1* |
| ENSG00000135677 | 12 | 65097225 | 65163227 | 105 | 18 | 41348 | 0.0257 | 0.48975 | *GNS* |

| | | | | | | | | | |
|---|---|---|---|---|---|---|---|---|---|
| ENSG00000177565 | 3 | 176727143 | 176925261 | 598 | 60 | 50272 | 0.02555 | 0.48981 | *TBL1XR1* |
| ENSG00000100207 | 22 | 42546019 | 42749622 | 509 | 67 | 45043 | 0.025534 | 0.48981 | *TCF20* |
| ENSG00000143001 | 1 | 55436465 | 55467966 | 107 | 11 | 32639 | 0.025017 | 0.49002 | *TMEM61* |
| ENSG00000043462 | 5 | 169663241 | 169735231 | 143 | 51 | 48710 | 0.024932 | 0.49005 | *LCP2* |
| ENSG00000173253 | 9 | 1040354 | 1067552 | 132 | 38 | 44930 | 0.024666 | 0.49016 | *DMRT2* |
| ENSG00000142459 | 19 | 7885119 | 7939862 | 178 | 22 | 48275 | 0.024665 | 0.49016 | *EVI5L* |
| ENSG00000175063 | 20 | 44431215 | 44455596 | 78 | 16 | 47729 | 0.024431 | 0.49025 | *UBE2C* |
| ENSG00000268821 | 2 | 172550844 | 172577314 | 67 | 14 | 43138 | 0.02431 | 0.4903 | *AC068039.1* |
| ENSG00000058799 | 1 | 54307392 | 54366407 | 231 | 21 | 53113 | 0.024229 | 0.49033 | *YIPF1* |
| ENSG00000205869 | 11 | 1595572 | 1616513 | 94 | 15 | 53503 | 0.02421 | 0.49034 | *KRTAP5-1* |
| ENSG00000184602 | 16 | 11752270 | 11783015 | 84 | 16 | 41909 | 0.024224 | 0.49034 | *SNN* |
| ENSG00000198216 | 1 | 181372238 | 181787219 | 1145 | 140 | 46626 | 0.024167 | 0.49036 | *CACNA1E* |
| ENSG00000228368 | 7 | 116595900 | 116617526 | 32 | 12 | 54264 | 0.024141 | 0.49037 | *AC106873.4* |
| ENSG00000165138 | 9 | 101483611 | 101569247 | 196 | 21 | 41371 | 0.023953 | 0.49044 | *ANKS6* |
| ENSG00000117691 | 1 | 212596229 | 212629714 | 125 | 21 | 41812 | 0.023901 | 0.49047 | *NENF* |
| ENSG00000171858 | 20 | 60952172 | 60973576 | 108 | 18 | 43215 | 0.023786 | 0.49051 | *RPS21* |
| ENSG00000198780 | 5 | 74063399 | 74172776 | 257 | 21 | 44420 | 0.02339 | 0.49067 | *FAM169A* |
| ENSG00000163374 | 1 | 155619237 | 155668791 | 53 | 13 | 38208 | 0.023333 | 0.49069 | *YY1AP1* |
| ENSG00000142867 | 1 | 85721931 | 85752773 | 133 | 24 | 55355 | 0.023298 | 0.49071 | *BCL10* |
| ENSG00000169836 | 4 | 104497188 | 104650973 | 533 | 56 | 35684 | 0.023177 | 0.49075 | *TACR3* |
| ENSG00000049283 | 17 | 48599904 | 48631111 | 128 | 21 | 41240 | 0.023106 | 0.49078 | *EPN3* |
| ENSG00000150779 | 11 | 111945524 | 111967522 | 24 | 8 | 42426 | 0.023068 | 0.4908 | *TIMM8B* |
| ENSG00000123395 | 12 | 52453030 | 52481278 | 101 | 19 | 48924 | 0.022817 | 0.4909 | *C12orf44* |
| ENSG00000186862 | 10 | 102757440 | 102800890 | 104 | 33 | 42039 | 0.022452 | 0.49104 | *PDZD7* |
| ENSG00000176244 | 10 | 15109522 | 15140775 | 161 | 33 | 45223 | 0.022149 | 0.49116 | *ACBD7* |
| ENSG00000179750 | 22 | 39368352 | 39398809 | 51 | 11 | 46994 | 0.02195 | 0.49124 | *APOBEC3B* |
| ENSG00000135372 | 11 | 34117149 | 34179217 | 129 | 10 | 53802 | 0.021854 | 0.49128 | *NAT10* |
| ENSG00000197977 | 6 | 10970992 | 11054547 | 178 | 20 | 34085 | 0.02176 | 0.49132 | *ELOVL2* |
| ENSG00000166391 | 11 | 75418864 | 75454003 | 95 | 17 | 41992 | 0.021698 | 0.49134 | *MOGAT2* |
| ENSG00000170748 | 11 | 7100165 | 7122379 | 110 | 14 | 45146 | 0.021659 | 0.49136 | *RBMXL2* |
| ENSG00000112769 | 6 | 112419963 | 112586141 | 475 | 59 | 43139 | 0.021595 | 0.49139 | *LAMA4* |
| ENSG00000183723 | 16 | 66638653 | 66740610 | 242 | 17 | 31278 | 0.021574 | 0.49139 | *CMTM4* |
| ENSG00000118557 | 16 | 72136056 | 72220777 | 252 | 30 | 44041 | 0.021518 | 0.49142 | *PMFBP1* |
| ENSG00000110696 | 11 | 16624679 | 16788428 | 306 | 30 | 43236 | 0.021489 | 0.49143 | *C11orf58* |
| ENSG00000120756 | 3 | 142305229 | 142442506 | 367 | 35 | 44413 | 0.021442 | 0.49145 | *PLS1* |
| ENSG00000168515 | 11 | 61947688 | 61971011 | 109 | 12 | 26267 | 0.021124 | 0.49157 | *SCGB1D1* |

| | | | | | | | | | |
|---|---|---|---|---|---|---|---|---|---|
| ENSG00000132965 | 13 | 31299645 | 31348556 | 209 | 38 | 44360 | 0.020921 | 0.49165 | *ALOX5AP* |
| ENSG00000134864 | 13 | 101173810 | 101251782 | 152 | 17 | 33700 | 0.020904 | 0.49166 | *GGACT* |
| ENSG00000006283 | 17 | 48628429 | 48714835 | 235 | 37 | 35361 | 0.020813 | 0.4917 | *CACNA1G* |
| ENSG00000170836 | 17 | 58667544 | 58751849 | 35 | 19 | 52633 | 0.020705 | 0.49174 | *PPM1D* |
| ENSG00000166509 | 16 | 78046412 | 78110658 | 552 | 31 | 35654 | 0.020557 | 0.4918 | *CLEC3A* |
| ENSG00000212659 | 17 | 39411591 | 39432893 | 29 | 7 | 34538 | 0.020333 | 0.49189 | *KRTAP9-6* |
| ENSG00000215704 | 1 | 15782404 | 15827895 | 192 | 24 | 48168 | 0.020232 | 0.49193 | *CELA2B* |
| ENSG00000197020 | 19 | 21895568 | 21960430 | 276 | 22 | 49783 | 0.020202 | 0.49194 | *ZNF100* |
| ENSG00000072163 | 2 | 128385956 | 128449360 | 260 | 50 | 44964 | 0.019821 | 0.49209 | *LIMS2* |
| ENSG00000241595 | 17 | 39395939 | 39416904 | 72 | 7 | 36719 | 0.019829 | 0.49209 | *KRTAP9-4* |
| ENSG00000197608 | 19 | 52557719 | 52609018 | 216 | 19 | 38202 | 0.019806 | 0.4921 | *ZNF841* |
| ENSG00000214842 | 2 | 17681851 | 17709706 | 95 | 9 | 47272 | 0.019415 | 0.49225 | *RAD51AP2* |
| ENSG00000184838 | 5 | 119789973 | 120033027 | 1067 | 93 | 45376 | 0.019432 | 0.49225 | *PRR16* |
| ENSG00000264364 | 17 | 56150776 | 56182897 | 69 | 14 | 40132 | 0.018812 | 0.4925 | *DYNLL2* |
| ENSG00000221874 | 19 | 53421728 | 53476076 | 217 | 38 | 38900 | 0.018681 | 0.49255 | *ZNF321P* |
| ENSG00000162458 | 1 | 16073102 | 16123089 | 149 | 25 | 48223 | 0.018547 | 0.4926 | *FBLIM1* |
| ENSG00000141452 | 18 | 21073473 | 21121746 | 99 | 13 | 41504 | 0.017993 | 0.49282 | *C18orf8* |
| ENSG00000111046 | 12 | 81091277 | 81113253 | 48 | 10 | 37469 | 0.01774 | 0.49292 | *MYF6* |
| ENSG00000172995 | 3 | 35670437 | 35845988 | 397 | 49 | 49182 | 0.017654 | 0.49296 | *ARPP21* |
| ENSG00000189057 | 11 | 58864658 | 58904883 | 67 | 15 | 51527 | 0.017473 | 0.49303 | *FAM111B* |
| ENSG00000258830 | 12 | 57633392 | 57700267 | 79 | 14 | 45009 | 0.017383 | 0.49307 | *RP11-123K3.4* |
| ENSG00000196966 | 6 | 26215383 | 26235844 | 85 | 8 | 55055 | 0.017309 | 0.49309 | *HIST1H3E* |
| ENSG00000132388 | 17 | 4162554 | 4279923 | 199 | 16 | 41316 | 0.017212 | 0.49313 | *UBE2G1* |
| ENSG00000198589 | 4 | 151175594 | 151946879 | 1485 | 95 | 35473 | 0.017096 | 0.49318 | *LRBA* |
| ENSG00000166669 | 16 | 10410291 | 10587495 | 670 | 62 | 42843 | 0.017106 | 0.49318 | *ATF7IP2* |
| ENSG00000108010 | 10 | 131924663 | 131992785 | 314 | 33 | 45427 | 0.01696 | 0.49323 | *GLRX3* |
| ENSG00000254413 | 22 | 50997298 | 51031394 | 168 | 28 | 29574 | 0.016907 | 0.49326 | *CHKB-CPT1B* |
| ENSG00000187908 | 10 | 124310181 | 124413252 | 438 | 25 | 49414 | 0.016612 | 0.49337 | *DMBT1* |
| ENSG00000166446 | 16 | 80621803 | 80848226 | 916 | 109 | 38576 | 0.016626 | 0.49337 | *CDYL2* |
| ENSG00000166035 | 15 | 58692768 | 58871151 | 649 | 122 | 46206 | 0.016599 | 0.49338 | *LIPC* |
| ENSG00000075914 | 3 | 45006733 | 45087558 | 209 | 19 | 30107 | 0.016404 | 0.49346 | *EXOSC7* |
| ENSG00000176444 | 1 | 155222659 | 155258282 | 63 | 7 | 32145 | 0.016372 | 0.49347 | *CLK2* |
| ENSG00000120802 | 12 | 98899290 | 98954157 | 174 | 26 | 43596 | 0.016354 | 0.49348 | *TMPO* |
| ENSG00000168746 | 20 | 43070624 | 43103984 | 74 | 21 | 41274 | 0.016291 | 0.4935 | *C20orf62* |
| ENSG00000064835 | 3 | 87298554 | 87335737 | 97 | 16 | 48598 | 0.016111 | 0.49357 | *POU1F1* |
| ENSG00000272762 | 3 | 51984317 | 52018032 | 43 | 15 | 33440 | 0.016074 | 0.49359 | *RP11-155D18.14* |

| | | | | | | | | | |
|---|---|---|---|---|---|---|---|---|---|
| ENSG00000069764 | 16 | 14756405 | 14798526 | 32 | 11 | 55525 | 0.015907 | 0.49365 | *PLA2G10* |
| ENSG00000100197 | 22 | 42512501 | 42536908 | 94 | 14 | 31379 | 0.015801 | 0.4937 | *CYP2D6* |
| ENSG00000169189 | 16 | 27226312 | 27290115 | 166 | 22 | 42316 | 0.015415 | 0.49385 | *NSMCE1* |
| ENSG00000006788 | 17 | 10191401 | 10286447 | 334 | 49 | 41824 | 0.015422 | 0.49385 | *MYH13* |
| ENSG00000005810 | 13 | 77608792 | 77911185 | 657 | 59 | 42314 | 0.015363 | 0.49387 | *MYCBP2* |
| ENSG00000261611 | 16 | 71471503 | 71533214 | 125 | 21 | 39051 | 0.01509 | 0.49398 | *AC010547.9* |
| ENSG00000122711 | 9 | 33208363 | 33258565 | 179 | 18 | 33817 | 0.014978 | 0.49402 | *SPINK4* |
| ENSG00000101353 | 20 | 35719629 | 35817991 | 289 | 26 | 50473 | 0.014712 | 0.49413 | *MROH8* |
| ENSG00000076258 | 1 | 171273347 | 171321223 | 101 | 17 | 39450 | 0.014531 | 0.4942 | *FMO4* |
| ENSG00000004809 | 6 | 110735890 | 110807844 | 365 | 56 | 42944 | 0.01447 | 0.49423 | *SLC22A16* |
| ENSG00000088305 | 20 | 31340191 | 31407162 | 220 | 21 | 33074 | 0.014413 | 0.49425 | *DNMT3B* |
| ENSG00000143450 | 1 | 151725445 | 151753808 | 81 | 11 | 46516 | 0.014353 | 0.49427 | *OAZ3* |
| ENSG00000183735 | 12 | 64835660 | 64905888 | 163 | 24 | 46053 | 0.014336 | 0.49428 | *TBK1* |
| ENSG00000064886 | 1 | 111733393 | 111796062 | 239 | 33 | 48570 | 0.01418 | 0.49434 | *CHI3L2* |
| ENSG00000257390 | 12 | 56141059 | 56231330 | 100 | 25 | 40918 | 0.014178 | 0.49434 | *RP11-762I7.5* |
| ENSG00000185176 | 2 | 241605835 | 241632323 | 105 | 17 | 50210 | 0.014079 | 0.49438 | *AQP12B* |
| ENSG00000132481 | 17 | 73860242 | 73884656 | 80 | 12 | 37491 | 0.014072 | 0.49439 | *TRIM47* |
| ENSG00000100304 | 22 | 43552628 | 43593139 | 231 | 37 | 46841 | 0.013951 | 0.49443 | *TTLL12* |
| ENSG00000010270 | 7 | 38207824 | 38280272 | 254 | 28 | 54915 | 0.013812 | 0.49449 | *STARD3NL* |
| ENSG00000157211 | 1 | 54588747 | 54629443 | 170 | 33 | 46433 | 0.013565 | 0.49459 | *CDCP2* |
| ENSG00000153201 | 2 | 109325937 | 109412267 | 161 | 12 | 45116 | 0.013499 | 0.49461 | *RANBP2* |
| ENSG00000116985 | 1 | 40212854 | 40264533 | 250 | 31 | 50753 | 0.01306 | 0.49479 | *BMP8B* |
| ENSG00000197721 | 1 | 207808458 | 207921761 | 515 | 43 | 42335 | 0.012976 | 0.49482 | *CR1L* |
| ENSG00000188816 | 10 | 124897638 | 124920188 | 41 | 9 | 48927 | 0.012694 | 0.49494 | *HMX2* |
| ENSG00000127928 | 7 | 93210885 | 93550577 | 1040 | 95 | 39183 | 0.012515 | 0.49501 | *GNGT1* |
| ENSG00000196277 | 3 | 6801688 | 7793215 | 3584 | 309 | 42870 | 0.012306 | 0.49509 | *GRM7* |
| ENSG00000066926 | 18 | 55205515 | 55264004 | 204 | 45 | 49502 | 0.012304 | 0.49509 | *FECH* |
| ENSG00000167281 | 17 | 77075427 | 77623550 | 1996 | 329 | 45461 | 0.012184 | 0.49514 | *RBFOX3* |
| ENSG00000165282 | 9 | 35078685 | 35106591 | 54 | 11 | 41986 | 0.012157 | 0.49515 | *PIGO* |
| ENSG00000169919 | 7 | 65415671 | 65457301 | 100 | 13 | 36610 | 0.012034 | 0.4952 | *GUSB* |
| ENSG00000084444 | 12 | 13187218 | 13305455 | 248 | 33 | 34513 | 0.011886 | 0.49526 | *KIAA1467* |
| ENSG00000166866 | 12 | 57412301 | 57454982 | 91 | 11 | 50601 | 0.011851 | 0.49527 | *MYO1A* |
| ENSG00000167094 | 9 | 130468345 | 130503879 | 71 | 12 | 46878 | 0.011753 | 0.49531 | *TTC16* |
| ENSG00000167994 | 11 | 61654773 | 61697741 | 107 | 25 | 34594 | 0.011763 | 0.49531 | *RAB3IL1* |
| ENSG00000120094 | 17 | 46595888 | 46618359 | 67 | 12 | 39268 | 0.011758 | 0.49531 | *HOXB1* |
| ENSG00000163736 | 4 | 74842755 | 74863914 | 55 | 9 | 30639 | 0.011696 | 0.49533 | *PPBP* |

| | | | | | | | | | |
|---|---|---|---|---|---|---|---|---|---|
| ENSG00000133063 | 1 | 203171955 | 203252769 | 254 | 39 | 38630 | 0.011675 | 0.49534 | *CHIT1* |
| ENSG00000156564 | 6 | 40349325 | 40565204 | 911 | 95 | 45163 | 0.011496 | 0.49541 | *LRFN2* |
| ENSG00000196642 | 9 | 139692374 | 139745639 | 82 | 13 | 50492 | 0.011334 | 0.49548 | *RABL6* |
| ENSG00000167656 | 8 | 143856296 | 143878008 | 124 | 12 | 43381 | 0.011282 | 0.4955 | *LY6D* |
| ENSG00000196247 | 7 | 64116511 | 64181404 | 200 | 26 | 48987 | 0.01121 | 0.49553 | *ZNF107* |
| ENSG00000117133 | 1 | 84934942 | 84973473 | 190 | 25 | 41870 | 0.010983 | 0.49562 | *RPF1* |
| ENSG00000215186 | 15 | 72937079 | 72968735 | 6 | 3 | 23100 | 0.01099 | 0.49562 | *GOLGA6B* |
| ENSG00000228672 | 5 | 138717635 | 138740885 | 29 | 5 | 26258 | 0.010903 | 0.49565 | *PROB1* |
| ENSG00000057704 | 12 | 94950900 | 95054338 | 435 | 76 | 44996 | 0.010764 | 0.49571 | *TMCC3* |
| ENSG00000166889 | 11 | 59394189 | 59446453 | 83 | 13 | 34888 | 0.010424 | 0.49584 | *PATL1* |
| ENSG00000166986 | 12 | 57859228 | 57921352 | 113 | 15 | 43303 | 0.010104 | 0.49597 | *MARS* |
| ENSG00000204147 | 10 | 52489078 | 52586255 | 257 | 20 | 31084 | 0.010079 | 0.49598 | *ASAH2B* |
| ENSG00000110171 | 11 | 6459843 | 6505689 | 88 | 20 | 49014 | 0.0099336 | 0.49604 | *TRIM3* |
| ENSG00000133475 | 22 | 21552262 | 21591926 | 8 | 6 | 39006 | 0.0098798 | 0.49606 | *GGT2* |
| ENSG00000106799 | 9 | 101856320 | 101926474 | 150 | 19 | 54071 | 0.0098281 | 0.49608 | *TGFBR1* |
| ENSG00000110958 | 12 | 57047127 | 57092159 | 125 | 21 | 44901 | 0.0098368 | 0.49608 | *PTGES3* |
| ENSG00000180257 | 19 | 53420388 | 53476164 | 224 | 40 | 38703 | 0.0098291 | 0.49608 | *ZNF816* |
| ENSG00000168329 | 3 | 39294985 | 39333226 | 158 | 35 | 48396 | 0.0097304 | 0.49612 | *CX3CR1* |
| ENSG00000188076 | 11 | 183080 | 204573 | 44 | 13 | 41976 | 0.0097362 | 0.49612 | *SCGB1C1* |
| ENSG00000185404 | 2 | 231181899 | 231278447 | 444 | 45 | 44002 | 0.009631 | 0.49616 | *SP140L* |
| ENSG00000129968 | 19 | 1866809 | 1895546 | 63 | 7 | 44997 | 0.0096181 | 0.49616 | *ABHD17A* |
| ENSG00000188643 | 1 | 153569362 | 153595621 | 63 | 17 | 40802 | 0.0095423 | 0.49619 | *S100A16* |
| ENSG00000163378 | 3 | 69014365 | 69073112 | 175 | 21 | 43222 | 0.0092142 | 0.49632 | *EOGT* |
| ENSG00000251184 | 9 | 131693757 | 131729300 | 53 | 7 | 49678 | 0.0091454 | 0.49635 | *RP11-101E3.5* |
| ENSG00000185942 | 8 | 63151150 | 63922211 | 2611 | 156 | 44998 | 0.0090701 | 0.49638 | *NKAIN3* |
| ENSG00000099364 | 16 | 30924376 | 30970104 | 52 | 7 | 22160 | 0.0089759 | 0.49642 | *FBXL19* |
| ENSG00000147650 | 8 | 105491459 | 105611252 | 240 | 26 | 45683 | 0.0084756 | 0.49662 | *LRP12* |
| ENSG00000162065 | 16 | 2515147 | 2565735 | 43 | 27 | 45346 | 0.0083207 | 0.49668 | *TBC1D24* |
| ENSG00000150776 | 11 | 111934810 | 111965874 | 42 | 12 | 34168 | 0.0081074 | 0.49677 | *C11orf57* |
| ENSG00000184108 | 4 | 189050573 | 189078897 | 200 | 34 | 32489 | 0.0080631 | 0.49678 | *TRIML1* |
| ENSG00000137962 | 1 | 94604544 | 94750624 | 305 | 43 | 51181 | 0.0080279 | 0.4968 | *ARHGAP29* |
| ENSG00000176714 | 2 | 27838506 | 27861879 | 63 | 12 | 49040 | 0.0079957 | 0.49681 | *CCDC121* |
| ENSG00000249209 | 21 | 34946993 | 35294635 | 711 | 118 | 47442 | 0.0079881 | 0.49681 | *AP000304.12* |
| ENSG00000204176 | 10 | 46945444 | 46981400 | 6 | 4 | 27763 | 0.0079479 | 0.49683 | *SYT15* |
| ENSG00000138293 | 10 | 51555108 | 51600734 | 49 | 14 | 47506 | 0.0078768 | 0.49686 | *NCOA4* |
| ENSG00000100867 | 14 | 24089324 | 24124848 | 50 | 24 | 45589 | 0.0078331 | 0.49688 | *DHRS2* |

| | | | | | | | | | |
|---|---|---|---|---|---|---|---|---|---|
| ENSG00000143674 | 1 | 233453514 | 233530894 | 334 | 31 | 54087 | 0.0076733 | 0.49694 | *MLK4* |
| ENSG00000175161 | 3 | 84998132 | 86133579 | 3365 | 116 | 35444 | 0.0076137 | 0.49696 | *CADM2* |
| ENSG00000260027 | 17 | 46674594 | 46720934 | 156 | 29 | 34606 | 0.0075808 | 0.49698 | *HOXB7* |
| ENSG00000141644 | 18 | 47783252 | 47818144 | 74 | 14 | 53992 | 0.0072292 | 0.49712 | *MBD1* |
| ENSG00000110955 | 12 | 57021959 | 57049798 | 57 | 12 | 45747 | 0.0070715 | 0.49718 | *ATP5B* |
| ENSG00000180228 | 2 | 179286141 | 179326239 | 65 | 12 | 35632 | 0.0069152 | 0.49724 | *PRKRA* |
| ENSG00000109189 | 4 | 53447138 | 53535502 | 114 | 10 | 56176 | 0.006718 | 0.49732 | *USP46* |
| ENSG00000255346 | 15 | 69212864 | 69365083 | 253 | 28 | 38817 | 0.0065862 | 0.49737 | *NOX5* |
| ENSG00000088053 | 19 | 55515073 | 55559632 | 291 | 33 | 47994 | 0.0062059 | 0.49752 | *GP6* |
| ENSG00000146350 | 6 | 121390640 | 121665891 | 1230 | 44 | 40971 | 0.0060638 | 0.49758 | *TBC1D32* |
| ENSG00000177181 | 1 | 42836466 | 42898249 | 215 | 15 | 52744 | 0.0060189 | 0.4976 | *RIMKLA* |
| ENSG00000174343 | 4 | 40327346 | 40367234 | 224 | 22 | 47048 | 0.0053449 | 0.49787 | *CHRNA9* |
| ENSG00000082898 | 2 | 61694984 | 61775761 | 179 | 18 | 52044 | 0.0053222 | 0.49788 | *XPO1* |
| ENSG00000034152 | 17 | 21177984 | 21228552 | 76 | 18 | 44747 | 0.0052343 | 0.49791 | *MAP2K3* |
| ENSG00000181631 | 3 | 151034100 | 151057336 | 82 | 18 | 55457 | 0.005211 | 0.49792 | *P2RY13* |
| ENSG00000242019 | 19 | 55225984 | 55258003 | 47 | 17 | 36659 | 0.0050973 | 0.49797 | *KIR3DL3* |
| ENSG00000268500 | 19 | 52105344 | 52160142 | 185 | 46 | 44502 | 0.0049915 | 0.49801 | *SIGLEC5* |
| ENSG00000224132 | 2 | 238155734 | 238176319 | 33 | 7 | 37137 | 0.0046651 | 0.49814 | *AC112715.2* |
| ENSG00000153786 | 16 | 84997787 | 85055141 | 313 | 34 | 39869 | 0.0044114 | 0.49824 | *ZDHHC7* |
| ENSG00000215131 | 16 | 3533484 | 3555480 | 102 | 19 | 38869 | 0.0043499 | 0.49826 | *C16orf90* |
| ENSG00000151623 | 4 | 148989913 | 149375850 | 1117 | 122 | 47134 | 0.0043064 | 0.49828 | *NR3C2* |
| ENSG00000175189 | 12 | 57818543 | 57854611 | 69 | 14 | 41509 | 0.0039238 | 0.49843 | *INHBC* |
| ENSG00000111231 | 12 | 110880289 | 110917073 | 106 | 14 | 34278 | 0.0038127 | 0.49848 | *GPN3* |
| ENSG00000159685 | 3 | 126413063 | 126689249 | 809 | 62 | 48934 | 0.0036837 | 0.49853 | *CHCHD6* |
| ENSG00000269417 | 2 | 85112209 | 85132345 | 71 | 4 | 31767 | 0.0035972 | 0.49856 | *AC022210.1* |
| ENSG00000110321 | 11 | 10808597 | 10840657 | 78 | 18 | 47322 | 0.0032128 | 0.49872 | *EIF4G2* |
| ENSG00000141503 | 17 | 4726683 | 4811356 | 250 | 29 | 43165 | 0.0031401 | 0.49875 | *MINK1* |
| ENSG00000159713 | 16 | 67413712 | 67437438 | 36 | 10 | 44032 | 0.0030242 | 0.49879 | *TPPP3* |
| ENSG00000183643 | 15 | 93004884 | 93054347 | 148 | 36 | 47831 | 0.0029698 | 0.49882 | *C15orf32* |
| ENSG00000012983 | 14 | 50875219 | 51037844 | 307 | 38 | 45293 | 0.002649 | 0.49894 | *MAP4K5* |
| ENSG00000269064 | 1 | 179433716 | 179467805 | 301 | 19 | 56141 | 0.0025295 | 0.49899 | *AL160286.1* |
| ENSG00000106344 | 7 | 127927738 | 127993962 | 162 | 28 | 45928 | 0.0024425 | 0.49903 | *RBM28* |
| ENSG00000103353 | 16 | 23558392 | 23595710 | 118 | 14 | 39202 | 0.0024251 | 0.49903 | *UBFD1* |
| ENSG00000112237 | 6 | 99980256 | 100026849 | 171 | 22 | 44264 | 0.0020875 | 0.49917 | *CCNC* |
| ENSG00000160791 | 3 | 46401633 | 46427697 | 84 | 14 | 33222 | 0.0017581 | 0.4993 | *CCR5* |
| ENSG00000180914 | 3 | 8782094 | 8821314 | 187 | 47 | 43575 | 0.0016032 | 0.49936 | *OXTR* |

| | | | | | | | | | |
|---|---|---|---|---|---|---|---|---|---|
| ENSG00000170122 | 9 | 106237 | 128417 | 18 | 3 | 37871 | 0.0016092 | 0.49936 | *FOXD4* |
| ENSG00000137312 | 6 | 30685486 | 30720510 | 195 | 16 | 23983 | 0.0015179 | 0.49939 | *FLOT1* |
| ENSG00000116212 | 1 | 54401750 | 54443841 | 80 | 15 | 36717 | 0.0014439 | 0.49942 | *LRRC42* |
| ENSG00000198400 | 1 | 156775432 | 156861642 | 195 | 42 | 43680 | 0.001399 | 0.49944 | *NTRK1* |
| ENSG00000108443 | 17 | 57960447 | 58037925 | 152 | 18 | 42397 | 0.0013262 | 0.49947 | *RPS6KB1* |
| ENSG00000242180 | 11 | 5353744 | 5374809 | 150 | 18 | 40528 | 0.0012686 | 0.49949 | *OR51B5* |
| ENSG00000153485 | 14 | 93641296 | 93663434 | 64 | 18 | 49932 | 0.0012312 | 0.49951 | *TMEM251* |
| ENSG00000248801 | 8 | 69205703 | 69253726 | 95 | 20 | 41857 | 0.0011855 | 0.49953 | *RP11-664D7.4* |
| ENSG00000269494 | 6 | 82252307 | 82272537 | 51 | 10 | 55011 | 0.0010907 | 0.49956 | *AL359693.1* |
| ENSG00000160654 | 11 | 118205059 | 118235876 | 76 | 12 | 36446 | 0.0010842 | 0.49957 | *CD3G* |
| ENSG00000187554 | 1 | 223272748 | 223326624 | 202 | 17 | 36936 | 0.0009799 | 0.49961 | *TLR5* |
| ENSG00000197044 | 19 | 11867815 | 11904893 | 132 | 13 | 48311 | 0.00098796 | 0.49961 | *ZNF441* |
| ENSG00000124177 | 20 | 40020741 | 40257133 | 583 | 43 | 41395 | 0.00095359 | 0.49962 | *CHD6* |
| ENSG00000112146 | 6 | 52906789 | 52975671 | 112 | 18 | 39732 | 0.00088146 | 0.49965 | *FBXO9* |
| ENSG00000106565 | 7 | 150478373 | 150508448 | 135 | 34 | 49554 | 0.00087464 | 0.49965 | *TMEM176B* |
| ENSG00000127578 | 16 | 669239 | 694116 | 71 | 12 | 51893 | 0.00073047 | 0.49971 | *WFIKKN1* |
| ENSG00000168137 | 3 | 9429299 | 9530924 | 218 | 28 | 45120 | 0.00070507 | 0.49972 | *SETD5* |
| ENSG00000168872 | 16 | 70370732 | 70417286 | 64 | 23 | 41207 | 0.00042304 | 0.49983 | *DDX19A* |
| ENSG00000163864 | 3 | 139269022 | 139406859 | 356 | 58 | 40696 | 0.00039323 | 0.49984 | *NMNAT3* |
| ENSG00000053702 | 12 | 2924514 | 2954710 | 78 | 9 | 25770 | 0.00019917 | 0.49992 | *NRIP2* |
| ENSG00000105501 | 19 | 52104781 | 52160151 | 187 | 46 | 44164 | -0.0001999 | 0.50008 | *SIGLEC5* |
| ENSG00000266997 | 18 | 47312163 | 47373993 | 171 | 21 | 44091 | -0.00042038 | 0.50017 | *RP11-886H22.1* |
| ENSG00000001084 | 6 | 53352139 | 53491768 | 491 | 86 | 46594 | -0.00048387 | 0.50019 | *GCLC* |
| ENSG00000221829 | 9 | 35063832 | 35090013 | 38 | 10 | 41365 | -0.00075841 | 0.5003 | *FANCG* |
| ENSG00000215397 | 20 | 632240 | 666823 | 93 | 22 | 44621 | -0.00074859 | 0.5003 | *SCRT2* |
| ENSG00000108523 | 17 | 4833303 | 4858517 | 63 | 18 | 39829 | -0.00077929 | 0.50031 | *RNF167* |
| ENSG00000214711 | 2 | 31385924 | 31466724 | 489 | 46 | 37532 | -0.00081277 | 0.50032 | *CAPN14* |
| ENSG00000156970 | 15 | 40443224 | 40523337 | 151 | 17 | 38930 | -0.00080553 | 0.50032 | *BUB1B* |
| ENSG00000115875 | 2 | 38960741 | 38988636 | 49 | 11 | 43294 | -0.00083405 | 0.50033 | *SRSF7* |
| ENSG00000257008 | 17 | 72353546 | 72378761 | 54 | 18 | 48929 | -0.0011248 | 0.50045 | *GPR142* |
| ENSG00000155034 | 7 | 5460966 | 5563429 | 457 | 65 | 50610 | -0.0015775 | 0.50063 | *FBXL18* |
| ENSG00000184560 | 17 | 7318934 | 7340887 | 48 | 15 | 41842 | -0.0016648 | 0.50066 | *C17orf74* |
| ENSG00000183784 | 9 | 203108 | 225893 | 142 | 20 | 53174 | -0.0016922 | 0.50068 | *C9orf66* |
| ENSG00000170178 | 2 | 176954458 | 176976408 | 54 | 12 | 38552 | -0.0017353 | 0.50069 | *HOXD12* |
| ENSG00000164299 | 5 | 79605444 | 79627661 | 110 | 16 | 37340 | -0.0017528 | 0.5007 | *SPZ1* |
| ENSG00000167461 | 19 | 16212439 | 16255044 | 146 | 16 | 49006 | -0.0022311 | 0.50089 | *RAB8A* |

| | | | | | | | | | |
|---|---|---|---|---|---|---|---|---|---|
| ENSG00000070526 | 17 | 74610843 | 74649920 | 168 | 21 | 40445 | -0.0027503 | 0.5011 | *ST6GALNAC1* |
| ENSG00000267918 | 14 | 101285066 | 101305537 | 89 | 22 | 49800 | -0.0028832 | 0.50115 | *AL117190.2* |
| ENSG00000087460 | 20 | 57404773 | 57496247 | 250 | 57 | 43302 | -0.0030371 | 0.50121 | *GNAS* |
| ENSG00000167360 | 11 | 5433341 | 5454436 | 175 | 19 | 39262 | -0.0036712 | 0.50146 | *OR51Q1* |
| ENSG00000197532 | 1 | 158506918 | 158527895 | 97 | 12 | 52505 | -0.0036871 | 0.50147 | *OR6Y1* |
| ENSG00000135736 | 16 | 57536090 | 57580511 | 185 | 33 | 45520 | -0.0038025 | 0.50152 | *CCDC102A* |
| ENSG00000120156 | 9 | 27099139 | 27240173 | 862 | 112 | 45396 | -0.0039889 | 0.50159 | *TEK* |
| ENSG00000132507 | 17 | 7200318 | 7225774 | 81 | 16 | 50505 | -0.0042181 | 0.50168 | *EIF5A* |
| ENSG00000188674 | 2 | 209020067 | 209064797 | 203 | 35 | 40199 | -0.0049511 | 0.50198 | *C2orf80* |
| ENSG00000157540 | 21 | 38728092 | 38899753 | 505 | 41 | 54701 | -0.0050044 | 0.502 | *DYRK1A* |
| ENSG00000138459 | 3 | 112270556 | 112314424 | 135 | 14 | 50417 | -0.0050391 | 0.50201 | *SLC35A5* |
| ENSG00000125848 | 20 | 14293634 | 14328262 | 91 | 20 | 46521 | -0.0051232 | 0.50204 | *FLRT3* |
| ENSG00000162643 | 1 | 85454830 | 85608821 | 529 | 61 | 48692 | -0.0051476 | 0.50205 | *WDR63* |
| ENSG00000268427 | 22 | 50704103 | 50724350 | 98 | 24 | 48282 | -0.0055573 | 0.50222 | *AL022328.1* |
| ENSG00000164794 | 8 | 110965874 | 110998076 | 68 | 13 | 35231 | -0.0055997 | 0.50223 | *KCNV1* |
| ENSG00000124383 | 2 | 71347444 | 71387231 | 157 | 22 | 45461 | -0.0056045 | 0.50224 | *MPHOSPH10* |
| ENSG00000100060 | 22 | 37855101 | 37892439 | 58 | 12 | 44635 | -0.0058004 | 0.50231 | *MFNG* |
| ENSG00000125046 | 3 | 8651086 | 8796726 | 530 | 130 | 44430 | -0.005806 | 0.50232 | *SSUH2* |
| ENSG00000168264 | 1 | 234730015 | 234755271 | 95 | 22 | 42457 | -0.0058327 | 0.50233 | *IRF2BP2* |
| ENSG00000104714 | 8 | 554746 | 698106 | 851 | 82 | 45860 | -0.0058845 | 0.50235 | *ERICH1* |
| ENSG00000122548 | 7 | 26562740 | 26588407 | 147 | 19 | 46806 | -0.006109 | 0.50244 | *KIAA0087* |
| ENSG00000165669 | 10 | 120055401 | 120111840 | 99 | 28 | 48109 | -0.0061414 | 0.50245 | *FAM204A* |
| ENSG00000104067 | 15 | 29981571 | 30271068 | 804 | 102 | 49386 | -0.0062145 | 0.50248 | *TJP1* |
| ENSG00000183426 | 16 | 15006659 | 15055915 | 17 | 8 | 43332 | -0.0064229 | 0.50256 | *NPIPA1* |
| ENSG00000258947 | 16 | 89977800 | 90015169 | 186 | 30 | 29588 | -0.0064354 | 0.50257 | *TUBB3* |
| ENSG00000144015 | 2 | 96247766 | 96275526 | 21 | 13 | 44126 | -0.0065 | 0.50259 | *TRIM43* |
| ENSG00000177301 | 1 | 111126202 | 111184096 | 148 | 28 | 33874 | -0.0068642 | 0.50274 | *KCNA2* |
| ENSG00000131503 | 5 | 139771399 | 139939163 | 182 | 27 | 50142 | -0.0068753 | 0.50274 | *ANKHD1* |
| ENSG00000254996 | 5 | 139771493 | 139939154 | 182 | 27 | 50142 | -0.0068753 | 0.50274 | *ANKHD1-EIF4EBP3* |
| ENSG00000121690 | 11 | 33027410 | 33065128 | 191 | 11 | 53671 | -0.0073508 | 0.50293 | *DEPDC7* |
| ENSG00000134249 | 1 | 120426156 | 120449118 | 46 | 12 | 32645 | -0.0074048 | 0.50295 | *ADAM30* |
| ENSG00000167380 | 19 | 44659226 | 44692534 | 35 | 11 | 43107 | -0.0074226 | 0.50296 | *ZNF226* |
| ENSG00000244362 | 21 | 31923194 | 31943633 | 66 | 7 | 54828 | -0.0076076 | 0.50303 | *KRTAP19-7* |
| ENSG00000140740 | 16 | 21953981 | 22004981 | 51 | 14 | 36818 | -0.0078062 | 0.50311 | *UQCRC2* |
| ENSG00000168502 | 18 | 8695659 | 8842776 | 539 | 101 | 45543 | -0.0078324 | 0.50312 | *SOGA2* |
| ENSG00000117115 | 1 | 17383256 | 17455948 | 347 | 27 | 49573 | -0.007863 | 0.50314 | *PADI2* |

| | | | | | | | | | |
|---|---|---|---|---|---|---|---|---|---|
| ENSG00000118518 | 6 | 127577755 | 127619712 | 83 | 13 | 48922 | -0.0078902 | 0.50315 | *RNF146* |
| ENSG00000135749 | 1 | 233109181 | 233441459 | 1273 | 125 | 45776 | -0.008112 | 0.50324 | *PCNXL2* |
| ENSG00000136694 | 2 | 113753038 | 113775621 | 46 | 10 | 39775 | -0.0088664 | 0.50354 | *IL36A* |
| ENSG00000221978 | 1 | 1311091 | 1344708 | 76 | 10 | 33898 | -0.00891 | 0.50355 | *CCNL2* |
| ENSG00000145349 | 4 | 114362188 | 114693083 | 906 | 99 | 43653 | -0.0089259 | 0.50356 | *CAMK2D* |
| ENSG00000138075 | 2 | 44029611 | 44076004 | 181 | 24 | 37060 | -0.0090108 | 0.50359 | *ABCG5* |
| ENSG00000226490 | 8 | 142514738 | 142538837 | 118 | 22 | 34707 | -0.0090632 | 0.50362 | *AC138647.1* |
| ENSG00000253459 | 14 | 50054193 | 50075882 | 74 | 9 | 47618 | -0.0090771 | 0.50362 | *AL139099.1* |
| ENSG00000103343 | 16 | 3441235 | 3469370 | 70 | 14 | 51170 | -0.0091182 | 0.50364 | *ZNF174* |
| ENSG00000237247 | 19 | 7020589 | 7043022 | 6 | 3 | 38165 | -0.0092244 | 0.50368 | *MBD3L5* |
| ENSG00000254995 | 20 | 57216490 | 57300466 | 261 | 37 | 43843 | -0.0092931 | 0.50371 | *STX16-NPEPL1* |
| ENSG00000182600 | 2 | 233711980 | 233753418 | 152 | 20 | 49038 | -0.0093415 | 0.50373 | *C2orf82* |
| ENSG00000161980 | 16 | 86407 | 113628 | 109 | 15 | 48648 | -0.0093766 | 0.50374 | *POLR3K* |
| ENSG00000090989 | 4 | 56709782 | 56781200 | 208 | 31 | 50129 | -0.0095376 | 0.5038 | *EXOC1* |
| ENSG00000157680 | 7 | 137055783 | 137541838 | 906 | 117 | 36922 | -0.0095236 | 0.5038 | *DGKI* |
| ENSG00000112394 | 6 | 111398781 | 111562397 | 340 | 37 | 46659 | -0.010053 | 0.50401 | *SLC16A10* |
| ENSG00000268550 | 19 | 14824747 | 14845121 | 84 | 17 | 50085 | -0.010049 | 0.50401 | *AC090427.1* |
| ENSG00000159164 | 1 | 149864870 | 149899434 | 72 | 11 | 46744 | -0.010119 | 0.50404 | *SV2A* |
| ENSG00000124839 | 2 | 238472965 | 238520257 | 311 | 42 | 35466 | -0.010209 | 0.50407 | *RAB17* |
| ENSG00000124181 | 20 | 39755600 | 39821629 | 95 | 20 | 46857 | -0.010252 | 0.50409 | *PLCG1* |
| ENSG00000166265 | 21 | 27828528 | 27955603 | 390 | 83 | 44253 | -0.01033 | 0.50412 | *CYYR1* |
| ENSG00000144959 | 3 | 172338039 | 172439008 | 322 | 60 | 45943 | -0.010398 | 0.50415 | *NCEH1* |
| ENSG00000134013 | 8 | 23144702 | 23292841 | 867 | 89 | 41877 | -0.010474 | 0.50418 | *LOXL2* |
| ENSG00000173480 | 19 | 58401664 | 58437978 | 108 | 15 | 48001 | -0.010476 | 0.50418 | *ZNF417* |
| ENSG00000182389 | 2 | 152679290 | 152965593 | 611 | 67 | 43693 | -0.01089 | 0.50434 | *CACNB4* |
| ENSG00000176246 | 14 | 20518204 | 20539142 | 149 | 15 | 41225 | -0.010896 | 0.50435 | *OR4L1* |
| ENSG00000121848 | 1 | 145601036 | 145706218 | 88 | 12 | 44259 | -0.010925 | 0.50436 | *RNF115* |
| ENSG00000176904 | 11 | 4870886 | 4891794 | 62 | 11 | 43464 | -0.011137 | 0.50444 | *OR51H1P* |
| ENSG00000184702 | 22 | 19691987 | 19722295 | 42 | 18 | 47133 | -0.011172 | 0.50446 | *Sep-05* |
| ENSG00000100813 | 14 | 23517773 | 23574823 | 165 | 27 | 49243 | -0.011462 | 0.50457 | *ACIN1* |
| ENSG00000261884 | 16 | 67953517 | 67979920 | 23 | 6 | 39503 | -0.011566 | 0.50461 | *CTC-479C5.12* |
| ENSG00000152749 | 13 | 95244157 | 95296899 | 123 | 15 | 42093 | -0.011662 | 0.50465 | *GPR180* |
| ENSG00000129277 | 17 | 34420983 | 34443014 | 108 | 12 | 46595 | -0.011873 | 0.50474 | *CCL4* |
| ENSG00000197629 | 11 | 58965983 | 58990424 | 65 | 11 | 53135 | -0.01262 | 0.50503 | *MPEG1* |
| ENSG00000125650 | 19 | 6365159 | 6389069 | 101 | 14 | 41654 | -0.012626 | 0.50504 | *PSPN* |
| ENSG00000175471 | 5 | 94029446 | 94630279 | 1666 | 202 | 48924 | -0.012677 | 0.50506 | *MCTP1* |

| | | | | | | | | | |
|---|---|---|---|---|---|---|---|---|---|
| ENSG00000141552 | 17 | 79838666 | 79868867 | 29 | 16 | 50628 | -0.013197 | 0.50526 | *ANAPC11* |
| ENSG00000138483 | 3 | 107086188 | 107107481 | 56 | 14 | 34404 | -0.013314 | 0.50531 | *CCDC54* |
| ENSG00000205336 | 16 | 57634564 | 57708944 | 340 | 82 | 44959 | -0.013465 | 0.50537 | *GPR56* |
| ENSG00000130714 | 9 | 134368289 | 134409193 | 127 | 11 | 30129 | -0.013733 | 0.50548 | *POMT1* |
| ENSG00000120733 | 5 | 137678285 | 137782717 | 267 | 28 | 44398 | -0.013771 | 0.50549 | *KDM3B* |
| ENSG00000171608 | 1 | 9701790 | 9799172 | 178 | 39 | 36447 | -0.013828 | 0.50552 | *PIK3CD* |
| ENSG00000089053 | 12 | 121736048 | 121847699 | 302 | 43 | 35759 | -0.014125 | 0.50563 | *ANAPC5* |
| ENSG00000106683 | 7 | 73487263 | 73546855 | 126 | 18 | 40345 | -0.014592 | 0.50582 | *LIMK1* |
| ENSG00000108557 | 17 | 17574787 | 17724767 | 227 | 39 | 38043 | -0.014845 | 0.50592 | *RAI1* |
| ENSG00000161609 | 19 | 49881475 | 49931251 | 164 | 15 | 53390 | -0.015092 | 0.50602 | *CCDC155* |
| ENSG00000133460 | 22 | 24188890 | 24238496 | 182 | 37 | 49209 | -0.015082 | 0.50602 | *SLC2A11* |
| ENSG00000006638 | 19 | 3584504 | 3616838 | 138 | 26 | 41398 | -0.015235 | 0.50608 | *TBXA2R* |
| ENSG00000135899 | 2 | 231022009 | 231100444 | 373 | 42 | 49253 | -0.015263 | 0.50609 | *SP110* |
| ENSG00000268107 | 19 | 57989086 | 58026749 | 224 | 11 | 52387 | -0.015274 | 0.50609 | *AC003005.4* |
| ENSG00000140557 | 15 | 92927058 | 93021958 | 293 | 52 | 41471 | -0.015331 | 0.50612 | *ST8SIA2* |
| ENSG00000131634 | 16 | 1568689 | 1615581 | 180 | 21 | 40781 | -0.015435 | 0.50616 | *TMEM204* |
| ENSG00000198019 | 1 | 120916979 | 120945937 | 10 | 4 | 43446 | -0.015461 | 0.50617 | *FCGR1B* |
| ENSG00000096746 | 10 | 70080931 | 70112948 | 87 | 14 | 39463 | -0.016222 | 0.50647 | *HNRNPH3* |
| ENSG00000262874 | 19 | 51881543 | 51903828 | 81 | 9 | 27677 | -0.016221 | 0.50647 | *C19orf84* |
| ENSG00000165061 | 8 | 40378109 | 40765352 | 1339 | 137 | 44539 | -0.01652 | 0.50659 | *ZMAT4* |
| ENSG00000116957 | 1 | 235520675 | 235622283 | 325 | 31 | 39189 | -0.016542 | 0.5066 | *TBCE* |
| ENSG00000044012 | 1 | 42609092 | 42631495 | 55 | 13 | 45116 | -0.016781 | 0.50669 | *GUCA2B* |
| ENSG00000163872 | 3 | 183405606 | 183540413 | 314 | 25 | 50833 | -0.01685 | 0.50672 | *YEATS2* |
| ENSG00000147853 | 9 | 4701155 | 4752043 | 251 | 33 | 36865 | -0.017012 | 0.50679 | *AK3* |
| ENSG00000267534 | 19 | 10324520 | 10351948 | 61 | 16 | 47263 | -0.017074 | 0.50681 | *S1PR2* |
| ENSG00000125835 | 20 | 2432280 | 2461499 | 64 | 12 | 47287 | -0.017379 | 0.50693 | *SNRPB* |
| ENSG00000135953 | 2 | 103322299 | 103363347 | 90 | 14 | 47791 | -0.018155 | 0.50724 | *MFSD9* |
| ENSG00000169504 | 1 | 25061848 | 25180815 | 160 | 20 | 44403 | -0.018377 | 0.50733 | *CLIC4* |
| ENSG00000027847 | 5 | 177017101 | 177047348 | 105 | 19 | 43340 | -0.018397 | 0.50734 | *B4GALT7* |
| ENSG00000269067 | 19 | 23148270 | 23195978 | 158 | 8 | 29615 | -0.019025 | 0.50759 | *ZNF728* |
| ENSG00000113719 | 5 | 172251278 | 172389688 | 445 | 91 | 46782 | -0.019113 | 0.50762 | *ERGIC1* |
| ENSG00000251258 | 6 | 112658532 | 112682498 | 65 | 13 | 45172 | -0.019182 | 0.50765 | *RFPL4B* |
| ENSG00000243943 | 2 | 27795897 | 27868041 | 160 | 21 | 48505 | -0.019353 | 0.50772 | *ZNF512* |
| ENSG00000158458 | 5 | 139216364 | 139432884 | 410 | 59 | 42031 | -0.019365 | 0.50773 | *NRG2* |
| ENSG00000107829 | 10 | 103360423 | 103465052 | 166 | 20 | 53620 | -0.01944 | 0.50775 | *FBXW4* |
| ENSG00000103184 | 16 | 4998318 | 5079159 | 404 | 47 | 41254 | -0.019561 | 0.5078 | *SEC14L5* |

| | | | | | | | | | |
|---|---|---|---|---|---|---|---|---|---|
| ENSG00000004139 | 17 | 26681378 | 26738065 | 112 | 13 | 35725 | -0.020048 | 0.508 | *SARM1* |
| ENSG00000128563 | 7 | 101994319 | 102077123 | 212 | 22 | 54292 | -0.020128 | 0.50803 | *PRKRIP1* |
| ENSG00000159263 | 21 | 38061433 | 38132218 | 192 | 62 | 47084 | -0.020402 | 0.50814 | *SIM2* |
| ENSG00000167767 | 12 | 52552780 | 52595784 | 190 | 32 | 48654 | -0.020501 | 0.50818 | *KRT80* |
| ENSG00000206384 | 3 | 130269178 | 130406999 | 255 | 42 | 38083 | -0.020685 | 0.50825 | *COL6A6* |
| ENSG00000135424 | 12 | 56068352 | 56119827 | 98 | 16 | 30742 | -0.020832 | 0.50831 | *ITGA7* |
| ENSG00000131471 | 17 | 40993201 | 41020147 | 20 | 13 | 53129 | -0.020893 | 0.50833 | *AOC3* |
| ENSG00000164185 | 5 | 121455208 | 121525312 | 117 | 31 | 39196 | -0.021514 | 0.50858 | *ZNF474* |
| ENSG00000142765 | 1 | 27658513 | 27690421 | 96 | 26 | 42248 | -0.021554 | 0.5086 | *SYTL1* |
| ENSG00000147408 | 8 | 19251672 | 19625540 | 1649 | 167 | 50132 | -0.02172 | 0.50866 | *CSGALNACT1* |
| ENSG00000140543 | 15 | 89044790 | 89099906 | 184 | 16 | 44793 | -0.021741 | 0.50867 | *DET1* |
| ENSG00000184814 | 3 | 138727873 | 138749768 | 43 | 10 | 42423 | -0.022123 | 0.50882 | *PRR23B* |
| ENSG00000122378 | 10 | 82157585 | 82202753 | 174 | 24 | 34704 | -0.02212 | 0.50882 | *FAM213A* |
| ENSG00000136891 | 9 | 103054359 | 103125221 | 185 | 16 | 35756 | -0.022338 | 0.50891 | *TEX10* |
| ENSG00000026652 | 6 | 161541011 | 161705093 | 537 | 90 | 44752 | -0.022662 | 0.50904 | *AGPAT4* |
| ENSG00000168309 | 3 | 58539844 | 58623337 | 329 | 53 | 41019 | -0.022675 | 0.50905 | *FAM107A* |
| ENSG00000140471 | 15 | 101089574 | 101153435 | 347 | 35 | 49331 | -0.022816 | 0.5091 | *LINS* |
| ENSG00000136099 | 13 | 53408109 | 53432775 | 64 | 15 | 48977 | -0.0229 | 0.50913 | *PCDH8* |
| ENSG00000206474 | 6 | 29397083 | 29418731 | 136 | 10 | 21712 | -0.023059 | 0.5092 | *OR10C1* |
| ENSG00000135469 | 12 | 56650642 | 56674750 | 35 | 6 | 55115 | -0.023059 | 0.5092 | *COQ10A* |
| ENSG00000267970 | 7 | 48877362 | 48901221 | 123 | 21 | 53046 | -0.023185 | 0.50925 | *AC004899.1* |
| ENSG00000204564 | 6 | 30604816 | 30630987 | 51 | 13 | 29686 | -0.023282 | 0.50929 | *C6orf136* |
| ENSG00000090266 | 7 | 140380577 | 140432590 | 99 | 14 | 33256 | -0.023478 | 0.50937 | *NDUFB2* |
| ENSG00000139233 | 12 | 66506842 | 66534548 | 52 | 10 | 44705 | -0.023502 | 0.50938 | *LLPH* |
| ENSG00000143921 | 2 | 44056103 | 44115605 | 398 | 24 | 46542 | -0.023592 | 0.50941 | *ABCG8* |
| ENSG00000154265 | 17 | 67230452 | 67333385 | 259 | 32 | 50978 | -0.023885 | 0.50953 | *ABCA5* |
| ENSG00000100359 | 22 | 40756595 | 40816293 | 100 | 16 | 48684 | -0.024088 | 0.50961 | *SGSM3* |
| ENSG00000117280 | 1 | 205727114 | 205754588 | 103 | 16 | 32643 | -0.024598 | 0.50981 | *RAB7L1* |
| ENSG00000108100 | 10 | 35525953 | 35870852 | 609 | 48 | 49295 | -0.024597 | 0.50981 | *CCNY* |
| ENSG00000268412 | 2 | 26241074 | 26261481 | 51 | 13 | 52289 | -0.024679 | 0.50984 | *AC013449.1* |
| ENSG00000068697 | 2 | 20222411 | 20261789 | 137 | 21 | 33027 | -0.025114 | 0.51002 | *LAPTM4A* |
| ENSG00000143554 | 1 | 153736830 | 153762633 | 52 | 16 | 37871 | -0.025594 | 0.51021 | *SLC27A3* |
| ENSG00000110848 | 12 | 9895082 | 9923497 | 151 | 18 | 49387 | -0.025606 | 0.51021 | *CD69* |
| ENSG00000161677 | 19 | 50999255 | 51024610 | 73 | 18 | 41957 | -0.025877 | 0.51032 | *JOSD2* |
| ENSG00000161573 | 17 | 34293529 | 34318532 | 110 | 21 | 43146 | -0.025996 | 0.51037 | *CCL16* |
| ENSG00000161103 | 22 | 18824324 | 18858562 | 8 | 4 | 42823 | -0.025992 | 0.51037 | *AC008132.13* |

| | | | | | | | | | |
|---|---|---|---|---|---|---|---|---|---|
| ENSG00000173041 | 7 | 63970262 | 64033484 | 254 | 16 | 51570 | -0.026054 | 0.51039 | *ZNF680* |
| ENSG00000091844 | 6 | 153315594 | 153462384 | 525 | 40 | 40208 | -0.026063 | 0.5104 | *RGS17* |
| ENSG00000188295 | 1 | 247253264 | 247277674 | 120 | 20 | 40205 | -0.026435 | 0.51054 | *ZNF669* |
| ENSG00000140279 | 15 | 45374848 | 45416542 | 73 | 16 | 41013 | -0.026472 | 0.51056 | *DUOX2* |
| ENSG00000065621 | 10 | 106018631 | 106074703 | 96 | 20 | 44822 | -0.026526 | 0.51058 | *GSTO2* |
| ENSG00000169214 | 1 | 247865045 | 247886105 | 70 | 14 | 42310 | -0.026635 | 0.51062 | *OR6F1* |
| ENSG00000105370 | 19 | 51873163 | 51901214 | 95 | 12 | 28420 | -0.026631 | 0.51062 | *LIM2* |
| ENSG00000162736 | 1 | 160303062 | 160338742 | 93 | 21 | 33458 | -0.026678 | 0.51064 | *NCSTN* |
| ENSG00000172137 | 16 | 71382616 | 71434341 | 173 | 25 | 45814 | -0.026694 | 0.51065 | *CALB2* |
| ENSG00000158516 | 7 | 129896667 | 129939638 | 104 | 19 | 42365 | -0.026735 | 0.51066 | *CPA2* |
| ENSG00000047410 | 1 | 186270784 | 186354825 | 218 | 21 | 54649 | -0.026972 | 0.51076 | *TPR* |
| ENSG00000205707 | 12 | 25338150 | 25372579 | 122 | 14 | 46086 | -0.027059 | 0.51079 | *LYRM5* |
| ENSG00000168385 | 2 | 242244515 | 242303442 | 177 | 25 | 51055 | -0.027086 | 0.5108 | *Sep-02* |
| ENSG00000244045 | 17 | 26674604 | 26718716 | 88 | 10 | 33705 | -0.027098 | 0.51081 | *TMEM199* |
| ENSG00000184524 | 11 | 777104 | 800123 | 54 | 14 | 45528 | -0.027473 | 0.51096 | *CEND1* |
| ENSG00000141367 | 17 | 57687219 | 57783671 | 162 | 15 | 36630 | -0.027905 | 0.51113 | *CLTC* |
| ENSG00000154118 | 16 | 87625441 | 87741762 | 597 | 100 | 48272 | -0.027976 | 0.51116 | *JPH3* |
| ENSG00000117410 | 1 | 44430159 | 44453967 | 35 | 9 | 34560 | -0.028019 | 0.51118 | *ATP6V0B* |
| ENSG00000267041 | 19 | 37195285 | 37273727 | 209 | 22 | 50660 | -0.028103 | 0.51121 | *ZNF850* |
| ENSG00000120278 | 6 | 150910999 | 151174799 | 1121 | 121 | 47102 | -0.028371 | 0.51132 | *PLEKHG1* |
| ENSG00000175213 | 11 | 46712368 | 46737462 | 39 | 10 | 40051 | -0.028376 | 0.51132 | *ZNF408* |
| ENSG00000175866 | 17 | 78998948 | 79101232 | 490 | 85 | 48682 | -0.028591 | 0.5114 | *BAIAP2* |
| ENSG00000142102 | 11 | 279135 | 306107 | 66 | 24 | 36197 | -0.028611 | 0.51141 | *ATHL1* |
| ENSG00000111110 | 12 | 63027762 | 63338817 | 1076 | 144 | 42844 | -0.029031 | 0.51158 | *PPM1H* |
| ENSG00000240720 | 7 | 91731473 | 91818845 | 166 | 12 | 49245 | -0.029171 | 0.51164 | *LRRD1* |
| ENSG00000155755 | 2 | 202474907 | 202518293 | 94 | 19 | 46926 | -0.029243 | 0.51166 | *TMEM237* |
| ENSG00000086061 | 9 | 33015209 | 33049905 | 173 | 14 | 51448 | -0.029386 | 0.51172 | *DNAJA1* |
| ENSG00000130950 | 9 | 97067605 | 97100926 | 90 | 24 | 41849 | -0.029493 | 0.51176 | *NUTM2F* |
| ENSG00000126091 | 1 | 44161495 | 44406831 | 605 | 45 | 38238 | -0.029576 | 0.5118 | *ST3GAL3* |
| ENSG00000161326 | 17 | 35839937 | 35883603 | 124 | 20 | 48240 | -0.029673 | 0.51184 | *DUSP14* |
| ENSG00000154727 | 21 | 27096881 | 27154771 | 123 | 18 | 42785 | -0.029984 | 0.51196 | *GABPA* |
| ENSG00000118245 | 2 | 217714181 | 217734787 | 75 | 16 | 43989 | -0.030139 | 0.51202 | *TNP1* |
| ENSG00000164823 | 8 | 90904087 | 90950116 | 101 | 16 | 38700 | -0.030297 | 0.51209 | *OSGIN2* |
| ENSG00000188042 | 2 | 235391685 | 235415697 | 75 | 26 | 49690 | -0.03037 | 0.51211 | *ARL4C* |
| ENSG00000204624 | 1 | 11529223 | 11607641 | 222 | 24 | 36817 | -0.030457 | 0.51215 | *PTCHD2* |
| ENSG00000178209 | 8 | 144979321 | 145060902 | 219 | 20 | 38874 | -0.030521 | 0.51217 | *PLEC* |

| | | | | | | | | | |
|---|---|---|---|---|---|---|---|---|---|
| ENSG00000128607 | 7 | 129700350 | 129785560 | 243 | 31 | 38266 | -0.030747 | 0.51226 | *KLHDC10* |
| ENSG00000269746 | 16 | 70229303 | 70249683 | 31 | 14 | 43317 | -0.030937 | 0.51234 | *AC009060.1* |
| ENSG00000145675 | 5 | 67501548 | 67607649 | 312 | 55 | 43628 | -0.031197 | 0.51244 | *PIK3R1* |
| ENSG00000119139 | 9 | 71726209 | 71880124 | 662 | 59 | 51810 | -0.031304 | 0.51249 | *TJP2* |
| ENSG00000177684 | 6 | 49917962 | 49941818 | 68 | 11 | 50832 | -0.031333 | 0.5125 | *DEFB114* |
| ENSG00000145216 | 4 | 54233810 | 55171439 | 3028 | 225 | 43750 | -0.031641 | 0.51262 | *FIP1L1* |
| ENSG00000007520 | 16 | 1389241 | 1411912 | 74 | 22 | 45632 | -0.031628 | 0.51262 | *TSR3* |
| ENSG00000108528 | 17 | 4830425 | 4853546 | 60 | 17 | 39208 | -0.031902 | 0.51273 | *SLC25A11* |
| ENSG00000100362 | 22 | 37186728 | 37225523 | 148 | 24 | 40723 | -0.031945 | 0.51274 | *PVALB* |
| ENSG00000174996 | 11 | 66014765 | 66045331 | 59 | 14 | 55090 | -0.032149 | 0.51282 | *KLC2* |
| ENSG00000221859 | 21 | 46047273 | 46068370 | 98 | 13 | 48308 | -0.032252 | 0.51286 | *KRTAP10-10* |
| ENSG00000189241 | 6 | 116587741 | 116611066 | 77 | 10 | 31981 | -0.032588 | 0.513 | *TSPYL1* |
| ENSG00000008394 | 12 | 16490076 | 16772193 | 603 | 89 | 45055 | -0.032833 | 0.5131 | *MGST1* |
| ENSG00000049246 | 1 | 7834380 | 7915237 | 352 | 42 | 47933 | -0.03335 | 0.5133 | *PER3* |
| ENSG00000167657 | 19 | 3948451 | 3981121 | 84 | 25 | 51040 | -0.033474 | 0.51335 | *DAPK3* |
| ENSG00000196154 | 1 | 153506089 | 153532612 | 60 | 7 | 29775 | -0.033548 | 0.51338 | *S100A4* |
| ENSG00000137968 | 1 | 75657816 | 76086801 | 1258 | 77 | 45728 | -0.033562 | 0.51339 | *SLC44A5* |
| ENSG00000189127 | 5 | 79842574 | 79876307 | 110 | 25 | 42045 | -0.033562 | 0.51339 | *ANKRD34B* |
| ENSG00000136167 | 13 | 46690055 | 46796006 | 318 | 55 | 37940 | -0.033617 | 0.51341 | *LCP1* |
| ENSG00000103546 | 16 | 55679516 | 55750104 | 243 | 47 | 47027 | -0.033758 | 0.51346 | *SLC6A2* |
| ENSG00000172183 | 15 | 89169384 | 89209714 | 165 | 30 | 38309 | -0.033772 | 0.51347 | *ISG20* |
| ENSG00000064393 | 7 | 139236316 | 139487577 | 485 | 76 | 41783 | -0.034138 | 0.51362 | *HIPK2* |
| ENSG00000162817 | 1 | 220853187 | 220882499 | 143 | 16 | 47273 | -0.034414 | 0.51373 | *C1orf115* |
| ENSG00000138892 | 22 | 50443552 | 50505364 | 304 | 52 | 51674 | -0.034441 | 0.51374 | *TTLL8* |
| ENSG00000104129 | 15 | 41050067 | 41109675 | 134 | 20 | 40242 | -0.034763 | 0.51387 | *DNAJC17* |
| ENSG00000078070 | 3 | 182723006 | 182843863 | 277 | 27 | 33169 | -0.034886 | 0.51391 | *MCCC1* |
| ENSG00000152661 | 6 | 121746838 | 121780873 | 141 | 23 | 45158 | -0.035073 | 0.51399 | *GJA1* |
| ENSG00000095587 | 10 | 98114363 | 98283675 | 548 | 84 | 50539 | -0.035147 | 0.51402 | *TLL2* |
| ENSG00000149257 | 11 | 75263101 | 75293828 | 90 | 22 | 37332 | -0.03514 | 0.51402 | *SERPINH1* |
| ENSG00000259112 | 11 | 77718017 | 77800911 | 344 | 33 | 50692 | -0.035173 | 0.51403 | *NDUFC2-KCTD14* |
| ENSG00000119669 | 14 | 77480888 | 77505034 | 80 | 18 | 43084 | -0.035258 | 0.51406 | *IRF2BPL* |
| ENSG00000115526 | 2 | 100998327 | 101044118 | 101 | 17 | 35465 | -0.035323 | 0.51409 | *CHST10* |
| ENSG00000232774 | 14 | 62027258 | 62135414 | 283 | 49 | 40681 | -0.03532 | 0.51409 | *RP11-47I22.3* |
| ENSG00000005981 | 7 | 95097756 | 95179544 | 267 | 46 | 46853 | -0.035771 | 0.51427 | *ASB4* |
| ENSG00000228049 | 7 | 102267474 | 102322076 | 3 | 2 | 42576 | -0.03583 | 0.51429 | *POLR2J2* |
| ENSG00000267645 | 7 | 102267496 | 102322088 | 3 | 2 | 42576 | -0.03583 | 0.51429 | *POLR2J2* |

| | | | | | | | | | |
|---|---|---|---|---|---|---|---|---|---|
| ENSG00000173678 | 7 | 102280772 | 102311847 | 3 | 2 | 42576 | -0.03583 | 0.51429 | *SPDYE2B* |
| ENSG00000270299 | 20 | 617270 | 666444 | 147 | 38 | 44285 | -0.035959 | 0.51434 | *RP5-850E9.3* |
| ENSG00000069431 | 12 | 21940335 | 22104336 | 376 | 54 | 48254 | -0.036093 | 0.5144 | *ABCC9* |
| ENSG00000123505 | 6 | 111185973 | 111226916 | 139 | 12 | 46760 | -0.036449 | 0.51454 | *AMD1* |
| ENSG00000178538 | 8 | 61089906 | 61203971 | 460 | 49 | 43509 | -0.036548 | 0.51458 | *CA8* |
| ENSG00000164309 | 5 | 78975700 | 79106063 | 389 | 66 | 41390 | -0.036639 | 0.51461 | *CMYA5* |
| ENSG00000060762 | 6 | 166768407 | 166806486 | 51 | 9 | 38742 | -0.03685 | 0.5147 | *MPC1* |
| ENSG00000148082 | 9 | 91618060 | 91803682 | 593 | 73 | 50369 | -0.036896 | 0.51472 | *SHC3* |
| ENSG00000146233 | 6 | 46507541 | 46630523 | 346 | 36 | 39075 | -0.036953 | 0.51474 | *CYP39A1* |
| ENSG00000094796 | 17 | 39539976 | 39563844 | 61 | 14 | 41231 | -0.037238 | 0.51485 | *KRT31* |
| ENSG00000187185 | 16 | 57822098 | 57860850 | 127 | 24 | 45964 | -0.037278 | 0.51487 | *CTD-2600O9.1* |
| ENSG00000116690 | 1 | 186255405 | 186293694 | 79 | 14 | 46678 | -0.037512 | 0.51496 | *PRG4* |
| ENSG00000177570 | 8 | 119191698 | 119644234 | 1353 | 136 | 39015 | -0.037544 | 0.51497 | *SAMD12* |
| ENSG00000140743 | 16 | 22347257 | 22458486 | 94 | 31 | 45452 | -0.037534 | 0.51497 | *CDR2* |
| ENSG00000163512 | 3 | 28346494 | 28400618 | 92 | 14 | 39085 | -0.037574 | 0.51499 | *AZI2* |
| ENSG00000188803 | 17 | 11134580 | 11477380 | 1225 | 162 | 46013 | -0.037579 | 0.51499 | *SHISA6* |
| ENSG00000198771 | 1 | 167589330 | 167685486 | 384 | 78 | 49106 | -0.037654 | 0.51502 | *RCSD1* |
| ENSG00000164746 | 7 | 48065108 | 48110901 | 159 | 20 | 40990 | -0.0377 | 0.51504 | *C7orf57* |
| ENSG00000179455 | 15 | 23800454 | 23883064 | 220 | 35 | 45140 | -0.037764 | 0.51506 | *MKRN3* |
| ENSG00000176879 | 11 | 4934531 | 4955637 | 113 | 26 | 50683 | -0.037804 | 0.51508 | *OR51G1* |
| ENSG00000125703 | 1 | 63239806 | 63341184 | 247 | 18 | 52305 | -0.037828 | 0.51509 | *ATG4C* |
| ENSG00000134760 | 18 | 28888052 | 28946992 | 297 | 22 | 37033 | -0.038061 | 0.51518 | *DSG1* |
| ENSG00000080910 | 1 | 196778898 | 196938356 | 319 | 37 | 29854 | -0.038087 | 0.51519 | *CFHR2* |
| ENSG00000151033 | 10 | 30890718 | 30928691 | 202 | 25 | 43323 | -0.038309 | 0.51528 | *LYZL2* |
| ENSG00000154997 | 7 | 55851237 | 55940482 | 267 | 39 | 36111 | -0.038355 | 0.5153 | *Sep-14* |
| ENSG00000119711 | 14 | 74513553 | 74561196 | 178 | 14 | 27726 | -0.038489 | 0.51535 | *ALDH6A1* |
| ENSG00000139946 | 14 | 56574532 | 56778244 | 764 | 37 | 46383 | -0.038716 | 0.51544 | *PELI2* |
| ENSG00000260092 | 16 | 75552430 | 75589326 | 114 | 13 | 48983 | -0.038992 | 0.51555 | *RP11-77K12.7* |
| ENSG00000140319 | 15 | 40317940 | 40341389 | 54 | 13 | 43650 | -0.039012 | 0.51556 | *SRP14* |
| ENSG00000144744 | 3 | 69093881 | 69139559 | 97 | 15 | 51964 | -0.039073 | 0.51558 | *UBA3* |
| ENSG00000167110 | 9 | 131008108 | 131048274 | 77 | 13 | 41794 | -0.039148 | 0.51561 | *GOLGA2* |
| ENSG00000196950 | 2 | 196430701 | 196612426 | 436 | 57 | 41882 | -0.039174 | 0.51562 | *SLC39A10* |
| ENSG00000173535 | 8 | 22931868 | 22984950 | 173 | 35 | 41911 | -0.039184 | 0.51563 | *TNFRSF10C* |
| ENSG00000213523 | 5 | 139906925 | 139947895 | 55 | 14 | 49736 | -0.039226 | 0.51565 | *SRA1* |
| ENSG00000099942 | 22 | 21261714 | 21318037 | 155 | 17 | 46166 | -0.039249 | 0.51565 | *CRKL* |
| ENSG00000167359 | 11 | 5451800 | 5472744 | 118 | 18 | 39951 | -0.039704 | 0.51584 | *OR51I1* |

| | | | | | | | | | |
|---|---|---|---|---|---|---|---|---|---|
| ENSG00000159348 | 1 | 202920997 | 202946408 | 58 | 14 | 37248 | -0.039792 | 0.51587 | *CYB5R1* |
| ENSG00000162706 | 1 | 159131399 | 159183103 | 121 | 30 | 41598 | -0.040152 | 0.51601 | *CADM3* |
| ENSG00000224870 | 1 | 1324902 | 1347426 | 41 | 8 | 32182 | -0.040169 | 0.51602 | *RP4-758J18.2* |
| ENSG00000140598 | 15 | 82412571 | 82565104 | 502 | 52 | 36637 | -0.040445 | 0.51613 | *EFTUD1* |
| ENSG00000267114 | 19 | 45443301 | 45467264 | 110 | 9 | 50331 | -0.040533 | 0.51617 | *CTB-129P6.11* |
| ENSG00000166938 | 15 | 66575555 | 66636236 | 190 | 35 | 53053 | -0.040685 | 0.51623 | *DIS3L* |
| ENSG00000134243 | 1 | 109842192 | 109950573 | 182 | 17 | 44216 | -0.040934 | 0.51633 | *SORT1* |
| ENSG00000155511 | 5 | 152859175 | 153203429 | 1340 | 114 | 44849 | -0.041031 | 0.51636 | *GRIA1* |
| ENSG00000123144 | 19 | 12831454 | 12855589 | 20 | 12 | 52682 | -0.041202 | 0.51643 | *C19orf43* |
| ENSG00000113083 | 5 | 121388890 | 121423980 | 83 | 17 | 50375 | -0.041606 | 0.51659 | *LOX* |
| ENSG00000267261 | 17 | 40261692 | 40316934 | 119 | 13 | 32965 | -0.041655 | 0.51661 | *CTD-2132N18.3* |
| ENSG00000107404 | 1 | 1260656 | 1294730 | 110 | 17 | 35459 | -0.041763 | 0.51666 | *DVL1* |
| ENSG00000269630 | 18 | 60495640 | 60515948 | 37 | 8 | 43476 | -0.041799 | 0.51667 | *AC015989.2* |
| ENSG00000196511 | 7 | 144139034 | 144543488 | 1159 | 86 | 33898 | -0.041852 | 0.51669 | *TPK1* |
| ENSG00000159733 | 4 | 2261309 | 2430390 | 743 | 83 | 47914 | -0.042032 | 0.51676 | *ZFYVE28* |
| ENSG00000164440 | 6 | 139551198 | 139623276 | 180 | 33 | 42776 | -0.042074 | 0.51678 | *TXLNB* |
| ENSG00000232382 | 3 | 98178324 | 98199420 | 55 | 13 | 42812 | -0.042178 | 0.51682 | *OR5K1* |
| ENSG00000025770 | 22 | 50936645 | 50971901 | 144 | 33 | 50383 | -0.042377 | 0.5169 | *NCAPH2* |
| ENSG00000177990 | 12 | 63942693 | 64072719 | 674 | 57 | 41499 | -0.042432 | 0.51692 | *DPY19L2* |
| ENSG00000104341 | 8 | 98777285 | 98875241 | 469 | 29 | 48228 | -0.04272 | 0.51704 | *LAPTM4B* |
| ENSG00000170516 | 4 | 46726844 | 46921252 | 523 | 32 | 53711 | -0.042739 | 0.51705 | *COX7B2* |
| ENSG00000121335 | 12 | 11534476 | 11663975 | 467 | 44 | 38119 | -0.042935 | 0.51712 | *PRB2* |
| ENSG00000102871 | 16 | 67178083 | 67204201 | 17 | 10 | 43523 | -0.042935 | 0.51712 | *TRADD* |
| ENSG00000124370 | 2 | 71326814 | 71367369 | 145 | 16 | 41064 | -0.043193 | 0.51723 | *MCEE* |
| ENSG00000108759 | 17 | 39606063 | 39633681 | 199 | 14 | 56287 | -0.043409 | 0.51731 | *KRT32* |
| ENSG00000105219 | 19 | 40718115 | 40742597 | 56 | 15 | 41640 | -0.043554 | 0.51737 | *CNTD2* |
| ENSG00000158023 | 12 | 122345768 | 122451833 | 314 | 33 | 33829 | -0.04358 | 0.51738 | *WDR66* |
| ENSG00000123815 | 19 | 41187434 | 41234112 | 148 | 25 | 52145 | -0.043667 | 0.51742 | *ADCK4* |
| ENSG00000169860 | 3 | 152542736 | 152565841 | 41 | 7 | 31054 | -0.043828 | 0.51748 | *P2RY1* |
| ENSG00000160208 | 21 | 45069429 | 45125958 | 129 | 11 | 44933 | -0.043986 | 0.51754 | *RRP1B* |
| ENSG00000173809 | 19 | 33200659 | 33330483 | 406 | 34 | 46664 | -0.044132 | 0.5176 | *TDRD12* |
| ENSG00000115339 | 2 | 166594101 | 166661192 | 119 | 23 | 42017 | -0.044159 | 0.51761 | *GALNT3* |
| ENSG00000172421 | 17 | 60437579 | 60503837 | 108 | 24 | 41190 | -0.044215 | 0.51763 | *EFCAB3* |
| ENSG00000183154 | 8 | 37582279 | 37604944 | 16 | 9 | 56555 | -0.044535 | 0.51776 | *RP11-863K10.7* |
| ENSG00000181744 | 3 | 143680640 | 143777561 | 368 | 24 | 34672 | -0.04461 | 0.51779 | *C3orf58* |
| ENSG00000269506 | 4 | 54917213 | 54937377 | 22 | 6 | 52853 | -0.044603 | 0.51779 | *AC110792.1* |

| | | | | | | | | | |
|---|---|---|---|---|---|---|---|---|---|
| ENSG00000171501 | 9 | 125305391 | 125326493 | 175 | 18 | 38904 | -0.04467 | 0.51781 | *OR1N2* |
| ENSG00000023734 | 12 | 16025325 | 16066412 | 38 | 17 | 49491 | -0.045404 | 0.51811 | *STRAP* |
| ENSG00000225485 | 17 | 36574662 | 36678628 | 334 | 58 | 44230 | -0.045405 | 0.51811 | *ARHGAP23* |
| ENSG00000143390 | 1 | 151303116 | 151329833 | 47 | 11 | 51911 | -0.045636 | 0.5182 | *RFX5* |
| ENSG00000185130 | 6 | 27765257 | 27785709 | 55 | 12 | 37200 | -0.045804 | 0.51827 | *HIST1H2BL* |
| ENSG00000139278 | 12 | 75864460 | 75907633 | 136 | 12 | 54835 | -0.045832 | 0.51828 | *GLIPR1* |
| ENSG00000163295 | 2 | 233310833 | 233334742 | 87 | 17 | 48531 | -0.045912 | 0.51831 | *ALPI* |
| ENSG00000183066 | 22 | 42384729 | 42464460 | 177 | 19 | 49065 | -0.045927 | 0.51832 | *WBP2NL* |
| ENSG00000196578 | 3 | 97796017 | 97816946 | 134 | 13 | 42997 | -0.045965 | 0.51833 | *OR5AC2* |
| ENSG00000114346 | 3 | 172458472 | 172549264 | 226 | 25 | 40868 | -0.045956 | 0.51833 | *ECT2* |
| ENSG00000066557 | 1 | 70600488 | 70681303 | 129 | 16 | 46134 | -0.045981 | 0.51834 | *LRRC40* |
| ENSG00000090863 | 16 | 74475856 | 74651012 | 612 | 45 | 49322 | -0.046203 | 0.51843 | *GLG1* |
| ENSG00000165699 | 9 | 135756735 | 135830020 | 179 | 23 | 51225 | -0.046241 | 0.51844 | *TSC1* |
| ENSG00000105894 | 7 | 136902088 | 137038611 | 387 | 69 | 44668 | -0.046413 | 0.51851 | *PTN* |
| ENSG00000268130 | 13 | 113745563 | 113766608 | 77 | 15 | 45578 | -0.046451 | 0.51852 | *AL137002.1* |
| ENSG00000214642 | 6 | 49926390 | 49947338 | 61 | 9 | 48318 | -0.0466 | 0.51858 | *DEFB113* |
| ENSG00000187288 | 3 | 9898398 | 9931938 | 93 | 19 | 50906 | -0.047019 | 0.51875 | *CIDEC* |
| ENSG00000105438 | 19 | 48875827 | 48904810 | 72 | 17 | 45138 | -0.047273 | 0.51885 | *KDELR1* |
| ENSG00000064703 | 1 | 112287867 | 112320638 | 126 | 16 | 40584 | -0.047509 | 0.51895 | *DDX20* |
| ENSG00000109684 | 4 | 10478019 | 10696489 | 879 | 84 | 44804 | -0.047834 | 0.51908 | *CLNK* |
| ENSG00000078808 | 1 | 1142288 | 1177411 | 188 | 15 | 49125 | -0.048399 | 0.5193 | *SDF4* |
| ENSG00000060237 | 12 | 851759 | 1030618 | 467 | 47 | 51051 | -0.048537 | 0.51936 | *WNK1* |
| ENSG00000136931 | 9 | 127233516 | 127279709 | 64 | 12 | 42154 | -0.048658 | 0.5194 | *NR5A1* |
| ENSG00000175336 | 12 | 56744353 | 56766607 | 22 | 5 | 52908 | -0.048675 | 0.51941 | *APOF* |
| ENSG00000186007 | 1 | 205340506 | 205435082 | 296 | 38 | 48030 | -0.048699 | 0.51942 | *LEMD1* |
| ENSG00000115840 | 2 | 172630880 | 172874766 | 632 | 52 | 42325 | -0.04897 | 0.51953 | *SLC25A12* |
| ENSG00000115902 | 2 | 65205611 | 65260999 | 200 | 36 | 50092 | -0.049384 | 0.51969 | *SLC1A4* |
| ENSG00000111676 | 12 | 7023626 | 7061484 | 62 | 21 | 43457 | -0.049483 | 0.51973 | *ATN1* |
| ENSG00000154310 | 3 | 170769128 | 171188197 | 1205 | 146 | 48133 | -0.049745 | 0.51984 | *TNIK* |
| ENSG00000171566 | 4 | 155446158 | 155481587 | 143 | 15 | 55392 | -0.049861 | 0.51988 | *PLRG1* |
| ENSG00000243135 | 2 | 234627754 | 234691945 | 259 | 32 | 40208 | -0.050298 | 0.52006 | *UGT1A3* |
| ENSG00000158055 | 1 | 24635812 | 24700972 | 168 | 38 | 43025 | -0.050949 | 0.52032 | *GRHL3* |
| ENSG00000120662 | 13 | 41780505 | 41847742 | 218 | 19 | 52412 | -0.051015 | 0.52034 | *MTRF1* |
| ENSG00000152193 | 13 | 79178426 | 79243314 | 172 | 16 | 46185 | -0.051041 | 0.52035 | *RNF219* |
| ENSG00000159961 | 17 | 3313862 | 3334827 | 54 | 11 | 32316 | -0.051143 | 0.52039 | *OR3A3* |
| ENSG00000159079 | 21 | 33954389 | 33995176 | 147 | 20 | 48140 | -0.05124 | 0.52043 | *C21orf59* |

| | | | | | | | | | |
|---|---|---|---|---|---|---|---|---|---|
| ENSG00000130193 | 8 | 143798621 | 143828345 | 87 | 13 | 50843 | -0.051352 | 0.52048 | *THEM6* |
| ENSG00000268182 | 19 | 57144513 | 57177134 | 118 | 21 | 47067 | -0.051358 | 0.52048 | *SMIM17* |
| ENSG00000169627 | 16 | 30194255 | 30215627 | 8 | 5 | 32944 | -0.05169 | 0.52061 | *BOLA2B* |
| ENSG00000144369 | 2 | 187548698 | 187640685 | 185 | 23 | 50135 | -0.051885 | 0.52069 | *FAM171B* |
| ENSG00000196859 | 17 | 39104669 | 39133144 | 133 | 15 | 34069 | -0.051991 | 0.52073 | *KRT39* |
| ENSG00000080608 | 9 | 2710469 | 2854241 | 457 | 81 | 45018 | -0.05206 | 0.52076 | *KIAA0020* |
| ENSG00000135898 | 2 | 231762033 | 231835781 | 226 | 44 | 39183 | -0.052298 | 0.52085 | *GPR55* |
| ENSG00000196747 | 6 | 27765899 | 27786429 | 57 | 12 | 37136 | -0.052402 | 0.5209 | *HIST1H2AI* |
| ENSG00000100342 | 22 | 36639056 | 36673576 | 129 | 23 | 47321 | -0.052506 | 0.52094 | *APOL1* |
| ENSG00000048828 | 9 | 96204004 | 96338397 | 431 | 40 | 49050 | -0.052661 | 0.521 | *FAM120A* |
| ENSG00000132471 | 17 | 73831780 | 73862588 | 119 | 17 | 36034 | -0.052807 | 0.52106 | *WBP2* |
| ENSG00000197429 | 1 | 46149996 | 46226322 | 186 | 9 | 54307 | -0.052971 | 0.52112 | *IPP* |
| ENSG00000104313 | 8 | 72099668 | 72284467 | 467 | 83 | 44822 | -0.052971 | 0.52112 | *EYA1* |
| ENSG00000163399 | 1 | 116905290 | 116962883 | 93 | 11 | 27639 | -0.053177 | 0.5212 | *ATP1A1* |
| ENSG00000234906 | 19 | 45439243 | 45462822 | 96 | 8 | 49418 | -0.053211 | 0.52122 | *APOC2* |
| ENSG00000155970 | 8 | 16874747 | 16990153 | 391 | 56 | 45148 | -0.053484 | 0.52133 | *MICU3* |
| ENSG00000197136 | 11 | 65373244 | 65414910 | 63 | 19 | 44601 | -0.053504 | 0.52133 | *PCNXL3* |
| ENSG00000123358 | 12 | 52406616 | 52463291 | 161 | 41 | 46891 | -0.053714 | 0.52142 | *NR4A1* |
| ENSG00000213463 | 14 | 70828148 | 70893778 | 286 | 31 | 44099 | -0.053828 | 0.52146 | *SYNJ2BP* |
| ENSG00000244274 | 20 | 44024697 | 44049250 | 58 | 9 | 43619 | -0.053807 | 0.52146 | *DBNDD2* |
| ENSG00000105376 | 19 | 10390657 | 10417454 | 86 | 18 | 39918 | -0.054266 | 0.52164 | *ICAM5* |
| ENSG00000237515 | 16 | 12985477 | 13344272 | 1551 | 181 | 46980 | -0.054519 | 0.52174 | *SHISA9* |
| ENSG00000182240 | 21 | 42529728 | 42664445 | 434 | 88 | 45901 | -0.054733 | 0.52182 | *BACE2* |
| ENSG00000155876 | 9 | 19039372 | 19061019 | 100 | 13 | 42424 | -0.054799 | 0.52185 | *RRAGA* |
| ENSG00000099246 | 10 | 27783197 | 27841143 | 140 | 17 | 51082 | -0.054942 | 0.52191 | *RAB18* |
| ENSG00000119227 | 3 | 196663214 | 196705931 | 153 | 31 | 35056 | -0.055014 | 0.52194 | *PIGZ* |
| ENSG00000205659 | 14 | 74541499 | 74677936 | 487 | 28 | 28850 | -0.055306 | 0.52205 | *LIN52* |
| ENSG00000102837 | 13 | 53592894 | 53636192 | 126 | 33 | 47625 | -0.055449 | 0.52211 | *OLFM4* |
| ENSG00000198454 | 9 | 139853695 | 139876442 | 95 | 13 | 46148 | -0.055605 | 0.52217 | *C9orf141* |
| ENSG00000131043 | 20 | 34814381 | 34868840 | 67 | 22 | 44901 | -0.055717 | 0.52222 | *AAR2* |
| ENSG00000101752 | 18 | 19274918 | 19460918 | 316 | 36 | 40212 | -0.055951 | 0.52231 | *MIB1* |
| ENSG00000242441 | 2 | 48834937 | 48970287 | 559 | 70 | 48309 | -0.05603 | 0.52234 | *GTF2A1L* |
| ENSG00000143612 | 1 | 154169182 | 154203104 | 40 | 11 | 42367 | -0.056257 | 0.52243 | *C1orf43* |
| ENSG00000140691 | 16 | 31459401 | 31488487 | 48 | 18 | 45748 | -0.056429 | 0.5225 | *ARMC5* |
| ENSG00000110063 | 11 | 126163647 | 126225644 | 190 | 30 | 43165 | -0.056487 | 0.52252 | *DCPS* |
| ENSG00000095951 | 6 | 11998995 | 12175232 | 569 | 72 | 47274 | -0.056562 | 0.52255 | *HIVEP1* |

| | | | | | | | | | |
|---|---|---|---|---|---|---|---|---|---|
| ENSG00000211456 | 3 | 45720548 | 45796916 | 277 | 19 | 51541 | -0.056569 | 0.52256 | *SACM1L* |
| ENSG00000267168 | 17 | 74719060 | 74744604 | 16 | 8 | 46017 | -0.056676 | 0.5226 | *RP11-318A15.7* |
| ENSG00000215712 | 6 | 157700418 | 157754633 | 139 | 18 | 47517 | -0.056759 | 0.52263 | *TMEM242* |
| ENSG00000197872 | 2 | 16720727 | 16857599 | 347 | 73 | 38569 | -0.056847 | 0.52267 | *FAM49A* |
| ENSG00000177257 | 8 | 7262382 | 7284385 | 6 | 3 | 42871 | -0.05696 | 0.52271 | *DEFB4B* |
| ENSG00000186572 | 8 | 7659242 | 7683238 | 2 | 1 | 42875 | -0.057067 | 0.52275 | *DEFB107A* |
| ENSG00000189013 | 19 | 55226002 | 55335972 | 169 | 52 | 29365 | -0.057564 | 0.52295 | *KIR2DL4* |
| ENSG00000173338 | 11 | 65350326 | 65373467 | 55 | 10 | 50704 | -0.057767 | 0.52303 | *KCNK7* |
| ENSG00000173918 | 17 | 77008896 | 77055870 | 130 | 22 | 39856 | -0.057833 | 0.52306 | *C1QTNF1* |
| ENSG00000004478 | 12 | 2894119 | 2924576 | 56 | 10 | 32806 | -0.058168 | 0.52319 | *FKBP4* |
| ENSG00000110090 | 11 | 68512088 | 68621878 | 261 | 41 | 39710 | -0.058263 | 0.52323 | *CPT1A* |
| ENSG00000205148 | 15 | 96821280 | 96841660 | 45 | 11 | 38373 | -0.058338 | 0.52326 | *AC016251.1* |
| ENSG00000170417 | 2 | 103343367 | 103470352 | 283 | 29 | 53236 | -0.05835 | 0.52327 | *TMEM182* |
| ENSG00000144451 | 2 | 214139113 | 215285225 | 3776 | 171 | 43043 | -0.058444 | 0.5233 | *SPAG16* |
| ENSG00000159224 | 17 | 47025916 | 47055958 | 115 | 15 | 28940 | -0.05846 | 0.52331 | *GIP* |
| ENSG00000164900 | 7 | 150835676 | 150881832 | 106 | 20 | 43992 | -0.058656 | 0.52339 | *GBX1* |
| ENSG00000007129 | 19 | 42045886 | 42103197 | 115 | 19 | 40903 | -0.058756 | 0.52343 | *CEACAM21* |
| ENSG00000258315 | 17 | 6907814 | 6930844 | 46 | 6 | 51627 | -0.058881 | 0.52348 | *C17orf49* |
| ENSG00000125772 | 20 | 5515085 | 5601672 | 243 | 29 | 46277 | -0.058892 | 0.52348 | *GPCPD1* |
| ENSG00000197694 | 9 | 131304866 | 131405941 | 147 | 17 | 29001 | -0.059134 | 0.52358 | *SPTAN1* |
| ENSG00000152760 | 1 | 67208142 | 67254470 | 155 | 21 | 45047 | -0.059173 | 0.52359 | *TCTEX1D1* |
| ENSG00000107831 | 10 | 103519899 | 103545854 | 29 | 12 | 51258 | -0.059186 | 0.5236 | *FGF8* |
| ENSG00000164933 | 8 | 104400863 | 104437417 | 135 | 13 | 55453 | -0.059238 | 0.52362 | *SLC25A32* |
| ENSG00000108591 | 17 | 17981200 | 18021285 | 90 | 11 | 29382 | -0.059235 | 0.52362 | *DRG2* |
| ENSG00000157881 | 1 | 2429972 | 2468039 | 103 | 14 | 32115 | -0.059447 | 0.5237 | *PANK4* |
| ENSG00000152464 | 10 | 15129179 | 15191817 | 284 | 39 | 40013 | -0.059512 | 0.52373 | *RPP38* |
| ENSG00000145794 | 5 | 126616523 | 126811429 | 722 | 56 | 45095 | -0.05955 | 0.52374 | *MEGF10* |
| ENSG00000165476 | 10 | 65271123 | 65394883 | 348 | 35 | 46508 | -0.059656 | 0.52379 | *REEP3* |
| ENSG00000188015 | 1 | 153509805 | 153531848 | 53 | 6 | 27834 | -0.059684 | 0.5238 | *S100A3* |
| ENSG00000135828 | 1 | 182532769 | 182568391 | 92 | 23 | 45941 | -0.059798 | 0.52384 | *RNASEL* |
| ENSG00000125877 | 20 | 3179514 | 3214516 | 146 | 25 | 47605 | -0.059817 | 0.52385 | *ITPA* |
| ENSG00000140829 | 16 | 72117461 | 72156811 | 139 | 14 | 38038 | -0.060006 | 0.52392 | *DHX38* |
| ENSG00000178372 | 10 | 5530658 | 5551533 | 73 | 18 | 44004 | -0.060039 | 0.52394 | *CALML5* |
| ENSG00000108474 | 17 | 16110505 | 16262115 | 358 | 29 | 44583 | -0.060373 | 0.52407 | *PIGL* |
| ENSG00000100012 | 22 | 30833946 | 30878036 | 184 | 21 | 37360 | -0.060881 | 0.52427 | *SEC14L3* |
| ENSG00000154370 | 1 | 228571374 | 228604541 | 61 | 12 | 45816 | -0.061333 | 0.52445 | *TRIM11* |

| | | | | | | | | | |
|---|---|---|---|---|---|---|---|---|---|
| ENSG00000197619 | 19 | 52484585 | 52521483 | 152 | 16 | 30950 | -0.061388 | 0.52447 | *ZNF615* |
| ENSG00000165934 | 14 | 92578281 | 92640755 | 241 | 17 | 38739 | -0.061605 | 0.52456 | *CPSF2* |
| ENSG00000066044 | 19 | 8013463 | 8080543 | 174 | 21 | 52614 | -0.06181 | 0.52464 | *ELAVL1* |
| ENSG00000104897 | 19 | 2226520 | 2258678 | 103 | 31 | 42412 | -0.062067 | 0.52475 | *SF3A2* |
| ENSG00000110104 | 11 | 60599544 | 60628554 | 126 | 26 | 45387 | -0.06223 | 0.52481 | *CCDC86* |
| ENSG00000119508 | 9 | 102574137 | 102639173 | 78 | 17 | 44657 | -0.062322 | 0.52485 | *NR4A3* |
| ENSG00000205595 | 4 | 75470629 | 75500486 | 28 | 9 | 42539 | -0.062345 | 0.52486 | *AREGB* |
| ENSG00000109956 | 11 | 134238398 | 134291812 | 226 | 32 | 51671 | -0.062364 | 0.52486 | *B3GAT1* |
| ENSG00000136842 | 9 | 100253462 | 100374030 | 317 | 63 | 51220 | -0.062469 | 0.52491 | *TMOD1* |
| ENSG00000132972 | 13 | 25328290 | 25464059 | 482 | 35 | 54844 | -0.062586 | 0.52495 | *RNF17* |
| ENSG00000130818 | 19 | 9628667 | 9659303 | 145 | 5 | 23775 | -0.062615 | 0.52496 | *ZNF426* |
| ENSG00000053328 | 6 | 110557131 | 110689475 | 400 | 47 | 37788 | -0.062727 | 0.52501 | *METTL24* |
| ENSG00000155980 | 12 | 57933781 | 57990415 | 103 | 29 | 40442 | -0.062996 | 0.52512 | *KIF5A* |
| ENSG00000180929 | 3 | 51979330 | 52001509 | 31 | 11 | 32693 | -0.063025 | 0.52513 | *GPR62* |
| ENSG00000166263 | 17 | 53036088 | 53251646 | 629 | 37 | 51493 | -0.063085 | 0.52515 | *STXBP4* |
| ENSG00000164134 | 4 | 140212609 | 140351187 | 358 | 30 | 50253 | -0.063229 | 0.52521 | *NAA15* |
| ENSG00000129009 | 15 | 74456012 | 74479213 | 60 | 11 | 26701 | -0.063409 | 0.52528 | *ISLR* |
| ENSG00000221955 | 3 | 124791480 | 125008021 | 803 | 130 | 46480 | -0.063503 | 0.52532 | *SLC12A8* |
| ENSG00000165629 | 10 | 7820092 | 7859778 | 155 | 20 | 48816 | -0.063757 | 0.52542 | *ATP5C1* |
| ENSG00000109390 | 4 | 140178034 | 140233705 | 152 | 19 | 51484 | -0.06378 | 0.52543 | *NDUFC1* |
| ENSG00000086730 | 7 | 73603982 | 73654161 | 58 | 20 | 47110 | -0.063821 | 0.52544 | *LAT2* |
| ENSG00000197697 | 6 | 26173958 | 26194454 | 76 | 9 | 48837 | -0.063926 | 0.52549 | *HIST1H2BE* |
| ENSG00000177599 | 19 | 11898482 | 11930437 | 76 | 11 | 34046 | -0.06397 | 0.5255 | *ZNF491* |
| ENSG00000110693 | 11 | 15977995 | 16771138 | 1592 | 104 | 46904 | -0.06409 | 0.52555 | *SOX6* |
| ENSG00000107745 | 10 | 74117098 | 74395899 | 471 | 22 | 25183 | -0.064101 | 0.52556 | *MICU1* |
| ENSG00000203546 | 14 | 31793518 | 31936647 | 272 | 29 | 41715 | -0.06411 | 0.52556 | *RP11-176H8.1* |
| ENSG00000261667 | 8 | 144151874 | 144174418 | 70 | 10 | 27035 | -0.064189 | 0.52559 | *RP11-520P18.5* |
| ENSG00000106819 | 9 | 95208487 | 95254788 | 116 | 12 | 36861 | -0.064184 | 0.52559 | *ASPN* |
| ENSG00000108296 | 17 | 36946687 | 36991734 | 119 | 21 | 40494 | -0.064206 | 0.5256 | *CWC25* |
| ENSG00000167580 | 12 | 50334524 | 50362664 | 72 | 20 | 32856 | -0.064392 | 0.52567 | *AQP2* |
| ENSG00000115446 | 2 | 99215042 | 99244978 | 57 | 6 | 55869 | -0.064772 | 0.52582 | *UNC50* |
| ENSG00000182154 | 9 | 140435651 | 140457007 | 49 | 19 | 45802 | -0.064907 | 0.52588 | *MRPL41* |
| ENSG00000145016 | 3 | 197388264 | 197486598 | 212 | 19 | 44745 | -0.065097 | 0.52595 | *KIAA0226* |
| ENSG00000184860 | 16 | 82021221 | 82055093 | 52 | 21 | 39358 | -0.065442 | 0.52609 | *SDR42E1* |
| ENSG00000235942 | 1 | 152805344 | 152826459 | 22 | 13 | 47243 | -0.065663 | 0.52618 | *LCE6A* |
| ENSG00000114573 | 3 | 113455866 | 113540903 | 129 | 8 | 52332 | -0.0658 | 0.52623 | *ATP6V1A* |

| | | | | | | | | | |
|---|---|---|---|---|---|---|---|---|---|
| ENSG00000182747 | 6 | 137233402 | 137256777 | 74 | 10 | 49223 | -0.065888 | 0.52627 | *SLC35D3* |
| ENSG00000239590 | 9 | 125271420 | 125292361 | 78 | 15 | 34634 | -0.065943 | 0.52629 | *OR1J4* |
| ENSG00000111341 | 12 | 15024115 | 15048860 | 54 | 5 | 54399 | -0.066246 | 0.52641 | *MGP* |
| ENSG00000234545 | 7 | 92180107 | 92229708 | 109 | 20 | 48955 | -0.066422 | 0.52648 | *FAM133B* |
| ENSG00000203827 | 1 | 148729443 | 148768311 | 3 | 1 | 43194 | -0.066448 | 0.52649 | *NBPF16* |
| ENSG00000134851 | 4 | 56252124 | 56329564 | 260 | 23 | 48335 | -0.066712 | 0.52659 | *TMEM165* |
| ENSG00000215067 | 17 | 6769954 | 6925668 | 419 | 80 | 47532 | -0.066947 | 0.52669 | *AC027763.2* |
| ENSG00000041802 | 3 | 194351517 | 194403206 | 280 | 24 | 46039 | -0.067122 | 0.52676 | *LSG1* |
| ENSG00000182782 | 12 | 123175840 | 123197890 | 81 | 8 | 34821 | -0.067246 | 0.52681 | *HCAR2* |
| ENSG00000186188 | 10 | 95316422 | 95374237 | 209 | 40 | 41452 | -0.067332 | 0.52684 | *FFAR4* |
| ENSG00000170264 | 2 | 62041989 | 62091278 | 158 | 19 | 54881 | -0.067406 | 0.52687 | *FAM161A* |
| ENSG00000213204 | 6 | 88107730 | 88231352 | 470 | 39 | 53358 | -0.067537 | 0.52692 | *C6orf165* |
| ENSG00000164983 | 8 | 125314231 | 125394933 | 247 | 26 | 51575 | -0.067672 | 0.52698 | *TMEM65* |
| ENSG00000004866 | 7 | 116583292 | 116880157 | 440 | 62 | 41792 | -0.067711 | 0.52699 | *ST7* |
| ENSG00000175600 | 7 | 40164575 | 40910362 | 1504 | 135 | 45468 | -0.067957 | 0.52709 | *SUGCT* |
| ENSG00000168894 | 2 | 85812848 | 85834736 | 46 | 12 | 38048 | -0.068061 | 0.52713 | *RNF181* |
| ENSG00000187871 | 6 | 55182267 | 55277291 | 751 | 51 | 46209 | -0.068249 | 0.52721 | *GFRAL* |
| ENSG00000012223 | 3 | 46467136 | 46536724 | 271 | 37 | 40312 | -0.068311 | 0.52723 | *LTF* |
| ENSG00000196236 | 22 | 41243081 | 41373838 | 134 | 29 | 43992 | -0.06849 | 0.5273 | *XPNPEP3* |
| ENSG00000184277 | 15 | 102151847 | 102202594 | 158 | 31 | 45723 | -0.068512 | 0.52731 | *TM2D3* |
| ENSG00000107551 | 10 | 45444855 | 45501339 | 177 | 40 | 44958 | -0.068611 | 0.52735 | *RASSF4* |
| ENSG00000138134 | 10 | 90629491 | 90744910 | 291 | 44 | 40639 | -0.068896 | 0.52746 | *STAMBPL1* |
| ENSG00000073849 | 3 | 186638274 | 186806341 | 704 | 134 | 48432 | -0.068986 | 0.5275 | *ST6GAL1* |
| ENSG00000180433 | 1 | 158714510 | 158735681 | 104 | 16 | 36202 | -0.069241 | 0.5276 | *OR6K6* |
| ENSG00000124575 | 6 | 26224440 | 26245216 | 72 | 9 | 51490 | -0.069644 | 0.52776 | *HIST1H1D* |
| ENSG00000100401 | 22 | 41631615 | 41692255 | 132 | 17 | 51862 | -0.069773 | 0.52781 | *RANGAP1* |
| ENSG00000117620 | 1 | 100425345 | 100502535 | 279 | 22 | 45662 | -0.070331 | 0.52803 | *SLC35A3* |
| ENSG00000136273 | 7 | 47725328 | 48029178 | 1441 | 117 | 47687 | -0.070355 | 0.52804 | *HUS1* |
| ENSG00000178222 | 4 | 1040038 | 1117350 | 388 | 23 | 40232 | -0.070427 | 0.52807 | *RNF212* |
| ENSG00000243749 | 1 | 35437136 | 35460954 | 26 | 9 | 31023 | -0.070483 | 0.5281 | *ZMYM6NB* |
| ENSG00000171827 | 19 | 37948487 | 37986260 | 66 | 13 | 35528 | -0.070539 | 0.52812 | *ZNF570* |
| ENSG00000137266 | 6 | 3259196 | 3467256 | 935 | 113 | 45000 | -0.070814 | 0.52823 | *SLC22A23* |
| ENSG00000164853 | 7 | 1262543 | 1286954 | 54 | 14 | 37342 | -0.070869 | 0.52825 | *UNCX* |
| ENSG00000063322 | 19 | 39871943 | 39901277 | 53 | 11 | 39266 | -0.070878 | 0.52825 | *MED29* |
| ENSG00000112242 | 6 | 20392398 | 20503941 | 254 | 57 | 45020 | -0.070899 | 0.52826 | *E2F3* |
| ENSG00000186086 | 1 | 108908421 | 109023624 | 80 | 13 | 40395 | -0.070955 | 0.52828 | *NBPF6* |

| | | | | | | | | | |
|---|---|---|---|---|---|---|---|---|---|
| ENSG00000168393 | 2 | 242605157 | 242636406 | 184 | 16 | 46605 | -0.071075 | 0.52833 | *DTYMK* |
| ENSG00000160404 | 9 | 130483803 | 130507604 | 64 | 11 | 48546 | -0.071934 | 0.52867 | *TOR2A* |
| ENSG00000197279 | 6 | 28038753 | 28067341 | 104 | 13 | 51565 | -0.072392 | 0.52885 | *ZNF165* |
| ENSG00000133740 | 8 | 86079460 | 86139387 | 106 | 13 | 50564 | -0.072449 | 0.52888 | *E2F5* |
| ENSG00000167286 | 11 | 118199669 | 118223459 | 59 | 13 | 42724 | -0.072661 | 0.52896 | *CD3D* |
| ENSG00000062038 | 16 | 68660092 | 68766519 | 354 | 34 | 48720 | -0.07274 | 0.52899 | *CDH3* |
| ENSG00000257950 | 17 | 3556357 | 3609488 | 193 | 32 | 44786 | -0.07281 | 0.52902 | *P2RX5-TAX1BP3* |
| ENSG00000105464 | 19 | 48888132 | 48958188 | 203 | 40 | 35834 | -0.073096 | 0.52914 | *GRIN2D* |
| ENSG00000122335 | 6 | 158520536 | 158599312 | 217 | 21 | 50596 | -0.073123 | 0.52915 | *SERAC1* |
| ENSG00000198862 | 21 | 30290466 | 30375270 | 213 | 29 | 45747 | -0.073314 | 0.52922 | *LTN1* |
| ENSG00000060491 | 20 | 61426187 | 61455352 | 150 | 29 | 49136 | -0.073354 | 0.52924 | *OGFR* |
| ENSG00000198826 | 15 | 32897345 | 32942150 | 100 | 27 | 48856 | -0.073395 | 0.52925 | *ARHGAP11A* |
| ENSG00000168505 | 2 | 237063879 | 237087012 | 56 | 12 | 42003 | -0.073867 | 0.52944 | *GBX2* |
| ENSG00000106028 | 7 | 141428121 | 141497722 | 186 | 8 | 52361 | -0.07401 | 0.5295 | *SSBP1* |
| ENSG00000205413 | 7 | 92718829 | 92757336 | 142 | 25 | 40610 | -0.074037 | 0.52951 | *SAMD9* |
| ENSG00000147533 | 8 | 41337915 | 41378499 | 165 | 25 | 50632 | -0.074254 | 0.5296 | *GOLGA7* |
| ENSG00000267901 | 14 | 71673031 | 71693462 | 82 | 16 | 44511 | -0.074313 | 0.52962 | *AC004817.1* |
| ENSG00000104320 | 8 | 90935564 | 91025456 | 226 | 19 | 41439 | -0.074442 | 0.52967 | *NBN* |
| ENSG00000183248 | 19 | 7923605 | 7949326 | 68 | 15 | 45824 | -0.074431 | 0.52967 | *CTD-3193O13.9* |
| ENSG00000165271 | 9 | 33451439 | 33483928 | 98 | 20 | 48440 | -0.074655 | 0.52976 | *NOL6* |
| ENSG00000174721 | 10 | 93656346 | 93679240 | 70 | 17 | 49849 | -0.074849 | 0.52983 | *FGFBP3* |
| ENSG00000129170 | 11 | 19193578 | 19242120 | 152 | 38 | 46733 | -0.074946 | 0.52987 | *CSRP3* |
| ENSG00000108578 | 17 | 28565218 | 28629074 | 123 | 18 | 42541 | -0.074948 | 0.52987 | *BLMH* |
| ENSG00000196466 | 19 | 12490830 | 12522085 | 57 | 13 | 30983 | -0.074938 | 0.52987 | *ZNF799* |
| ENSG00000214046 | 19 | 16731562 | 16781253 | 134 | 25 | 41181 | -0.074945 | 0.52987 | *SMIM7* |
| ENSG00000258472 | 17 | 26772770 | 26951215 | 291 | 45 | 49058 | -0.075025 | 0.5299 | *RP11-192H23.4* |
| ENSG00000155875 | 9 | 18917656 | 19043251 | 725 | 58 | 40684 | -0.075079 | 0.52992 | *FAM154A* |
| ENSG00000168828 | 9 | 35859375 | 35880461 | 59 | 14 | 45912 | -0.075255 | 0.52999 | *OR13J1* |
| ENSG00000162624 | 1 | 75584119 | 75637218 | 111 | 20 | 39926 | -0.075313 | 0.53002 | *LHX8* |
| ENSG00000131931 | 8 | 42681817 | 42708468 | 48 | 12 | 38008 | -0.075422 | 0.53006 | *THAP1* |
| ENSG00000224420 | 19 | 50181921 | 50203832 | 49 | 14 | 39371 | -0.075658 | 0.53015 | *ADM5* |
| ENSG00000141979 | 19 | 16579878 | 16749015 | 437 | 47 | 47868 | -0.075837 | 0.53023 | *CTD-3222D19.2* |
| ENSG00000102763 | 13 | 42130973 | 42545256 | 1217 | 91 | 47180 | -0.076325 | 0.53042 | *VWA8* |
| ENSG00000131737 | 17 | 39523902 | 39548655 | 47 | 15 | 43973 | -0.076564 | 0.53051 | *KRT34* |
| ENSG00000117971 | 15 | 78906461 | 79030096 | 389 | 42 | 39920 | -0.076571 | 0.53052 | *CHRNB4* |
| ENSG00000196812 | 6 | 28082338 | 28107860 | 83 | 10 | 48131 | -0.076951 | 0.53067 | *ZSCAN16* |

| | | | | | | | | | |
|---|---|---|---|---|---|---|---|---|---|
| ENSG00000134539 | 12 | 10368657 | 10479850 | 301 | 34 | 38447 | -0.076991 | 0.53068 | *KLRD1* |
| ENSG00000111752 | 12 | 9056492 | 9104063 | 150 | 18 | 41412 | -0.077265 | 0.53079 | *PHC1* |
| ENSG00000142634 | 1 | 15726391 | 15766839 | 127 | 34 | 48250 | -0.077846 | 0.53102 | *EFHD2* |
| ENSG00000104332 | 8 | 41109481 | 41177016 | 128 | 26 | 47730 | -0.078015 | 0.53109 | *SFRP1* |
| ENSG00000183421 | 21 | 43149529 | 43197266 | 225 | 33 | 53904 | -0.078217 | 0.53117 | *RIPK4* |
| ENSG00000137414 | 6 | 17590586 | 17621950 | 78 | 20 | 44296 | -0.078985 | 0.53148 | *FAM8A1* |
| ENSG00000102898 | 16 | 67870635 | 67916470 | 68 | 15 | 44928 | -0.079028 | 0.53149 | *NUTF2* |
| ENSG00000106524 | 7 | 16629401 | 16695442 | 215 | 32 | 49259 | -0.079352 | 0.53162 | *ANKMY2* |
| ENSG00000070388 | 19 | 629879 | 653703 | 135 | 38 | 42454 | -0.079435 | 0.53166 | *FGF22* |
| ENSG00000116014 | 19 | 907287 | 931015 | 72 | 20 | 44802 | -0.079474 | 0.53167 | *KISS1R* |
| ENSG00000071051 | 2 | 106351354 | 106520730 | 643 | 59 | 48979 | -0.079518 | 0.53169 | *NCK2* |
| ENSG00000152253 | 2 | 169680642 | 169779881 | 302 | 43 | 44381 | -0.079841 | 0.53182 | *SPC25* |
| ENSG00000037757 | 19 | 13865346 | 13895096 | 126 | 16 | 26899 | -0.079856 | 0.53182 | *MRI1* |
| ENSG00000225781 | 7 | 142739417 | 142760444 | 61 | 10 | 41036 | -0.080234 | 0.53197 | *OR6V1* |
| ENSG00000083720 | 5 | 41720167 | 41880621 | 248 | 23 | 50078 | -0.080286 | 0.532 | *OXCT1* |
| ENSG00000110315 | 11 | 10523225 | 10572777 | 207 | 28 | 40915 | -0.080437 | 0.53206 | *RNF141* |
| ENSG00000177291 | 10 | 35884338 | 35907863 | 73 | 16 | 37219 | -0.080524 | 0.53209 | *GJD4* |
| ENSG00000136193 | 7 | 29949719 | 30039905 | 266 | 31 | 55069 | -0.080549 | 0.5321 | *SCRN1* |
| ENSG00000115648 | 2 | 238384071 | 238473961 | 473 | 49 | 42913 | -0.08061 | 0.53212 | *MLPH* |
| ENSG00000150625 | 4 | 176544085 | 176933815 | 1315 | 149 | 45624 | -0.080655 | 0.53214 | *GPM6A* |
| ENSG00000115649 | 2 | 220026619 | 220052828 | 53 | 7 | 31231 | -0.080667 | 0.53215 | *CNPPD1* |
| ENSG00000174776 | 3 | 167186472 | 167381771 | 353 | 55 | 47012 | -0.080804 | 0.5322 | *WDR49* |
| ENSG00000180739 | 19 | 10613623 | 10638607 | 90 | 24 | 50411 | -0.080834 | 0.53221 | *S1PR5* |
| ENSG00000004948 | 7 | 93043799 | 93214042 | 471 | 55 | 40523 | -0.080953 | 0.53226 | *CALCR* |
| ENSG00000196792 | 14 | 31353005 | 31505607 | 421 | 36 | 43017 | -0.081651 | 0.53254 | *STRN3* |
| ENSG00000006007 | 16 | 19503015 | 19543467 | 162 | 13 | 34653 | -0.081725 | 0.53257 | *GDE1* |
| ENSG00000267467 | 19 | 45435495 | 45462820 | 100 | 9 | 49282 | -0.081756 | 0.53258 | *APOC4* |
| ENSG00000224916 | 19 | 45435495 | 45462822 | 100 | 9 | 49282 | -0.081756 | 0.53258 | *APOC4-APOC2* |
| ENSG00000187098 | 3 | 69778586 | 70027488 | 553 | 39 | 37988 | -0.081818 | 0.5326 | *MITF* |
| ENSG00000226887 | 4 | 53578785 | 53627807 | 95 | 8 | 56105 | -0.081993 | 0.53267 | *ERVMER34-1* |
| ENSG00000204138 | 1 | 28686114 | 28836881 | 552 | 19 | 29343 | -0.08202 | 0.53268 | *PHACTR4* |
| ENSG00000108219 | 10 | 82203922 | 82302879 | 291 | 37 | 44280 | -0.082314 | 0.5328 | *TSPAN14* |
| ENSG00000119862 | 2 | 64671103 | 64698515 | 73 | 22 | 45236 | -0.082445 | 0.53285 | *LGALSL* |
| ENSG00000150967 | 12 | 123395498 | 123476196 | 92 | 22 | 40890 | -0.082545 | 0.53289 | *ABCB9* |
| ENSG00000113658 | 5 | 135458534 | 135534435 | 197 | 8 | 39474 | -0.082569 | 0.5329 | *SMAD5* |
| ENSG00000035681 | 8 | 59486063 | 59582403 | 240 | 30 | 46340 | -0.082568 | 0.5329 | *NSMAF* |

| | | | | | | | | | |
|---|---|---|---|---|---|---|---|---|---|
| ENSG00000269741 | 19 | 51489274 | 51522837 | 140 | 18 | 40233 | -0.08281 | 0.533 | *KLK9* |
| ENSG00000132676 | 1 | 155647751 | 155718801 | 92 | 18 | 40330 | -0.083127 | 0.53312 | *DAP3* |
| ENSG00000171786 | 1 | 160326857 | 160352638 | 66 | 16 | 40502 | -0.08316 | 0.53314 | *NHLH1* |
| ENSG00000099617 | 19 | 1276153 | 1311430 | 66 | 21 | 41094 | -0.083201 | 0.53315 | *EFNA2* |
| ENSG00000167716 | 17 | 1609817 | 1651893 | 138 | 19 | 53383 | -0.083405 | 0.53324 | *WDR81* |
| ENSG00000186825 | 2 | 132542534 | 132569234 | 90 | 12 | 39637 | -0.083434 | 0.53325 | *C2orf27B* |
| ENSG00000091986 | 3 | 112313407 | 112378377 | 172 | 36 | 40980 | -0.083493 | 0.53327 | *CCDC80* |
| ENSG00000164520 | 6 | 150194511 | 150229238 | 127 | 10 | 42911 | -0.083583 | 0.53331 | *RAET1E* |
| ENSG00000142182 | 21 | 45656222 | 45692099 | 124 | 31 | 38314 | -0.083911 | 0.53344 | *DNMT3L* |
| ENSG00000168263 | 9 | 2707502 | 2740037 | 105 | 24 | 44959 | -0.083983 | 0.53346 | *KCNV2* |
| ENSG00000108953 | 17 | 1237566 | 1313672 | 355 | 43 | 47403 | -0.084018 | 0.53348 | *YWHAE* |
| ENSG00000171505 | 9 | 125278637 | 125299572 | 113 | 18 | 33384 | -0.084369 | 0.53362 | *OR1N1* |
| ENSG00000144229 | 2 | 137513115 | 138445287 | 3746 | 228 | 39013 | -0.084536 | 0.53368 | *THSD7B* |
| ENSG00000137731 | 11 | 117661559 | 117709413 | 161 | 34 | 47164 | -0.08459 | 0.53371 | *FXYD2* |
| ENSG00000164828 | 7 | 845528 | 946072 | 464 | 47 | 52149 | -0.084648 | 0.53373 | *SUN1* |
| ENSG00000162998 | 2 | 183688002 | 183741890 | 165 | 27 | 43283 | -0.085155 | 0.53393 | *FRZB* |
| ENSG00000213638 | 19 | 1895377 | 1923444 | 97 | 15 | 38438 | -0.085257 | 0.53397 | *ADAT3* |
| ENSG00000171360 | 17 | 39582698 | 39607173 | 128 | 15 | 52531 | -0.085349 | 0.53401 | *KRT38* |
| ENSG00000197702 | 11 | 12388732 | 12562348 | 572 | 92 | 41429 | -0.085457 | 0.53405 | *PARVA* |
| ENSG00000083093 | 16 | 23604488 | 23662631 | 222 | 25 | 49328 | -0.085724 | 0.53416 | *PALB2* |
| ENSG00000130640 | 10 | 135083135 | 135135841 | 165 | 23 | 42950 | -0.08622 | 0.53435 | *TUBGCP2* |
| ENSG00000134200 | 1 | 115562415 | 115586941 | 102 | 11 | 49539 | -0.086252 | 0.53437 | *TSHB* |
| ENSG00000105298 | 19 | 3600639 | 3636813 | 152 | 23 | 42651 | -0.086295 | 0.53438 | *CACTIN* |
| ENSG00000141540 | 17 | 72199653 | 72268155 | 230 | 31 | 48254 | -0.086301 | 0.53439 | *TTYH2* |
| ENSG00000104979 | 19 | 13874982 | 13899276 | 119 | 11 | 22484 | -0.086363 | 0.53441 | *C19orf53* |
| ENSG00000171792 | 12 | 2975424 | 3008626 | 117 | 7 | 38649 | -0.086405 | 0.53443 | *RHNO1* |
| ENSG00000070371 | 22 | 19156986 | 19289239 | 445 | 36 | 48806 | -0.086573 | 0.53449 | *CLTCL1* |
| ENSG00000127054 | 1 | 1236965 | 1270071 | 70 | 17 | 35948 | -0.086657 | 0.53453 | *CPSF3L* |
| ENSG00000073536 | 17 | 33445772 | 33479334 | 51 | 20 | 47461 | -0.086661 | 0.53453 | *NLE1* |
| ENSG00000159516 | 1 | 153112058 | 153133345 | 49 | 6 | 48398 | -0.086726 | 0.53456 | *SPRR2G* |
| ENSG00000100416 | 22 | 46716772 | 46763237 | 138 | 23 | 44506 | -0.086799 | 0.53458 | *TRMU* |
| ENSG00000185418 | 15 | 102183801 | 102274807 | 217 | 29 | 31349 | -0.086883 | 0.53462 | *TARSL2* |
| ENSG00000106018 | 7 | 158810866 | 158947649 | 585 | 63 | 43375 | -0.086924 | 0.53463 | *VIPR2* |
| ENSG00000268965 | 17 | 76412409 | 76432834 | 135 | 5 | 53464 | -0.087148 | 0.53472 | *AC061992.1* |
| ENSG00000268223 | 5 | 115377163 | 115405815 | 180 | 17 | 37958 | -0.087156 | 0.53473 | *ARL14EPL* |
| ENSG00000174327 | 17 | 6929394 | 6953440 | 62 | 14 | 40950 | -0.087591 | 0.5349 | *SLC16A13* |

| | | | | | | | | | |
|---|---|---|---|---|---|---|---|---|---|
| ENSG00000174903 | 11 | 66026004 | 66054963 | 51 | 11 | 54153 | -0.087691 | 0.53494 | *RAB1B* |
| ENSG00000138083 | 2 | 45158902 | 45183216 | 85 | 22 | 40152 | -0.088097 | 0.5351 | *SIX3* |
| ENSG00000055732 | 1 | 85473765 | 85524182 | 117 | 20 | 53532 | -0.088182 | 0.53513 | *MCOLN3* |
| ENSG00000164379 | 6 | 1302675 | 1324992 | 101 | 20 | 51873 | -0.088324 | 0.53519 | *FOXQ1* |
| ENSG00000146857 | 7 | 134906731 | 134953244 | 84 | 21 | 46480 | -0.088608 | 0.5353 | *STRA8* |
| ENSG00000254726 | 1 | 156031804 | 156061789 | 41 | 13 | 36053 | -0.088716 | 0.53535 | *MEX3A* |
| ENSG00000255439 | 16 | 31084760 | 31116277 | 40 | 7 | 21971 | -0.088724 | 0.53535 | *RP11-196G11.1* |
| ENSG00000100029 | 22 | 30962612 | 31013070 | 207 | 38 | 49703 | -0.088859 | 0.5354 | *PES1* |
| ENSG00000160767 | 1 | 155206996 | 155235274 | 57 | 6 | 28680 | -0.088871 | 0.53541 | *FAM189B* |
| ENSG00000243811 | 22 | 39400368 | 39439281 | 111 | 24 | 43761 | -0.088914 | 0.53542 | *APOBEC3D* |
| ENSG00000161649 | 17 | 41914516 | 41950997 | 76 | 13 | 41515 | -0.088954 | 0.53544 | *CD300LG* |
| ENSG00000183246 | 22 | 21889646 | 21915750 | 3 | 2 | 43070 | -0.089015 | 0.53546 | *RIMBP3C* |
| ENSG00000135605 | 4 | 48127800 | 48281881 | 603 | 62 | 42473 | -0.089143 | 0.53552 | *TEC* |
| ENSG00000185513 | 20 | 42126320 | 42189590 | 193 | 30 | 52194 | -0.089408 | 0.53562 | *L3MBTL1* |
| ENSG00000133731 | 8 | 82560196 | 82608928 | 160 | 26 | 47401 | -0.089428 | 0.53563 | *IMPA1* |
| ENSG00000101544 | 18 | 77856915 | 77915406 | 200 | 20 | 43088 | -0.089448 | 0.53564 | *ADNP2* |
| ENSG00000153802 | 4 | 68676594 | 68759750 | 138 | 36 | 50621 | -0.089618 | 0.5357 | *TMPRSS11D* |
| ENSG00000188487 | 11 | 15123970 | 15278754 | 554 | 91 | 42609 | -0.089607 | 0.5357 | *INSC* |
| ENSG00000180251 | 2 | 103079762 | 103160431 | 347 | 26 | 51240 | -0.089632 | 0.53571 | *SLC9A4* |
| ENSG00000119013 | 2 | 201926156 | 201960473 | 80 | 15 | 46901 | -0.089625 | 0.53571 | *NDUFB3* |
| ENSG00000164104 | 4 | 174242846 | 174266276 | 50 | 11 | 31695 | -0.089677 | 0.53573 | *HMGB2* |
| ENSG00000162771 | 1 | 212787789 | 212810120 | 70 | 22 | 42389 | -0.089763 | 0.53576 | *FAM71A* |
| ENSG00000257093 | 7 | 141346528 | 141411953 | 121 | 18 | 31673 | -0.089807 | 0.53578 | *KIAA1147* |
| ENSG00000167618 | 19 | 54999100 | 55031897 | 191 | 43 | 40939 | -0.089825 | 0.53579 | *LAIR2* |
| ENSG00000221932 | 11 | 124779146 | 124800573 | 83 | 26 | 44663 | -0.08998 | 0.53585 | *HEPN1* |
| ENSG00000050344 | 7 | 26181860 | 26236745 | 140 | 23 | 48747 | -0.090015 | 0.53586 | *NFE2L3* |
| ENSG00000137876 | 15 | 55463004 | 55499265 | 179 | 16 | 35630 | -0.090246 | 0.53595 | *RSL24D1* |
| ENSG00000180776 | 13 | 21940263 | 22043509 | 276 | 26 | 34318 | -0.090413 | 0.53602 | *ZDHHC20* |
| ENSG00000164916 | 7 | 4673388 | 4821074 | 502 | 81 | 45043 | -0.090435 | 0.53603 | *FOXK1* |
| ENSG00000055163 | 5 | 156683089 | 156832606 | 496 | 42 | 40904 | -0.090519 | 0.53606 | *CYFIP2* |
| ENSG00000109586 | 4 | 174079904 | 174255118 | 417 | 57 | 40035 | -0.09053 | 0.53607 | *GALNT7* |
| ENSG00000198729 | 6 | 150454212 | 150581493 | 538 | 93 | 39294 | -0.090601 | 0.5361 | *PPP1R14C* |
| ENSG00000101181 | 20 | 60748085 | 60788624 | 207 | 34 | 41711 | -0.090839 | 0.53619 | *MTG2* |
| ENSG00000184908 | 1 | 16360272 | 16393803 | 272 | 34 | 43987 | -0.090888 | 0.53621 | *CLCNKB* |
| ENSG00000111144 | 12 | 96384606 | 96447298 | 231 | 32 | 40612 | -0.090903 | 0.53622 | *LTA4H* |
| ENSG00000127804 | 17 | 2298856 | 2425185 | 352 | 53 | 41738 | -0.09092 | 0.53622 | *METTL16* |

| | | | | | | | | | |
|---|---|---|---|---|---|---|---|---|---|
| ENSG00000140534 | 15 | 90108713 | 90184287 | 218 | 19 | 44520 | -0.091132 | 0.53631 | *TICRR* |
| ENSG00000178927 | 17 | 80390465 | 80418705 | 114 | 15 | 41016 | -0.091566 | 0.53648 | *C17orf62* |
| ENSG00000186407 | 17 | 72596026 | 72629897 | 125 | 21 | 51632 | -0.091897 | 0.53661 | *CD300E* |
| ENSG00000167081 | 9 | 128499624 | 128739656 | 373 | 33 | 41386 | -0.091994 | 0.53665 | *PBX3* |
| ENSG00000214022 | 7 | 150055278 | 150081133 | 87 | 9 | 49514 | -0.09211 | 0.53669 | *REPIN1* |
| ENSG00000257327 | 12 | 104225229 | 104248442 | 129 | 19 | 48941 | -0.092549 | 0.53687 | *RP11-650K20.3* |
| ENSG00000177963 | 11 | 197511 | 225113 | 120 | 21 | 46619 | -0.09276 | 0.53695 | *RIC8A* |
| ENSG00000233436 | 11 | 57500986 | 57529253 | 39 | 9 | 43467 | -0.093272 | 0.53716 | *BTBD18* |
| ENSG00000165028 | 9 | 107516438 | 107549738 | 105 | 22 | 43711 | -0.093391 | 0.5372 | *NIPSNAP3B* |
| ENSG00000182521 | 14 | 55870259 | 55933444 | 294 | 27 | 44451 | -0.093374 | 0.5372 | *TBPL2* |
| ENSG00000111404 | 12 | 18223803 | 18483041 | 1011 | 76 | 51867 | -0.093429 | 0.53722 | *RERGL* |
| ENSG00000196689 | 17 | 3458738 | 3510392 | 210 | 35 | 44128 | -0.093468 | 0.53723 | *TRPV1* |
| ENSG00000261701 | 16 | 72078522 | 72121145 | 111 | 18 | 51258 | -0.093502 | 0.53725 | *HPR* |
| ENSG00000147419 | 8 | 27580835 | 27640170 | 239 | 18 | 49121 | -0.093709 | 0.53733 | *CCDC25* |
| ENSG00000260371 | 16 | 69358994 | 69400209 | 90 | 19 | 54389 | -0.094044 | 0.53746 | *RP11-343C2.9* |
| ENSG00000198522 | 2 | 27841114 | 27884375 | 79 | 14 | 43062 | -0.094117 | 0.53749 | *GPN1* |
| ENSG00000164125 | 4 | 159035626 | 159104470 | 192 | 18 | 35428 | -0.09418 | 0.53752 | *FAM198B* |
| ENSG00000179912 | 12 | 57633392 | 57834788 | 282 | 22 | 44172 | -0.094346 | 0.53758 | *R3HDM2* |
| ENSG00000256269 | 11 | 118945576 | 118974259 | 65 | 12 | 42929 | -0.094614 | 0.53769 | *HMBS* |
| ENSG00000010292 | 12 | 6592522 | 6651121 | 229 | 20 | 41045 | -0.094757 | 0.53775 | *NCAPD2* |
| ENSG00000203813 | 6 | 27767842 | 27788314 | 53 | 11 | 36425 | -0.094963 | 0.53783 | *HIST1H3H* |
| ENSG00000175756 | 1 | 1299110 | 1320875 | 71 | 11 | 37302 | -0.095254 | 0.53794 | *AURKAIP1* |
| ENSG00000106789 | 9 | 100873257 | 100964922 | 348 | 64 | 45514 | -0.09527 | 0.53795 | *CORO2A* |
| ENSG00000132182 | 3 | 13347737 | 13471809 | 454 | 31 | 33331 | -0.095364 | 0.53799 | *NUP210* |
| ENSG00000162437 | 1 | 65200778 | 65308915 | 247 | 32 | 43429 | -0.095388 | 0.538 | *RAVER2* |
| ENSG00000169575 | 22 | 22589087 | 22609927 | 109 | 18 | 47293 | -0.09551 | 0.53804 | *VPREB1* |
| ENSG00000237896 | 7 | 80794824 | 80838431 | 207 | 30 | 38259 | -0.095813 | 0.53817 | *AC005008.2* |
| ENSG00000164897 | 7 | 150768167 | 150790633 | 55 | 12 | 41015 | -0.095841 | 0.53818 | *TMUB1* |
| ENSG00000144476 | 2 | 237466430 | 237501001 | 88 | 18 | 37522 | -0.095973 | 0.53823 | *ACKR3* |
| ENSG00000164124 | 4 | 159112756 | 159186563 | 125 | 35 | 43274 | -0.096118 | 0.53829 | *TMEM144* |
| ENSG00000153391 | 18 | 33022194 | 33087955 | 147 | 25 | 53542 | -0.096298 | 0.53836 | *INO80C* |
| ENSG00000124374 | 2 | 71399869 | 71464213 | 284 | 32 | 42599 | -0.096338 | 0.53837 | *PAIP2B* |
| ENSG00000102978 | 16 | 57486299 | 57515922 | 97 | 22 | 52441 | -0.096447 | 0.53842 | *POLR2C* |
| ENSG00000099974 | 22 | 24299089 | 24324721 | 23 | 9 | 43357 | -0.096477 | 0.53843 | *DDTL* |
| ENSG00000151882 | 5 | 43366747 | 43422493 | 162 | 21 | 31495 | -0.096965 | 0.53862 | *CCL28* |
| ENSG00000160695 | 11 | 118928403 | 118962688 | 105 | 11 | 47042 | -0.096947 | 0.53862 | *VPS11* |

| | | | | | | | | | |
|---|---|---|---|---|---|---|---|---|---|
| ENSG00000108773 | 17 | 40255126 | 40283376 | 62 | 7 | 27410 | -0.096962 | 0.53862 | *KAT2A* |
| ENSG00000163251 | 2 | 208617310 | 208644287 | 58 | 18 | 41470 | -0.097084 | 0.53867 | *FZD5* |
| ENSG00000083842 | 19 | 58780317 | 58817254 | 134 | 21 | 43650 | -0.097315 | 0.53876 | *ZNF8* |
| ENSG00000273439 | 19 | 58780318 | 58817254 | 134 | 21 | 43650 | -0.097315 | 0.53876 | *ZNF8* |
| ENSG00000164128 | 4 | 164235113 | 164275984 | 138 | 28 | 39201 | -0.097326 | 0.53877 | *NPY1R* |
| ENSG00000104921 | 19 | 7743644 | 7777032 | 266 | 25 | 41097 | -0.09745 | 0.53882 | *FCER2* |
| ENSG00000163655 | 3 | 155578325 | 155671815 | 327 | 17 | 49419 | -0.097513 | 0.53884 | *GMPS* |
| ENSG00000139910 | 14 | 26902299 | 27076960 | 271 | 28 | 48050 | -0.097506 | 0.53884 | *NOVA1* |
| ENSG00000158615 | 1 | 204362515 | 204390919 | 61 | 20 | 37201 | -0.097563 | 0.53886 | *PPP1R15B* |
| ENSG00000142875 | 1 | 84533745 | 84714181 | 300 | 32 | 44395 | -0.097668 | 0.5389 | *PRKACB* |
| ENSG00000184384 | 11 | 95699762 | 96086344 | 1396 | 219 | 47543 | -0.097875 | 0.53898 | *MAML2* |
| ENSG00000197790 | 11 | 4556421 | 4577374 | 206 | 15 | 55429 | -0.097917 | 0.539 | *OR52M1* |
| ENSG00000259133 | 14 | 57105683 | 57207253 | 361 | 57 | 36262 | -0.098055 | 0.53906 | *RP11-1085N6.3* |
| ENSG00000158869 | 1 | 161175024 | 161200489 | 65 | 18 | 50976 | -0.098572 | 0.53926 | *FCER1G* |
| ENSG00000257482 | 7 | 63495821 | 63548927 | 235 | 14 | 38460 | -0.09875 | 0.53933 | *ZNF727* |
| ENSG00000142002 | 19 | 4665236 | 4734685 | 199 | 38 | 41316 | -0.098899 | 0.53939 | *DPP9* |
| ENSG00000198026 | 20 | 44567292 | 44610833 | 116 | 24 | 41558 | -0.098905 | 0.53939 | *ZNF335* |
| ENSG00000178115 | 15 | 30834183 | 30864704 | 1 | 1 | 43450 | -0.098922 | 0.5394 | *GOLGA8Q* |
| ENSG00000269866 | 16 | 70248261 | 70268641 | 21 | 10 | 43910 | -0.099009 | 0.53943 | *FKSG63* |
| ENSG00000109016 | 17 | 21016677 | 21106578 | 175 | 26 | 48693 | -0.098992 | 0.53943 | *DHRS7B* |
| ENSG00000126882 | 9 | 134123463 | 134161934 | 85 | 20 | 49888 | -0.099072 | 0.53946 | *FAM78A* |
| ENSG00000116266 | 1 | 109279296 | 109362148 | 294 | 15 | 34358 | -0.099192 | 0.53951 | *STXBP3* |
| ENSG00000111716 | 12 | 21778276 | 21920791 | 240 | 18 | 52721 | -0.09974 | 0.53972 | *LDHB* |
| ENSG00000141434 | 18 | 29755032 | 29810367 | 166 | 27 | 46081 | -0.10011 | 0.53987 | *MEP1B* |
| ENSG00000227151 | 13 | 57724766 | 57747787 | 7 | 4 | 35865 | -0.10015 | 0.53989 | *PRR20D* |
| ENSG00000168818 | 4 | 4407469 | 4554073 | 421 | 38 | 43642 | -0.10017 | 0.5399 | *STX18* |
| ENSG00000249853 | 6 | 114366750 | 114674209 | 706 | 106 | 53300 | -0.10024 | 0.53992 | *HS3ST5* |
| ENSG00000261609 | 16 | 81338557 | 81434489 | 403 | 54 | 40449 | -0.10025 | 0.53993 | *GAN* |
| ENSG00000154548 | 6 | 89795678 | 89837800 | 151 | 17 | 44091 | -0.10047 | 0.54002 | *SRSF12* |
| ENSG00000136754 | 10 | 27025522 | 27160016 | 456 | 28 | 33121 | -0.10052 | 0.54003 | *ABI1* |
| ENSG00000213022 | 19 | 51493806 | 51522890 | 123 | 17 | 40769 | -0.10109 | 0.54026 | *KLK9* |
| ENSG00000035499 | 5 | 59882739 | 60006017 | 255 | 24 | 38981 | -0.10115 | 0.54029 | *DEPDC1B* |
| ENSG00000149021 | 11 | 62162575 | 62200667 | 156 | 28 | 48595 | -0.1012 | 0.5403 | *SCGB1A1* |
| ENSG00000184988 | 17 | 41353854 | 41382061 | 49 | 5 | 47368 | -0.10118 | 0.5403 | *TMEM106A* |
| ENSG00000187535 | 16 | 1550428 | 1672111 | 438 | 39 | 40372 | -0.10122 | 0.54031 | *IFT140* |
| ENSG00000142444 | 19 | 11029409 | 11054211 | 56 | 9 | 54644 | -0.10166 | 0.54049 | *C19orf52* |

| | | | | | | | | | |
|---|---|---|---|---|---|---|---|---|---|
| ENSG00000154856 | 18 | 10444625 | 10499945 | 197 | 33 | 50172 | -0.10174 | 0.54052 | *APCDD1* |
| ENSG00000004975 | 17 | 7118660 | 7147864 | 37 | 9 | 53156 | -0.10188 | 0.54058 | *DVL2* |
| ENSG00000172239 | 5 | 43516369 | 43567860 | 150 | 16 | 36563 | -0.10198 | 0.54061 | *PAIP1* |
| ENSG00000173699 | 2 | 231850836 | 231899770 | 257 | 32 | 45469 | -0.10216 | 0.54068 | *SPATA3* |
| ENSG00000151881 | 5 | 43434354 | 43493995 | 189 | 18 | 32423 | -0.10214 | 0.54068 | *C5orf28* |
| ENSG00000081913 | 18 | 60372672 | 60657666 | 541 | 51 | 49765 | -0.1023 | 0.54074 | *PHLPP1* |
| ENSG00000186205 | 1 | 220950101 | 220997735 | 166 | 33 | 44998 | -0.10252 | 0.54083 | *Mar-01* |
| ENSG00000118094 | 11 | 118518026 | 118560399 | 146 | 8 | 54857 | -0.10264 | 0.54088 | *TREH* |
| ENSG00000140832 | 16 | 71650064 | 71686017 | 80 | 15 | 46777 | -0.10282 | 0.54095 | *MARVELD3* |
| ENSG00000181418 | 12 | 49378932 | 49403092 | 37 | 14 | 49908 | -0.10286 | 0.54096 | *DDN* |
| ENSG00000135040 | 9 | 88546061 | 88647213 | 136 | 36 | 46479 | -0.10292 | 0.54099 | *NAA35* |
| ENSG00000092108 | 14 | 31081318 | 31215018 | 368 | 37 | 38126 | -0.10316 | 0.54108 | *SCFD1* |
| ENSG00000184678 | 1 | 149846010 | 149868232 | 37 | 5 | 55079 | -0.1032 | 0.5411 | *HIST2H2BE* |
| ENSG00000173114 | 7 | 110721062 | 110775510 | 96 | 17 | 35528 | -0.10335 | 0.54116 | *LRRN3* |
| ENSG00000204300 | 11 | 123743633 | 123766349 | 79 | 15 | 55860 | -0.1035 | 0.54122 | *TMEM225* |
| ENSG00000204356 | 6 | 31909864 | 31936887 | 84 | 22 | 41563 | -0.10354 | 0.54123 | *NELFE* |
| ENSG00000214854 | 22 | 39646865 | 39667063 | 97 | 27 | 39666 | -0.10362 | 0.54126 | *AL031590.1* |
| ENSG00000114520 | 3 | 125155495 | 125249041 | 307 | 29 | 39187 | -0.1037 | 0.54129 | *SNX4* |
| ENSG00000120210 | 9 | 5121979 | 5195668 | 206 | 12 | 37019 | -0.10377 | 0.54133 | *INSL6* |
| ENSG00000214919 | 3 | 155449933 | 155471515 | 68 | 15 | 39207 | -0.10387 | 0.54137 | *AC104472.1* |
| ENSG00000129757 | 11 | 2894443 | 2917111 | 68 | 18 | 46633 | -0.10427 | 0.54152 | *CDKN1C* |
| ENSG00000169180 | 16 | 28099300 | 28233241 | 111 | 16 | 53710 | -0.10454 | 0.54163 | *XPO6* |
| ENSG00000188293 | 19 | 46723009 | 46744500 | 111 | 19 | 51659 | -0.10476 | 0.54172 | *IGFL1* |
| ENSG00000107521 | 10 | 100165955 | 100216684 | 234 | 25 | 38630 | -0.10496 | 0.54179 | *HPS1* |
| ENSG00000203630 | 20 | 1306228 | 1327555 | 64 | 11 | 34455 | -0.10551 | 0.54201 | *AL136531.1* |
| ENSG00000226894 | 4 | 69044242 | 69093798 | 138 | 7 | 29388 | -0.10557 | 0.54204 | *TMPRSS11BNL* |
| ENSG00000160446 | 9 | 131473148 | 131496406 | 25 | 12 | 44133 | -0.10558 | 0.54204 | *ZDHHC12* |
| ENSG00000120907 | 8 | 26595667 | 26734790 | 650 | 90 | 43152 | -0.10593 | 0.54218 | *ADRA1A* |
| ENSG00000119457 | 9 | 115631200 | 115663193 | 120 | 19 | 49650 | -0.10596 | 0.54219 | *SLC46A2* |
| ENSG00000170605 | 12 | 55513465 | 55534586 | 119 | 4 | 54400 | -0.10609 | 0.54224 | *OR9K2* |
| ENSG00000105889 | 7 | 22449063 | 22682544 | 824 | 111 | 48011 | -0.10636 | 0.54235 | *STEAP1B* |
| ENSG00000108465 | 17 | 46035176 | 46069140 | 111 | 17 | 47429 | -0.10647 | 0.5424 | *CDK5RAP3* |
| ENSG00000146555 | 7 | 3331080 | 4318632 | 4280 | 201 | 43356 | -0.10664 | 0.54246 | *SDK1* |
| ENSG00000147926 | 9 | 40690291 | 40716537 | 1 | 1 | 42379 | -0.10673 | 0.5425 | *SPATA31A3* |
| ENSG00000162641 | 1 | 109348520 | 109516106 | 679 | 79 | 46423 | -0.10679 | 0.54252 | *AKNAD1* |
| ENSG00000141232 | 17 | 48929584 | 48955339 | 58 | 13 | 43215 | -0.10685 | 0.54255 | *TOB1* |

| | | | | | | | | | |
|---|---|---|---|---|---|---|---|---|---|
| ENSG00000005884 | 17 | 48123332 | 48177845 | 157 | 25 | 33596 | -0.10713 | 0.54266 | *ITGA3* |
| ENSG00000027869 | 1 | 156766035 | 156796654 | 78 | 15 | 39783 | -0.10718 | 0.54268 | *SH2D2A* |
| ENSG00000119899 | 6 | 74293102 | 74373878 | 483 | 20 | 42258 | -0.10725 | 0.5427 | *SLC17A5* |
| ENSG00000085365 | 5 | 77646339 | 77786562 | 433 | 26 | 39678 | -0.10728 | 0.54272 | *SCAMP1* |
| ENSG00000139645 | 12 | 56621591 | 56662175 | 49 | 6 | 49948 | -0.10742 | 0.54277 | *ANKRD52* |
| ENSG00000172456 | 1 | 59752310 | 60243347 | 1295 | 122 | 42393 | -0.10752 | 0.54281 | *FGGY* |
| ENSG00000100448 | 14 | 25032728 | 25055466 | 77 | 15 | 53807 | -0.10766 | 0.54287 | *CTSG* |
| ENSG00000139508 | 13 | 29264201 | 29303107 | 83 | 15 | 49094 | -0.10772 | 0.54289 | *SLC46A3* |
| ENSG00000273395 | 1 | 48216979 | 48237290 | 64 | 19 | 51855 | -0.10776 | 0.54291 | *FLJ00388* |
| ENSG00000242485 | 1 | 1327288 | 1352693 | 55 | 7 | 28903 | -0.10797 | 0.54299 | *MRPL20* |
| ENSG00000139180 | 12 | 4748261 | 4808454 | 188 | 33 | 38221 | -0.10817 | 0.54307 | *NDUFA9* |
| ENSG00000057252 | 1 | 179252925 | 179337815 | 408 | 32 | 50629 | -0.10836 | 0.54314 | *SOAT1* |
| ENSG00000183853 | 1 | 157953063 | 158080052 | 426 | 82 | 43628 | -0.10855 | 0.54322 | *KIRREL* |
| ENSG00000157954 | 7 | 5219819 | 5283457 | 299 | 48 | 52102 | -0.10864 | 0.54325 | *WIPI2* |
| ENSG00000261427 | 16 | 19115324 | 19194900 | 148 | 36 | 37677 | -0.10885 | 0.54334 | *CTD-2349B8.1* |
| ENSG00000196844 | 11 | 125636008 | 125658714 | 85 | 13 | 54542 | -0.10916 | 0.54346 | *PATE2* |
| ENSG00000130770 | 1 | 28552620 | 28583417 | 120 | 11 | 40166 | -0.10921 | 0.54348 | *ATPIF1* |
| ENSG00000106331 | 7 | 127240346 | 127265982 | 60 | 10 | 34988 | -0.10941 | 0.54356 | *PAX4* |
| ENSG00000124772 | 6 | 36698552 | 36817778 | 489 | 88 | 50330 | -0.10967 | 0.54367 | *CPNE5* |
| ENSG00000175779 | 15 | 38978799 | 39002239 | 103 | 18 | 44062 | -0.10986 | 0.54374 | *C15orf53* |
| ENSG00000115758 | 2 | 10570094 | 10598630 | 77 | 14 | 49893 | -0.11002 | 0.54381 | *ODC1* |
| ENSG00000123689 | 1 | 209838765 | 209859733 | 43 | 11 | 45477 | -0.11039 | 0.54395 | *G0S2* |
| ENSG00000269295 | 9 | 71859331 | 71879642 | 106 | 35 | 54241 | -0.11055 | 0.54401 | *AL358113.1* |
| ENSG00000125731 | 19 | 6742171 | 6777599 | 100 | 17 | 49820 | -0.11055 | 0.54401 | *SH2D3A* |
| ENSG00000166930 | 11 | 60187062 | 60232687 | 168 | 25 | 48860 | -0.11083 | 0.54412 | *MS4A5* |
| ENSG00000143867 | 2 | 19541246 | 19568414 | 67 | 21 | 41745 | -0.11085 | 0.54413 | *OSR1* |
| ENSG00000147509 | 8 | 54754368 | 54881863 | 123 | 25 | 40442 | -0.11094 | 0.54417 | *RGS20* |
| ENSG00000243056 | 5 | 139917251 | 139939163 | 35 | 10 | 50435 | -0.11103 | 0.54421 | *EIF4EBP3* |
| ENSG00000176937 | 11 | 4814663 | 4835847 | 155 | 13 | 30932 | -0.1111 | 0.54423 | *OR52R1* |
| ENSG00000196184 | 1 | 159399512 | 159420600 | 86 | 19 | 40660 | -0.11114 | 0.54425 | *OR10J1* |
| ENSG00000174840 | 3 | 57532003 | 57562571 | 80 | 14 | 41446 | -0.11121 | 0.54427 | *PDE12* |
| ENSG00000159403 | 12 | 7177513 | 7255203 | 65 | 19 | 46815 | -0.11152 | 0.5444 | *C1R* |
| ENSG00000125430 | 17 | 14194400 | 14262721 | 254 | 70 | 43934 | -0.11173 | 0.54448 | *HS3ST3B1* |
| ENSG00000198788 | 11 | 1064875 | 1114419 | 259 | 44 | 38202 | -0.11188 | 0.54454 | *MUC2* |
| ENSG00000130338 | 6 | 158723692 | 158942860 | 852 | 54 | 42365 | -0.11198 | 0.54458 | *TULP4* |
| ENSG00000196323 | 11 | 130086572 | 130194581 | 342 | 25 | 30077 | -0.11199 | 0.54459 | *ZBTB44* |

| | | | | | | | | | |
|---|---|---|---|---|---|---|---|---|---|
| ENSG00000134489 | 18 | 22030593 | 22069921 | 125 | 29 | 52787 | -0.11201 | 0.54459 | *HRH4* |
| ENSG00000123307 | 12 | 55403729 | 55433798 | 64 | 12 | 38540 | -0.11225 | 0.54469 | *NEUROD4* |
| ENSG00000257017 | 16 | 72078491 | 72104954 | 60 | 10 | 53222 | -0.11253 | 0.5448 | *HP* |
| ENSG00000149582 | 11 | 118391756 | 118427995 | 85 | 8 | 28621 | -0.1128 | 0.54491 | *TMEM25* |
| ENSG00000078747 | 20 | 32941041 | 33109198 | 271 | 16 | 53285 | -0.11293 | 0.54496 | *ITCH* |
| ENSG00000150768 | 11 | 111885538 | 111945114 | 120 | 17 | 32814 | -0.11301 | 0.54499 | *DLAT* |
| ENSG00000145839 | 5 | 135217935 | 135241516 | 74 | 15 | 54733 | -0.11324 | 0.54508 | *IL9* |
| ENSG00000168748 | 16 | 66868282 | 66898056 | 45 | 7 | 42130 | -0.11326 | 0.54509 | *CA7* |
| ENSG00000169635 | 22 | 21761693 | 21815752 | 23 | 9 | 32910 | -0.11386 | 0.54533 | *HIC2* |
| ENSG00000206532 | 3 | 110597231 | 110622323 | 53 | 14 | 37172 | -0.11399 | 0.54538 | *RP11-553A10.1* |
| ENSG00000160181 | 21 | 43756466 | 43781237 | 137 | 27 | 46113 | -0.11399 | 0.54538 | *TFF2* |
| ENSG00000198342 | 19 | 12450185 | 12486719 | 37 | 15 | 41766 | -0.11418 | 0.54545 | *ZNF442* |
| ENSG00000187866 | 9 | 71384964 | 71408609 | 83 | 17 | 27668 | -0.11421 | 0.54546 | *FAM122A* |
| ENSG00000144791 | 3 | 45586886 | 45737830 | 608 | 60 | 51376 | -0.11434 | 0.54551 | *LIMD1* |
| ENSG00000105499 | 19 | 48541100 | 48624074 | 423 | 66 | 41269 | -0.11449 | 0.54558 | *PLA2G4C* |
| ENSG00000172575 | 15 | 38770304 | 38867776 | 259 | 33 | 50105 | -0.11465 | 0.54564 | *RASGRP1* |
| ENSG00000130755 | 19 | 39808993 | 39843012 | 87 | 18 | 44784 | -0.11479 | 0.54569 | *GMFG* |
| ENSG00000102981 | 16 | 67684849 | 67706681 | 24 | 10 | 45166 | -0.11507 | 0.5458 | *PARD6A* |
| ENSG00000159339 | 1 | 17624690 | 17700499 | 487 | 48 | 52611 | -0.11508 | 0.54581 | *PADI4* |
| ENSG00000165806 | 10 | 115428942 | 115500662 | 241 | 31 | 50356 | -0.11509 | 0.54581 | *CASP7* |
| ENSG00000179242 | 20 | 59817482 | 60525673 | 3098 | 322 | 44537 | -0.1151 | 0.54582 | *CDH4* |
| ENSG00000078898 | 20 | 31585406 | 31621515 | 127 | 33 | 46287 | -0.11523 | 0.54587 | *BPIFB2* |
| ENSG00000091009 | 5 | 145573113 | 145728814 | 336 | 27 | 50221 | -0.11526 | 0.54588 | *RBM27* |
| ENSG00000100393 | 22 | 41477790 | 41586081 | 214 | 25 | 46351 | -0.1153 | 0.5459 | *EP300* |
| ENSG00000186827 | 1 | 1136706 | 1159518 | 107 | 13 | 50211 | -0.11545 | 0.54595 | *TNFRSF4* |
| ENSG00000163818 | 3 | 45854808 | 45967534 | 230 | 34 | 38040 | -0.11548 | 0.54597 | *LZTFL1* |
| ENSG00000174233 | 12 | 49149975 | 49192820 | 80 | 12 | 40504 | -0.11552 | 0.54598 | *ADCY6* |
| ENSG00000103363 | 16 | 2811415 | 2837298 | 36 | 12 | 43232 | -0.11552 | 0.54598 | *TCEB2* |
| ENSG00000268656 | 2 | 27988851 | 28019186 | 43 | 12 | 39228 | -0.1156 | 0.54601 | *AC110084.1* |
| ENSG00000165782 | 14 | 20915878 | 20939771 | 70 | 14 | 52523 | -0.1157 | 0.54605 | *TMEM55B* |
| ENSG00000113905 | 3 | 186368005 | 186406029 | 143 | 17 | 46171 | -0.11584 | 0.54611 | *HRG* |
| ENSG00000140830 | 16 | 72068188 | 72138330 | 197 | 21 | 44141 | -0.11584 | 0.54611 | *TXNL4B* |
| ENSG00000129355 | 19 | 10667138 | 10689735 | 99 | 16 | 40177 | -0.11606 | 0.5462 | *CDKN2D* |
| ENSG00000161921 | 17 | 4626821 | 4653217 | 88 | 19 | 47001 | -0.11612 | 0.54622 | *CXCL16* |
| ENSG00000100312 | 22 | 51166624 | 51193762 | 54 | 15 | 45745 | -0.11649 | 0.54637 | *ACR* |
| ENSG00000104808 | 19 | 49426939 | 49458226 | 114 | 22 | 34889 | -0.1166 | 0.54641 | *DHDH* |

| | | | | | | | | | |
|---|---|---|---|---|---|---|---|---|---|
| ENSG00000099341 | 19 | 38855176 | 38884464 | 100 | 19 | 45266 | -0.11666 | 0.54644 | *PSMD8* |
| ENSG00000109323 | 4 | 103542660 | 103692151 | 348 | 29 | 46339 | -0.11672 | 0.54646 | *MANBA* |
| ENSG00000167765 | 19 | 52085036 | 52107630 | 132 | 27 | 34720 | -0.11676 | 0.54648 | *AC018755.1* |
| ENSG00000163737 | 4 | 74836794 | 74857841 | 59 | 10 | 30944 | -0.11717 | 0.54664 | *PF4* |
| ENSG00000183049 | 10 | 12381481 | 12887545 | 2067 | 264 | 47182 | -0.11723 | 0.54666 | *CAMK1D* |
| ENSG00000235863 | 6 | 33234917 | 33262609 | 95 | 17 | 32658 | -0.11733 | 0.5467 | *B3GALT4* |
| ENSG00000165795 | 14 | 21474922 | 21549031 | 228 | 73 | 46331 | -0.11761 | 0.54681 | *NDRG2* |
| ENSG00000153914 | 5 | 65425799 | 65489443 | 137 | 20 | 42272 | -0.11768 | 0.54684 | *SREK1* |
| ENSG00000180818 | 12 | 54368849 | 54394063 | 59 | 17 | 43004 | -0.11773 | 0.54686 | *HOXC10* |
| ENSG00000177943 | 9 | 139735395 | 139765249 | 43 | 10 | 51577 | -0.11782 | 0.5469 | *MAMDC4* |
| ENSG00000262660 | 17 | 79660401 | 79697569 | 161 | 25 | 48302 | -0.11813 | 0.54702 | *SLC25A10* |
| ENSG00000183048 | 17 | 79660404 | 79698042 | 161 | 25 | 48302 | -0.11813 | 0.54702 | *SLC25A10* |
| ENSG00000124491 | 6 | 6134318 | 6331246 | 752 | 122 | 47565 | -0.11862 | 0.54721 | *F13A1* |
| ENSG00000112249 | 6 | 100946070 | 101339248 | 1163 | 46 | 47015 | -0.11867 | 0.54723 | *ASCC3* |
| ENSG00000154781 | 3 | 14683271 | 14724166 | 124 | 14 | 39270 | -0.11881 | 0.54729 | *CCDC174* |
| ENSG00000144230 | 2 | 128393439 | 128420213 | 103 | 32 | 46834 | -0.11885 | 0.5473 | *GPR17* |
| ENSG00000102760 | 13 | 42021695 | 42055018 | 104 | 17 | 42810 | -0.11892 | 0.54733 | *RGCC* |
| ENSG00000188766 | 19 | 38869061 | 38896881 | 109 | 11 | 52327 | -0.11931 | 0.54748 | *SPRED3* |
| ENSG00000108684 | 17 | 31330105 | 32511983 | 4103 | 395 | 46588 | -0.11938 | 0.54751 | *ASIC2* |
| ENSG00000218672 | 7 | 155164771 | 155199051 | 207 | 37 | 45623 | -0.11941 | 0.54753 | *AC008060.7* |
| ENSG00000057468 | 1 | 76252567 | 76388923 | 564 | 42 | 46803 | -0.11955 | 0.54758 | *MSH4* |
| ENSG00000006327 | 16 | 3058446 | 3082384 | 54 | 14 | 51769 | -0.11977 | 0.54767 | *TNFRSF12A* |
| ENSG00000176956 | 8 | 144229331 | 144252128 | 49 | 14 | 41046 | -0.11991 | 0.54772 | *LY6H* |
| ENSG00000155761 | 1 | 118486484 | 118737846 | 515 | 36 | 31298 | -0.12036 | 0.5479 | *SPAG17* |
| ENSG00000153066 | 16 | 11762936 | 11846734 | 273 | 30 | 43814 | -0.12049 | 0.54795 | *TXNDC11* |
| ENSG00000170634 | 2 | 54187975 | 54542437 | 1179 | 79 | 49846 | -0.1211 | 0.54819 | *ACYP2* |
| ENSG00000134900 | 13 | 103239353 | 103341521 | 436 | 23 | 46606 | -0.12117 | 0.54822 | *TPP2* |
| ENSG00000204687 | 6 | 29444474 | 29465738 | 58 | 10 | 23043 | -0.12133 | 0.54829 | *MAS1L* |
| ENSG00000182704 | 11 | 76483295 | 76519198 | 102 | 18 | 38488 | -0.12144 | 0.54833 | *TSKU* |
| ENSG00000058085 | 1 | 183145373 | 183224035 | 308 | 31 | 41066 | -0.12146 | 0.54834 | *LAMC2* |
| ENSG00000131187 | 5 | 176819141 | 176846577 | 33 | 14 | 41861 | -0.12155 | 0.54837 | *F12* |
| ENSG00000013583 | 12 | 13117798 | 13163207 | 122 | 20 | 36126 | -0.12155 | 0.54837 | *HEBP1* |
| ENSG00000105290 | 19 | 36348801 | 36380693 | 72 | 5 | 26660 | -0.12192 | 0.54852 | *APLP1* |
| ENSG00000175393 | 11 | 7939180 | 7960209 | 114 | 12 | 52712 | -0.12215 | 0.54861 | *OR10A6* |
| ENSG00000196235 | 19 | 39916796 | 39977310 | 195 | 16 | 44388 | -0.12221 | 0.54863 | *SUPT5H* |
| ENSG00000105829 | 7 | 93582074 | 93643694 | 105 | 27 | 41516 | -0.12247 | 0.54874 | *BET1* |

| | | | | | | | | | |
|---|---|---|---|---|---|---|---|---|---|
| ENSG00000255482 | 11 | 102327986 | 102379532 | 158 | 28 | 53011 | -0.1225 | 0.54875 | *RP11-315O6.2* |
| ENSG00000273269 | 2 | 47283080 | 47413650 | 587 | 81 | 44395 | -0.12274 | 0.54884 | *RP11-761B3.1* |
| ENSG00000143199 | 1 | 167768625 | 167893453 | 444 | 76 | 47802 | -0.12276 | 0.54885 | *ADCY10* |
| ENSG00000148384 | 9 | 139313071 | 139344274 | 142 | 21 | 45302 | -0.12278 | 0.54886 | *INPP5E* |
| ENSG00000137507 | 11 | 76358568 | 76391791 | 75 | 23 | 50299 | -0.1228 | 0.54887 | *LRRC32* |
| ENSG00000249428 | 4 | 2441700 | 2474668 | 96 | 21 | 53302 | -0.12315 | 0.54901 | *RP11-503N18.3* |
| ENSG00000090054 | 9 | 94784281 | 94887666 | 350 | 30 | 36546 | -0.12317 | 0.54902 | *SPTLC1* |
| ENSG00000104147 | 15 | 41591466 | 41634819 | 117 | 14 | 41356 | -0.12341 | 0.54911 | *OIP5* |
| ENSG00000215298 | 8 | 23420157 | 23442974 | 151 | 19 | 52503 | -0.12358 | 0.54917 | *FP15737* |
| ENSG00000110427 | 11 | 33553618 | 33705648 | 436 | 67 | 48604 | -0.12384 | 0.54928 | *KIAA1549L* |
| ENSG00000105085 | 19 | 16675718 | 16749873 | 152 | 22 | 51271 | -0.12393 | 0.54932 | *MED26* |
| ENSG00000042832 | 8 | 133869203 | 134157147 | 1026 | 82 | 44014 | -0.12399 | 0.54934 | *TG* |
| ENSG00000112195 | 6 | 41148015 | 41178932 | 132 | 27 | 43534 | -0.12421 | 0.54942 | *TREML2* |
| ENSG00000137563 | 8 | 63917638 | 63961730 | 190 | 19 | 50560 | -0.1243 | 0.54946 | *GGH* |
| ENSG00000171931 | 17 | 18637326 | 18692662 | 152 | 25 | 47388 | -0.12469 | 0.54961 | *FBXW10* |
| ENSG00000183648 | 14 | 92572466 | 92598261 | 134 | 13 | 30086 | -0.1251 | 0.54978 | *NDUFB1* |
| ENSG00000211455 | 12 | 27386901 | 27488892 | 344 | 43 | 47438 | -0.12528 | 0.54985 | *STK38L* |
| ENSG00000181273 | 11 | 56746347 | 56767342 | 65 | 16 | 52980 | -0.12545 | 0.54992 | *OR5AK2* |
| ENSG00000258083 | 7 | 141608617 | 141629709 | 62 | 10 | 45851 | -0.12551 | 0.54994 | *OR9A4* |
| ENSG00000026025 | 10 | 17260258 | 17289592 | 52 | 16 | 37943 | -0.12563 | 0.54999 | *VIM* |
| ENSG00000124191 | 20 | 42533504 | 42708256 | 554 | 91 | 44658 | -0.12565 | 0.54999 | *TOX2* |
| ENSG00000267385 | 19 | 4518439 | 4550079 | 85 | 21 | 38565 | -0.12566 | 0.55 | *CTB-50L17.14* |
| ENSG00000134001 | 14 | 67816714 | 67863233 | 111 | 25 | 43726 | -0.12586 | 0.55008 | *EIF2S1* |
| ENSG00000026508 | 11 | 35150417 | 35263949 | 433 | 88 | 48237 | -0.12594 | 0.55011 | *CD44* |
| ENSG00000140798 | 16 | 48106884 | 48199929 | 85 | 20 | 43028 | -0.12624 | 0.55023 | *ABCC12* |
| ENSG00000196420 | 1 | 153499623 | 153524241 | 65 | 7 | 29381 | -0.12651 | 0.55034 | *S100A5* |
| ENSG00000167780 | 12 | 53487302 | 53528322 | 150 | 15 | 27532 | -0.12681 | 0.55045 | *SOAT2* |
| ENSG00000155313 | 21 | 17092344 | 17262377 | 361 | 25 | 43060 | -0.12718 | 0.5506 | *USP25* |
| ENSG00000115935 | 2 | 175414300 | 175557644 | 239 | 39 | 50537 | -0.12733 | 0.55066 | *WIPF1* |
| ENSG00000241598 | 11 | 1632188 | 1653368 | 176 | 23 | 54228 | -0.12745 | 0.55071 | *KRTAP5-4* |
| ENSG00000103495 | 16 | 29807427 | 29833649 | 29 | 8 | 49314 | -0.12751 | 0.55073 | *MAZ* |
| ENSG00000197301 | 12 | 66235120 | 66285947 | 85 | 22 | 44306 | -0.12753 | 0.55074 | *RP11-366L20.2* |
| ENSG00000184992 | 12 | 125468246 | 125525777 | 286 | 36 | 43386 | -0.12764 | 0.55078 | *BRI3BP* |
| ENSG00000178826 | 7 | 142967050 | 142995141 | 19 | 11 | 52048 | -0.12778 | 0.55084 | *TMEM139* |
| ENSG00000113328 | 5 | 162854575 | 162883157 | 41 | 13 | 39866 | -0.12782 | 0.55085 | *CCNG1* |
| ENSG00000107537 | 10 | 13309796 | 13354412 | 209 | 37 | 45304 | -0.1278 | 0.55085 | *PHYH* |

| | | | | | | | | | |
|---|---|---|---|---|---|---|---|---|---|
| ENSG00000181472 | 6 | 151675252 | 151722683 | 124 | 22 | 34045 | -0.12795 | 0.55091 | *ZBTB2* |
| ENSG00000153187 | 1 | 245004468 | 245037844 | 54 | 14 | 47772 | -0.12798 | 0.55092 | *HNRNPU* |
| ENSG00000176253 | 14 | 20492003 | 20512919 | 132 | 14 | 45626 | -0.12802 | 0.55094 | *OR4K13* |
| ENSG00000255104 | 17 | 15593006 | 15658092 | 181 | 30 | 42228 | -0.12806 | 0.55095 | *ZNF286A* |
| ENSG00000182334 | 11 | 7836584 | 7857519 | 116 | 11 | 38952 | -0.12823 | 0.55102 | *OR5P3* |
| ENSG00000169432 | 2 | 167041695 | 167242503 | 701 | 84 | 47545 | -0.12831 | 0.55105 | *SCN9A* |
| ENSG00000162733 | 1 | 162591163 | 162767190 | 476 | 57 | 38840 | -0.12839 | 0.55108 | *DDR2* |
| ENSG00000167925 | 17 | 40330817 | 40356531 | 19 | 11 | 50526 | -0.12855 | 0.55114 | *GHDC* |
| ENSG00000074842 | 19 | 4631386 | 4680382 | 186 | 26 | 40267 | -0.1286 | 0.55116 | *C19orf10* |
| ENSG00000125337 | 6 | 168386921 | 168455769 | 387 | 59 | 40526 | -0.12887 | 0.55127 | *KIF25* |
| ENSG00000188312 | 9 | 95077766 | 95392815 | 866 | 45 | 41580 | -0.12912 | 0.55137 | *CENPP* |
| ENSG00000253797 | 13 | 52588827 | 52617736 | 50 | 10 | 41039 | -0.12915 | 0.55138 | *UTP14C* |
| ENSG00000070748 | 10 | 50807141 | 50911925 | 296 | 56 | 40860 | -0.12927 | 0.55143 | *CHAT* |
| ENSG00000092094 | 14 | 20904570 | 20933264 | 118 | 17 | 50792 | -0.1293 | 0.55144 | *OSGEP* |
| ENSG00000205018 | 16 | 88996197 | 89027932 | 100 | 21 | 44010 | -0.12941 | 0.55148 | *RP11-830F9.6* |
| ENSG00000070404 | 19 | 666392 | 693385 | 166 | 29 | 49594 | -0.12946 | 0.5515 | *FSTL3* |
| ENSG00000105835 | 7 | 105878731 | 105936772 | 163 | 21 | 45586 | -0.12978 | 0.55163 | *NAMPT* |
| ENSG00000169258 | 5 | 176012803 | 176047134 | 71 | 15 | 40472 | -0.13004 | 0.55173 | *GPRIN1* |
| ENSG00000203756 | 6 | 130142389 | 130192692 | 304 | 32 | 40034 | -0.13022 | 0.5518 | *TMEM244* |
| ENSG00000184144 | 1 | 205002325 | 205057627 | 198 | 29 | 29442 | -0.13035 | 0.55186 | *CNTN2* |
| ENSG00000214643 | 6 | 49903814 | 49927157 | 82 | 15 | 49412 | -0.13043 | 0.55189 | *DEFB133* |
| ENSG00000183625 | 3 | 46195096 | 46318197 | 359 | 27 | 29261 | -0.13057 | 0.55194 | *CCR3* |
| ENSG00000151475 | 4 | 128641533 | 128705448 | 95 | 14 | 41928 | -0.13073 | 0.55201 | *SLC25A31* |
| ENSG00000170807 | 7 | 123285861 | 123314344 | 59 | 10 | 54819 | -0.13082 | 0.55204 | *LMOD2* |
| ENSG00000143315 | 1 | 159987462 | 160011783 | 66 | 9 | 51005 | -0.13092 | 0.55208 | *PIGM* |
| ENSG00000166750 | 17 | 33560055 | 33610674 | 206 | 32 | 43045 | -0.13091 | 0.55208 | *SLFN5* |
| ENSG00000166603 | 18 | 58028564 | 58050001 | 65 | 15 | 44389 | -0.13096 | 0.5521 | *MC4R* |
| ENSG00000259900 | 16 | 69359179 | 69395633 | 80 | 17 | 53991 | -0.13105 | 0.55213 | *RP11-343C2.7* |
| ENSG00000138018 | 2 | 26521415 | 26628759 | 272 | 39 | 32227 | -0.13109 | 0.55215 | *EPT1* |
| ENSG00000107282 | 9 | 72032446 | 72297222 | 721 | 64 | 45415 | -0.13116 | 0.55218 | *APBA1* |
| ENSG00000148671 | 10 | 88717949 | 88753691 | 147 | 25 | 49161 | -0.13135 | 0.55225 | *ADIRF* |
| ENSG00000165588 | 14 | 57257425 | 57287197 | 43 | 17 | 49849 | -0.13139 | 0.55226 | *OTX2* |
| ENSG00000127580 | 16 | 724622 | 750444 | 52 | 11 | 34744 | -0.13147 | 0.5523 | *WDR24* |
| ENSG00000136830 | 9 | 130257618 | 130351268 | 298 | 42 | 47363 | -0.1316 | 0.55235 | *FAM129B* |
| ENSG00000100033 | 22 | 18890294 | 18934066 | 97 | 27 | 44075 | -0.13164 | 0.55236 | *PRODH* |
| ENSG00000268181 | 7 | 62848414 | 62868860 | 1 | 1 | 42664 | -0.13173 | 0.5524 | *AC073188.1* |

| | | | | | | | | | |
|---|---|---|---|---|---|---|---|---|---|
| ENSG00000171811 | 10 | 134611896 | 134766327 | 913 | 79 | 47679 | -0.13189 | 0.55246 | *TTC40* |
| ENSG00000122085 | 2 | 242001584 | 242051747 | 171 | 19 | 36320 | -0.13225 | 0.55261 | *MTERFD2* |
| ENSG00000106400 | 7 | 100850949 | 100877471 | 64 | 15 | 47096 | -0.13228 | 0.55262 | *ZNHIT1* |
| ENSG00000127364 | 7 | 141468242 | 141489235 | 73 | 3 | 51950 | -0.13243 | 0.55268 | *TAS2R4* |
| ENSG00000157429 | 16 | 71488453 | 71608992 | 237 | 34 | 38898 | -0.13262 | 0.55275 | *ZNF19* |
| ENSG00000152439 | 19 | 58001283 | 58039772 | 249 | 8 | 54315 | -0.13272 | 0.55279 | *ZNF773* |
| ENSG00000170615 | 7 | 102983177 | 103096624 | 315 | 46 | 41946 | -0.13327 | 0.55301 | *SLC26A5* |
| ENSG00000240021 | 1 | 178472212 | 178527734 | 169 | 17 | 43426 | -0.13408 | 0.55333 | *TEX35* |
| ENSG00000173327 | 11 | 65355226 | 65392853 | 74 | 16 | 47691 | -0.13425 | 0.5534 | *MAP3K11* |
| ENSG00000136634 | 1 | 206930947 | 206955839 | 78 | 19 | 39564 | -0.13429 | 0.55341 | *IL10* |
| ENSG00000197798 | 11 | 126071309 | 126142881 | 173 | 24 | 45570 | -0.13461 | 0.55354 | *FAM118B* |
| ENSG00000196187 | 1 | 226023237 | 226080069 | 230 | 29 | 46586 | -0.13463 | 0.55355 | *TMEM63A* |
| ENSG00000167397 | 16 | 31092163 | 31117301 | 30 | 6 | 18997 | -0.13464 | 0.55355 | *VKORC1* |
| ENSG00000158710 | 1 | 159877897 | 159905522 | 76 | 22 | 38828 | -0.13477 | 0.5536 | *TAGLN2* |
| ENSG00000013275 | 19 | 40466912 | 40497348 | 84 | 14 | 37373 | -0.13508 | 0.55373 | *PSMC4* |
| ENSG00000116209 | 1 | 54487347 | 54529177 | 91 | 13 | 30441 | -0.13519 | 0.55377 | *TMEM59* |
| ENSG00000272195 | 1 | 245122652 | 245144390 | 43 | 12 | 48194 | -0.1352 | 0.55377 | *RP11-156E8.1* |
| ENSG00000268800 | 5 | 162856447 | 162883821 | 40 | 13 | 39440 | -0.13536 | 0.55384 | *AC112205.1* |
| ENSG00000156030 | 14 | 74171825 | 74266988 | 282 | 51 | 39036 | -0.13569 | 0.55397 | *ELMSAN1* |
| ENSG00000130340 | 6 | 158234296 | 158376109 | 510 | 29 | 48233 | -0.13601 | 0.55409 | *SNX9* |
| ENSG00000128908 | 15 | 41261078 | 41418552 | 189 | 26 | 43292 | -0.13608 | 0.55412 | *INO80* |
| ENSG00000203667 | 1 | 244988624 | 245018359 | 96 | 13 | 49651 | -0.13631 | 0.55421 | *COX20* |
| ENSG00000106541 | 7 | 16821435 | 16883057 | 360 | 38 | 42616 | -0.13639 | 0.55424 | *AGR2* |
| ENSG00000184923 | 10 | 88975205 | 89006665 | 30 | 8 | 35671 | -0.13646 | 0.55427 | *NUTM2A* |
| ENSG00000061938 | 3 | 195580235 | 195648816 | 224 | 47 | 43150 | -0.13674 | 0.55438 | *TNK2* |
| ENSG00000165478 | 11 | 124779089 | 124816308 | 139 | 33 | 41443 | -0.13689 | 0.55444 | *HEPACAM* |
| ENSG00000187189 | 6 | 116561151 | 116585261 | 72 | 10 | 30766 | -0.13697 | 0.55447 | *TSPYL4* |
| ENSG00000221963 | 22 | 36034442 | 36074456 | 161 | 20 | 36388 | -0.13715 | 0.55454 | *APOL6* |
| ENSG00000040531 | 17 | 3529762 | 3574836 | 195 | 26 | 36932 | -0.13736 | 0.55463 | *CTNS* |
| ENSG00000153093 | 2 | 111480150 | 111885799 | 1239 | 117 | 52195 | -0.13758 | 0.55471 | *ACOXL* |
| ENSG00000205726 | 21 | 35004706 | 35282165 | 582 | 102 | 46035 | -0.13784 | 0.55482 | *ITSN1* |
| ENSG00000118369 | 11 | 77889858 | 77935757 | 120 | 18 | 39179 | -0.13802 | 0.55489 | *USP35* |
| ENSG00000145088 | 3 | 121544030 | 121615373 | 227 | 19 | 51458 | -0.13811 | 0.55492 | *EAF2* |
| ENSG00000165970 | 11 | 20610946 | 20690831 | 340 | 53 | 50885 | -0.13836 | 0.55502 | *SLC6A5* |
| ENSG00000154723 | 21 | 27078815 | 27117984 | 75 | 15 | 39494 | -0.13873 | 0.55517 | *ATP5J* |
| ENSG00000196220 | 3 | 9012275 | 9414737 | 1320 | 207 | 47608 | -0.13877 | 0.55518 | *SRGAP3* |

| | | | | | | | | | |
|---|---|---|---|---|---|---|---|---|---|
| ENSG00000255398 | 12 | 123189303 | 123211439 | 85 | 7 | 44956 | -0.13875 | 0.55518 | *HCAR3* |
| ENSG00000197756 | 2 | 217352912 | 217453903 | 314 | 46 | 38910 | -0.13878 | 0.55519 | *RPL37A* |
| ENSG00000268076 | 12 | 25371515 | 25400840 | 114 | 18 | 50612 | -0.13878 | 0.55519 | *AC087239.1* |
| ENSG00000108417 | 17 | 39567005 | 39590775 | 88 | 13 | 53897 | -0.13882 | 0.5552 | *KRT37* |
| ENSG00000168397 | 2 | 242566628 | 242623272 | 255 | 27 | 43959 | -0.13884 | 0.55521 | *ATG4B* |
| ENSG00000137841 | 15 | 40560377 | 40610136 | 101 | 29 | 40940 | -0.13894 | 0.55525 | *PLCB2* |
| ENSG00000088808 | 14 | 104190089 | 104323927 | 355 | 21 | 48859 | -0.13935 | 0.55541 | *PPP1R13B* |
| ENSG00000184903 | 7 | 110293110 | 111212573 | 2075 | 141 | 37179 | -0.13981 | 0.55559 | *IMMP2L* |
| ENSG00000154240 | 17 | 63621656 | 64198202 | 2364 | 82 | 33657 | -0.13982 | 0.5556 | *CEP112* |
| ENSG00000214237 | 3 | 150578832 | 150622089 | 124 | 31 | 47858 | -0.14008 | 0.5557 | *FAM188B2* |
| ENSG00000196166 | 8 | 38358352 | 38396180 | 120 | 20 | 33290 | -0.1402 | 0.55575 | *C8orf86* |
| ENSG00000204392 | 6 | 31755173 | 31784761 | 84 | 27 | 36622 | -0.14029 | 0.55578 | *LSM2* |
| ENSG00000259458 | 15 | 62919371 | 62947490 | 76 | 21 | 44341 | -0.14058 | 0.5559 | *RP11-625H11.1* |
| ENSG00000107223 | 9 | 139746571 | 139770738 | 38 | 9 | 54099 | -0.14064 | 0.55592 | *EDF1* |
| ENSG00000143184 | 1 | 168535711 | 168561315 | 99 | 15 | 37182 | -0.14088 | 0.55602 | *XCL1* |
| ENSG00000120053 | 10 | 101146627 | 101200381 | 106 | 22 | 43017 | -0.14093 | 0.55604 | *GOT1* |
| ENSG00000073756 | 1 | 186630923 | 186659559 | 111 | 15 | 44199 | -0.14099 | 0.55606 | *PTGS2* |
| ENSG00000013364 | 16 | 29821715 | 29869355 | 63 | 6 | 49743 | -0.14117 | 0.55613 | *MVP* |
| ENSG00000041880 | 3 | 51966361 | 51992883 | 42 | 16 | 38082 | -0.1413 | 0.55618 | *PARP3* |
| ENSG00000091656 | 8 | 77583454 | 77789521 | 440 | 56 | 36888 | -0.14128 | 0.55618 | *ZFHX4* |
| ENSG00000203815 | 1 | 149665978 | 149686609 | 1 | 1 | 55578 | -0.14134 | 0.5562 | *FAM231D* |
| ENSG00000124532 | 6 | 24393153 | 24435810 | 287 | 24 | 45890 | -0.14138 | 0.55622 | *MRS2* |
| ENSG00000133612 | 7 | 150772918 | 150851523 | 213 | 37 | 43772 | -0.14147 | 0.55625 | *AGAP3* |
| ENSG00000183891 | 2 | 20086404 | 20111747 | 74 | 17 | 43946 | -0.14155 | 0.55628 | *TTC32* |
| ENSG00000128791 | 18 | 9324765 | 9412418 | 326 | 28 | 47677 | -0.14166 | 0.55633 | *TWSG1* |
| ENSG00000112984 | 5 | 137504408 | 137533404 | 49 | 14 | 52207 | -0.14169 | 0.55634 | *KIF20A* |
| ENSG00000140948 | 16 | 87429852 | 87535651 | 432 | 29 | 42804 | -0.14177 | 0.55637 | *ZCCHC14* |
| ENSG00000203661 | 1 | 248641885 | 248662898 | 4 | 2 | 49478 | -0.14187 | 0.55641 | *OR2T5* |
| ENSG00000130368 | 6 | 160317974 | 160339108 | 58 | 15 | 43232 | -0.14204 | 0.55648 | *MAS1* |
| ENSG00000166562 | 18 | 56796709 | 56836068 | 150 | 21 | 42088 | -0.14212 | 0.55651 | *SEC11C* |
| ENSG00000235169 | 1 | 3679352 | 3702546 | 128 | 18 | 46432 | -0.14214 | 0.55652 | *SMIM1* |
| ENSG00000170485 | 2 | 101426614 | 101623291 | 639 | 101 | 38624 | -0.14214 | 0.55652 | *NPAS2* |
| ENSG00000137449 | 4 | 14994298 | 15081777 | 201 | 25 | 43157 | -0.1423 | 0.55658 | *CPEB2* |
| ENSG00000189056 | 7 | 103102231 | 103639963 | 2377 | 191 | 44819 | -0.14282 | 0.55678 | *RELN* |
| ENSG00000087301 | 14 | 52887308 | 53029240 | 407 | 37 | 47775 | -0.14285 | 0.5568 | *TXNDC16* |
| ENSG00000061492 | 5 | 137409581 | 137438054 | 66 | 11 | 50839 | -0.14294 | 0.55683 | *WNT8A* |

| | | | | | | | | | |
|---|---|---|---|---|---|---|---|---|---|
| ENSG00000187187 | 19 | 40480041 | 40533514 | 90 | 10 | 39147 | -0.14296 | 0.55684 | *ZNF546* |
| ENSG00000173302 | 2 | 131476643 | 131497909 | 46 | 7 | 31127 | -0.14299 | 0.55685 | *GPR148* |
| ENSG00000179044 | 16 | 67208269 | 67234107 | 41 | 9 | 33333 | -0.14333 | 0.55698 | *EXOC3L1* |
| ENSG00000204889 | 17 | 39123968 | 39153387 | 237 | 18 | 28359 | -0.14333 | 0.55699 | *KRT40* |
| ENSG00000212128 | 12 | 11050525 | 11072161 | 78 | 6 | 28775 | -0.14336 | 0.557 | *TAS2R13* |
| ENSG00000163352 | 1 | 154956062 | 154976791 | 31 | 8 | 44464 | -0.14353 | 0.55706 | *LENEP* |
| ENSG00000089234 | 12 | 112069950 | 112133790 | 42 | 18 | 54334 | -0.14401 | 0.55726 | *BRAP* |
| ENSG00000138593 | 15 | 49270673 | 49348760 | 166 | 22 | 41767 | -0.14417 | 0.55732 | *SECISBP2L* |
| ENSG00000217442 | 22 | 50979541 | 51011334 | 208 | 23 | 30394 | -0.14431 | 0.55737 | *SYCE3* |
| ENSG00000100934 | 14 | 39491123 | 39588850 | 290 | 33 | 47496 | -0.14449 | 0.55744 | *SEC23A* |
| ENSG00000118965 | 2 | 20100021 | 20199892 | 258 | 27 | 51076 | -0.1446 | 0.55749 | *WDR35* |
| ENSG00000116574 | 1 | 228860824 | 228892416 | 113 | 15 | 41483 | -0.14476 | 0.55755 | *RHOU* |
| ENSG00000100528 | 14 | 54883654 | 54918149 | 92 | 9 | 31844 | -0.14486 | 0.55759 | *CNIH1* |
| ENSG00000144401 | 2 | 208435355 | 208500652 | 143 | 14 | 49898 | -0.14509 | 0.55768 | *METTL21A* |
| ENSG00000106853 | 9 | 114302002 | 114372135 | 278 | 49 | 46587 | -0.14512 | 0.55769 | *PTGR1* |
| ENSG00000108515 | 17 | 4841387 | 4870426 | 63 | 20 | 42319 | -0.14523 | 0.55774 | *ENO3* |
| ENSG00000168959 | 11 | 88227744 | 88809113 | 1909 | 107 | 35127 | -0.14542 | 0.55781 | *GRM5* |
| ENSG00000138039 | 2 | 48849428 | 48992880 | 603 | 79 | 47572 | -0.14562 | 0.55789 | *LHCGR* |
| ENSG00000110375 | 11 | 118785873 | 118839269 | 165 | 33 | 48147 | -0.1457 | 0.55792 | *UPK2* |
| ENSG00000182400 | 14 | 39607015 | 39649736 | 140 | 22 | 48882 | -0.1459 | 0.558 | *TRAPPC6B* |
| ENSG00000143933 | 2 | 47377221 | 47413740 | 127 | 20 | 36949 | -0.14593 | 0.55801 | *CALM2* |
| ENSG00000186318 | 11 | 117146402 | 117196975 | 122 | 31 | 48577 | -0.14602 | 0.55805 | *BACE1* |
| ENSG00000250588 | 3 | 158670024 | 159625155 | 2296 | 196 | 44005 | -0.14624 | 0.55814 | *IQCJ-SCHIP1* |
| ENSG00000143294 | 1 | 156710402 | 156780607 | 201 | 27 | 38948 | -0.14655 | 0.55826 | *PRCC* |
| ENSG00000119661 | 14 | 74101578 | 74180435 | 271 | 26 | 30588 | -0.14661 | 0.55828 | *DNAL1* |
| ENSG00000101189 | 20 | 61417805 | 61441945 | 102 | 22 | 47656 | -0.14681 | 0.55836 | *MRGBP* |
| ENSG00000164587 | 5 | 149812753 | 149839319 | 93 | 8 | 52068 | -0.14707 | 0.55846 | *RPS14* |
| ENSG00000253976 | 8 | 56037763 | 56064918 | 82 | 15 | 37349 | -0.14749 | 0.55863 | *RP11-386G21.1* |
| ENSG00000180198 | 1 | 28822455 | 28875812 | 151 | 14 | 31781 | -0.14761 | 0.55867 | *RCC1* |
| ENSG00000166295 | 10 | 73965787 | 74005618 | 82 | 13 | 32365 | -0.14765 | 0.55869 | *ANAPC16* |
| ENSG00000136542 | 2 | 158104110 | 158180723 | 133 | 24 | 37360 | -0.1477 | 0.55871 | *GALNT5* |
| ENSG00000266949 | 1 | 248892716 | 248913150 | 38 | 8 | 34528 | -0.14774 | 0.55873 | *LYPD8* |
| ENSG00000197712 | 4 | 38859298 | 38957360 | 383 | 42 | 41918 | -0.14815 | 0.55889 | *FAM114A1* |
| ENSG00000229415 | 14 | 36932493 | 36993034 | 116 | 30 | 40133 | -0.1483 | 0.55895 | *SFTA3* |
| ENSG00000004059 | 7 | 127218399 | 127241759 | 65 | 14 | 39268 | -0.14833 | 0.55896 | *ARF5* |
| ENSG00000249141 | 6 | 167261582 | 167379612 | 418 | 64 | 38367 | -0.14842 | 0.55899 | *RP11-514O12.4* |

| | | | | | | | | | |
|---|---|---|---|---|---|---|---|---|---|
| ENSG00000135702 | 16 | 75552433 | 75579145 | 84 | 11 | 47889 | -0.14856 | 0.55905 | *CHST5* |
| ENSG00000132016 | 19 | 13983161 | 14027265 | 96 | 6 | 27965 | -0.14874 | 0.55912 | *C19orf57* |
| ENSG00000148357 | 9 | 133036882 | 133319510 | 555 | 115 | 43989 | -0.14912 | 0.55927 | *HMCN2* |
| ENSG00000196419 | 22 | 42007123 | 42070044 | 126 | 19 | 34330 | -0.14934 | 0.55936 | *XRCC6* |
| ENSG00000185940 | 11 | 1641033 | 1662160 | 177 | 27 | 53548 | -0.14936 | 0.55937 | *KRTAP5-5* |
| ENSG00000164411 | 6 | 87982696 | 88048996 | 268 | 19 | 43373 | -0.14939 | 0.55938 | *GJB7* |
| ENSG00000180044 | 3 | 159933423 | 159955999 | 55 | 17 | 33522 | -0.14945 | 0.5594 | *C3orf80* |
| ENSG00000160224 | 21 | 45695721 | 45728531 | 174 | 31 | 44600 | -0.14944 | 0.5594 | *AIRE* |
| ENSG00000157856 | 2 | 26614784 | 26689579 | 131 | 33 | 37014 | -0.14951 | 0.55942 | *DRC1* |
| ENSG00000178904 | 19 | 32886449 | 32986801 | 257 | 34 | 42396 | -0.14967 | 0.55949 | *DPY19L3* |
| ENSG00000168374 | 3 | 57547090 | 57593947 | 142 | 18 | 45095 | -0.14976 | 0.55952 | *ARF4* |
| ENSG00000011028 | 17 | 60694762 | 60780958 | 148 | 29 | 41572 | -0.14987 | 0.55957 | *MRC2* |
| ENSG00000088682 | 16 | 57471337 | 57505187 | 114 | 24 | 52595 | -0.14993 | 0.55959 | *COQ9* |
| ENSG00000138386 | 2 | 191501472 | 191567492 | 183 | 23 | 46636 | -0.15005 | 0.55964 | *NAB1* |
| ENSG00000165731 | 10 | 43562475 | 43635799 | 302 | 33 | 50497 | -0.15011 | 0.55966 | *RET* |
| ENSG00000229474 | 15 | 44947930 | 45013514 | 55 | 24 | 43027 | -0.15017 | 0.55969 | *PATL2* |
| ENSG00000161243 | 19 | 39471354 | 39533425 | 374 | 25 | 46586 | -0.15022 | 0.55971 | *FBXO27* |
| ENSG00000104368 | 8 | 42022236 | 42075242 | 95 | 22 | 48163 | -0.15036 | 0.55976 | *PLAT* |
| ENSG00000159708 | 16 | 67350701 | 67429106 | 106 | 18 | 44731 | -0.15055 | 0.55983 | *LRRC36* |
| ENSG00000091592 | 17 | 5392747 | 5532744 | 674 | 48 | 46322 | -0.15063 | 0.55987 | *NLRP1* |
| ENSG00000111252 | 12 | 111833752 | 111899427 | 55 | 13 | 48226 | -0.15066 | 0.55988 | *SH2B3* |
| ENSG00000163492 | 2 | 179684484 | 179924813 | 777 | 96 | 42469 | -0.15103 | 0.56002 | *CCDC141* |
| ENSG00000260415 | 12 | 52193789 | 52216636 | 55 | 12 | 35302 | -0.15105 | 0.56003 | *AC068987.1* |
| ENSG00000273303 | 3 | 44597051 | 44645665 | 115 | 16 | 34160 | -0.15111 | 0.56006 | *RP11-944L7.5* |
| ENSG00000130038 | 12 | 3705799 | 3883985 | 738 | 100 | 51147 | -0.15166 | 0.56027 | *EFCAB4B* |
| ENSG00000105538 | 19 | 49213844 | 49253978 | 122 | 13 | 25633 | -0.15228 | 0.56052 | *RASIP1* |
| ENSG00000129493 | 14 | 31750994 | 31899788 | 234 | 28 | 37231 | -0.15235 | 0.56054 | *HEATR5A* |
| ENSG00000114115 | 3 | 139226276 | 139268671 | 143 | 21 | 33760 | -0.15243 | 0.56058 | *RBP1* |
| ENSG00000135083 | 5 | 159668659 | 159782403 | 340 | 54 | 39853 | -0.1525 | 0.5606 | *CCNJL* |
| ENSG00000153165 | 2 | 107011446 | 107094832 | 133 | 36 | 45751 | -0.1526 | 0.56064 | *RGPD3* |
| ENSG00000134058 | 5 | 68520668 | 68583250 | 204 | 19 | 50759 | -0.15261 | 0.56065 | *CDK7* |
| ENSG00000224051 | 1 | 1250136 | 1274277 | 67 | 16 | 38254 | -0.15264 | 0.56066 | *GLTPD1* |
| ENSG00000177151 | 1 | 248791588 | 248812559 | 9 | 2 | 43523 | -0.15264 | 0.56066 | *OR2T35* |
| ENSG00000138755 | 4 | 76912428 | 76938641 | 58 | 12 | 49732 | -0.1528 | 0.56072 | *CXCL9* |
| ENSG00000162385 | 1 | 53682564 | 53714282 | 63 | 14 | 41186 | -0.15281 | 0.56073 | *MAGOH* |
| ENSG00000142538 | 19 | 49915671 | 49936698 | 34 | 15 | 49972 | -0.15313 | 0.56085 | *PTH2* |

| | | | | | | | | | |
|---|---|---|---|---|---|---|---|---|---|
| ENSG00000188984 | 1 | 12766118 | 12798726 | 117 | 16 | 47286 | -0.15334 | 0.56094 | *AADACL3* |
| ENSG00000130477 | 19 | 17702137 | 17809401 | 427 | 90 | 41336 | -0.15357 | 0.56103 | *UNC13A* |
| ENSG00000140553 | 15 | 91463410 | 91507323 | 142 | 22 | 39191 | -0.15366 | 0.56106 | *UNC45A* |
| ENSG00000124523 | 6 | 13564816 | 13624790 | 109 | 17 | 50838 | -0.15368 | 0.56107 | *SIRT5* |
| ENSG00000064102 | 12 | 27048114 | 27101259 | 150 | 28 | 53716 | -0.15391 | 0.56116 | *ASUN* |
| ENSG00000127362 | 7 | 141453897 | 141474997 | 74 | 3 | 54512 | -0.15415 | 0.56125 | *TAS2R3* |
| ENSG00000186501 | 1 | 27638651 | 27672891 | 74 | 17 | 49773 | -0.15421 | 0.56128 | *TMEM222* |
| ENSG00000151388 | 5 | 33513640 | 33902297 | 1192 | 133 | 37510 | -0.15461 | 0.56143 | *ADAMTS12* |
| ENSG00000070814 | 5 | 149727202 | 149789871 | 132 | 23 | 53965 | -0.1546 | 0.56143 | *TCOF1* |
| ENSG00000120805 | 12 | 101776898 | 101811598 | 77 | 19 | 49000 | -0.15481 | 0.56151 | *ARL1* |
| ENSG00000086544 | 19 | 41213008 | 41256765 | 135 | 19 | 52445 | -0.1548 | 0.56151 | *ITPKC* |
| ENSG00000100596 | 14 | 77962340 | 78093116 | 590 | 69 | 47511 | -0.15524 | 0.56168 | *SPTLC2* |
| ENSG00000140067 | 14 | 94375240 | 94405954 | 97 | 25 | 42629 | -0.15528 | 0.5617 | *FAM181A* |
| ENSG00000167280 | 17 | 77061021 | 77094681 | 130 | 24 | 40335 | -0.15552 | 0.56179 | *ENGASE* |
| ENSG00000177602 | 17 | 3617211 | 3640067 | 95 | 11 | 33447 | -0.1556 | 0.56183 | *GSG2* |
| ENSG00000173769 | 3 | 44273378 | 44383590 | 252 | 16 | 44278 | -0.15588 | 0.56194 | *TOPAZ1* |
| ENSG00000224940 | 7 | 127980379 | 128011739 | 63 | 20 | 45940 | -0.15634 | 0.56212 | *PRRT4* |
| ENSG00000177989 | 22 | 50958139 | 50981009 | 95 | 20 | 48105 | -0.15686 | 0.56232 | *ODF3B* |
| ENSG00000173917 | 17 | 46608256 | 46633441 | 66 | 14 | 45836 | -0.15695 | 0.56236 | *HOXB2* |
| ENSG00000130764 | 1 | 3686784 | 3723068 | 202 | 27 | 45720 | -0.15705 | 0.5624 | *LRRC47* |
| ENSG00000100442 | 14 | 45574803 | 45614522 | 57 | 9 | 31040 | -0.15716 | 0.56244 | *FKBP3* |
| ENSG00000125903 | 20 | 197899 | 220527 | 88 | 13 | 42055 | -0.15727 | 0.56248 | *DEFB129* |
| ENSG00000137815 | 15 | 41690606 | 41785761 | 229 | 25 | 41700 | -0.15739 | 0.56253 | *RTF1* |
| ENSG00000139514 | 13 | 30073547 | 30179825 | 317 | 44 | 42118 | -0.15743 | 0.56255 | *SLC7A1* |
| ENSG00000124578 | 6 | 26236886 | 26257259 | 58 | 10 | 46914 | -0.1578 | 0.56269 | *HIST1H4G* |
| ENSG00000243147 | 2 | 27984584 | 28220954 | 402 | 29 | 28592 | -0.15789 | 0.56273 | *MRPL33* |
| ENSG00000261130 | 16 | 15479611 | 15619205 | 316 | 48 | 43969 | -0.15791 | 0.56273 | *RP11-1021N1.1* |
| ENSG00000168634 | 20 | 44320655 | 44347456 | 101 | 16 | 39265 | -0.15799 | 0.56277 | *WFDC13* |
| ENSG00000189269 | 22 | 23940639 | 23984508 | 150 | 30 | 46506 | -0.15811 | 0.56281 | *C22orf43* |
| ENSG00000107863 | 10 | 24862538 | 25022597 | 266 | 32 | 52545 | -0.15869 | 0.56304 | *ARHGAP21* |
| ENSG00000168778 | 12 | 124145660 | 124202948 | 277 | 14 | 44408 | -0.15868 | 0.56304 | *TCTN2* |
| ENSG00000181291 | 17 | 32897768 | 32976337 | 218 | 62 | 47773 | -0.15905 | 0.56318 | *TMEM132E* |
| ENSG00000105072 | 19 | 16597122 | 16642163 | 143 | 21 | 43922 | -0.15941 | 0.56333 | *C19orf44* |
| ENSG00000204842 | 12 | 111880018 | 112047480 | 129 | 20 | 46987 | -0.15948 | 0.56336 | *ATXN2* |
| ENSG00000205496 | 11 | 4966002 | 4986943 | 87 | 13 | 51419 | -0.15971 | 0.56344 | *OR51A2* |
| ENSG00000171862 | 10 | 89612870 | 89741687 | 196 | 41 | 54323 | -0.1598 | 0.56348 | *PTEN* |

| | | | | | | | | | |
|---|---|---|---|---|---|---|---|---|---|
| ENSG00000128714 | 2 | 176947619 | 176970666 | 62 | 13 | 38969 | -0.16004 | 0.56358 | *HOXD13* |
| ENSG00000165533 | 14 | 89280497 | 89354335 | 129 | 19 | 47161 | -0.16008 | 0.56359 | *TTC8* |
| ENSG00000085872 | 19 | 16618700 | 16663341 | 105 | 22 | 45456 | -0.1602 | 0.56364 | *CHERP* |
| ENSG00000077514 | 11 | 74194896 | 74390162 | 721 | 58 | 45754 | -0.16024 | 0.56365 | *POLD3* |
| ENSG00000167107 | 17 | 48493519 | 48562206 | 172 | 25 | 42511 | -0.16027 | 0.56367 | *ACSF2* |
| ENSG00000144120 | 2 | 120426743 | 120454083 | 127 | 12 | 44801 | -0.16073 | 0.56385 | *TMEM177* |
| ENSG00000227729 | 14 | 104396763 | 104418645 | 118 | 9 | 52568 | -0.1609 | 0.56391 | *RD3L* |
| ENSG00000072954 | 19 | 16761938 | 16810840 | 138 | 22 | 35548 | -0.16098 | 0.56394 | *TMEM38A* |
| ENSG00000161594 | 17 | 39981937 | 40014636 | 114 | 8 | 52343 | -0.161 | 0.56395 | *KLHL10* |
| ENSG00000130734 | 19 | 10644571 | 10674094 | 110 | 17 | 43228 | -0.1613 | 0.56407 | *ATG4D* |
| ENSG00000137648 | 11 | 117937753 | 118002605 | 234 | 36 | 46657 | -0.16139 | 0.56411 | *TMPRSS4* |
| ENSG00000128045 | 4 | 53718457 | 53743000 | 101 | 15 | 53513 | -0.16147 | 0.56414 | *RASL11B* |
| ENSG00000205339 | 11 | 9396169 | 9479673 | 194 | 18 | 40794 | -0.1617 | 0.56423 | *IPO7* |
| ENSG00000155158 | 9 | 15153620 | 15317358 | 616 | 87 | 44099 | -0.16191 | 0.56431 | *TTC39B* |
| ENSG00000092148 | 14 | 31559318 | 31687010 | 166 | 23 | 45917 | -0.16207 | 0.56437 | *HECTD1* |
| ENSG00000118513 | 6 | 135492453 | 135550311 | 119 | 23 | 46433 | -0.16219 | 0.56442 | *MYB* |
| ENSG00000142583 | 1 | 9085166 | 9158537 | 309 | 39 | 48762 | -0.16227 | 0.56445 | *SLC2A5* |
| ENSG00000189367 | 6 | 127751488 | 127790536 | 79 | 8 | 29657 | -0.16226 | 0.56445 | *KIAA0408* |
| ENSG00000131652 | 16 | 3064028 | 3087756 | 66 | 12 | 54262 | -0.16229 | 0.56446 | *THOC6* |
| ENSG00000139445 | 12 | 109705784 | 109757025 | 101 | 18 | 52699 | -0.16235 | 0.56449 | *FOXN4* |
| ENSG00000205138 | 19 | 36476090 | 36497220 | 41 | 12 | 48641 | -0.16245 | 0.56452 | *SDHAF1* |
| ENSG00000113721 | 5 | 149483400 | 149545435 | 210 | 44 | 45375 | -0.16259 | 0.56458 | *PDGFRB* |
| ENSG00000144567 | 2 | 220030947 | 220060201 | 59 | 8 | 30643 | -0.16295 | 0.56472 | *FAM134A* |
| ENSG00000143278 | 1 | 196998321 | 197046397 | 103 | 11 | 24543 | -0.16311 | 0.56478 | *F13B* |
| ENSG00000269559 | 4 | 75013327 | 75034085 | 9 | 6 | 31203 | -0.1631 | 0.56478 | *AC093677.1* |
| ENSG00000131370 | 3 | 15286360 | 15392875 | 298 | 58 | 43750 | -0.16322 | 0.56483 | *SH3BP5* |
| ENSG00000112115 | 6 | 52041185 | 52065436 | 129 | 13 | 31980 | -0.16347 | 0.56493 | *IL17A* |
| ENSG00000101342 | 20 | 35494524 | 35532638 | 103 | 23 | 41160 | -0.16365 | 0.565 | *TLDC2* |
| ENSG00000269042 | 11 | 108120255 | 108146480 | 35 | 8 | 45187 | -0.16376 | 0.56504 | *AP001925.1* |
| ENSG00000268040 | 21 | 47337679 | 47362477 | 85 | 13 | 33127 | -0.1641 | 0.56517 | *PRED62* |
| ENSG00000161031 | 19 | 15569456 | 15619767 | 154 | 28 | 49660 | -0.16415 | 0.56519 | *PGLYRP2* |
| ENSG00000227471 | 7 | 134223888 | 134274627 | 224 | 17 | 45064 | -0.16427 | 0.56524 | *AKR1B15* |
| ENSG00000163914 | 3 | 129237483 | 129264012 | 52 | 12 | 38774 | -0.16471 | 0.56541 | *RHO* |
| ENSG00000124357 | 2 | 71281474 | 71316935 | 148 | 20 | 49332 | -0.1648 | 0.56545 | *NAGK* |
| ENSG00000187990 | 6 | 26206428 | 26226872 | 99 | 9 | 54114 | -0.16494 | 0.5655 | *HIST1H2BG* |
| ENSG00000140538 | 15 | 88408230 | 88809999 | 936 | 125 | 45961 | -0.16504 | 0.56554 | *NTRK3* |

| | | | | | | | | | |
|---|---|---|---|---|---|---|---|---|---|
| ENSG00000103489 | 16 | 17185626 | 17574738 | 1284 | 154 | 43340 | -0.16516 | 0.56559 | *XYLT1* |
| ENSG00000131462 | 17 | 40751694 | 40777252 | 52 | 7 | 53673 | -0.16535 | 0.56567 | *TUBG1* |
| ENSG00000205309 | 17 | 17196649 | 17260977 | 232 | 38 | 44415 | -0.16555 | 0.56574 | *NT5M* |
| ENSG00000120306 | 5 | 139544227 | 139671637 | 289 | 18 | 53037 | -0.16558 | 0.56576 | *CYSTM1* |
| ENSG00000139044 | 12 | 559530 | 682675 | 546 | 122 | 40657 | -0.16565 | 0.56578 | *B4GALNT3* |
| ENSG00000139874 | 14 | 38667204 | 38692272 | 58 | 16 | 47748 | -0.16566 | 0.56579 | *SSTR1* |
| ENSG00000236279 | 7 | 139198602 | 139239730 | 77 | 13 | 36916 | -0.1659 | 0.56588 | *CLEC2L* |
| ENSG00000166961 | 11 | 60514426 | 60554205 | 91 | 41 | 45261 | -0.16594 | 0.5659 | *MS4A15* |
| ENSG00000154229 | 17 | 64288754 | 64816861 | 1994 | 213 | 49434 | -0.16597 | 0.56591 | *PRKCA* |
| ENSG00000137198 | 6 | 16228811 | 16305780 | 208 | 36 | 35729 | -0.16599 | 0.56592 | *GMPR* |
| ENSG00000122591 | 7 | 22970878 | 23063749 | 309 | 25 | 54308 | -0.16632 | 0.56605 | *FAM126A* |
| ENSG00000181804 | 3 | 142974064 | 143577373 | 2177 | 244 | 45781 | -0.16727 | 0.56642 | *SLC9A9* |
| ENSG00000170837 | 3 | 71793201 | 71815647 | 65 | 17 | 42347 | -0.1674 | 0.56647 | *GPR27* |
| ENSG00000075856 | 12 | 108906357 | 108965176 | 207 | 18 | 39356 | -0.16763 | 0.56656 | *SART3* |
| ENSG00000125735 | 19 | 6653148 | 6680599 | 108 | 32 | 52238 | -0.16763 | 0.56656 | *TNFSF14* |
| ENSG00000182985 | 11 | 115029938 | 115385675 | 714 | 103 | 37730 | -0.16774 | 0.5666 | *CADM1* |
| ENSG00000125652 | 19 | 6362444 | 6385042 | 98 | 15 | 38317 | -0.16774 | 0.56661 | *ALKBH7* |
| ENSG00000163902 | 3 | 128328817 | 128409918 | 276 | 23 | 54738 | -0.16791 | 0.56667 | *RPN1* |
| ENSG00000178982 | 19 | 39099735 | 39137595 | 58 | 20 | 43047 | -0.16795 | 0.56669 | *EIF3K* |
| ENSG00000065978 | 1 | 43138098 | 43178020 | 102 | 18 | 53692 | -0.1681 | 0.56675 | *YBX1* |
| ENSG00000107816 | 10 | 102746375 | 102777593 | 76 | 13 | 35511 | -0.16836 | 0.56685 | *LZTS2* |
| ENSG00000137090 | 9 | 831690 | 979090 | 869 | 109 | 37300 | -0.1684 | 0.56687 | *DMRT1* |
| ENSG00000171174 | 2 | 27994231 | 28123965 | 230 | 19 | 27409 | -0.16846 | 0.56689 | *RBKS* |
| ENSG00000148218 | 9 | 116138597 | 116173613 | 128 | 32 | 47490 | -0.16852 | 0.56691 | *ALAD* |
| ENSG00000178055 | 3 | 46861032 | 46885585 | 66 | 11 | 32116 | -0.16855 | 0.56692 | *PRSS42* |
| ENSG00000128340 | 22 | 37611301 | 37650488 | 207 | 25 | 49831 | -0.16853 | 0.56692 | *RAC2* |
| ENSG00000109339 | 4 | 86926276 | 87525284 | 1904 | 106 | 40044 | -0.1689 | 0.56706 | *MAPK10* |
| ENSG00000244509 | 22 | 39400088 | 39426357 | 82 | 21 | 45634 | -0.16888 | 0.56706 | *APOBEC3C* |
| ENSG00000204188 | 6 | 33541515 | 33566803 | 111 | 19 | 40021 | -0.16901 | 0.56711 | *GGNBP1* |
| ENSG00000213171 | 1 | 151762740 | 151788630 | 49 | 11 | 45905 | -0.16958 | 0.56733 | *LINGO4* |
| ENSG00000119865 | 2 | 68501303 | 68557183 | 136 | 33 | 35111 | -0.16976 | 0.5674 | *CNRIP1* |
| ENSG00000104081 | 15 | 40370091 | 40411093 | 78 | 24 | 44863 | -0.16976 | 0.5674 | *BMF* |
| ENSG00000266714 | 17 | 73574139 | 73632929 | 289 | 45 | 44203 | -0.16978 | 0.56741 | *MYO15B* |
| ENSG00000171603 | 1 | 9779084 | 9894584 | 178 | 32 | 38072 | -0.1699 | 0.56746 | *CLSTN1* |
| ENSG00000100425 | 22 | 50156931 | 50231160 | 248 | 30 | 38661 | -0.16995 | 0.56747 | *BRD1* |
| ENSG00000172775 | 16 | 57176378 | 57230028 | 115 | 21 | 45849 | -0.17029 | 0.56761 | *FAM192A* |

| | | | | | | | | | |
|---|---|---|---|---|---|---|---|---|---|
| ENSG00000180287 | 1 | 242236288 | 242697998 | 2056 | 197 | 45759 | -0.17039 | 0.56765 | *PLD5* |
| ENSG00000074855 | 19 | 17424032 | 17455638 | 126 | 17 | 45855 | -0.17065 | 0.56775 | *ANO8* |
| ENSG00000197140 | 8 | 38954509 | 39152430 | 490 | 34 | 50176 | -0.17076 | 0.56779 | *ADAM32* |
| ENSG00000104133 | 15 | 44844894 | 44965876 | 99 | 38 | 44739 | -0.17103 | 0.5679 | *SPG11* |
| ENSG00000089775 | 14 | 64905824 | 64981931 | 223 | 21 | 54832 | -0.17105 | 0.56791 | *ZBTB25* |
| ENSG00000198685 | 3 | 128280843 | 128304929 | 81 | 20 | 45869 | -0.17119 | 0.56796 | *C3orf27* |
| ENSG00000170054 | 14 | 94919054 | 94956026 | 185 | 32 | 46707 | -0.17122 | 0.56797 | *SERPINA9* |
| ENSG00000109079 | 17 | 26652628 | 26684035 | 39 | 10 | 47276 | -0.17169 | 0.56816 | *TNFAIP1* |
| ENSG00000198546 | 10 | 135111979 | 135176033 | 212 | 27 | 33146 | -0.17198 | 0.56827 | *ZNF511* |
| ENSG00000225899 | 10 | 135427399 | 135450299 | 69 | 12 | 27346 | -0.17199 | 0.56828 | *FRG2B* |
| ENSG00000177706 | 7 | 182969 | 310711 | 397 | 75 | 41926 | -0.17218 | 0.56835 | *FAM20C* |
| ENSG00000124508 | 6 | 26373324 | 26405102 | 167 | 20 | 40343 | -0.17227 | 0.56839 | *BTN2A2* |
| ENSG00000070061 | 9 | 111619797 | 111706396 | 395 | 46 | 44612 | -0.1724 | 0.56844 | *IKBKAP* |
| ENSG00000176422 | 12 | 56852301 | 56874763 | 40 | 11 | 44994 | -0.17256 | 0.5685 | *SPRYD4* |
| ENSG00000180269 | 16 | 20032807 | 20095239 | 247 | 35 | 45640 | -0.17258 | 0.56851 | *GPR139* |
| ENSG00000152207 | 13 | 49270951 | 49293498 | 57 | 17 | 44330 | -0.17272 | 0.56856 | *CYSLTR2* |
| ENSG00000164902 | 5 | 125925960 | 125972944 | 168 | 22 | 30637 | -0.17275 | 0.56858 | *PHAX* |
| ENSG00000147614 | 8 | 86989552 | 87176457 | 585 | 51 | 41570 | -0.17293 | 0.56865 | *ATP6V0D2* |
| ENSG00000158077 | 11 | 7031677 | 7102539 | 307 | 24 | 49156 | -0.1731 | 0.56871 | *NLRP14* |
| ENSG00000138735 | 4 | 120405550 | 120560146 | 532 | 29 | 55744 | -0.17323 | 0.56877 | *PDE5A* |
| ENSG00000037042 | 17 | 40801323 | 40829024 | 54 | 7 | 54136 | -0.17339 | 0.56883 | *TUBG2* |
| ENSG00000090402 | 3 | 164686686 | 164806283 | 304 | 20 | 46657 | -0.17383 | 0.569 | *SI* |
| ENSG00000186976 | 22 | 43914624 | 44218217 | 1009 | 55 | 50927 | -0.1739 | 0.56903 | *EFCAB6* |
| ENSG00000198894 | 14 | 77554440 | 77593630 | 91 | 17 | 49121 | -0.17409 | 0.5691 | *CIPC* |
| ENSG00000139112 | 12 | 10355057 | 10385727 | 129 | 23 | 30526 | -0.17434 | 0.5692 | *GABARAPL1* |
| ENSG00000254462 | 11 | 57470077 | 57569058 | 159 | 18 | 45524 | -0.17488 | 0.56941 | *TMX2-CTNND1* |
| ENSG00000111247 | 12 | 4637950 | 4679214 | 88 | 18 | 45535 | -0.17486 | 0.56941 | *RAD51AP1* |
| ENSG00000074803 | 15 | 48473861 | 48606275 | 167 | 20 | 32943 | -0.17489 | 0.56942 | *SLC12A1* |
| ENSG00000255748 | 12 | 133777246 | 133797772 | 84 | 9 | 43933 | -0.17498 | 0.56945 | *AC226150.4* |
| ENSG00000034510 | 2 | 85122749 | 85143795 | 68 | 10 | 38538 | -0.17521 | 0.56954 | *TMSB10* |
| ENSG00000100097 | 22 | 38061615 | 38085813 | 72 | 17 | 51364 | -0.17544 | 0.56963 | *LGALS1* |
| ENSG00000111886 | 6 | 89956927 | 90035018 | 306 | 66 | 43313 | -0.17553 | 0.56967 | *GABRR2* |
| ENSG00000164308 | 5 | 96201643 | 96265420 | 358 | 22 | 52000 | -0.17578 | 0.56977 | *ERAP2* |
| ENSG00000134551 | 12 | 11071835 | 11094940 | 78 | 11 | 47014 | -0.17607 | 0.56988 | *PRH2* |
| ENSG00000215305 | 20 | 2811349 | 2857378 | 125 | 16 | 28215 | -0.17616 | 0.56992 | *VPS16* |
| ENSG00000197081 | 6 | 160380131 | 160544539 | 445 | 65 | 43422 | -0.17659 | 0.57009 | *IGF2R* |

| | | | | | | | | | |
|---|---|---|---|---|---|---|---|---|---|
| ENSG00000135636 | 2 | 71670852 | 71923898 | 893 | 160 | 44850 | -0.17672 | 0.57014 | *DYSF* |
| ENSG00000130518 | 19 | 18357908 | 18395319 | 191 | 24 | 50115 | -0.17672 | 0.57014 | *KIAA1683* |
| ENSG00000167553 | 12 | 49572519 | 49677114 | 220 | 40 | 40160 | -0.17682 | 0.57017 | *TUBA1C* |
| ENSG00000269375 | 14 | 101349265 | 101370646 | 59 | 21 | 49748 | -0.17694 | 0.57022 | *AL117190.3* |
| ENSG00000168447 | 16 | 23279552 | 23402620 | 327 | 68 | 42502 | -0.1773 | 0.57036 | *SCNN1B* |
| ENSG00000179148 | 17 | 7989218 | 8032365 | 126 | 37 | 45304 | -0.17735 | 0.57038 | *ALOXE3* |
| ENSG00000129455 | 19 | 51489263 | 51515967 | 103 | 14 | 38127 | -0.17744 | 0.57042 | *KLK8* |
| ENSG00000184677 | 1 | 22768344 | 22867650 | 185 | 24 | 37006 | -0.17768 | 0.57051 | *ZBTB40* |
| ENSG00000172159 | 9 | 85847905 | 86163461 | 1351 | 141 | 46833 | -0.17776 | 0.57055 | *FRMD3* |
| ENSG00000176148 | 11 | 33050963 | 33137489 | 439 | 24 | 51861 | -0.17819 | 0.57071 | *TCP11L1* |
| ENSG00000092208 | 14 | 39573427 | 39616177 | 151 | 23 | 45557 | -0.17819 | 0.57071 | *GEMIN2* |
| ENSG00000162772 | 1 | 212728676 | 212804119 | 298 | 53 | 46245 | -0.17834 | 0.57077 | *ATF3* |
| ENSG00000198315 | 6 | 28099688 | 28137250 | 135 | 12 | 47651 | -0.17847 | 0.57082 | *ZKSCAN8* |
| ENSG00000171533 | 11 | 75287963 | 75390165 | 192 | 27 | 45956 | -0.17861 | 0.57088 | *MAP6* |
| ENSG00000170454 | 12 | 52807854 | 52838309 | 119 | 16 | 47267 | -0.17883 | 0.57096 | *KRT75* |
| ENSG00000184117 | 22 | 29940797 | 29987326 | 155 | 14 | 51441 | -0.17881 | 0.57096 | *NIPSNAP1* |
| ENSG00000167414 | 19 | 47127333 | 47147942 | 78 | 16 | 51350 | -0.17884 | 0.57097 | *GNG8* |
| ENSG00000105778 | 7 | 32525038 | 33088516 | 2194 | 84 | 46200 | -0.17928 | 0.57114 | *AVL9* |
| ENSG00000180616 | 17 | 71151151 | 71177185 | 66 | 19 | 42126 | -0.17952 | 0.57124 | *SSTR2* |
| ENSG00000169550 | 11 | 26570580 | 26603780 | 168 | 28 | 52494 | -0.17967 | 0.57129 | *MUC15* |
| ENSG00000166569 | 18 | 56932388 | 56995881 | 223 | 31 | 48556 | -0.17974 | 0.57132 | *CPLX4* |
| ENSG00000261221 | 19 | 56106771 | 56138635 | 72 | 22 | 52243 | -0.17981 | 0.57135 | *ZNF865* |
| ENSG00000188001 | 3 | 188655003 | 189053093 | 820 | 117 | 38973 | -0.18016 | 0.57149 | *TPRG1* |
| ENSG00000147526 | 8 | 38575704 | 38720546 | 329 | 40 | 52171 | -0.18024 | 0.57152 | *TACC1* |
| ENSG00000186448 | 3 | 44616380 | 44699963 | 130 | 17 | 35721 | -0.18027 | 0.57153 | *ZNF197* |
| ENSG00000182111 | 7 | 57499883 | 57543252 | 210 | 11 | 51706 | -0.18043 | 0.57159 | *ZNF716* |
| ENSG00000260903 | 20 | 30545805 | 30601190 | 108 | 26 | 43936 | -0.18088 | 0.57177 | *XKR7* |
| ENSG00000250486 | 4 | 165868100 | 165890273 | 62 | 12 | 44366 | -0.18094 | 0.57179 | *FAM218A* |
| ENSG00000187122 | 10 | 98747795 | 98955677 | 608 | 97 | 39972 | -0.181 | 0.57181 | *SLIT1* |
| ENSG00000141349 | 17 | 42138103 | 42163709 | 37 | 10 | 36079 | -0.18097 | 0.57181 | *G6PC3* |
| ENSG00000170214 | 5 | 159333790 | 159409551 | 217 | 31 | 50758 | -0.18154 | 0.57203 | *ADRA1B* |
| ENSG00000180660 | 13 | 36037926 | 36060832 | 47 | 16 | 48817 | -0.18165 | 0.57207 | *MAB21L1* |
| ENSG00000158828 | 1 | 20949948 | 20988004 | 176 | 20 | 53640 | -0.18173 | 0.5721 | *PINK1* |
| ENSG00000169087 | 3 | 122448846 | 122522671 | 280 | 30 | 47617 | -0.18178 | 0.57212 | *HSPBAP1* |
| ENSG00000259024 | 17 | 15329338 | 15476875 | 362 | 40 | 45203 | -0.18191 | 0.57217 | *TVP23C-CDRT4* |
| ENSG00000135049 | 9 | 88151455 | 88366944 | 688 | 37 | 47370 | -0.182 | 0.57221 | *AGTPBP1* |

| | | | | | | | | | |
|---|---|---|---|---|---|---|---|---|---|
| ENSG00000152669 | 5 | 54516980 | 54539508 | 71 | 9 | 53318 | -0.18207 | 0.57224 | *CCNO* |
| ENSG00000104886 | 19 | 2220002 | 2247703 | 88 | 22 | 40647 | -0.18216 | 0.57227 | *PLEKHJ1* |
| ENSG00000115977 | 2 | 69678532 | 69911481 | 350 | 43 | 39655 | -0.18218 | 0.57228 | *AAK1* |
| ENSG00000166363 | 11 | 6856883 | 6877936 | 148 | 8 | 52646 | -0.18229 | 0.57232 | *OR10A5* |
| ENSG00000205464 | 5 | 81565281 | 81692796 | 259 | 31 | 41433 | -0.1825 | 0.57241 | *ATP6AP1L* |
| ENSG00000163806 | 2 | 28995383 | 29083477 | 177 | 19 | 46766 | -0.18255 | 0.57242 | *SPDYA* |
| ENSG00000131473 | 17 | 40013161 | 40096795 | 206 | 19 | 45962 | -0.18256 | 0.57243 | *ACLY* |
| ENSG00000125170 | 16 | 57495863 | 57531239 | 109 | 25 | 52224 | -0.18261 | 0.57245 | *DOK4* |
| ENSG00000143333 | 1 | 182557758 | 182583543 | 50 | 14 | 41892 | -0.18293 | 0.57257 | *RGS16* |
| ENSG00000133858 | 12 | 71993252 | 72071505 | 130 | 14 | 45110 | -0.18315 | 0.57266 | *ZFC3H1* |
| ENSG00000122483 | 1 | 93635476 | 93754287 | 317 | 27 | 40021 | -0.18355 | 0.57282 | *CCDC18* |
| ENSG00000197329 | 2 | 64309786 | 64381588 | 203 | 25 | 44257 | -0.1836 | 0.57284 | *PELI1* |
| ENSG00000141002 | 16 | 89930000 | 89987792 | 236 | 26 | 39645 | -0.18361 | 0.57284 | *TCF25* |
| ENSG00000175518 | 11 | 5525623 | 5547935 | 77 | 17 | 43763 | -0.18364 | 0.57285 | *UBQLNL* |
| ENSG00000103342 | 16 | 11951985 | 12019939 | 180 | 23 | 29829 | -0.18364 | 0.57285 | *GSPT1* |
| ENSG00000214063 | 11 | 832808 | 877116 | 151 | 20 | 32190 | -0.18365 | 0.57286 | *TSPAN4* |
| ENSG00000185198 | 19 | 675521 | 705460 | 196 | 33 | 46316 | -0.18371 | 0.57288 | *PRSS57* |
| ENSG00000196476 | 20 | 241504 | 281390 | 203 | 37 | 43758 | -0.18373 | 0.57289 | *C20orf96* |
| ENSG00000204365 | 10 | 29125337 | 29180827 | 231 | 27 | 33866 | -0.18383 | 0.57293 | *C10orf126* |
| ENSG00000147485 | 8 | 52222138 | 52732005 | 2006 | 109 | 37839 | -0.18406 | 0.57302 | *PXDNL* |
| ENSG00000166484 | 17 | 19271034 | 19296857 | 18 | 6 | 47429 | -0.18408 | 0.57302 | *MAPK7* |
| ENSG00000100201 | 22 | 38869445 | 38913665 | 81 | 16 | 40164 | -0.18406 | 0.57302 | *DDX17* |
| ENSG00000169857 | 15 | 34148428 | 34341377 | 728 | 82 | 37736 | -0.18417 | 0.57306 | *AVEN* |
| ENSG00000103266 | 16 | 720224 | 742870 | 51 | 14 | 39591 | -0.18418 | 0.57306 | *STUB1* |
| ENSG00000268891 | 7 | 74989908 | 75015425 | 2 | 1 | 34665 | -0.18428 | 0.5731 | *AC006014.1* |
| ENSG00000155849 | 7 | 36883961 | 37498852 | 2127 | 220 | 44311 | -0.18438 | 0.57314 | *ELMO1* |
| ENSG00000196209 | 20 | 1441386 | 1482233 | 130 | 22 | 32842 | -0.18469 | 0.57326 | *SIRPB2* |
| ENSG00000148672 | 10 | 88800243 | 88864623 | 189 | 24 | 40157 | -0.18484 | 0.57332 | *GLUD1* |
| ENSG00000174306 | 20 | 39797088 | 39956312 | 317 | 32 | 41685 | -0.18491 | 0.57335 | *ZHX3* |
| ENSG00000004142 | 17 | 26663659 | 26694545 | 47 | 10 | 41456 | -0.18513 | 0.57344 | *POLDIP2* |
| ENSG00000221888 | 1 | 247910676 | 247931982 | 70 | 13 | 39931 | -0.18564 | 0.57364 | *OR1C1* |
| ENSG00000114646 | 3 | 47593729 | 47632282 | 51 | 11 | 33771 | -0.18583 | 0.57371 | *CSPG5* |
| ENSG00000185345 | 6 | 161758452 | 163158803 | 5425 | 380 | 45537 | -0.18589 | 0.57373 | *PARK2* |
| ENSG00000170412 | 17 | 72410990 | 72457792 | 164 | 25 | 31030 | -0.18611 | 0.57382 | *GPRC5C* |
| ENSG00000143183 | 1 | 165686032 | 165806992 | 257 | 36 | 45456 | -0.18617 | 0.57384 | *TMCO1* |
| ENSG00000180389 | 13 | 28509343 | 28529727 | 92 | 9 | 49971 | -0.18616 | 0.57384 | *ATP5EP2* |

| | | | | | | | | | |
|---|---|---|---|---|---|---|---|---|---|
| ENSG00000089505 | 16 | 66590294 | 66623040 | 77 | 6 | 27499 | -0.18621 | 0.57386 | *CMTM1* |
| ENSG00000167916 | 17 | 38844243 | 38870002 | 77 | 11 | 45783 | -0.18635 | 0.57392 | *KRT24* |
| ENSG00000204634 | 2 | 101614079 | 101879328 | 710 | 84 | 52569 | -0.18643 | 0.57395 | *TBC1D8* |
| ENSG00000166341 | 11 | 6632556 | 6687085 | 146 | 35 | 37607 | -0.18653 | 0.57398 | *DCHS1* |
| ENSG00000185610 | 12 | 45398455 | 45454882 | 167 | 19 | 40914 | -0.18661 | 0.57402 | *DBX2* |
| ENSG00000170819 | 3 | 133108839 | 133204066 | 361 | 69 | 44720 | -0.18711 | 0.57421 | *BFSP2* |
| ENSG00000172292 | 2 | 169302372 | 169641644 | 1113 | 105 | 39176 | -0.18729 | 0.57428 | *CERS6* |
| ENSG00000183011 | 17 | 7750003 | 7798556 | 132 | 25 | 37696 | -0.18738 | 0.57432 | *LSMD1* |
| ENSG00000235568 | 22 | 42766416 | 42838401 | 273 | 41 | 43111 | -0.18738 | 0.57432 | *NFAM1* |
| ENSG00000178184 | 18 | 77905115 | 78015429 | 330 | 32 | 39746 | -0.18772 | 0.57445 | *PARD6G* |
| ENSG00000067900 | 18 | 18516867 | 18701812 | 145 | 26 | 38547 | -0.18787 | 0.57451 | *ROCK1* |
| ENSG00000148942 | 11 | 26678566 | 26754974 | 301 | 52 | 47542 | -0.18835 | 0.5747 | *SLC5A12* |
| ENSG00000167230 | 17 | 35722928 | 35759662 | 65 | 21 | 45990 | -0.18852 | 0.57477 | *C17orf78* |
| ENSG00000214397 | 15 | 100245908 | 100268029 | 74 | 17 | 39505 | -0.18876 | 0.57486 | *DKFZP779J2370* |
| ENSG00000183324 | 15 | 73725499 | 73862355 | 319 | 36 | 44647 | -0.18907 | 0.57498 | *C15orf60* |
| ENSG00000134873 | 13 | 96075858 | 96242013 | 688 | 63 | 42928 | -0.18934 | 0.57509 | *CLDN10* |
| ENSG00000134256 | 1 | 117534382 | 117589167 | 162 | 29 | 48949 | -0.18982 | 0.57527 | *CD101* |
| ENSG00000124535 | 6 | 2755648 | 2797186 | 173 | 24 | 53253 | -0.18983 | 0.57528 | *WRNIP1* |
| ENSG00000267270 | 18 | 77895807 | 77946315 | 181 | 23 | 41011 | -0.18985 | 0.57529 | *AC139100.2* |
| ENSG00000215251 | 20 | 3117165 | 3150543 | 81 | 23 | 39314 | -0.18989 | 0.5753 | *FASTKD5* |
| ENSG00000187773 | 18 | 72092963 | 72135179 | 124 | 34 | 47134 | -0.19016 | 0.57541 | *FAM69C* |
| ENSG00000143196 | 1 | 168654697 | 168708502 | 259 | 38 | 44169 | -0.19027 | 0.57545 | *DPT* |
| ENSG00000132698 | 1 | 156020951 | 156050295 | 38 | 17 | 41057 | -0.19032 | 0.57547 | *RAB25* |
| ENSG00000152454 | 19 | 58442201 | 58469043 | 63 | 10 | 43095 | -0.19039 | 0.5755 | *ZNF256* |
| ENSG00000165443 | 10 | 60926350 | 61017534 | 184 | 17 | 32935 | -0.19081 | 0.57566 | *PHYHIPL* |
| ENSG00000171488 | 1 | 90088631 | 90245462 | 332 | 28 | 54038 | -0.19101 | 0.57574 | *LRRC8C* |
| ENSG00000183801 | 11 | 7496619 | 7542608 | 214 | 30 | 51882 | -0.19124 | 0.57583 | *OLFML1* |
| ENSG00000180638 | 17 | 19571601 | 19632292 | 201 | 29 | 50305 | -0.1915 | 0.57593 | *SLC47A2* |
| ENSG00000111328 | 12 | 123735528 | 123766881 | 126 | 16 | 43718 | -0.1916 | 0.57597 | *CDK2AP1* |
| ENSG00000159346 | 1 | 202899951 | 202937700 | 101 | 18 | 41693 | -0.19172 | 0.57602 | *ADIPOR1* |
| ENSG00000130758 | 19 | 40687651 | 40731482 | 79 | 19 | 41665 | -0.19182 | 0.57606 | *MAP3K10* |
| ENSG00000178307 | 17 | 21090593 | 21127937 | 90 | 20 | 47367 | -0.19221 | 0.57621 | *TMEM11* |
| ENSG00000153902 | 19 | 35605417 | 35643355 | 108 | 15 | 53741 | -0.19232 | 0.57625 | *LGI4* |
| ENSG00000163975 | 3 | 196705492 | 196766687 | 175 | 41 | 40792 | -0.19269 | 0.5764 | *MFI2* |
| ENSG00000136870 | 9 | 104151155 | 104182942 | 118 | 17 | 50514 | -0.19294 | 0.5765 | *ZNF189* |
| ENSG00000168065 | 11 | 64313098 | 64350347 | 51 | 11 | 35242 | -0.19304 | 0.57653 | *SLC22A11* |

| | | | | | | | | | |
|---|---|---|---|---|---|---|---|---|---|
| ENSG00000135387 | 11 | 34063230 | 34132703 | 242 | 11 | 54263 | -0.19305 | 0.57654 | *CAPRIN1* |
| ENSG00000042429 | 11 | 93507393 | 93557861 | 112 | 13 | 34762 | -0.19328 | 0.57663 | *MED17* |
| ENSG00000197971 | 18 | 74680783 | 74855639 | 843 | 174 | 44539 | -0.1933 | 0.57664 | *MBP* |
| ENSG00000213973 | 19 | 22929007 | 22976909 | 251 | 14 | 40645 | -0.19386 | 0.57686 | *ZNF99* |
| ENSG00000164929 | 8 | 104142938 | 104252533 | 406 | 60 | 53769 | -0.19395 | 0.57689 | *BAALC* |
| ENSG00000166351 | 21 | 14972498 | 15023906 | 3 | 2 | 32746 | -0.194 | 0.57691 | *POTED* |
| ENSG00000116141 | 1 | 220691568 | 220847803 | 342 | 31 | 48603 | -0.19404 | 0.57693 | *MARK1* |
| ENSG00000023228 | 2 | 206969541 | 207034327 | 216 | 18 | 38268 | -0.19408 | 0.57694 | *NDUFS1* |
| ENSG00000100053 | 22 | 25585817 | 25613330 | 125 | 25 | 45864 | -0.19411 | 0.57696 | *CRYBB3* |
| ENSG00000185306 | 12 | 64650217 | 64794972 | 415 | 34 | 39789 | -0.1944 | 0.57707 | *C12orf56* |
| ENSG00000161914 | 19 | 11584242 | 11626738 | 136 | 19 | 36752 | -0.19477 | 0.57721 | *ZNF653* |
| ENSG00000185149 | 4 | 156119781 | 156148230 | 102 | 20 | 38326 | -0.19483 | 0.57724 | *NPY2R* |
| ENSG00000143185 | 1 | 168500003 | 168523235 | 97 | 16 | 39305 | -0.19492 | 0.57727 | *XCL2* |
| ENSG00000080189 | 20 | 44968167 | 45003043 | 60 | 15 | 41271 | -0.19493 | 0.57728 | *SLC35C2* |
| ENSG00000003436 | 2 | 188318957 | 188440487 | 353 | 47 | 51457 | -0.19506 | 0.57733 | *TFPI* |
| ENSG00000175906 | 17 | 41466327 | 41488505 | 62 | 11 | 29238 | -0.19521 | 0.57738 | *ARL4D* |
| ENSG00000268991 | 1 | 17050447 | 17070956 | 1 | 1 | 43638 | -0.19525 | 0.5774 | *FAM231C* |
| ENSG00000212899 | 17 | 39139686 | 39160385 | 171 | 14 | 35292 | -0.1953 | 0.57742 | *KRTAP3-3* |
| ENSG00000178171 | 2 | 131503008 | 131535707 | 52 | 14 | 34643 | -0.19553 | 0.57751 | *AMER3* |
| ENSG00000186951 | 22 | 46536424 | 46649653 | 279 | 50 | 40201 | -0.19587 | 0.57765 | *PPARA* |
| ENSG00000165204 | 9 | 125552370 | 125573395 | 52 | 12 | 38719 | -0.1961 | 0.57773 | *OR1K1* |
| ENSG00000216921 | 2 | 242826136 | 242854702 | 109 | 16 | 29867 | -0.19622 | 0.57778 | *AC131097.4* |
| ENSG00000162913 | 1 | 228381207 | 228411365 | 76 | 11 | 50646 | -0.19657 | 0.57792 | *C1orf145* |
| ENSG00000189120 | 17 | 45912279 | 45943240 | 95 | 20 | 41730 | -0.19669 | 0.57797 | *SP6* |
| ENSG00000032742 | 13 | 21130585 | 21275503 | 463 | 32 | 43497 | -0.19677 | 0.57799 | *IFT88* |
| ENSG00000066248 | 2 | 233733396 | 233887982 | 669 | 93 | 45503 | -0.19693 | 0.57806 | *NGEF* |
| ENSG00000154920 | 17 | 48440581 | 48468844 | 101 | 12 | 48058 | -0.19694 | 0.57806 | *EME1* |
| ENSG00000136546 | 2 | 167250083 | 167360757 | 330 | 36 | 53655 | -0.19701 | 0.57809 | *SCN7A* |
| ENSG00000137807 | 15 | 69696585 | 69750764 | 54 | 15 | 33442 | -0.19707 | 0.57811 | *KIF23* |
| ENSG00000163288 | 4 | 46985740 | 47438461 | 1268 | 124 | 49988 | -0.19719 | 0.57816 | *GABRB1* |
| ENSG00000103145 | 16 | 3062621 | 3084287 | 56 | 11 | 54528 | -0.19755 | 0.5783 | *HCFC1R1* |
| ENSG00000043514 | 1 | 40296723 | 40359183 | 174 | 22 | 48449 | -0.19774 | 0.57838 | *TRIT1* |
| ENSG00000196979 | 9 | 132892889 | 132916498 | 116 | 21 | 38615 | -0.19783 | 0.57841 | *AL360004.1* |
| ENSG00000249860 | 10 | 57348750 | 57370488 | 68 | 8 | 46429 | -0.19803 | 0.57849 | *MTRNR2L5* |
| ENSG00000187109 | 12 | 76428670 | 76488813 | 89 | 17 | 41662 | -0.19817 | 0.57855 | *NAP1L1* |
| ENSG00000137745 | 11 | 102803724 | 102836463 | 103 | 19 | 51792 | -0.19883 | 0.5788 | *MMP13* |

| | | | | | | | | | |
|---|---|---|---|---|---|---|---|---|---|
| ENSG00000182118 | 1 | 231144704 | 231185992 | 200 | 41 | 44310 | -0.19886 | 0.57881 | *FAM89A* |
| ENSG00000268482 | 9 | 35351056 | 35371262 | 28 | 8 | 47926 | -0.19915 | 0.57893 | *AL160274.1* |
| ENSG00000175106 | 17 | 15331205 | 15476909 | 360 | 39 | 45136 | -0.19918 | 0.57894 | *TVP23C* |
| ENSG00000181786 | 19 | 8797751 | 8819172 | 71 | 12 | 37816 | -0.1993 | 0.57899 | *ACTL9* |
| ENSG00000169752 | 15 | 76218310 | 76362136 | 334 | 19 | 40777 | -0.1996 | 0.5791 | *NRG4* |
| ENSG00000188542 | 2 | 241489471 | 241513431 | 70 | 18 | 52257 | -0.19979 | 0.57918 | *DUSP28* |
| ENSG00000141425 | 18 | 33554350 | 33657539 | 240 | 24 | 52979 | -0.1998 | 0.57918 | *RPRD1A* |
| ENSG00000233816 | 9 | 21357371 | 21378075 | 89 | 12 | 52153 | -0.20048 | 0.57945 | *IFNA13* |
| ENSG00000204178 | 1 | 25747388 | 25836700 | 153 | 10 | 25092 | -0.20067 | 0.57952 | *TMEM57* |
| ENSG00000128709 | 2 | 176977088 | 176999853 | 48 | 15 | 42418 | -0.20068 | 0.57952 | *HOXD9* |
| ENSG00000197535 | 15 | 52589480 | 52831247 | 703 | 27 | 45994 | -0.20067 | 0.57952 | *MYO5A* |
| ENSG00000268790 | 19 | 16678453 | 16780926 | 241 | 39 | 45939 | -0.20074 | 0.57955 | *CTC-429P9.4* |
| ENSG00000135439 | 12 | 58108980 | 58145940 | 56 | 17 | 43681 | -0.20076 | 0.57956 | *AGAP2* |
| ENSG00000261739 | 15 | 23589895 | 23622378 | 11 | 7 | 40285 | -0.20095 | 0.57963 | *GOLGA8S* |
| ENSG00000176302 | 11 | 118832417 | 118862001 | 102 | 13 | 54180 | -0.20178 | 0.57995 | *FOXR1* |
| ENSG00000110799 | 12 | 6048040 | 6243936 | 785 | 102 | 40417 | -0.20204 | 0.58006 | *VWF* |
| ENSG00000152229 | 18 | 43553502 | 43662238 | 363 | 31 | 34662 | -0.20221 | 0.58012 | *PSTPIP2* |
| ENSG00000186038 | 3 | 183804852 | 183834783 | 85 | 13 | 44256 | -0.20245 | 0.58022 | *HTR3E* |
| ENSG00000140043 | 14 | 74308547 | 74363530 | 221 | 15 | 44335 | -0.20244 | 0.58022 | *PTGR2* |
| ENSG00000125398 | 17 | 70107161 | 70132561 | 54 | 12 | 31877 | -0.20245 | 0.58022 | *SOX9* |
| ENSG00000119844 | 2 | 64741465 | 64830139 | 181 | 16 | 40748 | -0.2025 | 0.58024 | *AFTPH* |
| ENSG00000268470 | 17 | 76484911 | 76509138 | 156 | 32 | 51912 | -0.20261 | 0.58028 | *DNAH17-AS1* |
| ENSG00000154143 | 11 | 124471386 | 124500252 | 31 | 16 | 48714 | -0.20279 | 0.58035 | *PANX3* |
| ENSG00000138675 | 4 | 81177753 | 81267834 | 263 | 44 | 35824 | -0.20306 | 0.58046 | *FGF5* |
| ENSG00000136717 | 2 | 127795603 | 127874931 | 378 | 53 | 52779 | -0.20327 | 0.58054 | *BIN1* |
| ENSG00000248727 | 5 | 55743622 | 55787773 | 246 | 36 | 42413 | -0.20339 | 0.58058 | *CTC-236F12.4* |
| ENSG00000175538 | 11 | 74155886 | 74188774 | 92 | 20 | 51233 | -0.20341 | 0.58059 | *KCNE3* |
| ENSG00000198719 | 6 | 170581294 | 170609561 | 80 | 23 | 41481 | -0.20355 | 0.58065 | *DLL1* |
| ENSG00000107021 | 9 | 131539483 | 131582711 | 142 | 17 | 41483 | -0.20365 | 0.58069 | *TBC1D13* |
| ENSG00000181894 | 19 | 58627619 | 58676477 | 203 | 21 | 38114 | -0.20386 | 0.58077 | *ZNF329* |
| ENSG00000157557 | 21 | 40167231 | 40206879 | 142 | 14 | 36989 | -0.20389 | 0.58078 | *ETS2* |
| ENSG00000148481 | 10 | 15810169 | 15912519 | 132 | 25 | 36396 | -0.204 | 0.58082 | *FAM188A* |
| ENSG00000146281 | 6 | 89845769 | 89885284 | 136 | 15 | 42459 | -0.20426 | 0.58093 | *PM20D2* |
| ENSG00000165887 | 10 | 99322198 | 99353641 | 77 | 17 | 41308 | -0.20429 | 0.58094 | *ANKRD2* |
| ENSG00000174469 | 7 | 145803453 | 148128090 | 9186 | 404 | 44336 | -0.20462 | 0.58106 | *CNTNAP2* |
| ENSG00000143702 | 1 | 243277730 | 243428650 | 258 | 28 | 42477 | -0.20492 | 0.58118 | *CEP170* |

| | | | | | | | | | |
|---|---|---|---|---|---|---|---|---|---|
| ENSG00000180479 | 19 | 38035684 | 38095673 | 145 | 16 | 34490 | -0.20531 | 0.58133 | *ZNF571* |
| ENSG00000204385 | 6 | 31820969 | 31856823 | 122 | 28 | 34165 | -0.20594 | 0.58158 | *SLC44A4* |
| ENSG00000167741 | 17 | 4450222 | 4474113 | 88 | 26 | 49355 | -0.20606 | 0.58163 | *GGT6* |
| ENSG00000182931 | 20 | 44303292 | 44343658 | 135 | 17 | 42262 | -0.20633 | 0.58173 | *WFDC10B* |
| ENSG00000165943 | 14 | 93638541 | 93661273 | 71 | 19 | 47561 | -0.20684 | 0.58193 | *MOAP1* |
| ENSG00000181036 | 1 | 159760301 | 159796041 | 122 | 18 | 42908 | -0.20704 | 0.58201 | *FCRL6* |
| ENSG00000140749 | 16 | 21642609 | 21673981 | 50 | 18 | 37348 | -0.20721 | 0.58208 | *IGSF6* |
| ENSG00000249693 | 4 | 57386775 | 57479673 | 338 | 53 | 41810 | -0.20728 | 0.5821 | *THEGL* |
| ENSG00000130244 | 19 | 38883775 | 38909728 | 113 | 15 | 50268 | -0.20735 | 0.58213 | *FAM98C* |
| ENSG00000081148 | 3 | 100931390 | 101049404 | 247 | 21 | 46525 | -0.2075 | 0.58219 | *IMPG2* |
| ENSG00000164180 | 5 | 87475450 | 87575293 | 180 | 27 | 44610 | -0.20753 | 0.5822 | *TMEM161B* |
| ENSG00000104221 | 8 | 37690786 | 37717422 | 40 | 12 | 52564 | -0.20791 | 0.58235 | *BRF2* |
| ENSG00000121879 | 3 | 178855902 | 178967881 | 343 | 31 | 40261 | -0.208 | 0.58238 | *PIK3CA* |
| ENSG00000168333 | 8 | 49956870 | 49998649 | 95 | 14 | 43326 | -0.20831 | 0.58251 | *C8orf22* |
| ENSG00000179038 | 11 | 66953391 | 66974638 | 19 | 9 | 38634 | -0.20843 | 0.58255 | *AP001885.1* |
| ENSG00000133195 | 17 | 70632088 | 71098851 | 1742 | 213 | 42045 | -0.2085 | 0.58258 | *SLC39A11* |
| ENSG00000162591 | 1 | 3396484 | 3538059 | 579 | 117 | 42455 | -0.2088 | 0.5827 | *MEGF6* |
| ENSG00000224712 | 16 | 14792801 | 14830195 | 1 | 1 | 44444 | -0.20881 | 0.5827 | *NPIPA3* |
| ENSG00000204532 | 19 | 56703670 | 56730821 | 183 | 28 | 41707 | -0.20894 | 0.58275 | *ZSCAN5C* |
| ENSG00000002746 | 7 | 43142198 | 43615600 | 1574 | 178 | 46234 | -0.20897 | 0.58276 | *HECW1* |
| ENSG00000255359 | 11 | 22858477 | 22891972 | 114 | 14 | 45257 | -0.2093 | 0.58289 | *CCDC179* |
| ENSG00000150773 | 11 | 111924734 | 111954998 | 46 | 11 | 34352 | -0.2096 | 0.58301 | *PIH1D2* |
| ENSG00000138378 | 2 | 191884302 | 192026322 | 305 | 48 | 46110 | -0.20979 | 0.58308 | *STAT4* |
| ENSG00000176566 | 8 | 88872973 | 88896296 | 37 | 13 | 45449 | -0.20981 | 0.58309 | *DCAF4L2* |
| ENSG00000258653 | 14 | 74308674 | 74408981 | 402 | 23 | 41417 | -0.20994 | 0.58314 | *RP5-1021I20.4* |
| ENSG00000182379 | 12 | 57600578 | 57630232 | 46 | 13 | 41408 | -0.21 | 0.58317 | *NXPH4* |
| ENSG00000197168 | 13 | 52601093 | 52713214 | 286 | 25 | 38687 | -0.21006 | 0.58319 | *NEK5* |
| ENSG00000168005 | 11 | 63570860 | 63605190 | 73 | 16 | 33462 | -0.21045 | 0.58334 | *C11orf84* |
| ENSG00000086065 | 9 | 33254940 | 33291977 | 43 | 9 | 48847 | -0.21077 | 0.58347 | *CHMP5* |
| ENSG00000183671 | 2 | 207030040 | 207092771 | 189 | 29 | 50127 | -0.21087 | 0.58351 | *GPR1* |
| ENSG00000186272 | 19 | 57912531 | 57943307 | 66 | 13 | 47619 | -0.21095 | 0.58354 | *ZNF17* |
| ENSG00000185974 | 13 | 114311470 | 114450708 | 125 | 20 | 41461 | -0.21135 | 0.58369 | *GRK1* |
| ENSG00000065526 | 1 | 16164359 | 16276955 | 222 | 27 | 50302 | -0.21169 | 0.58383 | *SPEN* |
| ENSG00000132406 | 4 | 4227269 | 4259950 | 199 | 14 | 51778 | -0.21196 | 0.58393 | *TMEM128* |
| ENSG00000104408 | 8 | 109203445 | 109457562 | 894 | 52 | 41208 | -0.21215 | 0.584 | *EIF3E* |
| ENSG00000166153 | 12 | 100587447 | 100670857 | 199 | 34 | 45443 | -0.21214 | 0.584 | *DEPDC4* |

| | | | | | | | | | |
|---|---|---|---|---|---|---|---|---|---|
| ENSG00000172742 | 11 | 59272386 | 59293330 | 34 | 6 | 55471 | -0.21218 | 0.58402 | *OR4D9* |
| ENSG00000017797 | 18 | 9465007 | 9548114 | 326 | 39 | 47436 | -0.21234 | 0.58408 | *RALBP1* |
| ENSG00000114487 | 3 | 108667086 | 108846989 | 641 | 35 | 47389 | -0.21238 | 0.58409 | *MORC1* |
| ENSG00000136933 | 9 | 127952821 | 128006437 | 142 | 19 | 50139 | -0.21239 | 0.5841 | *RABEPK* |
| ENSG00000171236 | 19 | 4526421 | 4550486 | 68 | 17 | 37574 | -0.21262 | 0.58419 | *LRG1* |
| ENSG00000165973 | 11 | 20681117 | 21607227 | 3472 | 320 | 44453 | -0.21297 | 0.58432 | *NELL1* |
| ENSG00000103876 | 15 | 80434832 | 80489288 | 207 | 31 | 52904 | -0.21303 | 0.58435 | *FAH* |
| ENSG00000131127 | 4 | 321603 | 388653 | 275 | 21 | 42499 | -0.21316 | 0.5844 | *ZNF141* |
| ENSG00000213199 | 7 | 150735379 | 150759843 | 54 | 15 | 54839 | -0.21335 | 0.58447 | *ASIC3* |
| ENSG00000176387 | 16 | 67454555 | 67481456 | 32 | 13 | 45514 | -0.21354 | 0.58455 | *HSD11B2* |
| ENSG00000127884 | 10 | 135165984 | 135197193 | 129 | 16 | 28951 | -0.21364 | 0.58459 | *ECHS1* |
| ENSG00000124571 | 6 | 43480072 | 43553812 | 129 | 17 | 41184 | -0.21369 | 0.58461 | *XPO5* |
| ENSG00000158486 | 16 | 20934433 | 21180762 | 736 | 76 | 52438 | -0.21376 | 0.58463 | *DNAH3* |
| ENSG00000121579 | 3 | 113425307 | 113475147 | 75 | 8 | 50046 | -0.2138 | 0.58465 | *NAA50* |
| ENSG00000186642 | 11 | 72277185 | 72395635 | 358 | 47 | 45592 | -0.2138 | 0.58465 | *PDE2A* |
| ENSG00000154645 | 21 | 19263580 | 19649690 | 1216 | 139 | 40551 | -0.21435 | 0.58486 | *CHODL* |
| ENSG00000176142 | 3 | 119138347 | 119197677 | 196 | 21 | 54083 | -0.21445 | 0.5849 | *TMEM39A* |
| ENSG00000198390 | 21 | 31758392 | 31779136 | 47 | 9 | 52855 | -0.21463 | 0.58497 | *KRTAP13-1* |
| ENSG00000183155 | 1 | 202838085 | 202868263 | 80 | 12 | 48744 | -0.2148 | 0.58504 | *RABIF* |
| ENSG00000179097 | 3 | 88029846 | 88052919 | 61 | 9 | 48385 | -0.21487 | 0.58506 | *HTR1F* |
| ENSG00000152380 | 5 | 79773788 | 79848382 | 191 | 31 | 43083 | -0.21493 | 0.58509 | *FAM151B* |
| ENSG00000182473 | 17 | 74067087 | 74127657 | 152 | 19 | 40873 | -0.21509 | 0.58515 | *EXOC7* |
| ENSG00000182132 | 5 | 169770491 | 170173636 | 1387 | 218 | 45618 | -0.21515 | 0.58518 | *KCNIP1* |
| ENSG00000173409 | 1 | 231104727 | 231146341 | 171 | 18 | 45116 | -0.21523 | 0.5852 | *ARV1* |
| ENSG00000113302 | 5 | 158731791 | 158767895 | 78 | 13 | 52721 | -0.21541 | 0.58528 | *IL12B* |
| ENSG00000173546 | 15 | 75956663 | 76015189 | 112 | 19 | 42754 | -0.21563 | 0.58536 | *CSPG4* |
| ENSG00000157456 | 15 | 59387277 | 59427244 | 81 | 12 | 48106 | -0.21577 | 0.58542 | *CCNB2* |
| ENSG00000187581 | 14 | 93803537 | 93824702 | 105 | 10 | 32993 | -0.21589 | 0.58546 | *COX8C* |
| ENSG00000090857 | 16 | 70137529 | 70205203 | 66 | 11 | 40717 | -0.21593 | 0.58548 | *PDPR* |
| ENSG00000179168 | 19 | 38864905 | 38888722 | 75 | 10 | 50318 | -0.21652 | 0.58571 | *GGN* |
| ENSG00000204682 | 10 | 21771587 | 21796191 | 28 | 9 | 47629 | -0.21654 | 0.58572 | *CASC10* |
| ENSG00000234776 | 11 | 45918085 | 45938833 | 23 | 8 | 54640 | -0.21664 | 0.58576 | *C11orf94* |
| ENSG00000189152 | 17 | 19020782 | 19072489 | 6 | 3 | 43590 | -0.21685 | 0.58584 | *GRAPL* |
| ENSG00000262180 | 1 | 186359704 | 186380587 | 51 | 14 | 53512 | -0.21689 | 0.58585 | *OCLM* |
| ENSG00000148584 | 10 | 52549169 | 52655435 | 184 | 32 | 42409 | -0.21709 | 0.58593 | *A1CF* |
| ENSG00000172673 | 6 | 128019217 | 128249776 | 515 | 59 | 45565 | -0.21731 | 0.58602 | *THEMIS* |

| | | | | | | | | | |
|---|---|---|---|---|---|---|---|---|---|
| ENSG00000168765 | 1 | 110188703 | 110218118 | 87 | 17 | 48797 | -0.21736 | 0.58603 | *GSTM4* |
| ENSG00000164362 | 5 | 1243262 | 1305184 | 156 | 55 | 45333 | -0.21734 | 0.58603 | *TERT* |
| ENSG00000110887 | 12 | 109242708 | 109304819 | 182 | 35 | 33854 | -0.21741 | 0.58606 | *DAO* |
| ENSG00000186442 | 12 | 53173469 | 53199901 | 93 | 23 | 38754 | -0.21765 | 0.58615 | *KRT3* |
| ENSG00000128250 | 22 | 29824572 | 29848444 | 83 | 13 | 51171 | -0.21783 | 0.58622 | *RFPL1* |
| ENSG00000143466 | 1 | 206633791 | 206680223 | 164 | 37 | 44467 | -0.21795 | 0.58627 | *IKBKE* |
| ENSG00000154478 | 10 | 125415871 | 125464123 | 177 | 36 | 49348 | -0.218 | 0.58628 | *GPR26* |
| ENSG00000214960 | 7 | 16120817 | 16470947 | 1226 | 91 | 43187 | -0.21808 | 0.58632 | *ISPD* |
| ENSG00000081181 | 14 | 68076515 | 68128437 | 121 | 18 | 36319 | -0.21809 | 0.58632 | *ARG2* |
| ENSG00000172845 | 2 | 174761187 | 174840430 | 281 | 30 | 52044 | -0.21884 | 0.58661 | *SP3* |
| ENSG00000166783 | 16 | 15678243 | 15747023 | 150 | 35 | 43167 | -0.21885 | 0.58662 | *KIAA0430* |
| ENSG00000123405 | 12 | 54675895 | 54704905 | 36 | 14 | 37896 | -0.21889 | 0.58663 | *NFE2* |
| ENSG00000187624 | 17 | 250118 | 283510 | 123 | 27 | 41573 | -0.21891 | 0.58664 | *C17orf97* |
| ENSG00000100292 | 22 | 35766354 | 35800207 | 114 | 19 | 43873 | -0.21916 | 0.58674 | *HMOX1* |
| ENSG00000254019 | 8 | 142254664 | 142276916 | 71 | 13 | 52165 | -0.21925 | 0.58677 | *RP11-10J21.3* |
| ENSG00000204700 | 6 | 29131311 | 29152351 | 127 | 14 | 33381 | -0.21973 | 0.58696 | *OR2J2* |
| ENSG00000076108 | 12 | 56979380 | 57040600 | 80 | 23 | 44161 | -0.21973 | 0.58696 | *BAZ2A* |
| ENSG00000224383 | 17 | 62065711 | 62091664 | 37 | 15 | 48901 | -0.21979 | 0.58698 | *C17orf72* |
| ENSG00000187258 | 7 | 34687851 | 34927944 | 971 | 95 | 47437 | -0.22013 | 0.58711 | *NPSR1* |
| ENSG00000120896 | 8 | 22392499 | 22443301 | 95 | 28 | 47148 | -0.22014 | 0.58712 | *SORBS3* |
| ENSG00000176974 | 17 | 18221187 | 18276856 | 181 | 18 | 51485 | -0.22024 | 0.58716 | *SHMT1* |
| ENSG00000101052 | 20 | 42209571 | 42285936 | 234 | 16 | 53617 | -0.22096 | 0.58744 | *IFT52* |
| ENSG00000006453 | 7 | 97910963 | 98040380 | 377 | 20 | 29158 | -0.22117 | 0.58752 | *BAIAP2L1* |
| ENSG00000163660 | 3 | 156854297 | 156888549 | 111 | 12 | 50297 | -0.22124 | 0.58755 | *CCNL1* |
| ENSG00000145996 | 6 | 20524688 | 21242635 | 2503 | 178 | 49772 | -0.22131 | 0.58757 | *CDKAL1* |
| ENSG00000073861 | 17 | 45800610 | 45833485 | 100 | 17 | 55013 | -0.22133 | 0.58758 | *TBX21* |
| ENSG00000234829 | 9 | 21217242 | 21238221 | 127 | 10 | 44062 | -0.22146 | 0.58763 | *IFNA17* |
| ENSG00000186918 | 8 | 28193102 | 28270218 | 124 | 24 | 40333 | -0.2216 | 0.58769 | *ZNF395* |
| ENSG00000121594 | 3 | 119233140 | 119288449 | 242 | 31 | 38765 | -0.22164 | 0.5877 | *CD80* |
| ENSG00000075407 | 10 | 38373264 | 38424472 | 207 | 12 | 54596 | -0.22181 | 0.58777 | *ZNF37A* |
| ENSG00000269476 | 19 | 58378133 | 58437959 | 170 | 24 | 45434 | -0.22205 | 0.58786 | *CTD-2583A14.9* |
| ENSG00000163689 | 3 | 58693092 | 59045810 | 584 | 41 | 35052 | -0.22218 | 0.58791 | *C3orf67* |
| ENSG00000140391 | 15 | 77326359 | 77386326 | 80 | 16 | 42283 | -0.22218 | 0.58791 | *TSPAN3* |
| ENSG00000272410 | 3 | 10281056 | 10337480 | 165 | 26 | 35682 | -0.22235 | 0.58798 | *RP11-438J1.1* |
| ENSG00000143319 | 1 | 156681683 | 156708591 | 58 | 18 | 41539 | -0.22247 | 0.58803 | *ISG20L2* |
| ENSG00000116785 | 1 | 196733925 | 196773203 | 13 | 8 | 40026 | -0.22313 | 0.58828 | *CFHR3* |

| | | | | | | | | | |
|---|---|---|---|---|---|---|---|---|---|
| ENSG00000158428 | 2 | 219211579 | 219242822 | 102 | 20 | 45978 | -0.22323 | 0.58832 | *C2orf62* |
| ENSG00000168092 | 11 | 117004983 | 117057610 | 181 | 25 | 40741 | -0.22352 | 0.58843 | *PAFAH1B2* |
| ENSG00000196646 | 19 | 12263879 | 12310064 | 89 | 16 | 38616 | -0.22367 | 0.58849 | *ZNF136* |
| ENSG00000182851 | 8 | 144285068 | 144309044 | 109 | 18 | 49129 | -0.22378 | 0.58854 | *GPIHBP1* |
| ENSG00000166200 | 15 | 49388268 | 49457858 | 199 | 26 | 39663 | -0.22391 | 0.58859 | *COPS2* |
| ENSG00000164109 | 4 | 120966763 | 120998229 | 102 | 18 | 52548 | -0.224 | 0.58862 | *MAD2L1* |
| ENSG00000168274 | 6 | 26207165 | 26227711 | 98 | 9 | 54085 | -0.22403 | 0.58863 | *HIST1H2AE* |
| ENSG00000136940 | 9 | 125550668 | 125600910 | 99 | 23 | 42335 | -0.22432 | 0.58875 | *PDCL* |
| ENSG00000151422 | 5 | 108073523 | 108542542 | 1996 | 104 | 52532 | -0.22436 | 0.58876 | *FER* |
| ENSG00000179833 | 2 | 64848755 | 64988139 | 295 | 37 | 48283 | -0.22439 | 0.58877 | *SERTAD2* |
| ENSG00000258987 | 14 | 94395901 | 94420475 | 85 | 13 | 39411 | -0.22439 | 0.58877 | *RP11-131H24.4* |
| ENSG00000159445 | 1 | 151836060 | 151892284 | 144 | 15 | 45031 | -0.22454 | 0.58883 | *THEM4* |
| ENSG00000141497 | 17 | 4633319 | 4659411 | 93 | 25 | 47957 | -0.22469 | 0.58889 | *ZMYND15* |
| ENSG00000171700 | 20 | 62694534 | 62721323 | 73 | 17 | 48980 | -0.22468 | 0.58889 | *RGS19* |
| ENSG00000255062 | 11 | 126154193 | 126184213 | 102 | 19 | 40607 | -0.22473 | 0.58891 | *RP11-712L6.5* |
| ENSG00000173465 | 11 | 65327901 | 65351413 | 42 | 10 | 47440 | -0.22487 | 0.58896 | *SSSCA1* |
| ENSG00000115484 | 2 | 62085224 | 62125939 | 139 | 18 | 53786 | -0.22514 | 0.58906 | *CCT4* |
| ENSG00000139194 | 12 | 7266280 | 7291538 | 67 | 16 | 47170 | -0.2253 | 0.58913 | *RBP5* |
| ENSG00000105366 | 19 | 51944101 | 51971710 | 61 | 15 | 43049 | -0.22575 | 0.5893 | *SIGLEC8* |
| ENSG00000213648 | 16 | 29457127 | 29486300 | 1 | 1 | 43583 | -0.226 | 0.5894 | *SULT1A4* |
| ENSG00000258150 | 16 | 29466288 | 29489154 | 1 | 1 | 43583 | -0.226 | 0.5894 | *RP11-345J4.3* |
| ENSG00000166634 | 18 | 61213393 | 61244244 | 89 | 10 | 27721 | -0.22603 | 0.58941 | *SERPINB12* |
| ENSG00000130675 | 7 | 156776745 | 156813345 | 150 | 34 | 42023 | -0.22609 | 0.58944 | *MNX1* |
| ENSG00000135506 | 12 | 58077738 | 58125340 | 97 | 13 | 39464 | -0.22621 | 0.58948 | *OS9* |
| ENSG00000088854 | 20 | 3219951 | 3398272 | 733 | 34 | 54967 | -0.22635 | 0.58953 | *C20orf194* |
| ENSG00000136783 | 9 | 107499969 | 107532403 | 110 | 21 | 45670 | -0.2265 | 0.58959 | *NIPSNAP3A* |
| ENSG00000156968 | 16 | 15479611 | 15517125 | 125 | 25 | 47164 | -0.22654 | 0.58961 | *MPV17L* |
| ENSG00000158747 | 1 | 19957048 | 19994945 | 142 | 32 | 47988 | -0.22656 | 0.58962 | *NBL1* |
| ENSG00000144504 | 2 | 241408839 | 241518626 | 408 | 63 | 50062 | -0.2266 | 0.58963 | *ANKMY1* |
| ENSG00000148346 | 9 | 130901350 | 130925734 | 46 | 13 | 33912 | -0.22676 | 0.58969 | *LCN2* |
| ENSG00000100564 | 14 | 68038672 | 68077004 | 145 | 26 | 41431 | -0.22687 | 0.58974 | *PIGH* |
| ENSG00000198792 | 22 | 38605298 | 38679040 | 191 | 34 | 44680 | -0.22701 | 0.58979 | *TMEM184B* |
| ENSG00000160882 | 8 | 143943772 | 143971262 | 122 | 12 | 30366 | -0.22725 | 0.58989 | *CYP11B1* |
| ENSG00000165185 | 9 | 115239127 | 115441677 | 515 | 49 | 51458 | -0.22779 | 0.5901 | *KIAA1958* |
| ENSG00000115266 | 19 | 1436300 | 1483243 | 110 | 34 | 41328 | -0.2283 | 0.59029 | *APC2* |
| ENSG00000166118 | 11 | 133700526 | 133725433 | 95 | 8 | 51167 | -0.22856 | 0.59039 | *SPATA19* |

| | | | | | | | | | |
|---|---|---|---|---|---|---|---|---|---|
| ENSG00000229314 | 9 | 117075336 | 117098755 | 53 | 20 | 47696 | -0.22861 | 0.59041 | *ORM1* |
| ENSG00000180448 | 19 | 1055922 | 1096627 | 189 | 37 | 44440 | -0.22865 | 0.59043 | *HMHA1* |
| ENSG00000105186 | 19 | 33077913 | 33177503 | 522 | 45 | 38915 | -0.22942 | 0.59073 | *ANKRD27* |
| ENSG00000010310 | 19 | 46161502 | 46196982 | 136 | 19 | 51881 | -0.22957 | 0.59079 | *GIPR* |
| ENSG00000188710 | 9 | 133758815 | 133779225 | 89 | 22 | 39215 | -0.2296 | 0.5908 | *QRFP* |
| ENSG00000166847 | 16 | 23642713 | 23691195 | 162 | 14 | 36266 | -0.23003 | 0.59096 | *DCTN5* |
| ENSG00000167840 | 17 | 4998836 | 5036411 | 109 | 24 | 41596 | -0.23003 | 0.59097 | *ZNF232* |
| ENSG00000142546 | 19 | 50048968 | 50103519 | 140 | 22 | 38701 | -0.2301 | 0.59099 | *NOSIP* |
| ENSG00000141568 | 17 | 80467589 | 80612538 | 666 | 84 | 47847 | -0.23026 | 0.59105 | *FOXK2* |
| ENSG00000171103 | 2 | 29062687 | 29103167 | 87 | 14 | 50595 | -0.23055 | 0.59117 | *TRMT61B* |
| ENSG00000139540 | 12 | 56613833 | 56641630 | 31 | 8 | 45964 | -0.23058 | 0.59118 | *SLC39A5* |
| ENSG00000075089 | 12 | 100582900 | 100645643 | 171 | 28 | 43760 | -0.23075 | 0.59125 | *ACTR6* |
| ENSG00000086666 | 15 | 80341910 | 80440735 | 234 | 28 | 47806 | -0.23079 | 0.59126 | *ZFAND6* |
| ENSG00000163032 | 2 | 17710393 | 17848285 | 509 | 35 | 44311 | -0.23137 | 0.59149 | *VSNL1* |
| ENSG00000163083 | 2 | 121093719 | 121119384 | 60 | 15 | 31760 | -0.23138 | 0.59149 | *INHBB* |
| ENSG00000231068 | 21 | 32080843 | 32101095 | 67 | 5 | 44517 | -0.23152 | 0.59155 | *KRTAP21-3* |
| ENSG00000100726 | 16 | 1533345 | 1570458 | 202 | 39 | 39770 | -0.23163 | 0.59159 | *TELO2* |
| ENSG00000154646 | 21 | 19631433 | 19868197 | 840 | 124 | 42863 | -0.23171 | 0.59162 | *TMPRSS15* |
| ENSG00000070269 | 14 | 56945072 | 57127324 | 698 | 96 | 41909 | -0.23178 | 0.59164 | *TMEM260* |
| ENSG00000177707 | 3 | 110778918 | 111004410 | 511 | 65 | 46222 | -0.23185 | 0.59167 | *PVRL3* |
| ENSG00000162493 | 1 | 13899960 | 13954452 | 175 | 43 | 41252 | -0.23201 | 0.59174 | *PDPN* |
| ENSG00000254535 | 4 | 135107488 | 135132903 | 51 | 12 | 34616 | -0.23203 | 0.59174 | *PABPC4L* |
| ENSG00000198286 | 7 | 2935775 | 3093579 | 698 | 139 | 45870 | -0.23206 | 0.59176 | *CARD11* |
| ENSG00000077616 | 11 | 89854683 | 89936062 | 401 | 26 | 41659 | -0.23214 | 0.59179 | *NAALAD2* |
| ENSG00000159398 | 16 | 55870066 | 55999943 | 387 | 61 | 42777 | -0.23244 | 0.5919 | *CES5A* |
| ENSG00000104899 | 19 | 2239308 | 2262072 | 79 | 28 | 41274 | -0.23242 | 0.5919 | *AMH* |
| ENSG00000185361 | 19 | 4629530 | 4665580 | 136 | 25 | 43119 | -0.23252 | 0.59193 | *TNFAIP8L1* |
| ENSG00000154529 | 9 | 43674902 | 43934049 | 21 | 8 | 42656 | -0.23264 | 0.59198 | *CNTNAP3B* |
| ENSG00000185686 | 22 | 22880123 | 22911768 | 143 | 17 | 35349 | -0.23292 | 0.59209 | *PRAME* |
| ENSG00000128536 | 7 | 105507242 | 105686877 | 681 | 128 | 47973 | -0.23315 | 0.59218 | *CDHR3* |
| ENSG00000168589 | 16 | 80564631 | 80594657 | 128 | 15 | 33923 | -0.23322 | 0.5922 | *DYNLRB2* |
| ENSG00000118816 | 4 | 77958311 | 78007158 | 105 | 23 | 41283 | -0.23345 | 0.59229 | *CCNI* |
| ENSG00000171476 | 4 | 57504155 | 57558065 | 225 | 39 | 40703 | -0.23372 | 0.5924 | *HOPX* |
| ENSG00000148826 | 10 | 134588297 | 134609556 | 59 | 19 | 50484 | -0.23416 | 0.59257 | *NKX6-2* |
| ENSG00000117625 | 1 | 211421719 | 211499727 | 285 | 35 | 46935 | -0.23425 | 0.59261 | *RCOR3* |
| ENSG00000197385 | 3 | 32013263 | 32043120 | 103 | 13 | 25298 | -0.23451 | 0.5927 | *ZNF860* |

| | | | | | | | | | |
|---|---|---|---|---|---|---|---|---|---|
| ENSG00000228474 | 2 | 27283340 | 27304641 | 35 | 9 | 41514 | -0.23462 | 0.59275 | *OST4* |
| ENSG00000179562 | 7 | 127210672 | 127243665 | 81 | 17 | 38520 | -0.23484 | 0.59284 | *GCC1* |
| ENSG00000197635 | 2 | 162838751 | 162941052 | 257 | 43 | 40967 | -0.23511 | 0.59294 | *DPP4* |
| ENSG00000131059 | 20 | 31795116 | 31825564 | 48 | 9 | 45225 | -0.23527 | 0.593 | *BPIFA3* |
| ENSG00000197599 | 16 | 1474384 | 1504557 | 168 | 27 | 44735 | -0.23537 | 0.59304 | *CCDC154* |
| ENSG00000147586 | 8 | 80820952 | 80952524 | 201 | 32 | 47548 | -0.23559 | 0.59312 | *MRPS28* |
| ENSG00000176922 | 11 | 4859427 | 4880514 | 82 | 12 | 49368 | -0.23586 | 0.59323 | *OR51S1* |
| ENSG00000066455 | 14 | 93250576 | 93316308 | 280 | 18 | 50057 | -0.23598 | 0.59328 | *GOLGA5* |
| ENSG00000158402 | 5 | 137610954 | 137684044 | 176 | 22 | 47203 | -0.2362 | 0.59336 | *CDC25C* |
| ENSG00000154783 | 3 | 14850469 | 14985895 | 625 | 57 | 49414 | -0.23662 | 0.59353 | *FGD5* |
| ENSG00000099785 | 19 | 8468154 | 8513901 | 165 | 33 | 41969 | -0.23725 | 0.59377 | *Mar-02* |
| ENSG00000127366 | 7 | 141480017 | 141501166 | 54 | 4 | 51021 | -0.2376 | 0.5939 | *TAS2R5* |
| ENSG00000074356 | 17 | 3704460 | 3759545 | 156 | 30 | 40678 | -0.2379 | 0.59402 | *C17orf85* |
| ENSG00000167232 | 19 | 23477793 | 23588362 | 387 | 25 | 45510 | -0.2379 | 0.59402 | *ZNF91* |
| ENSG00000162365 | 1 | 47593107 | 47625413 | 120 | 12 | 36992 | -0.23795 | 0.59404 | *CYP4A22* |
| ENSG00000134852 | 4 | 56284070 | 56423305 | 576 | 25 | 50015 | -0.23794 | 0.59404 | *CLOCK* |
| ENSG00000116745 | 1 | 68884505 | 68925642 | 135 | 24 | 40047 | -0.23798 | 0.59405 | *RPE65* |
| ENSG00000196345 | 3 | 44586685 | 44634975 | 129 | 15 | 31691 | -0.23801 | 0.59406 | *ZKSCAN7* |
| ENSG00000164742 | 7 | 45603739 | 45772715 | 542 | 79 | 47139 | -0.23804 | 0.59407 | *ADCY1* |
| ENSG00000122641 | 7 | 41714712 | 41752706 | 50 | 14 | 53237 | -0.23808 | 0.59409 | *INHBA* |
| ENSG00000142513 | 19 | 51283672 | 51308481 | 128 | 19 | 47880 | -0.23869 | 0.59433 | *ACPT* |
| ENSG00000119725 | 14 | 74343320 | 74409214 | 257 | 19 | 37779 | -0.23892 | 0.59442 | *ZNF410* |
| ENSG00000116194 | 1 | 178808840 | 178850187 | 88 | 16 | 39840 | -0.23907 | 0.59447 | *ANGPTL1* |
| ENSG00000116649 | 1 | 11104641 | 11130081 | 62 | 8 | 27359 | -0.23926 | 0.59455 | *SRM* |
| ENSG00000091157 | 18 | 54308574 | 54708828 | 1128 | 92 | 45020 | -0.23932 | 0.59457 | *WDR7* |
| ENSG00000130396 | 6 | 168217602 | 168382703 | 509 | 39 | 44944 | -0.23966 | 0.5947 | *MLLT4* |
| ENSG00000144320 | 2 | 176778620 | 176877567 | 296 | 32 | 49079 | -0.23979 | 0.59475 | *KIAA1715* |
| ENSG00000018869 | 19 | 56877413 | 56914914 | 121 | 33 | 44835 | -0.23977 | 0.59475 | *ZNF582* |
| ENSG00000160888 | 19 | 13251229 | 13275722 | 56 | 13 | 38084 | -0.23992 | 0.5948 | *IER2* |
| ENSG00000053524 | 3 | 182885831 | 183156566 | 851 | 100 | 40229 | -0.23996 | 0.59482 | *MCF2L2* |
| ENSG00000165233 | 9 | 95848500 | 95885565 | 128 | 21 | 34462 | -0.24002 | 0.59484 | *C9orf89* |
| ENSG00000127720 | 12 | 82742276 | 82883015 | 378 | 20 | 41298 | -0.24014 | 0.59489 | *METTL25* |
| ENSG00000076826 | 19 | 7650788 | 7693190 | 117 | 24 | 48376 | -0.24068 | 0.5951 | *CAMSAP3* |
| ENSG00000269305 | 9 | 4849120 | 4869260 | 96 | 16 | 48695 | -0.24077 | 0.59514 | *AL158147.2* |
| ENSG00000089006 | 20 | 17912241 | 17959623 | 179 | 15 | 54589 | -0.24078 | 0.59514 | *SNX5* |
| ENSG00000173868 | 17 | 47290724 | 47318128 | 80 | 19 | 45669 | -0.24102 | 0.59523 | *PHOSPHO1* |

| | | | | | | | | | |
|---|---|---|---|---|---|---|---|---|---|
| ENSG00000006555 | 1 | 55235385 | 55276940 | 138 | 29 | 51246 | -0.24113 | 0.59527 | *TTC22* |
| ENSG00000161652 | 19 | 50645805 | 50676452 | 83 | 18 | 50128 | -0.24116 | 0.59528 | *IZUMO2* |
| ENSG00000102781 | 13 | 30766767 | 30891621 | 459 | 68 | 53912 | -0.24172 | 0.5955 | *KATNAL1* |
| ENSG00000163104 | 4 | 95118762 | 95222443 | 260 | 17 | 51594 | -0.24174 | 0.59551 | *SMARCAD1* |
| ENSG00000119185 | 2 | 9533604 | 9573676 | 109 | 18 | 35385 | -0.24202 | 0.59562 | *ITGB1BP1* |
| ENSG00000120337 | 1 | 172999100 | 173030103 | 64 | 13 | 36065 | -0.24208 | 0.59564 | *TNFSF18* |
| ENSG00000084072 | 1 | 40147854 | 40239586 | 297 | 59 | 48379 | -0.24255 | 0.59582 | *PPIE* |
| ENSG00000109193 | 4 | 70666498 | 70735870 | 209 | 31 | 40712 | -0.24274 | 0.5959 | *SULT1E1* |
| ENSG00000140326 | 15 | 43005757 | 43039324 | 65 | 8 | 24586 | -0.24336 | 0.59614 | *CDAN1* |
| ENSG00000254827 | 11 | 2899010 | 2934970 | 119 | 30 | 47688 | -0.24344 | 0.59617 | *SLC22A18AS* |
| ENSG00000164934 | 8 | 104416942 | 104465681 | 149 | 12 | 54611 | -0.24349 | 0.59619 | *DCAF13* |
| ENSG00000197299 | 15 | 91250558 | 91368859 | 390 | 31 | 45639 | -0.24356 | 0.59621 | *BLM* |
| ENSG00000187559 | 9 | 70907276 | 70930000 | 1 | 1 | 55924 | -0.24456 | 0.5966 | *FOXD4L3* |
| ENSG00000108786 | 17 | 40691232 | 40717231 | 71 | 12 | 51245 | -0.24458 | 0.59661 | *HSD17B1* |
| ENSG00000187569 | 12 | 7854050 | 7880155 | 141 | 28 | 49994 | -0.24463 | 0.59663 | *DPPA3* |
| ENSG00000151532 | 10 | 114196756 | 114588503 | 903 | 161 | 42291 | -0.24494 | 0.59675 | *VTI1A* |
| ENSG00000145826 | 5 | 135247968 | 135300723 | 263 | 25 | 38289 | -0.24563 | 0.59702 | *LECT2* |
| ENSG00000186566 | 17 | 42462652 | 42590798 | 235 | 18 | 46646 | -0.24599 | 0.59715 | *GPATCH8* |
| ENSG00000153406 | 16 | 4501681 | 4555764 | 178 | 15 | 32254 | -0.24611 | 0.5972 | *NMRAL1* |
| ENSG00000112539 | 6 | 165683153 | 165733096 | 170 | 27 | 45807 | -0.24629 | 0.59727 | *C6orf118* |
| ENSG00000101438 | 20 | 37343105 | 37368015 | 89 | 16 | 52257 | -0.24634 | 0.59729 | *SLC32A1* |
| ENSG00000182208 | 11 | 1480687 | 1532477 | 205 | 18 | 54289 | -0.24651 | 0.59736 | *MOB2* |
| ENSG00000268012 | 5 | 78513714 | 78533911 | 44 | 8 | 32211 | -0.24679 | 0.59747 | *AC016559.1* |
| ENSG00000185112 | 3 | 194396622 | 194419762 | 116 | 20 | 45071 | -0.24696 | 0.59753 | *FAM43A* |
| ENSG00000171017 | 19 | 7943390 | 7976901 | 124 | 26 | 44966 | -0.24739 | 0.5977 | *LRRC8E* |
| ENSG00000172671 | 10 | 46101043 | 46178228 | 258 | 17 | 53911 | -0.24743 | 0.59771 | *ZFAND4* |
| ENSG00000074590 | 12 | 106447118 | 106543811 | 282 | 59 | 43213 | -0.2479 | 0.59789 | *NUAK1* |
| ENSG00000185479 | 12 | 52830435 | 52855910 | 101 | 11 | 47845 | -0.24831 | 0.59805 | *KRT6B* |
| ENSG00000145850 | 5 | 156336293 | 156400266 | 181 | 30 | 50190 | -0.24832 | 0.59806 | *TIMD4* |
| ENSG00000166887 | 15 | 42440899 | 42510514 | 137 | 18 | 31326 | -0.2484 | 0.59809 | *VPS39* |
| ENSG00000087157 | 17 | 76364721 | 76431195 | 336 | 29 | 50037 | -0.24851 | 0.59813 | *PGS1* |
| ENSG00000146426 | 6 | 155143831 | 155588857 | 1644 | 234 | 46923 | -0.24887 | 0.59827 | *TIAM2* |
| ENSG00000182575 | 17 | 47643220 | 47671189 | 52 | 21 | 45016 | -0.249 | 0.59832 | *NXPH3* |
| ENSG00000168754 | 2 | 97531620 | 97694175 | 290 | 37 | 31552 | -0.24903 | 0.59833 | *FAM178B* |
| ENSG00000196337 | 19 | 49547531 | 49572117 | 94 | 26 | 45592 | -0.24929 | 0.59843 | *CGB7* |
| ENSG00000163827 | 3 | 46546913 | 46631589 | 348 | 32 | 44110 | -0.24936 | 0.59846 | *LRRC2* |

| | | | | | | | | | |
|---|---|---|---|---|---|---|---|---|---|
| ENSG00000166596 | 17 | 9469944 | 9556776 | 139 | 33 | 44066 | -0.24969 | 0.59859 | *WDR16* |
| ENSG00000142751 | 1 | 27192624 | 27226788 | 61 | 10 | 53976 | -0.24992 | 0.59867 | *GPN2* |
| ENSG00000176407 | 2 | 85188216 | 85296595 | 221 | 27 | 40305 | -0.25006 | 0.59873 | *KCMF1* |
| ENSG00000005238 | 9 | 35094109 | 35126338 | 74 | 18 | 40026 | -0.25009 | 0.59874 | *FAM214B* |
| ENSG00000116521 | 1 | 155215770 | 155242221 | 52 | 5 | 28756 | -0.25011 | 0.59875 | *SCAMP3* |
| ENSG00000141448 | 18 | 19739404 | 19792491 | 64 | 22 | 48621 | -0.25034 | 0.59884 | *GATA6* |
| ENSG00000164342 | 4 | 186980306 | 187019223 | 165 | 32 | 45783 | -0.25046 | 0.59888 | *TLR3* |
| ENSG00000174780 | 4 | 57323081 | 57379839 | 219 | 21 | 40463 | -0.2505 | 0.5989 | *SRP72* |
| ENSG00000228144 | 12 | 66507697 | 66573765 | 161 | 22 | 38143 | -0.25049 | 0.5989 | *TMBIM4* |
| ENSG00000155957 | 12 | 66507709 | 66573852 | 161 | 22 | 38143 | -0.25049 | 0.5989 | *TMBIM4* |
| ENSG00000124920 | 11 | 61510114 | 61565990 | 106 | 26 | 34966 | -0.25074 | 0.59899 | *MYRF* |
| ENSG00000221953 | 1 | 247263462 | 247285719 | 95 | 19 | 42434 | -0.25078 | 0.59901 | *C1orf229* |
| ENSG00000147488 | 8 | 53013399 | 53383519 | 905 | 113 | 41545 | -0.25079 | 0.59901 | *ST18* |
| ENSG00000170430 | 10 | 131255448 | 131576271 | 1469 | 101 | 46429 | -0.25095 | 0.59907 | *MGMT* |
| ENSG00000184945 | 2 | 241621262 | 241647900 | 71 | 15 | 43199 | -0.25105 | 0.59911 | *AQP12A* |
| ENSG00000104983 | 19 | 46488339 | 46534576 | 188 | 29 | 48405 | -0.2511 | 0.59913 | *CCDC61* |
| ENSG00000188761 | 1 | 114410790 | 114440169 | 60 | 10 | 34751 | -0.2515 | 0.59929 | *BCL2L15* |
| ENSG00000171621 | 1 | 9342939 | 9439591 | 441 | 90 | 48189 | -0.25189 | 0.59944 | *SPSB1* |
| ENSG00000205846 | 12 | 8598522 | 8640926 | 189 | 6 | 47498 | -0.25204 | 0.5995 | *CLEC6A* |
| ENSG00000244752 | 22 | 25605489 | 25637836 | 143 | 37 | 42545 | -0.25211 | 0.59952 | *CRYBB2* |
| ENSG00000019102 | 11 | 124607368 | 124632134 | 50 | 13 | 41663 | -0.25228 | 0.59959 | *VSIG2* |
| ENSG00000150712 | 5 | 32217100 | 32323115 | 336 | 31 | 52724 | -0.25276 | 0.59977 | *MTMR12* |
| ENSG00000137941 | 1 | 84320711 | 84474833 | 500 | 28 | 44344 | -0.25283 | 0.5998 | *TTLL7* |
| ENSG00000062716 | 17 | 57774553 | 57929616 | 279 | 27 | 32285 | -0.25297 | 0.59985 | *VMP1* |
| ENSG00000239900 | 22 | 40732507 | 40796467 | 110 | 12 | 54197 | -0.25304 | 0.59988 | *ADSL* |
| ENSG00000185038 | 2 | 234674370 | 234752069 | 310 | 70 | 41766 | -0.25316 | 0.59993 | *MROH2A* |
| ENSG00000145919 | 5 | 173024517 | 173053663 | 97 | 12 | 34595 | -0.2532 | 0.59994 | *BOD1* |
| ENSG00000214860 | 17 | 18270976 | 18302961 | 78 | 8 | 42575 | -0.25329 | 0.59998 | *EVPLL* |
| ENSG00000166394 | 11 | 7676331 | 7708453 | 207 | 27 | 38330 | -0.25338 | 0.60001 | *CYB5R2* |
| ENSG00000139263 | 12 | 59255931 | 59324303 | 236 | 35 | 41151 | -0.2537 | 0.60014 | *LRIG3* |
| ENSG00000092529 | 15 | 42630301 | 42714516 | 249 | 17 | 32322 | -0.25382 | 0.60018 | *CAPN3* |
| ENSG00000184774 | 1 | 202779394 | 202806353 | 56 | 11 | 52968 | -0.25397 | 0.60024 | *RP11-480I12.4* |
| ENSG00000127533 | 19 | 16989671 | 17013417 | 66 | 17 | 46824 | -0.25443 | 0.60042 | *F2RL3* |
| ENSG00000197360 | 19 | 22563821 | 22725287 | 602 | 66 | 45183 | -0.25455 | 0.60047 | *ZNF98* |
| ENSG00000148229 | 9 | 116159515 | 116182952 | 61 | 10 | 56220 | -0.25461 | 0.60049 | *POLE3* |
| ENSG00000135925 | 2 | 219735085 | 219774303 | 39 | 11 | 36731 | -0.25467 | 0.60051 | *WNT10A* |

| | | | | | | | | | |
|---|---|---|---|---|---|---|---|---|---|
| ENSG00000180828 | 8 | 65482814 | 65506181 | 41 | 11 | 44308 | -0.25465 | 0.60051 | *BHLHE22* |
| ENSG00000145936 | 5 | 169791675 | 169826681 | 150 | 32 | 44307 | -0.25468 | 0.60052 | *KCNMB1* |
| ENSG00000179387 | 4 | 141435312 | 141484924 | 103 | 13 | 44222 | -0.25479 | 0.60056 | *ELMOD2* |
| ENSG00000106591 | 7 | 42961799 | 42998557 | 119 | 13 | 53843 | -0.25485 | 0.60058 | *MRPL32* |
| ENSG00000160951 | 19 | 14573278 | 14596174 | 117 | 25 | 38862 | -0.25488 | 0.60059 | *PTGER1* |
| ENSG00000130723 | 9 | 134259480 | 134385584 | 274 | 26 | 38135 | -0.2549 | 0.6006 | *PRRC2B* |
| ENSG00000087502 | 12 | 29480285 | 29544122 | 159 | 21 | 45134 | -0.25521 | 0.60072 | *ERGIC2* |
| ENSG00000121741 | 13 | 20522810 | 20675968 | 318 | 24 | 34069 | -0.25581 | 0.60095 | *ZMYM2* |
| ENSG00000256043 | 4 | 156835270 | 156885069 | 216 | 39 | 44023 | -0.25584 | 0.60096 | *CTSO* |
| ENSG00000156050 | 14 | 74388204 | 74427117 | 152 | 17 | 39575 | -0.25594 | 0.601 | *FAM161B* |
| ENSG00000044524 | 3 | 89146674 | 89541284 | 842 | 64 | 39809 | -0.25628 | 0.60113 | *EPHA3* |
| ENSG00000204866 | 19 | 46636363 | 46674556 | 93 | 13 | 46154 | -0.25638 | 0.60117 | *IGFL2* |
| ENSG00000179919 | 12 | 55604809 | 55625759 | 47 | 8 | 49936 | -0.25642 | 0.60119 | *OR10A7* |
| ENSG00000173093 | 12 | 111274573 | 111355339 | 279 | 38 | 45980 | -0.25654 | 0.60123 | *CCDC63* |
| ENSG00000159840 | 7 | 143068173 | 143098204 | 83 | 29 | 43996 | -0.25658 | 0.60125 | *ZYX* |
| ENSG00000104823 | 19 | 39296062 | 39332645 | 103 | 20 | 40055 | -0.25666 | 0.60128 | *ECH1* |
| ENSG00000267127 | 18 | 77784346 | 77864968 | 365 | 30 | 40405 | -0.25711 | 0.60145 | *RP11-795F19.5* |
| ENSG00000197208 | 5 | 131620136 | 131689899 | 213 | 18 | 35726 | -0.25738 | 0.60156 | *SLC22A4* |
| ENSG00000119636 | 14 | 74476056 | 74559566 | 327 | 24 | 29286 | -0.25744 | 0.60158 | *CCDC176* |
| ENSG00000177791 | 10 | 75381412 | 75411515 | 58 | 12 | 38836 | -0.25755 | 0.60162 | *MYOZ1* |
| ENSG00000171310 | 12 | 104839073 | 105165792 | 1179 | 155 | 46691 | -0.25779 | 0.60172 | *CHST11* |
| ENSG00000039068 | 16 | 68761128 | 68879451 | 422 | 45 | 41852 | -0.25794 | 0.60177 | *CDH1* |
| ENSG00000183647 | 19 | 58101253 | 58134090 | 78 | 14 | 52147 | -0.25797 | 0.60179 | *ZNF530* |
| ENSG00000185155 | 1 | 226401319 | 226424755 | 53 | 10 | 53757 | -0.25811 | 0.60184 | *MIXL1* |
| ENSG00000173226 | 3 | 121478610 | 121563926 | 334 | 18 | 54553 | -0.25817 | 0.60186 | *IQCB1* |
| ENSG00000124279 | 5 | 7849272 | 7879150 | 97 | 23 | 43490 | -0.25819 | 0.60187 | *FASTKD3* |
| ENSG00000109084 | 17 | 26636121 | 26665707 | 40 | 12 | 43462 | -0.25826 | 0.6019 | *TMEM97* |
| ENSG00000152291 | 2 | 85535147 | 85565548 | 102 | 15 | 41248 | -0.25834 | 0.60193 | *TGOLN2* |
| ENSG00000104879 | 19 | 45799672 | 45836235 | 136 | 29 | 40102 | -0.25842 | 0.60196 | *CKM* |
| ENSG00000121410 | 19 | 58846544 | 58874865 | 59 | 9 | 25881 | -0.25856 | 0.60201 | *A1BG* |
| ENSG00000269781 | 7 | 6512922 | 6534873 | 71 | 13 | 42310 | -0.25857 | 0.60202 | *FLJ20306* |
| ENSG00000145242 | 4 | 66175281 | 66546213 | 1566 | 143 | 48784 | -0.25871 | 0.60207 | *EPHA5* |
| ENSG00000197016 | 19 | 57068880 | 57110279 | 171 | 21 | 46591 | -0.25917 | 0.60225 | *ZNF470* |
| ENSG00000099139 | 9 | 78495560 | 78987255 | 2005 | 257 | 48379 | -0.25932 | 0.60231 | *PCSK5* |
| ENSG00000100368 | 22 | 37299670 | 37346491 | 207 | 39 | 41556 | -0.25934 | 0.60231 | *CSF2RB* |
| ENSG00000086548 | 19 | 42244885 | 42286113 | 84 | 21 | 49694 | -0.25958 | 0.60241 | *CEACAM6* |

| | | | | | | | | | |
|---|---|---|---|---|---|---|---|---|---|
| ENSG00000091483 | 1 | 241650903 | 241693061 | 87 | 16 | 45582 | -0.25989 | 0.60252 | *FH* |
| ENSG00000164687 | 8 | 82182598 | 82207012 | 17 | 6 | 47006 | -0.25986 | 0.60252 | *FABP5* |
| ENSG00000156414 | 14 | 104384799 | 104529004 | 561 | 35 | 47891 | -0.25994 | 0.60254 | *TDRD9* |
| ENSG00000158560 | 7 | 95391866 | 95749634 | 1013 | 128 | 46226 | -0.26009 | 0.6026 | *DYNC1I1* |
| ENSG00000149311 | 11 | 108083211 | 108249829 | 289 | 19 | 50769 | -0.26058 | 0.60279 | *ATM* |
| ENSG00000116171 | 1 | 53382901 | 53527375 | 457 | 25 | 27467 | -0.26071 | 0.60284 | *SCP2* |
| ENSG00000254788 | 16 | 66576490 | 66623038 | 117 | 8 | 25598 | -0.26074 | 0.60285 | *CKLF-CMTM1* |
| ENSG00000155749 | 2 | 202142994 | 202232121 | 261 | 28 | 40870 | -0.26104 | 0.60297 | *ALS2CR12* |
| ENSG00000104518 | 8 | 144625377 | 144655232 | 151 | 24 | 52420 | -0.26121 | 0.60303 | *GSDMD* |
| ENSG00000214872 | 11 | 57298979 | 57328108 | 78 | 10 | 55702 | -0.26163 | 0.6032 | *SMTNL1* |
| ENSG00000169122 | 8 | 58897068 | 59126838 | 668 | 91 | 40764 | -0.26167 | 0.60321 | *FAM110B* |
| ENSG00000138311 | 10 | 64123951 | 64441771 | 1077 | 112 | 45899 | -0.2619 | 0.6033 | *ZNF365* |
| ENSG00000256087 | 19 | 52524632 | 52608991 | 317 | 26 | 39385 | -0.26229 | 0.60345 | *ZNF432* |
| ENSG00000106328 | 7 | 127221463 | 127252198 | 85 | 14 | 37294 | -0.26262 | 0.60358 | *FSCN3* |
| ENSG00000131873 | 15 | 101705928 | 101802137 | 389 | 40 | 35974 | -0.26278 | 0.60364 | *CHSY1* |
| ENSG00000189129 | 10 | 81881438 | 81915115 | 210 | 34 | 41557 | -0.263 | 0.60372 | *PLAC9* |
| ENSG00000149300 | 11 | 111778756 | 111807596 | 49 | 11 | 34614 | -0.26305 | 0.60374 | *C11orf52* |
| ENSG00000107796 | 10 | 90684831 | 90761147 | 247 | 38 | 42041 | -0.26344 | 0.60389 | *ACTA2* |
| ENSG00000109132 | 4 | 41736099 | 41760987 | 96 | 15 | 38157 | -0.26349 | 0.60391 | *PHOX2B* |
| ENSG00000138207 | 10 | 95341444 | 95371501 | 152 | 27 | 43581 | -0.26354 | 0.60393 | *RBP4* |
| ENSG00000069943 | 15 | 55601158 | 55657846 | 209 | 27 | 35680 | -0.26395 | 0.60409 | *PIGB* |
| ENSG00000172766 | 13 | 41875341 | 41961166 | 237 | 16 | 50417 | -0.26398 | 0.6041 | *NAA16* |
| ENSG00000128185 | 22 | 20291799 | 20317603 | 35 | 8 | 47985 | -0.26421 | 0.60419 | *DGCR6L* |
| ENSG00000176198 | 14 | 20700898 | 20721979 | 86 | 22 | 36307 | -0.26437 | 0.60425 | *OR11H4* |
| ENSG00000055211 | 6 | 149877430 | 149922884 | 60 | 11 | 32201 | -0.26452 | 0.60431 | *GINM1* |
| ENSG00000214819 | 17 | 20473037 | 20494224 | 18 | 11 | 42134 | -0.26461 | 0.60434 | *CDRT15L2* |
| ENSG00000136918 | 9 | 127605755 | 127630160 | 54 | 8 | 51010 | -0.26481 | 0.60442 | *WDR38* |
| ENSG00000179299 | 4 | 40741914 | 40822002 | 221 | 28 | 51098 | -0.26482 | 0.60443 | *NSUN7* |
| ENSG00000197134 | 19 | 22225254 | 22284282 | 336 | 19 | 39712 | -0.26494 | 0.60447 | *ZNF257* |
| ENSG00000117751 | 1 | 28147289 | 28188187 | 31 | 13 | 39102 | -0.26495 | 0.60448 | *PPP1R8* |
| ENSG00000086102 | 9 | 33280509 | 33381155 | 137 | 28 | 53620 | -0.26536 | 0.60463 | *NFX1* |
| ENSG00000260970 | 9 | 122247653 | 122296403 | 201 | 28 | 45898 | -0.26568 | 0.60476 | *RP11-295D22.1* |
| ENSG00000116586 | 1 | 156014543 | 156038301 | 31 | 15 | 42115 | -0.26605 | 0.6049 | *LAMTOR2* |
| ENSG00000184557 | 17 | 76342864 | 76366158 | 59 | 17 | 40807 | -0.26605 | 0.6049 | *SOCS3* |
| ENSG00000197183 | 20 | 31020862 | 31182876 | 295 | 40 | 32979 | -0.26619 | 0.60495 | *C20orf112* |
| ENSG00000239282 | 22 | 30671106 | 30695616 | 61 | 20 | 42985 | -0.26626 | 0.60498 | *GATSL3* |

| | | | | | | | | | |
|---|---|---|---|---|---|---|---|---|---|
| ENSG00000268863 | 16 | 30986808 | 31007533 | 31 | 7 | 30147 | -0.26635 | 0.60502 | *AC135048.1* |
| ENSG00000106648 | 7 | 151643464 | 151727019 | 300 | 47 | 45191 | -0.2664 | 0.60503 | *GALNTL5* |
| ENSG00000257065 | 6 | 144118229 | 144175754 | 164 | 24 | 54464 | -0.26642 | 0.60504 | *RP3-468K18.5* |
| ENSG00000204246 | 9 | 107288030 | 107309137 | 61 | 11 | 37109 | -0.26675 | 0.60517 | *OR13C3* |
| ENSG00000136379 | 15 | 80962025 | 81057962 | 344 | 51 | 41844 | -0.26693 | 0.60524 | *ABHD17C* |
| ENSG00000212952 | 9 | 44390717 | 44412427 | 1 | 1 | 42798 | -0.26709 | 0.6053 | *BX088651.1* |
| ENSG00000117009 | 1 | 241685434 | 241768944 | 278 | 54 | 41535 | -0.26754 | 0.60547 | *KMO* |
| ENSG00000140090 | 14 | 92778925 | 92972596 | 873 | 121 | 45539 | -0.26755 | 0.60548 | *SLC24A4* |
| ENSG00000134398 | 16 | 23691647 | 23734821 | 125 | 26 | 37015 | -0.26762 | 0.6055 | *ERN2* |
| ENSG00000131504 | 5 | 140884583 | 141008622 | 250 | 36 | 51506 | -0.26784 | 0.60559 | *DIAPH1* |
| ENSG00000141404 | 18 | 11678955 | 11895684 | 768 | 73 | 37384 | -0.26855 | 0.60586 | *GNAL* |
| ENSG00000167723 | 17 | 3403796 | 3471289 | 293 | 55 | 40052 | -0.26858 | 0.60588 | *TRPV3* |
| ENSG00000161640 | 19 | 50442242 | 50474429 | 101 | 21 | 43385 | -0.2687 | 0.60592 | *SIGLEC11* |
| ENSG00000151006 | 16 | 31084746 | 31110949 | 37 | 7 | 22694 | -0.26877 | 0.60595 | *PRSS53* |
| ENSG00000156384 | 10 | 105871816 | 105896143 | 59 | 5 | 25078 | -0.26887 | 0.60598 | *SFR1* |
| ENSG00000172794 | 17 | 72656717 | 72753474 | 188 | 53 | 43958 | -0.26897 | 0.60602 | *RAB37* |
| ENSG00000233701 | 3 | 138750944 | 138773734 | 57 | 8 | 28389 | -0.26916 | 0.6061 | *PRR23C* |
| ENSG00000134538 | 12 | 21274136 | 21402180 | 572 | 50 | 36203 | -0.26942 | 0.6062 | *SLCO1B1* |
| ENSG00000130165 | 19 | 11651962 | 11680051 | 65 | 16 | 37454 | -0.26957 | 0.60626 | *ELOF1* |
| ENSG00000171509 | 4 | 159226463 | 159584524 | 1133 | 32 | 32526 | -0.26964 | 0.60628 | *RXFP1* |
| ENSG00000134830 | 19 | 47825404 | 47856606 | 157 | 15 | 49433 | -0.26992 | 0.60639 | *C5AR2* |
| ENSG00000172530 | 16 | 87972850 | 88120924 | 766 | 49 | 30896 | -0.26994 | 0.6064 | *BANP* |
| ENSG00000165698 | 9 | 135743414 | 135775588 | 82 | 19 | 44289 | -0.27007 | 0.60645 | *C9orf9* |
| ENSG00000170525 | 10 | 6176881 | 6287495 | 515 | 136 | 45091 | -0.27007 | 0.60645 | *PFKFB3* |
| ENSG00000236637 | 9 | 21176617 | 21197598 | 133 | 6 | 53752 | -0.27013 | 0.60647 | *IFNA4* |
| ENSG00000112293 | 6 | 24414793 | 24505433 | 721 | 67 | 44358 | -0.27022 | 0.6065 | *GPLD1* |
| ENSG00000197057 | 4 | 36273244 | 36357378 | 328 | 56 | 44718 | -0.27035 | 0.60655 | *DTHD1* |
| ENSG00000186063 | 1 | 222831355 | 222896552 | 128 | 22 | 44085 | -0.27046 | 0.6066 | *AIDA* |
| ENSG00000222047 | 10 | 75659727 | 75692535 | 67 | 10 | 33274 | -0.27057 | 0.60664 | *C10orf55* |
| ENSG00000125409 | 17 | 15197128 | 15254958 | 198 | 23 | 44612 | -0.27085 | 0.60675 | *TEKT3* |
| ENSG00000260537 | 16 | 70323097 | 70410163 | 102 | 35 | 44670 | -0.27103 | 0.60682 | *RP11-529K1.3* |
| ENSG00000163923 | 3 | 186828736 | 186908696 | 223 | 43 | 44435 | -0.27106 | 0.60683 | *RPL39L* |
| ENSG00000267216 | 19 | 58780332 | 58836563 | 178 | 30 | 39538 | -0.27127 | 0.60691 | *AC010642.1* |
| ENSG00000133104 | 13 | 36865775 | 36954317 | 294 | 20 | 51631 | -0.27135 | 0.60694 | *SPG20* |
| ENSG00000158714 | 1 | 159786540 | 159817039 | 86 | 22 | 41948 | -0.27148 | 0.60699 | *SLAMF8* |
| ENSG00000135414 | 12 | 56127064 | 56160911 | 29 | 10 | 40438 | -0.27172 | 0.60708 | *GDF11* |

| | | | | | | | | | |
|---|---|---|---|---|---|---|---|---|---|
| ENSG00000165684 | 9 | 139260029 | 139303249 | 200 | 23 | 55075 | -0.27176 | 0.6071 | *SNAPC4* |
| ENSG00000197241 | 1 | 9053359 | 9096404 | 177 | 36 | 48712 | -0.27214 | 0.60724 | *SLC2A7* |
| ENSG00000139835 | 13 | 113968505 | 114028446 | 218 | 34 | 45479 | -0.27231 | 0.60731 | *GRTP1* |
| ENSG00000188992 | 21 | 15471134 | 15593166 | 569 | 47 | 46546 | -0.27257 | 0.60741 | *LIPI* |
| ENSG00000167371 | 16 | 29813177 | 29837201 | 32 | 6 | 47399 | -0.27269 | 0.60745 | *PRRT2* |
| ENSG00000267508 | 19 | 44876459 | 44915774 | 146 | 30 | 34325 | -0.27295 | 0.60755 | *ZNF285* |
| ENSG00000142319 | 5 | 1382909 | 1455545 | 240 | 42 | 41991 | -0.27298 | 0.60757 | *SLC6A3* |
| ENSG00000136010 | 12 | 105403568 | 105488355 | 383 | 44 | 43051 | -0.27324 | 0.60767 | *ALDH1L2* |
| ENSG00000100348 | 22 | 36853083 | 36888077 | 172 | 25 | 33183 | -0.27326 | 0.60767 | *TXN2* |
| ENSG00000158481 | 1 | 158249576 | 158273420 | 30 | 10 | 38198 | -0.27349 | 0.60776 | *CD1C* |
| ENSG00000247596 | 3 | 52252626 | 52283276 | 54 | 14 | 52035 | -0.27361 | 0.60781 | *TWF2* |
| ENSG00000205269 | 6 | 11527938 | 11593757 | 253 | 27 | 50629 | -0.27363 | 0.60782 | *TMEM170B* |
| ENSG00000100104 | 22 | 26869843 | 26900624 | 111 | 22 | 43063 | -0.27365 | 0.60782 | *SRRD* |
| ENSG00000030110 | 6 | 33530329 | 33558019 | 109 | 21 | 39843 | -0.27368 | 0.60783 | *BAK1* |
| ENSG00000174326 | 17 | 6934949 | 6957242 | 62 | 16 | 35348 | -0.27383 | 0.60789 | *SLC16A11* |
| ENSG00000019582 | 5 | 149771200 | 149802492 | 91 | 30 | 49602 | -0.27399 | 0.60795 | *CD74* |
| ENSG00000109794 | 4 | 187015573 | 187103821 | 396 | 55 | 46334 | -0.27415 | 0.60801 | *FAM149A* |
| ENSG00000173267 | 10 | 88708375 | 88733017 | 99 | 17 | 49761 | -0.27421 | 0.60804 | *SNCG* |
| ENSG00000165637 | 10 | 76959912 | 77001206 | 112 | 22 | 39031 | -0.27424 | 0.60805 | *VDAC2* |
| ENSG00000161249 | 19 | 35978122 | 36014560 | 133 | 34 | 46142 | -0.27474 | 0.60824 | *DMKN* |
| ENSG00000101405 | 20 | 3042266 | 3063163 | 47 | 14 | 39838 | -0.27494 | 0.60832 | *OXT* |
| ENSG00000130558 | 9 | 137957268 | 138023025 | 365 | 65 | 44670 | -0.27511 | 0.60838 | *OLFM1* |
| ENSG00000148136 | 9 | 107278534 | 107299490 | 82 | 13 | 39973 | -0.27525 | 0.60844 | *OR13C4* |
| ENSG00000204403 | 11 | 104746445 | 104779397 | 42 | 5 | 32944 | -0.27529 | 0.60845 | *CASP12* |
| ENSG00000185652 | 12 | 5531278 | 5640702 | 322 | 68 | 51086 | -0.27549 | 0.60853 | *NTF3* |
| ENSG00000239605 | 2 | 47262677 | 47392517 | 609 | 90 | 45178 | -0.27664 | 0.60897 | *C2orf61* |
| ENSG00000139182 | 12 | 7272294 | 7321541 | 129 | 22 | 44474 | -0.27665 | 0.60898 | *CLSTN3* |
| ENSG00000185043 | 15 | 90763207 | 90787279 | 87 | 15 | 42193 | -0.27672 | 0.609 | *CIB1* |
| ENSG00000110107 | 11 | 60648202 | 60684060 | 73 | 12 | 43090 | -0.27687 | 0.60906 | *PRPF19* |
| ENSG00000121753 | 1 | 32182718 | 32240494 | 58 | 15 | 44278 | -0.27703 | 0.60912 | *BAI2* |
| ENSG00000100297 | 22 | 35786056 | 35831423 | 127 | 26 | 42859 | -0.27745 | 0.60928 | *MCM5* |
| ENSG00000123485 | 2 | 234732062 | 234773212 | 213 | 21 | 45238 | -0.27766 | 0.60936 | *HJURP* |
| ENSG00000179593 | 17 | 7932335 | 7962452 | 133 | 28 | 43106 | -0.27776 | 0.6094 | *ALOX15B* |
| ENSG00000164619 | 7 | 33934523 | 34205484 | 669 | 96 | 48671 | -0.27778 | 0.60941 | *BMPER* |
| ENSG00000112773 | 6 | 82191156 | 82472491 | 906 | 84 | 50791 | -0.2779 | 0.60945 | *FAM46A* |
| ENSG00000023839 | 10 | 101532489 | 101621949 | 270 | 29 | 52779 | -0.27806 | 0.60952 | *ABCC2* |

| | | | | | | | | | |
|---|---|---|---|---|---|---|---|---|---|
| ENSG00000168938 | 5 | 122348945 | 122382436 | 135 | 25 | 39082 | -0.27848 | 0.60968 | *PPIC* |
| ENSG00000116996 | 1 | 238035705 | 238064094 | 115 | 21 | 47784 | -0.27864 | 0.60974 | *ZP4* |
| ENSG00000166377 | 18 | 76819285 | 77148283 | 1183 | 60 | 42592 | -0.27866 | 0.60975 | *ATP9B* |
| ENSG00000164535 | 7 | 6438757 | 6533821 | 360 | 49 | 39852 | -0.27911 | 0.60992 | *DAGLB* |
| ENSG00000019144 | 11 | 118467155 | 118538741 | 128 | 22 | 50800 | -0.27925 | 0.60997 | *PHLDB1* |
| ENSG00000111796 | 12 | 9737147 | 9770482 | 164 | 23 | 44687 | -0.27945 | 0.61005 | *KLRB1* |
| ENSG00000148840 | 10 | 103882787 | 103920082 | 66 | 15 | 41200 | -0.27948 | 0.61006 | *PPRC1* |
| ENSG00000181374 | 17 | 32673471 | 32695629 | 84 | 18 | 50860 | -0.27949 | 0.61007 | *CCL13* |
| ENSG00000103356 | 16 | 23523335 | 23579052 | 207 | 18 | 43056 | -0.27963 | 0.61012 | *EARS2* |
| ENSG00000198965 | 1 | 158439668 | 158460675 | 93 | 10 | 47758 | -0.28005 | 0.61028 | *OR10R2* |
| ENSG00000263588 | 18 | 19658021 | 19688741 | 121 | 22 | 41720 | -0.2801 | 0.6103 | *RP11-595B24.2* |
| ENSG00000021300 | 11 | 73347223 | 73383864 | 140 | 26 | 40543 | -0.28018 | 0.61033 | *PLEKHB1* |
| ENSG00000108518 | 17 | 4838947 | 4862356 | 50 | 16 | 39931 | -0.28087 | 0.6106 | *PFN1* |
| ENSG00000214900 | 14 | 50438430 | 50484238 | 213 | 31 | 46515 | -0.28116 | 0.6107 | *C14orf182* |
| ENSG00000105122 | 19 | 15552435 | 15585382 | 57 | 17 | 53267 | -0.28116 | 0.61071 | *RASAL3* |
| ENSG00000168907 | 15 | 42423332 | 42458839 | 86 | 28 | 34756 | -0.28132 | 0.61077 | *PLA2G4F* |
| ENSG00000108848 | 17 | 48786905 | 48843574 | 135 | 26 | 50941 | -0.28135 | 0.61078 | *LUC7L3* |
| ENSG00000136932 | 9 | 100656771 | 100694852 | 106 | 11 | 37063 | -0.28141 | 0.6108 | *C9orf156* |
| ENSG00000167395 | 16 | 31075743 | 31105517 | 47 | 7 | 22524 | -0.28143 | 0.61081 | *ZNF646* |
| ENSG00000115425 | 2 | 216851052 | 216957678 | 354 | 55 | 40906 | -0.28217 | 0.61109 | *PECR* |
| ENSG00000273171 | 17 | 26681310 | 26705033 | 50 | 7 | 32540 | -0.28275 | 0.61132 | *CTB-96E2.2* |
| ENSG00000171970 | 19 | 2890896 | 2928474 | 249 | 23 | 29878 | -0.28294 | 0.61139 | *ZNF57* |
| ENSG00000184956 | 11 | 1002821 | 1046706 | 124 | 24 | 43331 | -0.28312 | 0.61146 | *MUC6* |
| ENSG00000111364 | 12 | 124076624 | 124115488 | 149 | 16 | 41545 | -0.28326 | 0.61151 | *DDX55* |
| ENSG00000117215 | 1 | 20428432 | 20456050 | 79 | 30 | 46609 | -0.28339 | 0.61156 | *PLA2G2D* |
| ENSG00000177596 | 13 | 74977094 | 75003252 | 136 | 24 | 37915 | -0.28355 | 0.61162 | *AL355390.1* |
| ENSG00000001631 | 7 | 91818283 | 91885480 | 78 | 17 | 41687 | -0.28368 | 0.61167 | *KRIT1* |
| ENSG00000204571 | 11 | 71282901 | 71324399 | 228 | 26 | 38900 | -0.2837 | 0.61168 | *KRTAP5-11* |
| ENSG00000159714 | 16 | 67418322 | 67460736 | 55 | 15 | 43040 | -0.28371 | 0.61168 | *ZDHHC1* |
| ENSG00000173065 | 17 | 27072996 | 27192250 | 226 | 13 | 47123 | -0.28372 | 0.61169 | *FAM222B* |
| ENSG00000142405 | 19 | 54286857 | 54337648 | 245 | 46 | 48163 | -0.28388 | 0.61175 | *NLRP12* |
| ENSG00000138382 | 2 | 170656591 | 170691441 | 128 | 22 | 46897 | -0.28482 | 0.61211 | *METTL5* |
| ENSG00000173567 | 2 | 26521041 | 26579685 | 157 | 33 | 34467 | -0.28514 | 0.61223 | *GPR113* |
| ENSG00000175130 | 1 | 32789433 | 32811980 | 11 | 8 | 56229 | -0.28519 | 0.61225 | *MARCKSL1* |
| ENSG00000148429 | 10 | 11485945 | 11663753 | 510 | 45 | 46725 | -0.28528 | 0.61229 | *USP6NL* |
| ENSG00000152942 | 5 | 68655120 | 68720628 | 133 | 14 | 41464 | -0.2855 | 0.61237 | *RAD17* |

| | | | | | | | | | |
|---|---|---|---|---|---|---|---|---|---|
| ENSG00000006282 | 17 | 48610419 | 48643213 | 108 | 18 | 31912 | -0.28589 | 0.61252 | *SPATA20* |
| ENSG00000099308 | 19 | 18198603 | 18272502 | 293 | 35 | 52227 | -0.28591 | 0.61252 | *MAST3* |
| ENSG00000076248 | 12 | 109525379 | 109558797 | 92 | 23 | 47432 | -0.28655 | 0.61277 | *UNG* |
| ENSG00000249961 | 16 | 66778879 | 66845523 | 145 | 12 | 33635 | -0.28675 | 0.61285 | *CCDC79* |
| ENSG00000185662 | 5 | 171202876 | 171231602 | 94 | 19 | 47590 | -0.28682 | 0.61288 | *SMIM23* |
| ENSG00000107104 | 9 | 460291 | 756105 | 1577 | 98 | 41120 | -0.28686 | 0.61289 | *KANK1* |
| ENSG00000112282 | 6 | 131885106 | 131959369 | 111 | 25 | 38825 | -0.28709 | 0.61298 | *MED23* |
| ENSG00000187037 | 7 | 37713399 | 37883390 | 565 | 66 | 45638 | -0.28709 | 0.61298 | *GPR141* |
| ENSG00000067191 | 17 | 37319709 | 37363956 | 93 | 30 | 46336 | -0.28712 | 0.61299 | *CACNB1* |
| ENSG00000231171 | 4 | 178639911 | 178921904 | 1034 | 109 | 42458 | -0.28725 | 0.61304 | *LINC01098* |
| ENSG00000163424 | 3 | 118854997 | 118888889 | 175 | 24 | 52108 | -0.28732 | 0.61306 | *C3orf30* |
| ENSG00000179886 | 8 | 144670074 | 144692485 | 91 | 13 | 35932 | -0.28746 | 0.61312 | *TIGD5* |
| ENSG00000119720 | 14 | 90732580 | 90808481 | 209 | 25 | 48788 | -0.28753 | 0.61315 | *NRDE2* |
| ENSG00000131196 | 18 | 77145856 | 77299325 | 741 | 85 | 42007 | -0.2878 | 0.61325 | *NFATC1* |
| ENSG00000187479 | 11 | 43936892 | 43975888 | 149 | 37 | 47168 | -0.28791 | 0.61329 | *C11orf96* |
| ENSG00000251139 | 4 | 185724773 | 185752443 | 104 | 20 | 46944 | -0.28796 | 0.61331 | *RP11-701P16.2* |
| ENSG00000132207 | 16 | 30195208 | 30218882 | 6 | 4 | 32203 | -0.28805 | 0.61335 | *SLX1A* |
| ENSG00000120318 | 5 | 141022968 | 141071788 | 178 | 47 | 51148 | -0.28834 | 0.61346 | *ARAP3* |
| ENSG00000132958 | 13 | 19987017 | 20120903 | 327 | 28 | 35804 | -0.28841 | 0.61348 | *TPTE2* |
| ENSG00000118307 | 12 | 25251354 | 25358096 | 403 | 34 | 43582 | -0.28857 | 0.61355 | *CASC1* |
| ENSG00000164007 | 1 | 43188764 | 43215925 | 72 | 15 | 55108 | -0.28868 | 0.61359 | *CLDN19* |
| ENSG00000168118 | 1 | 229396822 | 229451641 | 130 | 16 | 56288 | -0.28877 | 0.61362 | *RAB4A* |
| ENSG00000130176 | 19 | 11639532 | 11671138 | 72 | 16 | 41369 | -0.28876 | 0.61362 | *CNN1* |
| ENSG00000185633 | 12 | 57618686 | 57644498 | 30 | 12 | 42911 | -0.28885 | 0.61365 | *NDUFA4L2* |
| ENSG00000160392 | 19 | 40815443 | 40864434 | 135 | 19 | 38015 | -0.28899 | 0.61371 | *C19orf47* |
| ENSG00000104859 | 19 | 45532298 | 45584214 | 94 | 21 | 50627 | -0.28906 | 0.61373 | *CLASRP* |
| ENSG00000142156 | 21 | 47391651 | 47434964 | 192 | 30 | 44877 | -0.28935 | 0.61384 | *COL6A1* |
| ENSG00000206104 | 21 | 32005183 | 32025455 | 41 | 8 | 51988 | -0.28953 | 0.61391 | *KRTAP20-3* |
| ENSG00000176208 | 17 | 29148988 | 29232887 | 214 | 9 | 28088 | -0.28964 | 0.61396 | *ATAD5* |
| ENSG00000071073 | 2 | 99225569 | 99357589 | 302 | 39 | 40602 | -0.28976 | 0.614 | *MGAT4A* |
| ENSG00000163697 | 4 | 40802044 | 41228731 | 1721 | 145 | 44639 | -0.28998 | 0.61408 | *APBB2* |
| ENSG00000166368 | 11 | 6902721 | 6923830 | 75 | 16 | 43230 | -0.29021 | 0.61417 | *OR2D2* |
| ENSG00000137976 | 1 | 84854215 | 84890701 | 160 | 19 | 55824 | -0.29026 | 0.61419 | *DNASE2B* |
| ENSG00000169740 | 10 | 44129307 | 44154304 | 83 | 11 | 41561 | -0.29118 | 0.61454 | *ZNF32* |
| ENSG00000132768 | 1 | 44425672 | 44449041 | 37 | 8 | 28633 | -0.29132 | 0.6146 | *DPH2* |
| ENSG00000179841 | 14 | 64922217 | 64951221 | 76 | 11 | 55757 | -0.29133 | 0.6146 | *AKAP5* |

| | | | | | | | | | |
|---|---|---|---|---|---|---|---|---|---|
| ENSG00000220256 | 2 | 240674554 | 240734577 | 350 | 44 | 51528 | -0.2914 | 0.61463 | *AC093802.1* |
| ENSG00000143162 | 1 | 167488914 | 167533004 | 228 | 32 | 50977 | -0.29145 | 0.61465 | *CREG1* |
| ENSG00000231396 | 4 | 9202383 | 9223975 | 3 | 2 | 42709 | -0.29163 | 0.61472 | *USP17L10* |
| ENSG00000232399 | 4 | 9202383 | 9238214 | 3 | 2 | 42709 | -0.29163 | 0.61472 | *USP17L13* |
| ENSG00000233136 | 4 | 9207131 | 9228723 | 3 | 2 | 42709 | -0.29163 | 0.61472 | *USP17L11* |
| ENSG00000227551 | 4 | 9211878 | 9233470 | 3 | 2 | 42709 | -0.29163 | 0.61472 | *USP17L12* |
| ENSG00000132514 | 17 | 6967856 | 6993626 | 109 | 27 | 34564 | -0.29163 | 0.61472 | *CLEC10A* |
| ENSG00000132432 | 7 | 54809943 | 54837667 | 136 | 15 | 43623 | -0.29169 | 0.61474 | *SEC61G* |
| ENSG00000176399 | 9 | 22436840 | 22465739 | 99 | 15 | 50768 | -0.29177 | 0.61477 | *DMRTA1* |
| ENSG00000198513 | 14 | 50989227 | 51109786 | 263 | 30 | 47629 | -0.29204 | 0.61487 | *ATL1* |
| ENSG00000108349 | 17 | 38286571 | 38338436 | 96 | 14 | 39523 | -0.2922 | 0.61493 | *CASC3* |
| ENSG00000100226 | 22 | 39091728 | 39144304 | 118 | 16 | 42993 | -0.29225 | 0.61495 | *GTPBP1* |
| ENSG00000155530 | 7 | 133802052 | 133959343 | 506 | 24 | 42906 | -0.29267 | 0.61511 | *LRGUK* |
| ENSG00000174529 | 1 | 205042258 | 205063645 | 68 | 13 | 27276 | -0.29279 | 0.61516 | *TMEM81* |
| ENSG00000100380 | 22 | 41210539 | 41263026 | 93 | 20 | 42629 | -0.29295 | 0.61522 | *ST13* |
| ENSG00000244038 | 1 | 20968270 | 20998000 | 127 | 18 | 54626 | -0.29297 | 0.61523 | *DDOST* |
| ENSG00000140395 | 15 | 78560177 | 78602136 | 131 | 23 | 54683 | -0.29342 | 0.6154 | *WDR61* |
| ENSG00000002933 | 7 | 150487491 | 150512208 | 116 | 23 | 45045 | -0.29375 | 0.61552 | *TMEM176A* |
| ENSG00000136925 | 9 | 100352362 | 100405962 | 80 | 20 | 48141 | -0.29403 | 0.61563 | *TSTD2* |
| ENSG00000134901 | 13 | 103426631 | 103461357 | 96 | 13 | 44505 | -0.29413 | 0.61567 | *KDELC1* |
| ENSG00000176194 | 18 | 12244318 | 12287594 | 158 | 37 | 46429 | -0.29442 | 0.61578 | *CIDEA* |
| ENSG00000167703 | 17 | 1462561 | 1542180 | 271 | 67 | 47635 | -0.29458 | 0.61584 | *SLC43A2* |
| ENSG00000133103 | 13 | 40219764 | 40375802 | 526 | 31 | 55860 | -0.29473 | 0.6159 | *COG6* |
| ENSG00000213918 | 16 | 3651729 | 3740144 | 253 | 26 | 47899 | -0.29482 | 0.61594 | *DNASE1* |
| ENSG00000118620 | 19 | 21193426 | 21252856 | 164 | 15 | 35635 | -0.29501 | 0.61601 | *ZNF430* |
| ENSG00000185928 | 16 | 29817285 | 29851948 | 49 | 6 | 47241 | -0.29534 | 0.61613 | *PAGR1* |
| ENSG00000151023 | 10 | 25260908 | 25315089 | 148 | 20 | 36535 | -0.29548 | 0.61619 | *ENKUR* |
| ENSG00000180720 | 11 | 46396640 | 46418107 | 17 | 9 | 45340 | -0.29585 | 0.61633 | *CHRM4* |
| ENSG00000183160 | 12 | 108973622 | 109002096 | 83 | 19 | 44281 | -0.29616 | 0.61645 | *TMEM119* |
| ENSG00000090612 | 12 | 133697570 | 133793698 | 324 | 19 | 46736 | -0.29628 | 0.61649 | *ZNF268* |
| ENSG00000148110 | 9 | 97126833 | 97233324 | 399 | 28 | 48377 | -0.29629 | 0.6165 | *HIATL1* |
| ENSG00000125084 | 12 | 49362398 | 49385459 | 27 | 10 | 49932 | -0.29636 | 0.61652 | *WNT1* |
| ENSG00000125826 | 20 | 378142 | 421610 | 142 | 34 | 43803 | -0.29657 | 0.6166 | *RBCK1* |
| ENSG00000143125 | 1 | 110983822 | 111009976 | 90 | 25 | 45628 | -0.29667 | 0.61664 | *PROK1* |
| ENSG00000171817 | 19 | 38032308 | 38114998 | 186 | 20 | 35723 | -0.29676 | 0.61668 | *ZNF540* |
| ENSG00000119487 | 9 | 128189672 | 128479513 | 587 | 55 | 43839 | -0.29695 | 0.61675 | *MAPKAP1* |

| | | | | | | | | | |
|---|---|---|---|---|---|---|---|---|---|
| ENSG00000006695 | 17 | 13962813 | 14121994 | 521 | 66 | 51336 | -0.29741 | 0.61692 | *COX10* |
| ENSG00000113384 | 5 | 32114810 | 32184456 | 190 | 27 | 45111 | -0.29753 | 0.61697 | *GOLPH3* |
| ENSG00000196550 | 1 | 206126916 | 206165151 | 17 | 8 | 50320 | -0.29779 | 0.61707 | *FAM72A* |
| ENSG00000141458 | 18 | 21076148 | 21176862 | 207 | 23 | 39948 | -0.29779 | 0.61707 | *NPC1* |
| ENSG00000268519 | 12 | 77939297 | 77960568 | 161 | 13 | 52971 | -0.29785 | 0.61709 | *AC073528.1* |
| ENSG00000163624 | 4 | 85494132 | 85582491 | 165 | 23 | 48083 | -0.29795 | 0.61713 | *CDS1* |
| ENSG00000239665 | 10 | 13642047 | 13720299 | 322 | 33 | 47945 | -0.29816 | 0.61721 | *RP11-295P9.3* |
| ENSG00000135108 | 12 | 117571146 | 117638336 | 267 | 16 | 48106 | -0.29864 | 0.61739 | *FBXO21* |
| ENSG00000077312 | 19 | 41246542 | 41281294 | 112 | 21 | 47801 | -0.29864 | 0.61739 | *SNRPA* |
| ENSG00000204479 | 1 | 13706092 | 13729089 | 8 | 6 | 21926 | -0.29867 | 0.6174 | *PRAMEF17* |
| ENSG00000134193 | 1 | 120326641 | 120364283 | 123 | 9 | 52723 | -0.2987 | 0.61741 | *REG4* |
| ENSG00000105928 | 7 | 24727972 | 24819244 | 374 | 40 | 45081 | -0.29873 | 0.61743 | *DFNA5* |
| ENSG00000222007 | 2 | 236672499 | 236702031 | 144 | 5 | 53216 | -0.29919 | 0.6176 | *AC064874.1* |
| ENSG00000133597 | 7 | 140362953 | 140406061 | 94 | 11 | 28201 | -0.29927 | 0.61763 | *ADCK2* |
| ENSG00000163644 | 4 | 89168772 | 89215921 | 102 | 21 | 40293 | -0.29961 | 0.61776 | *PPM1K* |
| ENSG00000118997 | 2 | 196592427 | 196943536 | 806 | 65 | 44026 | -0.29977 | 0.61782 | *DNAH7* |
| ENSG00000169862 | 5 | 10961952 | 11914155 | 2809 | 306 | 46525 | -0.29986 | 0.61786 | *CTNND2* |
| ENSG00000103415 | 16 | 4514691 | 4570348 | 183 | 15 | 31012 | -0.29998 | 0.6179 | *HMOX2* |
| ENSG00000235608 | 4 | 1386720 | 1410119 | 72 | 15 | 40223 | -0.30005 | 0.61793 | *NKX1-1* |
| ENSG00000165895 | 11 | 100548384 | 100872668 | 1552 | 109 | 50273 | -0.30004 | 0.61793 | *ARHGAP42* |
| ENSG00000204174 | 10 | 47073534 | 47098320 | 13 | 4 | 43227 | -0.3006 | 0.61814 | *NPY4R* |
| ENSG00000108826 | 17 | 48435218 | 48460575 | 78 | 10 | 45832 | -0.30067 | 0.61817 | *MRPL27* |
| ENSG00000120451 | 11 | 130735331 | 130796404 | 326 | 22 | 50844 | -0.30102 | 0.6183 | *SNX19* |
| ENSG00000112763 | 6 | 26448150 | 26486849 | 130 | 20 | 39491 | -0.30119 | 0.61836 | *BTN2A1* |
| ENSG00000125691 | 17 | 36994118 | 37020096 | 94 | 20 | 45362 | -0.30163 | 0.61853 | *RPL23* |
| ENSG00000134815 | 19 | 47842538 | 47895961 | 228 | 23 | 43978 | -0.30208 | 0.6187 | *DHX34* |
| ENSG00000233822 | 6 | 27796323 | 27833487 | 68 | 15 | 36006 | -0.30208 | 0.61871 | *HIST1H2BN* |
| ENSG00000073282 | 3 | 189339205 | 189625068 | 1162 | 178 | 49769 | -0.3023 | 0.61879 | *TP63* |
| ENSG00000176171 | 10 | 133771578 | 133805435 | 148 | 14 | 53577 | -0.30245 | 0.61885 | *BNIP3* |
| ENSG00000164258 | 5 | 52846463 | 52989168 | 503 | 38 | 46812 | -0.30292 | 0.61902 | *NDUFS4* |
| ENSG00000067182 | 12 | 6427923 | 6461280 | 70 | 19 | 39994 | -0.30295 | 0.61904 | *TNFRSF1A* |
| ENSG00000141542 | 17 | 80602849 | 80666604 | 280 | 56 | 47319 | -0.30298 | 0.61905 | *RAB40B* |
| ENSG00000100644 | 14 | 62152231 | 62224976 | 171 | 25 | 32320 | -0.30305 | 0.61908 | *HIF1A* |
| ENSG00000188352 | 9 | 20648308 | 21005954 | 1123 | 70 | 54108 | -0.30354 | 0.61926 | *FOCAD* |
| ENSG00000167195 | 15 | 75540940 | 75572540 | 7 | 5 | 47121 | -0.30358 | 0.61928 | *GOLGA6C* |
| ENSG00000170667 | 7 | 102112892 | 102168228 | 34 | 10 | 38510 | -0.30368 | 0.61931 | *RASA4B* |

| | | | | | | | | | |
|---|---|---|---|---|---|---|---|---|---|
| ENSG00000182903 | 4 | 409604 | 502945 | 329 | 30 | 45794 | -0.30396 | 0.61942 | *ZNF721* |
| ENSG00000184162 | 19 | 19302225 | 19324233 | 42 | 13 | 49308 | -0.30399 | 0.61943 | *NR2C2AP* |
| ENSG00000256825 | 12 | 133697570 | 133788955 | 303 | 18 | 47316 | -0.30414 | 0.61949 | *CTD-2140B24.4* |
| ENSG00000165072 | 9 | 72648497 | 72851886 | 719 | 39 | 49449 | -0.30421 | 0.61952 | *MAMDC2* |
| ENSG00000144130 | 2 | 113469063 | 113510456 | 110 | 18 | 40387 | -0.30429 | 0.61955 | *NT5DC4* |
| ENSG00000224474 | 10 | 99169228 | 99195831 | 70 | 18 | 46613 | -0.30429 | 0.61955 | *AL355490.1* |
| ENSG00000163110 | 4 | 95363037 | 95599377 | 650 | 63 | 35385 | -0.30445 | 0.61961 | *PDLIM5* |
| ENSG00000130725 | 19 | 59057079 | 59080327 | 57 | 14 | 36415 | -0.30445 | 0.61961 | *UBE2M* |
| ENSG00000227057 | 6 | 33236885 | 33267304 | 98 | 18 | 32505 | -0.30448 | 0.61962 | *WDR46* |
| ENSG00000267477 | 19 | 11567055 | 11626513 | 200 | 26 | 33319 | -0.30476 | 0.61972 | *CTC-398G3.6* |
| ENSG00000115138 | 2 | 25373722 | 25401772 | 61 | 16 | 36799 | -0.30492 | 0.61979 | *POMC* |
| ENSG00000040731 | 5 | 24477209 | 24655087 | 734 | 62 | 42443 | -0.30512 | 0.61986 | *CDH10* |
| ENSG00000176239 | 11 | 5362738 | 5383676 | 254 | 14 | 46164 | -0.30554 | 0.62002 | *OR51B6* |
| ENSG00000153006 | 5 | 64003971 | 64074512 | 123 | 17 | 37383 | -0.30566 | 0.62007 | *SREK1IP1* |
| ENSG00000129596 | 5 | 115130430 | 115162651 | 56 | 20 | 49479 | -0.30591 | 0.62016 | *CDO1* |
| ENSG00000125872 | 20 | 6011424 | 6044695 | 103 | 18 | 50950 | -0.30609 | 0.62023 | *LRRN4* |
| ENSG00000091409 | 2 | 173282082 | 173381181 | 393 | 48 | 46312 | -0.30626 | 0.6203 | *ITGA6* |
| ENSG00000131148 | 16 | 85795364 | 85843214 | 118 | 18 | 34013 | -0.30644 | 0.62037 | *EMC8* |
| ENSG00000145642 | 5 | 63976135 | 64023917 | 75 | 12 | 36930 | -0.30653 | 0.6204 | *FAM159B* |
| ENSG00000137261 | 6 | 24534332 | 24656383 | 476 | 65 | 44739 | -0.30655 | 0.62041 | *KIAA0319* |
| ENSG00000118363 | 11 | 74650292 | 74700076 | 107 | 18 | 46131 | -0.30656 | 0.62041 | *SPCS2* |
| ENSG00000174429 | 8 | 107761711 | 107792473 | 90 | 19 | 48690 | -0.30672 | 0.62047 | *ABRA* |
| ENSG00000011114 | 14 | 93693896 | 93809438 | 378 | 32 | 38142 | -0.30699 | 0.62057 | *BTBD7* |
| ENSG00000117758 | 1 | 28089694 | 28160963 | 45 | 18 | 38766 | -0.30702 | 0.62059 | *STX12* |
| ENSG00000163661 | 3 | 157144578 | 157171417 | 72 | 12 | 40843 | -0.30711 | 0.62062 | *PTX3* |
| ENSG00000239732 | 3 | 52245096 | 52283183 | 58 | 15 | 52373 | -0.30753 | 0.62078 | *TLR9* |
| ENSG00000136560 | 2 | 161983419 | 162102732 | 169 | 24 | 50319 | -0.30805 | 0.62098 | *TANK* |
| ENSG00000197826 | 4 | 81246874 | 81894910 | 1669 | 90 | 33958 | -0.30838 | 0.62111 | *C4orf22* |
| ENSG00000157181 | 1 | 186334890 | 186400510 | 150 | 25 | 54352 | -0.30851 | 0.62115 | *C1orf27* |
| ENSG00000166582 | 17 | 16235848 | 16266970 | 80 | 11 | 41186 | -0.30854 | 0.62116 | *CENPV* |
| ENSG00000181852 | 12 | 56588285 | 56625717 | 27 | 12 | 44205 | -0.30855 | 0.62117 | *RNF41* |
| ENSG00000120049 | 10 | 103575731 | 103613677 | 68 | 16 | 53541 | -0.30878 | 0.62125 | *KCNIP2* |
| ENSG00000221972 | 3 | 133636992 | 133658656 | 51 | 16 | 50775 | -0.30883 | 0.62127 | *C3orf36* |
| ENSG00000146399 | 6 | 132956123 | 132977142 | 124 | 16 | 54188 | -0.30895 | 0.62132 | *TAAR1* |
| ENSG00000001629 | 7 | 91865548 | 92040698 | 217 | 25 | 44489 | -0.30942 | 0.6215 | *ANKIB1* |
| ENSG00000111432 | 12 | 130637004 | 130660285 | 97 | 27 | 51649 | -0.30959 | 0.62157 | *FZD10* |

| | | | | | | | | | |
|---|---|---|---|---|---|---|---|---|---|
| ENSG00000182584 | 20 | 32244304 | 32266331 | 28 | 10 | 47802 | -0.30982 | 0.62165 | *ACTL10* |
| ENSG00000130313 | 19 | 17612438 | 17642097 | 96 | 22 | 40657 | -0.31002 | 0.62173 | *PGLS* |
| ENSG00000119614 | 14 | 74696175 | 74739441 | 180 | 24 | 24925 | -0.31012 | 0.62176 | *VSX2* |
| ENSG00000166352 | 11 | 36606051 | 36704823 | 403 | 21 | 43011 | -0.31085 | 0.62204 | *C11orf74* |
| ENSG00000135837 | 1 | 179913873 | 180094015 | 511 | 33 | 43762 | -0.31132 | 0.62222 | *CEP350* |
| ENSG00000104915 | 19 | 13244872 | 13271197 | 56 | 12 | 40295 | -0.3115 | 0.62229 | *STX10* |
| ENSG00000124557 | 6 | 26491449 | 26520650 | 73 | 12 | 49919 | -0.31167 | 0.62235 | *BTN1A1* |
| ENSG00000249481 | 6 | 44300397 | 44354904 | 147 | 23 | 44308 | -0.31186 | 0.62243 | *SPATS1* |
| ENSG00000177606 | 1 | 59236465 | 59259785 | 46 | 12 | 42852 | -0.31194 | 0.62246 | *JUN* |
| ENSG00000170340 | 2 | 62413248 | 62461866 | 105 | 17 | 41777 | -0.31194 | 0.62246 | *B3GNT2* |
| ENSG00000196230 | 6 | 30677978 | 30703203 | 143 | 17 | 27858 | -0.31199 | 0.62248 | *TUBB* |
| ENSG00000109738 | 4 | 157987209 | 158103242 | 487 | 49 | 43743 | -0.31207 | 0.62251 | *GLRB* |
| ENSG00000132680 | 1 | 155872834 | 155914191 | 67 | 9 | 29615 | -0.3121 | 0.62252 | *KIAA0907* |
| ENSG00000142327 | 2 | 241495221 | 241530789 | 115 | 23 | 51872 | -0.31215 | 0.62254 | *RNPEPL1* |
| ENSG00000121898 | 10 | 125455723 | 125709783 | 1189 | 155 | 44848 | -0.31224 | 0.62257 | *CPXM2* |
| ENSG00000112679 | 6 | 281630 | 361355 | 41 | 16 | 43410 | -0.31227 | 0.62258 | *DUSP22* |
| ENSG00000115368 | 2 | 190296159 | 190350291 | 262 | 12 | 49790 | -0.31276 | 0.62277 | *WDR75* |
| ENSG00000144485 | 2 | 239136908 | 239159303 | 115 | 10 | 49484 | -0.31279 | 0.62278 | *HES6* |
| ENSG00000154845 | 18 | 9536789 | 9625238 | 349 | 41 | 44801 | -0.31283 | 0.62279 | *PPP4R1* |
| ENSG00000162398 | 1 | 55261736 | 55317925 | 274 | 22 | 48694 | -0.31285 | 0.6228 | *C1orf177* |
| ENSG00000138758 | 4 | 77860856 | 77971537 | 331 | 33 | 52054 | -0.31313 | 0.62291 | *Sep-11* |
| ENSG00000188342 | 13 | 45684650 | 45868237 | 384 | 28 | 40995 | -0.31313 | 0.62291 | *GTF2F2* |
| ENSG00000115419 | 2 | 191735553 | 191840278 | 188 | 25 | 43486 | -0.31357 | 0.62307 | *GLS* |
| ENSG00000078795 | 5 | 137213657 | 137288436 | 139 | 20 | 49282 | -0.31354 | 0.62307 | *PKD2L2* |
| ENSG00000131355 | 19 | 14719929 | 14810839 | 376 | 51 | 47563 | -0.3136 | 0.62309 | *EMR3* |
| ENSG00000215218 | 5 | 6438736 | 6505022 | 235 | 44 | 37443 | -0.31384 | 0.62318 | *UBE2QL1* |
| ENSG00000150394 | 16 | 61671146 | 62080939 | 1031 | 92 | 46474 | -0.31403 | 0.62325 | *CDH8* |
| ENSG00000076924 | 19 | 7674411 | 7704451 | 94 | 13 | 52464 | -0.31453 | 0.62344 | *XAB2* |
| ENSG00000054282 | 1 | 243409320 | 243673394 | 535 | 67 | 47064 | -0.31457 | 0.62346 | *SDCCAG8* |
| ENSG00000196700 | 20 | 62578055 | 62690113 | 289 | 33 | 48770 | -0.31471 | 0.62351 | *ZNF512B* |
| ENSG00000085741 | 11 | 75887369 | 75931780 | 133 | 27 | 48533 | -0.31475 | 0.62352 | *WNT11* |
| ENSG00000171872 | 1 | 44574522 | 44610812 | 127 | 28 | 47121 | -0.31517 | 0.62368 | *KLF17* |
| ENSG00000002016 | 12 | 1011243 | 1109219 | 332 | 54 | 47657 | -0.31519 | 0.62369 | *RAD52* |
| ENSG00000172086 | 2 | 88316724 | 88365248 | 105 | 18 | 48389 | -0.31522 | 0.6237 | *KRCC1* |
| ENSG00000126464 | 19 | 50084900 | 50139696 | 183 | 18 | 41249 | -0.31523 | 0.62371 | *PRR12* |
| ENSG00000254943 | 11 | 124751200 | 124772022 | 85 | 17 | 41596 | -0.31537 | 0.62376 | *RP11-664I21.6* |

| | | | | | | | | | |
|---|---|---|---|---|---|---|---|---|---|
| ENSG00000157637 | 17 | 79208800 | 79279347 | 269 | 45 | 41156 | -0.31541 | 0.62377 | *SLC38A10* |
| ENSG00000179134 | 19 | 39823036 | 39886350 | 154 | 24 | 44942 | -0.3154 | 0.62377 | *SAMD4B* |
| ENSG00000068305 | 15 | 100007370 | 100266671 | 722 | 83 | 38727 | -0.31546 | 0.62379 | *MEF2A* |
| ENSG00000136197 | 7 | 42938325 | 42961904 | 77 | 6 | 53541 | -0.3157 | 0.62388 | *C7orf25* |
| ENSG00000173285 | 1 | 158425272 | 158446344 | 90 | 14 | 41538 | -0.31627 | 0.6241 | *OR10K1* |
| ENSG00000243927 | 21 | 35435524 | 35525334 | 142 | 26 | 48448 | -0.31634 | 0.62413 | *MRPS6* |
| ENSG00000260272 | 16 | 2536033 | 2580219 | 37 | 23 | 47377 | -0.31636 | 0.62414 | *RP11-20I23.1* |
| ENSG00000112303 | 6 | 133055009 | 133094598 | 185 | 30 | 45115 | -0.31657 | 0.62421 | *VNN2* |
| ENSG00000229833 | 19 | 7684623 | 7706842 | 72 | 10 | 53476 | -0.31661 | 0.62423 | *PET100* |
| ENSG00000169903 | 3 | 149181761 | 149231068 | 257 | 27 | 40141 | -0.31678 | 0.6243 | *TM4SF4* |
| ENSG00000168653 | 1 | 39481990 | 39510308 | 76 | 15 | 44998 | -0.3169 | 0.62434 | *NDUFS5* |
| ENSG00000136866 | 9 | 115790660 | 115829039 | 100 | 20 | 35512 | -0.31694 | 0.62435 | *ZFP37* |
| ENSG00000100504 | 14 | 51314609 | 51421454 | 417 | 60 | 47876 | -0.31724 | 0.62447 | *PYGL* |
| ENSG00000162396 | 1 | 55212571 | 55240187 | 99 | 17 | 48554 | -0.31749 | 0.62456 | *PARS2* |
| ENSG00000271079 | 7 | 143258894 | 143281480 | 8 | 5 | 43112 | -0.31747 | 0.62456 | *CTAGE15* |
| ENSG00000174501 | 2 | 96504587 | 96667541 | 248 | 29 | 46876 | -0.31768 | 0.62464 | *ANKRD36C* |
| ENSG00000188508 | 19 | 35968226 | 35996460 | 102 | 21 | 47788 | -0.31775 | 0.62466 | *KRTDAP* |
| ENSG00000135211 | 7 | 77413045 | 77437897 | 67 | 15 | 42548 | -0.31791 | 0.62472 | *TMEM60* |
| ENSG00000172748 | 8 | 172137 | 207342 | 144 | 24 | 49834 | -0.31805 | 0.62478 | *ZNF596* |
| ENSG00000091138 | 7 | 107395912 | 107453670 | 177 | 33 | 44820 | -0.31834 | 0.62488 | *SLC26A3* |
| ENSG00000160013 | 19 | 47113725 | 47138375 | 101 | 16 | 43826 | -0.31841 | 0.62491 | *PTGIR* |
| ENSG00000184459 | 22 | 32799834 | 32870471 | 307 | 69 | 42661 | -0.3184 | 0.62491 | *BPIFC* |
| ENSG00000100147 | 22 | 42186683 | 42232303 | 117 | 17 | 45049 | -0.31841 | 0.62491 | *CCDC134* |
| ENSG00000164972 | 9 | 34369017 | 34407830 | 91 | 20 | 44434 | -0.31879 | 0.62506 | *C9orf24* |
| ENSG00000159063 | 11 | 77801982 | 77860706 | 319 | 13 | 35196 | -0.31884 | 0.62507 | *ALG8* |
| ENSG00000162630 | 1 | 193138175 | 193165784 | 70 | 11 | 54729 | -0.3193 | 0.62525 | *B3GALT2* |
| ENSG00000180190 | 8 | 429803 | 505781 | 383 | 48 | 48435 | -0.31954 | 0.62534 | *TDRP* |
| ENSG00000171962 | 17 | 17866127 | 17930203 | 82 | 16 | 37884 | -0.31969 | 0.6254 | *LRRC48* |
| ENSG00000178053 | 3 | 158278952 | 158335041 | 225 | 19 | 51791 | -0.31979 | 0.62544 | *MLF1* |
| ENSG00000171195 | 4 | 71286209 | 71358714 | 350 | 34 | 47965 | -0.32019 | 0.62559 | *MUC7* |
| ENSG00000166317 | 10 | 75394639 | 75433561 | 54 | 10 | 36265 | -0.32022 | 0.6256 | *SYNPO2L* |
| ENSG00000158555 | 11 | 75135685 | 75246948 | 295 | 40 | 41866 | -0.32024 | 0.62561 | *GDPD5* |
| ENSG00000141569 | 17 | 73866416 | 73903084 | 106 | 15 | 38471 | -0.32055 | 0.62573 | *TRIM65* |
| ENSG00000176798 | 11 | 5010213 | 5031160 | 93 | 12 | 53467 | -0.32085 | 0.62584 | *OR51L1* |
| ENSG00000072080 | 2 | 234949323 | 234995778 | 241 | 29 | 41369 | -0.32096 | 0.62588 | *SPP2* |
| ENSG00000002745 | 7 | 120955421 | 120991158 | 132 | 20 | 32441 | -0.32109 | 0.62593 | *WNT16* |

| | | | | | | | | | |
|---|---|---|---|---|---|---|---|---|---|
| ENSG00000122417 | 1 | 86804344 | 86872023 | 200 | 19 | 55813 | -0.32113 | 0.62594 | *ODF2L* |
| ENSG00000141753 | 17 | 38589702 | 38623983 | 86 | 20 | 48813 | -0.32121 | 0.62597 | *IGFBP4* |
| ENSG00000268744 | 19 | 12480003 | 12522088 | 74 | 16 | 34439 | -0.32123 | 0.62598 | *CTD-3105H18.14* |
| ENSG00000165131 | 7 | 142626440 | 142647955 | 57 | 11 | 29918 | -0.32146 | 0.62607 | *C7orf34* |
| ENSG00000089558 | 17 | 40298909 | 40343296 | 65 | 16 | 43577 | -0.32171 | 0.62617 | *KCNH4* |
| ENSG00000130699 | 20 | 60518525 | 60650866 | 396 | 37 | 40870 | -0.32199 | 0.62627 | *TAF4* |
| ENSG00000131966 | 14 | 58656798 | 58711750 | 102 | 17 | 47821 | -0.32236 | 0.62641 | *ACTR10* |
| ENSG00000139144 | 12 | 18390548 | 18811348 | 1405 | 114 | 49111 | -0.32243 | 0.62644 | *PIK3C2G* |
| ENSG00000100290 | 22 | 43496754 | 43535718 | 134 | 31 | 46891 | -0.32267 | 0.62653 | *BIK* |
| ENSG00000170734 | 6 | 43533887 | 43596701 | 110 | 18 | 42506 | -0.32304 | 0.62667 | *POLH* |
| ENSG00000149474 | 20 | 18108762 | 18179031 | 294 | 45 | 43871 | -0.32314 | 0.62671 | *CSRP2BP* |
| ENSG00000269620 | 1 | 159869884 | 159890159 | 46 | 17 | 41033 | -0.32319 | 0.62672 | *AL590560.1* |
| ENSG00000263136 | 16 | 29817832 | 29841438 | 35 | 6 | 45725 | -0.32322 | 0.62674 | *PAGR1* |
| ENSG00000171159 | 9 | 130912539 | 130936207 | 44 | 9 | 33794 | -0.32332 | 0.62677 | *C9orf16* |
| ENSG00000131435 | 5 | 131583364 | 131619147 | 111 | 16 | 29451 | -0.32334 | 0.62678 | *PDLIM4* |
| ENSG00000204695 | 6 | 29264403 | 29285519 | 92 | 16 | 47166 | -0.32335 | 0.62678 | *OR14J1* |
| ENSG00000109674 | 4 | 178220990 | 178294097 | 246 | 51 | 44164 | -0.32361 | 0.62688 | *NEIL3* |
| ENSG00000136636 | 1 | 215730735 | 215805149 | 54 | 23 | 41295 | -0.32363 | 0.62689 | *KCTD3* |
| ENSG00000272617 | 16 | 69352547 | 69378774 | 73 | 14 | 54189 | -0.32372 | 0.62693 | *RP11-343C2.12* |
| ENSG00000130803 | 19 | 9241056 | 9284100 | 194 | 25 | 44868 | -0.32373 | 0.62693 | *ZNF317* |
| ENSG00000197769 | 1 | 242148792 | 242172375 | 120 | 20 | 40117 | -0.32424 | 0.62712 | *MAP1LC3C* |
| ENSG00000166813 | 15 | 90142020 | 90208682 | 164 | 19 | 45773 | -0.32432 | 0.62715 | *KIF7* |
| ENSG00000204677 | 5 | 177423406 | 177492560 | 148 | 32 | 43097 | -0.32442 | 0.62719 | *FAM153C* |
| ENSG00000168495 | 8 | 22092617 | 22122113 | 89 | 12 | 48865 | -0.32452 | 0.62723 | *POLR3D* |
| ENSG00000136696 | 2 | 113769668 | 113820444 | 181 | 23 | 37761 | -0.32475 | 0.62732 | *IL36B* |
| ENSG00000111305 | 12 | 13226494 | 13266619 | 61 | 18 | 44528 | -0.32482 | 0.62734 | *GSG1* |
| ENSG00000162852 | 1 | 246719746 | 246841886 | 423 | 55 | 39676 | -0.325 | 0.62741 | *CNST* |
| ENSG00000085231 | 5 | 68636811 | 68675840 | 70 | 12 | 37818 | -0.32518 | 0.62748 | *TAF9* |
| ENSG00000152939 | 5 | 68700939 | 68750157 | 106 | 10 | 40222 | -0.32525 | 0.6275 | *MARVELD2* |
| ENSG00000175287 | 9 | 131673174 | 131714320 | 75 | 10 | 46443 | -0.32524 | 0.6275 | *PHYHD1* |
| ENSG00000151746 | 12 | 32249769 | 32546567 | 1431 | 118 | 49066 | -0.32535 | 0.62754 | *BICD1* |
| ENSG00000229972 | 3 | 51841620 | 51874876 | 40 | 6 | 24588 | -0.32577 | 0.6277 | *IQCF3* |
| ENSG00000095981 | 6 | 39272474 | 39300744 | 147 | 29 | 50887 | -0.32577 | 0.6277 | *KCNK16* |
| ENSG00000187735 | 8 | 54869112 | 54945089 | 85 | 17 | 35393 | -0.3258 | 0.62771 | *TCEA1* |
| ENSG00000122223 | 1 | 160789950 | 160842692 | 181 | 17 | 30623 | -0.32606 | 0.62781 | *CD244* |
| ENSG00000134291 | 12 | 48347352 | 48372661 | 84 | 18 | 50584 | -0.32607 | 0.62781 | *TMEM106C* |

| | | | | | | | | | |
|---|---|---|---|---|---|---|---|---|---|
| ENSG00000186970 | 21 | 31802597 | 31823070 | 67 | 4 | 56052 | -0.32611 | 0.62783 | *KRTAP15-1* |
| ENSG00000204052 | 6 | 43464707 | 43488424 | 44 | 9 | 45751 | -0.32628 | 0.62789 | *LRRC73* |
| ENSG00000104833 | 19 | 6484330 | 6512859 | 53 | 25 | 48517 | -0.32632 | 0.62791 | *TUBB4A* |
| ENSG00000155287 | 10 | 101360282 | 101390366 | 111 | 13 | 55884 | -0.32639 | 0.62793 | *SLC25A28* |
| ENSG00000163376 | 3 | 67038727 | 67071634 | 56 | 12 | 50943 | -0.3265 | 0.62798 | *KBTBD8* |
| ENSG00000054598 | 6 | 1600681 | 1624127 | 54 | 20 | 42409 | -0.3266 | 0.62801 | *FOXC1* |
| ENSG00000212725 | 17 | 39192793 | 39213568 | 83 | 11 | 37952 | -0.32662 | 0.62802 | *KRTAP2-1* |
| ENSG00000167600 | 19 | 41688954 | 41723444 | 127 | 21 | 40805 | -0.32674 | 0.62807 | *CYP2S1* |
| ENSG00000101400 | 20 | 31985761 | 32041698 | 71 | 12 | 51425 | -0.32676 | 0.62808 | *SNTA1* |
| ENSG00000248099 | 19 | 17917321 | 17942383 | 61 | 26 | 46843 | -0.32687 | 0.62812 | *INSL3* |
| ENSG00000169131 | 5 | 178128593 | 178167703 | 175 | 14 | 50937 | -0.32694 | 0.62814 | *ZNF354A* |
| ENSG00000105198 | 19 | 40083164 | 40108116 | 58 | 12 | 55622 | -0.32695 | 0.62815 | *LGALS13* |
| ENSG00000161634 | 12 | 55028375 | 55052277 | 73 | 18 | 50772 | -0.327 | 0.62816 | *DCD* |
| ENSG00000018189 | 4 | 71559921 | 71683032 | 386 | 29 | 28330 | -0.327 | 0.62817 | *RUFY3* |
| ENSG00000134748 | 1 | 52860236 | 52896511 | 38 | 11 | 39140 | -0.32723 | 0.62825 | *PRPF38A* |
| ENSG00000163464 | 2 | 219017568 | 219041718 | 54 | 11 | 40684 | -0.32802 | 0.62855 | *CXCR1* |
| ENSG00000148356 | 9 | 130203765 | 130275780 | 210 | 17 | 53479 | -0.32829 | 0.62865 | *LRSAM1* |
| ENSG00000060749 | 11 | 32904724 | 33024862 | 268 | 23 | 48528 | -0.32831 | 0.62866 | *QSER1* |
| ENSG00000064313 | 8 | 120733015 | 120855103 | 360 | 27 | 36331 | -0.3284 | 0.6287 | *TAF2* |
| ENSG00000165525 | 14 | 50239997 | 50329921 | 98 | 18 | 52202 | -0.32841 | 0.6287 | *NEMF* |
| ENSG00000187775 | 17 | 76409778 | 76583476 | 997 | 146 | 50665 | -0.32845 | 0.62872 | *DNAH17* |
| ENSG00000109099 | 17 | 15123095 | 15178643 | 133 | 24 | 48713 | -0.32871 | 0.62881 | *PMP22* |
| ENSG00000181218 | 1 | 228635065 | 228655560 | 25 | 9 | 52651 | -0.3289 | 0.62888 | *HIST3H2A* |
| ENSG00000196890 | 1 | 228635808 | 228656259 | 25 | 9 | 52651 | -0.3289 | 0.62888 | *HIST3H2BB* |
| ENSG00000144306 | 2 | 175250458 | 175304303 | 176 | 13 | 47713 | -0.3289 | 0.62888 | *SCRN3* |
| ENSG00000049860 | 5 | 73925848 | 74028472 | 375 | 63 | 48380 | -0.32928 | 0.62903 | *HEXB* |
| ENSG00000006530 | 7 | 141240989 | 141365044 | 220 | 25 | 34803 | -0.32927 | 0.62903 | *AGK* |
| ENSG00000133962 | 14 | 92037040 | 92257051 | 824 | 97 | 51214 | -0.32942 | 0.62908 | *CATSPERB* |
| ENSG00000143473 | 1 | 210846555 | 211317457 | 1711 | 163 | 48011 | -0.33009 | 0.62933 | *KCNH1* |
| ENSG00000100982 | 20 | 44553267 | 44586662 | 74 | 19 | 43733 | -0.3301 | 0.62934 | *PCIF1* |
| ENSG00000214029 | 12 | 133684740 | 133717059 | 107 | 13 | 49648 | -0.33041 | 0.62946 | *ZNF891* |
| ENSG00000132297 | 8 | 133063733 | 133133406 | 336 | 48 | 46091 | -0.33063 | 0.62954 | *HHLA1* |
| ENSG00000135720 | 16 | 66744796 | 66795701 | 114 | 9 | 33112 | -0.33063 | 0.62954 | *DYNC1LI2* |
| ENSG00000150760 | 10 | 128583978 | 129260781 | 2158 | 215 | 47293 | -0.33066 | 0.62955 | *DOCK1* |
| ENSG00000187742 | 9 | 91923421 | 91984557 | 183 | 30 | 50383 | -0.33079 | 0.6296 | *SECISBP2* |
| ENSG00000100151 | 22 | 38442318 | 38481708 | 99 | 19 | 48980 | -0.331 | 0.62968 | *PICK1* |

| | | | | | | | | | |
|---|---|---|---|---|---|---|---|---|---|
| ENSG00000106588 | 7 | 42946460 | 42981822 | 119 | 9 | 53377 | -0.33112 | 0.62972 | *PSMA2* |
| ENSG00000254531 | 4 | 102258937 | 102280040 | 30 | 10 | 41930 | -0.33113 | 0.62973 | *AP001816.1* |
| ENSG00000101152 | 20 | 62516518 | 62577384 | 193 | 26 | 34681 | -0.33118 | 0.62975 | *DNAJC5* |
| ENSG00000145901 | 5 | 150399506 | 150483138 | 312 | 55 | 39377 | -0.33121 | 0.62976 | *TNIP1* |
| ENSG00000156097 | 1 | 110072494 | 110101028 | 41 | 15 | 49056 | -0.33167 | 0.62993 | *GPR61* |
| ENSG00000177710 | 8 | 11178397 | 11199717 | 86 | 9 | 47709 | -0.33189 | 0.63001 | *SLC35G5* |
| ENSG00000124614 | 6 | 34375231 | 34403902 | 52 | 11 | 29239 | -0.332 | 0.63005 | *RPS10* |
| ENSG00000182700 | 5 | 139495521 | 139518391 | 19 | 7 | 50352 | -0.33215 | 0.63011 | *IGIP* |
| ENSG00000118705 | 20 | 35796813 | 35880022 | 278 | 20 | 50246 | -0.33219 | 0.63013 | *RPN2* |
| ENSG00000075303 | 7 | 87452883 | 87515672 | 143 | 17 | 48881 | -0.3328 | 0.63036 | *SLC25A40* |
| ENSG00000165630 | 10 | 13618927 | 13682868 | 196 | 23 | 50157 | -0.33288 | 0.63039 | *PRPF18* |
| ENSG00000143493 | 1 | 212103741 | 212218884 | 485 | 37 | 52294 | -0.33317 | 0.6305 | *INTS7* |
| ENSG00000107140 | 9 | 35595367 | 35620038 | 58 | 12 | 54412 | -0.33378 | 0.63073 | *TESK1* |
| ENSG00000125967 | 20 | 32234893 | 32272269 | 39 | 12 | 48991 | -0.33388 | 0.63077 | *NECAB3* |
| ENSG00000161999 | 16 | 721671 | 744529 | 48 | 13 | 39651 | -0.33403 | 0.63082 | *JMJD8* |
| ENSG00000162623 | 1 | 75188836 | 75242362 | 184 | 22 | 49075 | -0.33405 | 0.63083 | *TYW3* |
| ENSG00000100883 | 14 | 35441163 | 35508773 | 311 | 21 | 49749 | -0.33509 | 0.63122 | *SRP54* |
| ENSG00000116095 | 2 | 179335195 | 179391189 | 119 | 22 | 33571 | -0.33541 | 0.63134 | *PLEKHA3* |
| ENSG00000249679 | 4 | 186382670 | 186403417 | 59 | 16 | 29137 | -0.33548 | 0.63137 | *RP11-279O9.4* |
| ENSG00000141965 | 19 | 4781728 | 4805571 | 110 | 23 | 47872 | -0.33564 | 0.63143 | *FEM1A* |
| ENSG00000059377 | 7 | 139466850 | 139730125 | 670 | 127 | 42931 | -0.33568 | 0.63145 | *TBXAS1* |
| ENSG00000042317 | 14 | 88841268 | 88946694 | 172 | 28 | 41563 | -0.33569 | 0.63145 | *SPATA7* |
| ENSG00000171865 | 2 | 3582383 | 3616206 | 165 | 24 | 50305 | -0.33572 | 0.63146 | *RNASEH1* |
| ENSG00000186889 | 2 | 62717356 | 62749029 | 162 | 29 | 50466 | -0.33595 | 0.63155 | *TMEM17* |
| ENSG00000114450 | 3 | 179106990 | 179179378 | 169 | 28 | 51251 | -0.3363 | 0.63168 | *GNB4* |
| ENSG00000079691 | 6 | 25269306 | 25630758 | 1680 | 154 | 49951 | -0.33674 | 0.63184 | *LRRC16A* |
| ENSG00000196366 | 9 | 139367947 | 139390518 | 82 | 15 | 42868 | -0.33681 | 0.63187 | *C9orf163* |
| ENSG00000007516 | 16 | 1373602 | 1409439 | 148 | 27 | 45886 | -0.33697 | 0.63193 | *BAIAP3* |
| ENSG00000243772 | 19 | 55239980 | 55305776 | 56 | 28 | 33793 | -0.33699 | 0.63194 | *KIR2DL3* |
| ENSG00000213390 | 10 | 98968996 | 99062413 | 326 | 26 | 46416 | -0.33734 | 0.63207 | *ARHGAP19* |
| ENSG00000116147 | 1 | 175274330 | 175722906 | 1400 | 165 | 47743 | -0.33751 | 0.63213 | *TNR* |
| ENSG00000163126 | 2 | 97480263 | 97533671 | 59 | 11 | 37556 | -0.33776 | 0.63223 | *ANKRD23* |
| ENSG00000177888 | 1 | 197112810 | 197179672 | 118 | 15 | 27080 | -0.33817 | 0.63238 | *ZBTB41* |
| ENSG00000102904 | 16 | 67830668 | 67876051 | 72 | 21 | 39971 | -0.33832 | 0.63244 | *TSNAXIP1* |
| ENSG00000197766 | 19 | 849453 | 873453 | 97 | 28 | 41102 | -0.33845 | 0.63249 | *CFD* |
| ENSG00000198948 | 4 | 170897748 | 170964182 | 170 | 27 | 47182 | -0.3385 | 0.63251 | *MFAP3L* |

| | | | | | | | | | |
|---|---|---|---|---|---|---|---|---|---|
| ENSG00000142892 | 1 | 77544675 | 77695115 | 428 | 33 | 36040 | -0.33873 | 0.63259 | *PIGK* |
| ENSG00000205740 | 10 | 1007097 | 1044281 | 123 | 18 | 49301 | -0.33903 | 0.63271 | *AL359878.1* |
| ENSG00000267260 | 19 | 37254055 | 37277980 | 61 | 12 | 48906 | -0.33908 | 0.63273 | *CTD-2162K18.4* |
| ENSG00000117174 | 1 | 86105106 | 86184116 | 260 | 31 | 49710 | -0.33916 | 0.63275 | *ZNHIT6* |
| ENSG00000169946 | 8 | 106320920 | 106826760 | 1275 | 141 | 44313 | -0.33941 | 0.63285 | *ZFPM2* |
| ENSG00000124422 | 17 | 20892910 | 20957073 | 281 | 21 | 51028 | -0.33952 | 0.63289 | *USP22* |
| ENSG00000119919 | 10 | 101282690 | 101306278 | 92 | 10 | 53850 | -0.33965 | 0.63294 | *NKX2-3* |
| ENSG00000155330 | 16 | 46820519 | 46875323 | 44 | 9 | 31071 | -0.33975 | 0.63298 | *C16orf87* |
| ENSG00000172023 | 2 | 79302156 | 79325145 | 93 | 15 | 52212 | -0.34001 | 0.63307 | *REG1B* |
| ENSG00000169241 | 1 | 155097820 | 155121329 | 40 | 13 | 44258 | -0.34003 | 0.63308 | *SLC50A1* |
| ENSG00000121769 | 1 | 31828472 | 31859697 | 91 | 12 | 31254 | -0.34042 | 0.63323 | *FABP3* |
| ENSG00000011347 | 11 | 61272785 | 61358620 | 133 | 28 | 39472 | -0.34076 | 0.63336 | *SYT7* |
| ENSG00000138308 | 10 | 74684928 | 74724536 | 31 | 15 | 38232 | -0.3409 | 0.63341 | *PLA2G12B* |
| ENSG00000181495 | 5 | 32778945 | 32801819 | 50 | 10 | 46657 | -0.34103 | 0.63346 | *AC026703.1* |
| ENSG00000168301 | 3 | 58467841 | 58498087 | 66 | 16 | 48279 | -0.34135 | 0.63358 | *KCTD6* |
| ENSG00000077420 | 10 | 26717132 | 26866732 | 542 | 49 | 51697 | -0.34135 | 0.63358 | *APBB1IP* |
| ENSG00000109220 | 4 | 54865956 | 54940857 | 113 | 13 | 53411 | -0.34196 | 0.63381 | *CHIC2* |
| ENSG00000070669 | 7 | 97471430 | 97511854 | 170 | 24 | 43873 | -0.34199 | 0.63382 | *ASNS* |
| ENSG00000164885 | 7 | 150740899 | 150765617 | 63 | 17 | 55439 | -0.34201 | 0.63383 | *CDK5* |
| ENSG00000150893 | 13 | 39251266 | 39470074 | 610 | 74 | 42634 | -0.34208 | 0.63386 | *FREM2* |
| ENSG00000270898 | 2 | 53887430 | 54097297 | 725 | 59 | 45411 | -0.34213 | 0.63387 | *ASB3* |
| ENSG00000129347 | 19 | 10653761 | 10686713 | 124 | 21 | 43325 | -0.34268 | 0.63408 | *KRI1* |
| ENSG00000138032 | 2 | 44385108 | 44481523 | 242 | 28 | 49983 | -0.34273 | 0.6341 | *PPM1B* |
| ENSG00000102901 | 16 | 67852060 | 67891714 | 65 | 15 | 42833 | -0.34283 | 0.63414 | *CENPT* |
| ENSG00000135960 | 2 | 109500927 | 109615828 | 364 | 43 | 44697 | -0.34316 | 0.63426 | *EDAR* |
| ENSG00000167555 | 19 | 52891102 | 52931665 | 155 | 29 | 30145 | -0.34325 | 0.63429 | *ZNF528* |
| ENSG00000171714 | 11 | 22204722 | 22314903 | 220 | 25 | 40176 | -0.34362 | 0.63443 | *ANO5* |
| ENSG00000128512 | 7 | 111356166 | 111856466 | 1271 | 184 | 42228 | -0.34363 | 0.63444 | *DOCK4* |
| ENSG00000163518 | 1 | 157533539 | 157577870 | 160 | 17 | 42544 | -0.34376 | 0.63449 | *FCRL4* |
| ENSG00000105708 | 19 | 19811280 | 19853906 | 138 | 20 | 43642 | -0.34407 | 0.6346 | *ZNF14* |
| ENSG00000197969 | 9 | 79782269 | 80046457 | 880 | 57 | 55177 | -0.34454 | 0.63478 | *VPS13A* |
| ENSG00000137462 | 4 | 154612652 | 154636851 | 50 | 14 | 44641 | -0.34542 | 0.63511 | *TLR2* |
| ENSG00000164542 | 7 | 36353830 | 36439734 | 261 | 35 | 50025 | -0.34565 | 0.6352 | *KIAA0895* |
| ENSG00000258231 | 12 | 58927907 | 59216842 | 829 | 101 | 45891 | -0.34595 | 0.63531 | *RP11-362K2.2* |
| ENSG00000167768 | 12 | 53058520 | 53084191 | 101 | 16 | 55277 | -0.34602 | 0.63534 | *KRT1* |
| ENSG00000171049 | 19 | 52245279 | 52283779 | 165 | 32 | 43518 | -0.3462 | 0.6354 | *FPR2* |

| | | | | | | | | | |
|---|---|---|---|---|---|---|---|---|---|
| ENSG00000204381 | 11 | 111401005 | 111442264 | 77 | 14 | 43936 | -0.34648 | 0.63551 | *LAYN* |
| ENSG00000181072 | 7 | 136543416 | 136715002 | 556 | 56 | 38430 | -0.34666 | 0.63558 | *CHRM2* |
| ENSG00000213337 | 2 | 97492348 | 97533832 | 46 | 9 | 37764 | -0.34679 | 0.63563 | *ANKRD39* |
| ENSG00000269182 | 15 | 22446895 | 22470351 | 3 | 2 | 42731 | -0.3468 | 0.63563 | *AC010760.1* |
| ENSG00000027644 | 1 | 156799855 | 156838810 | 82 | 24 | 41940 | -0.34693 | 0.63568 | *INSRR* |
| ENSG00000196358 | 9 | 135027334 | 135129921 | 255 | 49 | 40476 | -0.3471 | 0.63574 | *NTNG2* |
| ENSG00000171794 | 10 | 135033778 | 135055062 | 104 | 22 | 44631 | -0.34709 | 0.63574 | *UTF1* |
| ENSG00000143416 | 1 | 151326778 | 151355209 | 74 | 20 | 45263 | -0.34715 | 0.63576 | *SELENBP1* |
| ENSG00000269165 | 10 | 52084298 | 52105876 | 57 | 13 | 47540 | -0.34718 | 0.63577 | *AC069547.1* |
| ENSG00000105556 | 19 | 295575 | 354798 | 280 | 28 | 32482 | -0.34727 | 0.63581 | *MIER2* |
| ENSG00000127081 | 9 | 95597874 | 95650304 | 176 | 25 | 47242 | -0.34733 | 0.63583 | *ZNF484* |
| ENSG00000118194 | 1 | 201318136 | 201356890 | 154 | 25 | 38232 | -0.34739 | 0.63585 | *TNNT2* |
| ENSG00000187223 | 1 | 152625872 | 152647135 | 34 | 7 | 27585 | -0.34741 | 0.63586 | *LCE2D* |
| ENSG00000156860 | 16 | 30659752 | 30692135 | 25 | 10 | 31418 | -0.34753 | 0.6359 | *FBRS* |
| ENSG00000261949 | 19 | 49917006 | 49942075 | 38 | 16 | 49252 | -0.34781 | 0.63601 | *GFY* |
| ENSG00000126775 | 14 | 55823110 | 55888576 | 233 | 22 | 47596 | -0.34822 | 0.63616 | *ATG14* |
| ENSG00000120837 | 12 | 104500855 | 104542067 | 113 | 21 | 51706 | -0.34829 | 0.63619 | *NFYB* |
| ENSG00000143344 | 1 | 183595220 | 183907665 | 1047 | 79 | 39687 | -0.34849 | 0.63626 | *RGL1* |
| ENSG00000075218 | 22 | 46682638 | 46736707 | 195 | 19 | 36287 | -0.34848 | 0.63626 | *GTSE1* |
| ENSG00000052126 | 12 | 19272648 | 19539334 | 593 | 67 | 42140 | -0.34877 | 0.63637 | *PLEKHA5* |
| ENSG00000212900 | 17 | 39145447 | 39166138 | 178 | 14 | 39422 | -0.34917 | 0.63652 | *KRTAP3-2* |
| ENSG00000108639 | 17 | 76154639 | 76179608 | 116 | 26 | 35453 | -0.35037 | 0.63697 | *SYNGR2* |
| ENSG00000256463 | 18 | 76730275 | 76772677 | 118 | 27 | 50325 | -0.35097 | 0.63719 | *SALL3* |
| ENSG00000120709 | 5 | 137657624 | 137695416 | 98 | 17 | 47643 | -0.35127 | 0.63731 | *FAM53C* |
| ENSG00000116833 | 1 | 199986730 | 200156552 | 638 | 96 | 44466 | -0.35129 | 0.63732 | *NR5A2* |
| ENSG00000169031 | 2 | 228019281 | 228189508 | 928 | 113 | 45640 | -0.35135 | 0.63734 | *COL4A3* |
| ENSG00000144671 | 3 | 38313785 | 38370066 | 246 | 16 | 49147 | -0.35183 | 0.63752 | *SLC22A14* |
| ENSG00000120697 | 13 | 37513912 | 37584398 | 138 | 20 | 41588 | -0.352 | 0.63758 | *ALG5* |
| ENSG00000205129 | 4 | 186337394 | 186380980 | 166 | 25 | 32425 | -0.35219 | 0.63765 | *C4orf47* |
| ENSG00000125726 | 19 | 6573194 | 6614114 | 192 | 36 | 46978 | -0.35227 | 0.63768 | *CD70* |
| ENSG00000166710 | 15 | 44993675 | 45021075 | 60 | 16 | 43154 | -0.35235 | 0.63771 | *B2M* |
| ENSG00000057663 | 6 | 106622351 | 106783666 | 492 | 38 | 52917 | -0.35237 | 0.63772 | *ATG5* |
| ENSG00000131037 | 19 | 55573388 | 55609291 | 143 | 25 | 52125 | -0.35276 | 0.63786 | *EPS8L1* |
| ENSG00000230891 | 3 | 25890024 | 25925191 | 85 | 10 | 31813 | -0.35277 | 0.63787 | *LINC00692* |
| ENSG00000111291 | 12 | 13083709 | 13115081 | 98 | 25 | 40170 | -0.35279 | 0.63788 | *GPRC5D* |
| ENSG00000109452 | 4 | 142934313 | 143778585 | 2646 | 175 | 42713 | -0.353 | 0.63796 | *INPP4B* |

| | | | | | | | | | |
|---|---|---|---|---|---|---|---|---|---|
| ENSG00000116396 | 1 | 110743965 | 110835722 | 312 | 39 | 45833 | -0.35319 | 0.63803 | *KCNC4* |
| ENSG00000134755 | 18 | 28635940 | 28692378 | 112 | 28 | 47097 | -0.35321 | 0.63804 | *DSC2* |
| ENSG00000180999 | 1 | 172379828 | 172447971 | 221 | 33 | 50179 | -0.35344 | 0.63812 | *C1orf105* |
| ENSG00000091732 | 7 | 129648126 | 129701291 | 172 | 20 | 46202 | -0.35355 | 0.63816 | *ZC3HC1* |
| ENSG00000253710 | 13 | 52576534 | 52613800 | 77 | 12 | 38564 | -0.35364 | 0.63819 | *ALG11* |
| ENSG00000140382 | 15 | 77702754 | 77787949 | 148 | 16 | 47506 | -0.35437 | 0.63847 | *HMG20A* |
| ENSG00000111254 | 12 | 4714674 | 4768213 | 221 | 27 | 38900 | -0.35441 | 0.63848 | *AKAP3* |
| ENSG00000159267 | 21 | 38113189 | 38372536 | 917 | 93 | 47452 | -0.35441 | 0.63848 | *HLCS* |
| ENSG00000171161 | 1 | 249122409 | 249153716 | 37 | 12 | 34511 | -0.35449 | 0.63851 | *ZNF672* |
| ENSG00000157349 | 16 | 70313566 | 70379186 | 69 | 31 | 48411 | -0.35457 | 0.63854 | *DDX19B* |
| ENSG00000067955 | 16 | 67053019 | 67144961 | 123 | 19 | 46448 | -0.35472 | 0.6386 | *CBFB* |
| ENSG00000133835 | 5 | 118778138 | 118982894 | 684 | 63 | 46709 | -0.35506 | 0.63873 | *HSD17B4* |
| ENSG00000139163 | 12 | 22768009 | 22853599 | 186 | 19 | 39594 | -0.35541 | 0.63886 | *ETNK1* |
| ENSG00000149182 | 11 | 47175848 | 47208676 | 69 | 14 | 47173 | -0.35547 | 0.63888 | *ARFGAP2* |
| ENSG00000163209 | 1 | 152964223 | 152986332 | 161 | 5 | 52779 | -0.35554 | 0.63891 | *SPRR3* |
| ENSG00000188488 | 14 | 95017779 | 95069457 | 311 | 42 | 46769 | -0.35568 | 0.63896 | *SERPINA5* |
| ENSG00000116857 | 1 | 201093900 | 201150702 | 237 | 46 | 46285 | -0.35605 | 0.6391 | *TMEM9* |
| ENSG00000162241 | 11 | 65132663 | 65161172 | 46 | 17 | 38076 | -0.35625 | 0.63917 | *SLC25A45* |
| ENSG00000197217 | 8 | 23233296 | 23325208 | 363 | 41 | 46341 | -0.35668 | 0.63933 | *ENTPD4* |
| ENSG00000105865 | 7 | 107193929 | 107228906 | 63 | 11 | 53644 | -0.35673 | 0.63935 | *DUS4L* |
| ENSG00000160588 | 11 | 118087409 | 118133065 | 172 | 15 | 39760 | -0.35674 | 0.63936 | *MPZL3* |
| ENSG00000173141 | 13 | 21740784 | 21763223 | 101 | 16 | 45104 | -0.35693 | 0.63943 | *MRP63* |
| ENSG00000188340 | 1 | 158736472 | 158757425 | 94 | 24 | 44526 | -0.35723 | 0.63954 | *OR6N2* |
| ENSG00000172171 | 17 | 29214354 | 29243838 | 84 | 7 | 25632 | -0.35724 | 0.63955 | *TEFM* |
| ENSG00000182944 | 22 | 29653998 | 29706515 | 94 | 23 | 51268 | -0.35725 | 0.63955 | *EWSR1* |
| ENSG00000182901 | 1 | 240921554 | 241530530 | 2263 | 222 | 45555 | -0.35734 | 0.63958 | *RGS7* |
| ENSG00000101109 | 20 | 43585115 | 43718600 | 282 | 21 | 54414 | -0.35771 | 0.63972 | *STK4* |
| ENSG00000166478 | 11 | 9471866 | 9560071 | 204 | 25 | 48821 | -0.35787 | 0.63978 | *ZNF143* |
| ENSG00000151466 | 4 | 129776076 | 130024764 | 619 | 47 | 47102 | -0.35801 | 0.63983 | *SCLT1* |
| ENSG00000180909 | 11 | 6163006 | 6183818 | 72 | 16 | 34076 | -0.35799 | 0.63983 | *OR52B1P* |
| ENSG00000215431 | 18 | 60481423 | 60502724 | 36 | 8 | 43516 | -0.35799 | 0.63983 | *AC015989.1* |
| ENSG00000100314 | 22 | 30106073 | 30137828 | 106 | 18 | 43927 | -0.35809 | 0.63986 | *CABP7* |
| ENSG00000160223 | 21 | 45632874 | 45670849 | 152 | 31 | 43777 | -0.35824 | 0.63992 | *ICOSLG* |
| ENSG00000124251 | 20 | 43992526 | 44046529 | 138 | 13 | 39538 | -0.35827 | 0.63993 | *TP53TG5* |
| ENSG00000268628 | 20 | 19728296 | 19748679 | 75 | 20 | 43379 | -0.35841 | 0.63998 | *AL121761.2* |
| ENSG00000185875 | 10 | 25295587 | 25325593 | 115 | 14 | 40269 | -0.35857 | 0.64004 | *THNSL1* |

| | | | | | | | | | |
|---|---|---|---|---|---|---|---|---|---|
| ENSG00000185761 | 19 | 1495017 | 1523603 | 94 | 22 | 40900 | -0.35877 | 0.64012 | *ADAMTSL5* |
| ENSG00000110047 | 11 | 64609114 | 64665768 | 117 | 20 | 36641 | -0.35884 | 0.64014 | *EHD1* |
| ENSG00000159579 | 16 | 57210049 | 57284387 | 161 | 22 | 40471 | -0.35885 | 0.64015 | *RSPRY1* |
| ENSG00000213029 | 1 | 229430129 | 229451248 | 47 | 8 | 55653 | -0.3593 | 0.64032 | *SPHAR* |
| ENSG00000057608 | 10 | 5797186 | 5894095 | 323 | 48 | 50317 | -0.3595 | 0.64039 | *GDI2* |
| ENSG00000156508 | 6 | 74215473 | 74243520 | 87 | 13 | 42583 | -0.35967 | 0.64045 | *EEF1A1* |
| ENSG00000176390 | 17 | 29086406 | 29161794 | 303 | 8 | 46358 | -0.35967 | 0.64045 | *CRLF3* |
| ENSG00000124882 | 4 | 75220860 | 75264468 | 206 | 20 | 43704 | -0.35977 | 0.64049 | *EREG* |
| ENSG00000079332 | 10 | 71897045 | 71940279 | 184 | 24 | 44198 | -0.35998 | 0.64057 | *SAR1A* |
| ENSG00000031691 | 6 | 49421091 | 49470820 | 122 | 17 | 51488 | -0.36021 | 0.64066 | *CENPQ* |
| ENSG00000100266 | 22 | 43221418 | 43421151 | 627 | 38 | 47731 | -0.36034 | 0.6407 | *PACSIN2* |
| ENSG00000161939 | 17 | 6905954 | 6930839 | 57 | 8 | 51261 | -0.36064 | 0.64082 | *RNASEK-C17orf49* |
| ENSG00000130487 | 22 | 50976462 | 50999451 | 158 | 16 | 28296 | -0.36082 | 0.64088 | *KLHDC7B* |
| ENSG00000136153 | 13 | 76184570 | 76444004 | 834 | 96 | 44538 | -0.36094 | 0.64093 | *LMO7* |
| ENSG00000054392 | 1 | 210491596 | 210859638 | 1539 | 186 | 50764 | -0.36109 | 0.64098 | *HHAT* |
| ENSG00000172765 | 3 | 129356635 | 129622419 | 323 | 34 | 48312 | -0.36144 | 0.64111 | *TMCC1* |
| ENSG00000121680 | 11 | 45921220 | 45950363 | 38 | 12 | 52004 | -0.36181 | 0.64125 | *PEX16* |
| ENSG00000268302 | 20 | 25197122 | 25217370 | 41 | 9 | 28924 | -0.36182 | 0.64126 | *AL035252.1* |
| ENSG00000189325 | 6 | 36273534 | 36314662 | 101 | 29 | 41972 | -0.36187 | 0.64127 | *C6orf222* |
| ENSG00000258555 | 22 | 24656866 | 24848324 | 537 | 47 | 41855 | -0.36201 | 0.64133 | *SPECC1L-ADORA2A* |
| ENSG00000168538 | 4 | 184570420 | 184644745 | 294 | 38 | 48464 | -0.36205 | 0.64134 | *TRAPPC11* |
| ENSG00000183111 | 5 | 148921510 | 149024531 | 328 | 37 | 41027 | -0.36216 | 0.64138 | *ARHGEF37* |
| ENSG00000168028 | 3 | 39438180 | 39464033 | 180 | 8 | 54051 | -0.36222 | 0.64141 | *RPSA* |
| ENSG00000173706 | 3 | 124674554 | 124784802 | 363 | 47 | 46525 | -0.36282 | 0.64163 | *HEG1* |
| ENSG00000122861 | 10 | 75658935 | 75687255 | 54 | 9 | 32772 | -0.36298 | 0.64169 | *PLAU* |
| ENSG00000171051 | 19 | 52238425 | 52317363 | 383 | 72 | 48679 | -0.363 | 0.6417 | *FPR1* |
| ENSG00000174279 | 2 | 176932200 | 176958641 | 64 | 16 | 43601 | -0.36304 | 0.64171 | *EVX2* |
| ENSG00000152192 | 13 | 79162497 | 79187673 | 61 | 12 | 49334 | -0.36321 | 0.64177 | *POU4F1* |
| ENSG00000169403 | 1 | 28463677 | 28530447 | 101 | 20 | 44660 | -0.36346 | 0.64187 | *PTAFR* |
| ENSG00000196503 | 4 | 57361509 | 57400614 | 138 | 21 | 40077 | -0.36349 | 0.64188 | *ARL9* |
| ENSG00000082512 | 1 | 211489957 | 211558288 | 189 | 32 | 42823 | -0.36363 | 0.64193 | *TRAF5* |
| ENSG00000116205 | 1 | 54509260 | 54588192 | 170 | 19 | 29926 | -0.36367 | 0.64195 | *TCEANC2* |
| ENSG00000198060 | 10 | 94040920 | 94123721 | 136 | 19 | 34217 | -0.36387 | 0.64202 | *Mar-05* |
| ENSG00000126215 | 14 | 104153946 | 104191841 | 89 | 18 | 47527 | -0.36394 | 0.64205 | *XRCC3* |
| ENSG00000237136 | 4 | 146591356 | 146702184 | 380 | 49 | 42738 | -0.36396 | 0.64206 | *C4orf51* |
| ENSG00000152766 | 10 | 90571889 | 90621575 | 136 | 26 | 45802 | -0.36409 | 0.64211 | *ANKRD22* |

| | | | | | | | | | |
|---|---|---|---|---|---|---|---|---|---|
| ENSG00000109072 | 17 | 26681290 | 26710110 | 60 | 7 | 30758 | -0.36419 | 0.64214 | *VTN* |
| ENSG00000223510 | 17 | 14128990 | 14150179 | 44 | 9 | 46630 | -0.36427 | 0.64217 | *CDRT15* |
| ENSG00000126698 | 1 | 28515967 | 28569536 | 177 | 15 | 41860 | -0.36476 | 0.64235 | *DNAJC8* |
| ENSG00000136098 | 13 | 52696775 | 52743996 | 142 | 19 | 41929 | -0.36489 | 0.6424 | *NEK3* |
| ENSG00000204928 | 5 | 145229296 | 145262531 | 93 | 26 | 36086 | -0.36501 | 0.64245 | *GRXCR2* |
| ENSG00000100565 | 14 | 77282715 | 77346645 | 235 | 48 | 51432 | -0.36508 | 0.64247 | *C14orf166B* |
| ENSG00000100154 | 22 | 28364004 | 29085853 | 1511 | 82 | 51426 | -0.36507 | 0.64247 | *TTC28* |
| ENSG00000167046 | 20 | 61262071 | 61284656 | 46 | 12 | 38938 | -0.36514 | 0.6425 | *RP11-93B14.6* |
| ENSG00000164715 | 7 | 97726197 | 97848945 | 285 | 42 | 43777 | -0.36517 | 0.64251 | *LMTK2* |
| ENSG00000040199 | 16 | 71661738 | 71768604 | 249 | 23 | 41602 | -0.36517 | 0.64251 | *PHLPP2* |
| ENSG00000181924 | 11 | 73573712 | 73598033 | 74 | 12 | 49547 | -0.36519 | 0.64252 | *COA4* |
| ENSG00000198791 | 8 | 17076737 | 17114387 | 96 | 18 | 40427 | -0.36536 | 0.64258 | *CNOT7* |
| ENSG00000116199 | 1 | 178984939 | 179055697 | 256 | 26 | 51626 | -0.36538 | 0.64259 | *FAM20B* |
| ENSG00000253457 | 8 | 30486117 | 30513581 | 78 | 15 | 57016 | -0.36542 | 0.6426 | *SMIM18* |
| ENSG00000212993 | 8 | 128416535 | 128442311 | 111 | 16 | 50293 | -0.36559 | 0.64267 | *POU5F1B* |
| ENSG00000142528 | 19 | 50518971 | 50566671 | 107 | 17 | 40240 | -0.3656 | 0.64267 | *ZNF473* |
| ENSG00000160716 | 1 | 154530257 | 154562502 | 65 | 18 | 49782 | -0.36578 | 0.64273 | *CHRNB2* |
| ENSG00000137210 | 6 | 10737992 | 10862986 | 283 | 55 | 49520 | -0.36585 | 0.64276 | *TMEM14B* |
| ENSG00000156639 | 6 | 37777275 | 38132400 | 1114 | 49 | 45484 | -0.36589 | 0.64278 | *ZFAND3* |
| ENSG00000115942 | 2 | 201763696 | 201838403 | 110 | 16 | 47391 | -0.36595 | 0.6428 | *ORC2* |
| ENSG00000137804 | 15 | 41614892 | 41683248 | 169 | 19 | 41777 | -0.36623 | 0.6429 | *NUSAP1* |
| ENSG00000160877 | 19 | 13218917 | 13261955 | 86 | 18 | 47536 | -0.36626 | 0.64291 | *NACC1* |
| ENSG00000183706 | 15 | 22372382 | 22393507 | 5 | 3 | 43426 | -0.36627 | 0.64292 | *OR4N4* |
| ENSG00000093072 | 22 | 17650194 | 17712879 | 315 | 53 | 48672 | -0.3665 | 0.643 | *CECR1* |
| ENSG00000186925 | 21 | 31903854 | 31924183 | 46 | 5 | 57064 | -0.36668 | 0.64307 | *KRTAP19-6* |
| ENSG00000206047 | 8 | 6825171 | 6847602 | 37 | 12 | 49597 | -0.36708 | 0.64322 | *DEFA1* |
| ENSG00000107036 | 9 | 5619025 | 5786557 | 532 | 44 | 48406 | -0.36746 | 0.64336 | *KIAA1432* |
| ENSG00000169688 | 16 | 56675811 | 56697116 | 135 | 20 | 46440 | -0.36784 | 0.64351 | *MT1B* |
| ENSG00000115271 | 2 | 163165350 | 163238105 | 65 | 21 | 37179 | -0.36801 | 0.64357 | *GCA* |
| ENSG00000198643 | 3 | 58609673 | 58662575 | 234 | 34 | 43625 | -0.36829 | 0.64367 | *FAM3D* |
| ENSG00000183019 | 19 | 7731514 | 7754712 | 128 | 21 | 47650 | -0.36855 | 0.64377 | *C19orf59* |
| ENSG00000164850 | 7 | 1111844 | 1143451 | 165 | 29 | 46246 | -0.36893 | 0.64391 | *GPER1* |
| ENSG00000259649 | 15 | 81178532 | 81212118 | 51 | 21 | 46149 | -0.36904 | 0.64395 | *RP11-351M8.1* |
| ENSG00000187180 | 1 | 152637771 | 152659050 | 44 | 7 | 28195 | -0.36915 | 0.64399 | *LCE2C* |
| ENSG00000197403 | 1 | 158725466 | 158746492 | 120 | 18 | 35241 | -0.36926 | 0.64403 | *OR6N1* |
| ENSG00000204495 | 1 | 13437414 | 13462656 | 2 | 1 | 42816 | -0.37051 | 0.6445 | *PRAMEF13* |

| | | | | | | | | | |
|---|---|---|---|---|---|---|---|---|---|
| ENSG00000177752 | 4 | 44614086 | 44690573 | 316 | 24 | 41299 | -0.37063 | 0.64454 | *YIPF7* |
| ENSG00000133317 | 11 | 63263556 | 63294246 | 43 | 14 | 38204 | -0.37073 | 0.64458 | *LGALS12* |
| ENSG00000137869 | 15 | 51490254 | 51640807 | 445 | 52 | 37808 | -0.37124 | 0.64477 | *CYP19A1* |
| ENSG00000101200 | 20 | 3053202 | 3075370 | 40 | 10 | 37496 | -0.3713 | 0.64479 | *AVP* |
| ENSG00000121446 | 1 | 182409256 | 182539734 | 447 | 31 | 45260 | -0.37132 | 0.6448 | *RGSL1* |
| ENSG00000067082 | 10 | 3808188 | 3837473 | 129 | 21 | 46788 | -0.37146 | 0.64485 | *KLF6* |
| ENSG00000146574 | 7 | 6823765 | 6876401 | 42 | 17 | 44538 | -0.37164 | 0.64492 | *CCZ1B* |
| ENSG00000090097 | 3 | 51981470 | 52012426 | 41 | 14 | 32715 | -0.37166 | 0.64493 | *PCBP4* |
| ENSG00000123364 | 12 | 54322535 | 54350328 | 65 | 12 | 32407 | -0.37172 | 0.64495 | *HOXC13* |
| ENSG00000146386 | 6 | 139339819 | 139374439 | 118 | 22 | 37643 | -0.3718 | 0.64498 | *ABRACL* |
| ENSG00000138119 | 10 | 95056186 | 95252074 | 798 | 92 | 39465 | -0.37217 | 0.64512 | *MYOF* |
| ENSG00000104231 | 8 | 82603569 | 82655138 | 202 | 20 | 38606 | -0.37245 | 0.64522 | *ZFAND1* |
| ENSG00000092929 | 17 | 73813306 | 73850798 | 117 | 19 | 34039 | -0.37255 | 0.64526 | *UNC13D* |
| ENSG00000234631 | 10 | 115664530 | 115686949 | 31 | 14 | 49062 | -0.37268 | 0.64531 | *AL162407.1* |
| ENSG00000112893 | 5 | 109015067 | 109215326 | 793 | 53 | 41768 | -0.37314 | 0.64548 | *MAN2A1* |
| ENSG00000222012 | 7 | 157396715 | 157421457 | 68 | 16 | 40478 | -0.3736 | 0.64565 | *AC005481.5* |
| ENSG00000160584 | 11 | 116704118 | 116979153 | 1056 | 48 | 36861 | -0.37366 | 0.64567 | *SIK3* |
| ENSG00000100109 | 22 | 26877191 | 26918471 | 153 | 32 | 46002 | -0.37379 | 0.64572 | *TFIP11* |
| ENSG00000070190 | 4 | 100727990 | 100801311 | 265 | 35 | 43228 | -0.3738 | 0.64573 | *DAPP1* |
| ENSG00000069345 | 16 | 46979299 | 47017699 | 48 | 9 | 34519 | -0.37388 | 0.64575 | *DNAJA2* |
| ENSG00000143207 | 1 | 175903967 | 176186629 | 865 | 62 | 42570 | -0.37398 | 0.64579 | *RFWD2* |
| ENSG00000230453 | 9 | 33514392 | 33583007 | 232 | 37 | 48051 | -0.37421 | 0.64588 | *ANKRD18B* |
| ENSG00000178163 | 4 | 10431498 | 10469034 | 111 | 26 | 44752 | -0.37428 | 0.6459 | *ZNF518B* |
| ENSG00000082212 | 18 | 48395419 | 48484691 | 212 | 21 | 47065 | -0.37426 | 0.6459 | *ME2* |
| ENSG00000165966 | 12 | 41572250 | 41978392 | 1539 | 93 | 44010 | -0.37432 | 0.64592 | *PDZRN4* |
| ENSG00000162761 | 1 | 165161104 | 165335952 | 613 | 96 | 48937 | -0.37435 | 0.64593 | *LMX1A* |
| ENSG00000164675 | 7 | 123082454 | 123185131 | 197 | 22 | 55954 | -0.3744 | 0.64595 | *IQUB* |
| ENSG00000177243 | 8 | 7276410 | 7297870 | 2 | 1 | 42795 | -0.37444 | 0.64596 | *DEFB103B* |
| ENSG00000106128 | 7 | 30968284 | 31042869 | 280 | 48 | 37354 | -0.37446 | 0.64597 | *GHRHR* |
| ENSG00000185873 | 4 | 69082371 | 69121438 | 82 | 7 | 33394 | -0.37449 | 0.64598 | *TMPRSS11B* |
| ENSG00000236609 | 7 | 6645248 | 6673921 | 106 | 22 | 44129 | -0.37485 | 0.64612 | *ZNF853* |
| ENSG00000090932 | 19 | 39979535 | 40009121 | 86 | 13 | 47291 | -0.37514 | 0.64622 | *DLL3* |
| ENSG00000144792 | 3 | 44609715 | 44651186 | 92 | 14 | 35255 | -0.37527 | 0.64627 | *ZNF660* |
| ENSG00000172346 | 22 | 41946767 | 41983745 | 69 | 9 | 49262 | -0.37551 | 0.64636 | *CSDC2* |
| ENSG00000176973 | 11 | 65329820 | 65351669 | 39 | 10 | 47798 | -0.37564 | 0.64641 | *FAM89B* |
| ENSG00000188493 | 19 | 41236761 | 41267458 | 89 | 16 | 53371 | -0.37573 | 0.64644 | *C19orf54* |

| | | | | | | | | | |
|---|---|---|---|---|---|---|---|---|---|
| ENSG00000101665 | 18 | 46436223 | 46487081 | 160 | 44 | 41269 | -0.37585 | 0.64649 | *SMAD7* |
| ENSG00000137699 | 11 | 119971983 | 120066237 | 260 | 40 | 38345 | -0.37589 | 0.6465 | *TRIM29* |
| ENSG00000151748 | 14 | 51088776 | 51145049 | 168 | 19 | 44703 | -0.37594 | 0.64652 | *SAV1* |
| ENSG00000130816 | 19 | 10234021 | 10351962 | 283 | 39 | 45756 | -0.37602 | 0.64655 | *DNMT1* |
| ENSG00000188037 | 7 | 143003219 | 143059176 | 136 | 30 | 50733 | -0.3761 | 0.64658 | *CLCN1* |
| ENSG00000015520 | 7 | 44542134 | 44590914 | 104 | 20 | 52413 | -0.37612 | 0.64659 | *NPC1L1* |
| ENSG00000073067 | 7 | 1012835 | 1039276 | 111 | 24 | 46220 | -0.37628 | 0.64664 | *CYP2W1* |
| ENSG00000170385 | 1 | 211734910 | 211762084 | 75 | 14 | 47406 | -0.37639 | 0.64669 | *SLC30A1* |
| ENSG00000180138 | 13 | 37667398 | 37689803 | 98 | 8 | 45846 | -0.37647 | 0.64672 | *CSNK1A1L* |
| ENSG00000256223 | 12 | 133697161 | 133746051 | 139 | 14 | 49528 | -0.37656 | 0.64675 | *ZNF10* |
| ENSG00000141699 | 17 | 40721531 | 40772641 | 107 | 13 | 52961 | -0.37664 | 0.64678 | *FAM134C* |
| ENSG00000112309 | 6 | 71556382 | 71676741 | 367 | 51 | 48836 | -0.37689 | 0.64687 | *B3GAT2* |
| ENSG00000153294 | 6 | 47643600 | 47699757 | 252 | 11 | 52386 | -0.37705 | 0.64693 | *GPR115* |
| ENSG00000143633 | 1 | 231349509 | 231386933 | 116 | 23 | 50121 | -0.37718 | 0.64698 | *C1orf131* |
| ENSG00000188177 | 2 | 113023171 | 113107640 | 204 | 17 | 34864 | -0.37727 | 0.64701 | *ZC3H6* |
| ENSG00000188452 | 2 | 182391403 | 182555392 | 490 | 42 | 38115 | -0.37739 | 0.64706 | *CERKL* |
| ENSG00000071246 | 14 | 77218532 | 77259354 | 130 | 38 | 49560 | -0.37757 | 0.64713 | *VASH1* |
| ENSG00000065427 | 16 | 75651622 | 75692541 | 117 | 19 | 34435 | -0.37762 | 0.64714 | *KARS* |
| ENSG00000166233 | 15 | 72756667 | 72889692 | 154 | 26 | 52261 | -0.37813 | 0.64733 | *ARIH1* |
| ENSG00000100505 | 14 | 51431980 | 51572779 | 615 | 83 | 42693 | -0.3782 | 0.64736 | *TRIM9* |
| ENSG00000138867 | 22 | 24926406 | 24961903 | 118 | 17 | 33386 | -0.37822 | 0.64737 | *GUCD1* |
| ENSG00000263417 | 18 | 68287755 | 68327980 | 152 | 24 | 45485 | -0.37838 | 0.64742 | *GTSCR1* |
| ENSG00000125775 | 20 | 1280619 | 1319883 | 149 | 37 | 44829 | -0.37855 | 0.64749 | *SDCBP2* |
| ENSG00000171606 | 19 | 58684396 | 58734928 | 157 | 26 | 46437 | -0.37931 | 0.64777 | *ZNF274* |
| ENSG00000170270 | 14 | 93659239 | 93683439 | 73 | 14 | 50396 | -0.37936 | 0.64779 | *C14orf142* |
| ENSG00000182810 | 16 | 68045179 | 68067770 | 31 | 9 | 49527 | -0.37948 | 0.64783 | *DDX28* |
| ENSG00000063046 | 12 | 53389942 | 53445993 | 145 | 11 | 25282 | -0.3795 | 0.64784 | *EIF4B* |
| ENSG00000085831 | 1 | 51742930 | 51820788 | 165 | 35 | 51330 | -0.37963 | 0.64789 | *TTC39A* |
| ENSG00000177575 | 12 | 7613409 | 7666489 | 124 | 26 | 41274 | -0.37965 | 0.6479 | *CD163* |
| ENSG00000166851 | 16 | 23678977 | 23711688 | 107 | 12 | 38106 | -0.37975 | 0.64794 | *PLK1* |
| ENSG00000169717 | 1 | 2928046 | 2949465 | 62 | 9 | 48604 | -0.37984 | 0.64797 | *ACTRT2* |
| ENSG00000108829 | 17 | 48442420 | 48484914 | 144 | 17 | 48748 | -0.37992 | 0.648 | *LRRC59* |
| ENSG00000100084 | 22 | 19308221 | 19445224 | 304 | 36 | 36928 | -0.37992 | 0.648 | *HIRA* |
| ENSG00000146263 | 6 | 97580037 | 97741093 | 360 | 26 | 36694 | -0.38025 | 0.64812 | *MMS22L* |
| ENSG00000062725 | 17 | 58510520 | 58613580 | 74 | 37 | 46657 | -0.38042 | 0.64818 | *APPBP2* |
| ENSG00000108823 | 17 | 48231575 | 48263292 | 79 | 19 | 42240 | -0.38062 | 0.64826 | *SGCA* |

| | | | | | | | | | |
|---|---|---|---|---|---|---|---|---|---|
| ENSG00000213741 | 14 | 50033390 | 50075408 | 108 | 11 | 44532 | -0.38073 | 0.6483 | *RPS29* |
| ENSG00000260869 | 16 | 30535548 | 30579600 | 52 | 19 | 35110 | -0.38101 | 0.6484 | *AC002310.13* |
| ENSG00000180758 | 1 | 9150364 | 9199229 | 143 | 24 | 50494 | -0.38109 | 0.64843 | *GPR157* |
| ENSG00000173404 | 20 | 20338765 | 20361590 | 38 | 14 | 41923 | -0.38114 | 0.64845 | *INSM1* |
| ENSG00000169679 | 2 | 111385275 | 111445691 | 68 | 16 | 37096 | -0.38122 | 0.64848 | *BUB1* |
| ENSG00000180483 | 20 | 29954967 | 29988406 | 98 | 18 | 43262 | -0.38125 | 0.64849 | *DEFB119* |
| ENSG00000177125 | 9 | 129612944 | 129658157 | 91 | 24 | 43138 | -0.38126 | 0.6485 | *ZBTB34* |
| ENSG00000183077 | 17 | 76173398 | 76213782 | 215 | 46 | 39536 | -0.3814 | 0.64855 | *AFMID* |
| ENSG00000127083 | 9 | 95166527 | 95196743 | 86 | 13 | 46490 | -0.38164 | 0.64864 | *OMD* |
| ENSG00000268351 | 11 | 69577254 | 69597718 | 80 | 22 | 42692 | -0.38219 | 0.64884 | *AP001888.1* |
| ENSG00000259066 | 14 | 93641318 | 93694932 | 174 | 30 | 50844 | -0.38223 | 0.64885 | *RP11-371E8.4* |
| ENSG00000205334 | 2 | 27918653 | 27948599 | 21 | 8 | 44798 | -0.38232 | 0.64889 | *AC074091.13* |
| ENSG00000119737 | 2 | 54070050 | 54097126 | 85 | 15 | 39859 | -0.38265 | 0.64901 | *GPR75* |
| ENSG00000162616 | 1 | 78434859 | 78493648 | 172 | 28 | 43943 | -0.38267 | 0.64902 | *DNAJB4* |
| ENSG00000117222 | 1 | 205045270 | 205101143 | 172 | 21 | 26805 | -0.38274 | 0.64904 | *RBBP5* |
| ENSG00000197933 | 19 | 11822080 | 11859824 | 90 | 16 | 36154 | -0.38284 | 0.64908 | *ZNF823* |
| ENSG00000059145 | 16 | 1403206 | 1474752 | 199 | 29 | 50954 | -0.38319 | 0.64921 | *UNKL* |
| ENSG00000272921 | 12 | 4704110 | 4781809 | 293 | 37 | 38447 | -0.38327 | 0.64924 | *RP11-500M8.7* |
| ENSG00000213639 | 2 | 28964506 | 29035806 | 161 | 15 | 44887 | -0.38332 | 0.64926 | *PPP1CB* |
| ENSG00000188263 | 22 | 50422942 | 50461088 | 145 | 23 | 52441 | -0.38348 | 0.64932 | *IL17REL* |
| ENSG00000177182 | 8 | 61959717 | 62424204 | 1416 | 118 | 37417 | -0.38377 | 0.64942 | *CLVS1* |
| ENSG00000181467 | 3 | 152870029 | 152896265 | 31 | 15 | 46722 | -0.38377 | 0.64943 | *RAP2B* |
| ENSG00000205678 | 4 | 65130975 | 65285186 | 598 | 36 | 35200 | -0.38392 | 0.64948 | *TECRL* |
| ENSG00000135824 | 1 | 182605239 | 182663711 | 155 | 23 | 41102 | -0.38416 | 0.64957 | *RGS8* |
| ENSG00000152465 | 10 | 15134583 | 15220692 | 340 | 49 | 39554 | -0.38415 | 0.64957 | *NMT2* |
| ENSG00000151779 | 2 | 15297032 | 15711454 | 1043 | 50 | 28835 | -0.38422 | 0.64959 | *NBAS* |
| ENSG00000269891 | 10 | 98747795 | 99062413 | 974 | 123 | 41783 | -0.38432 | 0.64963 | *ARHGAP19-SLIT1* |
| ENSG00000106477 | 7 | 130023612 | 130092274 | 168 | 34 | 52984 | -0.38434 | 0.64964 | *CEP41* |
| ENSG00000163249 | 2 | 208566264 | 208636563 | 190 | 12 | 29775 | -0.38448 | 0.64969 | *CCNYL1* |
| ENSG00000197008 | 7 | 64244766 | 64304054 | 261 | 22 | 45881 | -0.3845 | 0.6497 | *ZNF138* |
| ENSG00000154153 | 5 | 16463147 | 16627167 | 512 | 80 | 50400 | -0.38469 | 0.64977 | *FAM134B* |
| ENSG00000039123 | 5 | 54593588 | 54731409 | 354 | 14 | 36163 | -0.38473 | 0.64978 | *SKIV2L2* |
| ENSG00000109083 | 17 | 26645352 | 26672515 | 38 | 10 | 45467 | -0.38476 | 0.64979 | *IFT20* |
| ENSG00000168785 | 4 | 99381518 | 99589780 | 626 | 96 | 43649 | -0.38477 | 0.6498 | *TSPAN5* |
| ENSG00000155744 | 2 | 201828441 | 201946394 | 232 | 26 | 45347 | -0.38489 | 0.64984 | *FAM126B* |
| ENSG00000136448 | 17 | 43118978 | 43196384 | 266 | 28 | 52386 | -0.38488 | 0.64984 | *NMT1* |

| | | | | | | | | | |
|---|---|---|---|---|---|---|---|---|---|
| ENSG00000105323 | 19 | 41758401 | 41823503 | 116 | 15 | 54134 | -0.38525 | 0.64997 | *HNRNPUL1* |
| ENSG00000184731 | 2 | 28814 | 56870 | 72 | 13 | 28239 | -0.38526 | 0.64998 | *FAM110C* |
| ENSG00000167394 | 16 | 31062164 | 31095641 | 61 | 9 | 21755 | -0.38526 | 0.64998 | *ZNF668* |
| ENSG00000197102 | 14 | 102420865 | 102527129 | 273 | 30 | 35702 | -0.38541 | 0.65003 | *DYNC1H1* |
| ENSG00000122012 | 5 | 75368997 | 75659764 | 1026 | 115 | 45597 | -0.38575 | 0.65016 | *SV2C* |
| ENSG00000121864 | 3 | 179030779 | 179063323 | 81 | 16 | 52312 | -0.38616 | 0.65031 | *ZNF639* |
| ENSG00000151883 | 5 | 49951733 | 50152356 | 415 | 34 | 45231 | -0.38641 | 0.6504 | *PARP8* |
| ENSG00000173575 | 15 | 93416526 | 93581237 | 515 | 58 | 41190 | -0.38641 | 0.6504 | *CHD2* |
| ENSG00000107625 | 10 | 70651034 | 70716603 | 216 | 28 | 48964 | -0.38687 | 0.65057 | *DDX50* |
| ENSG00000004846 | 7 | 20644830 | 20826658 | 650 | 75 | 47242 | -0.38697 | 0.65061 | *ABCB5* |
| ENSG00000171860 | 12 | 8200898 | 8229067 | 58 | 18 | 44330 | -0.38698 | 0.65061 | *C3AR1* |
| ENSG00000128000 | 19 | 40524167 | 40572116 | 107 | 9 | 45572 | -0.38704 | 0.65064 | *ZNF780B* |
| ENSG00000175911 | 17 | 78967135 | 78988932 | 117 | 18 | 38868 | -0.38736 | 0.65076 | *AC127496.1* |
| ENSG00000258986 | 14 | 104931015 | 105081984 | 527 | 62 | 32702 | -0.38748 | 0.6508 | *TMEM179* |
| ENSG00000111729 | 12 | 8266228 | 8301203 | 56 | 14 | 36004 | -0.38762 | 0.65085 | *CLEC4A* |
| ENSG00000126804 | 14 | 64960430 | 65010408 | 160 | 15 | 56755 | -0.38762 | 0.65085 | *ZBTB1* |
| ENSG00000081052 | 2 | 227857427 | 228038829 | 612 | 63 | 45707 | -0.38777 | 0.65091 | *COL4A4* |
| ENSG00000141956 | 21 | 43208385 | 43309591 | 553 | 74 | 47290 | -0.38805 | 0.65101 | *PRDM15* |
| ENSG00000140262 | 15 | 57200821 | 57601479 | 1144 | 44 | 36739 | -0.38808 | 0.65102 | *TCF12* |
| ENSG00000100568 | 14 | 68103792 | 68151548 | 115 | 17 | 37303 | -0.3881 | 0.65103 | *VTI1B* |
| ENSG00000184544 | 17 | 9664751 | 9704614 | 186 | 32 | 48238 | -0.38819 | 0.65106 | *DHRS7C* |
| ENSG00000110169 | 11 | 6442279 | 6473847 | 60 | 10 | 53085 | -0.38831 | 0.65111 | *HPX* |
| ENSG00000167183 | 17 | 46019333 | 46045244 | 84 | 17 | 49915 | -0.38845 | 0.65116 | *PRR15L* |
| ENSG00000108669 | 17 | 76660130 | 76788379 | 322 | 30 | 38368 | -0.38858 | 0.65121 | *CYTH1* |
| ENSG00000269018 | 11 | 65348665 | 65369947 | 53 | 10 | 52101 | -0.38931 | 0.65148 | *AP001362.1* |
| ENSG00000168890 | 2 | 85815671 | 85840319 | 45 | 12 | 37984 | -0.38947 | 0.65154 | *TMEM150A* |
| ENSG00000217555 | 16 | 66576466 | 66610154 | 82 | 7 | 22267 | -0.38971 | 0.65162 | *CKLF* |
| ENSG00000236398 | 7 | 142870512 | 142891528 | 35 | 8 | 29442 | -0.3898 | 0.65166 | *TAS2R39* |
| ENSG00000163161 | 2 | 128004866 | 128061752 | 152 | 26 | 51929 | -0.38985 | 0.65168 | *ERCC3* |
| ENSG00000198833 | 6 | 90026344 | 90072567 | 117 | 30 | 46381 | -0.39008 | 0.65176 | *UBE2J1* |
| ENSG00000125107 | 16 | 58543855 | 58673790 | 444 | 27 | 38400 | -0.39027 | 0.65183 | *CNOT1* |
| ENSG00000187033 | 3 | 169619360 | 169666963 | 155 | 23 | 52340 | -0.39052 | 0.65192 | *SAMD7* |
| ENSG00000175787 | 9 | 97011593 | 97073736 | 257 | 32 | 44354 | -0.39066 | 0.65198 | *ZNF169* |
| ENSG00000070031 | 11 | 616431 | 637143 | 85 | 17 | 45106 | -0.39075 | 0.65201 | *SCT* |
| ENSG00000188921 | 9 | 20985306 | 21041635 | 201 | 23 | 53313 | -0.39086 | 0.65205 | *PTPLAD2* |
| ENSG00000119723 | 14 | 74406629 | 74440373 | 132 | 17 | 38823 | -0.391 | 0.6521 | *COQ6* |

| | | | | | | | | | |
|---|---|---|---|---|---|---|---|---|---|
| ENSG00000089737 | 14 | 94507266 | 94557591 | 166 | 17 | 43087 | -0.39129 | 0.65221 | *DDX24* |
| ENSG00000168062 | 11 | 64745415 | 64774517 | 64 | 15 | 42217 | -0.39146 | 0.65227 | *BATF2* |
| ENSG00000198722 | 9 | 35151999 | 35415335 | 427 | 43 | 49510 | -0.39194 | 0.65245 | *UNC13B* |
| ENSG00000155893 | 3 | 140937568 | 141023748 | 314 | 42 | 33348 | -0.39199 | 0.65247 | *ACPL2* |
| ENSG00000223547 | 19 | 12165514 | 12202380 | 96 | 11 | 28958 | -0.39199 | 0.65247 | *ZNF844* |
| ENSG00000268083 | 19 | 39298111 | 39340818 | 114 | 20 | 40159 | -0.39203 | 0.65248 | *AC104534.3* |
| ENSG00000119321 | 9 | 115913286 | 115993641 | 371 | 37 | 51541 | -0.3922 | 0.65254 | *FKBP15* |
| ENSG00000070808 | 5 | 149589054 | 149679854 | 232 | 46 | 41270 | -0.39255 | 0.65267 | *CAMK2A* |
| ENSG00000130304 | 19 | 17569578 | 17626977 | 167 | 30 | 43691 | -0.39265 | 0.65271 | *SLC27A1* |
| ENSG00000109265 | 4 | 57026361 | 57204791 | 629 | 117 | 45678 | -0.39269 | 0.65273 | *KIAA1211* |
| ENSG00000251695 | 12 | 69176125 | 69197743 | 58 | 20 | 39388 | -0.39284 | 0.65278 | *AC124890.1* |
| ENSG00000121542 | 3 | 122910774 | 123002977 | 358 | 11 | 52977 | -0.39307 | 0.65287 | *SEC22A* |
| ENSG00000223874 | 2 | 217725495 | 217746362 | 69 | 14 | 39716 | -0.39318 | 0.65291 | *AC007557.1* |
| ENSG00000100426 | 22 | 50237490 | 50293726 | 220 | 15 | 27851 | -0.39322 | 0.65292 | *ZBED4* |
| ENSG00000176381 | 6 | 166709168 | 166731936 | 108 | 14 | 47683 | -0.39337 | 0.65298 | *PRR18* |
| ENSG00000181616 | 11 | 5555719 | 5576779 | 124 | 6 | 22586 | -0.39346 | 0.65301 | *OR52H1* |
| ENSG00000143742 | 1 | 225955531 | 225988168 | 140 | 24 | 48597 | -0.39461 | 0.65343 | *SRP9* |
| ENSG00000006606 | 7 | 75388851 | 75429214 | 169 | 23 | 49429 | -0.39461 | 0.65343 | *CCL26* |
| ENSG00000164938 | 8 | 95928200 | 95971639 | 152 | 17 | 48258 | -0.39461 | 0.65344 | *TP53INP1* |
| ENSG00000105939 | 7 | 138718266 | 138804465 | 289 | 38 | 42269 | -0.39467 | 0.65346 | *ZC3HAV1* |
| ENSG00000075388 | 11 | 69577797 | 69600171 | 94 | 23 | 42521 | -0.39514 | 0.65363 | *FGF4* |
| ENSG00000126337 | 17 | 39632388 | 39658798 | 94 | 17 | 54595 | -0.39531 | 0.65369 | *KRT36* |
| ENSG00000187556 | 19 | 13962877 | 14001571 | 88 | 16 | 37969 | -0.39532 | 0.6537 | *NANOS3* |
| ENSG00000174007 | 3 | 196423148 | 196449164 | 111 | 17 | 47496 | -0.39536 | 0.65371 | *CEP19* |
| ENSG00000205922 | 19 | 1742372 | 1790987 | 137 | 39 | 45633 | -0.39535 | 0.65371 | *ONECUT3* |
| ENSG00000121766 | 1 | 31759842 | 31847783 | 245 | 19 | 32375 | -0.39561 | 0.65381 | *ZCCHC17* |
| ENSG00000182330 | 1 | 13376646 | 13400765 | 4 | 2 | 35118 | -0.39587 | 0.6539 | *PRAMEF8* |
| ENSG00000114279 | 3 | 191847184 | 192495553 | 1937 | 222 | 41549 | -0.3963 | 0.65406 | *FGF12* |
| ENSG00000144488 | 2 | 238998798 | 239051928 | 273 | 40 | 43557 | -0.39636 | 0.65408 | *ESPNL* |
| ENSG00000127324 | 12 | 71508865 | 71845678 | 1088 | 85 | 48531 | -0.39655 | 0.65415 | *TSPAN8* |
| ENSG00000181284 | 17 | 7328762 | 7350998 | 42 | 13 | 42409 | -0.39691 | 0.65428 | *TMEM102* |
| ENSG00000236669 | 7 | 157308477 | 157330019 | 89 | 13 | 47442 | -0.39693 | 0.65429 | *AC006372.1* |
| ENSG00000090554 | 19 | 49967464 | 49999488 | 100 | 15 | 31426 | -0.39696 | 0.6543 | *FLT3LG* |
| ENSG00000134438 | 18 | 56924267 | 56951318 | 124 | 15 | 50077 | -0.39711 | 0.65436 | *RAX* |
| ENSG00000078900 | 1 | 3559084 | 3662765 | 352 | 77 | 41291 | -0.3972 | 0.65439 | *TP73* |
| ENSG00000149534 | 11 | 59845734 | 59873444 | 95 | 15 | 36297 | -0.39722 | 0.6544 | *MS4A2* |

| | | | | | | | | | |
|---|---|---|---|---|---|---|---|---|---|
| ENSG00000127129 | 1 | 41934446 | 41960344 | 100 | 23 | 51664 | -0.39731 | 0.65443 | *EDN2* |
| ENSG00000068024 | 2 | 239959864 | 240333348 | 1347 | 160 | 45576 | -0.39737 | 0.65445 | *HDAC4* |
| ENSG00000157613 | 11 | 46289212 | 46352972 | 122 | 25 | 43237 | -0.39743 | 0.65448 | *CREB3L1* |
| ENSG00000196188 | 1 | 206307459 | 206342104 | 12 | 6 | 37665 | -0.39754 | 0.65451 | *CTSE* |
| ENSG00000114491 | 3 | 124439213 | 124474040 | 178 | 18 | 53752 | -0.39771 | 0.65458 | *UMPS* |
| ENSG00000126803 | 14 | 64992623 | 65019955 | 99 | 15 | 56436 | -0.39782 | 0.65462 | *HSPA2* |
| ENSG00000104356 | 8 | 99119525 | 99182062 | 282 | 36 | 50453 | -0.39789 | 0.65465 | *POP1* |
| ENSG00000177688 | 6 | 149711495 | 149732177 | 74 | 12 | 40532 | -0.39803 | 0.6547 | *SUMO4* |
| ENSG00000187607 | 17 | 15592891 | 15650874 | 159 | 27 | 42565 | -0.39806 | 0.65471 | *ZNF286A* |
| ENSG00000236320 | 17 | 33865144 | 33895117 | 143 | 25 | 45505 | -0.39819 | 0.65476 | *SLFN14* |
| ENSG00000138109 | 10 | 96688415 | 96759147 | 291 | 23 | 55875 | -0.39828 | 0.65479 | *CYP2C9* |
| ENSG00000134962 | 4 | 39398473 | 39463156 | 254 | 42 | 48382 | -0.39841 | 0.65484 | *KLB* |
| ENSG00000187175 | 21 | 46091491 | 46112078 | 186 | 15 | 56239 | -0.39845 | 0.65485 | *KRTAP12-1* |
| ENSG00000196152 | 9 | 130176661 | 130217651 | 98 | 8 | 54538 | -0.39864 | 0.65492 | *ZNF79* |
| ENSG00000163319 | 4 | 84367085 | 84400888 | 84 | 10 | 35458 | -0.39866 | 0.65493 | *MRPS18C* |
| ENSG00000171102 | 9 | 136070664 | 136094630 | 100 | 15 | 38260 | -0.39886 | 0.655 | *OBP2B* |
| ENSG00000162390 | 1 | 54997930 | 55114865 | 471 | 76 | 42552 | -0.39913 | 0.6551 | *ACOT11* |
| ENSG00000140854 | 16 | 57759642 | 57801162 | 129 | 29 | 44278 | -0.39938 | 0.65519 | *KATNB1* |
| ENSG00000104529 | 8 | 144651867 | 144691711 | 169 | 20 | 38992 | -0.39961 | 0.65528 | *EEF1D* |
| ENSG00000100307 | 22 | 39506172 | 39558679 | 197 | 26 | 43725 | -0.39972 | 0.65532 | *CBX7* |
| ENSG00000164190 | 5 | 36866861 | 37076515 | 313 | 40 | 47111 | -0.39983 | 0.65536 | *NIPBL* |
| ENSG00000145681 | 5 | 82923624 | 83027432 | 302 | 53 | 45978 | -0.3999 | 0.65539 | *HAPLN1* |
| ENSG00000096006 | 6 | 49685097 | 49722150 | 112 | 17 | 48260 | -0.40053 | 0.65562 | *CRISP3* |
| ENSG00000070423 | 19 | 637526 | 673277 | 161 | 41 | 45587 | -0.40055 | 0.65563 | *RNF126* |
| ENSG00000144810 | 3 | 99347319 | 99528070 | 484 | 56 | 53259 | -0.4006 | 0.65564 | *COL8A1* |
| ENSG00000149124 | 11 | 58397899 | 58509447 | 429 | 45 | 44172 | -0.40062 | 0.65565 | *GLYAT* |
| ENSG00000164674 | 6 | 159061046 | 159195908 | 497 | 70 | 43031 | -0.40099 | 0.65579 | *SYTL3* |
| ENSG00000112562 | 6 | 168831831 | 169083984 | 1048 | 119 | 41561 | -0.40109 | 0.65582 | *SMOC2* |
| ENSG00000137818 | 15 | 69735123 | 69758255 | 27 | 8 | 26140 | -0.40111 | 0.65583 | *RPLP1* |
| ENSG00000165025 | 9 | 93554069 | 93670831 | 585 | 84 | 49552 | -0.40156 | 0.656 | *SYK* |
| ENSG00000178295 | 2 | 17925125 | 17976632 | 143 | 21 | 40892 | -0.40172 | 0.65606 | *GEN1* |
| ENSG00000130544 | 19 | 7059455 | 7097979 | 128 | 26 | 45629 | -0.402 | 0.65616 | *ZNF557* |
| ENSG00000198818 | 6 | 166723216 | 166766079 | 119 | 13 | 51407 | -0.40212 | 0.6562 | *SFT2D1* |
| ENSG00000198892 | 1 | 201847808 | 201871434 | 44 | 13 | 52156 | -0.40219 | 0.65623 | *SHISA4* |
| ENSG00000176903 | 14 | 74168494 | 74191128 | 84 | 18 | 34432 | -0.40222 | 0.65624 | *PNMA1* |
| ENSG00000089177 | 20 | 16242749 | 16564078 | 1312 | 116 | 48723 | -0.40245 | 0.65632 | *KIF16B* |

| | | | | | | | | | |
|---|---|---|---|---|---|---|---|---|---|
| ENSG00000204566 | 10 | 23482745 | 23538745 | 80 | 19 | 45347 | -0.40255 | 0.65636 | *C10orf115* |
| ENSG00000101445 | 20 | 37424348 | 37561667 | 377 | 61 | 54809 | -0.40271 | 0.65642 | *PPP1R16B* |
| ENSG00000151353 | 2 | 657335 | 687439 | 104 | 19 | 44932 | -0.40293 | 0.6565 | *TMEM18* |
| ENSG00000151092 | 3 | 25750435 | 25841530 | 103 | 18 | 43247 | -0.40325 | 0.65662 | *NGLY1* |
| ENSG00000197410 | 4 | 155143399 | 155422930 | 880 | 103 | 42986 | -0.40359 | 0.65674 | *DCHS2* |
| ENSG00000185272 | 21 | 15578451 | 15610693 | 229 | 21 | 53166 | -0.40367 | 0.65677 | *RBM11* |
| ENSG00000196381 | 19 | 38148652 | 38193223 | 111 | 20 | 36491 | -0.4037 | 0.65678 | *ZNF781* |
| ENSG00000269515 | 10 | 44863687 | 44888739 | 120 | 20 | 53878 | -0.40387 | 0.65685 | *AL137026.1* |
| ENSG00000179774 | 10 | 69980386 | 70001871 | 83 | 18 | 40983 | -0.40389 | 0.65685 | *ATOH7* |
| ENSG00000151229 | 12 | 40138823 | 40509891 | 1085 | 102 | 49808 | -0.4039 | 0.65686 | *SLC2A13* |
| ENSG00000215595 | 20 | 1174098 | 1198918 | 106 | 16 | 46880 | -0.40392 | 0.65686 | *C20orf202* |
| ENSG00000172458 | 13 | 21266266 | 21307237 | 116 | 17 | 32856 | -0.40415 | 0.65695 | *IL17D* |
| ENSG00000185730 | 8 | 144361846 | 144390231 | 125 | 10 | 29711 | -0.40447 | 0.65707 | *ZNF696* |
| ENSG00000054277 | 1 | 241743404 | 241850678 | 372 | 83 | 48969 | -0.40451 | 0.65708 | *OPN3* |
| ENSG00000149090 | 11 | 35443370 | 35561848 | 447 | 65 | 47141 | -0.40454 | 0.65709 | *PAMR1* |
| ENSG00000180801 | 4 | 114811440 | 114910883 | 247 | 27 | 48144 | -0.40484 | 0.6572 | *ARSJ* |
| ENSG00000117519 | 1 | 95352507 | 95402834 | 185 | 39 | 51907 | -0.40489 | 0.65722 | *CNN3* |
| ENSG00000268553 | 5 | 172026245 | 172046436 | 70 | 23 | 42191 | -0.40548 | 0.65744 | *AC027309.1* |
| ENSG00000150051 | 10 | 27951804 | 28044989 | 223 | 30 | 46247 | -0.40572 | 0.65753 | *MKX* |
| ENSG00000007168 | 17 | 2486504 | 2598909 | 275 | 27 | 44300 | -0.40576 | 0.65754 | *PAFAH1B1* |
| ENSG00000126003 | 20 | 30770306 | 30805594 | 28 | 5 | 29754 | -0.40595 | 0.65761 | *PLAGL2* |
| ENSG00000168899 | 2 | 85801531 | 85830535 | 84 | 14 | 44027 | -0.40608 | 0.65766 | *VAMP5* |
| ENSG00000182315 | 19 | 7046220 | 7068651 | 19 | 11 | 48879 | -0.40607 | 0.65766 | *MBD3L3* |
| ENSG00000175264 | 11 | 45660427 | 45697172 | 115 | 19 | 41927 | -0.40621 | 0.65771 | *CHST1* |
| ENSG00000085982 | 2 | 234374166 | 234485428 | 238 | 18 | 50016 | -0.40659 | 0.65785 | *USP40* |
| ENSG00000215182 | 11 | 1141580 | 1232364 | 172 | 21 | 42403 | -0.40667 | 0.65787 | *MUC5AC* |
| ENSG00000206531 | 3 | 112524556 | 112575703 | 250 | 31 | 44980 | -0.40699 | 0.65799 | *CD200R1L* |
| ENSG00000256646 | 7 | 42938872 | 42981773 | 144 | 10 | 53788 | -0.40716 | 0.65805 | *PSMA2* |
| ENSG00000254206 | 16 | 29382675 | 29425350 | 47 | 18 | 38682 | -0.40728 | 0.6581 | *NPIPB11* |
| ENSG00000167257 | 11 | 117093341 | 117167161 | 213 | 33 | 43241 | -0.4074 | 0.65814 | *RNF214* |
| ENSG00000183010 | 17 | 79880260 | 79910288 | 63 | 12 | 44703 | -0.40751 | 0.65818 | *PYCR1* |
| ENSG00000138629 | 15 | 74728318 | 74763523 | 92 | 9 | 27721 | -0.40787 | 0.65832 | *UBL7* |
| ENSG00000073150 | 22 | 50599160 | 50628723 | 116 | 29 | 49900 | -0.40805 | 0.65838 | *PANX2* |
| ENSG00000004864 | 7 | 95739532 | 95961459 | 317 | 25 | 46667 | -0.4081 | 0.6584 | *SLC25A13* |
| ENSG00000149573 | 11 | 118114118 | 118145251 | 142 | 17 | 45491 | -0.40826 | 0.65846 | *MPZL2* |
| ENSG00000160710 | 1 | 154544538 | 154610475 | 148 | 16 | 49901 | -0.40845 | 0.65853 | *ADAR* |

| | | | | | | | | | |
|---|---|---|---|---|---|---|---|---|---|
| ENSG00000157985 | 2 | 236392733 | 237050444 | 1699 | 158 | 43861 | -0.4086 | 0.65858 | *AGAP1* |
| ENSG00000180998 | 14 | 53009866 | 53114431 | 277 | 37 | 47034 | -0.40898 | 0.65872 | *GPR137C* |
| ENSG00000109756 | 4 | 160015330 | 160291321 | 599 | 61 | 43131 | -0.40929 | 0.65884 | *RAPGEF2* |
| ENSG00000162510 | 1 | 31174124 | 31206434 | 141 | 13 | 38442 | -0.40943 | 0.65889 | *MATN1* |
| ENSG00000066135 | 1 | 44105829 | 44181186 | 146 | 20 | 36243 | -0.40956 | 0.65893 | *KDM4A* |
| ENSG00000188010 | 2 | 39093103 | 39166213 | 156 | 21 | 40238 | -0.40973 | 0.659 | *MORN2* |
| ENSG00000135778 | 1 | 233076351 | 233129628 | 151 | 14 | 54541 | -0.41015 | 0.65915 | *NTPCR* |
| ENSG00000222014 | 2 | 130727235 | 130750311 | 35 | 6 | 22864 | -0.41034 | 0.65922 | *RAB6C* |
| ENSG00000178502 | 17 | 39999797 | 40031684 | 103 | 9 | 52882 | -0.41044 | 0.65926 | *KLHL11* |
| ENSG00000176723 | 16 | 31435569 | 31464346 | 47 | 14 | 45399 | -0.41067 | 0.65934 | *ZNF843* |
| ENSG00000148377 | 10 | 1054847 | 1081799 | 119 | 23 | 49830 | -0.41105 | 0.65948 | *IDI2* |
| ENSG00000173559 | 2 | 192532794 | 192571385 | 82 | 18 | 52832 | -0.41152 | 0.65965 | *NABP1* |
| ENSG00000111319 | 12 | 6446009 | 6496896 | 127 | 46 | 38486 | -0.41155 | 0.65966 | *SCNN1A* |
| ENSG00000165621 | 13 | 97627973 | 97656984 | 82 | 20 | 41777 | -0.4116 | 0.65968 | *OXGR1* |
| ENSG00000145375 | 4 | 123834229 | 124250605 | 1563 | 68 | 33106 | -0.41172 | 0.65973 | *SPATA5* |
| ENSG00000233539 | 7 | 46703383 | 46809449 | 433 | 59 | 49990 | -0.41182 | 0.65977 | *AC011294.3* |
| ENSG00000167077 | 22 | 42085503 | 42205460 | 299 | 21 | 41715 | -0.41185 | 0.65978 | *MEI1* |
| ENSG00000109436 | 4 | 141531919 | 141687274 | 414 | 61 | 43468 | -0.41206 | 0.65985 | *TBC1D9* |
| ENSG00000184845 | 5 | 174857042 | 174881211 | 79 | 20 | 46816 | -0.41219 | 0.6599 | *DRD1* |
| ENSG00000012048 | 17 | 41186312 | 41287500 | 239 | 10 | 48613 | -0.41228 | 0.65993 | *BRCA1* |
| ENSG00000164073 | 4 | 128828960 | 128897150 | 64 | 24 | 43583 | -0.41263 | 0.66006 | *MFSD8* |
| ENSG00000267848 | 20 | 62575007 | 62595495 | 51 | 17 | 44806 | -0.41279 | 0.66012 | *AL118506.1* |
| ENSG00000186150 | 1 | 110645062 | 110666569 | 64 | 11 | 33427 | -0.41286 | 0.66014 | *UBL4B* |
| ENSG00000164822 | 8 | 6772215 | 6793598 | 130 | 31 | 48834 | -0.41312 | 0.66024 | *DEFA6* |
| ENSG00000173372 | 1 | 22952999 | 22976101 | 87 | 13 | 34891 | -0.41317 | 0.66026 | *C1QA* |
| ENSG00000148200 | 9 | 127269554 | 127543589 | 207 | 30 | 46691 | -0.4133 | 0.66031 | *NR6A1* |
| ENSG00000078081 | 3 | 182830001 | 182891627 | 282 | 33 | 44903 | -0.41358 | 0.66041 | *LAMP3* |
| ENSG00000164363 | 5 | 1215470 | 1256304 | 179 | 36 | 43458 | -0.41364 | 0.66043 | *SLC6A18* |
| ENSG00000125971 | 20 | 33094214 | 33138762 | 57 | 12 | 51702 | -0.41388 | 0.66052 | *DYNLRB1* |
| ENSG00000070614 | 5 | 149855381 | 149947773 | 233 | 29 | 40957 | -0.41439 | 0.66071 | *NDST1* |
| ENSG00000186723 | 19 | 15907817 | 15928936 | 105 | 15 | 46833 | -0.41453 | 0.66076 | *OR10H1* |
| ENSG00000134986 | 5 | 110988318 | 111343161 | 1021 | 98 | 37909 | -0.41468 | 0.66081 | *NREP* |
| ENSG00000170961 | 8 | 122614356 | 122663630 | 137 | 23 | 39762 | -0.41472 | 0.66083 | *HAS2* |
| ENSG00000186480 | 7 | 155079486 | 155111945 | 182 | 23 | 43580 | -0.41484 | 0.66087 | *INSIG1* |
| ENSG00000161682 | 17 | 42420583 | 42451243 | 47 | 17 | 47448 | -0.41495 | 0.66091 | *FAM171A2* |
| ENSG00000105255 | 19 | 4294597 | 4333840 | 150 | 32 | 51175 | -0.41512 | 0.66097 | *FSD1* |

| | | | | | | | | | |
|---|---|---|---|---|---|---|---|---|---|
| ENSG00000183576 | 14 | 99854083 | 99957216 | 164 | 15 | 49328 | -0.41516 | 0.66099 | *SETD3* |
| ENSG00000166987 | 12 | 57904493 | 57933931 | 35 | 5 | 48257 | -0.41566 | 0.66117 | *MBD6* |
| ENSG00000173068 | 9 | 16399501 | 16880841 | 1495 | 142 | 33981 | -0.41579 | 0.66122 | *BNC2* |
| ENSG00000138796 | 4 | 108900870 | 108966331 | 182 | 11 | 30291 | -0.41585 | 0.66124 | *HADH* |
| ENSG00000109501 | 4 | 6261576 | 6314992 | 259 | 20 | 51640 | -0.41627 | 0.66139 | *WFS1* |
| ENSG00000189090 | 10 | 47167204 | 47191681 | 1 | 1 | 42685 | -0.41629 | 0.6614 | *FAM25B* |
| ENSG00000163116 | 4 | 98095244 | 99074391 | 2945 | 86 | 49162 | -0.41644 | 0.66146 | *STPG2* |
| ENSG00000113319 | 5 | 80246491 | 80535975 | 991 | 154 | 50627 | -0.41665 | 0.66153 | *RASGRF2* |
| ENSG00000113492 | 5 | 34988206 | 35058198 | 265 | 40 | 45154 | -0.41672 | 0.66156 | *AGXT2* |
| ENSG00000219200 | 17 | 6905736 | 6927851 | 55 | 8 | 51848 | -0.41682 | 0.6616 | *RNASEK* |
| ENSG00000269871 | 17 | 6905928 | 6927755 | 55 | 8 | 51848 | -0.41682 | 0.6616 | *AC040977.1* |
| ENSG00000266967 | 17 | 41092543 | 41126515 | 32 | 10 | 34383 | -0.41694 | 0.66164 | *AARSD1* |
| ENSG00000134216 | 1 | 111823484 | 111873188 | 357 | 37 | 54181 | -0.41702 | 0.66167 | *CHIA* |
| ENSG00000108106 | 19 | 55902652 | 55929145 | 129 | 25 | 36849 | -0.41712 | 0.6617 | *UBE2S* |
| ENSG00000174574 | 1 | 39446895 | 39481731 | 131 | 16 | 36314 | -0.41748 | 0.66184 | *AKIRIN1* |
| ENSG00000154007 | 1 | 76374558 | 76408116 | 139 | 20 | 49073 | -0.41767 | 0.6619 | *ASB17* |
| ENSG00000132275 | 11 | 6606305 | 6634850 | 80 | 19 | 48379 | -0.41764 | 0.6619 | *RRP8* |
| ENSG00000169629 | 2 | 113117669 | 113202065 | 57 | 16 | 49096 | -0.41777 | 0.66194 | *RGPD8* |
| ENSG00000071205 | 4 | 148643214 | 149003931 | 683 | 92 | 34540 | -0.41785 | 0.66197 | *ARHGAP10* |
| ENSG00000187811 | 11 | 71282956 | 71328487 | 257 | 28 | 37517 | -0.41789 | 0.66198 | *AP000867.1* |
| ENSG00000187902 | 19 | 55930107 | 55964230 | 91 | 25 | 44613 | -0.41793 | 0.662 | *SHISA7* |
| ENSG00000140403 | 15 | 78546428 | 78584538 | 136 | 23 | 53493 | -0.41807 | 0.66205 | *DNAJA4* |
| ENSG00000197646 | 9 | 5500545 | 5581282 | 232 | 41 | 44583 | -0.41831 | 0.66214 | *PDCD1LG2* |
| ENSG00000156026 | 10 | 74441889 | 74657452 | 234 | 26 | 33534 | -0.4185 | 0.66221 | *MCU* |
| ENSG00000138623 | 15 | 74691630 | 74736808 | 130 | 21 | 29089 | -0.4185 | 0.66221 | *SEMA7A* |
| ENSG00000185344 | 12 | 124186865 | 124256302 | 257 | 21 | 45792 | -0.41855 | 0.66223 | *ATP6V0A2* |
| ENSG00000156853 | 16 | 30603879 | 30645333 | 43 | 7 | 25637 | -0.41903 | 0.6624 | *ZNF689* |
| ENSG00000016391 | 3 | 53836362 | 53890417 | 189 | 41 | 36247 | -0.41905 | 0.66241 | *CHDH* |
| ENSG00000154146 | 11 | 124599742 | 124627106 | 55 | 14 | 42296 | -0.41907 | 0.66242 | *NRGN* |
| ENSG00000101251 | 20 | 13819893 | 13987089 | 556 | 76 | 53465 | -0.41919 | 0.66246 | *SEL1L2* |
| ENSG00000119326 | 9 | 111694851 | 111785809 | 303 | 33 | 50347 | -0.41927 | 0.66249 | *CTNNAL1* |
| ENSG00000170848 | 19 | 43396231 | 43433715 | 101 | 11 | 24564 | -0.41948 | 0.66257 | *PSG6* |
| ENSG00000150787 | 11 | 112087088 | 112150678 | 221 | 11 | 51538 | -0.41973 | 0.66266 | *PTS* |
| ENSG00000142512 | 19 | 51903275 | 51931057 | 121 | 14 | 45766 | -0.41978 | 0.66268 | *SIGLEC10* |
| ENSG00000205307 | 7 | 100159855 | 100181270 | 47 | 11 | 51770 | -0.41994 | 0.66273 | *SAP25* |
| ENSG00000204152 | 10 | 51361390 | 51397768 | 4 | 2 | 27830 | -0.42004 | 0.66277 | *TIMM23B* |

| | | | | | | | | | |
|---|---|---|---|---|---|---|---|---|---|
| ENSG00000175691 | 19 | 2923216 | 2954969 | 107 | 33 | 43948 | -0.42031 | 0.66287 | *ZNF77* |
| ENSG00000137473 | 4 | 147617790 | 147877034 | 703 | 50 | 48265 | -0.42034 | 0.66288 | *TTC29* |
| ENSG00000065717 | 19 | 2987636 | 3057633 | 269 | 66 | 41871 | -0.42045 | 0.66292 | *TLE2* |
| ENSG00000205718 | 19 | 7027759 | 7050190 | 8 | 5 | 39290 | -0.42049 | 0.66294 | *MBD3L4* |
| ENSG00000169515 | 19 | 46903629 | 46926841 | 138 | 26 | 46227 | -0.4205 | 0.66294 | *CCDC8* |
| ENSG00000196312 | 9 | 99650348 | 99785862 | 134 | 27 | 38210 | -0.42071 | 0.66302 | *HIATL2* |
| ENSG00000100196 | 22 | 38854067 | 38889452 | 73 | 24 | 44288 | -0.42074 | 0.66303 | *KDELR3* |
| ENSG00000113269 | 5 | 179328651 | 179509118 | 512 | 59 | 43174 | -0.42124 | 0.66321 | *RNF130* |
| ENSG00000164808 | 8 | 48163167 | 48658868 | 469 | 59 | 39763 | -0.42144 | 0.66328 | *SPIDR* |
| ENSG00000140939 | 16 | 67194057 | 67219643 | 26 | 9 | 39878 | -0.42152 | 0.66331 | *NOL3* |
| ENSG00000188529 | 1 | 24281294 | 24317417 | 22 | 5 | 27811 | -0.4218 | 0.66342 | *SRSF10* |
| ENSG00000111802 | 6 | 24640205 | 24677261 | 155 | 25 | 47981 | -0.42202 | 0.66349 | *TDP2* |
| ENSG00000163848 | 3 | 124934405 | 125104198 | 366 | 18 | 49125 | -0.4221 | 0.66352 | *ZNF148* |
| ENSG00000178096 | 1 | 149849440 | 149882351 | 68 | 10 | 49373 | -0.42216 | 0.66355 | *BOLA1* |
| ENSG00000099260 | 1 | 100101499 | 100170097 | 207 | 31 | 38584 | -0.42242 | 0.66364 | *PALMD* |
| ENSG00000159593 | 16 | 66826778 | 66917159 | 170 | 18 | 41577 | -0.42251 | 0.66367 | *NAE1* |
| ENSG00000166845 | 18 | 51874287 | 51921588 | 216 | 13 | 50961 | -0.42263 | 0.66372 | *C18orf54* |
| ENSG00000172738 | 6 | 37169956 | 37235931 | 134 | 22 | 51904 | -0.42274 | 0.66376 | *TMEM217* |
| ENSG00000146090 | 5 | 179517795 | 179646153 | 598 | 118 | 47169 | -0.42295 | 0.66384 | *RASGEF1C* |
| ENSG00000244025 | 21 | 31853782 | 31874275 | 43 | 5 | 53807 | -0.42298 | 0.66385 | *KRTAP19-3* |
| ENSG00000163746 | 3 | 146099208 | 146223778 | 405 | 24 | 54419 | -0.42308 | 0.66388 | *PLSCR2* |
| ENSG00000184178 | 4 | 53729149 | 54242242 | 1421 | 120 | 45379 | -0.4231 | 0.66389 | *SCFD2* |
| ENSG00000153933 | 17 | 54901460 | 54956036 | 147 | 13 | 53252 | -0.42328 | 0.66396 | *DGKE* |
| ENSG00000151364 | 11 | 77716761 | 77767237 | 200 | 26 | 48670 | -0.4234 | 0.664 | *KCTD14* |
| ENSG00000160803 | 1 | 155995092 | 156033585 | 67 | 21 | 40478 | -0.42348 | 0.66403 | *UBQLN4* |
| ENSG00000187068 | 3 | 184785838 | 184880802 | 341 | 41 | 51091 | -0.42352 | 0.66404 | *C3orf70* |
| ENSG00000197150 | 7 | 150715510 | 150754869 | 109 | 26 | 54066 | -0.42357 | 0.66406 | *ABCB8* |
| ENSG00000083535 | 13 | 73346197 | 73600591 | 698 | 51 | 49606 | -0.42371 | 0.66411 | *PIBF1* |
| ENSG00000213213 | 9 | 139680802 | 139712193 | 90 | 14 | 53518 | -0.42375 | 0.66413 | *CCDC183* |
| ENSG00000108352 | 17 | 38323263 | 38361908 | 88 | 17 | 40109 | -0.42384 | 0.66416 | *RAPGEFL1* |
| ENSG00000171574 | 19 | 58902871 | 58939694 | 143 | 13 | 49156 | -0.42392 | 0.66419 | *ZNF584* |
| ENSG00000134899 | 13 | 103487194 | 103538345 | 126 | 20 | 46445 | -0.42412 | 0.66426 | *ERCC5* |
| ENSG00000170004 | 17 | 7778124 | 7826078 | 114 | 26 | 45166 | -0.42411 | 0.66426 | *CHD3* |
| ENSG00000160179 | 21 | 43609799 | 43727354 | 482 | 89 | 48614 | -0.42418 | 0.66428 | *ABCG1* |
| ENSG00000134765 | 18 | 28699199 | 28752819 | 237 | 21 | 42284 | -0.42442 | 0.66437 | *DSC1* |
| ENSG00000198133 | 14 | 67903801 | 68010456 | 420 | 73 | 44691 | -0.42462 | 0.66444 | *TMEM229B* |

| | | | | | | | | | |
|---|---|---|---|---|---|---|---|---|---|
| ENSG00000213886 | 6 | 29513292 | 29537702 | 85 | 19 | 37388 | -0.42463 | 0.66445 | *UBD* |
| ENSG00000188107 | 6 | 64419876 | 66427118 | 7023 | 254 | 48708 | -0.4251 | 0.66462 | *EYS* |
| ENSG00000105185 | 19 | 33061974 | 33088358 | 79 | 12 | 46268 | -0.4251 | 0.66462 | *PDCD5* |
| ENSG00000205236 | 7 | 102212970 | 102293316 | 4 | 2 | 43033 | -0.42513 | 0.66463 | *RP11-514P8.6* |
| ENSG00000113068 | 5 | 139614624 | 139692706 | 153 | 14 | 50348 | -0.42554 | 0.66478 | *PFDN1* |
| ENSG00000185482 | 12 | 57627236 | 57654976 | 25 | 9 | 45963 | -0.42553 | 0.66478 | *STAC3* |
| ENSG00000121871 | 3 | 164894508 | 164924897 | 74 | 13 | 47910 | -0.42566 | 0.66482 | *SLITRK3* |
| ENSG00000135932 | 2 | 231567560 | 231695790 | 270 | 28 | 40008 | -0.42618 | 0.66501 | *CAB39* |
| ENSG00000164488 | 6 | 168683510 | 168730434 | 216 | 41 | 40159 | -0.42628 | 0.66505 | *DACT2* |
| ENSG00000187792 | 22 | 24073769 | 24103279 | 90 | 19 | 44892 | -0.42659 | 0.66516 | *ZNF70* |
| ENSG00000268421 | 10 | 131298884 | 131319525 | 155 | 12 | 32726 | -0.42681 | 0.66524 | *AL355531.2* |
| ENSG00000122694 | 9 | 36126732 | 36173910 | 194 | 35 | 50215 | -0.42719 | 0.66538 | *GLIPR2* |
| ENSG00000147613 | 8 | 87050602 | 87110850 | 169 | 21 | 41258 | -0.42785 | 0.66562 | *PSKH2* |
| ENSG00000132141 | 17 | 33244878 | 33318097 | 101 | 22 | 49658 | -0.428 | 0.66567 | *CCT6B* |
| ENSG00000141295 | 17 | 45905058 | 45928699 | 74 | 15 | 33402 | -0.42818 | 0.66574 | *SCRN2* |
| ENSG00000184500 | 3 | 93581881 | 93702910 | 119 | 25 | 42555 | -0.42829 | 0.66578 | *PROS1* |
| ENSG00000188725 | 5 | 60443536 | 60468301 | 58 | 11 | 53253 | -0.42867 | 0.66592 | *SMIM15* |
| ENSG00000172954 | 2 | 30660092 | 30877091 | 881 | 48 | 46134 | -0.42877 | 0.66596 | *LCLAT1* |
| ENSG00000111816 | 6 | 116242312 | 116391921 | 373 | 38 | 40924 | -0.42909 | 0.66607 | *FRK* |
| ENSG00000159082 | 21 | 33991069 | 34110359 | 223 | 30 | 52487 | -0.42943 | 0.66619 | *SYNJ1* |
| ENSG00000175984 | 1 | 115115469 | 115223043 | 279 | 33 | 51843 | -0.4296 | 0.66626 | *DENND2C* |
| ENSG00000118271 | 18 | 29161689 | 29188974 | 54 | 11 | 46398 | -0.42963 | 0.66627 | *TTR* |
| ENSG00000159086 | 21 | 34096210 | 34154169 | 114 | 23 | 55362 | -0.42972 | 0.6663 | *PAXBP1* |
| ENSG00000083223 | 9 | 88892648 | 88979369 | 177 | 16 | 38938 | -0.42974 | 0.66631 | *ZCCHC6* |
| ENSG00000128310 | 22 | 38209389 | 38231502 | 43 | 10 | 44244 | -0.42992 | 0.66637 | *GALR3* |
| ENSG00000167945 | 16 | 845443 | 873861 | 137 | 43 | 34659 | -0.42995 | 0.66638 | *PRR25* |
| ENSG00000160072 | 1 | 1397143 | 1443228 | 80 | 23 | 40730 | -0.43011 | 0.66644 | *ATAD3B* |
| ENSG00000270136 | 1 | 19913473 | 19994549 | 259 | 43 | 47474 | -0.43027 | 0.6665 | *MINOS1-NBL1* |
| ENSG00000164692 | 7 | 94013873 | 94070544 | 156 | 34 | 51963 | -0.43042 | 0.66655 | *COL1A2* |
| ENSG00000164129 | 4 | 164255091 | 164283086 | 110 | 20 | 43743 | -0.43065 | 0.66664 | *NPY5R* |
| ENSG00000269093 | 19 | 32885915 | 32906466 | 54 | 12 | 37908 | -0.43074 | 0.66667 | *AC007773.3* |
| ENSG00000070495 | 17 | 74698919 | 74732866 | 80 | 12 | 45564 | -0.43076 | 0.66668 | *JMJD6* |
| ENSG00000221864 | 21 | 46076106 | 46096844 | 159 | 12 | 55479 | -0.4313 | 0.66687 | *KRTAP12-2* |
| ENSG00000206579 | 8 | 56004949 | 56464613 | 1402 | 116 | 41471 | -0.43136 | 0.6669 | *XKR4* |
| ENSG00000125508 | 20 | 62162163 | 62188857 | 72 | 22 | 38677 | -0.43145 | 0.66693 | *SRMS* |
| ENSG00000136874 | 9 | 102658915 | 102742618 | 170 | 16 | 40759 | -0.43151 | 0.66695 | *STX17* |

| | | | | | | | | | |
|---|---|---|---|---|---|---|---|---|---|
| ENSG00000164951 | 8 | 94860035 | 94948294 | 255 | 27 | 47969 | -0.43153 | 0.66696 | *PDP1* |
| ENSG00000104894 | 19 | 49828428 | 49856592 | 65 | 12 | 53280 | -0.43155 | 0.66697 | *CD37* |
| ENSG00000152952 | 3 | 145777227 | 145891440 | 390 | 23 | 53918 | -0.43167 | 0.66701 | *PLOD2* |
| ENSG00000123143 | 19 | 14533865 | 14592679 | 192 | 42 | 38400 | -0.43168 | 0.66701 | *PKN1* |
| ENSG00000102738 | 13 | 41293432 | 41355309 | 77 | 12 | 31287 | -0.43184 | 0.66707 | *MRPS31* |
| ENSG00000205929 | 21 | 34152985 | 34196053 | 116 | 11 | 49807 | -0.43247 | 0.6673 | *C21orf62* |
| ENSG00000272268 | 2 | 6101712 | 6124374 | 57 | 11 | 44754 | -0.4325 | 0.66731 | *DKFZP761K2322* |
| ENSG00000159166 | 1 | 201332372 | 201378736 | 173 | 33 | 41245 | -0.43254 | 0.66733 | *LAD1* |
| ENSG00000271793 | 6 | 86257696 | 86334952 | 135 | 24 | 38006 | -0.43257 | 0.66734 | *RP11-321N4.5* |
| ENSG00000163263 | 1 | 154161848 | 154188809 | 31 | 12 | 42349 | -0.43266 | 0.66737 | *C1orf189* |
| ENSG00000150337 | 1 | 149744227 | 149774074 | 19 | 5 | 30340 | -0.43281 | 0.66742 | *FCGR1A* |
| ENSG00000203326 | 19 | 53858946 | 53905470 | 215 | 40 | 41232 | -0.43291 | 0.66746 | *ZNF525* |
| ENSG00000100221 | 22 | 39071548 | 39107561 | 105 | 12 | 46540 | -0.43293 | 0.66747 | *JOSD1* |
| ENSG00000143106 | 1 | 109931653 | 109979062 | 80 | 11 | 45621 | -0.43308 | 0.66752 | *PSMA5* |
| ENSG00000126602 | 16 | 3691640 | 3777598 | 254 | 45 | 45917 | -0.43306 | 0.66752 | *TRAP1* |
| ENSG00000148737 | 10 | 114700009 | 114937437 | 542 | 99 | 42137 | -0.43309 | 0.66753 | *TCF7L2* |
| ENSG00000124743 | 6 | 53502699 | 53540506 | 220 | 19 | 53523 | -0.43366 | 0.66773 | *KLHL31* |
| ENSG00000171720 | 5 | 140990443 | 141026437 | 92 | 22 | 54231 | -0.43368 | 0.66774 | *HDAC3* |
| ENSG00000198597 | 19 | 30709197 | 31214445 | 1275 | 192 | 42314 | -0.43395 | 0.66784 | *ZNF536* |
| ENSG00000153495 | 13 | 111958531 | 112006596 | 245 | 43 | 47238 | -0.4343 | 0.66796 | *TEX29* |
| ENSG00000104884 | 19 | 45843095 | 45884176 | 131 | 19 | 36872 | -0.43462 | 0.66808 | *ERCC2* |
| ENSG00000120440 | 6 | 167728574 | 167782991 | 233 | 35 | 49583 | -0.43463 | 0.66809 | *TTLL2* |
| ENSG00000113648 | 5 | 134659590 | 134745604 | 148 | 24 | 39019 | -0.43466 | 0.6681 | *H2AFY* |
| ENSG00000163714 | 3 | 142673339 | 142789567 | 287 | 29 | 46363 | -0.43477 | 0.66814 | *U2SURP* |
| ENSG00000138080 | 2 | 27291435 | 27319271 | 33 | 6 | 43510 | -0.43481 | 0.66815 | *EMILIN1* |
| ENSG00000181609 | 11 | 5499915 | 5520979 | 100 | 16 | 48883 | -0.43482 | 0.66815 | *OR52D1* |
| ENSG00000099821 | 19 | 607223 | 643597 | 191 | 42 | 40947 | -0.4349 | 0.66818 | *POLRMT* |
| ENSG00000068976 | 11 | 64503861 | 64537769 | 47 | 11 | 36245 | -0.43492 | 0.66819 | *PYGM* |
| ENSG00000255330 | 6 | 127749551 | 127850146 | 243 | 34 | 37715 | -0.43498 | 0.66821 | *SOGA3* |
| ENSG00000165959 | 14 | 95638277 | 95796243 | 549 | 66 | 41307 | -0.43526 | 0.66831 | *CLMN* |
| ENSG00000111962 | 6 | 149058464 | 149408126 | 1122 | 158 | 48320 | -0.43547 | 0.66839 | *UST* |
| ENSG00000163219 | 2 | 68896733 | 69063965 | 760 | 90 | 45695 | -0.4355 | 0.6684 | *ARHGAP25* |
| ENSG00000144711 | 3 | 12928719 | 13124617 | 638 | 111 | 46709 | -0.43561 | 0.66844 | *IQSEC1* |
| ENSG00000148215 | 9 | 125541150 | 125562237 | 59 | 8 | 29377 | -0.43569 | 0.66847 | *OR5C1* |
| ENSG00000152382 | 1 | 166815747 | 166855564 | 113 | 13 | 54588 | -0.43571 | 0.66848 | *TADA1* |
| ENSG00000112378 | 6 | 138399642 | 138438648 | 146 | 30 | 48643 | -0.43577 | 0.6685 | *PERP* |

| | | | | | | | | | |
|---|---|---|---|---|---|---|---|---|---|
| ENSG00000073910 | 13 | 32595437 | 32880794 | 908 | 135 | 46926 | -0.43602 | 0.66859 | *FRY* |
| ENSG00000023516 | 13 | 42836289 | 42907396 | 198 | 22 | 47515 | -0.43603 | 0.66859 | *AKAP11* |
| ENSG00000183785 | 22 | 18583097 | 18639321 | 208 | 47 | 44583 | -0.43619 | 0.66865 | *TUBA8* |
| ENSG00000144645 | 3 | 31689382 | 32129072 | 1423 | 161 | 39404 | -0.43665 | 0.66882 | *OSBPL10* |
| ENSG00000185220 | 1 | 249190395 | 249224145 | 49 | 21 | 41969 | -0.43673 | 0.66885 | *PGBD2* |
| ENSG00000165688 | 9 | 139295110 | 139328213 | 195 | 25 | 48501 | -0.4371 | 0.66898 | *PMPCA* |
| ENSG00000074219 | 19 | 49833852 | 49875714 | 112 | 12 | 50223 | -0.43748 | 0.66912 | *TEAD2* |
| ENSG00000172878 | 2 | 172854490 | 172957158 | 268 | 41 | 42862 | -0.43782 | 0.66924 | *METAP1D* |
| ENSG00000092607 | 1 | 119415669 | 119542179 | 368 | 43 | 50427 | -0.43829 | 0.66941 | *TBX15* |
| ENSG00000165487 | 13 | 22056836 | 22188353 | 404 | 34 | 37180 | -0.43839 | 0.66945 | *MICU2* |
| ENSG00000139613 | 12 | 56546767 | 56593351 | 42 | 14 | 44525 | -0.43858 | 0.66952 | *SMARCC2* |
| ENSG00000174600 | 12 | 108671821 | 108743118 | 235 | 51 | 49456 | -0.43902 | 0.66968 | *CMKLR1* |
| ENSG00000135821 | 1 | 182340839 | 182371341 | 116 | 13 | 42347 | -0.43906 | 0.66969 | *GLUL* |
| ENSG00000123416 | 12 | 49511565 | 49535180 | 42 | 4 | 28194 | -0.43907 | 0.66969 | *TUBA1B* |
| ENSG00000108264 | 17 | 35756965 | 35849835 | 273 | 25 | 49278 | -0.43911 | 0.66971 | *TADA2A* |
| ENSG00000175595 | 16 | 14004014 | 14056202 | 148 | 17 | 50559 | -0.43923 | 0.66975 | *ERCC4* |
| ENSG00000101577 | 18 | 2906992 | 3023313 | 392 | 51 | 36898 | -0.43926 | 0.66976 | *LPIN2* |
| ENSG00000176533 | 19 | 2501217 | 2712707 | 939 | 153 | 43065 | -0.43937 | 0.6698 | *GNG7* |
| ENSG00000153498 | 13 | 113020633 | 113099003 | 354 | 40 | 38452 | -0.43983 | 0.66997 | *SPACA7* |
| ENSG00000132881 | 1 | 16548195 | 16573657 | 58 | 10 | 45243 | -0.43998 | 0.67002 | *RSG1* |
| ENSG00000064989 | 2 | 188197856 | 188323187 | 328 | 31 | 53329 | -0.44065 | 0.67027 | *CALCRL* |
| ENSG00000111530 | 12 | 67653061 | 67723731 | 253 | 22 | 42117 | -0.44086 | 0.67034 | *CAND1* |
| ENSG00000168792 | 17 | 27877565 | 27904155 | 42 | 18 | 48878 | -0.44099 | 0.67039 | *ABHD15* |
| ENSG00000186166 | 11 | 118858852 | 118896501 | 147 | 15 | 54893 | -0.44126 | 0.67049 | *CCDC84* |
| ENSG00000141959 | 21 | 45709934 | 45757259 | 177 | 25 | 46835 | -0.44129 | 0.6705 | *PFKL* |
| ENSG00000258945 | 14 | 85984943 | 86005730 | 66 | 14 | 45974 | -0.44133 | 0.67051 | *RP11-497E19.2* |
| ENSG00000122435 | 1 | 100588706 | 100626053 | 96 | 21 | 34969 | -0.44177 | 0.67067 | *TRMT13* |
| ENSG00000165471 | 10 | 54515140 | 54541460 | 134 | 20 | 41993 | -0.44202 | 0.67076 | *MBL2* |
| ENSG00000229676 | 19 | 22807126 | 22860472 | 361 | 12 | 32073 | -0.44214 | 0.6708 | *ZNF492* |
| ENSG00000186329 | 3 | 171551139 | 171666505 | 364 | 48 | 50930 | -0.44228 | 0.67086 | *TMEM212* |
| ENSG00000081237 | 1 | 198597801 | 198736545 | 444 | 49 | 47176 | -0.44233 | 0.67087 | *PTPRC* |
| ENSG00000170035 | 2 | 181821975 | 181951312 | 500 | 22 | 32936 | -0.44258 | 0.67097 | *UBE2E3* |
| ENSG00000128656 | 2 | 175654091 | 175880097 | 366 | 32 | 38877 | -0.44274 | 0.67102 | *CHN1* |
| ENSG00000113763 | 5 | 176227478 | 176317897 | 180 | 53 | 39804 | -0.44289 | 0.67108 | *UNC5A* |
| ENSG00000196214 | 19 | 52762824 | 52805977 | 237 | 28 | 43196 | -0.44333 | 0.67124 | *ZNF766* |
| ENSG00000075975 | 3 | 12588513 | 12635212 | 132 | 17 | 32241 | -0.44385 | 0.67142 | *MKRN2* |

| | | | | | | | | | |
|---|---|---|---|---|---|---|---|---|---|
| ENSG00000090061 | 14 | 99937506 | 100011381 | 130 | 25 | 44688 | -0.44397 | 0.67147 | *CCNK* |
| ENSG00000240764 | 5 | 140858808 | 140902546 | 119 | 25 | 46537 | -0.44407 | 0.6715 | *PCDHGC5* |
| ENSG00000102967 | 16 | 72032487 | 72068954 | 120 | 17 | 50304 | -0.44434 | 0.6716 | *DHODH* |
| ENSG00000156103 | 8 | 89034237 | 89350254 | 814 | 107 | 46233 | -0.44441 | 0.67163 | *MMP16* |
| ENSG00000166348 | 10 | 75247296 | 75395711 | 276 | 16 | 42929 | -0.44464 | 0.67171 | *USP54* |
| ENSG00000249471 | 19 | 58952971 | 58979200 | 57 | 12 | 44180 | -0.44463 | 0.67171 | *ZNF324B* |
| ENSG00000169918 | 15 | 31765329 | 32172992 | 1218 | 57 | 39867 | -0.44475 | 0.67175 | *OTUD7A* |
| ENSG00000090581 | 16 | 1391924 | 1423352 | 99 | 24 | 46201 | -0.44476 | 0.67175 | *GNPTG* |
| ENSG00000133454 | 22 | 26128111 | 26437007 | 1185 | 153 | 41956 | -0.44476 | 0.67175 | *MYO18B* |
| ENSG00000012963 | 14 | 93663401 | 93705561 | 138 | 24 | 49246 | -0.44484 | 0.67178 | *UBR7* |
| ENSG00000073598 | 17 | 33438598 | 33467751 | 36 | 15 | 42089 | -0.44509 | 0.67187 | *FNDC8* |
| ENSG00000180871 | 2 | 218980012 | 219011976 | 68 | 8 | 39649 | -0.44522 | 0.67192 | *CXCR2* |
| ENSG00000140992 | 16 | 2577965 | 2663189 | 47 | 28 | 46497 | -0.44525 | 0.67193 | *PDPK1* |
| ENSG00000183665 | 8 | 125453048 | 125484391 | 92 | 15 | 48160 | -0.44534 | 0.67196 | *TRMT12* |
| ENSG00000157423 | 16 | 70831281 | 71274625 | 526 | 66 | 42815 | -0.44539 | 0.67198 | *HYDIN* |
| ENSG00000115415 | 2 | 191819084 | 191895686 | 195 | 40 | 44546 | -0.44543 | 0.672 | *STAT1* |
| ENSG00000184650 | 17 | 8233182 | 8259364 | 149 | 11 | 40213 | -0.44544 | 0.672 | *ODF4* |
| ENSG00000119929 | 10 | 101452315 | 101525891 | 135 | 20 | 54572 | -0.44552 | 0.67203 | *CUTC* |
| ENSG00000157869 | 4 | 13352978 | 13495989 | 527 | 31 | 36559 | -0.44555 | 0.67204 | *RAB28* |
| ENSG00000171199 | 4 | 71253599 | 71285911 | 155 | 24 | 47418 | -0.4456 | 0.67206 | *PROL1* |
| ENSG00000165689 | 9 | 139286377 | 139315061 | 136 | 22 | 52954 | -0.44566 | 0.67208 | *SDCCAG3* |
| ENSG00000120685 | 13 | 39574003 | 39622252 | 117 | 14 | 40032 | -0.4457 | 0.67209 | *PROSER1* |
| ENSG00000055483 | 17 | 76773463 | 76847523 | 165 | 26 | 41478 | -0.44586 | 0.67215 | *USP36* |
| ENSG00000166881 | 12 | 57439426 | 57491846 | 116 | 17 | 48581 | -0.44609 | 0.67223 | *TMEM194A* |
| ENSG00000008853 | 8 | 22834930 | 22887712 | 150 | 33 | 46146 | -0.44628 | 0.6723 | *RHOBTB2* |
| ENSG00000204386 | 6 | 31815436 | 31840683 | 103 | 24 | 35017 | -0.44629 | 0.67231 | *NEU1* |
| ENSG00000159199 | 17 | 46960127 | 46983233 | 71 | 9 | 32238 | -0.44665 | 0.67244 | *ATP5G1* |
| ENSG00000137575 | 8 | 59455483 | 59505419 | 143 | 12 | 51254 | -0.44674 | 0.67247 | *SDCBP* |
| ENSG00000127412 | 7 | 142595267 | 142640905 | 120 | 18 | 28179 | -0.44681 | 0.67249 | *TRPV5* |
| ENSG00000100422 | 22 | 47070308 | 47144158 | 354 | 54 | 43279 | -0.44708 | 0.67259 | *CERK* |
| ENSG00000189144 | 19 | 38216734 | 38317940 | 229 | 32 | 40222 | -0.44714 | 0.67261 | *ZNF573* |
| ENSG00000269533 | 19 | 57891252 | 57941362 | 115 | 18 | 47029 | -0.44723 | 0.67265 | *AC003002.6* |
| ENSG00000156467 | 8 | 97228148 | 97257862 | 70 | 14 | 51284 | -0.44737 | 0.6727 | *UQCRB* |
| ENSG00000234769 | 16 | 54043 | 79452 | 4 | 2 | 42654 | -0.44748 | 0.67273 | *WASH4P* |
| ENSG00000204449 | 11 | 89754274 | 89816558 | 14 | 8 | 43496 | -0.44754 | 0.67276 | *TRIM49C* |
| ENSG00000116809 | 1 | 16258364 | 16312627 | 108 | 19 | 52145 | -0.44759 | 0.67278 | *ZBTB17* |

| | | | | | | | | | |
|---|---|---|---|---|---|---|---|---|---|
| ENSG00000175697 | 3 | 119873492 | 120013941 | 434 | 42 | 45934 | -0.44761 | 0.67278 | *GPR156* |
| ENSG00000253548 | 3 | 191168952 | 191189245 | 143 | 9 | 48612 | -0.44802 | 0.67293 | *PYDC2* |
| ENSG00000151490 | 12 | 15465331 | 15760333 | 723 | 90 | 45877 | -0.44821 | 0.673 | *PTPRO* |
| ENSG00000118729 | 1 | 116232628 | 116321402 | 427 | 34 | 44817 | -0.44824 | 0.67301 | *CASQ2* |
| ENSG00000125820 | 20 | 21481648 | 21504664 | 39 | 13 | 52915 | -0.44838 | 0.67306 | *NKX2-2* |
| ENSG00000102974 | 16 | 67586310 | 67683086 | 153 | 29 | 47006 | -0.4484 | 0.67307 | *CTCF* |
| ENSG00000129993 | 16 | 88931266 | 89053612 | 580 | 79 | 42044 | -0.44847 | 0.67309 | *CBFA2T3* |
| ENSG00000170920 | 19 | 9226688 | 9247626 | 91 | 6 | 26678 | -0.44861 | 0.67314 | *OR7G3* |
| ENSG00000114529 | 3 | 111795182 | 111859851 | 229 | 28 | 40583 | -0.44871 | 0.67318 | *C3orf52* |
| ENSG00000074276 | 5 | 175959512 | 176032975 | 250 | 31 | 40644 | -0.44874 | 0.67319 | *CDHR2* |
| ENSG00000186907 | 11 | 57218022 | 57255007 | 42 | 14 | 53389 | -0.44875 | 0.67319 | *RTN4RL2* |
| ENSG00000135597 | 6 | 139214630 | 139319398 | 396 | 26 | 31073 | -0.44878 | 0.6732 | *REPS1* |
| ENSG00000112294 | 6 | 24485080 | 24547435 | 332 | 42 | 43027 | -0.44889 | 0.67325 | *ALDH5A1* |
| ENSG00000234444 | 7 | 63757837 | 63825238 | 352 | 34 | 46593 | -0.44907 | 0.67331 | *ZNF736* |
| ENSG00000156587 | 11 | 57309129 | 57345757 | 99 | 13 | 53631 | -0.44924 | 0.67337 | *UBE2L6* |
| ENSG00000261247 | 15 | 30417352 | 30447770 | 5 | 3 | 33386 | -0.44935 | 0.67341 | *GOLGA8T* |
| ENSG00000169402 | 7 | 6783740 | 6848996 | 24 | 8 | 43059 | -0.44957 | 0.67349 | *RSPH10B2* |
| ENSG00000125815 | 20 | 23461738 | 23486655 | 79 | 12 | 40088 | -0.44956 | 0.67349 | *CST8* |
| ENSG00000105401 | 19 | 10491810 | 10540797 | 118 | 18 | 52084 | -0.44961 | 0.6735 | *CDC37* |
| ENSG00000112245 | 6 | 64221666 | 64303492 | 165 | 22 | 47967 | -0.44965 | 0.67352 | *PTP4A1* |
| ENSG00000103187 | 16 | 84589200 | 84661683 | 350 | 68 | 45025 | -0.44981 | 0.67358 | *COTL1* |
| ENSG00000198276 | 20 | 62561186 | 62597769 | 93 | 26 | 45580 | -0.4503 | 0.67375 | *UCKL1* |
| ENSG00000165702 | 9 | 135810932 | 135877083 | 265 | 61 | 46791 | -0.45052 | 0.67383 | *GFI1B* |
| ENSG00000172602 | 12 | 49240928 | 49269681 | 60 | 19 | 47253 | -0.45062 | 0.67387 | *RND1* |
| ENSG00000169379 | 3 | 93688983 | 93784512 | 89 | 20 | 44479 | -0.4508 | 0.67393 | *ARL13B* |
| ENSG00000182083 | 2 | 240958841 | 240979906 | 74 | 24 | 47970 | -0.45119 | 0.67407 | *OR6B2* |
| ENSG00000072274 | 3 | 195744054 | 195819060 | 331 | 45 | 49099 | -0.45118 | 0.67407 | *TFRC* |
| ENSG00000133980 | 14 | 74759772 | 74836711 | 191 | 39 | 40103 | -0.45122 | 0.67409 | *VRTN* |
| ENSG00000128463 | 15 | 34507200 | 34532357 | 108 | 16 | 50074 | -0.45136 | 0.67413 | *EMC4* |
| ENSG00000117594 | 1 | 209849510 | 209918295 | 129 | 17 | 41250 | -0.45156 | 0.67421 | *HSD11B1* |
| ENSG00000145365 | 4 | 113185698 | 113217059 | 65 | 7 | 29712 | -0.45167 | 0.67425 | *TIFA* |
| ENSG00000184260 | 1 | 149848525 | 149868961 | 38 | 6 | 53972 | -0.45184 | 0.67431 | *HIST2H2AC* |
| ENSG00000260362 | 16 | 9750742 | 9780700 | 188 | 19 | 45927 | -0.45196 | 0.67435 | *RP11-297M9.1* |
| ENSG00000204882 | 8 | 142356600 | 142387367 | 94 | 19 | 32355 | -0.452 | 0.67437 | *GPR20* |
| ENSG00000107937 | 10 | 1024338 | 1075876 | 181 | 24 | 49138 | -0.45201 | 0.67437 | *GTPBP4* |
| ENSG00000168906 | 2 | 85756288 | 85782403 | 90 | 12 | 48947 | -0.45235 | 0.67449 | *MAT2A* |

| | | | | | | | | | |
|---|---|---|---|---|---|---|---|---|---|
| ENSG00000165899 | 12 | 80593233 | 80782870 | 475 | 53 | 42526 | -0.45258 | 0.67457 | *OTOGL* |
| ENSG00000165434 | 11 | 74031363 | 74119518 | 291 | 28 | 43518 | -0.45265 | 0.6746 | *PGM2L1* |
| ENSG00000168528 | 1 | 31872412 | 31917525 | 172 | 15 | 29524 | -0.45268 | 0.67461 | *SERINC2* |
| ENSG00000141506 | 17 | 8772233 | 8879029 | 297 | 53 | 44666 | -0.45274 | 0.67463 | *PIK3R5* |
| ENSG00000196367 | 7 | 98465556 | 98620866 | 294 | 30 | 48371 | -0.45295 | 0.67471 | *TRRAP* |
| ENSG00000240344 | 2 | 201725630 | 201764026 | 70 | 16 | 48020 | -0.45307 | 0.67475 | *PPIL3* |
| ENSG00000177414 | 1 | 64659310 | 64743051 | 255 | 29 | 49370 | -0.45316 | 0.67478 | *UBE2U* |
| ENSG00000130159 | 19 | 11606731 | 11649989 | 109 | 17 | 38442 | -0.45315 | 0.67478 | *ECSIT* |
| ENSG00000181826 | 4 | 37582422 | 37697998 | 452 | 81 | 40139 | -0.45337 | 0.67486 | *RELL1* |
| ENSG00000140009 | 14 | 64540950 | 64814830 | 773 | 92 | 40559 | -0.45383 | 0.67502 | *ESR2* |
| ENSG00000215788 | 1 | 6511211 | 6536255 | 56 | 13 | 34845 | -0.45393 | 0.67506 | *TNFRSF25* |
| ENSG00000258869 | 14 | 98205601 | 98240326 | 115 | 18 | 47904 | -0.45425 | 0.67518 | *RP11-204N11.1* |
| ENSG00000257591 | 19 | 12241032 | 12277546 | 76 | 12 | 44402 | -0.45426 | 0.67518 | *ZNF625* |
| ENSG00000175768 | 9 | 37572643 | 37602639 | 77 | 12 | 49245 | -0.45496 | 0.67543 | *TOMM5* |
| ENSG00000269468 | 1 | 17645507 | 17686070 | 278 | 25 | 54139 | -0.45502 | 0.67545 | *AC004824.2* |
| ENSG00000090273 | 1 | 27216729 | 27283353 | 114 | 19 | 53386 | -0.45523 | 0.67553 | *NUDC* |
| ENSG00000163673 | 3 | 36743913 | 36791352 | 112 | 27 | 39312 | -0.45549 | 0.67562 | *DCLK3* |
| ENSG00000150455 | 11 | 126142960 | 126178740 | 114 | 21 | 42421 | -0.45589 | 0.67576 | *TIRAP* |
| ENSG00000184009 | 17 | 79466997 | 79500873 | 187 | 25 | 32481 | -0.45602 | 0.67581 | *ACTG1* |
| ENSG00000167642 | 19 | 38724675 | 38793254 | 231 | 32 | 52069 | -0.45601 | 0.67581 | *SPINT2* |
| ENSG00000162613 | 1 | 78399740 | 78454794 | 149 | 23 | 42878 | -0.45605 | 0.67582 | *FUBP1* |
| ENSG00000152213 | 13 | 50192435 | 50218008 | 96 | 22 | 31753 | -0.45632 | 0.67592 | *ARL11* |
| ENSG00000173366 | 3 | 52245097 | 52275206 | 44 | 13 | 54061 | -0.45637 | 0.67594 | *TLR9* |
| ENSG00000154305 | 1 | 222781428 | 222851354 | 182 | 21 | 40922 | -0.45655 | 0.676 | *MIA3* |
| ENSG00000198963 | 9 | 77102281 | 77318093 | 553 | 66 | 48478 | -0.4568 | 0.67609 | *RORB* |
| ENSG00000124490 | 6 | 49650073 | 49691274 | 140 | 20 | 44368 | -0.45683 | 0.6761 | *CRISP2* |
| ENSG00000198829 | 3 | 151581431 | 151612405 | 105 | 23 | 47959 | -0.45723 | 0.67625 | *SUCNR1* |
| ENSG00000198417 | 16 | 56681606 | 56704610 | 151 | 26 | 46958 | -0.4575 | 0.67634 | *MT1F* |
| ENSG00000186973 | 1 | 43600824 | 43632067 | 148 | 22 | 30379 | -0.45804 | 0.67654 | *FAM183A* |
| ENSG00000177692 | 21 | 34850497 | 34874027 | 69 | 13 | 52422 | -0.45823 | 0.67661 | *DNAJC28* |
| ENSG00000205930 | 21 | 34134411 | 34276043 | 368 | 36 | 52870 | -0.45844 | 0.67668 | *C21orf49* |
| ENSG00000214338 | 6 | 127749551 | 127850500 | 245 | 34 | 37821 | -0.45889 | 0.67684 | *SOGA3* |
| ENSG00000163558 | 3 | 169930153 | 170033769 | 392 | 28 | 45238 | -0.45892 | 0.67685 | *PRKCI* |
| ENSG00000159753 | 16 | 67668822 | 67701472 | 48 | 16 | 44562 | -0.4594 | 0.67703 | *RLTPR* |
| ENSG00000140961 | 16 | 83971887 | 84009937 | 236 | 38 | 40836 | -0.45995 | 0.67722 | *OSGIN1* |
| ENSG00000169213 | 1 | 52363628 | 52466436 | 256 | 32 | 38932 | -0.46018 | 0.6773 | *RAB3B* |

| | | | | | | | | | |
|---|---|---|---|---|---|---|---|---|---|
| ENSG00000120784 | 19 | 38094650 | 38193238 | 224 | 29 | 38313 | -0.46018 | 0.67731 | *ZFP30* |
| ENSG00000138744 | 4 | 76821809 | 76872204 | 212 | 24 | 36705 | -0.4604 | 0.67739 | *NAAA* |
| ENSG00000072682 | 5 | 131517531 | 131641008 | 326 | 40 | 32380 | -0.46083 | 0.67754 | *P4HA2* |
| ENSG00000138722 | 4 | 90790683 | 90885780 | 269 | 33 | 39671 | -0.46085 | 0.67755 | *MMRN1* |
| ENSG00000164920 | 8 | 99946631 | 99974332 | 34 | 14 | 51916 | -0.46093 | 0.67757 | *OSR2* |
| ENSG00000109519 | 4 | 7050633 | 7079924 | 137 | 19 | 43871 | -0.46101 | 0.67761 | *GRPEL1* |
| ENSG00000164070 | 4 | 128692976 | 128771888 | 70 | 19 | 47233 | -0.46127 | 0.6777 | *HSPA4L* |
| ENSG00000189195 | 1 | 92535862 | 92623393 | 182 | 18 | 46457 | -0.4617 | 0.67785 | *BTBD8* |
| ENSG00000129437 | 19 | 51570752 | 51597502 | 90 | 27 | 46438 | -0.46198 | 0.67795 | *KLK14* |
| ENSG00000140297 | 15 | 59877074 | 59942438 | 186 | 37 | 41132 | -0.46234 | 0.67808 | *GCNT3* |
| ENSG00000172379 | 15 | 80686692 | 80900278 | 602 | 110 | 46061 | -0.46245 | 0.67812 | *ARNT2* |
| ENSG00000154803 | 17 | 17105526 | 17150502 | 156 | 22 | 49297 | -0.4626 | 0.67817 | *FLCN* |
| ENSG00000130305 | 7 | 72706514 | 72732864 | 32 | 16 | 43088 | -0.46275 | 0.67823 | *NSUN5* |
| ENSG00000180921 | 8 | 144796103 | 144825971 | 97 | 17 | 35715 | -0.46296 | 0.6783 | *FAM83H* |
| ENSG00000173442 | 11 | 65333509 | 65370121 | 82 | 17 | 49450 | -0.46294 | 0.6783 | *EHBP1L1* |
| ENSG00000172519 | 19 | 15894761 | 15915892 | 80 | 13 | 53620 | -0.46305 | 0.67833 | *OR10H5* |
| ENSG00000160994 | 19 | 15111556 | 15144081 | 231 | 37 | 45684 | -0.46306 | 0.67834 | *CCDC105* |
| ENSG00000184032 | 21 | 31997555 | 32017938 | 40 | 8 | 50755 | -0.46333 | 0.67844 | *KRTAP20-2* |
| ENSG00000078114 | 10 | 21058902 | 21473116 | 1093 | 113 | 43259 | -0.4639 | 0.67864 | *NEBL* |
| ENSG00000106263 | 7 | 2383721 | 2430380 | 157 | 20 | 39335 | -0.46398 | 0.67867 | *EIF3B* |
| ENSG00000269027 | 5 | 35041012 | 35061198 | 59 | 16 | 36484 | -0.46451 | 0.67886 | *AC010368.2* |
| ENSG00000130414 | 2 | 240821867 | 240974819 | 740 | 85 | 49942 | -0.46479 | 0.67896 | *NDUFA10* |
| ENSG00000099960 | 22 | 21373007 | 21397129 | 81 | 18 | 45346 | -0.46486 | 0.67898 | *SLC7A4* |
| ENSG00000182013 | 19 | 46959748 | 46984820 | 142 | 20 | 51339 | -0.46493 | 0.67901 | *PNMAL1* |
| ENSG00000137441 | 4 | 15951866 | 15980932 | 108 | 25 | 46787 | -0.46497 | 0.67902 | *FGFBP2* |
| ENSG00000196787 | 6 | 27090832 | 27113070 | 41 | 17 | 45969 | -0.46514 | 0.67909 | *HIST1H2AG* |
| ENSG00000100138 | 22 | 42059934 | 42096508 | 83 | 10 | 40117 | -0.46573 | 0.6793 | *NHP2L1* |
| ENSG00000151164 | 12 | 110929460 | 110979891 | 132 | 16 | 31503 | -0.4663 | 0.6795 | *RAD9B* |
| ENSG00000167487 | 19 | 18737775 | 18791309 | 193 | 31 | 47355 | -0.4663 | 0.6795 | *KLHL26* |
| ENSG00000198408 | 10 | 103534200 | 103588696 | 80 | 16 | 52726 | -0.46632 | 0.67951 | *MGEA5* |
| ENSG00000134917 | 11 | 130264820 | 130308888 | 129 | 34 | 43483 | -0.46644 | 0.67955 | *ADAMTS8* |
| ENSG00000113966 | 3 | 97473365 | 97529953 | 119 | 17 | 41433 | -0.46663 | 0.67962 | *ARL6* |
| ENSG00000066279 | 1 | 197043258 | 197125824 | 160 | 13 | 23421 | -0.46666 | 0.67963 | *ASPM* |
| ENSG00000269383 | 2 | 100976705 | 100997007 | 40 | 7 | 40827 | -0.46698 | 0.67974 | *AC012493.2* |
| ENSG00000165716 | 9 | 139597022 | 139628502 | 125 | 22 | 48824 | -0.46697 | 0.67974 | *FAM69B* |
| ENSG00000138764 | 4 | 78068304 | 78364542 | 1829 | 47 | 47009 | -0.46716 | 0.67981 | *CCNG2* |

| | | | | | | | | | |
|---|---|---|---|---|---|---|---|---|---|
| ENSG00000026103 | 10 | 90740414 | 90785542 | 231 | 23 | 44488 | -0.4672 | 0.67982 | *FAS* |
| ENSG00000187630 | 14 | 24429148 | 24485617 | 213 | 33 | 23809 | -0.46723 | 0.67983 | *DHRS4L2* |
| ENSG00000135297 | 6 | 74161301 | 74228959 | 223 | 27 | 49908 | -0.46724 | 0.67984 | *MTO1* |
| ENSG00000099377 | 16 | 30986519 | 31010473 | 34 | 9 | 31210 | -0.46738 | 0.67989 | *HSD3B7* |
| ENSG00000067248 | 5 | 54542073 | 54613550 | 148 | 17 | 36584 | -0.46751 | 0.67993 | *DHX29* |
| ENSG00000198018 | 10 | 101409263 | 101475997 | 146 | 14 | 55272 | -0.46762 | 0.67997 | *ENTPD7* |
| ENSG00000176406 | 8 | 104502976 | 105278322 | 2825 | 80 | 46032 | -0.46789 | 0.68007 | *RIMS2* |
| ENSG00000268662 | 1 | 186354123 | 186375908 | 51 | 14 | 52630 | -0.46812 | 0.68015 | *AL596220.1* |
| ENSG00000186136 | 12 | 11328599 | 11349543 | 103 | 10 | 33778 | -0.46813 | 0.68015 | *TAS2R42* |
| ENSG00000178202 | 11 | 108332832 | 108379159 | 96 | 20 | 47710 | -0.46854 | 0.6803 | *KDELC2* |
| ENSG00000010278 | 12 | 6298881 | 6357425 | 99 | 23 | 41660 | -0.4687 | 0.68036 | *CD9* |
| ENSG00000108064 | 10 | 60134782 | 60168981 | 111 | 24 | 37097 | -0.46913 | 0.68051 | *TFAM* |
| ENSG00000145715 | 5 | 86553705 | 86697748 | 230 | 26 | 49567 | -0.46935 | 0.68059 | *RASA1* |
| ENSG00000186795 | 10 | 118947000 | 118979810 | 117 | 12 | 49736 | -0.46957 | 0.68067 | *KCNK18* |
| ENSG00000248487 | 3 | 51995442 | 52025212 | 33 | 13 | 34233 | -0.46964 | 0.6807 | *ABHD14A* |
| ENSG00000197006 | 16 | 21598539 | 21678794 | 106 | 28 | 38419 | -0.47009 | 0.68086 | *METTL9* |
| ENSG00000165512 | 10 | 45485923 | 45510774 | 62 | 15 | 45196 | -0.47032 | 0.68094 | *ZNF22* |
| ENSG00000127780 | 17 | 3326164 | 3347135 | 56 | 9 | 30394 | -0.47049 | 0.681 | *OR1E2* |
| ENSG00000169583 | 9 | 139879087 | 139901255 | 62 | 8 | 54242 | -0.47067 | 0.68106 | *CLIC3* |
| ENSG00000099889 | 22 | 19947419 | 20014331 | 285 | 41 | 43418 | -0.47092 | 0.68115 | *ARVCF* |
| ENSG00000147873 | 9 | 21294613 | 21315312 | 60 | 9 | 53930 | -0.47114 | 0.68123 | *IFNA5* |
| ENSG00000065675 | 10 | 6459105 | 6632263 | 557 | 93 | 42600 | -0.47118 | 0.68124 | *PRKCQ* |
| ENSG00000086696 | 16 | 82058609 | 82142139 | 170 | 33 | 36049 | -0.47138 | 0.68132 | *HSD17B2* |
| ENSG00000129195 | 17 | 6337735 | 6364789 | 142 | 29 | 40565 | -0.47166 | 0.68141 | *FAM64A* |
| ENSG00000163002 | 2 | 183972241 | 184036408 | 431 | 35 | 52209 | -0.47225 | 0.68163 | *NUP35* |
| ENSG00000149679 | 20 | 60953688 | 60992341 | 188 | 24 | 45055 | -0.47245 | 0.6817 | *CABLES2* |
| ENSG00000104427 | 8 | 79568282 | 79642000 | 198 | 17 | 52502 | -0.47256 | 0.68174 | *ZC2HC1A* |
| ENSG00000189058 | 3 | 195285573 | 195321076 | 173 | 34 | 43585 | -0.47267 | 0.68178 | *APOD* |
| ENSG00000122390 | 16 | 3483611 | 3546963 | 300 | 34 | 45685 | -0.47281 | 0.68183 | *NAA60* |
| ENSG00000105270 | 19 | 36495562 | 36534245 | 88 | 23 | 45244 | -0.47283 | 0.68183 | *CLIP3* |
| ENSG00000124635 | 6 | 27083676 | 27110541 | 53 | 18 | 46079 | -0.47291 | 0.68186 | *HIST1H2BJ* |
| ENSG00000131475 | 17 | 40915454 | 40941617 | 16 | 11 | 46155 | -0.47291 | 0.68186 | *VPS25* |
| ENSG00000065485 | 3 | 122775909 | 122954074 | 583 | 62 | 48310 | -0.47303 | 0.6819 | *PDIA5* |
| ENSG00000166435 | 11 | 74508784 | 74670245 | 320 | 39 | 44177 | -0.47331 | 0.682 | *XRRA1* |
| ENSG00000122643 | 7 | 33043742 | 33112409 | 281 | 17 | 50103 | -0.47359 | 0.6821 | *NT5C3A* |
| ENSG00000187912 | 19 | 14683896 | 14731969 | 181 | 32 | 48407 | -0.47377 | 0.68217 | *CLEC17A* |

| | | | | | | | | | |
|---|---|---|---|---|---|---|---|---|---|
| ENSG00000171855 | 9 | 21067104 | 21087943 | 74 | 16 | 56850 | -0.47385 | 0.6822 | *IFNB1* |
| ENSG00000104964 | 19 | 3042908 | 3073105 | 81 | 30 | 44895 | -0.47386 | 0.6822 | *AES* |
| ENSG00000182585 | 4 | 75164190 | 75191024 | 38 | 5 | 27087 | -0.47396 | 0.68223 | *EPGN* |
| ENSG00000249437 | 5 | 70254310 | 70330941 | 9 | 5 | 34632 | -0.47482 | 0.68254 | *NAIP* |
| ENSG00000213822 | 19 | 51969838 | 52004206 | 216 | 28 | 55423 | -0.47492 | 0.68258 | *CEACAM18* |
| ENSG00000119509 | 9 | 102851538 | 103073282 | 546 | 27 | 25860 | -0.47496 | 0.68259 | *INVS* |
| ENSG00000141499 | 17 | 7579389 | 7616820 | 100 | 19 | 30010 | -0.47504 | 0.68262 | *WRAP53* |
| ENSG00000268046 | 18 | 8326437 | 8347038 | 104 | 19 | 40970 | -0.47509 | 0.68264 | *AP001094.1* |
| ENSG00000134759 | 18 | 33699407 | 33767909 | 200 | 21 | 51144 | -0.47528 | 0.68271 | *ELP2* |
| ENSG00000105771 | 19 | 44225301 | 44269142 | 101 | 14 | 49263 | -0.47537 | 0.68274 | *SMG9* |
| ENSG00000144481 | 2 | 234816043 | 234938166 | 458 | 79 | 42561 | -0.47544 | 0.68276 | *TRPM8* |
| ENSG00000167261 | 16 | 68011297 | 68044489 | 52 | 13 | 43619 | -0.47549 | 0.68278 | *DPEP2* |
| ENSG00000165995 | 10 | 18419606 | 18840798 | 1844 | 209 | 48702 | -0.47565 | 0.68284 | *CACNB2* |
| ENSG00000267958 | 2 | 74408517 | 74428627 | 45 | 12 | 33287 | -0.47567 | 0.68285 | *CATX-2* |
| ENSG00000156875 | 1 | 100493653 | 100558933 | 217 | 25 | 43792 | -0.47581 | 0.6829 | *HIAT1* |
| ENSG00000259658 | 15 | 102267302 | 102295913 | 60 | 13 | 36144 | -0.47597 | 0.68295 | *RP11-89K11.1* |
| ENSG00000239886 | 17 | 39372900 | 39393904 | 60 | 13 | 50660 | -0.47614 | 0.68301 | *KRTAP9-2* |
| ENSG00000197414 | 15 | 22726246 | 22756002 | 19 | 5 | 44960 | -0.47624 | 0.68305 | *GOLGA6L1* |
| ENSG00000237521 | 19 | 9351606 | 9372820 | 70 | 10 | 41056 | -0.47648 | 0.68313 | *OR7E24* |
| ENSG00000153303 | 6 | 168446425 | 168492237 | 222 | 38 | 48630 | -0.47652 | 0.68315 | *FRMD1* |
| ENSG00000155304 | 21 | 15733436 | 15765805 | 136 | 21 | 49607 | -0.47655 | 0.68316 | *HSPA13* |
| ENSG00000148444 | 10 | 22594903 | 22619235 | 15 | 9 | 42363 | -0.47674 | 0.68323 | *COMMD3* |
| ENSG00000140157 | 15 | 22994684 | 23044427 | 139 | 18 | 45863 | -0.47681 | 0.68325 | *NIPA2* |
| ENSG00000115718 | 2 | 128166003 | 128196822 | 102 | 15 | 52898 | -0.47707 | 0.68335 | *PROC* |
| ENSG00000179914 | 1 | 160836329 | 160864960 | 116 | 7 | 32698 | -0.47724 | 0.6834 | *ITLN1* |
| ENSG00000171097 | 9 | 131585221 | 131654773 | 167 | 18 | 43090 | -0.47726 | 0.68341 | *CCBL1* |
| ENSG00000106052 | 7 | 27768950 | 27894183 | 374 | 45 | 51110 | -0.47728 | 0.68342 | *TAX1BP1* |
| ENSG00000138600 | 15 | 50989506 | 51068005 | 321 | 27 | 45600 | -0.47744 | 0.68348 | *SPPL2A* |
| ENSG00000177842 | 3 | 40537483 | 40570227 | 42 | 9 | 36841 | -0.47767 | 0.68356 | *ZNF620* |
| ENSG00000103034 | 16 | 58486750 | 58557532 | 200 | 51 | 43320 | -0.4777 | 0.68357 | *NDRG4* |
| ENSG00000047457 | 3 | 148870197 | 148949842 | 267 | 50 | 48478 | -0.47792 | 0.68365 | *CP* |
| ENSG00000158042 | 11 | 6692013 | 6714632 | 90 | 13 | 36939 | -0.47842 | 0.68382 | *MRPL17* |
| ENSG00000123329 | 12 | 57856038 | 57892597 | 80 | 16 | 37837 | -0.4784 | 0.68382 | *ARHGAP9* |
| ENSG00000156689 | 11 | 58591542 | 58681688 | 445 | 27 | 27759 | -0.47842 | 0.68383 | *GLYATL2* |
| ENSG00000188677 | 22 | 44385091 | 44578829 | 744 | 161 | 46918 | -0.47852 | 0.68386 | *PARVB* |
| ENSG00000155636 | 2 | 178967151 | 179013738 | 128 | 26 | 48844 | -0.47854 | 0.68387 | *RBM45* |

| | | | | | | | | | |
|---|---|---|---|---|---|---|---|---|---|
| ENSG00000160613 | 11 | 117065053 | 117113241 | 170 | 31 | 38731 | -0.47857 | 0.68388 | *PCSK7* |
| ENSG00000021574 | 2 | 32278680 | 32392706 | 356 | 26 | 49382 | -0.47885 | 0.68398 | *SPAST* |
| ENSG00000164729 | 17 | 33509539 | 33531412 | 72 | 15 | 33124 | -0.47906 | 0.68405 | *SLC35G3* |
| ENSG00000158089 | 2 | 31123333 | 31388068 | 980 | 122 | 46266 | -0.47965 | 0.68426 | *GALNT14* |
| ENSG00000141446 | 18 | 19099242 | 19190845 | 117 | 17 | 36534 | -0.47986 | 0.68434 | *ESCO1* |
| ENSG00000197213 | 19 | 56690939 | 56719289 | 164 | 28 | 43225 | -0.47988 | 0.68434 | *ZSCAN5B* |
| ENSG00000163623 | 4 | 85403140 | 85429603 | 48 | 13 | 42154 | -0.47993 | 0.68436 | *NKX6-1* |
| ENSG00000172172 | 8 | 121383000 | 121467642 | 159 | 17 | 34614 | -0.47992 | 0.68436 | *MRPL13* |
| ENSG00000253148 | 1 | 192276122 | 192346415 | 201 | 21 | 52144 | -0.48002 | 0.68439 | *RGS21* |
| ENSG00000253320 | 8 | 103866528 | 104000104 | 409 | 51 | 47833 | -0.48002 | 0.68439 | *KB-1507C5.2* |
| ENSG00000116874 | 1 | 119563839 | 119693294 | 387 | 35 | 48975 | -0.48016 | 0.68444 | *WARS2* |
| ENSG00000254806 | 20 | 43981840 | 44049250 | 173 | 15 | 40703 | -0.48055 | 0.68458 | *SYS1-DBNDD2* |
| ENSG00000172578 | 3 | 183195319 | 183283477 | 232 | 43 | 41432 | -0.48059 | 0.6846 | *KLHL6* |
| ENSG00000137491 | 11 | 74801608 | 74927594 | 413 | 38 | 37057 | -0.48064 | 0.68461 | *SLCO2B1* |
| ENSG00000237353 | 11 | 125693211 | 125719964 | 104 | 12 | 52817 | -0.48071 | 0.68464 | *PATE4* |
| ENSG00000197157 | 7 | 127282234 | 127742661 | 843 | 72 | 40697 | -0.48094 | 0.68472 | *SND1* |
| ENSG00000084628 | 1 | 31642592 | 31722401 | 301 | 29 | 28593 | -0.48106 | 0.68476 | *NKAIN1* |
| ENSG00000145495 | 5 | 10343815 | 10450500 | 87 | 35 | 42306 | -0.48112 | 0.68478 | *Mar-06* |
| ENSG00000258847 | 14 | 66414015 | 66481241 | 235 | 39 | 39920 | -0.48112 | 0.68479 | *CTD-2014B16.3* |
| ENSG00000152492 | 3 | 191036866 | 191126459 | 399 | 32 | 36608 | -0.48128 | 0.68484 | *CCDC50* |
| ENSG00000263002 | 19 | 44635710 | 44673156 | 61 | 11 | 41741 | -0.48138 | 0.68488 | *ZNF234* |
| ENSG00000163517 | 3 | 13511224 | 13557916 | 130 | 37 | 48516 | -0.48214 | 0.68515 | *HDAC11* |
| ENSG00000205670 | 21 | 35737779 | 35790164 | 149 | 21 | 43218 | -0.48227 | 0.68519 | *SMIM11* |
| ENSG00000183087 | 13 | 114513522 | 114577046 | 290 | 38 | 32968 | -0.48242 | 0.68525 | *GAS6* |
| ENSG00000114786 | 3 | 51999066 | 52033199 | 44 | 19 | 39959 | -0.48251 | 0.68528 | *ABHD14A-ACY1* |
| ENSG00000243989 | 3 | 51999066 | 52033213 | 44 | 19 | 39959 | -0.48251 | 0.68528 | *ACY1* |
| ENSG00000145861 | 5 | 159764758 | 159807648 | 127 | 23 | 48247 | -0.48255 | 0.68529 | *C1QTNF2* |
| ENSG00000173276 | 21 | 43396940 | 43440496 | 167 | 30 | 47616 | -0.48254 | 0.68529 | *ZBTB21* |
| ENSG00000170899 | 6 | 52832751 | 52870176 | 175 | 18 | 46272 | -0.48256 | 0.6853 | *GSTA4* |
| ENSG00000102924 | 16 | 49301828 | 49325742 | 72 | 19 | 45434 | -0.48287 | 0.6854 | *CBLN1* |
| ENSG00000078098 | 2 | 163017194 | 163111661 | 157 | 12 | 27550 | -0.48292 | 0.68542 | *FAP* |
| ENSG00000135390 | 12 | 54016510 | 54081192 | 214 | 18 | 39139 | -0.48297 | 0.68544 | *ATP5G2* |
| ENSG00000095015 | 5 | 56101401 | 56201979 | 294 | 26 | 40833 | -0.48339 | 0.68559 | *MAP3K1* |
| ENSG00000166856 | 12 | 57378230 | 57400468 | 36 | 8 | 51856 | -0.48369 | 0.6857 | *GPR182* |
| ENSG00000053372 | 1 | 19568033 | 19596622 | 154 | 21 | 48825 | -0.48376 | 0.68572 | *MRTO4* |
| ENSG00000122728 | 9 | 32619452 | 32645667 | 32 | 9 | 52918 | -0.48393 | 0.68578 | *TAF1L* |

| | | | | | | | | | |
|---|---|---|---|---|---|---|---|---|---|
| ENSG00000078246 | 12 | 2976389 | 3060306 | 318 | 12 | 45854 | -0.4843 | 0.68591 | *TULP3* |
| ENSG00000268657 | 14 | 92028788 | 92051383 | 128 | 10 | 50927 | -0.48463 | 0.68603 | *AL133373.1* |
| ENSG00000120332 | 1 | 175026994 | 175127202 | 432 | 49 | 47270 | -0.48508 | 0.68619 | *TNN* |
| ENSG00000160808 | 3 | 46889362 | 46933659 | 87 | 21 | 38989 | -0.48538 | 0.6863 | *MYL3* |
| ENSG00000237330 | 1 | 996346 | 1019687 | 59 | 10 | 45425 | -0.4859 | 0.68648 | *RNF223* |
| ENSG00000163214 | 2 | 39014871 | 39113075 | 216 | 20 | 40853 | -0.48608 | 0.68654 | *DHX57* |
| ENSG00000065491 | 6 | 37215548 | 37310746 | 204 | 20 | 52288 | -0.48613 | 0.68656 | *TBC1D22B* |
| ENSG00000255150 | 12 | 104687517 | 104708980 | 64 | 12 | 55277 | -0.48615 | 0.68657 | *EID3* |
| ENSG00000258875 | 14 | 91699103 | 91727426 | 93 | 13 | 41952 | -0.48629 | 0.68662 | *CTD-2547L24.3* |
| ENSG00000118160 | 19 | 47921274 | 47985425 | 146 | 30 | 40714 | -0.48628 | 0.68662 | *SLC8A2* |
| ENSG00000156475 | 5 | 145957936 | 146474347 | 1283 | 120 | 39471 | -0.48647 | 0.68668 | *PPP2R2B* |
| ENSG00000127954 | 7 | 87895744 | 87946206 | 99 | 11 | 48632 | -0.48656 | 0.68671 | *STEAP4* |
| ENSG00000198740 | 17 | 47356568 | 47449835 | 256 | 23 | 54164 | -0.48675 | 0.68678 | *ZNF652* |
| ENSG00000156521 | 10 | 71887737 | 71916432 | 125 | 24 | 43821 | -0.48681 | 0.6868 | *TYSND1* |
| ENSG00000132463 | 4 | 71671499 | 71715662 | 134 | 17 | 32873 | -0.48687 | 0.68682 | *GRSF1* |
| ENSG00000139597 | 13 | 32964861 | 33012315 | 104 | 23 | 37099 | -0.48694 | 0.68685 | *N4BP2L1* |
| ENSG00000197746 | 10 | 73566055 | 73621126 | 207 | 33 | 48818 | -0.48709 | 0.6869 | *PSAP* |
| ENSG00000101391 | 20 | 31936645 | 31999367 | 94 | 13 | 54198 | -0.48708 | 0.6869 | *CDK5RAP1* |
| ENSG00000142082 | 11 | 205458 | 246931 | 229 | 20 | 49365 | -0.4872 | 0.68694 | *SIRT3* |
| ENSG00000242259 | 22 | 19328891 | 19445755 | 244 | 32 | 36528 | -0.48756 | 0.68707 | *C22orf39* |
| ENSG00000160999 | 7 | 101918405 | 101972178 | 149 | 37 | 49993 | -0.48767 | 0.68711 | *SH2B2* |
| ENSG00000008277 | 7 | 87553458 | 87842204 | 577 | 50 | 37527 | -0.48787 | 0.68718 | *ADAM22* |
| ENSG00000248751 | 22 | 30671227 | 30705471 | 83 | 24 | 45401 | -0.48789 | 0.68719 | *RP1-130H16.18* |
| ENSG00000204060 | 1 | 41817594 | 41859262 | 134 | 34 | 48023 | -0.48806 | 0.68725 | *FOXO6* |
| ENSG00000075239 | 11 | 107982243 | 108028503 | 105 | 19 | 44516 | -0.48849 | 0.6874 | *ACAT1* |
| ENSG00000149968 | 11 | 102696532 | 102724534 | 93 | 14 | 50651 | -0.48859 | 0.68744 | *MMP3* |
| ENSG00000115687 | 2 | 242035514 | 242099679 | 249 | 32 | 36048 | -0.48869 | 0.68747 | *PASK* |
| ENSG00000269920 | 10 | 97723786 | 97802432 | 230 | 40 | 49209 | -0.48876 | 0.68749 | *RP11-690P14.4* |
| ENSG00000188649 | 10 | 97723786 | 97802441 | 230 | 40 | 49209 | -0.48876 | 0.68749 | *CC2D2B* |
| ENSG00000166415 | 15 | 53795938 | 54065075 | 1212 | 85 | 42448 | -0.48893 | 0.68755 | *WDR72* |
| ENSG00000079156 | 2 | 179049208 | 179274160 | 680 | 84 | 43707 | -0.48902 | 0.68759 | *OSBPL6* |
| ENSG00000099326 | 19 | 59063298 | 59094942 | 90 | 19 | 37402 | -0.48915 | 0.68763 | *MZF1* |
| ENSG00000136942 | 9 | 127610159 | 127634260 | 52 | 10 | 51020 | -0.48925 | 0.68767 | *RPL35* |
| ENSG00000188152 | 9 | 99680592 | 99712535 | 31 | 13 | 40653 | -0.48938 | 0.68771 | *NUTM2G* |
| ENSG00000198339 | 6 | 27097076 | 27118418 | 37 | 14 | 43577 | -0.48939 | 0.68772 | *HIST1H4I* |
| ENSG00000127241 | 3 | 186925942 | 187019810 | 328 | 58 | 44939 | -0.48958 | 0.68779 | *MASP1* |

| | | | | | | | | | |
|---|---|---|---|---|---|---|---|---|---|
| ENSG00000104325 | 8 | 91003633 | 91074320 | 183 | 20 | 32453 | -0.48966 | 0.68781 | *DECR1* |
| ENSG00000164896 | 7 | 150763711 | 150787953 | 69 | 18 | 42998 | -0.48993 | 0.68791 | *FASTK* |
| ENSG00000111087 | 12 | 57843918 | 57876045 | 77 | 18 | 37609 | -0.48999 | 0.68793 | *GLI1* |
| ENSG00000153046 | 6 | 4696393 | 4965785 | 795 | 120 | 46046 | -0.49003 | 0.68795 | *CDYL* |
| ENSG00000182318 | 19 | 58828385 | 58863698 | 88 | 6 | 22863 | -0.49009 | 0.68796 | *ZSCAN22* |
| ENSG00000182854 | 15 | 102348366 | 102369350 | 60 | 12 | 42919 | -0.49017 | 0.68799 | *OR4F15* |
| ENSG00000187513 | 1 | 35248599 | 35271348 | 91 | 15 | 40775 | -0.49043 | 0.68809 | *GJA4* |
| ENSG00000111490 | 12 | 65164589 | 65284812 | 187 | 40 | 44773 | -0.49083 | 0.68823 | *TBC1D30* |
| ENSG00000188931 | 1 | 161324521 | 161347664 | 133 | 5 | 22091 | -0.49111 | 0.68833 | *C1orf192* |
| ENSG00000198633 | 19 | 52922440 | 52965568 | 140 | 21 | 32109 | -0.49131 | 0.6884 | *ZNF534* |
| ENSG00000113889 | 3 | 186425065 | 186471743 | 303 | 33 | 52384 | -0.49149 | 0.68846 | *KNG1* |
| ENSG00000106144 | 7 | 142975308 | 143014789 | 36 | 16 | 48015 | -0.49164 | 0.68851 | *CASP2* |
| ENSG00000067596 | 17 | 41551233 | 41631831 | 167 | 10 | 53783 | -0.49168 | 0.68853 | *DHX8* |
| ENSG00000164904 | 5 | 125867533 | 125941110 | 233 | 42 | 42659 | -0.49178 | 0.68856 | *ALDH7A1* |
| ENSG00000167483 | 19 | 17624110 | 17674648 | 140 | 38 | 45032 | -0.49176 | 0.68856 | *FAM129C* |
| ENSG00000100281 | 22 | 35643445 | 35701800 | 161 | 8 | 27894 | -0.49215 | 0.68869 | *HMGXB4* |
| ENSG00000148834 | 10 | 105985114 | 106037217 | 130 | 28 | 47011 | -0.49226 | 0.68873 | *GSTO1* |
| ENSG00000269449 | 7 | 95161457 | 95183854 | 114 | 11 | 45403 | -0.49228 | 0.68874 | *AC002451.1* |
| ENSG00000112936 | 5 | 40899354 | 40993041 | 388 | 45 | 43647 | -0.49247 | 0.6888 | *C7* |
| ENSG00000047617 | 12 | 5631035 | 6065398 | 1570 | 157 | 48259 | -0.49323 | 0.68908 | *ANO2* |
| ENSG00000113231 | 5 | 76496274 | 76735632 | 768 | 101 | 42528 | -0.49339 | 0.68913 | *PDE8B* |
| ENSG00000196371 | 11 | 94267017 | 94293063 | 70 | 15 | 42888 | -0.49415 | 0.6894 | *FUT4* |
| ENSG00000139154 | 12 | 19546979 | 19883735 | 1829 | 142 | 43679 | -0.49458 | 0.68955 | *AEBP2* |
| ENSG00000167863 | 17 | 73024958 | 73053080 | 92 | 23 | 37768 | -0.49482 | 0.68964 | *ATP5H* |
| ENSG00000177383 | 3 | 184418156 | 184439836 | 51 | 14 | 42774 | -0.49498 | 0.68969 | *MAGEF1* |
| ENSG00000148362 | 9 | 139876870 | 139898436 | 59 | 8 | 54121 | -0.49544 | 0.68986 | *C9orf142* |
| ENSG00000122136 | 9 | 138427985 | 138451815 | 99 | 21 | 37777 | -0.49551 | 0.68988 | *OBP2A* |
| ENSG00000123066 | 12 | 116385711 | 116725143 | 652 | 56 | 52215 | -0.49552 | 0.68988 | *MED13L* |
| ENSG00000180817 | 10 | 71952586 | 72003667 | 211 | 30 | 33362 | -0.49564 | 0.68993 | *PPA1* |
| ENSG00000206561 | 3 | 15481640 | 15573258 | 310 | 31 | 43945 | -0.49571 | 0.68995 | *COLQ* |
| ENSG00000131469 | 17 | 41140290 | 41164976 | 46 | 9 | 49097 | -0.49577 | 0.68997 | *RPL27* |
| ENSG00000267873 | 12 | 52699128 | 52719790 | 146 | 13 | 54114 | -0.49612 | 0.69009 | *AC121757.1* |
| ENSG00000171408 | 6 | 136162834 | 136526712 | 714 | 103 | 43359 | -0.49639 | 0.69019 | *PDE7B* |
| ENSG00000204779 | 9 | 70165707 | 70188815 | 4 | 2 | 42743 | -0.49669 | 0.6903 | *FOXD4L5* |
| ENSG00000198799 | 1 | 113605831 | 113684882 | 157 | 29 | 43488 | -0.49687 | 0.69036 | *LRIG2* |
| ENSG00000182134 | 1 | 151732583 | 151773892 | 114 | 13 | 45212 | -0.49689 | 0.69037 | *TDRKH* |

| | | | | | | | | | |
|---|---|---|---|---|---|---|---|---|---|
| ENSG00000269738 | 9 | 125138344 | 125158928 | 69 | 10 | 34419 | -0.49691 | 0.69037 | *AL162424.1* |
| ENSG00000251380 | 5 | 134769905 | 134793038 | 93 | 17 | 35855 | -0.49705 | 0.69042 | *C5orf20* |
| ENSG00000226742 | 18 | 77714561 | 77740822 | 98 | 19 | 48046 | -0.4971 | 0.69044 | *HSBP1L1* |
| ENSG00000198612 | 2 | 237983955 | 238019109 | 89 | 18 | 38046 | -0.49718 | 0.69047 | *COPS8* |
| ENSG00000116747 | 1 | 193018552 | 193070907 | 98 | 13 | 52350 | -0.49734 | 0.69052 | *TROVE2* |
| ENSG00000182103 | 11 | 82433053 | 82454906 | 62 | 13 | 31575 | -0.49752 | 0.69059 | *FAM181B* |
| ENSG00000174788 | 19 | 7686497 | 7708634 | 69 | 9 | 53734 | -0.49752 | 0.69059 | *PCP2* |
| ENSG00000171121 | 3 | 178947530 | 178994790 | 190 | 31 | 39365 | -0.49786 | 0.69071 | *KCNMB3* |
| ENSG00000129116 | 4 | 169408217 | 169859608 | 1510 | 179 | 46126 | -0.49789 | 0.69072 | *PALLD* |
| ENSG00000128016 | 19 | 39887453 | 39910052 | 45 | 9 | 42486 | -0.49801 | 0.69076 | *ZFP36* |
| ENSG00000067334 | 1 | 94323373 | 94355474 | 151 | 17 | 42972 | -0.49849 | 0.69093 | *DNTTIP2* |
| ENSG00000104783 | 19 | 44260685 | 44295409 | 140 | 23 | 49860 | -0.49855 | 0.69095 | *KCNN4* |
| ENSG00000232748 | 16 | 31062813 | 31083451 | 43 | 8 | 21244 | -0.49915 | 0.69116 | *ZNF668* |
| ENSG00000141086 | 16 | 67951543 | 67976317 | 21 | 5 | 39992 | -0.4992 | 0.69118 | *CTRL* |
| ENSG00000196417 | 19 | 53883046 | 53940574 | 394 | 61 | 43051 | -0.49933 | 0.69123 | *ZNF765* |
| ENSG00000214248 | 19 | 7953689 | 7978427 | 90 | 18 | 44676 | -0.49973 | 0.69137 | *AC010336.1* |
| ENSG00000100650 | 14 | 70183617 | 70248722 | 163 | 25 | 46535 | -0.50032 | 0.69158 | *SRSF5* |
| ENSG00000147571 | 8 | 67078620 | 67100960 | 57 | 9 | 35358 | -0.50107 | 0.69184 | *CRH* |
| ENSG00000171060 | 8 | 10520147 | 10568103 | 116 | 25 | 36094 | -0.50123 | 0.6919 | *C8orf74* |
| ENSG00000189134 | 6 | 28217098 | 28238736 | 49 | 11 | 45294 | -0.50137 | 0.69194 | *NKAPL* |
| ENSG00000120820 | 12 | 104372762 | 104467961 | 322 | 55 | 48062 | -0.50143 | 0.69196 | *GLT8D2* |
| ENSG00000163825 | 3 | 46528981 | 46552439 | 72 | 10 | 40598 | -0.50156 | 0.69201 | *RTP3* |
| ENSG00000019186 | 20 | 52759988 | 52800512 | 221 | 35 | 43723 | -0.50222 | 0.69224 | *CYP24A1* |
| ENSG00000166086 | 11 | 133928820 | 134031896 | 369 | 29 | 48497 | -0.50292 | 0.69249 | *JAM3* |
| ENSG00000181754 | 1 | 110036797 | 110062360 | 69 | 13 | 45106 | -0.50302 | 0.69253 | *AMIGO1* |
| ENSG00000198900 | 20 | 39647458 | 39763127 | 187 | 33 | 49025 | -0.50309 | 0.69255 | *TOP1* |
| ENSG00000161920 | 17 | 4624723 | 4646905 | 86 | 18 | 47009 | -0.50315 | 0.69257 | *MED11* |
| ENSG00000171202 | 11 | 85349011 | 85377591 | 53 | 9 | 46066 | -0.50367 | 0.69275 | *TMEM126A* |
| ENSG00000137747 | 11 | 117761358 | 117810174 | 191 | 34 | 53601 | -0.50388 | 0.69283 | *TMPRSS13* |
| ENSG00000103154 | 16 | 83992237 | 84046381 | 413 | 46 | 34106 | -0.50394 | 0.69285 | *NECAB2* |
| ENSG00000126870 | 7 | 158639269 | 158759438 | 437 | 46 | 47655 | -0.50416 | 0.69293 | *WDR60* |
| ENSG00000204220 | 6 | 33247079 | 33276178 | 85 | 20 | 30533 | -0.50434 | 0.69299 | *PFDN6* |
| ENSG00000188486 | 11 | 118954564 | 118976177 | 53 | 11 | 33164 | -0.50455 | 0.69306 | *H2AFX* |
| ENSG00000136367 | 14 | 23980066 | 24035401 | 100 | 36 | 42674 | -0.50482 | 0.69316 | *ZFHX2* |
| ENSG00000176463 | 15 | 92386925 | 92725665 | 1227 | 148 | 46242 | -0.50485 | 0.69317 | *SLCO3A1* |
| ENSG00000189377 | 19 | 42922440 | 42957200 | 36 | 7 | 38412 | -0.50487 | 0.69317 | *CXCL17* |

| | | | | | | | | | |
|---|---|---|---|---|---|---|---|---|---|
| ENSG00000159210 | 17 | 46996678 | 47032479 | 133 | 11 | 28476 | -0.50496 | 0.69321 | *SNF8* |
| ENSG00000135413 | 12 | 55014595 | 55038679 | 50 | 13 | 48346 | -0.50525 | 0.69331 | *LACRT* |
| ENSG00000075429 | 17 | 64821235 | 64891603 | 296 | 72 | 51590 | -0.50525 | 0.69331 | *CACNG5* |
| ENSG00000161277 | 19 | 36515887 | 36555664 | 107 | 21 | 52108 | -0.50562 | 0.69344 | *THAP8* |
| ENSG00000165511 | 10 | 45483146 | 45506336 | 52 | 15 | 45284 | -0.50609 | 0.6936 | *C10orf25* |
| ENSG00000142330 | 2 | 241516133 | 241567122 | 227 | 28 | 50534 | -0.50619 | 0.69364 | *CAPN10* |
| ENSG00000204049 | 10 | 79616633 | 79639368 | 75 | 10 | 53651 | -0.50648 | 0.69374 | *AL391421.1* |
| ENSG00000182199 | 12 | 57613110 | 57638718 | 38 | 13 | 42712 | -0.50661 | 0.69379 | *SHMT2* |
| ENSG00000069020 | 5 | 65882176 | 66475423 | 1721 | 174 | 41739 | -0.50677 | 0.69384 | *MAST4* |
| ENSG00000184348 | 6 | 27795658 | 27816117 | 45 | 12 | 37227 | -0.50685 | 0.69387 | *HIST1H2AK* |
| ENSG00000114942 | 2 | 207014309 | 207037652 | 70 | 8 | 33142 | -0.50742 | 0.69407 | *EEF1B2* |
| ENSG00000165685 | 12 | 10313141 | 10354400 | 181 | 35 | 36590 | -0.5076 | 0.69413 | *TMEM52B* |
| ENSG00000051108 | 16 | 56955960 | 56987798 | 102 | 15 | 50149 | -0.50775 | 0.69419 | *HERPUD1* |
| ENSG00000150672 | 11 | 83156055 | 85348966 | 6845 | 374 | 47158 | -0.50788 | 0.69423 | *DLG2* |
| ENSG00000257138 | 7 | 141662431 | 141683573 | 16 | 8 | 50630 | -0.50798 | 0.69427 | *TAS2R38* |
| ENSG00000170542 | 6 | 2877500 | 2913514 | 134 | 26 | 51147 | -0.50805 | 0.69429 | *SERPINB9* |
| ENSG00000163528 | 3 | 14143580 | 14176370 | 70 | 17 | 44955 | -0.50836 | 0.6944 | *CHCHD4* |
| ENSG00000216490 | 19 | 18273972 | 18298927 | 68 | 13 | 52976 | -0.50865 | 0.6945 | *IFI30* |
| ENSG00000022355 | 5 | 161264197 | 161336975 | 206 | 24 | 43766 | -0.50866 | 0.69451 | *GABRA1* |
| ENSG00000198015 | 12 | 93851264 | 93907545 | 206 | 17 | 49251 | -0.50875 | 0.69454 | *MRPL42* |
| ENSG00000083544 | 13 | 60960591 | 61158012 | 518 | 40 | 55386 | -0.50884 | 0.69457 | *TDRD3* |
| ENSG00000108788 | 17 | 40709086 | 40735257 | 58 | 11 | 54192 | -0.50924 | 0.69471 | *MLX* |
| ENSG00000268716 | 12 | 52912915 | 52932980 | 77 | 13 | 33123 | -0.50926 | 0.69472 | *AC055736.1* |
| ENSG00000204920 | 19 | 44462014 | 44512477 | 198 | 27 | 35953 | -0.50929 | 0.69473 | *ZNF155* |
| ENSG00000175793 | 1 | 27179633 | 27200947 | 43 | 7 | 50128 | -0.50953 | 0.69481 | *SFN* |
| ENSG00000085377 | 6 | 105715440 | 105860959 | 454 | 61 | 37857 | -0.50978 | 0.6949 | *PREP* |
| ENSG00000174606 | 1 | 213155524 | 213199168 | 64 | 13 | 43761 | -0.50986 | 0.69493 | *ANGEL2* |
| ENSG00000134077 | 3 | 9394526 | 9438475 | 131 | 21 | 40192 | -0.5099 | 0.69494 | *THUMPD3* |
| ENSG00000262621 | 16 | 3405099 | 3546960 | 570 | 54 | 47871 | -0.51016 | 0.69503 | *NAA60* |
| ENSG00000106772 | 9 | 79216292 | 79531003 | 1077 | 119 | 43960 | -0.51032 | 0.69509 | *PRUNE2* |
| ENSG00000152409 | 5 | 78522012 | 78633038 | 262 | 24 | 32073 | -0.51051 | 0.69515 | *JMY* |
| ENSG00000164889 | 7 | 150744297 | 150783614 | 91 | 22 | 50009 | -0.51069 | 0.69522 | *SLC4A2* |
| ENSG00000169660 | 17 | 80366194 | 80410521 | 153 | 29 | 44283 | -0.51136 | 0.69545 | *HEXDC* |
| ENSG00000198356 | 19 | 12837953 | 12869137 | 25 | 17 | 48160 | -0.51143 | 0.69548 | *ASNA1* |
| ENSG00000129654 | 17 | 74122414 | 74147380 | 48 | 14 | 44776 | -0.51166 | 0.69556 | *FOXJ1* |
| ENSG00000237763 | 1 | 104187912 | 104217173 | 5 | 3 | 43505 | -0.51189 | 0.69563 | *AMY1A* |

| | | | | | | | | | |
|---|---|---|---|---|---|---|---|---|---|
| ENSG00000101096 | 20 | 49993494 | 50189370 | 605 | 128 | 44487 | -0.51231 | 0.69578 | *NFATC2* |
| ENSG00000164855 | 7 | 1571871 | 1610457 | 145 | 22 | 52419 | -0.51254 | 0.69586 | *TMEM184A* |
| ENSG00000214788 | 11 | 59695928 | 59722807 | 28 | 14 | 51879 | -0.51278 | 0.69595 | *OOSP1* |
| ENSG00000186965 | 21 | 31849362 | 31869755 | 36 | 4 | 53450 | -0.5134 | 0.69616 | *KRTAP19-2* |
| ENSG00000183401 | 19 | 11445360 | 11475620 | 67 | 14 | 37025 | -0.51354 | 0.69621 | *CCDC159* |
| ENSG00000205221 | 2 | 36913833 | 37051935 | 560 | 95 | 44175 | -0.51381 | 0.69631 | *VIT* |
| ENSG00000153575 | 15 | 22823395 | 22883892 | 93 | 26 | 45749 | -0.51403 | 0.69638 | *TUBGCP5* |
| ENSG00000189266 | 1 | 24275599 | 24299952 | 13 | 4 | 29125 | -0.51413 | 0.69642 | *PNRC2* |
| ENSG00000164332 | 5 | 158680089 | 158723044 | 93 | 17 | 50751 | -0.51423 | 0.69645 | *UBLCP1* |
| ENSG00000104164 | 15 | 45869321 | 45918197 | 87 | 14 | 48472 | -0.51425 | 0.69646 | *BLOC1S6* |
| ENSG00000198746 | 1 | 27206979 | 27236957 | 49 | 11 | 54595 | -0.51439 | 0.69651 | *GPATCH3* |
| ENSG00000106692 | 9 | 108310411 | 108413399 | 251 | 37 | 41350 | -0.51446 | 0.69653 | *FKTN* |
| ENSG00000081818 | 5 | 140491581 | 140515201 | 48 | 7 | 49606 | -0.51463 | 0.69659 | *PCDHB4* |
| ENSG00000101608 | 18 | 3237479 | 3266234 | 127 | 18 | 36665 | -0.51486 | 0.69667 | *MYL12A* |
| ENSG00000187699 | 2 | 190734335 | 191078210 | 857 | 113 | 40987 | -0.51491 | 0.69669 | *C2orf88* |
| ENSG00000000938 | 1 | 27928575 | 27971788 | 65 | 25 | 42586 | -0.5156 | 0.69693 | *FGR* |
| ENSG00000171428 | 8 | 18017986 | 18091198 | 418 | 54 | 42332 | -0.51574 | 0.69698 | *NAT1* |
| ENSG00000197555 | 14 | 71777166 | 72217946 | 719 | 58 | 50568 | -0.51575 | 0.69698 | *SIPA1L1* |
| ENSG00000077009 | 19 | 3923101 | 3952414 | 73 | 30 | 40013 | -0.51578 | 0.697 | *NMRK2* |
| ENSG00000124006 | 2 | 220405451 | 220446581 | 130 | 26 | 46616 | -0.51614 | 0.69712 | *OBSL1* |
| ENSG00000135447 | 12 | 54959171 | 54992443 | 72 | 21 | 43745 | -0.51614 | 0.69712 | *PPP1R1A* |
| ENSG00000166979 | 21 | 33774314 | 33897707 | 436 | 52 | 45228 | -0.51613 | 0.69712 | *EVA1C* |
| ENSG00000144847 | 3 | 118609404 | 118874915 | 831 | 85 | 44717 | -0.51621 | 0.69715 | *IGSF11* |
| ENSG00000251537 | 17 | 15464805 | 15564967 | 290 | 41 | 38406 | -0.5164 | 0.69721 | *RP11-385D13.1* |
| ENSG00000198160 | 1 | 67380578 | 67464302 | 221 | 19 | 48677 | -0.51648 | 0.69724 | *MIER1* |
| ENSG00000162614 | 1 | 78344198 | 78419580 | 170 | 26 | 42151 | -0.51673 | 0.69733 | *NEXN* |
| ENSG00000064195 | 17 | 48057369 | 48082588 | 84 | 20 | 39669 | -0.51719 | 0.69749 | *DLX3* |
| ENSG00000103544 | 16 | 19556562 | 19728115 | 580 | 66 | 40332 | -0.51783 | 0.69771 | *C16orf62* |
| ENSG00000116017 | 19 | 915781 | 985939 | 289 | 54 | 39425 | -0.51781 | 0.69771 | *ARID3A* |
| ENSG00000182858 | 22 | 50283877 | 50322106 | 228 | 15 | 30902 | -0.51799 | 0.69777 | *ALG12* |
| ENSG00000105792 | 7 | 89864488 | 89950377 | 219 | 26 | 43511 | -0.51809 | 0.6978 | *C7orf63* |
| ENSG00000214556 | 17 | 36981341 | 37007708 | 101 | 18 | 41772 | -0.51813 | 0.69782 | *C17orf98* |
| ENSG00000100014 | 22 | 24656786 | 24823708 | 474 | 41 | 42257 | -0.51851 | 0.69795 | *SPECC1L* |
| ENSG00000100418 | 22 | 41984032 | 42027100 | 75 | 12 | 34579 | -0.5186 | 0.69798 | *DESI1* |
| ENSG00000163377 | 3 | 68770917 | 68991761 | 740 | 55 | 43739 | -0.51874 | 0.69803 | *FAM19A4* |
| ENSG00000242419 | 5 | 140854741 | 140902546 | 127 | 26 | 45832 | -0.51877 | 0.69804 | *PCDHGC4* |

| | | | | | | | | | |
|---|---|---|---|---|---|---|---|---|---|
| ENSG00000175018 | 10 | 127255091 | 127381713 | 286 | 51 | 32427 | -0.51889 | 0.69808 | *TEX36* |
| ENSG00000204038 | 10 | 81999466 | 82023395 | 170 | 12 | 52597 | -0.5191 | 0.69816 | *AL359195.1* |
| ENSG00000135569 | 6 | 132899731 | 132920877 | 106 | 18 | 42648 | -0.51922 | 0.6982 | *TAAR5* |
| ENSG00000121931 | 1 | 111479807 | 111516701 | 86 | 13 | 55201 | -0.51934 | 0.69824 | *LRIF1* |
| ENSG00000101680 | 18 | 6931743 | 7127813 | 675 | 122 | 40913 | -0.51938 | 0.69825 | *LAMA1* |
| ENSG00000137337 | 6 | 30657584 | 30695666 | 225 | 21 | 35511 | -0.51944 | 0.69827 | *MDC1* |
| ENSG00000164236 | 5 | 10554442 | 10660308 | 391 | 43 | 46343 | -0.51957 | 0.69832 | *ANKRD33B* |
| ENSG00000105808 | 7 | 102210093 | 102267204 | 3 | 2 | 43106 | -0.51987 | 0.69842 | *RASA4* |
| ENSG00000213626 | 2 | 30444397 | 30556596 | 413 | 61 | 47312 | -0.52012 | 0.69851 | *LBH* |
| ENSG00000143494 | 1 | 213113862 | 213175379 | 168 | 33 | 42113 | -0.52027 | 0.69856 | *VASH2* |
| ENSG00000175707 | 1 | 27266053 | 27296897 | 40 | 8 | 54925 | -0.52061 | 0.69868 | *C1orf172* |
| ENSG00000170465 | 12 | 52852300 | 52877569 | 190 | 11 | 37792 | -0.52062 | 0.69869 | *KRT6C* |
| ENSG00000131969 | 14 | 51328878 | 51381688 | 171 | 30 | 48874 | -0.52113 | 0.69886 | *ABHD12B* |
| ENSG00000206535 | 3 | 100110037 | 100185163 | 119 | 27 | 42848 | -0.5212 | 0.69889 | *LNP1* |
| ENSG00000154175 | 3 | 100458000 | 100722359 | 827 | 87 | 40913 | -0.52143 | 0.69897 | *ABI3BP* |
| ENSG00000116754 | 1 | 70661365 | 70728735 | 100 | 13 | 52410 | -0.52187 | 0.69912 | *SRSF11* |
| ENSG00000156256 | 21 | 30386950 | 30436809 | 90 | 20 | 44811 | -0.52197 | 0.69915 | *USP16* |
| ENSG00000203730 | 1 | 182357252 | 182379751 | 86 | 12 | 46221 | -0.5221 | 0.6992 | *TEDDM1* |
| ENSG00000151136 | 12 | 107702190 | 108063419 | 1071 | 130 | 49126 | -0.52275 | 0.69943 | *BTBD11* |
| ENSG00000135070 | 9 | 88869461 | 88907676 | 82 | 10 | 41055 | -0.52284 | 0.69946 | *ISCA1* |
| ENSG00000175054 | 3 | 142158077 | 142307668 | 387 | 26 | 52072 | -0.52295 | 0.6995 | *ATR* |
| ENSG00000173812 | 17 | 39835137 | 39858920 | 50 | 9 | 36703 | -0.52305 | 0.69953 | *EIF1* |
| ENSG00000132704 | 1 | 157705523 | 157756922 | 112 | 24 | 49353 | -0.52339 | 0.69965 | *FCRL2* |
| ENSG00000187097 | 14 | 74414713 | 74496102 | 348 | 33 | 33891 | -0.52367 | 0.69974 | *ENTPD5* |
| ENSG00000180785 | 11 | 4654650 | 4686718 | 216 | 28 | 44338 | -0.52377 | 0.69978 | *OR51E1* |
| ENSG00000104827 | 19 | 49516126 | 49537590 | 91 | 14 | 49849 | -0.52385 | 0.69981 | *CGB* |
| ENSG00000165066 | 8 | 41492697 | 41518855 | 103 | 17 | 55343 | -0.52402 | 0.69987 | *NKX6-3* |
| ENSG00000099365 | 16 | 30990577 | 31031949 | 56 | 11 | 27208 | -0.52417 | 0.69992 | *STX1B* |
| ENSG00000183778 | 21 | 40918369 | 41055064 | 496 | 62 | 48990 | -0.52438 | 0.69999 | *B3GALT5* |
| ENSG00000178950 | 4 | 833064 | 936161 | 446 | 55 | 54134 | -0.52443 | 0.70001 | *GAK* |
| ENSG00000155827 | 9 | 104286133 | 104335622 | 130 | 23 | 54133 | -0.52451 | 0.70004 | *RNF20* |
| ENSG00000086827 | 11 | 113593909 | 113654533 | 194 | 25 | 51131 | -0.52462 | 0.70008 | *ZW10* |
| ENSG00000125871 | 20 | 17939556 | 17981765 | 170 | 15 | 54247 | -0.5251 | 0.70024 | *MGME1* |
| ENSG00000058866 | 3 | 185813457 | 186090026 | 674 | 127 | 43508 | -0.52553 | 0.70039 | *DGKG* |
| ENSG00000184113 | 22 | 19500547 | 19525068 | 84 | 24 | 45806 | -0.52568 | 0.70044 | *CLDN5* |
| ENSG00000125910 | 19 | 3162344 | 3190329 | 107 | 31 | 48007 | -0.5257 | 0.70045 | *S1PR4* |

| | | | | | | | | | |
|---|---|---|---|---|---|---|---|---|---|
| ENSG00000176108 | 17 | 78955398 | 78993317 | 186 | 29 | 42216 | -0.52587 | 0.70051 | *CHMP6* |
| ENSG00000184611 | 2 | 163217917 | 163705240 | 1024 | 87 | 37769 | -0.52591 | 0.70052 | *KCNH7* |
| ENSG00000178804 | 3 | 129252057 | 129280310 | 67 | 13 | 37241 | -0.52626 | 0.70065 | *H1FOO* |
| ENSG00000143569 | 1 | 154182655 | 154253986 | 129 | 18 | 45948 | -0.52677 | 0.70082 | *UBAP2L* |
| ENSG00000138356 | 2 | 201440591 | 201551787 | 360 | 48 | 48600 | -0.52697 | 0.70089 | *AOX1* |
| ENSG00000170881 | 8 | 125476979 | 125510155 | 83 | 17 | 48272 | -0.52708 | 0.70093 | *RNF139* |
| ENSG00000112462 | 6 | 29331200 | 29353068 | 114 | 17 | 36775 | -0.52717 | 0.70096 | *OR12D3* |
| ENSG00000114541 | 3 | 69209141 | 69601734 | 1398 | 138 | 44902 | -0.52729 | 0.701 | *FRMD4B* |
| ENSG00000163792 | 2 | 27361872 | 27386378 | 52 | 10 | 43711 | -0.52735 | 0.70103 | *TCF23* |
| ENSG00000134627 | 11 | 94267006 | 94364587 | 303 | 44 | 40661 | -0.52778 | 0.70117 | *PIWIL4* |
| ENSG00000109917 | 11 | 116638436 | 116668766 | 94 | 23 | 48817 | -0.52776 | 0.70117 | *ZNF259* |
| ENSG00000075240 | 22 | 46961909 | 47085688 | 648 | 66 | 40304 | -0.52819 | 0.70131 | *GRAMD4* |
| ENSG00000006712 | 19 | 39866492 | 39891835 | 51 | 14 | 43201 | -0.52832 | 0.70136 | *PAF1* |
| ENSG00000185811 | 7 | 50333720 | 50482799 | 348 | 60 | 49741 | -0.52845 | 0.70141 | *IKZF1* |
| ENSG00000198648 | 2 | 168800530 | 169114651 | 1031 | 82 | 36177 | -0.52878 | 0.70152 | *STK39* |
| ENSG00000110400 | 11 | 119484120 | 119609794 | 316 | 52 | 43710 | -0.52892 | 0.70157 | *PVRL1* |
| ENSG00000117054 | 1 | 76180036 | 76263260 | 346 | 30 | 39996 | -0.52895 | 0.70158 | *ACADM* |
| ENSG00000163328 | 2 | 175286966 | 175361822 | 190 | 25 | 41739 | -0.52966 | 0.70183 | *GPR155* |
| ENSG00000130638 | 22 | 46057678 | 46251187 | 537 | 53 | 52335 | -0.52967 | 0.70183 | *ATXN10* |
| ENSG00000196422 | 9 | 138360925 | 138390739 | 100 | 27 | 38788 | -0.5299 | 0.70191 | *PPP1R26* |
| ENSG00000184984 | 15 | 34250921 | 34367291 | 322 | 34 | 26231 | -0.5299 | 0.70191 | *CHRM5* |
| ENSG00000011478 | 19 | 46185741 | 46217247 | 82 | 18 | 50257 | -0.53001 | 0.70195 | *QPCTL* |
| ENSG00000125850 | 20 | 17927623 | 18049832 | 489 | 52 | 50952 | -0.53007 | 0.70197 | *OVOL2* |
| ENSG00000183748 | 10 | 17841362 | 17963178 | 38 | 12 | 23162 | -0.53028 | 0.70204 | *MRC1L1* |
| ENSG00000150281 | 16 | 30897928 | 30924881 | 31 | 11 | 35687 | -0.53042 | 0.70209 | *CTF1* |
| ENSG00000258466 | 14 | 68116537 | 68167084 | 124 | 18 | 37698 | -0.53067 | 0.70218 | *RP11-1012A1.4* |
| ENSG00000182077 | 10 | 27677116 | 27713297 | 178 | 19 | 28079 | -0.53075 | 0.7022 | *PTCHD3* |
| ENSG00000167315 | 18 | 47299869 | 47350330 | 142 | 20 | 45288 | -0.53079 | 0.70222 | *ACAA2* |
| ENSG00000133433 | 22 | 24289601 | 24313373 | 48 | 9 | 43665 | -0.53103 | 0.7023 | *GSTT2B* |
| ENSG00000132849 | 1 | 62198149 | 62639592 | 1494 | 171 | 46812 | -0.53113 | 0.70233 | *INADL* |
| ENSG00000205277 | 7 | 100602904 | 100672230 | 183 | 28 | 46704 | -0.53115 | 0.70234 | *MUC12* |
| ENSG00000014919 | 10 | 101461601 | 101501857 | 73 | 12 | 54779 | -0.53123 | 0.70237 | *COX15* |
| ENSG00000115414 | 2 | 216215163 | 216310895 | 349 | 41 | 39432 | -0.53128 | 0.70239 | *FN1* |
| ENSG00000170954 | 19 | 53601132 | 53646330 | 203 | 27 | 45383 | -0.53159 | 0.7025 | *ZNF415* |
| ENSG00000064270 | 16 | 84392133 | 84507793 | 859 | 131 | 47772 | -0.53181 | 0.70257 | *ATP2C2* |
| ENSG00000163866 | 1 | 35168338 | 35335417 | 629 | 99 | 47997 | -0.53186 | 0.70259 | *SMIM12* |

| | | | | | | | | | |
|---|---|---|---|---|---|---|---|---|---|
| ENSG00000198642 | 9 | 21319670 | 21345379 | 68 | 12 | 55330 | -0.53194 | 0.70262 | *KLHL9* |
| ENSG00000184164 | 22 | 50301815 | 50331188 | 209 | 17 | 35320 | -0.53205 | 0.70265 | *CRELD2* |
| ENSG00000184351 | 21 | 31842018 | 31862663 | 41 | 5 | 53924 | -0.5321 | 0.70267 | *KRTAP19-1* |
| ENSG00000172785 | 9 | 111041 | 198979 | 154 | 21 | 45108 | -0.53253 | 0.70282 | *CBWD1* |
| ENSG00000184507 | 15 | 34625516 | 34659938 | 136 | 19 | 50081 | -0.53336 | 0.70311 | *NUTM1* |
| ENSG00000174175 | 1 | 169548087 | 169609431 | 190 | 34 | 42202 | -0.53352 | 0.70316 | *SELP* |
| ENSG00000149531 | 20 | 29601857 | 29644010 | 125 | 20 | 46480 | -0.53359 | 0.70319 | *FRG1B* |
| ENSG00000171435 | 12 | 117880817 | 118416788 | 1927 | 256 | 42312 | -0.53379 | 0.70326 | *KSR2* |
| ENSG00000128917 | 15 | 41211538 | 41241237 | 46 | 13 | 31972 | -0.53393 | 0.70331 | *DLL4* |
| ENSG00000148335 | 9 | 132361163 | 132408209 | 160 | 27 | 46182 | -0.53398 | 0.70332 | *NTMT1* |
| ENSG00000124469 | 19 | 43074393 | 43109207 | 40 | 12 | 45577 | -0.53466 | 0.70356 | *CEACAM8* |
| ENSG00000137693 | 11 | 101971192 | 102114154 | 358 | 33 | 45375 | -0.53474 | 0.70359 | *YAP1* |
| ENSG00000143942 | 2 | 53984929 | 54012333 | 90 | 18 | 44204 | -0.53526 | 0.70376 | *CHAC2* |
| ENSG00000197110 | 19 | 39724246 | 39745646 | 76 | 14 | 41621 | -0.53531 | 0.70378 | *IFNL3* |
| ENSG00000182307 | 8 | 146267764 | 146291416 | 66 | 7 | 47246 | -0.5359 | 0.70399 | *C8orf33* |
| ENSG00000136869 | 9 | 120456610 | 120489149 | 90 | 21 | 48160 | -0.53601 | 0.70402 | *TLR4* |
| ENSG00000157131 | 1 | 57310479 | 57393894 | 361 | 42 | 35998 | -0.53637 | 0.70415 | *C8A* |
| ENSG00000171824 | 1 | 11116675 | 11169938 | 155 | 11 | 22602 | -0.53666 | 0.70425 | *EXOSC10* |
| ENSG00000164736 | 8 | 55360495 | 55383448 | 59 | 12 | 39269 | -0.53688 | 0.70432 | *SOX17* |
| ENSG00000163527 | 3 | 31564130 | 31689112 | 275 | 31 | 44268 | -0.53756 | 0.70456 | *STT3B* |
| ENSG00000270106 | 1 | 231654399 | 231964990 | 787 | 118 | 45903 | -0.53762 | 0.70458 | *TSNAX-DISC1* |
| ENSG00000111361 | 12 | 124094953 | 124128313 | 124 | 16 | 33436 | -0.53833 | 0.70483 | *EIF2B1* |
| ENSG00000184371 | 1 | 110442864 | 110483614 | 108 | 24 | 39715 | -0.53843 | 0.70486 | *CSF1* |
| ENSG00000166340 | 11 | 6624000 | 6650692 | 79 | 15 | 47913 | -0.53857 | 0.70491 | *TPP1* |
| ENSG00000186281 | 2 | 96677694 | 96715199 | 33 | 7 | 46291 | -0.53871 | 0.70496 | *GPAT2* |
| ENSG00000173269 | 10 | 88685297 | 88739238 | 192 | 33 | 49133 | -0.53885 | 0.705 | *MMRN2* |
| ENSG00000155545 | 5 | 56205429 | 56277502 | 273 | 29 | 43204 | -0.53893 | 0.70503 | *MIER3* |
| ENSG00000134853 | 4 | 55085264 | 55174414 | 228 | 19 | 47214 | -0.53931 | 0.70516 | *PDGFRA* |
| ENSG00000196739 | 9 | 116907840 | 117084791 | 680 | 143 | 46362 | -0.53986 | 0.70535 | *COL27A1* |
| ENSG00000141391 | 18 | 12397895 | 12442237 | 174 | 36 | 43928 | -0.53986 | 0.70535 | *SLMO1* |
| ENSG00000087903 | 19 | 5983175 | 6209583 | 693 | 68 | 36868 | -0.53987 | 0.70536 | *RFX2* |
| ENSG00000179915 | 2 | 50135643 | 51269674 | 3902 | 249 | 48680 | -0.54003 | 0.70541 | *NRXN1* |
| ENSG00000184321 | 11 | 5413827 | 5434777 | 214 | 14 | 31814 | -0.54014 | 0.70545 | *OR51J1* |
| ENSG00000104643 | 8 | 11131925 | 11195646 | 226 | 19 | 42521 | -0.54038 | 0.70553 | *MTMR9* |
| ENSG00000159202 | 17 | 46975731 | 47016418 | 138 | 11 | 30498 | -0.54094 | 0.70572 | *UBE2Z* |
| ENSG00000167664 | 19 | 4282229 | 4312428 | 115 | 26 | 50979 | -0.54097 | 0.70573 | *TMIGD2* |

| | | | | | | | | | |
|---|---|---|---|---|---|---|---|---|---|
| ENSG00000070729 | 16 | 57907503 | 58015020 | 387 | 84 | 43327 | -0.54145 | 0.7059 | *CNGB1* |
| ENSG00000106384 | 7 | 100828288 | 100854302 | 69 | 18 | 41092 | -0.54158 | 0.70595 | *MOGAT3* |
| ENSG00000136068 | 3 | 57984127 | 58167982 | 415 | 59 | 35193 | -0.54177 | 0.70601 | *FLNB* |
| ENSG00000135679 | 12 | 69191956 | 69249214 | 162 | 30 | 43984 | -0.54226 | 0.70618 | *MDM2* |
| ENSG00000152266 | 11 | 13503602 | 13527728 | 75 | 10 | 38476 | -0.54251 | 0.70627 | *PTH* |
| ENSG00000183130 | 1 | 248779420 | 248800491 | 7 | 3 | 43628 | -0.54355 | 0.70663 | *OR2T11* |
| ENSG00000144362 | 2 | 170540975 | 170568218 | 96 | 11 | 56617 | -0.54385 | 0.70673 | *PHOSPHO2* |
| ENSG00000185627 | 11 | 226546 | 262983 | 196 | 23 | 47732 | -0.54394 | 0.70676 | *PSMD13* |
| ENSG00000161681 | 19 | 51155084 | 51232707 | 197 | 58 | 45542 | -0.54408 | 0.70681 | *SHANK1* |
| ENSG00000013441 | 2 | 201707732 | 201739422 | 76 | 22 | 48453 | -0.54414 | 0.70683 | *CLK1* |
| ENSG00000187021 | 10 | 118339897 | 118378687 | 129 | 25 | 52479 | -0.54417 | 0.70684 | *PNLIPRP1* |
| ENSG00000099783 | 19 | 8499651 | 8563998 | 183 | 26 | 28985 | -0.54458 | 0.70698 | *HNRNPM* |
| ENSG00000258989 | 14 | 61985846 | 62134682 | 408 | 74 | 38981 | -0.5448 | 0.70705 | *RP11-47I22.4* |
| ENSG00000146109 | 6 | 26587180 | 26610278 | 78 | 14 | 48913 | -0.54488 | 0.70708 | *ABT1* |
| ENSG00000157500 | 3 | 57251765 | 57317496 | 119 | 17 | 41507 | -0.54501 | 0.70713 | *APPL1* |
| ENSG00000138413 | 2 | 209090951 | 209140798 | 132 | 28 | 40289 | -0.54517 | 0.70718 | *IDH1* |
| ENSG00000027075 | 14 | 61644277 | 62027694 | 1164 | 147 | 38170 | -0.54521 | 0.7072 | *PRKCH* |
| ENSG00000102595 | 13 | 96443834 | 96715736 | 644 | 43 | 47910 | -0.54526 | 0.70721 | *UGGT2* |
| ENSG00000104447 | 8 | 116410724 | 116831899 | 613 | 51 | 41868 | -0.54544 | 0.70727 | *TRPS1* |
| ENSG00000165572 | 13 | 41691705 | 41716882 | 22 | 7 | 42492 | -0.54549 | 0.70729 | *KBTBD6* |
| ENSG00000166860 | 12 | 57382618 | 57410230 | 48 | 9 | 55127 | -0.5457 | 0.70737 | *ZBTB39* |
| ENSG00000152689 | 2 | 33651391 | 33799817 | 545 | 110 | 46402 | -0.54577 | 0.70739 | *RASGRP3* |
| ENSG00000131591 | 1 | 1007198 | 1061741 | 211 | 22 | 35915 | -0.5458 | 0.7074 | *C1orf159* |
| ENSG00000135951 | 2 | 99603724 | 99781427 | 571 | 30 | 36989 | -0.54608 | 0.70749 | *TSGA10* |
| ENSG00000179133 | 10 | 23546124 | 23643774 | 170 | 22 | 39759 | -0.5461 | 0.7075 | *C10orf67* |
| ENSG00000198700 | 1 | 201788269 | 201863422 | 143 | 23 | 51629 | -0.54641 | 0.70761 | *IPO9* |
| ENSG00000174130 | 4 | 38815336 | 38868438 | 178 | 19 | 28573 | -0.54643 | 0.70761 | *TLR6* |
| ENSG00000228607 | 11 | 113640469 | 113661222 | 72 | 11 | 51106 | -0.54641 | 0.70761 | *CLDN25* |
| ENSG00000144857 | 3 | 112919850 | 113016303 | 355 | 62 | 43914 | -0.54653 | 0.70765 | *BOC* |
| ENSG00000166974 | 18 | 32546892 | 32733434 | 506 | 64 | 40792 | -0.54663 | 0.70768 | *MAPRE2* |
| ENSG00000187855 | 12 | 108158162 | 108180421 | 70 | 15 | 39281 | -0.54671 | 0.70771 | *ASCL4* |
| ENSG00000134333 | 11 | 18405935 | 18439972 | 151 | 15 | 42531 | -0.54701 | 0.70781 | *LDHA* |
| ENSG00000132294 | 8 | 132906335 | 133035889 | 440 | 42 | 50110 | -0.54726 | 0.7079 | *EFR3A* |
| ENSG00000268714 | 5 | 115348088 | 115404625 | 326 | 44 | 39810 | -0.5473 | 0.70791 | *CTD-2287O16.3* |
| ENSG00000173011 | 4 | 7033626 | 7069679 | 125 | 22 | 46641 | -0.54747 | 0.70797 | *TADA2B* |
| ENSG00000140941 | 16 | 87407601 | 87448385 | 145 | 31 | 42470 | -0.54753 | 0.70799 | *MAP1LC3B* |

| | | | | | | | | | |
|---|---|---|---|---|---|---|---|---|---|
| ENSG00000115592 | 2 | 219677106 | 219706809 | 24 | 11 | 51649 | -0.54763 | 0.70803 | *PRKAG3* |
| ENSG00000090376 | 12 | 66572659 | 66658402 | 189 | 39 | 40383 | -0.54764 | 0.70803 | *IRAK3* |
| ENSG00000119715 | 14 | 76766957 | 76978178 | 882 | 138 | 44979 | -0.54783 | 0.7081 | *ESRRB* |
| ENSG00000237172 | 16 | 67172008 | 67195117 | 17 | 10 | 44303 | -0.54793 | 0.70813 | *B3GNT9* |
| ENSG00000172497 | 5 | 80615824 | 80699998 | 310 | 52 | 47445 | -0.54836 | 0.70828 | *ACOT12* |
| ENSG00000117385 | 1 | 43202006 | 43242755 | 99 | 19 | 53922 | -0.54847 | 0.70831 | *LEPRE1* |
| ENSG00000171840 | 12 | 663462 | 782945 | 576 | 71 | 48334 | -0.54857 | 0.70835 | *NINJ2* |
| ENSG00000105641 | 19 | 17972782 | 18015983 | 127 | 20 | 41961 | -0.54872 | 0.7084 | *SLC5A5* |
| ENSG00000104413 | 8 | 95643302 | 95729694 | 331 | 22 | 33581 | -0.54894 | 0.70848 | *ESRP1* |
| ENSG00000268403 | 11 | 9471747 | 9492010 | 53 | 9 | 47305 | -0.54894 | 0.70848 | *AC132192.1* |
| ENSG00000111647 | 12 | 100412233 | 100546626 | 393 | 42 | 42372 | -0.54912 | 0.70854 | *UHRF1BP1L* |
| ENSG00000177889 | 12 | 93789449 | 93846038 | 113 | 30 | 50239 | -0.54914 | 0.70855 | *UBE2N* |
| ENSG00000111875 | 6 | 119205384 | 119240332 | 106 | 18 | 37631 | -0.54954 | 0.70868 | *ASF1A* |
| ENSG00000234972 | 17 | 34570902 | 34767050 | 7 | 4 | 42972 | -0.5496 | 0.7087 | *TBC1D3C* |
| ENSG00000116668 | 1 | 185116212 | 185270897 | 219 | 24 | 39413 | -0.54974 | 0.70875 | *SWT1* |
| ENSG00000055208 | 6 | 149529777 | 149742749 | 804 | 77 | 41885 | -0.54981 | 0.70877 | *TAB2* |
| ENSG00000123384 | 12 | 57512276 | 57617134 | 166 | 29 | 45369 | -0.55005 | 0.70886 | *LRP1* |
| ENSG00000060642 | 1 | 27103963 | 27134889 | 39 | 11 | 53476 | -0.55016 | 0.7089 | *PIGV* |
| ENSG00000136250 | 7 | 36542456 | 36774154 | 769 | 108 | 47264 | -0.55025 | 0.70892 | *AOAH* |
| ENSG00000166224 | 10 | 72565717 | 72650930 | 292 | 37 | 54846 | -0.5504 | 0.70898 | *SGPL1* |
| ENSG00000137558 | 8 | 75726772 | 75777264 | 156 | 20 | 54023 | -0.55042 | 0.70899 | *PI15* |
| ENSG00000189350 | 2 | 29169477 | 29294239 | 545 | 70 | 43560 | -0.55077 | 0.7091 | *FAM179A* |
| ENSG00000197614 | 12 | 8779942 | 8825484 | 129 | 13 | 49111 | -0.55096 | 0.70917 | *MFAP5* |
| ENSG00000165801 | 14 | 21528429 | 21568399 | 107 | 24 | 49330 | -0.55113 | 0.70923 | *ARHGEF40* |
| ENSG00000006116 | 16 | 24256874 | 24384122 | 439 | 88 | 51587 | -0.55141 | 0.70932 | *CACNG3* |
| ENSG00000176842 | 16 | 54954774 | 54978397 | 66 | 12 | 44221 | -0.5515 | 0.70935 | *IRX5* |
| ENSG00000105135 | 19 | 15215795 | 15246596 | 107 | 18 | 51821 | -0.55226 | 0.70962 | *ILVBL* |
| ENSG00000118217 | 1 | 161726084 | 161943860 | 739 | 40 | 30952 | -0.55267 | 0.70976 | *ATF6* |
| ENSG00000116329 | 1 | 29128654 | 29200208 | 230 | 32 | 37993 | -0.55282 | 0.70981 | *OPRD1* |
| ENSG00000118855 | 3 | 158439987 | 158557508 | 357 | 37 | 54355 | -0.55296 | 0.70985 | *MFSD1* |
| ENSG00000006744 | 17 | 12885708 | 12931504 | 261 | 17 | 52267 | -0.55305 | 0.70988 | *ELAC2* |
| ENSG00000100211 | 22 | 39042641 | 39079859 | 176 | 10 | 48784 | -0.55304 | 0.70988 | *CBY1* |
| ENSG00000116741 | 1 | 192768169 | 192791403 | 49 | 10 | 47841 | -0.55307 | 0.70989 | *RGS2* |
| ENSG00000176049 | 5 | 146957990 | 147172338 | 538 | 55 | 33978 | -0.55313 | 0.70991 | *JAKMIP2* |
| ENSG00000244617 | 2 | 70177226 | 70199397 | 34 | 5 | 17723 | -0.55336 | 0.70999 | *ASPRV1* |
| ENSG00000161551 | 19 | 52349055 | 52404203 | 283 | 20 | 29647 | -0.55335 | 0.70999 | *ZNF577* |

| | | | | | | | | | |
|---|---|---|---|---|---|---|---|---|---|
| ENSG00000212916 | 1 | 232930643 | 232956092 | 67 | 17 | 46560 | -0.55343 | 0.71002 | *MAP10* |
| ENSG00000170379 | 7 | 143308043 | 143437502 | 35 | 10 | 45743 | -0.55368 | 0.7101 | *FAM115C* |
| ENSG00000005961 | 17 | 42439548 | 42476873 | 48 | 10 | 51138 | -0.55403 | 0.71022 | *ITGA2B* |
| ENSG00000006831 | 12 | 1787740 | 1907844 | 384 | 37 | 50361 | -0.5541 | 0.71024 | *ADIPOR2* |
| ENSG00000163331 | 2 | 159641829 | 159729293 | 395 | 42 | 37737 | -0.5541 | 0.71025 | *DAPL1* |
| ENSG00000177627 | 12 | 48866286 | 48900295 | 131 | 17 | 31865 | -0.55438 | 0.71034 | *C12orf54* |
| ENSG00000178445 | 9 | 6522464 | 6655650 | 587 | 91 | 41854 | -0.55476 | 0.71047 | *GLDC* |
| ENSG00000149218 | 11 | 94812974 | 94875809 | 245 | 46 | 49123 | -0.55534 | 0.71067 | *ENDOD1* |
| ENSG00000080546 | 6 | 109297640 | 109426022 | 306 | 14 | 49549 | -0.55541 | 0.71069 | *SESN1* |
| ENSG00000173013 | 4 | 7032579 | 7054728 | 73 | 16 | 51888 | -0.5555 | 0.71072 | *CCDC96* |
| ENSG00000136021 | 12 | 100650918 | 100745502 | 186 | 26 | 51855 | -0.55613 | 0.71094 | *SCYL2* |
| ENSG00000230510 | 19 | 46974053 | 47114457 | 591 | 99 | 49306 | -0.55616 | 0.71095 | *PPP5D1* |
| ENSG00000169499 | 8 | 38748753 | 38841428 | 249 | 54 | 51689 | -0.55631 | 0.711 | *PLEKHA2* |
| ENSG00000143119 | 1 | 111405775 | 111452550 | 179 | 23 | 45097 | -0.55667 | 0.71112 | *CD53* |
| ENSG00000075035 | 12 | 108513248 | 108654314 | 408 | 59 | 49353 | -0.5567 | 0.71113 | *WSCD2* |
| ENSG00000078053 | 7 | 38413305 | 38681167 | 1262 | 126 | 45975 | -0.55676 | 0.71115 | *AMPH* |
| ENSG00000141437 | 18 | 29329525 | 29350843 | 76 | 15 | 44351 | -0.55745 | 0.71139 | *SLC25A52* |
| ENSG00000103313 | 16 | 3282028 | 3316627 | 102 | 17 | 43187 | -0.55787 | 0.71153 | *MEFV* |
| ENSG00000165548 | 14 | 77572911 | 77735838 | 688 | 89 | 44784 | -0.55794 | 0.71156 | *TMEM63C* |
| ENSG00000058804 | 1 | 54221133 | 54314533 | 181 | 23 | 44815 | -0.55813 | 0.71162 | *NDC1* |
| ENSG00000176435 | 14 | 38713308 | 38735574 | 37 | 9 | 46662 | -0.55846 | 0.71173 | *CLEC14A* |
| ENSG00000082641 | 17 | 46115691 | 46148849 | 107 | 17 | 49337 | -0.55861 | 0.71178 | *NFE2L1* |
| ENSG00000140563 | 15 | 94764767 | 95033632 | 893 | 148 | 48378 | -0.55871 | 0.71182 | *MCTP2* |
| ENSG00000183569 | 22 | 42939623 | 42980388 | 100 | 21 | 32549 | -0.55913 | 0.71196 | *SERHL2* |
| ENSG00000171204 | 11 | 85329629 | 85357580 | 53 | 10 | 47169 | -0.55938 | 0.71205 | *TMEM126B* |
| ENSG00000228570 | 10 | 81591114 | 81620632 | 43 | 11 | 37519 | -0.55946 | 0.71208 | *NUTM2E* |
| ENSG00000106733 | 9 | 77665489 | 77713133 | 217 | 30 | 55102 | -0.56012 | 0.7123 | *NMRK1* |
| ENSG00000166157 | 21 | 10896201 | 11039719 | 11 | 3 | 40108 | -0.56014 | 0.71231 | *TPTE* |
| ENSG00000173762 | 17 | 80262744 | 80285478 | 79 | 16 | 49446 | -0.56046 | 0.71242 | *CD7* |
| ENSG00000265690 | 16 | 67187288 | 67209718 | 16 | 8 | 48328 | -0.56061 | 0.71247 | *RP11-5A19.5* |
| ENSG00000103351 | 16 | 3540924 | 3599048 | 159 | 23 | 42775 | -0.56068 | 0.71249 | *CLUAP1* |
| ENSG00000185736 | 10 | 1218073 | 1789670 | 2595 | 278 | 44576 | -0.56083 | 0.71254 | *ADARB2* |
| ENSG00000185518 | 15 | 91633180 | 91854539 | 730 | 80 | 40632 | -0.56134 | 0.71272 | *SV2B* |
| ENSG00000164818 | 7 | 756338 | 839190 | 355 | 28 | 49653 | -0.5616 | 0.71281 | *HEATR2* |
| ENSG00000205791 | 12 | 12498342 | 12520001 | 75 | 19 | 47271 | -0.56166 | 0.71283 | *LOH12CR2* |
| ENSG00000173157 | 12 | 43737669 | 43955724 | 807 | 56 | 44616 | -0.56171 | 0.71284 | *ADAMTS20* |

| | | | | | | | | | |
|---|---|---|---|---|---|---|---|---|---|
| ENSG00000164182 | 5 | 60230956 | 60458853 | 624 | 38 | 51419 | -0.56174 | 0.71285 | *NDUFAF2* |
| ENSG00000181523 | 17 | 78170515 | 78204722 | 154 | 24 | 38690 | -0.56202 | 0.71295 | *SGSH* |
| ENSG00000176401 | 19 | 40011630 | 40033494 | 75 | 14 | 55725 | -0.5621 | 0.71298 | *EID2B* |
| ENSG00000232013 | 3 | 145772358 | 145794156 | 64 | 11 | 55217 | -0.56216 | 0.713 | *AC107021.1* |
| ENSG00000124249 | 20 | 43364421 | 43389675 | 68 | 13 | 51348 | -0.56223 | 0.71302 | *KCNK15* |
| ENSG00000254667 | 11 | 123291058 | 123316427 | 60 | 17 | 43958 | -0.56231 | 0.71305 | *AP000783.1* |
| ENSG00000255251 | 8 | 7387150 | 7409972 | 3 | 2 | 42926 | -0.5627 | 0.71318 | *PRR23D1* |
| ENSG00000165863 | 10 | 118413207 | 118439775 | 66 | 20 | 51485 | -0.56306 | 0.7133 | *C10orf82* |
| ENSG00000160352 | 19 | 21254965 | 21318073 | 286 | 20 | 35440 | -0.56321 | 0.71335 | *ZNF714* |
| ENSG00000178217 | 10 | 82287658 | 82416316 | 525 | 68 | 41804 | -0.56326 | 0.71337 | *SH2D4B* |
| ENSG00000177732 | 20 | 296207 | 320865 | 99 | 12 | 39861 | -0.56335 | 0.7134 | *SOX12* |
| ENSG00000135972 | 2 | 105644441 | 105726418 | 301 | 13 | 53343 | -0.56392 | 0.7136 | *MRPS9* |
| ENSG00000142556 | 19 | 52506018 | 52543493 | 120 | 16 | 36880 | -0.56412 | 0.71366 | *ZNF614* |
| ENSG00000100558 | 14 | 67843700 | 67888917 | 109 | 26 | 42300 | -0.56416 | 0.71368 | *PLEK2* |
| ENSG00000100146 | 22 | 38356693 | 38393429 | 47 | 10 | 44310 | -0.56448 | 0.71379 | *SOX10* |
| ENSG00000154134 | 11 | 124725282 | 124761366 | 116 | 28 | 44169 | -0.56452 | 0.7138 | *ROBO3* |
| ENSG00000177542 | 11 | 780475 | 808316 | 75 | 15 | 43589 | -0.56468 | 0.71385 | *SLC25A22* |
| ENSG00000167098 | 20 | 31561579 | 31602239 | 173 | 17 | 46505 | -0.56496 | 0.71395 | *SUN5* |
| ENSG00000138650 | 4 | 134060470 | 134139356 | 196 | 36 | 49787 | -0.565 | 0.71396 | *PCDH10* |
| ENSG00000185245 | 17 | 4825592 | 4848325 | 64 | 15 | 35697 | -0.56509 | 0.714 | *GP1BA* |
| ENSG00000175785 | 14 | 94174644 | 94264827 | 243 | 63 | 40805 | -0.56536 | 0.71409 | *PRIMA1* |
| ENSG00000130812 | 19 | 10193014 | 10223472 | 58 | 24 | 49607 | -0.56538 | 0.71409 | *ANGPTL6* |
| ENSG00000188581 | 17 | 39186811 | 39207713 | 95 | 13 | 37521 | -0.56588 | 0.71426 | *KRTAP1-1* |
| ENSG00000011451 | 19 | 15522319 | 15570762 | 58 | 21 | 49839 | -0.56672 | 0.71455 | *WIZ* |
| ENSG00000164944 | 8 | 95489921 | 95575757 | 376 | 28 | 42077 | -0.5669 | 0.71461 | *KIAA1429* |
| ENSG00000048545 | 6 | 42113144 | 42157794 | 184 | 29 | 44164 | -0.56745 | 0.7148 | *GUCA1A* |
| ENSG00000103037 | 16 | 58539383 | 58564431 | 84 | 12 | 37684 | -0.56764 | 0.71486 | *SETD6* |
| ENSG00000166439 | 11 | 74449913 | 74563458 | 245 | 26 | 39965 | -0.56768 | 0.71487 | *RNF169* |
| ENSG00000108784 | 17 | 40678190 | 40706467 | 77 | 17 | 46282 | -0.56777 | 0.71491 | *NAGLU* |
| ENSG00000163394 | 4 | 26473022 | 26502084 | 126 | 21 | 40019 | -0.56784 | 0.71493 | *CCKAR* |
| ENSG00000256206 | 11 | 14505329 | 14551890 | 90 | 13 | 54015 | -0.56795 | 0.71496 | *PSMA1* |
| ENSG00000149577 | 11 | 117039449 | 117078160 | 134 | 20 | 35634 | -0.56818 | 0.71504 | *SIDT2* |
| ENSG00000139549 | 12 | 49473204 | 49498602 | 39 | 13 | 46854 | -0.56843 | 0.71513 | *DHH* |
| ENSG00000094975 | 1 | 172491489 | 172590971 | 179 | 25 | 52849 | -0.56872 | 0.71523 | *SUCO* |
| ENSG00000162402 | 1 | 55522032 | 55690786 | 352 | 34 | 46204 | -0.56907 | 0.71535 | *USP24* |
| ENSG00000100276 | 22 | 29698922 | 29725668 | 52 | 15 | 49314 | -0.56912 | 0.71536 | *RASL10A* |

| | | | | | | | | | |
|---|---|---|---|---|---|---|---|---|---|
| ENSG00000178974 | 14 | 55728021 | 55838636 | 336 | 21 | 49190 | -0.56958 | 0.71552 | *FBXO34* |
| ENSG00000162391 | 1 | 55064855 | 55099229 | 140 | 27 | 48220 | -0.5696 | 0.71553 | *FAM151A* |
| ENSG00000057019 | 3 | 98504785 | 98630533 | 375 | 29 | 50948 | -0.56994 | 0.71564 | *DCBLD2* |
| ENSG00000123473 | 1 | 47705811 | 47789819 | 237 | 18 | 34687 | -0.57032 | 0.71577 | *STIL* |
| ENSG00000138400 | 2 | 207592487 | 207640271 | 169 | 18 | 27867 | -0.57034 | 0.71578 | *MDH1B* |
| ENSG00000173163 | 2 | 62105859 | 62384382 | 558 | 60 | 42979 | -0.57039 | 0.71579 | *COMMD1* |
| ENSG00000162702 | 1 | 200364068 | 200389184 | 49 | 11 | 51269 | -0.57069 | 0.7159 | *ZNF281* |
| ENSG00000197170 | 17 | 65324032 | 65372743 | 122 | 23 | 52168 | -0.57094 | 0.71598 | *PSMD12* |
| ENSG00000166220 | 10 | 72520995 | 72555157 | 206 | 35 | 51013 | -0.57118 | 0.71606 | *TBATA* |
| ENSG00000101323 | 20 | 7853628 | 7931121 | 224 | 38 | 34363 | -0.57132 | 0.71611 | *HAO1* |
| ENSG00000184221 | 21 | 34432450 | 34454726 | 41 | 17 | 46027 | -0.57138 | 0.71613 | *OLIG1* |
| ENSG00000269883 | 14 | 77597026 | 77619339 | 78 | 19 | 45603 | -0.57155 | 0.71619 | *AC007375.1* |
| ENSG00000165695 | 9 | 135590965 | 135764164 | 497 | 58 | 45900 | -0.57173 | 0.71625 | *AK8* |
| ENSG00000160321 | 19 | 22105760 | 22203751 | 424 | 28 | 46508 | -0.57199 | 0.71634 | *ZNF208* |
| ENSG00000123684 | 1 | 211906799 | 212014114 | 406 | 38 | 46199 | -0.5721 | 0.71637 | *LPGAT1* |
| ENSG00000142700 | 1 | 50873222 | 50899172 | 59 | 13 | 48643 | -0.57237 | 0.71646 | *DMRTA2* |
| ENSG00000175183 | 12 | 77242495 | 77282840 | 154 | 22 | 53070 | -0.57256 | 0.71653 | *CSRP2* |
| ENSG00000146416 | 6 | 143371633 | 143671441 | 583 | 89 | 38802 | -0.5726 | 0.71654 | *AIG1* |
| ENSG00000100225 | 22 | 32860663 | 32904818 | 189 | 19 | 45397 | -0.57266 | 0.71656 | *FBXO7* |
| ENSG00000180116 | 12 | 40009969 | 40312102 | 876 | 69 | 43686 | -0.57303 | 0.71669 | *C12orf40* |
| ENSG00000145217 | 4 | 962861 | 997228 | 130 | 29 | 50074 | -0.57318 | 0.71674 | *SLC26A1* |
| ENSG00000184809 | 21 | 40959074 | 40994748 | 109 | 24 | 52183 | -0.5732 | 0.71674 | *C21orf88* |
| ENSG00000175203 | 12 | 57913885 | 57951114 | 57 | 11 | 45900 | -0.57324 | 0.71676 | *DCTN2* |
| ENSG00000197562 | 16 | 629357 | 689272 | 225 | 16 | 54390 | -0.5733 | 0.71678 | *RAB40C* |
| ENSG00000128285 | 22 | 41064754 | 41088818 | 52 | 11 | 43627 | -0.57402 | 0.71702 | *MCHR1* |
| ENSG00000166448 | 7 | 98434111 | 98478394 | 100 | 20 | 44287 | -0.57405 | 0.71703 | *TMEM130* |
| ENSG00000118961 | 2 | 20873788 | 21032882 | 402 | 56 | 44456 | -0.57419 | 0.71708 | *C2orf43* |
| ENSG00000068079 | 17 | 41148742 | 41176473 | 47 | 9 | 48916 | -0.57447 | 0.71717 | *IFI35* |
| ENSG00000219481 | 1 | 16878814 | 16950057 | 19 | 9 | 42817 | -0.57458 | 0.71721 | *NBPF1* |
| ENSG00000139579 | 12 | 56605799 | 56633638 | 26 | 11 | 45433 | -0.57473 | 0.71726 | *NABP2* |
| ENSG00000164821 | 8 | 6783344 | 6805860 | 124 | 28 | 47460 | -0.5749 | 0.71732 | *DEFA4* |
| ENSG00000206127 | 15 | 32727307 | 32757835 | 3 | 2 | 42760 | -0.57518 | 0.71742 | *GOLGA8O* |
| ENSG00000106459 | 7 | 129241555 | 129406922 | 503 | 49 | 46637 | -0.57531 | 0.71746 | *NRF1* |
| ENSG00000174004 | 3 | 196356557 | 196398875 | 179 | 54 | 51871 | -0.57546 | 0.71751 | *NRROS* |
| ENSG00000104856 | 19 | 45494688 | 45551452 | 153 | 41 | 44378 | -0.57551 | 0.71753 | *RELB* |
| ENSG00000120885 | 8 | 27444434 | 27482548 | 103 | 25 | 38168 | -0.57554 | 0.71754 | *CLU* |

| | | | | | | | | | |
|---|---|---|---|---|---|---|---|---|---|
| ENSG00000197576 | 7 | 27158126 | 27180418 | 57 | 10 | 38836 | -0.57563 | 0.71757 | *HOXA4* |
| ENSG00000108602 | 17 | 19631297 | 19662256 | 69 | 15 | 36289 | -0.57573 | 0.7176 | *ALDH3A1* |
| ENSG00000101282 | 20 | 929095 | 992907 | 289 | 53 | 39900 | -0.57573 | 0.7176 | *RSPO4* |
| ENSG00000101161 | 20 | 62602488 | 62674453 | 173 | 17 | 50956 | -0.57594 | 0.71767 | *PRPF6* |
| ENSG00000188996 | 6 | 645939 | 666963 | 56 | 17 | 36716 | -0.57624 | 0.71778 | *HUS1B* |
| ENSG00000155189 | 8 | 6555878 | 6627184 | 286 | 36 | 44326 | -0.57667 | 0.71792 | *AGPAT5* |
| ENSG00000204856 | 12 | 110896169 | 110938190 | 117 | 15 | 33917 | -0.57705 | 0.71805 | *FAM216A* |
| ENSG00000198440 | 19 | 56899335 | 56957404 | 141 | 20 | 47842 | -0.57729 | 0.71813 | *ZNF583* |
| ENSG00000148803 | 10 | 135158658 | 135181529 | 95 | 9 | 25606 | -0.57773 | 0.71828 | *FUOM* |
| ENSG00000158246 | 1 | 27321511 | 27349327 | 27 | 10 | 44148 | -0.57821 | 0.71844 | *FAM46B* |
| ENSG00000132703 | 1 | 159547615 | 159568655 | 97 | 16 | 42194 | -0.57824 | 0.71845 | *APCS* |
| ENSG00000133818 | 11 | 14289472 | 14396052 | 156 | 26 | 55070 | -0.57852 | 0.71854 | *RRAS2* |
| ENSG00000121577 | 3 | 119345304 | 119394171 | 213 | 24 | 52292 | -0.57867 | 0.71859 | *POPDC2* |
| ENSG00000005801 | 11 | 3350491 | 3410448 | 192 | 18 | 47040 | -0.57875 | 0.71862 | *ZNF195* |
| ENSG00000141655 | 18 | 59982520 | 60068516 | 371 | 66 | 46091 | -0.57875 | 0.71862 | *TNFRSF11A* |
| ENSG00000114988 | 2 | 97361666 | 97415801 | 78 | 10 | 42750 | -0.57881 | 0.71864 | *LMAN2L* |
| ENSG00000135842 | 1 | 184749858 | 184953682 | 481 | 57 | 44029 | -0.57896 | 0.71869 | *FAM129A* |
| ENSG00000143748 | 1 | 224405036 | 224528089 | 281 | 32 | 42241 | -0.57895 | 0.71869 | *NVL* |
| ENSG00000173369 | 1 | 22969255 | 22998031 | 106 | 16 | 34270 | -0.57918 | 0.71876 | *C1QB* |
| ENSG00000160445 | 9 | 131482065 | 131544693 | 82 | 20 | 39597 | -0.57972 | 0.71895 | *ZER1* |
| ENSG00000182170 | 11 | 3229174 | 3250043 | 73 | 24 | 48013 | -0.5798 | 0.71898 | *MRGPRG* |
| ENSG00000205426 | 12 | 52669697 | 52695318 | 114 | 15 | 55205 | -0.57983 | 0.71899 | *KRT81* |
| ENSG00000126467 | 19 | 50233010 | 50276587 | 106 | 34 | 45050 | -0.5799 | 0.71901 | *TSKS* |
| ENSG00000158220 | 3 | 138143428 | 138210528 | 115 | 24 | 48777 | -0.57995 | 0.71903 | *ESYT3* |
| ENSG00000257599 | 12 | 29532227 | 29650421 | 494 | 47 | 44312 | -0.58002 | 0.71905 | *OVCH1-AS1* |
| ENSG00000203392 | 15 | 75960672 | 75983093 | 53 | 10 | 40850 | -0.5801 | 0.71908 | *AC105020.1* |
| ENSG00000100099 | 22 | 26829389 | 26889803 | 213 | 28 | 41821 | -0.58038 | 0.71917 | *HPS4* |
| ENSG00000141252 | 17 | 401908 | 634957 | 863 | 77 | 45848 | -0.58049 | 0.71921 | *VPS53* |
| ENSG00000112164 | 6 | 39006574 | 39065519 | 239 | 39 | 43780 | -0.58067 | 0.71927 | *GLP1R* |
| ENSG00000186930 | 21 | 31960909 | 31981219 | 43 | 8 | 55217 | -0.58088 | 0.71934 | *KRTAP6-2* |
| ENSG00000099977 | 22 | 24303554 | 24332660 | 19 | 8 | 41793 | -0.58103 | 0.71939 | *DDT* |
| ENSG00000114124 | 3 | 141486990 | 141547332 | 181 | 28 | 49930 | -0.58106 | 0.7194 | *GRK7* |
| ENSG00000048028 | 11 | 113658596 | 113756292 | 246 | 27 | 49738 | -0.58106 | 0.7194 | *USP28* |
| ENSG00000166337 | 11 | 6617526 | 6643898 | 74 | 15 | 48077 | -0.58129 | 0.71948 | *TAF10* |
| ENSG00000180425 | 11 | 114252165 | 114281139 | 66 | 13 | 47525 | -0.58139 | 0.71951 | *C11orf71* |
| ENSG00000154059 | 18 | 21996580 | 22043499 | 175 | 23 | 51581 | -0.58164 | 0.7196 | *IMPACT* |

| | | | | | | | | | |
|---|---|---|---|---|---|---|---|---|---|
| ENSG00000165023 | 9 | 93362114 | 93415386 | 220 | 41 | 52464 | -0.58174 | 0.71963 | *DIRAS2* |
| ENSG00000100167 | 22 | 42362276 | 42404225 | 109 | 14 | 47618 | -0.58174 | 0.71963 | *Sep-03* |
| ENSG00000145736 | 5 | 70320784 | 70373516 | 7 | 4 | 43375 | -0.58214 | 0.71977 | *GTF2H2* |
| ENSG00000107771 | 10 | 86078342 | 86288273 | 695 | 40 | 49047 | -0.58216 | 0.71977 | *CCSER2* |
| ENSG00000169347 | 16 | 20310894 | 20349130 | 111 | 22 | 31534 | -0.58228 | 0.71981 | *GP2* |
| ENSG00000186910 | 14 | 94898801 | 94929127 | 153 | 18 | 45069 | -0.58253 | 0.71989 | *SERPINA11* |
| ENSG00000272896 | 9 | 139675831 | 139705047 | 111 | 12 | 53235 | -0.5826 | 0.71992 | *RP11-216L13.17* |
| ENSG00000111843 | 6 | 10713148 | 10741362 | 51 | 9 | 39427 | -0.58381 | 0.72033 | *TMEM14C* |
| ENSG00000108242 | 10 | 96433251 | 96505947 | 207 | 20 | 49488 | -0.58405 | 0.72041 | *CYP2C18* |
| ENSG00000169359 | 3 | 155528813 | 155582218 | 108 | 20 | 42797 | -0.58445 | 0.72054 | *SLC33A1* |
| ENSG00000090905 | 16 | 24731016 | 24848953 | 271 | 36 | 51820 | -0.58474 | 0.72064 | *TNRC6A* |
| ENSG00000139200 | 12 | 6792958 | 6819966 | 43 | 17 | 53182 | -0.58484 | 0.72067 | *PIANP* |
| ENSG00000269026 | 19 | 58183413 | 58274943 | 225 | 32 | 45661 | -0.58499 | 0.72072 | *AC003006.7* |
| ENSG00000255181 | 8 | 144778864 | 144800279 | 55 | 15 | 33088 | -0.58516 | 0.72078 | *CCDC166* |
| ENSG00000144820 | 3 | 100318433 | 100424323 | 240 | 24 | 37188 | -0.58563 | 0.72094 | *GPR128* |
| ENSG00000125522 | 20 | 62727173 | 62748524 | 82 | 14 | 53660 | -0.58564 | 0.72094 | *NPBWR2* |
| ENSG00000010539 | 16 | 3262325 | 3296221 | 111 | 13 | 48229 | -0.58607 | 0.72109 | *ZNF200* |
| ENSG00000168306 | 3 | 58480863 | 58533046 | 116 | 28 | 40338 | -0.58616 | 0.72111 | *ACOX2* |
| ENSG00000165480 | 13 | 21717734 | 21760741 | 161 | 22 | 47736 | -0.58638 | 0.72119 | *SKA3* |
| ENSG00000101442 | 20 | 37367085 | 37410834 | 235 | 20 | 54788 | -0.5867 | 0.7213 | *ACTR5* |
| ENSG00000133246 | 19 | 8544940 | 8577996 | 108 | 17 | 35446 | -0.58692 | 0.72137 | *PRAM1* |
| ENSG00000188672 | 1 | 25678740 | 25766683 | 133 | 12 | 27518 | -0.58719 | 0.72146 | *RHCE* |
| ENSG00000119760 | 2 | 27863679 | 27896676 | 34 | 10 | 34003 | -0.58727 | 0.72149 | *SUPT7L* |
| ENSG00000155016 | 4 | 108842525 | 108884613 | 151 | 12 | 51474 | -0.58726 | 0.72149 | *CYP2U1* |
| ENSG00000156735 | 8 | 38024051 | 38080819 | 103 | 9 | 51617 | -0.58735 | 0.72152 | *BAG4* |
| ENSG00000168395 | 2 | 242631450 | 242678893 | 174 | 24 | 39506 | -0.58757 | 0.72159 | *ING5* |
| ENSG00000119888 | 2 | 47562297 | 47624740 | 225 | 46 | 41942 | -0.58766 | 0.72162 | *EPCAM* |
| ENSG00000088888 | 20 | 3817487 | 3859280 | 109 | 35 | 40271 | -0.58782 | 0.72167 | *MAVS* |
| ENSG00000133636 | 12 | 86258073 | 86286770 | 117 | 16 | 47318 | -0.58792 | 0.72171 | *NTS* |
| ENSG00000213265 | 7 | 130343488 | 130382268 | 101 | 19 | 47254 | -0.58798 | 0.72173 | *TSGA13* |
| ENSG00000169155 | 9 | 129557285 | 129610489 | 59 | 17 | 49044 | -0.58833 | 0.72185 | *ZBTB43* |
| ENSG00000133961 | 14 | 73731815 | 73940348 | 506 | 46 | 37977 | -0.58837 | 0.72186 | *NUMB* |
| ENSG00000179532 | 11 | 6508490 | 6624988 | 315 | 45 | 46872 | -0.58845 | 0.72189 | *DNHD1* |
| ENSG00000139625 | 12 | 53864274 | 53903847 | 43 | 17 | 51004 | -0.58861 | 0.72194 | *MAP3K12* |
| ENSG00000159189 | 1 | 22960123 | 22984603 | 110 | 15 | 32124 | -0.5891 | 0.7221 | *C1QC* |
| ENSG00000105793 | 7 | 89954537 | 90030769 | 297 | 32 | 50149 | -0.58918 | 0.72213 | *GTPBP10* |

| | | | | | | | | | |
|---|---|---|---|---|---|---|---|---|---|
| ENSG00000162434 | 1 | 65288912 | 65442187 | 454 | 47 | 35630 | -0.58948 | 0.72223 | *JAK1* |
| ENSG00000063127 | 19 | 49782895 | 49838482 | 97 | 16 | 49715 | -0.58983 | 0.72235 | *SLC6A16* |
| ENSG00000090316 | 4 | 1273639 | 1343935 | 267 | 25 | 36123 | -0.59016 | 0.72246 | *MAEA* |
| ENSG00000213145 | 14 | 105942654 | 105965284 | 18 | 10 | 34888 | -0.59016 | 0.72246 | *CRIP1* |
| ENSG00000139618 | 13 | 32879611 | 32983805 | 270 | 32 | 48382 | -0.59044 | 0.72255 | *BRCA2* |
| ENSG00000244094 | 1 | 153074590 | 153095991 | 47 | 13 | 38832 | -0.59053 | 0.72258 | *SPRR2F* |
| ENSG00000133401 | 5 | 31629517 | 32121037 | 1903 | 289 | 48634 | -0.59052 | 0.72258 | *PDZD2* |
| ENSG00000166342 | 18 | 70399549 | 70545381 | 560 | 62 | 47029 | -0.59075 | 0.72266 | *NETO1* |
| ENSG00000111879 | 6 | 119270928 | 119480552 | 742 | 87 | 44689 | -0.59087 | 0.7227 | *FAM184A* |
| ENSG00000173517 | 15 | 77390471 | 77722486 | 576 | 41 | 45467 | -0.59088 | 0.7227 | *PEAK1* |
| ENSG00000064225 | 3 | 98441080 | 98550045 | 315 | 33 | 51526 | -0.59109 | 0.72277 | *ST3GAL6* |
| ENSG00000166557 | 15 | 79593404 | 79714334 | 477 | 46 | 49963 | -0.59116 | 0.72279 | *TMED3* |
| ENSG00000169330 | 15 | 79714858 | 79774632 | 220 | 35 | 46738 | -0.59115 | 0.72279 | *KIAA1024* |
| ENSG00000132793 | 20 | 39959560 | 39999222 | 106 | 23 | 43694 | -0.59128 | 0.72284 | *LPIN3* |
| ENSG00000145700 | 5 | 74354100 | 74542703 | 579 | 30 | 29379 | -0.59167 | 0.72296 | *ANKRD31* |
| ENSG00000177340 | 12 | 31467250 | 31488879 | 13 | 6 | 35486 | -0.59167 | 0.72296 | *AC024940.1* |
| ENSG00000135919 | 2 | 224829829 | 224914036 | 279 | 42 | 38844 | -0.59189 | 0.72304 | *SERPINE2* |
| ENSG00000115085 | 2 | 98320023 | 98366325 | 97 | 19 | 37926 | -0.59209 | 0.72311 | *ZAP70* |
| ENSG00000175879 | 2 | 176984422 | 177007423 | 51 | 12 | 41213 | -0.59219 | 0.72314 | *HOXD8* |
| ENSG00000106049 | 7 | 27555061 | 27712614 | 640 | 35 | 49543 | -0.59225 | 0.72316 | *HIBADH* |
| ENSG00000197958 | 9 | 130199953 | 130223684 | 76 | 7 | 54929 | -0.59267 | 0.7233 | *RPL12* |
| ENSG00000006451 | 7 | 39653082 | 39757723 | 248 | 37 | 41794 | -0.59272 | 0.72331 | *RALA* |
| ENSG00000121318 | 12 | 10967916 | 10988957 | 38 | 9 | 35352 | -0.59281 | 0.72334 | *TAS2R10* |
| ENSG00000160180 | 21 | 43721777 | 43745761 | 92 | 22 | 41532 | -0.59287 | 0.72337 | *TFF3* |
| ENSG00000198858 | 19 | 886503 | 923240 | 104 | 37 | 43689 | -0.59316 | 0.72346 | *R3HDM4* |
| ENSG00000176542 | 3 | 113357232 | 113425493 | 93 | 11 | 48591 | -0.59334 | 0.72352 | *KIAA2018* |
| ENSG00000100122 | 22 | 26985242 | 27024052 | 130 | 21 | 46499 | -0.59339 | 0.72354 | *CRYBB1* |
| ENSG00000070778 | 14 | 88922122 | 89031077 | 269 | 23 | 41865 | -0.59387 | 0.7237 | *PTPN21* |
| ENSG00000197683 | 21 | 31681452 | 31702607 | 159 | 15 | 34970 | -0.59403 | 0.72376 | *KRTAP26-1* |
| ENSG00000215902 | 1 | 28517013 | 28537152 | 60 | 9 | 46499 | -0.5942 | 0.72381 | *AL353354.1* |
| ENSG00000168078 | 8 | 27657137 | 27705612 | 247 | 9 | 27541 | -0.59431 | 0.72385 | *PBK* |
| ENSG00000118482 | 6 | 64335725 | 64499229 | 307 | 32 | 48758 | -0.59446 | 0.7239 | *PHF3* |
| ENSG00000168672 | 8 | 127554687 | 127580638 | 48 | 14 | 42796 | -0.59475 | 0.72399 | *FAM84B* |
| ENSG00000204688 | 6 | 29414958 | 29442105 | 76 | 14 | 30643 | -0.59501 | 0.72408 | *OR2H1* |
| ENSG00000141294 | 17 | 45898993 | 45925079 | 97 | 17 | 33397 | -0.59516 | 0.72413 | *LRRC46* |
| ENSG00000264187 | 17 | 17095963 | 17150499 | 184 | 27 | 48672 | -0.59555 | 0.72426 | *RP11-45M22.4* |

| | | | | | | | | | |
|---|---|---|---|---|---|---|---|---|---|
| ENSG00000134371 | 1 | 193081147 | 193233031 | 411 | 28 | 54060 | -0.59559 | 0.72428 | *CDC73* |
| ENSG00000149609 | 20 | 32240090 | 32263425 | 22 | 8 | 51342 | -0.59561 | 0.72428 | *C20orf144* |
| ENSG00000109775 | 4 | 186310694 | 186357139 | 138 | 19 | 32058 | -0.59616 | 0.72447 | *UFSP2* |
| ENSG00000119138 | 9 | 72989503 | 73039540 | 107 | 25 | 42477 | -0.59706 | 0.72477 | *KLF9* |
| ENSG00000269469 | 19 | 49956009 | 49999488 | 142 | 23 | 31862 | -0.59708 | 0.72477 | *CTD-3148I10.9* |
| ENSG00000109762 | 4 | 186115391 | 186301339 | 676 | 58 | 47174 | -0.59719 | 0.72481 | *SNX25* |
| ENSG00000122477 | 1 | 100604409 | 100653771 | 117 | 23 | 34743 | -0.59727 | 0.72484 | *LRRC39* |
| ENSG00000168758 | 2 | 97515453 | 97546494 | 35 | 13 | 38350 | -0.59737 | 0.72487 | *SEMA4C* |
| ENSG00000189227 | 15 | 67803406 | 67829628 | 75 | 8 | 41772 | -0.59741 | 0.72488 | *C15orf61* |
| ENSG00000166523 | 12 | 8675901 | 8703559 | 117 | 18 | 48185 | -0.59778 | 0.72501 | *CLEC4E* |
| ENSG00000222036 | 14 | 19543365 | 19600078 | 2 | 1 | 42914 | -0.59791 | 0.72505 | *POTEG* |
| ENSG00000262304 | 17 | 3458738 | 3549543 | 382 | 61 | 38154 | -0.59799 | 0.72508 | *SHPK* |
| ENSG00000130643 | 10 | 135128927 | 135160475 | 87 | 20 | 41060 | -0.59813 | 0.72512 | *CALY* |
| ENSG00000010282 | 3 | 42724155 | 42754319 | 65 | 19 | 49460 | -0.59849 | 0.72525 | *HHATL* |
| ENSG00000083307 | 8 | 102494660 | 102691954 | 613 | 70 | 38161 | -0.59882 | 0.72535 | *GRHL2* |
| ENSG00000100216 | 22 | 39067953 | 39090818 | 71 | 8 | 47387 | -0.59897 | 0.7254 | *TOMM22* |
| ENSG00000170891 | 4 | 5006313 | 5031199 | 129 | 26 | 47968 | -0.59907 | 0.72544 | *CYTL1* |
| ENSG00000182107 | 14 | 61734088 | 61758558 | 66 | 17 | 34393 | -0.59907 | 0.72544 | *TMEM30B* |
| ENSG00000100852 | 14 | 32535320 | 32638934 | 103 | 37 | 50368 | -0.59911 | 0.72545 | *ARHGAP5* |
| ENSG00000179104 | 12 | 83070659 | 83538649 | 1419 | 140 | 43168 | -0.5992 | 0.72548 | *TMTC2* |
| ENSG00000188916 | 10 | 128923694 | 129004422 | 222 | 28 | 37763 | -0.59924 | 0.72549 | *FAM196A* |
| ENSG00000235978 | 3 | 4845978 | 4938977 | 303 | 43 | 41498 | -0.59934 | 0.72553 | *AC018816.3* |
| ENSG00000268533 | 19 | 57895551 | 57959825 | 147 | 26 | 47724 | -0.59989 | 0.72571 | *AC004076.7* |
| ENSG00000189001 | 19 | 36004269 | 36029253 | 95 | 20 | 46590 | -0.60005 | 0.72576 | *SBSN* |
| ENSG00000198673 | 12 | 62092040 | 62682931 | 1914 | 169 | 50715 | -0.60022 | 0.72582 | *FAM19A2* |
| ENSG00000081051 | 4 | 74286855 | 74331891 | 113 | 19 | 51514 | -0.60025 | 0.72583 | *AFP* |
| ENSG00000144909 | 3 | 125237702 | 125323934 | 354 | 30 | 45526 | -0.60029 | 0.72584 | *OSBPL11* |
| ENSG00000003987 | 8 | 17145539 | 17281037 | 646 | 83 | 36604 | -0.60064 | 0.72596 | *MTMR7* |
| ENSG00000067141 | 15 | 73334051 | 73607547 | 725 | 53 | 38808 | -0.60114 | 0.72613 | *NEO1* |
| ENSG00000169255 | 3 | 160791671 | 160833172 | 91 | 13 | 53447 | -0.60153 | 0.72626 | *B3GALNT1* |
| ENSG00000188785 | 19 | 57891218 | 57922786 | 76 | 15 | 46529 | -0.60177 | 0.72633 | *ZNF548* |
| ENSG00000253506 | 17 | 59657794 | 59678563 | 63 | 9 | 47353 | -0.60177 | 0.72634 | *NACA2* |
| ENSG00000146166 | 6 | 63975856 | 64039882 | 98 | 18 | 52334 | -0.60188 | 0.72637 | *LGSN* |
| ENSG00000171109 | 3 | 179055480 | 179122719 | 144 | 20 | 50945 | -0.60235 | 0.72653 | *MFN1* |
| ENSG00000145592 | 5 | 40815364 | 40845437 | 83 | 18 | 51848 | -0.60236 | 0.72653 | *RPL37* |
| ENSG00000151413 | 14 | 31949162 | 32340430 | 1403 | 59 | 44659 | -0.60257 | 0.7266 | *NUBPL* |

| | | | | | | | | | |
|---|---|---|---|---|---|---|---|---|---|
| ENSG00000136104 | 13 | 51473814 | 51554592 | 170 | 29 | 48247 | -0.60271 | 0.72665 | *RNASEH2B* |
| ENSG00000071127 | 4 | 10065963 | 10128573 | 325 | 30 | 46667 | -0.60292 | 0.72672 | *WDR1* |
| ENSG00000117748 | 1 | 28208035 | 28251257 | 141 | 12 | 38224 | -0.60321 | 0.72681 | *RPA2* |
| ENSG00000180509 | 21 | 35808988 | 35894573 | 252 | 46 | 46005 | -0.60333 | 0.72685 | *KCNE1* |
| ENSG00000130202 | 19 | 45339432 | 45402485 | 290 | 52 | 50293 | -0.60345 | 0.7269 | *PVRL2* |
| ENSG00000205497 | 11 | 4957355 | 4978356 | 127 | 13 | 51584 | -0.60386 | 0.72703 | *OR51A4* |
| ENSG00000079999 | 19 | 10586796 | 10624417 | 121 | 20 | 47576 | -0.60387 | 0.72704 | *KEAP1* |
| ENSG00000183814 | 1 | 226408850 | 226507570 | 181 | 24 | 53186 | -0.60403 | 0.72709 | *LIN9* |
| ENSG00000138449 | 2 | 190415305 | 190458484 | 109 | 21 | 38677 | -0.60411 | 0.72712 | *SLC40A1* |
| ENSG00000184698 | 11 | 5400607 | 5421664 | 204 | 18 | 52433 | -0.60436 | 0.7272 | *OR51M1* |
| ENSG00000053747 | 18 | 21259407 | 21545030 | 442 | 43 | 39519 | -0.6047 | 0.72731 | *LAMA3* |
| ENSG00000150556 | 2 | 149884621 | 150081776 | 675 | 64 | 44829 | -0.60487 | 0.72737 | *LYPD6B* |
| ENSG00000134548 | 12 | 21669241 | 21700311 | 127 | 22 | 43875 | -0.60545 | 0.72756 | *C12orf39* |
| ENSG00000197506 | 9 | 86880372 | 86965672 | 359 | 53 | 38710 | -0.60548 | 0.72757 | *SLC28A3* |
| ENSG00000205923 | 16 | 2567081 | 2591420 | 23 | 16 | 53077 | -0.60553 | 0.72759 | *CEMP1* |
| ENSG00000174749 | 4 | 113056553 | 113126412 | 157 | 20 | 44754 | -0.60592 | 0.72771 | *C4orf32* |
| ENSG00000134762 | 18 | 28559974 | 28632781 | 223 | 30 | 40940 | -0.60639 | 0.72787 | *DSC3* |
| ENSG00000101126 | 20 | 49495585 | 49557958 | 159 | 21 | 38553 | -0.60648 | 0.7279 | *ADNP* |
| ENSG00000240432 | 21 | 31787583 | 31808286 | 57 | 7 | 55586 | -0.6065 | 0.72791 | *KRTAP13-3* |
| ENSG00000187840 | 8 | 37877859 | 37927883 | 69 | 16 | 45843 | -0.60678 | 0.728 | *EIF4EBP1* |
| ENSG00000197408 | 19 | 41487204 | 41534303 | 206 | 24 | 34843 | -0.60706 | 0.72809 | *CYP2B6* |
| ENSG00000197181 | 8 | 22122810 | 22225076 | 283 | 23 | 42744 | -0.60723 | 0.72815 | *PIWIL2* |
| ENSG00000166128 | 15 | 63471668 | 63569981 | 210 | 35 | 41212 | -0.60723 | 0.72815 | *RAB8B* |
| ENSG00000139631 | 12 | 53541447 | 53585135 | 80 | 11 | 43301 | -0.6073 | 0.72817 | *CSAD* |
| ENSG00000269804 | 1 | 28517070 | 28537419 | 61 | 9 | 46682 | -0.60749 | 0.72824 | *AL353354.2* |
| ENSG00000145990 | 6 | 13348062 | 13497894 | 345 | 59 | 37758 | -0.60748 | 0.72824 | *GFOD1* |
| ENSG00000134198 | 1 | 115580632 | 115642121 | 270 | 35 | 50793 | -0.60764 | 0.72829 | *TSPAN2* |
| ENSG00000077454 | 7 | 100159855 | 100193776 | 58 | 12 | 52026 | -0.60799 | 0.7284 | *LRCH4* |
| ENSG00000006128 | 7 | 97351220 | 97379784 | 106 | 12 | 54924 | -0.608 | 0.72841 | *TAC1* |
| ENSG00000196123 | 16 | 67199505 | 67227943 | 34 | 9 | 36158 | -0.60873 | 0.72865 | *KIAA0895L* |
| ENSG00000060138 | 12 | 10841683 | 10885911 | 133 | 18 | 47063 | -0.60879 | 0.72867 | *YBX3* |
| ENSG00000213231 | 14 | 96142754 | 96168980 | 109 | 31 | 38481 | -0.60911 | 0.72878 | *TCL1B* |
| ENSG00000168491 | 4 | 186356336 | 186402913 | 175 | 35 | 33050 | -0.60934 | 0.72885 | *CCDC110* |
| ENSG00000205279 | 5 | 126974736 | 127004322 | 107 | 20 | 41972 | -0.60955 | 0.72892 | *CTXN3* |
| ENSG00000159131 | 21 | 34866238 | 34925797 | 110 | 18 | 53201 | -0.60966 | 0.72896 | *GART* |
| ENSG00000010379 | 12 | 319789 | 382039 | 205 | 40 | 41557 | -0.60971 | 0.72897 | *SLC6A13* |

| | | | | | | | | | |
|---|---|---|---|---|---|---|---|---|---|
| ENSG00000139223 | 12 | 48856448 | 48876843 | 50 | 10 | 34730 | -0.61001 | 0.72907 | *ANP32D* |
| ENSG00000215440 | 20 | 57254187 | 57304294 | 200 | 25 | 41497 | -0.61036 | 0.72919 | *NPEPL1* |
| ENSG00000238243 | 1 | 248021277 | 248070449 | 280 | 46 | 44107 | -0.6108 | 0.72934 | *OR2W3* |
| ENSG00000167080 | 17 | 47199822 | 47257351 | 237 | 31 | 49257 | -0.61092 | 0.72937 | *B4GALNT2* |
| ENSG00000206102 | 21 | 32400478 | 32420795 | 21 | 12 | 46714 | -0.61107 | 0.72942 | *KRTAP19-8* |
| ENSG00000205592 | 12 | 40777197 | 40974632 | 1136 | 61 | 44150 | -0.61114 | 0.72945 | *MUC19* |
| ENSG00000130035 | 12 | 4819507 | 4970277 | 551 | 64 | 45213 | -0.61158 | 0.72959 | *GALNT8* |
| ENSG00000146054 | 5 | 180610924 | 180642293 | 160 | 28 | 33541 | -0.61244 | 0.72988 | *TRIM7* |
| ENSG00000110074 | 11 | 126128950 | 126158026 | 82 | 16 | 41787 | -0.61247 | 0.72989 | *FOXRED1* |
| ENSG00000134042 | 18 | 48314574 | 48361772 | 199 | 40 | 47229 | -0.61254 | 0.72991 | *MRO* |
| ENSG00000186806 | 19 | 51824790 | 51855378 | 100 | 20 | 39984 | -0.61289 | 0.73002 | *VSIG10L* |
| ENSG00000261052 | 16 | 30195754 | 30225631 | 5 | 3 | 27239 | -0.61295 | 0.73005 | *SULT1A3* |
| ENSG00000205899 | 17 | 1163853 | 1184754 | 75 | 24 | 50588 | -0.61298 | 0.73005 | *BHLHA9* |
| ENSG00000166343 | 10 | 75173339 | 75203319 | 21 | 7 | 32342 | -0.61358 | 0.73025 | *MSS51* |
| ENSG00000084652 | 1 | 32635287 | 32673886 | 63 | 14 | 36521 | -0.61363 | 0.73027 | *TXLNA* |
| ENSG00000269647 | 1 | 147621967 | 147642428 | 1 | 1 | 12474 | -0.61372 | 0.7303 | *BX842679.1* |
| ENSG00000118242 | 2 | 216799213 | 216908819 | 433 | 87 | 43618 | -0.61387 | 0.73035 | *MREG* |
| ENSG00000083454 | 17 | 3565493 | 3609698 | 138 | 23 | 45720 | -0.61418 | 0.73045 | *P2RX5* |
| ENSG00000168702 | 2 | 140978992 | 142899270 | 8843 | 489 | 46949 | -0.61423 | 0.73047 | *LRP1B* |
| ENSG00000123349 | 12 | 53679075 | 53703234 | 41 | 11 | 42994 | -0.61445 | 0.73054 | *PFDN5* |
| ENSG00000126460 | 19 | 50073903 | 50104272 | 77 | 10 | 34996 | -0.6145 | 0.73056 | *PRRG2* |
| ENSG00000162620 | 1 | 74481699 | 74673871 | 389 | 28 | 36858 | -0.61526 | 0.73081 | *LRRIQ3* |
| ENSG00000106178 | 7 | 75430983 | 75462674 | 216 | 28 | 45118 | -0.61546 | 0.73087 | *CCL24* |
| ENSG00000116761 | 1 | 70866901 | 70915534 | 139 | 30 | 46227 | -0.6155 | 0.73089 | *CTH* |
| ENSG00000120925 | 8 | 42694780 | 42762433 | 114 | 20 | 48345 | -0.61553 | 0.7309 | *RNF170* |
| ENSG00000149150 | 11 | 57242007 | 57293259 | 91 | 18 | 50464 | -0.61574 | 0.73097 | *SLC43A1* |
| ENSG00000168904 | 15 | 99781567 | 99940934 | 567 | 52 | 41072 | -0.61614 | 0.7311 | *LRRC28* |
| ENSG00000141985 | 19 | 4350367 | 4410544 | 170 | 26 | 50245 | -0.61617 | 0.73111 | *SH3GL1* |
| ENSG00000170873 | 8 | 125553031 | 125750730 | 765 | 124 | 45709 | -0.61625 | 0.73113 | *MTSS1* |
| ENSG00000156413 | 19 | 5820621 | 5849742 | 116 | 22 | 37182 | -0.61663 | 0.73126 | *FUT6* |
| ENSG00000148848 | 10 | 127690950 | 128087024 | 1556 | 195 | 46754 | -0.61681 | 0.73132 | *ADAM12* |
| ENSG00000006047 | 17 | 7181571 | 7207934 | 64 | 17 | 45801 | -0.61688 | 0.73134 | *YBX2* |
| ENSG00000171916 | 17 | 18370051 | 18408259 | 4 | 2 | 45963 | -0.61709 | 0.73141 | *LGALS9C* |
| ENSG00000104728 | 8 | 1762142 | 1916807 | 884 | 156 | 49830 | -0.61714 | 0.73143 | *ARHGEF10* |
| ENSG00000213699 | 2 | 26977152 | 27014099 | 55 | 9 | 41044 | -0.61726 | 0.73147 | *SLC35F6* |
| ENSG00000204377 | 1 | 16545368 | 16566038 | 49 | 8 | 44827 | -0.61749 | 0.73154 | *C1orf134* |

| | | | | | | | | | |
|---|---|---|---|---|---|---|---|---|---|
| ENSG00000255713 | 17 | 56237017 | 56257940 | 74 | 15 | 42220 | -0.61747 | 0.73154 | *OR4D2* |
| ENSG00000214732 | 6 | 42113279 | 42141156 | 131 | 27 | 46804 | -0.61817 | 0.73177 | *RP1-139D8.6* |
| ENSG00000269113 | 1 | 48216200 | 48472567 | 749 | 78 | 49712 | -0.61826 | 0.7318 | *TRABD2B* |
| ENSG00000141576 | 17 | 74128534 | 74246454 | 291 | 41 | 42676 | -0.61829 | 0.73181 | *RNF157* |
| ENSG00000174194 | 10 | 51214681 | 51381321 | 6 | 3 | 32679 | -0.61843 | 0.73185 | *AGAP8* |
| ENSG00000125538 | 2 | 113577328 | 113604480 | 60 | 16 | 44474 | -0.61896 | 0.73203 | *IL1B* |
| ENSG00000146453 | 6 | 160211298 | 160251736 | 164 | 20 | 50397 | -0.61928 | 0.73213 | *PNLDC1* |
| ENSG00000182264 | 19 | 49234109 | 49260166 | 75 | 10 | 25351 | -0.61958 | 0.73223 | *IZUMO1* |
| ENSG00000222020 | 2 | 240313130 | 240334058 | 59 | 23 | 44376 | -0.61983 | 0.73231 | *AC062017.1* |
| ENSG00000150316 | 11 | 94685787 | 94716776 | 92 | 16 | 44780 | -0.62017 | 0.73243 | *CWC15* |
| ENSG00000267206 | 9 | 139628463 | 139652980 | 106 | 13 | 43908 | -0.62026 | 0.73246 | *LCN6* |
| ENSG00000206418 | 18 | 8599443 | 8649379 | 181 | 19 | 39032 | -0.62063 | 0.73258 | *RAB12* |
| ENSG00000182983 | 3 | 42937223 | 42970825 | 52 | 13 | 44959 | -0.62067 | 0.73259 | *ZNF662* |
| ENSG00000140859 | 16 | 57782129 | 57906957 | 447 | 68 | 46427 | -0.62076 | 0.73262 | *KIFC3* |
| ENSG00000168286 | 16 | 67866213 | 67888097 | 35 | 9 | 44482 | -0.62082 | 0.73264 | *THAP11* |
| ENSG00000131507 | 5 | 141478070 | 141544005 | 281 | 25 | 47888 | -0.6209 | 0.73267 | *NDFIP1* |
| ENSG00000101132 | 20 | 52814386 | 52854591 | 156 | 31 | 46070 | -0.62095 | 0.73268 | *PFDN4* |
| ENSG00000184508 | 15 | 91464148 | 91485799 | 68 | 13 | 40661 | -0.62101 | 0.7327 | *HDDC3* |
| ENSG00000105053 | 19 | 50469724 | 50539203 | 255 | 19 | 44491 | -0.6214 | 0.73283 | *VRK3* |
| ENSG00000141577 | 17 | 79153393 | 79206799 | 236 | 37 | 33397 | -0.62192 | 0.733 | *AZI1* |
| ENSG00000261594 | 11 | 74941950 | 74964742 | 41 | 10 | 36107 | -0.62206 | 0.73305 | *TPBGL* |
| ENSG00000184029 | 21 | 39313728 | 39503454 | 509 | 81 | 44378 | -0.6225 | 0.73319 | *DSCR4* |
| ENSG00000171163 | 1 | 249134205 | 249163343 | 36 | 9 | 34592 | -0.62252 | 0.7332 | *ZNF692* |
| ENSG00000145431 | 4 | 157671606 | 157902546 | 411 | 55 | 41920 | -0.62265 | 0.73324 | *PDGFC* |
| ENSG00000168454 | 18 | 9875763 | 9899272 | 96 | 21 | 52836 | -0.62267 | 0.73325 | *TXNDC2* |
| ENSG00000109158 | 4 | 46910917 | 47006424 | 357 | 34 | 54820 | -0.62278 | 0.73329 | *GABRA4* |
| ENSG00000259060 | 14 | 21041051 | 21087954 | 232 | 52 | 49256 | -0.62286 | 0.73331 | *RNASE11* |
| ENSG00000106809 | 9 | 95136249 | 95176978 | 100 | 13 | 45093 | -0.62326 | 0.73344 | *OGN* |
| ENSG00000122870 | 10 | 60262900 | 60601195 | 850 | 43 | 51073 | -0.62331 | 0.73346 | *BICC1* |
| ENSG00000182934 | 11 | 126122814 | 126149039 | 58 | 13 | 41963 | -0.62346 | 0.73351 | *SRPR* |
| ENSG00000146802 | 7 | 112392437 | 112440647 | 85 | 11 | 38275 | -0.62356 | 0.73354 | *TMEM168* |
| ENSG00000214402 | 9 | 139866356 | 139890862 | 67 | 11 | 53106 | -0.6247 | 0.73392 | *LCNL1* |
| ENSG00000257046 | 12 | 20958608 | 21253179 | 1214 | 49 | 32035 | -0.62484 | 0.73396 | *LST3* |
| ENSG00000233608 | 2 | 239746673 | 239805893 | 122 | 36 | 41453 | -0.62527 | 0.7341 | *TWIST2* |
| ENSG00000154188 | 8 | 108251721 | 108520283 | 957 | 110 | 49201 | -0.62559 | 0.73421 | *ANGPT1* |
| ENSG00000142677 | 1 | 24436261 | 24479611 | 149 | 28 | 50190 | -0.62578 | 0.73427 | *IL22RA1* |

| | | | | | | | | | |
|---|---|---|---|---|---|---|---|---|---|
| ENSG00000083814 | 19 | 58221119 | 58248992 | 76 | 13 | 45664 | -0.62647 | 0.7345 | *ZNF671* |
| ENSG00000118454 | 1 | 70716271 | 70830417 | 217 | 31 | 52356 | -0.62655 | 0.73452 | *ANKRD13C* |
| ENSG00000178028 | 1 | 44669127 | 44696353 | 38 | 13 | 50042 | -0.62695 | 0.73465 | *DMAP1* |
| ENSG00000119283 | 1 | 231287858 | 231367302 | 271 | 49 | 40797 | -0.62704 | 0.73468 | *TRIM67* |
| ENSG00000049130 | 12 | 88876570 | 88984628 | 223 | 16 | 41702 | -0.62723 | 0.73474 | *KITLG* |
| ENSG00000166959 | 11 | 60457047 | 60493284 | 39 | 7 | 44165 | -0.62736 | 0.73479 | *MS4A8* |
| ENSG00000055147 | 5 | 153359688 | 153428496 | 179 | 11 | 55349 | -0.62757 | 0.73486 | *FAM114A2* |
| ENSG00000135823 | 1 | 180931861 | 181002047 | 174 | 26 | 49591 | -0.6276 | 0.73487 | *STX6* |
| ENSG00000188305 | 19 | 2264631 | 2292181 | 106 | 19 | 26387 | -0.62761 | 0.73487 | *C19orf35* |
| ENSG00000147885 | 9 | 21206372 | 21227310 | 158 | 14 | 48848 | -0.62782 | 0.73494 | *IFNA16* |
| ENSG00000256892 | 10 | 37880366 | 37901859 | 92 | 10 | 55228 | -0.62811 | 0.73504 | *MTRNR2L7* |
| ENSG00000140015 | 14 | 63163287 | 63578755 | 1252 | 107 | 45994 | -0.62815 | 0.73505 | *KCNH5* |
| ENSG00000139746 | 13 | 79875962 | 79990612 | 362 | 14 | 46661 | -0.62832 | 0.7351 | *RBM26* |
| ENSG00000054654 | 14 | 64309683 | 64703165 | 1158 | 92 | 41183 | -0.62875 | 0.73524 | *SYNE2* |
| ENSG00000205439 | 21 | 46067849 | 46088258 | 123 | 13 | 51556 | -0.62874 | 0.73524 | *KRTAP12-3* |
| ENSG00000104671 | 8 | 30003813 | 30051156 | 95 | 15 | 39508 | -0.62889 | 0.73529 | *DCTN6* |
| ENSG00000085788 | 8 | 38072736 | 38143076 | 83 | 11 | 53581 | -0.62904 | 0.73534 | *DDHD2* |
| ENSG00000213023 | 19 | 51114564 | 51181651 | 192 | 46 | 44837 | -0.6292 | 0.73539 | *SYT3* |
| ENSG00000239857 | 7 | 906189 | 946073 | 197 | 43 | 51045 | -0.62934 | 0.73544 | *GET4* |
| ENSG00000019169 | 2 | 119689742 | 119762236 | 283 | 45 | 38019 | -0.62944 | 0.73547 | *MARCO* |
| ENSG00000175832 | 17 | 41595212 | 41666988 | 152 | 14 | 53352 | -0.62985 | 0.7356 | *ETV4* |
| ENSG00000147684 | 8 | 125541344 | 125590751 | 217 | 34 | 45607 | -0.62998 | 0.73565 | *NDUFB9* |
| ENSG00000175048 | 6 | 157792165 | 158109178 | 1235 | 118 | 49378 | -0.63054 | 0.73583 | *ZDHHC14* |
| ENSG00000169967 | 2 | 128046306 | 128156041 | 319 | 33 | 54396 | -0.63067 | 0.73587 | *MAP3K2* |
| ENSG00000269795 | 1 | 243378058 | 243405911 | 63 | 12 | 39728 | -0.63078 | 0.73591 | *AC092782.1* |
| ENSG00000102575 | 19 | 11675475 | 11699823 | 78 | 19 | 47558 | -0.63091 | 0.73595 | *ACP5* |
| ENSG00000185519 | 1 | 16374264 | 16410127 | 214 | 28 | 41671 | -0.63108 | 0.736 | *FAM131C* |
| ENSG00000198964 | 10 | 52055360 | 52394923 | 932 | 84 | 48075 | -0.63151 | 0.73615 | *SGMS1* |
| ENSG00000149091 | 11 | 46344455 | 46412104 | 93 | 24 | 45760 | -0.63157 | 0.73617 | *DGKZ* |
| ENSG00000125734 | 19 | 6719925 | 6747614 | 77 | 23 | 51023 | -0.63196 | 0.73629 | *GPR108* |
| ENSG00000229644 | 10 | 36800649 | 36823162 | 54 | 11 | 48376 | -0.6325 | 0.73647 | *NAMPTL* |
| ENSG00000205850 | 13 | 24885264 | 24905566 | 104 | 14 | 40580 | -0.63257 | 0.73649 | *AL359736.1* |
| ENSG00000100347 | 22 | 44341301 | 44416411 | 291 | 39 | 47522 | -0.63274 | 0.73655 | *SAMM50* |
| ENSG00000008128 | 1 | 1624169 | 1665766 | 21 | 15 | 39428 | -0.63281 | 0.73657 | *CDK11A* |
| ENSG00000165841 | 10 | 96437911 | 96623017 | 573 | 34 | 50262 | -0.6335 | 0.73679 | *CYP2C19* |
| ENSG00000134250 | 1 | 120444176 | 120622240 | 181 | 21 | 28163 | -0.63355 | 0.73681 | *NOTCH2* |

| | | | | | | | | | |
|---|---|---|---|---|---|---|---|---|---|
| ENSG00000128805 | 10 | 49644077 | 49874310 | 791 | 94 | 41354 | -0.63355 | 0.73681 | *ARHGAP22* |
| ENSG00000253309 | 13 | 51899909 | 51948871 | 118 | 12 | 54602 | -0.63362 | 0.73683 | *SERPINE3* |
| ENSG00000162851 | 1 | 246693862 | 246739626 | 259 | 34 | 36896 | -0.63367 | 0.73685 | *TFB2M* |
| ENSG00000143107 | 1 | 109245279 | 109295365 | 112 | 23 | 38598 | -0.63369 | 0.73686 | *FNDC7* |
| ENSG00000213593 | 11 | 57470072 | 57518445 | 75 | 11 | 44416 | -0.63368 | 0.73686 | *TMX2* |
| ENSG00000107099 | 9 | 204854 | 475259 | 1516 | 131 | 46032 | -0.63376 | 0.73688 | *DOCK8* |
| ENSG00000173464 | 14 | 21041054 | 21088043 | 233 | 52 | 49230 | -0.63379 | 0.73689 | *RNASE11* |
| ENSG00000164611 | 5 | 159838829 | 159865748 | 75 | 20 | 49269 | -0.6343 | 0.73706 | *PTTG1* |
| ENSG00000088451 | 13 | 95216308 | 95258511 | 122 | 11 | 45278 | -0.63433 | 0.73707 | *TGDS* |
| ENSG00000187527 | 3 | 192982579 | 193106632 | 456 | 51 | 48676 | -0.63457 | 0.73715 | *ATP13A5* |
| ENSG00000124275 | 5 | 7841299 | 7916138 | 383 | 45 | 46628 | -0.63483 | 0.73723 | *MTRR* |
| ENSG00000165912 | 11 | 47189076 | 47217994 | 60 | 13 | 45320 | -0.63525 | 0.73737 | *PACSIN3* |
| ENSG00000105877 | 7 | 21572833 | 21951457 | 1776 | 196 | 43297 | -0.63541 | 0.73742 | *DNAH11* |
| ENSG00000112419 | 6 | 143847982 | 144162322 | 1117 | 104 | 49266 | -0.63561 | 0.73748 | *PHACTR2* |
| ENSG00000110324 | 11 | 117847063 | 117882196 | 96 | 16 | 40687 | -0.63561 | 0.73749 | *IL10RA* |
| ENSG00000074706 | 6 | 154465631 | 154687926 | 863 | 119 | 40934 | -0.63566 | 0.7375 | *IPCEF1* |
| ENSG00000126453 | 19 | 50158823 | 50187173 | 42 | 11 | 38235 | -0.63664 | 0.73782 | *BCL2L12* |
| ENSG00000154133 | 11 | 124743587 | 124778396 | 124 | 25 | 41383 | -0.63667 | 0.73783 | *ROBO4* |
| ENSG00000186235 | 2 | 239123193 | 239150318 | 151 | 12 | 50131 | -0.63688 | 0.7379 | *AC016757.3* |
| ENSG00000146038 | 6 | 24161984 | 24368280 | 882 | 89 | 40329 | -0.63693 | 0.73792 | *DCDC2* |
| ENSG00000242631 | 9 | 109675630 | 109696455 | 40 | 14 | 50461 | -0.63694 | 0.73792 | *RP11-508N12.4* |
| ENSG00000166333 | 11 | 6614961 | 6642102 | 75 | 15 | 47422 | -0.63719 | 0.738 | *ILK* |
| ENSG00000106069 | 7 | 29151890 | 29563944 | 1511 | 196 | 45899 | -0.63745 | 0.73809 | *CHN2* |
| ENSG00000078304 | 14 | 102218135 | 102404326 | 631 | 71 | 40664 | -0.63762 | 0.73814 | *PPP2R5C* |
| ENSG00000205420 | 12 | 52870958 | 52897041 | 168 | 13 | 41606 | -0.63799 | 0.73826 | *KRT6A* |
| ENSG00000167654 | 19 | 3869862 | 3938077 | 211 | 57 | 38055 | -0.63807 | 0.73829 | *ATCAY* |
| ENSG00000125843 | 20 | 3791178 | 3815949 | 95 | 18 | 42061 | -0.63837 | 0.73838 | *AP5S1* |
| ENSG00000127472 | 1 | 20344672 | 20427683 | 296 | 40 | 45928 | -0.63909 | 0.73862 | *PLA2G5* |
| ENSG00000115084 | 2 | 114452588 | 114524400 | 280 | 23 | 42688 | -0.63909 | 0.73862 | *SLC35F5* |
| ENSG00000186583 | 8 | 145076582 | 145111933 | 38 | 16 | 45963 | -0.63937 | 0.73871 | *SPATC1* |
| ENSG00000205754 | 12 | 20958673 | 21255679 | 1220 | 50 | 32112 | -0.63989 | 0.73888 | *SLCO1B7* |
| ENSG00000129538 | 14 | 21259387 | 21281437 | 68 | 21 | 46028 | -0.6399 | 0.73888 | *RNASE1* |
| ENSG00000179755 | 16 | 34246507 | 34267278 | 13 | 9 | 44839 | -0.64078 | 0.73917 | *CTD-2144E22.5* |
| ENSG00000174123 | 4 | 38763860 | 38794611 | 174 | 18 | 20960 | -0.64096 | 0.73923 | *TLR10* |
| ENSG00000135272 | 7 | 114552209 | 114669256 | 246 | 48 | 42430 | -0.64113 | 0.73928 | *MDFIC* |
| ENSG00000013523 | 14 | 77243588 | 77302589 | 195 | 33 | 54934 | -0.6412 | 0.7393 | *ANGEL1* |

| | | | | | | | | | |
|---|---|---|---|---|---|---|---|---|---|
| ENSG00000125846 | 20 | 18259121 | 18307640 | 182 | 31 | 42581 | -0.64158 | 0.73943 | *ZNF133* |
| ENSG00000159618 | 16 | 57566333 | 57635593 | 250 | 35 | 38826 | -0.64178 | 0.73949 | *GPR114* |
| ENSG00000107317 | 9 | 139861956 | 139889887 | 90 | 15 | 51666 | -0.6421 | 0.7396 | *PTGDS* |
| ENSG00000124767 | 6 | 38633701 | 38680917 | 171 | 25 | 46119 | -0.64214 | 0.73961 | *GLO1* |
| ENSG00000099622 | 19 | 1249384 | 1284879 | 56 | 19 | 42472 | -0.64247 | 0.73972 | *CIRBP* |
| ENSG00000126524 | 7 | 66442664 | 66470588 | 96 | 27 | 43824 | -0.64251 | 0.73973 | *SBDS* |
| ENSG00000167083 | 17 | 47270153 | 47297936 | 90 | 17 | 48236 | -0.64252 | 0.73973 | *GNGT2* |
| ENSG00000135525 | 6 | 136653875 | 136881957 | 344 | 32 | 37901 | -0.64273 | 0.7398 | *MAP7* |
| ENSG00000259680 | 16 | 33637044 | 33657696 | 3 | 2 | 22665 | -0.64275 | 0.73981 | *RP11-812E19.9* |
| ENSG00000198887 | 9 | 72863937 | 72979804 | 346 | 28 | 53245 | -0.64299 | 0.73988 | *SMC5* |
| ENSG00000158258 | 3 | 139644027 | 140306239 | 2069 | 212 | 42453 | -0.64301 | 0.73989 | *CLSTN2* |
| ENSG00000092051 | 14 | 24027244 | 24058009 | 37 | 10 | 41987 | -0.64329 | 0.73998 | *JPH4* |
| ENSG00000257335 | 7 | 141597613 | 141816547 | 535 | 94 | 43614 | -0.64359 | 0.74008 | *MGAM* |
| ENSG00000147465 | 8 | 37991167 | 38018783 | 23 | 14 | 44466 | -0.6439 | 0.74018 | *STAR* |
| ENSG00000137221 | 6 | 43435261 | 43484294 | 102 | 12 | 46051 | -0.64397 | 0.7402 | *TJAP1* |
| ENSG00000104738 | 8 | 48862745 | 48900720 | 26 | 16 | 44135 | -0.64429 | 0.74031 | *MCM4* |
| ENSG00000109320 | 4 | 103412486 | 103548459 | 330 | 27 | 47287 | -0.64471 | 0.74044 | *NFKB1* |
| ENSG00000154493 | 10 | 128103566 | 128369079 | 871 | 142 | 49766 | -0.64532 | 0.74064 | *C10orf90* |
| ENSG00000104154 | 15 | 45761809 | 45825005 | 69 | 16 | 41633 | -0.64612 | 0.7409 | *SLC30A4* |
| ENSG00000180071 | 9 | 38530566 | 38630657 | 229 | 57 | 41293 | -0.64671 | 0.74109 | *ANKRD18A* |
| ENSG00000169047 | 2 | 227586033 | 227674475 | 220 | 25 | 34337 | -0.64687 | 0.74114 | *IRS1* |
| ENSG00000171243 | 7 | 16491106 | 16580205 | 399 | 52 | 49646 | -0.6472 | 0.74125 | *SOSTDC1* |
| ENSG00000268472 | 11 | 112120988 | 112142117 | 109 | 5 | 53785 | -0.64736 | 0.7413 | *AP002884.2* |
| ENSG00000224730 | 19 | 55137173 | 55160343 | 140 | 16 | 39652 | -0.6477 | 0.74141 | *AC009892.10* |
| ENSG00000149742 | 11 | 63127261 | 63187766 | 226 | 30 | 36315 | -0.64799 | 0.7415 | *SLC22A9* |
| ENSG00000161270 | 19 | 36306866 | 36370189 | 148 | 34 | 40677 | -0.648 | 0.74151 | *NPHS1* |
| ENSG00000170962 | 11 | 103767914 | 104045107 | 1064 | 95 | 49006 | -0.64836 | 0.74163 | *PDGFD* |
| ENSG00000163738 | 4 | 74969891 | 75178816 | 321 | 35 | 34947 | -0.64841 | 0.74164 | *MTHFD2L* |
| ENSG00000125863 | 20 | 10371657 | 10424870 | 246 | 16 | 29796 | -0.64845 | 0.74165 | *MKKS* |
| ENSG00000150764 | 11 | 111787868 | 111903308 | 198 | 24 | 34443 | -0.649 | 0.74183 | *DIXDC1* |
| ENSG00000187942 | 1 | 22128758 | 22161714 | 133 | 40 | 47289 | -0.64904 | 0.74185 | *LDLRAD2* |
| ENSG00000063241 | 19 | 55954352 | 55983710 | 113 | 34 | 45136 | -0.64924 | 0.74191 | *ISOC2* |
| ENSG00000090512 | 3 | 186343758 | 186380930 | 140 | 20 | 44901 | -0.64947 | 0.74198 | *FETUB* |
| ENSG00000126067 | 1 | 36057185 | 36117445 | 37 | 16 | 37739 | -0.64985 | 0.74211 | *PSMB2* |
| ENSG00000268154 | 1 | 161025655 | 161045943 | 45 | 14 | 47493 | -0.64991 | 0.74213 | *AL591806.1* |
| ENSG00000052723 | 1 | 115302100 | 115333306 | 39 | 13 | 51979 | -0.65012 | 0.74219 | *SIKE1* |

| | | | | | | | | | |
|---|---|---|---|---|---|---|---|---|---|
| ENSG00000170423 | 12 | 53221588 | 53252876 | 64 | 13 | 49612 | -0.65014 | 0.7422 | *KRT78* |
| ENSG00000128294 | 22 | 26911458 | 27002681 | 392 | 61 | 49923 | -0.65028 | 0.74224 | *TPST2* |
| ENSG00000119630 | 14 | 75398537 | 75432487 | 79 | 14 | 49303 | -0.65065 | 0.74236 | *PGF* |
| ENSG00000068912 | 2 | 54004181 | 54055956 | 147 | 21 | 42691 | -0.65075 | 0.7424 | *ERLEC1* |
| ENSG00000035928 | 4 | 39279076 | 39377995 | 195 | 16 | 34690 | -0.65088 | 0.74244 | *RFC1* |
| ENSG00000129221 | 17 | 6287013 | 6348519 | 373 | 74 | 44206 | -0.65091 | 0.74245 | *AIPL1* |
| ENSG00000186908 | 12 | 77147368 | 77257476 | 451 | 24 | 53457 | -0.65097 | 0.74247 | *ZDHHC17* |
| ENSG00000171456 | 20 | 30936155 | 31037122 | 158 | 11 | 47966 | -0.65103 | 0.74249 | *ASXL1* |
| ENSG00000111732 | 12 | 8744762 | 8775467 | 73 | 19 | 39325 | -0.65112 | 0.74252 | *AICDA* |
| ENSG00000228083 | 9 | 21229201 | 21249978 | 89 | 8 | 50141 | -0.65143 | 0.74261 | *IFNA14* |
| ENSG00000107130 | 9 | 132924857 | 133009583 | 325 | 59 | 46602 | -0.65145 | 0.74262 | *NCS1* |
| ENSG00000062282 | 11 | 75460557 | 75522579 | 83 | 10 | 39039 | -0.65162 | 0.74268 | *DGAT2* |
| ENSG00000182634 | 11 | 123898773 | 123919717 | 64 | 16 | 36556 | -0.65185 | 0.74275 | *OR10G7* |
| ENSG00000162836 | 1 | 147091453 | 147152618 | 183 | 28 | 44848 | -0.65191 | 0.74277 | *ACP6* |
| ENSG00000146926 | 7 | 150862785 | 150894919 | 91 | 19 | 44590 | -0.65213 | 0.74284 | *ASB10* |
| ENSG00000222005 | 2 | 47033807 | 47059796 | 105 | 23 | 45725 | -0.65224 | 0.74288 | *LINC01118* |
| ENSG00000005486 | 7 | 75461920 | 75528244 | 166 | 28 | 41872 | -0.6525 | 0.74296 | *RHBDD2* |
| ENSG00000166949 | 15 | 67346101 | 67497533 | 447 | 85 | 48095 | -0.65271 | 0.74303 | *SMAD3* |
| ENSG00000078549 | 7 | 31082076 | 31161089 | 234 | 57 | 42357 | -0.65296 | 0.74311 | *ADCYAP1R1* |
| ENSG00000212670 | 1 | 54559968 | 54581750 | 52 | 8 | 25786 | -0.65301 | 0.74312 | *AL161915.1* |
| ENSG00000163202 | 1 | 152541857 | 152562980 | 32 | 10 | 35296 | -0.65299 | 0.74312 | *LCE3D* |
| ENSG00000105143 | 19 | 15050846 | 15143802 | 632 | 77 | 45941 | -0.65334 | 0.74323 | *SLC1A6* |
| ENSG00000258729 | 14 | 96731383 | 96753060 | 85 | 19 | 49486 | -0.65338 | 0.74324 | *DKFZP434O1614* |
| ENSG00000133800 | 11 | 10568513 | 10643236 | 268 | 39 | 47910 | -0.65368 | 0.74334 | *LYVE1* |
| ENSG00000163431 | 1 | 201855580 | 201925715 | 177 | 34 | 46970 | -0.65369 | 0.74335 | *LMOD1* |
| ENSG00000110274 | 11 | 117175273 | 117293984 | 344 | 42 | 48707 | -0.65416 | 0.74349 | *CEP164* |
| ENSG00000114125 | 3 | 141447046 | 141476402 | 105 | 13 | 49579 | -0.6542 | 0.74351 | *RNF7* |
| ENSG00000181963 | 11 | 4460525 | 4481591 | 106 | 24 | 48191 | -0.65423 | 0.74352 | *OR52K2* |
| ENSG00000166311 | 11 | 6401655 | 6426228 | 71 | 16 | 51938 | -0.65424 | 0.74352 | *SMPD1* |
| ENSG00000155542 | 5 | 56195087 | 56231359 | 124 | 15 | 39830 | -0.65434 | 0.74355 | *SETD9* |
| ENSG00000116525 | 1 | 33601003 | 33657660 | 133 | 11 | 51507 | -0.65486 | 0.74372 | *TRIM62* |
| ENSG00000132010 | 19 | 12193658 | 12261222 | 133 | 22 | 39512 | -0.65499 | 0.74376 | *ZNF20* |
| ENSG00000156299 | 21 | 32480734 | 32942290 | 1474 | 182 | 42908 | -0.65497 | 0.74376 | *TIAM1* |
| ENSG00000111913 | 6 | 24787601 | 25052238 | 1024 | 99 | 39240 | -0.6551 | 0.7438 | *FAM65B* |
| ENSG00000126457 | 19 | 50169043 | 50202286 | 57 | 17 | 38956 | -0.65512 | 0.7438 | *PRMT1* |
| ENSG00000182974 | 15 | 22358478 | 22379561 | 4 | 2 | 43643 | -0.65514 | 0.74381 | *OR4M2* |

| | | | | | | | | | |
|---|---|---|---|---|---|---|---|---|---|
| ENSG00000175426 | 5 | 95716119 | 95779847 | 139 | 26 | 47403 | -0.65526 | 0.74385 | *PCSK1* |
| ENSG00000167302 | 17 | 79192077 | 79222891 | 116 | 21 | 33668 | -0.65528 | 0.74386 | *ENTHD2* |
| ENSG00000156858 | 16 | 30652038 | 30677761 | 18 | 8 | 30148 | -0.65582 | 0.74403 | *PRR14* |
| ENSG00000160182 | 21 | 43772391 | 43796703 | 150 | 31 | 43215 | -0.65621 | 0.74416 | *TFF1* |
| ENSG00000175782 | 12 | 69129886 | 69197508 | 188 | 40 | 44554 | -0.65626 | 0.74417 | *SLC35E3* |
| ENSG00000241322 | 17 | 15458798 | 15532826 | 235 | 33 | 38408 | -0.65674 | 0.74433 | *CDRT1* |
| ENSG00000213030 | 19 | 49540895 | 49562400 | 92 | 25 | 47765 | -0.65686 | 0.74436 | *CGB8* |
| ENSG00000206562 | 3 | 15412782 | 15492073 | 220 | 29 | 43754 | -0.65692 | 0.74439 | *METTL6* |
| ENSG00000065361 | 12 | 56463641 | 56507289 | 75 | 20 | 43921 | -0.6576 | 0.7446 | *ERBB3* |
| ENSG00000169548 | 22 | 22858060 | 22884613 | 81 | 21 | 43036 | -0.65763 | 0.74461 | *ZNF280A* |
| ENSG00000100284 | 22 | 35685268 | 35753985 | 209 | 21 | 33493 | -0.65765 | 0.74462 | *TOM1* |
| ENSG00000177994 | 2 | 54547171 | 54620879 | 236 | 38 | 42535 | -0.65803 | 0.74474 | *C2orf73* |
| ENSG00000112941 | 5 | 6704718 | 6767161 | 282 | 31 | 47561 | -0.65807 | 0.74475 | *PAPD7* |
| ENSG00000188771 | 11 | 112108876 | 112141583 | 127 | 7 | 53015 | -0.65809 | 0.74476 | *PLET1* |
| ENSG00000171246 | 17 | 78430948 | 78461643 | 83 | 24 | 47252 | -0.65812 | 0.74477 | *NPTX1* |
| ENSG00000269711 | 19 | 7733387 | 7757740 | 184 | 19 | 40370 | -0.65821 | 0.7448 | *CTD-3214H19.16* |
| ENSG00000147535 | 8 | 38110648 | 38136761 | 33 | 7 | 54362 | -0.6584 | 0.74486 | *PPAPDC1B* |
| ENSG00000165060 | 9 | 71640175 | 71725094 | 385 | 34 | 52453 | -0.65861 | 0.74493 | *FXN* |
| ENSG00000137806 | 15 | 41669551 | 41704717 | 77 | 13 | 38887 | -0.65946 | 0.7452 | *NDUFAF1* |
| ENSG00000124875 | 4 | 74692214 | 74724781 | 91 | 20 | 40115 | -0.65958 | 0.74524 | *CXCL6* |
| ENSG00000112130 | 6 | 37311748 | 37372514 | 76 | 29 | 44679 | -0.65964 | 0.74526 | *RNF8* |
| ENSG00000146374 | 6 | 127429749 | 127528910 | 212 | 19 | 52171 | -0.65995 | 0.74536 | *RSPO3* |
| ENSG00000196156 | 17 | 39313484 | 39334424 | 64 | 9 | 50323 | -0.66041 | 0.74551 | *KRTAP4-3* |
| ENSG00000117834 | 1 | 48678357 | 48724316 | 123 | 21 | 43061 | -0.66048 | 0.74553 | *SLC5A9* |
| ENSG00000179772 | 20 | 30422103 | 30443420 | 50 | 13 | 40058 | -0.66071 | 0.7456 | *FOXS1* |
| ENSG00000140577 | 15 | 91063157 | 91198577 | 480 | 42 | 43280 | -0.66092 | 0.74567 | *CRTC3* |
| ENSG00000170298 | 17 | 20342708 | 20380852 | 38 | 9 | 35202 | -0.66121 | 0.74576 | *LGALS9B* |
| ENSG00000145287 | 4 | 84001201 | 84068228 | 125 | 30 | 44603 | -0.66124 | 0.74577 | *PLAC8* |
| ENSG00000183128 | 10 | 105222561 | 105248997 | 76 | 14 | 45083 | -0.66132 | 0.7458 | *CALHM3* |
| ENSG00000162999 | 2 | 183933287 | 183974733 | 132 | 19 | 42001 | -0.66153 | 0.74586 | *DUSP19* |
| ENSG00000130559 | 9 | 138690333 | 138809074 | 226 | 27 | 46903 | -0.66159 | 0.74588 | *CAMSAP1* |
| ENSG00000242366 | 2 | 234658894 | 234691945 | 121 | 25 | 44228 | -0.6619 | 0.74598 | *UGT1A1* |
| ENSG00000180354 | 7 | 30164426 | 30212378 | 146 | 27 | 51545 | -0.66216 | 0.74607 | *MTURN* |
| ENSG00000179431 | 11 | 35629735 | 35652419 | 59 | 17 | 51518 | -0.66231 | 0.74611 | *FJX1* |
| ENSG00000135314 | 6 | 73941037 | 74030088 | 330 | 29 | 45531 | -0.6625 | 0.74617 | *KHDC1* |
| ENSG00000105675 | 19 | 36030945 | 36064560 | 83 | 25 | 46319 | -0.66249 | 0.74617 | *ATP4A* |

| | | | | | | | | | |
|---|---|---|---|---|---|---|---|---|---|
| ENSG00000148688 | 10 | 92621473 | 92678312 | 126 | 18 | 36486 | -0.66252 | 0.74618 | *RPP30* |
| ENSG00000156411 | 14 | 104368625 | 104404606 | 166 | 18 | 45622 | -0.66289 | 0.7463 | *C14orf2* |
| ENSG00000111536 | 12 | 68585131 | 68629601 | 149 | 25 | 33745 | -0.66312 | 0.74637 | *IL26* |
| ENSG00000170950 | 6 | 49743366 | 49765053 | 65 | 13 | 50205 | -0.66339 | 0.74646 | *PGK2* |
| ENSG00000133026 | 17 | 8367523 | 8544079 | 307 | 24 | 51749 | -0.66341 | 0.74647 | *MYH10* |
| ENSG00000131844 | 5 | 70873115 | 70964531 | 245 | 27 | 48760 | -0.6635 | 0.74649 | *MCCC2* |
| ENSG00000197124 | 19 | 20097867 | 20160315 | 265 | 21 | 39847 | -0.66355 | 0.74651 | *ZNF682* |
| ENSG00000104901 | 19 | 49855040 | 49888373 | 126 | 8 | 50983 | -0.664 | 0.74665 | *DKKL1* |
| ENSG00000182158 | 7 | 137549725 | 137696813 | 370 | 64 | 38879 | -0.6641 | 0.74669 | *CREB3L2* |
| ENSG00000100441 | 14 | 24888492 | 24920540 | 188 | 16 | 27645 | -0.66423 | 0.74673 | *KHNYN* |
| ENSG00000250506 | 17 | 73986987 | 74012080 | 65 | 17 | 46704 | -0.66453 | 0.74682 | *CDK3* |
| ENSG00000169951 | 16 | 30555085 | 30579819 | 27 | 11 | 34955 | -0.66458 | 0.74684 | *ZNF764* |
| ENSG00000118640 | 2 | 85778685 | 85819154 | 141 | 17 | 47586 | -0.66468 | 0.74687 | *VAMP8* |
| ENSG00000175097 | 11 | 36587124 | 36629829 | 116 | 14 | 46863 | -0.66468 | 0.74687 | *RAG2* |
| ENSG00000136936 | 9 | 100427191 | 100469639 | 61 | 15 | 43593 | -0.66495 | 0.74696 | *XPA* |
| ENSG00000184270 | 1 | 149849019 | 149869466 | 39 | 6 | 52926 | -0.6651 | 0.74701 | *HIST2H2AB* |
| ENSG00000196758 | 2 | 240489995 | 240517788 | 115 | 29 | 50322 | -0.66535 | 0.74709 | *AC079612.1* |
| ENSG00000185261 | 5 | 93478671 | 93964309 | 827 | 131 | 51004 | -0.66563 | 0.74718 | *KIAA0825* |
| ENSG00000130332 | 19 | 2311516 | 2338619 | 117 | 23 | 43848 | -0.66573 | 0.74721 | *LSM7* |
| ENSG00000149428 | 11 | 118904900 | 118937913 | 78 | 15 | 46781 | -0.66586 | 0.74725 | *HYOU1* |
| ENSG00000120688 | 13 | 41625410 | 41668137 | 55 | 19 | 44414 | -0.66605 | 0.74731 | *WBP4* |
| ENSG00000150977 | 12 | 123889936 | 123931264 | 126 | 30 | 42287 | -0.66632 | 0.7474 | *RILPL2* |
| ENSG00000185684 | 12 | 132558828 | 132623029 | 168 | 17 | 29067 | -0.66648 | 0.74745 | *EP400NL* |
| ENSG00000228835 | 18 | 30339758 | 30364376 | 49 | 10 | 32676 | -0.66673 | 0.74753 | *AC012123.1* |
| ENSG00000111700 | 12 | 20953636 | 21253040 | 1236 | 53 | 32410 | -0.66738 | 0.74774 | *SLCO1B3* |
| ENSG00000269403 | 19 | 51843627 | 51881353 | 135 | 24 | 36536 | -0.66771 | 0.74784 | *CTD-2616J11.11* |
| ENSG00000160050 | 1 | 32655987 | 32680988 | 39 | 10 | 36761 | -0.66904 | 0.74827 | *CCDC28B* |
| ENSG00000172986 | 3 | 72927224 | 73057289 | 443 | 55 | 38662 | -0.66928 | 0.74834 | *GXYLT2* |
| ENSG00000175520 | 11 | 5518530 | 5541215 | 66 | 17 | 43047 | -0.66958 | 0.74844 | *UBQLN3* |
| ENSG00000185900 | 8 | 42938658 | 42988577 | 94 | 18 | 46534 | -0.66984 | 0.74852 | *POMK* |
| ENSG00000138028 | 2 | 27311757 | 27351995 | 71 | 13 | 48192 | -0.6699 | 0.74854 | *CGREF1* |
| ENSG00000139318 | 12 | 89731009 | 89757048 | 100 | 16 | 47508 | -0.67004 | 0.74858 | *DUSP6* |
| ENSG00000075142 | 7 | 87824433 | 87866308 | 73 | 16 | 40238 | -0.67032 | 0.74867 | *SRI* |
| ENSG00000187922 | 9 | 139622619 | 139647808 | 96 | 15 | 47338 | -0.67079 | 0.74882 | *LCN10* |
| ENSG00000153237 | 2 | 159017593 | 159323265 | 1514 | 86 | 40519 | -0.67095 | 0.74887 | *CCDC148* |
| ENSG00000176928 | 5 | 74313289 | 74336724 | 78 | 17 | 43773 | -0.67211 | 0.74924 | *GCNT4* |

| | | | | | | | | | |
|---|---|---|---|---|---|---|---|---|---|
| ENSG00000186977 | 21 | 31863975 | 31884435 | 41 | 5 | 53908 | -0.67212 | 0.74925 | *KRTAP19-5* |
| ENSG00000106299 | 7 | 123311989 | 123399121 | 229 | 18 | 54818 | -0.67221 | 0.74928 | *WASL* |
| ENSG00000197928 | 19 | 53721577 | 53768151 | 218 | 18 | 41675 | -0.67234 | 0.74932 | *ZNF677* |
| ENSG00000138433 | 2 | 175202750 | 175270443 | 159 | 23 | 40351 | -0.67239 | 0.74933 | *CIR1* |
| ENSG00000033800 | 15 | 68336517 | 68493096 | 242 | 31 | 42636 | -0.67251 | 0.74937 | *PIAS1* |
| ENSG00000148513 | 10 | 37404785 | 37683039 | 734 | 46 | 47190 | -0.67313 | 0.74957 | *ANKRD30A* |
| ENSG00000179979 | 4 | 1375340 | 1399780 | 58 | 15 | 41092 | -0.6732 | 0.74959 | *CRIPAK* |
| ENSG00000155393 | 16 | 50089852 | 50150298 | 217 | 17 | 47117 | -0.67364 | 0.74973 | *HEATR3* |
| ENSG00000171953 | 17 | 17870723 | 17952523 | 103 | 16 | 35995 | -0.67387 | 0.7498 | *ATPAF2* |
| ENSG00000197641 | 18 | 61244223 | 61281873 | 59 | 12 | 47681 | -0.67388 | 0.7498 | *SERPINB13* |
| ENSG00000151834 | 4 | 46240444 | 46487247 | 620 | 46 | 48138 | -0.67394 | 0.74982 | *GABRA2* |
| ENSG00000197376 | 12 | 48891137 | 48933680 | 208 | 20 | 32679 | -0.67404 | 0.74986 | *OR8S1* |
| ENSG00000136514 | 3 | 187076120 | 187099864 | 91 | 23 | 49672 | -0.67446 | 0.74999 | *RTP4* |
| ENSG00000173436 | 1 | 19913477 | 19966314 | 158 | 25 | 47078 | -0.67497 | 0.75015 | *MINOS1* |
| ENSG00000181029 | 19 | 7735729 | 7757744 | 177 | 18 | 40955 | -0.67501 | 0.75016 | *TRAPPC5* |
| ENSG00000168242 | 6 | 26263144 | 26283622 | 72 | 13 | 39519 | -0.67506 | 0.75018 | *HIST1H2BI* |
| ENSG00000164626 | 6 | 39146749 | 39207226 | 218 | 46 | 47189 | -0.67526 | 0.75024 | *KCNK5* |
| ENSG00000170166 | 2 | 177005950 | 177027954 | 46 | 12 | 35835 | -0.6755 | 0.75032 | *HOXD4* |
| ENSG00000164066 | 4 | 128534426 | 128657892 | 229 | 27 | 49076 | -0.67584 | 0.75043 | *INTU* |
| ENSG00000100024 | 22 | 24853206 | 24934358 | 229 | 36 | 39258 | -0.67594 | 0.75046 | *UPB1* |
| ENSG00000235156 | 3 | 99894531 | 99923035 | 57 | 11 | 51647 | -0.67607 | 0.7505 | *TMEM30C* |
| ENSG00000171421 | 5 | 1788500 | 1811480 | 97 | 12 | 34819 | -0.67635 | 0.75059 | *MRPL36* |
| ENSG00000186967 | 21 | 31859142 | 31879451 | 37 | 4 | 54714 | -0.67665 | 0.75069 | *KRTAP19-4* |
| ENSG00000213931 | 11 | 5279582 | 5536847 | 1729 | 98 | 43721 | -0.67692 | 0.75077 | *HBE1* |
| ENSG00000133110 | 13 | 38126720 | 38182981 | 200 | 23 | 38755 | -0.67696 | 0.75079 | *POSTN* |
| ENSG00000002834 | 17 | 37016112 | 37088023 | 231 | 55 | 48165 | -0.67703 | 0.75081 | *LASP1* |
| ENSG00000170523 | 12 | 52698085 | 52725182 | 173 | 16 | 53969 | -0.67724 | 0.75087 | *KRT83* |
| ENSG00000049540 | 7 | 73432119 | 73494237 | 134 | 25 | 36772 | -0.67741 | 0.75093 | *ELN* |
| ENSG00000161911 | 6 | 41107080 | 41132075 | 22 | 5 | 50800 | -0.6775 | 0.75096 | *TREML1* |
| ENSG00000170876 | 3 | 14156440 | 14195179 | 96 | 21 | 47019 | -0.67754 | 0.75097 | *TMEM43* |
| ENSG00000171105 | 19 | 7102266 | 7304045 | 841 | 132 | 45305 | -0.67761 | 0.75099 | *INSR* |
| ENSG00000117713 | 1 | 27012524 | 27118595 | 165 | 12 | 54640 | -0.67781 | 0.75105 | *ARID1A* |
| ENSG00000172150 | 17 | 3090813 | 3111742 | 68 | 15 | 46459 | -0.67779 | 0.75105 | *OR1A2* |
| ENSG00000152147 | 2 | 38968676 | 39022142 | 126 | 17 | 40642 | -0.67802 | 0.75112 | *GEMIN6* |
| ENSG00000169021 | 19 | 29688173 | 29714448 | 104 | 15 | 53448 | -0.67807 | 0.75114 | *UQCRFS1* |
| ENSG00000167981 | 16 | 3476104 | 3503542 | 101 | 13 | 47271 | -0.67817 | 0.75117 | *ZNF597* |

| | | | | | | | | | |
|---|---|---|---|---|---|---|---|---|---|
| ENSG00000077943 | 10 | 15545948 | 15772124 | 769 | 75 | 41942 | -0.67862 | 0.75131 | *ITGA8* |
| ENSG00000186523 | 8 | 12029605 | 12061642 | 6 | 3 | 42829 | -0.67869 | 0.75133 | *FAM86B1* |
| ENSG00000268400 | 19 | 7684682 | 7717934 | 107 | 15 | 54060 | -0.67875 | 0.75135 | *CTD-3214H19.4* |
| ENSG00000267552 | 19 | 38170766 | 38220089 | 124 | 19 | 37610 | -0.67959 | 0.75162 | *CTD-2528L19.4* |
| ENSG00000242107 | 3 | 159723811 | 159759210 | 174 | 18 | 37566 | -0.6797 | 0.75165 | *LINC01100* |
| ENSG00000186924 | 21 | 31963414 | 31983612 | 48 | 9 | 55135 | -0.68003 | 0.75176 | *KRTAP22-1* |
| ENSG00000110079 | 11 | 60038014 | 60086445 | 141 | 24 | 51432 | -0.68009 | 0.75178 | *MS4A4A* |
| ENSG00000180535 | 7 | 97830739 | 97852291 | 65 | 18 | 39799 | -0.68042 | 0.75188 | *BHLHA15* |
| ENSG00000141030 | 17 | 17140141 | 17194607 | 185 | 16 | 52682 | -0.68044 | 0.75189 | *COPS3* |
| ENSG00000174407 | 20 | 61137660 | 61177971 | 184 | 18 | 46440 | -0.68064 | 0.75195 | *C20orf166* |
| ENSG00000269089 | 11 | 61725453 | 61746755 | 44 | 15 | 31303 | -0.68108 | 0.75209 | *AP003733.1* |
| ENSG00000204160 | 1 | 27143201 | 27194093 | 83 | 12 | 49850 | -0.6812 | 0.75213 | *ZDHHC18* |
| ENSG00000255833 | 5 | 134769908 | 134798089 | 104 | 18 | 35134 | -0.68127 | 0.75215 | *TIFAB* |
| ENSG00000124151 | 20 | 46120601 | 46295621 | 448 | 33 | 45208 | -0.68137 | 0.75218 | *NCOA3* |
| ENSG00000067113 | 5 | 54710682 | 54840878 | 380 | 19 | 32061 | -0.68147 | 0.75221 | *PPAP2A* |
| ENSG00000205549 | 9 | 16193933 | 16286311 | 393 | 70 | 41020 | -0.68154 | 0.75224 | *C9orf92* |
| ENSG00000143839 | 1 | 204113944 | 204145465 | 109 | 21 | 50863 | -0.68209 | 0.75241 | *REN* |
| ENSG00000163611 | 3 | 113151565 | 113244034 | 421 | 31 | 49924 | -0.68209 | 0.75241 | *SPICE1* |
| ENSG00000125510 | 20 | 62701526 | 62741996 | 118 | 23 | 51505 | -0.68234 | 0.75249 | *OPRL1* |
| ENSG00000172322 | 12 | 10093915 | 10158293 | 373 | 34 | 48013 | -0.68254 | 0.75255 | *CLEC12A* |
| ENSG00000205356 | 7 | 97833936 | 97891563 | 171 | 24 | 42312 | -0.6826 | 0.75257 | *TECPR1* |
| ENSG00000187398 | 11 | 24508516 | 25114150 | 2900 | 176 | 43074 | -0.68271 | 0.7526 | *LUZP2* |
| ENSG00000169826 | 10 | 43623934 | 43690756 | 188 | 19 | 44240 | -0.68274 | 0.75261 | *CSGALNACT2* |
| ENSG00000144868 | 3 | 132747235 | 133126636 | 1328 | 96 | 52799 | -0.68297 | 0.75269 | *TMEM108* |
| ENSG00000037241 | 5 | 172375732 | 172406774 | 147 | 23 | 34493 | -0.68363 | 0.7529 | *RPL26L1* |
| ENSG00000170949 | 19 | 53559859 | 53616689 | 278 | 40 | 52610 | -0.6839 | 0.75298 | *ZNF160* |
| ENSG00000000419 | 20 | 49541404 | 49585092 | 74 | 16 | 38909 | -0.6841 | 0.75304 | *DPM1* |
| ENSG00000162782 | 1 | 179550748 | 179670407 | 442 | 32 | 52671 | -0.68415 | 0.75306 | *TDRD5* |
| ENSG00000104824 | 19 | 39317028 | 39352987 | 95 | 13 | 36830 | -0.68423 | 0.75308 | *HNRNPL* |
| ENSG00000068078 | 4 | 1785034 | 1820599 | 81 | 27 | 40234 | -0.68477 | 0.75325 | *FGFR3* |
| ENSG00000212997 | 8 | 95548771 | 95570630 | 113 | 14 | 40282 | -0.68478 | 0.75326 | *AC023632.1* |
| ENSG00000134365 | 1 | 196809371 | 196898102 | 206 | 27 | 23326 | -0.68495 | 0.75331 | *CFHR4* |
| ENSG00000117148 | 1 | 18071808 | 18163558 | 261 | 38 | 41028 | -0.6854 | 0.75345 | *ACTL8* |
| ENSG00000256229 | 19 | 20268037 | 20321299 | 177 | 13 | 24267 | -0.68578 | 0.75357 | *ZNF486* |
| ENSG00000157693 | 9 | 117363486 | 117418702 | 287 | 29 | 51244 | -0.68598 | 0.75364 | *C9orf91* |
| ENSG00000104331 | 8 | 57860492 | 57916403 | 116 | 13 | 51912 | -0.6863 | 0.75374 | *IMPAD1* |

| | | | | | | | | | |
|---|---|---|---|---|---|---|---|---|---|
| ENSG00000173915 | 10 | 105138798 | 105166223 | 34 | 9 | 39633 | -0.68634 | 0.75375 | *USMG5* |
| ENSG00000126787 | 14 | 55604830 | 55668396 | 144 | 19 | 45853 | -0.68637 | 0.75376 | *DLGAP5* |
| ENSG00000113327 | 5 | 161484546 | 161592542 | 230 | 42 | 53247 | -0.68667 | 0.75385 | *GABRG2* |
| ENSG00000185621 | 3 | 197677071 | 197780591 | 333 | 29 | 45097 | -0.6869 | 0.75393 | *LMLN* |
| ENSG00000102539 | 13 | 49784474 | 49806513 | 25 | 5 | 37823 | -0.68718 | 0.75402 | *MLNR* |
| ENSG00000259164 | 14 | 77554654 | 77701805 | 603 | 79 | 45227 | -0.68724 | 0.75404 | *RP11-463C8.4* |
| ENSG00000164627 | 6 | 39287766 | 39703181 | 1331 | 108 | 43449 | -0.68728 | 0.75405 | *KIF6* |
| ENSG00000271043 | 5 | 79935819 | 79956855 | 132 | 6 | 55608 | -0.68741 | 0.75409 | *MTRNR2L2* |
| ENSG00000166736 | 11 | 113835603 | 113871035 | 95 | 23 | 45612 | -0.68764 | 0.75416 | *HTR3A* |
| ENSG00000171320 | 8 | 27619466 | 27680157 | 276 | 13 | 30838 | -0.68785 | 0.75423 | *ESCO2* |
| ENSG00000140368 | 15 | 77275700 | 77339673 | 150 | 28 | 40631 | -0.68785 | 0.75423 | *PSTPIP1* |
| ENSG00000235098 | 1 | 1343800 | 1367149 | 77 | 10 | 26913 | -0.6881 | 0.7543 | *ANKRD65* |
| ENSG00000103540 | 16 | 19525133 | 19574730 | 183 | 20 | 41906 | -0.68809 | 0.7543 | *CCP110* |
| ENSG00000158483 | 11 | 71488556 | 71522282 | 211 | 13 | 42912 | -0.68856 | 0.75445 | *FAM86C1* |
| ENSG00000138185 | 10 | 97461536 | 97647023 | 620 | 39 | 45712 | -0.68876 | 0.75451 | *ENTPD1* |
| ENSG00000105388 | 19 | 42202504 | 42243718 | 139 | 20 | 40582 | -0.68889 | 0.75455 | *CEACAM5* |
| ENSG00000268953 | 19 | 31630362 | 31650757 | 56 | 14 | 45917 | -0.6892 | 0.75465 | *AC020952.1* |
| ENSG00000204022 | 10 | 90336510 | 90376733 | 49 | 14 | 48788 | -0.68935 | 0.7547 | *LIPJ* |
| ENSG00000105552 | 19 | 49288319 | 49324286 | 137 | 26 | 42447 | -0.68939 | 0.75471 | *BCAT2* |
| ENSG00000108448 | 17 | 18591311 | 18649431 | 139 | 22 | 41039 | -0.68956 | 0.75476 | *TRIM16L* |
| ENSG00000206530 | 3 | 112995777 | 113170457 | 700 | 47 | 50249 | -0.68986 | 0.75486 | *WDR52* |
| ENSG00000163631 | 4 | 74252831 | 74297129 | 105 | 13 | 54608 | -0.68997 | 0.75489 | *ALB* |
| ENSG00000188784 | 1 | 20236502 | 20260110 | 93 | 22 | 48127 | -0.69017 | 0.75496 | *PLA2G2E* |
| ENSG00000167196 | 15 | 76186200 | 76237609 | 113 | 14 | 39940 | -0.69033 | 0.75501 | *FBXO22* |
| ENSG00000159111 | 17 | 45890638 | 45918900 | 110 | 16 | 32226 | -0.69049 | 0.75506 | *MRPL10* |
| ENSG00000182534 | 17 | 74658633 | 74717098 | 194 | 23 | 40784 | -0.69071 | 0.75513 | *MXRA7* |
| ENSG00000177697 | 11 | 822843 | 849831 | 125 | 14 | 30175 | -0.69079 | 0.75515 | *CD151* |
| ENSG00000134057 | 5 | 68452837 | 68484072 | 112 | 10 | 55075 | -0.69088 | 0.75518 | *CCNB1* |
| ENSG00000120235 | 9 | 21339834 | 21361377 | 76 | 13 | 55011 | -0.69155 | 0.75539 | *IFNA6* |
| ENSG00000076053 | 11 | 114260752 | 114294925 | 111 | 14 | 49955 | -0.69155 | 0.75539 | *RBM7* |
| ENSG00000171763 | 15 | 45684529 | 45723617 | 62 | 10 | 31152 | -0.69154 | 0.75539 | *SPATA5L1* |
| ENSG00000168172 | 8 | 42742075 | 42895682 | 354 | 28 | 49585 | -0.69197 | 0.75552 | *HOOK3* |
| ENSG00000183979 | 17 | 79848621 | 79870781 | 20 | 12 | 50912 | -0.69195 | 0.75552 | *NPB* |
| ENSG00000100764 | 14 | 90712839 | 90748968 | 155 | 8 | 49720 | -0.69217 | 0.75559 | *PSMC1* |
| ENSG00000070010 | 22 | 19427433 | 19476738 | 134 | 23 | 42600 | -0.69336 | 0.75596 | *UFD1L* |
| ENSG00000120437 | 6 | 160171360 | 160210144 | 141 | 17 | 46713 | -0.69339 | 0.75597 | *ACAT2* |

| | | | | | | | | | |
|---|---|---|---|---|---|---|---|---|---|
| ENSG00000108468 | 17 | 46137414 | 46188883 | 181 | 16 | 51394 | -0.69358 | 0.75603 | *CBX1* |
| ENSG00000167014 | 15 | 45238900 | 45281427 | 137 | 11 | 43756 | -0.69408 | 0.75619 | *C15orf43* |
| ENSG00000198182 | 19 | 38177264 | 38220691 | 105 | 17 | 36541 | -0.69434 | 0.75626 | *ZNF607* |
| ENSG00000165682 | 12 | 10128241 | 10176023 | 237 | 25 | 47254 | -0.69475 | 0.75639 | *CLEC1B* |
| ENSG00000188659 | 15 | 82545151 | 82587271 | 97 | 18 | 43183 | -0.69475 | 0.75639 | *FAM154B* |
| ENSG00000154274 | 4 | 37445563 | 37635117 | 853 | 117 | 43102 | -0.69485 | 0.75643 | *C4orf19* |
| ENSG00000079557 | 4 | 74337400 | 74379691 | 128 | 17 | 42233 | -0.69487 | 0.75643 | *AFM* |
| ENSG00000135317 | 6 | 86205214 | 86313874 | 203 | 27 | 36332 | -0.69538 | 0.75659 | *SNX14* |
| ENSG00000114978 | 2 | 74369655 | 74416025 | 102 | 12 | 33076 | -0.69581 | 0.75673 | *MOB1A* |
| ENSG00000149305 | 11 | 113765399 | 113827287 | 197 | 32 | 51503 | -0.69591 | 0.75676 | *HTR3B* |
| ENSG00000152127 | 2 | 134867554 | 135222192 | 1094 | 144 | 36170 | -0.69601 | 0.75679 | *MGAT5* |
| ENSG00000142408 | 19 | 54456294 | 54503469 | 162 | 34 | 48423 | -0.69603 | 0.7568 | *CACNG8* |
| ENSG00000101224 | 20 | 3757578 | 3796762 | 148 | 25 | 41052 | -0.6961 | 0.75682 | *CDC25B* |
| ENSG00000107562 | 10 | 44783038 | 44891941 | 466 | 71 | 45885 | -0.69621 | 0.75685 | *CXCL12* |
| ENSG00000105492 | 19 | 52012779 | 52045110 | 198 | 23 | 55278 | -0.69625 | 0.75686 | *SIGLEC6* |
| ENSG00000100731 | 14 | 71364122 | 71592099 | 703 | 48 | 47224 | -0.69643 | 0.75692 | *PCNX* |
| ENSG00000136695 | 2 | 113806215 | 113832959 | 155 | 18 | 39178 | -0.69654 | 0.75695 | *IL36RN* |
| ENSG00000148400 | 9 | 139378896 | 139450314 | 230 | 52 | 40120 | -0.69681 | 0.75704 | *NOTCH1* |
| ENSG00000150165 | 10 | 47147983 | 47184093 | 4 | 2 | 45942 | -0.69718 | 0.75715 | *ANXA8L1* |
| ENSG00000183475 | 15 | 101132739 | 101201910 | 221 | 45 | 43430 | -0.69716 | 0.75715 | *ASB7* |
| ENSG00000133101 | 13 | 36995967 | 37027019 | 72 | 17 | 55976 | -0.69745 | 0.75724 | *CCNA1* |
| ENSG00000110048 | 11 | 59331871 | 59393617 | 134 | 16 | 37580 | -0.69756 | 0.75727 | *OSBP* |
| ENSG00000162174 | 11 | 62094920 | 62170882 | 249 | 22 | 47141 | -0.69795 | 0.7574 | *ASRGL1* |
| ENSG00000106038 | 7 | 27272164 | 27300112 | 53 | 13 | 42165 | -0.69844 | 0.75755 | *EVX1* |
| ENSG00000137094 | 9 | 34979638 | 35008897 | 45 | 14 | 43095 | -0.69902 | 0.75773 | *DNAJB5* |
| ENSG00000125656 | 19 | 6351463 | 6378919 | 135 | 22 | 37173 | -0.70016 | 0.75809 | *CLPP* |
| ENSG00000180264 | 9 | 127203423 | 127249379 | 114 | 19 | 41451 | -0.70067 | 0.75824 | *GPR144* |
| ENSG00000109705 | 4 | 13532454 | 13556674 | 70 | 17 | 45022 | -0.70132 | 0.75845 | *NKX3-2* |
| ENSG00000135686 | 16 | 84672131 | 84711292 | 180 | 32 | 44230 | -0.70149 | 0.7585 | *KLHL36* |
| ENSG00000100985 | 20 | 44627547 | 44655200 | 99 | 17 | 44355 | -0.70163 | 0.75855 | *MMP9* |
| ENSG00000196565 | 11 | 5264420 | 5677019 | 2583 | 166 | 42076 | -0.70169 | 0.75856 | *HBG2* |
| ENSG00000073614 | 12 | 379295 | 508620 | 380 | 31 | 46236 | -0.70176 | 0.75859 | *KDM5A* |
| ENSG00000184060 | 17 | 29223362 | 29296340 | 103 | 14 | 31663 | -0.70184 | 0.75861 | *ADAP2* |
| ENSG00000104381 | 8 | 75223365 | 75411107 | 469 | 52 | 47073 | -0.70187 | 0.75862 | *GDAP1* |
| ENSG00000119559 | 19 | 1451142 | 1489555 | 99 | 34 | 40804 | -0.70195 | 0.75865 | *C19orf25* |
| ENSG00000163563 | 1 | 158791107 | 158829296 | 115 | 19 | 49939 | -0.70218 | 0.75872 | *MNDA* |

| | | | | | | | | | |
|---|---|---|---|---|---|---|---|---|---|
| ENSG00000152254 | 2 | 169747750 | 169776505 | 74 | 18 | 45590 | -0.70226 | 0.75874 | *G6PC2* |
| ENSG00000128567 | 7 | 131175021 | 131252976 | 230 | 56 | 48647 | -0.70231 | 0.75876 | *PODXL* |
| ENSG00000204003 | 9 | 139622619 | 139652905 | 131 | 17 | 45020 | -0.70246 | 0.7588 | *LCN6* |
| ENSG00000185507 | 11 | 602553 | 625999 | 104 | 15 | 41935 | -0.70255 | 0.75883 | *IRF7* |
| ENSG00000128050 | 4 | 57291907 | 57337534 | 149 | 20 | 41921 | -0.70295 | 0.75896 | *PAICS* |
| ENSG00000089063 | 20 | 5070486 | 5103749 | 76 | 11 | 27209 | -0.70297 | 0.75896 | *TMEM230* |
| ENSG00000101210 | 20 | 62109366 | 62140505 | 171 | 29 | 32982 | -0.7034 | 0.7591 | *EEF1A2* |
| ENSG00000256515 | 17 | 34512268 | 34534157 | 4 | 2 | 43471 | -0.70345 | 0.75911 | *CCL3L3* |
| ENSG00000141441 | 18 | 29694840 | 30060447 | 1100 | 101 | 43065 | -0.70371 | 0.75919 | *GAREM* |
| ENSG00000112599 | 6 | 42142139 | 42172654 | 118 | 9 | 33401 | -0.704 | 0.75928 | *GUCA1B* |
| ENSG00000214595 | 2 | 54940636 | 55209157 | 976 | 131 | 46214 | -0.70414 | 0.75933 | *EML6* |
| ENSG00000163812 | 3 | 44946749 | 45027677 | 134 | 14 | 33417 | -0.70419 | 0.75934 | *ZDHHC3* |
| ENSG00000221880 | 17 | 39180142 | 39201107 | 96 | 12 | 37495 | -0.70422 | 0.75935 | *KRTAP1-3* |
| ENSG00000105467 | 19 | 48857652 | 48889633 | 89 | 20 | 52935 | -0.70457 | 0.75946 | *SYNGR4* |
| ENSG00000125633 | 2 | 118663054 | 118781709 | 449 | 35 | 55870 | -0.7051 | 0.75963 | *CCDC93* |
| ENSG00000109534 | 4 | 110726666 | 110755893 | 82 | 13 | 35301 | -0.70532 | 0.7597 | *GAR1* |
| ENSG00000244187 | 9 | 139675807 | 139697709 | 98 | 10 | 53008 | -0.70562 | 0.75979 | *TMEM141* |
| ENSG00000068878 | 2 | 54081204 | 54207977 | 419 | 36 | 48045 | -0.70599 | 0.7599 | *PSME4* |
| ENSG00000135697 | 16 | 81262053 | 81334747 | 262 | 37 | 43998 | -0.70622 | 0.75997 | *BCMO1* |
| ENSG00000172007 | 4 | 140364386 | 140407763 | 120 | 16 | 52185 | -0.70629 | 0.76 | *RAB33B* |
| ENSG00000145332 | 4 | 88071255 | 88171466 | 237 | 33 | 48409 | -0.70638 | 0.76002 | *KLHL8* |
| ENSG00000154832 | 18 | 47798713 | 47824674 | 58 | 17 | 50898 | -0.70664 | 0.7601 | *CXXC1* |
| ENSG00000177023 | 8 | 7317830 | 7342604 | 2 | 1 | 42813 | -0.70667 | 0.76012 | *DEFB104B* |
| ENSG00000187082 | 8 | 7330026 | 7353922 | 2 | 1 | 42813 | -0.70667 | 0.76012 | *DEFB106B* |
| ENSG00000187609 | 9 | 140191348 | 140327714 | 397 | 90 | 42758 | -0.7069 | 0.76018 | *EXD3* |
| ENSG00000111639 | 12 | 6591150 | 6613007 | 110 | 8 | 36241 | -0.70767 | 0.76043 | *MRPL51* |
| ENSG00000220891 | 22 | 22891750 | 22919007 | 127 | 11 | 32521 | -0.70775 | 0.76045 | *LL22NC03-63E9.3* |
| ENSG00000198443 | 17 | 39330354 | 39351594 | 99 | 6 | 55222 | -0.70783 | 0.76047 | *KRTAP4-1* |
| ENSG00000122884 | 10 | 74756975 | 74866732 | 216 | 8 | 21939 | -0.70786 | 0.76048 | *P4HA1* |
| ENSG00000138685 | 4 | 123737863 | 123829391 | 400 | 55 | 36143 | -0.70808 | 0.76055 | *FGF2* |
| ENSG00000075886 | 2 | 132223666 | 132250507 | 136 | 9 | 22436 | -0.70813 | 0.76057 | *TUBA3D* |
| ENSG00000134644 | 1 | 31394353 | 31548838 | 350 | 30 | 35301 | -0.70821 | 0.76059 | *PUM1* |
| ENSG00000158477 | 1 | 158213927 | 158238059 | 70 | 17 | 34969 | -0.70822 | 0.7606 | *CD1A* |
| ENSG00000234560 | 11 | 123890297 | 123911341 | 71 | 15 | 41928 | -0.70826 | 0.76061 | *OR10G8* |
| ENSG00000142539 | 19 | 50909056 | 50941666 | 83 | 27 | 42299 | -0.70829 | 0.76062 | *CTD-2545M3.6* |
| ENSG00000175395 | 10 | 38228500 | 38275561 | 129 | 15 | 56806 | -0.70838 | 0.76065 | *ZNF25* |

| | | | | | | | | | |
|---|---|---|---|---|---|---|---|---|---|
| ENSG00000268324 | 3 | 46588888 | 46611178 | 130 | 15 | 47443 | -0.70867 | 0.76074 | *LUZPP1* |
| ENSG00000107290 | 9 | 135126743 | 135240372 | 405 | 32 | 28585 | -0.70876 | 0.76076 | *SETX* |
| ENSG00000171766 | 15 | 45643322 | 45704525 | 131 | 8 | 26991 | -0.70878 | 0.76077 | *GATM* |
| ENSG00000164713 | 7 | 97871691 | 97947162 | 232 | 20 | 34466 | -0.70924 | 0.76091 | *BRI3* |
| ENSG00000101049 | 20 | 42177608 | 42226877 | 160 | 36 | 49756 | -0.70923 | 0.76091 | *SGK2* |
| ENSG00000154945 | 17 | 48760551 | 48795285 | 73 | 15 | 51075 | -0.70991 | 0.76112 | *ANKRD40* |
| ENSG00000256018 | 6 | 26261146 | 26281612 | 73 | 13 | 40373 | -0.71033 | 0.76125 | *HIST1H3G* |
| ENSG00000225526 | 3 | 12546433 | 12612558 | 230 | 22 | 36252 | -0.71082 | 0.7614 | *C3orf83* |
| ENSG00000163739 | 4 | 74725110 | 74746959 | 92 | 15 | 32934 | -0.7108 | 0.7614 | *CXCL1* |
| ENSG00000130957 | 9 | 97311002 | 97366075 | 310 | 30 | 47465 | -0.7111 | 0.76149 | *FBP2* |
| ENSG00000107736 | 10 | 73146691 | 73585702 | 1410 | 201 | 47073 | -0.7116 | 0.76164 | *CDH23* |
| ENSG00000219607 | 6 | 5075720 | 5097455 | 67 | 16 | 36400 | -0.71166 | 0.76166 | *PPP1R3G* |
| ENSG00000169605 | 2 | 69191705 | 69218106 | 85 | 14 | 29360 | -0.7123 | 0.76186 | *GKN1* |
| ENSG00000144407 | 2 | 209214438 | 209729227 | 1762 | 124 | 41622 | -0.7124 | 0.76189 | *PTH2R* |
| ENSG00000269210 | 2 | 39176764 | 39197483 | 48 | 13 | 44505 | -0.71329 | 0.76217 | *AC019171.1* |
| ENSG00000136878 | 9 | 132586977 | 132654107 | 506 | 27 | 46603 | -0.71428 | 0.76247 | *USP20* |
| ENSG00000135316 | 6 | 86308053 | 86363510 | 99 | 20 | 33711 | -0.71471 | 0.76261 | *SYNCRIP* |
| ENSG00000165733 | 10 | 43268249 | 43340385 | 264 | 25 | 40107 | -0.71484 | 0.76265 | *BMS1* |
| ENSG00000269215 | 5 | 39095358 | 39117350 | 55 | 11 | 51518 | -0.71515 | 0.76274 | *AC008964.1* |
| ENSG00000176396 | 19 | 40018890 | 40040870 | 71 | 13 | 54599 | -0.71526 | 0.76278 | *EID2* |
| ENSG00000182591 | 21 | 32242966 | 32263874 | 46 | 10 | 50274 | -0.71531 | 0.76279 | *KRTAP11-1* |
| ENSG00000268133 | 19 | 57864934 | 57919952 | 148 | 24 | 47147 | -0.71541 | 0.76282 | *AC003002.4* |
| ENSG00000102189 | 12 | 93154413 | 93333107 | 417 | 31 | 40973 | -0.71548 | 0.76284 | *EEA1* |
| ENSG00000203618 | 22 | 19700468 | 19722294 | 25 | 11 | 44161 | -0.71548 | 0.76284 | *GP1BB* |
| ENSG00000160014 | 19 | 47094331 | 47124050 | 96 | 23 | 41304 | -0.71603 | 0.76301 | *CALM3* |
| ENSG00000133083 | 13 | 36335478 | 36715443 | 1311 | 156 | 43500 | -0.71617 | 0.76306 | *DCLK1* |
| ENSG00000151651 | 10 | 135065907 | 135100372 | 132 | 16 | 46101 | -0.71643 | 0.76314 | *ADAM8* |
| ENSG00000152240 | 18 | 43674298 | 43718299 | 157 | 17 | 36808 | -0.71687 | 0.76327 | *HAUS1* |
| ENSG00000137265 | 6 | 381739 | 421447 | 143 | 30 | 41518 | -0.71711 | 0.76335 | *IRF4* |
| ENSG00000108798 | 17 | 47277589 | 47310587 | 102 | 24 | 47489 | -0.71723 | 0.76339 | *ABI3* |
| ENSG00000176055 | 5 | 89744020 | 89780585 | 68 | 14 | 52483 | -0.71741 | 0.76344 | *MBLAC2* |
| ENSG00000257446 | 19 | 12144620 | 12177127 | 70 | 11 | 26040 | -0.718 | 0.76362 | *ZNF878* |
| ENSG00000081923 | 18 | 55303658 | 55480333 | 606 | 94 | 47482 | -0.71854 | 0.76379 | *ATP8B1* |
| ENSG00000163216 | 1 | 153002201 | 153024407 | 134 | 7 | 49165 | -0.7187 | 0.76384 | *SPRR2D* |
| ENSG00000128652 | 2 | 176991340 | 177047830 | 139 | 20 | 39840 | -0.71896 | 0.76392 | *HOXD3* |
| ENSG00000198203 | 2 | 108895095 | 108936371 | 126 | 30 | 48922 | -0.71909 | 0.76396 | *SULT1C2* |

| | | | | | | | | | |
|---|---|---|---|---|---|---|---|---|---|
| ENSG00000198039 | 7 | 64320550 | 64401344 | 361 | 22 | 48721 | -0.71918 | 0.76398 | *ZNF273* |
| ENSG00000157884 | 2 | 26794070 | 26874236 | 208 | 38 | 40138 | -0.71941 | 0.76406 | *CIB4* |
| ENSG00000105379 | 19 | 51838423 | 51879672 | 140 | 25 | 36988 | -0.71948 | 0.76408 | *ETFB* |
| ENSG00000188868 | 19 | 12418291 | 12454534 | 69 | 8 | 52849 | -0.71978 | 0.76417 | *ZNF563* |
| ENSG00000146576 | 7 | 6619648 | 6658357 | 137 | 18 | 48258 | -0.71988 | 0.7642 | *C7orf26* |
| ENSG00000268341 | 19 | 45832445 | 45852639 | 67 | 15 | 40008 | -0.72008 | 0.76426 | *L47234.1* |
| ENSG00000188100 | 10 | 88770063 | 88794487 | 110 | 26 | 47350 | -0.72014 | 0.76428 | *FAM25A* |
| ENSG00000062598 | 20 | 44984688 | 45071704 | 100 | 13 | 35749 | -0.72012 | 0.76428 | *ELMO2* |
| ENSG00000140873 | 16 | 77271710 | 77479011 | 1188 | 95 | 45585 | -0.72051 | 0.76439 | *ADAMTS18* |
| ENSG00000187950 | 12 | 29555407 | 29660619 | 460 | 47 | 43094 | -0.72083 | 0.76449 | *OVCH1* |
| ENSG00000166323 | 11 | 108169246 | 108348258 | 404 | 24 | 49933 | -0.72159 | 0.76473 | *C11orf65* |
| ENSG00000100592 | 14 | 59645364 | 59848123 | 688 | 61 | 48983 | -0.72183 | 0.7648 | *DAAM1* |
| ENSG00000198743 | 21 | 35435870 | 35488559 | 74 | 21 | 51574 | -0.72185 | 0.76481 | *SLC5A3* |
| ENSG00000272962 | 21 | 35435870 | 35488561 | 74 | 21 | 51574 | -0.72185 | 0.76481 | *SLC5A3* |
| ENSG00000176659 | 20 | 58620980 | 58658008 | 78 | 21 | 41726 | -0.72194 | 0.76483 | *C20orf197* |
| ENSG00000180592 | 10 | 21792407 | 21824611 | 32 | 11 | 50834 | -0.72197 | 0.76484 | *SKIDA1* |
| ENSG00000128713 | 2 | 176958944 | 176984722 | 60 | 14 | 39764 | -0.72203 | 0.76486 | *HOXD11* |
| ENSG00000125611 | 2 | 113331817 | 113356852 | 65 | 15 | 43954 | -0.72227 | 0.76494 | *CHCHD5* |
| ENSG00000147813 | 8 | 144646955 | 144670819 | 112 | 19 | 44979 | -0.72242 | 0.76498 | *NAPRT1* |
| ENSG00000119900 | 6 | 71988506 | 72028653 | 183 | 23 | 45142 | -0.72266 | 0.76506 | *OGFRL1* |
| ENSG00000165078 | 8 | 68324360 | 68668620 | 969 | 119 | 41535 | -0.72325 | 0.76524 | *CPA6* |
| ENSG00000267881 | 19 | 42202531 | 42275190 | 214 | 32 | 43649 | -0.7233 | 0.76525 | *CEA* |
| ENSG00000225830 | 10 | 50653414 | 50757584 | 249 | 27 | 38963 | -0.72386 | 0.76542 | *ERCC6* |
| ENSG00000227011 | 17 | 51052880 | 51075012 | 56 | 15 | 39764 | -0.72399 | 0.76546 | *C17orf112* |
| ENSG00000232056 | 2 | 10977703 | 11005081 | 157 | 23 | 40343 | -0.7243 | 0.76556 | *AC092687.4* |
| ENSG00000197417 | 17 | 3501556 | 3549616 | 196 | 32 | 32283 | -0.72459 | 0.76565 | *SHPK* |
| ENSG00000228300 | 19 | 1265026 | 1289248 | 34 | 10 | 39740 | -0.72468 | 0.76568 | *C19orf24* |
| ENSG00000180233 | 7 | 30313923 | 30462118 | 270 | 50 | 37628 | -0.72527 | 0.76586 | *ZNRF2* |
| ENSG00000183560 | 11 | 94028759 | 94050887 | 76 | 21 | 40620 | -0.72545 | 0.76591 | *FOLR4* |
| ENSG00000205476 | 14 | 99967603 | 100080363 | 452 | 40 | 39998 | -0.72573 | 0.766 | *CCDC85C* |
| ENSG00000090565 | 16 | 465619 | 583011 | 331 | 40 | 41016 | -0.72581 | 0.76602 | *RAB11FIP3* |
| ENSG00000097033 | 1 | 87160259 | 87223867 | 160 | 26 | 44459 | -0.72587 | 0.76604 | *SH3GLB1* |
| ENSG00000166396 | 18 | 61410169 | 61482604 | 254 | 33 | 41515 | -0.7259 | 0.76605 | *SERPINB7* |
| ENSG00000076984 | 19 | 7958728 | 7989363 | 99 | 22 | 43287 | -0.7261 | 0.76611 | *MAP2K7* |
| ENSG00000156642 | 15 | 73842355 | 73936475 | 202 | 26 | 49920 | -0.72645 | 0.76622 | *NPTN* |
| ENSG00000186732 | 22 | 43797202 | 43913728 | 481 | 81 | 39601 | -0.72647 | 0.76622 | *MPPED1* |

| | | | | | | | | | |
|---|---|---|---|---|---|---|---|---|---|
| ENSG00000164609 | 5 | 159818648 | 159858718 | 113 | 20 | 51017 | -0.72661 | 0.76627 | *SLU7* |
| ENSG00000170234 | 5 | 159478808 | 159556430 | 185 | 19 | 54452 | -0.72709 | 0.76641 | *PWWP2A* |
| ENSG00000269766 | 11 | 35140091 | 35160215 | 83 | 14 | 30908 | -0.72712 | 0.76642 | *AL356215.1* |
| ENSG00000188282 | 2 | 218889683 | 218965304 | 210 | 28 | 44050 | -0.72737 | 0.7665 | *RUFY4* |
| ENSG00000176454 | 15 | 34641106 | 34669479 | 103 | 29 | 48283 | -0.72766 | 0.76659 | *LPCAT4* |
| ENSG00000196267 | 19 | 52647117 | 52684896 | 117 | 28 | 44608 | -0.72771 | 0.7666 | *ZNF836* |
| ENSG00000152404 | 11 | 107187071 | 107338572 | 662 | 49 | 48586 | -0.728 | 0.76669 | *CWF19L2* |
| ENSG00000214042 | 9 | 21191468 | 21212204 | 150 | 16 | 49876 | -0.7285 | 0.76685 | *IFNA7* |
| ENSG00000196600 | 11 | 62921248 | 63007124 | 298 | 22 | 44840 | -0.72861 | 0.76688 | *SLC22A25* |
| ENSG00000123388 | 12 | 54356910 | 54381427 | 51 | 10 | 47045 | -0.72882 | 0.76694 | *HOXC11* |
| ENSG00000070731 | 17 | 74549792 | 74593038 | 180 | 28 | 43877 | -0.7288 | 0.76694 | *ST6GALNAC2* |
| ENSG00000197860 | 5 | 64951755 | 65028862 | 187 | 23 | 54523 | -0.72938 | 0.76711 | *SGTB* |
| ENSG00000136155 | 13 | 78099809 | 78229398 | 348 | 44 | 41865 | -0.72983 | 0.76725 | *SCEL* |
| ENSG00000204713 | 6 | 28860779 | 28901766 | 179 | 27 | 41001 | -0.73124 | 0.76768 | *TRIM27* |
| ENSG00000148450 | 10 | 23374435 | 23420942 | 102 | 20 | 48671 | -0.73151 | 0.76777 | *MSRB2* |
| ENSG00000206105 | 21 | 31982946 | 32003169 | 34 | 5 | 53467 | -0.73152 | 0.76777 | *KRTAP20-4* |
| ENSG00000039537 | 5 | 41132336 | 41271540 | 486 | 56 | 49356 | -0.73161 | 0.7678 | *C6* |
| ENSG00000095787 | 10 | 28811422 | 28922041 | 267 | 24 | 51492 | -0.73165 | 0.76781 | *WAC* |
| ENSG00000180423 | 11 | 46614411 | 46649459 | 29 | 7 | 50421 | -0.73173 | 0.76783 | *HARBI1* |
| ENSG00000148677 | 10 | 92661853 | 92691033 | 89 | 29 | 47620 | -0.73177 | 0.76784 | *ANKRD1* |
| ENSG00000169139 | 8 | 48910960 | 48987268 | 61 | 26 | 47536 | -0.73178 | 0.76785 | *UBE2V2* |
| ENSG00000066405 | 3 | 137707577 | 137762494 | 126 | 19 | 43594 | -0.73181 | 0.76786 | *CLDN18* |
| ENSG00000182793 | 6 | 52686453 | 52720893 | 107 | 12 | 45278 | -0.73192 | 0.76789 | *GSTA5* |
| ENSG00000123552 | 6 | 99870190 | 99979604 | 337 | 30 | 36470 | -0.7323 | 0.76801 | *USP45* |
| ENSG00000075275 | 22 | 46746731 | 46943067 | 907 | 126 | 43474 | -0.73233 | 0.76802 | *CELSR1* |
| ENSG00000144410 | 2 | 207794278 | 207844198 | 141 | 24 | 46177 | -0.73315 | 0.76827 | *CPO* |
| ENSG00000184148 | 1 | 152933142 | 152955050 | 36 | 9 | 47925 | -0.73342 | 0.76835 | *SPRR4* |
| ENSG00000021488 | 19 | 33311415 | 33370672 | 290 | 34 | 48034 | -0.73354 | 0.76838 | *SLC7A9* |
| ENSG00000204007 | 9 | 138505502 | 138541386 | 172 | 36 | 50124 | -0.73354 | 0.76839 | *GLT6D1* |
| ENSG00000111701 | 12 | 7791996 | 7828499 | 169 | 27 | 48526 | -0.73378 | 0.76846 | *APOBEC1* |
| ENSG00000141034 | 17 | 17932606 | 17981718 | 74 | 10 | 30234 | -0.73384 | 0.76848 | *GID4* |
| ENSG00000101115 | 20 | 50390581 | 50429059 | 189 | 33 | 49054 | -0.73402 | 0.76853 | *SALL4* |
| ENSG00000111678 | 12 | 7042141 | 7065166 | 40 | 18 | 42153 | -0.73418 | 0.76858 | *C12orf57* |
| ENSG00000187733 | 1 | 104283028 | 104311312 | 19 | 7 | 54399 | -0.73534 | 0.76893 | *AMY1C* |
| ENSG00000115488 | 2 | 233887382 | 233909767 | 76 | 20 | 45286 | -0.73579 | 0.76907 | *NEU2* |
| ENSG00000177551 | 1 | 116368998 | 116396538 | 106 | 15 | 45784 | -0.73593 | 0.76911 | *NHLH2* |

| | | | | | | | | | |
|---|---|---|---|---|---|---|---|---|---|
| ENSG00000166984 | 6 | 167549902 | 167620401 | 284 | 49 | 50820 | -0.73623 | 0.7692 | *TCP10L2* |
| ENSG00000204775 | 8 | 145307003 | 145341153 | 4 | 2 | 41946 | -0.73622 | 0.7692 | *KM-PA-2* |
| ENSG00000187786 | 8 | 145311517 | 145333045 | 4 | 2 | 41946 | -0.73622 | 0.7692 | *SCXB* |
| ENSG00000162819 | 1 | 222875895 | 222918538 | 76 | 17 | 45290 | -0.73627 | 0.76922 | *BROX* |
| ENSG00000100280 | 22 | 29713669 | 29829168 | 309 | 46 | 39509 | -0.73629 | 0.76922 | *AP1B1* |
| ENSG00000138698 | 4 | 99172535 | 99375012 | 529 | 29 | 33524 | -0.73672 | 0.76935 | *RAP1GDS1* |
| ENSG00000256060 | 19 | 57864879 | 57886721 | 68 | 14 | 49177 | -0.73686 | 0.7694 | *TRAPPC2P1* |
| ENSG00000116791 | 1 | 75161170 | 75209092 | 137 | 26 | 45484 | -0.73709 | 0.76947 | *CRYZ* |
| ENSG00000158488 | 1 | 158313254 | 158337343 | 67 | 15 | 39390 | -0.73728 | 0.76952 | *CD1E* |
| ENSG00000170776 | 15 | 85913802 | 86302586 | 1513 | 81 | 39416 | -0.7375 | 0.76959 | *AKAP13* |
| ENSG00000187626 | 6 | 28202401 | 28237011 | 80 | 14 | 44511 | -0.73769 | 0.76965 | *ZKSCAN4* |
| ENSG00000117419 | 1 | 44676742 | 44830932 | 253 | 30 | 47858 | -0.73793 | 0.76972 | *ERI3* |
| ENSG00000176200 | 11 | 59261049 | 59281984 | 47 | 8 | 53833 | -0.7387 | 0.76996 | *OR4D11* |
| ENSG00000128218 | 22 | 24084930 | 24106655 | 74 | 17 | 43943 | -0.73874 | 0.76997 | *VPREB3* |
| ENSG00000204612 | 9 | 79624571 | 79645869 | 72 | 21 | 46995 | -0.73878 | 0.76998 | *FOXB2* |
| ENSG00000026751 | 1 | 160699037 | 160734611 | 74 | 15 | 45104 | -0.73948 | 0.77019 | *SLAMF7* |
| ENSG00000102970 | 16 | 57428679 | 57459974 | 137 | 27 | 45313 | -0.73977 | 0.77028 | *CCL17* |
| ENSG00000149488 | 20 | 2507253 | 2632430 | 587 | 58 | 49203 | -0.73988 | 0.77031 | *TMC2* |
| ENSG00000005100 | 17 | 5334232 | 5382380 | 153 | 34 | 40729 | -0.74031 | 0.77044 | *DHX33* |
| ENSG00000131910 | 1 | 27227980 | 27250457 | 50 | 10 | 54875 | -0.74057 | 0.77052 | *NR0B2* |
| ENSG00000143756 | 1 | 224291789 | 224359749 | 198 | 17 | 48598 | -0.74077 | 0.77058 | *FBXO28* |
| ENSG00000077463 | 19 | 4164106 | 4192601 | 83 | 15 | 42282 | -0.74103 | 0.77066 | *SIRT6* |
| ENSG00000159173 | 1 | 201363625 | 201408994 | 114 | 35 | 41316 | -0.74114 | 0.7707 | *TNNI1* |
| ENSG00000075043 | 20 | 62027542 | 62113993 | 356 | 56 | 39766 | -0.74126 | 0.77073 | *KCNQ2* |
| ENSG00000136875 | 9 | 116027623 | 116065185 | 125 | 19 | 42075 | -0.74132 | 0.77075 | *PRPF4* |
| ENSG00000107447 | 10 | 98054085 | 98108321 | 146 | 15 | 45563 | -0.74136 | 0.77076 | *DNTT* |
| ENSG00000239998 | 19 | 55074387 | 55109027 | 198 | 21 | 40891 | -0.74157 | 0.77083 | *LILRA2* |
| ENSG00000143819 | 1 | 225987794 | 226043260 | 251 | 52 | 47508 | -0.7417 | 0.77086 | *EPHX1* |
| ENSG00000196136 | 14 | 95068714 | 95100392 | 110 | 33 | 46961 | -0.74239 | 0.77108 | *SERPINA3* |
| ENSG00000110680 | 11 | 14978214 | 15003900 | 45 | 6 | 50740 | -0.74268 | 0.77116 | *CALCA* |
| ENSG00000178397 | 7 | 6359040 | 6398612 | 172 | 34 | 40111 | -0.74285 | 0.77121 | *FAM220A* |
| ENSG00000076944 | 19 | 7691767 | 7722759 | 102 | 16 | 51563 | -0.74285 | 0.77121 | *STXBP2* |
| ENSG00000138030 | 2 | 27299615 | 27333640 | 44 | 9 | 47369 | -0.74285 | 0.77122 | *KHK* |
| ENSG00000125780 | 20 | 2266647 | 2331724 | 136 | 38 | 47665 | -0.74321 | 0.77132 | *TGM3* |
| ENSG00000112305 | 6 | 71367479 | 71581718 | 536 | 37 | 52700 | -0.74324 | 0.77133 | *SMAP1* |
| ENSG00000163687 | 3 | 58167984 | 58210424 | 164 | 26 | 48499 | -0.74337 | 0.77137 | *DNASE1L3* |

| | | | | | | | | | |
|---|---|---|---|---|---|---|---|---|---|
| ENSG00000136897 | 9 | 104139915 | 104170896 | 92 | 21 | 48801 | -0.74338 | 0.77137 | *MRPL50* |
| ENSG00000090920 | 19 | 40343963 | 40450533 | 289 | 53 | 46187 | -0.74341 | 0.77138 | *FCGBP* |
| ENSG00000050165 | 11 | 11974653 | 12041316 | 254 | 64 | 45426 | -0.74342 | 0.77139 | *DKK3* |
| ENSG00000104635 | 8 | 22214762 | 22301642 | 327 | 31 | 38166 | -0.74398 | 0.77156 | *SLC39A14* |
| ENSG00000153044 | 5 | 68475375 | 68516184 | 120 | 11 | 54748 | -0.74412 | 0.7716 | *CENPH* |
| ENSG00000163412 | 3 | 71714567 | 71813924 | 325 | 52 | 45308 | -0.74454 | 0.77173 | *EIF4E3* |
| ENSG00000004799 | 7 | 95202811 | 95235803 | 126 | 15 | 35236 | -0.74456 | 0.77173 | *PDK4* |
| ENSG00000072042 | 14 | 68133518 | 68172531 | 101 | 17 | 38036 | -0.74455 | 0.77173 | *RDH11* |
| ENSG00000100427 | 22 | 50487820 | 50534331 | 269 | 23 | 51743 | -0.74461 | 0.77175 | *MLC1* |
| ENSG00000127863 | 13 | 24134509 | 24260232 | 473 | 64 | 51477 | -0.7447 | 0.77177 | *TNFRSF19* |
| ENSG00000027001 | 13 | 24294328 | 24473558 | 575 | 50 | 46570 | -0.74496 | 0.77185 | *MIPEP* |
| ENSG00000155096 | 8 | 103828585 | 103916092 | 266 | 23 | 54474 | -0.74499 | 0.77186 | *AZIN1* |
| ENSG00000124467 | 19 | 43246838 | 43369843 | 605 | 53 | 26539 | -0.74506 | 0.77188 | *PSG8* |
| ENSG00000177600 | 11 | 799647 | 822880 | 64 | 14 | 41994 | -0.74525 | 0.77194 | *RPLP2* |
| ENSG00000213614 | 15 | 72625775 | 72678817 | 83 | 18 | 43929 | -0.74528 | 0.77195 | *HEXA* |
| ENSG00000101474 | 20 | 24933561 | 24983615 | 201 | 16 | 54110 | -0.74538 | 0.77198 | *APMAP* |
| ENSG00000186765 | 17 | 79485422 | 79514156 | 116 | 30 | 43050 | -0.74586 | 0.77212 | *FSCN2* |
| ENSG00000198105 | 10 | 38081751 | 38157034 | 309 | 17 | 56138 | -0.74641 | 0.77229 | *ZNF248* |
| ENSG00000164010 | 1 | 43272795 | 43320660 | 96 | 21 | 52438 | -0.74648 | 0.77231 | *ERMAP* |
| ENSG00000182968 | 13 | 112711913 | 112736020 | 35 | 13 | 37036 | -0.7469 | 0.77244 | *SOX1* |
| ENSG00000141622 | 18 | 43896772 | 44053103 | 420 | 82 | 48320 | -0.74771 | 0.77268 | *RNF165* |
| ENSG00000196526 | 4 | 7750441 | 7951653 | 1088 | 69 | 40578 | -0.748 | 0.77277 | *AFAP1* |
| ENSG00000168530 | 2 | 211144874 | 211189914 | 117 | 13 | 53765 | -0.74803 | 0.77278 | *MYL1* |
| ENSG00000002919 | 17 | 46170719 | 46210436 | 146 | 14 | 50357 | -0.74856 | 0.77294 | *SNX11* |
| ENSG00000128346 | 22 | 38329528 | 38359676 | 40 | 10 | 43212 | -0.74866 | 0.77297 | *C22orf23* |
| ENSG00000186106 | 8 | 101511980 | 101582012 | 190 | 31 | 47739 | -0.74875 | 0.773 | *ANKRD46* |
| ENSG00000121413 | 19 | 58585205 | 58639794 | 253 | 21 | 34345 | -0.74902 | 0.77308 | *ZSCAN18* |
| ENSG00000083812 | 19 | 58968423 | 58994945 | 46 | 12 | 47311 | -0.74904 | 0.77308 | *ZNF324* |
| ENSG00000166747 | 16 | 71752913 | 71853104 | 268 | 26 | 44018 | -0.74941 | 0.77319 | *AP1G1* |
| ENSG00000131981 | 14 | 55580828 | 55622126 | 108 | 14 | 47597 | -0.74942 | 0.7732 | *LGALS3* |
| ENSG00000082068 | 5 | 37369314 | 37763537 | 967 | 59 | 41389 | -0.74959 | 0.77325 | *WDR70* |
| ENSG00000269404 | 19 | 50912195 | 50944570 | 88 | 27 | 44075 | -0.75036 | 0.77348 | *SPIB* |
| ENSG00000148985 | 11 | 3808954 | 3857601 | 172 | 27 | 46263 | -0.75041 | 0.7735 | *PGAP2* |
| ENSG00000186838 | 19 | 39995753 | 40021326 | 70 | 21 | 51660 | -0.75088 | 0.77364 | *SELV* |
| ENSG00000171502 | 1 | 86184916 | 86632626 | 1842 | 136 | 47823 | -0.75138 | 0.77379 | *COL24A1* |
| ENSG00000099834 | 11 | 606565 | 636078 | 124 | 17 | 42091 | -0.75218 | 0.77403 | *CDHR5* |

| | | | | | | | | | |
|---|---|---|---|---|---|---|---|---|---|
| ENSG00000163444 | 1 | 202966514 | 203003976 | 86 | 19 | 44600 | -0.7527 | 0.77418 | *TMEM183A* |
| ENSG00000163285 | 4 | 46027786 | 46136098 | 382 | 27 | 39936 | -0.7529 | 0.77424 | *GABRG1* |
| ENSG00000258838 | 10 | 50713247 | 50757584 | 119 | 18 | 36503 | -0.75304 | 0.77429 | *ERCC6-PGBD3* |
| ENSG00000170419 | 7 | 54600018 | 54648773 | 181 | 25 | 40552 | -0.75346 | 0.77441 | *VSTM2A* |
| ENSG00000164136 | 4 | 142547752 | 142665140 | 270 | 29 | 53398 | -0.75367 | 0.77448 | *IL15* |
| ENSG00000169474 | 1 | 152946557 | 152968289 | 77 | 5 | 53088 | -0.75386 | 0.77453 | *SPRR1A* |
| ENSG00000123268 | 12 | 51147493 | 51224905 | 156 | 18 | 45255 | -0.75392 | 0.77455 | *ATF1* |
| ENSG00000171314 | 10 | 99175917 | 99203198 | 56 | 14 | 46879 | -0.75396 | 0.77456 | *PGAM1* |
| ENSG00000183137 | 6 | 109406313 | 109495135 | 134 | 16 | 51588 | -0.75432 | 0.77467 | *CEP57L1* |
| ENSG00000211445 | 5 | 150390124 | 150418554 | 88 | 19 | 32000 | -0.75439 | 0.77469 | *GPX3* |
| ENSG00000144597 | 3 | 15458862 | 15494120 | 101 | 20 | 37644 | -0.75442 | 0.7747 | *EAF1* |
| ENSG00000173406 | 1 | 57450451 | 59022406 | 5110 | 378 | 44889 | -0.75446 | 0.77471 | *DAB1* |
| ENSG00000115163 | 2 | 26977157 | 27033935 | 121 | 15 | 44324 | -0.75466 | 0.77477 | *CENPA* |
| ENSG00000115904 | 2 | 39198537 | 39361486 | 365 | 23 | 41406 | -0.75465 | 0.77477 | *SOS1* |
| ENSG00000244537 | 17 | 39323698 | 39344460 | 113 | 6 | 55100 | -0.75464 | 0.77477 | *KRTAP4-2* |
| ENSG00000188878 | 17 | 73895655 | 73947221 | 107 | 18 | 38157 | -0.75486 | 0.77483 | *FBF1* |
| ENSG00000054690 | 14 | 67990018 | 68066329 | 248 | 48 | 46310 | -0.755 | 0.77488 | *PLEKHH1* |
| ENSG00000162006 | 16 | 809428 | 843370 | 178 | 34 | 37084 | -0.75587 | 0.77514 | *MSLNL* |
| ENSG00000111790 | 12 | 27081316 | 27129583 | 144 | 33 | 52028 | -0.75598 | 0.77517 | *FGFR1OP2* |
| ENSG00000268552 | 1 | 28167085 | 28187255 | 14 | 7 | 34180 | -0.75616 | 0.77522 | *AL109927.1* |
| ENSG00000214189 | 19 | 12193078 | 12258050 | 129 | 21 | 38979 | -0.75615 | 0.77522 | *ZNF788* |
| ENSG00000270181 | 13 | 103449704 | 103534748 | 244 | 31 | 47947 | -0.75635 | 0.77528 | *BIVM-ERCC5* |
| ENSG00000179058 | 9 | 132364504 | 132393055 | 106 | 20 | 45316 | -0.75639 | 0.77529 | *C9orf50* |
| ENSG00000244624 | 21 | 31978750 | 31999003 | 41 | 6 | 53735 | -0.75647 | 0.77532 | *KRTAP20-1* |
| ENSG00000197265 | 8 | 30425835 | 30525768 | 246 | 26 | 55076 | -0.75659 | 0.77535 | *GTF2E2* |
| ENSG00000104998 | 19 | 14132560 | 14173743 | 118 | 23 | 36120 | -0.75666 | 0.77537 | *IL27RA* |
| ENSG00000177595 | 11 | 789179 | 819753 | 75 | 17 | 44242 | -0.75681 | 0.77542 | *PIDD* |
| ENSG00000178750 | 3 | 93723213 | 93757454 | 35 | 13 | 44798 | -0.75693 | 0.77545 | *STX19* |
| ENSG00000265096 | 17 | 77005291 | 77033737 | 56 | 16 | 42045 | -0.75694 | 0.77546 | *C1QTNF1-AS1* |
| ENSG00000164414 | 6 | 88170341 | 88232054 | 222 | 25 | 54312 | -0.75792 | 0.77575 | *SLC35A1* |
| ENSG00000258792 | 14 | 90085265 | 90119015 | 112 | 14 | 50082 | -0.7582 | 0.77584 | *RP11-944C7.1* |
| ENSG00000152433 | 19 | 57864845 | 57900933 | 108 | 20 | 46334 | -0.7583 | 0.77586 | *ZNF547* |
| ENSG00000235268 | 11 | 94748422 | 94770760 | 69 | 15 | 41824 | -0.75858 | 0.77595 | *KDM4E* |
| ENSG00000248469 | 5 | 175466681 | 175499024 | 12 | 7 | 41670 | -0.75914 | 0.77612 | *RP11-826N14.2* |
| ENSG00000049541 | 7 | 73635829 | 73678774 | 73 | 16 | 44917 | -0.75922 | 0.77614 | *RFC2* |
| ENSG00000186265 | 3 | 112172815 | 112228408 | 99 | 17 | 28477 | -0.75944 | 0.77621 | *BTLA* |

| | | | | | | | | | |
|---|---|---|---|---|---|---|---|---|---|
| ENSG00000181031 | 17 | 52293 | 246045 | 894 | 193 | 42837 | -0.75948 | 0.77622 | *RPH3AL* |
| ENSG00000176641 | 18 | 59465296 | 59571480 | 450 | 52 | 52266 | -0.75956 | 0.77624 | *RNF152* |
| ENSG00000173431 | 14 | 21515981 | 21536614 | 59 | 17 | 45854 | -0.76009 | 0.7764 | *RNASE8* |
| ENSG00000075234 | 22 | 46653858 | 46699905 | 175 | 15 | 29453 | -0.7603 | 0.77646 | *TTC38* |
| ENSG00000232196 | 16 | 3411053 | 3432283 | 93 | 10 | 54512 | -0.76101 | 0.77667 | *MTRNR2L4* |
| ENSG00000143845 | 1 | 204090190 | 204131307 | 121 | 36 | 47401 | -0.76103 | 0.77668 | *ETNK2* |
| ENSG00000151849 | 13 | 25447171 | 25507018 | 219 | 29 | 52421 | -0.76117 | 0.77672 | *CENPJ* |
| ENSG00000184724 | 21 | 31975750 | 31996249 | 38 | 5 | 54600 | -0.76123 | 0.77674 | *KRTAP6-1* |
| ENSG00000134107 | 3 | 5010801 | 5037008 | 88 | 18 | 47500 | -0.76137 | 0.77678 | *BHLHE40* |
| ENSG00000161896 | 6 | 33679444 | 33724762 | 211 | 27 | 41190 | -0.76147 | 0.77681 | *IP6K3* |
| ENSG00000213901 | 2 | 219930051 | 220045549 | 227 | 20 | 37843 | -0.76159 | 0.77685 | *SLC23A3* |
| ENSG00000243251 | 10 | 50713151 | 50757086 | 118 | 18 | 36702 | -0.76173 | 0.77689 | *PGBD3* |
| ENSG00000141574 | 17 | 80268900 | 80301950 | 127 | 17 | 51499 | -0.76191 | 0.77694 | *SECTM1* |
| ENSG00000101104 | 20 | 43528703 | 43597676 | 140 | 25 | 51153 | -0.76199 | 0.77697 | *PABPC1L* |
| ENSG00000008294 | 17 | 49029535 | 49208226 | 276 | 29 | 44262 | -0.76218 | 0.77702 | *SPAG9* |
| ENSG00000214518 | 17 | 39200750 | 39221482 | 99 | 13 | 28452 | -0.7623 | 0.77706 | *KRTAP2-2* |
| ENSG00000076043 | 11 | 114300108 | 114331001 | 111 | 20 | 45484 | -0.76254 | 0.77713 | *REXO2* |
| ENSG00000009790 | 1 | 209919377 | 209965668 | 122 | 15 | 38817 | -0.76288 | 0.77723 | *TRAF3IP3* |
| ENSG00000068120 | 17 | 40703485 | 40728295 | 60 | 9 | 54383 | -0.76296 | 0.77725 | *COASY* |
| ENSG00000077984 | 20 | 24919866 | 24950564 | 192 | 14 | 54361 | -0.76294 | 0.77725 | *CST7* |
| ENSG00000188257 | 1 | 20291925 | 20316932 | 93 | 22 | 48864 | -0.76314 | 0.77731 | *PLA2G2A* |
| ENSG00000152578 | 11 | 105470721 | 105862819 | 926 | 85 | 50685 | -0.7632 | 0.77733 | *GRIA4* |
| ENSG00000070718 | 8 | 42000464 | 42039191 | 70 | 11 | 51798 | -0.76334 | 0.77737 | *AP3M2* |
| ENSG00000112319 | 6 | 133551736 | 133863258 | 984 | 107 | 41377 | -0.76338 | 0.77738 | *EYA4* |
| ENSG00000198198 | 1 | 43845553 | 43928321 | 133 | 15 | 37131 | -0.76362 | 0.77745 | *SZT2* |
| ENSG00000198551 | 19 | 11660189 | 11739950 | 265 | 33 | 48933 | -0.76369 | 0.77747 | *ZNF627* |
| ENSG00000158683 | 7 | 47804250 | 47998088 | 812 | 82 | 44357 | -0.76375 | 0.77749 | *PKD1L1* |
| ENSG00000268926 | 9 | 98524605 | 98547013 | 77 | 18 | 41604 | -0.76382 | 0.77751 | *DKFZP434H0512* |
| ENSG00000110888 | 12 | 30852486 | 30917885 | 200 | 21 | 54169 | -0.76387 | 0.77753 | *CAPRIN2* |
| ENSG00000181085 | 8 | 144788429 | 144814628 | 81 | 17 | 34145 | -0.76439 | 0.77768 | *MAPK15* |
| ENSG00000156282 | 21 | 31528241 | 31548971 | 82 | 10 | 37047 | -0.7644 | 0.77769 | *CLDN17* |
| ENSG00000162367 | 1 | 47671962 | 47707892 | 101 | 13 | 37237 | -0.76474 | 0.77779 | *TAL1* |
| ENSG00000196814 | 9 | 129079128 | 129279320 | 589 | 60 | 50330 | -0.76496 | 0.77785 | *MVB12B* |
| ENSG00000185046 | 12 | 99110235 | 100388432 | 3476 | 280 | 44537 | -0.76505 | 0.77788 | *ANKS1B* |
| ENSG00000185818 | 4 | 2051239 | 2080816 | 47 | 9 | 32042 | -0.76514 | 0.77791 | *NAT8L* |
| ENSG00000104219 | 8 | 17003538 | 17092308 | 263 | 47 | 39983 | -0.76518 | 0.77792 | *ZDHHC2* |

| | | | | | | | | | |
|---|---|---|---|---|---|---|---|---|---|
| ENSG00000116667 | 1 | 184346192 | 184608154 | 730 | 77 | 46443 | -0.76521 | 0.77793 | *C1orf21* |
| ENSG00000168522 | 8 | 42879337 | 42950931 | 140 | 23 | 47519 | -0.76522 | 0.77793 | *FNTA* |
| ENSG00000167346 | 11 | 4716157 | 5023659 | 1418 | 93 | 44454 | -0.76537 | 0.77797 | *MMP26* |
| ENSG00000213922 | 19 | 39920212 | 39942082 | 73 | 9 | 44740 | -0.76579 | 0.7781 | *AC011500.1* |
| ENSG00000165828 | 10 | 135150650 | 135176187 | 115 | 11 | 27267 | -0.76583 | 0.77811 | *PRAP1* |
| ENSG00000124253 | 20 | 56126136 | 56151513 | 121 | 29 | 49333 | -0.76589 | 0.77813 | *PCK1* |
| ENSG00000100802 | 14 | 23446110 | 23489375 | 67 | 18 | 49883 | -0.76596 | 0.77815 | *C14orf93* |
| ENSG00000111799 | 6 | 75784042 | 75925767 | 229 | 44 | 34760 | -0.76618 | 0.77822 | *COL12A1* |
| ENSG00000153253 | 2 | 165934032 | 166070577 | 321 | 33 | 41333 | -0.76623 | 0.77823 | *SCN3A* |
| ENSG00000034053 | 15 | 29119629 | 29420518 | 498 | 76 | 31027 | -0.76685 | 0.77841 | *APBA2* |
| ENSG00000135148 | 12 | 112553305 | 112601407 | 47 | 12 | 40947 | -0.76717 | 0.77851 | *TRAFD1* |
| ENSG00000127463 | 1 | 19532158 | 19588046 | 201 | 27 | 50092 | -0.76735 | 0.77856 | *EMC1* |
| ENSG00000092009 | 14 | 24964559 | 24987471 | 96 | 17 | 44366 | -0.76734 | 0.77856 | *CMA1* |
| ENSG00000100842 | 14 | 23815611 | 23844961 | 77 | 15 | 39585 | -0.76752 | 0.77861 | *EFS* |
| ENSG00000241258 | 7 | 65569591 | 65629555 | 181 | 17 | 44142 | -0.76807 | 0.77878 | *CRCP* |
| ENSG00000126456 | 19 | 50152826 | 50179132 | 40 | 10 | 38923 | -0.76843 | 0.77888 | *IRF3* |
| ENSG00000189167 | 13 | 32867837 | 32899481 | 119 | 16 | 53707 | -0.76882 | 0.779 | *ZAR1L* |
| ENSG00000165076 | 7 | 141526086 | 141551287 | 97 | 8 | 56672 | -0.76905 | 0.77907 | *PRSS37* |
| ENSG00000163964 | 3 | 196356646 | 196472878 | 448 | 92 | 48288 | -0.76956 | 0.77922 | *PIGX* |
| ENSG00000166426 | 15 | 78622666 | 78650572 | 77 | 24 | 45129 | -0.76959 | 0.77923 | *CRABP1* |
| ENSG00000176256 | 1 | 34316076 | 34340392 | 48 | 14 | 43715 | -0.7699 | 0.77932 | *HMGB4* |
| ENSG00000185019 | 20 | 3078219 | 3150842 | 184 | 32 | 40603 | -0.77006 | 0.77937 | *UBOX5* |
| ENSG00000258429 | 16 | 69352524 | 69374498 | 59 | 12 | 54160 | -0.77035 | 0.77945 | *PDF* |
| ENSG00000141255 | 17 | 3333313 | 3427146 | 383 | 40 | 37574 | -0.77055 | 0.77951 | *SPATA22* |
| ENSG00000169251 | 3 | 160812484 | 160981320 | 445 | 43 | 46365 | -0.77065 | 0.77954 | *NMD3* |
| ENSG00000113593 | 5 | 64849063 | 64893376 | 160 | 12 | 50555 | -0.77071 | 0.77956 | *PPWD1* |
| ENSG00000153820 | 2 | 228834666 | 229056361 | 867 | 57 | 54623 | -0.77142 | 0.77977 | *SPHKAP* |
| ENSG00000131686 | 1 | 8995926 | 9045151 | 191 | 54 | 43442 | -0.7718 | 0.77988 | *CA6* |
| ENSG00000169429 | 4 | 74596223 | 74619433 | 53 | 8 | 47856 | -0.77198 | 0.77994 | *IL8* |
| ENSG00000198062 | 22 | 16246441 | 16297937 | 5 | 3 | 42713 | -0.77226 | 0.78002 | *POTEH* |
| ENSG00000126259 | 19 | 36336763 | 36368011 | 58 | 10 | 31437 | -0.77246 | 0.78008 | *KIRREL2* |
| ENSG00000183779 | 8 | 37543269 | 37567537 | 14 | 8 | 54151 | -0.77281 | 0.78018 | *ZNF703* |
| ENSG00000141580 | 17 | 80562438 | 80616429 | 241 | 40 | 49911 | -0.77312 | 0.78028 | *WDR45B* |
| ENSG00000160856 | 1 | 157634111 | 157680647 | 127 | 18 | 45503 | -0.77318 | 0.78029 | *FCRL3* |
| ENSG00000184155 | 1 | 159494793 | 159515842 | 70 | 12 | 36934 | -0.7732 | 0.7803 | *OR10J5* |
| ENSG00000184040 | 10 | 18031218 | 18099855 | 3 | 2 | 43742 | -0.77334 | 0.78034 | *TMEM236* |

| | | | | | | | | | |
|---|---|---|---|---|---|---|---|---|---|
| ENSG00000128596 | 7 | 128420811 | 128472186 | 132 | 15 | 49216 | -0.7736 | 0.78042 | *CCDC136* |
| ENSG00000180438 | 3 | 13968756 | 14134311 | 414 | 29 | 53470 | -0.77383 | 0.78049 | *TPRXL* |
| ENSG00000104237 | 8 | 55518627 | 55553394 | 85 | 15 | 43487 | -0.77412 | 0.78057 | *RP1* |
| ENSG00000125816 | 20 | 21366005 | 21388666 | 55 | 15 | 53228 | -0.77432 | 0.78063 | *NKX2-4* |
| ENSG00000115486 | 2 | 85761846 | 85798670 | 129 | 18 | 47465 | -0.77443 | 0.78066 | *GGCX* |
| ENSG00000164430 | 6 | 74113238 | 74171999 | 136 | 38 | 48924 | -0.77445 | 0.78067 | *MB21D1* |
| ENSG00000133302 | 5 | 93944052 | 94085141 | 446 | 31 | 51676 | -0.77451 | 0.78069 | *ANKRD32* |
| ENSG00000100417 | 22 | 41962898 | 41995894 | 56 | 9 | 36323 | -0.77452 | 0.78069 | *PMM1* |
| ENSG00000175899 | 12 | 9210260 | 9278825 | 188 | 14 | 41811 | -0.77469 | 0.78074 | *A2M* |
| ENSG00000161618 | 19 | 49946426 | 49984305 | 129 | 17 | 29079 | -0.77495 | 0.78081 | *ALDH16A1* |
| ENSG00000214414 | 11 | 89433467 | 89461040 | 79 | 6 | 53451 | -0.77502 | 0.78084 | *TRIM77* |
| ENSG00000118231 | 2 | 208976331 | 208999225 | 117 | 20 | 47009 | -0.77516 | 0.78088 | *CRYGD* |
| ENSG00000197405 | 19 | 47783280 | 47835323 | 199 | 20 | 40444 | -0.77546 | 0.78097 | *C5AR1* |
| ENSG00000214575 | 15 | 83201951 | 83327612 | 333 | 33 | 44456 | -0.7758 | 0.78107 | *CPEB1* |
| ENSG00000170242 | 11 | 11852970 | 11990870 | 323 | 33 | 48786 | -0.77597 | 0.78112 | *USP47* |
| ENSG00000067064 | 10 | 1075848 | 1105110 | 94 | 20 | 49178 | -0.77604 | 0.78114 | *IDI1* |
| ENSG00000109475 | 4 | 109531722 | 109561568 | 88 | 10 | 54317 | -0.7769 | 0.78139 | *RPL34* |
| ENSG00000175324 | 8 | 38010839 | 38044248 | 63 | 10 | 45274 | -0.77695 | 0.7814 | *LSM1* |
| ENSG00000143624 | 1 | 153690543 | 153756555 | 125 | 21 | 38681 | -0.77708 | 0.78144 | *INTS3* |
| ENSG00000141642 | 18 | 48484361 | 48524491 | 57 | 10 | 45651 | -0.77719 | 0.78148 | *ELAC1* |
| ENSG00000257062 | 12 | 21190113 | 21339829 | 578 | 45 | 30947 | -0.7779 | 0.78169 | *RP11-125O5.2* |
| ENSG00000114779 | 3 | 51992526 | 52027425 | 43 | 15 | 34452 | -0.77859 | 0.78189 | *ABHD14B* |
| ENSG00000214694 | 2 | 39107021 | 39212590 | 204 | 33 | 41722 | -0.77896 | 0.782 | *ARHGEF33* |
| ENSG00000204256 | 6 | 32926437 | 32959282 | 259 | 38 | 48389 | -0.77973 | 0.78223 | *BRD2* |
| ENSG00000185052 | 20 | 19183290 | 19713581 | 1701 | 183 | 44203 | -0.78029 | 0.78239 | *SLC24A3* |
| ENSG00000105650 | 19 | 18308771 | 18376229 | 223 | 29 | 44855 | -0.7804 | 0.78242 | *PDE4C* |
| ENSG00000196290 | 2 | 201744050 | 201778655 | 51 | 10 | 46839 | -0.78057 | 0.78247 | *NIF3L1* |
| ENSG00000232838 | 20 | 18108517 | 18133813 | 102 | 23 | 48286 | -0.78107 | 0.78262 | *PET117* |
| ENSG00000185739 | 16 | 4229375 | 4302081 | 295 | 52 | 43365 | -0.78109 | 0.78263 | *SRL* |
| ENSG00000115685 | 2 | 242078991 | 242133067 | 185 | 27 | 42727 | -0.78117 | 0.78265 | *PPP1R7* |
| ENSG00000132953 | 13 | 21341469 | 21487187 | 348 | 31 | 32015 | -0.78197 | 0.78288 | *XPO4* |
| ENSG00000157326 | 14 | 24412795 | 24448488 | 167 | 9 | 20433 | -0.78214 | 0.78293 | *DHRS4* |
| ENSG00000187272 | 17 | 39384297 | 39405256 | 103 | 7 | 44509 | -0.78231 | 0.78298 | *KRTAP9-8* |
| ENSG00000141646 | 18 | 48484410 | 48621415 | 171 | 19 | 52161 | -0.78271 | 0.7831 | *SMAD4* |
| ENSG00000079435 | 19 | 42895659 | 42941578 | 37 | 14 | 39649 | -0.7828 | 0.78313 | *LIPE* |
| ENSG00000106615 | 7 | 151153098 | 151227206 | 158 | 23 | 46755 | -0.7829 | 0.78316 | *RHEB* |

| | | | | | | | | | |
|---|---|---|---|---|---|---|---|---|---|
| ENSG00000104888 | 19 | 49922658 | 49955617 | 70 | 13 | 34587 | -0.78359 | 0.78336 | *SLC17A7* |
| ENSG00000184058 | 22 | 19734226 | 19781116 | 151 | 27 | 47344 | -0.78413 | 0.78352 | *TBX1* |
| ENSG00000101782 | 18 | 21022787 | 21076567 | 170 | 24 | 37763 | -0.78458 | 0.78365 | *RIOK3* |
| ENSG00000167862 | 17 | 72998765 | 73027356 | 127 | 20 | 37674 | -0.78471 | 0.78369 | *ICT1* |
| ENSG00000189375 | 17 | 18528319 | 18575263 | 82 | 21 | 38191 | -0.7848 | 0.78371 | *TBC1D28* |
| ENSG00000197905 | 12 | 3058496 | 3159839 | 350 | 51 | 34027 | -0.78549 | 0.78392 | *TEAD4* |
| ENSG00000153666 | 15 | 23245242 | 23278823 | 30 | 13 | 42897 | -0.78555 | 0.78393 | *GOLGA8I* |
| ENSG00000183018 | 17 | 4392133 | 4452330 | 181 | 48 | 44949 | -0.78566 | 0.78397 | *SPNS2* |
| ENSG00000184208 | 22 | 42074943 | 42104140 | 58 | 7 | 40074 | -0.78623 | 0.78413 | *C22orf46* |
| ENSG00000109927 | 11 | 120961882 | 121072202 | 341 | 30 | 35135 | -0.78628 | 0.78415 | *TECTA* |
| ENSG00000196418 | 1 | 247275277 | 247345318 | 305 | 36 | 40620 | -0.78653 | 0.78422 | *ZNF124* |
| ENSG00000184344 | 12 | 7832378 | 7858372 | 144 | 31 | 50253 | -0.78699 | 0.78436 | *GDF3* |
| ENSG00000196754 | 1 | 153523584 | 153550366 | 51 | 13 | 34812 | -0.78715 | 0.7844 | *S100A2* |
| ENSG00000122008 | 5 | 74797581 | 74906969 | 188 | 24 | 49407 | -0.78714 | 0.7844 | *POLK* |
| ENSG00000172315 | 20 | 45303004 | 45328418 | 67 | 15 | 41467 | -0.78714 | 0.7844 | *TP53RK* |
| ENSG00000107968 | 10 | 30712866 | 30760762 | 100 | 17 | 34982 | -0.78749 | 0.7845 | *MAP3K8* |
| ENSG00000162676 | 1 | 92930319 | 92962433 | 60 | 10 | 27659 | -0.78768 | 0.78456 | *GFI1* |
| ENSG00000214013 | 15 | 42555431 | 42655864 | 318 | 18 | 33095 | -0.78781 | 0.78459 | *GANC* |
| ENSG00000003402 | 2 | 201970827 | 202051410 | 215 | 25 | 50731 | -0.78781 | 0.7846 | *CFLAR* |
| ENSG00000188000 | 19 | 9286279 | 9309493 | 99 | 14 | 36849 | -0.78785 | 0.78461 | *OR7D2* |
| ENSG00000102878 | 16 | 67187288 | 67213848 | 22 | 10 | 44775 | -0.78832 | 0.78474 | *HSF4* |
| ENSG00000182108 | 16 | 11012748 | 11046317 | 97 | 26 | 44324 | -0.78839 | 0.78477 | *DEXI* |
| ENSG00000196131 | 19 | 53751545 | 53775090 | 118 | 16 | 48273 | -0.78858 | 0.78482 | *VN1R2* |
| ENSG00000169327 | 14 | 21613026 | 21634222 | 101 | 24 | 51535 | -0.78862 | 0.78483 | *OR5AU1* |
| ENSG00000161381 | 17 | 37209556 | 37320647 | 457 | 62 | 52756 | -0.78865 | 0.78484 | *PLXDC1* |
| ENSG00000103148 | 16 | 124273 | 198859 | 205 | 35 | 36785 | -0.78877 | 0.78488 | *NPRL3* |
| ENSG00000047056 | 10 | 1085478 | 1188237 | 334 | 47 | 46591 | -0.78889 | 0.78491 | *WDR37* |
| ENSG00000259517 | 16 | 54269456 | 54314658 | 197 | 18 | 46738 | -0.78916 | 0.78499 | *RP11-324D17.1* |
| ENSG00000115310 | 2 | 55189325 | 55349757 | 551 | 59 | 42124 | -0.78922 | 0.78501 | *RTN4* |
| ENSG00000241935 | 10 | 99334080 | 99382559 | 123 | 31 | 43513 | -0.78925 | 0.78502 | *HOGA1* |
| ENSG00000254004 | 19 | 36991597 | 37029562 | 115 | 15 | 50462 | -0.79011 | 0.78527 | *ZNF260* |
| ENSG00000198178 | 12 | 7872011 | 7914201 | 211 | 27 | 48000 | -0.79059 | 0.78541 | *CLEC4C* |
| ENSG00000156804 | 8 | 124500129 | 124563446 | 186 | 48 | 46610 | -0.79109 | 0.78555 | *FBXO32* |
| ENSG00000106066 | 7 | 29024847 | 29245067 | 840 | 122 | 45879 | -0.79151 | 0.78568 | *CPVL* |
| ENSG00000110651 | 11 | 2387407 | 2428649 | 92 | 25 | 37703 | -0.79154 | 0.78569 | *CD81* |
| ENSG00000178404 | 17 | 76856992 | 76909299 | 256 | 36 | 42638 | -0.79158 | 0.7857 | *DDC8* |

| | | | | | | | | | |
|---|---|---|---|---|---|---|---|---|---|
| ENSG00000197079 | 17 | 39622941 | 39647392 | 135 | 11 | 56669 | -0.79179 | 0.78576 | *KRT35* |
| ENSG00000090447 | 16 | 4297187 | 4333076 | 79 | 13 | 52133 | -0.79273 | 0.78603 | *TFAP4* |
| ENSG00000123999 | 2 | 220423884 | 220450435 | 78 | 20 | 48036 | -0.79306 | 0.78613 | *INHA* |
| ENSG00000130540 | 22 | 44210389 | 44268398 | 226 | 23 | 54751 | -0.79312 | 0.78615 | *SULT4A1* |
| ENSG00000169484 | 14 | 20472419 | 20493352 | 110 | 17 | 40759 | -0.79316 | 0.78616 | *OR4K14* |
| ENSG00000172380 | 1 | 68157149 | 68309150 | 411 | 51 | 49231 | -0.79331 | 0.7862 | *GNG12* |
| ENSG00000176887 | 2 | 5822799 | 5851516 | 72 | 17 | 49683 | -0.79386 | 0.78636 | *SOX11* |
| ENSG00000171804 | 19 | 38365463 | 38407317 | 82 | 19 | 37326 | -0.79387 | 0.78637 | *WDR87* |
| ENSG00000152413 | 5 | 78658459 | 78820040 | 372 | 46 | 37362 | -0.7942 | 0.78646 | *HOMER1* |
| ENSG00000178860 | 8 | 72743784 | 72766703 | 69 | 13 | 37707 | -0.79419 | 0.78646 | *MSC* |
| ENSG00000111450 | 12 | 131264145 | 131333811 | 317 | 24 | 43234 | -0.79442 | 0.78652 | *STX2* |
| ENSG00000157119 | 3 | 42717011 | 42744036 | 61 | 21 | 46526 | -0.79482 | 0.78664 | *KLHL40* |
| ENSG00000127377 | 7 | 151115921 | 151147899 | 114 | 23 | 34124 | -0.79539 | 0.78681 | *CRYGN* |
| ENSG00000150753 | 5 | 10240033 | 10276524 | 190 | 13 | 23506 | -0.79551 | 0.78684 | *CCT5* |
| ENSG00000188060 | 1 | 28908712 | 28931955 | 33 | 13 | 48624 | -0.79601 | 0.78699 | *RAB42* |
| ENSG00000184949 | 22 | 38964125 | 39062634 | 271 | 18 | 46599 | -0.79612 | 0.78702 | *FAM227A* |
| ENSG00000065970 | 12 | 8175299 | 8218099 | 76 | 21 | 41308 | -0.7965 | 0.78713 | *FOXJ2* |
| ENSG00000205864 | 11 | 1708425 | 1728985 | 110 | 19 | 45521 | -0.79659 | 0.78716 | *KRTAP5-6* |
| ENSG00000160207 | 21 | 44939072 | 45089374 | 317 | 21 | 46047 | -0.79669 | 0.78718 | *HSF2BP* |
| ENSG00000150687 | 11 | 86492101 | 86673952 | 723 | 96 | 45904 | -0.79719 | 0.78733 | *PRSS23* |
| ENSG00000236981 | 11 | 123883720 | 123904655 | 71 | 14 | 45582 | -0.79728 | 0.78736 | *OR10G9* |
| ENSG00000158234 | 3 | 138317448 | 138362218 | 66 | 9 | 42724 | -0.79758 | 0.78744 | *FAIM* |
| ENSG00000197123 | 7 | 63678852 | 63737309 | 274 | 30 | 46804 | -0.79766 | 0.78747 | *ZNF679* |
| ENSG00000156261 | 21 | 30418126 | 30456118 | 72 | 17 | 44112 | -0.79781 | 0.78751 | *CCT8* |
| ENSG00000142949 | 1 | 43980858 | 44099343 | 323 | 30 | 41401 | -0.79789 | 0.78753 | *PTPRF* |
| ENSG00000153789 | 16 | 85121965 | 85156114 | 211 | 46 | 49363 | -0.79804 | 0.78758 | *FAM92B* |
| ENSG00000113240 | 5 | 178019665 | 178067616 | 117 | 16 | 31007 | -0.79812 | 0.7876 | *CLK4* |
| ENSG00000126838 | 12 | 9291436 | 9370966 | 215 | 31 | 51753 | -0.79827 | 0.78764 | *PZP* |
| ENSG00000118276 | 18 | 29192210 | 29275799 | 178 | 30 | 45544 | -0.79864 | 0.78775 | *B4GALT6* |
| ENSG00000204278 | 17 | 76217391 | 76247068 | 146 | 30 | 54106 | -0.79906 | 0.78787 | *TMEM235* |
| ENSG00000268538 | 22 | 30804212 | 30824469 | 49 | 12 | 42111 | -0.7995 | 0.788 | *KIAA1658* |
| ENSG00000172568 | 5 | 156758608 | 156782729 | 91 | 8 | 30660 | -0.79988 | 0.78811 | *FNDC9* |
| ENSG00000188280 | 22 | 20682438 | 20726938 | 41 | 17 | 40774 | -0.80077 | 0.78837 | *FAM230A* |
| ENSG00000123933 | 4 | 2239159 | 2274021 | 95 | 16 | 33186 | -0.80086 | 0.78839 | *MXD4* |
| ENSG00000091704 | 7 | 130010180 | 130037955 | 93 | 15 | 54999 | -0.80088 | 0.7884 | *CPA1* |
| ENSG00000182768 | 15 | 90798891 | 90826463 | 62 | 12 | 39400 | -0.80089 | 0.7884 | *NGRN* |

| | | | | | | | | | |
|---|---|---|---|---|---|---|---|---|---|
| ENSG00000174950 | 1 | 27695666 | 27719870 | 33 | 9 | 39716 | -0.80097 | 0.78842 | *CD164L2* |
| ENSG00000243452 | 1 | 148545979 | 148606267 | 12 | 5 | 43409 | -0.801 | 0.78843 | *NBPF15* |
| ENSG00000163701 | 3 | 9934296 | 9968086 | 108 | 12 | 31986 | -0.80105 | 0.78845 | *IL17RE* |
| ENSG00000203785 | 1 | 153055611 | 153088660 | 98 | 22 | 31449 | -0.8013 | 0.78852 | *SPRR2E* |
| ENSG00000186026 | 19 | 44566297 | 44603766 | 77 | 10 | 25488 | -0.8013 | 0.78852 | *ZNF284* |
| ENSG00000205116 | 1 | 1351508 | 1373167 | 59 | 11 | 29646 | -0.80148 | 0.78857 | *TMEM88B* |
| ENSG00000099219 | 9 | 5755076 | 5843117 | 236 | 24 | 44454 | -0.80174 | 0.78865 | *ERMP1* |
| ENSG00000198874 | 7 | 66450160 | 66714501 | 1616 | 38 | 49411 | -0.80212 | 0.78876 | *TYW1* |
| ENSG00000166796 | 11 | 18423854 | 18483605 | 300 | 23 | 48485 | -0.80237 | 0.78883 | *LDHC* |
| ENSG00000183722 | 13 | 39907029 | 40187665 | 901 | 130 | 45468 | -0.80281 | 0.78896 | *LHFP* |
| ENSG00000205423 | 16 | 50048321 | 50080999 | 100 | 15 | 47296 | -0.80286 | 0.78897 | *CNEP1R1* |
| ENSG00000029559 | 4 | 88710733 | 88743074 | 115 | 27 | 44296 | -0.80296 | 0.789 | *IBSP* |
| ENSG00000128298 | 22 | 38470896 | 38516677 | 125 | 21 | 48233 | -0.80312 | 0.78905 | *BAIAP2L2* |
| ENSG00000188474 | 19 | 12193078 | 12235491 | 89 | 15 | 36326 | -0.80418 | 0.78935 | *ZNF788* |
| ENSG00000177700 | 11 | 827356 | 852545 | 121 | 13 | 30430 | -0.8042 | 0.78936 | *POLR2L* |
| ENSG00000162729 | 1 | 160051130 | 160078733 | 74 | 14 | 48629 | -0.80456 | 0.78946 | *IGSF8* |
| ENSG00000132196 | 1 | 162750492 | 162792607 | 116 | 27 | 39322 | -0.80481 | 0.78954 | *HSD17B7* |
| ENSG00000154479 | 2 | 170491935 | 170560943 | 275 | 17 | 56298 | -0.80512 | 0.78963 | *CCDC173* |
| ENSG00000100739 | 14 | 96712161 | 96745304 | 129 | 31 | 51867 | -0.80544 | 0.78972 | *BDKRB1* |
| ENSG00000165102 | 8 | 42985556 | 43067998 | 113 | 23 | 40666 | -0.80556 | 0.78975 | *HGSNAT* |
| ENSG00000139438 | 12 | 110142033 | 110218312 | 182 | 34 | 52356 | -0.8058 | 0.78982 | *FAM222A* |
| ENSG00000174125 | 4 | 38782298 | 38868438 | 312 | 32 | 26045 | -0.80589 | 0.78985 | *TLR1* |
| ENSG00000048544 | 6 | 42164539 | 42195603 | 127 | 9 | 32179 | -0.80624 | 0.78995 | *MRPS10* |
| ENSG00000160226 | 21 | 45738827 | 45769285 | 79 | 21 | 45486 | -0.80635 | 0.78998 | *C21orf2* |
| ENSG00000187054 | 4 | 68765103 | 68839858 | 240 | 33 | 42121 | -0.80682 | 0.79012 | *TMPRSS11A* |
| ENSG00000135678 | 12 | 69225977 | 69375350 | 486 | 93 | 43115 | -0.80723 | 0.79023 | *CPM* |
| ENSG00000153064 | 4 | 102322443 | 103005969 | 2044 | 164 | 44719 | -0.80733 | 0.79026 | *BANK1* |
| ENSG00000217702 | 2 | 74351599 | 74372521 | 62 | 13 | 36992 | -0.80743 | 0.79029 | *MGC10955* |
| ENSG00000178690 | 18 | 52244988 | 52276724 | 79 | 13 | 42096 | -0.80753 | 0.79032 | *DYNAP* |
| ENSG00000204257 | 6 | 32906390 | 32946871 | 285 | 32 | 50449 | -0.80787 | 0.79042 | *HLA-DMA* |
| ENSG00000122490 | 18 | 77652420 | 77721664 | 227 | 35 | 49083 | -0.80803 | 0.79046 | *PQLC1* |
| ENSG00000145623 | 5 | 38835960 | 38955698 | 350 | 62 | 50169 | -0.80814 | 0.7905 | *OSMR* |
| ENSG00000203727 | 6 | 147820063 | 148068683 | 1160 | 110 | 44215 | -0.8084 | 0.79057 | *SAMD5* |
| ENSG00000157087 | 3 | 10355707 | 10759716 | 1527 | 182 | 40689 | -0.80858 | 0.79062 | *ATP2B2* |
| ENSG00000006377 | 7 | 96624860 | 96650351 | 45 | 15 | 54822 | -0.80878 | 0.79068 | *DLX6* |
| ENSG00000159055 | 21 | 33630530 | 33661380 | 53 | 13 | 44627 | -0.80885 | 0.7907 | *MIS18A* |

| | | | | | | | | | |
|---|---|---|---|---|---|---|---|---|---|
| ENSG00000104290 | 8 | 28341729 | 28441775 | 220 | 14 | 52295 | -0.80897 | 0.79073 | *FZD3* |
| ENSG00000249242 | 4 | 83394323 | 83493510 | 251 | 54 | 43332 | -0.80909 | 0.79077 | *TMEM150C* |
| ENSG00000149591 | 11 | 117060037 | 117085498 | 94 | 17 | 35320 | -0.80918 | 0.79079 | *TAGLN* |
| ENSG00000198431 | 12 | 104599557 | 104754061 | 558 | 53 | 48740 | -0.80917 | 0.79079 | *TXNRD1* |
| ENSG00000177673 | 2 | 232447575 | 232468994 | 52 | 14 | 44289 | -0.80947 | 0.79088 | *C2orf57* |
| ENSG00000111877 | 6 | 119124605 | 119266327 | 546 | 36 | 41060 | -0.80976 | 0.79096 | *MCM9* |
| ENSG00000149654 | 20 | 44792372 | 44947137 | 352 | 73 | 45736 | -0.80992 | 0.79101 | *CDH22* |
| ENSG00000182447 | 3 | 161204596 | 161231730 | 134 | 10 | 47762 | -0.81026 | 0.7911 | *OTOL1* |
| ENSG00000113811 | 3 | 53908437 | 53936015 | 86 | 13 | 32403 | -0.81032 | 0.79112 | *SELK* |
| ENSG00000139269 | 12 | 57836106 | 57863063 | 60 | 16 | 34919 | -0.81038 | 0.79114 | *INHBE* |
| ENSG00000152726 | 10 | 47884023 | 47959412 | 1 | 1 | 42478 | -0.81059 | 0.7912 | *FAM21B* |
| ENSG00000107651 | 10 | 121642223 | 121712014 | 264 | 16 | 35140 | -0.81065 | 0.79122 | *SEC23IP* |
| ENSG00000111837 | 6 | 10752956 | 10848764 | 201 | 37 | 52013 | -0.81148 | 0.79146 | *MAK* |
| ENSG00000268275 | 6 | 41643964 | 41664127 | 81 | 15 | 54343 | -0.81202 | 0.79161 | *AL035588.1* |
| ENSG00000149289 | 11 | 109954087 | 110052566 | 367 | 31 | 46735 | -0.81224 | 0.79167 | *ZC3H12C* |
| ENSG00000154864 | 18 | 10656480 | 11158587 | 2066 | 207 | 46579 | -0.81224 | 0.79167 | *PIEZO2* |
| ENSG00000105204 | 19 | 40305990 | 40334841 | 78 | 12 | 54747 | -0.81319 | 0.79195 | *DYRK1B* |
| ENSG00000084090 | 2 | 96840597 | 96884563 | 69 | 13 | 50750 | -0.81328 | 0.79197 | *STARD7* |
| ENSG00000134480 | 5 | 86677311 | 86718836 | 54 | 9 | 48429 | -0.81333 | 0.79199 | *CCNH* |
| ENSG00000155380 | 1 | 113444469 | 113509635 | 110 | 22 | 37410 | -0.81338 | 0.792 | *SLC16A1* |
| ENSG00000099810 | 9 | 21792542 | 21941646 | 344 | 43 | 37393 | -0.81339 | 0.792 | *MTAP* |
| ENSG00000150636 | 18 | 66372446 | 66732426 | 1209 | 113 | 37707 | -0.81338 | 0.792 | *CCDC102B* |
| ENSG00000150456 | 13 | 21292870 | 21358097 | 228 | 23 | 32821 | -0.81386 | 0.79214 | *N6AMT2* |
| ENSG00000105374 | 19 | 51864860 | 51885969 | 65 | 17 | 37844 | -0.81412 | 0.79221 | *NKG7* |
| ENSG00000062822 | 19 | 50877461 | 50931273 | 134 | 20 | 35510 | -0.81418 | 0.79223 | *POLD1* |
| ENSG00000180901 | 17 | 73018670 | 73071984 | 178 | 33 | 38645 | -0.81432 | 0.79227 | *KCTD2* |
| ENSG00000204548 | 20 | 29982648 | 30010641 | 96 | 12 | 36521 | -0.81464 | 0.79236 | *DEFB121* |
| ENSG00000165188 | 9 | 116049373 | 116075656 | 75 | 15 | 41506 | -0.81487 | 0.79243 | *RNF183* |
| ENSG00000101350 | 20 | 30855467 | 30932814 | 136 | 12 | 45488 | -0.81493 | 0.79244 | *KIF3B* |
| ENSG00000173905 | 3 | 167716465 | 167823763 | 266 | 33 | 37417 | -0.81516 | 0.79251 | *GOLIM4* |
| ENSG00000013016 | 2 | 31446880 | 31502313 | 158 | 37 | 44966 | -0.81532 | 0.79256 | *EHD3* |
| ENSG00000144746 | 3 | 69124095 | 69165217 | 86 | 16 | 42697 | -0.81601 | 0.79275 | *ARL6IP5* |
| ENSG00000134460 | 10 | 6042652 | 6114288 | 335 | 66 | 48483 | -0.81622 | 0.79281 | *IL2RA* |
| ENSG00000176788 | 5 | 17055707 | 17286943 | 894 | 135 | 44335 | -0.81662 | 0.79293 | *BASP1* |
| ENSG00000101158 | 20 | 57546263 | 57580188 | 118 | 18 | 54830 | -0.81711 | 0.79307 | *NELFCD* |
| ENSG00000113070 | 5 | 139702428 | 139736216 | 66 | 16 | 54073 | -0.81738 | 0.79314 | *HBEGF* |

| | | | | | | | | | |
|---|---|---|---|---|---|---|---|---|---|
| ENSG00000010219 | 12 | 4661370 | 4733325 | 228 | 42 | 40170 | -0.81749 | 0.79318 | *DYRK4* |
| ENSG00000267426 | 17 | 73884726 | 73936210 | 111 | 19 | 38696 | -0.81761 | 0.79321 | *RP11-552F3.12* |
| ENSG00000104872 | 19 | 49939555 | 49966754 | 92 | 11 | 30116 | -0.8178 | 0.79326 | *PIH1D1* |
| ENSG00000171495 | 5 | 40988119 | 41081444 | 460 | 55 | 52711 | -0.81841 | 0.79344 | *MROH2B* |
| ENSG00000177108 | 14 | 77587613 | 77619077 | 107 | 26 | 47396 | -0.81847 | 0.79346 | *ZDHHC22* |
| ENSG00000006075 | 17 | 34405602 | 34427515 | 108 | 12 | 47228 | -0.81887 | 0.79357 | *CCL3* |
| ENSG00000170310 | 17 | 9143788 | 9489908 | 998 | 173 | 47023 | -0.81938 | 0.79372 | *STX8* |
| ENSG00000178409 | 6 | 107376386 | 107446473 | 201 | 41 | 43856 | -0.81963 | 0.79379 | *BEND3* |
| ENSG00000186049 | 12 | 52991354 | 53022343 | 121 | 23 | 50786 | -0.81966 | 0.79379 | *KRT73* |
| ENSG00000176225 | 18 | 67661029 | 67883181 | 565 | 53 | 41070 | -0.81981 | 0.79384 | *RTTN* |
| ENSG00000130433 | 19 | 54485542 | 54525923 | 198 | 38 | 45259 | -0.82001 | 0.79389 | *CACNG6* |
| ENSG00000150756 | 5 | 10216442 | 10260009 | 229 | 25 | 28139 | -0.82041 | 0.79401 | *FAM173B* |
| ENSG00000112077 | 6 | 49562871 | 49614552 | 218 | 22 | 27985 | -0.8204 | 0.79401 | *RHAG* |
| ENSG00000116117 | 2 | 205400516 | 206494886 | 2948 | 221 | 43889 | -0.82107 | 0.7942 | *PARD3B* |
| ENSG00000181464 | 17 | 15458797 | 15479590 | 40 | 9 | 35667 | -0.82116 | 0.79422 | *CDRT1* |
| ENSG00000212999 | 8 | 93885865 | 93908013 | 58 | 8 | 28175 | -0.82126 | 0.79425 | *AC117834.1* |
| ENSG00000198521 | 19 | 21977752 | 22044927 | 222 | 27 | 34901 | -0.82208 | 0.79448 | *ZNF43* |
| ENSG00000165935 | 12 | 27609743 | 27665118 | 256 | 22 | 35541 | -0.82243 | 0.79458 | *SMCO2* |
| ENSG00000132964 | 13 | 26818276 | 26989375 | 446 | 39 | 40685 | -0.82271 | 0.79466 | *CDK8* |
| ENSG00000102606 | 13 | 111756906 | 111968084 | 702 | 91 | 49005 | -0.82273 | 0.79467 | *ARHGEF7* |
| ENSG00000145494 | 5 | 1791514 | 1826719 | 161 | 17 | 34350 | -0.82284 | 0.7947 | *NDUFS6* |
| ENSG00000180219 | 12 | 100031531 | 100053824 | 57 | 14 | 38414 | -0.82287 | 0.79471 | *FAM71C* |
| ENSG00000138668 | 4 | 83263651 | 83305656 | 132 | 20 | 40255 | -0.82299 | 0.79474 | *HNRNPD* |
| ENSG00000260001 | 19 | 7971030 | 7993982 | 63 | 25 | 42675 | -0.82298 | 0.79474 | *TGFBR3L* |
| ENSG00000228032 | 14 | 104700541 | 104722473 | 119 | 15 | 50155 | -0.82348 | 0.79488 | *C14orf144* |
| ENSG00000167476 | 19 | 2242251 | 2279758 | 138 | 29 | 32378 | -0.82363 | 0.79493 | *JSRP1* |
| ENSG00000254673 | 8 | 42863567 | 42942477 | 150 | 24 | 47483 | -0.82475 | 0.79524 | *RP11-598P20.5* |
| ENSG00000142178 | 21 | 44824395 | 44857008 | 80 | 25 | 43928 | -0.82523 | 0.79538 | *SIK1* |
| ENSG00000180316 | 6 | 36200980 | 36286372 | 224 | 61 | 48689 | -0.8254 | 0.79543 | *PNPLA1* |
| ENSG00000143954 | 2 | 79242812 | 79265631 | 134 | 12 | 55814 | -0.82543 | 0.79544 | *REG3G* |
| ENSG00000175895 | 8 | 96136032 | 96178912 | 130 | 16 | 46153 | -0.82548 | 0.79545 | *PLEKHF2* |
| ENSG00000132669 | 20 | 19857165 | 19993101 | 495 | 91 | 41292 | -0.82571 | 0.79551 | *RIN2* |
| ENSG00000170632 | 7 | 102705328 | 102750205 | 53 | 19 | 46437 | -0.82685 | 0.79584 | *ARMC10* |
| ENSG00000173218 | 1 | 116174574 | 116250845 | 274 | 43 | 44612 | -0.82739 | 0.79599 | *VANGL1* |
| ENSG00000111237 | 12 | 110918902 | 110949922 | 72 | 12 | 33434 | -0.82773 | 0.79609 | *VPS29* |
| ENSG00000267884 | 1 | 166016679 | 166038709 | 74 | 15 | 51193 | -0.82809 | 0.79619 | *AL626787.1* |

| | | | | | | | | | |
|---|---|---|---|---|---|---|---|---|---|
| ENSG00000186439 | 6 | 123527483 | 123968238 | 1492 | 116 | 42496 | -0.82816 | 0.79621 | *TRDN* |
| ENSG00000174885 | 11 | 268365 | 295359 | 59 | 24 | 38117 | -0.82835 | 0.79626 | *NLRP6* |
| ENSG00000135111 | 12 | 115098059 | 115131969 | 82 | 27 | 50124 | -0.82846 | 0.7963 | *TBX3* |
| ENSG00000080293 | 2 | 120187419 | 120292070 | 398 | 54 | 48199 | -0.82859 | 0.79633 | *SCTR* |
| ENSG00000164880 | 7 | 1499913 | 1555489 | 174 | 11 | 34584 | -0.82892 | 0.79642 | *INTS1* |
| ENSG00000119913 | 10 | 114033493 | 114074793 | 138 | 25 | 43856 | -0.8289 | 0.79642 | *TECTB* |
| ENSG00000080007 | 6 | 74094471 | 74137292 | 89 | 20 | 53824 | -0.82913 | 0.79649 | *DDX43* |
| ENSG00000178586 | 2 | 240974494 | 240995489 | 120 | 21 | 50739 | -0.8294 | 0.79656 | *OR6B3* |
| ENSG00000175582 | 11 | 73376683 | 73482182 | 308 | 32 | 48015 | -0.82944 | 0.79657 | *RAB6A* |
| ENSG00000178585 | 1 | 9898334 | 9980394 | 117 | 30 | 44502 | -0.82947 | 0.79658 | *CTNNBIP1* |
| ENSG00000214645 | 9 | 110529471 | 110550419 | 72 | 14 | 48894 | -0.82945 | 0.79658 | *AL162389.1* |
| ENSG00000095303 | 9 | 125122824 | 125167982 | 146 | 26 | 40767 | -0.82962 | 0.79662 | *PTGS1* |
| ENSG00000114251 | 3 | 55489743 | 55533973 | 130 | 27 | 38329 | -0.82977 | 0.79667 | *WNT5A* |
| ENSG00000189298 | 6 | 28307691 | 28346947 | 166 | 22 | 40777 | -0.83062 | 0.79691 | *ZKSCAN3* |
| ENSG00000269633 | 2 | 207210463 | 207230843 | 54 | 10 | 49890 | -0.83066 | 0.79692 | *AC017081.1* |
| ENSG00000183615 | 1 | 32702834 | 32724457 | 42 | 7 | 28955 | -0.83103 | 0.79702 | *FAM167B* |
| ENSG00000180440 | 13 | 37238049 | 37281976 | 93 | 30 | 49075 | -0.83101 | 0.79702 | *SERTM1* |
| ENSG00000136436 | 17 | 46898350 | 46953884 | 168 | 14 | 25883 | -0.83101 | 0.79702 | *CALCOCO2* |
| ENSG00000151743 | 12 | 31814071 | 31892108 | 233 | 38 | 42426 | -0.83156 | 0.79717 | *AMN1* |
| ENSG00000254858 | 19 | 18293992 | 18317758 | 83 | 14 | 50307 | -0.83188 | 0.79726 | *MPV17L2* |
| ENSG00000162650 | 1 | 110016101 | 110045426 | 46 | 10 | 45817 | -0.832 | 0.79729 | *ATXN7L2* |
| ENSG00000005194 | 16 | 57452081 | 57491440 | 137 | 24 | 54020 | -0.83209 | 0.79732 | *CIAPIN1* |
| ENSG00000168679 | 1 | 110895470 | 110943704 | 130 | 21 | 48278 | -0.83236 | 0.7974 | *SLC16A4* |
| ENSG00000185950 | 13 | 110396184 | 110448915 | 176 | 27 | 52516 | -0.83277 | 0.79751 | *IRS2* |
| ENSG00000170370 | 10 | 119291955 | 119319056 | 99 | 15 | 38266 | -0.83313 | 0.79762 | *EMX2* |
| ENSG00000108771 | 17 | 40243422 | 40274751 | 71 | 11 | 32896 | -0.8332 | 0.79763 | *DHX58* |
| ENSG00000215187 | 9 | 35551828 | 35573896 | 48 | 12 | 46609 | -0.83356 | 0.79774 | *FAM166B* |
| ENSG00000112038 | 6 | 154321631 | 154578001 | 853 | 100 | 45133 | -0.83375 | 0.79779 | *OPRM1* |
| ENSG00000197457 | 20 | 62261061 | 62294780 | 121 | 23 | 38260 | -0.83394 | 0.79784 | *STMN3* |
| ENSG00000182247 | 3 | 23234511 | 23643284 | 1592 | 52 | 51229 | -0.83433 | 0.79795 | *UBE2E2* |
| ENSG00000069974 | 15 | 55485164 | 55621311 | 542 | 57 | 39357 | -0.83464 | 0.79804 | *RAB27A* |
| ENSG00000137338 | 6 | 28239314 | 28280326 | 109 | 19 | 31334 | -0.83486 | 0.7981 | *PGBD1* |
| ENSG00000125534 | 20 | 62142077 | 62163559 | 99 | 23 | 38338 | -0.83511 | 0.79817 | *PPDPF* |
| ENSG00000188039 | 19 | 16820787 | 16938774 | 309 | 47 | 42144 | -0.83532 | 0.79823 | *NWD1* |
| ENSG00000224854 | 9 | 21957137 | 21977738 | 37 | 12 | 45650 | -0.83594 | 0.79841 | *C9orf53* |
| ENSG00000152990 | 4 | 22336694 | 22527677 | 717 | 86 | 40734 | -0.83695 | 0.79869 | *GPR125* |

| | | | | | | | | | |
|---|---|---|---|---|---|---|---|---|---|
| ENSG00000167711 | 17 | 1636130 | 1668562 | 140 | 22 | 52375 | -0.83701 | 0.79871 | *SERPINF2* |
| ENSG00000124302 | 19 | 34102861 | 34274414 | 615 | 101 | 49515 | -0.83732 | 0.79879 | *CHST8* |
| ENSG00000151665 | 2 | 46798076 | 46854258 | 149 | 20 | 38742 | -0.83783 | 0.79894 | *PIGF* |
| ENSG00000150459 | 13 | 21704653 | 21733221 | 97 | 15 | 55448 | -0.83827 | 0.79906 | *SAP18* |
| ENSG00000127529 | 19 | 15042301 | 15063260 | 108 | 18 | 44868 | -0.83861 | 0.79915 | *OR7C2* |
| ENSG00000139656 | 13 | 44707679 | 44745393 | 69 | 14 | 34163 | -0.83891 | 0.79924 | *SMIM2* |
| ENSG00000261740 | 16 | 29444551 | 29475972 | 2 | 1 | 34442 | -0.83897 | 0.79926 | *RP11-345J4.5* |
| ENSG00000162139 | 11 | 74689179 | 74739938 | 107 | 19 | 39471 | -0.83942 | 0.79938 | *NEU3* |
| ENSG00000172164 | 8 | 121537985 | 121835513 | 806 | 87 | 37454 | -0.83985 | 0.7995 | *SNTB1* |
| ENSG00000197724 | 9 | 96328689 | 96451869 | 621 | 61 | 46470 | -0.83988 | 0.79951 | *PHF2* |
| ENSG00000186280 | 11 | 94696845 | 94742682 | 114 | 20 | 44150 | -0.84016 | 0.79959 | *KDM4D* |
| ENSG00000159374 | 2 | 74775010 | 74885465 | 105 | 17 | 34671 | -0.84033 | 0.79964 | *M1AP* |
| ENSG00000167157 | 9 | 132417920 | 132494875 | 294 | 51 | 40923 | -0.84032 | 0.79964 | *PRRX2* |
| ENSG00000268162 | 12 | 57380745 | 57401335 | 36 | 8 | 53077 | -0.84037 | 0.79965 | *HBCBP* |
| ENSG00000166501 | 16 | 23837322 | 24241932 | 1340 | 172 | 48624 | -0.84042 | 0.79966 | *PRKCB* |
| ENSG00000015532 | 17 | 48413453 | 48450499 | 153 | 15 | 38999 | -0.84085 | 0.79978 | *XYLT2* |
| ENSG00000065135 | 1 | 110081233 | 110146975 | 107 | 19 | 46939 | -0.84095 | 0.79981 | *GNAI3* |
| ENSG00000215343 | 8 | 11951898 | 11983025 | 3 | 2 | 42983 | -0.84158 | 0.79999 | *ZNF705D* |
| ENSG00000165409 | 14 | 81411333 | 81622646 | 843 | 88 | 42033 | -0.84163 | 0.8 | *TSHR* |
| ENSG00000197903 | 6 | 27096073 | 27124619 | 54 | 19 | 42498 | -0.84182 | 0.80006 | *HIST1H2BK* |
| ENSG00000133107 | 13 | 38200773 | 38454562 | 942 | 114 | 48461 | -0.84252 | 0.80025 | *TRPC4* |
| ENSG00000179241 | 11 | 35955531 | 36263686 | 1168 | 150 | 46608 | -0.84266 | 0.80029 | *LDLRAD3* |
| ENSG00000072803 | 5 | 171278553 | 171443877 | 228 | 39 | 42133 | -0.84303 | 0.80039 | *FBXW11* |
| ENSG00000100804 | 14 | 23475752 | 23514439 | 74 | 18 | 40917 | -0.84301 | 0.80039 | *PSMB5* |
| ENSG00000258436 | 14 | 21048352 | 21068982 | 97 | 25 | 51931 | -0.84306 | 0.8004 | *RNASE12* |
| ENSG00000108825 | 17 | 41092543 | 41142545 | 52 | 12 | 34681 | -0.84379 | 0.80061 | *PTGES3L-AARSD1* |
| ENSG00000229809 | 16 | 30570667 | 30594055 | 17 | 9 | 40626 | -0.84384 | 0.80062 | *ZNF688* |
| ENSG00000213416 | 17 | 39269343 | 39290419 | 115 | 8 | 25948 | -0.84384 | 0.80062 | *KRTAP4-12* |
| ENSG00000168398 | 14 | 96661016 | 96720666 | 265 | 54 | 49492 | -0.84389 | 0.80063 | *BDKRB2* |
| ENSG00000196597 | 9 | 99568754 | 99647905 | 81 | 22 | 38952 | -0.84416 | 0.80071 | *ZNF782* |
| ENSG00000184999 | 11 | 62895339 | 63147190 | 844 | 54 | 43971 | -0.84415 | 0.80071 | *SLC22A10* |
| ENSG00000177694 | 3 | 174146363 | 175533428 | 4927 | 353 | 44675 | -0.84458 | 0.80083 | *NAALADL2* |
| ENSG00000113812 | 3 | 53891093 | 53926229 | 133 | 18 | 35687 | -0.84481 | 0.80089 | *ACTR8* |
| ENSG00000187701 | 1 | 248803232 | 248824185 | 29 | 9 | 44848 | -0.84502 | 0.80095 | *OR2T27* |
| ENSG00000071991 | 18 | 64158320 | 64281375 | 301 | 37 | 43591 | -0.84504 | 0.80096 | *CDH19* |
| ENSG00000182568 | 3 | 18376879 | 18497080 | 274 | 46 | 45351 | -0.84513 | 0.80098 | *SATB1* |

| | | | | | | | | | |
|---|---|---|---|---|---|---|---|---|---|
| ENSG00000182544 | 12 | 53635035 | 53658189 | 66 | 13 | 41533 | -0.84525 | 0.80102 | *MFSD5* |
| ENSG00000070961 | 12 | 89971828 | 90113077 | 263 | 31 | 49944 | -0.84607 | 0.80124 | *ATP2B1* |
| ENSG00000101290 | 20 | 5097432 | 5188533 | 273 | 32 | 46841 | -0.84616 | 0.80127 | *CDS2* |
| ENSG00000156253 | 21 | 30366705 | 30401699 | 92 | 17 | 45486 | -0.84616 | 0.80127 | *RWDD2B* |
| ENSG00000228716 | 5 | 79912047 | 79960802 | 276 | 10 | 54887 | -0.84655 | 0.80138 | *DHFR* |
| ENSG00000144583 | 2 | 217112588 | 217246750 | 361 | 54 | 40132 | -0.84802 | 0.80179 | *Mar-04* |
| ENSG00000112486 | 6 | 167515295 | 167563184 | 197 | 38 | 45448 | -0.84826 | 0.80185 | *CCR6* |
| ENSG00000167769 | 19 | 6296153 | 6343640 | 288 | 31 | 42526 | -0.84922 | 0.80212 | *ACER1* |
| ENSG00000136628 | 1 | 220131943 | 220230000 | 375 | 23 | 48431 | -0.84932 | 0.80215 | *EPRS* |
| ENSG00000188994 | 6 | 87852551 | 87983914 | 389 | 23 | 35621 | -0.84939 | 0.80217 | *ZNF292* |
| ENSG00000123191 | 13 | 52496809 | 52595630 | 267 | 30 | 40587 | -0.84943 | 0.80218 | *ATP7B* |
| ENSG00000212938 | 21 | 31954759 | 31975394 | 43 | 10 | 53959 | -0.84945 | 0.80218 | *KRTAP6-3* |
| ENSG00000187736 | 2 | 219930039 | 220035587 | 204 | 18 | 38243 | -0.84946 | 0.80219 | *NHEJ1* |
| ENSG00000127870 | 13 | 26696253 | 26806791 | 432 | 59 | 49179 | -0.84998 | 0.80233 | *RNF6* |
| ENSG00000196876 | 12 | 51974050 | 52216648 | 472 | 47 | 33188 | -0.85009 | 0.80236 | *SCN8A* |
| ENSG00000237441 | 6 | 33249431 | 33277101 | 79 | 19 | 31631 | -0.85016 | 0.80238 | *RGL2* |
| ENSG00000175329 | 22 | 35452129 | 35493380 | 222 | 17 | 40218 | -0.85024 | 0.8024 | *ISX* |
| ENSG00000118246 | 2 | 207620081 | 207667233 | 138 | 18 | 34775 | -0.8503 | 0.80242 | *FASTKD2* |
| ENSG00000124783 | 6 | 7258539 | 7357679 | 601 | 57 | 43267 | -0.85041 | 0.80245 | *SSR1* |
| ENSG00000180730 | 13 | 26608735 | 26635169 | 138 | 22 | 36667 | -0.85048 | 0.80247 | *SHISA2* |
| ENSG00000267060 | 17 | 41110105 | 41142545 | 40 | 10 | 33650 | -0.85087 | 0.80258 | *PTGES3L* |
| ENSG00000269690 | 1 | 201582013 | 201602738 | 43 | 15 | 42262 | -0.85091 | 0.80259 | *AC096677.1* |
| ENSG00000165643 | 9 | 138575253 | 138601374 | 122 | 22 | 33364 | -0.85109 | 0.80264 | *SOHLH1* |
| ENSG00000249319 | 7 | 65542756 | 65627228 | 258 | 21 | 44323 | -0.85114 | 0.80265 | *AC068533.7* |
| ENSG00000145687 | 5 | 80698840 | 81057616 | 817 | 79 | 38971 | -0.85125 | 0.80268 | *SSBP2* |
| ENSG00000075131 | 15 | 66618544 | 66689084 | 232 | 33 | 50563 | -0.85123 | 0.80268 | *TIPIN* |
| ENSG00000134056 | 5 | 68503587 | 68535956 | 109 | 11 | 52919 | -0.85136 | 0.80271 | *MRPS36* |
| ENSG00000146267 | 6 | 99709045 | 99807938 | 381 | 44 | 43325 | -0.85179 | 0.80284 | *FAXC* |
| ENSG00000004399 | 3 | 129264018 | 129335661 | 234 | 35 | 42624 | -0.85193 | 0.80287 | *PLXND1* |
| ENSG00000100490 | 14 | 50786310 | 50893179 | 307 | 41 | 49391 | -0.85193 | 0.80287 | *CDKL1* |
| ENSG00000105642 | 19 | 18052102 | 18120889 | 191 | 41 | 48153 | -0.85207 | 0.80291 | *KCNN1* |
| ENSG00000138772 | 4 | 79462673 | 79541597 | 225 | 33 | 41825 | -0.85249 | 0.80303 | *ANXA3* |
| ENSG00000146282 | 6 | 88214096 | 88309721 | 375 | 27 | 54438 | -0.85256 | 0.80305 | *RARS2* |
| ENSG00000019991 | 7 | 81318322 | 81409754 | 198 | 24 | 51420 | -0.85279 | 0.80311 | *HGF* |
| ENSG00000006025 | 17 | 45874738 | 45909200 | 128 | 21 | 34677 | -0.85276 | 0.80311 | *OSBPL7* |
| ENSG00000151131 | 12 | 105370088 | 105453515 | 353 | 35 | 39106 | -0.85296 | 0.80316 | *C12orf45* |

| | | | | | | | | | |
|---|---|---|---|---|---|---|---|---|---|
| ENSG00000269190 | 19 | 39422044 | 39476550 | 191 | 31 | 44691 | -0.853 | 0.80317 | *FBXO17* |
| ENSG00000158604 | 7 | 44607493 | 44631886 | 78 | 13 | 32594 | -0.85306 | 0.80319 | *TMED4* |
| ENSG00000138379 | 2 | 190910423 | 190937455 | 62 | 9 | 27576 | -0.8531 | 0.8032 | *MSTN* |
| ENSG00000268467 | 11 | 107452471 | 107473949 | 70 | 9 | 29483 | -0.85369 | 0.80336 | *AP000889.3* |
| ENSG00000253251 | 5 | 64910592 | 64936718 | 106 | 12 | 51296 | -0.85398 | 0.80344 | *CTC-534A2.2* |
| ENSG00000177459 | 8 | 99066539 | 99115838 | 132 | 33 | 48096 | -0.85399 | 0.80345 | *C8orf47* |
| ENSG00000179335 | 15 | 74880841 | 74942057 | 88 | 13 | 33970 | -0.8544 | 0.80356 | *CLK3* |
| ENSG00000151093 | 3 | 25814408 | 25846025 | 39 | 9 | 39108 | -0.85501 | 0.80373 | *OXSM* |
| ENSG00000221826 | 19 | 43215790 | 43254721 | 153 | 18 | 45798 | -0.85529 | 0.8038 | *PSG3* |
| ENSG00000197162 | 16 | 30575061 | 30607092 | 27 | 11 | 33672 | -0.85592 | 0.80398 | *ZNF785* |
| ENSG00000198863 | 17 | 41122582 | 41155707 | 44 | 9 | 39609 | -0.85613 | 0.80404 | *RUNDC1* |
| ENSG00000152292 | 2 | 85635844 | 85674152 | 142 | 29 | 44177 | -0.85662 | 0.80417 | *SH2D6* |
| ENSG00000122545 | 7 | 35830542 | 35954917 | 242 | 19 | 44160 | -0.85741 | 0.80439 | *Sep-07* |
| ENSG00000131016 | 6 | 151551134 | 151689692 | 553 | 92 | 41602 | -0.85755 | 0.80443 | *AKAP12* |
| ENSG00000104343 | 8 | 74682332 | 74801145 | 188 | 31 | 46763 | -0.85756 | 0.80443 | *UBE2W* |
| ENSG00000141837 | 19 | 13307256 | 13744804 | 1624 | 239 | 40088 | -0.85776 | 0.80449 | *CACNA1A* |
| ENSG00000132031 | 2 | 20181872 | 20222455 | 113 | 22 | 45294 | -0.85785 | 0.80451 | *MATN3* |
| ENSG00000185864 | 16 | 21835890 | 21902148 | 16 | 8 | 47861 | -0.85794 | 0.80454 | *NPIPB4* |
| ENSG00000163235 | 2 | 70664412 | 70791325 | 443 | 65 | 49271 | -0.85872 | 0.80475 | *TGFA* |
| ENSG00000204839 | 8 | 144638357 | 144665141 | 131 | 23 | 49586 | -0.8588 | 0.80477 | *MROH6* |
| ENSG00000101605 | 18 | 3056805 | 3230106 | 771 | 125 | 46201 | -0.85886 | 0.80479 | *MYOM1* |
| ENSG00000164326 | 5 | 71004990 | 71026875 | 81 | 15 | 42344 | -0.85948 | 0.80496 | *CARTPT* |
| ENSG00000082269 | 6 | 71112644 | 71280877 | 402 | 20 | 48665 | -0.85972 | 0.80503 | *FAM135A* |
| ENSG00000172869 | 5 | 118363467 | 118594833 | 403 | 67 | 46221 | -0.85998 | 0.8051 | *DMXL1* |
| ENSG00000183038 | 22 | 20356206 | 20378028 | 1 | 1 | 42968 | -0.85998 | 0.8051 | *GGTLC3* |
| ENSG00000198815 | 1 | 42632210 | 42811548 | 372 | 24 | 44974 | -0.8601 | 0.80513 | *FOXJ3* |
| ENSG00000144867 | 3 | 133492877 | 133554616 | 181 | 30 | 51104 | -0.86008 | 0.80513 | *SRPRB* |
| ENSG00000143507 | 1 | 221864766 | 221925518 | 138 | 33 | 44330 | -0.86015 | 0.80515 | *DUSP10* |
| ENSG00000215271 | 14 | 23731666 | 23778656 | 110 | 29 | 45049 | -0.8603 | 0.80519 | *HOMEZ* |
| ENSG00000137478 | 11 | 72537790 | 72863306 | 382 | 63 | 49970 | -0.86077 | 0.80532 | *FCHSD2* |
| ENSG00000106701 | 9 | 108200077 | 108324714 | 302 | 37 | 45700 | -0.86093 | 0.80536 | *FSD1L* |
| ENSG00000138190 | 10 | 94580935 | 94829250 | 718 | 53 | 40784 | -0.86093 | 0.80536 | *EXOC6* |
| ENSG00000128683 | 2 | 171659723 | 171727661 | 201 | 31 | 46151 | -0.86102 | 0.80539 | *GAD1* |
| ENSG00000146540 | 7 | 1026622 | 1187896 | 906 | 78 | 45114 | -0.86111 | 0.80541 | *C7orf50* |
| ENSG00000163565 | 1 | 158959758 | 159034945 | 264 | 27 | 39890 | -0.86193 | 0.80564 | *IFI16* |
| ENSG00000138738 | 4 | 121596074 | 121854025 | 1130 | 62 | 47605 | -0.8625 | 0.80579 | *PRDM5* |

| | | | | | | | | | |
|---|---|---|---|---|---|---|---|---|---|
| ENSG00000120690 | 13 | 41496056 | 41645576 | 340 | 30 | 36344 | -0.86272 | 0.80585 | *ELF1* |
| ENSG00000066382 | 11 | 30396040 | 30618419 | 607 | 73 | 41751 | -0.86354 | 0.80608 | *MPPED2* |
| ENSG00000110448 | 11 | 60859867 | 60905324 | 118 | 20 | 34527 | -0.86365 | 0.80611 | *CD5* |
| ENSG00000102543 | 13 | 49812047 | 49877618 | 120 | 10 | 45922 | -0.86414 | 0.80625 | *CDADC1* |
| ENSG00000157064 | 1 | 183207372 | 183397737 | 549 | 51 | 43870 | -0.8645 | 0.80634 | *NMNAT2* |
| ENSG00000114948 | 2 | 207298263 | 207495851 | 463 | 67 | 53524 | -0.86457 | 0.80636 | *ADAM23* |
| ENSG00000122484 | 1 | 92754522 | 92877613 | 164 | 19 | 27255 | -0.86486 | 0.80644 | *RPAP2* |
| ENSG00000115919 | 2 | 143625067 | 143809890 | 544 | 59 | 35801 | -0.86532 | 0.80657 | *KYNU* |
| ENSG00000197324 | 14 | 23330822 | 23360789 | 57 | 14 | 48438 | -0.86546 | 0.80661 | *LRP10* |
| ENSG00000073737 | 2 | 169911299 | 169962677 | 76 | 19 | 47467 | -0.86563 | 0.80665 | *DHRS9* |
| ENSG00000121350 | 12 | 21580549 | 21633300 | 350 | 18 | 53454 | -0.86586 | 0.80672 | *PYROXD1* |
| ENSG00000172661 | 10 | 46212648 | 46298409 | 99 | 12 | 36855 | -0.86655 | 0.80691 | *FAM21C* |
| ENSG00000167034 | 8 | 23526206 | 23550440 | 77 | 15 | 46611 | -0.86699 | 0.80703 | *NKX3-1* |
| ENSG00000106351 | 7 | 100126834 | 100175842 | 78 | 12 | 51859 | -0.86703 | 0.80704 | *AGFG2* |
| ENSG00000088356 | 20 | 30522758 | 30549895 | 42 | 7 | 39239 | -0.86776 | 0.80724 | *PDRG1* |
| ENSG00000126106 | 1 | 45090910 | 45150227 | 125 | 13 | 35937 | -0.8679 | 0.80728 | *TMEM53* |
| ENSG00000167311 | 11 | 3649733 | 3673546 | 105 | 25 | 34221 | -0.86794 | 0.80729 | *ART5* |
| ENSG00000085662 | 7 | 134117102 | 134154036 | 102 | 26 | 41797 | -0.86871 | 0.8075 | *AKR1B1* |
| ENSG00000184156 | 8 | 133123108 | 133503200 | 1456 | 162 | 47503 | -0.86896 | 0.80757 | *KCNQ3* |
| ENSG00000215547 | 20 | 29835467 | 29857435 | 65 | 13 | 40440 | -0.87005 | 0.80786 | *DEFB115* |
| ENSG00000075568 | 2 | 98362799 | 98622388 | 600 | 41 | 44437 | -0.8701 | 0.80788 | *TMEM131* |
| ENSG00000168658 | 2 | 98693579 | 98939762 | 725 | 48 | 44795 | -0.87031 | 0.80793 | *VWA3B* |
| ENSG00000135517 | 12 | 56833286 | 56872950 | 76 | 19 | 45338 | -0.87036 | 0.80795 | *MIP* |
| ENSG00000203972 | 6 | 49457671 | 49504760 | 133 | 14 | 42073 | -0.87115 | 0.80816 | *GLYATL3* |
| ENSG00000151360 | 2 | 3695785 | 3760261 | 274 | 41 | 47708 | -0.87124 | 0.80819 | *ALLC* |
| ENSG00000184402 | 20 | 60708822 | 60767540 | 265 | 24 | 33484 | -0.87128 | 0.8082 | *SS18L1* |
| ENSG00000078237 | 12 | 4420371 | 4472338 | 136 | 24 | 46542 | -0.87154 | 0.80827 | *C12orf5* |
| ENSG00000167900 | 17 | 76160160 | 76193314 | 167 | 31 | 34241 | -0.87153 | 0.80827 | *TK1* |
| ENSG00000156738 | 11 | 60213225 | 60248233 | 105 | 20 | 50942 | -0.87156 | 0.80828 | *MS4A1* |
| ENSG00000050730 | 4 | 122042563 | 122158621 | 360 | 36 | 46098 | -0.87171 | 0.80832 | *TNIP3* |
| ENSG00000099817 | 19 | 1076594 | 1105598 | 167 | 40 | 45318 | -0.87184 | 0.80835 | *POLR2E* |
| ENSG00000156603 | 11 | 57461186 | 57489693 | 51 | 9 | 45185 | -0.87228 | 0.80847 | *MED19* |
| ENSG00000106397 | 7 | 100839258 | 100871701 | 83 | 23 | 45549 | -0.87235 | 0.80849 | *PLOD3* |
| ENSG00000134490 | 18 | 20767108 | 21027925 | 584 | 75 | 42164 | -0.87235 | 0.80849 | *TMEM241* |
| ENSG00000139287 | 12 | 72322626 | 72590398 | 738 | 70 | 42749 | -0.87253 | 0.80854 | *TPH2* |
| ENSG00000186803 | 9 | 21196180 | 21217142 | 155 | 15 | 50248 | -0.87277 | 0.80861 | *IFNA10* |

| | | | | | | | | | |
|---|---|---|---|---|---|---|---|---|---|
| ENSG00000198121 | 9 | 113625543 | 113810981 | 615 | 76 | 42463 | -0.87287 | 0.80863 | *LPAR1* |
| ENSG00000158485 | 1 | 158287741 | 158311321 | 67 | 15 | 38339 | -0.87302 | 0.80867 | *CD1B* |
| ENSG00000212933 | 21 | 46064130 | 46084576 | 105 | 13 | 49952 | -0.87323 | 0.80873 | *KRTAP12-4* |
| ENSG00000161638 | 12 | 54779045 | 54823244 | 84 | 13 | 30092 | -0.87349 | 0.8088 | *ITGA5* |
| ENSG00000186866 | 21 | 46673843 | 46717813 | 180 | 15 | 51289 | -0.87362 | 0.80884 | *POFUT2* |
| ENSG00000082497 | 1 | 210396144 | 210429976 | 59 | 14 | 48648 | -0.87372 | 0.80887 | *SERTAD4* |
| ENSG00000203832 | 1 | 148240249 | 148357506 | 6 | 3 | 42605 | -0.87416 | 0.80898 | *NBPF20* |
| ENSG00000232560 | 21 | 18801208 | 18831503 | 114 | 42 | 44983 | -0.87418 | 0.80899 | *C21orf37* |
| ENSG00000158186 | 3 | 138056539 | 138134375 | 197 | 19 | 53687 | -0.8744 | 0.80905 | *MRAS* |
| ENSG00000173598 | 12 | 93761659 | 93807024 | 132 | 23 | 50689 | -0.87467 | 0.80912 | *NUDT4* |
| ENSG00000143768 | 1 | 226114298 | 226139189 | 56 | 16 | 50170 | -0.87493 | 0.80919 | *LEFTY2* |
| ENSG00000132623 | 20 | 9956774 | 10047410 | 369 | 50 | 51100 | -0.87503 | 0.80922 | *ANKEF1* |
| ENSG00000136872 | 9 | 104172860 | 104208105 | 127 | 19 | 51862 | -0.87551 | 0.80935 | *ALDOB* |
| ENSG00000138182 | 10 | 91451367 | 91544700 | 353 | 18 | 33480 | -0.87552 | 0.80935 | *KIF20B* |
| ENSG00000130147 | 2 | 235850617 | 235974358 | 325 | 77 | 40532 | -0.87552 | 0.80936 | *SH3BP4* |
| ENSG00000182952 | 6 | 26528633 | 26556482 | 77 | 9 | 52064 | -0.87603 | 0.80949 | *HMGN4* |
| ENSG00000215545 | 20 | 29881015 | 29906388 | 85 | 16 | 42508 | -0.87655 | 0.80963 | *DEFB116* |
| ENSG00000151458 | 4 | 125575207 | 125643887 | 166 | 27 | 53493 | -0.87662 | 0.80965 | *ANKRD50* |
| ENSG00000164325 | 5 | 72459022 | 72480970 | 52 | 16 | 47807 | -0.87697 | 0.80975 | *TMEM174* |
| ENSG00000161533 | 17 | 73927588 | 73985515 | 214 | 30 | 45240 | -0.87726 | 0.80983 | *ACOX1* |
| ENSG00000160593 | 11 | 118054455 | 118105809 | 161 | 16 | 39048 | -0.87736 | 0.80985 | *AMICA1* |
| ENSG00000124217 | 20 | 49565363 | 49587820 | 39 | 11 | 41350 | -0.87763 | 0.80993 | *MOCS3* |
| ENSG00000172215 | 3 | 45972425 | 45999845 | 76 | 12 | 31560 | -0.87775 | 0.80996 | *CXCR6* |
| ENSG00000124762 | 6 | 36634305 | 36665116 | 121 | 16 | 35314 | -0.87819 | 0.81008 | *CDKN1A* |
| ENSG00000051825 | 12 | 123626867 | 123738561 | 342 | 20 | 34600 | -0.87866 | 0.81021 | *MPHOSPH9* |
| ENSG00000162244 | 3 | 52017616 | 52039958 | 29 | 14 | 44995 | -0.8789 | 0.81027 | *RPL29* |
| ENSG00000224470 | 16 | 71869894 | 71929171 | 131 | 19 | 44975 | -0.87904 | 0.81031 | *ATXN1L* |
| ENSG00000178789 | 17 | 72507313 | 72537613 | 111 | 13 | 40801 | -0.87906 | 0.81031 | *CD300LB* |
| ENSG00000115239 | 2 | 53749810 | 54097170 | 1272 | 119 | 46037 | -0.87924 | 0.81036 | *GPR75-ASB3* |
| ENSG00000092820 | 6 | 159176773 | 159250444 | 331 | 24 | 31340 | -0.87937 | 0.8104 | *EZR* |
| ENSG00000177047 | 9 | 21130631 | 21152144 | 75 | 18 | 49736 | -0.87946 | 0.81042 | *IFNW1* |
| ENSG00000189180 | 10 | 38289578 | 38364016 | 233 | 22 | 54802 | -0.87962 | 0.81047 | *ZNF33A* |
| ENSG00000143028 | 1 | 109999180 | 110034759 | 93 | 9 | 45442 | -0.88042 | 0.81068 | *SYPL2* |
| ENSG00000159692 | 4 | 1195236 | 1253741 | 191 | 25 | 32064 | -0.88081 | 0.81079 | *CTBP1* |
| ENSG00000104892 | 19 | 45826692 | 45864778 | 123 | 24 | 42207 | -0.88112 | 0.81087 | *KLC3* |
| ENSG00000256660 | 12 | 10153226 | 10181218 | 121 | 18 | 51047 | -0.88196 | 0.8111 | *CLEC12B* |

| | | | | | | | | | |
|---|---|---|---|---|---|---|---|---|---|
| ENSG00000267699 | 18 | 48484389 | 48594514 | 143 | 17 | 51856 | -0.88221 | 0.81117 | *RP11-729L2.2* |
| ENSG00000080824 | 14 | 102537075 | 102616036 | 227 | 28 | 37479 | -0.88241 | 0.81122 | *HSP90AA1* |
| ENSG00000156469 | 8 | 97241626 | 97283838 | 81 | 17 | 49898 | -0.88272 | 0.81131 | *MTERFD1* |
| ENSG00000066027 | 1 | 212448879 | 212545200 | 278 | 33 | 45778 | -0.88291 | 0.81136 | *PPP2R5A* |
| ENSG00000169994 | 2 | 128283378 | 128405304 | 389 | 54 | 41511 | -0.88341 | 0.81149 | *MYO7B* |
| ENSG00000183770 | 3 | 138653066 | 138675982 | 35 | 11 | 40130 | -0.88416 | 0.81169 | *FOXL2* |
| ENSG00000148841 | 10 | 106061894 | 106108162 | 92 | 16 | 42942 | -0.88461 | 0.81182 | *ITPRIP* |
| ENSG00000180861 | 12 | 13514023 | 13550101 | 142 | 15 | 52306 | -0.88482 | 0.81187 | *C12orf36* |
| ENSG00000174672 | 11 | 1401129 | 1493919 | 459 | 62 | 45597 | -0.88502 | 0.81193 | *BRSK2* |
| ENSG00000141934 | 19 | 271040 | 301393 | 83 | 31 | 40088 | -0.88504 | 0.81193 | *PPAP2C* |
| ENSG00000156042 | 10 | 75003517 | 75128617 | 367 | 6 | 19417 | -0.88537 | 0.81202 | *TTC18* |
| ENSG00000273045 | 2 | 99747948 | 99777950 | 93 | 11 | 45283 | -0.88544 | 0.81204 | *C2ORF15* |
| ENSG00000230522 | 19 | 7039332 | 7061746 | 5 | 3 | 43029 | -0.88576 | 0.81213 | *MBD3L2* |
| ENSG00000181481 | 17 | 29285803 | 29336929 | 30 | 20 | 42761 | -0.88599 | 0.81219 | *RNF135* |
| ENSG00000152894 | 6 | 128279924 | 128851870 | 1121 | 117 | 45944 | -0.88604 | 0.8122 | *PTPRK* |
| ENSG00000151208 | 10 | 79540549 | 79696378 | 490 | 37 | 53866 | -0.88676 | 0.8124 | *DLG5* |
| ENSG00000184471 | 16 | 1130005 | 1156244 | 127 | 28 | 41659 | -0.88711 | 0.81249 | *C1QTNF8* |
| ENSG00000253368 | 1 | 27310198 | 27337389 | 37 | 9 | 48631 | -0.8873 | 0.81254 | *TRNP1* |
| ENSG00000131068 | 20 | 29946421 | 29971726 | 62 | 15 | 42421 | -0.88745 | 0.81258 | *DEFB118* |
| ENSG00000170144 | 2 | 178067291 | 178098686 | 39 | 8 | 36869 | -0.88758 | 0.81262 | *HNRNPA3* |
| ENSG00000126883 | 9 | 133990948 | 134120057 | 304 | 30 | 52838 | -0.88781 | 0.81268 | *NUP214* |
| ENSG00000187140 | 1 | 63778730 | 63800797 | 63 | 25 | 52874 | -0.88833 | 0.81282 | *FOXD3* |
| ENSG00000123219 | 5 | 64803593 | 64868998 | 250 | 16 | 53169 | -0.88846 | 0.81285 | *CENPK* |
| ENSG00000101363 | 20 | 35908041 | 35955663 | 126 | 19 | 48773 | -0.88849 | 0.81286 | *MANBAL* |
| ENSG00000174448 | 18 | 51840728 | 51894334 | 191 | 15 | 46478 | -0.88863 | 0.8129 | *STARD6* |
| ENSG00000142655 | 1 | 10522345 | 10700815 | 482 | 52 | 44510 | -0.88875 | 0.81293 | *PEX14* |
| ENSG00000166922 | 15 | 32923877 | 32999299 | 253 | 67 | 44112 | -0.88907 | 0.81302 | *SCG5* |
| ENSG00000170442 | 12 | 52633084 | 52712947 | 346 | 40 | 52609 | -0.8891 | 0.81303 | *KRT86* |
| ENSG00000119912 | 10 | 94201441 | 94343833 | 403 | 42 | 51858 | -0.88916 | 0.81304 | *IDE* |
| ENSG00000125885 | 20 | 5921298 | 5985852 | 248 | 33 | 40459 | -0.88927 | 0.81307 | *MCM8* |
| ENSG00000110876 | 12 | 109005686 | 109037735 | 75 | 25 | 42007 | -0.88939 | 0.8131 | *SELPLG* |
| ENSG00000179361 | 15 | 74823518 | 74900472 | 139 | 13 | 29945 | -0.88958 | 0.81316 | *ARID3B* |
| ENSG00000196539 | 1 | 248626627 | 248647634 | 28 | 3 | 45040 | -0.88986 | 0.81323 | *OR2T3* |
| ENSG00000188306 | 3 | 169529710 | 169565563 | 99 | 13 | 50827 | -0.88994 | 0.81325 | *LRRIQ4* |
| ENSG00000007080 | 19 | 18033825 | 18064800 | 94 | 23 | 44787 | -0.89019 | 0.81332 | *CCDC124* |
| ENSG00000102710 | 13 | 37573449 | 37643850 | 120 | 15 | 42384 | -0.89053 | 0.81341 | *SUPT20H* |

| | | | | | | | | | |
|---|---|---|---|---|---|---|---|---|---|
| ENSG00000167995 | 11 | 61707293 | 61742987 | 84 | 22 | 29764 | -0.89091 | 0.81351 | *BEST1* |
| ENSG00000135451 | 12 | 49707019 | 49735514 | 96 | 15 | 48100 | -0.89137 | 0.81363 | *TROAP* |
| ENSG00000232850 | 9 | 130880612 | 130901481 | 28 | 8 | 27136 | -0.89149 | 0.81367 | *AL590708.2* |
| ENSG00000196363 | 9 | 136990487 | 137035093 | 314 | 31 | 43947 | -0.89149 | 0.81367 | *WDR5* |
| ENSG00000132024 | 19 | 14007014 | 14051692 | 80 | 13 | 34744 | -0.89163 | 0.81371 | *CC2D1A* |
| ENSG00000090020 | 1 | 27415306 | 27503472 | 134 | 10 | 54007 | -0.89181 | 0.81375 | *SLC9A1* |
| ENSG00000164603 | 7 | 112449202 | 112589971 | 259 | 35 | 44084 | -0.89179 | 0.81375 | *C7orf60* |
| ENSG00000268206 | 17 | 26628667 | 26652323 | 39 | 13 | 44926 | -0.89217 | 0.81385 | *AC061975.10* |
| ENSG00000198796 | 18 | 56138479 | 56306189 | 694 | 103 | 49118 | -0.89232 | 0.81389 | *ALPK2* |
| ENSG00000102802 | 13 | 31470328 | 31509709 | 134 | 27 | 53706 | -0.89342 | 0.81418 | *MEDAG* |
| ENSG00000172146 | 17 | 3108915 | 3129844 | 152 | 12 | 51330 | -0.89371 | 0.81426 | *OR1A1* |
| ENSG00000113595 | 5 | 64875507 | 64931802 | 199 | 16 | 50201 | -0.89401 | 0.81434 | *TRIM23* |
| ENSG00000112299 | 6 | 132992729 | 133045188 | 255 | 28 | 37587 | -0.89408 | 0.81436 | *VNN1* |
| ENSG00000130943 | 22 | 46641560 | 46669219 | 117 | 10 | 26188 | -0.89413 | 0.81437 | *PKDREJ* |
| ENSG00000167996 | 11 | 61717190 | 61745132 | 71 | 18 | 29792 | -0.89522 | 0.81467 | *FTH1* |
| ENSG00000177425 | 12 | 79958759 | 80094877 | 121 | 26 | 46553 | -0.89529 | 0.81468 | *PAWR* |
| ENSG00000075702 | 19 | 36535783 | 36606008 | 212 | 28 | 54346 | -0.89531 | 0.81469 | *WDR62* |
| ENSG00000156170 | 8 | 95897995 | 96138683 | 822 | 67 | 48040 | -0.89558 | 0.81476 | *NDUFAF6* |
| ENSG00000165300 | 13 | 88314870 | 88341871 | 69 | 14 | 46580 | -0.89679 | 0.81508 | *SLITRK5* |
| ENSG00000103226 | 16 | 16316352 | 16398668 | 78 | 9 | 43381 | -0.89683 | 0.8151 | *NOMO3* |
| ENSG00000116918 | 1 | 231654399 | 231712270 | 105 | 23 | 42315 | -0.89689 | 0.81511 | *TSNAX* |
| ENSG00000113407 | 5 | 33430802 | 33479644 | 152 | 20 | 36204 | -0.89696 | 0.81513 | *TARS* |
| ENSG00000118520 | 6 | 131884284 | 131915472 | 62 | 16 | 38720 | -0.89742 | 0.81525 | *ARG1* |
| ENSG00000170860 | 3 | 14209858 | 14252619 | 87 | 17 | 43975 | -0.89773 | 0.81534 | *LSM3* |
| ENSG00000268028 | 12 | 65080189 | 65100329 | 53 | 15 | 49808 | -0.89854 | 0.81555 | *AC025262.1* |
| ENSG00000147573 | 8 | 67029131 | 67097720 | 168 | 18 | 35530 | -0.89866 | 0.81558 | *TRIM55* |
| ENSG00000090975 | 12 | 123458027 | 123644562 | 249 | 37 | 43231 | -0.89885 | 0.81563 | *PITPNM2* |
| ENSG00000257726 | 12 | 115790817 | 115812550 | 55 | 12 | 44519 | -0.89972 | 0.81587 | *RP11-116D17.1* |
| ENSG00000256771 | 19 | 19966695 | 20015483 | 177 | 22 | 33440 | -0.90007 | 0.81596 | *ZNF253* |
| ENSG00000099795 | 19 | 14666890 | 14692874 | 74 | 16 | 49710 | -0.90018 | 0.81599 | *NDUFB7* |
| ENSG00000083838 | 19 | 58975384 | 59002597 | 61 | 13 | 49606 | -0.90035 | 0.81603 | *ZNF446* |
| ENSG00000077327 | 10 | 22624399 | 22753153 | 130 | 35 | 44865 | -0.90046 | 0.81606 | *SPAG6* |
| ENSG00000159763 | 7 | 142819170 | 142846839 | 79 | 13 | 27007 | -0.90072 | 0.81613 | *PIP* |
| ENSG00000170803 | 11 | 6796248 | 6817245 | 96 | 12 | 53568 | -0.90145 | 0.81633 | *OR2AG1* |
| ENSG00000162426 | 1 | 8367886 | 8414227 | 141 | 26 | 46497 | -0.90157 | 0.81636 | *SLC45A1* |
| ENSG00000105193 | 19 | 39913847 | 39936588 | 73 | 9 | 41111 | -0.90174 | 0.8164 | *RPS16* |

| | | | | | | | | | |
|---|---|---|---|---|---|---|---|---|---|
| ENSG00000108187 | 10 | 70032417 | 70102806 | 255 | 21 | 39987 | -0.90176 | 0.81641 | *PBLD* |
| ENSG00000180777 | 18 | 14718271 | 14862737 | 367 | 35 | 45237 | -0.90244 | 0.81659 | *ANKRD30B* |
| ENSG00000174373 | 14 | 35997558 | 36288510 | 400 | 30 | 37728 | -0.90248 | 0.8166 | *RALGAPA1* |
| ENSG00000184613 | 12 | 44892058 | 45325631 | 1151 | 98 | 47116 | -0.90267 | 0.81665 | *NELL2* |
| ENSG00000188386 | 9 | 104343897 | 104367283 | 70 | 16 | 53205 | -0.90273 | 0.81667 | *PPP3R2* |
| ENSG00000111728 | 12 | 22206707 | 22599975 | 1263 | 119 | 45445 | -0.90326 | 0.81681 | *ST8SIA1* |
| ENSG00000164985 | 9 | 15454064 | 15521017 | 206 | 28 | 36852 | -0.90349 | 0.81687 | *PSIP1* |
| ENSG00000113368 | 5 | 126102315 | 126182712 | 325 | 40 | 54101 | -0.90398 | 0.817 | *LMNB1* |
| ENSG00000133657 | 3 | 194113401 | 194229093 | 325 | 45 | 30733 | -0.9044 | 0.81711 | *ATP13A3* |
| ENSG00000113597 | 5 | 64910543 | 64972060 | 228 | 17 | 52692 | -0.90486 | 0.81723 | *TRAPPC13* |
| ENSG00000172349 | 15 | 81441916 | 81615104 | 482 | 61 | 39377 | -0.90525 | 0.81733 | *IL16* |
| ENSG00000064205 | 20 | 43333485 | 43367150 | 72 | 21 | 51777 | -0.90556 | 0.81742 | *WISP2* |
| ENSG00000240386 | 1 | 152738848 | 152759445 | 64 | 10 | 40222 | -0.90579 | 0.81748 | *LCE1F* |
| ENSG00000169245 | 4 | 76932273 | 76954650 | 106 | 6 | 52020 | -0.90609 | 0.81756 | *CXCL10* |
| ENSG00000092054 | 14 | 23871947 | 23914927 | 128 | 22 | 39709 | -0.90624 | 0.8176 | *MYH7* |
| ENSG00000102984 | 16 | 71883583 | 71939239 | 124 | 19 | 44877 | -0.90657 | 0.81768 | *ZNF821* |
| ENSG00000167468 | 19 | 1093936 | 1116787 | 119 | 24 | 47504 | -0.90661 | 0.81769 | *GPX4* |
| ENSG00000084112 | 12 | 109166466 | 109261366 | 196 | 31 | 38643 | -0.90736 | 0.81789 | *SSH1* |
| ENSG00000197771 | 10 | 121578972 | 121662068 | 325 | 20 | 35693 | -0.90793 | 0.81804 | *MCMBP* |
| ENSG00000168875 | 3 | 137473579 | 137494396 | 47 | 12 | 52442 | -0.90856 | 0.81821 | *SOX14* |
| ENSG00000113163 | 5 | 74654311 | 74817963 | 335 | 34 | 46694 | -0.9087 | 0.81825 | *COL4A3BP* |
| ENSG00000243440 | 21 | 15954251 | 16041142 | 300 | 33 | 42194 | -0.90918 | 0.81837 | *AF165138.7* |
| ENSG00000140465 | 15 | 75001883 | 75027951 | 33 | 10 | 30179 | -0.90992 | 0.81857 | *CYP1A1* |
| ENSG00000268172 | 1 | 144979309 | 145001365 | 1 | 1 | 42683 | -0.91004 | 0.8186 | *AL590452.1* |
| ENSG00000134184 | 1 | 110220436 | 110261661 | 85 | 17 | 29568 | -0.91015 | 0.81863 | *GSTM1* |
| ENSG00000155115 | 6 | 111269763 | 111299093 | 141 | 8 | 28639 | -0.91028 | 0.81866 | *GTF3C6* |
| ENSG00000122515 | 7 | 44778180 | 44819477 | 75 | 15 | 42616 | -0.91049 | 0.81872 | *ZMIZ2* |
| ENSG00000204703 | 6 | 29043985 | 29065090 | 74 | 16 | 40908 | -0.91076 | 0.81879 | *OR2B3* |
| ENSG00000182636 | 15 | 23920565 | 23942450 | 72 | 20 | 45576 | -0.9116 | 0.81901 | *NDN* |
| ENSG00000188157 | 1 | 945503 | 1001496 | 164 | 21 | 33259 | -0.91168 | 0.81903 | *AGRN* |
| ENSG00000131470 | 17 | 40714333 | 40739849 | 48 | 9 | 54469 | -0.91188 | 0.81908 | *PSMC3IP* |
| ENSG00000111249 | 12 | 111461828 | 111798358 | 419 | 73 | 36777 | -0.91191 | 0.81909 | *CUX2* |
| ENSG00000120438 | 6 | 160189530 | 160220781 | 148 | 14 | 47905 | -0.91245 | 0.81923 | *TCP1* |
| ENSG00000151657 | 10 | 7782925 | 7839990 | 208 | 37 | 46187 | -0.913 | 0.81938 | *KIN* |
| ENSG00000197951 | 19 | 57096632 | 57147849 | 180 | 39 | 44144 | -0.91302 | 0.81938 | *ZNF71* |
| ENSG00000129071 | 3 | 129139787 | 129168878 | 79 | 10 | 33647 | -0.91353 | 0.81952 | *MBD4* |

| | | | | | | | | | |
|---|---|---|---|---|---|---|---|---|---|
| ENSG00000258691 | 14 | 96661181 | 96740266 | 350 | 65 | 50277 | -0.91379 | 0.81959 | *RP11-404P21.8* |
| ENSG00000174842 | 1 | 92701959 | 92774544 | 93 | 11 | 25997 | -0.91413 | 0.81968 | *GLMN* |
| ENSG00000184220 | 3 | 99526678 | 99907447 | 550 | 44 | 53714 | -0.91473 | 0.81983 | *CMSS1* |
| ENSG00000185640 | 12 | 53205194 | 53238079 | 110 | 23 | 45469 | -0.9151 | 0.81993 | *KRT79* |
| ENSG00000023445 | 11 | 102178215 | 102220134 | 70 | 15 | 55167 | -0.91515 | 0.81994 | *BIRC3* |
| ENSG00000188227 | 19 | 37987841 | 38049664 | 143 | 26 | 35348 | -0.91549 | 0.82003 | *ZNF793* |
| ENSG00000148835 | 10 | 105117724 | 105158822 | 58 | 11 | 37305 | -0.91573 | 0.82009 | *TAF5* |
| ENSG00000054611 | 22 | 47148518 | 47581336 | 1695 | 158 | 43599 | -0.91572 | 0.82009 | *TBC1D22A* |
| ENSG00000197323 | 1 | 114925399 | 115063781 | 414 | 33 | 53514 | -0.91574 | 0.8201 | *TRIM33* |
| ENSG00000106948 | 9 | 117086436 | 117166685 | 175 | 29 | 43899 | -0.91604 | 0.82018 | *AKNA* |
| ENSG00000258744 | 14 | 24960800 | 24987894 | 121 | 19 | 42183 | -0.91614 | 0.8202 | *RP11-80A15.1* |
| ENSG00000165732 | 10 | 70705884 | 70754829 | 132 | 21 | 43965 | -0.91624 | 0.82023 | *DDX21* |
| ENSG00000114757 | 3 | 179502746 | 179764841 | 820 | 92 | 47193 | -0.91644 | 0.82028 | *PEX5L* |
| ENSG00000162604 | 1 | 62136718 | 62201095 | 171 | 23 | 44452 | -0.91661 | 0.82033 | *TM2D1* |
| ENSG00000242265 | 7 | 94275637 | 94309007 | 58 | 12 | 53391 | -0.91666 | 0.82034 | *PEG10* |
| ENSG00000113712 | 5 | 148861760 | 148941007 | 147 | 23 | 39496 | -0.9168 | 0.82038 | *CSNK1A1* |
| ENSG00000170633 | 12 | 121827844 | 121878389 | 170 | 12 | 32244 | -0.91711 | 0.82046 | *RNF34* |
| ENSG00000145545 | 5 | 6623456 | 6679675 | 240 | 32 | 50779 | -0.91779 | 0.82064 | *SRD5A1* |
| ENSG00000128645 | 2 | 177043307 | 177065688 | 38 | 10 | 50342 | -0.91798 | 0.82069 | *HOXD1* |
| ENSG00000186860 | 17 | 39461173 | 39481947 | 83 | 10 | 50494 | -0.91809 | 0.82072 | *KRTAP17-1* |
| ENSG00000239388 | 3 | 57292375 | 57336710 | 96 | 15 | 40695 | -0.91849 | 0.82082 | *ASB14* |
| ENSG00000204873 | 17 | 39378715 | 39399705 | 101 | 8 | 45992 | -0.91852 | 0.82083 | *KRTAP9-3* |
| ENSG00000119986 | 10 | 99427181 | 99457080 | 115 | 18 | 44968 | -0.91942 | 0.82106 | *AVPI1* |
| ENSG00000082293 | 6 | 70566463 | 70929679 | 1017 | 104 | 43944 | -0.9195 | 0.82108 | *COL19A1* |
| ENSG00000215695 | 1 | 15976364 | 15998217 | 55 | 6 | 48137 | -0.91972 | 0.82114 | *RSC1A1* |
| ENSG00000130768 | 1 | 28251504 | 28295668 | 157 | 19 | 40237 | -0.91994 | 0.8212 | *SMPDL3B* |
| ENSG00000132854 | 1 | 62692651 | 62795085 | 443 | 55 | 45470 | -0.91993 | 0.8212 | *KANK4* |
| ENSG00000145730 | 5 | 102079685 | 102376809 | 848 | 61 | 50175 | -0.91995 | 0.8212 | *PAM* |
| ENSG00000153395 | 5 | 1446595 | 1534092 | 329 | 61 | 44136 | -0.92095 | 0.82146 | *LPCAT1* |
| ENSG00000176907 | 8 | 40000989 | 40022821 | 58 | 13 | 38598 | -0.92123 | 0.82154 | *C8orf4* |
| ENSG00000164830 | 8 | 107272473 | 107774922 | 1546 | 125 | 46085 | -0.92125 | 0.82154 | *OXR1* |
| ENSG00000112110 | 6 | 160200844 | 160229468 | 137 | 11 | 48362 | -0.92129 | 0.82155 | *MRPL18* |
| ENSG00000172410 | 1 | 67253424 | 67276939 | 81 | 14 | 47810 | -0.92164 | 0.82164 | *INSL5* |
| ENSG00000185787 | 15 | 79092829 | 79200475 | 438 | 37 | 47993 | -0.92174 | 0.82167 | *MORF4L1* |
| ENSG00000268948 | 19 | 58193868 | 58214128 | 56 | 15 | 50550 | -0.92185 | 0.8217 | *AC004017.1* |
| ENSG00000049759 | 18 | 55701599 | 56078772 | 1345 | 152 | 47168 | -0.92193 | 0.82172 | *NEDD4L* |

| | | | | | | | | | |
|---|---|---|---|---|---|---|---|---|---|
| ENSG00000146463 | 1 | 35724568 | 35897659 | 129 | 33 | 42908 | -0.92213 | 0.82177 | *ZMYM4* |
| ENSG00000223524 | 8 | 86737543 | 86759570 | 3 | 1 | 43374 | -0.92221 | 0.82179 | *REXO1L11P* |
| ENSG00000255940 | 8 | 86746718 | 86768745 | 3 | 1 | 43374 | -0.92221 | 0.82179 | *REXO1L10P* |
| ENSG00000172238 | 4 | 94740042 | 94761221 | 62 | 14 | 42558 | -0.92289 | 0.82197 | *ATOH1* |
| ENSG00000132846 | 5 | 76357897 | 76393148 | 102 | 22 | 44557 | -0.92318 | 0.82204 | *ZBED3* |
| ENSG00000111581 | 12 | 69070514 | 69146785 | 205 | 29 | 47292 | -0.92316 | 0.82204 | *NUP107* |
| ENSG00000104818 | 19 | 49525169 | 49546495 | 76 | 15 | 51956 | -0.92327 | 0.82207 | *CGB2* |
| ENSG00000267941 | 7 | 65929543 | 65950709 | 64 | 10 | 52842 | -0.92331 | 0.82208 | *AC008267.1* |
| ENSG00000179023 | 1 | 18797424 | 18822478 | 125 | 20 | 50127 | -0.9236 | 0.82215 | *KLHDC7A* |
| ENSG00000163170 | 2 | 74352525 | 74385121 | 89 | 17 | 36169 | -0.92362 | 0.82216 | *BOLA3* |
| ENSG00000172167 | 8 | 121447640 | 121564373 | 287 | 21 | 29791 | -0.92365 | 0.82216 | *MTBP* |
| ENSG00000174953 | 3 | 153980335 | 154052286 | 144 | 18 | 44945 | -0.92378 | 0.8222 | *DHX36* |
| ENSG00000111266 | 12 | 12618829 | 12725317 | 293 | 37 | 51305 | -0.92386 | 0.82222 | *DUSP16* |
| ENSG00000170248 | 3 | 33829844 | 33921194 | 341 | 20 | 49272 | -0.92423 | 0.82232 | *PDCD6IP* |
| ENSG00000130037 | 12 | 5143085 | 5165949 | 79 | 25 | 49396 | -0.92426 | 0.82232 | *KCNA5* |
| ENSG00000125266 | 13 | 107132079 | 107197462 | 156 | 39 | 49047 | -0.92478 | 0.82246 | *EFNB2* |
| ENSG00000249590 | 22 | 30795244 | 30834977 | 108 | 19 | 42693 | -0.92492 | 0.8225 | *RP4-539M6.19* |
| ENSG00000138160 | 10 | 94343043 | 94425150 | 355 | 31 | 38361 | -0.92566 | 0.82269 | *KIF11* |
| ENSG00000237440 | 19 | 20708631 | 20758615 | 337 | 21 | 51663 | -0.92582 | 0.82273 | *ZNF737* |
| ENSG00000157542 | 21 | 38969678 | 39298749 | 1140 | 128 | 43652 | -0.92608 | 0.8228 | *KCNJ6* |
| ENSG00000176386 | 9 | 116008115 | 116047869 | 142 | 27 | 43906 | -0.92637 | 0.82287 | *CDC26* |
| ENSG00000087095 | 17 | 26358763 | 26533407 | 107 | 44 | 51138 | -0.92686 | 0.823 | *NLK* |
| ENSG00000168418 | 16 | 84245823 | 84283356 | 169 | 36 | 43361 | -0.9271 | 0.82306 | *KCNG4* |
| ENSG00000243489 | 21 | 46056331 | 46077564 | 118 | 17 | 48455 | -0.92742 | 0.82314 | *KRTAP10-11* |
| ENSG00000153898 | 1 | 85381268 | 85472796 | 369 | 32 | 51547 | -0.92758 | 0.82319 | *MCOLN2* |
| ENSG00000185972 | 9 | 36159389 | 36181329 | 86 | 11 | 46327 | -0.92762 | 0.8232 | *CCIN* |
| ENSG00000260240 | 16 | 59733897 | 59798752 | 210 | 35 | 49394 | -0.92763 | 0.8232 | *RP11-105C20.2* |
| ENSG00000175110 | 3 | 138714648 | 139086065 | 907 | 93 | 43745 | -0.92807 | 0.82331 | *MRPS22* |
| ENSG00000198000 | 9 | 95049640 | 95097918 | 110 | 13 | 46018 | -0.92842 | 0.8234 | *NOL8* |
| ENSG00000203685 | 1 | 226726501 | 226806915 | 254 | 44 | 49883 | -0.92847 | 0.82342 | *C1orf95* |
| ENSG00000110484 | 11 | 62027627 | 62050628 | 67 | 10 | 40549 | -0.92858 | 0.82345 | *SCGB2A2* |
| ENSG00000184489 | 8 | 142392093 | 142451620 | 197 | 33 | 43572 | -0.92886 | 0.82352 | *PTP4A3* |
| ENSG00000120254 | 6 | 151176685 | 151433023 | 1185 | 115 | 45170 | -0.92963 | 0.82372 | *MTHFD1L* |
| ENSG00000158691 | 6 | 28336732 | 28377511 | 175 | 16 | 45008 | -0.92995 | 0.8238 | *ZSCAN12* |
| ENSG00000167676 | 19 | 4492204 | 4527716 | 124 | 30 | 43636 | -0.92996 | 0.8238 | *PLIN4* |
| ENSG00000174339 | 5 | 180156042 | 180177099 | 70 | 19 | 40747 | -0.93022 | 0.82387 | *OR2Y1* |

| | | | | | | | | | |
|---|---|---|---|---|---|---|---|---|---|
| ENSG00000163501 | 2 | 219909142 | 219935189 | 84 | 12 | 45541 | -0.93031 | 0.82389 | *IHH* |
| ENSG00000150477 | 18 | 34399069 | 34822135 | 891 | 41 | 31963 | -0.93033 | 0.8239 | *KIAA1328* |
| ENSG00000182551 | 2 | 3491132 | 3533507 | 122 | 24 | 43528 | -0.93041 | 0.82392 | *ADI1* |
| ENSG00000112902 | 5 | 9025138 | 9556187 | 1857 | 237 | 45427 | -0.93043 | 0.82393 | *SEMA5A* |
| ENSG00000182218 | 14 | 100101447 | 100156906 | 171 | 28 | 49236 | -0.93116 | 0.82411 | *HHIPL1* |
| ENSG00000136895 | 9 | 129976544 | 130165939 | 333 | 40 | 49390 | -0.93137 | 0.82417 | *GARNL3* |
| ENSG00000095596 | 10 | 94823232 | 94847647 | 61 | 18 | 41675 | -0.93345 | 0.82471 | *CYP26A1* |
| ENSG00000178175 | 5 | 71728479 | 71813554 | 223 | 24 | 52534 | -0.93421 | 0.8249 | *ZNF366* |
| ENSG00000148459 | 10 | 26976588 | 27045727 | 293 | 28 | 45497 | -0.93431 | 0.82493 | *PDSS1* |
| ENSG00000123352 | 12 | 49750367 | 49931205 | 431 | 32 | 41339 | -0.9343 | 0.82493 | *SPATS2* |
| ENSG00000130724 | 19 | 59052933 | 59076491 | 50 | 14 | 37353 | -0.93547 | 0.82523 | *CHMP2A* |
| ENSG00000188290 | 1 | 924342 | 945552 | 91 | 16 | 39357 | -0.93594 | 0.82535 | *HES4* |
| ENSG00000179151 | 15 | 74912899 | 74998633 | 85 | 17 | 36412 | -0.93602 | 0.82537 | *EDC3* |
| ENSG00000111358 | 12 | 124108375 | 124157153 | 150 | 20 | 31965 | -0.93623 | 0.82542 | *GTF2H3* |
| ENSG00000250317 | 4 | 25853452 | 25941435 | 264 | 47 | 48328 | -0.93689 | 0.82559 | *SMIM20* |
| ENSG00000183960 | 3 | 19179946 | 19587138 | 648 | 98 | 47345 | -0.937 | 0.82562 | *KCNH8* |
| ENSG00000132932 | 13 | 25936209 | 26609989 | 2318 | 208 | 42425 | -0.93709 | 0.82564 | *ATP8A2* |
| ENSG00000269267 | 5 | 60401538 | 60421762 | 64 | 13 | 52029 | -0.93722 | 0.82568 | *AC008498.1* |
| ENSG00000165837 | 13 | 46098657 | 46199874 | 287 | 44 | 38011 | -0.93728 | 0.82569 | *FAM194B* |
| ENSG00000122692 | 9 | 33031762 | 33086665 | 206 | 24 | 51819 | -0.9376 | 0.82577 | *SMU1* |
| ENSG00000215193 | 22 | 18550689 | 18623905 | 239 | 52 | 45393 | -0.93765 | 0.82579 | *PEX26* |
| ENSG00000042445 | 2 | 85559211 | 85591743 | 86 | 18 | 39871 | -0.93815 | 0.82592 | *RETSAT* |
| ENSG00000179826 | 11 | 18132502 | 18170027 | 116 | 18 | 46150 | -0.93829 | 0.82595 | *MRGPRX3* |
| ENSG00000140379 | 15 | 80243231 | 80273788 | 61 | 10 | 48676 | -0.93841 | 0.82598 | *BCL2A1* |
| ENSG00000273259 | 14 | 95048395 | 95100983 | 260 | 47 | 40957 | -0.93856 | 0.82602 | *RP11-986E7.7* |
| ENSG00000268279 | 3 | 14166717 | 14199548 | 87 | 20 | 49455 | -0.93874 | 0.82607 | *RP11-434D12.1* |
| ENSG00000118181 | 11 | 118876422 | 118899401 | 88 | 10 | 54468 | -0.93876 | 0.82607 | *RPS25* |
| ENSG00000006576 | 7 | 77418122 | 77596818 | 457 | 42 | 41528 | -0.93908 | 0.82616 | *PHTF2* |
| ENSG00000036828 | 3 | 121892530 | 122015342 | 505 | 48 | 46572 | -0.93928 | 0.82621 | *CASR* |
| ENSG00000181830 | 11 | 45815623 | 45844566 | 88 | 24 | 41762 | -0.9393 | 0.82621 | *SLC35C1* |
| ENSG00000169248 | 4 | 76944835 | 76972568 | 253 | 6 | 52565 | -0.93969 | 0.82631 | *CXCL11* |
| ENSG00000166012 | 11 | 93453114 | 93527557 | 191 | 17 | 33299 | -0.9397 | 0.82631 | *TAF1D* |
| ENSG00000009724 | 1 | 11076580 | 11117290 | 85 | 15 | 39189 | -0.93984 | 0.82635 | *MASP2* |
| ENSG00000141429 | 18 | 33151081 | 33301798 | 510 | 30 | 50390 | -0.94038 | 0.82649 | *GALNT1* |
| ENSG00000268141 | 1 | 154575067 | 154595190 | 50 | 7 | 50421 | -0.94047 | 0.82651 | *AL606500.1* |
| ENSG00000151116 | 11 | 18541156 | 18620294 | 261 | 29 | 51330 | -0.94074 | 0.82658 | *UEVLD* |

| | | | | | | | | | |
|---|---|---|---|---|---|---|---|---|---|
| ENSG00000168246 | 5 | 171626644 | 171721075 | 331 | 21 | 50779 | -0.94118 | 0.82669 | *UBTD2* |
| ENSG00000158292 | 1 | 6297406 | 6331035 | 99 | 18 | 41911 | -0.94138 | 0.82674 | *GPR153* |
| ENSG00000143297 | 1 | 157473167 | 157532310 | 207 | 21 | 36853 | -0.94149 | 0.82677 | *FCRL5* |
| ENSG00000114405 | 3 | 62294648 | 62331888 | 52 | 21 | 49906 | -0.94164 | 0.82681 | *C3orf14* |
| ENSG00000175868 | 11 | 14916543 | 15113888 | 383 | 22 | 42295 | -0.94166 | 0.82682 | *CALCB* |
| ENSG00000197705 | 18 | 30242634 | 30363025 | 316 | 27 | 46507 | -0.94224 | 0.82697 | *KLHL14* |
| ENSG00000132563 | 5 | 137764706 | 137792658 | 78 | 16 | 46319 | -0.94284 | 0.82712 | *REEP2* |
| ENSG00000139737 | 13 | 78262023 | 78348377 | 229 | 36 | 45411 | -0.94289 | 0.82713 | *SLAIN1* |
| ENSG00000164458 | 6 | 166561144 | 166592188 | 191 | 23 | 46157 | -0.94296 | 0.82715 | *T* |
| ENSG00000108255 | 17 | 27563881 | 27591512 | 77 | 16 | 46550 | -0.9432 | 0.82721 | *CRYBA1* |
| ENSG00000166913 | 20 | 43504317 | 43547173 | 102 | 10 | 54386 | -0.94326 | 0.82723 | *YWHAB* |
| ENSG00000169902 | 7 | 65660186 | 65895530 | 543 | 30 | 50946 | -0.9437 | 0.82734 | *TPST1* |
| ENSG00000134897 | 13 | 103441399 | 103503885 | 195 | 24 | 47202 | -0.94391 | 0.82739 | *BIVM* |
| ENSG00000105679 | 19 | 36014314 | 36046218 | 105 | 20 | 48788 | -0.94458 | 0.82756 | *GAPDHS* |
| ENSG00000135336 | 6 | 88289839 | 88387169 | 291 | 30 | 53763 | -0.94497 | 0.82766 | *ORC3* |
| ENSG00000167658 | 19 | 3966054 | 3995467 | 74 | 28 | 50469 | -0.94537 | 0.82776 | *EEF2* |
| ENSG00000156959 | 3 | 9533481 | 9605486 | 264 | 28 | 47777 | -0.94556 | 0.82781 | *LHFPL4* |
| ENSG00000163421 | 3 | 71810807 | 71844357 | 90 | 26 | 41728 | -0.94555 | 0.82781 | *PROK2* |
| ENSG00000213297 | 19 | 12232932 | 12277546 | 95 | 16 | 43708 | -0.94562 | 0.82783 | *ZNF625-ZNF20* |
| ENSG00000143753 | 1 | 224353458 | 224391143 | 135 | 18 | 43580 | -0.94586 | 0.82789 | *DEGS1* |
| ENSG00000139292 | 12 | 71823550 | 71990090 | 640 | 49 | 49593 | -0.94608 | 0.82795 | *LGR5* |
| ENSG00000204709 | 6 | 28901654 | 28922314 | 106 | 20 | 38452 | -0.94621 | 0.82798 | *C6orf100* |
| ENSG00000188778 | 8 | 37810516 | 37834483 | 38 | 7 | 51507 | -0.94663 | 0.82809 | *ADRB3* |
| ENSG00000183291 | 1 | 87318132 | 87390107 | 206 | 16 | 36517 | -0.94709 | 0.8282 | *Sep-15* |
| ENSG00000168763 | 2 | 97471982 | 97509648 | 49 | 11 | 34999 | -0.94707 | 0.8282 | *CNNM3* |
| ENSG00000172404 | 22 | 41245553 | 41268130 | 33 | 13 | 39790 | -0.94753 | 0.82831 | *DNAJB7* |
| ENSG00000178597 | 4 | 7422022 | 7446700 | 173 | 41 | 49153 | -0.94756 | 0.82832 | *PSAPL1* |
| ENSG00000092978 | 1 | 217590334 | 217814424 | 683 | 88 | 43810 | -0.9476 | 0.82833 | *GPATCH2* |
| ENSG00000259288 | 15 | 40499629 | 40578597 | 162 | 51 | 42352 | -0.94761 | 0.82833 | *RP11-133K1.2* |
| ENSG00000266086 | 17 | 56056399 | 56092614 | 77 | 13 | 49530 | -0.94761 | 0.82834 | *RP11-159D12.5* |
| ENSG00000181315 | 6 | 26626518 | 26669980 | 141 | 15 | 50684 | -0.94766 | 0.82835 | *ZNF322* |
| ENSG00000089248 | 12 | 112441120 | 112471255 | 46 | 6 | 32760 | -0.94843 | 0.82854 | *ERP29* |
| ENSG00000165868 | 10 | 118420703 | 118512085 | 342 | 38 | 50444 | -0.94875 | 0.82863 | *HSPA12A* |
| ENSG00000162066 | 16 | 2560358 | 2591423 | 26 | 18 | 51854 | -0.9489 | 0.82866 | *AMDHD2* |
| ENSG00000167470 | 19 | 1238552 | 1269142 | 57 | 29 | 46239 | -0.94919 | 0.82874 | *MIDN* |
| ENSG00000169783 | 15 | 77895369 | 78123242 | 770 | 84 | 42457 | -0.94923 | 0.82875 | *LINGO1* |

| | | | | | | | | | |
|---|---|---|---|---|---|---|---|---|---|
| ENSG00000158470 | 20 | 48239482 | 48340415 | 283 | 24 | 55183 | -0.94948 | 0.82881 | *B4GALT5* |
| ENSG00000100302 | 22 | 35926915 | 35960048 | 85 | 19 | 45129 | -0.94967 | 0.82886 | *RASD2* |
| ENSG00000162909 | 1 | 223879295 | 223973720 | 306 | 36 | 36542 | -0.94972 | 0.82887 | *CAPN2* |
| ENSG00000080823 | 14 | 102680837 | 102781537 | 299 | 31 | 45023 | -0.94972 | 0.82887 | *MOK* |
| ENSG00000108813 | 17 | 48036334 | 48062321 | 68 | 21 | 48987 | -0.94995 | 0.82893 | *DLX4* |
| ENSG00000185055 | 7 | 105195567 | 105251322 | 145 | 29 | 42364 | -0.95033 | 0.82903 | *EFCAB10* |
| ENSG00000187553 | 10 | 94811021 | 94838454 | 61 | 14 | 34382 | -0.95071 | 0.82912 | *CYP26C1* |
| ENSG00000006042 | 17 | 31244928 | 31282124 | 139 | 24 | 48568 | -0.95069 | 0.82912 | *TMEM98* |
| ENSG00000105419 | 19 | 47896381 | 47932780 | 104 | 31 | 43242 | -0.95077 | 0.82914 | *MEIS3* |
| ENSG00000261567 | 16 | 85381069 | 85403055 | 84 | 21 | 52006 | -0.95088 | 0.82917 | *RP11-680G10.1* |
| ENSG00000051382 | 3 | 138362860 | 138563780 | 312 | 17 | 44132 | -0.95099 | 0.82919 | *PIK3CB* |
| ENSG00000165490 | 11 | 82601017 | 82679319 | 223 | 25 | 48909 | -0.95108 | 0.82922 | *C11orf82* |
| ENSG00000108679 | 17 | 76957320 | 76986191 | 93 | 30 | 45497 | -0.95162 | 0.82936 | *LGALS3BP* |
| ENSG00000118200 | 1 | 200698686 | 200839832 | 397 | 36 | 41196 | -0.95213 | 0.82948 | *CAMSAP2* |
| ENSG00000163071 | 4 | 52907497 | 52973458 | 143 | 17 | 29195 | -0.95245 | 0.82957 | *SPATA18* |
| ENSG00000124786 | 6 | 8403301 | 8445794 | 196 | 19 | 40139 | -0.95252 | 0.82958 | *SLC35B3* |
| ENSG00000141337 | 17 | 66245323 | 66428872 | 523 | 103 | 42887 | -0.95258 | 0.8296 | *ARSG* |
| ENSG00000115008 | 2 | 113521492 | 113552167 | 91 | 8 | 51554 | -0.9529 | 0.82968 | *IL1A* |
| ENSG00000140263 | 15 | 45305302 | 45379383 | 171 | 29 | 42704 | -0.95314 | 0.82974 | *SORD* |
| ENSG00000163599 | 2 | 204722509 | 204748683 | 55 | 10 | 53948 | -0.9533 | 0.82978 | *CTLA4* |
| ENSG00000111707 | 12 | 118804185 | 118865840 | 169 | 17 | 51551 | -0.95353 | 0.82984 | *SUDS3* |
| ENSG00000127955 | 7 | 79753271 | 79858718 | 308 | 49 | 48634 | -0.95424 | 0.83002 | *GNAI1* |
| ENSG00000111554 | 12 | 68656223 | 68736161 | 188 | 35 | 49088 | -0.9546 | 0.83011 | *MDM1* |
| ENSG00000133048 | 1 | 203138059 | 203165877 | 88 | 22 | 36311 | -0.9549 | 0.83019 | *CHI3L1* |
| ENSG00000257949 | 17 | 73965301 | 74006667 | 144 | 24 | 49258 | -0.95496 | 0.8302 | *TEN1* |
| ENSG00000182446 | 17 | 79513913 | 79625495 | 598 | 39 | 39803 | -0.95505 | 0.83022 | *NPLOC4* |
| ENSG00000089094 | 12 | 121856900 | 122028920 | 371 | 45 | 36086 | -0.95614 | 0.8305 | *KDM2B* |
| ENSG00000123977 | 2 | 228725770 | 228799060 | 344 | 25 | 53441 | -0.95637 | 0.83056 | *DAW1* |
| ENSG00000130921 | 12 | 123707463 | 123752506 | 175 | 15 | 37438 | -0.95676 | 0.83066 | *C12orf65* |
| ENSG00000143157 | 1 | 166798681 | 166835581 | 101 | 11 | 53777 | -0.95704 | 0.83073 | *POGK* |
| ENSG00000136943 | 9 | 99784940 | 99811925 | 43 | 6 | 29477 | -0.95759 | 0.83086 | *CTSV* |
| ENSG00000129197 | 17 | 5312961 | 5346196 | 152 | 14 | 26764 | -0.95918 | 0.83127 | *RPAIN* |
| ENSG00000152234 | 18 | 43654110 | 43694300 | 177 | 16 | 45945 | -0.95934 | 0.83131 | *ATP5A1* |
| ENSG00000168386 | 3 | 99538985 | 99843357 | 462 | 39 | 53708 | -0.95939 | 0.83132 | *FILIP1L* |
| ENSG00000248993 | 6 | 32895141 | 32930899 | 213 | 32 | 50575 | -0.9598 | 0.83142 | *XXbac-BPG181M17.5* |
| ENSG00000269185 | 7 | 108530389 | 108550655 | 99 | 9 | 51439 | -0.95988 | 0.83144 | *FLJ00325* |

| | | | | | | | | | |
|---|---|---|---|---|---|---|---|---|---|
| ENSG00000173272 | 2 | 132212473 | 132260316 | 238 | 13 | 21060 | -0.95995 | 0.83146 | *MZT2A* |
| ENSG00000115761 | 2 | 10700892 | 10840101 | 657 | 28 | 29256 | -0.95998 | 0.83147 | *NOL10* |
| ENSG00000145491 | 5 | 10431636 | 10482141 | 64 | 19 | 34833 | -0.96029 | 0.83155 | *ROPN1L* |
| ENSG00000197302 | 16 | 31714550 | 31816190 | 216 | 16 | 34795 | -0.96065 | 0.83164 | *ZNF720* |
| ENSG00000140548 | 15 | 90534624 | 90635438 | 239 | 49 | 39125 | -0.96069 | 0.83165 | *ZNF710* |
| ENSG00000215790 | 1 | 1646277 | 1687431 | 34 | 21 | 42068 | -0.96126 | 0.83179 | *SLC35E2* |
| ENSG00000163510 | 2 | 180799603 | 180881840 | 259 | 22 | 52755 | -0.96142 | 0.83183 | *CWC22* |
| ENSG00000213240 | 1 | 145199119 | 145301972 | 6 | 4 | 38135 | -0.96158 | 0.83187 | *NOTCH2NL* |
| ENSG00000160318 | 19 | 51860352 | 51882257 | 66 | 16 | 35889 | -0.96171 | 0.8319 | *CLDND2* |
| ENSG00000110665 | 11 | 2306875 | 2334279 | 122 | 25 | 43232 | -0.96197 | 0.83197 | *C11orf21* |
| ENSG00000154174 | 3 | 100072275 | 100130242 | 108 | 18 | 39749 | -0.9621 | 0.832 | *TOMM70A* |
| ENSG00000158417 | 2 | 99943816 | 100027789 | 235 | 22 | 50837 | -0.96253 | 0.83211 | *EIF5B* |
| ENSG00000042088 | 14 | 90411283 | 90521106 | 291 | 25 | 27628 | -0.9627 | 0.83215 | *TDP1* |
| ENSG00000204316 | 17 | 73884724 | 73915899 | 78 | 16 | 39974 | -0.96356 | 0.83237 | *MRPL38* |
| ENSG00000240240 | 9 | 42399989 | 42484239 | 1 | 1 | 42793 | -0.96369 | 0.8324 | *RP11-146D12.2* |
| ENSG00000058729 | 5 | 96486571 | 96528964 | 162 | 27 | 45399 | -0.96422 | 0.83253 | *RIOK2* |
| ENSG00000178358 | 11 | 6932205 | 6953264 | 112 | 9 | 33049 | -0.96461 | 0.83263 | *OR2D3* |
| ENSG00000186971 | 21 | 31792572 | 31813216 | 57 | 6 | 55730 | -0.96465 | 0.83264 | *KRTAP13-4* |
| ENSG00000023697 | 12 | 16054106 | 16200220 | 241 | 43 | 46709 | -0.96472 | 0.83266 | *DERA* |
| ENSG00000048707 | 1 | 12280124 | 12582099 | 657 | 44 | 43641 | -0.96477 | 0.83267 | *VPS13D* |
| ENSG00000108559 | 17 | 5254258 | 5333480 | 375 | 19 | 27579 | -0.96573 | 0.83291 | *NUP88* |
| ENSG00000072778 | 17 | 7110444 | 7138592 | 43 | 10 | 49579 | -0.96585 | 0.83294 | *ACADVL* |
| ENSG00000115109 | 2 | 120760581 | 120946695 | 392 | 23 | 39255 | -0.9659 | 0.83295 | *EPB41L5* |
| ENSG00000160200 | 21 | 44463301 | 44507053 | 129 | 29 | 43669 | -0.96594 | 0.83296 | *CBS* |
| ENSG00000104763 | 8 | 17903934 | 17952494 | 317 | 35 | 47297 | -0.96608 | 0.833 | *ASAH1* |
| ENSG00000124784 | 6 | 7379729 | 7428270 | 171 | 18 | 36462 | -0.96716 | 0.83327 | *RIOK1* |
| ENSG00000186017 | 19 | 36926021 | 36990804 | 205 | 20 | 53804 | -0.96716 | 0.83327 | *ZNF566* |
| ENSG00000146909 | 7 | 156732417 | 156775876 | 322 | 27 | 54479 | -0.96737 | 0.83332 | *NOM1* |
| ENSG00000126522 | 7 | 65530785 | 65568545 | 109 | 14 | 45107 | -0.96779 | 0.83342 | *ASL* |
| ENSG00000234237 | 1 | 180931695 | 180959868 | 87 | 12 | 50962 | -0.96784 | 0.83344 | *AL162431.1* |
| ENSG00000154258 | 17 | 66960629 | 67067205 | 258 | 32 | 35771 | -0.96803 | 0.83349 | *ABCA9* |
| ENSG00000173728 | 1 | 244505937 | 244562965 | 228 | 38 | 45927 | -0.96818 | 0.83352 | *C1orf100* |
| ENSG00000094914 | 12 | 53691240 | 53728648 | 56 | 12 | 39694 | -0.96872 | 0.83366 | *AAAS* |
| ENSG00000100300 | 22 | 43537520 | 43569248 | 93 | 18 | 38147 | -0.96958 | 0.83387 | *TSPO* |
| ENSG00000172269 | 11 | 118957213 | 118989041 | 75 | 12 | 33126 | -0.96961 | 0.83388 | *DPAGT1* |
| ENSG00000163608 | 3 | 112711287 | 112748708 | 79 | 21 | 40528 | -0.96977 | 0.83392 | *C3orf17* |

| | | | | | | | | | |
|---|---|---|---|---|---|---|---|---|---|
| ENSG00000177666 | 11 | 808902 | 835573 | 86 | 21 | 35827 | -0.97116 | 0.83427 | *PNPLA2* |
| ENSG00000115665 | 2 | 108592979 | 108640450 | 133 | 33 | 50660 | -0.97147 | 0.83434 | *SLC5A7* |
| ENSG00000113790 | 3 | 184898412 | 185009778 | 217 | 26 | 33169 | -0.97191 | 0.83445 | *EHHADH* |
| ENSG00000111276 | 12 | 12857992 | 12885305 | 70 | 14 | 47902 | -0.9722 | 0.83452 | *CDKN1B* |
| ENSG00000162512 | 1 | 31332314 | 31391608 | 162 | 46 | 46443 | -0.97247 | 0.83459 | *SDC3* |
| ENSG00000120563 | 10 | 29567990 | 29617257 | 286 | 42 | 51291 | -0.97253 | 0.83461 | *LYZL1* |
| ENSG00000129187 | 4 | 183801213 | 183849089 | 237 | 35 | 45096 | -0.97321 | 0.83477 | *DCTD* |
| ENSG00000174238 | 17 | 1411012 | 1476110 | 241 | 32 | 50601 | -0.97327 | 0.83479 | *PITPNA* |
| ENSG00000184792 | 22 | 31079769 | 31313811 | 729 | 96 | 47986 | -0.97352 | 0.83485 | *OSBP2* |
| ENSG00000188343 | 8 | 94700789 | 94753755 | 104 | 19 | 55235 | -0.97448 | 0.83509 | *FAM92A1* |
| ENSG00000184254 | 15 | 101407919 | 101466831 | 123 | 24 | 39256 | -0.9747 | 0.83514 | *ALDH1A3* |
| ENSG00000137843 | 15 | 40499629 | 40579688 | 163 | 51 | 42445 | -0.97574 | 0.8354 | *PAK6* |
| ENSG00000163607 | 3 | 112699765 | 112743907 | 105 | 25 | 40338 | -0.97587 | 0.83544 | *GTPBP8* |
| ENSG00000170498 | 1 | 204149469 | 204175614 | 95 | 19 | 48690 | -0.97598 | 0.83546 | *KISS1* |
| ENSG00000267335 | 19 | 49516210 | 49550152 | 136 | 22 | 49389 | -0.97637 | 0.83556 | *CTB-60B18.6* |
| ENSG00000162368 | 1 | 47789469 | 47854511 | 183 | 18 | 38463 | -0.9768 | 0.83567 | *CMPK1* |
| ENSG00000245017 | 12 | 98869322 | 98907633 | 130 | 23 | 45958 | -0.97728 | 0.83578 | *RP11-181C3.1* |
| ENSG00000187730 | 1 | 1940780 | 1972192 | 82 | 16 | 44807 | -0.9781 | 0.83599 | *GABRD* |
| ENSG00000163743 | 4 | 76394247 | 76449974 | 146 | 17 | 51394 | -0.97838 | 0.83606 | *RCHY1* |
| ENSG00000226209 | 10 | 13340145 | 13360447 | 103 | 19 | 46552 | -0.97845 | 0.83607 | *AL138764.1* |
| ENSG00000075790 | 7 | 107210422 | 107279615 | 108 | 17 | 52582 | -0.97872 | 0.83614 | *BCAP29* |
| ENSG00000123094 | 12 | 26101962 | 26242825 | 652 | 67 | 49177 | -0.97879 | 0.83616 | *RASSF8* |
| ENSG00000213077 | 17 | 18417880 | 18440160 | 2 | 1 | 42944 | -0.97937 | 0.8363 | *FAM106A* |
| ENSG00000123360 | 12 | 54933134 | 54983023 | 124 | 36 | 45036 | -0.98042 | 0.83656 | *PDE1B* |
| ENSG00000090924 | 19 | 39893225 | 39929054 | 86 | 19 | 41541 | -0.98059 | 0.8366 | *PLEKHG2* |
| ENSG00000105825 | 7 | 93504709 | 93530303 | 105 | 9 | 31938 | -0.98137 | 0.8368 | *TFPI2* |
| ENSG00000170852 | 7 | 32897784 | 32943743 | 111 | 13 | 37378 | -0.98172 | 0.83688 | *KBTBD2* |
| ENSG00000125755 | 19 | 46308668 | 46376548 | 284 | 20 | 52047 | -0.98188 | 0.83692 | *SYMPK* |
| ENSG00000157851 | 2 | 27060615 | 27183219 | 255 | 20 | 48134 | -0.98239 | 0.83705 | *DPYSL5* |
| ENSG00000176102 | 11 | 33088734 | 33193917 | 339 | 29 | 40959 | -0.98293 | 0.83718 | *CSTF3* |
| ENSG00000240542 | 17 | 39334697 | 39356891 | 92 | 7 | 54281 | -0.98349 | 0.83732 | *KRTAP9-1* |
| ENSG00000100399 | 22 | 41615517 | 41646938 | 63 | 13 | 45124 | -0.98348 | 0.83732 | *CHADL* |
| ENSG00000197312 | 1 | 15933995 | 16005539 | 193 | 11 | 47044 | -0.98368 | 0.83736 | *DDI2* |
| ENSG00000138279 | 10 | 75124859 | 75183834 | 112 | 7 | 24197 | -0.98421 | 0.83749 | *ANXA7* |
| ENSG00000104967 | 19 | 46426992 | 46486804 | 128 | 33 | 47857 | -0.98437 | 0.83753 | *NOVA2* |
| ENSG00000177868 | 1 | 43262723 | 43292954 | 80 | 16 | 54713 | -0.98441 | 0.83754 | *CCDC23* |

| | | | | | | | | | |
|---|---|---|---|---|---|---|---|---|---|
| ENSG00000198369 | 2 | 65527985 | 65669771 | 444 | 44 | 33744 | -0.98498 | 0.83768 | *SPRED2* |
| ENSG00000156265 | 21 | 30439792 | 30558210 | 439 | 38 | 44688 | -0.98501 | 0.83769 | *MAP3K7CL* |
| ENSG00000142684 | 1 | 26486362 | 26508551 | 70 | 13 | 38041 | -0.98505 | 0.8377 | *ZNF593* |
| ENSG00000171119 | 19 | 5813813 | 5838335 | 111 | 20 | 39225 | -0.98541 | 0.83779 | *NRTN* |
| ENSG00000141858 | 19 | 14188652 | 14211848 | 32 | 14 | 48054 | -0.98559 | 0.83783 | *SAMD1* |
| ENSG00000163114 | 4 | 96751239 | 96772625 | 111 | 12 | 49415 | -0.98611 | 0.83796 | *PDHA2* |
| ENSG00000231274 | 19 | 56042023 | 56066909 | 129 | 34 | 42656 | -0.98614 | 0.83797 | *SBK3* |
| ENSG00000127903 | 19 | 57164020 | 57193151 | 141 | 23 | 41107 | -0.98632 | 0.83801 | *ZNF835* |
| ENSG00000169344 | 16 | 20334374 | 20377623 | 144 | 28 | 39935 | -0.98663 | 0.83809 | *UMOD* |
| ENSG00000156876 | 1 | 100539119 | 100608511 | 208 | 26 | 36152 | -0.9871 | 0.8382 | *SASS6* |
| ENSG00000111199 | 12 | 110210890 | 110281212 | 221 | 42 | 48662 | -0.98708 | 0.8382 | *TRPV4* |
| ENSG00000187833 | 2 | 74001316 | 74054274 | 32 | 8 | 42182 | -0.98734 | 0.83826 | *C2orf78* |
| ENSG00000100003 | 22 | 30782846 | 30831305 | 140 | 23 | 43420 | -0.98745 | 0.83829 | *SEC14L2* |
| ENSG00000180332 | 13 | 45756988 | 45785175 | 58 | 9 | 42803 | -0.9875 | 0.8383 | *KCTD4* |
| ENSG00000138463 | 3 | 122503642 | 122609986 | 280 | 32 | 42695 | -0.9879 | 0.8384 | *DIRC2* |
| ENSG00000186105 | 5 | 61864562 | 61887275 | 35 | 10 | 48700 | -0.98815 | 0.83846 | *LRRC70* |
| ENSG00000198331 | 11 | 125743509 | 125780543 | 136 | 19 | 44553 | -0.98847 | 0.83854 | *HYLS1* |
| ENSG00000064787 | 20 | 52543316 | 52697304 | 748 | 90 | 46095 | -0.98853 | 0.83855 | *BCAS1* |
| ENSG00000155282 | 9 | 67958793 | 67997998 | 1 | 1 | 42758 | -0.98913 | 0.8387 | *RP11-195B21.3* |
| ENSG00000169914 | 1 | 20199006 | 20249438 | 163 | 26 | 54465 | -0.98928 | 0.83874 | *OTUD3* |
| ENSG00000203877 | 6 | 84552985 | 84577234 | 36 | 7 | 32296 | -0.98933 | 0.83875 | *RIPPLY2* |
| ENSG00000148153 | 9 | 115436206 | 115490516 | 140 | 25 | 45648 | -0.99012 | 0.83894 | *INIP* |
| ENSG00000105875 | 7 | 134858590 | 134906316 | 143 | 9 | 53125 | -0.99029 | 0.83898 | *WDR91* |
| ENSG00000099999 | 22 | 30763835 | 30827760 | 195 | 27 | 46270 | -0.99031 | 0.83899 | *RNF215* |
| ENSG00000109321 | 4 | 75300851 | 75330726 | 35 | 11 | 36644 | -0.99045 | 0.83902 | *AREG* |
| ENSG00000269490 | 6 | 33653196 | 33673474 | 101 | 11 | 27845 | -0.99056 | 0.83905 | *SBP1* |
| ENSG00000166326 | 11 | 35674353 | 35839775 | 238 | 43 | 46932 | -0.9916 | 0.8393 | *TRIM44* |
| ENSG00000204503 | 1 | 13318196 | 13341692 | 6 | 2 | 42825 | -0.99185 | 0.83936 | *PRAMEF3* |
| ENSG00000181061 | 3 | 42788669 | 42856023 | 142 | 23 | 50669 | -0.99185 | 0.83936 | *HIGD1A* |
| ENSG00000164778 | 7 | 155240824 | 155267526 | 101 | 28 | 47481 | -0.99231 | 0.83948 | *EN2* |
| ENSG00000168614 | 1 | 144801744 | 144840413 | 2 | 1 | 42648 | -0.99239 | 0.8395 | *NBPF9* |
| ENSG00000146842 | 7 | 129794555 | 129857610 | 162 | 25 | 49584 | -0.99243 | 0.83951 | *TMEM209* |
| ENSG00000198090 | 17 | 39285685 | 39306739 | 57 | 12 | 34385 | -0.99306 | 0.83966 | *KRTAP4-6* |
| ENSG00000143228 | 1 | 163226366 | 163335554 | 564 | 53 | 52316 | -0.99315 | 0.83968 | *NUF2* |
| ENSG00000255663 | 11 | 114261404 | 114324654 | 227 | 28 | 47886 | -0.99322 | 0.8397 | *RP11-212D19.4* |
| ENSG00000114315 | 3 | 193843934 | 193866521 | 71 | 16 | 49004 | -0.99339 | 0.83974 | *HES1* |

| | | | | | | | | | |
|---|---|---|---|---|---|---|---|---|---|
| ENSG00000078902 | 11 | 1285601 | 1340884 | 199 | 31 | 50549 | -0.99361 | 0.83979 | *TOLLIP* |
| ENSG00000224997 | 14 | 104167607 | 104189149 | 43 | 12 | 49544 | -0.99384 | 0.83985 | *AL049840.1* |
| ENSG00000156006 | 8 | 18238755 | 18268728 | 219 | 20 | 50364 | -0.99396 | 0.83988 | *NAT2* |
| ENSG00000136881 | 9 | 104112699 | 104155801 | 162 | 11 | 52095 | -0.99406 | 0.8399 | *BAAT* |
| ENSG00000135404 | 12 | 56109107 | 56133491 | 37 | 15 | 38788 | -0.99452 | 0.84001 | *CD63* |
| ENSG00000267631 | 19 | 49528831 | 49562006 | 130 | 30 | 49062 | -0.99467 | 0.84005 | *CGB1* |
| ENSG00000095564 | 10 | 93673526 | 93800082 | 268 | 22 | 31847 | -0.99646 | 0.84049 | *BTAF1* |
| ENSG00000136750 | 10 | 26495236 | 26603487 | 367 | 27 | 42126 | -0.9966 | 0.84052 | *GAD2* |
| ENSG00000113013 | 5 | 137880571 | 137921133 | 84 | 13 | 44166 | -0.99683 | 0.84058 | *HSPA9* |
| ENSG00000169599 | 2 | 69612882 | 69674760 | 166 | 13 | 38806 | -0.99695 | 0.84061 | *NFU1* |
| ENSG00000114113 | 3 | 139161726 | 139209589 | 127 | 14 | 26801 | -0.99704 | 0.84063 | *RBP2* |
| ENSG00000227921 | 9 | 40018620 | 40042417 | 2 | 1 | 42892 | -0.99708 | 0.84064 | *AL353791.1* |
| ENSG00000186352 | 4 | 186307175 | 186331782 | 73 | 13 | 33589 | -0.99724 | 0.84068 | *ANKRD37* |
| ENSG00000008311 | 7 | 121705701 | 121794334 | 192 | 25 | 46709 | -0.9975 | 0.84074 | *AASS* |
| ENSG00000257743 | 7 | 141801549 | 141932124 | 399 | 67 | 48536 | -0.99751 | 0.84074 | *RP11-1220K2.2* |
| ENSG00000047365 | 4 | 35939843 | 36256131 | 997 | 122 | 46871 | -0.99753 | 0.84075 | *ARAP2* |
| ENSG00000189114 | 19 | 45672003 | 45695059 | 81 | 19 | 49196 | -0.99763 | 0.84077 | *BLOC1S3* |
| ENSG00000181026 | 15 | 89154527 | 89185513 | 112 | 21 | 40933 | -0.99882 | 0.84106 | *AEN* |
| ENSG00000170113 | 15 | 23033277 | 23110005 | 109 | 32 | 45192 | -0.9991 | 0.84113 | *NIPA1* |
| ENSG00000042286 | 10 | 71847979 | 71902690 | 205 | 47 | 39749 | -0.99924 | 0.84116 | *AIFM2* |
| ENSG00000123453 | 9 | 136518682 | 136615077 | 358 | 62 | 44084 | -1.0003 | 0.84143 | *SARDH* |
| ENSG00000094661 | 19 | 15187791 | 15209032 | 122 | 22 | 39759 | -1.0008 | 0.84155 | *OR1I1* |
| ENSG00000105137 | 19 | 15208214 | 15235799 | 80 | 18 | 46060 | -1.001 | 0.8416 | *SYDE1* |
| ENSG00000134884 | 13 | 107184021 | 107230512 | 76 | 14 | 51264 | -1.0012 | 0.84165 | *ARGLU1* |
| ENSG00000133878 | 8 | 33438856 | 33467624 | 70 | 12 | 48679 | -1.0013 | 0.84167 | *DUSP26* |
| ENSG00000179041 | 8 | 67331263 | 67352966 | 81 | 18 | 36557 | -1.0013 | 0.84167 | *RRS1* |
| ENSG00000181396 | 17 | 80337099 | 80386513 | 191 | 25 | 45691 | -1.0015 | 0.84171 | *OGFOD3* |
| ENSG00000108256 | 17 | 27572854 | 27631136 | 166 | 21 | 45311 | -1.0017 | 0.84176 | *NUFIP2* |
| ENSG00000101346 | 20 | 30785683 | 30836470 | 58 | 11 | 36685 | -1.0019 | 0.8418 | *POFUT1* |
| ENSG00000176624 | 18 | 48690920 | 48754674 | 161 | 14 | 36050 | -1.0019 | 0.84181 | *MEX3C* |
| ENSG00000188266 | 15 | 78789906 | 78839714 | 142 | 13 | 34383 | -1.0021 | 0.84185 | *HYKK* |
| ENSG00000061676 | 2 | 183763843 | 183913586 | 410 | 42 | 43431 | -1.0021 | 0.84186 | *NCKAP1* |
| ENSG00000138769 | 4 | 76493215 | 76565900 | 189 | 17 | 37288 | -1.0031 | 0.84211 | *CDKL2* |
| ENSG00000242715 | 13 | 36791182 | 36881977 | 487 | 12 | 55154 | -1.0034 | 0.84216 | *CCDC169* |
| ENSG00000268838 | 15 | 78820023 | 78841288 | 73 | 6 | 29759 | -1.0039 | 0.84228 | *AC027228.1* |
| ENSG00000007541 | 16 | 606995 | 644136 | 138 | 13 | 50667 | -1.0039 | 0.84228 | *PIGQ* |

| | | | | | | | | | |
|---|---|---|---|---|---|---|---|---|---|
| ENSG00000188811 | 13 | 39602443 | 39634246 | 68 | 12 | 42912 | -1.0041 | 0.84233 | *NHLRC3* |
| ENSG00000167088 | 18 | 19182228 | 19220417 | 52 | 9 | 31546 | -1.0043 | 0.84238 | *SNRPD1* |
| ENSG00000163132 | 4 | 4851393 | 4875663 | 113 | 30 | 46082 | -1.0048 | 0.84251 | *MSX1* |
| ENSG00000127530 | 19 | 14899958 | 14921023 | 80 | 14 | 56489 | -1.0049 | 0.84253 | *OR7C1* |
| ENSG00000136240 | 7 | 6475584 | 6533873 | 201 | 34 | 41160 | -1.005 | 0.84255 | *KDELR2* |
| ENSG00000125498 | 19 | 55271263 | 55305774 | 30 | 15 | 33700 | -1.0053 | 0.84262 | *KIR2DL1* |
| ENSG00000164532 | 7 | 35232042 | 35303758 | 237 | 14 | 52582 | -1.0059 | 0.84276 | *TBX20* |
| ENSG00000038945 | 8 | 15955387 | 16434999 | 1778 | 141 | 35901 | -1.0061 | 0.84283 | *MSR1* |
| ENSG00000146243 | 6 | 79567189 | 79666157 | 220 | 22 | 52580 | -1.0069 | 0.84302 | *IRAK1BP1* |
| ENSG00000268402 | 6 | 143749575 | 143774295 | 84 | 5 | 54786 | -1.0073 | 0.84311 | *AL031320.1* |
| ENSG00000147536 | 8 | 41376725 | 41412565 | 150 | 26 | 50809 | -1.0082 | 0.84331 | *GINS4* |
| ENSG00000161850 | 12 | 52777715 | 52810139 | 156 | 23 | 40163 | -1.0083 | 0.84333 | *KRT82* |
| ENSG00000174151 | 1 | 110026674 | 110055554 | 58 | 13 | 46645 | -1.0087 | 0.84345 | *CYB561D1* |
| ENSG00000169925 | 9 | 136885427 | 136943657 | 252 | 23 | 44252 | -1.0087 | 0.84345 | *BRD3* |
| ENSG00000212724 | 17 | 39205495 | 39226344 | 90 | 13 | 27247 | -1.0088 | 0.84347 | *KRTAP2-3* |
| ENSG00000217236 | 2 | 175189674 | 175213220 | 71 | 19 | 35189 | -1.0089 | 0.84348 | *SP9* |
| ENSG00000212998 | 8 | 94742349 | 94763001 | 45 | 5 | 55258 | -1.0099 | 0.84373 | *RBM12B-AS1* |
| ENSG00000160932 | 8 | 144089399 | 144115249 | 81 | 16 | 38018 | -1.0105 | 0.84388 | *LY6E* |
| ENSG00000169314 | 22 | 24095208 | 24118048 | 77 | 18 | 43679 | -1.0108 | 0.84394 | *C22orf15* |
| ENSG00000254737 | 11 | 123876282 | 123897217 | 88 | 15 | 47734 | -1.0111 | 0.84403 | *OR10G4* |
| ENSG00000138398 | 2 | 170430850 | 170507916 | 338 | 25 | 55460 | -1.0114 | 0.84408 | *PPIG* |
| ENSG00000093010 | 22 | 19919130 | 19967498 | 227 | 41 | 46077 | -1.0115 | 0.84412 | *COMT* |
| ENSG00000165219 | 9 | 128014073 | 128139486 | 218 | 17 | 45942 | -1.012 | 0.84423 | *GAPVD1* |
| ENSG00000237452 | 19 | 46226509 | 46277792 | 81 | 15 | 51034 | -1.012 | 0.84423 | *AC074212.3* |
| ENSG00000116560 | 1 | 35631979 | 35668749 | 17 | 8 | 31075 | -1.0122 | 0.84428 | *SFPQ* |
| ENSG00000196832 | 14 | 20655495 | 20676605 | 108 | 31 | 53658 | -1.0128 | 0.84443 | *OR11G2* |
| ENSG00000235109 | 6 | 28282470 | 28334048 | 186 | 29 | 33161 | -1.0135 | 0.84459 | *ZSCAN31* |
| ENSG00000072201 | 4 | 54315468 | 54577572 | 988 | 114 | 47162 | -1.0135 | 0.8446 | *LNX1* |
| ENSG00000197520 | 1 | 222900549 | 222934147 | 59 | 15 | 48774 | -1.0141 | 0.84474 | *FAM177B* |
| ENSG00000163873 | 1 | 37251128 | 37509730 | 575 | 86 | 46743 | -1.0143 | 0.84477 | *GRIK3* |
| ENSG00000177938 | 12 | 18881045 | 18902121 | 76 | 14 | 44222 | -1.0145 | 0.84483 | *CAPZA3* |
| ENSG00000129749 | 11 | 3676817 | 3702614 | 92 | 19 | 41290 | -1.0148 | 0.8449 | *CHRNA10* |
| ENSG00000175344 | 15 | 32312691 | 32474722 | 361 | 50 | 42369 | -1.0157 | 0.84511 | *CHRNA7* |
| ENSG00000171490 | 16 | 11919056 | 11955442 | 110 | 17 | 31177 | -1.0162 | 0.84524 | *RSL1D1* |
| ENSG00000142252 | 19 | 45572453 | 45604782 | 144 | 14 | 51985 | -1.0163 | 0.84525 | *GEMIN7* |
| ENSG00000177807 | 1 | 159997257 | 160050038 | 119 | 25 | 47995 | -1.017 | 0.84542 | *KCNJ10* |

| | | | | | | | | | |
|---|---|---|---|---|---|---|---|---|---|
| ENSG00000117650 | 1 | 211826114 | 211858960 | 111 | 21 | 50127 | -1.017 | 0.84543 | *NEK2* |
| ENSG00000016602 | 1 | 87002761 | 87056437 | 142 | 29 | 49063 | -1.018 | 0.84565 | *CLCA4* |
| ENSG00000159455 | 1 | 152637771 | 152669877 | 99 | 10 | 31449 | -1.0181 | 0.84568 | *LCE2B* |
| ENSG00000166902 | 11 | 59563608 | 59588345 | 72 | 10 | 34063 | -1.0188 | 0.84584 | *MRPL16* |
| ENSG00000099992 | 22 | 30677979 | 30733035 | 113 | 21 | 47298 | -1.0191 | 0.84592 | *TBC1D10A* |
| ENSG00000184432 | 3 | 139064442 | 139118574 | 139 | 15 | 30683 | -1.0196 | 0.84603 | *COPB2* |
| ENSG00000233434 | 9 | 67914791 | 67935210 | 6 | 2 | 42618 | -1.0196 | 0.84604 | *BX649567.1* |
| ENSG00000121897 | 4 | 39450620 | 39489273 | 204 | 24 | 49864 | -1.0204 | 0.84622 | *LIAS* |
| ENSG00000113318 | 5 | 79940467 | 80182279 | 809 | 49 | 50950 | -1.0204 | 0.84624 | *MSH3* |
| ENSG00000137185 | 6 | 28182664 | 28211260 | 79 | 14 | 39664 | -1.0205 | 0.84626 | *ZSCAN9* |
| ENSG00000100395 | 22 | 41591209 | 41637275 | 135 | 10 | 36841 | -1.0217 | 0.84655 | *L3MBTL2* |
| ENSG00000154330 | 9 | 70933224 | 71155977 | 469 | 77 | 37908 | -1.023 | 0.84684 | *PGM5* |
| ENSG00000170624 | 5 | 155287354 | 156204799 | 2911 | 208 | 46805 | -1.023 | 0.84685 | *SGCD* |
| ENSG00000147687 | 8 | 125490726 | 125561699 | 175 | 25 | 44432 | -1.023 | 0.84685 | *TATDN1* |
| ENSG00000142871 | 1 | 86036444 | 86059645 | 52 | 15 | 53856 | -1.0232 | 0.84688 | *CYR61* |
| ENSG00000147889 | 9 | 21957751 | 22005300 | 89 | 21 | 46782 | -1.0232 | 0.8469 | *CDKN2A* |
| ENSG00000165152 | 9 | 104225453 | 104305819 | 297 | 41 | 48030 | -1.0236 | 0.847 | *TMEM246* |
| ENSG00000180011 | 18 | 72897063 | 72931303 | 126 | 17 | 47372 | -1.0239 | 0.84706 | *ZADH2* |
| ENSG00000198125 | 22 | 35992811 | 36043998 | 196 | 25 | 39900 | -1.024 | 0.84709 | *MB* |
| ENSG00000136827 | 9 | 132565223 | 132596413 | 134 | 11 | 53029 | -1.0249 | 0.8473 | *TOR1A* |
| ENSG00000157617 | 21 | 43295221 | 43383999 | 379 | 97 | 47865 | -1.025 | 0.84732 | *C2CD2* |
| ENSG00000115844 | 2 | 172954167 | 172977628 | 60 | 17 | 51219 | -1.0253 | 0.8474 | *DLX2* |
| ENSG00000135903 | 2 | 223054607 | 223173715 | 399 | 55 | 50354 | -1.0255 | 0.84743 | *PAX3* |
| ENSG00000137393 | 6 | 18358779 | 18479105 | 677 | 63 | 49938 | -1.0257 | 0.84748 | *RNF144B* |
| ENSG00000105851 | 7 | 106495723 | 106557590 | 150 | 26 | 53491 | -1.0264 | 0.84764 | *PIK3CG* |
| ENSG00000105649 | 19 | 18297594 | 18324884 | 78 | 14 | 50456 | -1.0267 | 0.84772 | *RAB3A* |
| ENSG00000103355 | 16 | 2823954 | 2847949 | 26 | 7 | 44602 | -1.027 | 0.84779 | *PRSS33* |
| ENSG00000259202 | 15 | 67425072 | 67449277 | 72 | 22 | 45333 | -1.0279 | 0.84801 | *RP11-342M21.2* |
| ENSG00000108381 | 17 | 3365668 | 3416713 | 190 | 32 | 36490 | -1.0281 | 0.84804 | *ASPA* |
| ENSG00000221886 | 5 | 159810155 | 159837104 | 77 | 12 | 53760 | -1.0285 | 0.84814 | *C5orf54* |
| ENSG00000104299 | 8 | 28615178 | 28757759 | 260 | 33 | 42194 | -1.0288 | 0.84822 | *INTS9* |
| ENSG00000165985 | 10 | 16545742 | 16574004 | 129 | 25 | 39354 | -1.0291 | 0.84829 | *C1QL3* |
| ENSG00000104660 | 8 | 29942914 | 30044724 | 248 | 29 | 36625 | -1.0296 | 0.8484 | *LEPROTL1* |
| ENSG00000122254 | 16 | 22815498 | 22937659 | 395 | 77 | 44035 | -1.03 | 0.84849 | *HS3ST2* |
| ENSG00000138131 | 10 | 99997447 | 100038007 | 98 | 17 | 47251 | -1.0303 | 0.84856 | *LOXL4* |
| ENSG00000155011 | 4 | 107832959 | 108214963 | 1073 | 132 | 48571 | -1.0304 | 0.84859 | *DKK2* |

| | | | | | | | | | |
|---|---|---|---|---|---|---|---|---|---|
| ENSG00000179938 | 15 | 30365158 | 30395702 | 36 | 9 | 29099 | -1.0308 | 0.84869 | *GOLGA8J* |
| ENSG00000141564 | 17 | 78508619 | 78950171 | 2002 | 121 | 45642 | -1.0311 | 0.84876 | *RPTOR* |
| ENSG00000198324 | 12 | 111788455 | 111816925 | 40 | 9 | 28159 | -1.0315 | 0.84884 | *FAM109A* |
| ENSG00000151304 | 5 | 121287656 | 121421265 | 282 | 26 | 51342 | -1.0319 | 0.84895 | *SRFBP1* |
| ENSG00000166321 | 10 | 74860217 | 74901586 | 114 | 5 | 20888 | -1.0321 | 0.84898 | *NUDT13* |
| ENSG00000113100 | 5 | 26870709 | 27131257 | 662 | 68 | 39661 | -1.0325 | 0.84909 | *CDH9* |
| ENSG00000138315 | 10 | 74643339 | 74702791 | 47 | 18 | 41935 | -1.0326 | 0.8491 | *OIT3* |
| ENSG00000111261 | 12 | 12472198 | 12513475 | 208 | 30 | 49546 | -1.0331 | 0.84922 | *MANSC1* |
| ENSG00000183978 | 17 | 40937165 | 40960722 | 15 | 8 | 56457 | -1.0341 | 0.84946 | *COA3* |
| ENSG00000162433 | 1 | 65603232 | 65707828 | 246 | 38 | 37115 | -1.0347 | 0.84959 | *AK4* |
| ENSG00000174799 | 4 | 56805037 | 56909529 | 243 | 32 | 51924 | -1.0349 | 0.84964 | *CEP135* |
| ENSG00000114767 | 3 | 51957446 | 51985957 | 41 | 16 | 42989 | -1.0351 | 0.84968 | *RRP9* |
| ENSG00000114062 | 15 | 25572381 | 25694128 | 295 | 28 | 42717 | -1.0356 | 0.84981 | *UBE3A* |
| ENSG00000171136 | 19 | 14128960 | 14151854 | 65 | 11 | 35552 | -1.0357 | 0.84984 | *RLN3* |
| ENSG00000184313 | 1 | 55097463 | 55185939 | 326 | 58 | 48362 | -1.0361 | 0.84993 | *MROH7* |
| ENSG00000134531 | 12 | 13339650 | 13379708 | 108 | 27 | 48979 | -1.0364 | 0.84999 | *EMP1* |
| ENSG00000175646 | 16 | 11364693 | 11385207 | 97 | 16 | 45603 | -1.0365 | 0.85001 | *PRM1* |
| ENSG00000124092 | 20 | 56061035 | 56110708 | 176 | 37 | 51350 | -1.0369 | 0.8501 | *CTCFL* |
| ENSG00000196772 | 1 | 247968102 | 247989031 | 138 | 11 | 27232 | -1.0382 | 0.85041 | *OR14A16* |
| ENSG00000171813 | 10 | 134200672 | 134241367 | 142 | 37 | 44310 | -1.0382 | 0.85041 | *PWWP2B* |
| ENSG00000196588 | 22 | 40796285 | 41042706 | 496 | 29 | 53152 | -1.0383 | 0.85044 | *MKL1* |
| ENSG00000129084 | 11 | 14505329 | 14675181 | 275 | 38 | 52455 | -1.039 | 0.85059 | *PSMA1* |
| ENSG00000137288 | 6 | 33652070 | 33689504 | 178 | 15 | 29092 | -1.0393 | 0.85066 | *UQCC2* |
| ENSG00000152592 | 4 | 88561459 | 88595513 | 109 | 15 | 50251 | -1.0397 | 0.85076 | *DMP1* |
| ENSG00000213417 | 17 | 39211369 | 39232131 | 76 | 11 | 25363 | -1.0397 | 0.85076 | *KRTAP2-4* |
| ENSG00000138821 | 4 | 103162198 | 103362415 | 722 | 68 | 42301 | -1.0398 | 0.85078 | *SLC39A8* |
| ENSG00000185340 | 22 | 29692572 | 29718774 | 46 | 13 | 51727 | -1.0398 | 0.85079 | *GAS2L1* |
| ENSG00000099284 | 10 | 71802552 | 71882036 | 289 | 47 | 44263 | -1.0399 | 0.8508 | *H2AFY2* |
| ENSG00000161992 | 16 | 600422 | 625528 | 67 | 19 | 44722 | -1.0402 | 0.85087 | *C16orf11* |
| ENSG00000184811 | 17 | 1172957 | 1214281 | 270 | 47 | 48328 | -1.0402 | 0.85088 | *TUSC5* |
| ENSG00000141485 | 17 | 6578032 | 6626886 | 187 | 46 | 42302 | -1.0405 | 0.85093 | *SLC13A5* |
| ENSG00000186635 | 11 | 72386114 | 72514644 | 360 | 41 | 48728 | -1.0407 | 0.85099 | *ARAP1* |
| ENSG00000173391 | 12 | 10300902 | 10334737 | 152 | 17 | 31098 | -1.0407 | 0.85099 | *OLR1* |
| ENSG00000142173 | 21 | 47508011 | 47562763 | 287 | 44 | 39965 | -1.0411 | 0.85108 | *COL6A2* |
| ENSG00000206262 | 3 | 138656076 | 138682293 | 38 | 11 | 40311 | -1.0414 | 0.85115 | *C3orf72* |
| ENSG00000152208 | 4 | 93215550 | 94705707 | 4365 | 244 | 45104 | -1.0417 | 0.85122 | *GRID2* |

| | | | | | | | | | |
|---|---|---|---|---|---|---|---|---|---|
| ENSG00000204887 | 17 | 39175949 | 39196356 | 90 | 12 | 38142 | -1.0418 | 0.85125 | *KRTAP1-4* |
| ENSG00000116748 | 1 | 115205719 | 115248239 | 114 | 25 | 49271 | -1.0426 | 0.85142 | *AMPD1* |
| ENSG00000167685 | 19 | 56633968 | 56682262 | 160 | 25 | 37896 | -1.0429 | 0.85149 | *ZNF444* |
| ENSG00000065615 | 6 | 84559362 | 84687143 | 195 | 20 | 35576 | -1.0432 | 0.85157 | *CYB5R4* |
| ENSG00000130377 | 19 | 6125258 | 6203112 | 372 | 28 | 31862 | -1.0441 | 0.85179 | *ACSBG2* |
| ENSG00000106436 | 7 | 101246605 | 101282576 | 150 | 30 | 47951 | -1.0442 | 0.85181 | *MYL10* |
| ENSG00000094916 | 12 | 54614724 | 54683886 | 74 | 14 | 36640 | -1.0447 | 0.85192 | *CBX5* |
| ENSG00000198876 | 9 | 34076385 | 34137397 | 235 | 23 | 50459 | -1.0451 | 0.85201 | *DCAF12* |
| ENSG00000118473 | 1 | 66989066 | 67223982 | 904 | 61 | 35815 | -1.0452 | 0.85204 | *SGIP1* |
| ENSG00000134202 | 1 | 110266554 | 110294384 | 73 | 17 | 35432 | -1.046 | 0.85221 | *GSTM3* |
| ENSG00000116133 | 1 | 55305306 | 55362891 | 235 | 25 | 48605 | -1.046 | 0.85223 | *DHCR24* |
| ENSG00000221855 | 7 | 143164966 | 143185889 | 71 | 15 | 55693 | -1.046 | 0.85223 | *TAS2R41* |
| ENSG00000100652 | 14 | 70232134 | 70274006 | 92 | 11 | 44071 | -1.0463 | 0.85229 | *SLC10A1* |
| ENSG00000143994 | 2 | 27336682 | 27363680 | 56 | 14 | 50171 | -1.0469 | 0.85243 | *ABHD1* |
| ENSG00000149403 | 11 | 120372468 | 120869613 | 1488 | 193 | 44243 | -1.0481 | 0.8527 | *GRIK4* |
| ENSG00000108839 | 17 | 6889384 | 6924055 | 107 | 13 | 50627 | -1.0491 | 0.85294 | *ALOX12* |
| ENSG00000165325 | 11 | 93053137 | 93181653 | 464 | 37 | 46217 | -1.0502 | 0.85318 | *CCDC67* |
| ENSG00000231925 | 6 | 33257471 | 33292164 | 71 | 14 | 33161 | -1.0511 | 0.8534 | *TAPBP* |
| ENSG00000197706 | 12 | 55630982 | 55652086 | 62 | 5 | 44333 | -1.0513 | 0.85344 | *OR6C74* |
| ENSG00000161671 | 19 | 50969657 | 50996608 | 89 | 19 | 39850 | -1.0513 | 0.85344 | *EMC10* |
| ENSG00000221961 | 2 | 240971230 | 240992399 | 118 | 21 | 52080 | -1.0514 | 0.85347 | *PRR21* |
| ENSG00000114107 | 3 | 138203186 | 138323380 | 206 | 16 | 43585 | -1.0521 | 0.85362 | *CEP70* |
| ENSG00000197122 | 20 | 35963088 | 36044453 | 221 | 38 | 39571 | -1.0522 | 0.85366 | *SRC* |
| ENSG00000165650 | 10 | 119030000 | 119144978 | 356 | 21 | 39031 | -1.0529 | 0.8538 | *PDZD8* |
| ENSG00000115539 | 2 | 101169152 | 101203197 | 117 | 15 | 32984 | -1.0534 | 0.85391 | *PDCL3* |
| ENSG00000137270 | 6 | 52981762 | 53023627 | 177 | 28 | 42979 | -1.0534 | 0.85391 | *GCM1* |
| ENSG00000023041 | 10 | 114180058 | 114216672 | 70 | 20 | 45715 | -1.0537 | 0.85399 | *ZDHHC6* |
| ENSG00000224226 | 17 | 34483061 | 34513996 | 9 | 6 | 34479 | -1.0541 | 0.85407 | *TBC1D3B* |
| ENSG00000067533 | 1 | 218448629 | 218521325 | 131 | 15 | 41400 | -1.0547 | 0.85422 | *RRP15* |
| ENSG00000077238 | 16 | 27314989 | 27386099 | 253 | 40 | 46956 | -1.0547 | 0.85422 | *IL4R* |
| ENSG00000146858 | 7 | 138700452 | 138730775 | 133 | 10 | 47200 | -1.0549 | 0.85427 | *ZC3HAV1L* |
| ENSG00000164400 | 5 | 131399483 | 131421859 | 48 | 10 | 28649 | -1.0552 | 0.85434 | *CSF2* |
| ENSG00000178279 | 16 | 11351605 | 11373390 | 93 | 17 | 40511 | -1.0552 | 0.85434 | *TNP2* |
| ENSG00000272442 | 6 | 44230931 | 44356694 | 378 | 58 | 43391 | -1.0555 | 0.8544 | *RP11-444E17.6* |
| ENSG00000116353 | 1 | 29509385 | 29567454 | 174 | 23 | 34439 | -1.0555 | 0.85441 | *MECR* |
| ENSG00000258881 | 2 | 71156448 | 71232466 | 261 | 62 | 45805 | -1.0558 | 0.85448 | *AC007040.11* |

| | | | | | | | | | |
|---|---|---|---|---|---|---|---|---|---|
| ENSG00000050393 | 6 | 13776789 | 13824800 | 115 | 29 | 43344 | -1.0562 | 0.85457 | *MCUR1* |
| ENSG00000185813 | 17 | 79848841 | 79879340 | 25 | 16 | 50290 | -1.0573 | 0.85482 | *PCYT2* |
| ENSG00000166069 | 15 | 38204140 | 38269925 | 131 | 21 | 47924 | -1.0577 | 0.85491 | *TMCO5A* |
| ENSG00000213859 | 17 | 7245208 | 7268258 | 44 | 19 | 44154 | -1.0589 | 0.85517 | *KCTD11* |
| ENSG00000099860 | 19 | 2466120 | 2488257 | 96 | 28 | 50016 | -1.059 | 0.85521 | *GADD45B* |
| ENSG00000236104 | 6 | 33272183 | 33295719 | 46 | 8 | 34154 | -1.0591 | 0.85523 | *ZBTB22* |
| ENSG00000123407 | 12 | 54338618 | 54362740 | 52 | 13 | 47891 | -1.0591 | 0.85523 | *HOXC12* |
| ENSG00000176136 | 18 | 13814154 | 13836861 | 159 | 17 | 44884 | -1.0592 | 0.85524 | *MC5R* |
| ENSG00000204533 | 19 | 56652314 | 56673250 | 81 | 16 | 37429 | -1.0597 | 0.85535 | *AC024580.1* |
| ENSG00000103274 | 16 | 10827643 | 10873208 | 139 | 18 | 42213 | -1.0598 | 0.85538 | *NUBP1* |
| ENSG00000144285 | 2 | 166835670 | 166994523 | 547 | 49 | 47086 | -1.0603 | 0.85549 | *SCN1A* |
| ENSG00000107295 | 9 | 17569080 | 17807127 | 1212 | 107 | 45642 | -1.0605 | 0.85553 | *SH3GL2* |
| ENSG00000164442 | 6 | 139683393 | 139705757 | 69 | 16 | 43218 | -1.0605 | 0.85555 | *CITED2* |
| ENSG00000164399 | 5 | 131386222 | 131408897 | 44 | 8 | 28031 | -1.0609 | 0.85563 | *IL3* |
| ENSG00000197651 | 12 | 91289399 | 91358953 | 200 | 28 | 36489 | -1.0609 | 0.85564 | *CCER1* |
| ENSG00000095397 | 9 | 117154360 | 117277730 | 418 | 47 | 45867 | -1.0615 | 0.85578 | *DFNB31* |
| ENSG00000151806 | 4 | 44670444 | 44712943 | 101 | 10 | 36365 | -1.0619 | 0.85585 | *GUF1* |
| ENSG00000049167 | 5 | 60159658 | 60250900 | 270 | 28 | 47282 | -1.0619 | 0.85585 | *ERCC8* |
| ENSG00000273170 | 9 | 38530566 | 38587204 | 128 | 45 | 39582 | -1.062 | 0.85589 | *ANKRD18A* |
| ENSG00000136868 | 9 | 115973808 | 116038674 | 231 | 30 | 46924 | -1.0624 | 0.85597 | *SLC31A1* |
| ENSG00000099822 | 19 | 579893 | 627159 | 270 | 64 | 37688 | -1.0627 | 0.85603 | *HCN2* |
| ENSG00000171316 | 8 | 61581337 | 61789465 | 481 | 46 | 40044 | -1.0629 | 0.8561 | *CHD7* |
| ENSG00000041357 | 15 | 78822747 | 78851604 | 96 | 9 | 31710 | -1.0631 | 0.85613 | *PSMA4* |
| ENSG00000213366 | 1 | 110200644 | 110262171 | 144 | 30 | 37056 | -1.0639 | 0.85632 | *GSTM2* |
| ENSG00000133393 | 16 | 15949577 | 15992482 | 298 | 10 | 54022 | -1.0643 | 0.8564 | *FOPNL* |
| ENSG00000101298 | 20 | 1236960 | 1299972 | 227 | 45 | 46893 | -1.0645 | 0.85644 | *SNPH* |
| ENSG00000268434 | 19 | 46272695 | 46299231 | 43 | 11 | 50995 | -1.0647 | 0.85649 | *AC011530.4* |
| ENSG00000152763 | 1 | 67268568 | 67400570 | 303 | 52 | 47749 | -1.065 | 0.85656 | *WDR78* |
| ENSG00000214510 | 5 | 147637743 | 147675817 | 45 | 16 | 41668 | -1.0654 | 0.85665 | *SPINK13* |
| ENSG00000135486 | 12 | 54663977 | 54690872 | 31 | 9 | 33903 | -1.0664 | 0.85688 | *HNRNPA1* |
| ENSG00000127920 | 7 | 93541011 | 93567922 | 101 | 17 | 50877 | -1.0665 | 0.8569 | *GNG11* |
| ENSG00000127588 | 16 | 838041 | 860733 | 127 | 28 | 38330 | -1.0665 | 0.8569 | *GNG13* |
| ENSG00000134809 | 11 | 57285936 | 57308276 | 47 | 12 | 53790 | -1.067 | 0.85701 | *TIMM10* |
| ENSG00000149294 | 11 | 112821997 | 113159158 | 1084 | 94 | 42998 | -1.0671 | 0.85705 | *NCAM1* |
| ENSG00000111684 | 12 | 7075348 | 7135814 | 100 | 12 | 35122 | -1.0672 | 0.85706 | *LPCAT3* |
| ENSG00000172062 | 5 | 70210768 | 70259769 | 4 | 2 | 35065 | -1.0674 | 0.8571 | *SMN1* |

| | | | | | | | | | |
|---|---|---|---|---|---|---|---|---|---|
| ENSG00000165029 | 9 | 107533283 | 107700518 | 721 | 124 | 43824 | -1.0678 | 0.85719 | *ABCA1* |
| ENSG00000174255 | 3 | 113943483 | 113966425 | 103 | 24 | 51153 | -1.0688 | 0.85742 | *ZNF80* |
| ENSG00000168010 | 11 | 72515353 | 72564719 | 77 | 18 | 53240 | -1.0699 | 0.85767 | *ATG16L2* |
| ENSG00000243678 | 17 | 49220920 | 49259108 | 148 | 24 | 49566 | -1.0702 | 0.85774 | *NME1-NME2* |
| ENSG00000011052 | 17 | 49220951 | 49259105 | 148 | 24 | 49566 | -1.0702 | 0.85774 | *NME2* |
| ENSG00000101220 | 20 | 3724155 | 3759034 | 126 | 38 | 50215 | -1.0703 | 0.85776 | *C20orf27* |
| ENSG00000170647 | 11 | 100852811 | 100874663 | 99 | 23 | 45960 | -1.0704 | 0.85778 | *TMEM133* |
| ENSG00000122912 | 10 | 70227756 | 70297231 | 312 | 28 | 37121 | -1.0705 | 0.8578 | *SLC25A16* |
| ENSG00000152558 | 11 | 102257063 | 102351115 | 228 | 35 | 53318 | -1.0705 | 0.8578 | *TMEM123* |
| ENSG00000159248 | 15 | 35033233 | 35057166 | 87 | 19 | 47006 | -1.0707 | 0.85784 | *GJD2* |
| ENSG00000241127 | 7 | 39595975 | 39659919 | 230 | 33 | 41374 | -1.0708 | 0.85787 | *YAE1D1* |
| ENSG00000122224 | 1 | 160755864 | 160808051 | 161 | 17 | 43214 | -1.071 | 0.85791 | *LY9* |
| ENSG00000242247 | 22 | 43182508 | 43264112 | 298 | 23 | 48210 | -1.0711 | 0.85793 | *ARFGAP3* |
| ENSG00000224877 | 17 | 79203039 | 79225081 | 80 | 19 | 35254 | -1.0727 | 0.85829 | *C17orf89* |
| ENSG00000104904 | 19 | 2259485 | 2283487 | 98 | 16 | 24710 | -1.0728 | 0.85831 | *OAZ1* |
| ENSG00000066735 | 14 | 104595060 | 104657231 | 260 | 55 | 47472 | -1.0731 | 0.85839 | *KIF26A* |
| ENSG00000170364 | 3 | 4334988 | 4369251 | 93 | 19 | 29927 | -1.0733 | 0.85843 | *SETMAR* |
| ENSG00000214285 | 10 | 129337613 | 129360935 | 195 | 20 | 36230 | -1.0746 | 0.85873 | *NPS* |
| ENSG00000196805 | 1 | 153032700 | 153123927 | 275 | 31 | 32260 | -1.0748 | 0.85876 | *SPRR2B* |
| ENSG00000128342 | 22 | 30626436 | 30652840 | 42 | 16 | 49560 | -1.0749 | 0.85879 | *LIF* |
| ENSG00000170471 | 20 | 37091459 | 37217504 | 223 | 30 | 37050 | -1.0751 | 0.85883 | *RALGAPB* |
| ENSG00000213281 | 1 | 115237090 | 115269515 | 48 | 13 | 48008 | -1.0752 | 0.85885 | *NRAS* |
| ENSG00000132475 | 17 | 73762515 | 73791974 | 54 | 11 | 32375 | -1.0753 | 0.85887 | *H3F3B* |
| ENSG00000121481 | 1 | 185004496 | 185081740 | 103 | 20 | 47438 | -1.0757 | 0.85898 | *RNF2* |
| ENSG00000179869 | 7 | 48201055 | 48697092 | 2102 | 129 | 47481 | -1.076 | 0.85903 | *ABCA13* |
| ENSG00000100206 | 22 | 38904954 | 38976291 | 127 | 24 | 42103 | -1.0764 | 0.85913 | *DMC1* |
| ENSG00000173335 | 20 | 23573047 | 23596513 | 77 | 7 | 43091 | -1.0767 | 0.8592 | *CST9* |
| ENSG00000060982 | 12 | 24954295 | 25112393 | 690 | 93 | 48851 | -1.077 | 0.85925 | *BCAT1* |
| ENSG00000184470 | 22 | 19853040 | 19939341 | 415 | 55 | 45377 | -1.0773 | 0.85934 | *TXNRD2* |
| ENSG00000108950 | 17 | 66521254 | 66607530 | 227 | 45 | 53298 | -1.0777 | 0.85941 | *FAM20A* |
| ENSG00000139648 | 12 | 52927693 | 52956931 | 130 | 24 | 37727 | -1.0778 | 0.85945 | *KRT71* |
| ENSG00000135917 | 2 | 228539926 | 228592728 | 240 | 29 | 36570 | -1.0779 | 0.85947 | *SLC19A3* |
| ENSG00000124140 | 20 | 44640356 | 44698784 | 190 | 25 | 49322 | -1.0781 | 0.85952 | *SLC12A5* |
| ENSG00000173124 | 10 | 96943957 | 96998685 | 210 | 16 | 43104 | -1.0789 | 0.85968 | *C10orf129* |
| ENSG00000060140 | 12 | 10761538 | 10836917 | 220 | 21 | 46849 | -1.079 | 0.85972 | *STYK1* |
| ENSG00000105647 | 19 | 18253928 | 18291350 | 101 | 20 | 51665 | -1.0791 | 0.85973 | *PIK3R2* |

| | | | | | | | | | |
|---|---|---|---|---|---|---|---|---|---|
| ENSG00000072041 | 12 | 85243492 | 85317394 | 137 | 20 | 41242 | -1.0792 | 0.85975 | *SLC6A15* |
| ENSG00000163166 | 2 | 128183783 | 128294462 | 310 | 36 | 41698 | -1.0794 | 0.85979 | *IWS1* |
| ENSG00000144677 | 3 | 37893451 | 38035960 | 357 | 47 | 48032 | -1.0794 | 0.8598 | *CTDSPL* |
| ENSG00000136247 | 7 | 6607065 | 6639005 | 106 | 15 | 49301 | -1.0798 | 0.85988 | *ZDHHC4* |
| ENSG00000186111 | 19 | 3620181 | 3710477 | 275 | 47 | 45952 | -1.0799 | 0.85991 | *PIP5K1C* |
| ENSG00000001461 | 1 | 24732284 | 24809466 | 226 | 26 | 47085 | -1.0805 | 0.86004 | *NIPAL3* |
| ENSG00000204704 | 6 | 29001990 | 29023017 | 108 | 15 | 39289 | -1.0808 | 0.86011 | *OR2W1* |
| ENSG00000122068 | 3 | 197454050 | 197524467 | 156 | 24 | 40063 | -1.0808 | 0.86012 | *FYTTD1* |
| ENSG00000196873 | 9 | 70846397 | 70924929 | 2 | 1 | 49271 | -1.0815 | 0.86025 | *CBWD3* |
| ENSG00000106404 | 7 | 100865373 | 100892101 | 60 | 11 | 44755 | -1.0819 | 0.86036 | *CLDN15* |
| ENSG00000105968 | 7 | 44856390 | 44897682 | 88 | 18 | 38783 | -1.082 | 0.86037 | *H2AFV* |
| ENSG00000171561 | 11 | 74789758 | 74810799 | 42 | 10 | 45316 | -1.0826 | 0.86051 | *OR2AT4* |
| ENSG00000111696 | 12 | 104154231 | 104244975 | 456 | 37 | 45066 | -1.0827 | 0.86053 | *NT5DC3* |
| ENSG00000123095 | 12 | 26262959 | 26288060 | 60 | 16 | 46775 | -1.0832 | 0.86064 | *BHLHE41* |
| ENSG00000142192 | 21 | 27242861 | 27553446 | 898 | 120 | 41152 | -1.0832 | 0.86064 | *APP* |
| ENSG00000171487 | 19 | 56501092 | 56583179 | 657 | 113 | 48750 | -1.0837 | 0.86076 | *NLRP5* |
| ENSG00000187257 | 7 | 77315760 | 77422339 | 327 | 28 | 37091 | -1.0843 | 0.86088 | *RSBN1L* |
| ENSG00000127990 | 7 | 94204542 | 94295521 | 146 | 18 | 52182 | -1.0846 | 0.86095 | *SGCE* |
| ENSG00000108479 | 17 | 73737675 | 73771792 | 72 | 26 | 41607 | -1.0846 | 0.86096 | *GALK1* |
| ENSG00000188997 | 11 | 77872295 | 77909868 | 85 | 18 | 50393 | -1.0848 | 0.861 | *KCTD21* |
| ENSG00000268173 | 19 | 18253968 | 18298927 | 120 | 23 | 51314 | -1.0849 | 0.86102 | *PIK3R2* |
| ENSG00000029725 | 17 | 5175558 | 5299129 | 513 | 46 | 37274 | -1.085 | 0.86104 | *RABEP1* |
| ENSG00000186306 | 1 | 158358312 | 158379256 | 118 | 23 | 44076 | -1.0854 | 0.86114 | *OR10T2* |
| ENSG00000119125 | 9 | 74719511 | 74882109 | 545 | 53 | 46711 | -1.0857 | 0.86118 | *GDA* |
| ENSG00000197461 | 7 | 526895 | 569933 | 165 | 33 | 43843 | -1.0869 | 0.86147 | *PDGFA* |
| ENSG00000204306 | 11 | 120372511 | 120395732 | 72 | 23 | 42307 | -1.0878 | 0.86166 | *AP002348.1* |
| ENSG00000120158 | 9 | 4782869 | 4895917 | 426 | 58 | 39110 | -1.0881 | 0.86172 | *RCL1* |
| ENSG00000139515 | 13 | 28484157 | 28510368 | 65 | 13 | 48683 | -1.0883 | 0.86177 | *PDX1* |
| ENSG00000163428 | 3 | 120033356 | 120078186 | 145 | 19 | 43421 | -1.0888 | 0.86189 | *LRRC58* |
| ENSG00000133115 | 13 | 39530062 | 39575203 | 232 | 25 | 48097 | -1.089 | 0.86192 | *STOML3* |
| ENSG00000001460 | 1 | 24673489 | 24753424 | 242 | 28 | 48657 | -1.0896 | 0.86206 | *STPG1* |
| ENSG00000186314 | 5 | 144841362 | 145224932 | 1015 | 63 | 36078 | -1.0898 | 0.86209 | *PRELID2* |
| ENSG00000099984 | 22 | 24312339 | 24336106 | 26 | 12 | 43367 | -1.0898 | 0.8621 | *GSTT2* |
| ENSG00000188910 | 1 | 35236790 | 35261970 | 95 | 18 | 44776 | -1.0901 | 0.86216 | *GJB3* |
| ENSG00000108561 | 17 | 5326097 | 5362150 | 128 | 24 | 37576 | -1.091 | 0.86235 | *C1QBP* |
| ENSG00000007255 | 19 | 45656186 | 45691495 | 94 | 21 | 53105 | -1.0916 | 0.86251 | *TRAPPC6A* |

| | | | | | | | | | |
|---|---|---|---|---|---|---|---|---|---|
| ENSG00000174243 | 12 | 49213547 | 49256625 | 82 | 20 | 50844 | -1.0917 | 0.86252 | *DDX23* |
| ENSG00000248789 | 5 | 51293865 | 51386821 | 329 | 22 | 42980 | -1.0922 | 0.86262 | *CTD-2203A3.1* |
| ENSG00000220575 | 7 | 154848777 | 154873267 | 85 | 12 | 45052 | -1.0923 | 0.86264 | *HTR5A-AS1* |
| ENSG00000136003 | 12 | 108946358 | 108973160 | 106 | 14 | 33658 | -1.0927 | 0.86274 | *ISCU* |
| ENSG00000023318 | 9 | 102731461 | 102871322 | 275 | 25 | 31815 | -1.0932 | 0.86285 | *ERP44* |
| ENSG00000182885 | 16 | 57692099 | 57733975 | 147 | 38 | 38551 | -1.0939 | 0.863 | *GPR97* |
| ENSG00000161204 | 3 | 183893811 | 183921800 | 72 | 13 | 56017 | -1.0944 | 0.86311 | *ABCF3* |
| ENSG00000162775 | 1 | 110871128 | 110899299 | 64 | 11 | 47468 | -1.0947 | 0.86318 | *RBM15* |
| ENSG00000177335 | 8 | 144110626 | 144151359 | 142 | 14 | 36784 | -1.0949 | 0.86321 | *C8orf31* |
| ENSG00000258130 | 16 | 30205637 | 30230392 | 2 | 1 | 42773 | -1.095 | 0.86324 | *RP11-347C12.3* |
| ENSG00000260456 | 16 | 87107168 | 87361022 | 1216 | 128 | 39618 | -1.095 | 0.86324 | *C16orf95* |
| ENSG00000204070 | 20 | 43980577 | 44015438 | 92 | 9 | 41107 | -1.0951 | 0.86325 | *SYS1* |
| ENSG00000023330 | 3 | 52222102 | 52258343 | 39 | 8 | 56040 | -1.0956 | 0.86338 | *ALAS1* |
| ENSG00000073734 | 2 | 169769448 | 169897832 | 450 | 55 | 45266 | -1.0958 | 0.86342 | *ABCB11* |
| ENSG00000166527 | 12 | 8652071 | 8684962 | 83 | 10 | 32459 | -1.0963 | 0.86352 | *CLEC4D* |
| ENSG00000170820 | 2 | 49179296 | 49391676 | 904 | 71 | 45567 | -1.0969 | 0.86365 | *FSHR* |
| ENSG00000129515 | 14 | 35020300 | 35109389 | 261 | 26 | 41840 | -1.0971 | 0.8637 | *SNX6* |
| ENSG00000135503 | 12 | 52335451 | 52400862 | 130 | 24 | 49735 | -1.0976 | 0.86382 | *ACVR1B* |
| ENSG00000035862 | 17 | 76839059 | 76931469 | 385 | 56 | 45571 | -1.0986 | 0.86403 | *TIMP2* |
| ENSG00000165832 | 10 | 116687952 | 116747430 | 163 | 21 | 32422 | -1.0993 | 0.86419 | *TRUB1* |
| ENSG00000164645 | 7 | 88413420 | 88435035 | 78 | 13 | 37518 | -1.0994 | 0.8642 | *C7orf62* |
| ENSG00000188243 | 13 | 76089350 | 76133575 | 144 | 23 | 46168 | -1.0994 | 0.86421 | *COMMD6* |
| ENSG00000188558 | 1 | 248674916 | 248695964 | 39 | 5 | 33998 | -1.1 | 0.86434 | *OR2G6* |
| ENSG00000196240 | 1 | 248606077 | 248627130 | 11 | 2 | 38552 | -1.1001 | 0.86436 | *OR2T2* |
| ENSG00000215045 | 7 | 6527093 | 6601067 | 220 | 47 | 43089 | -1.1003 | 0.8644 | *GRID2IP* |
| ENSG00000092096 | 14 | 23805515 | 23832121 | 73 | 19 | 41279 | -1.1004 | 0.86441 | *SLC22A17* |
| ENSG00000175894 | 21 | 45907775 | 46141495 | 985 | 101 | 45734 | -1.1004 | 0.86443 | *TSPEAR* |
| ENSG00000164919 | 8 | 100875428 | 100916290 | 99 | 13 | 42216 | -1.1005 | 0.86444 | *COX6C* |
| ENSG00000225885 | 8 | 119284378 | 119422844 | 468 | 47 | 35737 | -1.1005 | 0.86444 | *AC023590.1* |
| ENSG00000155249 | 14 | 20393767 | 20414842 | 19 | 6 | 43958 | -1.1012 | 0.86459 | *OR4K1* |
| ENSG00000173572 | 19 | 56393065 | 56453702 | 278 | 44 | 50114 | -1.1013 | 0.86462 | *NLRP13* |
| ENSG00000130775 | 1 | 28189055 | 28223196 | 104 | 19 | 41712 | -1.1015 | 0.86467 | *THEMIS2* |
| ENSG00000268241 | 2 | 175190601 | 175212151 | 66 | 19 | 35549 | -1.1017 | 0.8647 | *AC018470.1* |
| ENSG00000269879 | 10 | 24760273 | 24780632 | 66 | 9 | 51747 | -1.1017 | 0.86471 | *AL353583.1* |
| ENSG00000268730 | 15 | 99849300 | 99869828 | 74 | 14 | 46817 | -1.1019 | 0.86474 | *AC022819.2* |
| ENSG00000136986 | 8 | 124015404 | 124064663 | 156 | 27 | 46885 | -1.1021 | 0.86479 | *DERL1* |

| | | | | | | | | | |
|---|---|---|---|---|---|---|---|---|---|
| ENSG00000164323 | 4 | 186070819 | 186140658 | 287 | 20 | 50288 | -1.1021 | 0.8648 | *KIAA1430* |
| ENSG00000259471 | 15 | 66864528 | 66988132 | 570 | 84 | 46410 | -1.1022 | 0.86482 | *RP11-321F6.1* |
| ENSG00000091137 | 7 | 107291080 | 107368254 | 153 | 30 | 49911 | -1.1034 | 0.86507 | *SLC26A4* |
| ENSG00000257108 | 16 | 606996 | 629495 | 69 | 14 | 45833 | -1.1037 | 0.86514 | *NHLRC4* |
| ENSG00000158406 | 6 | 26271283 | 26295762 | 105 | 14 | 30877 | -1.1047 | 0.86535 | *HIST1H4H* |
| ENSG00000136828 | 9 | 129667053 | 129995445 | 647 | 46 | 40373 | -1.1047 | 0.86535 | *RALGPS1* |
| ENSG00000168158 | 16 | 3395889 | 3416924 | 106 | 13 | 54603 | -1.1047 | 0.86535 | *OR2C1* |
| ENSG00000174348 | 1 | 53517854 | 53561174 | 178 | 25 | 41851 | -1.1048 | 0.86537 | *PODN* |
| ENSG00000170627 | 12 | 54839734 | 54877386 | 122 | 14 | 47297 | -1.1048 | 0.86537 | *GTSF1* |
| ENSG00000130201 | 19 | 45705879 | 45747469 | 211 | 27 | 36915 | -1.1057 | 0.86558 | *EXOC3L2* |
| ENSG00000185899 | 7 | 143130546 | 143151502 | 44 | 10 | 47605 | -1.1058 | 0.8656 | *TAS2R60* |
| ENSG00000124608 | 6 | 44257391 | 44291063 | 118 | 22 | 41688 | -1.1063 | 0.86571 | *AARS2* |
| ENSG00000136451 | 17 | 56038910 | 56075620 | 79 | 13 | 51557 | -1.1069 | 0.86583 | *VEZF1* |
| ENSG00000049245 | 1 | 7821329 | 7851492 | 103 | 21 | 53416 | -1.107 | 0.86585 | *VAMP3* |
| ENSG00000156650 | 10 | 76575340 | 76802380 | 317 | 39 | 26770 | -1.108 | 0.86606 | *KAT6B* |
| ENSG00000025800 | 1 | 32563639 | 32652169 | 134 | 16 | 33058 | -1.1081 | 0.8661 | *KPNA6* |
| ENSG00000197745 | 11 | 62053754 | 62076536 | 108 | 10 | 24500 | -1.1083 | 0.86613 | *SCGB1D4* |
| ENSG00000126749 | 12 | 7062408 | 7115520 | 79 | 13 | 38176 | -1.1084 | 0.86616 | *EMG1* |
| ENSG00000059573 | 10 | 97355696 | 97426463 | 279 | 30 | 37301 | -1.1085 | 0.86617 | *ALDH18A1* |
| ENSG00000163840 | 3 | 122273085 | 122304050 | 138 | 18 | 53938 | -1.1087 | 0.86622 | *DTX3L* |
| ENSG00000180543 | 8 | 98275717 | 98300176 | 135 | 17 | 37158 | -1.109 | 0.86629 | *TSPYL5* |
| ENSG00000101074 | 20 | 42955626 | 42989875 | 134 | 12 | 54658 | -1.1099 | 0.86648 | *R3HDML* |
| ENSG00000127616 | 19 | 11061598 | 11186071 | 377 | 47 | 51491 | -1.1102 | 0.86654 | *SMARCA4* |
| ENSG00000180155 | 8 | 143835752 | 143869640 | 131 | 24 | 44696 | -1.1109 | 0.86669 | *LYNX1* |
| ENSG00000185527 | 17 | 79607489 | 79640142 | 256 | 21 | 32421 | -1.1119 | 0.86691 | *PDE6G* |
| ENSG00000125868 | 20 | 17540508 | 17600564 | 170 | 24 | 28243 | -1.112 | 0.86694 | *DSTN* |
| ENSG00000173890 | 3 | 169745717 | 169813191 | 177 | 19 | 49934 | -1.1123 | 0.86699 | *GPR160* |
| ENSG00000203690 | 6 | 167760521 | 167807998 | 203 | 37 | 50094 | -1.1138 | 0.86733 | *TCP10* |
| ENSG00000152270 | 11 | 14655269 | 14902350 | 391 | 39 | 45489 | -1.1139 | 0.86735 | *PDE3B* |
| ENSG00000068724 | 2 | 47133296 | 47313276 | 816 | 105 | 47321 | -1.1146 | 0.86749 | *TTC7A* |
| ENSG00000171759 | 12 | 103220663 | 103362188 | 515 | 53 | 44810 | -1.1146 | 0.86749 | *PAH* |
| ENSG00000142733 | 1 | 27671675 | 27703383 | 79 | 27 | 39601 | -1.1151 | 0.86759 | *MAP3K6* |
| ENSG00000169607 | 2 | 113483930 | 113532254 | 127 | 17 | 46266 | -1.1157 | 0.86773 | *CKAP2L* |
| ENSG00000064419 | 7 | 128584948 | 128705198 | 312 | 30 | 52332 | -1.1158 | 0.86775 | *TNPO3* |
| ENSG00000182149 | 16 | 71869899 | 71972913 | 260 | 28 | 45204 | -1.1161 | 0.86781 | *IST1* |
| ENSG00000213424 | 17 | 38800917 | 38831433 | 66 | 8 | 50127 | -1.1161 | 0.86781 | *KRT222* |

| | | | | | | | | | |
|---|---|---|---|---|---|---|---|---|---|
| ENSG00000023909 | 1 | 94340761 | 94384966 | 168 | 23 | 41130 | -1.1162 | 0.86783 | *GCLM* |
| ENSG00000182783 | 1 | 248711784 | 248732797 | 4 | 2 | 35314 | -1.117 | 0.868 | *OR2T29* |
| ENSG00000188559 | 20 | 20360196 | 20703131 | 497 | 43 | 46334 | -1.117 | 0.86801 | *RALGAPA2* |
| ENSG00000107779 | 10 | 88506407 | 88702595 | 596 | 53 | 34073 | -1.1175 | 0.86811 | *BMPR1A* |
| ENSG00000198271 | 17 | 39295178 | 39316054 | 54 | 13 | 36122 | -1.118 | 0.86821 | *KRTAP4-5* |
| ENSG00000165092 | 9 | 75505578 | 75705358 | 405 | 51 | 42112 | -1.1182 | 0.86827 | *ALDH1A1* |
| ENSG00000099385 | 16 | 30834947 | 30916281 | 105 | 15 | 28752 | -1.119 | 0.86843 | *BCL7C* |
| ENSG00000111727 | 12 | 104448235 | 104508639 | 164 | 30 | 50868 | -1.1195 | 0.86854 | *HCFC2* |
| ENSG00000171302 | 17 | 76977799 | 77015949 | 138 | 24 | 43454 | -1.1211 | 0.86887 | *CANT1* |
| ENSG00000173200 | 3 | 122286449 | 122367894 | 329 | 41 | 46995 | -1.1211 | 0.86888 | *PARP15* |
| ENSG00000103310 | 16 | 21198773 | 21235831 | 114 | 22 | 50269 | -1.1214 | 0.86895 | *ZP2* |
| ENSG00000204519 | 19 | 58183337 | 58238669 | 146 | 27 | 48295 | -1.1215 | 0.86897 | *ZNF551* |
| ENSG00000072135 | 2 | 131103580 | 131142982 | 57 | 14 | 39658 | -1.1221 | 0.86909 | *PTPN18* |
| ENSG00000166912 | 15 | 31221144 | 31293810 | 234 | 23 | 42594 | -1.1221 | 0.86909 | *MTMR10* |
| ENSG00000176563 | 17 | 40940810 | 40973605 | 21 | 11 | 51751 | -1.1228 | 0.86923 | *CNTD1* |
| ENSG00000151552 | 4 | 17451884 | 17523857 | 306 | 47 | 50091 | -1.1228 | 0.86924 | *QDPR* |
| ENSG00000148908 | 10 | 121249340 | 121312220 | 208 | 38 | 46981 | -1.1228 | 0.86924 | *RGS10* |
| ENSG00000107890 | 10 | 27270843 | 27399421 | 485 | 38 | 40292 | -1.1231 | 0.86929 | *ANKRD26* |
| ENSG00000215009 | 12 | 7446880 | 7491320 | 107 | 17 | 47266 | -1.1231 | 0.86931 | *ACSM4* |
| ENSG00000124721 | 6 | 38673117 | 39008301 | 1329 | 93 | 47934 | -1.124 | 0.86949 | *DNAH8* |
| ENSG00000161849 | 12 | 52761596 | 52789436 | 160 | 15 | 31043 | -1.1242 | 0.86955 | *KRT84* |
| ENSG00000197992 | 12 | 10173276 | 10228565 | 94 | 28 | 41401 | -1.1246 | 0.86962 | *CLEC9A* |
| ENSG00000125879 | 20 | 16719003 | 16760707 | 198 | 36 | 39267 | -1.1246 | 0.86963 | *OTOR* |
| ENSG00000109832 | 11 | 125763271 | 125803158 | 137 | 27 | 47599 | -1.1248 | 0.86966 | *DDX25* |
| ENSG00000134152 | 15 | 34422875 | 34512297 | 363 | 27 | 46596 | -1.1252 | 0.86975 | *KATNBL1* |
| ENSG00000212721 | 17 | 39263433 | 39284606 | 152 | 7 | 24272 | -1.1259 | 0.8699 | *KRTAP4-11* |
| ENSG00000269657 | 17 | 66529369 | 66552970 | 69 | 12 | 55663 | -1.1263 | 0.86998 | *AC079210.1* |
| ENSG00000183303 | 11 | 7807452 | 7828520 | 86 | 8 | 37958 | -1.1264 | 0.87 | *OR5P2* |
| ENSG00000222001 | 2 | 233867324 | 233890595 | 73 | 20 | 39406 | -1.1268 | 0.87009 | *AC106876.2* |
| ENSG00000169302 | 5 | 146604526 | 146777415 | 749 | 79 | 47253 | -1.1268 | 0.87009 | *STK32A* |
| ENSG00000175643 | 16 | 11333476 | 11455619 | 533 | 48 | 46358 | -1.1271 | 0.87015 | *RMI2* |
| ENSG00000107758 | 10 | 75186186 | 75265782 | 78 | 16 | 36674 | -1.1272 | 0.87017 | *PPP3CB* |
| ENSG00000185608 | 22 | 19409425 | 19433598 | 51 | 14 | 41457 | -1.1273 | 0.87018 | *MRPL40* |
| ENSG00000008118 | 1 | 209747062 | 209797283 | 178 | 29 | 42418 | -1.1275 | 0.87022 | *CAMK1G* |
| ENSG00000185728 | 8 | 64071112 | 64135346 | 112 | 27 | 42365 | -1.1278 | 0.87031 | *YTHDF3* |
| ENSG00000122304 | 16 | 11359496 | 11380337 | 93 | 16 | 41623 | -1.1279 | 0.87033 | *PRM2* |

| | | | | | | | | | |
|---|---|---|---|---|---|---|---|---|---|
| ENSG00000104213 | 8 | 17423942 | 17511580 | 674 | 95 | 40907 | -1.129 | 0.87055 | *PDGFRL* |
| ENSG00000142549 | 19 | 51805102 | 51843608 | 105 | 20 | 37384 | -1.129 | 0.87055 | *IGLON5* |
| ENSG00000086475 | 10 | 13349424 | 13400297 | 196 | 26 | 45516 | -1.1308 | 0.87094 | *SEPHS1* |
| ENSG00000197584 | 3 | 177980720 | 178572217 | 1943 | 171 | 45375 | -1.1309 | 0.87096 | *KCNMB2* |
| ENSG00000103888 | 15 | 81061684 | 81254117 | 400 | 104 | 44729 | -1.1311 | 0.87099 | *KIAA1199* |
| ENSG00000115514 | 2 | 99925445 | 99967165 | 116 | 16 | 49663 | -1.1318 | 0.87114 | *TXNDC9* |
| ENSG00000139133 | 12 | 34165216 | 34192629 | 62 | 10 | 37285 | -1.1318 | 0.87114 | *ALG10* |
| ENSG00000115165 | 2 | 158261131 | 158355473 | 229 | 41 | 43428 | -1.1325 | 0.87129 | *CYTIP* |
| ENSG00000129667 | 17 | 74456973 | 74507872 | 200 | 40 | 37840 | -1.1325 | 0.87129 | *RHBDF2* |
| ENSG00000077044 | 2 | 234253153 | 234390750 | 311 | 38 | 43978 | -1.1328 | 0.87135 | *DGKD* |
| ENSG00000198785 | 9 | 104321635 | 104510862 | 769 | 55 | 41822 | -1.1332 | 0.87143 | *GRIN3A* |
| ENSG00000197111 | 12 | 53825525 | 53884946 | 80 | 20 | 44523 | -1.1336 | 0.87151 | *PCBP2* |
| ENSG00000174928 | 3 | 155470401 | 155534140 | 188 | 27 | 43743 | -1.1337 | 0.87153 | *C3orf33* |
| ENSG00000128591 | 7 | 128460431 | 128509328 | 131 | 24 | 43118 | -1.134 | 0.87161 | *FLNC* |
| ENSG00000134812 | 11 | 59586741 | 59622974 | 58 | 15 | 40356 | -1.1348 | 0.87176 | *GIF* |
| ENSG00000134443 | 18 | 56877400 | 56908006 | 128 | 20 | 49621 | -1.1349 | 0.87179 | *GRP* |
| ENSG00000070159 | 9 | 112127746 | 112270590 | 424 | 49 | 49967 | -1.1352 | 0.87186 | *PTPN3* |
| ENSG00000148702 | 10 | 115300596 | 115359361 | 261 | 46 | 42105 | -1.1356 | 0.87194 | *HABP2* |
| ENSG00000250479 | 22 | 24098021 | 24120630 | 72 | 17 | 45828 | -1.1357 | 0.87197 | *CHCHD10* |
| ENSG00000144962 | 3 | 172597148 | 172869058 | 817 | 95 | 46590 | -1.136 | 0.87202 | *SPATA16* |
| ENSG00000130766 | 1 | 28576038 | 28619002 | 118 | 17 | 44054 | -1.1362 | 0.87206 | *SESN2* |
| ENSG00000134153 | 15 | 34366218 | 34404149 | 74 | 13 | 32670 | -1.1371 | 0.87224 | *EMC7* |
| ENSG00000119878 | 2 | 46833555 | 46862881 | 88 | 13 | 40266 | -1.1374 | 0.8723 | *CRIPT* |
| ENSG00000134265 | 18 | 10515902 | 10562758 | 131 | 12 | 50144 | -1.1378 | 0.8724 | *NAPG* |
| ENSG00000110060 | 11 | 125753381 | 125783116 | 108 | 17 | 43775 | -1.1381 | 0.87245 | *PUS3* |
| ENSG00000121552 | 3 | 122034091 | 122070819 | 122 | 21 | 37237 | -1.1381 | 0.87247 | *CSTA* |
| ENSG00000156463 | 5 | 145306142 | 145471354 | 440 | 31 | 43884 | -1.1382 | 0.87249 | *SH3RF2* |
| ENSG00000164506 | 6 | 147515561 | 147716866 | 356 | 34 | 54593 | -1.1387 | 0.87258 | *STXBP5* |
| ENSG00000146247 | 6 | 79635584 | 79797953 | 365 | 33 | 53187 | -1.1388 | 0.8726 | *PHIP* |
| ENSG00000126562 | 17 | 40922696 | 40958954 | 17 | 10 | 53936 | -1.1404 | 0.87295 | *WNK4* |
| ENSG00000064115 | 12 | 27116128 | 27177367 | 195 | 40 | 53277 | -1.141 | 0.87306 | *TM7SF3* |
| ENSG00000100124 | 22 | 38216862 | 38255334 | 59 | 14 | 47574 | -1.1411 | 0.87309 | *ANKRD54* |
| ENSG00000157796 | 4 | 39174024 | 39297430 | 260 | 17 | 40194 | -1.1419 | 0.87325 | *WDR19* |
| ENSG00000140153 | 14 | 102595840 | 102701184 | 298 | 37 | 38261 | -1.1419 | 0.87326 | *WDR20* |
| ENSG00000134758 | 18 | 29661818 | 29721524 | 147 | 21 | 48600 | -1.1427 | 0.87342 | *RNF138* |
| ENSG00000139988 | 14 | 68158603 | 68211169 | 169 | 21 | 38503 | -1.1428 | 0.87344 | *RDH12* |

| | | | | | | | | | |
|---|---|---|---|---|---|---|---|---|---|
| ENSG00000115107 | 2 | 119971384 | 120033228 | 265 | 45 | 48024 | -1.1432 | 0.87352 | *STEAP3* |
| ENSG00000112246 | 6 | 100822891 | 100922805 | 282 | 37 | 50545 | -1.1434 | 0.87357 | *SIM1* |
| ENSG00000125375 | 14 | 50769044 | 50812276 | 137 | 9 | 52742 | -1.1435 | 0.87358 | *ATP5S* |
| ENSG00000197816 | 9 | 99990765 | 100150806 | 508 | 29 | 39213 | -1.1437 | 0.87362 | *CCDC180* |
| ENSG00000183644 | 11 | 111375510 | 111417756 | 81 | 17 | 45509 | -1.1442 | 0.87373 | *C11orf88* |
| ENSG00000185591 | 12 | 53763960 | 53820230 | 129 | 13 | 46128 | -1.1443 | 0.87376 | *SP1* |
| ENSG00000179066 | 19 | 35586873 | 35608141 | 63 | 12 | 44632 | -1.1444 | 0.87377 | *AC020907.1* |
| ENSG00000165046 | 8 | 38233725 | 38277045 | 68 | 7 | 53673 | -1.1447 | 0.87383 | *LETM2* |
| ENSG00000186517 | 1 | 161006736 | 161049760 | 103 | 19 | 42612 | -1.1448 | 0.87386 | *ARHGAP30* |
| ENSG00000184985 | 4 | 7184265 | 7754554 | 3269 | 318 | 45037 | -1.1452 | 0.87393 | *SORCS2* |
| ENSG00000170917 | 4 | 123799852 | 123854123 | 228 | 29 | 34445 | -1.1461 | 0.87411 | *NUDT6* |
| ENSG00000136504 | 17 | 47855917 | 47916458 | 73 | 23 | 44127 | -1.1461 | 0.87413 | *KAT7* |
| ENSG00000164849 | 7 | 1074212 | 1108897 | 222 | 24 | 49759 | -1.1468 | 0.87426 | *GPR146* |
| ENSG00000185163 | 12 | 132611139 | 132638880 | 82 | 14 | 37312 | -1.1474 | 0.87439 | *DDX51* |
| ENSG00000169129 | 10 | 116044583 | 116174515 | 369 | 61 | 35478 | -1.1475 | 0.87441 | *AFAP1L2* |
| ENSG00000154237 | 15 | 101449420 | 101620317 | 654 | 78 | 45401 | -1.1475 | 0.87441 | *LRRK1* |
| ENSG00000110436 | 11 | 35262753 | 35451610 | 647 | 68 | 48775 | -1.1475 | 0.87442 | *SLC1A2* |
| ENSG00000137656 | 11 | 116608886 | 116653704 | 164 | 28 | 47136 | -1.1481 | 0.87454 | *BUD13* |
| ENSG00000115255 | 19 | 1481165 | 1507926 | 75 | 14 | 42729 | -1.1483 | 0.87459 | *REEP6* |
| ENSG00000177685 | 11 | 816144 | 841991 | 122 | 16 | 31031 | -1.1508 | 0.87509 | *EFCAB4A* |
| ENSG00000176293 | 19 | 58560607 | 58607677 | 137 | 21 | 52099 | -1.151 | 0.87514 | *ZNF135* |
| ENSG00000218823 | 7 | 4887364 | 4911625 | 97 | 14 | 40376 | -1.1512 | 0.87518 | *PAPOLB* |
| ENSG00000131152 | 16 | 87326482 | 87377879 | 245 | 37 | 38976 | -1.152 | 0.87534 | *RP11-178L8.4* |
| ENSG00000188171 | 19 | 20792867 | 20854402 | 421 | 30 | 42990 | -1.1521 | 0.87535 | *ZNF626* |
| ENSG00000080166 | 13 | 95079558 | 95141936 | 209 | 28 | 49047 | -1.1525 | 0.87544 | *DCT* |
| ENSG00000173239 | 10 | 90552487 | 90590303 | 111 | 25 | 44011 | -1.1525 | 0.87545 | *LIPM* |
| ENSG00000198498 | 4 | 164405594 | 164451691 | 128 | 15 | 44115 | -1.1532 | 0.87559 | *TMA16* |
| ENSG00000142748 | 1 | 27685603 | 27711315 | 42 | 17 | 35074 | -1.1533 | 0.87561 | *FCN3* |
| ENSG00000111325 | 12 | 123449127 | 123474590 | 27 | 9 | 42685 | -1.1533 | 0.87561 | *OGFOD2* |
| ENSG00000111262 | 12 | 5009071 | 5050527 | 110 | 21 | 50444 | -1.1534 | 0.87562 | *KCNA1* |
| ENSG00000196932 | 10 | 63156401 | 63223208 | 189 | 27 | 41210 | -1.1535 | 0.87564 | *TMEM26* |
| ENSG00000088930 | 20 | 21273942 | 21380463 | 247 | 24 | 51510 | -1.1536 | 0.87567 | *XRN2* |
| ENSG00000070882 | 7 | 24826158 | 25031253 | 692 | 124 | 45164 | -1.1542 | 0.8758 | *OSBPL3* |
| ENSG00000157017 | 3 | 10317359 | 10344631 | 85 | 18 | 38113 | -1.155 | 0.87596 | *GHRL* |
| ENSG00000140968 | 16 | 85922409 | 85966215 | 173 | 35 | 44761 | -1.155 | 0.87596 | *IRF8* |
| ENSG00000182326 | 12 | 7086351 | 7188336 | 180 | 21 | 35736 | -1.156 | 0.87617 | *C1S* |

| | | | | | | | | | |
|---|---|---|---|---|---|---|---|---|---|
| ENSG00000110713 | 11 | 3682313 | 3829022 | 513 | 48 | 48317 | -1.1562 | 0.87621 | *NUP98* |
| ENSG00000131381 | 3 | 15101580 | 15150670 | 128 | 12 | 52131 | -1.1566 | 0.87628 | *ZFYVE20* |
| ENSG00000242574 | 6 | 32892406 | 32918847 | 168 | 26 | 52218 | -1.1567 | 0.87631 | *HLA-DMB* |
| ENSG00000126934 | 19 | 4080319 | 4134126 | 134 | 30 | 44408 | -1.1575 | 0.87646 | *MAP2K2* |
| ENSG00000183486 | 21 | 42723870 | 42791317 | 206 | 49 | 31358 | -1.1581 | 0.87659 | *MX2* |
| ENSG00000106012 | 7 | 2588632 | 2664368 | 390 | 52 | 51096 | -1.1582 | 0.87661 | *IQCE* |
| ENSG00000188277 | 15 | 41052159 | 41074643 | 42 | 10 | 40391 | -1.1582 | 0.87661 | *C15orf62* |
| ENSG00000108852 | 17 | 41942725 | 41997068 | 190 | 23 | 40627 | -1.1583 | 0.87662 | *MPP2* |
| ENSG00000197362 | 7 | 148756735 | 148797874 | 120 | 16 | 37816 | -1.1584 | 0.87665 | *ZNF786* |
| ENSG00000089356 | 19 | 35596732 | 35625228 | 113 | 15 | 51690 | -1.1587 | 0.87672 | *FXYD3* |
| ENSG00000198690 | 15 | 31186055 | 31245311 | 184 | 25 | 45349 | -1.1593 | 0.87684 | *FAN1* |
| ENSG00000183208 | 15 | 90767040 | 90795315 | 116 | 13 | 38147 | -1.1598 | 0.87693 | *GDPGP1* |
| ENSG00000143156 | 1 | 169091769 | 169347205 | 917 | 44 | 44310 | -1.1599 | 0.87695 | *NME7* |
| ENSG00000123338 | 12 | 54881495 | 54947726 | 198 | 45 | 43539 | -1.1602 | 0.87702 | *NCKAP1L* |
| ENSG00000150656 | 18 | 72191675 | 72264448 | 240 | 56 | 43204 | -1.1608 | 0.87714 | *CNDP1* |
| ENSG00000136002 | 2 | 131584489 | 131814836 | 667 | 87 | 35063 | -1.1629 | 0.87757 | *ARHGEF4* |
| ENSG00000169469 | 1 | 152993678 | 153015376 | 132 | 5 | 49900 | -1.1646 | 0.87791 | *SPRR1B* |
| ENSG00000165309 | 10 | 23206953 | 23336518 | 356 | 45 | 43093 | -1.1648 | 0.87795 | *ARMC3* |
| ENSG00000248874 | 5 | 23941457 | 24188372 | 1077 | 46 | 48017 | -1.1649 | 0.87797 | *C5orf17* |
| ENSG00000160199 | 21 | 44384620 | 44463691 | 438 | 34 | 46757 | -1.1657 | 0.87813 | *PKNOX1* |
| ENSG00000138115 | 10 | 96786530 | 96839254 | 226 | 23 | 50594 | -1.1658 | 0.87814 | *CYP2C8* |
| ENSG00000222028 | 14 | 23501376 | 23523269 | 49 | 14 | 39630 | -1.1669 | 0.87837 | *PSMB11* |
| ENSG00000145907 | 5 | 151140606 | 151202346 | 171 | 20 | 49353 | -1.1685 | 0.8787 | *G3BP1* |
| ENSG00000183808 | 8 | 94731584 | 94763245 | 65 | 9 | 55327 | -1.1692 | 0.87885 | *RBM12B* |
| ENSG00000196604 | 2 | 130821108 | 130896795 | 103 | 14 | 38881 | -1.1706 | 0.87912 | *POTEF* |
| ENSG00000149599 | 20 | 30425440 | 30468550 | 106 | 26 | 42330 | -1.1706 | 0.87913 | *DUSP15* |
| ENSG00000168876 | 11 | 94216795 | 94242749 | 66 | 15 | 45621 | -1.1709 | 0.87918 | *ANKRD49* |
| ENSG00000229571 | 1 | 13206356 | 13229581 | 1 | 1 | 42665 | -1.171 | 0.8792 | *PRAMEF26* |
| ENSG00000100884 | 14 | 24530046 | 24557309 | 42 | 19 | 48520 | -1.1716 | 0.87931 | *CPNE6* |
| ENSG00000262543 | 6 | 138689042 | 138714212 | 119 | 7 | 24978 | -1.1717 | 0.87933 | *RP3-422G23.4* |
| ENSG00000167291 | 17 | 77896142 | 78019647 | 459 | 112 | 49781 | -1.1719 | 0.87939 | *TBC1D16* |
| ENSG00000127329 | 12 | 70900630 | 71041220 | 575 | 94 | 45506 | -1.172 | 0.8794 | *PTPRB* |
| ENSG00000188906 | 12 | 40580546 | 40773087 | 753 | 66 | 45306 | -1.1727 | 0.87954 | *LRRK2* |
| ENSG00000185453 | 19 | 48663949 | 48710877 | 208 | 30 | 41356 | -1.1728 | 0.87955 | *C19orf68* |
| ENSG00000222022 | 2 | 238320063 | 238343919 | 67 | 13 | 44932 | -1.173 | 0.8796 | *AC112721.1* |
| ENSG00000125817 | 20 | 3754498 | 3777337 | 67 | 15 | 47941 | -1.1745 | 0.8799 | *CENPB* |

| | | | | | | | | | |
|---|---|---|---|---|---|---|---|---|---|
| ENSG00000158769 | 1 | 160955001 | 161001138 | 151 | 15 | 48948 | -1.1747 | 0.87994 | *F11R* |
| ENSG00000080345 | 2 | 152256397 | 152374527 | 311 | 33 | 38472 | -1.1747 | 0.87995 | *RIF1* |
| ENSG00000136275 | 7 | 47824889 | 47869445 | 165 | 24 | 56115 | -1.1754 | 0.88008 | *C7orf69* |
| ENSG00000123612 | 2 | 158373279 | 158495517 | 185 | 36 | 42264 | -1.1758 | 0.88017 | *ACVR1C* |
| ENSG00000137710 | 11 | 110035605 | 110177447 | 309 | 25 | 50736 | -1.1759 | 0.88019 | *RDX* |
| ENSG00000164008 | 1 | 43222940 | 43273968 | 107 | 18 | 53508 | -1.176 | 0.8802 | *C1orf50* |
| ENSG00000196372 | 10 | 5670830 | 5718558 | 291 | 49 | 48074 | -1.1768 | 0.88035 | *ASB13* |
| ENSG00000204033 | 10 | 85970254 | 85995345 | 89 | 12 | 40507 | -1.1774 | 0.88049 | *LRIT2* |
| ENSG00000184635 | 19 | 20001722 | 20056384 | 181 | 24 | 36038 | -1.178 | 0.88061 | *ZNF93* |
| ENSG00000126581 | 17 | 40952152 | 40995367 | 16 | 10 | 47959 | -1.179 | 0.8808 | *BECN1* |
| ENSG00000160191 | 21 | 44063746 | 44205619 | 657 | 91 | 44219 | -1.179 | 0.8808 | *PDE9A* |
| ENSG00000267545 | 19 | 45673080 | 45715702 | 185 | 41 | 42172 | -1.1793 | 0.88087 | *AC005779.2* |
| ENSG00000178257 | 16 | 11357144 | 11377452 | 86 | 16 | 41754 | -1.1801 | 0.88103 | *PRM3* |
| ENSG00000133704 | 12 | 30771922 | 30858920 | 276 | 18 | 51054 | -1.1807 | 0.88113 | *IPO8* |
| ENSG00000106631 | 7 | 44168463 | 44190931 | 76 | 15 | 46508 | -1.1809 | 0.88117 | *MYL7* |
| ENSG00000139726 | 12 | 123227321 | 123265611 | 80 | 11 | 24490 | -1.1816 | 0.88132 | *DENR* |
| ENSG00000053770 | 14 | 57725627 | 57766797 | 37 | 13 | 39391 | -1.1817 | 0.88134 | *AP5M1* |
| ENSG00000127951 | 7 | 76812688 | 76839143 | 62 | 17 | 45318 | -1.1819 | 0.88138 | *FGL2* |
| ENSG00000117899 | 15 | 81229667 | 81292219 | 163 | 16 | 47889 | -1.1824 | 0.88148 | *MESDC2* |
| ENSG00000123091 | 1 | 51691943 | 51749127 | 77 | 14 | 49659 | -1.1826 | 0.88151 | *RNF11* |
| ENSG00000105486 | 19 | 48608702 | 48683860 | 336 | 27 | 36430 | -1.1826 | 0.88151 | *LIG1* |
| ENSG00000156304 | 21 | 33033346 | 33114388 | 171 | 26 | 47696 | -1.1837 | 0.88172 | *SCAF4* |
| ENSG00000111859 | 6 | 11173531 | 11392581 | 779 | 138 | 48666 | -1.1844 | 0.88186 | *NEDD9* |
| ENSG00000175143 | 1 | 248559296 | 248580405 | 115 | 7 | 21251 | -1.1844 | 0.88188 | *OR2T1* |
| ENSG00000188375 | 12 | 31934123 | 31955175 | 107 | 22 | 46122 | -1.1849 | 0.88197 | *H3F3C* |
| ENSG00000172375 | 11 | 118962908 | 118999252 | 87 | 11 | 27829 | -1.1853 | 0.88205 | *C2CD2L* |
| ENSG00000196116 | 9 | 100164232 | 100268407 | 240 | 26 | 46177 | -1.1864 | 0.88227 | *TDRD7* |
| ENSG00000137700 | 11 | 118884824 | 118911616 | 92 | 16 | 47377 | -1.1865 | 0.88229 | *SLC37A4* |
| ENSG00000174371 | 1 | 242001269 | 242068450 | 316 | 34 | 48181 | -1.1866 | 0.8823 | *EXO1* |
| ENSG00000203786 | 1 | 152720506 | 152744529 | 64 | 8 | 43265 | -1.1871 | 0.88241 | *KPRP* |
| ENSG00000139637 | 12 | 53683470 | 53710961 | 43 | 10 | 36695 | -1.1873 | 0.88244 | *C12orf10* |
| ENSG00000069712 | 1 | 92622542 | 92660280 | 71 | 11 | 43939 | -1.1885 | 0.88268 | *KIAA1107* |
| ENSG00000106484 | 7 | 130116012 | 130156133 | 103 | 15 | 36218 | -1.1889 | 0.88275 | *MEST* |
| ENSG00000179698 | 8 | 145152629 | 145183218 | 73 | 8 | 37279 | -1.1889 | 0.88276 | *KIAA1875* |
| ENSG00000179820 | 19 | 54359477 | 54389691 | 49 | 14 | 38036 | -1.1889 | 0.88276 | *MYADM* |
| ENSG00000167508 | 16 | 88708343 | 88739569 | 132 | 29 | 46730 | -1.1893 | 0.88283 | *MVD* |

| | | | | | | | | | |
|---|---|---|---|---|---|---|---|---|---|
| ENSG00000158109 | 1 | 3531566 | 3556691 | 101 | 20 | 42559 | -1.1896 | 0.8829 | *TPRG1L* |
| ENSG00000183741 | 22 | 39247455 | 39278319 | 73 | 20 | 38351 | -1.1902 | 0.88302 | *CBX6* |
| ENSG00000116661 | 1 | 11698424 | 11725842 | 77 | 25 | 45320 | -1.1907 | 0.88312 | *FBXO2* |
| ENSG00000100345 | 22 | 36667327 | 36794063 | 379 | 56 | 38202 | -1.1907 | 0.88312 | *MYH9* |
| ENSG00000149131 | 11 | 57354860 | 57392326 | 92 | 14 | 48726 | -1.1924 | 0.88344 | *SERPING1* |
| ENSG00000182022 | 10 | 125757184 | 125863206 | 281 | 51 | 46303 | -1.1926 | 0.88348 | *CHST15* |
| ENSG00000268955 | 8 | 8036158 | 8056331 | 1 | 1 | 43280 | -1.1927 | 0.8835 | *LRLE1* |
| ENSG00000141316 | 17 | 31287394 | 31334895 | 214 | 38 | 41732 | -1.193 | 0.88357 | *SPACA3* |
| ENSG00000136826 | 9 | 110237133 | 110262763 | 81 | 14 | 45950 | -1.1935 | 0.88366 | *KLF4* |
| ENSG00000159784 | 7 | 143040493 | 143069863 | 86 | 26 | 53739 | -1.1939 | 0.88373 | *FAM131B* |
| ENSG00000164825 | 8 | 6718097 | 6745544 | 276 | 29 | 48244 | -1.1939 | 0.88374 | *DEFB1* |
| ENSG00000143341 | 1 | 185693683 | 186170085 | 770 | 77 | 38251 | -1.194 | 0.88376 | *HMCN1* |
| ENSG00000183844 | 21 | 42666139 | 42739358 | 245 | 46 | 42234 | -1.194 | 0.88376 | *FAM3B* |
| ENSG00000124172 | 20 | 57590522 | 57617437 | 51 | 14 | 54872 | -1.1945 | 0.88386 | *ATP5E* |
| ENSG00000104972 | 19 | 55075346 | 55158979 | 523 | 45 | 43606 | -1.1947 | 0.88389 | *LILRB1* |
| ENSG00000163312 | 4 | 84318496 | 84387009 | 155 | 23 | 37222 | -1.1953 | 0.88402 | *HELQ* |
| ENSG00000078668 | 8 | 42239142 | 42273415 | 72 | 12 | 27999 | -1.1956 | 0.88408 | *VDAC3* |
| ENSG00000187867 | 19 | 14154177 | 14179971 | 79 | 20 | 42006 | -1.1965 | 0.88424 | *PALM3* |
| ENSG00000198131 | 19 | 58729960 | 58798815 | 275 | 29 | 43545 | -1.1966 | 0.88427 | *ZNF544* |
| ENSG00000143149 | 1 | 165621453 | 165678100 | 226 | 28 | 50917 | -1.1967 | 0.88428 | *ALDH9A1* |
| ENSG00000003989 | 8 | 17344597 | 17438082 | 539 | 119 | 46303 | -1.1968 | 0.88432 | *SLC7A2* |
| ENSG00000162728 | 1 | 160041360 | 160070353 | 78 | 15 | 47354 | -1.197 | 0.88434 | *KCNJ9* |
| ENSG00000205045 | 17 | 33781268 | 33874880 | 317 | 67 | 44353 | -1.197 | 0.88435 | *SLFN12L* |
| ENSG00000142039 | 19 | 41806094 | 41840788 | 71 | 9 | 55223 | -1.1972 | 0.88439 | *CCDC97* |
| ENSG00000198911 | 22 | 42219109 | 42313312 | 257 | 37 | 42643 | -1.1976 | 0.88446 | *SREBF2* |
| ENSG00000176769 | 10 | 132880654 | 133119984 | 1292 | 183 | 47267 | -1.1979 | 0.88452 | *TCERG1L* |
| ENSG00000009307 | 1 | 115249534 | 115311297 | 125 | 17 | 51593 | -1.198 | 0.88455 | *CSDE1* |
| ENSG00000187860 | 22 | 30742624 | 30784647 | 147 | 14 | 50696 | -1.1993 | 0.88479 | *CCDC157* |
| ENSG00000139354 | 12 | 100957461 | 101032064 | 170 | 23 | 39033 | -1.1994 | 0.88481 | *GAS2L3* |
| ENSG00000212658 | 17 | 39448078 | 39469103 | 86 | 8 | 50720 | -1.1996 | 0.88486 | *KRTAP29-1* |
| ENSG00000093009 | 22 | 19456982 | 19518135 | 191 | 32 | 43076 | -1.2004 | 0.88501 | *CDC45* |
| ENSG00000107165 | 9 | 12675439 | 12720290 | 183 | 21 | 42241 | -1.2008 | 0.88508 | *TYRP1* |
| ENSG00000099624 | 19 | 1231749 | 1254824 | 53 | 19 | 49492 | -1.2008 | 0.88508 | *ATP5D* |
| ENSG00000204849 | 9 | 39345699 | 39371956 | 1 | 1 | 42880 | -1.2014 | 0.8852 | *SPATA31A1* |
| ENSG00000196139 | 10 | 5067546 | 5159878 | 587 | 64 | 44976 | -1.2017 | 0.88526 | *AKR1C3* |
| ENSG00000126391 | 11 | 65144070 | 65190996 | 58 | 19 | 37447 | -1.2026 | 0.88544 | *FRMD8* |

| | | | | | | | | | |
|---|---|---|---|---|---|---|---|---|---|
| ENSG00000268217 | 5 | 72884203 | 72906845 | 66 | 11 | 46156 | -1.2029 | 0.88549 | *AC008387.1* |
| ENSG00000139572 | 12 | 54746229 | 54768271 | 39 | 8 | 49196 | -1.2038 | 0.88567 | *GPR84* |
| ENSG00000184933 | 11 | 6805756 | 6827128 | 74 | 11 | 54737 | -1.2039 | 0.88569 | *OR6A2* |
| ENSG00000108946 | 17 | 66497921 | 66557460 | 167 | 24 | 55675 | -1.205 | 0.8859 | *PRKAR1A* |
| ENSG00000166002 | 11 | 93201638 | 93286674 | 292 | 40 | 53062 | -1.2061 | 0.8861 | *SMCO4* |
| ENSG00000183638 | 8 | 10453859 | 10579697 | 415 | 76 | 42166 | -1.2061 | 0.88611 | *RP1L1* |
| ENSG00000184881 | 11 | 5334541 | 5355582 | 142 | 16 | 39317 | -1.2068 | 0.88624 | *OR51B2* |
| ENSG00000133313 | 18 | 72153051 | 72198366 | 157 | 26 | 47526 | -1.207 | 0.88628 | *CNDP2* |
| ENSG00000110786 | 11 | 18739475 | 18824268 | 264 | 59 | 45077 | -1.2072 | 0.88632 | *PTPN5* |
| ENSG00000100711 | 14 | 104172067 | 104210005 | 77 | 21 | 47329 | -1.2072 | 0.88632 | *ZFYVE21* |
| ENSG00000165269 | 9 | 33374765 | 33412643 | 91 | 22 | 51699 | -1.2075 | 0.88637 | *AQP7* |
| ENSG00000095585 | 10 | 97941458 | 98041344 | 270 | 67 | 40292 | -1.2081 | 0.88649 | *BLNK* |
| ENSG00000187510 | 12 | 93105281 | 93176231 | 171 | 21 | 36284 | -1.2091 | 0.88669 | *PLEKHG7* |
| ENSG00000100445 | 14 | 24898972 | 24922111 | 169 | 10 | 23765 | -1.2099 | 0.88685 | *SDR39U1* |
| ENSG00000175221 | 19 | 857962 | 903218 | 187 | 33 | 42693 | -1.2099 | 0.88685 | *MED16* |
| ENSG00000160188 | 21 | 43882596 | 43926464 | 128 | 33 | 45622 | -1.2101 | 0.88688 | *RSPH1* |
| ENSG00000196118 | 16 | 30758744 | 30784031 | 25 | 12 | 46215 | -1.2105 | 0.88696 | *C16orf93* |
| ENSG00000258405 | 19 | 52946829 | 53025407 | 325 | 33 | 47446 | -1.2119 | 0.88722 | *ZNF578* |
| ENSG00000116544 | 1 | 35321037 | 35405186 | 130 | 36 | 39656 | -1.2122 | 0.88728 | *DLGAP3* |
| ENSG00000240563 | 1 | 62650503 | 62688000 | 315 | 41 | 44493 | -1.2123 | 0.8873 | *L1TD1* |
| ENSG00000165006 | 9 | 34169003 | 34262521 | 218 | 20 | 36022 | -1.2128 | 0.88739 | *UBAP1* |
| ENSG00000136802 | 9 | 131634391 | 131690318 | 127 | 19 | 47599 | -1.2131 | 0.88745 | *LRRC8A* |
| ENSG00000189403 | 13 | 31022884 | 31201734 | 362 | 75 | 39148 | -1.2131 | 0.88746 | *HMGB1* |
| ENSG00000175536 | 11 | 74192757 | 74214778 | 84 | 11 | 50644 | -1.214 | 0.88763 | *LIPT2* |
| ENSG00000186081 | 12 | 52898359 | 52924471 | 127 | 14 | 47431 | -1.2151 | 0.88783 | *KRT5* |
| ENSG00000056736 | 3 | 53870607 | 53909827 | 150 | 29 | 35290 | -1.2163 | 0.88807 | *IL17RB* |
| ENSG00000267561 | 1 | 87448692 | 87644884 | 475 | 47 | 44638 | -1.2164 | 0.88809 | *RP5-1052I5.2* |
| ENSG00000096696 | 6 | 7531808 | 7596950 | 230 | 40 | 48425 | -1.2171 | 0.88821 | *DSP* |
| ENSG00000149243 | 11 | 75123438 | 75151674 | 42 | 11 | 44416 | -1.2173 | 0.88826 | *KLHL35* |
| ENSG00000197043 | 5 | 150470273 | 150547443 | 312 | 59 | 44252 | -1.2187 | 0.88853 | *ANXA6* |
| ENSG00000269545 | 19 | 58730831 | 58808173 | 291 | 30 | 44037 | -1.2194 | 0.88865 | *CTD-3138B18.4* |
| ENSG00000139651 | 12 | 53564484 | 53594596 | 56 | 9 | 47763 | -1.2198 | 0.88872 | *ZNF740* |
| ENSG00000196455 | 3 | 130387779 | 130475673 | 174 | 9 | 48148 | -1.2199 | 0.88874 | *PIK3R4* |
| ENSG00000212901 | 17 | 39154773 | 39185576 | 164 | 17 | 45368 | -1.2199 | 0.88876 | *KRTAP3-1* |
| ENSG00000070601 | 9 | 37640997 | 37756901 | 469 | 72 | 46985 | -1.2204 | 0.88885 | *FRMPD1* |
| ENSG00000105048 | 19 | 55634162 | 55670722 | 93 | 30 | 36683 | -1.2214 | 0.88903 | *TNNT1* |

| | | | | | | | | | |
|---|---|---|---|---|---|---|---|---|---|
| ENSG00000163530 | 3 | 109002635 | 109045364 | 135 | 20 | 31758 | -1.2229 | 0.88932 | *DPPA2* |
| ENSG00000197775 | 14 | 24397940 | 24419196 | 116 | 6 | 23708 | -1.2232 | 0.88938 | *DHRS4-AS1* |
| ENSG00000139880 | 14 | 23506271 | 23536747 | 84 | 19 | 44638 | -1.2246 | 0.88963 | *CDH24* |
| ENSG00000174482 | 9 | 27938076 | 28680283 | 2916 | 318 | 44755 | -1.2248 | 0.88968 | *LINGO2* |
| ENSG00000157020 | 3 | 10324815 | 10372862 | 152 | 24 | 39038 | -1.225 | 0.8897 | *SEC13* |
| ENSG00000174951 | 19 | 49241268 | 49268647 | 80 | 14 | 29516 | -1.225 | 0.8897 | *FUT1* |
| ENSG00000148334 | 9 | 130872972 | 130900741 | 40 | 10 | 30599 | -1.2255 | 0.88981 | *PTGES2* |
| ENSG00000177201 | 1 | 248447918 | 248468880 | 106 | 7 | 49262 | -1.2259 | 0.88989 | *OR2T12* |
| ENSG00000184825 | 6 | 27104861 | 27125317 | 38 | 14 | 38886 | -1.226 | 0.88989 | *HIST1H2AH* |
| ENSG00000165516 | 14 | 50224326 | 50259909 | 46 | 13 | 48332 | -1.227 | 0.89009 | *KLHDC2* |
| ENSG00000156873 | 16 | 30749591 | 30782490 | 26 | 13 | 41528 | -1.2272 | 0.89013 | *PHKG2* |
| ENSG00000196091 | 12 | 101952131 | 102089796 | 589 | 84 | 44593 | -1.2273 | 0.89014 | *MYBPC1* |
| ENSG00000196990 | 9 | 136434079 | 136461319 | 72 | 17 | 41241 | -1.2278 | 0.89024 | *FAM163B* |
| ENSG00000213462 | 7 | 64441187 | 64477121 | 151 | 19 | 46743 | -1.2279 | 0.89027 | *ERV3-1* |
| ENSG00000138768 | 4 | 76639777 | 76745382 | 280 | 19 | 36553 | -1.228 | 0.89028 | *USO1* |
| ENSG00000158764 | 1 | 160904815 | 160934589 | 109 | 22 | 48561 | -1.2284 | 0.89034 | *ITLN2* |
| ENSG00000144182 | 2 | 99761418 | 99789620 | 88 | 11 | 42248 | -1.2284 | 0.89036 | *LIPT1* |
| ENSG00000212657 | 17 | 39453952 | 39475505 | 92 | 9 | 50869 | -1.2291 | 0.89049 | *KRTAP16-1* |
| ENSG00000101365 | 20 | 2629041 | 2654865 | 113 | 19 | 49824 | -1.2297 | 0.8906 | *IDH3B* |
| ENSG00000153936 | 1 | 87370331 | 87612334 | 584 | 35 | 40894 | -1.2298 | 0.89061 | *HS2ST1* |
| ENSG00000139641 | 12 | 56502034 | 56548455 | 72 | 26 | 37537 | -1.2302 | 0.89069 | *ESYT1* |
| ENSG00000168275 | 1 | 234499202 | 234529795 | 94 | 13 | 48040 | -1.2314 | 0.8909 | *COA6* |
| ENSG00000163281 | 4 | 44674217 | 44738612 | 170 | 13 | 33871 | -1.2315 | 0.89093 | *GNPDA2* |
| ENSG00000154358 | 1 | 228385831 | 228576577 | 559 | 45 | 52315 | -1.2321 | 0.89105 | *OBSCN* |
| ENSG00000107643 | 10 | 49504698 | 49657403 | 379 | 36 | 50413 | -1.2321 | 0.89105 | *MAPK8* |
| ENSG00000103043 | 16 | 70711342 | 70845064 | 355 | 53 | 41354 | -1.2326 | 0.89113 | *VAC14* |
| ENSG00000118197 | 1 | 200583024 | 200649097 | 279 | 29 | 42088 | -1.2329 | 0.8912 | *DDX59* |
| ENSG00000162600 | 1 | 58871056 | 59022474 | 698 | 62 | 45350 | -1.2332 | 0.89124 | *OMA1* |
| ENSG00000105171 | 19 | 30084924 | 30118144 | 156 | 26 | 40111 | -1.2332 | 0.89124 | *POP4* |
| ENSG00000138495 | 3 | 119363360 | 119406301 | 186 | 18 | 54284 | -1.2336 | 0.89132 | *COX17* |
| ENSG00000198092 | 4 | 68908916 | 69005598 | 322 | 17 | 37759 | -1.2337 | 0.89134 | *TMPRSS11F* |
| ENSG00000126458 | 19 | 50128549 | 50153458 | 62 | 11 | 37115 | -1.2342 | 0.89144 | *RRAS* |
| ENSG00000188373 | 10 | 85923494 | 85955050 | 84 | 18 | 53234 | -1.2345 | 0.89149 | *C10orf99* |
| ENSG00000148602 | 10 | 85981349 | 86011217 | 104 | 18 | 41366 | -1.2357 | 0.89172 | *LRIT1* |
| ENSG00000117069 | 1 | 77323126 | 77541396 | 632 | 81 | 41711 | -1.2363 | 0.89182 | *ST6GALNAC5* |
| ENSG00000174109 | 16 | 1459745 | 1489345 | 107 | 21 | 46127 | -1.2363 | 0.89182 | *C16orf91* |

| | | | | | | | | | |
|---|---|---|---|---|---|---|---|---|---|
| ENSG00000115756 | 2 | 10433015 | 10577743 | 622 | 109 | 46442 | -1.2369 | 0.89193 | *HPCAL1* |
| ENSG00000058335 | 15 | 79242289 | 79393115 | 508 | 94 | 46464 | -1.2372 | 0.89199 | *RASGRF1* |
| ENSG00000100373 | 22 | 45670863 | 45701755 | 131 | 18 | 31075 | -1.2373 | 0.89202 | *UPK3A* |
| ENSG00000104974 | 19 | 55095047 | 55123555 | 208 | 27 | 45052 | -1.2374 | 0.89203 | *LILRA1* |
| ENSG00000131437 | 5 | 132018320 | 132083330 | 86 | 8 | 28619 | -1.2389 | 0.89231 | *KIF3A* |
| ENSG00000150630 | 4 | 177594689 | 177723881 | 308 | 26 | 30910 | -1.2391 | 0.89235 | *VEGFC* |
| ENSG00000105176 | 19 | 30404551 | 30516611 | 245 | 28 | 37238 | -1.2392 | 0.89237 | *URI1* |
| ENSG00000074319 | 11 | 18479883 | 18558779 | 246 | 26 | 51748 | -1.2396 | 0.89243 | *TSG101* |
| ENSG00000115421 | 2 | 60973365 | 61039220 | 190 | 21 | 54471 | -1.2401 | 0.89253 | *PAPOLG* |
| ENSG00000135437 | 12 | 56104151 | 56128489 | 35 | 14 | 37723 | -1.2411 | 0.89272 | *RDH5* |
| ENSG00000170703 | 17 | 46829597 | 46904576 | 243 | 31 | 43677 | -1.2419 | 0.89287 | *TTLL6* |
| ENSG00000113196 | 5 | 153844532 | 153867824 | 70 | 20 | 56554 | -1.2423 | 0.89294 | *HAND1* |
| ENSG00000185338 | 16 | 11338262 | 11360036 | 73 | 14 | 42670 | -1.2423 | 0.89294 | *SOCS1* |
| ENSG00000136819 | 9 | 132579569 | 132608142 | 169 | 12 | 50479 | -1.2426 | 0.89299 | *C9orf78* |
| ENSG00000178951 | 19 | 4034362 | 4076943 | 121 | 27 | 40844 | -1.2428 | 0.89304 | *ZBTB7A* |
| ENSG00000104722 | 8 | 24760525 | 24786607 | 54 | 15 | 44725 | -1.2433 | 0.89313 | *NEFM* |
| ENSG00000123975 | 9 | 91916113 | 91941618 | 71 | 20 | 52887 | -1.2434 | 0.89315 | *CKS2* |
| ENSG00000167987 | 11 | 60887728 | 60939089 | 168 | 13 | 26939 | -1.2436 | 0.89317 | *VPS37C* |
| ENSG00000111057 | 12 | 53332655 | 53356685 | 55 | 15 | 46510 | -1.2438 | 0.89321 | *KRT18* |
| ENSG00000167880 | 17 | 73990583 | 74033533 | 98 | 28 | 44282 | -1.2441 | 0.89326 | *EVPL* |
| ENSG00000268950 | 1 | 31892192 | 31912614 | 80 | 11 | 25521 | -1.2441 | 0.89327 | *AC114494.1* |
| ENSG00000138095 | 2 | 44103647 | 44233144 | 439 | 34 | 32061 | -1.2442 | 0.89329 | *LRPPRC* |
| ENSG00000213694 | 9 | 91596362 | 91629925 | 102 | 27 | 46922 | -1.2442 | 0.8933 | *S1PR3* |
| ENSG00000179270 | 2 | 29273842 | 29307127 | 126 | 25 | 44283 | -1.2446 | 0.89335 | *C2orf71* |
| ENSG00000130726 | 19 | 59045458 | 59072089 | 47 | 14 | 40455 | -1.2461 | 0.89364 | *TRIM28* |
| ENSG00000111058 | 12 | 81321594 | 81660533 | 1255 | 73 | 37692 | -1.2462 | 0.89365 | *ACSS3* |
| ENSG00000109929 | 11 | 121153162 | 121189403 | 88 | 15 | 48157 | -1.2465 | 0.8937 | *SC5D* |
| ENSG00000174567 | 1 | 204157288 | 204193220 | 111 | 26 | 50233 | -1.2474 | 0.89388 | *GOLT1A* |
| ENSG00000013293 | 3 | 170167372 | 170313863 | 367 | 57 | 45923 | -1.2476 | 0.89391 | *SLC7A14* |
| ENSG00000213762 | 19 | 58115601 | 58144724 | 96 | 13 | 47317 | -1.2476 | 0.89391 | *ZNF134* |
| ENSG00000161048 | 7 | 102730223 | 102800007 | 171 | 18 | 48051 | -1.2478 | 0.89394 | *NAPEPLD* |
| ENSG00000128242 | 22 | 30940622 | 30980574 | 115 | 34 | 50255 | -1.2485 | 0.89408 | *GAL3ST1* |
| ENSG00000179909 | 19 | 58198735 | 58230579 | 93 | 20 | 49897 | -1.2498 | 0.89432 | *ZNF154* |
| ENSG00000154743 | 3 | 12515931 | 12591122 | 325 | 26 | 36046 | -1.2499 | 0.89433 | *TSEN2* |
| ENSG00000152359 | 5 | 74959949 | 75023313 | 278 | 20 | 46424 | -1.25 | 0.89434 | *POC5* |
| ENSG00000215203 | 4 | 42885284 | 43042675 | 553 | 67 | 45591 | -1.2501 | 0.89437 | *GRXCR1* |

| | | | | | | | | | |
|---|---|---|---|---|---|---|---|---|---|
| ENSG00000170477 | 12 | 53190333 | 53218335 | 127 | 28 | 41635 | -1.2511 | 0.89454 | *KRT4* |
| ENSG00000110422 | 11 | 33268218 | 33388569 | 298 | 31 | 48759 | -1.2511 | 0.89456 | *HIPK3* |
| ENSG00000102580 | 13 | 96319393 | 96457243 | 198 | 34 | 44304 | -1.2512 | 0.89457 | *DNAJC3* |
| ENSG00000139574 | 12 | 53890474 | 53911422 | 17 | 9 | 53953 | -1.2521 | 0.89474 | *NPFF* |
| ENSG00000181045 | 17 | 78183498 | 78237299 | 225 | 37 | 44949 | -1.2521 | 0.89474 | *SLC26A11* |
| ENSG00000248458 | 1 | 67121547 | 67152710 | 118 | 17 | 38928 | -1.2529 | 0.89488 | *AL139147.1* |
| ENSG00000100344 | 22 | 44309619 | 44370368 | 224 | 24 | 51432 | -1.2534 | 0.89498 | *PNPLA3* |
| ENSG00000258474 | 14 | 32404059 | 32429588 | 130 | 24 | 50924 | -1.2538 | 0.89503 | *RP11-187E13.1* |
| ENSG00000120992 | 8 | 54948938 | 55024577 | 137 | 12 | 31466 | -1.2538 | 0.89504 | *LYPLA1* |
| ENSG00000214415 | 7 | 80077987 | 80151336 | 188 | 30 | 39723 | -1.2545 | 0.89517 | *GNAT3* |
| ENSG00000104866 | 19 | 45584654 | 45661335 | 293 | 30 | 52497 | -1.2546 | 0.89518 | *PPP1R37* |
| ENSG00000075073 | 10 | 71153659 | 71186623 | 117 | 17 | 50501 | -1.2555 | 0.89536 | *TACR2* |
| ENSG00000197935 | 6 | 28952562 | 28983093 | 103 | 20 | 41073 | -1.2558 | 0.89541 | *ZNF311* |
| ENSG00000109272 | 4 | 74708906 | 74729872 | 75 | 16 | 42476 | -1.2559 | 0.89542 | *PF4V1* |
| ENSG00000114354 | 3 | 100418205 | 100477810 | 106 | 11 | 45364 | -1.2563 | 0.8955 | *TFG* |
| ENSG00000136271 | 7 | 44595016 | 44624650 | 94 | 14 | 36225 | -1.2572 | 0.89565 | *DDX56* |
| ENSG00000171303 | 2 | 26905619 | 26966288 | 142 | 23 | 36328 | -1.2579 | 0.89578 | *KCNK3* |
| ENSG00000102870 | 16 | 30779778 | 30808523 | 30 | 10 | 42692 | -1.2581 | 0.89583 | *ZNF629* |
| ENSG00000077684 | 4 | 129720779 | 129806379 | 140 | 27 | 42328 | -1.2583 | 0.89586 | *JADE1* |
| ENSG00000091128 | 7 | 107653993 | 107780801 | 223 | 28 | 42030 | -1.2585 | 0.89589 | *LAMB4* |
| ENSG00000153179 | 12 | 64994293 | 65101347 | 328 | 38 | 51647 | -1.2591 | 0.896 | *RASSF3* |
| ENSG00000101337 | 20 | 30687309 | 30765061 | 157 | 12 | 45146 | -1.2592 | 0.89602 | *TM9SF4* |
| ENSG00000077585 | 1 | 236295832 | 236395165 | 372 | 40 | 36312 | -1.2598 | 0.89613 | *GPR137B* |
| ENSG00000198954 | 10 | 70738487 | 70786738 | 185 | 24 | 42317 | -1.2602 | 0.8962 | *KIAA1279* |
| ENSG00000101040 | 20 | 45827859 | 45995567 | 318 | 54 | 41388 | -1.2608 | 0.89631 | *ZMYND8* |
| ENSG00000268818 | 22 | 36013040 | 36041181 | 97 | 19 | 37720 | -1.2611 | 0.89637 | *AL049747.1* |
| ENSG00000117597 | 1 | 209991352 | 210040910 | 199 | 21 | 54704 | -1.2615 | 0.89644 | *DIEXF* |
| ENSG00000181585 | 3 | 46732823 | 46762376 | 82 | 20 | 42247 | -1.2617 | 0.89646 | *TMIE* |
| ENSG00000168066 | 11 | 64522078 | 64556258 | 44 | 10 | 36922 | -1.2628 | 0.89667 | *SF1* |
| ENSG00000163254 | 2 | 208982861 | 209004554 | 108 | 17 | 49361 | -1.2629 | 0.89669 | *CRYGC* |
| ENSG00000268735 | 13 | 24312243 | 24333901 | 66 | 13 | 46817 | -1.2635 | 0.8968 | *AL139080.1* |
| ENSG00000214253 | 7 | 100872739 | 100905597 | 156 | 9 | 28943 | -1.2639 | 0.89688 | *FIS1* |
| ENSG00000111481 | 12 | 54684986 | 54755633 | 89 | 13 | 47195 | -1.2657 | 0.89718 | *COPZ1* |
| ENSG00000164331 | 5 | 72838160 | 72871511 | 110 | 12 | 44756 | -1.2664 | 0.89732 | *ANKRA2* |
| ENSG00000157551 | 21 | 39519128 | 39689279 | 535 | 48 | 35333 | -1.268 | 0.89759 | *KCNJ15* |
| ENSG00000139546 | 12 | 53884705 | 53910215 | 19 | 9 | 53501 | -1.2688 | 0.89775 | *TARBP2* |

| | | | | | | | | | |
|---|---|---|---|---|---|---|---|---|---|
| ENSG00000138061 | 2 | 38284116 | 38347044 | 258 | 33 | 38516 | -1.2691 | 0.8978 | *CYP1B1* |
| ENSG00000171224 | 10 | 71380007 | 71403352 | 128 | 19 | 45531 | -1.2693 | 0.89783 | *C10orf35* |
| ENSG00000180773 | 11 | 92867341 | 92941130 | 181 | 22 | 46246 | -1.2705 | 0.89804 | *SLC36A4* |
| ENSG00000242221 | 19 | 43558363 | 43597197 | 273 | 24 | 20071 | -1.2704 | 0.89804 | *PSG2* |
| ENSG00000185164 | 16 | 18501182 | 18583533 | 18 | 13 | 26681 | -1.2706 | 0.89806 | *NOMO2* |
| ENSG00000164953 | 8 | 94757072 | 94841462 | 96 | 18 | 50296 | -1.2708 | 0.8981 | *TMEM67* |
| ENSG00000212743 | 10 | 6382278 | 6404723 | 65 | 15 | 44174 | -1.2713 | 0.89819 | *DKFZP667F0711* |
| ENSG00000120742 | 3 | 150249781 | 150331015 | 281 | 31 | 45796 | -1.2718 | 0.89828 | *SERP1* |
| ENSG00000111052 | 12 | 81176299 | 81341704 | 517 | 48 | 37796 | -1.2719 | 0.8983 | *LIN7A* |
| ENSG00000158716 | 1 | 159740722 | 159762333 | 68 | 14 | 48295 | -1.272 | 0.89832 | *DUSP23* |
| ENSG00000106511 | 7 | 15640837 | 15736437 | 363 | 40 | 47461 | -1.2724 | 0.89838 | *MEOX2* |
| ENSG00000068784 | 2 | 45605819 | 45849304 | 795 | 60 | 38313 | -1.2725 | 0.89841 | *SRBD1* |
| ENSG00000147434 | 8 | 42597763 | 42661535 | 157 | 17 | 35465 | -1.2726 | 0.89842 | *CHRNA6* |
| ENSG00000212722 | 17 | 39251584 | 39272740 | 189 | 9 | 28159 | -1.2736 | 0.89859 | *KRTAP4-9* |
| ENSG00000101280 | 20 | 843296 | 906977 | 310 | 57 | 40826 | -1.2736 | 0.89861 | *ANGPT4* |
| ENSG00000185414 | 2 | 99787542 | 99824089 | 96 | 14 | 42905 | -1.2737 | 0.89862 | *MRPL30* |
| ENSG00000160055 | 1 | 32670073 | 32697972 | 43 | 8 | 32044 | -1.2746 | 0.89878 | *TMEM234* |
| ENSG00000111269 | 12 | 12754761 | 12808042 | 102 | 18 | 48751 | -1.2748 | 0.89881 | *CREBL2* |
| ENSG00000273155 | 2 | 99761461 | 99821761 | 177 | 17 | 42498 | -1.2749 | 0.89882 | *MRPL30* |
| ENSG00000102962 | 16 | 57382684 | 57410102 | 123 | 28 | 46181 | -1.2755 | 0.89893 | *CCL22* |
| ENSG00000151240 | 10 | 310130 | 745683 | 1056 | 78 | 40003 | -1.2755 | 0.89894 | *DIP2C* |
| ENSG00000214324 | 3 | 126901974 | 126927025 | 83 | 25 | 48884 | -1.276 | 0.89903 | *C3orf56* |
| ENSG00000165804 | 14 | 21548205 | 21582881 | 96 | 24 | 43852 | -1.276 | 0.89903 | *ZNF219* |
| ENSG00000137273 | 6 | 1380069 | 1405832 | 83 | 37 | 45598 | -1.2769 | 0.89919 | *FOXF2* |
| ENSG00000196262 | 7 | 44826279 | 44874163 | 119 | 17 | 42786 | -1.277 | 0.8992 | *PPIA* |
| ENSG00000171757 | 3 | 169501216 | 169540774 | 88 | 12 | 46904 | -1.2771 | 0.89922 | *LRRC34* |
| ENSG00000111679 | 12 | 7045631 | 7080479 | 55 | 25 | 43048 | -1.2776 | 0.89931 | *PTPN6* |
| ENSG00000115353 | 2 | 75263590 | 75436826 | 504 | 87 | 40028 | -1.2779 | 0.89936 | *TACR1* |
| ENSG00000226974 | 13 | 64310934 | 64331323 | 51 | 12 | 41939 | -1.2786 | 0.89948 | *AL445989.1* |
| ENSG00000112144 | 6 | 52856077 | 52936600 | 160 | 24 | 47001 | -1.2788 | 0.89951 | *ICK* |
| ENSG00000074657 | 18 | 56519832 | 56663712 | 366 | 43 | 41592 | -1.2789 | 0.89953 | *ZNF532* |
| ENSG00000130348 | 6 | 107067453 | 107126292 | 232 | 28 | 45609 | -1.279 | 0.89955 | *QRSL1* |
| ENSG00000254521 | 19 | 51984611 | 52015043 | 247 | 37 | 53166 | -1.2793 | 0.8996 | *SIGLEC12* |
| ENSG00000134769 | 18 | 32063254 | 32481808 | 1105 | 69 | 50857 | -1.2806 | 0.89983 | *DTNA* |
| ENSG00000117707 | 1 | 214146524 | 214224595 | 213 | 36 | 47462 | -1.2809 | 0.89989 | *PROX1* |
| ENSG00000074695 | 18 | 56985055 | 57037194 | 191 | 21 | 51505 | -1.2812 | 0.89994 | *LMAN1* |

| | | | | | | | | | |
|---|---|---|---|---|---|---|---|---|---|
| ENSG00000105954 | 7 | 25254189 | 25278105 | 110 | 23 | 42905 | -1.2817 | 0.90002 | *NPVF* |
| ENSG00000135847 | 1 | 180234515 | 180482089 | 767 | 55 | 42504 | -1.2822 | 0.90012 | *ACBD6* |
| ENSG00000145246 | 4 | 47477305 | 47605503 | 453 | 57 | 51849 | -1.2826 | 0.90019 | *ATP10D* |
| ENSG00000140406 | 15 | 81283295 | 81306342 | 66 | 9 | 45877 | -1.2828 | 0.90021 | *MESDC1* |
| ENSG00000205106 | 11 | 45782983 | 45803909 | 68 | 16 | 37045 | -1.2829 | 0.90023 | *CTD-2210P24.4* |
| ENSG00000143320 | 1 | 156659398 | 156685608 | 61 | 28 | 44737 | -1.2832 | 0.90028 | *CRABP2* |
| ENSG00000197991 | 13 | 61973818 | 62012220 | 118 | 18 | 35617 | -1.2845 | 0.90051 | *PCDH20* |
| ENSG00000181378 | 2 | 219857568 | 219916249 | 108 | 18 | 35756 | -1.2846 | 0.90053 | *CCDC108* |
| ENSG00000154639 | 21 | 18874700 | 18975897 | 294 | 41 | 46906 | -1.285 | 0.9006 | *CXADR* |
| ENSG00000090661 | 19 | 8261620 | 8337305 | 346 | 76 | 47939 | -1.2854 | 0.90067 | *CERS4* |
| ENSG00000231824 | 18 | 5135284 | 5207502 | 227 | 55 | 42447 | -1.2869 | 0.90094 | *C18orf42* |
| ENSG00000108753 | 17 | 36036435 | 36115237 | 224 | 63 | 46248 | -1.287 | 0.90096 | *HNF1B* |
| ENSG00000038219 | 4 | 13560362 | 13639347 | 163 | 23 | 41441 | -1.2875 | 0.90105 | *BOD1L1* |
| ENSG00000122882 | 10 | 74879913 | 74938813 | 162 | 5 | 18577 | -1.2886 | 0.90123 | *ECD* |
| ENSG00000185049 | 4 | 1974441 | 2053630 | 136 | 22 | 44176 | -1.2886 | 0.90124 | *NELFA* |
| ENSG00000183092 | 14 | 100993486 | 101063750 | 172 | 26 | 34616 | -1.289 | 0.9013 | *BEGAIN* |
| ENSG00000244607 | 3 | 42724155 | 42824745 | 228 | 28 | 53159 | -1.2898 | 0.90144 | *CCDC13* |
| ENSG00000132424 | 6 | 99835927 | 99883207 | 134 | 17 | 41441 | -1.2899 | 0.90145 | *PNISR* |
| ENSG00000092841 | 12 | 56541945 | 56567280 | 24 | 11 | 43991 | -1.2904 | 0.90154 | *MYL6* |
| ENSG00000182896 | 17 | 7248442 | 7270538 | 54 | 21 | 42472 | -1.2905 | 0.90156 | *TMEM95* |
| ENSG00000170486 | 12 | 52969373 | 53005322 | 171 | 23 | 53115 | -1.2907 | 0.90159 | *KRT72* |
| ENSG00000173852 | 7 | 34958488 | 35087883 | 496 | 30 | 53744 | -1.2908 | 0.90161 | *DPY19L1* |
| ENSG00000196946 | 12 | 8280733 | 8351133 | 165 | 13 | 34788 | -1.291 | 0.90164 | *ZNF705A* |
| ENSG00000095970 | 6 | 41116244 | 41140924 | 21 | 10 | 39260 | -1.2914 | 0.90171 | *TREM2* |
| ENSG00000008952 | 3 | 169674423 | 169726161 | 111 | 14 | 43845 | -1.2921 | 0.90183 | *SEC62* |
| ENSG00000100162 | 22 | 42324725 | 42353168 | 107 | 18 | 37888 | -1.2921 | 0.90184 | *CENPM* |
| ENSG00000006468 | 7 | 13920853 | 14041050 | 511 | 82 | 48870 | -1.2922 | 0.90186 | *ETV1* |
| ENSG00000109182 | 4 | 48978264 | 49074098 | 180 | 14 | 31227 | -1.2928 | 0.90196 | *CWH43* |
| ENSG00000185585 | 9 | 127529437 | 127587164 | 83 | 26 | 49829 | -1.2934 | 0.90206 | *OLFML2A* |
| ENSG00000177468 | 6 | 137803336 | 137825531 | 83 | 21 | 43495 | -1.2935 | 0.90209 | *OLIG3* |
| ENSG00000231122 | 10 | 46300876 | 46323036 | 8 | 5 | 16700 | -1.2939 | 0.90215 | *FAM25E* |
| ENSG00000126856 | 16 | 90112974 | 90168480 | 147 | 27 | 39786 | -1.2944 | 0.90223 | *PRDM7* |
| ENSG00000144895 | 3 | 150254465 | 150312029 | 195 | 25 | 47340 | -1.2947 | 0.90229 | *EIF2A* |
| ENSG00000024862 | 6 | 139084657 | 139124456 | 144 | 21 | 41365 | -1.295 | 0.90234 | *CCDC28A* |
| ENSG00000132646 | 20 | 5085599 | 5117272 | 78 | 8 | 27871 | -1.295 | 0.90234 | *PCNA* |
| ENSG00000121775 | 1 | 32527632 | 32578467 | 78 | 14 | 34742 | -1.2952 | 0.90238 | *TMEM39B* |

| | | | | | | | | | |
|---|---|---|---|---|---|---|---|---|---|
| ENSG00000214087 | 17 | 79638204 | 79660954 | 135 | 16 | 50857 | -1.2953 | 0.90239 | *ARL16* |
| ENSG00000107954 | 10 | 105243736 | 105362309 | 340 | 53 | 48047 | -1.2973 | 0.90273 | *NEURL1* |
| ENSG00000147883 | 9 | 21992902 | 22019362 | 65 | 14 | 42163 | -1.2975 | 0.90277 | *CDKN2B* |
| ENSG00000024048 | 6 | 42521800 | 42671242 | 359 | 31 | 54500 | -1.2979 | 0.90283 | *UBR2* |
| ENSG00000138496 | 3 | 122236771 | 122293424 | 141 | 21 | 52861 | -1.2988 | 0.90299 | *PARP9* |
| ENSG00000169760 | 3 | 173104074 | 174014434 | 2709 | 217 | 41342 | -1.2989 | 0.90301 | *NLGN1* |
| ENSG00000128710 | 2 | 176963518 | 176994670 | 66 | 20 | 42452 | -1.2995 | 0.90312 | *HOXD10* |
| ENSG00000255168 | 1 | 145199145 | 145329796 | 8 | 5 | 39298 | -1.2998 | 0.90317 | *RP11-458D21.5* |
| ENSG00000139433 | 12 | 110278748 | 110328293 | 125 | 21 | 44103 | -1.2999 | 0.90318 | *GLTP* |
| ENSG00000164434 | 6 | 123090620 | 123115219 | 64 | 22 | 39082 | -1.3022 | 0.90357 | *FABP7* |
| ENSG00000189052 | 19 | 49537141 | 49558568 | 81 | 21 | 47796 | -1.3023 | 0.90359 | *CGB5* |
| ENSG00000258928 | 14 | 52372863 | 52395289 | 68 | 19 | 37923 | -1.3025 | 0.90363 | *RP11-463J10.2* |
| ENSG00000164199 | 5 | 89815161 | 90470038 | 1770 | 146 | 43720 | -1.3028 | 0.90368 | *GPR98* |
| ENSG00000106628 | 7 | 44144286 | 44173957 | 83 | 16 | 47734 | -1.304 | 0.90388 | *POLD2* |
| ENSG00000069122 | 6 | 46810249 | 46932680 | 475 | 58 | 37545 | -1.3043 | 0.90393 | *GPR116* |
| ENSG00000131849 | 19 | 58934181 | 58961589 | 72 | 11 | 44325 | -1.3044 | 0.90395 | *ZNF132* |
| ENSG00000038382 | 5 | 14133811 | 14542235 | 1100 | 94 | 44460 | -1.3047 | 0.90401 | *TRIO* |
| ENSG00000133773 | 12 | 82607460 | 82762584 | 447 | 43 | 47352 | -1.305 | 0.90406 | *CCDC59* |
| ENSG00000162543 | 1 | 20502578 | 20532541 | 57 | 12 | 46264 | -1.3052 | 0.90408 | *UBXN10* |
| ENSG00000136758 | 10 | 27389383 | 27454195 | 281 | 23 | 37805 | -1.3056 | 0.90416 | *YME1L1* |
| ENSG00000163781 | 3 | 133307019 | 133390737 | 270 | 25 | 51010 | -1.3057 | 0.90417 | *TOPBP1* |
| ENSG00000157193 | 1 | 53701217 | 53803742 | 245 | 44 | 37142 | -1.3058 | 0.9042 | *LRP8* |
| ENSG00000163380 | 3 | 69146023 | 69182183 | 75 | 20 | 42947 | -1.3063 | 0.90427 | *LMOD3* |
| ENSG00000181733 | 19 | 8831316 | 8852371 | 38 | 9 | 34901 | -1.3068 | 0.90435 | *OR2Z1* |
| ENSG00000166573 | 18 | 74952505 | 74990858 | 216 | 37 | 49392 | -1.3074 | 0.90447 | *GALR1* |
| ENSG00000101166 | 20 | 57598200 | 57627964 | 60 | 13 | 56663 | -1.3076 | 0.90449 | *SLMO2* |
| ENSG00000113946 | 3 | 190030330 | 190139932 | 574 | 60 | 52545 | -1.3076 | 0.9045 | *CLDN16* |
| ENSG00000198843 | 3 | 150310662 | 150358222 | 143 | 21 | 46311 | -1.3079 | 0.90455 | *SELT* |
| ENSG00000126262 | 19 | 35924809 | 35952669 | 106 | 35 | 51011 | -1.3084 | 0.90463 | *FFAR2* |
| ENSG00000138399 | 2 | 170376259 | 170440385 | 228 | 34 | 48814 | -1.3086 | 0.90466 | *FASTKD1* |
| ENSG00000172296 | 20 | 12979627 | 13157411 | 902 | 95 | 50771 | -1.3088 | 0.9047 | *SPTLC3* |
| ENSG00000165389 | 14 | 34891995 | 34941562 | 148 | 21 | 40396 | -1.31 | 0.9049 | *SPTSSA* |
| ENSG00000198064 | 16 | 30224258 | 30275831 | 1 | 1 | 43270 | -1.31 | 0.9049 | *RP11-347C12.1* |
| ENSG00000081386 | 9 | 99508147 | 99550411 | 32 | 12 | 39448 | -1.3102 | 0.90493 | *ZNF510* |
| ENSG00000084623 | 1 | 32677529 | 32707205 | 51 | 8 | 30283 | -1.3107 | 0.90502 | *EIF3I* |
| ENSG00000134452 | 10 | 5921535 | 5989556 | 295 | 40 | 50082 | -1.3108 | 0.90503 | *FBXO18* |

| | | | | | | | | | |
|---|---|---|---|---|---|---|---|---|---|
| ENSG00000270149 | 1 | 160957747 | 161018752 | 193 | 21 | 46515 | -1.3114 | 0.90514 | *RP11-544M22.13* |
| ENSG00000166788 | 11 | 18081482 | 18137638 | 169 | 15 | 48572 | -1.3117 | 0.90519 | *SAAL1* |
| ENSG00000135205 | 7 | 76741751 | 76968850 | 812 | 77 | 45260 | -1.3119 | 0.90523 | *CCDC146* |
| ENSG00000170315 | 17 | 16274112 | 16296059 | 49 | 11 | 41751 | -1.313 | 0.9054 | *UBB* |
| ENSG00000100353 | 22 | 36896897 | 36935483 | 196 | 28 | 39789 | -1.3134 | 0.90547 | *EIF3D* |
| ENSG00000044574 | 9 | 127987132 | 128013609 | 43 | 11 | 44925 | -1.3138 | 0.90554 | *HSPA5* |
| ENSG00000051523 | 16 | 88699691 | 88727560 | 114 | 37 | 43707 | -1.3139 | 0.90556 | *CYBA* |
| ENSG00000196655 | 11 | 118879142 | 118906164 | 104 | 15 | 48989 | -1.3143 | 0.90562 | *TRAPPC4* |
| ENSG00000073754 | 1 | 157790704 | 157878046 | 285 | 30 | 41363 | -1.3147 | 0.9057 | *CD5L* |
| ENSG00000164695 | 8 | 82634669 | 82681750 | 155 | 27 | 44127 | -1.3158 | 0.90588 | *CHMP4C* |
| ENSG00000116903 | 1 | 231458480 | 231483598 | 58 | 9 | 47067 | -1.3159 | 0.9059 | *EXOC8* |
| ENSG00000062582 | 7 | 43896157 | 43919492 | 64 | 12 | 49359 | -1.3166 | 0.90602 | *MRPS24* |
| ENSG00000223443 | 8 | 11984677 | 12006586 | 10 | 4 | 42889 | -1.3171 | 0.90609 | *USP17L2* |
| ENSG00000119328 | 9 | 111686461 | 111723024 | 115 | 18 | 50769 | -1.3172 | 0.90612 | *FAM206A* |
| ENSG00000171443 | 19 | 56101388 | 56124504 | 57 | 18 | 49681 | -1.3179 | 0.90624 | *ZNF524* |
| ENSG00000163322 | 4 | 84372092 | 84454501 | 272 | 18 | 35513 | -1.3189 | 0.9064 | *FAM175A* |
| ENSG00000132938 | 13 | 29588748 | 30087892 | 1954 | 155 | 42769 | -1.3189 | 0.9064 | *MTUS2* |
| ENSG00000092203 | 14 | 21934756 | 21977319 | 145 | 17 | 50253 | -1.3197 | 0.90653 | *TOX4* |
| ENSG00000121351 | 12 | 21497893 | 21542912 | 196 | 28 | 46326 | -1.3198 | 0.90654 | *IAPP* |
| ENSG00000064012 | 2 | 202088166 | 202162434 | 220 | 34 | 34390 | -1.3203 | 0.90664 | *CASP8* |
| ENSG00000174444 | 15 | 66780355 | 66826870 | 145 | 22 | 55050 | -1.3204 | 0.90664 | *RPL4* |
| ENSG00000204740 | 10 | 19482779 | 20089330 | 2759 | 179 | 47587 | -1.322 | 0.90691 | *MALRD1* |
| ENSG00000121486 | 1 | 185077220 | 185136204 | 107 | 10 | 29462 | -1.3222 | 0.90694 | *TRMT1L* |
| ENSG00000130303 | 19 | 17503748 | 17526457 | 74 | 15 | 34505 | -1.3226 | 0.90702 | *BST2* |
| ENSG00000112280 | 6 | 70914764 | 71022786 | 509 | 67 | 44494 | -1.3229 | 0.90706 | *COL9A1* |
| ENSG00000104941 | 19 | 46288968 | 46328577 | 142 | 20 | 53477 | -1.3237 | 0.90719 | *RSPH6A* |
| ENSG00000137502 | 11 | 82674175 | 82792965 | 286 | 36 | 45941 | -1.3237 | 0.9072 | *RAB30* |
| ENSG00000186897 | 12 | 49716200 | 49740971 | 58 | 14 | 47016 | -1.3241 | 0.90727 | *C1QL4* |
| ENSG00000132164 | 3 | 10847885 | 10992419 | 477 | 74 | 48281 | -1.3243 | 0.9073 | *SLC6A11* |
| ENSG00000105254 | 19 | 36595191 | 36626849 | 103 | 20 | 51229 | -1.3245 | 0.90733 | *TBCB* |
| ENSG00000185905 | 16 | 29743784 | 29767327 | 45 | 6 | 44383 | -1.3259 | 0.90757 | *C16orf54* |
| ENSG00000165140 | 9 | 97355415 | 97412531 | 254 | 28 | 50013 | -1.3262 | 0.90762 | *FBP1* |
| ENSG00000172264 | 20 | 13966015 | 16043842 | 6965 | 501 | 43468 | -1.3263 | 0.90763 | *MACROD2* |
| ENSG00000203857 | 1 | 120039821 | 120067681 | 59 | 8 | 30569 | -1.3267 | 0.90769 | *HSD3B1* |
| ENSG00000097096 | 1 | 85612556 | 85676729 | 219 | 25 | 40888 | -1.3271 | 0.90776 | *SYDE2* |
| ENSG00000169340 | 16 | 20360492 | 20426059 | 213 | 38 | 42673 | -1.3274 | 0.90782 | *PDILT* |

| | | | | | | | | | |
|---|---|---|---|---|---|---|---|---|---|
| ENSG00000203876 | 10 | 111691029 | 111723649 | 69 | 5 | 19671 | -1.3283 | 0.90797 | *RP11-451M19.3* |
| ENSG00000239474 | 2 | 170356212 | 170392772 | 102 | 22 | 51505 | -1.329 | 0.90808 | *KLHL41* |
| ENSG00000185630 | 1 | 164514821 | 164878533 | 1067 | 169 | 47406 | -1.3293 | 0.90813 | *PBX1* |
| ENSG00000179709 | 19 | 56449198 | 56509995 | 348 | 56 | 52323 | -1.3293 | 0.90813 | *NLRP8* |
| ENSG00000136111 | 13 | 75848808 | 76066250 | 606 | 90 | 37700 | -1.3295 | 0.90816 | *TBC1D4* |
| ENSG00000174446 | 15 | 66772473 | 66800151 | 90 | 18 | 54464 | -1.3306 | 0.90834 | *SNAPC5* |
| ENSG00000178038 | 3 | 46700487 | 46745194 | 140 | 24 | 46921 | -1.3332 | 0.90877 | *ALS2CL* |
| ENSG00000221852 | 17 | 39172278 | 39193454 | 92 | 12 | 38561 | -1.3341 | 0.90891 | *KRTAP1-5* |
| ENSG00000177675 | 12 | 7489281 | 7642493 | 373 | 55 | 36808 | -1.3343 | 0.90894 | *CD163L1* |
| ENSG00000101076 | 20 | 42974340 | 43071485 | 261 | 41 | 46860 | -1.3345 | 0.90898 | *HNF4A* |
| ENSG00000175931 | 17 | 74375532 | 74459288 | 263 | 34 | 38614 | -1.3346 | 0.909 | *UBE2O* |
| ENSG00000127586 | 16 | 828046 | 860737 | 170 | 34 | 38045 | -1.3347 | 0.90901 | *CHTF18* |
| ENSG00000204209 | 6 | 33276335 | 33307046 | 62 | 11 | 37348 | -1.3352 | 0.9091 | *DAXX* |
| ENSG00000136697 | 2 | 113815547 | 113843427 | 226 | 27 | 34770 | -1.3354 | 0.90913 | *IL1F10* |
| ENSG00000185674 | 2 | 99848709 | 99881745 | 90 | 20 | 48722 | -1.3366 | 0.90933 | *LYG2* |
| ENSG00000136770 | 10 | 22035466 | 22302698 | 495 | 44 | 43058 | -1.3369 | 0.90937 | *DNAJC1* |
| ENSG00000112697 | 6 | 75952640 | 76004684 | 106 | 14 | 30914 | -1.3377 | 0.9095 | *TMEM30A* |
| ENSG00000177469 | 17 | 40544470 | 40585535 | 135 | 18 | 38113 | -1.3377 | 0.9095 | *PTRF* |
| ENSG00000102977 | 16 | 67681415 | 67704713 | 26 | 9 | 43378 | -1.3377 | 0.90951 | *ACD* |
| ENSG00000158411 | 2 | 99767890 | 99807521 | 114 | 12 | 41325 | -1.3379 | 0.90954 | *MITD1* |
| ENSG00000101057 | 20 | 42285754 | 42355136 | 191 | 19 | 46466 | -1.3382 | 0.90958 | *MYBL2* |
| ENSG00000115705 | 2 | 1367995 | 1557483 | 864 | 97 | 45267 | -1.3386 | 0.90965 | *TPO* |
| ENSG00000266958 | 19 | 45680646 | 45730191 | 250 | 41 | 35642 | -1.3405 | 0.90995 | *AC006126.3* |
| ENSG00000240871 | 17 | 39230459 | 39251396 | 175 | 6 | 49922 | -1.3407 | 0.90999 | *KRTAP4-7* |
| ENSG00000178229 | 19 | 57821877 | 57852144 | 156 | 25 | 52793 | -1.3407 | 0.90999 | *ZNF543* |
| ENSG00000268865 | 12 | 25045243 | 25067615 | 91 | 22 | 51298 | -1.3419 | 0.91018 | *AC026310.1* |
| ENSG00000189169 | 21 | 46107087 | 46127959 | 124 | 21 | 45623 | -1.343 | 0.91036 | *KRTAP10-12* |
| ENSG00000169744 | 4 | 16493164 | 16910432 | 1233 | 177 | 48250 | -1.3434 | 0.91043 | *LDB2* |
| ENSG00000105677 | 19 | 36026497 | 36048428 | 56 | 15 | 49864 | -1.344 | 0.91052 | *TMEM147* |
| ENSG00000187955 | 8 | 121062019 | 121394275 | 909 | 99 | 43407 | -1.3447 | 0.91064 | *COL14A1* |
| ENSG00000102468 | 13 | 47395685 | 47481169 | 308 | 51 | 49142 | -1.3449 | 0.91067 | *HTR2A* |
| ENSG00000078403 | 10 | 21813094 | 22042559 | 256 | 50 | 46640 | -1.3455 | 0.91076 | *MLLT10* |
| ENSG00000198783 | 17 | 33278549 | 33300205 | 37 | 12 | 49233 | -1.3455 | 0.91076 | *ZNF830* |
| ENSG00000161642 | 12 | 54752917 | 54795082 | 64 | 16 | 37633 | -1.3457 | 0.91081 | *ZNF385A* |
| ENSG00000021852 | 1 | 57384883 | 57441813 | 241 | 37 | 46234 | -1.346 | 0.91084 | *C8B* |
| ENSG00000178531 | 19 | 7979372 | 8001051 | 75 | 16 | 50131 | -1.3465 | 0.91094 | *CTXN1* |

| | | | | | | | | | |
|---|---|---|---|---|---|---|---|---|---|
| ENSG00000153885 | 19 | 34276838 | 34316668 | 120 | 29 | 49519 | -1.3466 | 0.91094 | *KCTD15* |
| ENSG00000204702 | 6 | 29058386 | 29079658 | 77 | 14 | 39695 | -1.3477 | 0.91113 | *OR2J1* |
| ENSG00000174948 | 3 | 154045461 | 154157504 | 347 | 46 | 49096 | -1.3478 | 0.91114 | *GPR149* |
| ENSG00000129518 | 14 | 34975135 | 35018916 | 134 | 18 | 43319 | -1.3484 | 0.91123 | *EAPP* |
| ENSG00000070985 | 11 | 2415745 | 2454275 | 128 | 28 | 43817 | -1.3502 | 0.91153 | *TRPM5* |
| ENSG00000166840 | 11 | 58662871 | 58821000 | 481 | 35 | 50386 | -1.3506 | 0.91158 | *GLYATL1* |
| ENSG00000101182 | 20 | 60701791 | 60728496 | 74 | 9 | 34438 | -1.3507 | 0.91161 | *PSMA7* |
| ENSG00000160202 | 21 | 44579118 | 44602915 | 49 | 12 | 43930 | -1.3512 | 0.91169 | *CRYAA* |
| ENSG00000148488 | 10 | 17350382 | 17506329 | 939 | 88 | 48812 | -1.3518 | 0.91179 | *ST8SIA6* |
| ENSG00000007376 | 16 | 824974 | 848397 | 123 | 24 | 39048 | -1.3522 | 0.91184 | *RPUSD1* |
| ENSG00000139767 | 12 | 119409300 | 119610856 | 827 | 91 | 49972 | -1.3528 | 0.91194 | *SRRM4* |
| ENSG00000185800 | 19 | 46276205 | 46306060 | 58 | 14 | 52276 | -1.3531 | 0.91199 | *DMWD* |
| ENSG00000186104 | 11 | 14889553 | 14923798 | 42 | 14 | 48321 | -1.3535 | 0.91206 | *CYP2R1* |
| ENSG00000146910 | 7 | 155256901 | 155336557 | 254 | 72 | 46539 | -1.3537 | 0.91209 | *CNPY1* |
| ENSG00000205456 | 16 | 32254040 | 32277240 | 10 | 6 | 42302 | -1.3545 | 0.91221 | *TP53TG3D* |
| ENSG00000176971 | 11 | 27005628 | 27028630 | 110 | 19 | 51153 | -1.3546 | 0.91224 | *FIBIN* |
| ENSG00000163064 | 2 | 119589747 | 119615254 | 55 | 15 | 41065 | -1.3548 | 0.91226 | *EN1* |
| ENSG00000163534 | 1 | 157754193 | 157799895 | 104 | 15 | 51131 | -1.3554 | 0.91235 | *FCRL1* |
| ENSG00000130700 | 20 | 61028553 | 61061026 | 91 | 24 | 42848 | -1.3555 | 0.91237 | *GATA5* |
| ENSG00000213020 | 19 | 53196066 | 53248326 | 240 | 30 | 47453 | -1.3561 | 0.91247 | *ZNF611* |
| ENSG00000133256 | 4 | 609373 | 674571 | 220 | 22 | 53004 | -1.3566 | 0.91254 | *PDE6B* |
| ENSG00000173545 | 5 | 16441628 | 16475901 | 76 | 14 | 52480 | -1.3568 | 0.91258 | *ZNF622* |
| ENSG00000139636 | 12 | 49480919 | 49514683 | 47 | 9 | 34470 | -1.3582 | 0.91279 | *LMBR1L* |
| ENSG00000206075 | 18 | 61133994 | 61182318 | 231 | 22 | 35751 | -1.361 | 0.91324 | *SERPINB5* |
| ENSG00000130940 | 1 | 10686661 | 10866707 | 489 | 118 | 44030 | -1.362 | 0.9134 | *CASZ1* |
| ENSG00000163531 | 1 | 204787779 | 205001950 | 579 | 91 | 38996 | -1.364 | 0.91372 | *NFASC* |
| ENSG00000142188 | 21 | 34794792 | 34862318 | 234 | 17 | 38301 | -1.3644 | 0.91378 | *TMEM50B* |
| ENSG00000129673 | 17 | 74439433 | 74476199 | 127 | 29 | 38345 | -1.3651 | 0.91389 | *AANAT* |
| ENSG00000175265 | 15 | 34661269 | 34890704 | 283 | 42 | 47017 | -1.3658 | 0.91399 | *GOLGA8A* |
| ENSG00000158623 | 7 | 130136089 | 130363598 | 323 | 32 | 40057 | -1.3663 | 0.91408 | *COPG2* |
| ENSG00000183629 | 15 | 28754757 | 28788160 | 3 | 1 | 43288 | -1.3663 | 0.91408 | *GOLGA8G* |
| ENSG00000166676 | 16 | 10844776 | 10922651 | 147 | 39 | 36611 | -1.367 | 0.91419 | *TVP23A* |
| ENSG00000259518 | 15 | 82370935 | 82400035 | 137 | 36 | 35168 | -1.3682 | 0.91438 | *RP11-597K23.2* |
| ENSG00000130479 | 19 | 17820051 | 17855325 | 147 | 20 | 37651 | -1.3692 | 0.91453 | *MAP1S* |
| ENSG00000096395 | 6 | 33752450 | 33781788 | 111 | 15 | 53965 | -1.3693 | 0.91454 | *MLN* |
| ENSG00000152137 | 12 | 119606447 | 119668936 | 171 | 38 | 46910 | -1.3708 | 0.91478 | *HSPB8* |

| | | | | | | | | | |
|---|---|---|---|---|---|---|---|---|---|
| ENSG00000137573 | 8 | 70368859 | 70583150 | 586 | 136 | 47423 | -1.371 | 0.91481 | *SULF1* |
| ENSG00000148331 | 9 | 132389171 | 132414444 | 69 | 19 | 48479 | -1.3714 | 0.91488 | *ASB6* |
| ENSG00000148484 | 10 | 16622610 | 16869527 | 880 | 121 | 42260 | -1.3721 | 0.91498 | *RSU1* |
| ENSG00000104976 | 19 | 7975201 | 7998135 | 68 | 22 | 46851 | -1.3721 | 0.91499 | *SNAPC2* |
| ENSG00000154822 | 3 | 16834159 | 17142086 | 1014 | 81 | 48580 | -1.3726 | 0.91506 | *PLCL2* |
| ENSG00000135476 | 12 | 53652083 | 53697427 | 109 | 18 | 41631 | -1.3733 | 0.91517 | *ESPL1* |
| ENSG00000136816 | 9 | 132555432 | 132583560 | 102 | 11 | 53671 | -1.3736 | 0.91521 | *TOR1B* |
| ENSG00000269446 | 7 | 156892674 | 156913764 | 88 | 13 | 42239 | -1.3739 | 0.91527 | *AC006967.1* |
| ENSG00000142731 | 4 | 128792016 | 128830350 | 29 | 13 | 45112 | -1.3744 | 0.91534 | *PLK4* |
| ENSG00000100483 | 14 | 50565350 | 50593318 | 100 | 16 | 51718 | -1.3754 | 0.9155 | *VCPKMT* |
| ENSG00000136381 | 15 | 78719773 | 78803798 | 202 | 22 | 45333 | -1.3754 | 0.9155 | *IREB2* |
| ENSG00000196878 | 1 | 209778215 | 209835811 | 208 | 49 | 44755 | -1.3763 | 0.91564 | *LAMB3* |
| ENSG00000185385 | 19 | 14981138 | 15002264 | 128 | 21 | 42696 | -1.3766 | 0.91568 | *OR7A17* |
| ENSG00000117983 | 11 | 1234296 | 1293406 | 354 | 28 | 51149 | -1.3779 | 0.91588 | *MUC5B* |
| ENSG00000139190 | 12 | 6561403 | 6590153 | 133 | 16 | 51582 | -1.3779 | 0.91588 | *VAMP1* |
| ENSG00000166024 | 10 | 99884387 | 100014654 | 270 | 23 | 53713 | -1.3786 | 0.91598 | *R3HCC1L* |
| ENSG00000023572 | 1 | 193055598 | 193085244 | 77 | 12 | 51963 | -1.3787 | 0.916 | *GLRX2* |
| ENSG00000145781 | 5 | 115410688 | 115758459 | 1848 | 69 | 36763 | -1.3791 | 0.91607 | *COMMD10* |
| ENSG00000049449 | 11 | 31823939 | 32137301 | 793 | 115 | 49528 | -1.3792 | 0.91609 | *RCN1* |
| ENSG00000088766 | 20 | 5976736 | 6030699 | 193 | 27 | 47454 | -1.3804 | 0.91626 | *CRLS1* |
| ENSG00000175003 | 6 | 160532821 | 160589750 | 179 | 44 | 43213 | -1.3805 | 0.91628 | *SLC22A1* |
| ENSG00000224130 | 11 | 58662897 | 58686071 | 74 | 15 | 37164 | -1.3806 | 0.9163 | *AP001652.1* |
| ENSG00000146221 | 6 | 44236480 | 44275458 | 141 | 18 | 36081 | -1.3811 | 0.91638 | *TCTE1* |
| ENSG00000196305 | 9 | 94962489 | 95066038 | 306 | 23 | 46001 | -1.3815 | 0.91644 | *IARS* |
| ENSG00000268942 | 5 | 61798070 | 61818309 | 35 | 8 | 54498 | -1.3834 | 0.91672 | *CKS1B* |
| ENSG00000183310 | 1 | 248727022 | 248748080 | 3 | 2 | 43173 | -1.3838 | 0.9168 | *OR2T34* |
| ENSG00000269810 | 15 | 100028447 | 100048578 | 98 | 13 | 42255 | -1.3839 | 0.9168 | *AC015660.1* |
| ENSG00000152944 | 12 | 27165479 | 27229276 | 242 | 26 | 39883 | -1.3842 | 0.91685 | *MED21* |
| ENSG00000242950 | 7 | 92087694 | 92117300 | 80 | 15 | 49318 | -1.3843 | 0.91686 | *ERVW-1* |
| ENSG00000148700 | 10 | 111746126 | 111905323 | 251 | 29 | 28681 | -1.3843 | 0.91687 | *ADD3* |
| ENSG00000146278 | 6 | 89780470 | 89804879 | 78 | 12 | 51067 | -1.3853 | 0.91702 | *PNRC1* |
| ENSG00000204701 | 6 | 29069668 | 29090603 | 81 | 16 | 39401 | -1.3857 | 0.91709 | *OR2J3* |
| ENSG00000160185 | 21 | 43814008 | 43877791 | 337 | 46 | 47335 | -1.3858 | 0.91709 | *UBASH3A* |
| ENSG00000265352 | 18 | 69389851 | 69465364 | 433 | 33 | 46189 | -1.386 | 0.91713 | *RP11-723G8.2* |
| ENSG00000086619 | 1 | 236368855 | 236455319 | 461 | 25 | 39381 | -1.3861 | 0.91714 | *ERO1LB* |
| ENSG00000267950 | 11 | 1285809 | 1306033 | 64 | 16 | 55216 | -1.3863 | 0.91717 | *AC136297.1* |

| | | | | | | | | | |
|---|---|---|---|---|---|---|---|---|---|
| ENSG00000232070 | 14 | 21557096 | 21581883 | 63 | 18 | 40973 | -1.3866 | 0.91722 | *TMEM253* |
| ENSG00000155307 | 21 | 15847549 | 15965723 | 461 | 69 | 43143 | -1.3866 | 0.91722 | *SAMSN1* |
| ENSG00000205571 | 5 | 69335350 | 69384349 | 2 | 1 | 43295 | -1.3873 | 0.91733 | *SMN2* |
| ENSG00000178922 | 1 | 43906824 | 43929660 | 37 | 8 | 37874 | -1.3876 | 0.91738 | *HYI* |
| ENSG00000176281 | 14 | 20378741 | 20399818 | 11 | 3 | 44166 | -1.3877 | 0.91739 | *OR4K5* |
| ENSG00000123213 | 5 | 65008023 | 65177553 | 603 | 69 | 51696 | -1.3879 | 0.91742 | *NLN* |
| ENSG00000115596 | 2 | 219714544 | 219748955 | 48 | 13 | 37227 | -1.3884 | 0.91749 | *WNT6* |
| ENSG00000172216 | 20 | 48797376 | 48819212 | 26 | 9 | 49290 | -1.3886 | 0.91753 | *CEBPB* |
| ENSG00000111537 | 12 | 68538548 | 68563527 | 68 | 13 | 33602 | -1.3897 | 0.91769 | *IFNG* |
| ENSG00000153487 | 13 | 111355083 | 111383421 | 93 | 11 | 34458 | -1.3898 | 0.9177 | *ING1* |
| ENSG00000164304 | 6 | 7316889 | 7399976 | 331 | 35 | 42241 | -1.3902 | 0.91777 | *CAGE1* |
| ENSG00000135622 | 2 | 74871355 | 74919186 | 29 | 15 | 53250 | -1.391 | 0.91789 | *SEMA4F* |
| ENSG00000174804 | 11 | 86646721 | 86676433 | 83 | 16 | 48526 | -1.3913 | 0.91794 | *FZD4* |
| ENSG00000178233 | 6 | 44228203 | 44285243 | 194 | 25 | 40368 | -1.3917 | 0.91799 | *TMEM151B* |
| ENSG00000139131 | 12 | 32870424 | 32918836 | 172 | 11 | 49502 | -1.3925 | 0.91811 | *YARS2* |
| ENSG00000169777 | 5 | 9619109 | 9722490 | 296 | 55 | 51140 | -1.3934 | 0.91826 | *TAS2R1* |
| ENSG00000178814 | 8 | 145096167 | 145128735 | 45 | 12 | 42247 | -1.3937 | 0.9183 | *OPLAH* |
| ENSG00000007047 | 19 | 45572546 | 45818541 | 896 | 114 | 45525 | -1.3938 | 0.91831 | *MARK4* |
| ENSG00000136603 | 3 | 170065466 | 170124623 | 174 | 26 | 46020 | -1.3939 | 0.91832 | *SKIL* |
| ENSG00000188069 | 11 | 4780209 | 4801168 | 167 | 13 | 39124 | -1.3947 | 0.91845 | *OR51F1* |
| ENSG00000177272 | 1 | 111204310 | 111227655 | 58 | 12 | 53410 | -1.3955 | 0.91856 | *KCNA3* |
| ENSG00000077232 | 2 | 183570999 | 183669191 | 204 | 16 | 44707 | -1.3958 | 0.91862 | *DNAJC10* |
| ENSG00000197498 | 6 | 111293218 | 111359466 | 353 | 28 | 43017 | -1.3966 | 0.91873 | *RPF2* |
| ENSG00000170484 | 12 | 52949566 | 52977609 | 159 | 34 | 47174 | -1.3971 | 0.9188 | *KRT74* |
| ENSG00000196465 | 12 | 56536040 | 56563431 | 25 | 11 | 49304 | -1.3972 | 0.91882 | *MYL6B* |
| ENSG00000127952 | 7 | 75615656 | 75687322 | 181 | 26 | 40181 | -1.3973 | 0.91884 | *STYXL1* |
| ENSG00000162757 | 1 | 209945661 | 209967904 | 52 | 12 | 53572 | -1.3986 | 0.91903 | *C1orf74* |
| ENSG00000251655 | 12 | 11494757 | 11558500 | 114 | 41 | 43901 | -1.399 | 0.91909 | *PRB1* |
| ENSG00000134775 | 18 | 33867677 | 34370018 | 1374 | 155 | 46380 | -1.4001 | 0.91925 | *FHOD3* |
| ENSG00000169020 | 4 | 656225 | 678127 | 56 | 12 | 51213 | -1.4001 | 0.91926 | *ATP5I* |
| ENSG00000125733 | 19 | 6727936 | 6761537 | 96 | 30 | 49880 | -1.4001 | 0.91926 | *TRIP10* |
| ENSG00000131482 | 17 | 41042814 | 41075386 | 69 | 14 | 44910 | -1.4008 | 0.91936 | *G6PC* |
| ENSG00000128915 | 15 | 60701808 | 60781359 | 176 | 21 | 42300 | -1.4014 | 0.91945 | *NARG2* |
| ENSG00000137509 | 11 | 82524544 | 82691626 | 503 | 41 | 44898 | -1.4019 | 0.91953 | *PRCP* |
| ENSG00000166387 | 11 | 7524529 | 7688358 | 789 | 110 | 45619 | -1.4024 | 0.9196 | *PPFIBP2* |
| ENSG00000183726 | 1 | 25654408 | 25698852 | 36 | 11 | 38753 | -1.4025 | 0.91962 | *TMEM50A* |

| | | | | | | | | | |
|---|---|---|---|---|---|---|---|---|---|
| ENSG00000203697 | 1 | 223701349 | 223863436 | 393 | 55 | 48896 | -1.4025 | 0.91962 | *CAPN8* |
| ENSG00000139626 | 12 | 53575102 | 53611091 | 64 | 16 | 42567 | -1.4033 | 0.91974 | *ITGB7* |
| ENSG00000173376 | 4 | 121946768 | 122004176 | 170 | 25 | 38729 | -1.4035 | 0.91977 | *NDNF* |
| ENSG00000105261 | 19 | 36592104 | 36614613 | 69 | 14 | 53434 | -1.404 | 0.91984 | *OVOL3* |
| ENSG00000151612 | 4 | 146668779 | 146869787 | 427 | 69 | 41237 | -1.4041 | 0.91985 | *ZNF827* |
| ENSG00000161904 | 6 | 33728979 | 33766913 | 180 | 17 | 49372 | -1.4045 | 0.91992 | *LEMD2* |
| ENSG00000106278 | 7 | 121503143 | 121712090 | 542 | 59 | 45073 | -1.4051 | 0.92001 | *PTPRZ1* |
| ENSG00000020922 | 11 | 94142895 | 94237074 | 272 | 31 | 44293 | -1.4052 | 0.92003 | *MRE11A* |
| ENSG00000203705 | 1 | 212955170 | 212999968 | 234 | 15 | 50387 | -1.4055 | 0.92006 | *TATDN3* |
| ENSG00000105258 | 19 | 36594612 | 36616248 | 68 | 14 | 52953 | -1.4065 | 0.92021 | *POLR2I* |
| ENSG00000135334 | 6 | 88374790 | 88421927 | 141 | 22 | 47293 | -1.4066 | 0.92022 | *AKIRIN2* |
| ENSG00000189007 | 6 | 143733969 | 143781810 | 137 | 8 | 53991 | -1.4067 | 0.92024 | *ADAT2* |
| ENSG00000137872 | 15 | 47466298 | 48076420 | 1932 | 141 | 42834 | -1.4067 | 0.92024 | *SEMA6D* |
| ENSG00000164211 | 5 | 110821731 | 110858288 | 75 | 23 | 47921 | -1.4077 | 0.92039 | *STARD4* |
| ENSG00000136856 | 9 | 130149421 | 130180703 | 70 | 10 | 54431 | -1.4079 | 0.92042 | *SLC2A8* |
| ENSG00000148604 | 10 | 85994809 | 86029716 | 131 | 26 | 40774 | -1.4083 | 0.92048 | *RGR* |
| ENSG00000204518 | 1 | 12694566 | 12737097 | 117 | 16 | 35234 | -1.4092 | 0.92062 | *AADACL4* |
| ENSG00000125571 | 2 | 113660548 | 113686459 | 124 | 6 | 48511 | -1.4096 | 0.92067 | *IL37* |
| ENSG00000242802 | 7 | 4805253 | 4843943 | 242 | 27 | 37022 | -1.4103 | 0.92078 | *AP5Z1* |
| ENSG00000197714 | 19 | 57781419 | 57814937 | 157 | 21 | 45314 | -1.4104 | 0.92079 | *ZNF460* |
| ENSG00000130309 | 19 | 17656403 | 17703971 | 144 | 29 | 49650 | -1.4106 | 0.92081 | *COLGALT1* |
| ENSG00000267022 | 19 | 44519506 | 44601471 | 177 | 16 | 27306 | -1.4109 | 0.92087 | *ZNF223* |
| ENSG00000182180 | 10 | 74996510 | 75022451 | 72 | 3 | 18368 | -1.4117 | 0.92098 | *MRPS16* |
| ENSG00000172992 | 17 | 43090708 | 43148473 | 201 | 23 | 53943 | -1.4119 | 0.92101 | *DCAKD* |
| ENSG00000262814 | 17 | 79660387 | 79684556 | 111 | 15 | 54721 | -1.412 | 0.92103 | *MRPL12* |
| ENSG00000101162 | 20 | 57584309 | 57611709 | 64 | 15 | 54874 | -1.4126 | 0.92112 | *TUBB1* |
| ENSG00000197430 | 10 | 98092973 | 98129092 | 107 | 18 | 41609 | -1.4134 | 0.92123 | *OPALIN* |
| ENSG00000112559 | 6 | 41594620 | 41631984 | 139 | 37 | 52181 | -1.4141 | 0.92133 | *MDFI* |
| ENSG00000185359 | 17 | 79640356 | 79680168 | 212 | 21 | 53288 | -1.4142 | 0.92135 | *HGS* |
| ENSG00000184281 | 11 | 2411718 | 2435106 | 51 | 29 | 37941 | -1.4144 | 0.92137 | *TSSC4* |
| ENSG00000162631 | 1 | 107672629 | 108036080 | 861 | 143 | 46007 | -1.4148 | 0.92144 | *NTNG1* |
| ENSG00000164252 | 5 | 76315076 | 76371059 | 169 | 25 | 50500 | -1.4151 | 0.92149 | *AGGF1* |
| ENSG00000214530 | 11 | 72455774 | 72514726 | 157 | 20 | 48131 | -1.4155 | 0.92154 | *STARD10* |
| ENSG00000010438 | 9 | 33740515 | 33809230 | 232 | 31 | 48534 | -1.4162 | 0.92164 | *PRSS3* |
| ENSG00000235531 | 8 | 72730402 | 73040628 | 1356 | 132 | 48113 | -1.4166 | 0.92169 | *RP11-383H13.1* |
| ENSG00000108932 | 17 | 66253167 | 66297408 | 122 | 23 | 51671 | -1.4169 | 0.92175 | *SLC16A6* |

| | | | | | | | | | |
|---|---|---|---|---|---|---|---|---|---|
| ENSG00000158158 | 2 | 97416639 | 97487628 | 111 | 16 | 38247 | -1.4172 | 0.92179 | *CNNM4* |
| ENSG00000170684 | 19 | 45564758 | 45589846 | 69 | 7 | 51131 | -1.4172 | 0.92179 | *ZNF296* |
| ENSG00000214700 | 12 | 27223991 | 27245447 | 86 | 15 | 38385 | -1.4174 | 0.92181 | *C12orf71* |
| ENSG00000186468 | 5 | 81559177 | 81584396 | 75 | 16 | 47358 | -1.4178 | 0.92187 | *RPS23* |
| ENSG00000188086 | 3 | 46773582 | 46864048 | 173 | 16 | 29217 | -1.4194 | 0.9221 | *PRSS45* |
| ENSG00000165323 | 11 | 92075262 | 92639618 | 1399 | 92 | 43510 | -1.4195 | 0.92212 | *FAT3* |
| ENSG00000140307 | 15 | 59920261 | 59959740 | 117 | 11 | 41712 | -1.4194 | 0.92212 | *GTF2A2* |
| ENSG00000171773 | 19 | 17556234 | 17581763 | 70 | 12 | 45671 | -1.4212 | 0.92238 | *NXNL1* |
| ENSG00000167641 | 19 | 38731877 | 38757231 | 109 | 16 | 52588 | -1.4215 | 0.92242 | *PPP1R14A* |
| ENSG00000109854 | 11 | 20375231 | 20415329 | 105 | 19 | 46006 | -1.4226 | 0.92257 | *HTATIP2* |
| ENSG00000145743 | 5 | 107184736 | 107727799 | 1429 | 92 | 45659 | -1.4232 | 0.92265 | *FBXL17* |
| ENSG00000075618 | 7 | 5622439 | 5656286 | 161 | 35 | 47300 | -1.4237 | 0.92273 | *FSCN1* |
| ENSG00000163568 | 1 | 159022274 | 159126886 | 177 | 37 | 39882 | -1.4247 | 0.92288 | *AIM2* |
| ENSG00000167759 | 19 | 51549463 | 51578371 | 85 | 28 | 46856 | -1.4247 | 0.92288 | *KLK13* |
| ENSG00000215021 | 12 | 7064490 | 7089988 | 38 | 12 | 39068 | -1.4258 | 0.92304 | *PHB2* |
| ENSG00000215252 | 15 | 34807408 | 34890704 | 156 | 19 | 48392 | -1.4265 | 0.92314 | *GOLGA8B* |
| ENSG00000006210 | 16 | 57396370 | 57428960 | 111 | 31 | 45347 | -1.427 | 0.92321 | *CX3CL1* |
| ENSG00000105948 | 7 | 138808490 | 138886732 | 60 | 16 | 38308 | -1.4272 | 0.92324 | *TTC26* |
| ENSG00000222018 | 21 | 35762615 | 35783370 | 87 | 13 | 44279 | -1.4275 | 0.92328 | *C21orf140* |
| ENSG00000156482 | 8 | 99027079 | 99068697 | 123 | 25 | 45590 | -1.4281 | 0.92336 | *RPL30* |
| ENSG00000102743 | 13 | 41353548 | 41394247 | 119 | 19 | 43936 | -1.4285 | 0.92343 | *SLC25A15* |
| ENSG00000159882 | 19 | 44497100 | 44528078 | 69 | 10 | 35480 | -1.43 | 0.92365 | *ZNF230* |
| ENSG00000105518 | 19 | 11443452 | 11467194 | 56 | 12 | 35939 | -1.4306 | 0.92372 | *TMEM205* |
| ENSG00000020577 | 14 | 55023815 | 55270033 | 701 | 144 | 46657 | -1.4306 | 0.92373 | *SAMD4A* |
| ENSG00000160183 | 21 | 43781999 | 43826955 | 250 | 59 | 43564 | -1.4307 | 0.92374 | *TMPRSS3* |
| ENSG00000203782 | 1 | 153222176 | 153244598 | 78 | 10 | 49457 | -1.4314 | 0.92384 | *LOR* |
| ENSG00000135318 | 6 | 86149809 | 86215500 | 136 | 15 | 41977 | -1.4327 | 0.92403 | *NT5E* |
| ENSG00000213079 | 6 | 155044459 | 155165192 | 283 | 27 | 43193 | -1.4333 | 0.92411 | *SCAF8* |
| ENSG00000168288 | 2 | 150416148 | 150454330 | 110 | 12 | 34282 | -1.4341 | 0.92423 | *MMADHC* |
| ENSG00000186283 | 1 | 179040512 | 179077158 | 122 | 23 | 54515 | -1.4346 | 0.9243 | *TOR3A* |
| ENSG00000185883 | 16 | 2553871 | 2580219 | 19 | 12 | 49908 | -1.4346 | 0.9243 | *ATP6V0C* |
| ENSG00000255804 | 14 | 23092673 | 23113716 | 131 | 7 | 52015 | -1.4353 | 0.9244 | *OR6J1* |
| ENSG00000186591 | 7 | 129460572 | 129602789 | 583 | 31 | 50907 | -1.4353 | 0.92441 | *UBE2H* |
| ENSG00000116786 | 1 | 16000827 | 16071264 | 216 | 19 | 38448 | -1.4358 | 0.92447 | *PLEKHM2* |
| ENSG00000168497 | 2 | 192689028 | 192721981 | 67 | 22 | 50572 | -1.4363 | 0.92454 | *SDPR* |
| ENSG00000131238 | 1 | 40528379 | 40573375 | 161 | 19 | 41408 | -1.4378 | 0.92476 | *PPT1* |

| | | | | | | | | | |
|---|---|---|---|---|---|---|---|---|---|
| ENSG00000185594 | 15 | 97316619 | 97338845 | 74 | 30 | 51147 | -1.4379 | 0.92477 | *SPATA8* |
| ENSG00000128604 | 7 | 128567666 | 128600089 | 113 | 21 | 49521 | -1.4407 | 0.92516 | *IRF5* |
| ENSG00000164338 | 5 | 72851268 | 72887794 | 105 | 12 | 41881 | -1.4425 | 0.92542 | *UTP15* |
| ENSG00000171262 | 15 | 38736328 | 38789911 | 216 | 22 | 53248 | -1.4425 | 0.92542 | *FAM98B* |
| ENSG00000101751 | 18 | 51785774 | 51857636 | 252 | 17 | 51652 | -1.4428 | 0.92545 | *POLI* |
| ENSG00000116962 | 1 | 236129130 | 236238462 | 294 | 44 | 39204 | -1.4429 | 0.92548 | *NID1* |
| ENSG00000121104 | 17 | 47777694 | 47876542 | 109 | 32 | 44785 | -1.4431 | 0.9255 | *FAM117A* |
| ENSG00000007216 | 17 | 26790311 | 26834799 | 109 | 15 | 50806 | -1.4432 | 0.92552 | *SLC13A2* |
| ENSG00000144214 | 2 | 99890701 | 99931205 | 135 | 17 | 48936 | -1.4452 | 0.92579 | *LYG1* |
| ENSG00000198054 | 21 | 39483545 | 39570110 | 213 | 27 | 33834 | -1.4452 | 0.9258 | *DSCR8* |
| ENSG00000075643 | 18 | 33757482 | 33862120 | 369 | 56 | 50220 | -1.4456 | 0.92585 | *MOCOS* |
| ENSG00000178386 | 19 | 44545520 | 44582144 | 76 | 10 | 28184 | -1.4465 | 0.92599 | *ZNF223* |
| ENSG00000105953 | 7 | 44636171 | 44758665 | 212 | 32 | 46799 | -1.4477 | 0.92615 | *OGDH* |
| ENSG00000060688 | 1 | 31722417 | 31779662 | 176 | 15 | 28869 | -1.448 | 0.92619 | *SNRNP40* |
| ENSG00000206069 | 22 | 25321077 | 25352662 | 59 | 14 | 46925 | -1.4482 | 0.92622 | *TMEM211* |
| ENSG00000116906 | 1 | 231366953 | 231423719 | 152 | 20 | 53171 | -1.4491 | 0.92634 | *GNPAT* |
| ENSG00000187821 | 4 | 185929995 | 185951958 | 106 | 27 | 49102 | -1.4491 | 0.92635 | *HELT* |
| ENSG00000161203 | 3 | 183882477 | 183911879 | 68 | 16 | 52150 | -1.4504 | 0.92653 | *AP2M1* |
| ENSG00000116213 | 1 | 3537331 | 3579325 | 135 | 28 | 41497 | -1.4507 | 0.92656 | *WRAP73* |
| ENSG00000103319 | 16 | 22207603 | 22308554 | 210 | 35 | 40712 | -1.4506 | 0.92656 | *EEF2K* |
| ENSG00000159231 | 21 | 37497210 | 37528864 | 140 | 14 | 49993 | -1.4513 | 0.92665 | *CBR3* |
| ENSG00000110330 | 11 | 102207942 | 102259401 | 125 | 17 | 54896 | -1.4516 | 0.92669 | *BIRC2* |
| ENSG00000079385 | 19 | 43001456 | 43075386 | 64 | 10 | 39768 | -1.4524 | 0.9268 | *CEACAM1* |
| ENSG00000185271 | 14 | 20886970 | 20913801 | 144 | 36 | 50581 | -1.4526 | 0.92683 | *KLHL33* |
| ENSG00000151079 | 12 | 4908342 | 4970277 | 303 | 38 | 45949 | -1.4554 | 0.92723 | *KCNA6* |
| ENSG00000183145 | 21 | 38368450 | 38401959 | 132 | 18 | 53227 | -1.4555 | 0.92723 | *RIPPLY3* |
| ENSG00000182405 | 15 | 34384274 | 34406591 | 35 | 8 | 33628 | -1.4561 | 0.92731 | *PGBD4* |
| ENSG00000154760 | 17 | 33752115 | 33785856 | 131 | 29 | 32060 | -1.4567 | 0.92741 | *SLFN13* |
| ENSG00000104980 | 19 | 7981603 | 8018805 | 129 | 16 | 53328 | -1.4577 | 0.92754 | *TIMM44* |
| ENSG00000205147 | 19 | 4031127 | 4053154 | 71 | 15 | 38435 | -1.458 | 0.92758 | *AC016586.1* |
| ENSG00000152926 | 7 | 64422154 | 64477062 | 224 | 20 | 44049 | -1.4583 | 0.92762 | *ZNF117* |
| ENSG00000197616 | 14 | 23841199 | 23887486 | 141 | 31 | 41559 | -1.4599 | 0.92784 | *MYH6* |
| ENSG00000176358 | 17 | 47905671 | 47935379 | 32 | 11 | 48509 | -1.46 | 0.92786 | *TAC4* |
| ENSG00000112118 | 6 | 52118807 | 52159679 | 135 | 27 | 47650 | -1.4609 | 0.92798 | *MCM3* |
| ENSG00000129744 | 11 | 3656358 | 3695646 | 195 | 34 | 36147 | -1.4616 | 0.92807 | *ART1* |
| ENSG00000259784 | 16 | 2553966 | 2587956 | 26 | 17 | 51298 | -1.4621 | 0.92815 | *ATP6C* |

| | | | | | | | | | |
|---|---|---|---|---|---|---|---|---|---|
| ENSG00000108828 | 17 | 41156622 | 41187140 | 49 | 12 | 51230 | -1.4622 | 0.92816 | *VAT1* |
| ENSG00000222046 | 1 | 32664695 | 32691797 | 43 | 10 | 33492 | -1.4635 | 0.92833 | *DCDC2B* |
| ENSG00000273173 | 15 | 25190133 | 25233729 | 95 | 18 | 42485 | -1.4635 | 0.92833 | *SNURF* |
| ENSG00000111241 | 12 | 4527321 | 4564780 | 116 | 19 | 42168 | -1.464 | 0.9284 | *FGF6* |
| ENSG00000204237 | 17 | 79622066 | 79643665 | 160 | 14 | 37674 | -1.464 | 0.9284 | *OXLD1* |
| ENSG00000196636 | 7 | 96735902 | 96821075 | 187 | 19 | 54977 | -1.4655 | 0.92861 | *ACN9* |
| ENSG00000244588 | 20 | 1196700 | 1287065 | 296 | 45 | 47528 | -1.4657 | 0.92864 | *RAD21L1* |
| ENSG00000130783 | 12 | 123248874 | 123322075 | 260 | 22 | 24955 | -1.466 | 0.92868 | *CCDC62* |
| ENSG00000049323 | 2 | 33162039 | 33634576 | 2223 | 177 | 47556 | -1.4667 | 0.92878 | *LTBP1* |
| ENSG00000163820 | 3 | 45949396 | 46047316 | 292 | 27 | 29302 | -1.467 | 0.92881 | *FYCO1* |
| ENSG00000132792 | 20 | 36312408 | 36510531 | 388 | 35 | 50884 | -1.4671 | 0.92883 | *CTNNBL1* |
| ENSG00000133055 | 1 | 203126939 | 203154941 | 98 | 21 | 35278 | -1.4678 | 0.92892 | *MYBPH* |
| ENSG00000117598 | 1 | 99345806 | 99480588 | 539 | 69 | 54557 | -1.4683 | 0.92899 | *LPPR5* |
| ENSG00000267893 | 17 | 70026164 | 70046872 | 87 | 11 | 47802 | -1.4692 | 0.92912 | *AC007461.1* |
| ENSG00000197056 | 1 | 35515387 | 35591460 | 69 | 18 | 35987 | -1.4704 | 0.92927 | *ZMYM1* |
| ENSG00000138757 | 4 | 76557966 | 76659709 | 287 | 26 | 42281 | -1.4708 | 0.92933 | *G3BP2* |
| ENSG00000120948 | 1 | 11062414 | 11095796 | 87 | 13 | 38249 | -1.4709 | 0.92934 | *TARDBP* |
| ENSG00000188234 | 10 | 46311042 | 46359323 | 15 | 10 | 19659 | -1.4709 | 0.92934 | *AGAP4* |
| ENSG00000176605 | 14 | 99167950 | 99194098 | 125 | 22 | 46094 | -1.4711 | 0.92937 | *C14orf177* |
| ENSG00000172689 | 11 | 60542821 | 60578778 | 96 | 21 | 40346 | -1.4712 | 0.92938 | *MS4A10* |
| ENSG00000168081 | 8 | 28164503 | 28210872 | 172 | 18 | 29104 | -1.4722 | 0.92952 | *PNOC* |
| ENSG00000136213 | 7 | 2433223 | 2484242 | 131 | 24 | 51646 | -1.4723 | 0.92953 | *CHST12* |
| ENSG00000189181 | 1 | 248834628 | 248855629 | 84 | 8 | 38624 | -1.4728 | 0.9296 | *OR14I1* |
| ENSG00000140025 | 14 | 90251013 | 90431121 | 542 | 60 | 32428 | -1.4736 | 0.9297 | *EFCAB11* |
| ENSG00000138079 | 2 | 44492599 | 44558633 | 175 | 22 | 41620 | -1.4739 | 0.92974 | *SLC3A1* |
| ENSG00000010072 | 1 | 231462850 | 231500769 | 100 | 9 | 41378 | -1.4741 | 0.92977 | *SPRTN* |
| ENSG00000165527 | 14 | 50349810 | 50371490 | 30 | 9 | 51095 | -1.4741 | 0.92977 | *ARF6* |
| ENSG00000174111 | 17 | 36498111 | 36566019 | 142 | 18 | 44138 | -1.4744 | 0.92981 | *SOCS7* |
| ENSG00000127947 | 7 | 77156592 | 77279388 | 394 | 30 | 43712 | -1.4746 | 0.92984 | *PTPN12* |
| ENSG00000003400 | 2 | 202037604 | 202104129 | 131 | 19 | 43795 | -1.4756 | 0.92998 | *CASP10* |
| ENSG00000204880 | 17 | 39243233 | 39264393 | 212 | 11 | 39596 | -1.4757 | 0.92998 | *KRTAP4-8* |
| ENSG00000132478 | 17 | 73770681 | 73831886 | 137 | 22 | 35117 | -1.4762 | 0.93006 | *UNK* |
| ENSG00000164976 | 9 | 34356668 | 34386851 | 62 | 13 | 42416 | -1.4764 | 0.93009 | *KIAA1161* |
| ENSG00000123096 | 12 | 26264924 | 26462223 | 687 | 96 | 41341 | -1.4771 | 0.93018 | *SSPN* |
| ENSG00000188032 | 19 | 14182431 | 14206687 | 35 | 14 | 47512 | -1.4773 | 0.93021 | *C19orf67* |
| ENSG00000127515 | 19 | 14941760 | 14962689 | 117 | 28 | 45673 | -1.4777 | 0.93025 | *OR7A10* |

| | | | | | | | | | |
|---|---|---|---|---|---|---|---|---|---|
| ENSG00000143575 | 1 | 154234987 | 154258351 | 41 | 10 | 51268 | -1.4778 | 0.93026 | *HAX1* |
| ENSG00000086189 | 5 | 61673081 | 61709766 | 96 | 16 | 50578 | -1.4787 | 0.93039 | *DIMT1* |
| ENSG00000196196 | 9 | 35896189 | 35917138 | 60 | 18 | 51506 | -1.4787 | 0.93039 | *HRCT1* |
| ENSG00000163682 | 4 | 39445744 | 39470568 | 106 | 17 | 50721 | -1.4793 | 0.93047 | *RPL9* |
| ENSG00000268423 | 19 | 47040313 | 47114118 | 233 | 46 | 49019 | -1.4802 | 0.93059 | *AC011551.3* |
| ENSG00000196431 | 22 | 27007928 | 27036636 | 112 | 17 | 49000 | -1.4804 | 0.93061 | *CRYBA4* |
| ENSG00000109771 | 4 | 186275032 | 186327053 | 158 | 28 | 42340 | -1.4806 | 0.93065 | *LRP2BP* |
| ENSG00000162601 | 1 | 59110411 | 59175764 | 305 | 20 | 46537 | -1.4807 | 0.93066 | *MYSM1* |
| ENSG00000125503 | 19 | 55592281 | 55638927 | 107 | 31 | 46517 | -1.4813 | 0.93074 | *PPP1R12C* |
| ENSG00000145888 | 5 | 151192074 | 151314403 | 297 | 23 | 49027 | -1.4821 | 0.93085 | *GLRA1* |
| ENSG00000164647 | 7 | 89773689 | 89804143 | 85 | 14 | 43004 | -1.4822 | 0.93085 | *STEAP1* |
| ENSG00000178896 | 8 | 145123529 | 145145550 | 36 | 5 | 31141 | -1.4822 | 0.93086 | *EXOSC4* |
| ENSG00000179817 | 11 | 18184384 | 18205827 | 106 | 7 | 50492 | -1.4838 | 0.93107 | *MRGPRX4* |
| ENSG00000104936 | 19 | 46262975 | 46295810 | 57 | 14 | 52317 | -1.4839 | 0.93109 | *DMPK* |
| ENSG00000205643 | 22 | 46629908 | 46656576 | 105 | 13 | 35983 | -1.4848 | 0.93121 | *CDPF1* |
| ENSG00000129083 | 11 | 14454986 | 14531573 | 178 | 17 | 53542 | -1.4865 | 0.93143 | *COPB1* |
| ENSG00000198435 | 9 | 140184083 | 140206703 | 40 | 15 | 38081 | -1.4874 | 0.93155 | *NRARP* |
| ENSG00000126461 | 19 | 50135382 | 50171899 | 65 | 16 | 36491 | -1.4887 | 0.93172 | *SCAF1* |
| ENSG00000006074 | 17 | 34381640 | 34409392 | 94 | 18 | 45360 | -1.4891 | 0.93177 | *CCL18* |
| ENSG00000140519 | 15 | 89988680 | 90049844 | 113 | 18 | 47661 | -1.4899 | 0.93187 | *RHCG* |
| ENSG00000160221 | 21 | 45543487 | 45575605 | 39 | 15 | 41130 | -1.4902 | 0.93191 | *C21orf33* |
| ENSG00000167851 | 17 | 72452555 | 72490933 | 204 | 36 | 34674 | -1.4903 | 0.93193 | *CD300A* |
| ENSG00000163945 | 4 | 1331054 | 1391837 | 223 | 29 | 36737 | -1.4905 | 0.93196 | *UVSSA* |
| ENSG00000135945 | 2 | 100006938 | 100116497 | 296 | 21 | 51858 | -1.4919 | 0.93213 | *REV1* |
| ENSG00000136206 | 7 | 44030488 | 44059721 | 68 | 12 | 50762 | -1.492 | 0.93215 | *SPDYE1* |
| ENSG00000072952 | 11 | 10584638 | 10725535 | 530 | 70 | 43668 | -1.4926 | 0.93223 | *MRVI1* |
| ENSG00000136011 | 12 | 103971051 | 104170505 | 781 | 122 | 44659 | -1.493 | 0.93229 | *STAB2* |
| ENSG00000165120 | 7 | 129837700 | 129866683 | 88 | 18 | 41515 | -1.4937 | 0.93237 | *SSMEM1* |
| ENSG00000184900 | 21 | 46215532 | 46248694 | 131 | 25 | 42928 | -1.4939 | 0.93239 | *SUMO3* |
| ENSG00000102804 | 13 | 44997655 | 45161283 | 391 | 29 | 53615 | -1.4947 | 0.93251 | *TSC22D1* |
| ENSG00000172348 | 6 | 46178475 | 46469709 | 657 | 59 | 36680 | -1.4962 | 0.9327 | *RCAN2* |
| ENSG00000132341 | 12 | 131346424 | 131372223 | 99 | 15 | 53542 | -1.4965 | 0.93274 | *RAN* |
| ENSG00000151835 | 13 | 23892965 | 24017841 | 466 | 68 | 49190 | -1.4977 | 0.93289 | *SACS* |
| ENSG00000184182 | 2 | 238865469 | 238961236 | 305 | 20 | 49488 | -1.4984 | 0.93299 | *UBE2F* |
| ENSG00000118434 | 6 | 88747507 | 88786550 | 65 | 9 | 48710 | -1.4988 | 0.93304 | *SPACA1* |
| ENSG00000185298 | 17 | 79623394 | 79650934 | 189 | 15 | 40341 | -1.5003 | 0.93324 | *CCDC137* |

| | | | | | | | | | |
|---|---|---|---|---|---|---|---|---|---|
| ENSG00000197591 | 1 | 247994199 | 248015254 | 113 | 14 | 33555 | -1.5009 | 0.93331 | *OR11L1* |
| ENSG00000020129 | 1 | 36013074 | 36042875 | 14 | 7 | 46781 | -1.5011 | 0.93333 | *NCDN* |
| ENSG00000162366 | 1 | 47639265 | 47666716 | 50 | 18 | 40764 | -1.5015 | 0.93339 | *PDZK1IP1* |
| ENSG00000164334 | 5 | 118955254 | 118981517 | 137 | 9 | 53552 | -1.5016 | 0.9334 | *FAM170A* |
| ENSG00000104970 | 19 | 55033909 | 55067053 | 149 | 34 | 49116 | -1.5016 | 0.9334 | *KIR3DX1* |
| ENSG00000114013 | 3 | 121764213 | 121849983 | 206 | 45 | 46281 | -1.5019 | 0.93344 | *CD86* |
| ENSG00000173212 | 1 | 116644376 | 116687861 | 138 | 24 | 40257 | -1.5021 | 0.93346 | *MAB21L3* |
| ENSG00000120800 | 12 | 101663887 | 101790394 | 281 | 35 | 46206 | -1.5028 | 0.93355 | *UTP20* |
| ENSG00000164181 | 5 | 60037618 | 60150216 | 303 | 34 | 41817 | -1.5028 | 0.93356 | *ELOVL7* |
| ENSG00000197024 | 7 | 148813508 | 148890116 | 261 | 20 | 39024 | -1.5037 | 0.93367 | *ZNF398* |
| ENSG00000242114 | 22 | 30811518 | 30835045 | 58 | 14 | 43674 | -1.5047 | 0.9338 | *MTFP1* |
| ENSG00000166016 | 11 | 34162535 | 34389555 | 742 | 133 | 43171 | -1.5051 | 0.93386 | *ABTB2* |
| ENSG00000160051 | 1 | 32661236 | 32684288 | 38 | 12 | 38523 | -1.5058 | 0.93394 | *IQCC* |
| ENSG00000145734 | 5 | 70741442 | 70873649 | 361 | 18 | 54184 | -1.506 | 0.93397 | *BDP1* |
| ENSG00000142661 | 1 | 24372525 | 24448665 | 294 | 40 | 44076 | -1.5069 | 0.93408 | *MYOM3* |
| ENSG00000100485 | 14 | 50573847 | 50708276 | 477 | 37 | 47930 | -1.5071 | 0.9341 | *SOS2* |
| ENSG00000180525 | 10 | 685888 | 721109 | 55 | 14 | 40496 | -1.5088 | 0.93433 | *PRR26* |
| ENSG00000136859 | 9 | 129839611 | 129895162 | 86 | 16 | 44424 | -1.5098 | 0.93445 | *ANGPTL2* |
| ENSG00000250709 | 13 | 36732931 | 36881979 | 657 | 25 | 54933 | -1.511 | 0.93461 | *CCDC169-SOHLH2* |
| ENSG00000206549 | 3 | 46743605 | 46864064 | 275 | 26 | 32601 | -1.5112 | 0.93463 | *PRSS50* |
| ENSG00000251209 | 15 | 98275448 | 98427780 | 700 | 81 | 47039 | -1.5132 | 0.93489 | *LINC00923* |
| ENSG00000103051 | 16 | 70504471 | 70567468 | 100 | 18 | 37388 | -1.514 | 0.93499 | *COG4* |
| ENSG00000135443 | 12 | 52743790 | 52771265 | 113 | 20 | 46209 | -1.5154 | 0.93516 | *KRT85* |
| ENSG00000148344 | 9 | 132490610 | 132525326 | 163 | 28 | 42555 | -1.5159 | 0.93522 | *PTGES* |
| ENSG00000176170 | 17 | 74362665 | 74393941 | 96 | 22 | 36512 | -1.5161 | 0.93525 | *SPHK1* |
| ENSG00000147548 | 8 | 38117215 | 38249790 | 206 | 16 | 54514 | -1.5163 | 0.93528 | *WHSC1L1* |
| ENSG00000269948 | 10 | 97699725 | 97754785 | 182 | 29 | 42068 | -1.5166 | 0.93532 | *RP11-248J23.6* |
| ENSG00000122696 | 9 | 37869400 | 37914350 | 93 | 18 | 53165 | -1.5171 | 0.93538 | *SLC25A51* |
| ENSG00000170608 | 19 | 46357247 | 46387055 | 102 | 22 | 49823 | -1.5173 | 0.9354 | *FOXA3* |
| ENSG00000164418 | 6 | 101836664 | 102527958 | 2365 | 217 | 42778 | -1.5174 | 0.93542 | *GRIK2* |
| ENSG00000162670 | 1 | 190056792 | 190456759 | 866 | 68 | 41759 | -1.5177 | 0.93545 | *BRINP3* |
| ENSG00000230778 | 15 | 40563645 | 40584787 | 32 | 13 | 43824 | -1.5187 | 0.93558 | *ANKRD63* |
| ENSG00000177669 | 8 | 29979340 | 30012202 | 74 | 10 | 32765 | -1.5191 | 0.93563 | *MBOAT4* |
| ENSG00000122359 | 10 | 81900645 | 81975328 | 321 | 34 | 47136 | -1.5191 | 0.93563 | *ANXA11* |
| ENSG00000131845 | 19 | 57852675 | 57881266 | 108 | 21 | 53106 | -1.5197 | 0.9357 | *ZNF304* |
| ENSG00000119977 | 10 | 97413158 | 97463900 | 147 | 21 | 39882 | -1.5214 | 0.93592 | *TCTN3* |

| | | | | | | | | | |
|---|---|---|---|---|---|---|---|---|---|
| ENSG00000182256 | 15 | 27206429 | 27788373 | 1818 | 221 | 39188 | -1.5219 | 0.93598 | *GABRG3* |
| ENSG00000188959 | 9 | 112942328 | 112980469 | 154 | 27 | 43373 | -1.5221 | 0.936 | *C9orf152* |
| ENSG00000175202 | 15 | 72958124 | 72988490 | 48 | 17 | 42905 | -1.5221 | 0.93601 | *HIGD2B* |
| ENSG00000267819 | 12 | 131770067 | 131792763 | 50 | 15 | 19647 | -1.523 | 0.93612 | *AC092850.1* |
| ENSG00000120669 | 13 | 36732345 | 36881979 | 659 | 25 | 54939 | -1.5231 | 0.93614 | *SOHLH2* |
| ENSG00000168135 | 22 | 38812332 | 38861205 | 155 | 28 | 38044 | -1.5234 | 0.93617 | *KCNJ4* |
| ENSG00000141971 | 19 | 17506531 | 17554533 | 144 | 28 | 38392 | -1.5239 | 0.93624 | *MVB12A* |
| ENSG00000112619 | 6 | 42654340 | 42700312 | 167 | 19 | 54053 | -1.5241 | 0.93626 | *PRPH2* |
| ENSG00000009780 | 1 | 28042490 | 28099633 | 32 | 19 | 46604 | -1.5248 | 0.93635 | *FAM76A* |
| ENSG00000185477 | 4 | 90147537 | 90239161 | 247 | 44 | 41589 | -1.5256 | 0.93644 | *GPRIN3* |
| ENSG00000213983 | 14 | 24018774 | 24047279 | 38 | 12 | 43705 | -1.5264 | 0.93655 | *AP1G2* |
| ENSG00000136888 | 9 | 117340026 | 117370653 | 131 | 21 | 45878 | -1.527 | 0.93662 | *ATP6V1G1* |
| ENSG00000130707 | 9 | 133310316 | 133386661 | 217 | 48 | 45188 | -1.528 | 0.93674 | *ASS1* |
| ENSG00000133872 | 8 | 29910528 | 29950723 | 105 | 15 | 42014 | -1.5288 | 0.93685 | *TMEM66* |
| ENSG00000142168 | 21 | 33021935 | 33051244 | 62 | 20 | 49816 | -1.529 | 0.93687 | *SOD1* |
| ENSG00000188716 | 10 | 76787594 | 76828272 | 93 | 18 | 23057 | -1.5291 | 0.93688 | *DUPD1* |
| ENSG00000087299 | 14 | 50694281 | 50789266 | 261 | 21 | 48212 | -1.5307 | 0.93708 | *L2HGDH* |
| ENSG00000082684 | 3 | 122618041 | 122757452 | 457 | 73 | 40807 | -1.5309 | 0.9371 | *SEMA5B* |
| ENSG00000083290 | 17 | 19664142 | 19781249 | 162 | 20 | 35554 | -1.5329 | 0.93734 | *ULK2* |
| ENSG00000180822 | 6 | 3221637 | 3313607 | 437 | 34 | 34606 | -1.533 | 0.93736 | *PSMG4* |
| ENSG00000240230 | 7 | 928415 | 1025235 | 475 | 92 | 45096 | -1.5332 | 0.93738 | *COX19* |
| ENSG00000261147 | 15 | 90782788 | 90825440 | 140 | 13 | 32209 | -1.5334 | 0.93741 | *RP11-697E2.6* |
| ENSG00000128641 | 2 | 192099911 | 192300115 | 368 | 23 | 49440 | -1.5339 | 0.93748 | *MYO1B* |
| ENSG00000111300 | 12 | 112454500 | 112556826 | 103 | 24 | 42376 | -1.5347 | 0.93757 | *NAA25* |
| ENSG00000106723 | 9 | 90993334 | 91103609 | 245 | 12 | 27674 | -1.5355 | 0.93767 | *SPIN1* |
| ENSG00000119574 | 19 | 59014897 | 59060278 | 80 | 19 | 43376 | -1.5358 | 0.93771 | *ZBTB45* |
| ENSG00000164074 | 4 | 128876461 | 128970866 | 88 | 26 | 43145 | -1.5364 | 0.93778 | *C4orf29* |
| ENSG00000185352 | 13 | 96733093 | 97495671 | 2006 | 145 | 46127 | -1.5364 | 0.93778 | *HS6ST3* |
| ENSG00000137440 | 4 | 15927192 | 15950363 | 78 | 19 | 48896 | -1.5367 | 0.93782 | *FGFBP1* |
| ENSG00000113312 | 5 | 159426120 | 159502550 | 202 | 21 | 54261 | -1.5374 | 0.93791 | *TTC1* |
| ENSG00000137760 | 11 | 107363452 | 107446472 | 292 | 33 | 40518 | -1.5379 | 0.93797 | *ALKBH8* |
| ENSG00000204947 | 7 | 148789876 | 148833438 | 132 | 13 | 35498 | -1.5387 | 0.93806 | *ZNF425* |
| ENSG00000176299 | 14 | 20238401 | 20259516 | 7 | 4 | 42851 | -1.5402 | 0.93824 | *OR4M1* |
| ENSG00000203970 | 6 | 49966851 | 49999694 | 74 | 11 | 39913 | -1.5404 | 0.93827 | *DEFB110* |
| ENSG00000134874 | 13 | 96220457 | 96306957 | 172 | 32 | 45962 | -1.5407 | 0.93831 | *DZIP1* |
| ENSG00000144228 | 2 | 139249371 | 139341117 | 158 | 24 | 40996 | -1.541 | 0.93834 | *SPOPL* |

| | | | | | | | | | |
|---|---|---|---|---|---|---|---|---|---|
| ENSG00000185523 | 1 | 212993483 | 213030991 | 213 | 16 | 49175 | -1.542 | 0.93847 | *C1orf227* |
| ENSG00000152910 | 16 | 76301176 | 76603135 | 1317 | 89 | 37120 | -1.5424 | 0.93851 | *CNTNAP4* |
| ENSG00000137077 | 9 | 34699002 | 34720121 | 70 | 17 | 34751 | -1.5426 | 0.93853 | *CCL21* |
| ENSG00000130787 | 12 | 123309000 | 123357507 | 177 | 16 | 32103 | -1.5428 | 0.93855 | *HIP1R* |
| ENSG00000170571 | 5 | 49682026 | 49749082 | 153 | 12 | 54659 | -1.5436 | 0.93866 | *EMB* |
| ENSG00000164978 | 9 | 34319504 | 34353709 | 89 | 16 | 36869 | -1.5436 | 0.93866 | *NUDT2* |
| ENSG00000170374 | 12 | 53710362 | 53749099 | 67 | 17 | 43030 | -1.5446 | 0.93878 | *SP7* |
| ENSG00000077498 | 11 | 88900620 | 89038927 | 394 | 29 | 26592 | -1.546 | 0.93894 | *TYR* |
| ENSG00000198854 | 1 | 152681998 | 152702905 | 89 | 5 | 48162 | -1.5461 | 0.93896 | *C1orf68* |
| ENSG00000139505 | 13 | 25792307 | 25872147 | 166 | 24 | 48867 | -1.5486 | 0.93927 | *MTMR6* |
| ENSG00000101489 | 18 | 34813010 | 35156000 | 906 | 159 | 45588 | -1.549 | 0.93931 | *CELF4* |
| ENSG00000144648 | 3 | 42836244 | 42939466 | 277 | 34 | 43495 | -1.5503 | 0.93947 | *ACKR2* |
| ENSG00000173578 | 3 | 46048516 | 46079234 | 113 | 12 | 28172 | -1.5521 | 0.93968 | *XCR1* |
| ENSG00000240747 | 3 | 42840975 | 42994284 | 355 | 44 | 43299 | -1.5524 | 0.93972 | *KRBOX1* |
| ENSG00000179846 | 19 | 45643008 | 45673408 | 73 | 21 | 52458 | -1.5531 | 0.9398 | *NKPD1* |
| ENSG00000137561 | 8 | 63951112 | 64008612 | 205 | 25 | 38335 | -1.5536 | 0.93986 | *TTPA* |
| ENSG00000257704 | 19 | 47768142 | 47788979 | 76 | 13 | 46372 | -1.5542 | 0.93993 | *PRR24* |
| ENSG00000213471 | 15 | 90782762 | 90818199 | 119 | 11 | 30110 | -1.5551 | 0.94004 | *TTLL13* |
| ENSG00000152595 | 4 | 88732563 | 88777969 | 224 | 35 | 48837 | -1.5554 | 0.94007 | *MEPE* |
| ENSG00000165059 | 9 | 71617469 | 71639039 | 82 | 19 | 46272 | -1.5568 | 0.94024 | *PRKACG* |
| ENSG00000132330 | 2 | 238959530 | 239018054 | 251 | 26 | 44363 | -1.5579 | 0.94037 | *SCLY* |
| ENSG00000177733 | 5 | 137077075 | 137100039 | 25 | 10 | 49282 | -1.5581 | 0.9404 | *HNRNPA0* |
| ENSG00000178828 | 1 | 20130523 | 20151771 | 113 | 29 | 43733 | -1.559 | 0.9405 | *RNF186* |
| ENSG00000234511 | 5 | 169649451 | 169689579 | 64 | 19 | 51938 | -1.559 | 0.9405 | *C5orf58* |
| ENSG00000249459 | 17 | 18551742 | 18595575 | 124 | 18 | 43766 | -1.5612 | 0.94076 | *ZNF286B* |
| ENSG00000241644 | 7 | 30727601 | 30807218 | 270 | 56 | 43014 | -1.5613 | 0.94077 | *INMT* |
| ENSG00000117475 | 1 | 169327208 | 169375778 | 164 | 19 | 44219 | -1.5631 | 0.94099 | *BLZF1* |
| ENSG00000099625 | 19 | 1218286 | 1248026 | 92 | 18 | 50764 | -1.5636 | 0.94104 | *C19orf26* |
| ENSG00000064692 | 5 | 121637049 | 121809914 | 292 | 35 | 52314 | -1.5638 | 0.94106 | *SNCAIP* |
| ENSG00000235162 | 12 | 105619068 | 105799875 | 695 | 72 | 50108 | -1.5644 | 0.94113 | *C12orf75* |
| ENSG00000182035 | 20 | 37199838 | 37227106 | 70 | 16 | 30625 | -1.5652 | 0.94123 | *ADIG* |
| ENSG00000198477 | 22 | 22828767 | 22873505 | 149 | 18 | 34904 | -1.5655 | 0.94127 | *ZNF280B* |
| ENSG00000183273 | 12 | 119762517 | 119988852 | 658 | 101 | 47415 | -1.5678 | 0.94153 | *CCDC60* |
| ENSG00000121417 | 19 | 58131761 | 58164147 | 155 | 10 | 48459 | -1.5683 | 0.94159 | *ZNF211* |
| ENSG00000060656 | 1 | 29553028 | 29663325 | 295 | 49 | 38930 | -1.5701 | 0.9418 | *PTPRU* |
| ENSG00000172752 | 3 | 130054359 | 130213688 | 538 | 60 | 41873 | -1.5729 | 0.94213 | *COL6A5* |

| | | | | | | | | | |
|---|---|---|---|---|---|---|---|---|---|
| ENSG00000082482 | 1 | 215169118 | 215420436 | 851 | 70 | 31658 | -1.5752 | 0.94239 | *KCNK2* |
| ENSG00000121743 | 13 | 20702394 | 20745188 | 162 | 29 | 34161 | -1.5752 | 0.9424 | *GJA3* |
| ENSG00000122497 | 1 | 147993642 | 148035863 | 5 | 3 | 42848 | -1.5757 | 0.94245 | *NBPF14* |
| ENSG00000183034 | 17 | 72910370 | 72940007 | 56 | 19 | 38537 | -1.5768 | 0.94258 | *OTOP2* |
| ENSG00000101463 | 20 | 24439835 | 24657252 | 624 | 79 | 48065 | -1.5788 | 0.94281 | *SYNDIG1* |
| ENSG00000135441 | 12 | 56099820 | 56123871 | 40 | 15 | 34579 | -1.579 | 0.94284 | *BLOC1S1* |
| ENSG00000127334 | 12 | 68032118 | 68069186 | 82 | 14 | 54457 | -1.5792 | 0.94286 | *DYRK2* |
| ENSG00000152315 | 14 | 90518109 | 90662201 | 582 | 71 | 38976 | -1.5807 | 0.94303 | *KCNK13* |
| ENSG00000269700 | 10 | 52142766 | 52164321 | 68 | 14 | 44097 | -1.5811 | 0.94307 | *AC069547.2* |
| ENSG00000120699 | 13 | 37562953 | 37593750 | 40 | 9 | 42804 | -1.5813 | 0.9431 | *EXOSC8* |
| ENSG00000163913 | 3 | 129148968 | 129249198 | 243 | 18 | 34527 | -1.5833 | 0.94333 | *IFT122* |
| ENSG00000196460 | 2 | 102003823 | 102101165 | 415 | 29 | 39769 | -1.5835 | 0.94335 | *RFX8* |
| ENSG00000132142 | 17 | 35431923 | 35776909 | 587 | 60 | 35395 | -1.5841 | 0.94341 | *ACACA* |
| ENSG00000185070 | 14 | 85986488 | 86105034 | 424 | 64 | 47429 | -1.5845 | 0.94346 | *FLRT2* |
| ENSG00000172123 | 17 | 33728079 | 33770302 | 148 | 28 | 26578 | -1.585 | 0.94352 | *SLFN12* |
| ENSG00000145214 | 4 | 942675 | 990683 | 155 | 30 | 49758 | -1.5852 | 0.94354 | *DGKQ* |
| ENSG00000172273 | 11 | 118982297 | 119016752 | 94 | 10 | 23660 | -1.5852 | 0.94354 | *HINFP* |
| ENSG00000205403 | 4 | 110651852 | 110733335 | 201 | 25 | 46348 | -1.5856 | 0.94358 | *CFI* |
| ENSG00000151693 | 2 | 9336894 | 9555812 | 501 | 52 | 44515 | -1.5862 | 0.94366 | *ASAP2* |
| ENSG00000112695 | 6 | 75937391 | 75970039 | 69 | 9 | 24182 | -1.5877 | 0.94383 | *COX7A2* |
| ENSG00000168260 | 14 | 50540369 | 50569361 | 105 | 18 | 49926 | -1.588 | 0.94386 | *C14orf183* |
| ENSG00000267496 | 17 | 41984576 | 42005326 | 88 | 13 | 37778 | -1.5883 | 0.94389 | *FAM215A* |
| ENSG00000203836 | 1 | 147564428 | 147644886 | 3 | 2 | 12600 | -1.5887 | 0.94394 | *NBPF24* |
| ENSG00000175093 | 3 | 140760244 | 140877453 | 246 | 37 | 39753 | -1.5893 | 0.944 | *SPSB4* |
| ENSG00000114030 | 3 | 122130796 | 122243792 | 328 | 29 | 51223 | -1.5905 | 0.94414 | *KPNA1* |
| ENSG00000205445 | 21 | 45960240 | 45981388 | 70 | 12 | 50596 | -1.5909 | 0.94419 | *KRTAP10-2* |
| ENSG00000144283 | 2 | 159303476 | 159549391 | 851 | 53 | 39194 | -1.5917 | 0.94428 | *PKP4* |
| ENSG00000179913 | 19 | 17895637 | 17933891 | 127 | 31 | 48838 | -1.592 | 0.9443 | *B3GNT3* |
| ENSG00000241794 | 1 | 153018589 | 153040013 | 73 | 11 | 28858 | -1.593 | 0.94441 | *SPRR2A* |
| ENSG00000137073 | 9 | 33911691 | 34058947 | 518 | 32 | 47779 | -1.5946 | 0.9446 | *UBAP2* |
| ENSG00000205441 | 21 | 46010497 | 46032091 | 96 | 8 | 27889 | -1.595 | 0.94464 | *KRTAP10-7* |
| ENSG00000121289 | 19 | 33359902 | 33472897 | 514 | 44 | 51462 | -1.5955 | 0.9447 | *CEP89* |
| ENSG00000182718 | 15 | 60629333 | 60705082 | 214 | 37 | 47313 | -1.5961 | 0.94477 | *ANXA2* |
| ENSG00000116350 | 1 | 29464255 | 29518499 | 130 | 23 | 39009 | -1.5974 | 0.94492 | *SRSF4* |
| ENSG00000268098 | 21 | 37392368 | 37427818 | 123 | 14 | 53881 | -1.5975 | 0.94492 | *AP000688.1* |
| ENSG00000112561 | 6 | 41641716 | 41713997 | 233 | 45 | 51581 | -1.5981 | 0.94498 | *TFEB* |

| | | | | | | | | | |
|---|---|---|---|---|---|---|---|---|---|
| ENSG00000182836 | 5 | 41297056 | 41520730 | 600 | 62 | 39397 | -1.5982 | 0.945 | *PLCXD3* |
| ENSG00000072858 | 3 | 113241143 | 113358425 | 390 | 49 | 41448 | -1.599 | 0.94509 | *SIDT1* |
| ENSG00000268688 | 2 | 37058327 | 37078530 | 41 | 11 | 33579 | -1.6002 | 0.94522 | *AC007382.1* |
| ENSG00000103723 | 15 | 83318033 | 83388666 | 184 | 27 | 42597 | -1.6006 | 0.94526 | *AP3B2* |
| ENSG00000198380 | 2 | 69536905 | 69624382 | 181 | 12 | 36579 | -1.6012 | 0.94534 | *GFPT1* |
| ENSG00000172289 | 11 | 59470296 | 59491337 | 67 | 8 | 26786 | -1.6013 | 0.94535 | *OR10V1* |
| ENSG00000086967 | 19 | 50926160 | 50979578 | 168 | 34 | 46421 | -1.6029 | 0.94553 | *MYBPC2* |
| ENSG00000156486 | 8 | 99429250 | 99455076 | 36 | 12 | 52190 | -1.6045 | 0.94569 | *KCNS2* |
| ENSG00000075539 | 4 | 48489378 | 48792339 | 463 | 36 | 47348 | -1.6048 | 0.94573 | *FRYL* |
| ENSG00000263053 | 17 | 79359275 | 79385658 | 74 | 19 | 37495 | -1.6048 | 0.94573 | *RP11-1055B8.6* |
| ENSG00000151474 | 10 | 13675706 | 14514141 | 3738 | 356 | 45685 | -1.6059 | 0.94586 | *FRMD4A* |
| ENSG00000241241 | 17 | 39247754 | 39268461 | 182 | 11 | 34193 | -1.6071 | 0.94598 | *KRTAP4-16P* |
| ENSG00000261603 | 3 | 46751073 | 46787919 | 160 | 14 | 29879 | -1.6075 | 0.94603 | *PRSS46* |
| ENSG00000040275 | 5 | 169000638 | 169041782 | 145 | 15 | 51452 | -1.6089 | 0.94618 | *SPDL1* |
| ENSG00000173334 | 8 | 126432563 | 126460647 | 55 | 20 | 37729 | -1.6089 | 0.94618 | *TRIB1* |
| ENSG00000215845 | 1 | 160997421 | 161018780 | 64 | 15 | 41300 | -1.6093 | 0.94623 | *TSTD1* |
| ENSG00000131669 | 9 | 95873771 | 95906570 | 139 | 24 | 38525 | -1.6118 | 0.9465 | *NINJ1* |
| ENSG00000080371 | 12 | 72138654 | 72194699 | 112 | 15 | 34305 | -1.6132 | 0.94665 | *RAB21* |
| ENSG00000134996 | 9 | 77693459 | 77772181 | 326 | 48 | 51579 | -1.6162 | 0.94698 | *OSTF1* |
| ENSG00000100294 | 22 | 43518212 | 43549400 | 59 | 20 | 40133 | -1.6169 | 0.94705 | *MCAT* |
| ENSG00000130561 | 2 | 234206462 | 234265701 | 207 | 30 | 42969 | -1.6184 | 0.94721 | *SAG* |
| ENSG00000179632 | 8 | 145149402 | 145172514 | 48 | 7 | 32889 | -1.6209 | 0.94748 | *MAF1* |
| ENSG00000238227 | 9 | 138996427 | 139020731 | 88 | 22 | 50513 | -1.6211 | 0.9475 | *C9orf69* |
| ENSG00000128408 | 22 | 45799572 | 45838376 | 145 | 14 | 30746 | -1.6214 | 0.94753 | *RIBC2* |
| ENSG00000222031 | 2 | 151847734 | 151915288 | 274 | 29 | 38858 | -1.6214 | 0.94754 | *AC023469.1* |
| ENSG00000099958 | 22 | 24166690 | 24191315 | 154 | 19 | 45715 | -1.6241 | 0.94782 | *DERL3* |
| ENSG00000120910 | 8 | 22288332 | 22408652 | 264 | 20 | 48099 | -1.6243 | 0.94784 | *PPP3CC* |
| ENSG00000033122 | 1 | 70024081 | 70627628 | 1901 | 142 | 47291 | -1.6257 | 0.94799 | *LRRC7* |
| ENSG00000177212 | 1 | 248426073 | 248447138 | 77 | 5 | 50177 | -1.6269 | 0.94812 | *OR2T33* |
| ENSG00000157219 | 7 | 154852034 | 154887459 | 143 | 15 | 47616 | -1.6269 | 0.94812 | *HTR5A* |
| ENSG00000160293 | 9 | 136617016 | 136867726 | 1237 | 142 | 41139 | -1.6271 | 0.94814 | *VAV2* |
| ENSG00000166900 | 11 | 59470929 | 59583354 | 363 | 18 | 28765 | -1.6285 | 0.94829 | *STX3* |
| ENSG00000121570 | 3 | 109034988 | 109066419 | 112 | 10 | 21423 | -1.6307 | 0.94852 | *DPPA4* |
| ENSG00000168710 | 1 | 110517308 | 110576357 | 53 | 19 | 39056 | -1.6307 | 0.94853 | *AHCYL1* |
| ENSG00000101349 | 20 | 9508036 | 9829689 | 1118 | 118 | 43786 | -1.632 | 0.94866 | *PAK7* |
| ENSG00000180872 | 6 | 50001288 | 50026364 | 69 | 12 | 38795 | -1.6329 | 0.94875 | *DEFB112* |

| | | | | | | | | | |
|---|---|---|---|---|---|---|---|---|---|
| ENSG00000188611 | 10 | 51932538 | 52018370 | 106 | 15 | 20665 | -1.633 | 0.94877 | *ASAH2* |
| ENSG00000107897 | 10 | 27474146 | 27541059 | 411 | 26 | 38280 | -1.6336 | 0.94883 | *ACBD5* |
| ENSG00000271321 | 7 | 143442182 | 143464789 | 4 | 2 | 42915 | -1.6341 | 0.94888 | *CTAGE6* |
| ENSG00000173085 | 4 | 84172689 | 84216067 | 111 | 14 | 41372 | -1.6341 | 0.94889 | *COQ2* |
| ENSG00000140463 | 15 | 72968527 | 73040817 | 164 | 26 | 43583 | -1.6351 | 0.94899 | *BBS4* |
| ENSG00000081041 | 4 | 74952752 | 74975010 | 35 | 12 | 35821 | -1.6352 | 0.949 | *CXCL2* |
| ENSG00000120436 | 6 | 167559759 | 167581817 | 53 | 12 | 53695 | -1.6354 | 0.94902 | *GPR31* |
| ENSG00000181409 | 17 | 79081095 | 79149877 | 333 | 62 | 42631 | -1.6356 | 0.94904 | *AATK* |
| ENSG00000134248 | 1 | 110933871 | 110960564 | 81 | 11 | 50939 | -1.6361 | 0.94909 | *LAMTOR5* |
| ENSG00000154079 | 6 | 71266620 | 71309272 | 156 | 12 | 51429 | -1.6361 | 0.9491 | *C6orf57* |
| ENSG00000182899 | 3 | 197666858 | 197693481 | 70 | 10 | 36124 | -1.6366 | 0.94914 | *RPL35A* |
| ENSG00000162722 | 1 | 248010501 | 248051507 | 236 | 36 | 44912 | -1.6371 | 0.94919 | *TRIM58* |
| ENSG00000089723 | 14 | 94482675 | 94525276 | 161 | 23 | 46373 | -1.6375 | 0.94924 | *OTUB2* |
| ENSG00000152642 | 3 | 32137181 | 32220205 | 361 | 34 | 30410 | -1.6412 | 0.94962 | *GPD1L* |
| ENSG00000157927 | 7 | 4826686 | 4933350 | 530 | 61 | 41385 | -1.6425 | 0.94975 | *RADIL* |
| ENSG00000127318 | 12 | 68632022 | 68657387 | 118 | 16 | 49095 | -1.6435 | 0.94986 | *IL22* |
| ENSG00000163069 | 4 | 52876872 | 52914648 | 54 | 11 | 35744 | -1.6443 | 0.94994 | *SGCB* |
| ENSG00000136689 | 2 | 113854791 | 113901593 | 198 | 23 | 43583 | -1.6451 | 0.95003 | *IL1RN* |
| ENSG00000165819 | 14 | 21956277 | 21989517 | 106 | 17 | 52475 | -1.6455 | 0.95006 | *METTL3* |
| ENSG00000164576 | 5 | 153815517 | 153850614 | 87 | 26 | 52627 | -1.6461 | 0.95013 | *SAP30L* |
| ENSG00000105321 | 19 | 47749237 | 47785210 | 117 | 15 | 48168 | -1.6461 | 0.95013 | *CCDC9* |
| ENSG00000112116 | 6 | 52091479 | 52119335 | 93 | 18 | 47113 | -1.6475 | 0.95028 | *IL17F* |
| ENSG00000157259 | 7 | 92066767 | 92098150 | 81 | 13 | 48390 | -1.6478 | 0.9503 | *GATAD1* |
| ENSG00000182601 | 16 | 25693347 | 26159009 | 1811 | 261 | 49152 | -1.6484 | 0.95036 | *HS3ST4* |
| ENSG00000213096 | 19 | 24206276 | 24322654 | 450 | 19 | 50336 | -1.6484 | 0.95037 | *ZNF254* |
| ENSG00000184787 | 21 | 46178955 | 46231934 | 226 | 29 | 47679 | -1.6499 | 0.95052 | *UBE2G2* |
| ENSG00000254349 | 8 | 75502010 | 75745548 | 696 | 81 | 41588 | -1.6502 | 0.95055 | *RP11-758M4.1* |
| ENSG00000104613 | 8 | 19664651 | 19719594 | 170 | 28 | 52134 | -1.6504 | 0.95057 | *INTS10* |
| ENSG00000197437 | 1 | 247825320 | 247846365 | 140 | 9 | 54818 | -1.6506 | 0.95059 | *OR13G1* |
| ENSG00000186212 | 4 | 77806082 | 77829002 | 72 | 9 | 54259 | -1.6518 | 0.95071 | *SOWAHB* |
| ENSG00000089195 | 20 | 5907881 | 5941182 | 165 | 27 | 36298 | -1.6528 | 0.95081 | *TRMT6* |
| ENSG00000087495 | 20 | 58142564 | 58432766 | 940 | 131 | 42526 | -1.6528 | 0.95081 | *PHACTR3* |
| ENSG00000138286 | 10 | 74917924 | 75014262 | 240 | 7 | 19905 | -1.6536 | 0.9509 | *FAM149B1* |
| ENSG00000170421 | 12 | 53280977 | 53353738 | 291 | 31 | 50335 | -1.6541 | 0.95094 | *KRT8* |
| ENSG00000177045 | 19 | 46258043 | 46282484 | 38 | 9 | 51245 | -1.6558 | 0.95112 | *SIX5* |
| ENSG00000105866 | 7 | 21457652 | 21564440 | 382 | 51 | 39576 | -1.656 | 0.95114 | *SP4* |

| | | | | | | | | | |
|---|---|---|---|---|---|---|---|---|---|
| ENSG00000177483 | 2 | 238697032 | 238761451 | 167 | 23 | 39134 | -1.6574 | 0.95128 | *RBM44* |
| ENSG00000111845 | 6 | 10684928 | 10720015 | 25 | 16 | 53733 | -1.6573 | 0.95128 | *PAK1IP1* |
| ENSG00000213132 | 3 | 187886331 | 187908598 | 85 | 22 | 40781 | -1.6579 | 0.95133 | *AC022498.1* |
| ENSG00000139173 | 12 | 44219770 | 44793545 | 1616 | 83 | 36579 | -1.6579 | 0.95133 | *TMEM117* |
| ENSG00000243130 | 19 | 43501808 | 43540664 | 79 | 18 | 27261 | -1.6585 | 0.95139 | *PSG11* |
| ENSG00000105550 | 19 | 49248816 | 49271587 | 71 | 21 | 32814 | -1.6613 | 0.95167 | *FGF21* |
| ENSG00000221845 | 7 | 47684842 | 47711246 | 95 | 18 | 54804 | -1.6642 | 0.95196 | *C7orf65* |
| ENSG00000151239 | 12 | 44177526 | 44210178 | 89 | 17 | 48586 | -1.6642 | 0.95197 | *TWF1* |
| ENSG00000169026 | 4 | 665618 | 693230 | 70 | 21 | 50520 | -1.6664 | 0.95218 | *MFSD7* |
| ENSG00000241962 | 2 | 99747948 | 99949204 | 574 | 46 | 45393 | -1.6672 | 0.95226 | *C2orf15* |
| ENSG00000170967 | 11 | 103897308 | 103919922 | 60 | 13 | 45175 | -1.6672 | 0.95226 | *DDI1* |
| ENSG00000118939 | 13 | 76113619 | 76190085 | 263 | 24 | 38912 | -1.6681 | 0.95235 | *UCHL3* |
| ENSG00000073146 | 22 | 50518308 | 50610119 | 539 | 27 | 50979 | -1.6684 | 0.95238 | *MOV10L1* |
| ENSG00000176619 | 19 | 2417636 | 2466994 | 218 | 28 | 47155 | -1.6698 | 0.95252 | *LMNB2* |
| ENSG00000068796 | 5 | 61591989 | 61843076 | 611 | 42 | 50616 | -1.6701 | 0.95255 | *KIF2A* |
| ENSG00000205108 | 9 | 34713052 | 34739464 | 81 | 14 | 35139 | -1.6702 | 0.95256 | *FAM205A* |
| ENSG00000174196 | 10 | 51177938 | 51202032 | 1 | 1 | 42696 | -1.6706 | 0.9526 | *FAM21D* |
| ENSG00000182919 | 11 | 93464757 | 93507915 | 104 | 9 | 31987 | -1.6723 | 0.95277 | *C11orf54* |
| ENSG00000170293 | 3 | 32270171 | 32421817 | 577 | 113 | 46454 | -1.675 | 0.95303 | *CMTM8* |
| ENSG00000142197 | 21 | 37519080 | 37676572 | 701 | 90 | 48237 | -1.6753 | 0.95306 | *DOPEY2* |
| ENSG00000174227 | 4 | 482989 | 543985 | 277 | 17 | 34053 | -1.6754 | 0.95307 | *PIGG* |
| ENSG00000175746 | 15 | 39532885 | 39557046 | 85 | 20 | 47436 | -1.6756 | 0.95309 | *C15orf54* |
| ENSG00000187766 | 21 | 46021996 | 46042871 | 86 | 12 | 30489 | -1.6759 | 0.95312 | *KRTAP10-8* |
| ENSG00000100129 | 22 | 38234875 | 38295414 | 89 | 15 | 44277 | -1.6763 | 0.95316 | *EIF3L* |
| ENSG00000182223 | 4 | 48482269 | 48506406 | 57 | 9 | 51271 | -1.6774 | 0.95327 | *ZAR1* |
| ENSG00000084453 | 12 | 21407534 | 21582528 | 711 | 83 | 45585 | -1.6777 | 0.9533 | *SLCO1A2* |
| ENSG00000165617 | 14 | 59090685 | 59125039 | 103 | 20 | 50556 | -1.6794 | 0.95346 | *DACT1* |
| ENSG00000183696 | 7 | 48118225 | 48158330 | 75 | 17 | 45463 | -1.6797 | 0.95349 | *UPP1* |
| ENSG00000105278 | 19 | 3794022 | 3879030 | 288 | 37 | 49307 | -1.6803 | 0.95355 | *ZFR2* |
| ENSG00000033627 | 17 | 40600862 | 40684629 | 154 | 23 | 48372 | -1.685 | 0.95401 | *ATP6V0A1* |
| ENSG00000185504 | 17 | 79496911 | 79530987 | 136 | 26 | 48857 | -1.6854 | 0.95404 | *C17orf70* |
| ENSG00000131941 | 19 | 33459499 | 33565794 | 456 | 52 | 35171 | -1.686 | 0.9541 | *RHPN2* |
| ENSG00000095777 | 10 | 26213196 | 26511456 | 1544 | 43 | 50232 | -1.6865 | 0.95415 | *MYO3A* |
| ENSG00000188051 | 19 | 17536318 | 17569376 | 87 | 19 | 44349 | -1.687 | 0.9542 | *TMEM221* |
| ENSG00000162692 | 1 | 101175298 | 101214601 | 65 | 23 | 42253 | -1.6878 | 0.95428 | *VCAM1* |
| ENSG00000105963 | 7 | 927540 | 1005043 | 397 | 83 | 45698 | -1.6879 | 0.95429 | *ADAP1* |

| | | | | | | | | | |
|---|---|---|---|---|---|---|---|---|---|
| ENSG00000264545 | 9 | 21792635 | 22042985 | 572 | 81 | 39389 | -1.6915 | 0.95463 | *RP11-145E5.5* |
| ENSG00000258873 | 19 | 57655389 | 57688811 | 142 | 25 | 35956 | -1.6915 | 0.95463 | *DUXA* |
| ENSG00000162461 | 1 | 16052900 | 16077891 | 60 | 12 | 43715 | -1.6921 | 0.95468 | *SLC25A34* |
| ENSG00000116237 | 1 | 6271253 | 6306032 | 91 | 16 | 35060 | -1.6932 | 0.9548 | *ICMT* |
| ENSG00000258984 | 2 | 238865656 | 239018053 | 552 | 33 | 47180 | -1.6934 | 0.95481 | *UBE2F-SCLY* |
| ENSG00000120549 | 10 | 23973675 | 24846772 | 2787 | 269 | 45135 | -1.6941 | 0.95487 | *KIAA1217* |
| ENSG00000105639 | 19 | 17925589 | 17968880 | 111 | 31 | 43556 | -1.6943 | 0.95489 | *JAK3* |
| ENSG00000185305 | 5 | 53169775 | 53616412 | 1572 | 148 | 45524 | -1.6951 | 0.95497 | *ARL15* |
| ENSG00000177105 | 11 | 3838208 | 3872213 | 89 | 20 | 48653 | -1.6978 | 0.95522 | *RHOG* |
| ENSG00000182196 | 12 | 123454607 | 123477456 | 22 | 7 | 41337 | -1.6984 | 0.95528 | *ARL6IP4* |
| ENSG00000100836 | 14 | 23780498 | 23805394 | 60 | 17 | 45461 | -1.6992 | 0.95536 | *PABPN1* |
| ENSG00000135406 | 12 | 49677035 | 49702465 | 114 | 11 | 46601 | -1.7005 | 0.95548 | *PRPH* |
| ENSG00000118526 | 6 | 134200276 | 134226691 | 79 | 15 | 51718 | -1.7014 | 0.95556 | *TCF21* |
| ENSG00000251569 | 2 | 170325990 | 170392432 | 200 | 31 | 48550 | -1.7044 | 0.95585 | *RP11-724O16.1* |
| ENSG00000184937 | 11 | 32399321 | 32467176 | 284 | 29 | 29325 | -1.7047 | 0.95588 | *WT1* |
| ENSG00000168314 | 3 | 39498689 | 39580970 | 366 | 19 | 22934 | -1.7054 | 0.95594 | *MOBP* |
| ENSG00000196774 | 9 | 67916761 | 67979840 | 8 | 3 | 42619 | -1.7072 | 0.95611 | *ANKRD20A1* |
| ENSG00000136052 | 12 | 105186331 | 105362522 | 600 | 47 | 41463 | -1.7081 | 0.95619 | *SLC41A2* |
| ENSG00000100139 | 22 | 38291664 | 38348829 | 105 | 22 | 46088 | -1.7096 | 0.95633 | *MICALL1* |
| ENSG00000134461 | 10 | 5893580 | 5941869 | 250 | 24 | 43932 | -1.7098 | 0.95635 | *ANKRD16* |
| ENSG00000104228 | 8 | 27132404 | 27178836 | 93 | 14 | 41253 | -1.7114 | 0.9565 | *TRIM35* |
| ENSG00000243137 | 19 | 43686854 | 43721451 | 139 | 21 | 22771 | -1.7146 | 0.95679 | *PSG4* |
| ENSG00000205457 | 16 | 33194156 | 33274727 | 3 | 1 | 42891 | -1.7155 | 0.95688 | *TP53TG3C* |
| ENSG00000089009 | 12 | 112832994 | 112866642 | 35 | 8 | 34301 | -1.7159 | 0.95691 | *RPL6* |
| ENSG00000117697 | 1 | 212889495 | 212975124 | 453 | 24 | 46399 | -1.718 | 0.9571 | *NSL1* |
| ENSG00000183850 | 19 | 23248012 | 23340021 | 239 | 22 | 46220 | -1.719 | 0.95719 | *ZNF730* |
| ENSG00000138678 | 4 | 84447067 | 84537028 | 318 | 26 | 49757 | -1.7191 | 0.9572 | *AGPAT9* |
| ENSG00000167535 | 12 | 49197577 | 49232724 | 57 | 16 | 50113 | -1.7198 | 0.95727 | *CACNB3* |
| ENSG00000131697 | 1 | 5912871 | 6062533 | 517 | 45 | 54184 | -1.721 | 0.95737 | *NPHP4* |
| ENSG00000188501 | 15 | 66829517 | 66868317 | 165 | 30 | 53610 | -1.7213 | 0.9574 | *LCTL* |
| ENSG00000205609 | 16 | 28380900 | 28425200 | 1 | 1 | 43203 | -1.7213 | 0.9574 | *EIF3CL* |
| ENSG00000163093 | 2 | 170325688 | 170392432 | 201 | 31 | 48595 | -1.7233 | 0.95758 | *BBS5* |
| ENSG00000181656 | 1 | 100993693 | 101017574 | 64 | 10 | 36962 | -1.7234 | 0.95759 | *GPR88* |
| ENSG00000140332 | 15 | 70330129 | 70400515 | 194 | 47 | 44527 | -1.7245 | 0.95769 | *TLE3* |
| ENSG00000093134 | 6 | 133033926 | 133065904 | 137 | 20 | 43856 | -1.7246 | 0.9577 | *VNN3* |
| ENSG00000171135 | 3 | 9922238 | 9946033 | 81 | 17 | 42471 | -1.7262 | 0.95784 | *JAGN1* |

| | | | | | | | | | |
|---|---|---|---|---|---|---|---|---|---|
| ENSG00000134779 | 18 | 34349987 | 34419158 | 131 | 11 | 50495 | -1.7291 | 0.95811 | *TPGS2* |
| ENSG00000177556 | 5 | 151111877 | 151162093 | 126 | 15 | 44120 | -1.7305 | 0.95823 | *ATOX1* |
| ENSG00000134138 | 15 | 37171406 | 37403504 | 504 | 119 | 46410 | -1.7322 | 0.95838 | *MEIS2* |
| ENSG00000170509 | 4 | 88214941 | 88254058 | 132 | 24 | 49685 | -1.7325 | 0.95841 | *HSD17B13* |
| ENSG00000180432 | 3 | 42887497 | 42927633 | 102 | 14 | 43772 | -1.733 | 0.95846 | *CYP8B1* |
| ENSG00000269396 | 7 | 292918 | 314070 | 41 | 16 | 31125 | -1.7353 | 0.95865 | *AC187652.1* |
| ENSG00000146676 | 7 | 44905896 | 44934960 | 45 | 10 | 45077 | -1.7357 | 0.95869 | *PURB* |
| ENSG00000253857 | 8 | 56064043 | 56087287 | 54 | 11 | 41933 | -1.7356 | 0.95869 | *RP11-386G21.2* |
| ENSG00000213551 | 10 | 74933120 | 75018620 | 213 | 6 | 19504 | -1.7375 | 0.95885 | *DNAJC9* |
| ENSG00000197487 | 19 | 56677389 | 56707144 | 167 | 36 | 41913 | -1.7395 | 0.95903 | *GALP* |
| ENSG00000102531 | 13 | 49540048 | 49793915 | 360 | 31 | 45672 | -1.7411 | 0.95917 | *FNDC3A* |
| ENSG00000166091 | 14 | 23836017 | 23858981 | 71 | 21 | 45522 | -1.7419 | 0.95924 | *CMTM5* |
| ENSG00000162627 | 1 | 99117236 | 99236056 | 409 | 28 | 47850 | -1.7428 | 0.95931 | *SNX7* |
| ENSG00000065600 | 1 | 212527273 | 212598243 | 194 | 33 | 41785 | -1.7433 | 0.95936 | *TMEM206* |
| ENSG00000174442 | 15 | 66787297 | 66852115 | 218 | 27 | 55777 | -1.7453 | 0.95953 | *ZWILCH* |
| ENSG00000158773 | 1 | 160999041 | 161025767 | 74 | 15 | 38838 | -1.7469 | 0.95968 | *USF1* |
| ENSG00000103264 | 16 | 87350593 | 87435748 | 311 | 50 | 40526 | -1.7476 | 0.95973 | *FBXO31* |
| ENSG00000103522 | 16 | 27403483 | 27472115 | 246 | 50 | 49166 | -1.7496 | 0.95991 | *IL21R* |
| ENSG00000120738 | 5 | 137791179 | 137815004 | 60 | 13 | 45555 | -1.7502 | 0.95995 | *EGR1* |
| ENSG00000258677 | 8 | 74590926 | 74801089 | 472 | 47 | 48685 | -1.7502 | 0.95996 | *RP11-463D19.2* |
| ENSG00000260300 | 16 | 83931738 | 83995058 | 392 | 48 | 47696 | -1.7508 | 0.96001 | *RP11-505K9.4* |
| ENSG00000159128 | 21 | 34765202 | 34861655 | 308 | 25 | 37169 | -1.7523 | 0.96014 | *IFNGR2* |
| ENSG00000158286 | 1 | 6255535 | 6291359 | 93 | 17 | 34127 | -1.7551 | 0.96037 | *RNF207* |
| ENSG00000215375 | 4 | 657369 | 685822 | 72 | 16 | 51258 | -1.7557 | 0.96043 | *MYL5* |
| ENSG00000013588 | 12 | 13033716 | 13080871 | 136 | 28 | 37686 | -1.756 | 0.96045 | *GPRC5A* |
| ENSG00000110675 | 11 | 107451817 | 107547505 | 289 | 49 | 39339 | -1.7562 | 0.96047 | *ELMOD1* |
| ENSG00000165124 | 9 | 113117531 | 113352160 | 1091 | 105 | 49227 | -1.7567 | 0.96052 | *SVEP1* |
| ENSG00000178401 | 12 | 49730700 | 49761309 | 66 | 10 | 26832 | -1.7583 | 0.96065 | *DNAJC22* |
| ENSG00000011275 | 7 | 5649678 | 5831370 | 714 | 58 | 44950 | -1.7612 | 0.9609 | *RNF216* |
| ENSG00000113296 | 5 | 79277134 | 79389110 | 319 | 26 | 49677 | -1.7617 | 0.96094 | *THBS4* |
| ENSG00000120008 | 10 | 122600687 | 122679036 | 371 | 22 | 52946 | -1.7625 | 0.96101 | *WDR11* |
| ENSG00000213923 | 22 | 38676697 | 38804527 | 253 | 52 | 45753 | -1.7637 | 0.96111 | *CSNK1E* |
| ENSG00000197322 | 17 | 32891142 | 32916388 | 99 | 22 | 50486 | -1.7658 | 0.96128 | *C17orf102* |
| ENSG00000179943 | 19 | 56092746 | 56123336 | 95 | 17 | 51667 | -1.766 | 0.9613 | *FIZ1* |
| ENSG00000185238 | 11 | 20399076 | 20540840 | 291 | 36 | 46913 | -1.7667 | 0.96136 | *PRMT3* |
| ENSG00000105229 | 19 | 3997644 | 4049384 | 164 | 22 | 43863 | -1.7667 | 0.96136 | *PIAS4* |

| | | | | | | | | | |
|---|---|---|---|---|---|---|---|---|---|
| ENSG00000135482 | 12 | 56501943 | 56526278 | 42 | 14 | 33184 | -1.768 | 0.96147 | *ZC3H10* |
| ENSG00000179119 | 11 | 18617948 | 18666338 | 172 | 28 | 39123 | -1.7687 | 0.96153 | *SPTY2D1* |
| ENSG00000188993 | 4 | 52849868 | 52893786 | 81 | 12 | 35152 | -1.7688 | 0.96154 | *LRRC66* |
| ENSG00000157214 | 7 | 89786904 | 89877451 | 279 | 25 | 44099 | -1.7691 | 0.96156 | *STEAP2* |
| ENSG00000138081 | 2 | 48006455 | 48142932 | 431 | 35 | 52574 | -1.7701 | 0.96164 | *FBXO11* |
| ENSG00000078967 | 7 | 43956037 | 44005735 | 169 | 23 | 53753 | -1.7704 | 0.96167 | *UBE2D4* |
| ENSG00000110880 | 12 | 109028885 | 109135372 | 270 | 31 | 41538 | -1.7715 | 0.96176 | *CORO1C* |
| ENSG00000117595 | 1 | 209949036 | 209989465 | 121 | 15 | 52839 | -1.7728 | 0.96187 | *IRF6* |
| ENSG00000204640 | 2 | 101076944 | 101109742 | 219 | 14 | 39259 | -1.7732 | 0.9619 | *NMS* |
| ENSG00000229117 | 12 | 56500370 | 56521727 | 37 | 14 | 34698 | -1.7738 | 0.96195 | *RPL41* |
| ENSG00000114021 | 3 | 100043545 | 100085710 | 88 | 22 | 41738 | -1.7746 | 0.96202 | *NIT2* |
| ENSG00000106571 | 7 | 41990548 | 42287469 | 954 | 139 | 47151 | -1.7747 | 0.96203 | *GLI3* |
| ENSG00000179526 | 8 | 145143536 | 145173027 | 59 | 7 | 30689 | -1.7772 | 0.96223 | *SHARPIN* |
| ENSG00000184647 | 8 | 10373056 | 10421676 | 308 | 35 | 53650 | -1.7779 | 0.96229 | *PRSS55* |
| ENSG00000178287 | 8 | 7695398 | 7736389 | 7 | 2 | 44066 | -1.7792 | 0.9624 | *SPAG11A* |
| ENSG00000182613 | 5 | 180571943 | 180592890 | 128 | 17 | 29730 | -1.7799 | 0.96246 | *OR2V2* |
| ENSG00000176753 | 15 | 40532865 | 40555170 | 54 | 20 | 40551 | -1.7813 | 0.96257 | *C15orf56* |
| ENSG00000109184 | 4 | 52699166 | 52793003 | 124 | 15 | 33962 | -1.7829 | 0.9627 | *DCUN1D4* |
| ENSG00000083807 | 19 | 58980879 | 59033780 | 124 | 20 | 47435 | -1.7852 | 0.96289 | *SLC27A5* |
| ENSG00000136450 | 17 | 56070721 | 56094707 | 48 | 14 | 48274 | -1.7857 | 0.96293 | *SRSF1* |
| ENSG00000257411 | 12 | 56485115 | 56513073 | 49 | 18 | 40567 | -1.786 | 0.96295 | *RP11-603J24.9* |
| ENSG00000106714 | 9 | 39062764 | 39298312 | 220 | 20 | 40199 | -1.7866 | 0.963 | *CNTNAP3* |
| ENSG00000143850 | 1 | 204177979 | 204356793 | 598 | 66 | 49333 | -1.7872 | 0.96305 | *PLEKHA6* |
| ENSG00000232040 | 6 | 28529407 | 28593989 | 173 | 27 | 40463 | -1.7873 | 0.96306 | *SCAND3* |
| ENSG00000186354 | 9 | 91595778 | 91621055 | 72 | 20 | 52454 | -1.7876 | 0.96308 | *C9orf47* |
| ENSG00000103549 | 16 | 30763066 | 30797628 | 36 | 14 | 44742 | -1.7881 | 0.96312 | *RNF40* |
| ENSG00000075624 | 7 | 5556782 | 5613415 | 234 | 48 | 44589 | -1.7885 | 0.96315 | *ACTB* |
| ENSG00000100916 | 14 | 36285524 | 36411531 | 212 | 33 | 35622 | -1.7889 | 0.96318 | *BRMS1L* |
| ENSG00000182150 | 9 | 98627983 | 98786842 | 458 | 40 | 53401 | -1.7891 | 0.9632 | *ERCC6L2* |
| ENSG00000107341 | 9 | 33807565 | 33930402 | 371 | 29 | 53016 | -1.7926 | 0.96348 | *UBE2R2* |
| ENSG00000101336 | 20 | 30629991 | 30699659 | 127 | 24 | 38444 | -1.7958 | 0.96374 | *HCK* |
| ENSG00000128731 | 15 | 28346186 | 28577298 | 143 | 30 | 34870 | -1.7978 | 0.96389 | *HERC2* |
| ENSG00000113396 | 5 | 127863706 | 128379335 | 2360 | 95 | 47549 | -1.7997 | 0.96405 | *SLC27A6* |
| ENSG00000196954 | 11 | 104803593 | 104850163 | 122 | 20 | 46873 | -1.8003 | 0.96409 | *CASP4* |
| ENSG00000145725 | 5 | 102445853 | 102558500 | 279 | 23 | 48903 | -1.801 | 0.96415 | *PPIP5K2* |
| ENSG00000115257 | 19 | 1471427 | 1500751 | 83 | 19 | 40193 | -1.8022 | 0.96424 | *PCSK4* |

| | | | | | | | | | |
|---|---|---|---|---|---|---|---|---|---|
| ENSG00000074966 | 4 | 48058410 | 48146273 | 242 | 34 | 45719 | -1.8026 | 0.96428 | *TXK* |
| ENSG00000198093 | 19 | 52382477 | 52418293 | 158 | 15 | 31364 | -1.8043 | 0.96441 | *ZNF649* |
| ENSG00000268461 | 19 | 20037769 | 20058639 | 75 | 15 | 33216 | -1.8054 | 0.9645 | *AC007204.1* |
| ENSG00000132376 | 17 | 1387865 | 1430182 | 183 | 28 | 53343 | -1.8061 | 0.96455 | *INPP5K* |
| ENSG00000268702 | 14 | 23780839 | 23801484 | 48 | 14 | 46249 | -1.8064 | 0.96457 | *AL049829.1* |
| ENSG00000259494 | 15 | 88992707 | 89020650 | 14 | 8 | 47634 | -1.8068 | 0.9646 | *MRPL46* |
| ENSG00000132423 | 6 | 99807276 | 99852080 | 146 | 22 | 47836 | -1.8078 | 0.96468 | *COQ3* |
| ENSG00000149657 | 20 | 60687517 | 60720434 | 69 | 9 | 35102 | -1.8088 | 0.96476 | *LSM14B* |
| ENSG00000100376 | 22 | 45694849 | 45747836 | 182 | 15 | 31483 | -1.8092 | 0.96479 | *FAM118A* |
| ENSG00000187010 | 1 | 25588884 | 25666936 | 35 | 21 | 45487 | -1.8097 | 0.96483 | *RHD* |
| ENSG00000120658 | 13 | 43777654 | 44371044 | 1625 | 124 | 38870 | -1.8123 | 0.96503 | *ENOX1* |
| ENSG00000162383 | 1 | 53542855 | 53618289 | 393 | 54 | 46565 | -1.8127 | 0.96506 | *SLC1A7* |
| ENSG00000166090 | 14 | 23832018 | 23855612 | 70 | 21 | 44988 | -1.8133 | 0.96511 | *IL25* |
| ENSG00000259495 | 15 | 80627195 | 80705917 | 406 | 36 | 54011 | -1.8149 | 0.96523 | *RP11-210M15.2* |
| ENSG00000218891 | 19 | 56078899 | 56102211 | 102 | 14 | 53920 | -1.8154 | 0.96527 | *ZNF579* |
| ENSG00000040341 | 8 | 74322604 | 74669943 | 1171 | 88 | 41944 | -1.8171 | 0.9654 | *STAU2* |
| ENSG00000117616 | 1 | 25558728 | 25674704 | 76 | 37 | 46106 | -1.8179 | 0.96546 | *C1orf63* |
| ENSG00000132591 | 17 | 27171956 | 27198085 | 49 | 7 | 36948 | -1.8182 | 0.96548 | *ERAL1* |
| ENSG00000120539 | 10 | 27433753 | 27485853 | 239 | 24 | 46739 | -1.8186 | 0.96552 | *MASTL* |
| ENSG00000095637 | 10 | 97061528 | 97331171 | 1049 | 119 | 47063 | -1.8197 | 0.96559 | *SORBS1* |
| ENSG00000138756 | 4 | 79687496 | 79847526 | 306 | 20 | 29184 | -1.8199 | 0.96561 | *BMP2K* |
| ENSG00000177034 | 5 | 79265584 | 79297082 | 69 | 9 | 55294 | -1.8205 | 0.96566 | *MTX3* |
| ENSG00000135312 | 6 | 78161948 | 78183490 | 53 | 13 | 47904 | -1.822 | 0.96578 | *HTR1B* |
| ENSG00000205318 | 6 | 10623993 | 10657501 | 49 | 12 | 41457 | -1.8222 | 0.96579 | *GCNT6* |
| ENSG00000077782 | 8 | 38258656 | 38336352 | 148 | 27 | 48583 | -1.8225 | 0.96581 | *FGFR1* |
| ENSG00000258311 | 12 | 56099820 | 56128487 | 45 | 16 | 36056 | -1.8227 | 0.96582 | *RP11-644F5.10* |
| ENSG00000170515 | 12 | 56488103 | 56517691 | 47 | 17 | 39855 | -1.8235 | 0.96589 | *PA2G4* |
| ENSG00000154767 | 3 | 14176647 | 14230283 | 118 | 21 | 50929 | -1.8238 | 0.96591 | *XPC* |
| ENSG00000204941 | 19 | 43660408 | 43700688 | 260 | 24 | 16868 | -1.8241 | 0.96593 | *PSG5* |
| ENSG00000181722 | 3 | 114046941 | 114876118 | 1691 | 191 | 42980 | -1.8254 | 0.96603 | *ZBTB20* |
| ENSG00000235194 | 14 | 23754852 | 23782057 | 56 | 19 | 48687 | -1.8256 | 0.96605 | *PPP1R3E* |
| ENSG00000159885 | 19 | 44519494 | 44547265 | 59 | 9 | 28318 | -1.8274 | 0.96618 | *ZNF222* |
| ENSG00000181215 | 4 | 5948845 | 6001545 | 381 | 50 | 43023 | -1.8296 | 0.96634 | *C4orf50* |
| ENSG00000205495 | 11 | 5057756 | 5078691 | 211 | 8 | 52818 | -1.8298 | 0.96636 | *OR52J3* |
| ENSG00000182459 | 17 | 80307123 | 80331652 | 104 | 13 | 53167 | -1.8297 | 0.96636 | *TEX19* |
| ENSG00000186638 | 9 | 34242379 | 34339198 | 241 | 23 | 33193 | -1.8319 | 0.96652 | *KIF24* |

| | | | | | | | | | |
|---|---|---|---|---|---|---|---|---|---|
| ENSG00000113580 | 5 | 142647496 | 142825077 | 297 | 36 | 50878 | -1.833 | 0.9666 | *NR3C1* |
| ENSG00000104321 | 8 | 72922152 | 72997852 | 348 | 40 | 51515 | -1.8342 | 0.96669 | *TRPA1* |
| ENSG00000255470 | 11 | 18151733 | 18221042 | 350 | 20 | 49942 | -1.8352 | 0.96676 | *RP11-113D6.6* |
| ENSG00000143891 | 2 | 38883052 | 38978379 | 240 | 35 | 35551 | -1.8354 | 0.96677 | *GALM* |
| ENSG00000121964 | 2 | 144685635 | 145100135 | 630 | 73 | 44925 | -1.8368 | 0.96688 | *GTDC1* |
| ENSG00000101361 | 20 | 2622791 | 2649039 | 161 | 23 | 50233 | -1.8399 | 0.96711 | *NOP56* |
| ENSG00000139722 | 12 | 123339882 | 123390991 | 99 | 15 | 38580 | -1.8406 | 0.96716 | *VPS37B* |
| ENSG00000198825 | 10 | 121475609 | 121598652 | 346 | 30 | 38141 | -1.841 | 0.96719 | *INPP5F* |
| ENSG00000137434 | 6 | 10661651 | 10705030 | 31 | 19 | 57321 | -1.8412 | 0.9672 | *C6orf52* |
| ENSG00000188124 | 11 | 6779218 | 6800286 | 110 | 16 | 50868 | -1.843 | 0.96733 | *OR2AG2* |
| ENSG00000163291 | 4 | 79798281 | 79870592 | 195 | 14 | 28144 | -1.8448 | 0.96746 | *PAQR3* |
| ENSG00000132879 | 1 | 11704432 | 11733384 | 96 | 26 | 47121 | -1.845 | 0.96748 | *FBXO44* |
| ENSG00000162972 | 2 | 200810040 | 200883263 | 154 | 23 | 49360 | -1.8452 | 0.9675 | *C2orf47* |
| ENSG00000244694 | 6 | 47835764 | 48046425 | 762 | 73 | 54659 | -1.8476 | 0.96767 | *PTCHD4* |
| ENSG00000146701 | 7 | 75667369 | 75706826 | 116 | 14 | 35326 | -1.8485 | 0.96774 | *MDH2* |
| ENSG00000196704 | 17 | 66233715 | 66263297 | 118 | 20 | 41729 | -1.8487 | 0.96775 | *AMZ2* |
| ENSG00000117501 | 1 | 170894612 | 171043906 | 571 | 54 | 50217 | -1.8501 | 0.96785 | *MROH9* |
| ENSG00000148386 | 9 | 138545168 | 138568268 | 104 | 22 | 48400 | -1.8506 | 0.96789 | *LCN9* |
| ENSG00000111602 | 12 | 56800903 | 56853187 | 126 | 15 | 43690 | -1.8509 | 0.96791 | *TIMELESS* |
| ENSG00000153094 | 2 | 111866955 | 111936024 | 175 | 22 | 49099 | -1.8575 | 0.96838 | *BCL2L11* |
| ENSG00000169710 | 17 | 80026214 | 80066208 | 127 | 26 | 38600 | -1.8598 | 0.96855 | *FASN* |
| ENSG00000133997 | 14 | 71037974 | 71077384 | 169 | 13 | 27067 | -1.8602 | 0.96857 | *MED6* |
| ENSG00000175267 | 16 | 22093859 | 22178287 | 117 | 29 | 40062 | -1.8628 | 0.96876 | *VWA3A* |
| ENSG00000106624 | 7 | 44133960 | 44164161 | 79 | 16 | 48702 | -1.8644 | 0.96887 | *AEBP1* |
| ENSG00000001036 | 6 | 143805948 | 143842827 | 85 | 18 | 41755 | -1.8645 | 0.96888 | *FUCA2* |
| ENSG00000243449 | 4 | 2033689 | 2055697 | 36 | 9 | 37192 | -1.8711 | 0.96933 | *C4orf48* |
| ENSG00000115042 | 2 | 96058469 | 96092364 | 27 | 9 | 36980 | -1.8736 | 0.96951 | *FAHD2A* |
| ENSG00000134504 | 18 | 24024874 | 24247365 | 669 | 124 | 45284 | -1.8742 | 0.96955 | *KCTD1* |
| ENSG00000197037 | 7 | 99204569 | 99240030 | 36 | 14 | 49639 | -1.875 | 0.96961 | *ZSCAN25* |
| ENSG00000168566 | 6 | 7580432 | 7622200 | 138 | 22 | 44526 | -1.8793 | 0.9699 | *SNRNP48* |
| ENSG00000212935 | 21 | 45967673 | 45988643 | 70 | 12 | 51189 | -1.8797 | 0.96993 | *KRTAP10-3* |
| ENSG00000187531 | 17 | 79859815 | 79889199 | 24 | 16 | 46904 | -1.8831 | 0.97016 | *SIRT7* |
| ENSG00000151005 | 4 | 164382257 | 164405047 | 50 | 10 | 51190 | -1.884 | 0.97022 | *TKTL2* |
| ENSG00000169032 | 15 | 66669155 | 66794650 | 425 | 39 | 48953 | -1.8845 | 0.97025 | *MAP2K1* |
| ENSG00000182054 | 15 | 90616277 | 90655736 | 130 | 19 | 38898 | -1.8863 | 0.97038 | *IDH2* |
| ENSG00000160124 | 3 | 122068438 | 122112078 | 145 | 22 | 42861 | -1.8879 | 0.97048 | *CCDC58* |

| | | | | | | | | | |
|---|---|---|---|---|---|---|---|---|---|
| ENSG00000198650 | 16 | 71589563 | 71621033 | 55 | 13 | 41773 | -1.8894 | 0.97058 | *TAT* |
| ENSG00000214265 | 15 | 25190181 | 25255423 | 170 | 21 | 43877 | -1.8906 | 0.97066 | *SNURF* |
| ENSG00000137285 | 6 | 3214495 | 3241964 | 31 | 11 | 41107 | -1.8914 | 0.97071 | *TUBB2B* |
| ENSG00000156931 | 3 | 184519931 | 184780402 | 567 | 42 | 38093 | -1.8939 | 0.97088 | *VPS8* |
| ENSG00000160551 | 17 | 27707482 | 27888922 | 392 | 29 | 41366 | -1.8947 | 0.97093 | *TAOK1* |
| ENSG00000206106 | 21 | 31952424 | 31972716 | 33 | 9 | 54274 | -1.8949 | 0.97094 | *KRTAP22-2* |
| ENSG00000198363 | 8 | 62403116 | 62637155 | 642 | 71 | 45050 | -1.8964 | 0.97104 | *ASPH* |
| ENSG00000160256 | 21 | 46349925 | 46406904 | 228 | 25 | 43406 | -1.8985 | 0.97118 | *FAM207A* |
| ENSG00000212807 | 7 | 143919004 | 143939936 | 1 | 1 | 56198 | -1.8987 | 0.9712 | *OR2A42* |
| ENSG00000155719 | 16 | 21679835 | 21782050 | 149 | 30 | 43178 | -1.8993 | 0.97124 | *OTOA* |
| ENSG00000198300 | 19 | 57311445 | 57362096 | 89 | 18 | 51673 | -1.8998 | 0.97127 | *PEG3* |
| ENSG00000091527 | 3 | 133282574 | 133319105 | 110 | 21 | 42645 | -1.9018 | 0.9714 | *CDV3* |
| ENSG00000125834 | 20 | 2072257 | 2167684 | 282 | 39 | 38395 | -1.902 | 0.97141 | *STK35* |
| ENSG00000103494 | 16 | 53621595 | 53747850 | 325 | 33 | 32657 | -1.9034 | 0.9715 | *RPGRIP1L* |
| ENSG00000182938 | 17 | 72921814 | 72956087 | 83 | 28 | 40800 | -1.9034 | 0.97151 | *OTOP3* |
| ENSG00000101439 | 20 | 23598534 | 23629110 | 122 | 9 | 50689 | -1.9047 | 0.97159 | *CST3* |
| ENSG00000117477 | 1 | 169354108 | 169439907 | 246 | 28 | 42920 | -1.9067 | 0.97172 | *CCDC181* |
| ENSG00000196981 | 3 | 122120715 | 122144882 | 69 | 14 | 51340 | -1.9068 | 0.97173 | *WDR5B* |
| ENSG00000131044 | 20 | 30448505 | 30542764 | 214 | 39 | 46357 | -1.9069 | 0.97173 | *TTLL9* |
| ENSG00000162769 | 1 | 213021597 | 213082705 | 405 | 25 | 46211 | -1.9112 | 0.97201 | *FLVCR1* |
| ENSG00000120705 | 5 | 137831784 | 137888989 | 123 | 20 | 42452 | -1.9115 | 0.97203 | *ETF1* |
| ENSG00000132549 | 8 | 100015494 | 100899808 | 1879 | 58 | 46911 | -1.9116 | 0.97204 | *VPS13B* |
| ENSG00000270099 | 10 | 97610305 | 97694601 | 267 | 34 | 45394 | -1.9128 | 0.97211 | *RP11-248J23.7* |
| ENSG00000131408 | 19 | 50822910 | 50896239 | 156 | 22 | 48088 | -1.9128 | 0.97212 | *NR1H2* |
| ENSG00000147570 | 8 | 66923795 | 67022751 | 328 | 43 | 39613 | -1.9165 | 0.97235 | *DNAJC5B* |
| ENSG00000214787 | 11 | 59958726 | 60020561 | 229 | 22 | 53173 | -1.9173 | 0.9724 | *MS4A4E* |
| ENSG00000116670 | 1 | 11724537 | 11761707 | 135 | 30 | 44285 | -1.9217 | 0.97268 | *MAD2L2* |
| ENSG00000141526 | 17 | 80176273 | 80229005 | 117 | 28 | 38984 | -1.9266 | 0.97298 | *SLC16A3* |
| ENSG00000198668 | 14 | 90852846 | 90884605 | 60 | 14 | 40409 | -1.93 | 0.9732 | *CALM1* |
| ENSG00000117479 | 1 | 169423147 | 169465241 | 101 | 25 | 43563 | -1.9308 | 0.97325 | *SLC19A2* |
| ENSG00000177051 | 19 | 46203887 | 46244162 | 79 | 21 | 47163 | -1.9326 | 0.97336 | *FBXO46* |
| ENSG00000127511 | 19 | 16930211 | 17001164 | 176 | 28 | 48404 | -1.9332 | 0.97339 | *SIN3B* |
| ENSG00000128524 | 7 | 128492880 | 128515898 | 55 | 15 | 36342 | -1.9364 | 0.97359 | *ATP6V1F* |
| ENSG00000205020 | 17 | 34528310 | 34550280 | 1 | 1 | 43140 | -1.9366 | 0.9736 | *CCL4L1* |
| ENSG00000181927 | 11 | 55395834 | 55416772 | 11 | 6 | 44072 | -1.9372 | 0.97364 | *OR4P4* |
| ENSG00000151176 | 12 | 113786371 | 113837203 | 104 | 16 | 52175 | -1.9394 | 0.97378 | *PLBD2* |

| | | | | | | | | | |
|---|---|---|---|---|---|---|---|---|---|
| ENSG00000124570 | 6 | 2938393 | 2982090 | 193 | 29 | 45237 | -1.9397 | 0.97379 | *SERPINB6* |
| ENSG00000169085 | 8 | 67362238 | 67440759 | 242 | 39 | 41949 | -1.9422 | 0.97394 | *C8orf46* |
| ENSG00000196542 | 3 | 161052580 | 161100668 | 218 | 30 | 46639 | -1.9423 | 0.97395 | *SPTSSB* |
| ENSG00000268852 | 17 | 80204131 | 80226080 | 21 | 11 | 47939 | -1.9427 | 0.97397 | *AC132872.2* |
| ENSG00000166920 | 15 | 45712727 | 45750959 | 55 | 12 | 34261 | -1.9429 | 0.97398 | *C15orf48* |
| ENSG00000128585 | 7 | 130784855 | 131191395 | 1101 | 142 | 46685 | -1.9481 | 0.9743 | *MKLN1* |
| ENSG00000176018 | 5 | 89801428 | 89835401 | 87 | 12 | 49770 | -1.9509 | 0.97446 | *LYSMD3* |
| ENSG00000108830 | 17 | 41167258 | 41194057 | 37 | 5 | 48353 | -1.9508 | 0.97446 | *RND2* |
| ENSG00000205089 | 5 | 132073137 | 132099856 | 10 | 6 | 49903 | -1.9542 | 0.97466 | *CCNI2* |
| ENSG00000106078 | 7 | 51073909 | 51394515 | 745 | 91 | 43375 | -1.9546 | 0.97468 | *COBL* |
| ENSG00000142530 | 19 | 50960042 | 50990010 | 93 | 20 | 40055 | -1.9574 | 0.97485 | *FAM71E1* |
| ENSG00000099953 | 22 | 24100413 | 24136503 | 106 | 23 | 47805 | -1.9574 | 0.97485 | *MMP11* |
| ENSG00000196083 | 3 | 190221840 | 190385843 | 534 | 93 | 41422 | -1.958 | 0.97488 | *IL1RAP* |
| ENSG00000198382 | 11 | 75516212 | 75864239 | 649 | 51 | 34861 | -1.9586 | 0.97492 | *UVRAG* |
| ENSG00000205442 | 9 | 24532950 | 24555944 | 73 | 7 | 51788 | -1.9588 | 0.97493 | *IZUMO3* |
| ENSG00000139193 | 12 | 6544033 | 6570884 | 95 | 22 | 49940 | -1.9587 | 0.97493 | *CD27* |
| ENSG00000267110 | 19 | 55656125 | 55682039 | 87 | 19 | 32321 | -1.96 | 0.975 | *CTD-2587H24.4* |
| ENSG00000181458 | 3 | 100201463 | 100306288 | 313 | 24 | 38508 | -1.9624 | 0.97514 | *TMEM45A* |
| ENSG00000162542 | 1 | 19998706 | 20136438 | 471 | 74 | 47191 | -1.9634 | 0.9752 | *TMCO4* |
| ENSG00000148600 | 10 | 85944410 | 85989377 | 150 | 23 | 45385 | -1.9663 | 0.97537 | *CDHR1* |
| ENSG00000256980 | 6 | 73923267 | 73945493 | 73 | 13 | 50413 | -1.9678 | 0.97545 | *KHDC1L* |
| ENSG00000135063 | 9 | 71929488 | 72017371 | 368 | 46 | 50740 | -1.9693 | 0.97554 | *FAM189A2* |
| ENSG00000101213 | 20 | 62149778 | 62178723 | 103 | 25 | 35182 | -1.9698 | 0.97557 | *PTK6* |
| ENSG00000080493 | 4 | 72043003 | 72447804 | 742 | 83 | 31467 | -1.9724 | 0.97572 | *SLC4A4* |
| ENSG00000214782 | 11 | 60486789 | 60521685 | 83 | 17 | 48250 | -1.9734 | 0.97578 | *MS4A18* |
| ENSG00000197063 | 17 | 79866146 | 79895590 | 24 | 15 | 47473 | -1.9749 | 0.97586 | *MAFG* |
| ENSG00000100697 | 14 | 95542565 | 95634347 | 163 | 24 | 42871 | -1.9758 | 0.97591 | *DICER1* |
| ENSG00000160345 | 9 | 138377027 | 138403580 | 95 | 23 | 43015 | -1.9765 | 0.97595 | *C9orf116* |
| ENSG00000197858 | 8 | 145127493 | 145151119 | 41 | 5 | 31038 | -1.9788 | 0.97608 | *GPAA1* |
| ENSG00000182652 | 14 | 20205587 | 20226528 | 8 | 4 | 43564 | -1.9793 | 0.97611 | *OR4Q3* |
| ENSG00000180016 | 17 | 3290398 | 3311704 | 57 | 12 | 33430 | -1.9794 | 0.97612 | *OR1E1* |
| ENSG00000259431 | 14 | 24015216 | 24039480 | 36 | 12 | 44176 | -1.981 | 0.9762 | *THTPA* |
| ENSG00000107957 | 10 | 105338285 | 105625301 | 792 | 114 | 44866 | -1.9834 | 0.97634 | *SH3PXD2A* |
| ENSG00000267913 | 21 | 45577818 | 45603580 | 34 | 20 | 43684 | -1.9849 | 0.97642 | *AP001055.1* |
| ENSG00000176294 | 14 | 20177174 | 20306531 | 48 | 22 | 43352 | -1.9858 | 0.97647 | *OR4N2* |
| ENSG00000177186 | 1 | 248476894 | 248497899 | 108 | 4 | 55719 | -1.9885 | 0.97662 | *OR2M7* |

| | | | | | | | | | |
|---|---|---|---|---|---|---|---|---|---|
| ENSG00000162460 | 1 | 16058917 | 16084477 | 71 | 15 | 47144 | -1.9893 | 0.97666 | *TMEM82* |
| ENSG00000177174 | 1 | 248502077 | 248523015 | 162 | 9 | 49627 | -1.9893 | 0.97667 | *OR14C36* |
| ENSG00000205174 | 7 | 108514032 | 108534644 | 73 | 13 | 44734 | -1.993 | 0.97687 | *C7orf66* |
| ENSG00000168811 | 3 | 159696537 | 159723806 | 86 | 17 | 40420 | -1.9939 | 0.97692 | *IL12A* |
| ENSG00000196533 | 1 | 206228872 | 206298647 | 78 | 20 | 46405 | -1.9966 | 0.97707 | *C1orf186* |
| ENSG00000111731 | 12 | 22591517 | 22707480 | 202 | 13 | 31679 | -1.9969 | 0.97708 | *C2CD5* |
| ENSG00000163625 | 4 | 85580704 | 85897544 | 423 | 44 | 48518 | -1.9981 | 0.97715 | *WDFY3* |
| ENSG00000104375 | 8 | 99403631 | 99965055 | 941 | 86 | 53165 | -2.0036 | 0.97744 | *STK3* |
| ENSG00000105643 | 19 | 18101941 | 18134911 | 125 | 27 | 47154 | -2.0043 | 0.97748 | *ARRDC2* |
| ENSG00000139370 | 12 | 129267739 | 129318528 | 235 | 24 | 48966 | -2.0075 | 0.97765 | *SLC15A4* |
| ENSG00000116663 | 1 | 11714181 | 11744411 | 117 | 24 | 46851 | -2.0081 | 0.97769 | *FBXO6* |
| ENSG00000167193 | 17 | 1313983 | 1376456 | 246 | 31 | 47291 | -2.0087 | 0.97771 | *CRK* |
| ENSG00000138709 | 4 | 128972423 | 129154086 | 420 | 22 | 40570 | -2.0102 | 0.97779 | *LARP1B* |
| ENSG00000269035 | 19 | 17537420 | 17581613 | 123 | 22 | 45778 | -2.0108 | 0.97783 | *CTD-2521M24.10* |
| ENSG00000131944 | 19 | 33453115 | 33479128 | 108 | 27 | 50219 | -2.011 | 0.97784 | *C19orf40* |
| ENSG00000060566 | 19 | 4143598 | 4183051 | 105 | 22 | 42094 | -2.0153 | 0.97806 | *CREB3L3* |
| ENSG00000241186 | 3 | 46606045 | 46678033 | 350 | 42 | 46803 | -2.0164 | 0.97812 | *TDGF1* |
| ENSG00000213967 | 19 | 24087678 | 24137961 | 175 | 23 | 53184 | -2.0177 | 0.97819 | *ZNF726* |
| ENSG00000113657 | 5 | 146760374 | 146899619 | 209 | 44 | 50038 | -2.0187 | 0.97824 | *DPYSL3* |
| ENSG00000165275 | 9 | 37743804 | 37788969 | 148 | 21 | 54451 | -2.022 | 0.97841 | *TRMT10B* |
| ENSG00000248905 | 15 | 33047747 | 33496897 | 2237 | 214 | 46784 | -2.0223 | 0.97843 | *FMN1* |
| ENSG00000186260 | 16 | 14155178 | 14370630 | 428 | 34 | 30023 | -2.0223 | 0.97843 | *MKL2* |
| ENSG00000171695 | 20 | 62704733 | 62725712 | 49 | 15 | 49494 | -2.0267 | 0.97865 | *C20orf201* |
| ENSG00000267954 | 22 | 24114467 | 24136145 | 52 | 11 | 48350 | -2.0272 | 0.97868 | *AP000349.1* |
| ENSG00000183281 | 2 | 87219682 | 87258975 | 4 | 2 | 27658 | -2.0284 | 0.97874 | *PLGLB1* |
| ENSG00000133985 | 14 | 71098504 | 71152077 | 177 | 48 | 47148 | -2.0315 | 0.9789 | *TTC9* |
| ENSG00000172809 | 17 | 72189721 | 72216794 | 116 | 30 | 45024 | -2.0345 | 0.97905 | *RPL38* |
| ENSG00000196944 | 1 | 248514883 | 248535929 | 159 | 9 | 44244 | -2.0356 | 0.9791 | *OR2T4* |
| ENSG00000161202 | 3 | 183863176 | 183901398 | 85 | 17 | 52000 | -2.0364 | 0.97914 | *DVL3* |
| ENSG00000121005 | 8 | 75886750 | 75956793 | 267 | 33 | 43488 | -2.0367 | 0.97916 | *CRISPLD1* |
| ENSG00000150637 | 18 | 67488394 | 67639039 | 464 | 71 | 46453 | -2.037 | 0.97917 | *CD226* |
| ENSG00000051620 | 6 | 138714668 | 138753334 | 97 | 13 | 32598 | -2.0377 | 0.97921 | *HEBP2* |
| ENSG00000187170 | 1 | 152670636 | 152691967 | 63 | 10 | 39624 | -2.04 | 0.97932 | *LCE4A* |
| ENSG00000151025 | 10 | 25453991 | 25901155 | 1939 | 109 | 32003 | -2.0399 | 0.97932 | *GPR158* |
| ENSG00000138078 | 2 | 44533420 | 44599001 | 197 | 24 | 44572 | -2.04 | 0.97933 | *PREPL* |
| ENSG00000186001 | 3 | 197508097 | 197625307 | 292 | 42 | 39009 | -2.0499 | 0.97981 | *LRCH3* |

| | | | | | | | | | |
|---|---|---|---|---|---|---|---|---|---|
| ENSG00000089169 | 12 | 112998184 | 113346686 | 699 | 107 | 44349 | -2.0505 | 0.97984 | *RPH3A* |
| ENSG00000103995 | 15 | 48995125 | 49113343 | 218 | 33 | 39685 | -2.0507 | 0.97985 | *CEP152* |
| ENSG00000089199 | 20 | 5882076 | 5916007 | 163 | 22 | 45228 | -2.051 | 0.97987 | *CHGB* |
| ENSG00000139352 | 12 | 103341464 | 103364294 | 63 | 19 | 41784 | -2.0529 | 0.97996 | *ASCL1* |
| ENSG00000112406 | 6 | 139446249 | 139511939 | 100 | 13 | 48886 | -2.0545 | 0.98004 | *HECA* |
| ENSG00000138829 | 5 | 127583601 | 128004878 | 1365 | 71 | 43701 | -2.0555 | 0.98009 | *FBN2* |
| ENSG00000198517 | 7 | 1560350 | 1592679 | 146 | 10 | 55822 | -2.0558 | 0.9801 | *MAFK* |
| ENSG00000118046 | 19 | 1179406 | 1238428 | 196 | 44 | 48122 | -2.0573 | 0.98017 | *STK11* |
| ENSG00000127948 | 7 | 75518518 | 75626173 | 214 | 44 | 43256 | -2.0611 | 0.98035 | *POR* |
| ENSG00000154553 | 4 | 186412851 | 186466766 | 150 | 24 | 35136 | -2.0618 | 0.98039 | *PDLIM3* |
| ENSG00000037474 | 5 | 6589352 | 6643404 | 284 | 31 | 48536 | -2.0629 | 0.98044 | *NSUN2* |
| ENSG00000133794 | 11 | 13288199 | 13418813 | 380 | 58 | 40498 | -2.0646 | 0.98052 | *ARNTL* |
| ENSG00000101222 | 20 | 3748151 | 3772095 | 76 | 20 | 50159 | -2.0676 | 0.98066 | *SPEF1* |
| ENSG00000130475 | 19 | 17848527 | 17909377 | 229 | 35 | 37643 | -2.069 | 0.98073 | *FCHO1* |
| ENSG00000129991 | 19 | 55653137 | 55679141 | 82 | 19 | 32417 | -2.0706 | 0.9808 | *TNNI3* |
| ENSG00000087470 | 12 | 32822134 | 32908486 | 403 | 11 | 51365 | -2.0768 | 0.98109 | *DNM1L* |
| ENSG00000138073 | 2 | 27343624 | 27367543 | 43 | 11 | 49763 | -2.08 | 0.98124 | *PREB* |
| ENSG00000243501 | 6 | 73909469 | 73982919 | 314 | 32 | 50437 | -2.0843 | 0.98143 | *RP11-257K9.8* |
| ENSG00000166800 | 11 | 18467371 | 18511147 | 139 | 28 | 49350 | -2.0858 | 0.9815 | *LDHAL6A* |
| ENSG00000106100 | 7 | 30454143 | 30528400 | 187 | 27 | 43362 | -2.0868 | 0.98155 | *NOD1* |
| ENSG00000203734 | 6 | 139107063 | 139235207 | 472 | 73 | 41912 | -2.0886 | 0.98163 | *ECT2L* |
| ENSG00000269709 | 1 | 48559306 | 48579878 | 47 | 14 | 52377 | -2.0895 | 0.98167 | *AL109659.1* |
| ENSG00000145248 | 4 | 48475360 | 48501213 | 85 | 12 | 54099 | -2.0902 | 0.9817 | *SLC10A4* |
| ENSG00000134905 | 13 | 111283759 | 111375950 | 440 | 38 | 41815 | -2.0902 | 0.9817 | *CARS2* |
| ENSG00000100503 | 14 | 51176481 | 51307839 | 340 | 60 | 47616 | -2.0904 | 0.98171 | *NIN* |
| ENSG00000269964 | 6 | 78390375 | 78646691 | 965 | 56 | 45369 | -2.0914 | 0.98176 | *MEI4* |
| ENSG00000109689 | 4 | 26849300 | 27037003 | 458 | 59 | 48858 | -2.092 | 0.98178 | *STIM2* |
| ENSG00000158748 | 1 | 19981780 | 20016055 | 116 | 17 | 46578 | -2.0982 | 0.98206 | *HTR6* |
| ENSG00000077235 | 16 | 27460876 | 27571234 | 164 | 20 | 48407 | -2.0997 | 0.98212 | *GTF3C1* |
| ENSG00000113356 | 5 | 89757565 | 89820370 | 141 | 23 | 50914 | -2.1123 | 0.98267 | *POLR3G* |
| ENSG00000108270 | 17 | 35296175 | 35424171 | 206 | 30 | 41750 | -2.1164 | 0.98285 | *AATF* |
| ENSG00000184350 | 11 | 3238928 | 3263616 | 127 | 26 | 51908 | -2.1174 | 0.98289 | *MRGPRE* |
| ENSG00000100721 | 14 | 96166304 | 96190533 | 75 | 11 | 27655 | -2.1186 | 0.98294 | *TCL1A* |
| ENSG00000115841 | 2 | 38140330 | 38304285 | 487 | 43 | 37235 | -2.1221 | 0.98308 | *RMDN2* |
| ENSG00000109180 | 4 | 48797229 | 48873834 | 110 | 17 | 51212 | -2.1228 | 0.98312 | *OCIAD1* |
| ENSG00000099995 | 22 | 30717977 | 30762936 | 134 | 15 | 49700 | -2.1228 | 0.98312 | *SF3A1* |

| | | | | | | | | | |
|---|---|---|---|---|---|---|---|---|---|
| ENSG00000101187 | 20 | 61263797 | 61327137 | 234 | 58 | 47964 | -2.1232 | 0.98313 | *SLCO4A1* |
| ENSG00000184378 | 3 | 169474709 | 169497683 | 48 | 9 | 42679 | -2.1237 | 0.98315 | *ACTRT3* |
| ENSG00000139151 | 12 | 18826103 | 18900991 | 279 | 38 | 43783 | -2.1305 | 0.98343 | *PLCZ1* |
| ENSG00000143322 | 1 | 179058462 | 179208819 | 416 | 33 | 55716 | -2.1324 | 0.98351 | *ABL2* |
| ENSG00000175065 | 18 | 28946740 | 29004875 | 135 | 17 | 45456 | -2.1326 | 0.98352 | *DSG4* |
| ENSG00000181991 | 15 | 89000684 | 89032222 | 15 | 10 | 47223 | -2.1338 | 0.98357 | *MRPS11* |
| ENSG00000172175 | 18 | 56328618 | 56427371 | 283 | 30 | 35150 | -2.1374 | 0.98372 | *MALT1* |
| ENSG00000127419 | 4 | 916175 | 962444 | 148 | 32 | 51254 | -2.1392 | 0.98379 | *TMEM175* |
| ENSG00000114023 | 3 | 122093023 | 122141181 | 163 | 24 | 45885 | -2.1459 | 0.98406 | *FAM162A* |
| ENSG00000239672 | 17 | 49220897 | 49249789 | 114 | 19 | 49652 | -2.1484 | 0.98416 | *NME1* |
| ENSG00000156052 | 9 | 80321003 | 80656374 | 628 | 55 | 43680 | -2.1506 | 0.98424 | *GNAQ* |
| ENSG00000157335 | 16 | 70197225 | 70231264 | 30 | 14 | 43055 | -2.1511 | 0.98426 | *CLEC18C* |
| ENSG00000101160 | 20 | 57560240 | 57592302 | 104 | 17 | 54942 | -2.151 | 0.98426 | *CTSZ* |
| ENSG00000047578 | 16 | 27551454 | 27801690 | 367 | 34 | 49721 | -2.1589 | 0.98457 | *KIAA0556* |
| ENSG00000173889 | 3 | 169794520 | 169909537 | 210 | 23 | 52461 | -2.1597 | 0.9846 | *PHC3* |
| ENSG00000080603 | 16 | 30699530 | 30765602 | 47 | 13 | 35311 | -2.1616 | 0.98467 | *SRCAP* |
| ENSG00000036565 | 8 | 19992366 | 20050717 | 240 | 37 | 49443 | -2.1675 | 0.9849 | *SLC18A1* |
| ENSG00000179295 | 12 | 112846155 | 112957717 | 113 | 18 | 39067 | -2.1675 | 0.9849 | *PTPN11* |
| ENSG00000147576 | 8 | 67332420 | 67393836 | 248 | 33 | 39575 | -2.171 | 0.98503 | *ADHFE1* |
| ENSG00000171863 | 2 | 3612795 | 3638509 | 118 | 25 | 50549 | -2.1726 | 0.9851 | *RPS7* |
| ENSG00000153904 | 1 | 85774164 | 86053933 | 887 | 92 | 49706 | -2.1735 | 0.98513 | *DDAH1* |
| ENSG00000214167 | 17 | 65017509 | 65038198 | 62 | 20 | 45704 | -2.1746 | 0.98517 | *AC005544.1* |
| ENSG00000139985 | 14 | 70908874 | 70936622 | 122 | 21 | 37405 | -2.1779 | 0.98529 | *ADAM21* |
| ENSG00000107789 | 10 | 89254632 | 89323217 | 155 | 28 | 48961 | -2.1782 | 0.9853 | *MINPP1* |
| ENSG00000189045 | 5 | 74897284 | 74977671 | 230 | 31 | 33971 | -2.1799 | 0.98537 | *ANKDD1B* |
| ENSG00000128513 | 7 | 124452440 | 124580037 | 444 | 16 | 52977 | -2.1828 | 0.98547 | *POT1* |
| ENSG00000167646 | 19 | 55660031 | 55688090 | 132 | 19 | 30055 | -2.1833 | 0.98549 | *DNAAF3* |
| ENSG00000135940 | 2 | 98252503 | 98274846 | 36 | 10 | 31995 | -2.1867 | 0.98562 | *COX5B* |
| ENSG00000131236 | 1 | 40495905 | 40548321 | 185 | 11 | 46692 | -2.188 | 0.98567 | *CAP1* |
| ENSG00000198129 | 8 | 7343368 | 7376833 | 4 | 2 | 42697 | -2.1922 | 0.98582 | *DEFB107B* |
| ENSG00000136718 | 2 | 131089798 | 131115383 | 33 | 13 | 37972 | -2.1927 | 0.98583 | *IMP4* |
| ENSG00000140575 | 15 | 90921450 | 91055475 | 439 | 30 | 32444 | -2.2018 | 0.98616 | *IQGAP1* |
| ENSG00000100156 | 22 | 38464141 | 38490100 | 54 | 15 | 49941 | -2.2023 | 0.98618 | *SLC16A8* |
| ENSG00000184292 | 1 | 59031099 | 59053166 | 131 | 15 | 46385 | -2.2039 | 0.98624 | *TACSTD2* |
| ENSG00000139192 | 12 | 6550856 | 6585683 | 155 | 25 | 49960 | -2.2081 | 0.98638 | *TAPBPL* |
| ENSG00000165714 | 12 | 12500013 | 12629840 | 457 | 52 | 38931 | -2.2088 | 0.98641 | *LOH12CR1* |

| | | | | | | | | | |
|---|---|---|---|---|---|---|---|---|---|
| ENSG00000171940 | 20 | 52173604 | 52236446 | 174 | 37 | 45219 | -2.2096 | 0.98644 | *ZNF217* |
| ENSG00000122140 | 9 | 138381830 | 138406519 | 86 | 19 | 44465 | -2.2121 | 0.98652 | *MRPS2* |
| ENSG00000221900 | 7 | 53093349 | 53114617 | 105 | 19 | 41864 | -2.2123 | 0.98653 | *POM121L12* |
| ENSG00000091513 | 3 | 133454800 | 133507850 | 195 | 28 | 51559 | -2.2154 | 0.98663 | *TF* |
| ENSG00000181867 | 5 | 121177650 | 121198519 | 65 | 20 | 40802 | -2.2215 | 0.98684 | *FTMT* |
| ENSG00000115073 | 2 | 98262431 | 98290570 | 45 | 11 | 26149 | -2.2229 | 0.98689 | *ACTR1B* |
| ENSG00000110492 | 11 | 46392306 | 46415375 | 20 | 7 | 41147 | -2.2269 | 0.98702 | *MDK* |
| ENSG00000164975 | 9 | 15412702 | 15475951 | 208 | 25 | 34135 | -2.228 | 0.98706 | *SNAPC3* |
| ENSG00000072786 | 5 | 171459077 | 171625390 | 683 | 85 | 40356 | -2.2297 | 0.98712 | *STK10* |
| ENSG00000126709 | 1 | 27982572 | 28008729 | 34 | 17 | 41675 | -2.2301 | 0.98713 | *IFI6* |
| ENSG00000131400 | 19 | 50851734 | 50879087 | 70 | 9 | 54112 | -2.232 | 0.98719 | *NAPSA* |
| ENSG00000186143 | 2 | 27349723 | 27372278 | 47 | 10 | 45462 | -2.2324 | 0.98721 | *C2orf53* |
| ENSG00000132640 | 20 | 11861371 | 11917257 | 157 | 14 | 48328 | -2.2403 | 0.98746 | *BTBD3* |
| ENSG00000114473 | 3 | 197605946 | 197697013 | 279 | 29 | 37351 | -2.2444 | 0.9876 | *IQCG* |
| ENSG00000167037 | 22 | 25192236 | 25333545 | 608 | 108 | 47155 | -2.2453 | 0.98763 | *SGSM1* |
| ENSG00000142794 | 1 | 21756621 | 21821498 | 260 | 27 | 43285 | -2.2483 | 0.98772 | *NBPF3* |
| ENSG00000077935 | 22 | 45729944 | 45819500 | 278 | 17 | 30636 | -2.2494 | 0.98776 | *SMC1B* |
| ENSG00000071203 | 11 | 60250251 | 60284903 | 210 | 16 | 51067 | -2.2501 | 0.98778 | *MS4A12* |
| ENSG00000135409 | 12 | 53807639 | 53835318 | 53 | 6 | 43537 | -2.2567 | 0.98799 | *AMHR2* |
| ENSG00000213995 | 13 | 111257881 | 111302340 | 222 | 25 | 43416 | -2.263 | 0.98818 | *CARKD* |
| ENSG00000107438 | 10 | 96987329 | 97060781 | 278 | 38 | 44115 | -2.2647 | 0.98823 | *PDLIM1* |
| ENSG00000204752 | 19 | 7001514 | 7023338 | 96 | 15 | 51955 | -2.2654 | 0.98826 | *AC025278.1* |
| ENSG00000249967 | 10 | 99334131 | 99443667 | 278 | 37 | 44688 | -2.2683 | 0.98834 | *PI4K2A* |
| ENSG00000136295 | 7 | 2661585 | 2714436 | 159 | 33 | 46101 | -2.2695 | 0.98838 | *TTYH3* |
| ENSG00000181333 | 11 | 93744527 | 93856917 | 271 | 34 | 44498 | -2.2719 | 0.98845 | *HEPHL1* |
| ENSG00000161542 | 17 | 74295567 | 74390602 | 293 | 44 | 36149 | -2.2752 | 0.98855 | *PRPSAP1* |
| ENSG00000139220 | 12 | 81642045 | 82163332 | 1412 | 125 | 44691 | -2.2754 | 0.98856 | *PPFIA2* |
| ENSG00000204979 | 11 | 60272886 | 60320194 | 199 | 24 | 41686 | -2.277 | 0.98861 | *MS4A13* |
| ENSG00000107372 | 9 | 74956341 | 74990163 | 92 | 11 | 46370 | -2.2777 | 0.98863 | *ZFAND5* |
| ENSG00000145723 | 5 | 102411704 | 102465855 | 219 | 14 | 52952 | -2.2827 | 0.98878 | *GIN1* |
| ENSG00000109536 | 4 | 190851943 | 190894359 | 118 | 6 | 45159 | -2.2853 | 0.98885 | *FRG1* |
| ENSG00000085978 | 2 | 234108697 | 234214320 | 335 | 40 | 46733 | -2.2913 | 0.98903 | *ATG16L1* |
| ENSG00000197893 | 10 | 115338475 | 115433886 | 372 | 40 | 47163 | -2.2928 | 0.98907 | *NRAP* |
| ENSG00000147852 | 9 | 2611834 | 2670053 | 207 | 40 | 42188 | -2.2946 | 0.98912 | *VLDLR* |
| ENSG00000143801 | 1 | 227047885 | 227093806 | 127 | 19 | 49555 | -2.2967 | 0.98918 | *PSEN2* |
| ENSG00000141854 | 19 | 14173348 | 14195874 | 64 | 12 | 45704 | -2.2971 | 0.98919 | *hsa-mir-1199* |

| | | | | | | | | | |
|---|---|---|---|---|---|---|---|---|---|
| ENSG00000157978 | 1 | 25860071 | 25905377 | 116 | 20 | 39364 | -2.2988 | 0.98924 | *LDLRAP1* |
| ENSG00000134007 | 14 | 70979075 | 71011732 | 108 | 16 | 32969 | -2.3043 | 0.9894 | *ADAM20* |
| ENSG00000198104 | 1 | 248540910 | 248561836 | 109 | 8 | 24772 | -2.3054 | 0.98943 | *OR2T6* |
| ENSG00000154065 | 18 | 21168890 | 21252849 | 157 | 34 | 42326 | -2.3097 | 0.98955 | *ANKRD29* |
| ENSG00000270617 | 7 | 43896157 | 43956188 | 183 | 24 | 52361 | -2.3111 | 0.98959 | *URGCP-MRPS24* |
| ENSG00000187173 | 1 | 152660840 | 152681918 | 81 | 11 | 35873 | -2.32 | 0.98983 | *LCE2A* |
| ENSG00000085274 | 3 | 169480619 | 169517504 | 74 | 10 | 42456 | -2.3223 | 0.98989 | *MYNN* |
| ENSG00000268365 | 1 | 240619183 | 240653504 | 137 | 25 | 45690 | -2.3269 | 0.99002 | *AL646016.1* |
| ENSG00000155252 | 10 | 99334131 | 99446191 | 287 | 38 | 45067 | -2.3277 | 0.99004 | *PI4K2A* |
| ENSG00000241945 | 21 | 45517171 | 45561063 | 42 | 16 | 41295 | -2.331 | 0.99012 | *PWP2* |
| ENSG00000086200 | 5 | 61689799 | 61934409 | 521 | 35 | 53863 | -2.3313 | 0.99013 | *IPO11* |
| ENSG00000159496 | 22 | 24020323 | 24051363 | 107 | 16 | 53503 | -2.3359 | 0.99025 | *RGL4* |
| ENSG00000004700 | 12 | 21611845 | 21664603 | 266 | 34 | 50981 | -2.3385 | 0.99032 | *RECQL* |
| ENSG00000181444 | 7 | 149451271 | 149480568 | 105 | 18 | 34829 | -2.3389 | 0.99033 | *ZNF467* |
| ENSG00000198915 | 10 | 43679983 | 43772367 | 356 | 40 | 41214 | -2.3394 | 0.99034 | *RASGEF1A* |
| ENSG00000258643 | 14 | 23766044 | 23804578 | 87 | 23 | 45910 | -2.34 | 0.99036 | *BCL2L2-PABPN1* |
| ENSG00000170255 | 11 | 18945360 | 18971054 | 31 | 11 | 38316 | -2.3405 | 0.99037 | *MRGPRX1* |
| ENSG00000268344 | 13 | 26442422 | 26462679 | 30 | 7 | 50438 | -2.3413 | 0.99039 | *AL138815.2* |
| ENSG00000174982 | 11 | 55408380 | 55429315 | 14 | 8 | 44187 | -2.3437 | 0.99045 | *OR4S2* |
| ENSG00000183813 | 3 | 32983066 | 33007841 | 53 | 9 | 32059 | -2.3444 | 0.99047 | *CCR4* |
| ENSG00000205352 | 12 | 53825389 | 53850429 | 36 | 8 | 40529 | -2.3449 | 0.99048 | *PRR13* |
| ENSG00000173064 | 12 | 112587992 | 112829896 | 221 | 29 | 40506 | -2.3495 | 0.9906 | *HECTD4* |
| ENSG00000122574 | 7 | 29836102 | 29966682 | 493 | 63 | 51137 | -2.35 | 0.99061 | *WIPF3* |
| ENSG00000159322 | 15 | 73033710 | 73088187 | 127 | 19 | 38641 | -2.3514 | 0.99065 | *ADPGK* |
| ENSG00000249008 | 4 | 178153693 | 178179927 | 79 | 20 | 45692 | -2.352 | 0.99066 | *RP11-487E13.1* |
| ENSG00000178695 | 13 | 77444312 | 77470540 | 79 | 23 | 41676 | -2.352 | 0.99066 | *KCTD12* |
| ENSG00000188800 | 1 | 40701619 | 40727363 | 39 | 14 | 48800 | -2.357 | 0.99079 | *TMCO2* |
| ENSG00000170955 | 11 | 6330176 | 6351877 | 108 | 22 | 45325 | -2.3601 | 0.99086 | *PRKCDBP* |
| ENSG00000106608 | 7 | 43905493 | 43976010 | 226 | 26 | 53870 | -2.3615 | 0.9909 | *URGCP* |
| ENSG00000155130 | 6 | 114168541 | 114194648 | 54 | 17 | 47272 | -2.3625 | 0.99092 | *MARCKS* |
| ENSG00000133119 | 13 | 34382186 | 34550695 | 432 | 42 | 25920 | -2.3713 | 0.99114 | *RFC3* |
| ENSG00000185917 | 21 | 37396839 | 37461687 | 262 | 28 | 54384 | -2.3781 | 0.9913 | *SETD4* |
| ENSG00000101435 | 20 | 23535369 | 23559386 | 113 | 7 | 52290 | -2.3786 | 0.99131 | *CST9L* |
| ENSG00000183150 | 12 | 12803825 | 12859141 | 147 | 31 | 50906 | -2.3789 | 0.99132 | *GPR19* |
| ENSG00000066583 | 5 | 128420444 | 128459721 | 158 | 24 | 36271 | -2.3886 | 0.99154 | *ISOC1* |
| ENSG00000071242 | 6 | 166812852 | 167329939 | 1829 | 224 | 42700 | -2.389 | 0.99155 | *RPS6KA2* |

| | | | | | | | | | |
|---|---|---|---|---|---|---|---|---|---|
| ENSG00000183921 | 16 | 22167618 | 22213075 | 75 | 15 | 37538 | -2.396 | 0.99171 | *SDR42E2* |
| ENSG00000099800 | 19 | 2415622 | 2437892 | 103 | 16 | 45013 | -2.3959 | 0.99171 | *TIMM13* |
| ENSG00000034693 | 6 | 143761944 | 143821147 | 119 | 14 | 45985 | -2.3983 | 0.99177 | *PEX3* |
| ENSG00000144355 | 2 | 172939468 | 172964405 | 57 | 19 | 49869 | -2.402 | 0.99185 | *DLX1* |
| ENSG00000171450 | 2 | 219814377 | 219836876 | 52 | 9 | 32424 | -2.4061 | 0.99194 | *CDK5R2* |
| ENSG00000173088 | 10 | 97657360 | 97708480 | 192 | 26 | 41790 | -2.412 | 0.99207 | *C10orf131* |
| ENSG00000197879 | 17 | 1357392 | 1406106 | 192 | 50 | 49454 | -2.4156 | 0.99215 | *MYO1C* |
| ENSG00000152348 | 5 | 81257844 | 81582676 | 635 | 63 | 44105 | -2.4196 | 0.99223 | *ATG10* |
| ENSG00000164588 | 5 | 45249349 | 45706253 | 652 | 58 | 48033 | -2.4264 | 0.99238 | *HCN1* |
| ENSG00000110218 | 11 | 93852094 | 93925138 | 231 | 35 | 53454 | -2.4338 | 0.99253 | *PANX1* |
| ENSG00000273291 | 3 | 42840938 | 43107363 | 657 | 69 | 42853 | -2.4361 | 0.99258 | *KRBOX1* |
| ENSG00000159958 | 22 | 42311045 | 42332822 | 63 | 12 | 42623 | -2.4383 | 0.99262 | *TNFRSF13C* |
| ENSG00000057149 | 18 | 61312431 | 61339197 | 41 | 11 | 49776 | -2.4422 | 0.9927 | *SERPINB3* |
| ENSG00000113369 | 5 | 90654541 | 90689176 | 56 | 10 | 31536 | -2.4443 | 0.99274 | *ARRDC3* |
| ENSG00000134453 | 10 | 6120950 | 6169420 | 195 | 40 | 46977 | -2.4499 | 0.99286 | *RBM17* |
| ENSG00000214360 | 5 | 171611176 | 171640456 | 89 | 23 | 47292 | -2.4506 | 0.99287 | *EFCAB9* |
| ENSG00000109790 | 4 | 39036659 | 39138477 | 378 | 18 | 51920 | -2.4631 | 0.99311 | *KLHL5* |
| ENSG00000268856 | 21 | 46482796 | 46502927 | 88 | 21 | 53313 | -2.4633 | 0.99312 | *AP001579.1* |
| ENSG00000099956 | 22 | 24119150 | 24186703 | 383 | 26 | 48091 | -2.4674 | 0.9932 | *SMARCB1* |
| ENSG00000178074 | 2 | 200765979 | 200830658 | 159 | 22 | 51234 | -2.4692 | 0.99323 | *C2orf69* |
| ENSG00000175874 | 2 | 101952013 | 102014057 | 188 | 19 | 45268 | -2.4714 | 0.99327 | *CREG2* |
| ENSG00000156345 | 9 | 90571356 | 90599668 | 52 | 15 | 49497 | -2.4811 | 0.99345 | *CDK20* |
| ENSG00000196648 | 15 | 83088710 | 83118085 | 1 | 1 | 42839 | -2.4838 | 0.9935 | *GOLGA6L20* |
| ENSG00000136261 | 7 | 16675756 | 16756148 | 240 | 46 | 44911 | -2.492 | 0.99365 | *BZW2* |
| ENSG00000176787 | 11 | 5069880 | 5090857 | 201 | 12 | 53401 | -2.4938 | 0.99368 | *OR52E2* |
| ENSG00000162992 | 2 | 182527815 | 182555603 | 54 | 9 | 27567 | -2.4965 | 0.99373 | *NEUROD1* |
| ENSG00000129990 | 19 | 55672567 | 55701809 | 171 | 22 | 32244 | -2.4987 | 0.99377 | *SYT5* |
| ENSG00000204814 | 9 | 44857571 | 44880528 | 12 | 4 | 45010 | -2.5021 | 0.99383 | *RP11-160N1.10* |
| ENSG00000269699 | 19 | 57275920 | 57362097 | 156 | 29 | 51677 | -2.5045 | 0.99387 | *ZIM2* |
| ENSG00000183250 | 21 | 46342729 | 46369828 | 107 | 17 | 38243 | -2.507 | 0.99391 | *C21orf67* |
| ENSG00000136710 | 2 | 131085814 | 131109922 | 35 | 12 | 36572 | -2.527 | 0.99425 | *CCDC115* |
| ENSG00000114248 | 3 | 169546967 | 169597718 | 148 | 20 | 52661 | -2.5424 | 0.99449 | *LRRC31* |
| ENSG00000144043 | 2 | 71203068 | 71232075 | 88 | 19 | 49212 | -2.5445 | 0.99453 | *TEX261* |
| ENSG00000171160 | 10 | 99364310 | 99403344 | 86 | 15 | 43006 | -2.5446 | 0.99453 | *MORN4* |
| ENSG00000102471 | 13 | 80045287 | 80140210 | 164 | 19 | 36639 | -2.5443 | 0.99453 | *NDFIP2* |
| ENSG00000176155 | 17 | 80049336 | 80180706 | 620 | 16 | 26585 | -2.5476 | 0.99458 | *CCDC57* |

| | | | | | | | | | |
|---|---|---|---|---|---|---|---|---|---|
| ENSG00000107371 | 9 | 37756975 | 37811434 | 161 | 18 | 54872 | -2.5543 | 0.99468 | *EXOSC3* |
| ENSG00000127311 | 12 | 66686325 | 66747423 | 158 | 15 | 48697 | -2.5736 | 0.99497 | *HELB* |
| ENSG00000238083 | 17 | 44578877 | 44643016 | 2 | 1 | 43331 | -2.5777 | 0.99503 | *LRRC37A2* |
| ENSG00000137673 | 11 | 102381239 | 102411484 | 92 | 25 | 47185 | -2.5822 | 0.99509 | *MMP7* |
| ENSG00000144649 | 3 | 43010759 | 43111703 | 279 | 34 | 42005 | -2.5845 | 0.99512 | *FAM198A* |
| ENSG00000167674 | 19 | 4462284 | 4512223 | 248 | 31 | 47263 | -2.5859 | 0.99514 | *HDGFRP2* |
| ENSG00000163393 | 1 | 116509119 | 116622675 | 299 | 40 | 44425 | -2.5866 | 0.99515 | *SLC22A15* |
| ENSG00000146757 | 7 | 64828712 | 64876038 | 179 | 13 | 18828 | -2.5953 | 0.99527 | *ZNF92* |
| ENSG00000172469 | 6 | 96015419 | 96067333 | 207 | 15 | 55748 | -2.5971 | 0.9953 | *MANEA* |
| ENSG00000120149 | 5 | 174141536 | 174167896 | 35 | 17 | 45040 | -2.5993 | 0.99533 | *MSX2* |
| ENSG00000180694 | 8 | 91624223 | 91813860 | 418 | 26 | 47659 | -2.5999 | 0.99534 | *TMEM64* |
| ENSG00000129473 | 14 | 23757999 | 23790968 | 70 | 23 | 48288 | -2.6089 | 0.99546 | *BCL2L2* |
| ENSG00000100311 | 22 | 39609364 | 39650756 | 138 | 20 | 51923 | -2.6093 | 0.99546 | *PDGFB* |
| ENSG00000166664 | 15 | 30643443 | 30696052 | 3 | 1 | 42519 | -2.6154 | 0.99554 | *CHRFAM7A* |
| ENSG00000135540 | 6 | 138733180 | 139023708 | 825 | 118 | 41930 | -2.6171 | 0.99557 | *NHSL1* |
| ENSG00000188269 | 19 | 14893302 | 14956188 | 252 | 49 | 49436 | -2.6189 | 0.99559 | *OR7A5* |
| ENSG00000164040 | 4 | 129180397 | 129219984 | 75 | 14 | 52426 | -2.6408 | 0.99586 | *PGRMC2* |
| ENSG00000084073 | 1 | 40713779 | 40769856 | 105 | 25 | 41684 | -2.6483 | 0.99595 | *ZMPSTE24* |
| ENSG00000215549 | 13 | 26432061 | 26465095 | 59 | 14 | 47576 | -2.668 | 0.99618 | *AL138815.1* |
| ENSG00000234965 | 22 | 42297297 | 42320570 | 65 | 9 | 44593 | -2.6767 | 0.99628 | *SHISA8* |
| ENSG00000151778 | 13 | 44937801 | 44981850 | 74 | 14 | 51798 | -2.6822 | 0.99634 | *SERP2* |
| ENSG00000135625 | 2 | 73508058 | 73530833 | 21 | 6 | 33942 | -2.6855 | 0.99638 | *EGR4* |
| ENSG00000186599 | 8 | 7335191 | 7357115 | 2 | 1 | 42633 | -2.6874 | 0.9964 | *DEFB105B* |
| ENSG00000180098 | 1 | 28869597 | 28915051 | 84 | 15 | 46731 | -2.6906 | 0.99643 | *TRNAU1AP* |
| ENSG00000108878 | 17 | 65030706 | 65062909 | 94 | 24 | 44351 | -2.7183 | 0.99672 | *CACNG1* |
| ENSG00000159228 | 21 | 37432239 | 37455464 | 113 | 20 | 53898 | -2.722 | 0.99676 | *CBR1* |
| ENSG00000179091 | 8 | 145139930 | 145162428 | 45 | 6 | 29327 | -2.7246 | 0.99678 | *CYC1* |
| ENSG00000103381 | 16 | 12746919 | 12907874 | 606 | 75 | 41530 | -2.7303 | 0.99684 | *CPPED1* |
| ENSG00000267857 | 21 | 46510728 | 46544873 | 120 | 19 | 55762 | -2.7372 | 0.9969 | *PRED58* |
| ENSG00000120586 | 10 | 18088352 | 18210091 | 45 | 17 | 16470 | -2.7417 | 0.99694 | *MRC1* |
| ENSG00000105640 | 19 | 17960685 | 17984962 | 83 | 12 | 37981 | -2.7439 | 0.99696 | *RPL18A* |
| ENSG00000162971 | 2 | 200784698 | 200830459 | 107 | 18 | 50851 | -2.7447 | 0.99697 | *TYW5* |
| ENSG00000170231 | 5 | 159604374 | 159675742 | 371 | 40 | 46904 | -2.7499 | 0.99702 | *FABP6* |
| ENSG00000214215 | 12 | 93086619 | 93112325 | 51 | 8 | 29535 | -2.7499 | 0.99702 | *C12orf74* |
| ENSG00000268805 | 21 | 46501596 | 46525493 | 77 | 13 | 54821 | -2.7541 | 0.99706 | *PRED57* |
| ENSG00000206073 | 18 | 61294493 | 61321532 | 26 | 6 | 54396 | -2.7681 | 0.99718 | *SERPINB4* |

| | | | | | | | | | |
|---|---|---|---|---|---|---|---|---|---|
| ENSG00000120696 | 13 | 41753969 | 41778702 | 22 | 10 | 51122 | -2.7777 | 0.99726 | *KBTBD7* |
| ENSG00000117000 | 1 | 40617045 | 40716593 | 138 | 27 | 49096 | -2.7906 | 0.99737 | *RLF* |
| ENSG00000105509 | 19 | 52206365 | 52237247 | 149 | 36 | 50041 | -2.7922 | 0.99738 | *HAS1* |
| ENSG00000156869 | 1 | 100164259 | 100242187 | 299 | 14 | 23797 | -2.7981 | 0.99743 | *FRRS1* |
| ENSG00000123901 | 11 | 94100477 | 94144585 | 209 | 30 | 49097 | -2.8054 | 0.99749 | *GPR83* |
| ENSG00000181751 | 5 | 102584403 | 102624361 | 110 | 15 | 49516 | -2.8491 | 0.99781 | *C5orf30* |
| ENSG00000135709 | 16 | 85051375 | 85137836 | 466 | 58 | 40690 | -2.8845 | 0.99804 | *KIAA0513* |
| ENSG00000173673 | 1 | 6294252 | 6315638 | 55 | 13 | 39975 | -2.908 | 0.99818 | *HES3* |
| ENSG00000177426 | 18 | 3401606 | 3468409 | 185 | 53 | 43854 | -2.9258 | 0.99828 | *TGIF1* |
| ENSG00000197381 | 21 | 46483768 | 46656478 | 555 | 56 | 50142 | -2.9322 | 0.99832 | *ADARB1* |
| ENSG00000204595 | 19 | 54125310 | 54150263 | 53 | 19 | 40628 | -2.9601 | 0.99846 | *DPRX* |
| ENSG00000122741 | 9 | 37790499 | 37877663 | 294 | 13 | 55562 | -2.975 | 0.99854 | *DCAF10* |
| ENSG00000103150 | 16 | 83922731 | 83959787 | 197 | 25 | 48078 | -2.9844 | 0.99858 | *MLYCD* |
| ENSG00000181408 | 17 | 80322153 | 80343462 | 115 | 11 | 54998 | -3.0232 | 0.99875 | *UTS2R* |
| ENSG00000166923 | 15 | 33000175 | 33036870 | 125 | 26 | 39429 | -3.0258 | 0.99876 | *GREM1* |
| ENSG00000144031 | 2 | 71195510 | 71222626 | 73 | 18 | 44905 | -3.0277 | 0.99877 | *ANKRD53* |
| ENSG00000160255 | 21 | 46295868 | 46361904 | 317 | 65 | 43105 | -3.04 | 0.99882 | *ITGB2* |
| ENSG00000183255 | 21 | 46259500 | 46303752 | 228 | 30 | 46118 | -3.0489 | 0.99885 | *PTTG1IP* |
| ENSG00000173083 | 4 | 84203614 | 84266306 | 161 | 19 | 47655 | -3.0594 | 0.99889 | *HPSE* |
| ENSG00000111711 | 12 | 21644715 | 21681342 | 167 | 27 | 49008 | -3.1144 | 0.99908 | *GOLT1B* |
| ENSG00000198049 | 1 | 206213976 | 206241639 | 52 | 12 | 45880 | -3.1334 | 0.99914 | *AVPR1B* |
| ENSG00000099985 | 22 | 30648818 | 30672829 | 43 | 17 | 49853 | -3.1337 | 0.99914 | *OSM* |
| ENSG00000238244 | 15 | 90880819 | 90902669 | 158 | 14 | 38877 | -3.1464 | 0.99917 | *GABARAPL3* |
| ENSG00000108039 | 10 | 111614524 | 111693311 | 146 | 24 | 30813 | -3.1833 | 0.99927 | *XPNPEP1* |
| ENSG00000141551 | 17 | 80186899 | 80241607 | 83 | 37 | 41983 | -3.2246 | 0.99937 | *CSNK1D* |
| ENSG00000105879 | 7 | 107374142 | 107412112 | 100 | 13 | 51152 | -3.4035 | 0.99967 | *CBLL1* |
| ENSG00000150361 | 13 | 70264726 | 70692591 | 2137 | 130 | 43475 | -3.4292 | 0.9997 | *KLHL1* |
| ENSG00000123685 | 1 | 212849760 | 212883327 | 143 | 28 | 54146 | -3.549 | 0.99981 | *BATF3* |
| ENSG00000196391 | 15 | 90885477 | 90919324 | 145 | 15 | 38176 | -3.6855 | 0.99989 | *ZNF774* |
| ENSG00000080031 | 19 | 55682616 | 55730874 | 236 | 40 | 36686 | -3.7756 | 0.99992 | *PTPRH* |
| ENSG00000198081 | 18 | 5279018 | 5307052 | 98 | 24 | 52245 | -3.9332 | 0.99996 | *ZBTB14* |
| ENSG00000005700 | 6 | 82869700 | 82967471 | 278 | 17 | 53082 | -4.1554 | 0.99998 | *IBTK* |
