## Supplementary material for "A genome-wide association study identified 10 novel genomic loci associated with intrinsic capacity": List of panther and reactome pathways where the IC prioritized genes have shown statistically signficant enrichment

| Pathways | #Genes | #Genes enriched | expected | Fold Enrichment | +/- | raw P value | Pathway databse |
|---|---|---|---|---|---|---|---|
| Activation of APC/C and APC/C:Cdc20 mediated degradation of mitotic proteins | 76 | 3 | 0.47 | 6.4 | + | 0.0116 | Reactome |
| Activation, myristolyation of BID and translocation to mitochondria | 4 | 1 | 0.02 | 40.51 | + | 0.0245 | Reactome |
| Alzheimer disease-presenilin pathway | 127 | 3 | 0.78 | 3.83 | + | 0.0441 | Panther pathway |
| Antigen processing-Cross presentation | 106 | 3 | 0.65 | 4.59 | + | 0.028 | Reactome |
| APC/C:Cdc20 mediated degradation of mitotic proteins | 75 | 3 | 0.46 | 6.48 | + | 0.0112 | Reactome |
| APC/C:Cdc20 mediated degradation of Securin | 67 | 3 | 0.41 | 7.26 | + | 0.00825 | Reactome |
| APC/C:Cdh1 mediated degradation of Cdc20 and other APC/C:Cdh1 targeted proteins in late mitosis/early G1 | 73 | 3 | 0.45 | 6.66 | + | 0.0104 | Reactome |
| APC/C-mediated degradation of cell cycle proteins | 87 | 3 | 0.54 | 5.59 | + | 0.0167 | Reactome |
| APC:Cdc20 mediated degradation of cell cycle proteins prior to satisfation of the cell cycle checkpoint | 73 | 3 | 0.45 | 6.66 | + | 0.0104 | Reactome |
| Apoptosis | 173 | 4 | 1.07 | 3.75 | + | 0.0224 | Reactome |
| Aryl hydrocarbon receptor signalling | 7 | 1 | 0.04 | 23.15 | + | 0.0424 | Reactome |
| Assembly of the pre-replicative complex | 110 | 3 | 0.68 | 4.42 | + | 0.0307 | Reactome |
| Asymmetric localization of PCP proteins | 63 | 3 | 0.39 | 7.72 | + | 0.00696 | Reactome |
| AUF1 (hnRNP D0) binds and destabilizes mRNA | 53 | 2 | 0.33 | 6.11 | + | 0.0424 | Reactome |
| Autodegradation of Cdh1 by Cdh1:APC/C | 63 | 3 | 0.39 | 7.72 | + | 0.00696 | Reactome |
| Autodegradation of the E3 ubiquitin ligase COP1 | 51 | 2 | 0.31 | 6.35 | + | 0.0396 | Reactome |
| Cdc20:Phospho-APC/C mediated degradation of Cyclin A | 72 | 3 | 0.44 | 6.75 | + | 0.01 | Reactome |
| CDC42 GTPase cycle | 154 | 4 | 0.95 | 4.21 | + | 0.0153 | Reactome |
| CDK-mediated phosphorylation and removal of Cdc6 | 72 | 3 | 0.44 | 6.75 | + | 0.01 | Reactome |
| Cell cycle | 22 | 2 | 0.14 | 14.73 | + | 0.00805 | Panther pathway |
| Cellular response to hypoxia | 74 | 3 | 0.46 | 6.57 | + | 0.0108 | Reactome |
| Citric acid cycle (TCA cycle) | 22 | 2 | 0.14 | 14.73 | + | 0.00805 | Reactome |
| Class B/2 (Secretin family receptors) | 93 | 3 | 0.57 | 5.23 | + | 0.0199 | Reactome |
| CLEC7A (Dectin-1) signaling | 96 | 3 | 0.59 | 5.06 | + | 0.0216 | Reactome |
| Coenzyme A biosynthesis | 7 | 1 | 0.04 | 23.15 | + | 0.0424 | Reactome |
| Coenzyme A biosynthesis | 8 | 1 | 0.05 | 20.26 | + | 0.0483 | Panther pathway |
| Cross-presentation of soluble exogenous antigens (endosomes) | 49 | 2 | 0.3 | 6.61 | + | 0.0368 | Reactome |
| C-type lectin receptors (CLRs) | 138 | 4 | 0.85 | 4.7 | + | 0.0106 | Reactome |
| Dectin-1 mediated noncanonical NF-kB signaling | 59 | 3 | 0.36 | 8.24 | + | 0.0058 | Reactome |
| Degradation of AXIN | 54 | 2 | 0.33 | 6 | + | 0.0439 | Reactome |
| Degradation of DVL | 56 | 2 | 0.35 | 5.79 | + | 0.0469 | Reactome |
| DNA Replication Pre-Initiation | 127 | 3 | 0.78 | 3.83 | + | 0.0441 | Reactome |
| FBXL7 down-regulates AURKA during mitotic entry and in early mitosis | 54 | 2 | 0.33 | 6 | + | 0.0439 | Reactome |
| GSK3B and BTRC:CUL1-mediated-degradation of NFE2L2 | 51 | 2 | 0.31 | 6.35 | + | 0.0396 | Reactome |
| Hedgehog 'on' state | 84 | 3 | 0.52 | 5.79 | + | 0.0152 | Reactome |
| Hh mutants abrogate ligand secretion | 58 | 2 | 0.36 | 5.59 | + | 0.0499 | Reactome |
| Hh mutants are degraded by ERAD | 55 | 2 | 0.34 | 5.89 | + | 0.0454 | Reactome |
| Host Interactions of HIV factors | 125 | 3 | 0.77 | 3.89 | + | 0.0424 | Reactome |
| Interferon alpha/beta signaling | 76 | 3 | 0.47 | 6.4 | + | 0.0116 | Reactome |
| Interleukin-1 signaling | 112 | 3 | 0.69 | 4.34 | + | 0.0322 | Reactome |
| Intra-Golgi traffic | 43 | 2 | 0.27 | 7.54 | + | 0.0289 | Reactome |
| IRE1alpha activates chaperones | 49 | 2 | 0.3 | 6.61 | + | 0.0368 | Reactome |
| Ligand-receptor interactions | 8 | 1 | 0.05 | 20.26 | + | 0.0483 | Reactome |
| Metabolism of polyamines | 57 | 2 | 0.35 | 5.69 | + | 0.0484 | Reactome |
| mRNA Splicing | 212 | 4 | 1.31 | 3.06 | + | 0.0425 | Reactome |
| mRNA splicing | 7 | 1 | 0.04 | 23.15 | + | 0.0424 | Panther pathway |
| mRNA Splicing - Major Pathway | 204 | 4 | 1.26 | 3.18 | + | 0.0378 | Reactome |
| mRNA Splicing - Minor Pathway | 50 | 2 | 0.31 | 6.48 | + | 0.0382 | Reactome |
| Negative regulation of NOTCH4 signaling | 52 | 2 | 0.32 | 6.23 | + | 0.041 | Reactome |
| NIK-->noncanonical NF-kB signaling | 58 | 3 | 0.36 | 8.38 | + | 0.00553 | Reactome |
| Orexin and neuropeptides FF and QRFP bind to their respective receptors | 8 | 1 | 0.05 | 20.26 | + | 0.0483 | Reactome |
| p53-Independent DNA Damage Response | 51 | 2 | 0.31 | 6.35 | + | 0.0396 | Reactome |
| p53-Independent G1/S DNA damage checkpoint | 51 | 2 | 0.31 | 6.35 | + | 0.0396 | Reactome |
| PCP/CE pathway | 91 | 3 | 0.56 | 5.34 | + | 0.0188 | Reactome |
| Programmed Cell Death | 205 | 4 | 1.27 | 3.16 | + | 0.0384 | Reactome |
| PTK6 Regulates Proteins Involved in RNA Processing | 5 | 1 | 0.03 | 32.41 | + | 0.0305 | Reactome |
| Pyruvate metabolism and Citric Acid (TCA) cycle | 54 | 2 | 0.33 | 6 | + | 0.0439 | Reactome |
| RAC1 GTPase cycle | 183 | 4 | 1.13 | 3.54 | + | 0.0269 | Reactome |
| Regulation of activated PAK-2p34 by proteasome mediated degradation | 49 | 2 | 0.3 | 6.61 | + | 0.0368 | Reactome |
| Regulation of APC/C activators between G1/S and early anaphase | 80 | 3 | 0.49 | 6.08 | + | 0.0134 | Reactome |
| Regulation of Apoptosis | 52 | 2 | 0.32 | 6.23 | + | 0.041 | Reactome |
| Regulation of mitotic cell cycle | 87 | 3 | 0.54 | 5.59 | + | 0.0167 | Reactome |
| Regulation of ornithine decarboxylase (ODC) | 50 | 2 | 0.31 | 6.48 | + | 0.0382 | Reactome |
| Regulation of RUNX3 expression and activity | 53 | 2 | 0.33 | 6.11 | + | 0.0424 | Reactome |
| Replication of the SARS-CoV-1 genome | 4 | 1 | 0.02 | 40.51 | + | 0.0245 | Reactome |
| Replication of the SARS-CoV-2 genome | 4 | 1 | 0.02 | 40.51 | + | 0.0245 | Reactome |
| RMTs methylate histone arginines | 48 | 2 | 0.3 | 6.75 | + | 0.0354 | Reactome |
| RUNX1 interacts with co-factors whose precise effect on RUNX1 targets is not known | 36 | 2 | 0.22 | 9 | + | 0.0208 | Reactome |
| RUNX1 regulates transcription of genes involved in differentiation of keratinocytes | 8 | 1 | 0.05 | 20.26 | + | 0.0483 | Reactome |
| SARS-CoV-1 Genome Replication and Transcription | 4 | 1 | 0.02 | 40.51 | + | 0.0245 | Reactome |
| SARS-CoV-2 Genome Replication and Transcription | 4 | 1 | 0.02 | 40.51 | + | 0.0245 | Reactome |
| SCF-beta-TrCP mediated degradation of Emi1 | 54 | 2 | 0.33 | 6 | + | 0.0439 | Reactome |
| Somitogenesis | 54 | 2 | 0.33 | 6 | + | 0.0439 | Reactome |
| Stabilization of p53 | 55 | 2 | 0.34 | 5.89 | + | 0.0454 | Reactome |
| Switching of origins to a post-replicative state | 91 | 3 | 0.56 | 5.34 | + | 0.0188 | Reactome |
| Synaptic vesicle trafficking | 30 | 2 | 0.19 | 10.8 | + | 0.0147 | Panther pathway |
| Synthesis of DNA | 120 | 3 | 0.74 | 4.05 | + | 0.0383 | Reactome |
| TCF dependent signaling in response to WNT | 200 | 4 | 1.23 | 3.24 | + | 0.0355 | Reactome |
| TNFR2 non-canonical NF-kB pathway | 98 | 3 | 0.6 | 4.96 | + | 0.0228 | Reactome |
| Toxicity of botulinum toxin type C (botC) | 3 | 1 | 0.02 | 54.02 | + | 0.0184 | Reactome |
| Trafficking and processing of endosomal TLR | 13 | 2 | 0.08 | 24.93 | + | 0.00282 | Reactome |
| Transcriptional regulation by RUNX1 | 202 | 5 | 1.25 | 4.01 | + | 0.0084 | Reactome |
| Ubiquitin Mediated Degradation of Phosphorylated Cdc25A | 51 | 2 | 0.31 | 6.35 | + | 0.0396 | Reactome |
| Ubiquitin proteasome pathway | 58 | 2 | 0.36 | 5.59 | + | 0.0499 | Panther pathway |
| Ubiquitin-dependent degradation of Cyclin D | 51 | 2 | 0.31 | 6.35 | + | 0.0396 | Reactome |
| Vif-mediated degradation of APOBEC3G | 52 | 2 | 0.32 | 6.23 | + | 0.041 | Reactome |
| Vpr-mediated induction of apoptosis by mitochondrial outer membrane permeabilization | 3 | 1 | 0.02 | 54.02 | + | 0.0184 | Reactome |
| Vpu mediated degradation of CD4 | 51 | 2 | 0.31 | 6.35 | + | 0.0396 | Reactome |
