## Supplementary material for "A genome-wide association study identified 10 novel genomic loci associated with intrinsic capacity": Development of IC score in the Canadian longitudinal study of aging

### **Intrinsic capacity score in the CLSA**

The Canadian Longitudinal Study on Aging (CLSA) is a significant research initiative aimed at comprehensively examining the aging process and its implications for health and well-being over time. Initiated in 2010, the CLSA represents one of Canada's largest studies on aging, involving a diverse sample of approximately 50,000 individuals aged 45 to 85 years at baseline. Participants are drawn from various regions across Canada, encompassing both urban and rural settings. The study comprises two primary cohorts: a tracking cohort (~20,000 participants) and a comprehensive cohort (~30,000 participants), each of which is longitudinally followed for 20 years. Data collection includes detailed interviews, physical assessments, and biological samples, providing valuable insights into health determinants and aging trajectories among Canadians. Currently, genomic data has been collected and genotyping performed for the comprehensive cohort (n=26,622 individuals), directing our focus to this cohort for genetic analyses[1, 2]. For more detailed information, please refer to the official CLSA website(<https://bmjopen.bmj.com/content/12/3/e059021>).

For this study, we developed the Intrinsic Capacity (IC) score within the CLSA using a methodology similar to that we employed in the UK Biobank study[3]. Initially, variables analogous to those used in the UK Biobank and commonly employed to assess the five domains of intrinsic capacity were selected. Subsequently, through exploratory analyses, 15 variables demonstrating factor loadings of 0.2 or higher and exhibiting good fit within the bifactor model were tested in confirmatory models. Thus, a model comprising these 15 variables was utilized to generate IC scores in the CLSA study. The steps undertaken, along with results from exploratory and confirmatory factor analyses and preliminary tests of adequacy, are detailed herein.

| **Variable** | **Name in the CLSA** | **Measurement** |
| --- | --- | --- |
| Immediate Recall | COG_REYI_SCORE_COM | Measured using Rey Auditory Verbal Learning Test. Number of words (or variants) correctly recalled in 90 seconds - Immediate Recall. |
| Delayed Recall | COG_REYII_SCORE_COM | Measured using Rey Auditory Verbal Learning Test. Number of words (or variants) correctly recalled in 90 seconds - Delayed Recall. |
| Verbal fluency (FAS-score) | Sum of:  FAS_F_SCORE_COM,  FAS_A_SCORE_COM,  FAS_S_SCORE_COM | This variable measures verbal fluency using the Controlled Oral Word Association Test (COWAT), specifically summing responses for the letters F, A, and S. Each sum represents the total number of words associated with each letter within a designated time frame. |
| Standing Balance | BAL_BEST_COM | This is measured by the best attained time for the accomplishing standing balance test in seconds (given 60 seconds maximum and sopped beyond this). |
| Walking Pace | WLK_TIME_COM | This is a timed 4-meter Walk Test measured in total time required to complete 4mWalk (in seconds). |
| Anxiety disorder | CCC_ANXI_COM | Self-reported (Yes/No) question |
| CESD-10 Score | DEP_CESD10_COM | Center for Epidemiological Studies Short Depression Scale (CES-D 10) score. |
| Mood disorder | CCC_MOOD_COM | Self-reported (yes/no) |
| Hearing rating | HRG_HRG_COM | Self-rated hearing (rated 1-5) |
| Hearing aid use | HRG_AID_COM | Self-reported use of any hearing aid |
| Hearing difficulty with background noise | HRG_NOIS_COM | Self-reported hearing difficulty in the presence of background noise |
| Haemoglobin concentration | BLD_Hgb_COM | Haemoglobin concentration in grams per decilitre |
| Hand grip strength | GS_EXAM_AVG_COM | Average hand grip strength of left and right hand |
| FEV1 | Maximum | Forced expiratory volume in 1 second, maximum of all trails made |

**Table 1:** Description of the items used to measure IC

#### **Factorability (Factor adequacy) test**

Though factor adequacy for items commonly used to measure IC is tested for their factorability in different previous studies, we checked this before proceeding to factor analyses to confirm if it is still maintained for the items used in the CLSA. This is usually the first step in factor analyses to determine how suited the data is for factor analysis. We used three methods: the Kaiser-Meyer-Olkin (KMO) Test (should be at least 0.6; mediocre), Bartlett’s test of sphericity (a significant test indicates that factor analysis may be worthwhile for the data set), and the determinant of matrix tests (correlation matrix should be > 0.00001)[4]. According to this our tests using the 15 variables used to develop the IC construct indicate factorability: the overall KMO =0.65, P-value < 0.001 for Bartlett’s test of sphericity and determinant of matrix score = 0.084.

#### **Determining the number of factors**

Following factor adequacy testing, we utilized the parallel analysis method—a precise yet underutilized approach previously employed in our study within the UK Biobank—to determine the optimal number of factors for retention. Using this method, we identified five factors in our analysis. [3, 5, 6].

#### **Exploratory factor analysis (EFA)**

We conducted both correlated five-factor (conventional) Exploratory Factor Analysis (EFA) and bifactor EFA. The conventional EFA utilized the "psych" package with the "factanal" function, employing the "ProMax" oblique rotation method. For the bifactor EFA, we selected the "bifactor" rotation method. In both analyses, the maximum likelihood (ML) method was chosen for factor extraction [7]. The correlated factors EFA revealed distinct loading patterns of variables across five factors. Similarly, the bifactor EFA demonstrated clear loading of variables onto the five specific domains as well as the general intrinsic capacity domain. The goodness of fit statistics for the bifactor EFA indicated a good model fit, with a Tucker Lewis index (TLI) of 0.995, root mean square error of approximation (RMSEA) of 0.012, and root mean square residual (RMSR) of 0.01.

| **Variable** | **Factor1** | **Factor2** | **Factor3** | **Factor4** | **Factor5** |
| --- | --- | --- | --- | --- | --- |
| Immediate Recall |  | 0.979 |  |  |  |
| Delayed Recall |  | 0.724 |  |  |  |
| Verbal fluency (FAS-score) |  | 0.218 |  |  | 0.169 |
| Standing Balance |  |  |  |  | 0.712 |
| Walking Pace |  |  |  |  | -0.402 |
| Anxiety disorder |  |  |  | 0.483 |  |
| CESD-10 Score |  |  |  | -0.416 |  |
| Mood disorder |  |  |  | 0.673 |  |
| Hearing rating |  |  | -0.611 |  |  |
| Hearing aid use |  |  | 0.258 |  | 0.108 |
| Hearing difficulty with background noise |  |  | 0.706 |  |  |
| Haemoglobin concentration | 0.538 |  |  |  | -0.139 |
| Hand grip strength | 0.903 |  |  |  |  |
| FEV1 | 0.715 |  |  |  | 0.179 |

**Table 2**: Conventional EFA

| **Variable** | **General** | **Factor1** | **Factor2** | **Factor3** | **Factor4** | **Factor5** |
| --- | --- | --- | --- | --- | --- | --- |
| Immediate Recall | 0.45 | 0.00 | 0.01 | -0.01 | 0.81 | -0.01 |
| Delayed Recall | 0.43 | -0.02 | -0.01 | -0.01 | 0.62 | 0.03 |
| Verbal fluency (FAS-score) | 0.27 | 0.05 | 0.00 | -0.03 | 0.19 | -0.09 |
| Standing Balance | 0.55 | 0.33 | -0.03 | 0.03 | -0.04 | -0.01 |
| Walking Pace | -0.34 | -0.25 | 0.00 | -0.05 | 0.04 | 0.24 |
| Anxiety disorder | -0.02 | 0.04 | 0.02 | 0.47 | -0.01 | 0.02 |
| CESD-10 Score | -0.03 | -0.12 | -0.12 | -0.42 | -0.01 | 0.20 |
| Mood disorder | -0.05 | 0.06 | 0.04 | 0.67 | -0.03 | 0.03 |
| Hearing rating | -0.22 | 0.02 | -0.57 | -0.02 | -0.01 | 0.01 |
| Hearing aid use | 0.20 | 0.04 | 0.24 | -0.02 | 0.01 | 0.12 |
| Hearing difficulty with background noise | 0.14 | -0.04 | 0.67 | 0.07 | 0.00 | 0.00 |
| Haemoglobin concentration | -0.20 | 0.47 | 0.00 | 0.03 | -0.01 | -0.02 |
| Hand grip strength | -0.11 | 0.88 | 0.00 | 0.03 | 0.00 | -0.03 |
| FEV1 | 0.09 | 0.79 | -0.02 | 0.01 | 0.01 | 0.05 |

**Table 3:** Bifactor EFA

#### **Confirmatory factor analysis**

We did confirmatory factor analysis using the bifactor approach analogous to the one done in the UK biobank and using this bifactor model under the SEM framework we developed a score for intrinsic capacity. The “lavaan” package was used for the CFA, employing the Goemin rotation method[8]. The CFA has then confirmed the structure and dimensionality of the IC observed in the EFA (with the goodness of fit statistics; CFI=0.992, TLI=0.985, RMSEA =0.020, and RMSR=0.011). Below are findings from the bifactor CFA.

| **Variable** | **General** | **Factor1** | **Factor2** | **Factor3** | **Factor4** | **Factor5** |
| --- | --- | --- | --- | --- | --- | --- |
| Hand grip strength | -0.11 | 0.88 | 0.00 | 0.03 | 0.00 | -0.03 |
| FEV1 | 0.09 | 0.79 | -0.02 | 0.01 | 0.01 | 0.05 |
| Haemoglobin concentration | -0.20 | 0.47 | 0.00 | 0.03 | -0.01 | -0.02 |
| Hearing aid use | 0.20 | 0.04 | 0.24 | -0.02 | 0.01 | 0.12 |
| Hearing rating | -0.22 | 0.02 | -0.57 | -0.02 | -0.01 | 0.01 |
| Hearing difficulty with background noise | 0.14 | -0.04 | 0.67 | 0.07 | 0.00 | 0.00 |
| Anxiety disorder | -0.02 | 0.04 | 0.02 | 0.47 | -0.01 | 0.02 |
| CESD-10 Score | -0.03 | -0.12 | -0.12 | -0.42 | -0.01 | 0.20 |
| Mood disorder | -0.05 | 0.06 | 0.04 | 0.67 | -0.03 | 0.03 |
| Verbal fluency (FAS-score) | 0.27 | 0.05 | 0.00 | -0.03 | 0.19 | -0.09 |
| Immediate Recall | 0.45 | 0.00 | 0.01 | -0.01 | 0.81 | -0.01 |
| Delayed Recall | 0.43 | -0.02 | -0.01 | -0.01 | 0.62 | 0.03 |
| Standing Balance | 0.55 | 0.33 | -0.03 | 0.03 | -0.04 | -0.01 |
| Walking Pace | -0.34 | -0.25 | 0.00 | -0.05 | 0.04 | 0.24 |

**Table 4**: Factor loadings from confirmatory factor analysis

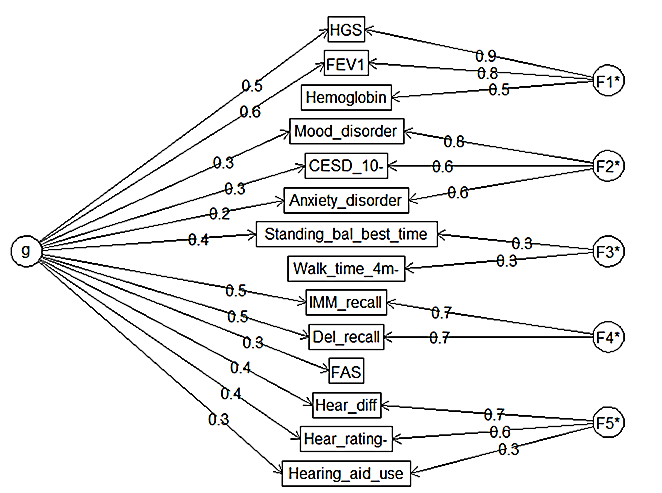

**Figure 1**. Bifactor CFA model

#### **Distribution of IC score**

After generating an intrinsic capacity score for the general and specific domains, we further explored their distributions, pattern of IC with age, and differences in IC score between the two sexes as well as

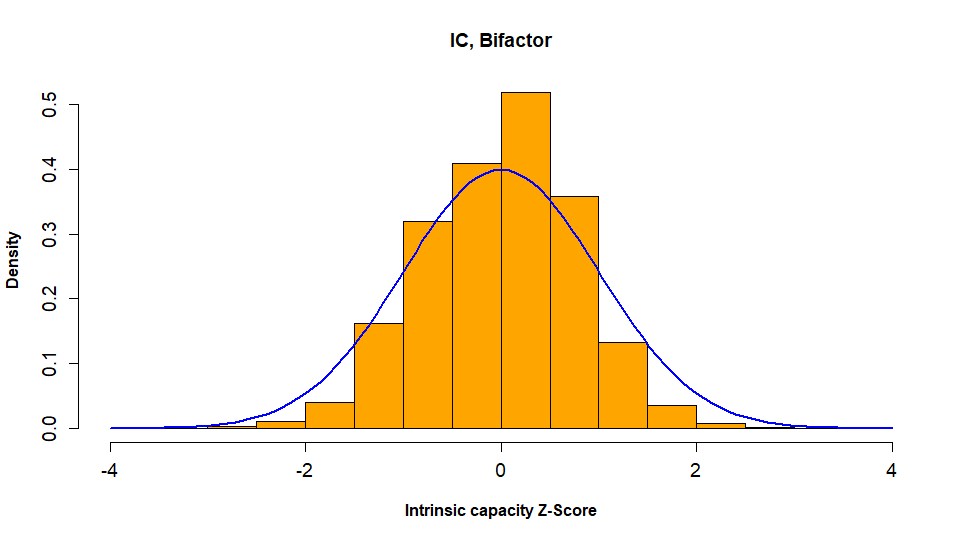

**Figure 2**: Histogram with Normal Curve for the Intrinsic Capacity General Score. This histogram, overlaid with a normal curve, shows that the intrinsic capacity general score constructed from the bifactor confirmatory model has an approximately normal distribution.

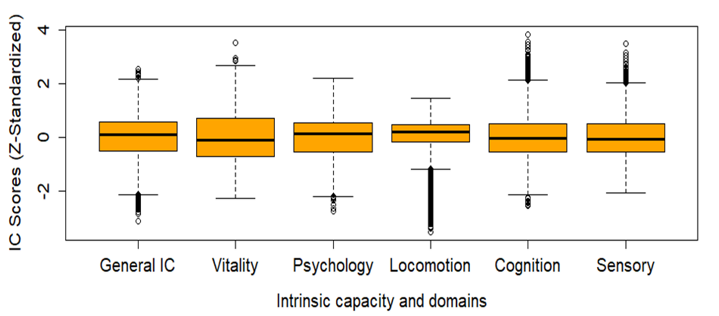

**Figure 3:** Box plots for the Intrinsic Capacity General Score and Domain-Specific Scores Developed from the Bifactor Confirmatory Model. These box plots illustrate that the distributions of both the general score and domain-specific scores are approximately normal, with no severe outlier values observed.

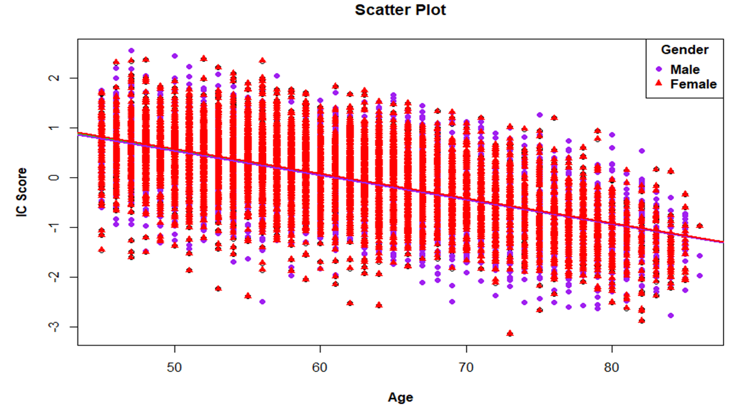

**Figure 4:** Scatter plot of IC with age with linear regression line for males and females. This figure illustrates a scatter plot with a fitted linear regression line. It demonstrates a consistent decrease in the intrinsic capacity score with increasing age. Furthermore, the distribution of IC scores between males and females exhibits minor differences across age groups.

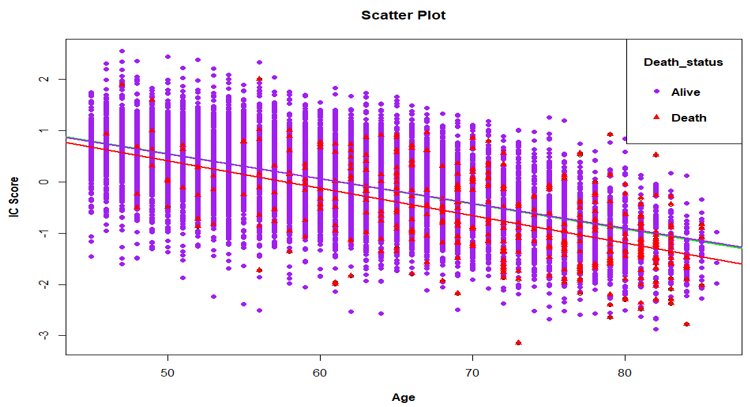

**Figure 5:** Scatter plot of IC with age with linear regression lines for deceased and alive. The figure illustrates that, on average, the baseline IC score for the dead is lower than that for the alive.

The logistic regression analysis for exploring if intrinsic capacity predicts death or not showed that the risk of death is higher for people with lower intrinsic capacity. People with a one-unit higher baseline IC Z-score have, on average, about 40% lower risk of death at the end of the second follow-up (OR =0.5951; 95% CI: 0.505-0.701). The death record at the second follow-up (Data released in September 2022) was used for ascertaining death status in this analysis.

|  |  | **IC Quartiles** | | | |
| --- | --- | --- | --- | --- | --- |
|  |  | Q1 | Q2 | Q3 | Q4 |
| **Death Status** | Alive | 3039 | 3192 | 3238 | 3238 |
|  | Dead | 239 | 86 | 40 | 40 |

**Table 5:** The distribution of death in the four quartiles of intrinsic capacity. As can be seen from the distribution, more than half (59%) of the deaths are in the first quartile of intrinsic capacity, followed by the next highest death in the second quartile of IC, whereas the third and fourth quartiles have almost the same number of deaths.
