## Supplementary material for "A genome-wide association study identified 10 novel genomic loci associated with intrinsic capacity": partiotion of variation in IC due to genetic varaints tested and other factors along with SNP based heritability estimate

**Supplementary table 1:** SNP-based heritability estimates of IC using GCTA - REML in the UKB and CLSA

| **SNP-based heritability statistics, UKB** | | | **SNP-based heritability statistics, CLSA** | | |
| --- | --- | --- | --- | --- | --- |
| **Source** | **Variance** | **SE** | **Source** | **Variance** | **SE** |
| V(G) | 0.062475 | 0.002669 | V(G) | 0.071038 | 0.010012 |
| V(e) | 0.185960 | 0.002598 | V(e) | 0.293082 | 0.010202 |
| Vp | 0.248435 | 0.001701 | Vp | 0.364120 | 0.004526 |
| ***V(G)/Vp*** | ***0.251474*** | 0.010249 | ***V(G)/Vp*** | ***0.195095*** | 0.027156 |
| logL | 9112.565 |  | logL | 24.187 |  |
| logL0 | 8726.085 |  | logL0 | -12.497 |  |
| LRT | 772.960 |  | LRT | 73.368 |  |
| df | 1 |  | df | 1 |  |
| Pval | 0.0000e+00 |  | Pval | 0.0000e+00 |  |
| n | 44631 |  | n | 13085 |  |
