## Supplementary material for "A genome-wide association study identified 10 novel genomic loci associated with intrinsic capacity": Data management and analysis codes

############################  A. QC UKB ##################################################

### SNP QC ###

Step-1: Extract the SNPS from the main bgen file (from the 22 chromosomes) by info score (infoscore >=0.3); chromosome by chromsome 

# First imputation qaulity score [Imputation minor allelle frequncy and infromation scores] shall be downloaded from the uk biobank resource (Resource code 1967).
Or simply it is the resource part of the field with ID: 100319 which says Impuation. 

For that copy and run this wget link on phoenix (HPC): wget  -nd  biobank.ndph.ox.ac.uk/ukb/ukb/auxdata/ukb_imp_mfi.tgz

# Then extract the zip file by: tar -xvzf file_name
#Then go to R on phoenix and among the extracted info score for each of the 22 chromosomes, prepare a text file that conatins only those SNPS with info score of 0.3. 


#Read file on R (call)
a=read.table("ukb_mfi_chr22_v3.txt", header=F, sep='\t')

# extract only those with info score>=0.3
chr22_info=subset(a, V8>=0.3)

#slect only SNP and info score columns
chr22_info=chr22_info[,c(2,8)] #To have only list of SNPS and info score columns

#Finally, on R, prepare a file containg SNPS and info score to take to phoenix for extraction from the main bgen files as; (eg. for chr22)
write.table(chr22_info, 'SNP_list_info_chr22.txt', row.names=F, col.names=F, quote=F, sep='\t')

Then quit R and go back to phoenix and extract those from the main geno file. 

(Alternatively as done below the SNP list for chromosomes 1-22 with imputation quality >=0.3 can be merged and used for all). 

Step-2: Extract the bgen file using the the list of SNPs prepared by screening using info score >=0.3 for each chromosome.

# Using Combined SNP list for chromosomes 1-22, extract bgen file from each chromosome as:
./plink2 --threads 8 --bgen ukb22828_c20_b0_v3.bgen ref-first --sample ukb22828_c22_b0_v3_s487160.sample --extract SNP_list_info_chr1_chr22.txt --set-missing-var-ids @:#[b37]\$r,\$a --export bgen-1.2 --out ukb_chr20_post_impute
./plink2 --threads 8 --bgen ukb22828_c21_b0_v3.bgen ref-first --sample ukb22828_c22_b0_v3_s487160.sample --extract SNP_list_info_chr1_chr22.txt --set-missing-var-ids @:#[b37]\$r,\$a --export bgen-1.2 --out ukb_chr21_post_impute
./plink2 --threads 8 --bgen ukb22828_c22_b0_v3.bgen ref-first --sample ukb22828_c22_b0_v3_s487160.sample --extract SNP_list_info_chr1_chr22.txt --set-missing-var-ids @:#[b37]\$r,\$a --export bgen-1.2 --out ukb_chr22_post_impute


Step-3: extract using the maf 0.01, geno 0.05, mind 0.05, hwe 1e-7 and max-alleles 2.
./plink2 --threads 6 --bgen ukb_chr20_post_impute.bgen ref-first --sample ukb_chr20_post_impute.sample --maf 0.01 --geno 0.05 --mind 0.05 --hwe 1e-7  --max-alleles 2 --export bgen-1.2 --out ukb_chr20_post_impute_1
./plink2 --threads 6 --bgen ukb_chr21_post_impute.bgen ref-first --sample ukb_chr21_post_impute.sample --maf 0.01 --geno 0.05 --mind 0.05 --hwe 1e-7  --max-alleles 2 --export bgen-1.2 --out ukb_chr21_post_impute_1
./plink2 --threads 6 --bgen ukb_chr22_post_impute.bgen ref-first --sample ukb_chr22_post_impute.sample --maf 0.01 --geno 0.05 --mind 0.05 --hwe 1e-7  --max-alleles 2 --export bgen-1.2 --out ukb_chr22_post_impute_1


Step_4: make ".bed" files from the .bgen files
./plink2 --bgen ukb_chr20_post_impute_1.bgen ref-first --sample  ukb_chr20_post_impute_1.sample --rm-dup --make-bed --out ukb_chr20_post_impute_2
./plink2 --bgen ukb_chr20_post_impute_1.bgen ref-first --sample  ukb_chr20_post_impute_1.sample --exclude ukb_chr20_post_impute_2.rmdup.mismatch --make-bed --out ukb_chr20_post_impute_2

./plink2 --bgen ukb_chr21_post_impute_1.bgen ref-first --sample  ukb_chr21_post_impute_1.sample --rm-dup --make-bed --out ukb_chr21_post_impute_2
./plink2 --bgen ukb_chr21_post_impute_1.bgen ref-first --sample  ukb_chr21_post_impute_1.sample --exclude ukb_chr21_post_impute_2.rmdup.mismatch --make-bed --out ukb_chr21_post_impute_2


./plink2 --bgen ukb_chr22_post_impute_1.bgen ref-first --sample  ukb_chr22_post_impute_1.sample --rm-dup --make-bed --out ukb_chr22_post_impute_2
./plink2 --bgen ukb_chr22_post_impute_1.bgen ref-first --sample  ukb_chr22_post_impute_1.sample --exclude ukb_chr22_post_impute_2.rmdup.mismatch --make-bed --out ukb_chr22_post_impute_2


### Individuaul (sample) QC ###

Step-5: exclude those individuals with sex aneuploidy ==1. The varaible/field sex_aneuoploidy is extracted from UK biobank and using the ID list of individulas with sex aneuploidy value =1, screen out those individuals. Use the following command done for chromsome 22 as example and duplicate for all chromosome bfiles.

### Do this for all chrs or loop as below
./plink2 --bfile ukb_chr22_post_impute_2 --remove sex_aneuploidy.txt --make-bed --out ukb_chr22_post_impute_3


### Loop code to execute the task for the chromosomes 1-22 ###
for (( i=1; i<=22; i++ )); do
    ./plink2 --bfile ukb_chr"${i}"_post_impute_2 --remove sex_aneuploidy.txt --make-bed --out ukb_chr"${i}"_post_impute_3
done


#Step-6: Sex-Mismatch. UK biobank genetic sex field is extracted and recorded sex from questionnaire is there. So, exclude those with mismatch or maintain those which match. command is done for chromosome 22 as example and duplicate it for all chromosome. by taking the base file immediately created before this step (i.e. ukb_chr22_post_impute_4), it will filter out those with sex mismatch.  example for chr22. 
 
./plink2 --bfile ukb_chr22_post_impute_3 --remove sex_mismatch_m.txt --make-bed --out ukb_chr22_post_impute_4


### Loop code to execute this task for the chromosomes 1-22 ###
for (( i=1; i<=22; i++ )); do
    ./plink2 --bfile ukb_chr"${i}"_post_impute_3 --remove sex_mismatch_m.txt --make-bed --out ukb_chr"${i}"_post_impute_4
done


Step-7: Hetrozygosity.    
./plink2 --bfile ukb_chr22_post_impute_4 --het --out R_check_22                ["R_check" can be arranged for chr 1-22 or use loop code]
 
(loop code: for (( i=1; i<=22; i++ )); do
    ./plink2 --bfile ukb_chr"${i}"_post_impute_4 --het --out R_check_"${i}"
done)

#Then go to R on linux (module load R, R)

het <- read.delim("R_check.het", head=TRUE)
pdf("heterozygosity.pdf")

het$HET_RATE = (het$"E.HOM." - het$"O.HOM.")/het$"E.HOM."

het$HET_RATE=(het$X51077-het$X49388.8)/het$X51077 [I used this for my UKB data instead of the above. It gives me HET_RATE =1- O(HOM)/E(HOM)

hist(het$HET_RATE, xlab="Heterozygosity Rate", ylab="Frequency", main= "Heterozygosity Rate")
dev.off()


het_fail = subset(het, (het$HET_RATE < mean(het$HET_RATE)-3*sd(het$HET_RATE)) | (het$HET_RATE > mean(het$HET_RATE)+3*sd(het$HET_RATE)));
het_fail$HET_DST = (het_fail$HET_RATE-mean(het$HET_RATE))/sd(het$HET_RATE);
write.table(het_fail, "fail-het-qc.txt", row.names=FALSE)                           [have to adjust "fail_het_qc_#" for chr you are doing]


#Then back to phoenix: 
sed 's/"// g' fail-het-qc.txt | awk '{print$1, $2}'> het_fail_ind.txt           

### Loop option (chr1-22)
for (( i=1; i<=22; i++ )); do
    sed 's/"//g' fail-het-qc_"${i}".txt | awk '{print $1, $2}' > het_fail_ind_"${i}".txt
done

### Then; 
./plink2 --bfile ukb_chr22_post_impute_4 --remove het_fail_ind.txt --make-bed --out ukb_chr22_post_impute_5

####loop option (chr 1-22)
for ((i=1; i<=22; i++)); do
    ./plink2 --bfile ukb_chr"${i}"_post_impute_4 --remove het_fail_ind_"${i}".txt --make-bed --out ukb_chr"${i}"_post_impute_5
done

### Step-7: Merge the final binary file for Analysis ###

./plink1.9 --bfile ukb_chr1_post_impute_5 --threads 72 --merge-list all_files_4merging.txt --make-bed --out merged_data


### WE THEN USED THIS FINAL "mereged_data" FOR GENETIC ANALYSES!


###################################################### B. QC CLSA ###########################################################################################

### SNP QC

#1. Prepare list of SNIPs with info score >=0.3 (two columns; SNPs ID and infoscore) and save to file name "clsa_info_score_0.3.txt "     ["can do this in R"]

#2. Extract bgen by the info score file (clsa_info_score_0.3.txt )created.

for chrom in {1..22}; do
    ./plink2 --threads 8 --bgen clsa_imp_${chrom}_v3.bgen ref-first --sample clsa_imp_v3.sample --extract clsa_info_score_0.3.txt --export bgen-1.2 --out clsa_imp_${chrom}_v3_post_impute
done
###

#3. Apply SNP based QC filtering 

for chr in {1..22}; do
    ./plink2 --threads 6 --bgen clsa_imp_${chr}_v3_post_impute.bgen ref-first --sample clsa_imp_${chr}_v3_post_impute.sample --maf 0.01 --geno 0.05 --mind 0.05 --hwe 1e-7 --max-alleles 2 --export bgen-1.2 --out clsa_imp_${chr}_v3_post_impute_1
done
###

###4. Idenitfy and Remove duplicates and make bed files 

for chr in {1..22}; do
    ./plink2 --bgen clsa_imp_${chr}_v3_post_impute_1.bgen ref-first --sample clsa_imp_${chr}_v3_post_impute_1.sample --rm-dup --make-bed --out clsa_imp_${chr}_v3_post_impute_2
done

### Clean using the duplicate and mismatch file created
for chr in {1..22}; do
./plink2 --bgen clsa_imp_${chr}_v3_post_impute_1.bgen ref-first --sample  clsa_imp_${chr}_v3_post_impute_1.sample --exclude clsa_imp_${chr}_v3_post_impute_2.rmdup.mismatch --make-bed --out clsa_imp_${chr}_v3_post_impute_2
done


###5. Merege the bed files (chr 1-22)
/hpcfs/users/a1877115/UKB_QC_M/plink1.9 --bfile clsa_imp_1_v3_post_impute_2 --threads 12 --merge-list all_files_4merging.txt --make-bed --out clsa_SNP_QC_merged_data


### Individual QC using the merged bed file ###
6.Sex Chromosome anneuploid 
./plink2 --bfile clsa_SNP_QC_merged_data --remove Sex_chromosome_anneploidy.tab --make-bed --out clsa_SNP_QC_merged_data_1

7. Exclude those with sex mismatch between reported and chromosomal sex
./plink2 --bfile clsa_SNP_QC_merged_data_1 --remove Sex_mismatch_ids.tab --make-bed --out clsa_SNP_QC_merged_data_2

8. Hetrozygosity
 ./plink2 --bfile clsa_SNP_QC_merged_data_2 --remove Hetrozygosity.tab --make-bed --out clsa_SNP_QC_merged_data_3

### WE THEN USED THIS FINAL "clsa_SNP_QC_merged_data_3" FOR GENETIC ANALYSES!


####################################### C. Heritability UKB ##############################################################

### First prepare files and upload - IC score file, qualitative covariates and quantiative covaraites in separte files (quantitaive covaraties including the ten PCs have been scales/convereted to z score).
### Produce the Genetic relationship matrix (GRM) 

### GRM (linux code)
#!/bin/bash
#SBATCH -p icelake
#SBATCH -N 4
#SBATCH -n 72
#SBATCH --time=72:00:00
#SBATCH --mem=160GB
#SBATCH --mail-type=END
#SBATCH --mail-type=BEGIN
#SBATCH --mail-type=FAIL
#SBATCH --mail-user=

/hpcfs/users/a1877115/UKB_QC_M/gcta-1.94.1 --bfile /hpcfs/users/a1877115/UKB_QC_M/merged_chr1-22_45K --autosome --make-grm --out GRM_GCTA_UKB_chr1_22_45Ksamples


### Heritability analysis

!/bin/bash
#SBATCH -p icelake,skylake,a100
#SBATCH -N 2
#SBATCH -n 36
#SBATCH --time=4:00:00
#SBATCH --mem=160GB
#SBATCH --mail-type=END
#SBATCH --mail-type=BEGIN
#SBATCH --mail-type=FAIL
#SBATCH --mail-user=

/hpcfs/users/a1877115/UKB_QC_M/gcta-1.94.1 \
--grm /hpcfs/users/a1877115/UKB_QC_M/GRM_GCTA_UKB_All_45Ksamples/GRM_GCTA_UKB_chr1_22_45Ksamples \
--pheno /hpcfs/users/a1877115/UKB_QC_M/IC_bifactor_score.tab \
--reml \
--covar /hpcfs/users/a1877115/UKB_QC_M/Quali_cov.tab \
--qcovar /hpcfs/users/a1877115/UKB_QC_M/Quanti_cov_z_pc10.tab \
--out /hpcfs/users/a1877115/UKB_QC_M/Heritability_GCTA_UKB_IC_All_45Ksamples/Heritability_estimate_GCTA_UKB_All_45Ksamples \
--thread-num 10 \

####################################### D. Heritability CLSA ##############################################################

### Heritability in CLSA - exactly same steps followed as in UKB
### First prepare files and upload - IC score file for CLSA, qualitative covariates and quantiative covaraites in separte files (quantitaive covaraties including the ten PCs have been scales/convereted to z score).
### Produce the Genetic relationship matrix (GRM)

### GRM (linux code)
#!/bin/bash
#SBATCH -p icelake
#SBATCH -N 2
#SBATCH -n 16
#SBATCH --time=48:00:00
#SBATCH --mem=64GB
#SBATCH --mail-type=END
#SBATCH --mail-type=BEGIN
#SBATCH --mail-type=FAIL
#SBATCH --mail-user=

/hpcfs/users/a1877115/UKB_QC_M/gcta-1.94.1 --bfile /hpcfs/users/a1877115/CLSA/CLSA_BGEN_CHR1-22/clsa_SNP_QC_merged_data_3 --autosome --make-grm --out GRM_GCTA_CLSA_All_26KSamples


### Heritability CLSA
!/bin/bash
#SBATCH -p icelake,skylake,a100
#SBATCH -N 2
#SBATCH -n 36
#SBATCH --time=4:00:00
#SBATCH --mem=160GB
#SBATCH --mail-type=END
#SBATCH --mail-type=BEGIN
#SBATCH --mail-type=FAIL
#SBATCH --mail-user=


/hpcfs/users/a1877115/UKB_QC_M/gcta-1.94.1 \
--grm GRM_GCTA_CLSA_All_26KSamples
--pheno IC_only_bifactor_score_CLSA.txt
--reml
--covar Quali_Cov.tab
--qcovar Quanti_cov_Zscored_10PCs.tab
--out Heritability_estimate_GCTA_CLSA_All_26Ksamples
--thread-num 10
### Done ####


####################################################### E. GWAS UKB ###############################################################################################################

### for fastGWA-mlm create sparse GRM from dense GRM produced above 

/hpcfs/users/a1877115/UKB_QC_M/gcta-1.94.1 --grm GRM_GCTA_UKB_chr1_22_45Ksamples --make-bK-sparse 0.05 --out sparse_grm_output_gcta_all_45Ksamles 


### GWAS UKB (fastGWA-mlm)

/hpcfs/users/a1877115/UKB_QC_M/gcta-1.94.1 \
--fastGWA-mlm 
--bfile /hpcfs/users/a1877115/UKB_QC_M/merged_chr1-22_45K 
--grm-sparse /hpcfs/users/a1877115/UKB_QC_M/sparse_grm_output_all_45Ksamples 
--pheno /hpcfs/users/a1877115/UKB_QC_M/IC_bifactor_score.tab 
--covar /hpcfs/users/a1877115/UKB_QC_M/Quali_cov.tab 
--qcovar /hpcfs/users/a1877115/UKB_QC_M/Quanti_cov_z_pc10.tab 
--out GWAS_Summary_UKB_IC_GCTA_fastGWA-mlm_All_45ksamples 
--thread-num 11 


####################################################### F. GWAS CLSA ###############################################################################################################

### Sparse GRM
/hpcfs/users/a1877115/UKB_QC_M/gcta-1.94.1 --grm GRM_GCTA_CLSA_All_26KSamples --make-bK-sparse 0.05 --out sparse_grm_gcta_clsa_26Ksamles


### GWAS CLSA (fastGWA-mlm)

/hpcfs/users/a1877115/UKB_QC_M/gcta-1.94.1 \
--fastGWA-mlm 
--bfile /hpcfs/users/a1877115/CLSA/CLSA_BGEN_CHR1-22/clsa_SNP_QC_merged_data_3 
--grm-sparse sparse_grm_gcta_clsa_26Ksamles 
--pheno IC_only_bifactor_score_CLSA.txt 
--covar Quali_Cov.tab 
--qcovar Quanti_cov_Zscored_10PCs.tab 
--out GWAS_Summary_CLSA_IC_GCTA_fastGWA-mlm_All_26ksamples 
--thread-num 11 


####################################################### G. Meta GWAS - Metal ###############################################################################################################

### We used metal package (checkthe github/documentation at:  https://genome.sph.umich.edu/wiki/METAL_Documentation)

### METAL is a command line tool.It is typically run from a Linux, Unix or DOS prompt by invoking the command metal (./metal) like we do with plink as "./plink2") 
### We prepare the Metal script first and saved it as "metal_SE_weighted_GRCH37.txt" (can be any other name)
### Upload the the summary statistics to be meta anlysed (need to be formated same) - for this anlaysis - UKB and CLSA summary statistics 
### the Metal scirpt has paramters to set (read the documentation).For this anlysis we have set parameters as below; 


### code in our script file metal_SE_weighted_GRCH37.txt (our gwas summarries are from these two directories in linux- "/hpcfs/users/a1877115/meta_GWAS/GWAS_UKB_GRCH37.tab" and "/hpcfs/users/a1877115/meta_GWAS/GWAS_CLSA_GRCH37.tab").

SCHEME STDERR
SEPARATOR  TAB
WEIGHT N
WEIGHTLABEL N
MARKER SNP
ALLELE A1 A2
EFFECT BETA
STDERR SE
STDERRLABEL SE
PVALUE P
PROCESS /hpcfs/users/a1877115/meta_GWAS/GWAS_UKB_GRCH37.tab
PROCESS /hpcfs/users/a1877115/meta_GWAS/GWAS_CLSA_GRCH37.tab
ANALYZE
ANALYZE HETEROGENEITY
QUIT

### Then saving the script file and going back to linux we run the metal script 
./metal metal_SE_weighted_GRCH37.txt

### Additional by swapping SCHEME to SAMPLESIZE, i edited script file and save as "metal_n_weighted_GRCH37.txt" and run the meta- i did this to get N in the meta as it is not produced under STDERR option
 
SCHEME SAMPLESIZE
SEPARATOR  TAB
WEIGHT N
WEIGHTLABEL N
MARKER SNP
ALLELE A1 A2
EFFECT BETA
STDERR SE
STDERRLABEL SE
PVALUE P
PROCESS /hpcfs/users/a1877115/meta_GWAS/GWAS_UKB_GRCH37.tab
PROCESS /hpcfs/users/a1877115/meta_GWAS/GWAS_CLSA_GRCH37.tab
ANALYZE
ANALYZE HETEROGENEITY
QUIT

### Then saving the script file and going back to linux we run the metal script 
./metal metal_n_weighted_GRCH37.txt


####################################################### H. Polygenic score analysis ###############################################################################################################
# github_link: https://github.com/getian107/PRScs

# Up loaded files
# GWAS summary for UKB (columns required and naming - format to: CHR, SNP, A1, A2, BETA, P)
# LD reference panel - EUR reference (download and untar from the github page)
# Path to final QCed renamed clsa binary file ("renamed_new_filtered_clsa_SNP_QC_merged_data_3")


#1.Install PRS-CS. Available from GitHub
 
git clone https://github.com/getian107/PRScs.git

cd PRScs


#2. Create pynthon environement its dependcies - PRS-CS requires a specific Python environment. 
module load python v...
module load Anaconda v...

conda create -n prscs_env python=3.7

conda activate prscs_env

pip install h5py

pip install numpy scipy pandas matplotlib scikit-learn


### 3. Post-estimation analysis PRScs (Can be in job submission format too)

python PRScs.py --ref_dir=/scratchdata1/users/a1877115/PRS_UKB_discovery/PRScs/ldblk_1kg_eur --bim_prefix=/scratchdata1/users/a1877115/PRS_UKB_discovery/PRScs/renamed_new_filtered_clsa_SNP_QC_merged_data_3 --sst_file=/scratchdata1/users/a1877115/PRS_UKB_discovery/PRScs/GWAS_UKB_GRCH38_Final_PB_new.tab --n_gwas=44631 --phi=1e-2 --chr=1,2,3,4,5,6,7,8,9,10,11,12,13,14,15,16,17,18,19,20,21,22 --out_dir=/hpcfs/users/a1877115/PRS/PRScs/UKB_PRS_post_eff_IC

### Concatenate the post estimate effect sizes for the 22 chromosomes
cat UKB_PRS_post_eff_IC_pst_eff_a1_b0.5_phi1e-02_chr{1..22}.txt > UKB_PRS_post_eff_IC_pst_eff_a1_b0.5_phi1e-02_all.txt

### Developing PRS (first remove duplicate and makebed - renamed_new_filtered_clsa_SNP_QC_merged_data_4 and then proceed to PRS scores creation)
/hpcfs/users/a1877115/UKB_QC_M/plink2 --bfile /scratchdata1/users/a1877115/PRS_UKB_discovery/PRScs/renamed_new_filtered_clsa_SNP_QC_merged_data_3 --rm-dup force-first --make-bed --out /scratchdata1/users/a1877115/PRS_UKB_discovery/PRScs/renamed_new_filtered_clsa_SNP_QC_merged_data_4

/hpcfs/users/a1877115/UKB_QC_M/plink2 --bfile /scratchdata1/users/a1877115/PRS_UKB_discovery/PRScs/renamed_new_filtered_clsa_SNP_QC_merged_data_4 --score /scratchdata1/users/a1877115/PRS_UKB_discovery/PRScs/UKB_PRS_post_eff_IC_pst_eff_a1_b0.5_phi1e-02_all.txt 2 4 6 --out /scratchdata1/users/a1877115/PRS_UKB_discovery/PRScs/PRScs_Scores/simplex


############################################# Association testing (linear regression using R)######################################
#Go to R software and open the data that I already mereged with PRS scores and all varaibles required for regression analysis (All_vars_PRScs_CLSA.tab)
#calculate R2 using full covaraites including PRS
#calculate R2 using covaraites except PRS
#calculate the difference in R2 which tell you how much varaibaility the PRS explains


### Load data to R
All_vars_PRScs<- read.table("//uofa/users$/users5/a1877115/Desktop/PRS/All_vars_PRScs_CLSA.tab", header = TRUE)

# Models
model1 <- lm(IC_Zscore ~ Age + PC1 + PC2 + PC3 + PC4 + PC5 + PC6 + PC7 + PC8 + PC9 + PC10 + as.factor(Sex) + as.factor(Batch) + PRScs_Zscore, data = All_vars_PRScs)
model2 <- lm(IC_Zscore ~ Age + PC1 + PC2 + PC3 + PC4 + PC5 + PC6 + PC7 + PC8 + PC9 + PC10 + as.factor(Sex) + as.factor(Batch), data = All_vars_PRScs)

summary(model1)
conf_intervals <- confint(model1, level = 0.95)
print(conf_intervals)

summary(model2)


##################################### Association tetsing using PGS deciles ###############################
All_vars_PRScs$PRS_deciles <- cut(All_vars_PRScs$PRScs_Zscore, breaks = quantile(All_vars_PRScs$PRScs_Zscore, probs = 0:10 / 10), labels = FALSE, include.lowest = TRUE)

model_lm_deciles <- lm(IC_Zscore ~ Age + PC1 + PC2 + PC3 + PC4 + PC5 + PC6 + PC7 + PC8 + PC9 + PC10 + as.factor(Sex) + as.factor(Batch) + as.factor(PRS_deciles), data = All_vars_PRScs)
conf_intervals <- confint(model_lm_deciles, level = 0.95)


####################################################### I. Genetic correlation analysis ###############################################################################################################

### github link and Youtube tutorial how to do LDSC genetic correlation analysis
Github: https://github.com/bulik/ldsc
Youtube: https://www.youtube.com/watch?v=DQITrRe693I

### Instal LDSC
git clone https://github.com/bulik/ldsc.git
cd ldsc

### Instal Python and Anaconda softwares 
module load Python/3.11.3-GCCcore-12.3.0
module load Anaconda3/2024.06-1

### Create Python environment 
conda env create --file environment.yml
source activate ldsc

### Download Reference LD Scores (I used sent files on box from Dr.Azmeraw) 
(The link on github seems down- the following link obtained in google working group: wget -r -np -nH --cut-dirs=3 -R index.html https://ibg.colorado.edu/cdrom2021/Day06-nivard/GenomicSEM_practical/eur_w_ld_chr/
vtar -xvjf eur_w_ld_chr.tar.bz2


### Prepare GWAS Summary Statistics for Trait 1 (IC)
#!/bin/bash

# Directory containing the munged summary statistics files
trait_dir="./GWAS_Summary_statistics for genetic correlation"

# Path to the already munged Intrinsic Capacity (IC) summary statistics
ic_sumstats="${trait_dir}/Intrinsic_Capacity_gwas.sumstats.gz"

# Check if the IC summary statistics file exists
if [ ! -f "$ic_sumstats" ]; then
  echo "Intrinsic Capacity summary statistics file not found: ${ic_sumstats}"
  exit 1
fi

# Loop through each munged summary statistics file in the folder
for file in "${trait_dir}"/*_gwas.sumstats.gz; do
  # Extract the trait name from the file name
  trait=$(basename "${file}" _gwas.sumstats.gz)

  echo "Calculating genetic correlation for trait: ${trait}"

  # Calculate genetic correlation between IC and each trait
  python2 ldsc.py \
  --rg "${ic_sumstats},${file}" \
  --ref-ld-chr eur_w_ld_chr/ \
  --w-ld-chr eur_w_ld_chr/ \
  --out "${trait_dir}/IC_${trait}"

  # Check if the genetic correlation calculation was successful
  if [ $? -ne 0 ]; then
    echo "Error calculating genetic correlation for trait: ${trait}"
  fi
done

############ Loop menging through each summary statistics file in the folder

#!/bin/bash

# Directory containing the trait summary statistics files
trait_dir="./GWAS_Summary_statistics for genetic correlation"

# List the files in the directory for debugging
echo "Listing files in ${trait_dir}:"
ls -l "${trait_dir}"

# Loop through each summary statistics file in the folder
for file in "${trait_dir}"/*.tab; do
  # Check if the file exists
  if [ ! -f "$file" ]; then
    echo "No .tab files found in the directory."
    continue
  fi

  # Extract the trait name from the file name
  trait=$(basename "${file}" .tab)

  echo "Munging summary statistics for trait: ${trait}"

  # Munge summary statistics for each trait
  ./munge_sumstats.py \
  --out "${trait_dir}/${trait}_gwas" \
  --merge-alleles w_hm3.snplist \
  --N-col N \
  --sumstats "${file}" \
  --a1 A1 \
  --a2 A2 \
  --snp SNP \
  --p P \
  --signed-sumstats BETA,0 \
  --chunksize 50000

  # Check if the munging was successful
  if [ $? -ne 0 ]; then
    echo "Error munging summary statistics for trait: ${trait}"
    continue
  fi
done

################## loop GC analysis through each trait with IC

#!/bin/bash

# Directory containing the munged summary statistics files
trait_dir="./GWAS_Summary_statistics for genetic correlation"

# Loop through each munged summary statistics file in the folder
for file in "${trait_dir}"/*_gwas.sumstats.gz; do
  # Extract the trait name from the file name
  trait=$(basename "${file}" _gwas.sumstats.gz)

  echo "Calculating genetic correlation for trait: ${trait}"

  # Calculate genetic correlation between IC and each trait
  python2 ldsc.py \
  --rg "${trait_dir}/Intrinsic_Capacity_gwas.sumstats.gz,${file}" \
  --ref-ld-chr eur_w_ld_chr/ \
  --w-ld-chr eur_w_ld_chr/ \
  --out "${trait_dir}/IC_${trait}"

  # Check if the genetic correlation calculation was successful
  if [ $? -ne 0 ]; then
    echo "Error calculating genetic correlation for trait: ${trait}"
  fi
done

#############################
